# Comparative genomics and characterization of SARS-CoV-2 P.1 (Gamma) Variant of Concern (VOC) from Amazonas, Brazil

**DOI:** 10.1101/2021.10.30.21265694

**Authors:** Ricardo Ariel Zimerman, Patrícia Aline Gröhs Ferrareze, Flávio Adsuara Cadegiani, Carlos Gustavo Wambier, Daniel do Nascimento Fonseca, Andrea Roberto de Souza, Andy Goren, Liane Nanci Rotta, Zhihua Ren, Claudia Elizabeth Thompson

## Abstract

**Background:** P.1 lineage (Gamma) was first described in the State of Amazonas, northern Brazil, in the end of 2020, and has emerged as a very important variant of concern (VOC) of SARS-CoV-2 worldwide. P.1 has been linked to increased infectivity, higher mortality and immune evasion, leading to reinfections and potentially reduced efficacy of vaccines and neutralizing antibodies.

**Methods:** The samples of 276 patients from the State of Amazonas were sent to a central referral laboratory for sequencing by gold standard techniques, through Illumina MiSeq platform. Both global and regional phylogenetic analyses of the successfully sequenced genomes were conducted through maximum likelihood method. Multiple alignments were obtained including previously obtained unique human SARS-CoV-2 sequences. The evolutionary histories of spike and non-structural proteins from ORF1a of northern genomes were described and their molecular evolution was analyzed for detection of positive (FUBAR, FEL, and MEME) and negative (FEL and SLAC) selective pressures. To further evaluate the possible pathways of evolution leading to the emergence of P.1, we performed specific analysis for copy-choice recombination events. A global phylogenomic analysis with subsampled P.1 and B.1.1.28 genomes was applied to evaluate the relationship among samples.

**Results:** Forty-four samples from the State of Amazonas were successfully sequenced and confirmed as P.1 (Gamma) lineage. In addition to previously described P.1 characteristic mutations, we find evidence of continuous diversification of SARS-CoV-2, as rare and previously unseen P.1 mutations were detected in spike and non-structural protein from ORF1a. No evidence of recombination was found. Several sites were demonstrated to be under positive and negative selection, with various mutations identified mostly in P.1 lineage. According to the Pango assignment, phylogenomic analyses indicate all samples as belonging to the P.1 lineage.

**Conclusion:** P.1 has shown continuous evolution after its emergence. The lack of clear evidence for recombination and the positive selection demonstrated for several sites suggest that this lineage emergence resulted mainly from strong evolutionary forces and progressive accumulation of a favorable signature set of mutations.

## INTRODUCTION

The Severe acute respiratory syndrome coronavirus 2 (SARS-CoV-2) P.1 (Gamma) lineage has accounted for the majority of the genomes sequenced in Brazil during the second wave of the COVID-19 pandemic (1). It is considered one of the most relevant “Variants Of Concern” (VOC) worldwide. It carries 10 non-synonymous mutations in the spike protein, including three located at the Receptor Binding Domain (RBD: K417T, N501Y, and E484K). Remarkably, N501Y has demonstrated enhanced affinity for ACE-2 binding, potentially resulting in higher viral loads (2). The E484K substitution was previously demonstrated by our group to have arisen and maintained in four genetically diverse lineages in Brazil (3). It can induce immune escape with consequent reinfection and is associated with convalescent plasma therapy failure (4). We have previously shown that both of these substitutions are under strong positive selection (5). Since these mutations are present in other VOCs around the world, their emergence is evidence of convergent SARS-CoV-2 evolution (5,6). K417T occurs in an important foot-print for class I anti-RBD antibodies and has been associated to decrease neutralization activity for some monoclonal antibodies (mAb). Despite the mutations in spike predominate, P.1 also presents substitutions at other sites potentially relevant. Mutations in non-structural proteins may alter their function related to transcription, duplication and immune modulation. In fact, it was suggested that at least emerging lineages may induce weaker antiviral interferon activity (7). The association of higher infectivity related to spike mutations with possible innate immune evasion could be associated with higher viral load. An earlier study describing P.1 in northern Brazil has linked this lineage to lower Cycle Threshold (Ct) values (8). This may be of major significance, since in studies testing mAb (9) persistently higher viral loads after one week of symptom initiation were independently associated with worse outcomes.

P.1 has been tied to higher case fatality rate (8) and this phenotypic trait may be related to its genetic background. The origin of the unique set of approximately 35 amino acid substitutions that characterize P.1 remain largely unknown. Reassortment of entire segments of genomes by “copy-choice” recombinations are well described among coronaviruses (10). Alternatively, the high seroprevalence of anti-SARS-CoV-2 antibodies in northern Brazil citizens could indicate that strong selective pressure was responsible for the new lineage origin. Accordingly, B.1.351 (Beta) has also emerged in a heavily previously exposed population at Mandela’s Bay, South Africa (11). Antigenic drift “by chance” appears less likely, because intense ongoing transmission relieves evolutionary bottlenecks required for this phenomenon. Due to intense viral turnover, we expected to find signs of diversification of the original P.1 in clinical samples obtained during the highs of the second wave in north Brazil.

In this study, we describe the full-length SARS-CoV-2 genomes of 44 clinical samples from Amazonas, Brazil, sequenced and analyzed by our group, and compare them to previously described P.1 from Brazil and worldwide. In order to better understand the evolutionary forces driving the P.1 emergence and evolution in the North region from Brazil, we conducted tests for recombination, phylogenomics, phylogenetic analyses of spike and non-structural proteins from ORF1a, and the detection of selective pressures acting on these sequences. New or uncommon Single Nucleotide Polymorphisms (SNPs) were also described and their potential relevance discussed.

## MATERIALS AND METHODS

In total, 276 nasopharyngeal swab samples of patients from Manaus, Parintins, and Itacoatiara (Amazonas, Brazil) collected for SARS-CoV-2 Quantitative Reverse Transcription Polymerase Reaction (RT-qPCR), which would be discarded by the laboratory, were initially included in this study. Patient SARS-CoV-2 status was determined by RT-qPCR testing following the Cobas SARS-CoV-2 RT-qPCR kit test protocol (Roche, USA).

### SARS-CoV-2 genome sequencing and assembly

For the genome sequencing, confirmed SARS-CoV-2 patients (by RT-qPCR test result) had their clinical samples submitted to a second RT-qPCR, which was performed by BiomeHub (Florianópolis, Santa Catarina, Brazil), with a *charite-berlin* protocol. Samples with quantification cycle (Cq) up to 30 for at least one primer were selected for SARS-CoV-2 genome sequencing and assembly by the BiomeHub laboratory.

The total RNAs were prepared according to reference protocol (*dx*.*doi*.*org/10*.*17504/protocols*.*io*.*befyjbpw*), with the cDNA synthesized with SuperScript IV (Invitrogen) and the DNA amplified with Platinum Taq High Fidelity (Invitrogen). The library preparation was performed with Nextera Flex (Illumina) and quantification was performed with Picogreen and Collibri Library Quantification Kit (Invitrogen). The genome sequencing was generated on Illumina MiSeq Platform by 150×150 runs with 500xSARS-CoV-2 coverage (50-100 mil reads/per sample).

For the genome assembly (BiomeHub in-house script), the adapters removal and read trimming for 150 nt read sequences were performed by *fastqtools*.*py*. The alignment of the sequenced reads to the reference SARS-CoV-2 genome (GenBank ID: NC_045512.2) was performed by Bowtie v2.4.2 (12) and additional parameters as *end-to-end* and *very-sensitive*. The analysis of the sequencing coverage and depth was generated by samtools v1.11 (13) with minimum base quality per base (Q) ≥ 30. Finally, the consensus sequence for each SARS-CoV-2 genome was generated by a bcftools pipeline (14), including the commands *mpileup* (parameters: Q ≥ 30; depth (d) ≤ 1,000), *filter* (parameters: DP>50) and *consensus*.

### Genomic analyses

The assignment of the SARS-CoV-2 lineages (Pango lineages assignment for September, 2021) for each assembled genome was obtained with the Pangolin v2.3.8 web server (https://github.com/cov-lineages/pangolin). The clade assignment, mutation calling and sequence quality check were performed by the Nextclade web server (https://clades.nextstrain.org/). The Snippy variant calling and core genome alignment pipeline v4.6.0 (https://github.com/tseemann/snippy) identified Single Nucleotide Polymorphisms (SNPs) and insertions/deletions from each genome sequence. For the genome map and SNP histogram, the genome alignment was performed by the MAFFT v7.475 (15) web server followed by the running of the *msastats*.*py* script, and the *plotAlignment* and *plotSNPHist* functions in RStudio (16). The SARS-CoV-2 genome with GenBank ID NC_045512.2 was used as reference.

### Recombination analysis of northern samples

Initially, 2,485 complete and high covered genomes from the northern states in Brazil (57 genomes from Acre, 298 from Amapá, 1,511 from Amazonas, 394 from Pará, 32 from Rondônia, 38 from Roraima, and 155 from Tocantins) were retrieved from the GISAID database (submission up to September 12, 2021) and added to our 44 sequenced genomes. The genome alignment was performed using the Wuhan genome NC_045512.2 as a reference sequence in the MAFFT web server (15) with 1PAM / κ=2 scoring matrix. The resulting alignment was trimmed in 266 positions for the 5’ end and 264 positions in the 3’ end with UGENE (17) v39.0, followed by the deletion of sequence duplicates, maintaining 1,931 unique genomes in the sequence set.

For the recombination analysis, the aligned sequence set was tested by the Recombination Detection Program (RDP) software v4.101 (18). The methods RDP (19), GENECONV (20), Chimaera (21), MaxChi (22), and 3Seq (23) were applied for the primary scan, followed by BootScan (24) and SiScan (25) for the second scan. After the detection of genomic breakpoints and possible recombination events, the results were re-evaluated with the available methods RDP, GENECONV, BootScan, Chimaera, MaxChi, SiScan, 3Seq, LARD (26), and Phylpro (27). Recombination events identified after Bonferroni correction at a p-value ≤ 0.05 were accepted. All analyses were performed with default parameters. The genetic distance plot comparison of the 44 sequenced genomes from this study with the NC_045512.2 reference was performed with the Python package recan using a window of 200 nt and a shift of 50 nt as parameters (28).

### Phylogenomic analysis of northern samples

For the phylogenetic analysis of the northern genomes, the aligned set of 1,931 unique sequences also used in the recombination analysis was selected. The best evolutionary model was inferred as GTR+R4 by ModelTest-ng (29). The phylogenetic tree was built by the Maximum Likelihood method using the IQTREE software (30) with a Shimodaira-Hasegawa-like approximate likelihood ratio test of 1,000 replicates added to 1,000 replicated of a ultrafast bootstrap, 2,000 iterations and the optimization of the UFBoot trees by NNI on bootstrap alignment. The tree visualization and editing was generated by the FigTree software (http://tree.bio.ed.ac.uk/software/figtree/).

### Phylogenetic analyses of spike and non-structural proteins from ORF1a of northern samples

Considering that spike and ORF1a were the genomic regions in the northern Brazilian samples with higher number of mutations identified in our genomic analyses, we performed phylogenetic analyses of those regions to better understand the specific patterns of molecular evolution. The nucleotide sequence alignments for the spike and non-structural proteins from the ORF1a sequence were recovered from the alignment of 1,931 northern Brazilian genome samples, according to the CDS coordinates of the reference genome (NC_045512.2). In total, eleven alignment sets (spike, NSP1, NSP2, NSP3, NSP4, NSP5, NSP6, NSP7, NSP8, NSP9, and NSP10) had their duplicated sequences removed and followed the same Modeltest-ng / IQTREE / FigTree workflow method previously described.

### Molecular evolution of spike and non-structural proteins from ORF1a of northern samples

Selection tests were performed with HyPhy v2.5.32 (31) using the nucleotide sequence alignment and the maximum likelihood tree (previously described) for spike and non-structural proteins from ORF1a. The FUBAR (32), FEL (33), and MEME (34) methods were applied with default parameters for the analysis of positive selection, while FEL (33) and SLAC (33) methods were used with default parameters to evaluate if those regions were submitted to negative selection.

### Global phylogenomic analyses

A set of 54,214 P.1 and 2,081 B.1.1.28 completed, dated and high-covered genomes, available on GISAID, with collection date from January 01, 2020 and submission date until September 12, 2021, were date-ordered and had their duplicated sequences removed, keeping the first occurrence of each sequence. Subsequently, the software Augur (35), implemented a probabilistic subsampling based on the sample location (country) in order to select 5,000 P.1 and 100 B.1.1.28 genomes. The multiple sequence alignment with the P.1, B.1.1.28, the 44 sequenced genomes from this study, and the SARS-CoV-2 reference genome NC_045512.2 was generated by the MAFFT web server (1PAM / κ=2 scoring matrix), while the alignment trimming (deletion of 265 and 259 nucleotides at 5’ and 3’ ends, respectively) was performed with UGENE (17). A second round of sequence duplicates exclusion was applied in order to keep the first occurrence of each unique sequence. In case of sequence identity between a genome from this study and genomes downloaded from GISAID, the date was disregarded and our genomes were kept as representative sequences.

The global phylogenetic analysis was then performed with the forty-four genomes sequenced in this study added to 4,953 GISAID data, totalizing 4,997 unique genomes. The 4,953 GISAID genomes were assigned to the B (SARS-CoV-2 reference genome), P.1 (4,856 genomes), and B.1.1.28 (96 genomes) lineages. The best evolutionary model was inferred as GTR+R4 by ModelTest-ng (29). The phylogenetic tree was built by the Maximum Likelihood method using the IQTREE software (30) with Shimodaira-Hasegawa-like approximate likelihood ratio test of 1,000 replicates added to 1,000 replicated of a ultrafast bootstrap, 2,000 iterations and the optimization of the UFBoot trees by NNI on bootstrap alignment. The tree visualization and editing was generated by the FigTree software (http://tree.bio.ed.ac.uk/software/figtree/).

In all phylogenetic analyses, including northern samples, spike, non-structural proteins from ORF1a and the global phylogenomics, a group was considered monophyletic when satisfied the criteria of ≥80% for SH-aLRT (36) and ≥95% for ultrafast bootstrap branch support values (37). However, we also always described all cases whose branch support values for both statistical tests were ≥80%, considering the previously cited criteria seem to conservative.

## RESULTS

The flowchart of SARS-CoV-2 genome sequencing is presented in Figure 1. The nasopharyngeal swab samples (n=276) were firstly analyzed to detect SARS-CoV-2 using RT-qPCR in a private lab and, subsequently, sent to the reference sequencing center. Twenty-five of them were excluded from this study due to their poor conditions, such as low volume. After the first RT-qPCR, 120 samples that tested positive for SARS-CoV-2 were submitted to a second RT-qPCR for Cq values verification. Only 45 samples that had Cq below 30 for at least one primer were submitted to sequencing. In total, 44 patients had their SARS-CoV-2 positive samples successfully genotyped.

**Figure 1.**
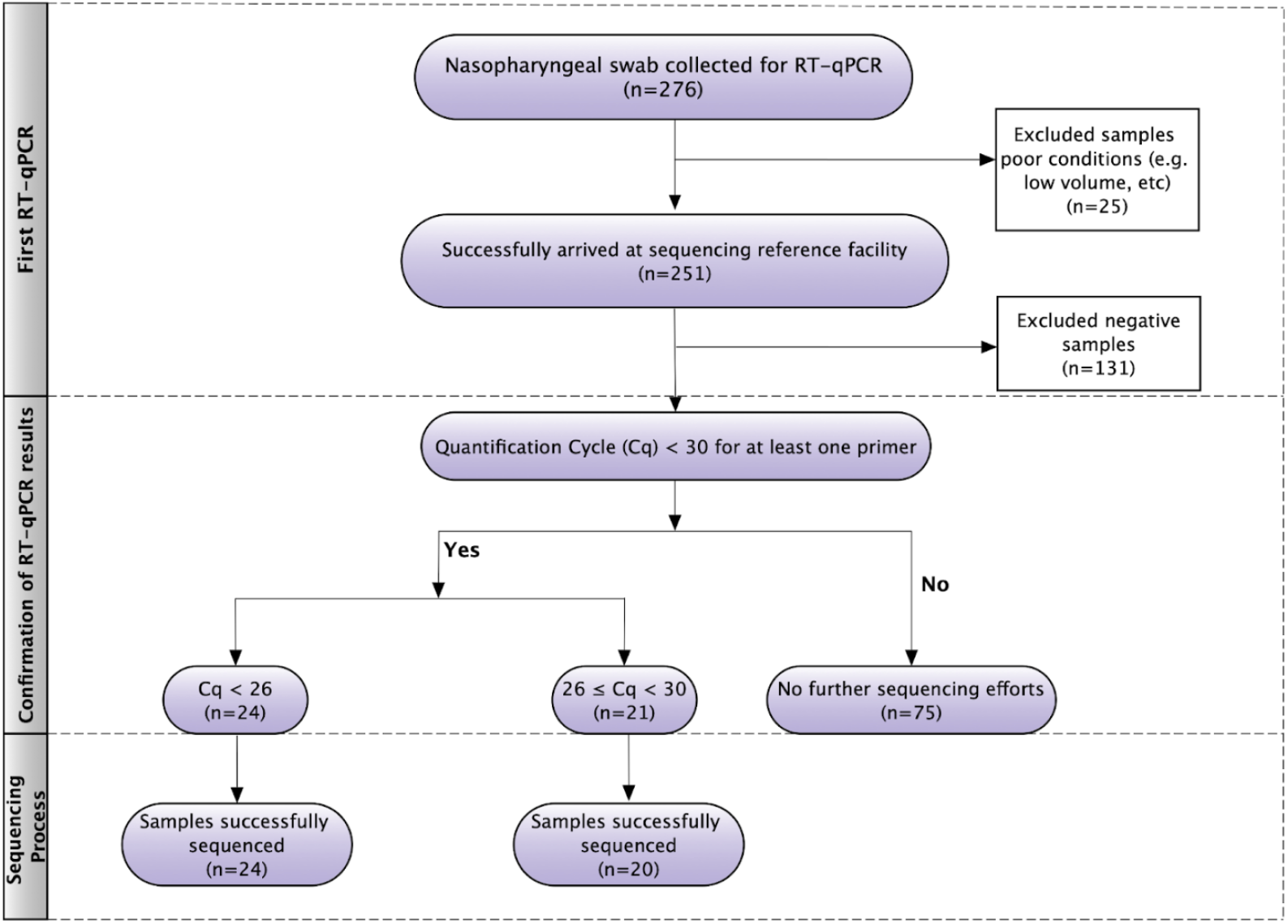
Flowchart of SARS-CoV-2 RT-qPCR and genome sequencing.

### Genomic analysis

Forty-four successfully sequenced SARS-CoV-2 genomes were obtained from samples taken from February, 18 to March, 10, 2021, representing patients from three different cities: Manaus (n=42), Parintins (n=1), and Itatiacoara (n=1). The sequence coverage for the forty-four assemblies ranged between 95.26 up to 99.79% of the 29,903 bp of NC_045512.2 reference genome (average mean of 99.50%). The mean sequencing depth was calculated to 771.79x, with a range variation between 499 and 1,060x (Supplementary File 1). At least 83.21% of the sequence positions achieved a coverage depth ≥ 51x (max = 99.64%, mean = 96.04%). According to the Pango Lineages assignment, the forty-four samples were characterized as belonging to the P.1 lineage (subclade 20J, Gamma, V3). Figure 2 shows the frequency of SNPs per SARS-CoV-2 genome position along the 44 sequenced genomes. In summary, in this study, the sequenced genomes from Amazonas showed between 21 and 26 amino acid mutations and two up to five deletions. A set of 12 genomes present a four nucleotides insertion (AACA) after the genomic position 28269.

**Figure 2.**
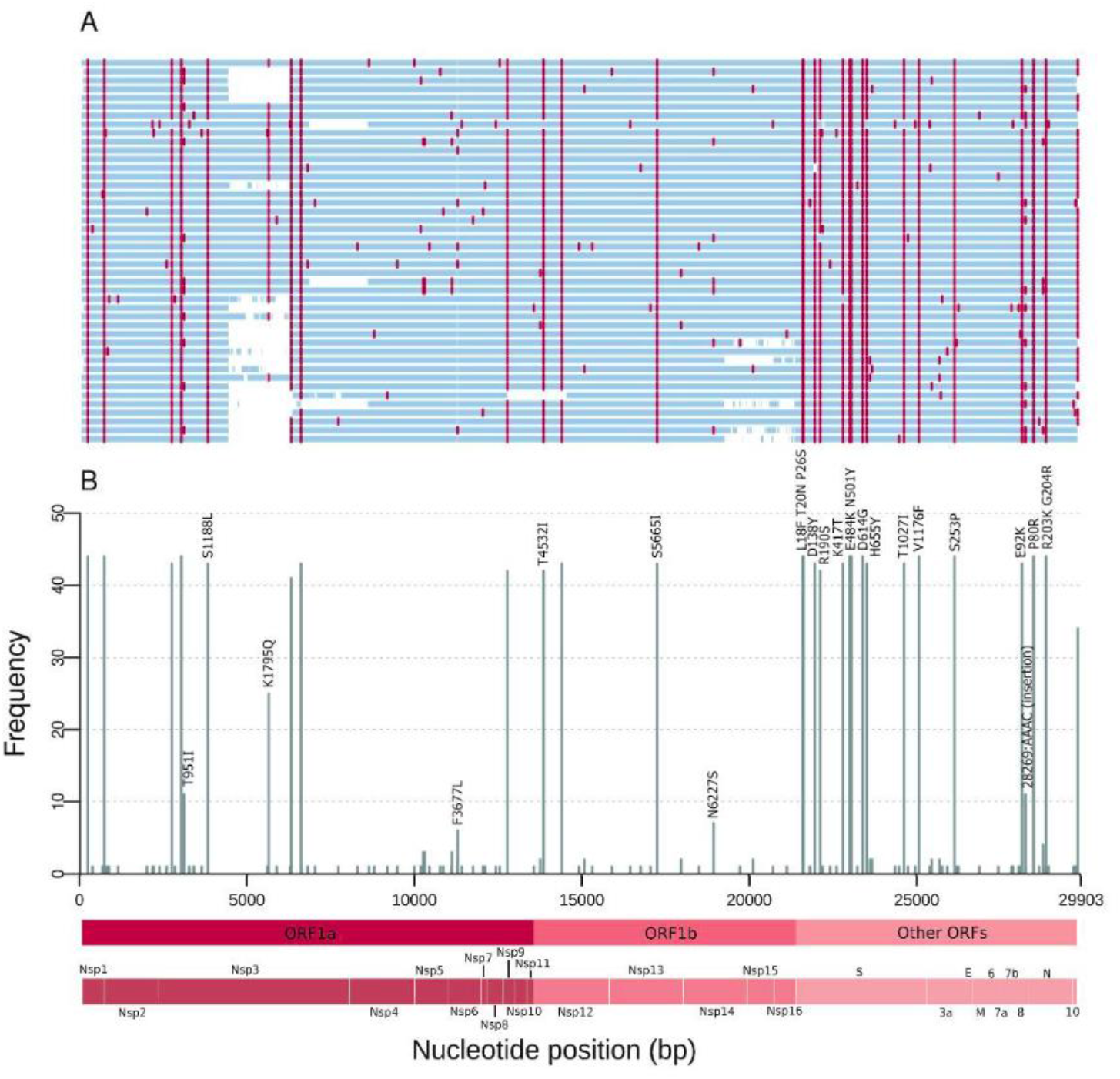
Mutations of the SARS-CoV-2 genomes from the Amazonas state, Northern Brazil sampled in February and March 2021. (A) Genome map for the 44 genomes sequenced. Nucleotide substitutions are colored in red and blank regions represent low sequencing coverage. (B) Frequency of SNPs per SARS-CoV-2 genome position along the 44 genomes. These mutations are corresponding to the red lines in (A) and only missense substitutions represented by >10 sequences have their respective amino acid changes indicated above the bars. Main Open Reading Frames (ORFs) and SARS-CoV-2 proteins are indicated at the bottom to allow a rapid visualization of the viral proteins affected.

Beyond the constellation of mutations expected from the P.1 lineage, we have found some unusual and/or uncertain deletions and substitutions. After automatic alignment using MAFFT v7.475 and visual inspection, we identified six nucleotide deletions of amino acid residues N188 and L189 from the spike protein in three genomic sequences followed by the R190S substitution in two of them. However, by observing the alignment data is not possible to completely discard the occurrence of a N188S substitution followed by deletion of the residues L189 and R190, since in both cases (N188S/R190S) the codons are being mutated to a serine residue, which is the pattern commonly seen on GISAID. It is important to note that there is no previous evidence on the GISAID database of a coupled N188-L189 deletion with the conservation of the neighbor residues. In 22 genome sequences (P.1.10 and B.1.36, mostly) available on GISAID from Belgium, Egypt, India, Germany, Greece, Suriname, and USA, there is a deletion of the adjacent positions (e.g. 186, 187, and 190), in addition to N188-L189 deletion. Nevertheless, the finding of 15 P.1 genomes from Brazil and Taiwan presenting N188S/L189-R190del does not exclude the possibility of generalized alignment errors in these positions due to the mistaken AGT codon mapping, which is probably the case.

The frequency of spike mutations identified in the P.1 sequenced genomes (this study) compared with sequences from Amazonas, Brazil and World (up to September 12, 2021) available on GISAID is shown in Table 1. Interestingly, the P209H mutation at spike NTD, identified in 2.27% of our genomes, was not previously detected in the P.1 samples sequenced around the world. Actually, this mutation was identified in several lineages, including B.1.1.7 (Alpha; England, Italy, and USA), B.1.351 (Beta; South Africa and Sweden), and B.1.617.2 (Delta; England and Indonesia). The deletions S:A243 and S:L244 in the spike protein were simultaneously found in 43 P.1 genomes, excluding those sequenced in our study, from Brazil (Amazonas, Distrito Federal, Rio de Janeiro, Sergipe, and São Paulo), Belgium, Canada, Chile, French Guiana, Malta, Peru, and USA. Another example is the substitution S:T1066A, which was only found in genomes from the lineages AY.4, AY.7.1, B.1.1.7, and B.1.258.10 in USA and European countries. This substitution has been identified for the first time in a P.1 genome sequenced in this study. The S:D215G mutation was described in another eight P.1 Amazonas genomes after our sample collection, as well as in P.1 samples from the states of Goiás (first occurrence in Brazil), Paraná, Santa Catarina, São Paulo and also in Chile, Mexico, Netherlands, and USA. Therefore, one genome sequenced in this study and one additional genome were the first sequences identified carrying this mutation in P.1 lineage in Amazonas. The spike mutations L18F, P26S, E484K, N501Y, D614G, and V1176 were identified in all 44 genomes sequenced in this study, followed by the recognition of T20N, D138Y, H655Y, K417T, and T1027I in 43 of them. Eight spike mutations were observed in three sequences or less (N188-L189del, P209H, D215G, A243-L244del, A688V, and T1066A).

**Table 1.**
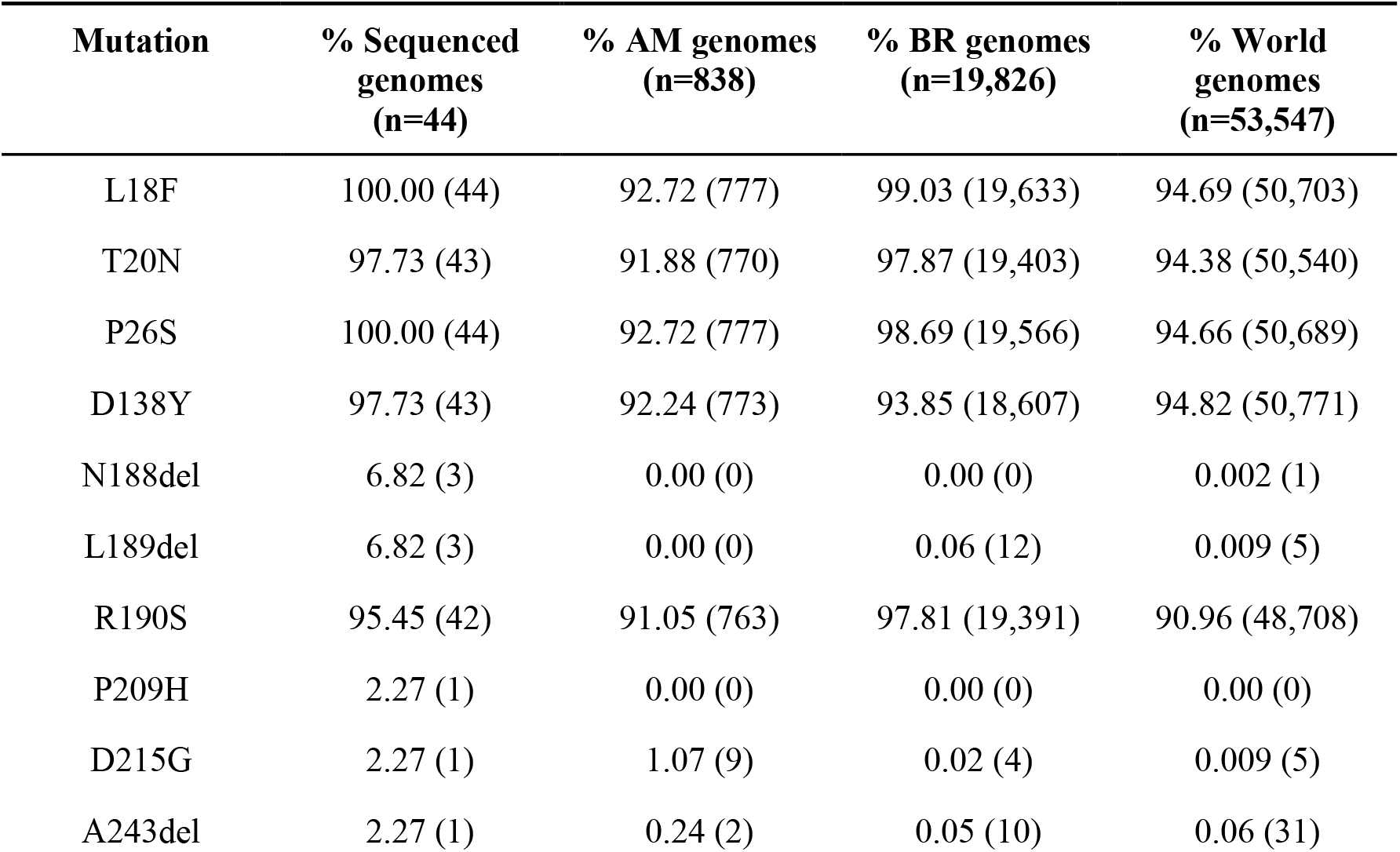
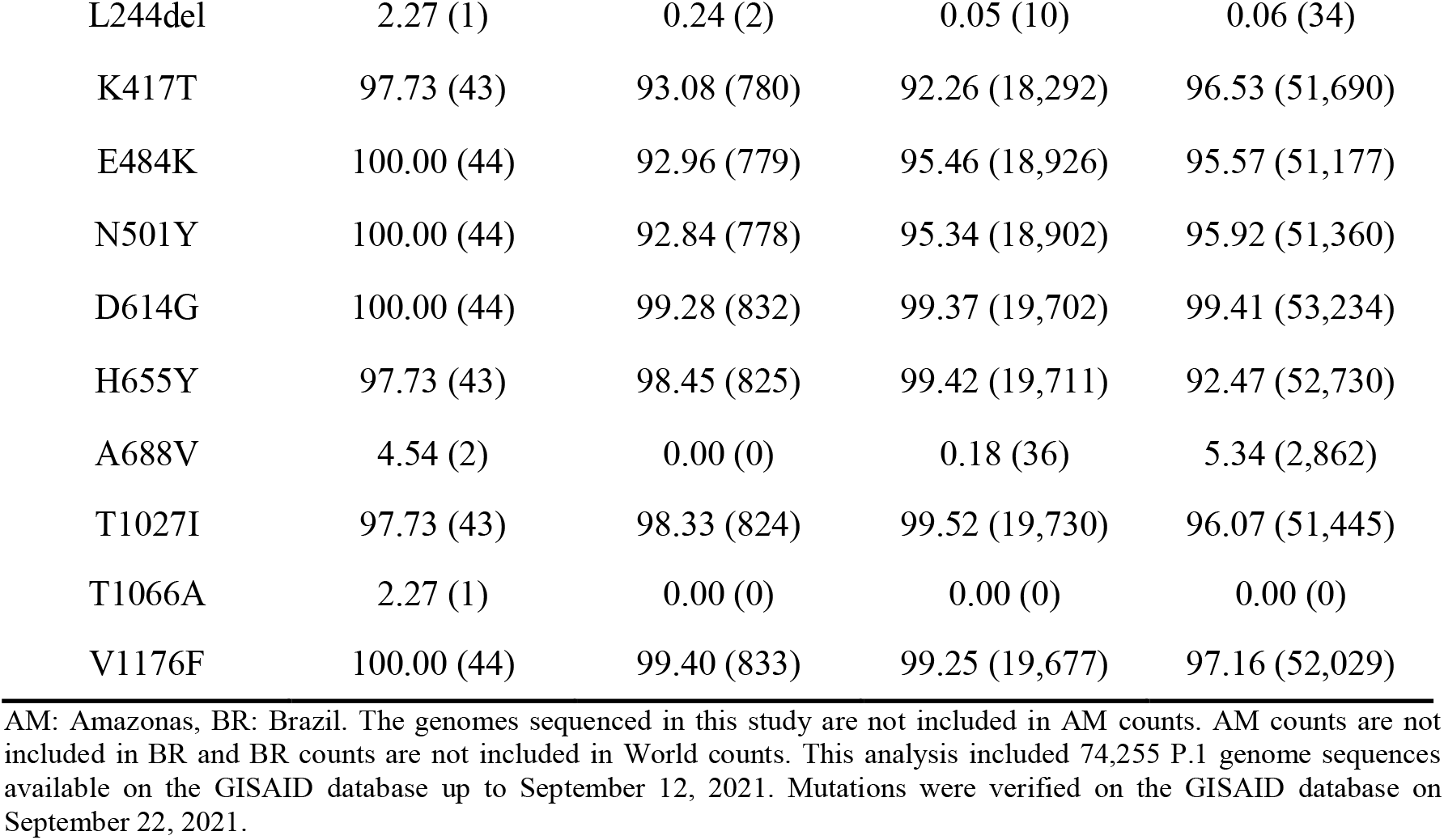
Frequency of spike mutations identified in the P.1 sequenced genomes (this study) compared with sequences from Amazonas, Brazil and World (up to September 12, 2021) available on GISAID.

| **Mutation** | **% Sequenced**<br>**genomes**<br>**(n=44)** | **% AM genomes**<br>**(n=838)** | **% BR genomes**<br>**(n=19,826)** | **% World**<br>**genomes**<br>**(n=53,547)** |
| --- | --- | --- | --- | --- |
| L18F | 100.00 (44) | 92.72 (777) | 99.03 (19,633) | 94.69 (50,703) |
| T20N | 97.73 (43) | 91.88 (770) | 97.87 (19,403) | 94.38 (50,540) |
| P26S | 100.00 (44) | 92.72 (777) | 98.69 (19,566) | 94.66 (50,689) |
| D138Y | 97.73 (43) | 92.24 (773) | 93.85 (18,607) | 94.82 (50,771) |
| N188del | 6.82 (3) | 0.00 (0) | 0.00 (0) | 0.002 (1) |
| L189del | 6.82 (3) | 0.00 (0) | 0.06 (12) | 0.009 (5) |
| R190S | 95.45 (42) | 91.05 (763) | 97.81 (19,391) | 90.96 (48,708) |
| P209H | 2.27 (1) | 0.00 (0) | 0.00 (0) | 0.00 (0) |
| D215G | 2.27 (1) | 1.07 (9) | 0.02 (4) | 0.009 (5) |
| A243del | 2.27 (1) | 0.24 (2) | 0.05 (10) | 0.06 (31) |
| L244del | 2.27 (1) | 0.24 (2) | 0.05 (10) | 0.06 (34) |
| K417T | 97.73 (43) | 93.08 (780) | 92.26 (18,292) | 96.53 (51,690) |
| E484K | 100.00 (44) | 92.96 (779) | 95.46 (18,926) | 95.57 (51,177) |
| N501Y | 100.00 (44) | 92.84 (778) | 95.34 (18,902) | 95.92 (51,360) |
| D614G | 100.00 (44) | 99.28 (832) | 99.37 (19,702) | 99.41 (53,234) |
| H655Y | 97.73 (43) | 98.45 (825) | 99.42 (19,711) | 92.47 (52,730) |
| A688V | 4.54 (2) | 0.00 (0) | 0.18 (36) | 5.34 (2,862) |
| T1027I | 97.73 (43) | 98.33 (824) | 99.52 (19,730) | 96.07 (51,445) |
| T1066A | 2.27 (1) | 0.00 (0) | 0.00 (0) | 0.00 (0) |
| V1176F | 100.00 (44) | 99.40 (833) | 99.25 (19,677) | 97.16 (52,029) |

AM: Amazonas, BR: Brazil. The genomes sequenced in this study are not included in AM counts. AM counts are not included in BR and BR counts are not included in World counts. This analysis included 74,255 P.1 genome sequences available on the GISAID database up to September 12, 2021. Mutations were verified on the GISAID database on September 22, 2021.

The frequency of mutations in non-structural proteins (NSPs) and N protein identified in the P.1 sequenced genomes (this study) compared with sequences from Amazonas, Brazil and World (up to September 12, 2021) available on GISAID is shown in Table 2. The N protein sequence exhibited the substitutions P80R, R203K, and G204R in all 44 genomes, which are highly frequent mutations in the genomes sequenced in Brazil and World, and two of them also presented - separately - the mutations P199L and T135I. A high number of mutations was identified for ORF1a, totalizing 29 possible amino acid substitutions and three deletions. The most frequent ones include K1795Q (NSP3:K977Q), T951I (NSP3:T133I), A3333V (NSP5:A70V), V3349I (NSP5:V86I), A3620V (NSP6:A51V), G3676S (NSP6:G107S), F3677L (NSP6:F108L), and S3675/G3676/F3677del (NSP6:S106/G107/F108del). The S1188L substitution in ORF1a (NSP3:S370L) was identified in 43 genomes, with the exception of one sequence presenting the combination K636R (NSP2:K456R), T1004P (NSP3:T186P), K1795Q (NSP3:K977Q), T2007I (NSP3:T1189I), and V3718A (NSP6:V149A). The N189S (NSP2:N9S) and V2793A (NSP4:V30A) mutations in ORF1a were identified for the first time in P.1 genomes sequenced in this study. ORF1b is majoritarily represented by the substitutions P314L (NSP12:P323L) and E1264D (NSP13:E341D), which were described in 43 genomes each. Other nine amino acid substitutions were also found. The S253P (NS3:S253P) mutation in ORF3a occurred in 44 sequences, occasionally followed by other substitutions such as P25L, P1045S, S117I, W131C, and E181V. Finally, the mutations ORF8:E92K (NS8:E92K) and ORF9b:Q77E (accessory protein, alternative ORF from nucleocapsid - N - gene) were assigned to 43 and 44 genome sequences, respectively. Other substitutions include E59D in ORF8 (NS8:E59D), as well as ORF7a:E22D (NS7a:E22D) and ORF7b:S31L (NS7b:S31L).

**Table 2.**
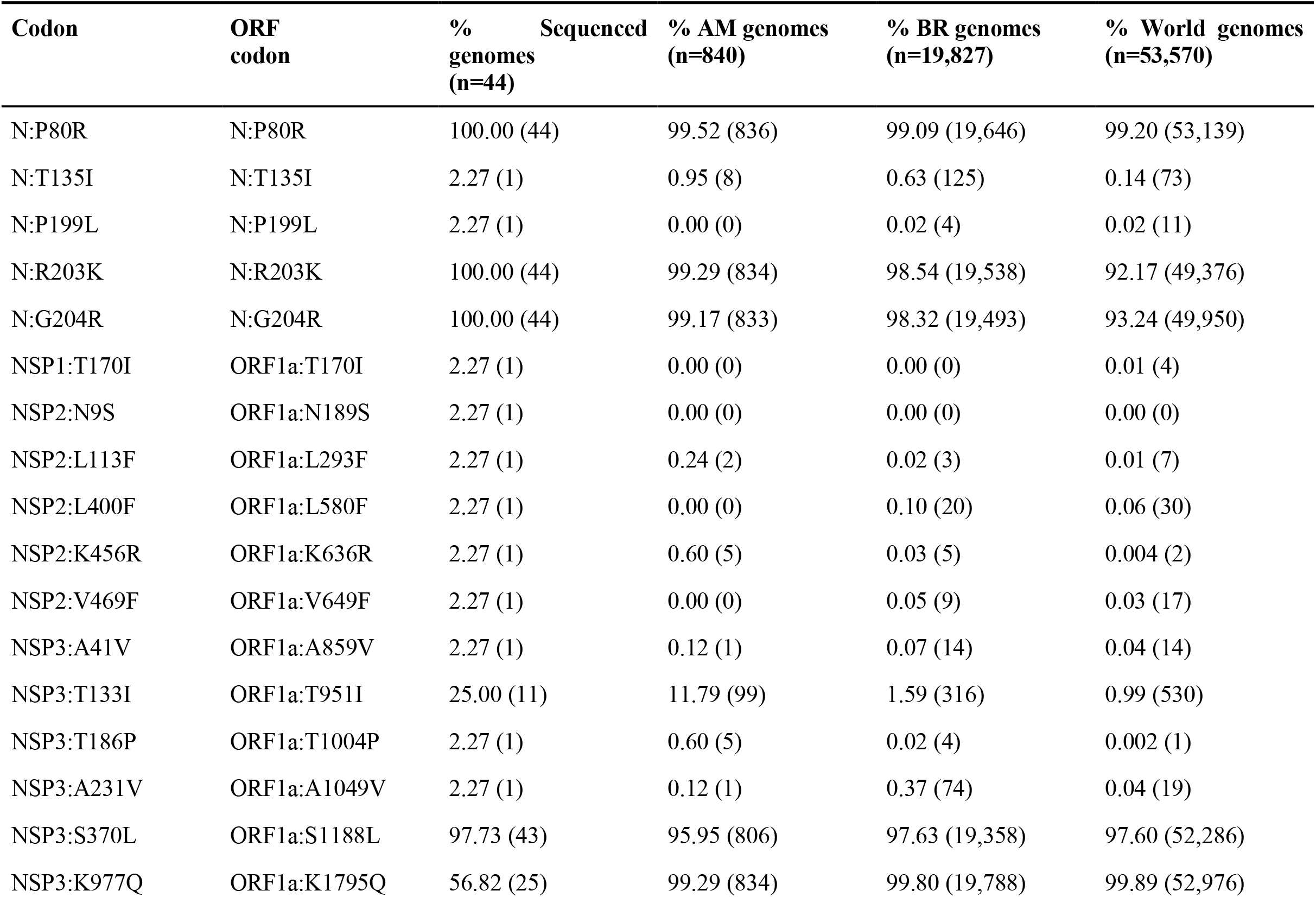
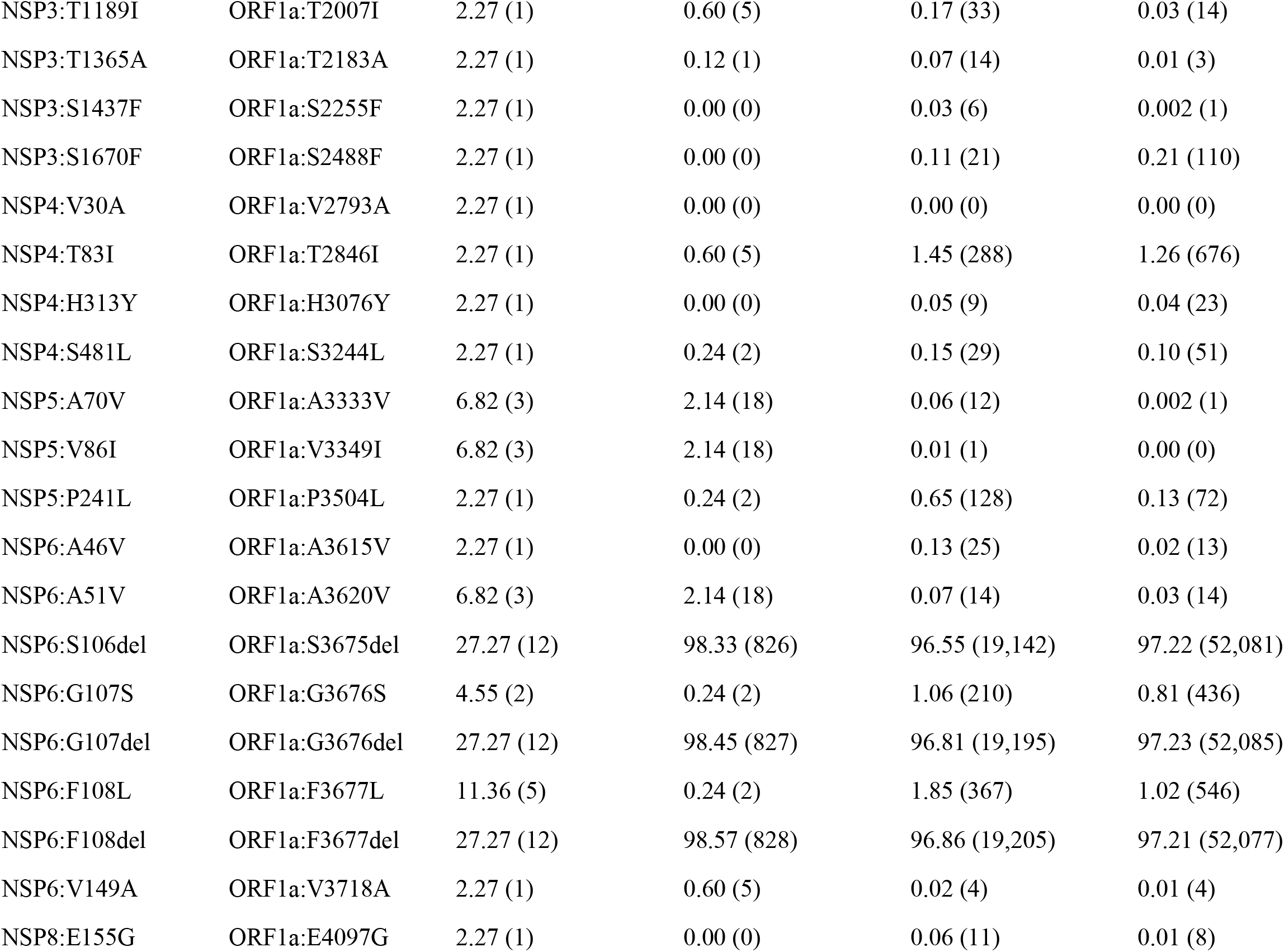
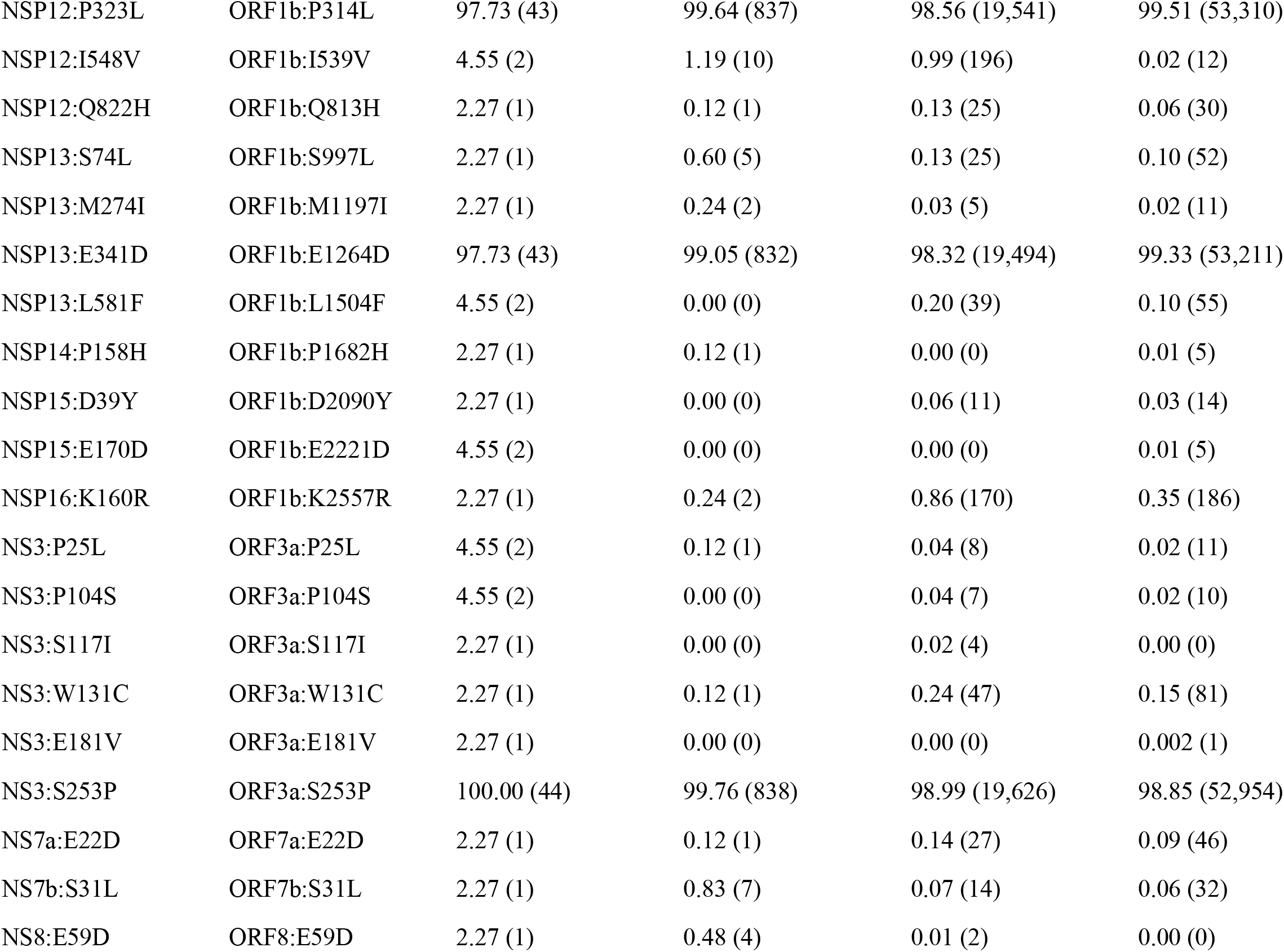
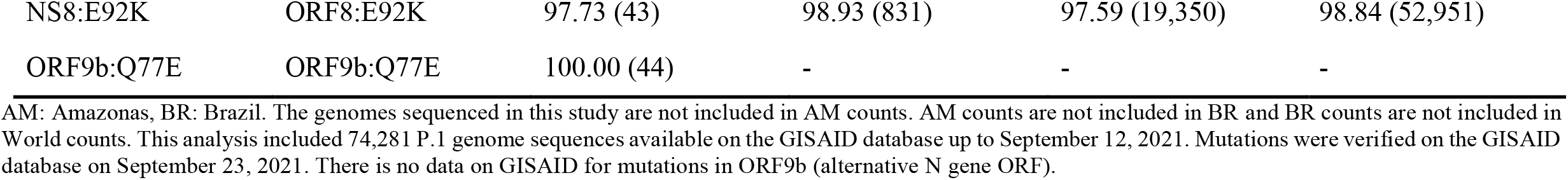
Frequency of mutations identified in the P.1 sequenced genomes (this study) compared with sequences from Amazonas, Brazil and World (up to September 12, 2021) available on GISAID.

| **Codon** | **ORF codon** | **% Sequenced genomes (n=44)** | **% AM genomes (n=840)** | **% BR genomes (n=19,827)** | **% World genomes (n=53,570)** |
| --- | --- | --- | --- | --- | --- |
| N:P80R | N:P80R | 100.00 (44) | 99.52 (836) | 99.09 (19,646) | 99.20 (53,139) |
| N:T135I | N:T135I | 2.27 (1) | 0.95 (8) | 0.63 (125) | 0.14 (73) |
| N:P199L | N:P199L | 2.27 (1) | 0.00 (0) | 0.02 (4) | 0.02 (11) |
| N:R203K | N:R203K | 100.00 (44) | 99.29 (834) | 98.54 (19,538) | 92.17 (49,376) |
| N:G204R | N:G204R | 100.00 (44) | 99.17 (833) | 98.32 (19,493) | 93.24 (49,950) |
| NSP1:T170I | ORF1a:T170I | 2.27 (1) | 0.00 (0) | 0.00 (0) | 0.01 (4) |
| NSP2:N9S | ORF1a:N189S | 2.27 (1) | 0.00 (0) | 0.00 (0) | 0.00 (0) |
| NSP2:L113F | ORF1a:L293F | 2.27 (1) | 0.24 (2) | 0.02 (3) | 0.01 (7) |
| NSP2:L400F | ORF1a:L580F | 2.27 (1) | 0.00 (0) | 0.10 (20) | 0.06 (30) |
| NSP2:K456R | ORF1a:K636R | 2.27 (1) | 0.60 (5) | 0.03 (5) | 0.004 (2) |
| NSP2:V469F | ORF1a:V649F | 2.27 (1) | 0.00 (0) | 0.05 (9) | 0.03 (17) |
| NSP3:A41V | ORF1a:A859V | 2.27 (1) | 0.12 (1) | 0.07 (14) | 0.04 (14) |
| NSP3:T133I | ORF1a:T951I | 25.00 (11) | 11.79 (99) | 1.59 (316) | 0.99 (530) |
| NSP3:T186P | ORF1a:T1004P | 2.27 (1) | 0.60 (5) | 0.02 (4) | 0.002 (1) |
| NSP3:A231V | ORF1a:A1049V | 2.27 (1) | 0.12 (1) | 0.37 (74) | 0.04 (19) |
| NSP3:S370L | ORF1a:S1188L | 97.73 (43) | 95.95 (806) | 97.63 (19,358) | 97.60 (52,286) |
| NSP3:K977Q | ORF1a:K1795Q | 56.82 (25) | 99.29 (834) | 99.80 (19,788) | 99.89 (52,976) |
| NSP3:T1189I | ORF1a:T2007I | 2.27 (1) | 0.60 (5) | 0.17 (33) | 0.03 (14) |
| NSP3:T1365A | ORF1a:T2183A | 2.27 (1) | 0.12 (1) | 0.07 (14) | 0.01 (3) |
| NSP3:S1437F | ORF1a:S2255F | 2.27 (1) | 0.00 (0) | 0.03 (6) | 0.002 (1) |
| NSP3:S1670F | ORF1a:S2488F | 2.27 (1) | 0.00 (0) | 0.11 (21) | 0.21 (110) |
| NSP4:V30A | ORF1a:V2793A | 2.27 (1) | 0.00 (0) | 0.00 (0) | 0.00 (0) |
| NSP4:T83I | ORF1a:T2846I | 2.27 (1) | 0.60 (5) | 1.45 (288) | 1.26 (676) |
| NSP4:H313Y | ORF1a:H3076Y | 2.27 (1) | 0.00 (0) | 0.05 (9) | 0.04 (23) |
| NSP4:S481L | ORF1a:S3244L | 2.27 (1) | 0.24 (2) | 0.15 (29) | 0.10 (51) |
| NSP5:A70V | ORF1a:A3333V | 6.82 (3) | 2.14 (18) | 0.06 (12) | 0.002 (1) |
| NSP5:V86I | ORF1a:V3349I | 6.82 (3) | 2.14 (18) | 0.01 (1) | 0.00 (0) |
| NSP5:P241L | ORF1a:P3504L | 2.27 (1) | 0.24 (2) | 0.65 (128) | 0.13 (72) |
| NSP6:A46V | ORF1a:A3615V | 2.27 (1) | 0.00 (0) | 0.13 (25) | 0.02 (13) |
| NSP6:A51V | ORF1a:A3620V | 6.82 (3) | 2.14 (18) | 0.07 (14) | 0.03 (14) |
| NSP6:S106del | ORF1a:S3675del | 27.27 (12) | 98.33 (826) | 96.55 (19,142) | 97.22 (52,081) |
| NSP6:G107S | ORF1a:G3676S | 4.55 (2) | 0.24 (2) | 1.06 (210) | 0.81 (436) |
| NSP6:G107del | ORF1a:G3676del | 27.27 (12) | 98.45 (827) | 96.81 (19,195) | 97.23 (52,085) |
| NSP6:F108L | ORF1a:F3677L | 11.36 (5) | 0.24 (2) | 1.85 (367) | 1.02 (546) |
| NSP6:F108del | ORF1a:F3677del | 27.27 (12) | 98.57 (828) | 96.86 (19,205) | 97.21 (52,077) |
| NSP6:V149A | ORF1a:V3718A | 2.27 (1) | 0.60 (5) | 0.02 (4) | 0.01 (4) |
| NSP8:E155G | ORF1a:E4097G | 2.27 (1) | 0.00 (0) | 0.06 (11) | 0.01 (8) |
| NSP12:P323L | ORF1b:P314L | 97.73 (43) | 99.64 (837) | 98.56 (19,541) | 99.51 (53,310) |
| NSP12:I548V | ORF1b:I539V | 4.55 (2) | 1.19 (10) | 0.99 (196) | 0.02 (12) |
| NSP12:Q822H | ORF1b:Q813H | 2.27 (1) | 0.12 (1) | 0.13 (25) | 0.06 (30) |
| NSP13:S74L | ORF1b:S997L | 2.27 (1) | 0.60 (5) | 0.13 (25) | 0.10 (52) |
| NSP13:M274I | ORF1b:M1197I | 2.27 (1) | 0.24 (2) | 0.03 (5) | 0.02 (11) |
| NSP13:E341D | ORF1b:E1264D | 97.73 (43) | 99.05 (832) | 98.32 (19,494) | 99.33 (53,211) |
| NSP13:L581F | ORF1b:L1504F | 4.55 (2) | 0.00 (0) | 0.20 (39) | 0.10 (55) |
| NSP14:P158H | ORF1b:P1682H | 2.27 (1) | 0.12 (1) | 0.00 (0) | 0.01 (5) |
| NSP15:D39Y | ORF1b:D2090Y | 2.27 (1) | 0.00 (0) | 0.06 (11) | 0.03 (14) |
| NSP15:E170D | ORF1b:E2221D | 4.55 (2) | 0.00 (0) | 0.00 (0) | 0.01 (5) |
| NSP16:K160R | ORF1b:K2557R | 2.27 (1) | 0.24 (2) | 0.86 (170) | 0.35 (186) |
| NS3:P25L | ORF3a:P25L | 4.55 (2) | 0.12 (1) | 0.04 (8) | 0.02 (11) |
| NS3:P104S | ORF3a:P104S | 4.55 (2) | 0.00 (0) | 0.04 (7) | 0.02 (10) |
| NS3:S117I | ORF3a:S117I | 2.27 (1) | 0.00 (0) | 0.02 (4) | 0.00 (0) |
| NS3:W131C | ORF3a:W131C | 2.27 (1) | 0.12 (1) | 0.24 (47) | 0.15 (81) |
| NS3:E181V | ORF3a:E181V | 2.27 (1) | 0.00 (0) | 0.00 (0) | 0.002 (1) |
| NS3:S253P | ORF3a:S253P | 100.00 (44) | 99.76 (838) | 98.99 (19,626) | 98.85 (52,954) |
| NS7a:E22D | ORF7a:E22D | 2.27 (1) | 0.12 (1) | 0.14 (27) | 0.09 (46) |
| NS7b:S31L | ORF7b:S31L | 2.27 (1) | 0.83 (7) | 0.07 (14) | 0.06 (32) |
| NS8:E59D | ORF8:E59D | 2.27 (1) | 0.48 (4) | 0.01 (2) | 0.00 (0) |
| NS8:E92K | ORF8:E92K | 97.73 (43) | 98.93 (831) | 97.59 (19,350) | 98.84 (52,951) |
| ORF9b:Q77E | ORF9b:Q77E | 100.00 (44) | - | - | - |

AM: Amazonas, BR: Brazil. The genomes sequenced in this study are not included in AM counts. AM counts are not included in BR and BR counts are not included in World counts. This analysis included 74,281 P.1 genome sequences available on the GISAID database up to September 12, 2021. Mutations were verified on the GISAID database on September 23, 2021. There is no data on GISAID for mutations in ORF9b (alternative N gene ORF).

According to the collection date registered on GISAID, the first occurrence for each spike mutation identified in all sequenced genomes in this study is shown in Table 3. Five mutations have their first occurrence in Brazilian P.1 genomes sequenced in this study: A243/L244del, N188del, P209H, and T1066A. Specifically, the A243/L244 double deletion identified in our samples is the first occurrence of this combination in P.1 genomes worldwide. Before February 22, 2021, these deletions were mostly found in B.1.351 genomes (n= 6,461 of 8,009). The case of N188del is not clear, since several genomes may contain alignment errors and, consequently, not being possible to identify this mutation on the GISAID data. The P209H and T1066A substitutions, which are present in lineages such as B.1.1.7 and AY.4 (B.1.617.2 derivative), are only represented by our Amazonas genomes in P.1 data. Moreover, the genome sequences in this study represent the first occurrence of mutations A688V, D215G, and L189 del in P.1 genomes from Amazonas.

**Table 3.** First occurrence (by collection date) registered on GISAID for spike mutations identified in the sequenced genomes from this study.

| **Mutation** | **Amazonas** | **Brazil** | **Brazil (outside Amazonas)** | **World (outside Brazil)** |
| --- | --- | --- | --- | --- |
| L18F | 2020-12-03 | 2020-09-22 / PE | 2020-09-22 / PE | 2020-12-17 / Peru* |
| T20N | 2020-12-03 | 2020-09-22 / PE | 2020-09-22 / PE | 2020-12-17 / Peru* |
| P26S | 2020-12-03 | 2020-09-22 / PE | 2020-09-22 / PE | 2020-12-17 / Peru* |
| D138Y | 2020-12-03 | 2020-10-01 / PB | 2020-10-01 / PB | 2020-12-17 / Peru* |
| N188del | **2021-02-23** | **2021-02-23 / AM** | - | 2021-05-25 / Suriname |
| L189del | **2021-02-23** | 2021-01-27 / AL | 2021-01-27 / AL | 2021-01-28 / Taiwan |
| R190S | 2020-12-03 | 2020-10-01 / PB | 2020-10-01 / PB | 2020-12-17 / Peru* |
| P209H | **2020-02-25** | **2021-02-25 / AM** | - | - |
| D215G | 2021-03-10 | 2021-03-08 / GO | 2021-03-08 / GO | 2021-04-07 / USA |
| A243del | **2021-02-22** | **2021-02-22 / AM** | 2021-02-23 / SE | 2021-03-23 / USA |
| L244del | **2021-02-22** | **2021-02-22 / AM** | 2021-02-23 / SE | 2021-03-23 / USA |
| K417T | 2020-12-03 | 2020-09-22 / PE | 2020-09-22 / PE | 2020-12-17 / Peru* |
| E484K | 2020-12-03 | 2020-10-01 / PB | 2020-10-01 / PB | 2020-11-11 / USA* |
| N501Y | 2020-12-03 | 2020-10-01 / PB | 2020-10-01 / PB | 2020-12-17 / Peru* |
| D614G | 2020-12-03 | 2020-09-11 / SP | 2020-09-11 / SP | 2020-11-11 / USA* |
| H655Y | 2020-12-03 | 2020-09-11 / SP | 2020-09-11 / SP | 2020-12-17 / Peru* |
| A688V | 2021-02-22 | 2021-01-26 / SP | 2021-01-26 / SP | 2021-02-15 / Peru* |
| T1027I | 2020-12-03 | 2020-09-11 / SP | 2020-09-11 / SP | 2020-11-11 / USA* |
| T1066A | **2020-02-25** | **2021-02-25 / AM** | - | - |
| V1176F | 2020-12-03 | 2020-09-11 / SP | 2020-09-11 / SP | 2020-12-17 / Peru* |

Only complete collection dates were considered for this analysis. AL: Alagoas, AM: Amazonas, GO: Goiás, PB: Paraíba, PE: Pernambuco, SE: Sergipe, SP: São Paulo. Date verification on September 23, 2021. The first occurrence in the world of some mutations that were identified in the genomes sequenced in this study are highlighted in bold. \*According to the GISAID database, there are three P.1 genomes from the USA carrying these mutations with collection dates from April and May, 2020 (before their first occurrence in Brazil). However, we do not find any publication confirming this USA origin of P.1 lineage.

Following the collection dates on Supplementary File 2, it is possible to observe that the first occurrence for the mutations N:P199L, NSP3:T1365A, NSP3:S1437F, NSP3:S1670F, NSP4:S481L, NSP5:A70V, NSP6:A51V, NSP8:E155G, NSP13:M274I, NSP15:D39Y, NSP16:K160R, and NS3:P104S in P.1 sequences from Brazil is related to the sequenced genomes from this study. Except for N:P199L and NSP16:K160R, previously reported in the USA and Italy, all others have their first appearance for P.1 genomes in the world with the genomes from this study. Before February 22, 2021, N:P199L is found in more than 100,000 genomes worldwide, majoritarily in lineages such as B.1.2, B.1.221, and B.1.596, despite being present in several other lineages. Other substitutions, such as NSP1:T170I, NSP14:P158H, NSP15:E170D, have their only P.1 occurrence in Brazil identified in the genomes from this study, despite other countries posteriorly reporting their presence. A third group was formed by the mutations NSP2:N9S, NSP4:V30A, and NS3:E181V, which were not found in any other place from Brazil or the World. For 28 mutations, our genomes represent their first occurrence in P.1 Amazonas samples.

### Recombination analysis of northern samples

The analysis performed by the RDP software did not find any statistically supported recombination event in the set of 1,931 northern genomes from Brazil. Similarly, the genetic distance plot inferred by the recan package did not suggest any potential breakpoints in the evaluated sequences. The indicated peaks of genetic distance are related to the presence of undefined genomic regions in the sequenced samples (Figure 3).

**Figure 3.**
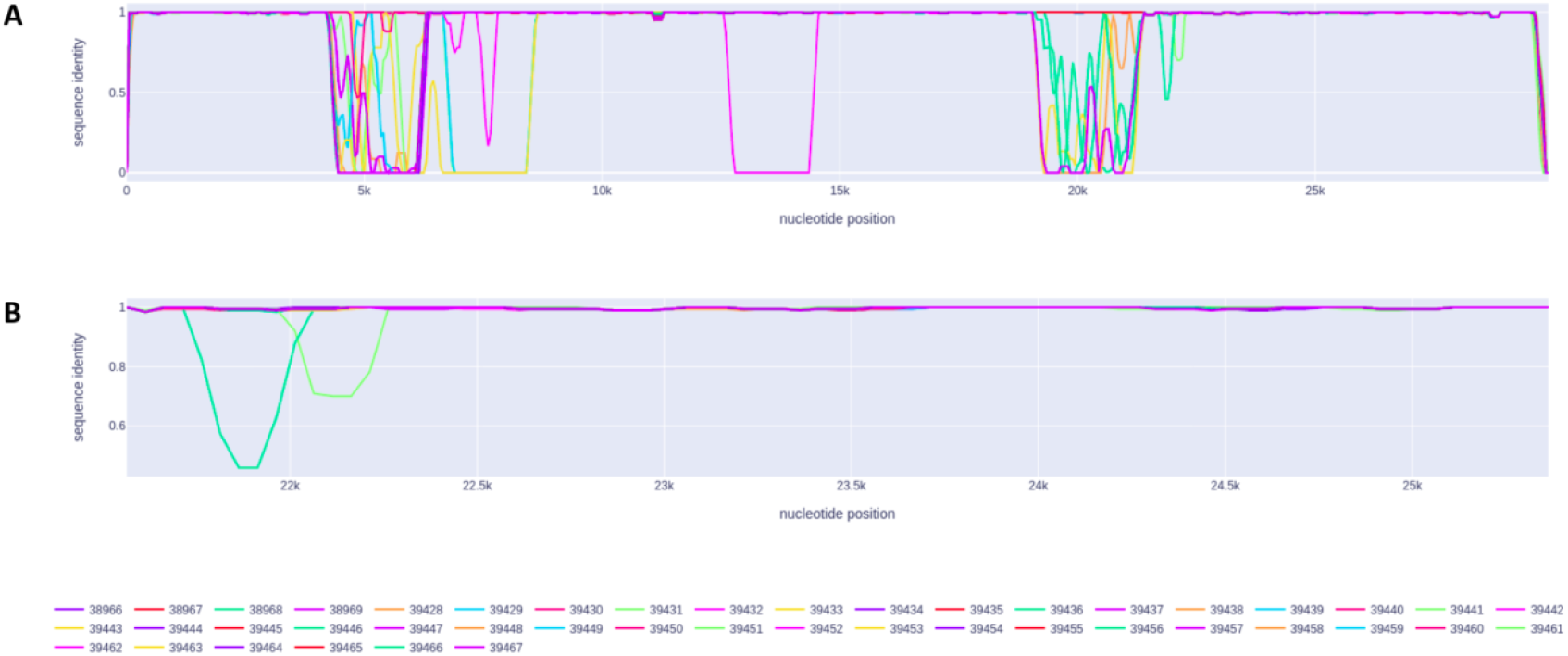
Genetic distance plot of the 44 sequenced genomes in comparison with the NC_045512.2 reference genome (Wuhan). (A) Genomic sequence identity. (B) Sequence identity for the spike gene region.

### Phylogenomic analysis of northern samples

The phylogenetic analysis of the 1,931 genomes obtained by the maximum likelihood method showed the formation of statistically supported clades for the lineages P.1 (100% of branch support by the SH-aLTR test and ultrafast bootstrap) and P.2 (93% of branch support by the SH-aLRT test and 96% by the ultrafast bootstrap) as derivatives of the B.1.1.28 lineage (Figure 4). All samples from this study were included in the P.1 monophyletic group.

**Figure 4.**
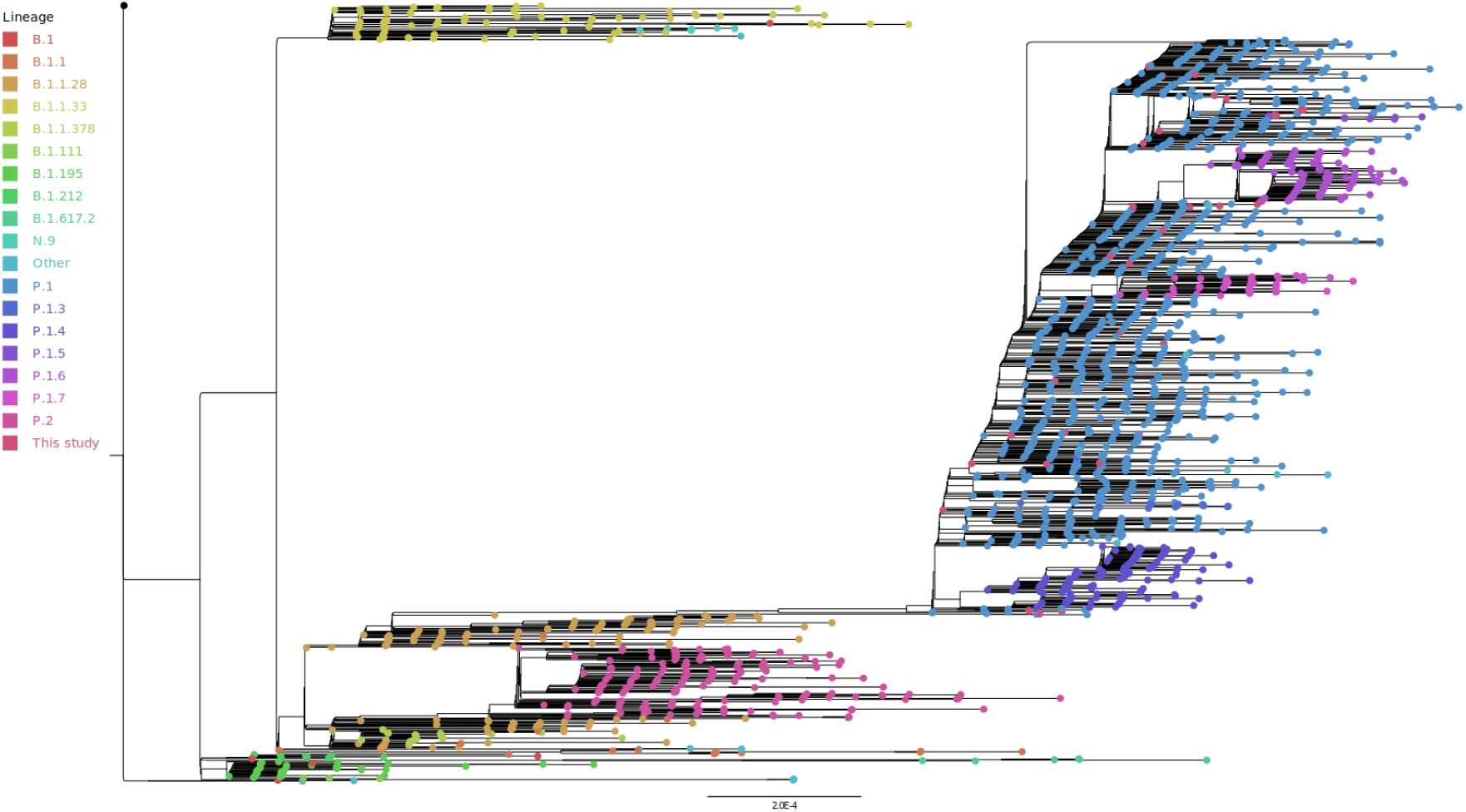
Maximum Likelihood tree of 1,887 unique genomes from the North region - Brazil, added to the 44 sequenced samples of this study. Pangolin lineages represented by < five genomes are labeled as “Other”.

Inside the P.1 clade was identified a group of P.1.7 genomes, with 90% of statistical support by SH-aLRT and 83% by the ultrafast bootstrap, as well as a group of P.1.6 genomes with statistical support of 78.7% by the SH-aLRT and 78% by the ultrafast bootstrap test. P.1.4 formed a group with 77.8 and 71% for SH-aLRT and ultrafast bootstrap branch support, respectively. Both P.1.4 and P.1.6 groups did not reach the minimal branch support cutoff of 80% in both statistical tests. Two P.1.7 genomes were found outside the P.1.7 group, clustering with P.1 sequences. The analysis of these two genomes showed the absence of the P.1.7 lineage defining mutation P681H for the spike sequence.

As expected, B.1.1.33 and N.9 sequences were clustered, despite the absence of statistical validation when considering both methods together (SH-aLRT branch support of 91.9 and 43% by the SH-aLRT and ultrafast bootstrap). However, the N.9 subgroup had 98.5 and 100% of branch support - SH-aLRT and ultrafast bootstrap test, validating it as a clade. Sequences from the lineages B.1.195, B.1.212, B.1.617.2, and B.1.111 formed their own groups in the B.1 related most external branch (>75% of statistical support by all tests). Two B.1 sequences were found inside the B.1.195 and B.1.212 groups (one in each cluster).

### Phylogeny of spike and non-structural proteins of northern samples

Since the spike and ORF1a regions accumulated a higher number of mutations in the sequenced genomes, phylogenetic analyses were performed for these sequences, allowing to compare the clusterization of the North samples according to specific protein profiles. The best evolutionary model was predicted for each sequence alignment, as follows: spike - GTR+G4, NSP1 - TrNef, NSP2 - TIM1, NSP3 - GTR+G4, NSP4 - TrN, NSP5 - TrN, NSP6 - TIM1+G4, NSP7 - HKY, NSP8 - TrN, NSP9 - TrN+I, and NSP10 - TIM1. In fact, the exclusion of sequence duplicates reduced the sample data set, leading to a lack of sequence variability for some ORF1a regions. The evident difference in sequence conservation among NSPs as well as the inference of different evolutionary models denote the various selective pressures possible acting on these non-structural proteins.

Interestingly, in the spike phylogeny, the sequences clustered according to the assigned genome lineages (Figure 5A). According to the spike mutational pattern, which is reflected in the phylogenetic tree of 584 unique sequences, P.1, P.1.4, P.1.6, and P.1.7 formed a monophyletic group, with 96.6 and 99% of branch support by the SH-aLRT and ultrafast bootstrap tests, inside the P.2 cluster. The group including P.1, P.2, and B.1.1.28 genomes showed 85.4 and 27% of branch support by SH-aLRT and ultrafast bootstrap tests, which indicates it does not form a clade.

**Figure 5.**
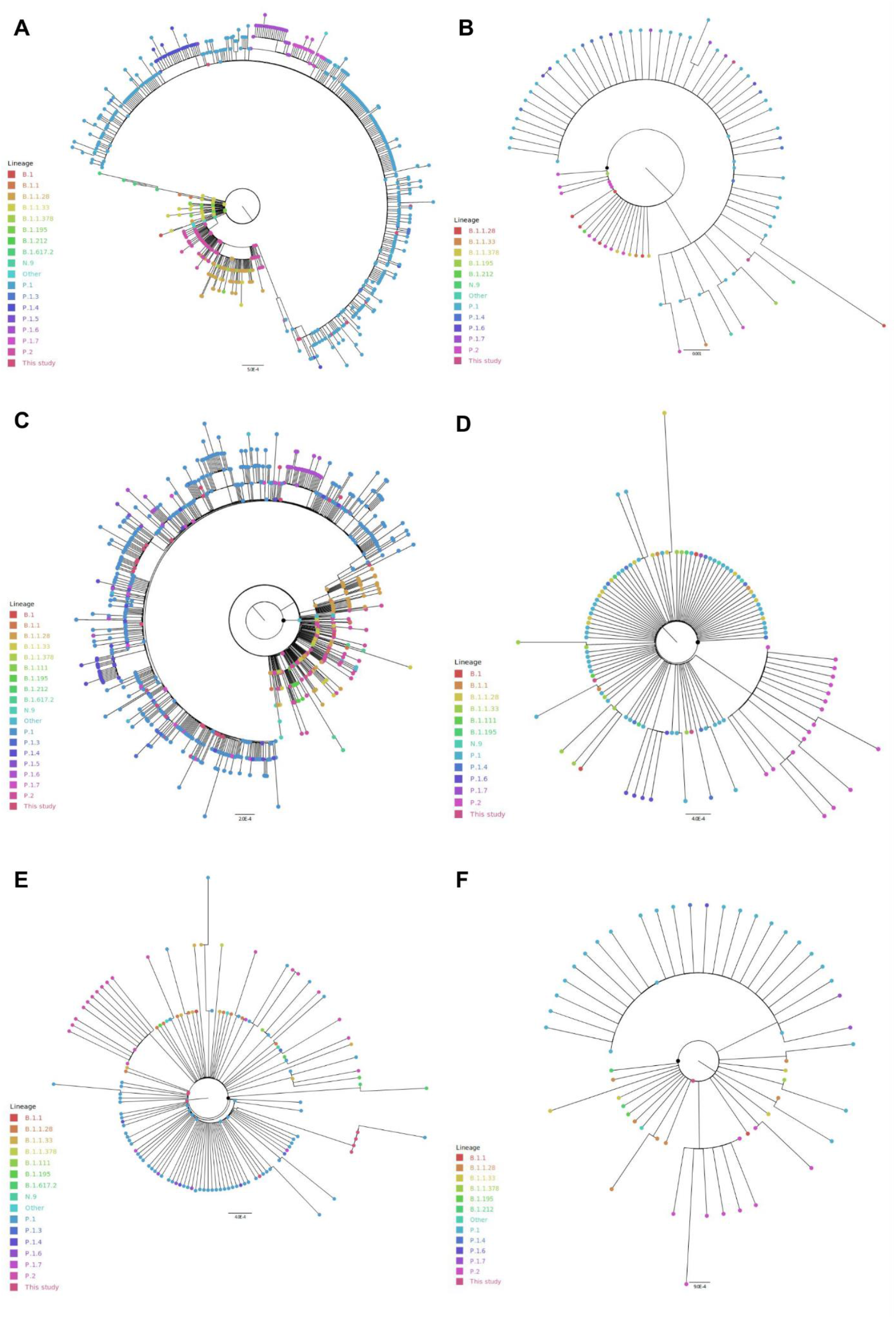
Phylogenetic analyses of spike and non-structural proteins from ORF1a including northern Brazilian SARS-CoV-2 genomes. (A) spike, (B) NSP1, (C) NSP3, (D) NSP5, (E) NSP6, and (F) NSP9.

For the non-structural protein 1 (NSP1 - 86 unique sequences), the P.1 sequences and their derivatives (P.1.4, P.1.6, and P.1.7) were clustered together, without statistical validation (84.6% of branch support by the SH-aLRT test and 13% by ultrafast bootstrap). A few random samples from different lineages such as B.1.1.28, B.1.1.33, P.2, N.9, and B.1.195 were found in this cluster (Figure 5B). In NSP3 phylogeny (747 unique sequences), P.1, P.1.4, P.1.5, P.1.6, and P.1.7 sequences also formed statistically non validated group (86.3 and 77% of branch support by the SH-aLRT and ultrafast bootstrap tests) (Figure 5C). B.1.195 formed a cluster with support of 86% and 18% by aLRT and ultrafast bootstrap, which is not statistically well-supported. NSP5 (116 unique sequences) presented a group of P.2 sequences with 86.3 and 50% of branch support by the SH-aLRT and ultrafast bootstrap tests, respectively (Figure 5D). Additionally, there is a P.1.6 group with 84.2 and 63% of branch support by SH-aLRT and ultrafast bootstrap. Both groups achieved branch support values below the cutoff considered for clade validation. For NSP6 phylogeny (140 unique sequences), P.1 sequences (and their derivatives) majoritarily clustered together. However, the cluster is not statistically supported (Figure 5E). The same pattern was observed for P.2 sequences. A similar model was obtained for NSP9 (58 unique sequences), where most of all P.1 sequences and P.2 sequences clustered without statistical support (Figure 5F).

In all these phylogenies, it was observed the dispersion of lineages along the tree by the formation of some statistically well-supported groups including different lineages, which may suggest a low genetic divergence among them in the ORF1a. In this way, it was not possible to identify a sequence clustering based on the expected similarity intralineage for the non-structural proteins 2 (329 unique sequences), 4 (172 unique sequences), 7 (29 unique sequences), 8 (58 unique sequences), and 10 (38 unique sequences) (Supplementary File 3). Considering the SH-aLRT and ultrafast bootstrap cutoff values (≥80% for SH-aLRT and ≥95% for ultrafast bootstrap), the formation of a monophyletic group was only supported for P.1 lineage and their derivatives (P.1.4, P.1.6, and P.1.7) in the spike phylogenetic tree.

### Molecular evolution of spike and non-structural proteins of northern samples

The selection tests performed by the FUBAR method using the spike sequence alignment detected 23 sites potentially under pervasive diversifying selective pressure (Table 4). Of these, five (5, 138, 417, 681, and 1264) were also indicated by the FEL method. Interestingly, the sites 138 and 417 were observed in MEME results for episodic diversifying selection. A number of 78 sites were found to be under purifying selective pressure by FEL, of which 18 were also predicted by SLAC. The full set of results, including FEL, MEME, and SLAC methods, is available on the Supplementary File 4.

**Table 4.** Fast Unbiased AppRoximate Bayesian (FUBAR) analysis of pervasive diversifying selection on spike protein.

| codon | alpha | beta | posterior probability |
| --- | --- | --- | --- |
| <u>5</u> | <u>0.883</u> | <u>8.954</u> | <u>0.9688</u> |
| **20** | **1.823** | **12.403** | **0.9267** |
| 29 | 0.917 | 6.771 | 0.9244 |
| 49 | 1.045 | 8.832 | 0.9502 |
| 67 | 0.932 | 5.622 | 0.9036 |
| 95 | 0.920 | 6.822 | 0.9248 |
| 96 | 1.246 | 7.541 | 0.9075 |
| **<u>138</u>** | **<u>1.100</u>** | **<u>19.873</u>** | **<u>0.9865</u>** |
| 222 | 0.894 | 8.019 | 0.9562 |
| 257 | 0.922 | 6.516 | 0.9204 |
| **<u>417</u>** | **<u>0.863</u>** | **<u>10.051</u>** | **<u>0.9702</u>** |
| **484** | **2.688** | **29.850** | **0.9627** |
| **501** | **1.062** | **9.105** | **0.9363** |
| 613 | 1.266 | 12.121 | 0.9555 |
| 653 | 0.932 | 5.622 | 0.9036 |
| **655** | **1.066** | **7.230** | **0.9168** |
| 677 | 1.246 | 8.376 | 0.9173 |
| <u>681</u> | <u>0.887</u> | <u>8.815</u> | <u>0.9678</u> |
| 689 | 1.067 | 6.999 | 0.9136 |
| 769 | 1.361 | 8.428 | 0.9111 |
| 1174 | 0.932 | 5.623 | 0.9037 |
| **1176** | **1.010** | **11.358** | **0.9726** |
| <u>1264</u> | <u>0.799</u> | <u>5.875</u> | <u>0.9244</u> |

Underlined data indicate codons also identified by FEL/MEME with positive selection evidence. Highlighted in bold are the mutations found in the 44 genomes of this study.

About the FUBAR identified sites, seven are largely found in P.1 genomes (as lineage-defining mutations), including in those 44 sequenced in this study. This mutation set, which comprised substitutions in sites 20, 138, 417, 484, 501, 655, and 1176, is found in more than 90% of the P.1 genomes in the world (n = 62,863), as well in other 9,282 genomes currently assigned in lineages such as B.1, B.1.1, B.1.628, N.9, and P.1-derivatives (e.g.: P.1.2, P.1.1.6, P.1.10). Mutations such as L5F are mostly found in the lineages B.1.526 and B.1.1.7 worldwide. In Brazil, its presence is majoritarily identified in P.1 lineage. Substitutions such as T29I, H49Y/L, A67S/T, T95I, E96D/G, A222V/S, G257D/S, Q613H/E, A653S/V, Q677H, S689I, G769V/R, A1174V/S, and V1264L were found in a wide range of brazilian lineages (average mean of seven lineages per mutation, 50% with at least five lineages), despite their low frequency (average mean of 68 sequences per mutation, 50% of them with at least 13 sequences), including VOC and VOI lineages. However, the amino acid substitutions P681H/R/T present a higher frequency. The P681R mutation is found in more than 2,800 genomes and 35 lineages. The variation P681H exceeds 2,000 genomes in 21 different lineages.

In the constellation of possible substitutions occurring in the selected sites, it was observed that the mutation H49L is only found in one P.1 genome from Amazonas. About 40.30% of sequences with the prevalent mutation H49Y were identified in genomes from Amazonas and Tocantins, comprising the P.1, B.1.1.28, and B.1.195 lineages. Similarly, P681T has its unique appearance in a P.1.4 genome from Amazonas. Of the eight genomes presenting the A1174V substitution, seven are from the North region (Amapá and Pará) including P.1 and P.1.7 lineages.

In relation to mutations identified in ORF1a by the selection methods applied, NSP1 (three sites), NSP3 (six sites), and NSP9 (two sites) achieved the higher number of sites suggested to be under adaptive pressure (Table 5). For NSP1, the sites V5A/I, R43C/H, and R77Q/L presented evidence of pervasive diversifying selection by the FUBAR analysis. Specifically, site 5 was also identified by the FEL method. Interestingly, 136 genomes from 16 lineages in the world, including P.1 and B.1.617.2, carry the V5A mutation. In Brazil, this substitution was previously detected in only one genome (in Amazonas) from the P.1.4 lineage. The V5I substitution, identified in 224 sequences in the world (two in Brazil), was detected in a set of 27 lineages; the Amapá sample, specifically, belongs to P.1. In turn, R43C is part of 348 genomes from 53 lineages. In Brazil, three genomes (two of them from Amazonas) belonging to the lineages P.1, B.1.1, and B.1.1.33 present this mutation. The variant R43H is found in 279 sequences in the world, comprising 34 lineages, of which P.1 is represented by a sample from Pará. Finally, R77Q, which was found in 280 genomes in the world and 29 lineages, has in Pará its Brazilian representative for the P.1.7 lineage. The least prevalent of the NSP1 substitutions is R77L, with 33 sequences belonging to 15 lineages. The sample from Amapá (P.1) is the only one in Brazilian territory. No sites in NSP1 were found to be under episodic selection by MEME analysis. By FEL method, additional four sites in NSP1 were also detected with evidence of purifying selection: 36, 42, 74, and 156. Of these, site 156 was identified by SLAC as well.

**Table 5.**
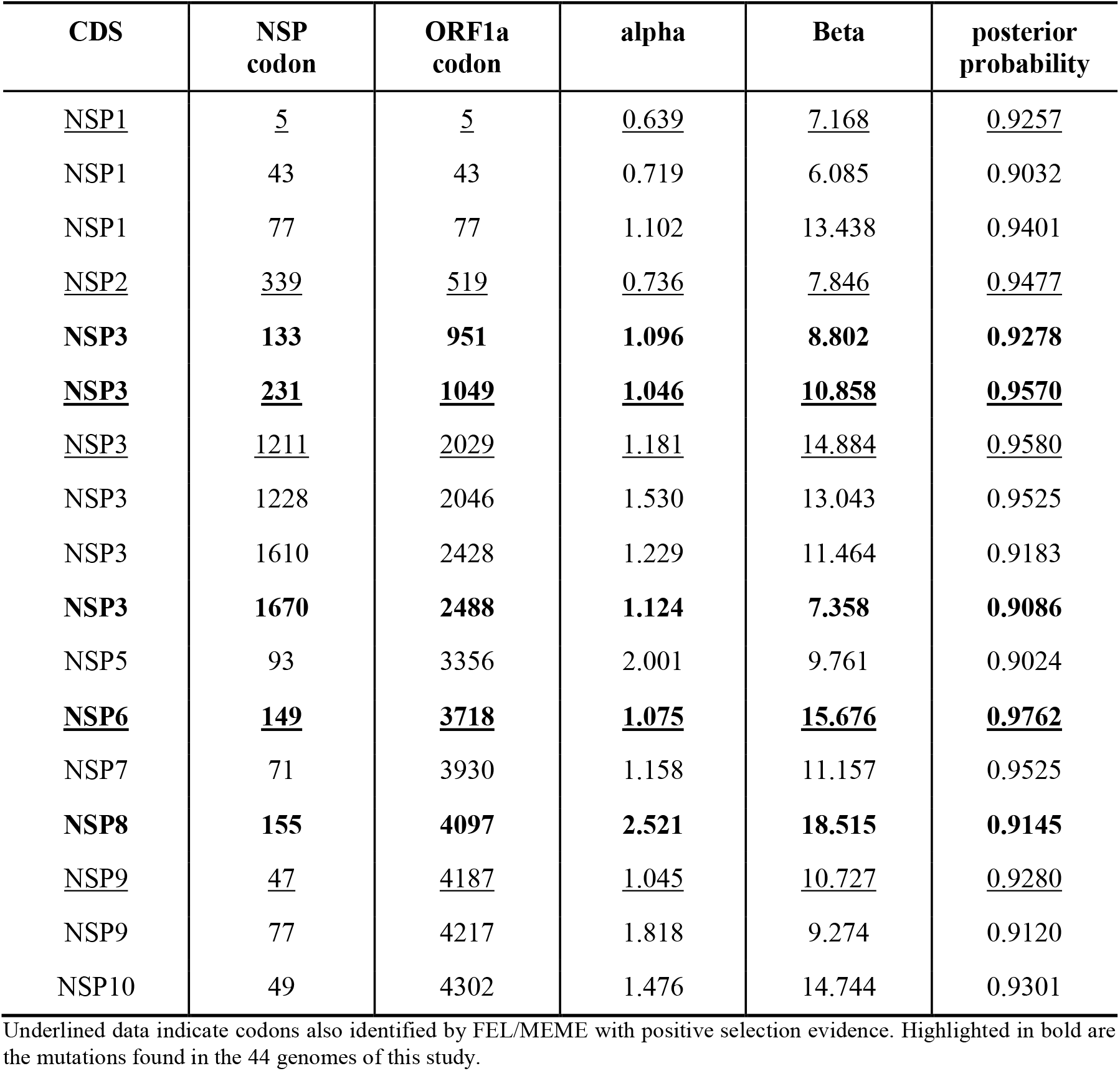
Fast Unbiased AppRoximate Bayesian (FUBAR) analysis of pervasive diversifying selection on non-structural proteins (NSPs) from ORF1a.

| CDS | NSP codon | ORF1a codon | alpha | Beta | posterior probability |
| --- | --- | --- | --- | --- | --- |
| <u>NSP1</u> | <u>5</u> | <u>5</u> | <u>0.639</u> | <u>7.168</u> | <u>0.9257</u> |
| NSP1 | 43 | 43 | 0.719 | 6.085 | 0.9032 |
| NSP1 | 77 | 77 | 1.102 | 13.438 | 0.9401 |
| <u>NSP2</u> | <u>339</u> | <u>519</u> | <u>0.736</u> | <u>7.846</u> | <u>0.9477</u> |
| **NSP3** | **133** | **951** | **1.096** | **8.802** | **0.9278** |
| **<u>NSP3</u>** | **<u>231</u>** | **<u>1049</u>** | **<u>1.046</u>** | **<u>10.858</u>** | **<u>0.9570</u>** |
| <u>NSP3</u> | <u>1211</u> | <u>2029</u> | <u>1.181</u> | <u>14.884</u> | <u>0.9580</u> |
| NSP3 | 1228 | 2046 | 1.530 | 13.043 | 0.9525 |
| NSP3 | 1610 | 2428 | 1.229 | 11.464 | 0.9183 |
| **NSP3** | **1670** | **2488** | **1.124** | **7.358** | **0.9086** |
| NSP5 | 93 | 3356 | 2.001 | 9.761 | 0.9024 |
| **<u>NSP6</u>** | **<u>149</u>** | **<u>3718</u>** | **<u>1.075</u>** | **<u>15.676</u>** | **<u>0.9762</u>** |
| NSP7 | 71 | 3930 | 1.158 | 11.157 | 0.9525 |
| **NSP8** | **155** | **4097** | **2.521** | **18.515** | **0.9145** |
| <u>NSP9</u> | <u>47</u> | <u>4187</u> | <u>1.045</u> | <u>10.727</u> | <u>0.9280</u> |
| NSP9 | 77 | 4217 | 1.818 | 9.274 | 0.9120 |
| NSP10 | 49 | 4302 | 1.476 | 14.744 | 0.9301 |

Underlined data indicate codons also identified by FEL/MEME with positive selection evidence. Highlighted in bold are the mutations found in the 44 genomes of this study.

The codon 519 from ORF1a, related to the position 339 in the NSP2 CDS, presented two possible amino acid substitutions in our analyzed data. G339S was found in 4.0% (13 of 329 unique NSP2 sequences) of our aligned data, while G339D was identified in only one genome. Similarly, the GISAID database shows that 25,389 genomes in the world present the G339S mutation, including several lineages. In Brazil, a total of 247 sequences carry the G339S substitution in 13 lineages. By the FEL method, other three sites were suggested to be under pervasive adaptive pressure in NSP2: 136, 165, and 447. Moreover, MEME has detected episodic diversifying selection in the 165, 339, 447, and 458. All sites identified as purifying selection targets are described in the Supplementary File 4.

A total of six sites were identified to be under positive selection in NSP3. About 5.9% (44 of 746 unique sequences) of the evaluated North genomes presented the amino acid substitution T133I in the NSP3 sequence alignment. In the world, 3,306 sequences are sampled with this substitution, of which 431 are from Brazil and include lineages such as B.1.1.33, P.1, P.1.5, P.1.9, and P.7. A231V corresponds to 97 Brazilian samples (of 5,281 in the world pool) from nine different lineages in GISAID, while its frequency is about 0.9% of the unique NSP2 sequences in our sequenced data. Despite the lower occurrence, a variant A231T is also present in the analyzed genomes. Another example is K1211N/E, which is mutated in 0.4% of the unique NSP3 sequences from North genomes. The K1211N substitution is found in 50 samples from Brazil, occurring in eight lineages. In the world, 3,253 sequences carry this mutation. The P1228L/S mutations are part of 2.0% of the unique NSP3 sequences (1.2 and 0.8%, respectively). Considering 2,815 deposited genomes in Brazil (and genomes in the world), the P1228L mutation comprises 27 lineages, mostly from the AY group of B.1.617.2 derivatives. Accountable for 1% of the unique sequences, the mutations N1610K/S (0.4/0.1%) and S1670F (0.5%) are found in low numbers (n = 3 / 3 / 43) in Brazil (S1670F has a higher number at world level), despite the diversified set of lineages (2, 2 and 9, respectively). In Brazil, N1610K is only found in Amapá (n = 2) and Roraima (n = 1). The sites 231, 618, 727, 1211, 1303, and 1440 were also detected by FEL/MEME methods. A number of 88 sites were suggested to be under negative selection pressure by FEL, 18 of them were also suggested by SLAC inference.

The substitutions NSP5:T93I/A (1.7/0.9%), NSP6:V149F/A/I (2.1/1.4/0.7%), NSP8:E155G/D (1.7/1.7%), NSP9:D47F (1.7%), NSP9:T77I/A (3.4/1.7%), and NSP10:T49I (5.3%) are found in one up to three unique sequences of their alignment sets. The NSP5 T93 substitution to an isoleucine residue is found in 47 genomes from Brazil (one sequence from Rondônia), including six lineages. In the world, this event is found in 1,690 GISAID genomes. However, the variant T93A only occurs in one Amapá sample from P.2, besides other 54 sequences in the world, in a set of 19 lineages. By the FEL results, the NSP5 sequence presents 15 sites under purifying selection, while the SLAC method only detected site 151. The NSP6:V149F amino acid change was identified in 16,214 genomes, of which 1,090 are from Brazil (16 lineages). The variants V149A/I are associated with a lower number of sequences and lineages when compared to the others. However, this site was also detected by the FEL method. For NSP6, the MEME method also detected evidence of episodic diversifying selection in the sites 106 and 194. Six sites (118, 120, 200, 262, 270, 284, and 286) were identified in the FEL/SLAC analysis for negative selection.

NSP7:L71F (27.6% of the unique sequences from North genomes) reaches 10,569 world samples on the GISAID database; of these, 4,317 (22 lineages) are Brazilian. A low frequency pattern can be observed in the mutated site E155G/D (NSP8). The NSP8 sequence analysis by FEL indicated seven sites under negative selection: 18, 91, 111, 121, 137, 162, and 183. For NSP9, the substitution D47F is only found in one sample from Roraima (B.1.1.33). The substitution NSP9:T77I can be located in 783 genomes, 10 of them from Brazilian samples (six lineages). In NSP9, FEL detected evidence of pervasive selection in site 33, while the MEME method detected in site 47. The sites 6, 28, 31, 42, 68, 93, 103, and 112 were inferred by FEL as submitted to negative selection, while the SLAC method detected the sites 31 and 95. In relation to NSP10:T49I, it is found in 521 genomes in the world, of which 11 (in five lineages) are from Brazil. Six sites NSP10 (30, 59, 64, 65, 66, and 83) were identified to be under purifying selective pressure by FEL.

It is important to notice that nearly all substitutions potentially maintained by adaptive pressure are found in VOC lineages such as P.1, B.1.1.7, and B.1.617.2, besides their derivatives and other lineages of interest and/or monitoring. Despite their low prevalence, the high number of lineages carrying these mutations may also indicate a positive selective pressure acting on these sites, maintaining mutational events in ORF1a.

### Global phylogenomic analyses

The phylogenetic analysis of 4,952 worldwide P.1 and B.1.1.28 genomes with the 44 genome sequences of this study (and the NC_045512.2 SARS-CoV-2 reference genome) showed the formation of a P.1 group (87.9 and 94% of statistical support by the SH-aLRT and ultrafast bootstrap tests) with the B.1.1.28 sequences in the basal branches. One additional B.1.1.28 sequence from Turkey was found inside the P.1 group. However, the analysis of the genomic mutations suggest that this sequence probably belongs to the P.1 lineage due to the presence of substitutions such as L18F, T20N, P26S, E484K, N501Y, and H655Y. Interestingly, this genome is one of the two B.1.1.28 genomes from GISAID presenting the mutation N440K in the spike protein. The other one, also from Turkey, presents this same mutation set and is located at the basis of the P.1 group, separately clustering with a P.1 genome from South Korea also presenting this mutation (Figure 6A).

**Figure 6.**
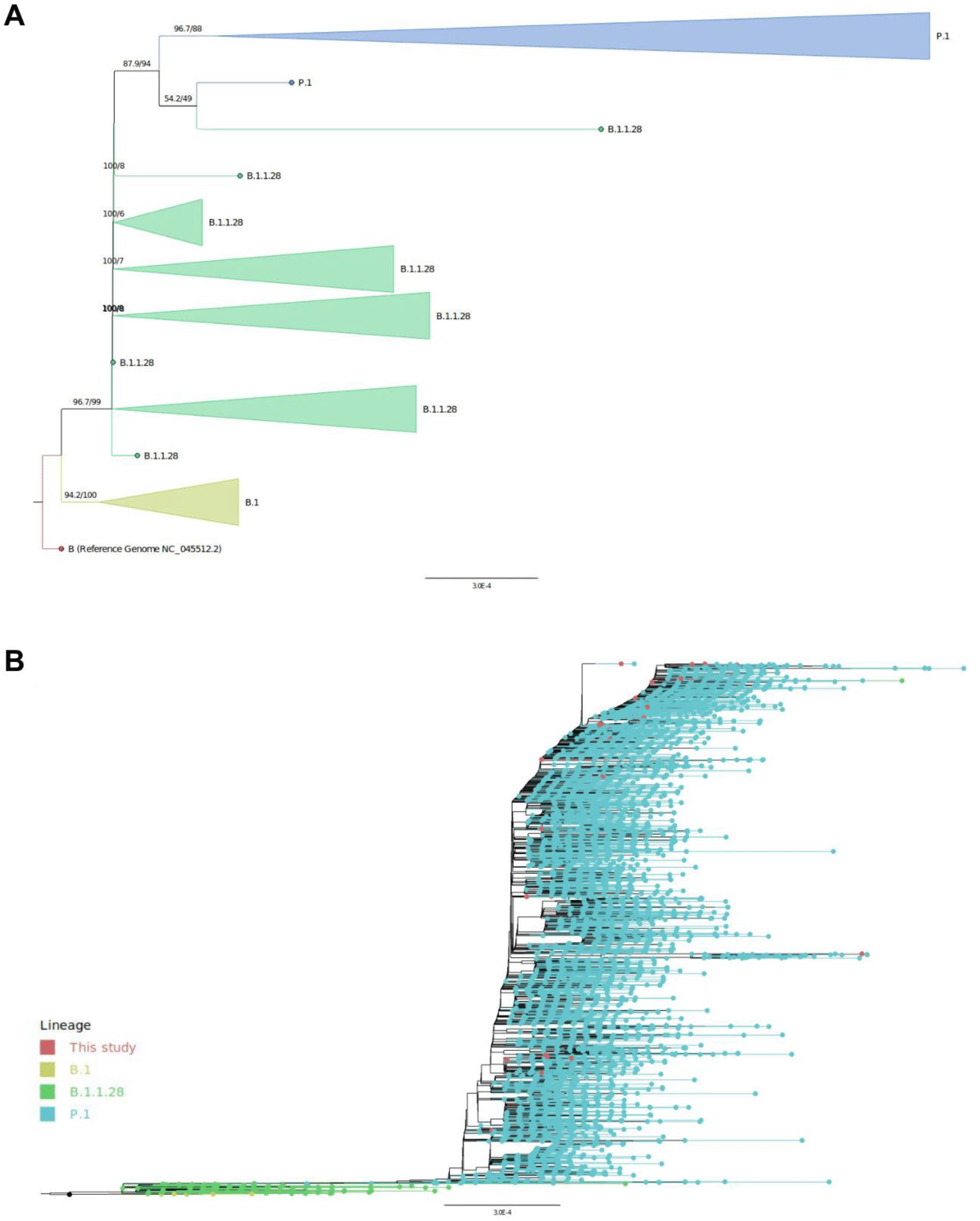
Global phylogenomic analyses. (A) (B) Global phylogeny of 4,997 subsampled genomes from P.1 and B.1.1.28 lineages. P.1 genomes are labeled by pink and B.1.1.28 by blue tips.

Despite only P.1 and B.1.1.28 sequences have been selected on GISAID (at September 12, 2021), six genomes from Turkey were posteriorly reassigned (at September 28, 2021) as B.1, corroborating the formation of a basal clade with 94.2 and 100% of branch support by the SH-aLRT and ultrafast bootstrap tests (Figure 6A). The large monophyletic group formed by B.1.1.28 and P.1 genomes is validated by 96.7 and 99% of statistical support in both tests, evidencing the ancestor-descendent relationship between B.1.1.28 (ancestor) and P.1 (descendant) lineages. As expected, the 44 sequenced genomes from the Amazonas were in the P.1 cluster (Figure 6B), despite their location in different subgroups along the tree.

Eighteen sequences (38966, 39448, 39454, 39432, 39443, 39438, 39428, 39436, 39452, 39440, 39465, 39437, 38967, 39444, 39456, 39433, 39449, and 39450) grouped with brazilian genomes. Eight of them (39432, 39443, 39436, 39452, 39431, 39433, 39449, and 39450) only with Amazonas genomes (Figure 7). Only four sequences (39431, 39433, 39449, and 39450) were found in statistically validated clades with the subsampled genomes. The genome 39431 (presenting the double deletion of the spike A243-L244 residues and the absence of the lineage-defining mutations ORF1a:S1188L, ORF1b:E1264D, S:T20N, R190S, S:H655Y, and S:T1027I) formed a fully supported clade (100% for both tests) with a genome from Amazonas. The monophyletic group with the sequences 39433, 39449, and 39450 (that share mutations such as ORF1a:T951I, A3333V, V3349I, and A3620V) and brazilian genomes from Amazonas was supported by 95.6 and 96% of branch support by the SH-aLRT and ultrafast bootstrap tests. The sequences 38969 and 39459 (sharing two ORF1b substitutions: I539V and E2221D) form their own clade with 93.4 and 96% of branch support by the SH-aLRT and ultrafast bootstrap, respectively.

**Figure 7.**
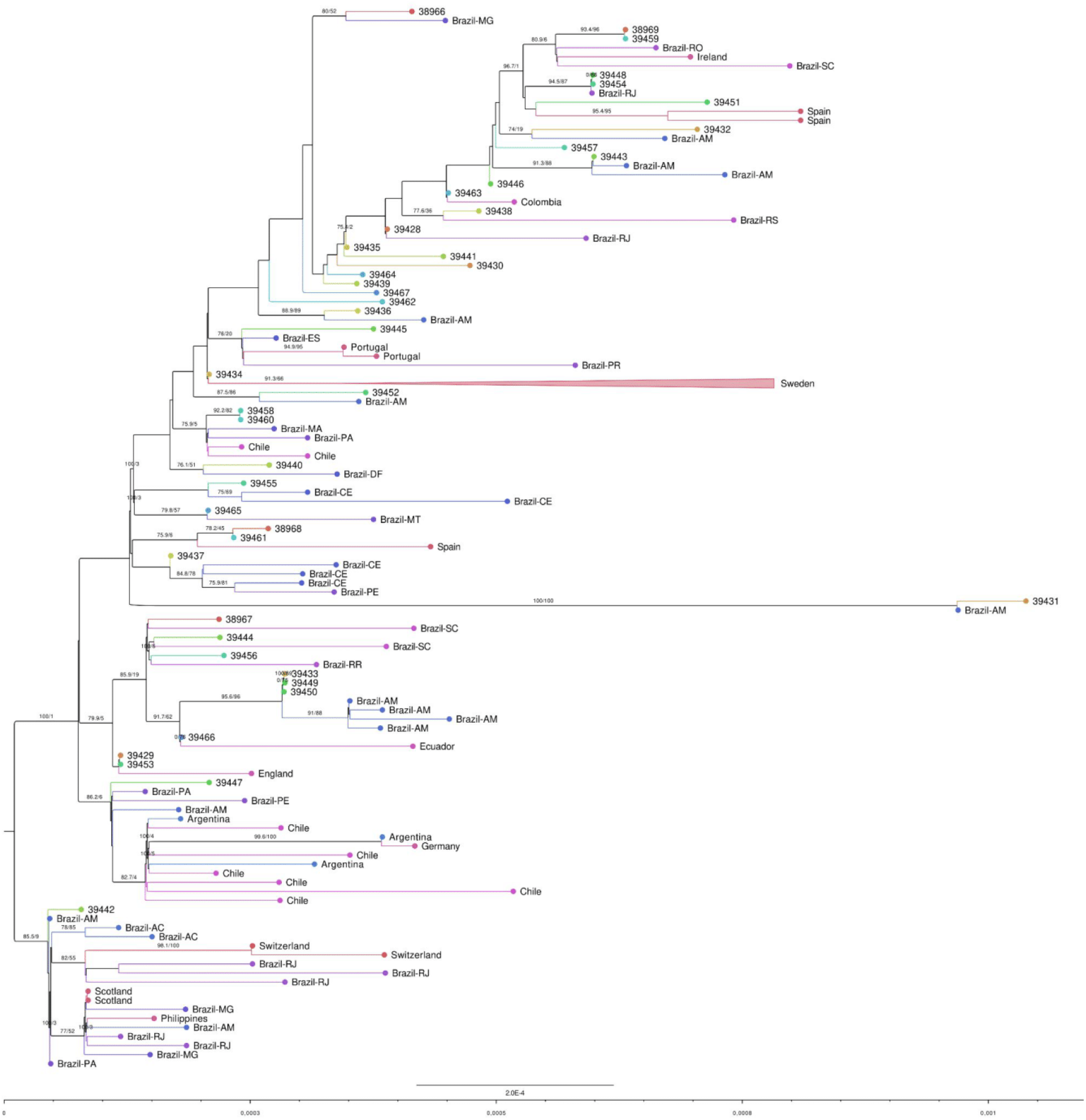
Representative subtree of the 44 sequenced samples of this study and their nearest grouped neighbors inside the P.1 clade. The branch support values are represented by the SH-aLRT and ultrafast bootstrap percentages ≥70.

Other clusters were also found despite the absence of statistical evidence as monophyletic groups by both tests (only the SH-aLRT test achieved values above the cutoff of 80%). The closest neighbours of the genomes 38969 and 39459 are Brazilian genomes from Rondonia and Santa Catarina, as well as a genome from Ireland (80.9 and 6% of branch support by the SH-aLRT and ultrafast bootstrap). A group between the genomes 39448, 39454 (that share a L1504F substitution in ORF1b) and a brazilian sequence from Rio de Janeiro is found with 94.5/87% of branch support by the SH-aLRT/ultrafast bootstrap tests. The sequence 39443 was located in a group (91.3/88% of branch support by the SH-aLRT/ultrafast bootstrap) with two Amazonas genomes. The genome 39438 clustered with a brazilian sequence from Rio Grande do Sul (77.6 and 56% of branch support by the SH-aLRT and ultrafast bootstrap). The sequence 39436 formed a group with a Brazilian genome from Amazonas with 88.9/89% of statistical support by the tests. The sequence 39452 was grouped with an Amazonas genome (87.5/86% of statistical support by the SH-aLRT/ultrafast bootstrap). The sequence 39447 (presenting the substitutions T2183I, H3076Y, G3676S, and F3677L in ORF1a) grouped with genomes from Brazil (Pará, Pernambuco, and Amazonas), Argentina, Chile and Germany with 86.2% of branch support by SH-aLRT test (6% by the ultrafast bootstrap). Inside the P.1 group, the sequence 39442 formed a separate cluster (85.5% of branch support by the aLRT test and 9% by the ultrafast bootstrap) in relation to the previously described sequences, clustering with genomes from Brazil (Amazonas, Acre, Rio de Janeiro, Minas Gerais and Pará), Switzerland, Scotland, and Philippines.

The genomes 39457, 39446, 39435, 39441, 39430, 39464, 39439, 39467, and 39462 grouped as external branches, unrelated to other subsampled genomes. The genomes 39451, 39432, 39463, 39445 (including the residue alterations ORF1a:F3677L and ORF1b:P1682H), 39434, 39458 and 39460 (carrying the mutations S:A688V and ORF3a:P104S), 39440 (presenting all the P.1 lineage-defining mutations added to the substitutions ORF1a:S2255F and F3677L), 39465, 38968 and 39461 (sharing the mutations ORF1a:T951I and ORF3a:P25L), 39437, 39466, 39429 and 39453 formed clusters with branch support values below 80% for both tests.

## DISCUSSION

P.1 (Gamma) lineage is arguably the most concerning VOC emerged thus far. Originally described in 6 January, Japan, from four returning travelers from Manaus, the lineage exhibited accelerated dissemination from December 2021. Thirty-five SNPs, including ten located at the spike, were initially characterized (38). We have demonstrated signs of diversification from the original P1. More specifically, we have found mutations that were either rare in this lineage (such as deletions at sites 188 and 189 coupled with substitutions at site 190 and deletions at 243 and 244) or previously not described at all (P209H), all located at the spike NTD. Despite this site has not been the target of much attention as RBD, it is in fact a hotspot for mutations, including “indels” and other missense substitutions, which may affect the activity of neutralizing antibodies and/or lead fitness recovery activity (39, 40). Deletions at amino acids 243 and 244 are common in B.1.351 (Beta), but are infrequent in worldwide P.1 (0.06%). These mutations occur at the exposed antigenic supersite from NTD and abolish the activity of some anti-NTD neutralizing antibodies (e.g 4A8) (41). Besides being rare to date, to our knowledge, our sequences are the first to describe the presence of del 243-244 in P.1 lineage. The P209H substitution replaces a proline, which has a rigid impact on peptide structure, for a positively charged, imidazole-containing, histidine. We project that this could have an effect on the resulting NTD structure as well. Since approximately 10 to 50% of neutralizing antibodies are directed towards NTD, it is noteworthy that we have found additional mutations at this site (41). We hypothesized that they could have had a critical role in allowing the emergence of a new lineage (P.1) despite a populational background of 75% previous seroprevalence. Similarly, S:T1066A is a low frequent mutation between the heptad repeat (HR1 and HR2) regions of the spike subunit 2, which was previously found in lineages such as B.1.258.10 and B.1.1.7, being firstly detected in the Brazilian territory in our P.1 genome. However, we were not able to demonstrate that any of these newly described mutations were being actively selected at this moment. On the other hand, canonical substitutions present in the P.1 lineage, such as E484K, K417T, N501Y, and H655Y were shown to be positively selected. E484K is important in immune evasion, particularly against class II anti-RBD antibodies, while K417T may be strongly related to immune evasion against class I anti-RBD antibodies (41). N501Y has minimal impact on antigenic evasion, but results in an important increase in binding affinity to hACE-2 (42, 4). Substitutions near the Furin Binding Site (FCS), such as H655Y, may augment FCS activity, further increasing infectivity or may be involved in dynamics S1 changes through increased hydrophobicity in the Gear-like Domain (43). The combination of the known phenotypic effects of these mutations with our positive selection results strongly support the idea that P.1 emergence is the result of intense selective pressure, which also explains the appearance of mutations at these sites in different lineages (convergent evolution).

Substitutions in genome regions outside the S gene may be critical for the infection severity, with potential impact on the modulation of innate immune response. For example, ORF9b, which is an alternative open reading frame within the nucleocapsid (N) gene, may have a important role in SARS-CoV-2-human interactions and has been demonstrated to significantly inhibit the IFN-I production as a result of targeting mitochondria. Moreover, the analyses of the sera of convalescent SARS-CoV or SARS-CoV-2 patients showed antibodies against ORF9b (44, 45). It is interesting to note that mutations at this site were found in the vast majority of our sequences.

In relation to the non-structural proteins from ORF1a, only NSP4 did not present evidence of sites under diversifying selection pressure. In NSP1, none of the sites involved in the translation inhibition (first and second C-terminal helix residues 153-160 / 166-178) (46) or host mRNA cleavage (R124 and K125) (47) were detected by the positive selection tests. However, site 156 was found to be under purifying selection pressure by FEL and SLAC methods. The NSP3 residue 1031, belonging to the canonical cysteine protease catalytic triad (related to the Papain-like viral protease site D286) on the active site (48) was also detected by FEL and SLAC, which indicates it is submitted to negative selection. Coronaviral proteases as Papain-like protease (PLpro) are essential for viral replication and modulate host immune response to viral infection (48), being known as an IFN antagonist (49). All positively selected sites identified by the FUBAR method are associated with NSP3 interdomain regions. Site 618, detected as positively selected by FEL and MEME, belongs to the Single-stranded poly(A) binding domain (SUD-M), and is involved in the recognition of Guanine-rich non-canonical nucleic acid structures (G-quadruplexes or G4), being essential for SARS-CoV replication (50); whereas site 727, also positively selected, is part of the Coronavirus polyprotein cleavage domain (Nsp3_PL2pro). At NSP5 C-terminal autocleavage sequence (S301 – Q306) (51), the residue T304 is under purifying selection. Both NSP7 and NSP8 have essential functions for the RNA synthesis machinery orchestrated by the NSP12 RNA-dependent RNA polymerase. In NSP7, evidence of positive selection was found for site 71, located at the interface between helix α1 and α4 in the NSP7– NSP8 complex (52). The NSP12 residues contact the NSP8 N-terminal region (77–126) where the primase activity lies. Of these, sites 91, 111, and 121 were identified to be under purifying selection. Specifically, site 91 from NSP8 helix α1 is involved in the NSP7-NSP8 complex formation (52, 53). NSP9 sites 6, 31, 103 detected under negative selection interact with peptides by hydrogen and Van der Waals bonds at the hydrophobic cavity in a region close to the dimer interface, with potential impact on the juxtapositioning of the monomers within the homodimer (54). In NSP10, site 83 in the zinc binding site located between the helices α2 and α3 (55) is under purifying selection, possibly due to its importance to stabilize the structural conformation.

All sites indicated as negatively selected in our study probably have an important role for the viral infection mechanisms, consequently being conserved among SARS-CoV-2 genomes. However, our results also indicated that positive selection is leading to an increase in amino acid variability in some viral sites, which results in an amplified potential to viral adaptation and evolutionary success. While a first and important mechanism for viral evolution is the action of positive selection, another possible mechanism for rapid acquisition of new substitutions would be the occurrence of “copy choice recombinations”, a well described mechanism of recombination for coronavirus, which displays discontinuous replication process, favoring frame-shifting. In fact, recombination between B.1.1.7 (Alpha) and other lineages have been characterized (56), but the evidence of the process of selecting new lineages is scarce. However, we were not able to find consistent evidence for copy choice recombination between full genomes from northern Brazil. This process could have explained the fast acquisition of a plethora of distinctive mutations and the apparent lack of recombination in our study strengthens the hypothesis of intense and continuous evolution under strong selective forces. Alternatively, additional substitutions found in our P.1 sequences may be simply the consequence of a matter of chance related, for instance, to “bottlenecks’’ or other evolutionary constraints acting as “drifters”.

In the phylogenomic analysis of north samples, it was possible to observe the formation of monophyletic groups for P.1 and P.2 genomes (E484K-presenting lineages derivative from B.1.1.28), as well as for the N.9 lineage (E484K presenting lineage derivative from B.1.1.3). Similarly, the phylogenetic analysis of spike sequences showed that P.1 genomes formed a clade. In the global phylogenomics, B.1.1.28 and P.1 were recognized as a clade, exposing their ancestor-descendent relationship. The clade formation for P.1 genomes in the phylogenomic analysis of north samples, as well as in the spike sequences, indicates the importance of the spike protein for SARS-CoV-2 evolution and lineage definition of these genomes. Sequences of non-structural proteins from ORF1a accumulated multiple substitutions without evidence of diversifying selection pressure in these sites. Considering the limitations related to a subsampled global phylogeny analysis, it is not possible to know if the absence of well-supported monophyletic groups is due to the missing branches and nodes representing the most related sequences and their ancestors or some technical limitation inherent to the application of the branch support statistical tests to the SARS-CoV-2 genomic sequences, since bootstrapping approaches require multiple sites supporting a clade to infer strong support value in near-perfect trees (57). In fact, SARS-CoV-2 genomes present a low number of informative sites, which may generate topology with low statistical support and ambiguous clustering of large data sets (58). Additionally, both SH-aLRT and ultrafast bootstrap methods rely on bootstrap resampling (57). Considering all aspects discussed above and the divergence among sequences, it is expected that the P.1 genomes (specially in the spike sequence analysis) cluster together and phylogenetic trees including multiple lineages present better statistical support values inter-lineages in comparison with intra-lineages.

In summary, the association of diversification of P.1 sequences, the known phenotypic consequences of some signature mutations, the confirmation of positive selection acting on some sites and the absence of evidence for recombinations, all suggest that the main driving force in the evolution of P.1 viruses was selective pressure.

## Data Availability

Consensus genomes generated in this study were deposited on the GISAID database under Accession IDs: EPI_ISL_3149974 to EPI_ISL_3150017.

## DATA AVAILABILITY STATEMENT

Full tables acknowledging the authors and corresponding labs submitting sequencing data used in this study can be found in Supplementary File 5. Consensus genomes generated in this study were deposited on the GISAID database under Accession IDs: EPI_ISL_3149974 to EPI_ISL_3150017.

## ETHICS STATEMENT

The molecular analyses and genome sequencing of the samples were performed and the study obtained a waiver of informed consent with approval of Comitê de Ética em Pesquisa em Seres Humanos da Universidade Federal de Ciências da Saúde de Porto Alegre (CEP - UFCSPA) under process number CAAE 35083220.2.0000.5345.

## AUTHOR CONTRIBUTIONS

CET, RAZ: conceived the study. CET, PAGF: designed and performed the formal bioinformatics analyses. CET, PAGF, RAZ: interpretation of genomic data. CET, LNR: resources, supervision and project administration related to the molecular and bioinformatics analyses. ARS, DNF: resources related to the clinical samples. AG, CGW, FAC, RAZ, ZR: funding acquisition. CET, PAGF, RAZ: wrote the original draft. All authors contributed to the final reviewed manuscript. All authors have read and approved the manuscript.

## ACKNOWLEDGMENTS

We thank the administrators of the GISAID database and research groups across the world for supporting the rapid and transparent sharing of genomic data during the COVID-19 pandemic.

## SUPPLEMENTARY FILES

**Supplementary File 1**. Sequencing depth and coverage plots for the forty-four sequenced genomes from this study.

**Supplementary File 2**. Collection dates (first occurrence) on GISAID database for SARS-CoV-2 mutations identified in the forty-four sequenced genomes from this study.

**Supplementary File 3**. Phylogenetic analysis of spike and non-structural proteins from ORF1a. (A) Spike, (B) NSP1, (C) NSP2, (D) NSP3, (E) NSP4, (F) NSP5, (G) NSP6, (H) NSP7, (I) NSP8, (J) NSP9, and (K) NSP10. Branch support values below 70 for both SH-aLRT and ultrafast bootstrap tests were not labeled in the trees.

**Supplementary File 4**. Sites under adaptive or purifying pressure according to the HyPhy tests FUBAR, FEL, MEME, and SLAC.

**Supplementary File 5**. Authors acknowledgement lists for GISAID genomes used in the genomic and phylogenomic analyses.

