## Supplementary File 2 for "Comparative genomics and characterization of SARS-CoV-2 P.1 (Gamma) Variant of Concern (VOC) from Amazonas, Brazil"

**Supplementary File 2. First occurrence (by collection date) registered on GISAID for SARS-CoV-2 mutations identified in the sequenced genomes from this study.**

| Mutation | Amazonas | Brazil | Brazil (outside Amazonas) | World (outside Brazil) |
|---|---|---|---|---|
| N:P80R | 2020-12-03 | 2020-09-11 / SP | 2020-09-11 / SP | 2020-12-17 / Peru |
| N:T135I | 2021-01-20 | 2021-01-10 / AP | 2021-01-10 / AP | 2021-01-02 / Peru |
| N:P199L | **2021-02-22** | **2021-02-22 / AM** | 2021-03-09 / SE | 2021-02-11 / USA |
| N:R203K | 2020-12-03 | 2020-09-11 / SP | 2020-09-11 / SP | 2020-12-17 / Peru |
| N:G204R | 2020-12-03 | 2020-09-11 / SP | 2020-09-11 / SP | 2020-12-17 / Peru |
| NSP1:T170I | **2021-02-25** | **2021-02-25 / AM** | -- | 2021-06-28 / Canada |
| NSP2:N9S | **2021-02-25** | **2021-02-25 / AM** | -- | -- |
| NSP2:L113F | 2021-02-11 | 2021-02-11 / AM | 2021-04-01 / PR | 2021-04-23 / Paraguay |
| NSP2:L400F | **2021-03-05** | 2021-02-26 / SE | 2021-02-26 / SE | 2021-04-10 / USA |
| NSP2:K456R | 2020-12-23 | 2020-12-23 / AM | 2021-01-21 / PR | 2021-01-18 / Guyana |
| NSP2:V469F | **2021-02-25** | 2021-02-22 / SP | 2021-02-22 / SP | 2021-03-27 / Colombia |
| NSP3:A41V | 2021-02-11 | 2021-02-11 / AM | 2021-02-23 / SP | 2021-04-11 / USA |
| NSP3:T133I | 2020-12-29 | 2020-12-29 / AM | 2021-01-03 / PA | 2021-01-18 / Italy |
| NSP3:T186P | 2020-12-23 | 2020-12-23 / AM | 2021-01-21 / PR | 2021-01-18 / Guyana |
| NSP3:A231V | **2021-03-01** | 2021-02-22 / RJ | 2021-02-22 / RJ | 2021-06-07 / USA |
| NSP3:S370L | 2020-12-03 | 2020-09-11 / SP | 2020-09-11 / SP | 2020-12-17 / Peru |
| NSP3:K977Q | 2020-12-03 | 2020-09-11 / SP | 2020-09-11 / SP | 2020-12-17 / Peru |
| NSP3:T1189I | 2020-12-23 | 2020-12-23 / AM | 2021-01-21 / PR | 2021-01-18 / Guyana |
| NSP3:T1365A | **2021-02-24** | **2021-02-24 / AM** | 2021-03-23 / SP | 2021-06-04 / Mexico |
| NSP3:S1437F | **2021-03-01** | **2021-03-01 / AM** | 2021-03-25 / RJ | 2021-06-21 / Mexico |
| NSP3:S1670F | **2021-03-08** | **2021-03-08 / AM** | 2021-03-17 / SP | 2021-03-17 / Peru |
| NSP4:V30A | **2021-02-23** | **2021-02-23 / AM** | -- | -- |
| NSP4:T83I | 2020-12-23 | 2020-12-23 / AM | 2021-01-07 / PB | 2021-01-06 / Colombia |
| NSP4:H313Y | **2021-02-26** | 2021-02-19 / PA | 2021-02-19 / PA | 2021-02-20 / USA |
| NSP4:S481L | **2021-02-23** | **2021-02-23 / AM** | 2021-03-31 / RJ | 2021-03-18 / Canada |
| NSP5:A70V | **2021-02-23** | **2021-02-23 / AM** | 2021-03-25 / RJ | 2021-04-19 / Argentina |
| NSP5:V86I | **2021-02-23** | 2021-02-22 / TO | 2021-02-22 / TO | -- |
| NSP5:P241L | 2021-01-07 | 2021-01-07 / AM | 2021-01-28 / AP | 2021-01-22 / Italy |
| NSP6:A46V | **2021-03-01** | 2021-02-19 / SC | 2021-02-19 / SC | 2021-05-09 / Colombia |
| NSP6:A51V | **2021-02-23** | **2021-02-23 / AM** | 2021-03-04 / GO | 2021-04-27 / USA |
| NSP6:S106del | 2020-12-03 | 2020-09-11 / SP | 2020-09-11 / SP | 2020-11-11 / USA |
| NSP6:G107S | 2020-12-21 | 2020-12-21 / AM | 2021-01-11 / RJ | 2021-01-04 / Italy |
| NSP6:G107del | 2020-12-03 | 2020-09-11 / SP | 2020-09-11 / SP | 2020-11-11 / USA |
| NSP6:F108L | 2020-12-03 | 2020-09-11 / SP | 2020-09-11 / SP | 2020-11-11 / USA |
| NSP6:F108del | 2020-12-21 | 2020-12-21 / AM | 2021-01-11 / RJ | 2020-12-17 / Peru |
| NSP6:V149A | 2020-12-23 | 2020-12-23 / AM | 2021-01-21 / PR | 2021-01-18 / Guyana |
| NSP8:E155G | **2021-02-23** | **2021-02-23 / AM** | 2021-03-05 / PR | 2021-04-22 / USA |
| NSP12:P323L | 2020-12-03 | 2020-09-11 / SP | 2020-09-11 / SP | 2020-11-11 / USA |
| NSP12:I548V | 2021-01-04 | 2021-01-04 / AM | 2021-01-27 / AC | 2021-03-04 / Ireland |
| NSP12:Q822H | **2021-03-01** | 2021-02-01 / RN | 2021-02-01 / RN | 2021-01-23 / Venezuela |
| NSP13:S74L | 2020-12-23 | 2020-12-23 / AM | 2021-01-21 / PR | 2021-01-18 / Guyana |
| NSP13:M274I | **2021-03-01** | **2021-03-01 / AM** | 2021-04-19 / SP | 2021-04-16 / Colombia |
| NSP13:E341D | 2020-12-03 | 2020-09-11 / SP | 2020-09-11 / SP | 2020-12-17 / Peru |
| NSP13:L581F | **2021-02-22** | 2021-02-11 / RJ | 2021-02-11 / RJ | 2021-03-26 / USA |
| NSP14:P158H | **2021-02-23** | **2021-02-23 / AM** | -- | 2021-06-28 / Chile |

| | | | | |
|---|---|---|---|---|
| NSP15:D39Y | **2021-02-23** | **2021-02-23 / AM** | 2021-03-23 / SP | 2021-03-08 / Switzerland |
| NSP15:E170D | **2021-02-22** | **2021-02-22 / AM** | -- | 2021-05-14 / USA |
| NSP16:K160R | **2021-02-22** | **2021-02-22 / AM** | 2021-02-22 / PA | 2021-01-04 / Italy |
| NS3:P25L | **2021-02-25** | 2021-01-27 / AL | 2021-01-27 / AL | 2021-03-30 / Germany |
| NS3:P104S | **2021-02-22** | **2021-02-22 / AM** | 2021-03-21 / GO | 2021-03-29 / USA |
| NS3:S117I | **2021-02-22** | 2021-01-02 / PR | 2021-01-02 / PR | -- |
| NS3:W131C | 2021-01-19 | 2021-01-19 / AM | 2021-03-08 / RS | 2021-04-09 / USA |
| NS3:E181V | **2021-02-25** | **2021-02-25 / AM** | -- | -- |
| NS3:S253P | 2020-12-03 | 2020-09-11 / SP | 2020-09-11 / SP | 2020-12-17 / Peru |
| NS7a:E22D | 2021-02-02 | 2021-02-02 / AM | 2021-02-23 / CE | 2021-04-14 / USA |
| NS7b:S31L | 2021-02-21 | 2021-02-21 / AM | 2021-05-06 / MS | 2021-03-31 / USA |
| NS8:E59D | 2021-02-21 | 2021-02-21 / AM | 2021-03-04 / CE | -- |
| NS8:E92K | 2020-12-03 | 2020-09-11 / SP | 2020-09-11 / SP | 2021-04-07 / USA |

Only complete collection dates were considered for this analysis. Collection dates related to genomes from this study are labeled in bold. AC: Acre, AL: Alagoas, AM: Amazonas, AP: Amapá, CE: Ceará, GO: Goiás, MS: Mato Grosso do Sul, PB: Paraíba, PR: Paraná, SC: Santa Catarina, SE: Sergipe, SP: São Paulo, RJ: Rio de Janeiro, RN: Rio Grande do Norte, RS: Rio Grande do Sul, TO: Tocantins. Date verification on September 24, 2021. *According to the GISAID database there are three P.1 genomes from the USA carrying these mutations with collection dates from april and may, 2020 (before its first occurrence in Brazil), however we do not find any publication confirming this USA origin of P.1 lineage.
