## Supplementary File 4 for "Comparative genomics and characterization of SARS-CoV-2 P.1 (Gamma) Variant of Concern (VOC) from Amazonas, Brazil"

### SPIKE

#### FEL

| Codon | alpha | beta | LRT | Selection detected? |
|---|---|---|---|---|
| 4 | 6.741 | 0.000 | 3.571 | Neg. p = 0.0588 |
| 5 | 0.000 | 7.119 | 2.805 | Pos. p = 0.0940 |
| 11 | 4.037 | 0.000 | 3.268 | Neg. p = 0.0706 |
| 32 | 6.982 | 0.000 | 5.919 | Neg. p = 0.0150 |
| 43 | 4.605 | 0.000 | 3.940 | Neg. p = 0.0471 |
| 66 | 6.694 | 0.000 | 3.139 | Neg. p = 0.0764 |
| 84 | 5.302 | 0.000 | 3.026 | Neg. p = 0.0820 |
| 130 | 6.097 | 0.000 | 4.921 | Neg. p = 0.0265 |
| 138 | 0.000 | 13.185 | 3.371 | Pos. p = 0.0664 |
| 168 | 6.694 | 0.000 | 3.578 | Neg. p = 0.0585 |
| 170 | 6.694 | 0.000 | 2.751 | Neg. p = 0.0972 |
| 198 | 13.486 | 0.000 | 5.844 | Neg. p = 0.0156 |
| 224 | 23.025 | 0.000 | 4.176 | Neg. p = 0.0410 |
| 287 | 6.741 | 0.000 | 2.919 | Neg. p = 0.0876 |
| 291 | 6.788 | 0.000 | 3.485 | Neg. p = 0.0619 |
| 293 | 8.465 | 0.000 | 5.612 | Neg. p = 0.0178 |
| 295 | 4.369 | 0.000 | 2.836 | Neg. p = 0.0922 |
| 296 | 8.080 | 0.000 | 7.085 | Neg. p = 0.0078 |
| 300 | 17.587 | 0.000 | 6.824 | Neg. p = 0.0090 |
| 306 | 4.645 | 0.000 | 3.943 | Neg. p = 0.0471 |
| 313 | 6.741 | 0.000 | 2.753 | Neg. p = 0.0971 |
| 351 | 6.694 | 0.000 | 2.753 | Neg. p = 0.0971 |
| 364 | 6.741 | 0.000 | 2.926 | Neg. p = 0.0872 |
| 369 | 6.694 | 0.000 | 2.753 | Neg. p = 0.0971 |
| 375 | 4.025 | 0.000 | 3.424 | Neg. p = 0.0643 |
| 392 | 6.694 | 0.000 | 3.573 | Neg. p = 0.0587 |
| 395 | 4.028 | 0.000 | 3.273 | Neg. p = 0.0704 |
| 413 | 10.812 | 0.000 | 7.648 | Neg. p = 0.0057 |
| 417 | 0.000 | 7.902 | 3.469 | Pos. p = 0.0625 |
| 421 | 6.694 | 0.000 | 2.753 | Neg. p = 0.0971 |
| 432 | 6.788 | 0.000 | 5.515 | Neg. p = 0.0189 |
| 441 | 8.756 | 0.000 | 5.617 | Neg. p = 0.0178 |
| 469 | 10.392 | 0.000 | 3.947 | Neg. p = 0.0470 |
| 491 | 4.333 | 0.000 | 2.835 | Neg. p = 0.0922 |
| 507 | 10.465 | 0.000 | 4.277 | Neg. p = 0.0386 |
| 518 | 5.320 | 0.000 | 3.024 | Neg. p = 0.0821 |
| 562 | 4.605 | 0.000 | 3.939 | Neg. p = 0.0472 |
| 578 | 6.694 | 0.000 | 2.919 | Neg. p = 0.0876 |
| 618 | 10.328 | 0.000 | 3.645 | Neg. p = 0.0562 |
| 673 | 6.694 | 0.000 | 3.077 | Neg. p = 0.0794 |
| 680 | 4.333 | 0.000 | 2.736 | Neg. p = 0.0981 |
| 681 | 0.000 | 7.119 | 2.777 | Pos. p = 0.0956 |

#### SLAC

| Codon | S | N | dS | dN | Selection detected? |
|---|---|---|---|---|---|
| 32 | 3.000 | 0.000 | 3.575 | 0.000 | Neg. p = 0.022 |
| 43 | 2.000 | 0.000 | 2.382 | 0.000 | Neg. p = 0.078 |
| 130 | 3.000 | 0.000 | 3.000 | 0.000 | Neg. p = 0.037 |
| 146 | 3.000 | 1.000 | 3.576 | 0.463 | Neg. p = 0.069 |
| 198 | 2.000 | 0.000 | 3.005 | 0.000 | Neg. p = 0.049 |
| 296 | 4.000 | 0.000 | 4.000 | 0.000 | Neg. p = 0.012 |
| 300 | 2.000 | 0.000 | 5.077 | 0.000 | Neg. p = 0.019 |
| 306 | 2.000 | 0.000 | 2.382 | 0.000 | Neg. p = 0.078 |
| 354 | 3.000 | 1.000 | 3.576 | 0.463 | Neg. p = 0.069 |
| 413 | 4.000 | 0.000 | 4.000 | 0.000 | Neg. p = 0.012 |
| 432 | 3.000 | 0.000 | 3.573 | 0.000 | Neg. p = 0.024 |
| 543 | 3.000 | 1.000 | 3.573 | 0.463 | Neg. p = 0.069 |
| 562 | 2.000 | 0.000 | 2.381 | 0.000 | Neg. p = 0.078 |
| 682 | 3.000 | 0.000 | 2.707 | 0.000 | Neg. p = 0.050 |
| 692 | 3.000 | 1.000 | 3.171 | 0.487 | Neg. p = 0.096 |
| 707 | 5.000 | 0.000 | 5.958 | 0.000 | Neg. p = 0.002 |
| 821 | 5.000 | 0.000 | 2.722 | 0.000 | Neg. p = 0.086 |
| 856 | 2.000 | 0.000 | 2.382 | 0.000 | Neg. p = 0.078 |
| 936 | 2.000 | 0.000 | 2.382 | 0.000 | Neg. p = 0.078 |
| 960 | 2.000 | 0.000 | 2.384 | 0.000 | Neg. p = 0.078 |
| 1018 | 2.000 | 0.000 | 2.109 | 0.000 | Neg. p = 0.100 |
| 1101 | 2.000 | 0.000 | 2.382 | 0.000 | Neg. p = 0.078 |
| 1178 | 6.000 | 0.000 | 7.161 | 0.000 | Neg. p = 0.000 |
| 1215 | 3.000 | 0.000 | 3.575 | 0.000 | Neg. p = 0.026 |

Significant sites at p <= 0.1

#### MEME

| Codon | alpha | beta+ | p+ | LRT | Episodic selection detected? | # branches | Most common codon substitutions at this site |
|---|---|---|---|---|---|---|---|
| 138 | 0.000 | 13.028 | 1.000 | 3.371 | Yes, p = 0.0879 | 6 | [4]TAT>GAT \| [2]GAT>TAT |
| 417 | 0.000 | 7.889 | 0.990 | 3.471 | Yes, p = 0.0834 | 4 | [2]ACG>AAG \| [1]AAG>ACG,ACG>ATG |

Significant sites at p <= 0.1

| | | | | |
|---|---|---|---|---|
| 682 | 6.982 | 0.000 | 5.473 | Neg. p = 0.0193 |
| 707 | 11.629 | 0.000 | 7.037 | Neg. p = 0.0080 |
| 718 | 6.694 | 0.000 | 3.578 | Neg. p = 0.0585 |
| 743 | 6.694 | 0.000 | 3.477 | Neg. p = 0.0622 |
| 756 | 6.694 | 0.000 | 2.751 | Neg. p = 0.0972 |
| 793 | 10.392 | 0.000 | 4.265 | Neg. p = 0.0389 |
| 803 | 10.262 | 0.000 | 3.935 | Neg. p = 0.0473 |
| 817 | 6.694 | 0.000 | 3.576 | Neg. p = 0.0586 |
| 818 | 5.935 | 0.000 | 2.907 | Neg. p = 0.0882 |
| 819 | 23.146 | 0.000 | 4.177 | Neg. p = 0.0410 |
| 821 | 13.347 | 0.000 | 7.579 | Neg. p = 0.0059 |
| 881 | 10.392 | 0.000 | 3.653 | Neg. p = 0.0560 |
| 893 | 10.392 | 0.000 | 4.068 | Neg. p = 0.0437 |
| 897 | 20.554 | 0.000 | 8.491 | Neg. p = 0.0036 |
| 934 | 5.935 | 0.000 | 2.907 | Neg. p = 0.0882 |
| 936 | 4.605 | 0.000 | 2.978 | Neg. p = 0.0844 |
| 948 | 4.273 | 0.000 | 2.804 | Neg. p = 0.0940 |
| 972 | 10.392 | 0.000 | 4.080 | Neg. p = 0.0434 |
| 979 | 6.694 | 0.000 | 2.916 | Neg. p = 0.0877 |
| 988 | 8.684 | 0.000 | 3.419 | Neg. p = 0.0645 |
| 1018 | 4.154 | 0.000 | 2.859 | Neg. p = 0.0908 |
| 1030 | 10.392 | 0.000 | 3.947 | Neg. p = 0.0470 |
| 1034 | 4.333 | 0.000 | 2.807 | Neg. p = 0.0939 |
| 1047 | 6.694 | 0.000 | 2.753 | Neg. p = 0.0971 |
| 1055 | 20.205 | 0.000 | 7.888 | Neg. p = 0.0050 |
| 1067 | 6.694 | 0.000 | 2.753 | Neg. p = 0.0971 |
| 1070 | 10.262 | 0.000 | 4.068 | Neg. p = 0.0437 |
| 1087 | 10.262 | 0.000 | 4.068 | Neg. p = 0.0437 |
| 1101 | 4.617 | 0.000 | 3.293 | Neg. p = 0.0696 |
| 1122 | 5.278 | 0.000 | 3.919 | Neg. p = 0.0477 |
| 1129 | 10.262 | 0.000 | 4.032 | Neg. p = 0.0446 |
| 1136 | 10.262 | 0.000 | 3.641 | Neg. p = 0.0564 |
| 1159 | 6.694 | 0.000 | 3.144 | Neg. p = 0.0762 |
| 1175 | 10.324 | 0.000 | 3.931 | Neg. p = 0.0474 |
| 1178 | 13.995 | 0.000 | 6.911 | Neg. p = 0.0086 |
| 1196 | 4.333 | 0.000 | 2.736 | Neg. p = 0.0981 |
| 1213 | 10.245 | 0.000 | 4.261 | Neg. p = 0.0390 |
| 1215 | 6.788 | 0.000 | 4.225 | Neg. p = 0.0398 |
| 1239 | 6.694 | 0.000 | 3.077 | Neg. p = 0.0794 |
| 1264 | 0.000 | 4.771 | 2.827 | Pos. p = 0.0927 |

Significant sites at p  <= 0.1

### NSP1

| FEL | | | | |
|---|---|---|---|---|
| Codon | alpha | beta | LRT | Selection detected? |
| 5 | 0 | 6.771 | 3.021 | Pos. p = 0.0822 |
| 36 | 11.333 | 0 | 2.814 | Neg. p = 0.0934 |
| 42 | 8.352 | 0 | 2.914 | Neg. p = 0.0878 |
| 74 | 6.097 | 0 | 2.999 | Neg. p = 0.0833 |
| 156 | 31.027 | 3.885 | 5.935 | Neg. p = 0.0148 |
| Significant sites at p <= 0.1 | | | | |

| SLAC | | | | | |
|---|---|---|---|---|---|
| Codon | S | N | dS | dN | Selection detected? |
| 156 | 8.000 | 1.000 | 9.188 | 0.470 | Neg. p = 0.000 |
| Significant sites at p <= 0.1 | | | | | |

| MEME | | | | | | | |
|---|---|---|---|---|---|---|---|
| Codon | alpha | beta+ | p+ | LRT | Episodic selection detected? | # branches | Most common codon substitutions at this site |
| No sites found at p <= 0.1 | | | | | | | |

### NSP2

**FEL**

| Codon | alpha | beta | LRT | Selection detected? |
|---|---|---|---|---|
| 86 | 4.698 | 0 | 3.263 | Neg. p = 0.0709 |
| 136 | 0 | 5.146 | 2.988 | Pos. p = 0.0839 |
| 165 | 0 | 6.407 | 3.718 | Pos. p = 0.0538 |
| 205 | 4.223 | 0 | 2.899 | Neg. p = 0.0886 |
| 207 | 7.056 | 0 | 3.392 | Neg. p = 0.0655 |
| 256 | 10.891 | 0 | 3.749 | Neg. p = 0.0528 |
| 339 | 0 | 9.624 | 3.236 | Pos. p = 0.0720 |
| 347 | 18.887 | 0 | 3.710 | Neg. p = 0.0541 |
| 369 | 13.010 | 0 | 9.759 | Neg. p = 0.0018 |
| 414 | 10.812 | 0 | 3.644 | Neg. p = 0.0563 |
| 446 | 4.210 | 0 | 3.602 | Neg. p = 0.0577 |
| 447 | 0 | 4.804 | 3.045 | Pos. p = 0.0810 |
| 473 | 11.034 | 0 | 3.856 | Neg. p = 0.0496 |
| 483 | 18.887 | 0 | 3.079 | Neg. p = 0.0793 |
| 569 | 10.922 | 0 | 3.757 | Neg. p = 0.0526 |
| 597 | 10.934 | 0 | 4.300 | Neg. p = 0.0381 |
| 636 | 18.887 | 0 | 3.705 | Neg. p = 0.0543 |

Significant sites at p <= 0.1

**SLAC**

| Codon | S | N | dS | dN | Selection detected? |
|---|---|---|---|---|---|
| 8 | 2.000 | 0 | 2.253 | 0 | Neg. p = 0.088 |
| 40 | 2.000 | 0 | 2.253 | 0 | Neg. p = 0.088 |
| 86 | 2.000 | 0 | 2.254 | 0 | Neg. p = 0.088 |
| 207 | 3.000 | 0 | 3.381 | 0 | Neg. p = 0.029 |
| 369 | 6.000 | 0 | 6.000 | 0 | Neg. p = 0.001 |
| 414 | 2.000 | 0 | 2.359 | 0 | Neg. p = 0.080 |
| 606 | 2.000 | 0 | 2.257 | 0 | Neg. p = 0.087 |

Significant sites at p <= 0.1

**MEME**

| Codon | alpha | beta+ | p+ | LRT | Episodic selection detected? | # branches | Most common codon substitutions at this site |
|---|---|---|---|---|---|---|---|
| 165 | 0 | 6.341 | 0.990 | 3.718 | Yes, p = 0.0733 | 2 | [1]GGC>AGC,GGC>GAC |
| 339 | 0 | 10.038 | 0.960 | 3.236 | Yes, p = 0.0943 | 3 | [2]GGT>AGT \| [1]GGT>GAT |
| 447 | 0 | 1407.863 | 0.005 | 11.733 | Yes, p = 0.0012 | 1 | [1]GTC>TCC |
| 458 | 0 | 790.947 | 0.006 | 3.376 | Yes, p = 0.0876 | 1 | [1]GGT>GCT |

Significant sites at p <= 0.1

### NSP3

#### FEL

| Codon | alpha | beta | LRT | Selection detected? |
|---|---|---|---|---|
| 5 | 10.392 | 0 | 5.360 | Neg. p = 0.0206 |
| 21 | 7.464 | 0 | 4.384 | Neg. p = 0.0363 |
| 27 | 5.357 | 0 | 3.056 | Neg. p = 0.0804 |
| 48 | 11.754 | 0 | 3.918 | Neg. p = 0.0478 |
| 100 | 11.754 | 0 | 4.133 | Neg. p = 0.0420 |
| 106 | 7.212 | 0 | 3.417 | Neg. p = 0.0645 |
| 146 | 5.473 | 0 | 4.319 | Neg. p = 0.0377 |
| 231 | 0 | 8.571 | 2.794 | Pos. p = 0.0946 |
| 266 | 34.200 | 0 | 4.801 | Neg. p = 0.0284 |
| 268 | 31.517 | 0 | 6.458 | Neg. p = 0.0110 |
| 300 | 7.401 | 0 | 5.447 | Neg. p = 0.0196 |
| 329 | 11.754 | 0 | 4.490 | Neg. p = 0.0341 |
| 348 | 11.754 | 0 | 3.970 | Neg. p = 0.0463 |
| 368 | 5.357 | 0 | 3.057 | Neg. p = 0.0804 |
| 422 | 11.754 | 0 | 3.929 | Neg. p = 0.0474 |
| 446 | 7.238 | 0 | 3.027 | Neg. p = 0.0819 |
| 472 | 5.428 | 0 | 2.712 | Neg. p = 0.0996 |
| 497 | 15.762 | 0 | 3.282 | Neg. p = 0.0700 |
| 504 | 11.754 | 0 | 3.916 | Neg. p = 0.0478 |
| 512 | 7.464 | 0 | 5.206 | Neg. p = 0.0225 |
| 524 | 11.754 | 0 | 4.122 | Neg. p = 0.0423 |
| 527 | 11.148 | 0 | 6.575 | Neg. p = 0.0103 |
| 528 | 5.357 | 0 | 3.058 | Neg. p = 0.0804 |
| 538 | 7.401 | 0 | 3.792 | Neg. p = 0.0515 |
| 564 | 11.754 | 0 | 4.110 | Neg. p = 0.0426 |
| 597 | 7.466 | 0 | 4.382 | Neg. p = 0.0363 |
| 613 | 11.754 | 0 | 3.970 | Neg. p = 0.0463 |
| 618 | 0 | 7.272 | 4.236 | Pos. p = 0.0396 |
| 620 | 5.357 | 0 | 3.056 | Neg. p = 0.0804 |
| 631 | 11.754 | 0 | 4.490 | Neg. p = 0.0341 |
| 665 | 5.402 | 0 | 2.712 | Neg. p = 0.0996 |
| 680 | 15.762 | 0 | 3.258 | Neg. p = 0.0711 |
| 685 | 15.762 | 0 | 3.259 | Neg. p = 0.0710 |
| 692 | 3.663 | 0 | 3.040 | Neg. p = 0.0813 |
| 707 | 43.945 | 1.881 | 3.704 | Neg. p = 0.0543 |
| 727 | 0 | 4.230 | 2.955 | Pos. p = 0.0856 |
| 827 | 6.097 | 0 | 2.907 | Neg. p = 0.0882 |
| 850 | 15.762 | 0 | 3.282 | Neg. p = 0.0701 |
| 885 | 6.097 | 0 | 2.907 | Neg. p = 0.0882 |
| 903 | 11.754 | 0 | 3.918 | Neg. p = 0.0478 |
| 955 | 11.754 | 0 | 3.905 | Neg. p = 0.0482 |
| 970 | 11.257 | 1.929 | 2.792 | Neg. p = 0.0947 |
| 973 | 15.762 | 0 | 3.277 | Neg. p = 0.0702 |

#### SLAC

| Codon | S | N | dS | dN | Selection detected? |
|---|---|---|---|---|---|
| 146 | 3.000 | 0.000 | 3.000 | 0.000 | Neg. p = 0.037 |
| 262 | 2.000 | 0.000 | 2.150 | 0.000 | Neg. p = 0.096 |
| 268 | 2.000 | 0.000 | 2.528 | 0.000 | Neg. p = 0.088 |
| 300 | 4.000 | 0.000 | 4.000 | 0.000 | Neg. p = 0.012 |
| 356 | 3.000 | 1.000 | 3.225 | 0.500 | Neg. p = 0.097 |
| 455 | 2.000 | 0.000 | 2.152 | 0.000 | Neg. p = 0.096 |
| 527 | 3.000 | 0.000 | 3.000 | 0.000 | Neg. p = 0.037 |
| 621 | 2.000 | 0.000 | 2.150 | 0.000 | Neg. p = 0.096 |
| 736 | 2.000 | 0.000 | 2.152 | 0.000 | Neg. p = 0.096 |
| 760 | 2.000 | 0.000 | 2.150 | 0.000 | Neg. p = 0.096 |
| 855 | 3.000 | 0.000 | 3.226 | 0.000 | Neg. p = 0.030 |
| 931 | 2.000 | 0.000 | 2.150 | 0.000 | Neg. p = 0.096 |
| 953 | 0.000 | 2.000 | 0.000 | 0.667 | Neg. p = 1.000 |
| 1031 | 3.000 | 0.000 | 3.226 | 0.000 | Neg. p = 0.030 |
| 1038 | 3.000 | 0.000 | 3.000 | 0.000 | Neg. p = 0.037 |
| 1089 | 2.000 | 0.000 | 2.150 | 0.000 | Neg. p = 0.096 |
| 1099 | 2.000 | 0.000 | 2.150 | 0.000 | Neg. p = 0.096 |
| 1104 | 2.000 | 0.000 | 2.150 | 0.000 | Neg. p = 0.096 |
| 1107 | 3.000 | 1.000 | 3.230 | 0.483 | Neg. p = 0.091 |
| 1117 | 4.000 | 0.000 | 4.055 | 0.000 | Neg. p = 0.012 |
| 1228 | 0.000 | 7.000 | 0.000 | 3.519 | Pos. p = 0.056 |
| 1238 | 3.000 | 0.000 | 3.000 | 0.000 | Neg. p = 0.037 |
| 1298 | 3.000 | 0.000 | 3.000 | 0.000 | Neg. p = 0.037 |
| 1329 | 3.000 | 0.000 | 3.226 | 0.000 | Neg. p = 0.030 |
| 1354 | 3.000 | 0.000 | 3.226 | 0.000 | Neg. p = 0.030 |
| 1516 | 2.000 | 0.000 | 2.150 | 0.000 | Neg. p = 0.096 |
| 1567 | 3.000 | 0.000 | 3.042 | 0.000 | Neg. p = 0.036 |
| 1603 | 6.000 | 0.000 | 6.000 | 0.000 | Neg. p = 0.001 |
| 1705 | 2.000 | 0.000 | 2.150 | 0.000 | Neg. p = 0.096 |
| 1713 | 2.000 | 0.000 | 2.150 | 0.000 | Neg. p = 0.096 |
| 1739 | 3.000 | 0.000 | 3.000 | 0.000 | Neg. p = 0.037 |
| 1820 | 4.000 | 0.000 | 3.788 | 0.000 | Neg. p = 0.015 |
| 1858 | 3.000 | 0.000 | 3.000 | 0.000 | Neg. p = 0.037 |
| 1860 | 2.000 | 0.000 | 2.150 | 0.000 | Neg. p = 0.096 |
| 1875 | 2.000 | 0.000 | 2.150 | 0.000 | Neg. p = 0.096 |
| 1891 | 2.000 | 0.000 | 2.406 | 0.000 | Neg. p = 0.077 |

Significant sites at p <= 0.1

#### MEME

| Codon | alpha | beta+ | p+ | LRT | Episodic selection detected? | # branches | Most common codon substitutions at this site |
|---|---|---|---|---|---|---|---|
| 618 | 0.000 | 7.275 | 0.990 | 4.237 | Yes, p = 0.0560 | 2 | [2]AAC>AGC |

Significant sites at p <= 0.1

| | | | | |
|---|---|---|---|---|
| 1024 | 15.762 | 0 | 3.283 | Neg. p = 0.0700 |
| 1031 | 5.829 | 0 | 3.005 | Neg. p = 0.0830 |
| 1038 | 5.647 | 0 | 4.656 | Neg. p = 0.0310 |
| 1060 | 15.762 | 0 | 3.283 | Neg. p = 0.0700 |
| 1065 | 15.762 | 0 | 3.281 | Neg. p = 0.0701 |
| 1089 | 3.890 | 0 | 3.115 | Neg. p = 0.0776 |
| 1117 | 7.401 | 0 | 5.049 | Neg. p = 0.0246 |
| 1118 | 15.762 | 0 | 3.283 | Neg. p = 0.0700 |
| 1159 | 3.629 | 0 | 3.341 | Neg. p = 0.0676 |
| 1207 | 7.215 | 0 | 3.422 | Neg. p = 0.0643 |
| 1211 | 0 | 11.198 | 2.737 | Pos. p = 0.0981 |
| 1222 | 3.646 | 0 | 2.887 | Neg. p = 0.0893 |
| 1235 | 15.762 | 0 | 3.259 | Neg. p = 0.0710 |
| 1238 | 5.580 | 0 | 4.055 | Neg. p = 0.0440 |
| 1249 | 15.791 | 0 | 3.257 | Neg. p = 0.0711 |
| 1298 | 14.732 | 0 | 7.731 | Neg. p = 0.0054 |
| 1299 | 13.165 | 0 | 5.889 | Neg. p = 0.0152 |
| 1303 | 0 | 3.871 | 2.899 | Pos. p = 0.0886 |
| 1354 | 5.795 | 0 | 4.679 | Neg. p = 0.0305 |
| 1368 | 23.920 | 0 | 7.858 | Neg. p = 0.0051 |
| 1440 | 0 | 4.296 | 3.118 | Pos. p = 0.0774 |
| 1470 | 7.277 | 0 | 3.207 | Neg. p = 0.0733 |
| 1487 | 15.762 | 0 | 3.259 | Neg. p = 0.0710 |
| 1491 | 6.694 | 0 | 2.976 | Neg. p = 0.0845 |
| 1494 | 11.754 | 0 | 4.137 | Neg. p = 0.0419 |
| 1497 | 15.762 | 0 | 3.279 | Neg. p = 0.0702 |
| 1516 | 3.890 | 0 | 3.118 | Neg. p = 0.0774 |
| 1519 | 7.277 | 0 | 3.419 | Neg. p = 0.0644 |
| 1544 | 5.357 | 0 | 2.940 | Neg. p = 0.0864 |
| 1558 | 7.464 | 0 | 5.201 | Neg. p = 0.0226 |
| 1567 | 5.529 | 0 | 3.791 | Neg. p = 0.0515 |
| 1569 | 7.186 | 0 | 3.414 | Neg. p = 0.0646 |
| 1572 | 7.215 | 0 | 3.422 | Neg. p = 0.0643 |
| 1603 | 11.034 | 0 | 8.168 | Neg. p = 0.0043 |
| 1634 | 7.186 | 0 | 3.205 | Neg. p = 0.0734 |
| 1665 | 5.357 | 0 | 3.156 | Neg. p = 0.0757 |
| 1728 | 15.762 | 0 | 3.282 | Neg. p = 0.0700 |
| 1739 | 11.148 | 0 | 6.900 | Neg. p = 0.0086 |
| 1799 | 15.762 | 0 | 3.259 | Neg. p = 0.0710 |
| 1807 | 3.649 | 0 | 3.039 | Neg. p = 0.0813 |
| 1820 | 13.654 | 0 | 8.737 | Neg. p = 0.0031 |
| 1858 | 5.473 | 0 | 3.986 | Neg. p = 0.0459 |
| 1891 | 13.679 | 0 | 5.999 | Neg. p = 0.0143 |
| 1914 | 5.357 | 0 | 2.826 | Neg. p = 0.0928 |
| 1915 | 5.357 | 0 | 2.826 | Neg. p = 0.0928 |
| Significant sites at p <= 0.1 | | | | |

### NSP4

#### FEL

| Codon | alpha | beta | LRT | Selection detected? |
|---|---|---|---|---|
| 69 | 7.402 | 0.000 | 2.887 | Neg. p = 0.0893 |
| 76 | 8.670 | 0.049 | 3.715 | Neg. p = 0.0539 |
| 121 | 5.487 | 0.000 | 2.847 | Neg. p = 0.0916 |
| 145 | 8.670 | 0.000 | 2.899 | Neg. p = 0.0886 |
| 154 | 7.402 | 0.000 | 2.887 | Neg. p = 0.0893 |
| 213 | 14.541 | 0.000 | 3.671 | Neg. p = 0.0554 |
| 214 | 14.409 | 0.000 | 3.893 | Neg. p = 0.0485 |
| 228 | 18.887 | 0.000 | 3.768 | Neg. p = 0.0522 |
| 229 | 14.541 | 0.000 | 3.834 | Neg. p = 0.0502 |
| 252 | 14.523 | 0.000 | 4.473 | Neg. p = 0.0344 |
| 258 | 14.541 | 0.000 | 3.671 | Neg. p = 0.0554 |
| 298 | 5.170 | 0.000 | 3.026 | Neg. p = 0.0820 |
| 363 | 8.251 | 0.000 | 2.718 | Neg. p = 0.0992 |
| 390 | 5.484 | 0.000 | 2.855 | Neg. p = 0.0911 |
| 413 | 21.710 | 0.000 | 3.685 | Neg. p = 0.0549 |
| 456 | 14.702 | 0.000 | 3.253 | Neg. p = 0.0713 |
| 475 | 5.170 | 0.000 | 3.315 | Neg. p = 0.0686 |
| 489 | 14.418 | 0.000 | 4.465 | Neg. p = 0.0346 |

Significant sites at p  <= 0.1

#### SLAC

| Codon | S | N | dS | dN | Selection detected? |
|---|---|---|---|---|---|
| 76 | 3.000 | 0.000 | 3.227 | 0.000 | Neg. p = 0.030 |
| 121 | 2.000 | 0.000 | 2.150 | 0.000 | Neg. p = 0.096 |
| 390 | 2.000 | 0.000 | 2.151 | 0.000 | Neg. p = 0.096 |

Significant sites at p  <= 0.1

#### MEME

| Codon | alpha | beta+ | p+ | LRT | Episodic selection detected? | # branches | Most common codon substitutions at this site |
|---|---|---|---|---|---|---|---|

No sites found at p <= 0.1

### NSP5

#### FEL

| Codon | alpha | beta | LRT | Selection detected? |
|---|---|---|---|---|
| 3 | 6.426 | 0.000 | 3.204 | Neg. p = 0.0735 |
| 18 | 11.754 | 0.000 | 3.784 | Neg. p = 0.0517 |
| 24 | 11.754 | 0.000 | 3.972 | Neg. p = 0.0463 |
| 55 | 17.723 | 0.000 | 3.553 | Neg. p = 0.0594 |
| 105 | 3.464 | 0.000 | 2.741 | Neg. p = 0.0978 |
| 132 | 3.464 | 0.000 | 3.291 | Neg. p = 0.0697 |
| 134 | 3.649 | 0.000 | 2.982 | Neg. p = 0.0842 |
| 178 | 17.723 | 0.000 | 3.553 | Neg. p = 0.0594 |
| 194 | 5.233 | 0.000 | 2.743 | Neg. p = 0.0977 |
| 226 | 5.293 | 0.000 | 2.712 | Neg. p = 0.0996 |
| 269 | 17.723 | 0.000 | 3.665 | Neg. p = 0.0556 |
| 272 | 5.491 | 0.000 | 2.918 | Neg. p = 0.0876 |
| 285 | 5.083 | 0.000 | 2.718 | Neg. p = 0.0992 |
| 293 | 5.205 | 0.000 | 3.123 | Neg. p = 0.0772 |
| 304 | 5.293 | 0.000 | 2.712 | Neg. p = 0.0996 |

Significant sites at p <= 0.1

#### SLAC

| Codon | S | N | dS | dN | Selection detected? |
|---|---|---|---|---|---|
| 151 | 3.000 | 0 | 3.162 | 0 | Neg. p = 0.032 |

Significant sites at p <= 0.1

#### MEME

| Codon | alpha | beta+ | p+ | LRT | Episodic selection detected? | # branches | Most common codon substitutions at this site |
|---|---|---|---|---|---|---|---|

No sites found at p <= 0.1

### NSP6

#### FEL

| Codon | alpha | beta | LRT | Selection detected? |
|---|---|---|---|---|
| 118 | 7.536 | 0.000 | 3.556 | Neg. p = 0.0593 |
| 120 | 7.400 | 0.000 | 3.184 | Neg. p = 0.0743 |
| 149 | 0.000 | 7.689 | 2.739 | Pos. p = 0.0980 |
| 200 | 4.238 | 0.000 | 2.984 | Neg. p = 0.0841 |
| 262 | 12.358 | 0.000 | 12.426 | Neg. p = 0.0004 |
| 270 | 17.723 | 0.000 | 5.302 | Neg. p = 0.0213 |
| 284 | 18.001 | 0.000 | 10.773 | Neg. p = 0.0010 |
| 286 | 7.627 | 0.000 | 3.203 | Neg. p = 0.0735 |

Significant sites at p <= 0.1

#### SLAC

| Codon | S | N | dS | dN | Selection detected? |
|---|---|---|---|---|---|
| 200 | 2.000 | 0.000 | 2.140 | 0.000 | Neg. p = 0.097 |
| 262 | 6.000 | 0.000 | 6.000 | 0.000 | Neg. p = 0.001 |
| 270 | 2.000 | 0.000 | 2.409 | 0.000 | Neg. p = 0.078 |
| 284 | 8.000 | 0.000 | 8.259 | 0.000 | Neg. p = 0.000 |

Significant sites at p <= 0.1

#### MEME

| Codon | alpha | beta+ | p+ | LRT | Episodic selection detected? | # branches | Most common codon substitutions at this site |
|---|---|---|---|---|---|---|---|
| 106 | 0.000 | 2198.575 | 0.021 | 4.790 | Yes, p = 0.0420 | 1 | [1]TCT>ACT,TCT>TNT |
| 194 | 0.020 | 216.548 | 0.015 | 7.676 | Yes, p = 0.0096 | 1 | [1]GTT>ACT |

Significant sites at p <= 0.1

| NSP7 | | | | | | | | | | | | | | | | | | | | |
|---|---|---|---|---|---|---|---|---|---|---|---|---|---|---|---|---|---|---|---|---|
| **FEL** | | | | | | **SLAC** | | | | | | | **MEME** | | | | | | | |
| **Codon** | **alpha** | **beta** | **LRT** | **Selection detected?** | | **Codon** | **S** | **N** | **dS** | **dN** | **Selection detected?** | | **Codon** | **alpha** | **beta+** | **p+** | **LRT** | **Episodic selection detected?** | **# branches** | **Most common codon substitutions at this site** |
| No sites found at p <= 0.1 | | | | | | No sites found at p <= 0.1 | | | | | | | No sites found at p <= 0.1 | | | | | | | |

### NSP8

**FEL**

| Codon | alpha | beta | LRT | Selection detected? |
|---|---|---|---|---|
| 18 | 7.186 | 0 | 2.861 | Neg. p = 0.0908 |
| 91 | 7.186 | 0 | 3.166 | Neg. p = 0.0752 |
| 111 | 13.785 | 0 | 3.219 | Neg. p = 0.0728 |
| 121 | 7.401 | 0 | 3.245 | Neg. p = 0.0717 |
| 137 | 11.754 | 0 | 3.567 | Neg. p = 0.0590 |
| 162 | 11.754 | 0 | 3.681 | Neg. p = 0.0550 |
| 183 | 7.186 | 0 | 3.209 | Neg. p = 0.0732 |

Significant sites at p <= 0.1

**SLAC**

| Codon | S | N | dS | dN | Selection detected? |
|---|---|---|---|---|---|

No sites found at p <= 0.1

**MEME**

| Codon | alpha | beta+ | p+ | LRT | Episodic selection detected? | # branches | Most common codon substitutions at this site |
|---|---|---|---|---|---|---|---|

No sites found at p <= 0.1

### NSP9

| FEL | | | | |
|---|---|---|---|---|
| Codon | alpha | beta | LRT | Selection detected? |
| 6 | 3.886 | 0 | 2.989 | Neg. p = 0.0838 |
| 28 | 4.605 | 0 | 2.739 | Neg. p = 0.0979 |
| 31 | 8.729 | 0 | 5.680 | Neg. p = 0.0172 |
| 33 | 0 | 4.341 | 2.850 | Pos. p = 0.0914 |
| 42 | 8.077 | 0 | 6.241 | Neg. p = 0.0125 |
| 68 | 18.887 | 0 | 3.519 | Neg. p = 0.0607 |
| 93 | 9.695 | 0 | 2.781 | Neg. p = 0.0954 |
| 103 | 3.906 | 0 | 3.104 | Neg. p = 0.0781 |
| 112 | 3.951 | 0 | 2.900 | Neg. p = 0.0886 |

Significant sites at p <= 0.1

| SLAC | | | | | |
|---|---|---|---|---|---|
| Codon | S | N | dS | dN | Selection detected? |
| 31 | 4.000 | 0.000 | 4.366 | 0.000 | Neg. p = 0.010 |
| 95 | 3.000 | 0.000 | 3.087 | 0.000 | Neg. p = 0.034 |

Significant sites at p <= 0.1

| MEME | | | | | | | |
|---|---|---|---|---|---|---|---|
| Codon | alpha | beta+ | p+ | LRT | Episodic selection detected? | # branches | Most common codon substitutions at this site |
| 47 | 0.010 | 987.683 | 0.020 | 8.976 | Yes, p = 0.0049 | 1 | [1]GAT>TTT |

Significant sites at p <= 0.1

### NSP10

**FEL**

| Codon | alpha | beta | LRT | Selection detected? |
|---|---|---|---|---|
| 30 | 9.738 | 0 | 2.853 | Neg. p = 0.0912 |
| 59 | 8.624 | 0 | 3.993 | Neg. p = 0.0457 |
| 64 | 9.738 | 0 | 2.762 | Neg. p = 0.0965 |
| 65 | 32.365 | 0 | 3.516 | Neg. p = 0.0608 |
| 66 | 30.699 | 0 | 3.907 | Neg. p = 0.0481 |
| 83 | 9.738 | 0 | 2.993 | Neg. p = 0.0836 |

Significant sites at p <= 0.1

**SLAC**

| Codon | S | N | dS | dN | Selection detected? |
|---|---|---|---|---|---|

No sites found at p <= 0.1

**MEME**

| Codon | alpha | beta+ | p+ | LRT | Episodic selection detected? | # branches | Most common codon substitutions at this site |
|---|---|---|---|---|---|---|---|

No sites found at p <= 0.1
