## Supplementary File 5 for "Comparative genomics and characterization of SARS-CoV-2 P.1 (Gamma) Variant of Concern (VOC) from Amazonas, Brazil"

We gratefully acknowledge the following Authors from the Originating laboratories responsible for obtaining the specimens, as well as the Submitting laboratories where the genome data were generated and shared via GISAID, on which this research is based.

All Submitters of data may be contacted directly via www.gisaid.org

Authors are sorted alphabetically.

| Accession ID | Originating Laboratory | Submitting Laboratory | Authors |
|---|---|---|---|
| EPI_ISL_1061031, EPI_ISL_1061032 | CDL Laboratorio Santos e Vidal LTDA. | Instituto de Medicina Tropical de Sao Paulo | Brazil-UK Centre for Arbovirus Discovery Diagnosis Genomics and Epidemiology (CADDE) Genomic Network - Instituto de Medicina Tropical |
| EPI_ISL_833167, EPI_ISL_833168, EPI_ISL_833169, EPI_ISL_833170, EPI_ISL_833171, EPI_ISL_833172, EPI_ISL_833173, EPI_ISL_833174, EPI_ISL_833175, EPI_ISL_833176 | | | |
| see above | DB Diagnosticos do Brasil | Instituto Adolfo Lutz, Interdiciplinary Procedures Center, Strategic Laboratory | Claudia Regina Gonçalves; Claudio Tavares Sacchi; Erica Valessa Ramos Gomes; Karoline Rodrigues Campos |
| EPI_ISL_1708318, EPI_ISL_2499748 | Fundação Hospitalar de Hematologia e Hemoterapia do Amazonas | Laboratório de Estudos de Vírus Emergentes | Adriana S. S. Duarte; Alessandro S. Farias; Arilson Bernardo S. P. Gomes; Audrey B. Zangirolami; Bruno D. Benites; Camila A.M. Silva; Camila L. Simeoni; Camila Lopes Simeoni; Carolina Costa-Lima; Cecilia C. Camilo; Chieh-Hsi Wu; Christopher Dye; Clarice W. Arns; Daniel A. Toledo-Teixeira; Daniel Augusto de Toledo-Teixeira; Darlan S. Candido; Erika R. Manuli; Esmenia C. Rocha; Ester C. Sabino; Ester Cerdeira Sabino; Fabia; Fabiana Granja; Fernando R. Spilki; Flavia C. Sales; Gabriela Fabiano de Souza; Giulia M. Ferreira; Grazielle C. Maktura; Henrique Marques-Souza; Ingra M. Claro; Jaqueline G. de Jesus; José Luiz Proença Módena; Julia Forato; Karina Bispo dos Santos; Karina Bispo-dos-Santos; Lais D. Coimbra; Laís Durço Coimbra; Leandro M. Souza; Lucas A.M. Franco; Lucas I. Buscaratti; Luciana S. Mofatto; Luciana Souto Mofatto; Magnun N.N. Santos; Marcelo A.S. Mori; Marcelo Addas-Carvalho; Maria L. Moretti; Maria Luiza Moretti; Mariana C. Pinho; Mariana S. Ramundo; Mariene R. Amorim; Mariene Ribeiro Amorim; Michael S. Diamond; Myuki A.E. Crispim; Natalia S. Brunetti; Nelson Gaburo; Nuno R. Faria; Nuno Rodrigues Faria; Oliver G. Pybus; Pamela S. Andrade; Pierina L. Parise; Pierina Lorencini Parise; Priscilla P. Barbosa; Priscilla Paschoal Barbosa; Rafael E. Marques; Renata Sesti-Costa; Rodrigo N. Angerami; Rodrigo Nogueira Angerami; Stefanie Primon Muraro; Thais M. Coletti; Vitor A. Costa; William M. Souza; William Marciel de Souza |
| EPI_ISL_3149997, EPI_ISL_3149998, EPI_ISL_3149999, EPI_ISL_3150000, EPI_ISL_3150001, EPI_ISL_3150002, EPI_ISL_3150003, EPI_ISL_3150004, EPI_ISL_3150005, EPI_ISL_3150006, EPI_ISL_3150007, EPI_ISL_3150008, EPI_ISL_3150009, EPI_ISL_3150010, EPI_ISL_3150011, EPI_ISL_3150012, EPI_ISL_3150013, EPI_ISL_3150014, EPI_ISL_3150015, EPI_ISL_3150016, EPI_ISL_3150017 | | | |
| see above | Hermes Pardini | Universidade Federal de Ciencias da Saude de Porto Alegre | Andrea Roberto de Souza; Andy Goren; Carlos Gustavo Wambier; Claudia Elizabeth Thompson; Daniel do Nascimento Fonseca; Emilyn Oliveira Guerreiro; Flávio Adsuara Cadegiani; Liane Nanci Rotta; Patrícia Aline Gröhs Ferrareze; Ricardo Ariel Zimerman; Zhihua Ren |
| EPI_ISL_906080, EPI_ISL_906081 | Hospital Beneficiencia Portuguesa | Instituto Adolfo Lutz, Interdiciplinary Procedures Center, Strategic Laboratory | Claudia Regina Gonçalves; Claudio Tavares Sacchi; Erica Valessa Ramos Gomes; Karoline Rodrigues Campos |
| EPI_ISL_940626, EPI_ISL_940627 | Hospital Central Sao Caetano do Sul | Instituto Adolfo Lutz, Interdiciplinary Procedures Center, Strategic Laboratory | Claudia Regina Gonçalves; Claudio Tavares Sacchi; Erica Valessa Ramos Gomes; Karoline Rodrigues Campos |
| EPI_ISL_906075 | Hospital Geral de Vila Penteado Dr Jose Pangella Sao Paulo | Instituto Adolfo Lutz, Interdiciplinary Procedures Center, Strategic Laboratory | Claudia Regina Gonçalves; Claudio Tavares Sacchi; Erica Valessa Ramos Gomes; Karoline Rodrigues Campos |
| EPI_ISL_940620, EPI_ISL_940623, EPI_ISL_940624 | Hospital Sao Joaquim - Beneficiencia Portuguesa | Instituto Adolfo Lutz, Interdiciplinary Procedures Center, Strategic Laboratory | Claudia Regina Gonçalves; Claudio Tavares Sacchi; Erica Valessa Ramos Gomes; Karoline Rodrigues Campos |
| EPI_ISL_906076, EPI_ISL_906077 | Hospital Sao Luiz Sao Caetano | Instituto Adolfo Lutz, Interdiciplinary Procedures Center, Strategic Laboratory | Claudia Regina Gonçalves; Claudio Tavares Sacchi; Erica Valessa Ramos Gomes; Karoline Rodrigues Campos |
| EPI_ISL_906069 | Instituto Adolfo Lutz - Regional de Campinas | Instituto Adolfo Lutz, Interdiciplinary Procedures Center, Strategic Laboratory | Claudia Regina Gonçalves; Claudio Tavares Sacchi; Erica Valessa Ramos Gomes; Karoline Rodrigues Campos |
| EPI_ISL_925846, EPI_ISL_925916, EPI_ISL_926446 | LACEN - Laboratório Central de Saúde Pública do Amazonas | Evandro Chagas Institute Virology | A.M.; Barbagelata; E.C.; E.M.A.; Ferreira; J.A.; Junior; K.C.; L.C.; L.S.; M.C.; P.S.; Pinheiro; Santos; Silva; Sousa; Sousa Junior; W.D.C.; da Silva |
| EPI_ISL_918499, EPI_ISL_918500, EPI_ISL_918501, EPI_ISL_918502, EPI_ISL_918503, EPI_ISL_918504, EPI_ISL_918505, EPI_ISL_918506, EPI_ISL_918507, EPI_ISL_918508, EPI_ISL_918509, EPI_ISL_918510, EPI_ISL_918511, EPI_ISL_918512, EPI_ISL_918531, EPI_ISL_918532, EPI_ISL_918533, EPI_ISL_918534, EPI_ISL_918535, EPI_ISL_1261683, EPI_ISL_1261685, EPI_ISL_1261690, EPI_ISL_1261694, EPI_ISL_2488774, EPI_ISL_2488775 | | | |
| see above | LACEN - Laboratório Central de Saúde Pública do Amazonas | Evandro Chagas Institute | A.M.; Barbagelata; E.C.; E.M.A.; Ferreira; J.A.; Junior; K.C.; L.C.; L.S.; M.C.; P.S.; Pinheiro; Santos; Silva; Sousa; Sousa Junior; W.D.C.; da Silva |
| EPI_ISL_1166615 | LACEN - Laboratório Central de Saúde Pública do Rio Grande do Norte | Evandro Chagas Institute Virology | A.M.; Barbagelata; E.C.; E.M.A.; Ferreira; J.A.; Junior; K.C.; L.C.; L.S.; M.C.; P.S.; Pinheiro; Santos; Silva; Sousa; Sousa Junior; W.D.C.; da Silva |
| EPI_ISL_943990 | LACEN do Estado de Goias | Instituto Adolfo Lutz, Interdiciplinary Procedures Center, Strategic Laboratory | Claudia Regina Gonçalves; Claudio Tavares Sacchi; Erica Valessa Ramos Gomes; Karoline Rodrigues Campos |
| EPI_ISL_906071, EPI_ISL_940614, EPI_ISL_940615, EPI_ISL_940616, EPI_ISL_940617, EPI_ISL_940618 | LACEN-PI DR. Costa Alvarenga | Instituto Adolfo Lutz, Interdiciplinary Procedures Center, Strategic Laboratory | Claudia Regina Gonçalves; Claudio Tavares Sacchi; Erica Valessa Ramos Gomes; Karoline Rodrigues Campos |
| EPI_ISL_1213190, EPI_ISL_1213202, EPI_ISL_1213204, EPI_ISL_1213206 | LBM/UFPB | Bioinformatics Laboratory / LNCC | Alessandra P Lamarca; Alexandra L Gerber; Ana Paula Melo Mariano; Ana Paula de C Guimarães; Ana Tereza R Vasconcelos; Angela Maria Guimarães Santos; Bianca Mendes Maciel; Danielle Angst Secco; Eduardo Sérgio Soares Sousa; Eloiza Helena Campana; Francisco Paulo Freire Neto; George Rego Albuquerque; Kátia Castanho Scortecci; Lucymara Fassarella Agnez Lima; Luiz G P de Almeida; Luís Cristóvão Porto; Otavio J. Brustolini; Paulo Ricardo Nascimento; Ronaldo da Silva Francisco Jr; Sandra Rocha Gadelha; Selma Maria Bezerra Jeronimo; Vinícius Pietta Perez |
| EPI_ISL_4030305, EPI_ISL_4030306, EPI_ISL_4030311, EPI_ISL_4030312, EPI_ISL_4030313, EPI_ISL_4030318, EPI_ISL_4030320, EPI_ISL_4030321, EPI_ISL_4030322, EPI_ISL_4030323, EPI_ISL_4030368, EPI_ISL_4030369, EPI_ISL_4030370, EPI_ISL_4030371, EPI_ISL_4030372, EPI_ISL_4030373, EPI_ISL_4030374, EPI_ISL_4030375, EPI_ISL_4030376, EPI_ISL_4030377, EPI_ISL_4030378, EPI_ISL_4030379, EPI_ISL_4030380, EPI_ISL_4030381, EPI_ISL_4030382 | | | |
| see above | Laboratorio Central de Saude Publica do Amazonas - LACEN-AM | Laboratorio de Ecologia de Doencas Transmissiveis na Amazonia, Instituto Leonidas e Maria Deane - Fiocruz Amazonia | André Corado; Debora Duarte; Felipe Naveca; Fernanda Nascimento; George Silva; Karina Pessoa; Luciana Gonçalves; Maria Júlia Brandão; Matilde Mejía; Michele Jesus; Valdinete Nascimento; Victor Souza; Ágatha Costa |
| EPI_ISL_417034, EPI_ISL_792560, EPI_ISL_801386, EPI_ISL_801387, EPI_ISL_801388, EPI_ISL_801389, EPI_ISL_801390, EPI_ISL_801391, EPI_ISL_801392, EPI_ISL_801393, EPI_ISL_801394, EPI_ISL_801395, EPI_ISL_801396, EPI_ISL_811148, EPI_ISL_811149, EPI_ISL_833131, EPI_ISL_833132, EPI_ISL_833133, EPI_ISL_833134, EPI_ISL_833135, EPI_ISL_833136, EPI_ISL_833137, EPI_ISL_833138, EPI_ISL_833139, EPI_ISL_833140, EPI_ISL_1034304, EPI_ISL_1034305, EPI_ISL_1034306, EPI_ISL_1068078, EPI_ISL_1068079, EPI_ISL_1068080, EPI_ISL_1068081, EPI_ISL_1068082, EPI_ISL_1068083, EPI_ISL_1068084, EPI_ISL_1068085, EPI_ISL_1068086, EPI_ISL_1068087, EPI_ISL_1068088, EPI_ISL_1068089, EPI_ISL_1068090, EPI_ISL_1068091, EPI_ISL_1068092, EPI_ISL_1068093, EPI_ISL_1068094, EPI_ISL_1068095, EPI_ISL_1068096, EPI_ISL_1068097, EPI_ISL_1068098, EPI_ISL_1068099, EPI_ISL_1068100, EPI_ISL_1068101, EPI_ISL_1068102, EPI_ISL_1068103, EPI_ISL_1068104, EPI_ISL_1068105, EPI_ISL_1068106, EPI_ISL_1068107, EPI_ISL_1068108, EPI_ISL_1068109, EPI_ISL_1068110, EPI_ISL_1068111, EPI_ISL_1068112, EPI_ISL_1068113, EPI_ISL_1068114, EPI_ISL_1068115, EPI_ISL_1068117, EPI_ISL_1068118, EPI_ISL_1068119, EPI_ISL_1068120, EPI_ISL_1068121, EPI_ISL_1068122, EPI_ISL_1068123, EPI_ISL_1068124, EPI_ISL_1068125, EPI_ISL_1068126, EPI_ISL_1068127, EPI_ISL_1068128, EPI_ISL_1068129, EPI_ISL_1068130, EPI_ISL_1068131, EPI_ISL_1068132, EPI_ISL_1068133, EPI_ISL_1068134, EPI_ISL_1068135, EPI_ISL_1068136, EPI_ISL_1068137, EPI_ISL_1068138, EPI_ISL_1068139, EPI_ISL_1068140, EPI_ISL_1068141, EPI_ISL_1068142, EPI_ISL_1068143, EPI_ISL_1068144, EPI_ISL_1068145, EPI_ISL_1068146, EPI_ISL_1068147, EPI_ISL_1068148, EPI_ISL_1068149, EPI_ISL_1068150, EPI_ISL_1068151, EPI_ISL_1068152, EPI_ISL_1068153, EPI_ISL_1068154, EPI_ISL_1068155, EPI_ISL_1068156, EPI_ISL_1068157, EPI_ISL_1068158, EPI_ISL_1068159, EPI_ISL_1068160, EPI_ISL_1068161, EPI_ISL_1068162, EPI_ISL_1068163, EPI_ISL_1068164, EPI_ISL_1068165, EPI_ISL_1068166, EPI_ISL_1068167, EPI_ISL_1068168, EPI_ISL_1068169, EPI_ISL_1068170, EPI_ISL_1068171, EPI_ISL_1068172, EPI_ISL_1068173, EPI_ISL_1068174, EPI_ISL_1068175, EPI_ISL_1068176, EPI_ISL_1068177, EPI_ISL_1068178, EPI_ISL_1068179, EPI_ISL_1068180, EPI_ISL_1068181, EPI_ISL_1068182, EPI_ISL_1068183, EPI_ISL_1068184, EPI_ISL_1068185, EPI_ISL_1068186, EPI_ISL_1068187, EPI_ISL_1068188, EPI_ISL_1068189, EPI_ISL_1068190, EPI_ISL_1068191, EPI_ISL_1068192, EPI_ISL_1068193, EPI_ISL_1068194, EPI_ISL_1068195, EPI_ISL_1068196, EPI_ISL_1068197, EPI_ISL_1068198, EPI_ISL_1068201, EPI_ISL_1068203, EPI_ISL_1068204, EPI_ISL_1068205, EPI_ISL_1068206, EPI_ISL_1068207, EPI_ISL_1068208, EPI_ISL_1068209, EPI_ISL_1068210, EPI_ISL_1068211, EPI_ISL_1068212, EPI_ISL_1068213, EPI_ISL_1068214, EPI_ISL_1068215, EPI_ISL_1068216, EPI_ISL_1068217, EPI_ISL_1068218, EPI_ISL_1068219, EPI_ISL_1068221, EPI_ISL_1068222, EPI_ISL_1068223, EPI_ISL_1068224, EPI_ISL_1068225, EPI_ISL_1068226, EPI_ISL_1068227, EPI_ISL_1068228, EPI_ISL_1068229, EPI_ISL_1068230, EPI_ISL_1068231, EPI_ISL_1068232, EPI_ISL_1068233, EPI_ISL_1068234, EPI_ISL_1068235, EPI_ISL_1068236, EPI_ISL_1068237, EPI_ISL_1068238, EPI_ISL_1068239, EPI_ISL_1068240, EPI_ISL_1068241, EPI_ISL_1068242, EPI_ISL_1068243, EPI_ISL_1068244, EPI_ISL_1068245, EPI_ISL_1068246, EPI_ISL_1068247, EPI_ISL_1068248, EPI_ISL_1068249, EPI_ISL_1068250, EPI_ISL_1068251, EPI_ISL_1068252, EPI_ISL_1068253, EPI_ISL_1068254, EPI_ISL_1068255, EPI_ISL_1068256, EPI_ISL_1068257, EPI_ISL_1068258, EPI_ISL_1068259, EPI_ISL_1068260, EPI_ISL_1068261, EPI_ISL_1068262, EPI_ISL_1068263, EPI_ISL_1068264, EPI_ISL_1068265, EPI_ISL_1068266, EPI_ISL_1068268, EPI_ISL_1068269, EPI_ISL_1068270, EPI_ISL_1068271, EPI_ISL_1068272, EPI_ISL_1068273, EPI_ISL_1068274, EPI_ISL_1068275, EPI_ISL_1068276, EPI_ISL_1068277, EPI_ISL_1068278, EPI_ISL_1068279, EPI_ISL_1068280, EPI_ISL_1068281, EPI_ISL_1068282, EPI_ISL_1068283, EPI_ISL_1068284, EPI_ISL_1068285, EPI_ISL_1068286, EPI_ISL_1068287, EPI_ISL_1068288, EPI_ISL_1068289, EPI_ISL_1068290, EPI_ISL_1068291, EPI_ISL_1068292, EPI_ISL_1261122, EPI_ISL_1261123, EPI_ISL_1533609, EPI_ISL_1661250, EPI_ISL_1661251, EPI_ISL_1661252, EPI_ISL_2777360, EPI_ISL_2777361, EPI_ISL_2777362, EPI_ISL_2777363, EPI_ISL_2777364, EPI_ISL_2777365, EPI_ISL_2777366, EPI_ISL_2777367, EPI_ISL_2777368, EPI_ISL_2777369, EPI_ISL_2777370, EPI_ISL_2777371, EPI_ISL_2777372, EPI_ISL_2777373, EPI_ISL_2777374, EPI_ISL_2777375, EPI_ISL_2777376, EPI_ISL_2777377, EPI_ISL_2777378, EPI_ISL_2777379, EPI_ISL_2777380, EPI_ISL_2777381, EPI_ISL_2777382, EPI_ISL_2777383, EPI_ISL_2777384, EPI_ISL_2777385, EPI_ISL_2777386, EPI_ISL_2777387, EPI_ISL_2777388, EPI_ISL_2777389, EPI_ISL_2777390, EPI_ISL_2777391, EPI_ISL_2777392, EPI_ISL_2777393, EPI_ISL_2777394, EPI_ISL_2777395, EPI_ISL_2777396, EPI_ISL_2777397, EPI_ISL_2777398, EPI_ISL_2777399, EPI_ISL_2777400, EPI_ISL_2777401, EPI_ISL_2777402, EPI_ISL_2777403, EPI_ISL_2777404, EPI_ISL_2777405, EPI_ISL_2777406, EPI_ISL_2777407, EPI_ISL_2777408, EPI_ISL_2777409, EPI_ISL_2777410, EPI_ISL_2777411, EPI_ISL_2777412, EPI_ISL_2777413, EPI_ISL_2777414, EPI_ISL_2777415, EPI_ISL_2777416, EPI_ISL_2777417, EPI_ISL_2777418, EPI_ISL_2777419, EPI_ISL_2777420, EPI_ISL_2777421, EPI_ISL_2777423, EPI_ISL_2777424, EPI_ISL_2777425, EPI_ISL_2777426, EPI_ISL_2777427, EPI_ISL_2777428, EPI_ISL_2777429, EPI_ISL_2777430, EPI_ISL_2777431, EPI_ISL_2777432, EPI_ISL_2777433, EPI_ISL_2777436, EPI_ISL_2777437, EPI_ISL_2777438, EPI_ISL_2777439, EPI_ISL_2777440, EPI_ISL_2777441, EPI_ISL_2777442, EPI_ISL_2777443, EPI_ISL_2777444, EPI_ISL_2777445, EPI_ISL_2777446, EPI_ISL_2777447, EPI_ISL_2777448, EPI_ISL_2777449, EPI_ISL_2777450, EPI_ISL_2777451, EPI_ISL_2777452, EPI_ISL_2777453, EPI_ISL_2777454, EPI_ISL_2777455, EPI_ISL_2777456, EPI_ISL_2777457, EPI_ISL_2777458, EPI_ISL_2777459, EPI_ISL_2777460, EPI_ISL_2777461, EPI_ISL_2777462, EPI_ISL_2777463, EPI_ISL_2777464, EPI_ISL_2777465, EPI_ISL_2777466, EPI_ISL_2777467, EPI_ISL_2777468, EPI_ISL_2777469, EPI_ISL_2777470, EPI_ISL_2777471, EPI_ISL_2777472, EPI_ISL_2777473, EPI_ISL_2777474, EPI_ISL_2777475, EPI_ISL_2777476, EPI_ISL_2777477, EPI_ISL_2777478, EPI_ISL_2777479, EPI_ISL_2777480, EPI_ISL_2777481, EPI_ISL_2777482, EPI_ISL_2777483, EPI_ISL_2777484, EPI_ISL_2777485, EPI_ISL_2777486, EPI_ISL_2777487, EPI_ISL_2777488, EPI_ISL_2777489, EPI_ISL_2777490, EPI_ISL_2777491, EPI_ISL_2777492, EPI_ISL_2777493, EPI_ISL_2777494, EPI_ISL_2777995, | | | |

| | | | |
|---|---|---|---|
| EPI_ISL_2777996, EPI_ISL_2777997, EPI_ISL_2777998, EPI_ISL_2777999, EPI_ISL_2778000, EPI_ISL_2778001, EPI_ISL_2778003, EPI_ISL_3050307, EPI_ISL_3050457, EPI_ISL_3050515, EPI_ISL_3050516, EPI_ISL_3050517, EPI_ISL_3050574, EPI_ISL_3050591, EPI_ISL_3050615, EPI_ISL_3050616, EPI_ISL_3050617, EPI_ISL_3050618, EPI_ISL_3050641, EPI_ISL_4030308, EPI_ISL_4030309, EPI_ISL_4030310, EPI_ISL_4030314, EPI_ISL_4030315, EPI_ISL_4030316, EPI_ISL_4030317, EPI_ISL_4030319, EPI_ISL_4030324, EPI_ISL_4030325, EPI_ISL_4030326, EPI_ISL_4030327, EPI_ISL_4030328, EPI_ISL_4030329, EPI_ISL_4030330, EPI_ISL_4030331, EPI_ISL_4030332, EPI_ISL_4030333, EPI_ISL_4030334, EPI_ISL_4030335, EPI_ISL_4030336, EPI_ISL_4030337, EPI_ISL_4030338, EPI_ISL_4030339, EPI_ISL_4030340, EPI_ISL_4030341, EPI_ISL_4030342, EPI_ISL_4030343, EPI_ISL_4030344, EPI_ISL_4030345, EPI_ISL_4030346, EPI_ISL_4030347, EPI_ISL_4030348, EPI_ISL_4030349, EPI_ISL_4030350, EPI_ISL_4030351, EPI_ISL_4030352, EPI_ISL_4030353, EPI_ISL_4030354, EPI_ISL_4030355, EPI_ISL_4030356, EPI_ISL_4030357, EPI_ISL_4030358, EPI_ISL_4030359, EPI_ISL_4030360, EPI_ISL_4030361, EPI_ISL_4030362, EPI_ISL_4030363, EPI_ISL_4030364, EPI_ISL_4030365, EPI_ISL_4030366, EPI_ISL_4030367 | | | |
| see above | Laboratorio de Ecologia de Doencas Transmissiveis na Amazonia, Instituto Leonidas e Maria Deane - Fiocruz Amazonia | Laboratorio de Ecologia de Doencas Transmissiveis na Amazonia, Instituto Leonidas e Maria Deane - Fiocruz Amazonia | Alex Martins; André Corado; Debora Duarte; Fabiola Mendonça da Silva Chui; Felipe Naveca; Felipe Naveca on behalf of the Fiocruz COVID-19 Genomic Surveillance Network; Fernanda Nascimento; Fernando Fonseca de Almeida Val; George Silva; Gisely Cardoso de Melo; Karina Pessoa; Luciana Gonçalves; Marcus Vinicius Guimarães de Lacerda; Maria Júlia Brandão; Maria Paula Gomes Mourão; Mariana Xavier Simão; Matilde Mejía; Michele Jesus; Sérgio Luz; Valdinete Nascimento; Vanderson de Souza Sampaio; Victor Souza; Ágatha Costa |
| EPI_ISL_1402431 | Laboratory of Respiratory Viruses and Measles, Oswaldo Cruz Institute, FIOCRUZ | Laboratory of Respiratory Viruses and Measles, Oswaldo Cruz Institute, FIOCRUZ | Alex Pauvolid-Correa; Alice Sampaio Rocha; Ana Beatriz Machado Lima; Ana Carolina Mendonca; Anna Carolina Paixao; Felipe Naveca; Fernando Motta; Luciana Appolinario; Marilda Siqueira on behalf of the Fiocruz COVID-19 Genomic Surveillance Network; Mia Ferreira Araujo; Paola Resende; Renata Serrano Lopes |
| EPI_ISL_2241532, EPI_ISL_2241605 | Laboratório Central de Saúde Pública de Pernambuco | Coordenação Geral de Laboratórios de Saúde Pública (CGLAB/DAEVS/SVS/MS) | Vagner Fonseca; et al. |
| EPI_ISL_2298850, EPI_ISL_2298852, EPI_ISL_2298854, EPI_ISL_2298856, EPI_ISL_2298859, EPI_ISL_2298861, EPI_ISL_2298863, EPI_ISL_2298865, EPI_ISL_2298867, EPI_ISL_2298869, EPI_ISL_2298871 | | | |
| see above | Laboratório Central de Saúde Pública do Amazonas | Coordenação Geral de Laboratórios de Saúde Pública (CGLAB/DAEVS/SVS/MS) | Vagner Fonseca; et al. |
| EPI_ISL_2777236, EPI_ISL_2777237, EPI_ISL_2777238, EPI_ISL_2777239, EPI_ISL_2777240, EPI_ISL_2777241, EPI_ISL_2777242, EPI_ISL_2777243, EPI_ISL_2777244, EPI_ISL_2777245, EPI_ISL_2777246, EPI_ISL_2777247, EPI_ISL_2777248, EPI_ISL_2777249, EPI_ISL_2777250, EPI_ISL_2777251, EPI_ISL_2777252, EPI_ISL_2777253, EPI_ISL_2777254, EPI_ISL_2777255, EPI_ISL_2777256, EPI_ISL_2777257, EPI_ISL_2777258, EPI_ISL_2777259, EPI_ISL_2777260, EPI_ISL_2777261, EPI_ISL_2777262, EPI_ISL_2777263, EPI_ISL_2777264, EPI_ISL_2777265, EPI_ISL_2777266, EPI_ISL_2777267, EPI_ISL_2777268, EPI_ISL_2777269, EPI_ISL_2777270, EPI_ISL_2777271, EPI_ISL_2777272, EPI_ISL_2777273, EPI_ISL_2777274, EPI_ISL_2777275, EPI_ISL_2777276, EPI_ISL_2777277, EPI_ISL_2777278, EPI_ISL_2777279, EPI_ISL_2777280, EPI_ISL_2777281, EPI_ISL_2777282, EPI_ISL_2777283, EPI_ISL_2777284, EPI_ISL_2777285, EPI_ISL_2777287, EPI_ISL_2777288, EPI_ISL_2777290, EPI_ISL_2777292, EPI_ISL_2777293, EPI_ISL_2777295, EPI_ISL_2777297, EPI_ISL_2777299, EPI_ISL_2777301, EPI_ISL_2777302, EPI_ISL_2777304, EPI_ISL_2777306, EPI_ISL_2777309, EPI_ISL_2777311, EPI_ISL_2777313, EPI_ISL_2777314, EPI_ISL_2777316, EPI_ISL_2777318, EPI_ISL_2777320, EPI_ISL_2777321, EPI_ISL_2777324, EPI_ISL_2777327, EPI_ISL_2777328, EPI_ISL_2777329, EPI_ISL_2777330, EPI_ISL_2777331, EPI_ISL_2777332, EPI_ISL_2777333, EPI_ISL_2777334, EPI_ISL_2777335, EPI_ISL_2777337, EPI_ISL_2777338, EPI_ISL_2777339, EPI_ISL_2777340, EPI_ISL_2777341, EPI_ISL_2777342, EPI_ISL_2777343, EPI_ISL_2777344, EPI_ISL_2777345, EPI_ISL_2777346, EPI_ISL_2777347, EPI_ISL_2777348, EPI_ISL_2777349, EPI_ISL_2777350, EPI_ISL_2777351, EPI_ISL_2777352, EPI_ISL_2777353, EPI_ISL_2777354, EPI_ISL_2777355, EPI_ISL_2777356, EPI_ISL_2777357, EPI_ISL_2777358, EPI_ISL_2777359, EPI_ISL_2777495, EPI_ISL_2777496, EPI_ISL_2777497, EPI_ISL_2777498, EPI_ISL_2777499, EPI_ISL_2777500, EPI_ISL_2777501, EPI_ISL_2777502, EPI_ISL_2777503, EPI_ISL_2777504, EPI_ISL_2777505, EPI_ISL_2777506, EPI_ISL_2777507, EPI_ISL_2777508, EPI_ISL_2777509, EPI_ISL_2777510, EPI_ISL_2777511, EPI_ISL_2777512, EPI_ISL_2777513, EPI_ISL_2777514, EPI_ISL_2777515, EPI_ISL_2777516, EPI_ISL_2777517, EPI_ISL_2777518, EPI_ISL_2777519, EPI_ISL_2777520, EPI_ISL_2777521, EPI_ISL_2777522, EPI_ISL_2777523, EPI_ISL_2777524, EPI_ISL_2777525, EPI_ISL_2777526, EPI_ISL_2777527, EPI_ISL_2777528, EPI_ISL_2777529, EPI_ISL_2777530, EPI_ISL_2777531, EPI_ISL_2777532, EPI_ISL_2777533, EPI_ISL_2777534, EPI_ISL_2777535, EPI_ISL_2777536, EPI_ISL_2777537, EPI_ISL_2777538, EPI_ISL_2777539, EPI_ISL_2777540, EPI_ISL_2777541, EPI_ISL_2777542, EPI_ISL_2777543, EPI_ISL_2777544, EPI_ISL_2777545, EPI_ISL_2777546, EPI_ISL_2777547, EPI_ISL_2777548, EPI_ISL_2777549, EPI_ISL_2777550, EPI_ISL_2777551, EPI_ISL_2777552, EPI_ISL_2777553, EPI_ISL_2777554, EPI_ISL_2777555, EPI_ISL_2777556, EPI_ISL_2777557, EPI_ISL_2777558, EPI_ISL_2777559, EPI_ISL_2777560, EPI_ISL_2777561, EPI_ISL_2777562, EPI_ISL_2777563, EPI_ISL_2777564, EPI_ISL_2777565, EPI_ISL_2777566, EPI_ISL_2777567, EPI_ISL_2777568, EPI_ISL_2777569, EPI_ISL_2777570, EPI_ISL_2777571, EPI_ISL_2777572, EPI_ISL_2777573, EPI_ISL_2777574, EPI_ISL_2777575, EPI_ISL_2777576, EPI_ISL_2777577, EPI_ISL_2777578, EPI_ISL_2777579, EPI_ISL_2777580, EPI_ISL_2777581, EPI_ISL_2777582, EPI_ISL_2777583, EPI_ISL_2777584, EPI_ISL_2777585, EPI_ISL_2777586, EPI_ISL_2777587, EPI_ISL_2777588, EPI_ISL_2777589, EPI_ISL_2777590, EPI_ISL_2777591, EPI_ISL_2777592, EPI_ISL_2777593, EPI_ISL_2777594, EPI_ISL_2777596, EPI_ISL_2777597, EPI_ISL_2777598, EPI_ISL_2777599, EPI_ISL_2777600, EPI_ISL_2777601, EPI_ISL_2777602, EPI_ISL_2777603, EPI_ISL_2777604, EPI_ISL_2777605, EPI_ISL_2777606, EPI_ISL_2777607, EPI_ISL_2777608, EPI_ISL_2777609, EPI_ISL_2777610, EPI_ISL_2777611, EPI_ISL_2777612, EPI_ISL_2777613, EPI_ISL_2777614, EPI_ISL_2777615, EPI_ISL_2777616, EPI_ISL_2777617, EPI_ISL_2777618, EPI_ISL_2777619, EPI_ISL_2777620, EPI_ISL_2777621, EPI_ISL_2777622, EPI_ISL_2777623, EPI_ISL_2777624, EPI_ISL_2777625, EPI_ISL_2777626, EPI_ISL_2777627, EPI_ISL_2777628, EPI_ISL_2777629, EPI_ISL_2777630, EPI_ISL_2777631, EPI_ISL_2777632, EPI_ISL_2777633, EPI_ISL_2777634, EPI_ISL_2777635, EPI_ISL_2777636, EPI_ISL_2777637, EPI_ISL_2777638, EPI_ISL_2777639, EPI_ISL_2777640, EPI_ISL_2777641, EPI_ISL_2777642, EPI_ISL_2777643, EPI_ISL_2777644, EPI_ISL_2777645, EPI_ISL_2777646, EPI_ISL_2777648, EPI_ISL_2777649, EPI_ISL_2777650, EPI_ISL_2777651, EPI_ISL_2777652, EPI_ISL_2777653, EPI_ISL_2777654, EPI_ISL_2777655, EPI_ISL_2777656, EPI_ISL_2777657, EPI_ISL_2777658, EPI_ISL_2777659, EPI_ISL_2777660, EPI_ISL_2777661, EPI_ISL_2777662, EPI_ISL_2777663, EPI_ISL_2777664, EPI_ISL_2777665, EPI_ISL_2777666, EPI_ISL_2777667, EPI_ISL_2777668, EPI_ISL_2777669, EPI_ISL_2777670, EPI_ISL_2777671, EPI_ISL_2777672, EPI_ISL_2777673, EPI_ISL_2777674, EPI_ISL_2777675, EPI_ISL_2777676, EPI_ISL_2777677, EPI_ISL_2777678, EPI_ISL_2777679, EPI_ISL_2777680, EPI_ISL_2777681, EPI_ISL_2777682, EPI_ISL_2777683, EPI_ISL_2777684, EPI_ISL_2777685, EPI_ISL_2777686, EPI_ISL_2777687, EPI_ISL_2777688, EPI_ISL_2777689, EPI_ISL_2777690, EPI_ISL_2777691, EPI_ISL_2777692, EPI_ISL_2777693, EPI_ISL_2777694, EPI_ISL_2777695, EPI_ISL_2777696, EPI_ISL_2777697, EPI_ISL_2777698, EPI_ISL_2777699, EPI_ISL_2777700, EPI_ISL_2777701, EPI_ISL_2777702, EPI_ISL_2777703, EPI_ISL_2777704, EPI_ISL_2777705, EPI_ISL_2777706, EPI_ISL_2777707, EPI_ISL_2777708, EPI_ISL_2777709, EPI_ISL_2777710, EPI_ISL_2777711, EPI_ISL_2777712, EPI_ISL_2777713, EPI_ISL_2777714, EPI_ISL_2777715, EPI_ISL_2777716, EPI_ISL_2777717, EPI_ISL_2777718, EPI_ISL_2777719, EPI_ISL_2777720, EPI_ISL_2777721, EPI_ISL_2777722, EPI_ISL_2777723, EPI_ISL_2777724, EPI_ISL_2777725, EPI_ISL_2777726, EPI_ISL_2777727, EPI_ISL_2777728, EPI_ISL_2777729, EPI_ISL_2777730, EPI_ISL_2777731, EPI_ISL_2777732, EPI_ISL_2777733, EPI_ISL_2777734, EPI_ISL_2777735, EPI_ISL_2777736, EPI_ISL_2777737, EPI_ISL_2777738, EPI_ISL_2777739, EPI_ISL_2777740, EPI_ISL_2777741, EPI_ISL_2777742, EPI_ISL_2777743, EPI_ISL_2777744, EPI_ISL_2777745, EPI_ISL_2777746, EPI_ISL_2777747, EPI_ISL_2777748, EPI_ISL_2777749, EPI_ISL_2777750, EPI_ISL_2777751, EPI_ISL_2777752, EPI_ISL_2777753, EPI_ISL_2777754, EPI_ISL_2777755, EPI_ISL_2777756, EPI_ISL_2777757, EPI_ISL_2777758, EPI_ISL_2777759, EPI_ISL_2777760, EPI_ISL_2777761, EPI_ISL_2777762, EPI_ISL_2777763, EPI_ISL_2777764, EPI_ISL_2777765, EPI_ISL_2777766, EPI_ISL_2777767, EPI_ISL_2777768, EPI_ISL_2777769, EPI_ISL_2777770, EPI_ISL_2777771, EPI_ISL_2777772, EPI_ISL_2777773, EPI_ISL_2777774, EPI_ISL_2777775, EPI_ISL_2777776, EPI_ISL_2777777, EPI_ISL_2777778, EPI_ISL_2777779, EPI_ISL_2777780, EPI_ISL_2777781, EPI_ISL_2777782, EPI_ISL_2777783, EPI_ISL_2777784, EPI_ISL_2777785, EPI_ISL_2777786, EPI_ISL_2777787, EPI_ISL_2777788, EPI_ISL_2777789, EPI_ISL_2777790, EPI_ISL_2777791, EPI_ISL_2777792, EPI_ISL_2777793, EPI_ISL_2777794, EPI_ISL_2777795, EPI_ISL_2777796, EPI_ISL_2777797, EPI_ISL_2777798, EPI_ISL_2777799, EPI_ISL_2777800, EPI_ISL_2777801, EPI_ISL_2777802, EPI_ISL_2777803, EPI_ISL_2777804, EPI_ISL_2777805, EPI_ISL_2777806, EPI_ISL_2777807, EPI_ISL_2777808, EPI_ISL_2777809, EPI_ISL_2777810, EPI_ISL_2777811, EPI_ISL_2777812, EPI_ISL_2777813, EPI_ISL_2777814, EPI_ISL_2777815, EPI_ISL_2777816, EPI_ISL_2777817, EPI_ISL_2777818, EPI_ISL_2777819, EPI_ISL_2777820, EPI_ISL_2777821, EPI_ISL_2777822, EPI_ISL_2777823, EPI_ISL_2777824, EPI_ISL_2777825, EPI_ISL_2777826, EPI_ISL_2777827, EPI_ISL_2777828, EPI_ISL_2777829, EPI_ISL_2777830, EPI_ISL_2777831, EPI_ISL_2777832, EPI_ISL_2777833, EPI_ISL_2777834, EPI_ISL_2777835, EPI_ISL_2777836, EPI_ISL_2777837, EPI_ISL_2777838, EPI_ISL_2777839, EPI_ISL_2777840, EPI_ISL_2777841, EPI_ISL_2777842, EPI_ISL_2777843, EPI_ISL_2777844, EPI_ISL_2777845, EPI_ISL_2777846, EPI_ISL_2777847, EPI_ISL_2777848, EPI_ISL_2777849, EPI_ISL_2777850, EPI_ISL_2777851, EPI_ISL_2777852, EPI_ISL_2777853, EPI_ISL_2777854, EPI_ISL_2777855, EPI_ISL_2777856, EPI_ISL_2777857, EPI_ISL_2777858, EPI_ISL_2777859, EPI_ISL_2777860, EPI_ISL_2777861, EPI_ISL_2777862, EPI_ISL_2777863, EPI_ISL_2777864, EPI_ISL_2777865, EPI_ISL_2777866, EPI_ISL_2777867, EPI_ISL_2777868, EPI_ISL_2777869, EPI_ISL_2777870, EPI_ISL_2777871, EPI_ISL_2777872, EPI_ISL_2777873, EPI_ISL_2777874, EPI_ISL_2777875, EPI_ISL_2777876, EPI_ISL_2777877, EPI_ISL_2777878, EPI_ISL_2777879, EPI_ISL_2777880, EPI_ISL_2777881, EPI_ISL_2777882, EPI_ISL_2777883, EPI_ISL_2777884, EPI_ISL_2777885, EPI_ISL_2777886, EPI_ISL_2777887, EPI_ISL_2777888, EPI_ISL_2777889, EPI_ISL_2777890, EPI_ISL_2777891, EPI_ISL_2777892, EPI_ISL_2777893, EPI_ISL_2777894, EPI_ISL_2777895, EPI_ISL_2777896, EPI_ISL_2777897, EPI_ISL_2777898, EPI_ISL_2777899, EPI_ISL_2777900, EPI_ISL_2777901, EPI_ISL_2777902, EPI_ISL_2777903, EPI_ISL_2777904, EPI_ISL_2777905, EPI_ISL_2777906, EPI_ISL_2777907, EPI_ISL_2777908, EPI_ISL_2777909, EPI_ISL_2777910, EPI_ISL_2777911, EPI_ISL_2777912, EPI_ISL_2777913, EPI_ISL_2777914, EPI_ISL_2777915, EPI_ISL_2777916, EPI_ISL_2777917, EPI_ISL_2777918, EPI_ISL_2777919, EPI_ISL_2777920, EPI_ISL_2777921, EPI_ISL_2777922, EPI_ISL_2777923, EPI_ISL_2777924, EPI_ISL_2777925, EPI_ISL_2777926, EPI_ISL_2777927, EPI_ISL_2777928, EPI_ISL_2777929, EPI_ISL_2777930, EPI_ISL_2777931, EPI_ISL_2777932, EPI_ISL_2777933, EPI_ISL_2777934, EPI_ISL_2777935, EPI_ISL_2777936, EPI_ISL_2777937, EPI_ISL_2777938, EPI_ISL_2777939, EPI_ISL_2777940, EPI_ISL_2777941, EPI_ISL_2777942, EPI_ISL_2777943, EPI_ISL_2777944, EPI_ISL_2777945, EPI_ISL_2777946, EPI_ISL_2777947, EPI_ISL_2777948, EPI_ISL_2777949, EPI_ISL_2777950, EPI_ISL_2777951, EPI_ISL_2777952, EPI_ISL_2777953, EPI_ISL_2777954, EPI_ISL_2777955, EPI_ISL_2777956, EPI_ISL_2777957, EPI_ISL_2777958, EPI_ISL_2777959, EPI_ISL_2777960, EPI_ISL_2777961, EPI_ISL_2777962, EPI_ISL_2777963, EPI_ISL_2777964, EPI_ISL_2777965, EPI_ISL_2777966, EPI_ISL_2777967, EPI_ISL_2777968, EPI_ISL_2777969, EPI_ISL_2777970, EPI_ISL_2777971, EPI_ISL_2777972, EPI_ISL_2777973, EPI_ISL_2777974, EPI_ISL_2777975, EPI_ISL_2777976, EPI_ISL_2777977, EPI_ISL_2777978, EPI_ISL_2777979, EPI_ISL_2777980, EPI_ISL_2777981, EPI_ISL_2777982, EPI_ISL_2777983, EPI_ISL_2777984, EPI_ISL_2777985, EPI_ISL_2777986, EPI_ISL_2777987, EPI_ISL_2777988, EPI_ISL_2777989, EPI_ISL_2777990, EPI_ISL_2777991, EPI_ISL_2777992, EPI_ISL_2777993, EPI_ISL_2777994, EPI_ISL_2778002, EPI_ISL_2778004, EPI_ISL_2778005, EPI_ISL_3050295, EPI_ISL_3050296, EPI_ISL_3050297, EPI_ISL_3050298, EPI_ISL_3050299, EPI_ISL_3050300, EPI_ISL_3050302, EPI_ISL_3050303, EPI_ISL_3050304, EPI_ISL_3050305, EPI_ISL_3050306, EPI_ISL_3050308, EPI_ISL_3050309, EPI_ISL_3050310, EPI_ISL_3050311, EPI_ISL_3050312, EPI_ISL_3050313, EPI_ISL_3050314, EPI_ISL_3050315, EPI_ISL_3050316, EPI_ISL_3050317, EPI_ISL_3050318, EPI_ISL_3050319, EPI_ISL_3050320, EPI_ISL_3050321, EPI_ISL_3050322, EPI_ISL_3050323, EPI_ISL_3050324, EPI_ISL_3050325, EPI_ISL_3050326, EPI_ISL_3050327, EPI_ISL_3050328, EPI_ISL_3050329, EPI_ISL_3050330, EPI_ISL_3050331, EPI_ISL_3050332, EPI_ISL_3050333, EPI_ISL_3050334, EPI_ISL_3050335, EPI_ISL_3050336, EPI_ISL_3050337, EPI_ISL_3050338, EPI_ISL_3050339, EPI_ISL_3050340, EPI_ISL_3050341, EPI_ISL_3050342, EPI_ISL_3050343, EPI_ISL_3050344, EPI_ISL_3050345, EPI_ISL_3050346, EPI_ISL_3050347, EPI_ISL_3050348, EPI_ISL_3050349, EPI_ISL_3050350, EPI_ISL_3050351, EPI_ISL_3050352, EPI_ISL_3050353, EPI_ISL_3050354, EPI_ISL_3050355, EPI_ISL_3050356, EPI_ISL_3050358, EPI_ISL_3050359, EPI_ISL_3050360, EPI_ISL_3050361, EPI_ISL_3050362, EPI_ISL_3050363, EPI_ISL_3050364, EPI_ISL_3050365, EPI_ISL_3050366, EPI_ISL_3050367, EPI_ISL_3050368, EPI_ISL_3050369, EPI_ISL_3050370, EPI_ISL_3050371, EPI_ISL_3050372, EPI_ISL_3050373, EPI_ISL_3050374, EPI_ISL_3050375, EPI_ISL_3050376, EPI_ISL_3050377, EPI_ISL_3050378, EPI_ISL_3050379, EPI_ISL_3050380, EPI_ISL_3050381, EPI_ISL_3050382, EPI_ISL_3050383, EPI_ISL_3050384, EPI_ISL_3050385, EPI_ISL_3050386, EPI_ISL_3050387, EPI_ISL_3050388, EPI_ISL_3050389, EPI_ISL_3050390, EPI_ISL_3050391, EPI_ISL_3050392, EPI_ISL_3050393, EPI_ISL_3050394, EPI_ISL_3050395, EPI_ISL_3050396, EPI_ISL_3050397, EPI_ISL_3050398, EPI_ISL_3050399, EPI_ISL_3050400, EPI_ISL_3050401, EPI_ISL_3050402, EPI_ISL_3050403, EPI_ISL_3050404, EPI_ISL_3050405, EPI_ISL_3050406, EPI_ISL_3050407, EPI_ISL_3050408, EPI_ISL_3050409, EPI_ISL_3050410, EPI_ISL_3050411, EPI_ISL_3050412, EPI_ISL_3050413, EPI_ISL_3050414, EPI_ISL_3050415, EPI_ISL_3050416, EPI_ISL_3050417, EPI_ISL_3050418, EPI_ISL_3050419, EPI_ISL_3050420, EPI_ISL_3050421, EPI_ISL_3050422, EPI_ISL_3050423, EPI_ISL_3050424, EPI_ISL_3050425, EPI_ISL_3050426, EPI_ISL_3050427, EPI_ISL_3050428, EPI_ISL_3050429, EPI_ISL_3050430, EPI_ISL_3050431, EPI_ISL_3050432, EPI_ISL_3050433, EPI_ISL_3050434, EPI_ISL_3050435, EPI_ISL_3050436, EPI_ISL_3050437, EPI_ISL_3050438, EPI_ISL_3050439, EPI_ISL_3050440, EPI_ISL_3050441, EPI_ISL_3050442, EPI_ISL_3050443, EPI_ISL_3050444, EPI_ISL_3050445, EPI_ISL_3050446, EPI_ISL_3050447, EPI_ISL_3050448, EPI_ISL_3050449, EPI_ISL_3050450, EPI_ISL_3050451, EPI_ISL_3050452, EPI_ISL_3050453, EPI_ISL_3050454, EPI_ISL_3050455, EPI_ISL_3050456, EPI_ISL_3050458, EPI_ISL_3050459, EPI_ISL_3050460, EPI_ISL_3050461, EPI_ISL_3050462, EPI_ISL_3050463, EPI_ISL_3050464, EPI_ISL_3050465, EPI_ISL_3050466, EPI_ISL_3050467, EPI_ISL_3050468, EPI_ISL_3050469, EPI_ISL_3050470, EPI_ISL_3050471, EPI_ISL_3050472, EPI_ISL_3050473, EPI_ISL_3050474, EPI_ISL_3050475, EPI_ISL_3050476, EPI_ISL_3050477, EPI_ISL_3050478, EPI_ISL_3050479, EPI_ISL_3050480, EPI_ISL_3050481, EPI_ISL_3050482, EPI_ISL_3050483, EPI_ISL_3050484, EPI_ISL_3050485, EPI_ISL_3050486, EPI_ISL_3050487, EPI_ISL_3050488, EPI_ISL_3050489, EPI_ISL_3050490, EPI_ISL_3050491, EPI_ISL_3050492, EPI_ISL_3050493, EPI_ISL_3050494, EPI_ISL_3050495, EPI_ISL_3050496, EPI_ISL_3050497, EPI_ISL_3050498, EPI_ISL_3050499, EPI_ISL_3050500, EPI_ISL_3050501, EPI_ISL_3050502, EPI_ISL_3050503, EPI_ISL_3050504, EPI_ISL_3050505, EPI_ISL_3050506, EPI_ISL_3050507, EPI_ISL_3050508, EPI_ISL_3050509, EPI_ISL_3050510, EPI_ISL_3050511, EPI_ISL_3050512, EPI_ISL_3050513, EPI_ISL_3050514, EPI_ISL_3050518, EPI_ISL_3050519, EPI_ISL_3050520, EPI_ISL_3050521, EPI_ISL_3050522, EPI_ISL_3050523, EPI_ISL_3050524, EPI_ISL_3050525, EPI_ISL_3050526, EPI_ISL_3050527, EPI_ISL_3050528, EPI_ISL_3050529, EPI_ISL_3050530, EPI_ISL_3050531, EPI_ISL_3050532, EPI_ISL_3050533, EPI_ISL_3050534, EPI_ISL_3050535, EPI_ISL_3050536, EPI_ISL_3050537, EPI_ISL_3050538, EPI_ISL_3050539, EPI_ISL_3050540, EPI_ISL_3050541, EPI_ISL_3050542, EPI_ISL_3050543, EPI_ISL_3050544, EPI_ISL_3050545, EPI_ISL_3050546, EPI_ISL_3050547, EPI_ISL_3050548, EPI_ISL_3050549, EPI_ISL_3050550, EPI_ISL_3050551, EPI_ISL_3050552, EPI_ISL_3050553, EPI_ISL_3050554, EPI_ISL_3050555, EPI_ISL_3050556, EPI_ISL_3050557, EPI_ISL_3050558, EPI_ISL_3050559, EPI_ISL_3050560, EPI_ISL_3050561, EPI_ISL_3050562, EPI_ISL_3050563, EPI_ISL_3050564, EPI_ISL_3050565, EPI_ISL_3050566, EPI_ISL_3050567, EPI_ISL_3050568, EPI_ISL_3050569, EPI_ISL_3050570, EPI_ISL_3050571, EPI_ISL_3050572, EPI_ISL_3050573, EPI_ISL_3050575, EPI_ISL_3050576, EPI_ISL_3050577, EPI_ISL_3050578, EPI_ISL_3050579, EPI_ISL_3050580, EPI_ISL_3050581, EPI_ISL_3050582, EPI_ISL_3050583, EPI_ISL_3050584, EPI_ISL_3050585, EPI_ISL_3050586, EPI_ISL_3050587, EPI_ISL_3050588, EPI_ISL_3050589, EPI_ISL_3050590, EPI_ISL_3050592, EPI_ISL_3050593, EPI_ISL_3050594, EPI_ISL_3050595, EPI_ISL_3050596, EPI_ISL_3050597, EPI_ISL_3050598, EPI_ISL_3050599, EPI_ISL_3050600, EPI_ISL_3050601, EPI_ISL_3050602, EPI_ISL_3050603, EPI_ISL_3050604, EPI_ISL_3050605, EPI_ISL_3050606, EPI_ISL_3050607, EPI_ISL_3050608, EPI_ISL_3050609, EPI_ISL_3050610, EPI_ISL_3050611, EPI_ISL_3050612, EPI_ISL_3050613, EPI_ISL_3050614, EPI_ISL_3050619, EPI_ISL_3050620, EPI_ISL_3050621, EPI_ISL_3050622, EPI_ISL_3050623, EPI_ISL_3050624, EPI_ISL_3050625, EPI_ISL_3050626, EPI_ISL_3050627, EPI_ISL_3050628, EPI_ISL_3050629, EPI_ISL_3050630, EPI_ISL_3050631, EPI_ISL_3050632, EPI_ISL_3050633, EPI_ISL_3050634, EPI_ISL_3050635, EPI_ISL_3050636, EPI_ISL_3050637, EPI_ISL_3050638, EPI_ISL_3050639, EPI_ISL_3050640, EPI_ISL_3050642, EPI_ISL_3050643, EPI_ISL_3050644, EPI_ISL_3050645, EPI_ISL_3050646, EPI_ISL_3050647, EPI_ISL_3050648, EPI_ISL_3050649, EPI_ISL_3050650 | | | |
| see above | Laboratório Central de Saúde Pública do Amazonas - LACEN-AM | Laboratorio de Ecologia de Doencas Transmissiveis na Amazonia, Instituto Leonidas e Maria Deane - Fiocruz Amazonia | André Corado; Debora Duarte; Felipe Naveca; Fernanda Nascimento; George Silva; Karina Pessoa; Luciana Gonçalves; Maria Júlia Brandão; Matilde Mejía; Michele Jesus; Valdinete Nascimento; Victor Souza; Ágatha Costa |
| EPI_ISL_1239012 | Laboratório Central de Saúde Pública do Estado de Pernambuco (LACEN-PE) | WallauLab, Aggeu Magalhaes Institute | Alexandre Freitas da Silva; Antonio Mauro Rezende; Constância Flávia Junqueira Ayres; Cássia Docena; Duschinka Ribeiro Duarte Guedes; Elisama Helvecio; Filipe Zimmer Dezordi; Gabriel Luz Wallau on behalf of the Fiocruz COVID-19 Genomic Surveillance Network; Larissa Krokovsky; Laís Ceschini Machado; Marcelo Henrique dos Santos Paiva; Matheus Filgueira Bezerra; Sinval Pinto Brandão Filho |
| EPI_ISL_801397, EPI_ISL_801398, EPI_ISL_801399, EPI_ISL_801400, EPI_ISL_801401, EPI_ISL_801402, EPI_ISL_801403 | | | |
| see above | Laboratório Central de Saúde Pública do Estado do Amazonas (LACEN-AM) | Laboratorio de Ecologia de Doencas Transmissiveis na Amazonia, Instituto Leonidas e Maria Deane - Fiocruz Amazonia | André Corado; Debora Duarte; Felipe Naveca on behalf of the Fiocruz COVID-19 Genomic Surveillance Network; Fernanda Nascimento; George Silva; Luciana Gonçalves; Maria Júlia Brandão; Michele Jesus; Valdinete Nascimento; Victor Souza; Ágatha Costa |
| EPI_ISL_2466265, EPI_ISL_2466266, EPI_ISL_2466267, EPI_ISL_3061877 | Laboratório de Ecologia de Doenças Transmissíveis na Amazônia (EDTA), Instituto Leônidas e Maria Deane, FIOCRUZ, Manaus, Amazonas, Brazil. | Laboratory of Respiratory Viruses and Measles, Oswaldo Cruz Institute, FIOCRUZ | Alex Pauvolid-Corrêa; Alice Sampaio Rocha; Ana Beatriz Machado Lima; Ana Carolina Mendonca; Anna Carolina Paixao; Elisa Cavalcante Pereira; Felipe Naveca; Fernando Motta; Luciana Appolinario; Marilda Siqueira on behalf of the Fiocruz COVID-19 Genomic Surveillance Network; Mia Ferreira de Araujo; Paola Resende; Renata Serrano Lopes; Taina Venas |
| EPI_ISL_2233906 | Scientific Platform Pasteur USP, University of Sao Paulo | Scientific Platform Pasteur USP, University of Sao Paulo | Cunha; M.P.; Muxel; S.M. and Minoprio, P. |
| EPI_ISL_906070 | UPA Dr. Akira Tada | Instituto Adolfo Lutz, Interdiciplinary Procedures Center, Strategic Laboratory | Claudia Regina Gonçalves; Claudio Tavares Sacchi; Erica Valessa Ramos Gomes; Karoline Rodrigues Campos |
| EPI_ISL_2535159 | Unidade de apoio ao diagnostico da COVID - UNADIG | Bioinformatics Laboratory / LNCC | Alessandra P Lamarca; Alexandra L Gerber; Amilcar Tanuri; Ana Paula de C Guimaraes; Ana Tereza R Vasconcelos; Andrea Cony Cavalcanti; Caio Luiz Pereira Ribeiro; Cassia Alves; Cintia Policarpo; Claudia Maria Braga de Mello; Cristiane Gomes da Silva; Diana Mariani; Douglas Terra Machado; Flavio Dias da Silva; Gleidson da Silva de Oliveira; Leandro Magalhaes de Souza; Liliane Cavalcante; Luiz G P de Almeida; Marcio Henrique de Oliveira Garcia; Mario Sergio Ribeiro; Ronaldo da Silva F Jr; Silvia Carvalho |

We gratefully acknowledge the following Authors from the Originating laboratories responsible for obtaining the specimens, as well as the Submitting laboratories where the genome data were generated and shared via GISAID, on which this research is based.

All Submitters of data may be contacted directly via www.gisaid.org

Authors are sorted alphabetically.

| Accession ID | Originating Laboratory | Submitting Laboratory | Authors |
|---|---|---|---|
| EPI_ISL_2612324, EPI_ISL_2612325, EPI_ISL_2612326, EPI_ISL_2612328, EPI_ISL_2612330, EPI_ISL_2612343, EPI_ISL_2612347, EPI_ISL_2612348, EPI_ISL_2612349, EPI_ISL_2612350, EPI_ISL_2612351, EPI_ISL_2612352, EPI_ISL_2612353, EPI_ISL_2612355, EPI_ISL_2612358, EPI_ISL_2612360, EPI_ISL_2612361, EPI_ISL_2612362, EPI_ISL_2612363, EPI_ISL_2612364, EPI_ISL_2612365, EPI_ISL_2612366, EPI_ISL_2612372, EPI_ISL_2612373, EPI_ISL_2612377, EPI_ISL_2612378, EPI_ISL_2612379, EPI_ISL_2612380, EPI_ISL_2612391, EPI_ISL_2612392, EPI_ISL_2612393, EPI_ISL_2612394, EPI_ISL_2612395, EPI_ISL_2612396, EPI_ISL_2612398, EPI_ISL_2612399, EPI_ISL_2612400, EPI_ISL_2612402, EPI_ISL_2612407, EPI_ISL_2612408 | | | |
| see above | Centro de Infectologia Charles Mérieux/ Laboratório Rodolphe Mérieux, FUNDHACRE | Bioinformatics Laboratory / LNCC | Alessandra P Lamarca; Alexandra L Gerber; Ana Paula de C Guimarães; Ana Tereza R Vasconcelos; Andreas Stocker; Cirley Maria de Oliveira Lobato; Douglas Terra Machado; Luiz Fellype Alves de Souza; Luiz G P de Almeida; Ronaldo da Silva F Jr |
| EPI_ISL_2245062, EPI_ISL_2248780, EPI_ISL_2248781 | Laboratório Central de Saúde Pública do Acre | Coordenação Geral de Laboratórios de Saúde Pública (CGLAB/DAEVS/SVS/MS) | Vagner Fonseca; et al. |
| EPI_ISL_1251222, EPI_ISL_1251223, EPI_ISL_1251224, EPI_ISL_1251225 | Raimundo Reginaldo de Almeida | MOA Lab | Marcelo U. Ferreira; Priscila T. Rodrigues; Vanessa C. Nicolete |
| EPI_ISL_458139, EPI_ISL_848573, EPI_ISL_848574, EPI_ISL_848599, EPI_ISL_848600, EPI_ISL_848601, EPI_ISL_848602, EPI_ISL_848612, EPI_ISL_848613, EPI_ISL_848627 | | | |
| see above | Evandro Chagas Institute | Evandro Chagas Institute | A.M.; Barbagelata; E.C.; E.M.A.; Ferreira; G.M.R; H.R; J.A.; Junior; K.C.; L.C.; L.S.; M.C.; Martins; P.S.; Pinheiro; Resque; Santos; Silva; Sousa; Sousa Junior; Viana; W.D.C.; da Silva |

We gratefully acknowledge the following Authors from the Originating laboratories responsible for obtaining the specimens, as well as the Submitting laboratories where the genome data were generated and shared via GISAID, on which this research is based.

All Submitters of data may be contacted directly via www.gisaid.org

Authors are sorted alphabetically.

| Accession ID | Originating Laboratory | Submitting Laboratory | Authors |
|---|---|---|---|
| EPI_ISL_450873 | Evandro Chagas Institute | Evandro Chagas Institute | A.M.; Barbagelata; E.C.; E.M.A.; Ferreira; G.M.R; J.A.; Junior; L.C.; L.S.; M.C.; Martins; P.S.; Santos; Silva; Sousa; Sousa Junior; Viana; W.D.C.; da Silva |
| EPI_ISL_918551, EPI_ISL_918553, EPI_ISL_918554, EPI_ISL_918555, EPI_ISL_918556, EPI_ISL_918557, EPI_ISL_918558, EPI_ISL_918559, EPI_ISL_918560, EPI_ISL_918561 | | | |
| see above | LACEN - Laboratório Central de Saúde Pública do Amapa | Evandro Chagas Institute | A.M.; Barbagelata; E.C.; E.M.A.; Ferreira; J.A.; Junior; K.C.; L.C.; L.S.; M.C.; P.S.; Pinheiro; Santos; Silva; Sousa; Sousa Junior; W.D.C.; da Silva |
| EPI_ISL_1164976, EPI_ISL_1164981, EPI_ISL_1164982, EPI_ISL_1164984, EPI_ISL_1164985, EPI_ISL_1261686, EPI_ISL_1261688, EPI_ISL_1261689, EPI_ISL_1261692, EPI_ISL_1261696, EPI_ISL_2488776, EPI_ISL_2488777, EPI_ISL_2488792, EPI_ISL_2488793, EPI_ISL_2488794, EPI_ISL_2488804, EPI_ISL_2488805, EPI_ISL_2488806, EPI_ISL_2488807 | | | |
| see above | LACEN - Laboratório Central de Saúde Pública do Amapá | Evandro Chagas Institute | A.M.; Barbagelata; E.C.; E.M.A.; Ferreira; J.A.; Junior; K.C.; L.C.; L.S.; M.C.; P.S.; Pinheiro; Santos; Silva; Sousa; Sousa Junior; W.D.C.; da Silva |
| EPI_ISL_4030307 | Laboratorio Central de Saude Publica do Amazonas - LACEN-AM | Laboratorio de Ecologia de Doencas Transmissiveis na Amazonia, Instituto Leonidas e Maria Deane - Fiocruz Amazonia | André Corado; Debora Duarte; Felipe Naveca; Fernanda Nascimento; George Silva; Karina Pessoa; Luciana Gonçalves; Maria Júlia Brandão; Matilde Mejía; Michele Jesus; Valdinete Nascimento; Victor Souza; Ágatha Costa |
| EPI_ISL_2983086, EPI_ISL_2983087, EPI_ISL_2983088, EPI_ISL_2983089, EPI_ISL_2983090, EPI_ISL_2983091, EPI_ISL_2983092, EPI_ISL_2983093, EPI_ISL_2983094, EPI_ISL_2983095, EPI_ISL_2983096, EPI_ISL_2983097, EPI_ISL_2983098, EPI_ISL_2983099, EPI_ISL_2983100, EPI_ISL_2983101, EPI_ISL_2983102, EPI_ISL_2983103, EPI_ISL_2983104, EPI_ISL_2983105, EPI_ISL_2983106, EPI_ISL_2983107, EPI_ISL_2983108, EPI_ISL_2983109, EPI_ISL_2983110, EPI_ISL_2983111, EPI_ISL_2983112, EPI_ISL_2983113, EPI_ISL_2983114, EPI_ISL_2983116, EPI_ISL_2983117, EPI_ISL_2983118, EPI_ISL_2983119, EPI_ISL_2983120, EPI_ISL_2983121, EPI_ISL_2983122, EPI_ISL_2983123, EPI_ISL_2983124, EPI_ISL_2983125, EPI_ISL_2983126, EPI_ISL_2983128, EPI_ISL_2983129, EPI_ISL_2983131, EPI_ISL_2983132, EPI_ISL_2983133, EPI_ISL_2983134, EPI_ISL_2983135, EPI_ISL_2983136, EPI_ISL_2983137, EPI_ISL_2983139, EPI_ISL_2983140, EPI_ISL_2983141, EPI_ISL_2983142, EPI_ISL_2983143, EPI_ISL_2983144, EPI_ISL_2983145, EPI_ISL_2983146, EPI_ISL_2983147, EPI_ISL_2983149, EPI_ISL_2983150, EPI_ISL_2983151, EPI_ISL_2983152, EPI_ISL_2983153, EPI_ISL_2983154, EPI_ISL_2983155, EPI_ISL_2983156, EPI_ISL_2983157, EPI_ISL_2983158, EPI_ISL_2983159, EPI_ISL_2983160, EPI_ISL_2983162, EPI_ISL_2983163, EPI_ISL_2983164, EPI_ISL_2983165, EPI_ISL_2983166, EPI_ISL_2983167, EPI_ISL_2983169, EPI_ISL_2983170, EPI_ISL_2983171, EPI_ISL_2983172, EPI_ISL_2983173, EPI_ISL_2983174, EPI_ISL_2983175, EPI_ISL_2983177, EPI_ISL_2983178, EPI_ISL_2983476, EPI_ISL_3434898, EPI_ISL_3434899, EPI_ISL_3434900, EPI_ISL_3434901, EPI_ISL_3434902, EPI_ISL_3434903, EPI_ISL_3434904, EPI_ISL_3434948, EPI_ISL_3434956, EPI_ISL_3434957, EPI_ISL_3434958, EPI_ISL_3434959, EPI_ISL_3434960, EPI_ISL_3434961, EPI_ISL_3434962, EPI_ISL_3434963, EPI_ISL_3434964, EPI_ISL_3434965, EPI_ISL_3434966, EPI_ISL_3434967, EPI_ISL_3434968, EPI_ISL_3434969, EPI_ISL_3434970, EPI_ISL_3434971, EPI_ISL_3434972, EPI_ISL_3434973, EPI_ISL_3434974, EPI_ISL_3434975, EPI_ISL_3434976, EPI_ISL_3434977, EPI_ISL_3434978, EPI_ISL_3434979, EPI_ISL_3434980, EPI_ISL_3434981, EPI_ISL_3434982, EPI_ISL_3434983, EPI_ISL_3435067, EPI_ISL_3539742, EPI_ISL_3539743, EPI_ISL_3539744, EPI_ISL_3539745, EPI_ISL_3539746, EPI_ISL_3539747, EPI_ISL_3539748, EPI_ISL_3539749, EPI_ISL_3539795, EPI_ISL_3539796, EPI_ISL_3539797, EPI_ISL_3539798, EPI_ISL_3539799, EPI_ISL_3539800, EPI_ISL_3539801, EPI_ISL_3539802, EPI_ISL_3539803, EPI_ISL_3539804, EPI_ISL_3539805, EPI_ISL_3539806, EPI_ISL_3539807, EPI_ISL_3539808, EPI_ISL_3539809, EPI_ISL_3539811, EPI_ISL_3539812, EPI_ISL_3539813, EPI_ISL_3539814, EPI_ISL_3539815, EPI_ISL_3539816, EPI_ISL_3539817, EPI_ISL_3539818, EPI_ISL_3539819, EPI_ISL_3539820, EPI_ISL_3539821, EPI_ISL_3539822, EPI_ISL_3539823, EPI_ISL_3539824, EPI_ISL_3539825, EPI_ISL_3539826, EPI_ISL_3539827, EPI_ISL_3539828, EPI_ISL_3539931, EPI_ISL_3539932, EPI_ISL_4080931, EPI_ISL_4080932, EPI_ISL_4080933, EPI_ISL_4081029, EPI_ISL_4081030, EPI_ISL_4081031, EPI_ISL_4081032, EPI_ISL_4081033, EPI_ISL_4081034, EPI_ISL_4081035, EPI_ISL_4081036, EPI_ISL_4081037, EPI_ISL_4081038, EPI_ISL_4081039, EPI_ISL_4081040, EPI_ISL_4081041, EPI_ISL_4081042, EPI_ISL_4081043, EPI_ISL_4081044, EPI_ISL_4081045, EPI_ISL_4081046, EPI_ISL_4081047, EPI_ISL_4081048, EPI_ISL_4081049, EPI_ISL_4081050, EPI_ISL_4081051, EPI_ISL_4081052, EPI_ISL_4081053, EPI_ISL_4081054, EPI_ISL_4081055, EPI_ISL_4081056, EPI_ISL_4081057, EPI_ISL_4081058, EPI_ISL_4081059, EPI_ISL_4081060, EPI_ISL_4081061, EPI_ISL_4081062, EPI_ISL_4081063, EPI_ISL_4081064, EPI_ISL_4081065, EPI_ISL_4081066 | | | |
| see above | Laboratorio Central de Saude Publica do Estado do Amapa (LACEN/AP) | Laboratory of Respiratory Viruses and Measles, Oswaldo Cruz Institute, FIOCRUZ | Agatha Cristinne Prudencio; Alice Sampaio Rocha; Ana Carolina Mendonca; Andreia Santos Costa; Anna Carolina Paixao; Anne Caroline da Silva Soledade; Elisa Cavalcante Pereira; Fernando Motta; Ighor Leonardo Arantes Gomes; Lindomar dos Anjos Silva; Luciana Appolinario; Marcia Socorro Pereira Cavalcante; Marilda Siqueira on behalf of the Fiocruz COVID-19 Genomic Surveillance Network; Paola Resende; Renata Serrano Lopes; Taina Venas |
| EPI_ISL_2241547 | Laboratório Central de Saúde Pública de Pernambuco | Coordenação Geral de Laboratórios de Saúde Pública (CGLAB/DAEVS/SVS/MS) | Vagner Fonseca; et al. |
| EPI_ISL_2245064, EPI_ISL_2245065, EPI_ISL_2245066, EPI_ISL_2245067, EPI_ISL_2245068, EPI_ISL_2245069, EPI_ISL_2245070, EPI_ISL_2245071, EPI_ISL_2245072, EPI_ISL_2245073, EPI_ISL_2245074, EPI_ISL_2245075, EPI_ISL_2245076, EPI_ISL_2245077, EPI_ISL_2245078, EPI_ISL_2245079, EPI_ISL_2248784, EPI_ISL_2248785, EPI_ISL_2248786, EPI_ISL_2248787, EPI_ISL_2298747, EPI_ISL_2298760, EPI_ISL_2298790, EPI_ISL_2298791, EPI_ISL_2298793, EPI_ISL_2298795, EPI_ISL_2298797, EPI_ISL_2298799, EPI_ISL_2298800, EPI_ISL_2298801, EPI_ISL_2298803, EPI_ISL_2298804, EPI_ISL_2298806, EPI_ISL_2298808, EPI_ISL_2298816, EPI_ISL_2298817, EPI_ISL_2298819, EPI_ISL_2298821, EPI_ISL_2298823, EPI_ISL_2298824, EPI_ISL_2298826, EPI_ISL_2298828, EPI_ISL_2298830, EPI_ISL_2298832 | | | |
| see above | Laboratório Central de Saúde Pública do Amapá | Coordenação Geral de Laboratórios de Saúde Pública (CGLAB/DAEVS/SVS/MS) | Vagner Fonseca; et al. |
| EPI_ISL_458138, EPI_ISL_458142, EPI_ISL_458143, EPI_ISL_458144, EPI_ISL_458145, EPI_ISL_524784, EPI_ISL_524792, EPI_ISL_524793, EPI_ISL_524795, EPI_ISL_524796, EPI_ISL_848575, EPI_ISL_848576, EPI_ISL_848577, EPI_ISL_848578, EPI_ISL_848579, EPI_ISL_848584 | | | |
| see above | Evandro Chagas Institute | Evandro Chagas Institute | A.M.; Barbagelata; E.C.; E.M.A.; Ferreira; G.M.R; H.R; J.A.; Junior; K.C.; L.C.; L.S.; M.C.; Martins; P.S.; Pinheiro; Resque; Santos; Silva; Sousa; Sousa Junior; Viana; W.D.C.; da Silva |

We gratefully acknowledge the following Authors from the Originating laboratories responsible for obtaining the specimens, as well as the Submitting laboratories where the genome data were generated and shared via GISAID, on which this research is based.

All Submitters of data may be contacted directly via www.gisaid.org

Authors are sorted alphabetically.

| Accession ID | Originating Laboratory | Submitting Laboratory | Authors |
|---|---|---|---|
| EPI_ISL_1293052, EPI_ISL_1293053, EPI_ISL_1293054, EPI_ISL_1293055, EPI_ISL_1303499, EPI_ISL_1303500, EPI_ISL_1303501, EPI_ISL_1303502, EPI_ISL_1303503, EPI_ISL_1303504, EPI_ISL_1303505 | | | |
| see above | LACEN de Rondonia | Instituto Adolfo Lutz, Interdiciplinary Procedures Center, Strategic Laboratory | Caio Vinicius Dias Lopes; Claudia Regina Gonçalves; Claudio Tavares Sacchi; Erica Valessa Ramos Gomes; Karoline Rodrigues Campos |
| EPI_ISL_1493578, EPI_ISL_1493579, EPI_ISL_1493584, EPI_ISL_1493595, EPI_ISL_1493596, EPI_ISL_1493597, EPI_ISL_1493598, EPI_ISL_1493599, EPI_ISL_1493600, EPI_ISL_1494924, EPI_ISL_1520107, EPI_ISL_1520108, EPI_ISL_1520109 | | | |
| see above | LACEN do Estado de Rondonia | Instituto Adolfo Lutz, Interdiciplinary Procedures Center, Strategic Laboratory | Caio Vinicius Dias Lopes; Claudia Regina Gonçalves; Claudio Tavares Sacchi; Erica Valessa Ramos Gomes; Karoline Rodrigues Campos |
| EPI_ISL_514131, EPI_ISL_514132, EPI_ISL_514133, EPI_ISL_514134, EPI_ISL_514135, EPI_ISL_514136, EPI_ISL_514137, EPI_ISL_514138 | | | |
| see above | Rondônia Central Public Health Laboratory (LACEN/RO), vinculated to State Health Secretariat of Rondônia (SESAU/RO) | Molecular Virology Laboratory of Oswaldo Cruz Foundation of Rondônia | Adriana Cristina Salvador Maia; Alcione de Oliveira dos Santos; Alice Paula Di Sabatino Guimarães; Aline Linhares Ferreira de Melo Mendonça; Caio Henrique Nemeth Santos; Camila Flávia Gomes Azzi; Celina Aparecida Bertoni Lugtenburg; Cicileia Correia da Silva; Felipe Gomes Naveca; Felipe Souza Nogueira-Lima; Fernando Rodrigues Máximo; Jansen Fernandes de Medeiros; Juan Miguel Vilallobos-Salcedo and Deusilene Souza Vieira; Juan Miguel Vilallobos-Salcedo and Deusilene Souza Vieira.; Juan Miguel Vilallobos-Salcedo and Deusilene Souza Vieira1; Juliana Loca Furtado; Luan Felipe Botelho-Souza; Suelen Cavalcante; Tárcio Peixoto Roca; Rita de Cássia Pontello Rampazzo |

We gratefully acknowledge the following Authors from the Originating laboratories responsible for obtaining the specimens, as well as the Submitting laboratories where the genome data were generated and shared via GISAID, on which this research is based.

All Submitters of data may be contacted directly via www.gisaid.org

Authors are sorted alphabetically.

| Accession ID | Originating Laboratory | Submitting Laboratory | Authors |
| --- | --- | --- | --- |
| EPI_ISL_940630, EPI_ISL_943967, EPI_ISL_943968, EPI_ISL_943969, EPI_ISL_943970, EPI_ISL_943971 | Hospital Geral de Sao Paulo | Instituto Adolfo Lutz, Interdiciplinary Procedures Center, Strategic Laboratory | Claudia Regina Gonçalves; Claudio Tavares Sacchi; Erica Valessa Ramos Gomes; Karoline Rodrigues Campos |
| EPI_ISL_1261687 | LACEN - Laboratório Central de Saúde Pública de Roraima | Evandro Chagas Institute | A.M.; Barbagelata; E.C.; E.M.A.; Ferreira; J.A.; Junior; K.C.; L.C.; L.S.; M.C.; P.S.; Pinheiro; Santos; Silva; Sousa; Sousa Junior; W.D.C.; da Silva |
| EPI_ISL_918513, EPI_ISL_2488772, EPI_ISL_2488780, EPI_ISL_2488781, EPI_ISL_2488782, EPI_ISL_2488783, EPI_ISL_2488784, EPI_ISL_2488785, EPI_ISL_2488786 | | | |
| see above | LACEN - Laboratório Central de Saúde Pública do Roraima | Evandro Chagas Institute | A.M.; Barbagelata; E.C.; E.M.A.; Ferreira; J.A.; Junior; K.C.; L.C.; L.S.; M.C.; P.S.; Pinheiro; Santos; Silva; Sousa; Sousa Junior; W.D.C.; da Silva |
| EPI_ISL_2245107, EPI_ISL_2245108, EPI_ISL_2245109, EPI_ISL_2245110, EPI_ISL_2245111, EPI_ISL_2245112, EPI_ISL_2245113, EPI_ISL_2298748, EPI_ISL_2298762, EPI_ISL_2298764, EPI_ISL_2298765, EPI_ISL_2298767, EPI_ISL_2298769, EPI_ISL_2298788, EPI_ISL_2298847, EPI_ISL_2298849 | | | |
| see above | Laboratório Central de Saúde Pública de Roraima | Coordenação Geral de Laboratórios de Saúde Pública (CGLAB/DAEVS/SVS/MS) | Vagner Fonseca; et al. |
| EPI_ISL_2778007, EPI_ISL_2778008 | Laboratório Central de Saúde Pública do Amazonas - LACEN-AM | Laboratorio de Ecologia de Doencas Transmissiveis na Amazonia, Instituto Leonidas e Maria Deane - Fiocruz Amazonia | André Corado; Debora Duarte; Felipe Naveca; Fernanda Nascimento; George Silva; Karina Pessoa; Luciana Gonçalves; Maria Júlia Brandão; Matilde Mejía; Michele Jesus; Valdinete Nascimento; Victor Souza; Ágatha Costa |
| EPI_ISL_848556, EPI_ISL_848586, EPI_ISL_848610, EPI_ISL_848616 | Evandro Chagas Institute | Evandro Chagas Institute | A.M.; Barbagelata; E.C.; E.M.A.; Ferreira; J.A.; Junior; K.C.; L.C.; L.S.; M.C.; P.S.; Pinheiro; Santos; Silva; Sousa; Sousa Junior; W.D.C.; da Silva |

We gratefully acknowledge the following Authors from the Originating laboratories responsible for obtaining the specimens, as well as the Submitting laboratories where the genome data were generated and shared via GISAID, on which this research is based.

All Submitters of data may be contacted directly via www.gisaid.org

Authors are sorted alphabetically.

| Accession ID | Originating Laboratory | Submitting Laboratory | Authors |
|---|---|---|---|
| EPI_ISL_3663878, EPI_ISL_3663887, EPI_ISL_3663893, EPI_ISL_3664065, EPI_ISL_3664168, EPI_ISL_3664170 | DASA | DASA | Adriano Bonaldi; Angelica Hristov; Annelise Lopes; Bianca Cota; Cristina Oliveira; Jose Levi; Lidia Yamamoto; Paulo Pierry; Rodrigo Guarischi; Rodrigo Salazar |
| EPI_ISL_2756435, EPI_ISL_2756436, EPI_ISL_2756437, EPI_ISL_2756438, EPI_ISL_2756439, EPI_ISL_2756440, EPI_ISL_2756441, EPI_ISL_2756442, EPI_ISL_2756443, EPI_ISL_2756444, EPI_ISL_2756445, EPI_ISL_2756446, EPI_ISL_2756447, EPI_ISL_2756448, EPI_ISL_2756449, EPI_ISL_2756450, EPI_ISL_2756451, EPI_ISL_2756452, EPI_ISL_2756453, EPI_ISL_2756454, EPI_ISL_2756455, EPI_ISL_2756456, EPI_ISL_2756457, EPI_ISL_2756458, EPI_ISL_2756459, EPI_ISL_2756460, EPI_ISL_2756461, EPI_ISL_2756462, EPI_ISL_2756463, EPI_ISL_2756464, EPI_ISL_2756465, EPI_ISL_2756466, EPI_ISL_2756467, EPI_ISL_2756468, EPI_ISL_2756469, EPI_ISL_2756470, EPI_ISL_2756471 | | | |
| see above | Instituto Adolfo Lutz Central | Instituto Adolfo Lutz, Interdiciplinary Procedures Center, Strategic Laboratory | Caio Vinicius Dias Lopes; Claudia Regina Gonçalves; Claudio Tavares Sacchi; Erica Valessa Ramos Gomes; Karoline Rodrigues Campos; Leonardo Jose Tadeu de Araujo |
| EPI_ISL_943973, EPI_ISL_943974, EPI_ISL_943975, EPI_ISL_943976, EPI_ISL_943977, EPI_ISL_943978, EPI_ISL_943979, EPI_ISL_943980, EPI_ISL_943981, EPI_ISL_943982, EPI_ISL_943983, EPI_ISL_943985, EPI_ISL_943986, EPI_ISL_943987, EPI_ISL_943991, EPI_ISL_1303509, EPI_ISL_2919205, EPI_ISL_2919206, EPI_ISL_2919207, EPI_ISL_2919208, EPI_ISL_2919209, EPI_ISL_2919210, EPI_ISL_2919211, EPI_ISL_2919212, EPI_ISL_2919213, EPI_ISL_2919214, EPI_ISL_2919215, EPI_ISL_2919216, EPI_ISL_2919217, EPI_ISL_2919218, EPI_ISL_2919219, EPI_ISL_2919220, EPI_ISL_2919221, EPI_ISL_2919222, EPI_ISL_2919223, EPI_ISL_2919224, EPI_ISL_2919225, EPI_ISL_2919226, EPI_ISL_2919227, EPI_ISL_2958854, EPI_ISL_3316155, EPI_ISL_3316213 | | | |
| see above | LACEN do Estado de Tocantins | Instituto Adolfo Lutz, Interdiciplinary Procedures Center, Strategic Laboratory | Caio Vinicius Dias Lopes; Claudia Regina Gonçalves; Claudio Tavares Sacchi; Erica Valessa Ramos Gomes; Karoline Rodrigues Campos; Leonardo Tadeu de Araujo; Marlon Benedito Nascomento Santos |
| EPI_ISL_3235304, EPI_ISL_3235305, EPI_ISL_3235306, EPI_ISL_3235307, EPI_ISL_3235308, EPI_ISL_3235310, EPI_ISL_3235311, EPI_ISL_3235312, EPI_ISL_3235313, EPI_ISL_3235314, EPI_ISL_4080794, EPI_ISL_4080795, EPI_ISL_4080796, EPI_ISL_4080797, EPI_ISL_4080798, EPI_ISL_4080799, EPI_ISL_4080800, EPI_ISL_4080801, EPI_ISL_4080802, EPI_ISL_4080803, EPI_ISL_4080804, EPI_ISL_4080805 | | | |
| see above | Laboratorio Central de Saude Publica do Estado do Tocantins (LACEN/TO) | Laboratory of Respiratory Viruses and Measles, Oswaldo Cruz Institute, FIOCRUZ | Agatha Soares; Alice Sampaio Rocha; Ana Carolina Mendonca; Anna Carolina Paixao; Elisa Cavalcante Pereira; Fernando Motta; Ighor Arantes; Jucimaria Dantas Galvao; Luciana Appolinario; Marilda Siqueira on behalf of the Fiocruz COVID-19 Genomic Surveillance Network; Paola Resende; Renata Serrano Lopes; Taina Venas |
| EPI_ISL_3426438, EPI_ISL_3584949, EPI_ISL_3585952, EPI_ISL_3587057, EPI_ISL_3587490, EPI_ISL_3602957, EPI_ISL_3602958, EPI_ISL_3602960, EPI_ISL_3602961, EPI_ISL_3602964, EPI_ISL_3610172, EPI_ISL_3610352, EPI_ISL_3610353, EPI_ISL_3610354, EPI_ISL_3610355, EPI_ISL_3610356, EPI_ISL_3610357, EPI_ISL_3610358, EPI_ISL_3610359 | | | |
| see above | Laboratório Central de Saúde Pública do Estado do Tocantins (LACEN/TO) | Bioinformatics and Biotechnology Laboratory (Labinftec/Federal University of Tocantins, Campus of Gurupi) | Bergmann Morais Ribeiro; Chácaras 69/72; Fabrício Souza Campos; Fernando Lucas Melo; Gurupi/TO Ueric José Borges de Souza; Jucimária Dantas Galvão; Laboratório de Bioinformática e Biotecnologia; Lote 7; Raíssa Nunes dos Santos; Rua Badejós; Ueric José Borges de Souza; Zona Rural \| 77.402-970\| UFT Campus de Gurupi |
| EPI_ISL_2645507, EPI_ISL_2645508, EPI_ISL_2645509, EPI_ISL_2645510, EPI_ISL_2645511, EPI_ISL_2645512, EPI_ISL_2645513, EPI_ISL_2863595, EPI_ISL_2863596, EPI_ISL_2863598, EPI_ISL_2863600, EPI_ISL_2983228, EPI_ISL_2983229, EPI_ISL_2983417, EPI_ISL_2983418, EPI_ISL_2983419, EPI_ISL_2983420, EPI_ISL_2983421, EPI_ISL_2983422, EPI_ISL_2983423, EPI_ISL_3190409, EPI_ISL_3190410, EPI_ISL_3190411 | | | |
| see above | Labortorio Central de Saude Publica do Estado do Tocantins (LACEN/TO) | Laboratory of Respiratory Viruses and Measles, Oswaldo Cruz Institute, FIOCRUZ | Agatha Soares; Alice Sampaio Rocha; Ana Carolina Mendonca; Anna Carolina Paixao; Elisa Cavalcante Pereira; Fernando Motta; Ighor Arantes; Jucimaria Dantas Galvao; Luciana Appolinario; Marilda Siqueira on behalf of the Fiocruz COVID-19 Genomic Surveillance Network; Paola Resende; Renata Serrano Lopes; Taina Venas |
| EPI_ISL_1358300, EPI_ISL_1358301, EPI_ISL_1358302, EPI_ISL_1358303 | Lacen de Tocantins | Instituto Adolfo Lutz, Interdiciplinary Procedures Center, Strategic Laboratory | Caio Vinicius Dias Lopes; Claudia Regina Gonçalves; Claudio Tavares Sacchi; Erica Valessa Ramos Gomes; Karoline Rodrigues Campos |
| EPI_ISL_3014467 | Unidade de apoio ao diagnostico da COVID - UNADIG | Bioinformatics Laboratory / LNCC | Alessandra P Lamarca; Alexandra L Gerber; Amilcar Tanuri; Ana Paula de C Guimaraes; Ana Tereza R Vasconcelos; Andrea Cony Cavalcanti; Caio Luiz Pereira Ribeiro; Cassia Alves; Cintia Policarpo; Claudia Maria Braga de Mello; Cristiane Gomes da Silva; Diana Mariani; Douglas Terra Machado; Flavio Dias da Silva; Gleidson da Silva de Oliveira; Leandro Magalhaes de Souza; Liliane Cavalcante; Luiz G P de Almeida; Marcio Henrique de Oliveira Garcia; Mario Sergio Ribeiro; Ronaldo da Silva F Jr; Silvia Carvalho |
| EPI_ISL_1272236 | Universidade Federal do Norte do Tocantins (UFNT) | Laboratório de Bioinformática e Biotecnologia (Labinftec/UFT) | Bergmann Morais Ribeiro; Fabrício Souza Campos; Fernando Lucas Melo; José Carlos Ribeiro Júnior; Monike da Silva Oliveira; Raíssa Nunes dos Santos; Rogério Fernandes Carvalho; Ueric José Borges de Souza |

We gratefully acknowledge the following Authors from the Originating laboratories responsible for obtaining the specimens, as well as the Submitting laboratories where the genome data were generated and shared via GISAID, on which this research is based.

All Submitters of data may be contacted directly via www.gisaid.org

Authors are sorted alphabetically.

| Accession ID | Originating Laboratory | Submitting Laboratory | Authors |
|---|---|---|---|
| EPI_ISL_1499114, EPI_ISL_1499205, EPI_ISL_1499298, EPI_ISL_1499299, EPI_ISL_1499300 | Associação Fundo de Incentivo à Pesquisa (AFIP) | Associação Fundo de Incentivo à Pesquisa (AFIP) | Debora R. Ramadan; Erika Rodrigues de Oliveira; Juliana Nogueira Martins Rodrigues; Priscila Farias Tempaku; Sergio Tufik.; Soraya Sgambatti de Andrade |
| EPI_ISL_3663810, EPI_ISL_3663939, EPI_ISL_3663955, EPI_ISL_3663957, EPI_ISL_3663959, EPI_ISL_3663968, EPI_ISL_3663969, EPI_ISL_3663977, EPI_ISL_3663978, EPI_ISL_3663980, EPI_ISL_3663982, EPI_ISL_3663991, EPI_ISL_3663997, EPI_ISL_3664007, EPI_ISL_3664027, EPI_ISL_3664028, EPI_ISL_3664030, EPI_ISL_3664048, EPI_ISL_3664049, EPI_ISL_3664058, EPI_ISL_3664077, EPI_ISL_3664084, EPI_ISL_3664087, EPI_ISL_3664097, EPI_ISL_3664106, EPI_ISL_3664107, EPI_ISL_3664108, EPI_ISL_3664109, EPI_ISL_3664110, EPI_ISL_3664111, EPI_ISL_3664113, EPI_ISL_3664114, EPI_ISL_3664132, EPI_ISL_3664133, EPI_ISL_3664134, EPI_ISL_3664135, EPI_ISL_3664136, EPI_ISL_3664137, EPI_ISL_3664138, EPI_ISL_3664139, EPI_ISL_3664140, EPI_ISL_3664141, EPI_ISL_3664142, EPI_ISL_3664143, EPI_ISL_3664144, EPI_ISL_3664148, EPI_ISL_3664149, EPI_ISL_3664150, EPI_ISL_3664151, EPI_ISL_3664154, EPI_ISL_3664155, EPI_ISL_3664158, EPI_ISL_3664159, EPI_ISL_3664161, EPI_ISL_3664222 | | | |
| see above | DASA | DASA | Adriano Bonaldi; Angelica Hristov; Annelise Lopes; Bianca Cota; Cristina Oliveira; Jose Levi; Lidia Yamamoto; Paulo Pierry; Rodrigo Guarischi; Rodrigo Salazar |
| EPI_ISL_476282 | DB Diagnósticos do Brasil | Instituto de Medicina Tropical da Univesidade de São Paulo | Camila Alves Maia da Silva; Darlan da Silva Candido; Erika Regina Manuli; Ester Sabino; Flavia Cristina da Silva Sales; Giula Magalhaes Ferreira; Jaqueline Goes de Jesus; Julien Theze; Mariana Severo Ramundo; Nuno Faria; Samples: Nelson Gaburo Jr; Sequencing: Ingra Morales Claro; Thais de Moura Coletti |
| EPI_ISL_450874 | Evandro Chagas Institute | Evandro Chagas Institute | A.M.; Barbagelata; E.C.; E.M.A.; Ferreira; G.M.R; J.A.; Junior; L.C.; L.S.; M.C.; Martins; P.S.; Santos; Silva; Sousa; Sousa Junior; Viana; W.D.C.; da Silva |
| EPI_ISL_1063789 | Evandro Chagas Institute | Evandro Chagas Institute Virology | A.M.; Barbagelata; E.C.; E.M.A.; Ferreira; J.A.; Junior; K.C.; L.C.; L.S.; M.C.; P.S.; Pinheiro; Santos; Silva; Sousa; Sousa Junior; W.D.C.; da Silva |
| EPI_ISL_2758675, EPI_ISL_2758676, EPI_ISL_2758692, EPI_ISL_2758698, EPI_ISL_2758699, EPI_ISL_2758700, EPI_ISL_2758702, EPI_ISL_2758703, EPI_ISL_2758704, EPI_ISL_2758706, EPI_ISL_2758707, EPI_ISL_2758708, EPI_ISL_2758709, EPI_ISL_2758710, EPI_ISL_2758711, EPI_ISL_2758712, EPI_ISL_2758767, EPI_ISL_2758768, EPI_ISL_2758769, EPI_ISL_2758770, EPI_ISL_2758772, EPI_ISL_2758773, EPI_ISL_2758774, EPI_ISL_2758775 | | | |
| see above | HUEM/IBMP | IPEC Guarapuava | NAPI-Genômica (Novos Arranjo de Pesquisa e Inovação em Genômica): Ademar Dantas da Cunha Júnior Adriano Ferrasa Adriano Mondini Aldo Przybysz Alessandra Lourenço Cecchini Armani Alex Sandro Jorge Alexandra Ivo de Medeiros Alexandre Maller Aline Cristina Batista Rodrigues Johann Ana Lucia Ferreira Ana Marisa Fusco Almeida Anderson Joel Martino Andrade André Luís Laforga Vanzela Andrea Duarte Doetzer Andrea Name Colado Simao Andressa Pereira de Souza Anelisa Ramão Angelica Beate Winter Boldt Anna Hermínia Castro Gomes de Amorim Anna Silvia Penteado Setti da Rocha Antonio Camilo da Silva Filho Antonio Stabelini Neto Arthur Hirata Bertachi Barbara Mendes Paz Chao Betty Cristiane Kuhn Bruno Ambrozio Galindo Bruno Ribeiro Cruz Camilla Reginatto De Pierri Carla Fredrichsen Moya Araujo Carla Fredrichsen Moya Araujo Carlos Alberto Oliveira de Biagi Junior Carlos Augusto Nassar Carlos Eduardo Buss Carlos Gilberto Carlotti Junior Carlos Henrique Schneider Carolina Panis Carolina Weigert Galvão Caroline de Jesus Coelho Donha Caroline Guisantes de Salvo Toni Caryna Eurich Mazur Catiuscie Cabreira da Silva Tortorella Celso F. D. Doliveira Cesar Luiz Boguszewski Christiane Pienna Soares Chung Man Chin Claudia Moro Cleverson Busso Cristiane Cominetti Daiane Priscila Simão-Silva Dalila Luciola Zanette Daniel de Paula Daniel de Paula Daniel Rech Daniela Fiori Gradia Daniela Pretti da Cunha Tirapelli Daniela Viganó Zanoti Jeronymo Daniele Ukan Danielle Malheiros Ferreira Danielle Venturini Deborah Catharine de Assis Leite Deivid Calebe de Souza Dennis Armando Bertolini Edenir Inez Pamero Edna Maria Vissoci Reiche Edson Roberto Arpini Miguel Eduardo José de Almeida Araújo Eliana Carolina Vespero Eliandro Reis Tavares Elza Kimura Grimshaw Emanuel Maltempi de Souza Emanuele Cristina Gustani Buss Émerson Carraro Emiliana Cristina Melo ENILze Maria de Souza Fonseca Ribeiro Enilze Maria de Souza Fonseca Ribeiro Erika Izumi Erika Seki Kioshima Cotica Evani Marques Pereira Fabio Negretti Fábio Rodrigues Ferreira Seiva Felipe Dunin dos Santos Felipe Tuon Fernanda Andreia Rosa Fernanda Cestaro Prado Cortez Fernanda Ivanski Fernanda Maris Peria Flavia Regina Oliveira de Barros Franciele Aní Caovilla Follador Franciele Mara Lucca Zanardo Bohm Francinete Ramos Campos Fulviana Silva Nishiyama GABRIEL RIBEIRO CORDEIRO Gabriela Datsch Bennemann Gisele Santos de Oliveira Glaucio Valdameri Glauco Akelinghton Freire Vitiello Glauco Vieira Miranda Glaura Scantamburlo ALves Fernandes Guilherme Ferreira Silveira Gustavo Bianchini Porfírio Gustavo Lenci Marques Hélito Volpato Hildebrando Masshiro Nagai Huei Diana Lee Ilce Mara de Syllos Cólus Iris Rabinovich Israel Gomy Jackson Kawakami Jacques Duílio Brancher Jaime Luís Lopes Rocha Jaqueline Carvalho de Oliveira Jean Henrique da Silva Rodrigues Jean Leandro dos Santos Jeane Eliete Laguila Visentainer João Paulo Bianchi Ximenez Joaquim Manoel da Silva Jociani Ascari Joel Donazzolo Jorge Luis Maria Ruiz Jose Knopfholz José Luis da Conceição Silva José Sebastião dos Santos Joseane Carla Schabarum Juliana Cheleski Wiggers Juliana Mara Serpeloni Juliana Morini Küpper Cardoso Perseguini Karen Brajão de Oliveira Karin Braun Prado Karine Aparecida de Lima Katiany Rizzieri Caleffi Ferracioli Katiuscia de Oliveira Francisco Gabriel Kelvinson Fernandes Viana Larissa Beatriz Cossalter Larissa Danielle Bahls Pinto Laurival Antonio Vilas Boas Léia Carolina Lucio Líbero Mezzadri Neto Ligia Carla Faccin Galhardi Lirane Elize Defante Ferreto Luciana Furlaneto Maia Luciana Oliveira de Fariña Luciana Reis Azevedo Alanis Luciane Regina Cavalli Lucy Megumi Yamauchi Lioni Luis Paulo Gomes Mascarenhas Luis Paulo Gomes Mascarenhas Luís Paulo Mascarenhas Lupe Furtado Alle Lyvia Regina Biagi Silva Bertachi Mara Antonia Ramos Costa Mara L. Cordeiro Marcela Maria Birolim Marcelo Ricardo Vicari Marcia Edilaine Lopes Consolaro Marcia Holsbach Beltrame Marcia Regina Eches Perugini Marcos Abdo Arbex Marcos Pileggi MARCOS TADEU GRZELCZAK Marcus Peikriszwili Tartaruga Maria Angelica Ehara Watanabe Maria Antonia Ramos Costa Maria Claudia Gross Maria José Soares Mendes Giannini Maria Leandra Terencio Maria Lúcia Bonfleur Maria Luiza Guimarães de Oliveira Maria Luiza Petzl-Erler Mariana Abe Vicente Cavagnari Marina Kimiko Kadowaki Marise Fonseca dos Santos Marla Karine Amarante Maurício Turkiewicz Mauro Antonio Alves Castro Michel Rodrigo Zambrano Passarini Michele Potrich Michelle Orane Schemberger Milena Massumi Kozonoe Mônica Degraf Cavallin Monica Tereza Suldofski Mucio Luiz de Assis Cirino Nadia Graciele Krohn Najeh Maissar Khalil Nédia de Castilhos Ghisi Neide Tomimura Costa Neiva Leite Neyva Maria Lopes Romeiro Patrícia Amâncio da Rosa Patricia Dayane Carvalho Schaker Patricia Oehlmeyer Nassar Patrícia Savio de Araújo-Souza Patricia Silva Lucio Paulo Henrique Couto Souza Paulo Roberto Donadio Percy Nohama Quirino Alves de Lima Neto Rafael Deminice Rafael dos Santos Bezerra Raquel Alves dos Santos Renan Manozzo Galante Renata Ernlund Freitas de Macedo Rita de Cássia Garcia Simão Roberta Losi Guembarovski Roberto H. Herai Roberto Rosati Rodrigo Ferreira Rodrigo Rodrigues Matiello Rogério Neri Shinsato Rogério Pincela Mateus Rosane Aparecida Ribeiro Rosilene Fressatti Cardoso Rosilene Fressatti Cardoso Sandra Mara Guse Scós Venske Selene Elifio Esposito Sérgio Ossamu Ioshii Silvana Giuliatti Silvia Mara de Souza Halick Silvio Henrique Maia de Almeida Simone Neumann Wendt Spencer Luiz Marques Payão Stefan Wolanski Negrão Stephane Janaina de Moura Escobar Sueli Fumie Yamada Ogatta SUELI PERCIO QUINAIA Taciane Finatto Tatiana Mayumi Veiga Iriyoda Tayza Katelline Danilau Ostroski Tony Alexander Hild Valeria Valente Vanessa Nascimento Kozak Vanessa Santos Sotomaior Victor Breno Pedrosa Victoria Zeghbi Cochenski Borba Vivian Rotuno Moure Valdameri Wander Rogerio Pavanelli Weber Cláudio Francisco Nunes da Silva Willian Augusto de Melo Yohandra Reyes Torres |
| EPI_ISL_2758771, EPI_ISL_2758797, EPI_ISL_2758798 | HUEM/LACEN | IPEC Guarapuava | NAPI-Genômica (Novos Arranjo de Pesquisa e Inovação em Genômica): Ademar Dantas da Cunha Júnior Adriano Ferrasa Adriano Mondini Aldo Przybysz Alessandra Lourenço Cecchini Armani Alex Sandro Jorge Alexandra Ivo de Medeiros Alexandre Maller Aline Cristina Batista Rodrigues Johann Ana Lucia Ferreira Ana Marisa Fusco Almeida Anderson Joel Martino Andrade André Luís Laforga Vanzela Andrea Duarte Doetzer Andrea Name Colado Simao Andressa Pereira de Souza Anelisa Ramão Angelica Beate Winter Boldt Anna Hermínia Castro Gomes de Amorim Anna Silvia Penteado Setti da Rocha Antonio Camilo da Silva Filho Antonio Stabelini Neto Arthur Hirata Bertachi Barbara Mendes Paz Chao Betty Cristiane Kuhn Bruno Ambrozio Galindo Bruno Ribeiro Cruz Camilla Reginatto De Pierri Carla Fredrichsen Moya Araujo Carla Fredrichsen Moya Araujo Carlos Alberto Oliveira de Biagi Junior Carlos Augusto Nassar Carlos Eduardo Buss Carlos Gilberto Carlotti Junior Carlos Henrique Schneider Carolina Panis Carolina Weigert Galvão Caroline de Jesus Coelho Donha Caroline Guisantes de Salvo Toni Caryna Eurich Mazur Catiuscie Cabreira da Silva Tortorella Celso F. D. Doliveira Cesar Luiz Boguszewski Christiane Pienna Soares Chung Man Chin Claudia Moro Cleverson Busso Cristiane Cominetti Daiane Priscila Simão-Silva Dalila Luciola Zanette Daniel de Paula Daniel de Paula Daniel Rech Daniela Fiori Gradia Daniela Pretti da Cunha Tirapelli Daniela Viganó Zanoti Jeronymo Daniele Ukan Danielle Malheiros Ferreira Danielle Venturini Deborah Catharine de Assis Leite Deivid Calebe de Souza Dennis Armando Bertolini Edenir Inez Pamero Edna Maria Vissoci Reiche Edson Roberto Arpini Miguel Eduardo José de Almeida Araújo Eliana Carolina Vespero Eliandro Reis Tavares Elza Kimura Grimshaw Emanuel Maltempi de Souza Emanuele Cristina Gustani Buss Émerson Carraro Emiliana Cristina Melo ENILze Maria de Souza Fonseca Ribeiro Enilze Maria de Souza Fonseca Ribeiro Erika Izumi Erika Seki Kioshima Cotica Evani Marques Pereira Fabio Negretti Fábio Rodrigues Ferreira Seiva Felipe Dunin dos Santos Felipe Tuon Fernanda Andreia Rosa Fernanda Cestaro Prado Cortez Fernanda Ivanski Fernanda Maris Peria Flavia Regina Oliveira de Barros Franciele Aní Caovilla Follador Franciele Mara Lucca Zanardo Bohm Francinete Ramos Campos Fulviana Silva Nishiyama GABRIEL RIBEIRO CORDEIRO Gabriela Datsch Bennemann Gisele Santos de Oliveira Glaucio Valdameri Glauco Akelinghton Freire Vitiello Glauco Vieira Miranda Glaura Scantamburlo ALves Fernandes Guilherme Ferreira Silveira Gustavo Bianchini Porfírio Gustavo Lenci Marques Hélito Volpato Hildebrando Masshiro Nagai Huei Diana Lee Ilce Mara de Syllos Cólus Iris Rabinovich Israel Gomy Jackson Kawakami Jacques Duílio Brancher Jaime Luís Lopes Rocha Jaqueline Carvalho de Oliveira Jean Henrique da Silva Rodrigues Jean Leandro dos Santos Jeane Eliete Laguila Visentainer João Paulo Bianchi Ximenez Joaquim Manoel da Silva Jociani Ascari Joel Donazzolo Jorge Luis Maria Ruiz Jose Knopfholz José Luis da Conceição Silva José Sebastião dos Santos Joseane Carla Schabarum Juliana Cheleski Wiggers Juliana Mara Serpeloni Juliana Morini Küpper Cardoso Perseguini Karen Brajão de Oliveira Karin Braun Prado Karine Aparecida de Lima Katiany Rizzieri Caleffi Ferracioli Katiuscia de Oliveira Francisco Gabriel Kelvinson Fernandes Viana Larissa Beatriz Cossalter Larissa Danielle Bahls Pinto Laurival Antonio Vilas Boas Léia Carolina Lucio Líbero Mezzadri Neto Ligia Carla Faccin Galhardi Lirane Elize Defante Ferreto Luciana Furlaneto Maia Luciana Oliveira de Fariña Luciana Reis Azevedo Alanis Luciane Regina Cavalli Lucy Megumi Yamauchi Lioni Luis Paulo Gomes Mascarenhas Luis Paulo Gomes Mascarenhas Luís Paulo Mascarenhas Lupe Furtado Alle Lyvia Regina Biagi Silva Bertachi Mara Antonia Ramos Costa Mara L. Cordeiro Marcela Maria Birolim Marcelo Ricardo Vicari Marcia Edilaine Lopes Consolaro Marcia Holsbach Beltrame Marcia Regina Eches Perugini Marcos Abdo Arbex Marcos Pileggi MARCOS TADEU GRZELCZAK Marcus Peikriszwili Tartaruga Maria Angelica Ehara Watanabe Maria Antonia Ramos Costa Maria Claudia Gross Maria José Soares Mendes Giannini Maria Leandra Terencio Maria Lúcia Bonfleur Maria Luiza Guimarães de Oliveira Maria Luiza Petzl-Erler Mariana Abe Vicente Cavagnari Marina Kimiko Kadowaki Marise Fonseca dos Santos Marla Karine Amarante Maurício Turkiewicz Mauro Antonio Alves Castro Michel Rodrigo Zambrano Passarini Michele Potrich Michelle Orane Schemberger Milena Massumi Kozonoe Mônica Degraf Cavallin Monica Tereza Suldofski Mucio Luiz de Assis Cirino Nadia Graciele Krohn Najeh Maissar Khalil Nédia de Castilhos Ghisi Neide Tomimura Costa Neiva Leite Neyva Maria Lopes Romeiro Patrícia Amâncio da Rosa Patricia Dayane Carvalho Schaker Patricia Oehlmeyer Nassar Patrícia Savio de Araújo-Souza Patricia Silva Lucio Paulo Henrique Couto Souza Paulo Roberto Donadio Percy Nohama Quirino Alves de Lima Neto Rafael Deminice Rafael dos Santos Bezerra Raquel Alves dos Santos Renan Manozzo Galante Renata Ernlund Freitas de Macedo Rita de Cássia Garcia Simão Roberta Losi Guembarovski Roberto H. Herai Roberto Rosati Rodrigo Ferreira Rodrigo Rodrigues Matiello Rogério Neri Shinsato Rogério Pincela Mateus Rosane Aparecida Ribeiro Rosilene Fressatti Cardoso Rosilene Fressatti Cardoso Sandra Mara Guse Scós Venske Selene Elifio Esposito Sérgio Ossamu Ioshii Silvana Giuliatti Silvia Mara de Souza Halick Silvio Henrique Maia de Almeida Simone Neumann Wendt Spencer Luiz Marques Payão Stefan Wolanski Negrão Stephane Janaina de Moura Escobar Sueli Fumie Yamada Ogatta SUELI PERCIO QUINAIA Taciane Finatto Tatiana Mayumi Veiga Iriyoda Tayza Katelline Danilau Ostroski Tony Alexander Hild Valeria Valente Vanessa Nascimento Kozak Vanessa Santos Sotomaior Victor Breno Pedrosa Victoria Zeghbi Cochenski Borba Vivian Rotuno Moure Valdameri Wander Rogerio Pavanelli Weber Cláudio Francisco Nunes da Silva Willian Augusto de Melo Yohandra Reyes Torres |
| EPI_ISL_2758668, EPI_ISL_2758669, EPI_ISL_2758670, EPI_ISL_2758671, EPI_ISL_2758672, EPI_ISL_2758806 | IPEC Guarapuava | IPEC Guarapuava | NAPI-Genômica (Novos Arranjo de Pesquisa e Inovação em Genômica): Ademar Dantas da Cunha Júnior Adriano Ferrasa Adriano Mondini Aldo Przybysz Alessandra Lourenço Cecchini Armani Alex Sandro Jorge Alexandra Ivo de Medeiros Alexandre Maller Aline Cristina Batista Rodrigues Johann Ana Lucia Ferreira Ana Marisa Fusco Almeida Anderson Joel Martino Andrade André Luís Laforga Vanzela Andrea Duarte Doetzer Andrea Name Colado Simao Andressa Pereira de Souza Anelisa Ramão Angelica Beate Winter Boldt Anna Hermínia Castro Gomes de Amorim Anna Silvia Penteado Setti da Rocha Antonio Camilo da Silva Filho Antonio Stabelini Neto Arthur Hirata Bertachi Barbara Mendes Paz Chao Betty Cristiane Kuhn Bruno Ambrozio Galindo Bruno Ribeiro Cruz Camilla Reginatto De Pierri Carla Fredrichsen Moya Araujo Carla Fredrichsen Moya Araujo Carlos Alberto Oliveira de Biagi Junior Carlos Augusto Nassar Carlos Eduardo Buss Carlos Gilberto Carlotti Junior Carlos Henrique Schneider Carolina Panis Carolina Weigert Galvão Caroline de Jesus Coelho Donha Caroline Guisantes de Salvo Toni Caryna Eurich Mazur Catiuscie Cabreira da Silva Tortorella Celso F. D. Doliveira Cesar Luiz Boguszewski Christiane Pienna Soares Chung Man Chin Claudia Moro Cleverson Busso Cristiane Cominetti Daiane Priscila Simão-Silva Dalila Luciola Zanette Daniel de Paula Daniel de Paula Daniel Rech Daniela Fiori Gradia Daniela Pretti da Cunha Tirapelli Daniela Viganó Zanoti Jeronymo Daniele Ukan Danielle Malheiros Ferreira Danielle Venturini Deborah Catharine de Assis Leite Deivid Calebe de Souza Dennis Armando Bertolini Edenir Inez Pamero Edna Maria Vissoci Reiche Edson Roberto Arpini Miguel Eduardo José de Almeida Araújo Eliana Carolina Vespero Eliandro Reis Tavares Elza Kimura Grimshaw Emanuel Maltempi de Souza Emanuele Cristina Gustani Buss Émerson Carraro Emiliana Cristina Melo ENILze Maria de Souza Fonseca Ribeiro Enilze Maria de Souza Fonseca Ribeiro Erika Izumi Erika Seki Kioshima Cotica Evani Marques Pereira Fabio Negretti Fábio Rodrigues Ferreira Seiva Felipe Dunin dos Santos Felipe Tuon Fernanda Andreia Rosa Fernanda Cestaro Prado Cortez Fernanda Ivanski Fernanda Maris Peria Flavia Regina Oliveira de Barros Franciele Aní Caovilla Follador Franciele Mara Lucca Zanardo Bohm Francinete Ramos Campos Fulviana Silva Nishiyama GABRIEL RIBEIRO CORDEIRO Gabriela Datsch Bennemann Gisele Santos de Oliveira Glaucio Valdameri Glauco Akelinghton Freire Vitiello Glauco Vieira Miranda Glaura Scantamburlo ALves Fernandes Guilherme Ferreira Silveira Gustavo Bianchini Porfírio Gustavo Lenci Marques Hélito Volpato Hildebrando Masshiro Nagai Huei Diana Lee Ilce Mara de Syllos Cólus Iris Rabinovich Israel Gomy Jackson Kawakami Jacques Duílio Brancher Jaime Luís Lopes Rocha Jaqueline Carvalho de Oliveira Jean Henrique da Silva Rodrigues Jean Leandro dos Santos Jeane Eliete Laguila Visentainer João Paulo Bianchi Ximenez Joaquim Manoel da Silva Jociani Ascari Joel Donazzolo Jorge Luis Maria Ruiz Jose Knopfholz José Luis da Conceição Silva José Sebastião dos Santos Joseane Carla Schabarum Juliana Cheleski Wiggers Juliana Mara Serpeloni Juliana Morini Küpper Cardoso Perseguini Karen Brajão de Oliveira Karin Braun Prado Karine Aparecida de Lima Katiany Rizzieri Caleffi Ferracioli Katiuscia de Oliveira Francisco Gabriel Kelvinson Fernandes Viana Larissa Beatriz Cossalter Larissa Danielle Bahls Pinto Laurival Antonio Vilas Boas Léia Carolina Lucio Líbero Mezzadri Neto Ligia Carla Faccin Galhardi Lirane Elize Defante Ferreto Luciana Furlaneto Maia Luciana Oliveira de Fariña Luciana Reis Azevedo Alanis Luciane Regina Cavalli Lucy Megumi Yamauchi Lioni Luis Paulo Gomes Mascarenhas Luis Paulo Gomes Mascarenhas Luís Paulo Mascarenhas Lupe Furtado Alle Lyvia Regina Biagi Silva Bertachi Mara Antonia Ramos Costa Mara L. Cordeiro Marcela Maria Birolim Marcelo Ricardo Vicari Marcia Edilaine Lopes Consolaro Marcia Holsbach Beltrame Marcia Regina Eches Perugini Marcos Abdo Arbex Marcos Pileggi MARCOS TADEU GRZELCZAK Marcus Peikriszwili Tartaruga Maria Angelica Ehara Watanabe Maria Antonia Ramos Costa Maria Claudia Gross Maria José Soares Mendes Giannini Maria Leandra Terencio Maria Lúcia Bonfleur Maria Luiza Guimarães de Oliveira Maria Luiza Petzl-Erler Mariana Abe Vicente Cavagnari Marina Kimiko Kadowaki Marise Fonseca dos Santos Marla Karine Amarante Maurício Turkiewicz Mauro Antonio Alves Castro Michel Rodrigo Zambrano Passarini Michele Potrich Michelle Orane Schemberger Milena Massumi Kozonoe Mônica Degraf Cavallin Monica Tereza Suldofski Mucio Luiz de Assis Cirino Nadia Graciele Krohn Najeh Maissar Khalil Nédia de Castilhos Ghisi Neide Tomimura Costa Neiva Leite Neyva Maria Lopes Romeiro Patrícia Amâncio da Rosa Patricia Dayane Carvalho Schaker Patricia Oehlmeyer Nassar Patrícia Savio de Araújo-Souza Patricia Silva Lucio Paulo Henrique Couto Souza Paulo Roberto Donadio Percy Nohama Quirino Alves de Lima Neto Rafael Deminice Rafael dos Santos Bezerra Raquel Alves dos Santos Renan Manozzo Galante Renata Ernlund Freitas de Macedo Rita de Cássia Garcia Simão Roberta Losi Guembarovski Roberto H. Herai Roberto Rosati Rodrigo Ferreira Rodrigo Rodrigues Matiello Rogério Neri Shinsato Rogério Pincela Mateus Rosane Aparecida Ribeiro Rosilene Fressatti Cardoso Rosilene Fressatti Cardoso Sandra Mara Guse Scós Venske Selene Elifio Esposito Sérgio Ossamu Ioshii Silvana Giuliatti Silvia Mara de Souza Halick Silvio Henrique Maia de Almeida Simone Neumann Wendt Spencer Luiz Marques Payão Stefan Wolanski Negrão Stephane Janaina de Moura Escobar Sueli Fumie Yamada Ogatta SUELI PERCIO QUINAIA Taciane Finatto Tatiana Mayumi Veiga Iriyoda Tayza Katelline Danilau Ostroski Tony Alexander Hild Valeria Valente Vanessa Nascimento Kozak Vanessa Santos Sotomaior Victor Breno Pedrosa Victoria Zeghbi Cochenski Borba Vivian Rotuno Moure Valdameri Wander Rogerio Pavanelli Weber Cláudio Francisco Nunes da Silva Willian Augusto de Melo Yohandra Reyes Torres |
| EPI_ISL_2344420, EPI_ISL_2344421, EPI_ISL_2344425, EPI_ISL_2344426, EPI_ISL_2344428, EPI_ISL_2344431, EPI_ISL_2344440, EPI_ISL_2344450, EPI_ISL_2344451, EPI_ISL_2344455, EPI_ISL_2344456, EPI_ISL_2344457, EPI_ISL_2344458 | | | |
| see above | Instituto Butantan | Instituto de Medicina Tropical de Sao Paulo | Brazil-UK Centre for Arbovirus Discovery Diagnosis Genomics and Epidemiology (CADDE) Genomic Network - Instituto de Medicina Tropical |
| EPI_ISL_3418670, EPI_ISL_3418671, EPI_ISL_3418673, EPI_ISL_3418676, EPI_ISL_3418678, EPI_ISL_3418679, EPI_ISL_3418680, EPI_ISL_3418681, EPI_ISL_3418682, EPI_ISL_3418683 | | | |
| see above | Instituto Carlos Chagas - ICC, FIOCRUZ Parana | Instituto Carlos Chagas - ICC, FIOCRUZ Parana | A.A.; A.M.; A.R.; Aguiar; Albrecht, L.; Alves; Avila; Balsanelli, E.; Becker, G.; Blanes, L.; Dallagiovanna, B.; Debur; E.M.; F.K.; F.O.; Faoro, H.; Graef, T.; H.G.; I.N.; L.G.; L.R.; M.M.; M.O.; Marchini; Md.C.; Morello; Nardeli; Oliveira; P.C.; Passetti, F.; Pedrosa; Resende; Riediger; S.C.; Schemberger; Suzukawa; V.A.; Zanette, D.; de Baura; de Souza; dos Santos |
| EPI_ISL_2758987, EPI_ISL_2758988, EPI_ISL_2758992, EPI_ISL_2758994, EPI_ISL_2758995, EPI_ISL_2758997, EPI_ISL_2759001, EPI_ISL_2759002, EPI_ISL_2759004, EPI_ISL_2759006, EPI_ISL_2759008, EPI_ISL_2759011, EPI_ISL_2759012, EPI_ISL_2759016, EPI_ISL_2759017, EPI_ISL_2759020, EPI_ISL_2759021, EPI_ISL_2759022, EPI_ISL_2759024, EPI_ISL_2759025, EPI_ISL_2759026, EPI_ISL_2759028, EPI_ISL_2759030, EPI_ISL_2759033, EPI_ISL_2759035, EPI_ISL_2759037, EPI_ISL_2759040, EPI_ISL_2759041, EPI_ISL_2759045, EPI_ISL_2759046, EPI_ISL_2759047, EPI_ISL_2759050, EPI_ISL_2759051, EPI_ISL_2759052, EPI_ISL_2759054, EPI_ISL_2759056, EPI_ISL_2759057, EPI_ISL_2759059, EPI_ISL_2759061, EPI_ISL_2759063, EPI_ISL_2759065, EPI_ISL_2759066, EPI_ISL_2759068, EPI_ISL_2775419 | | | |
| see above | Instituto de Biologia Molecular do Paraná (LAC) | Instituto Carlos Chagas - Fiocruz | Alessandra De Melo Aguiar; Andreia Akemi Suzukawa; Andréa Rodrigues Ávila; Bruno Dallagiovanna; Dalila Zanette; Eduardo Balsanelli; Emanuel Maltempi de Souza; Fabio Passetti; Fabricio Klerynton Marchini; Fábio de Oliveira Pedrosa; Guilherme Becker; Helisson Faoro; Hellen Geremias dos Santos; Irina Nastassja Riediger; Letusa Albrecht; Lucas Blanes; Luis Gustavo Morello; Lysangela Ronalte Alves; Maria do Carmo Debur; Mauro de Medeiros Oliveira; Michelle Orane Schemberger; Paola Cristina Resende; Sheila Cristina Nardeli; Tiago Gräf; Valter Antônio de Baura |

EPI_ISL_918515, EPI_ISL_918516, EPI_ISL_918517, EPI_ISL_918522, EPI_ISL_918523, EPI_ISL_918524, EPI_ISL_918525, EPI_ISL_918526, EPI_ISL_918527, EPI_ISL_918528, EPI_ISL_918529, EPI_ISL_918530, EPI_ISL_918545, EPI_ISL_918546, EPI_ISL_918547, EPI_ISL_918548, EPI_ISL_918549, EPI_ISL_918550, EPI_ISL_918552, EPI_ISL_2488814, EPI_ISL_2488815, EPI_ISL_2488816, EPI_ISL_2488817, EPI_ISL_2488818, EPI_ISL_2488819

| | | | |
|---|---|---|---|
| see above | LACEN - Laboratório Central de Saúde Pública do Para | Evandro Chagas Institute | A.M.; Barbagelata; E.C.; E.M.A.; Ferreira; J.A.; Junior; K.C.; L.C.; L.S.; M.C.; P.S.; Pinheiro; Santos; Silva; Sousa; Sousa Junior; W.D.C.; da Silva |

EPI_ISL_1086377, EPI_ISL_1164989, EPI_ISL_1164990, EPI_ISL_1164991, EPI_ISL_1164992, EPI_ISL_1164994, EPI_ISL_1164999

| | | | |
|---|---|---|---|
| see above | LACEN - Laboratório Central de Saúde Pública do Paraiba | Evandro Chagas Institute | A.M.; Barbagelata; E.C.; E.M.A.; Ferreira; J.A.; Junior; K.C.; L.C.; L.S.; M.C.; P.S.; Pinheiro; Santos; Silva; Sousa; Sousa Junior; W.D.C.; da Silva |

EPI_ISL_904120, EPI_ISL_904121, EPI_ISL_1164972, EPI_ISL_1164974, EPI_ISL_1164975, EPI_ISL_1164978, EPI_ISL_1164983

| | | | |
|---|---|---|---|
| see above | LACEN - Laboratório Central de Saúde Pública do Pará | Evandro Chagas Institute | A.M.; Barbagelata; E.C.; E.M.A.; Ferreira; J.A.; Junior; K.C.; L.C.; L.S.; M.C.; P.S.; Pinheiro; Santos; Silva; Sousa; Sousa Junior; W.D.C.; da Silva |

EPI_ISL_3835195, EPI_ISL_3835196, EPI_ISL_3835197, EPI_ISL_3835198, EPI_ISL_3835199, EPI_ISL_3835200, EPI_ISL_3835201, EPI_ISL_3835202, EPI_ISL_3835204, EPI_ISL_3835205, EPI_ISL_3835209, EPI_ISL_3835210, EPI_ISL_3835211, EPI_ISL_3835212, EPI_ISL_3835213, EPI_ISL_3835214, EPI_ISL_3835215, EPI_ISL_3835216, EPI_ISL_3835217, EPI_ISL_3835218, EPI_ISL_3835219, EPI_ISL_3835220, EPI_ISL_3835222, EPI_ISL_3835224, EPI_ISL_3835225, EPI_ISL_3835228, EPI_ISL_3835229, EPI_ISL_3835230, EPI_ISL_3835231, EPI_ISL_3835232, EPI_ISL_3835233, EPI_ISL_3835234, EPI_ISL_3835235, EPI_ISL_3835237, EPI_ISL_3835238, EPI_ISL_3835239

| | | | |
|---|---|---|---|
| see above | LACEN/PB | WallauLab on behalf of Fiocruz COVID-19 Genomic Surveillance Network | Alexandre Freitas da Silva; Cassia Docena; Constância Flávia Junqueira Ayres; Filipe Zimmer Dezordi; Gabriel Luz Wallau; Gustavo Barbosa de Lima; Lais Ceschini Machado; Lilian Caroliny Amorim Silva; Maira Galdino da Rocha Pitta; Marcelo Henrique dos Santos Paiva; Matheus Filgueira Bezerra; Michelly Cristiny Pereira; Rômulo Pessoa e Silva; Sinval Pinto Brandão Filho |

EPI_ISL_1213160, EPI_ISL_1213161, EPI_ISL_1213189, EPI_ISL_1213201, EPI_ISL_1213207, EPI_ISL_1213213, EPI_ISL_1213336, EPI_ISL_1213343, EPI_ISL_1213344, EPI_ISL_1213367, EPI_ISL_1213369, EPI_ISL_1213370, EPI_ISL_1213388, EPI_ISL_1213392, EPI_ISL_1213404, EPI_ISL_1213406, EPI_ISL_1213427, EPI_ISL_1213429, EPI_ISL_1213431, EPI_ISL_1213433, EPI_ISL_1213434, EPI_ISL_1213436, EPI_ISL_1213438, EPI_ISL_1213439, EPI_ISL_1213441, EPI_ISL_1213443, EPI_ISL_1213444, EPI_ISL_1213446, EPI_ISL_1213448, EPI_ISL_1213449, EPI_ISL_1213451, EPI_ISL_1213453, EPI_ISL_1213454, EPI_ISL_1213456, EPI_ISL_1213458, EPI_ISL_1213459, EPI_ISL_1213461, EPI_ISL_1213463

| | | | |
|---|---|---|---|
| see above | LBM/UFPB | Bioinformatics Laboratory / LNCC | Alessandra P Lamarca; Alexandra L Gerber; Ana Paula Melo Mariano; Ana Paula de C Guimarães; Ana Tereza R Vasconcelos; Angela Maria Guimarães Santos; Bianca Mendes Maciel; Danielle Angst Secco; Eduardo Sérgio Soares Sousa; Eloiza Helena Campana; Francisco Paulo Freire Neto; George Rego Albuquerque; Kátia Castanho Scortecci; Lucymara Fassarella Agnez Lima; Luiz G P de Almeida; Luís Cristóvão Porto; Otavio J. Brustolini; Paulo Ricardo Nascimento; Ronaldo da Silva Francisco Jr; Sandra Rocha Gadelha; Selma Maria Bezerra Jeronimo; Vinícius Pietta Perez |

EPI_ISL_2196243, EPI_ISL_2196281, EPI_ISL_2196282, EPI_ISL_2196283, EPI_ISL_2196284, EPI_ISL_2196285, EPI_ISL_2196286, EPI_ISL_2196287, EPI_ISL_2196288, EPI_ISL_2196289, EPI_ISL_2196290, EPI_ISL_2196291, EPI_ISL_2196292, EPI_ISL_2196293, EPI_ISL_2196294, EPI_ISL_2196295, EPI_ISL_2196296, EPI_ISL_2196297, EPI_ISL_2196298, EPI_ISL_2196299, EPI_ISL_2196300, EPI_ISL_2196301, EPI_ISL_2196302, EPI_ISL_2196303, EPI_ISL_2196304, EPI_ISL_2196305, EPI_ISL_2443557, EPI_ISL_2443558, EPI_ISL_2443559, EPI_ISL_2443560, EPI_ISL_2443561, EPI_ISL_2443562, EPI_ISL_2443563, EPI_ISL_2443564, EPI_ISL_2645421, EPI_ISL_2645422, EPI_ISL_2645423, EPI_ISL_2645424, EPI_ISL_2645425, EPI_ISL_2645426, EPI_ISL_2645427, EPI_ISL_2645428, EPI_ISL_2645429, EPI_ISL_2645430, EPI_ISL_2645431, EPI_ISL_2645432, EPI_ISL_2645433, EPI_ISL_2645434, EPI_ISL_2645435, EPI_ISL_2645436, EPI_ISL_2645437, EPI_ISL_2645438, EPI_ISL_2645439, EPI_ISL_2645819, EPI_ISL_2645820, EPI_ISL_2645821, EPI_ISL_2645822, EPI_ISL_2645823, EPI_ISL_2645824, EPI_ISL_2645825, EPI_ISL_2645826, EPI_ISL_2645827, EPI_ISL_2645828, EPI_ISL_2645829, EPI_ISL_2645830, EPI_ISL_2645831, EPI_ISL_2645832, EPI_ISL_2645833, EPI_ISL_2645834, EPI_ISL_2645835, EPI_ISL_2645836, EPI_ISL_2645837, EPI_ISL_2645838, EPI_ISL_2645839, EPI_ISL_2645840, EPI_ISL_2645841, EPI_ISL_2645842, EPI_ISL_2645843, EPI_ISL_2645844, EPI_ISL_2645845, EPI_ISL_2645846, EPI_ISL_2645847, EPI_ISL_2645848, EPI_ISL_2645849, EPI_ISL_2645850, EPI_ISL_2645851, EPI_ISL_2645852, EPI_ISL_2645853, EPI_ISL_2645854, EPI_ISL_2645855, EPI_ISL_2645856, EPI_ISL_2645857, EPI_ISL_2645858, EPI_ISL_2645859, EPI_ISL_2645860, EPI_ISL_2645861, EPI_ISL_2645862, EPI_ISL_2645863, EPI_ISL_2645864, EPI_ISL_2645865, EPI_ISL_2645866, EPI_ISL_2645867, EPI_ISL_2645868, EPI_ISL_2645869, EPI_ISL_2645870, EPI_ISL_2645871, EPI_ISL_2645872, EPI_ISL_2645873, EPI_ISL_2645874, EPI_ISL_2645875, EPI_ISL_2645876, EPI_ISL_2645877, EPI_ISL_2645878, EPI_ISL_2645879, EPI_ISL_2645880, EPI_ISL_2645882, EPI_ISL_2645883, EPI_ISL_2645884, EPI_ISL_2645886, EPI_ISL_2645887, EPI_ISL_2645888, EPI_ISL_2645889, EPI_ISL_2645890, EPI_ISL_2645891, EPI_ISL_2645892, EPI_ISL_2645893, EPI_ISL_2645894, EPI_ISL_2645895, EPI_ISL_2645896, EPI_ISL_2645897, EPI_ISL_2645898, EPI_ISL_2645899, EPI_ISL_2645900, EPI_ISL_2645901, EPI_ISL_2645902, EPI_ISL_2645903, EPI_ISL_2645904, EPI_ISL_2645905, EPI_ISL_2645906, EPI_ISL_2645907, EPI_ISL_2645908, EPI_ISL_2645909, EPI_ISL_2645910, EPI_ISL_2645911, EPI_ISL_2645912, EPI_ISL_2645913, EPI_ISL_2645914, EPI_ISL_2645915, EPI_ISL_2645916, EPI_ISL_2645917, EPI_ISL_2645918, EPI_ISL_2645919, EPI_ISL_2645920, EPI_ISL_2645921, EPI_ISL_2645922, EPI_ISL_2645923, EPI_ISL_2645924, EPI_ISL_2645925, EPI_ISL_2645926, EPI_ISL_2645927, EPI_ISL_2645928, EPI_ISL_2645929, EPI_ISL_2645930, EPI_ISL_2645931, EPI_ISL_2645932, EPI_ISL_2731453, EPI_ISL_2731454, EPI_ISL_2731455, EPI_ISL_2731456, EPI_ISL_2863578, EPI_ISL_2863580, EPI_ISL_2863582, EPI_ISL_2863706, EPI_ISL_2863707, EPI_ISL_2863709, EPI_ISL_2863711, EPI_ISL_2863801, EPI_ISL_2863802, EPI_ISL_2863803, EPI_ISL_2863804, EPI_ISL_2863805, EPI_ISL_2863806, EPI_ISL_2863807, EPI_ISL_2863808, EPI_ISL_2863809, EPI_ISL_2863810, EPI_ISL_2863811, EPI_ISL_2863812, EPI_ISL_2863813, EPI_ISL_2863814, EPI_ISL_2863815, EPI_ISL_2863816, EPI_ISL_2863817, EPI_ISL_2863818, EPI_ISL_2863819, EPI_ISL_2863820, EPI_ISL_2863821, EPI_ISL_2863822, EPI_ISL_2863823, EPI_ISL_2863824, EPI_ISL_2863825, EPI_ISL_2863826, EPI_ISL_2863827, EPI_ISL_2863828, EPI_ISL_2863829, EPI_ISL_2863830, EPI_ISL_2863831, EPI_ISL_2863832, EPI_ISL_2863833, EPI_ISL_2863834, EPI_ISL_2863835, EPI_ISL_2863836, EPI_ISL_2863837, EPI_ISL_2863838, EPI_ISL_2863839, EPI_ISL_2863840, EPI_ISL_2863841, EPI_ISL_2863842, EPI_ISL_2863843, EPI_ISL_2863844, EPI_ISL_2863845, EPI_ISL_2863846, EPI_ISL_2863847, EPI_ISL_2863848, EPI_ISL_2863849, EPI_ISL_2863850, EPI_ISL_2863851, EPI_ISL_2863852, EPI_ISL_2863853, EPI_ISL_2863854, EPI_ISL_2863894, EPI_ISL_3539238, EPI_ISL_3539239, EPI_ISL_3539240, EPI_ISL_3801916, EPI_ISL_3802965, EPI_ISL_3802966, EPI_ISL_4061287, EPI_ISL_4061289, EPI_ISL_4061290, EPI_ISL_4061291, EPI_ISL_4061293, EPI_ISL_4061295, EPI_ISL_4080766, EPI_ISL_4080767, EPI_ISL_4080768, EPI_ISL_4080769, EPI_ISL_4080770

| | | | |
|---|---|---|---|
| see above | Laboratorio Central de Saude Publica do Estado do Para (LACEN/PA) | Laboratory of Respiratory Viruses and Measles, Oswaldo Cruz Institute, FIOCRUZ | Agatha Cristinne Prudencio; Agatha Cristinne Prudencio Soares; Agatha Soares; Alice Sampaio Rocha; Ana Carolina Mendonca; Anna Carolina Paixao; Elisa Cavalcante Pereira; Fernando Motta; Ighor Arantes; Ighor Leonardo Arantes Gomes; Luciana Appolinario; Marilda Siqueira on behalf of the Fiocruz COVID-19 Genomic Surveillance Network; Paola Resende; Renata Serrano Lopes; Taina Venas; Valnete Andrade |
| EPI_ISL_2535017 | Laboratorio Central Noel Nutels | Bioinformatics Laboratory / LNCC | Alessandra P Lamarca; Alexandra L Gerber; Amilcar Tanuri; Ana Paula de C Guimaraes; Ana Tereza R Vasconcelos; Andrea Cony Cavalcanti; Caio Luiz Pereira Ribeiro; Cassia Alves; Cintia Policarpo; Claudia Maria Braga de Mello; Cristiane Gomes da Silva; Diana Mariani; Douglas Terra Machado; Flavio Dias da Silva; Gleidson da Silva de Oliveira; Leandro Magalhaes de Souza; Liliane Cavalcante; Luiz G P de Almeida; Marcio Henrique de Oliveira Garcia; Mario Sergio Ribeiro; Ronaldo da Silva F Jr; Silvia Carvalho |

EPI_ISL_1181416, EPI_ISL_1181422, EPI_ISL_2157379, EPI_ISL_2157380, EPI_ISL_2157383, EPI_ISL_2157394, EPI_ISL_2157395, EPI_ISL_2157401, EPI_ISL_2157425, EPI_ISL_2157550, EPI_ISL_2157578, EPI_ISL_2157581, EPI_ISL_2157582, EPI_ISL_2157589, EPI_ISL_2157591, EPI_ISL_2274092, EPI_ISL_2274093, EPI_ISL_2536303, EPI_ISL_2536304, EPI_ISL_2536305, EPI_ISL_2536306, EPI_ISL_2536307, EPI_ISL_2536308, EPI_ISL_2536309, EPI_ISL_2536310, EPI_ISL_2536311, EPI_ISL_2536312, EPI_ISL_2536313, EPI_ISL_2536314, EPI_ISL_2536315, EPI_ISL_2536316, EPI_ISL_2536317, EPI_ISL_2536318, EPI_ISL_2536319, EPI_ISL_2536320, EPI_ISL_2536321, EPI_ISL_2536322, EPI_ISL_2536323, EPI_ISL_2536324, EPI_ISL_2536325, EPI_ISL_2536326, EPI_ISL_2536327, EPI_ISL_2536328, EPI_ISL_2536329, EPI_ISL_2536330, EPI_ISL_2536331, EPI_ISL_2536332, EPI_ISL_2536333, EPI_ISL_2536334, EPI_ISL_2536335, EPI_ISL_2536336, EPI_ISL_2536337, EPI_ISL_2536338, EPI_ISL_2536339, EPI_ISL_2536340, EPI_ISL_2536341, EPI_ISL_2536342, EPI_ISL_2536343, EPI_ISL_2536344, EPI_ISL_2536345, EPI_ISL_2536346, EPI_ISL_2536347, EPI_ISL_2536348, EPI_ISL_2536349, EPI_ISL_2536350, EPI_ISL_2536351, EPI_ISL_2536352, EPI_ISL_2536353, EPI_ISL_2731457, EPI_ISL_3061880, EPI_ISL_3061881, EPI_ISL_3061882, EPI_ISL_3061883, EPI_ISL_3061884, EPI_ISL_3061885, EPI_ISL_3061886, EPI_ISL_3061887, EPI_ISL_3061888, EPI_ISL_3061889, EPI_ISL_3061890, EPI_ISL_3061891

| | | | |
|---|---|---|---|
| see above | Laboratorio Central de Saude Publica do Estado da Paraiba (LACEN-PB) | Laboratory of Respiratory Viruses and Measles, Oswaldo Cruz Institute, FIOCRUZ | Alice Sampaio Rocha; Ana Carolina Mendonca; Anna Carolina Paixao; Dalane Loudal Florentino Teixeira; Elisa Cavalcante Pereira; Fernando Motta; Irina Riediger; Joao Felipe Bezerra; Luciana Appolinario; Marilda Siqueira on behalf of the Fiocruz COVID-19 Genomic Surveillance Network; Paola Resende; Renata Serrano Lopes; Taina Venas |

EPI_ISL_3190174, EPI_ISL_3190175, EPI_ISL_3190176, EPI_ISL_3190178, EPI_ISL_3190179, EPI_ISL_3190180, EPI_ISL_3190181, EPI_ISL_3190182, EPI_ISL_3190183, EPI_ISL_3190184, EPI_ISL_3190185, EPI_ISL_3190186, EPI_ISL_3190187, EPI_ISL_3190188, EPI_ISL_3190189, EPI_ISL_3235238, EPI_ISL_3235239, EPI_ISL_3235240, EPI_ISL_3235241, EPI_ISL_3235242, EPI_ISL_3235243, EPI_ISL_3235244, EPI_ISL_3235245, EPI_ISL_3235246, EPI_ISL_3235247, EPI_ISL_3235248, EPI_ISL_3235249, EPI_ISL_3235250, EPI_ISL_3235251, EPI_ISL_3235252, EPI_ISL_3235253, EPI_ISL_3235254, EPI_ISL_3235255, EPI_ISL_3235256, EPI_ISL_3434726, EPI_ISL_3434759, EPI_ISL_3434760, EPI_ISL_3434761, EPI_ISL_3434762, EPI_ISL_3434763, EPI_ISL_3434764, EPI_ISL_3434765, EPI_ISL_3434766, EPI_ISL_3434767, EPI_ISL_3434768, EPI_ISL_3434769, EPI_ISL_3434770, EPI_ISL_3434771, EPI_ISL_3434772, EPI_ISL_3434773, EPI_ISL_3434774, EPI_ISL_3434775, EPI_ISL_3434776, EPI_ISL_3434777, EPI_ISL_3434778, EPI_ISL_3434779, EPI_ISL_3434780, EPI_ISL_3434781, EPI_ISL_3434782, EPI_ISL_3434783, EPI_ISL_3434784, EPI_ISL_3434785, EPI_ISL_3434786, EPI_ISL_3434787, EPI_ISL_3434788, EPI_ISL_3434789, EPI_ISL_3434802, EPI_ISL_3434905, EPI_ISL_3539936, EPI_ISL_3832350, EPI_ISL_3832351, EPI_ISL_3832352, EPI_ISL_3832353, EPI_ISL_3832354, EPI_ISL_3832355, EPI_ISL_3832356, EPI_ISL_3832357, EPI_ISL_3832358, EPI_ISL_3832359, EPI_ISL_3832360, EPI_ISL_3832361, EPI_ISL_3832362, EPI_ISL_3832363, EPI_ISL_3832364, EPI_ISL_3832365, EPI_ISL_3832394, EPI_ISL_3832397, EPI_ISL_4080930, EPI_ISL_4080936, EPI_ISL_4080937, EPI_ISL_4080938, EPI_ISL_4080939, EPI_ISL_4080940, EPI_ISL_4080941, EPI_ISL_4080942, EPI_ISL_4080943, EPI_ISL_4080944, EPI_ISL_4080945, EPI_ISL_4080946, EPI_ISL_4080947, EPI_ISL_4080948, EPI_ISL_4080949, EPI_ISL_4080950, EPI_ISL_4080951, EPI_ISL_4080952, EPI_ISL_4080953, EPI_ISL_4080954, EPI_ISL_4080955, EPI_ISL_4080956, EPI_ISL_4080957, EPI_ISL_4080958, EPI_ISL_4080959, EPI_ISL_4080960, EPI_ISL_4080961, EPI_ISL_4080962, EPI_ISL_4080963, EPI_ISL_4080964, EPI_ISL_4080965, EPI_ISL_4080966, EPI_ISL_4080967, EPI_ISL_4080968, EPI_ISL_4080969, EPI_ISL_4080970, EPI_ISL_4080971, EPI_ISL_4080972, EPI_ISL_4080973, EPI_ISL_4080974, EPI_ISL_4080975, EPI_ISL_4080976, EPI_ISL_4080977, EPI_ISL_4080978, EPI_ISL_4080979, EPI_ISL_4080980, EPI_ISL_4080981, EPI_ISL_4080982, EPI_ISL_4080983, EPI_ISL_4080984, EPI_ISL_4080985, EPI_ISL_4080986, EPI_ISL_4080987, EPI_ISL_4080988, EPI_ISL_4080989, EPI_ISL_4080990, EPI_ISL_4080991, EPI_ISL_4080992, EPI_ISL_4080993, EPI_ISL_4080994, EPI_ISL_4080995, EPI_ISL_4080996, EPI_ISL_4080997, EPI_ISL_4080998, EPI_ISL_4080999, EPI_ISL_4081000, EPI_ISL_4081001, EPI_ISL_4081002, EPI_ISL_4081003, EPI_ISL_4081004, EPI_ISL_4081005, EPI_ISL_4081006, EPI_ISL_4081007, EPI_ISL_4081008, EPI_ISL_4081009, EPI_ISL_4081010, EPI_ISL_4081011, EPI_ISL_4081012, EPI_ISL_4081013, EPI_ISL_4081014, EPI_ISL_4081015, EPI_ISL_4081016, EPI_ISL_4081017, EPI_ISL_4081018, EPI_ISL_4081019, EPI_ISL_4081020, EPI_ISL_4081021, EPI_ISL_4081022, EPI_ISL_4081023, EPI_ISL_4081024, EPI_ISL_4081025, EPI_ISL_4081026, EPI_ISL_4081027, EPI_ISL_4081069, EPI_ISL_4081070, EPI_ISL_4081071, EPI_ISL_4081072, EPI_ISL_4081073, EPI_ISL_4081074, EPI_ISL_4081075, EPI_ISL_4081076, EPI_ISL_4081077, EPI_ISL_4081078, EPI_ISL_4081079, EPI_ISL_4081080, EPI_ISL_4081081, EPI_ISL_4081082, EPI_ISL_4081083, EPI_ISL_4081084, EPI_ISL_4081085, EPI_ISL_4081086, EPI_ISL_4081087, EPI_ISL_4081088, EPI_ISL_4081089, EPI_ISL_4081091, EPI_ISL_4081092, EPI_ISL_4081093, EPI_ISL_4081094, EPI_ISL_4081095, EPI_ISL_4081097, EPI_ISL_4081098, EPI_ISL_4081099, EPI_ISL_4081100, EPI_ISL_4081101

| | | | |
|---|---|---|---|
| see above | Laboratorio Central de Saude Publica do Estado da Paraiba (LACEN/PB) | Laboratory of Respiratory Viruses and Measles, Oswaldo Cruz Institute, FIOCRUZ | Alice Sampaio Rocha; Ana Carolina Mendonca; Anna Carolina Paixao; Dalane Loudal Florentino Teixeira; Elisa Cavalcante Pereira; Fernando Motta; Joao Felipe Bezerra; Luciana Appolinario; Marilda Siqueira on behalf of the Fiocruz COVID-19 Genomic Surveillance Network; Paola Resende; Renata Serrano Lopes; Taina Venas |
| EPI_ISL_2536354 | Laboratorio Central de Saude Publica do Estado do Para (LACEN/PA) | Laboratory of Respiratory Viruses and Measles, Oswaldo Cruz Institute, FIOCRUZ | Alice Sampaio Rocha; Ana Carolina Mendonca; Anna Carolina Paixao; Elisa Cavalcante Pereira; Fernando Motta; Luciana Appolinario; Marilda Siqueira on behalf of the Fiocruz COVID-19 Genomic Surveillance Network; Paola Resende; Renata Serrano Lopes; Taina Venas; Valnete Andrade |

EPI_ISL_2758939, EPI_ISL_2758940, EPI_ISL_2758942, EPI_ISL_2758945, EPI_ISL_2758947, EPI_ISL_2758948, EPI_ISL_2758949, EPI_ISL_2758950, EPI_ISL_2758951, EPI_ISL_2758952, EPI_ISL_2758956, EPI_ISL_2758958, EPI_ISL_2758964, EPI_ISL_2758990, EPI_ISL_2758998, EPI_ISL_2759007, EPI_ISL_2759010, EPI_ISL_2759013, EPI_ISL_2759023, EPI_ISL_2759027, EPI_ISL_2759029, EPI_ISL_2759032, EPI_ISL_2759036, EPI_ISL_2759038, EPI_ISL_2759039, EPI_ISL_2759042, EPI_ISL_2759044, EPI_ISL_2759053, EPI_ISL_2759058, EPI_ISL_2759064, EPI_ISL_2759069, EPI_ISL_2759070, EPI_ISL_2759071, EPI_ISL_2775416, EPI_ISL_2775417, EPI_ISL_2775423, EPI_ISL_2775424, EPI_ISL_2775425, EPI_ISL_2775426, EPI_ISL_2775427, EPI_ISL_2775428, EPI_ISL_2775429, EPI_ISL_2775430, EPI_ISL_2775431, EPI_ISL_2775432, EPI_ISL_2775433, EPI_ISL_2775434, EPI_ISL_2775435, EPI_ISL_2775436, EPI_ISL_2775437, EPI_ISL_2775438, EPI_ISL_2775439, EPI_ISL_2775440, EPI_ISL_2775441, EPI_ISL_2775442, EPI_ISL_2775443, EPI_ISL_2775444, EPI_ISL_2775445, EPI_ISL_2775446, EPI_ISL_2775447, EPI_ISL_2775448, EPI_ISL_2775449, EPI_ISL_2775450, EPI_ISL_2775451, EPI_ISL_2775452, EPI_ISL_2775453, EPI_ISL_2775454, EPI_ISL_2775455, EPI_ISL_2775456, EPI_ISL_2775457, EPI_ISL_2775458, EPI_ISL_2775459, EPI_ISL_2775460, EPI_ISL_2775461, EPI_ISL_2775462, EPI_ISL_2775463, EPI_ISL_2775464, EPI_ISL_2775465, EPI_ISL_2775466, EPI_ISL_2775467, EPI_ISL_2775468, EPI_ISL_2775469, EPI_ISL_2775470, EPI_ISL_2775471, EPI_ISL_2775472, EPI_ISL_2775473, EPI_ISL_2775474, EPI_ISL_2775475, EPI_ISL_2775476, EPI_ISL_2775477, EPI_ISL_2775478, EPI_ISL_2775479, EPI_ISL_2775480, EPI_ISL_2775481, EPI_ISL_2775482, EPI_ISL_2775483, EPI_ISL_2775485, EPI_ISL_2775486, EPI_ISL_2775487, EPI_ISL_2775488, EPI_ISL_2775489, EPI_ISL_2775490, EPI_ISL_2775491, EPI_ISL_2775492, EPI_ISL_2775493, EPI_ISL_2775494, EPI_ISL_2775495, EPI_ISL_2775496, EPI_ISL_2775497, EPI_ISL_2775498, EPI_ISL_2775499

| | | | |
|---|---|---|---|
| see above | Laboratorio Central de Saude Publica do Estado do Parana (Instituto de | Instituto Carlos Chagas - Fiocruz | Alessandra De Melo Aguiar; Andreia Akemi Suzukawa; Andréa Rodrigues Ávila; Bruno Dallagiovanna; Dalila Zanette; Eduardo Balsanelli; Emanuel Maltempi de Souza; Fabio Passetti; Fabricio Klerynton Marchini; Fábio de Oliveira Pedrosa; Guilherme Becker; Helisson Faoro; Hellen Geremias dos Santos; Irina Nastassja Riediger; Letusa Albrecht; Lucas Blanes; Luis Gustavo Morello; Lysangela Ronalte Alves; Maria do Carmo Debur; Mauro de Medeiros Oliveira; Michelle Orane Schemberger; Paola Cristina Resende; Sheila Cristina Nardeli; Tiago Gräf; Valter Antônio de Baura |

| | | | |
|---|---|---|---|
| | Biologia Molecular do Paraná (LAC)EN-PR) | | |
| EPI_ISL_3913651, EPI_ISL_3913656, EPI_ISL_3913660, EPI_ISL_3913664, EPI_ISL_3913669, EPI_ISL_3913672, EPI_ISL_3913676, EPI_ISL_3913681, EPI_ISL_3913685, EPI_ISL_3913690, EPI_ISL_3913694, EPI_ISL_3913698, EPI_ISL_3913703, EPI_ISL_3913707, EPI_ISL_3913712, EPI_ISL_3913715, EPI_ISL_3913720, EPI_ISL_3913724, EPI_ISL_3913729, EPI_ISL_3913733, EPI_ISL_3913738, EPI_ISL_3913741, EPI_ISL_3913746, EPI_ISL_3913751, EPI_ISL_3913755, EPI_ISL_3913759, EPI_ISL_3913764, EPI_ISL_3913767, EPI_ISL_3913772, EPI_ISL_3913775, EPI_ISL_3913780, EPI_ISL_3913784, EPI_ISL_3913789, EPI_ISL_3913792, EPI_ISL_3913797, EPI_ISL_3913801, EPI_ISL_3913806, EPI_ISL_3913809, EPI_ISL_3913815, EPI_ISL_3913818, EPI_ISL_3913823, EPI_ISL_3913827, EPI_ISL_3913832, EPI_ISL_3913836, EPI_ISL_3913841, EPI_ISL_3913845, EPI_ISL_3913849, EPI_ISL_3913854, EPI_ISL_3913858, EPI_ISL_3913862, EPI_ISL_3913866, EPI_ISL_3913870, EPI_ISL_3913874, EPI_ISL_3913879, EPI_ISL_3913882, EPI_ISL_3913891, EPI_ISL_3913896, EPI_ISL_3913900, EPI_ISL_3913905, EPI_ISL_3913909, EPI_ISL_3913913, EPI_ISL_3913917, EPI_ISL_3913921, EPI_ISL_3913925, EPI_ISL_3913930, EPI_ISL_3913934, EPI_ISL_3913938, EPI_ISL_3913943, EPI_ISL_3913947, EPI_ISL_3913951, EPI_ISL_3913955, EPI_ISL_3913960, EPI_ISL_3913964, EPI_ISL_3913969, EPI_ISL_3913973, EPI_ISL_3913977, EPI_ISL_3913981, EPI_ISL_3913985, EPI_ISL_3913989, EPI_ISL_3913994, EPI_ISL_3913997, EPI_ISL_3914002, EPI_ISL_3914007, EPI_ISL_3914010, EPI_ISL_3914014, EPI_ISL_3914019, EPI_ISL_3914023, EPI_ISL_3914027, EPI_ISL_3914032, EPI_ISL_3914036, EPI_ISL_3914040, EPI_ISL_3914045, EPI_ISL_3914048, EPI_ISL_3914053, EPI_ISL_3914056, EPI_ISL_3914061, EPI_ISL_3914064, EPI_ISL_3914069, EPI_ISL_3914073, EPI_ISL_3914078, EPI_ISL_3914082, EPI_ISL_3914090, EPI_ISL_3914095, EPI_ISL_3914099, EPI_ISL_3914103, EPI_ISL_3914107, EPI_ISL_3914111, EPI_ISL_3914116, EPI_ISL_3914119, EPI_ISL_3914124, EPI_ISL_3914129, EPI_ISL_3914134, EPI_ISL_3914138, EPI_ISL_3914142, EPI_ISL_3914147, EPI_ISL_3914150, EPI_ISL_3914155, EPI_ISL_3914158, EPI_ISL_3914163, EPI_ISL_3914166, EPI_ISL_3914171, EPI_ISL_3914175, EPI_ISL_3914179, EPI_ISL_3914184, EPI_ISL_3914193, EPI_ISL_3914198, EPI_ISL_3914202, EPI_ISL_3914206, EPI_ISL_3914211, EPI_ISL_3914214, EPI_ISL_3914219, EPI_ISL_3914223, EPI_ISL_3914228, EPI_ISL_3914232, EPI_ISL_3914236, EPI_ISL_3914240, EPI_ISL_3914244, EPI_ISL_3914249, EPI_ISL_3914254, EPI_ISL_3914258, EPI_ISL_3914262, EPI_ISL_3914267, EPI_ISL_3914271, EPI_ISL_3914275, EPI_ISL_3914279, EPI_ISL_3914282, EPI_ISL_3914287, EPI_ISL_3914291, EPI_ISL_3914296, EPI_ISL_3914300, EPI_ISL_3914304, EPI_ISL_3914308, EPI_ISL_3914313, EPI_ISL_3914317, EPI_ISL_3914321, EPI_ISL_3914326, EPI_ISL_3914331, EPI_ISL_3914334, EPI_ISL_3914339, EPI_ISL_3914342, EPI_ISL_3914347, EPI_ISL_3914351, EPI_ISL_3914355, EPI_ISL_3914359, EPI_ISL_3914363, EPI_ISL_3914368, EPI_ISL_3914372, EPI_ISL_3914381, EPI_ISL_3914384, EPI_ISL_3914389, EPI_ISL_3914394, EPI_ISL_3914397, EPI_ISL_3914402, EPI_ISL_3914407, EPI_ISL_3914410, EPI_ISL_3914415, EPI_ISL_3914418, EPI_ISL_3914423, EPI_ISL_3914427, EPI_ISL_3914430, EPI_ISL_3914434 | | | |
| see above | Laboratorio Central de Saude Publica do Estado do Parana (Instituto de Biologia Molecular do Paraná (LACEN-PR) | Instituto Carlos Chagas - Fiocruz | Alessandra De Melo Aguiar; Andreia Akemi Suzukawa; Andréa Rodrigues Ávila; Bruno Dallagiovanna; Dalila Zanette; Eduardo Balsanelli; Emanuel Maltempi de Souza; Fabio Passetti; Fabricio Klerynton Marchini; Fábio de Oliveira Pedrosa; Guilherme Becker; Helisson Faoro; Hellen Geremias dos Santos; Irina Nastassja Riediger; Letusa Albrecht; Lucas Blanes; Luis Gustavo Morello; Lysangela Ronalte Alves; Maria do Carmo Debur; Mauro de Medeiros Oliveira; Michelle Orane Schemberger; Paola Cristina Resende; Sheila Cristina Nardeli; Tiago Gräf; Valter Antônio de Baura |
| EPI_ISL_1181390, EPI_ISL_1181392, EPI_ISL_1181394, EPI_ISL_1181474, EPI_ISL_1181498, EPI_ISL_1181517, EPI_ISL_1181526, EPI_ISL_1181527, EPI_ISL_1181528, EPI_ISL_1181530, EPI_ISL_1181533, EPI_ISL_1181534, EPI_ISL_1181536, EPI_ISL_1181537, EPI_ISL_1181538, EPI_ISL_1181539, EPI_ISL_1181540, EPI_ISL_1181541, EPI_ISL_1181542, EPI_ISL_1181543, EPI_ISL_1181544, EPI_ISL_1181545, EPI_ISL_1181551, EPI_ISL_1181552, EPI_ISL_1181553, EPI_ISL_1181554, EPI_ISL_1181555, EPI_ISL_1181556, EPI_ISL_1181557, EPI_ISL_1181558, EPI_ISL_1181559, EPI_ISL_1181563, EPI_ISL_1181564, EPI_ISL_1181565, EPI_ISL_1181566, EPI_ISL_1181568, EPI_ISL_1181570, EPI_ISL_1181576, EPI_ISL_1181580, EPI_ISL_1181581, EPI_ISL_1181584, EPI_ISL_1181605, EPI_ISL_1181609, EPI_ISL_1181610, EPI_ISL_1181614, EPI_ISL_1181615, EPI_ISL_1181616, EPI_ISL_1181617, EPI_ISL_1181618, EPI_ISL_1181619, EPI_ISL_1181623, EPI_ISL_1219132, EPI_ISL_1219133, EPI_ISL_1533978, EPI_ISL_1533999, EPI_ISL_1534000, EPI_ISL_1534001, EPI_ISL_2038955 | | | |
| see above | Laboratorio Central de Saude Publica do Estado do Parana (LACEN-PR) | Laboratory of Respiratory Viruses and Measles, Oswaldo Cruz Institute, FIOCRUZ | Alice Sampaio Rocha; Ana Carolina Mendonca; Anna Carolina Paixao; Elisa Cavalcante Pereira; Fernando Motta; Irina Nastassja Riediger; Irina Riediger; Luciana Appolinario; Maria do Carmo Debur; Marilda Siqueira on behalf of the Fiocruz COVID-19 Genomic Surveillance Network; Paola Resende; Renata Serrano Lopes; Taina Venas |
| EPI_ISL_2157482, EPI_ISL_2645445, EPI_ISL_2645446, EPI_ISL_2645447, EPI_ISL_2645448, EPI_ISL_2645449, EPI_ISL_2645450, EPI_ISL_2645451, EPI_ISL_2645452, EPI_ISL_2645453, EPI_ISL_2645454, EPI_ISL_2645455, EPI_ISL_2645456, EPI_ISL_2645457, EPI_ISL_2645458, EPI_ISL_2645459, EPI_ISL_2645460, EPI_ISL_2645461, EPI_ISL_2645462, EPI_ISL_2645463, EPI_ISL_2645464, EPI_ISL_2645465, EPI_ISL_2645466, EPI_ISL_2645467, EPI_ISL_2645468, EPI_ISL_2645469, EPI_ISL_2645470, EPI_ISL_2645471, EPI_ISL_2645472, EPI_ISL_2645473, EPI_ISL_2645474, EPI_ISL_2645475, EPI_ISL_2645476, EPI_ISL_2645477, EPI_ISL_2645478, EPI_ISL_2645479, EPI_ISL_2645480, EPI_ISL_2645481, EPI_ISL_2645482, EPI_ISL_2645483, EPI_ISL_2645484, EPI_ISL_2645485, EPI_ISL_2645486, EPI_ISL_2645487, EPI_ISL_2645491, EPI_ISL_2645492, EPI_ISL_2645493, EPI_ISL_2645494, EPI_ISL_2645495, EPI_ISL_2645496, EPI_ISL_2645497, EPI_ISL_2645498, EPI_ISL_2645499, EPI_ISL_2645500, EPI_ISL_2645501, EPI_ISL_2645502, EPI_ISL_2645503, EPI_ISL_2645504, EPI_ISL_2731458, EPI_ISL_2731459, EPI_ISL_2731461, EPI_ISL_2731466, EPI_ISL_2731467, EPI_ISL_2983373, EPI_ISL_2983426, EPI_ISL_2983427, EPI_ISL_2983428, EPI_ISL_2983430, EPI_ISL_2983431, EPI_ISL_2983432, EPI_ISL_2983433, EPI_ISL_2983434, EPI_ISL_2983435, EPI_ISL_2983437, EPI_ISL_2983438, EPI_ISL_2983439, EPI_ISL_2983440, EPI_ISL_2983441, EPI_ISL_2983442, EPI_ISL_2983443, EPI_ISL_2983444, EPI_ISL_2983445, EPI_ISL_2983446, EPI_ISL_2983447, EPI_ISL_2983448, EPI_ISL_2983449, EPI_ISL_2983450, EPI_ISL_2983451, EPI_ISL_2983452, EPI_ISL_3061892, EPI_ISL_3061893, EPI_ISL_3190190, EPI_ISL_3190191, EPI_ISL_3190192, EPI_ISL_3190264, EPI_ISL_3190266, EPI_ISL_3190267, EPI_ISL_3190268, EPI_ISL_3190360, EPI_ISL_3190361, EPI_ISL_3190362, EPI_ISL_3190363, EPI_ISL_3190364, EPI_ISL_3190365, EPI_ISL_3190366, EPI_ISL_3190367, EPI_ISL_3190368, EPI_ISL_3190369, EPI_ISL_3190370, EPI_ISL_3190371, EPI_ISL_3190372, EPI_ISL_3190373, EPI_ISL_3190374, EPI_ISL_3190375, EPI_ISL_3190376, EPI_ISL_3190377, EPI_ISL_3190378, EPI_ISL_3190379, EPI_ISL_3190380, EPI_ISL_3190381, EPI_ISL_3190382, EPI_ISL_3190383, EPI_ISL_3190384, EPI_ISL_3190385, EPI_ISL_3190386, EPI_ISL_3190387, EPI_ISL_3190388, EPI_ISL_3190389, EPI_ISL_3190390, EPI_ISL_3190391, EPI_ISL_3235257, EPI_ISL_3243095, EPI_ISL_3243096, EPI_ISL_3243097, EPI_ISL_3243098, EPI_ISL_3243099, EPI_ISL_3243100, EPI_ISL_3243101, EPI_ISL_3243102, EPI_ISL_3243103, EPI_ISL_3243104, EPI_ISL_3243105, EPI_ISL_3243106, EPI_ISL_3243107, EPI_ISL_3243108, EPI_ISL_3243109, EPI_ISL_3243110, EPI_ISL_3243111, EPI_ISL_3243112, EPI_ISL_3435048, EPI_ISL_3540001, EPI_ISL_3540002, EPI_ISL_3801799, EPI_ISL_3801800, EPI_ISL_3801801, EPI_ISL_3801802, EPI_ISL_3801823, EPI_ISL_3801824, EPI_ISL_3801825, EPI_ISL_3801839, EPI_ISL_3801840, EPI_ISL_3801841, EPI_ISL_3801842, EPI_ISL_3801843, EPI_ISL_3801844, EPI_ISL_3801852, EPI_ISL_3801853, EPI_ISL_3801854, EPI_ISL_3801855, EPI_ISL_3801856, EPI_ISL_3801857, EPI_ISL_3801858, EPI_ISL_3801859, EPI_ISL_3801860, EPI_ISL_3801861, EPI_ISL_3801862, EPI_ISL_3801863, EPI_ISL_3801864, EPI_ISL_3801865, EPI_ISL_3801866, EPI_ISL_3801867, EPI_ISL_3801868, EPI_ISL_3801869, EPI_ISL_3801870, EPI_ISL_3801875, EPI_ISL_3801920, EPI_ISL_3801921, EPI_ISL_3801922, EPI_ISL_3801923, EPI_ISL_3801925, EPI_ISL_3801926, EPI_ISL_3801927, EPI_ISL_3801928, EPI_ISL_3801929, EPI_ISL_3801930, EPI_ISL_3801931, EPI_ISL_3802976, EPI_ISL_3802977, EPI_ISL_3827988, EPI_ISL_3827989, EPI_ISL_3827992, EPI_ISL_3828024, EPI_ISL_3828025, EPI_ISL_3828026, EPI_ISL_3832366, EPI_ISL_3832367, EPI_ISL_3832368, EPI_ISL_3832369, EPI_ISL_3832370, EPI_ISL_3832371, EPI_ISL_3832372, EPI_ISL_3832373, EPI_ISL_3832374, EPI_ISL_3832375, EPI_ISL_3832376, EPI_ISL_3832377, EPI_ISL_3832378, EPI_ISL_3832379, EPI_ISL_3832380, EPI_ISL_3832381, EPI_ISL_3832382, EPI_ISL_3832393, EPI_ISL_4061328, EPI_ISL_4061329, EPI_ISL_4061330, EPI_ISL_4061424, EPI_ISL_4061425, EPI_ISL_4061426, EPI_ISL_4061427, EPI_ISL_4061428, EPI_ISL_4061429, EPI_ISL_4061430, EPI_ISL_4061431, EPI_ISL_4061432, EPI_ISL_4061433, EPI_ISL_4061437, EPI_ISL_4061438 | | | |
| see above | Laboratorio Central de Saude Publica do Estado do Parana (LACEN/PR) | Laboratory of Respiratory Viruses and Measles, Oswaldo Cruz Institute, FIOCRUZ | Agatha Soares; Alice Sampaio Rocha; Ana Carolina Mendonca; Anna Carolina Paixao; Elisa Cavalcante Pereira; Fernando Motta; Ighor Arantes; Ighor Leonardo Arantes; Irina Riediger; Luciana Appolinario; Marilda Siqueira on behalf of the Fiocruz COVID-19 Genomic Surveillance Network; Paola Resende; Renata Serrano Lopes; Taina Venas |
| EPI_ISL_3997110, EPI_ISL_3997227, EPI_ISL_3997228, EPI_ISL_3997230, EPI_ISL_3997240, EPI_ISL_3997739, EPI_ISL_3997833, EPI_ISL_3997877, EPI_ISL_3998018, EPI_ISL_3998051, EPI_ISL_3998169, EPI_ISL_3999620, EPI_ISL_4000067, EPI_ISL_4000069, EPI_ISL_4000229, EPI_ISL_4000270, EPI_ISL_4000384, EPI_ISL_4000695, EPI_ISL_4000717, EPI_ISL_4000718, EPI_ISL_4000719, EPI_ISL_4000801, EPI_ISL_4000802, EPI_ISL_4000803, EPI_ISL_4000804, EPI_ISL_4001402, EPI_ISL_4001403, EPI_ISL_4001404, EPI_ISL_4001405, EPI_ISL_4001562, EPI_ISL_4001708, EPI_ISL_4009398 | | | |
| see above | Laboratorio Central do Estado do Parana | Laboratorio Central do Estado do Parana | Guilherme Nardi Becker; Irina Nastassja Riediger; Maria do Carmo Debur Rossa; Mayra Presibella Giacomini; Nelson Quallio Marques |
| EPI_ISL_1402429, EPI_ISL_2731462, EPI_ISL_2731463, EPI_ISL_2731464, EPI_ISL_2731465 | Laboratory of Respiratory Viruses and Measles, Oswaldo Cruz Institute, FIOCRUZ | Laboratory of Respiratory Viruses and Measles, Oswaldo Cruz Institute, FIOCRUZ | Alex Pauvolid-Correa; Alice Sampaio Rocha; Ana Beatriz Machado Lima; Ana Carolina Mendonca; Anna Carolina Paixao; Elisa Cavalcante Pereira; Fernando Motta; Luciana Appolinario; Marilda Siqueira on behalf of the Fiocruz COVID-19 Genomic Surveillance Network; Mia Ferreira Araujo; Paola Resende; Renata Serrano Lopes; Taina Venas |
| EPI_ISL_2157506, EPI_ISL_2157516, EPI_ISL_2157517 | Laboratório Central de Saude Publica do Estado do Paraná (LACEN/PR) | Laboratory of Respiratory Viruses and Measles, Oswaldo Cruz Institute, FIOCRUZ | Alice Sampaio Rocha; Ana Carolina Mendonca; Anna Carolina Paixao; Elisa Cavalcante Pereira; Fernando Motta; Irina Nastassja Riediger; Luciana Appolinario; Maria do Carmo Debur; Marilda Siqueira on behalf of the Fiocruz COVID-19 Genomic Surveillance Network; Paola Resende; Renata Serrano Lopes; Taina Venas |
| EPI_ISL_2241495, EPI_ISL_2241496, EPI_ISL_2241497, EPI_ISL_2241498, EPI_ISL_2241499, EPI_ISL_2241514, EPI_ISL_2241516, EPI_ISL_2241517, EPI_ISL_2241518, EPI_ISL_2241519, EPI_ISL_2241520, EPI_ISL_2241522, EPI_ISL_2241523, EPI_ISL_2241525, EPI_ISL_2241526, EPI_ISL_2241527, EPI_ISL_2241529, EPI_ISL_2241556, EPI_ISL_2241558, EPI_ISL_2241559, EPI_ISL_2241561, EPI_ISL_2241562, EPI_ISL_2241563, EPI_ISL_2241564, EPI_ISL_2241566, EPI_ISL_2241567, EPI_ISL_2241568, EPI_ISL_2241569, EPI_ISL_2241570, EPI_ISL_2241571, EPI_ISL_2241572, EPI_ISL_2241573, EPI_ISL_2241576, EPI_ISL_2241581, EPI_ISL_2241583, EPI_ISL_2241585, EPI_ISL_2241586, EPI_ISL_2241588, EPI_ISL_2241594, EPI_ISL_2241596, EPI_ISL_2246285 | | | |
| see above | Laboratório Central de Saúde Pública da Paraiba | Coordenação Geral de Laboratórios de Saúde Pública (CGLAB/DAEVS/SVS/MS) | Vagner Fonseca; et al. |
| EPI_ISL_792561, EPI_ISL_792562, EPI_ISL_792563, EPI_ISL_792564, EPI_ISL_792565, EPI_ISL_792566, EPI_ISL_792567, EPI_ISL_792568, EPI_ISL_792569, EPI_ISL_792570, EPI_ISL_792571, EPI_ISL_792572, EPI_ISL_792573, EPI_ISL_792574, EPI_ISL_792575, EPI_ISL_792576, EPI_ISL_792577, EPI_ISL_792578, EPI_ISL_792579, EPI_ISL_792580, EPI_ISL_792581, EPI_ISL_792582, EPI_ISL_792583, EPI_ISL_792584, EPI_ISL_792585, EPI_ISL_792586, EPI_ISL_792587, EPI_ISL_792588, EPI_ISL_792589, EPI_ISL_792590, EPI_ISL_792591, EPI_ISL_792592, EPI_ISL_792593, EPI_ISL_792594, EPI_ISL_792595, EPI_ISL_792596, EPI_ISL_792597, EPI_ISL_792598, EPI_ISL_792599, EPI_ISL_792600, EPI_ISL_792601, EPI_ISL_792602, EPI_ISL_792603, EPI_ISL_792604, EPI_ISL_792605, EPI_ISL_792606, EPI_ISL_792607, EPI_ISL_792608, EPI_ISL_792609, EPI_ISL_792610, EPI_ISL_792611, EPI_ISL_792612, EPI_ISL_792613, EPI_ISL_792614, EPI_ISL_792615, EPI_ISL_792616, EPI_ISL_792617, EPI_ISL_792618, EPI_ISL_792619, EPI_ISL_792620, EPI_ISL_792621, EPI_ISL_792622, EPI_ISL_792623, EPI_ISL_792624, EPI_ISL_792625, EPI_ISL_792626, EPI_ISL_792627, EPI_ISL_792628, EPI_ISL_792629, EPI_ISL_792630, EPI_ISL_792631, EPI_ISL_792632, EPI_ISL_792633, EPI_ISL_792634, EPI_ISL_792635, EPI_ISL_792636, EPI_ISL_792637, EPI_ISL_792638 | | | |
| see above | Laboratório Central de Saúde Pública do Estado da Paraíba (LACEN-PB) | Laboratory of Respiratory Viruses and Measles, Oswaldo Cruz Institute, FIOCRUZ | Ana Carolina Mendonca; Anna Carolina Paixao; Dalane Loudal Florentino Teixeira; Fernando Motta; João Felipe Bezerra; Luciana Appolinario; Marilda Siqueira on behalf of the Fiocruz COVID-19 Genomic Surveillance Network; Paola Resende; Romero Henrique Teixeira de Vasconcelos; Thiago Franco de Oliveira Carneiro |
| EPI_ISL_541340, EPI_ISL_541341, EPI_ISL_541342, EPI_ISL_541343, EPI_ISL_541344, EPI_ISL_541345, EPI_ISL_541346, EPI_ISL_792645, EPI_ISL_792646, EPI_ISL_792647, EPI_ISL_792648, EPI_ISL_792649, EPI_ISL_792650, EPI_ISL_792651, EPI_ISL_792652, EPI_ISL_792653, EPI_ISL_792654 | | | |
| see above | Laboratório Central de Saúde Pública do Estado do Paraná (LACEN-PR) | Laboratory of Respiratory Viruses and Measles, Oswaldo Cruz Institute, FIOCRUZ | Ana Carolina Mendonca; Ana Carolina Mendonça; Anna Carolina Paixao; Anna Carolina Paixão; Fernando Motta; Irina Nastassja Riediger; Irina Riediger; Jonathan Lopes; Luciana Appolinario; Maria do Carmo Debur; Marilda Siqueira on behalf of the Fiocruz COVID-19 Genomic Surveillance Network; Paola Resende |
| EPI_ISL_2249446, EPI_ISL_2249447, EPI_ISL_2249448, EPI_ISL_2249449, EPI_ISL_2249450, EPI_ISL_2249451, EPI_ISL_2249452, EPI_ISL_2249453, EPI_ISL_2249454, EPI_ISL_2249455, EPI_ISL_2249456, EPI_ISL_2249457 | | | |
| see above | Laboratório | Coordenação Geral de | Vagner Fonseca; et al. |

| | | | |
|---|---|---|---|
| | Central de Saúde Pública do Paraá | Laboratórios de Saúde Pública (CGLAB/DAEVS/SVS/MS) | |
| EPI_ISL_2245087, EPI_ISL_2245089, EPI_ISL_2245090, EPI_ISL_2245092, EPI_ISL_2245093, EPI_ISL_2245094, EPI_ISL_2245095, EPI_ISL_2245098, EPI_ISL_2245101, EPI_ISL_2245102, EPI_ISL_2245103, EPI_ISL_2245104, EPI_ISL_2245105, EPI_ISL_2248758, EPI_ISL_2248759, EPI_ISL_2248760, EPI_ISL_2248761, EPI_ISL_2248762, EPI_ISL_2248763, EPI_ISL_2248764, EPI_ISL_2248765, EPI_ISL_2248766, EPI_ISL_2248767, EPI_ISL_2248782, EPI_ISL_2248783, EPI_ISL_2298758, EPI_ISL_2298783, EPI_ISL_2298787, EPI_ISL_2298810, EPI_ISL_2298812, EPI_ISL_2298814 | | | |
| see above | Laboratório Central de Saúde Pública do Pará | Coordenação Geral de Laboratórios de Saúde Pública (CGLAB/DAEVS/SVS/MS) | Vagner Fonseca; et al. |
| EPI_ISL_1182546, EPI_ISL_1182562, EPI_ISL_1182563, EPI_ISL_1182565, EPI_ISL_1182567, EPI_ISL_1182568, EPI_ISL_1182571, EPI_ISL_1182572, EPI_ISL_1182575, EPI_ISL_1182576, EPI_ISL_1182580, EPI_ISL_1182581, EPI_ISL_1182582, EPI_ISL_1182583, EPI_ISL_1182584, EPI_ISL_1182588, EPI_ISL_1182589, EPI_ISL_1182592, EPI_ISL_1182594, EPI_ISL_1182595, EPI_ISL_1182596, EPI_ISL_1182603, EPI_ISL_1182604, EPI_ISL_1182613, EPI_ISL_1182614, EPI_ISL_1182617, EPI_ISL_1182622, EPI_ISL_1182624 | | | |
| see above | Laboratório Central do Estado do Paraná | Coordenação Geral de Laboratórios de Saúde Pública (CGLAB/DAEVS/SVS/MS) | Vagner Fonseca; et al. |
| EPI_ISL_1133256 | Laboratório Hermes Pardini | Laboratório de Biologia Integrativa, Instituto de Ciências Biológicas, Universidade Federal de Minas Gerais | Alessandro Clayton de Souza Ferreira; Aline Brito de Lima; Carolina Moreira Voloch; Daniel Costa Queiroz; Danielle Alves Gomes Zauli; Diego Menezes Bonfim; Filipe Romero Rebello Moreira; Frederico Scott Varella Malta; Joice do Prado Silva; Rafael Marques de Souza; Renan Pedra de Souza; Renato Santana Aguiar.; Rennan Garcias Moreira; Victor Cavalcanti Pardini; Victor Emmanuel Viana Geddes; Wagner Carlos Santos Magalhaes; Walyson Coelho Costa |
| EPI_ISL_1261691 | Laboratório Paulo C. Azevedo | Evandro Chagas Institute | A.M.; Barbagelata; E.C.; E.M.A.; Ferreira; J.A.; Junior; K.C.; L.C.; L.S.; M.C.; P.S.; Pinheiro; Santos; Silva; Sousa; Sousa Junior; W.D.C.; da Silva |
| EPI_ISL_2157343, EPI_ISL_2157386, EPI_ISL_2157387, EPI_ISL_2157388, EPI_ISL_2157397, EPI_ISL_2157402, EPI_ISL_2157403, EPI_ISL_2157404, EPI_ISL_2157408, EPI_ISL_2157413, EPI_ISL_2157416, EPI_ISL_2157417, EPI_ISL_2157419, EPI_ISL_2157420, EPI_ISL_2157421, EPI_ISL_2157424, EPI_ISL_2157431, EPI_ISL_2157452, EPI_ISL_2157479, EPI_ISL_2157483, EPI_ISL_2157485, EPI_ISL_2157486, EPI_ISL_2157487, EPI_ISL_2157488, EPI_ISL_2157492, EPI_ISL_2157496, EPI_ISL_2157504, EPI_ISL_2157527, EPI_ISL_2157593, EPI_ISL_2157595, EPI_ISL_2157597, EPI_ISL_2196244, EPI_ISL_2196252, EPI_ISL_2196259, EPI_ISL_2196306, EPI_ISL_2196307, EPI_ISL_2196308, EPI_ISL_2196309, EPI_ISL_2196310, EPI_ISL_2196311, EPI_ISL_2196312, EPI_ISL_2196362, EPI_ISL_2274094, EPI_ISL_2274095, EPI_ISL_2274096, EPI_ISL_2443528, EPI_ISL_2443530, EPI_ISL_2443531, EPI_ISL_2443532, EPI_ISL_2443533, EPI_ISL_2443534, EPI_ISL_2443535, EPI_ISL_2443536, EPI_ISL_2443537, EPI_ISL_2443538, EPI_ISL_2443539, EPI_ISL_2443540, EPI_ISL_2443541, EPI_ISL_2443542, EPI_ISL_2443543, EPI_ISL_2443544, EPI_ISL_2443545, EPI_ISL_2443546, EPI_ISL_2443547, EPI_ISL_2443548, EPI_ISL_2443549, EPI_ISL_2443550, EPI_ISL_2443553, EPI_ISL_2443554, EPI_ISL_2443565, EPI_ISL_2443566, EPI_ISL_2443567, EPI_ISL_2443568, EPI_ISL_2443569, EPI_ISL_2443570, EPI_ISL_2443571, EPI_ISL_2443572, EPI_ISL_2443573, EPI_ISL_2443574, EPI_ISL_2443575, EPI_ISL_2443576, EPI_ISL_2443577, EPI_ISL_2557359, EPI_ISL_2557360, EPI_ISL_2557361, EPI_ISL_2557362, EPI_ISL_2557363, EPI_ISL_2557364, EPI_ISL_2557365, EPI_ISL_2557366, EPI_ISL_2557367, EPI_ISL_2557368, EPI_ISL_2557369, EPI_ISL_2557370, EPI_ISL_2557371, EPI_ISL_2557372, EPI_ISL_2557373, EPI_ISL_2557374, EPI_ISL_2557375, EPI_ISL_2603424, EPI_ISL_2603425, EPI_ISL_2603426, EPI_ISL_2603427, EPI_ISL_2603428, EPI_ISL_2603429, EPI_ISL_2603430, EPI_ISL_2603431, EPI_ISL_2603437, EPI_ISL_2603438, EPI_ISL_2603439, EPI_ISL_2603440, EPI_ISL_2603451, EPI_ISL_2603452, EPI_ISL_2603453, EPI_ISL_2603454, EPI_ISL_2603455, EPI_ISL_2603456, EPI_ISL_2603457, EPI_ISL_2603458, EPI_ISL_2603459, EPI_ISL_2603460, EPI_ISL_2603461, EPI_ISL_2603462, EPI_ISL_2603463, EPI_ISL_2603464, EPI_ISL_2603465, EPI_ISL_2603466, EPI_ISL_2603467, EPI_ISL_2603468, EPI_ISL_2603469, EPI_ISL_2603470, EPI_ISL_2603471, EPI_ISL_2603543, EPI_ISL_2603544, EPI_ISL_2603545, EPI_ISL_2603546, EPI_ISL_2613809, EPI_ISL_2613810, EPI_ISL_2613811, EPI_ISL_2613812, EPI_ISL_2613813, EPI_ISL_2613814, EPI_ISL_2613815, EPI_ISL_2613816, EPI_ISL_2613817, EPI_ISL_2613819, EPI_ISL_2613820, EPI_ISL_2613821, EPI_ISL_2613822, EPI_ISL_2613823, EPI_ISL_2613824, EPI_ISL_2645440, EPI_ISL_2645441, EPI_ISL_2645442, EPI_ISL_2645443, EPI_ISL_2645488, EPI_ISL_2645489, EPI_ISL_2645490, EPI_ISL_2863571, EPI_ISL_2863576, EPI_ISL_2863682, EPI_ISL_2863684, EPI_ISL_2863685, EPI_ISL_2863687, EPI_ISL_2863725, EPI_ISL_2863795, EPI_ISL_2863855, EPI_ISL_2863856, EPI_ISL_2863857, EPI_ISL_2863858, EPI_ISL_2863859, EPI_ISL_2863860, EPI_ISL_2863861, EPI_ISL_2863862, EPI_ISL_2863863, EPI_ISL_2863864, EPI_ISL_2863865, EPI_ISL_2863866, EPI_ISL_2863867, EPI_ISL_2863868, EPI_ISL_2863869, EPI_ISL_2863870, EPI_ISL_2863871, EPI_ISL_2863872, EPI_ISL_2863873, EPI_ISL_2863874, EPI_ISL_2863875, EPI_ISL_2863876, EPI_ISL_2863877, EPI_ISL_2863878, EPI_ISL_2863879, EPI_ISL_2863880, EPI_ISL_2863881, EPI_ISL_2863882, EPI_ISL_2863883, EPI_ISL_2863884, EPI_ISL_2863885, EPI_ISL_2863886, EPI_ISL_2863887, EPI_ISL_2863888, EPI_ISL_2863889, EPI_ISL_2863890, EPI_ISL_2863893, EPI_ISL_2863895, EPI_ISL_2863896, EPI_ISL_2863898, EPI_ISL_2863899 | | | |
| see above | Lboratorio Central de Saude Publica do Estado do Parana (LACEN/PR) | Laboratory of Respiratory Viruses and Measles, Oswaldo Cruz Institute, FIOCRUZ | Agatha Cristinne Prudencio; Agatha Cristinne Prudencio Soares; Alice Sampaio Rocha; Ana Carolina Mendonca; Anna Carolina Paixao; Elisa Cavalcante Pereira; Fernando Motta; Ighor Leonardo Arantes Gomes; Irina Riediger; Luciana Appolinario; Marilda Siqueira on behalf of the Fiocruz COVID-19 Genomic Surveillance Network; Paola Resende; Renata Serrano Lopes; Taina Venas |
| EPI_ISL_510535 | Molecular Virology, Instituto Carlos Chagas / Fiocruz Paraná | Universidade Federal do Parana (UFPR) | Duarte dos Santos, C.; Raboni, S.; Suzukawa, A.; Tscha, M.; Zanluca, C. |
| EPI_ISL_3982748, EPI_ISL_3982751, EPI_ISL_3982756, EPI_ISL_3982761, EPI_ISL_3982764, EPI_ISL_3982767, EPI_ISL_3982769, EPI_ISL_3982771 | | | |
| see above | Parana | LACEN PR | Guilherme Nardi Becker; Irina Nastassja Riediger; Maria do Carmo Debur Rossa; Mayra Presibella Giacomini; Nelson Quallio Marques |
| EPI_ISL_3982759, EPI_ISL_3982768 | Parana | Lacen-PR | Guilherme Nardi Becker; Irina Nastassja Riediger; Maria do Carmo Debur Rossa; Mayra Presibella Giacomini; Nelson Quallio Marques |
| EPI_ISL_3982749 | Parana | Lacen/PR | Guilherme Nardi Becker; Irina Nastassja Riediger; Maria do Carmo Debur Rossa; Mayra Presibella Giacomini; Nelson Quallio Marques |
| EPI_ISL_3982745, EPI_ISL_3982763 | Parana | Parana | Guilherme Nardi Becker; Irina Nastassja Riediger; Maria do Carmo Debur Rossa; Mayra Presibella Giacomini; Nelson Quallio Marques |
| EPI_ISL_2758648, EPI_ISL_2758649, EPI_ISL_2758650, EPI_ISL_2758651, EPI_ISL_2758652, EPI_ISL_2758653, EPI_ISL_2758654, EPI_ISL_2758655, EPI_ISL_2758656, EPI_ISL_2758657, EPI_ISL_2758658, EPI_ISL_2758659, EPI_ISL_2758660, EPI_ISL_2758661, EPI_ISL_2758662, EPI_ISL_2758663, EPI_ISL_2758664, EPI_ISL_2758665, EPI_ISL_2758666, EPI_ISL_2758667, EPI_ISL_2758673, EPI_ISL_2758674, EPI_ISL_2758677, EPI_ISL_2758678, EPI_ISL_2758679, EPI_ISL_2758680, EPI_ISL_2758681, EPI_ISL_2758682, EPI_ISL_2758683, EPI_ISL_2758684, EPI_ISL_2758685, EPI_ISL_2758686, EPI_ISL_2758687, EPI_ISL_2758688, EPI_ISL_2758689, EPI_ISL_2758690, EPI_ISL_2758691, EPI_ISL_2758693, EPI_ISL_2758694, EPI_ISL_2758695, EPI_ISL_2758696, EPI_ISL_2758697, EPI_ISL_2758701, EPI_ISL_2758705, EPI_ISL_2758713, EPI_ISL_2758714, EPI_ISL_2758715, EPI_ISL_2758716, EPI_ISL_2758717, EPI_ISL_2758718, EPI_ISL_2758719, EPI_ISL_2758720, EPI_ISL_2758721, EPI_ISL_2758722, EPI_ISL_2758723, EPI_ISL_2758724, EPI_ISL_2758725, EPI_ISL_2758726, EPI_ISL_2758727, EPI_ISL_2758728, EPI_ISL_2758729, EPI_ISL_2758730, EPI_ISL_2758731, EPI_ISL_2758732, EPI_ISL_2758733, EPI_ISL_2758734, EPI_ISL_2758735, EPI_ISL_2758736, EPI_ISL_2758737, EPI_ISL_2758738, EPI_ISL_2758739, EPI_ISL_2758740, EPI_ISL_2758741, EPI_ISL_2758742, EPI_ISL_2758743, EPI_ISL_2758744, EPI_ISL_2758745, EPI_ISL_2758746, EPI_ISL_2758747, EPI_ISL_2758748, EPI_ISL_2758749, EPI_ISL_2758750, EPI_ISL_2758751, EPI_ISL_2758752, EPI_ISL_2758753, EPI_ISL_2758754, EPI_ISL_2758755, EPI_ISL_2758756, EPI_ISL_2758757, EPI_ISL_2758758, EPI_ISL_2758759, EPI_ISL_2758760, EPI_ISL_2758761, EPI_ISL_2758762, EPI_ISL_2758763, EPI_ISL_2758764, EPI_ISL_2758765, EPI_ISL_2758766, EPI_ISL_2758776, EPI_ISL_2758777, EPI_ISL_2758778, EPI_ISL_2758779, EPI_ISL_2758780, EPI_ISL_2758781, EPI_ISL_2758782, EPI_ISL_2758783, EPI_ISL_2758784, EPI_ISL_2758785, EPI_ISL_2758786, EPI_ISL_2758787, EPI_ISL_2758788, EPI_ISL_2758789, EPI_ISL_2758790, EPI_ISL_2758791, EPI_ISL_2758792, EPI_ISL_2758793, EPI_ISL_2758794, EPI_ISL_2758795, EPI_ISL_2758796, EPI_ISL_2758799, EPI_ISL_2758801, EPI_ISL_2758802, EPI_ISL_2758803, EPI_ISL_2758804, EPI_ISL_2758805 | | | |
| see above | UEL | IPEC Guarapuava | NAPI-Genômica (Novos Arranjo de Pesquisa e Inovação em Genômica): Ademar Dantas da Cunha Júnior Adriano Ferrasa Adriano Mondini Aldo Przybysz Alessandra Lourenço Cecchini Armani Alex Sandro Jorge Alexandra Ivo de Medeiros Alexandre Maller Aline Cristina Batista Rodrigues Johann Ana Lucia Ferreira Ana Marisa Fusco Almeida Anderson Joel Martino Andrade André Luís Laforga Vanzela Andrea Duarte Doetzer Andrea Name Colado Simao Andressa Pereira de Souza Anelisa Ramão Angelica Beate Winter Boldt Anna Hermínia Castro Gomes de Amorim Anna Silvia Penteado Setti da Rocha Antonio Camilo da Silva Filho Antonio Stabelini Neto Arthur Hirata Bertachi Barbara Mendes Paz Chao Betty Cristiane Kuhn Bruno Ambrozio Galindo Bruno Ribeiro Cruz Camilla Reginatto De Pierri Carla Fredrichsen Moya Araujo Carla Fredrichsen Moya Araujo Carlos Alberto Oliveira de Biagi Junior Carlos Augusto Nassar Carlos Eduardo Buss Carlos Gilberto Carlotti Junior Carlos Henrique Schneider Carolina Panis Carolina Weigert Galvão Caroline de Jesus Coelho Donha Caroline Guisantes de Salvo Toni Caryna Eurich Mazur Catiuscie Cabreira da Silva Tortorella Celso F. D. Doliveira Cesar Luiz Boguszewski Christiane Pienna Soares Chung Man Chin Claudia Moro Cleverson Busso Cristiane Cominetti Daiane Priscila Simão-Silva Dalila Luciola Zanette Daniel de Paula Daniel de Paula Daniel Rech Daniela Fiori Gradia Daniela Pretti da Cunha Tirapelli Daniela Viganó Zanoti Jeronymo Daniele Ukan Danielle Malheiros Ferreira Danielle Venturini Deborah Catharine de Assis Leite Deivid Calebe de Souza Dennis Armando Bertolini Edenir Inez Pamero Edna Maria Vissoci Reiche Edson Roberto Arpini Miguel Eduardo José de Almeida Araújo Eliana Carolina Vespero Eliandro Reis Tavares Elza Kimura Grimshaw Emanuel Maltempi de Souza Emanuele Cristina Gustani Buss Émerson Carraro Emiliana Cristina Melo ENILze Maria de Souza Fonseca Ribeiro Enilze Maria de Souza Fonseca Ribeiro Erika Izumi Erika Seki Kioshima Cotica Evani Marques Pereira Fabio Negretti Fábio Rodrigues Ferreira Seiva Felipe Dunin dos Santos Felipe Tuon Fernanda Andreia Rosa Fernanda Cestaro Prado Cortez Fernanda Ivanski Fernanda Maris Peria Flavia Regina Oliveira de Barros Franciele Aní Caovilla Follador Franciele Mara Lucca Zanardo Bohm Francinete Ramos Campos Fulviana Silva Nishiyama GABRIEL RIBEIRO CORDEIRO Gabriela Datsch Bennemann Gisele Santos de Oliveira Glaucio Valdameri Glauco Akelinghton Freire Vitiello Glauco Vieira Miranda Glaura Scantamburlo ALves Fernandes Guilherme Ferreira Silveira Gustavo Bianchini Porfírio Gustavo Lenci Marques Hélito Volpato Hildebrando Masshiro Nagai Huei Diana Lee Ilce Mara de Syllos Cólus Iris Rabinovich Israel Gomy Jackson Kawakami Jacques Duílio Brancher Jaime Luís Lopes Rocha Jaqueline Carvalho de Oliveira Jean Henrique da Silva Rodrigues Jean Leandro dos Santos Jeane Eliete Laguila Visentainer João Paulo Bianchi Ximenez Joaquim Manoel da Silva Jociani Ascari Joel Donazzolo Jorge Luis Maria Ruiz Jose Knopfholz José Luis da Conceição Silva José Sebastião dos Santos Joseane Carla Schabarum Juliana Cheleski Wiggers Juliana Mara Serpeloni Juliana Morini Küpper Cardoso Perseguini Karen Brajão de Oliveira Karin Braun Prado Karine Aparecida de Lima Katiany Rizzieri Caleffi Ferracioli Katiuscia de Oliveira Francisco Gabriel Kelvinson Fernandes Viana Larissa Beatriz Cossalter Larissa Danielle Bahls Pinto Laurival Antonio Vilas Boas Léia Carolina Lucio Líbero Mezzadri Neto Ligia Carla Faccin Galhardi Lirane Elize Defante Ferreto Luciana Furlaneto Maia Luciana Oliveira de Fariña Luciana Reis Azevedo Alanis Luciane Regina Cavalli Lucy Megumi Yamauchi Lioni Luis Paulo Gomes Mascarenhas Luis Paulo Gomes Mascarenhas Luís Paulo Mascarenhas Lupe Furtado Alle Lyvia Regina Biagi Silva Bertachi Mara Antonia Ramos Costa Mara L. Cordeiro Marcela Maria Birolim Marcelo Ricardo Vicari Marcia Edilaine Lopes Consolaro Marcia Holsbach Beltrame Marcia Regina Eches Perugini Marcos Abdo Arbex Marcos Pileggi MARCOS TADEU GRZELCZAK Marcus Peikriszwili Tartaruga Maria Angelica Ehara Watanabe Maria Antonia Ramos Costa Maria Claudia Gross Maria José Soares Mendes Giannini Maria Leandra Terencio Maria Lúcia Bonfleur Maria Luiza Guimarães de Oliveira Maria Luiza Petzl-Erler Mariana Abe Vicente Cavagnari Marina Kimiko Kadowaki Marise Fonseca dos Santos Marla Karine Amarante Maurício Turkiewicz Mauro Antonio Alves Castro Michel Rodrigo Zambrano Passarini Michele Potrich Michelle Orane Schemberger Milena Massumi Kozonoe Mônica Degraf Cavallin Monica Tereza Suldofski Mucio Luiz de Assis Cirino Nadia Graciele Krohn Najeh Maissar Khalil Nédia de Castilhos Ghisi Neide Tomimura Costa Neiva Leite Neyva Maria Lopes Romeiro Patrícia Amâncio da Rosa Patricia Dayane Carvalho Schaker Patricia Oehlmeyer Nassar Patrícia Savio de Araújo-Souza Patricia Silva Lucio Paulo Henrique Couto Souza Paulo Roberto Donadio Percy Nohama Quirino Alves de Lima Neto Rafael Deminice Rafael dos Santos Bezerra Raquel Alves dos Santos Renan Manozzo Galante Renata Ernlund Freitas de Macedo Rita de Cássia Garcia Simão Roberta Losi Guembarovski Roberto H. Herai Roberto Rosati Rodrigo Ferreira Rodrigo Rodrigues Matiello Rogério Neri Shinsato Rogério Pincela Mateus Rosane Aparecida Ribeiro Rosilene Fressatti Cardoso Rosilene Fressatti Cardoso Sandra Mara Guse Scós Venske Selene Elifio Esposito Sérgio Ossamu Ioshii Silvana Giuliatti Silvia Mara de Souza Halick Silvio Henrique Maia de Almeida Simone Neumann Wendt Spencer Luiz Marques Payão Stefan Wolanski Negrão Stephane Janaina de Moura Escobar Sueli Fumie Yamada Ogatta SUELI PERCIO QUINAIA Taciane Finatto Tatiana Mayumi Veiga Iriyoda Tayza Katelline Danilau Ostroski Tony Alexander Hild Valeria Valente Vanessa Nascimento Kozak Vanessa Santos Sotomaior Victor Breno Pedrosa Victoria Zeghbi Cochenski Borba Vivian Rotuno Moure Valdameri Wander Rogerio Pavanelli Weber Cláudio Francisco Nunes da Silva Willian Augusto de Melo Yohandra Reyes Torres |
| EPI_ISL_3369438, EPI_ISL_3369439, EPI_ISL_3369440, EPI_ISL_3369441 | UFPA- Federal University of Pará | ITV-Vale Institute of Technology | Amanda Vidal; Guilherme Oliveira; Tatianne Costa Negri; Ândrea Kely Campos Ribeiro dos Santos |
| EPI_ISL_3072192 | Unidade de apoio ao diagnostico da COVID - UNADIG | Bioinformatics Laboratory / LNCC | Alessandra P Lamarca; Alexandra L Gerber; Amilcar Tanuri; Ana Paula de C Guimaraes; Ana Tereza R Vasconcelos; Andrea Cony Cavalcanti; Caio Luiz Pereira Ribeiro; Cassia Alves; Cintia Policarpo; Claudia Maria Braga de Mello; Cristiane Gomes da Silva; Diana Mariani; Douglas Terra Machado; Flavio Dias da Silva; Gleidson da Silva de Oliveira; Leandro Magalhaes de Souza; Liliane Cavalcante; Luiz G P de Almeida; Marcio Henrique de Oliveira Garcia; Mario Sergio Ribeiro; Ronaldo da Silva F Jr; Silvia Carvalho |
| EPI_ISL_458140, EPI_ISL_458141, EPI_ISL_458146, EPI_ISL_458147, EPI_ISL_458148, EPI_ISL_524783, EPI_ISL_524785, EPI_ISL_524786, EPI_ISL_524787, EPI_ISL_524800, EPI_ISL_524801, EPI_ISL_848555, EPI_ISL_848557, EPI_ISL_848561, EPI_ISL_848562, EPI_ISL_848563, EPI_ISL_848564, EPI_ISL_848565, EPI_ISL_848566, EPI_ISL_848567, EPI_ISL_848568, EPI_ISL_848569, EPI_ISL_848570, EPI_ISL_848571, EPI_ISL_848572, EPI_ISL_848582, EPI_ISL_848583, EPI_ISL_848585, EPI_ISL_848587, EPI_ISL_848588, EPI_ISL_848589, EPI_ISL_848590, EPI_ISL_848591, EPI_ISL_848592, EPI_ISL_848593, EPI_ISL_848594, EPI_ISL_848595, EPI_ISL_848596, EPI_ISL_848597, EPI_ISL_848598, EPI_ISL_848606, EPI_ISL_848607, EPI_ISL_848608, EPI_ISL_848609, EPI_ISL_848611, EPI_ISL_848614, EPI_ISL_848615, EPI_ISL_848617, EPI_ISL_848618, EPI_ISL_848619, EPI_ISL_848620, EPI_ISL_848621, EPI_ISL_848622, EPI_ISL_848623, EPI_ISL_848624, EPI_ISL_848625, EPI_ISL_848628, EPI_ISL_848629, EPI_ISL_918514, EPI_ISL_918518, EPI_ISL_918519, EPI_ISL_918520, EPI_ISL_918521, EPI_ISL_2488795, EPI_ISL_2488796, EPI_ISL_2488813 | | | |
| see above | Evandro Chagas Institute | Evandro Chagas Institute | A.M.; Barbagelata; E.C.; E.M.A.; Ferreira; G.M.R; H.R; J.A.; Junior; K.C.; L.C.; L.S.; M.C.; Martins; P.S.; Pinheiro; Resque; Santos; Silva; Sousa; Sousa Junior; Viana; W.D.C.; da Silva |

We gratefully acknowledge the following Authors from the Originating laboratories responsible for obtaining the specimens, as well as the Submitting laboratories where the genome data were generated and shared via GISAID, on which this research is based.

All Submitters of data may be contacted directly via www.gisaid.org

Authors are sorted alphabetically.

| Accession ID | Originating Laboratory | Submitting Laboratory | Authors |
|---|---|---|---|
| EPI_ISL_2645298, EPI_ISL_2645301, EPI_ISL_2645304, EPI_ISL_2645306, EPI_ISL_2645307 | "CO Dept. of Public Health and Environment, Lab Services Division" | Centers for Disease Control and Prevention Division of Viral Diseases, Pathogen Discovery | Alison Laufer Halpin; Ben L. Rambo-Martin; Clinton R. Paden; Dakota Howard; Darlene Wagner; Dave Wentworth; Dhwani Batra; Jasmine Padilla; Justin Lee; Katie Dillon; Krista Queen; Kristen Knipe; Kristine Lacek; Mark Burroughs; Matthew Schmerer; Mili Sheth; Peter Cook; Sam Shepard; Sarah Nobles; Shoshona Le; Suxiang Tong; Vivien Dugan; Yvette Unoarumhi |
| EPI_ISL_471546 | AMA DR Jose Soares Hungria | Instituto Adolfo Lutz, Interdiciplinary Procedures Center, Strategic Laboratory | Claudia Regina Gonçalves; Claudio Tavares Sacchi; Erica Valessa Ramos Gomes |
| EPI_ISL_523989 | AMA Jardim Joamar | Instituto Adolfo Lutz, Interdiciplinary Procedures Center, Strategic Laboratory | Claudia Regina Gonçalves; Claudio Tavares Sacchi; Erica Valessa Ramos Gomes |
| EPI_ISL_523990 | AMA Jardim Peri | Instituto Adolfo Lutz, Interdiciplinary Procedures Center, Strategic Laboratory | Claudia Regina Gonçalves; Claudio Tavares Sacchi; Erica Valessa Ramos Gomes |
| EPI_ISL_861652 | AMA Wamberto Dias da Costa | Instituto Adolfo Lutz, Interdiciplinary Procedures Center, Strategic Laboratory | Claudia Regina Gonçalves; Claudio Tavares Sacchi; Erica Valessa Ramos Gomes; Karoline Rodrigues Campos |
| EPI_ISL_1966699 | AMBULATORIO MEDICO DE ESPECIALIDADES DE PERUIBE | Instituto Butantan / Mendelics | Antonio Jorge Martins; Bianca Cechetto Carlos. Mendelics: Bibiana Santos; Claudia Renata dos Santos Barros; Cíntia Bittar; David Schlesinger. Hemocentro Ribeirão Preto: Simone Kashima; Debora Botequio Moretti; Elaine Cristina Marqueze; Elaine Vieira dos Santos; Elisangela Chicaroni Mattos; Erika Freitas; Evandra Strazza Rodrigues; Felipe Allan da Silva da Costa; Flavia Aburjaile; Fábio Sossai Possebon; Guilherme Campos; Guilherme Targino Valente; Heidge Fukumasu. USP-Botucatu: Rejane Maria Tommasini Grotto; Helena Lage Ferreira; Instituto Butantan: Dimas Tadeu Covas; Jardelina de Souza Todao Bernardino; Jayme A. Souza-Neto; Jessika Cristina Chagas Lesbon; Jorge A. Petroli Marchesi; José Salvatore Leister Patané; João Paulo Kitajima; João Pessoa Araújo Jr.; Leila Sabrina Ullmann; Loyze Paola Oliveira de Lima; Luiz Aurelio de Campos Crispin. Centro de Genômica Funcional da ESALQ: Luiz Lehmann Coutinho; Luiz Carlos Junior de Alcantara; Lívia Sacchetto; Maisa C. Pereira Parra; Maria Carolina Elias; Marta Giovanetti; Marília Moraes; Maurício Lacerda Nogueira. Prefeitura de Sao Paulo: Melissa Palmieri.; Patricia Akemi Assato; Paula Rahal; Paulo Inacio da Costa; Rafael dos Santos Bezerra; Raquel de Lello Rocha Campos Cassano. NGS Soluções Genômicas: Pilar Drummond Sampaio Corrêa Mariani. FZEA-USP Pirassununga: Mirele Daiana Poleti; Raul Machado Neto; Ricardo Augusto Brassaloti; Ricardo Haddad; Rodrigo Tocantins Calado. FAMERP-SJRP: Cecília Artico Banho; Sandra Coccuzzo Sampaio; Svetoslav Nanev Slavov; Vagner Fonseca; Vincent Louis Viala |
| EPI_ISL_2150721, EPI_ISL_2421940 | Aegis Sciences Corporation | Centers for Disease Control and Prevention Division of Viral Diseases, Pathogen Discovery | Adrian Paskey; Alec Vest; Benjamin Rambo-Martin; Christopher Gulvick; Clinton R. Paden; Cyndi Clark; Dakota Howard; Darlene Wagner; Dhwani Batra; Dillon Nall; Duncan MacCannell; Ethan Sanders; Holly Houdeshell; Jason Caravas; Kara Moser; Matthew Hardison; Matthew Schmerer; Ola Kvalvaag; Patrick Campbell; Peter W. Cook; Rob Case; Scott Sammons; Shatavia Morrison; Shaun Westlund; Vikramsinha Ghorpade; Yvette Unoarumhi |
| EPI_ISL_1201886, EPI_ISL_1219028, EPI_ISL_1219032 | Aeroporto Internacional de Guarulhos | Instituto Adolfo Lutz, Interdiciplinary Procedures Center, Strategic Laboratory | Caio Vinicius Dias Lopes; Claudia Regina Gonçalves; Claudio Tavares Sacchi; Erica Valessa Ramos Gomes; Karoline Rodrigues Campos |
| EPI_ISL_549084 | Akershus University Hospital, Department for Microbiology and Infectious Disease Control | Norwegian Institute of Public Health, Department of Virology | Hilde Elshaug; Hilde Synnøve Vollan; Kamilla Heddeland Instefjord; Karoline Bragstad; Kathrine Stene-Johansen; Olav Hungnes; Rasmus Riis Kopperud |
| EPI_ISL_515544, EPI_ISL_523984, EPI_ISL_523986 | Ama Dr Jose Soares Hungria | Instituto Adolfo Lutz, Interdiciplinary Procedures Center, Strategic Laboratory | Claudia Regina Gonçalves; Claudio Tavares Sacchi; Erica Valessa Ramos Gomes |
| EPI_ISL_2156551 | Amosup Seamen's Hospital | Philippine Genome Center | Alethea R. de Guzman; Anna Ong-Lim; Arianne A. Zamora; Asia Louisa U. Chong; Benedict A. Maralit; Candice Francheska B. Tambaoan; Carlo M. Lapid; Celia Carlos; Devon Ray Pacial; Edsel Maurice Salvaña; El King D. Morado; Elcid Aaron R. Pangilinan; Eva Maria Cutiongco-de la Paz; Francis A. Tablizo; Irish Coleen A. Asin; Jaime C. Montoya; Jan Michael C. Yap; Jo-Hannah S. Llames; John Q. Wong; Joshua Gregor A. Dizon; Juan Antonio R. Magalang; Karol Sophia Agape R. Padilla; Kenneth M. Kim; Kris P. Punayan; Marc Edsel C. Ayes; Marc Jerrone R. Castro; Maria Rosario Singh-Vergeire and Cynthia P. Saloma; Maria Sofia L. Yangzon; Marissa Alejandria; Razel Nikka M. Hao; Rianna Patricia S. Cruz; Sheila Mae M. Araiza |
| EPI_ISL_509500, EPI_ISL_527019, EPI_ISL_527032 | Area of Virology, Serology and Virology Division (SAViD), New South Wales Health Pathology Randwick | Area of Virology, Serology and Virology Division (SAViD), New South Wales Health Pathology Randwick | Rawlinson, W. |
| EPI_ISL_678320 | Area of Virology, Serology and Virology Division (SAViD), New South Wales Health Pathology Randwick | Virology Research Laboratory; Area of Virology, Serology and Virology Division (SAViD), New South Wales Health Pathology Randwick | Au, J.; Bull, R.; Deveson, I.; Foster, C.; Rawlinson, W.; Ruiz Silva, M.; Van Hal, S. |
| EPI_ISL_2105513, EPI_ISL_2105518, EPI_ISL_2105520 | Army General Hospital Molecular Laboratory | Philippine Genome Center | Alethea R. de Guzman; Anna Ong-Lim; Arianne A. Zamora; Asia Louisa U. Chong; Benedict A. Maralit; Candice Francheska B. Tambaoan; Carlo M. Lapid; Celia Carlos; Devon Ray Pacial; Edsel Maurice Salvaña; El King D. Morado; Eva Maria Cutiongco-de la Paz; Francis A. Tablizo; Irish Coleen A. Asin; Jaime C. Montoya; Jan Michael C. Yap; Jo-Hannah S. Llames; John Q. Wong; Joshua Gregor A. Dizon; Juan Antonio R. Magalang; Karol Sophia Agape R. Padilla; Kenneth M. Kim; Kris P. Punayan; Marc Edsel C. Ayes; Marc Jerrone R. Castro; Maria Rosario Singh-Vergeire and Cynthia P. Saloma; Maria Sofia L. Yangzon; Marissa Alejandria; Razel Nikka M. Hao; Rianna Patricia S. Cruz; Sheila Mae M. Araiza |
| EPI_ISL_1499020, EPI_ISL_1499297 | Associação Fundo de Incentivo à Pesquisa (AFIP) | Associação Fundo de Incentivo à Pesquisa (AFIP) | Debora R. Ramadan; Erika Rodrigues de Oliveira; Juliana Nogueira Martins Rodrigues; Priscila Farias Tempaku; Sergio Tufik.; Soraya Sgambatti de Andrade |
| EPI_ISL_2499718, EPI_ISL_2499719 | BCCDC Public Health Laboratory | BCCDC Public Health Laboratory | Ana Pacagnella; Corrinne Ng; Dan Fornika; John Tyson; Kim Macdonald; Kimia Kamelian; Linda Hoang; Loretta Janz; Mel Krajden; Prystajecky Natalie; Robert Azana; Shannon Russell |
| EPI_ISL_2156319 | BULACAN MEDICAL CENTER | Philippine Genome Center | Alethea R. de Guzman; Anna Ong-Lim; Arianne A. Zamora; Asia Louisa U. Chong; Benedict A. Maralit; Candice Francheska B. Tambaoan; Carlo M. Lapid; Celia Carlos; Devon Ray Pacial; Edsel Maurice Salvaña; El King D. Morado; Elcid Aaron R. Pangilinan; Eva Maria Cutiongco-de la Paz; Francis A. Tablizo; Irish Coleen A. Asin; Jaime C. Montoya; Jan Michael C. Yap; Jo-Hannah S. Llames; John Q. Wong; Joshua Gregor A. Dizon; Juan Antonio R. Magalang; Karol Sophia Agape R. Padilla; Kenneth M. Kim; Kris P. Punayan; Marc Edsel C. Ayes; Marc Jerrone R. Castro; Maria Rosario Singh-Vergeire and Cynthia P. Saloma; Maria Sofia L. Yangzon; Marissa Alejandria; Razel Nikka M. Hao; Rianna Patricia S. Cruz; Sheila Mae M. Araiza |
| EPI_ISL_2105421, EPI_ISL_2188197 | Baguio General Hospital Medical Center (BGHMC) | Philippine Genome Center | Alethea R. de Guzman; Anna Ong-Lim; Arianne A. Zamora; Asia Louisa U. Chong; Benedict A. Maralit; Candice Francheska B. Tambaoan; Carlo M. Lapid; Celia Carlos; Devon Ray Pacial; Edsel Maurice Salvaña; El King D. Morado; Elcid Aaron R. Pangilinan; Eva Maria Cutiongco-de la Paz; Francis A. Tablizo; Irish Coleen A. Asin; Jaime C. Montoya; Jan Michael C. Yap; Jo-Hannah S. Llames; John Q. Wong; Joshua Gregor A. Dizon; Juan Antonio R. Magalang; Karol Sophia Agape R. Padilla; Kenneth M. Kim; Kris P. Punayan; Marc Edsel C. Ayes; Marc Jerrone R. Castro; Maria Rosario Singh-Vergeire and Cynthia P. Saloma; Maria Sofia L. Yangzon; Marissa Alejandria; Razel Nikka M. Hao; Rianna Patricia S. Cruz; Sheila Mae M. Araiza |
| EPI_ISL_2156577 | Bataan General Hospital and Medical Center | Philippine Genome Center | Alethea R. de Guzman; Anna Ong-Lim; Arianne A. Zamora; Asia Louisa U. Chong; Benedict A. Maralit; Candice Francheska B. Tambaoan; Carlo M. Lapid; Celia Carlos; Devon Ray Pacial; Edsel Maurice Salvaña; El King D. Morado; Elcid Aaron R. Pangilinan; Eva Maria Cutiongco-de la Paz; Francis A. Tablizo; Irish Coleen A. Asin; Jaime C. Montoya; Jan Michael C. Yap; Jo-Hannah S. Llames; John Q. Wong; Joshua Gregor A. Dizon; Juan Antonio R. Magalang; Karol Sophia Agape R. Padilla; Kenneth M. Kim; Kris P. Punayan; Marc Edsel C. Ayes; Marc Jerrone R. Castro; Maria Rosario Singh-Vergeire and Cynthia P. Saloma; Maria Sofia L. Yangzon; Marissa Alejandria; Razel Nikka M. Hao; Rianna Patricia S. Cruz; Sheila Mae M. Araiza |
| EPI_ISL_2551528 | Belo Horizonte center-south emergency care unit - UPA-BH | Laboratório de Virologia Clínica e Molecular | Alex Fiorini; Ana Paula Salles Fernandes; Bruna Larotonda Telezynski; Danielle Bruna Leal Oliveira; Edison Luiz Durigon; Erick Gustavo Dorlass; Flavio Fonseca e Santuza Teixeira; Guilherme Pereira Scagion; Helena Perez Coelho; Hugo Sato; Karine Lima Lourenço; Luciano Matsumiya Thomazelli; Renata Peixoto; Rubens Daniel Miserani Magalhães; Tatiana Ometto |
| EPI_ISL_2698098 | Biology, UFLA, Universidade Federal de Lavras | Biology, UFLA | Barcante, J.; Cherem, J.; Fernandes, G.; Luciano, P.; Melo, D.; Pylro, V. |
| EPI_ISL_3364406 | Biotia | Biotia | Christopher Mason; David Danko; Dorottya Nagy-Szakal; Mara Couto-Rodriguez; Marilyne Debieu; Niamh O'Hara; Xavier Jirau Serrano |
| EPI_ISL_976950, EPI_ISL_976998, EPI_ISL_1121645, EPI_ISL_1121665, EPI_ISL_1121671, EPI_ISL_1121679, EPI_ISL_1121680 | | | |
| see above | Broad Institute | Infectious Disease | Adams, G.; B.L.; B.W.; Bauer, M.; Birren; Carter, A.; Chaluvadi, S.; D.J.; DeRuff, K.; Gallagher, G.; Gladden-Young, A.; J.E.; K.J.; Lagerborg, K.; Lemieux; Loreth, C.; MacInnis; Normandin, E.; P.C.; Park; Reilly, S.; Rudy, M.; Siddle; Smole, S.; Tomkins-Tinch, C.; and Sabeti |

| | | | |
|---|---|---|---|
| | Clinical Research Sequencing Platform | Program, Broad Institute of Harvard and MIT | |
| EPI_ISL_2105414, EPI_ISL_2105456, EPI_ISL_2105522, EPI_ISL_2105524, EPI_ISL_2105544, EPI_ISL_2105548, EPI_ISL_2156340, EPI_ISL_2156343, EPI_ISL_2156380, EPI_ISL_2156397, EPI_ISL_2156422, EPI_ISL_2156445, EPI_ISL_2188097, EPI_ISL_2188994, EPI_ISL_2189115 | | | |
| see above | Butuan Medical Center | Philippine Genome Center | Alethea R. de Guzman; Anna Ong-Lim; Arianne A. Zamora; Asia Louisa U. Chong; Benedict A. Maralit; Candice Francheska B. Tambaoan; Carlo M. Lapid; Celia Carlos; Devon Ray Pacial; Edsel Maurice Salvaña; El King D. Morado; Elcid Aaron R. Pangilinan; Eva Maria Cutiongco-de la Paz; Francis A. Tablizo; Irish Coleen A. Asin; Jaime C. Montoya; Jan Michael C. Yap; Jo-Hannah S. Llames; John Q. Wong; Joshua Gregor A. Dizon; Juan Antonio R. Magalang; Karol Sophia Agape R. Padilla; Kenneth M. Kim; Kris P. Punayan; Marc Edsel C. Ayes; Marc Jerrone R. Castro; Maria Rosario Singh-Vergeire and Cynthia P. Saloma; Maria Sofia L. Yangzon; Marissa Alejandria; Razel Nikka M. Hao; Renato Jacinto Q. Mantaring; Rianna Patricia S. Cruz; Sheila Mae M. Araiza |
| EPI_ISL_2797603 | CDPH VBL | California Department of Public Health | CDPH-COVIDNet; UCSF Center for Advanced Technology |
| EPI_ISL_3912410 | CENTRO DE ESPECIALIDADES MEDICAS DR NESTOR DE PAULA PESSOA | Analytical Competence Molecular Epidemiology Lab/ACME, Oswaldo Cruz Foundation, Ceara (FIOCRUZ CE) | Cleber Furtado Aksenen; Fabio Miyajima; Fernando Braga Stehling; Francisco Eder de Moura Lopes; Jamille Maria Mendes Bezerra; Joaquim Cesar do Nascimento Sousa Junior; Pedro Miguel Carneiro Jeronimo; Suzana Porto Almeida & Lucas Delerino on behalf of COVID-19 FIOCRUZ Genomic Network; Thais Ferreira de Oliveira; Thais de Oliveira Costa; Ticiane Cavalcante de Souza; Veridiana Pessoa Miyajima |
| EPI_ISL_1469658 | CENTRO DE ESPECIALIDADES TRIUNFO | Epiclin | Ana Paula Muterle; Carolina Comerlato; Eliana Márcia Da Ros Wendland; Fernando Hayashi Sant'Anna; Janira Prichula; Juliana Comerlato |
| EPI_ISL_1469642, EPI_ISL_1469737 | CENTRO DE REFERENCIA EM SINDROMES GRIPAIS | Epiclin | Ana Paula Muterle; Carolina Comerlato; Eliana Márcia Da Ros Wendland; Fernando Hayashi Sant'Anna; Janira Prichula; Juliana Comerlato |
| EPI_ISL_1445195, EPI_ISL_1966219 | CENTRO DE SAUDE DE BORA | Instituto Butantan / Mendelics | Antonio Jorge Martins; Bianca Cechetto Carlos. Mendelics: Bibiana Santos; Bibiana Santos; Claudia Renata dos Santos Barros; Cíntia Bittar; David Schlesinger; David Schlesinger. Hemocentro Ribeirão Preto: Simone Kashima; Debora Botequio Moretti; Dimas Tadeu Covas; Elaine Cristina Marqueze; Elaine Vieira dos Santos; Elisangela Chicaroni Mattos; Erika Freitas; Evandra Strazza Rodrigues; Felipe Allan da Silva da Costa; Flavia Aburjaile; Fábio Sossai Possebon; Guilherme Campos; Guilherme Targino Valente; Heidge Fukumasu. USP-Botucatu: Rejane Maria Tommasini Grotto; Helena Lage Ferreira; Instituto Butantan: Dimas Tadeu Covas; Jardelina de Souza Todao Bernardino; Jayme A. Souza-Neto; Jessika Cristina Chagas Lesbon; Jorge A. Petroli Marchesi; José Salvatore Leister Patané; João Paulo Kitajima; João Pessoa Araújo Jr.; Leila Sabrina Ullmann; Loyze Paola Oliveira de Lima; Luiz Aurelio de Campos Crispin. Centro de Genômica Funcional da ESALQ: Luiz Lehmann Coutinho; Luiz Carlos Junior de Alcantara; Lívia Sacchetto; Maisa C. Pereira Parra; Maria Carolina Elias; Marta Giovanetti; Marília Moraes; Maurício Lacerda Nogueira. Prefeitura de Sao Paulo: Melissa Palmieri.; Patricia Akemi Assato; Paula Rahal; Paulo Inacio da Costa; Rafael dos Santos Bezerra; Raquel de Lello Rocha Campos Cassano. NGS Soluções Genômicas: Pilar Drummond Sampaio Corrêa Mariani. FZEA-USP Pirassununga: Mirele Daiana Poleti; Raul Machado Neto; Ricardo Augusto Brassaloti; Ricardo Haddad; Rodrigo Tocantins Calado.; Rodrigo Tocantins Calado. FAMERP-SJRP: Cecília Artico Banho; Sandra Coccuzzo Sampaio; Simone Kashima; Svetoslav Nanev Slavov; Vagner Fonseca; Vincent Louis Viala |
| EPI_ISL_1966239 | CENTRO DE SAUDE DR MARIO DIAS DE AGUIAR CAPIVARI | Instituto Butantan / Mendelics | Antonio Jorge Martins; Bianca Cechetto Carlos. Mendelics: Bibiana Santos; Claudia Renata dos Santos Barros; Cíntia Bittar; David Schlesinger. Hemocentro Ribeirão Preto: Simone Kashima; Debora Botequio Moretti; Elaine Cristina Marqueze; Elaine Vieira dos Santos; Elisangela Chicaroni Mattos; Erika Freitas; Evandra Strazza Rodrigues; Felipe Allan da Silva da Costa; Flavia Aburjaile; Fábio Sossai Possebon; Guilherme Campos; Guilherme Targino Valente; Heidge Fukumasu. USP-Botucatu: Rejane Maria Tommasini Grotto; Helena Lage Ferreira; Instituto Butantan: Dimas Tadeu Covas; Jardelina de Souza Todao Bernardino; Jayme A. Souza-Neto; Jessika Cristina Chagas Lesbon; Jorge A. Petroli Marchesi; José Salvatore Leister Patané; João Paulo Kitajima; João Pessoa Araújo Jr.; Leila Sabrina Ullmann; Loyze Paola Oliveira de Lima; Luiz Aurelio de Campos Crispin. Centro de Genômica Funcional da ESALQ: Luiz Lehmann Coutinho; Luiz Carlos Junior de Alcantara; Lívia Sacchetto; Maisa C. Pereira Parra; Maria Carolina Elias; Marta Giovanetti; Marília Moraes; Maurício Lacerda Nogueira. Prefeitura de Sao Paulo: Melissa Palmieri.; Patricia Akemi Assato; Paula Rahal; Paulo Inacio da Costa; Rafael dos Santos Bezerra; Raquel de Lello Rocha Campos Cassano. NGS Soluções Genômicas: Pilar Drummond Sampaio Corrêa Mariani. FZEA-USP Pirassununga: Mirele Daiana Poleti; Raul Machado Neto; Ricardo Augusto Brassaloti; Ricardo Haddad; Rodrigo Tocantins Calado. FAMERP-SJRP: Cecília Artico Banho; Sandra Coccuzzo Sampaio; Svetoslav Nanev Slavov; Vagner Fonseca; Vincent Louis Viala |
| EPI_ISL_3912412 | CENTRO DE SAUDE DR MIRANDA TAVARES | Analytical Competence Molecular Epidemiology Lab/ACME, Oswaldo Cruz Foundation, Ceara (FIOCRUZ CE) | Cleber Furtado Aksenen; Fabio Miyajima; Fernando Braga Stehling; Francisco Eder de Moura Lopes; Jamille Maria Mendes Bezerra; Joaquim Cesar do Nascimento Sousa Junior; Pedro Miguel Carneiro Jeronimo; Suzana Porto Almeida & Lucas Delerino on behalf of COVID-19 FIOCRUZ Genomic Network; Thais Ferreira de Oliveira; Thais de Oliveira Costa; Ticiane Cavalcante de Souza; Veridiana Pessoa Miyajima |
| EPI_ISL_1445089, EPI_ISL_1966131 | CENTRO DE SAUDE II MAIRINQUE MAIRINQUE | Instituto Butantan / Mendelics | Antonio Jorge Martins; Bianca Cechetto Carlos. Mendelics: Bibiana Santos; Bibiana Santos; Claudia Renata dos Santos Barros; Cíntia Bittar; David Schlesinger; David Schlesinger. Hemocentro Ribeirão Preto: Simone Kashima; Debora Botequio Moretti; Dimas Tadeu Covas; Elaine Cristina Marqueze; Elaine Vieira dos Santos; Elisangela Chicaroni Mattos; Erika Freitas; Evandra Strazza Rodrigues; Felipe Allan da Silva da Costa; Flavia Aburjaile; Fábio Sossai Possebon; Guilherme Campos; Guilherme Targino Valente; Heidge Fukumasu. USP-Botucatu: Rejane Maria Tommasini Grotto; Helena Lage Ferreira; Instituto Butantan: Dimas Tadeu Covas; Jardelina de Souza Todao Bernardino; Jayme A. Souza-Neto; Jessika Cristina Chagas Lesbon; Jorge A. Petroli Marchesi; José Salvatore Leister Patané; João Paulo Kitajima; João Pessoa Araújo Jr.; Leila Sabrina Ullmann; Loyze Paola Oliveira de Lima; Luiz Aurelio de Campos Crispin. Centro de Genômica Funcional da ESALQ: Luiz Lehmann Coutinho; Luiz Carlos Junior de Alcantara; Lívia Sacchetto; Maisa C. Pereira Parra; Maria Carolina Elias; Marta Giovanetti; Marília Moraes; Maurício Lacerda Nogueira. Prefeitura de Sao Paulo: Melissa Palmieri.; Patricia Akemi Assato; Paula Rahal; Paulo Inacio da Costa; Rafael dos Santos Bezerra; Raquel de Lello Rocha Campos Cassano. NGS Soluções Genômicas: Pilar Drummond Sampaio Corrêa Mariani. FZEA-USP Pirassununga: Mirele Daiana Poleti; Raul Machado Neto; Ricardo Augusto Brassaloti; Ricardo Haddad; Rodrigo Tocantins Calado.; Rodrigo Tocantins Calado. FAMERP-SJRP: Cecília Artico Banho; Sandra Coccuzzo Sampaio; Simone Kashima; Svetoslav Nanev Slavov; Vagner Fonseca; Vincent Louis Viala |
| EPI_ISL_3912414 | CENTRO DE SAUDE OTAVIO LOBO | Analytical Competence Molecular Epidemiology Lab/ACME, Oswaldo Cruz Foundation, Ceara (FIOCRUZ CE) | Cleber Furtado Aksenen; Fabio Miyajima; Fernando Braga Stehling; Francisco Eder de Moura Lopes; Jamille Maria Mendes Bezerra; Joaquim Cesar do Nascimento Sousa Junior; Pedro Miguel Carneiro Jeronimo; Suzana Porto Almeida & Lucas Delerino on behalf of COVID-19 FIOCRUZ Genomic Network; Thais Ferreira de Oliveira; Thais de Oliveira Costa; Ticiane Cavalcante de Souza; Veridiana Pessoa Miyajima |
| EPI_ISL_1469584, EPI_ISL_1469629 | CENTRO MUNICIPAL DE SAUDE DE ROLANTE | Epiclin | Ana Paula Muterle; Carolina Comerlato; Eliana Márcia Da Ros Wendland; Fernando Hayashi Sant'Anna; Janira Prichula; Juliana Comerlato |
| EPI_ISL_3912415 | CENTRO ODONTOLOGICO DA POLICIA MILITAR DO CEARA CEOP PMCE | Analytical Competence Molecular Epidemiology Lab/ACME, Oswaldo Cruz Foundation, Ceara (FIOCRUZ CE) | Cleber Furtado Aksenen; Fabio Miyajima; Fernando Braga Stehling; Francisco Eder de Moura Lopes; Jamille Maria Mendes Bezerra; Joaquim Cesar do Nascimento Sousa Junior; Pedro Miguel Carneiro Jeronimo; Suzana Porto Almeida & Lucas Delerino on behalf of COVID-19 FIOCRUZ Genomic Network; Thais Ferreira de Oliveira; Thais de Oliveira Costa; Ticiane Cavalcante de Souza; Veridiana Pessoa Miyajima |
| EPI_ISL_732252 | CH Porto - H Sto Antonio | Instituto Nacional de Saude (INSA) and Instituto Gulbenkian de Ciencia (IGC) | Borges et al |
| EPI_ISL_2017667, EPI_ISL_2516562, EPI_ISL_2516565, EPI_ISL_2516577 | CLILAB | Microbiology Department | Aida Gonzalez-Diaz; Carmen Ardanuy; Jordi Camara; Jordi Niubó; Laura Calatayud; M Angeles Domínguez; Miguel Fernandez-Huerta; Sara Marti |
| EPI_ISL_2383909, EPI_ISL_2383915 | CO Dept. of Public Health and Environment, Lab Services Division | Centers for Disease Control and Prevention Division of Viral Diseases, Pathogen Discovery | Alison Laufer Halpin; Ben L. Rambo-Martin; Clinton R. Paden; Dakota Howard; Darlene Wagner; Dave Wentworth; Dhwani Batra; Jasmine Padilla; Justin Lee; Katie Dillon; Krista Queen; Kristen Knipe; Kristine Lacek; Mark Burroughs; Matthew Schmerer; Mili Sheth; Peter Cook; Sam Shepard; Sarah Nobles; Shoshona Le; Suxiang Tong; Vivien Dugan; Yvette Unoarumhi |
| EPI_ISL_1469604, EPI_ISL_1469608, EPI_ISL_1469636, EPI_ISL_1469641 | COORDENADORIA GERAL DE VIGILANCIA EM SAUDE | Epiclin | Ana Paula Muterle; Carolina Comerlato; Eliana Márcia Da Ros Wendland; Fernando Hayashi Sant'Anna; Janira Prichula; Juliana Comerlato |
| EPI_ISL_735408 | COVID 19 Centro de Combate ao Coronavirus CCC Jandira | Instituto Adolfo Lutz, Interdiciplinary Procedures Center, Strategic Laboratory | Claudia Regina Gonçalves; Claudio Tavares Sacchi; Erica Valessa Ramos Gomes; Karoline Rodrigues Campos |
| EPI_ISL_2170967 | CS DE NIPOA | Instituto Butantan / Mendelics | Antonio Jorge Martins; Bianca Cechetto Carlos. Mendelics: Bibiana Santos; Claudia Renata dos Santos Barros; Cíntia Bittar; David Schlesinger. Hemocentro Ribeirão Preto: Simone Kashima; Debora Botequio Moretti; Elaine Cristina Marqueze; Elaine Vieira dos Santos; Elisangela Chicaroni Mattos; Erika Freitas; Evandra Strazza Rodrigues; Felipe Allan da Silva da Costa; Flavia Aburjaile; Fábio Sossai Possebon; Guilherme Campos; Guilherme Targino Valente; Heidge Fukumasu. USP-Botucatu: Rejane Maria Tommasini Grotto; Helena Lage Ferreira; Instituto Butantan: Dimas Tadeu Covas; Jardelina de Souza Todao Bernardino; Jayme A. Souza-Neto; Jessika Cristina Chagas Lesbon; Jorge A. Petroli Marchesi; José Salvatore Leister Patané; João Paulo Kitajima; João Pessoa Araújo Jr.; Leila Sabrina Ullmann; Loyze Paola Oliveira de Lima; Luiz Aurelio de Campos Crispin. Centro de Genômica Funcional da ESALQ: Luiz Lehmann Coutinho; Luiz Carlos Junior de Alcantara; Lívia Sacchetto; Maisa C. Pereira Parra; Maria Carolina Elias; Marta Giovanetti; Marília Moraes; Maurício Lacerda Nogueira. Prefeitura de Sao Paulo: Melissa Palmieri.; Patricia Akemi Assato; Paula Rahal; Paulo Inacio da Costa; Rafael dos Santos Bezerra; Raquel de Lello Rocha Campos Cassano. NGS Soluções Genômicas: Pilar Drummond Sampaio Corrêa Mariani. FZEA-USP Pirassununga: Mirele Daiana Poleti; Raul Machado Neto; Ricardo Augusto Brassaloti; Ricardo Haddad; Rodrigo Tocantins Calado. FAMERP-SJRP: Cecília Artico Banho; Sandra Coccuzzo Sampaio; Svetoslav Nanev Slavov; Vagner Fonseca; Vincent Louis Viala |
| EPI_ISL_2344853 | CS DE PALMARES PAULISTA | Instituto Butantan / ESALQ-Piracicaba | Antonio Jorge Martins; Claudia Renata dos Santos Barros; David Schlesinger; Debora Botequio Moretti; Dimas Tadeu Covas; Elaine Cristina Marqueze; Elaine Vieira Santos; Evandra Strazza Rodrigues; Heidge Fukumasu; Jayme Augusto de Souza-Neto; José Salvatore Leister Patané; Luiz Alcantara; Luiz Lehmann Coutinho; Maria Carolina Elias; Maurício Lacerda Nogueira; Rafael dos Santos Bezerra; Raul Machado Neto; Rejane Maria Tommasini Grotto; Ricardo Haddad; Sandra Coccuzzo Sampaio Vessoni; Simone Kashima; Svetoslav Nanev Slavov; Vincent Louis Viala |
| EPI_ISL_583494 | CS II Dr. Antonio Vicoso Moreira de Rezende Sumare | Instituto Adolfo Lutz, Interdiciplinary Procedures Center, Strategic Laboratory | Claudia Regina Gonçalves; Claudio Tavares Sacchi; Erica Valessa Ramos Gomes; Karoline Rodrigues Campos |
| EPI_ISL_468314, EPI_ISL_583503 | CTA Centro de Testagem e Aconselhamento | Instituto Adolfo Lutz, Interdiciplinary Procedures Center, Strategic Laboratory | Claudia Regina Gonçalves; Claudio Tavares Sacchi; Erica Valessa Ramos Gomes; Karoline Rodrigues Campos |
| EPI_ISL_2105218, EPI_ISL_2105225, EPI_ISL_2105226, EPI_ISL_2105231, EPI_ISL_2105242, EPI_ISL_2105245, EPI_ISL_2105253, EPI_ISL_2105254, EPI_ISL_2105255, EPI_ISL_2105270, EPI_ISL_2105276, EPI_ISL_2105282, EPI_ISL_2105291, EPI_ISL_2105299, EPI_ISL_2105322, EPI_ISL_2105409, EPI_ISL_2105448, EPI_ISL_2105453, EPI_ISL_2105529, EPI_ISL_2105532, EPI_ISL_2156415, EPI_ISL_2156431, EPI_ISL_2156432, EPI_ISL_2156433, EPI_ISL_2156460, EPI_ISL_2156515, EPI_ISL_2156553, EPI_ISL_2156581, EPI_ISL_2156625, EPI_ISL_2156650, EPI_ISL_2156679, EPI_ISL_2156700, EPI_ISL_2156705, EPI_ISL_2156712, EPI_ISL_2156733, EPI_ISL_2188061 | | | |
| see above | Cagayan Valley Medical Center Molecular Laboratory | Philippine Genome Center | Alethea R. de Guzman; Anna Ong-Lim; Arianne A. Zamora; Asia Louisa U. Chong; Benedict A. Maralit; Candice Francheska B. Tambaoan; Carlo M. Lapid; Celia Carlos; Devon Ray Pacial; Edsel Maurice Salvaña; El King D. Morado; Elcid Aaron R. Pangilinan; Eva Maria Cutiongco-de la Paz; Francis A. Tablizo; Irish Coleen A. Asin; Jaime C. Montoya; Jan Michael C. Yap; Jo-Hannah S. Llames; John Q. Wong; Joshua Gregor A. Dizon; Juan Antonio R. Magalang; Karol Sophia Agape R. Padilla; Kenneth M. Kim; Kris P. Punayan; Marc Edsel C. Ayes; Marc Jerrone R. Castro; Maria Rosario Singh-Vergeire and Cynthia P. Saloma; Maria Sofia L. Yangzon; Marissa Alejandria; Razel Nikka M. Hao; Rianna Patricia S. Cruz; Sheila Mae M. Araiza |
| EPI_ISL_4005510 | California | California Department | CDPH IDLB COVIDNet et al |

| | | | |
|---|---|---|---|
| | Department of Public Health | of Public Health | |
| EPI_ISL_2837812 | California Department of Public Health Valencia Branch Laboratory (CDPH VBL) | California Department of Public Health | CDPH-COVIDNet |
| EPI_ISL_579220 | Canterbury Health Laboratories | Institute of Environmental Science and Research (ESR) | Anja Werno; Antje van der Linden; Arlo Upton; Chris Mansell; David Hammer; Dragana Drinkovic; Erasmus Smit; Gary McAuliffe; Hana Sofia Andersson; Hermes Perez; James Ussher; Jill Sherwood; Jing Wang; Joep de Ligt; Josh Freeman; Julia Howard; Juliet Elvy; Lauren Jelly; Mary DeAlmeida; Matt Blakiston; Matt Storey; Matthew Rogers; Max Bloomfield; Michael Addidle; Michelle Balm; Muhammad Faisal; Nikki Freed; Olin Silander; Sally Roberts; Sarah Jefferies; Sharmini Muttaiyah; Susan Morpeth; Susan Taylor; Timothy Blackmore; Vani Sathyendran; Veronica Playle; Virginia Hope; Xiaoyun Ren |
| EPI_ISL_583504, EPI_ISL_583505 | Casa de Saude Stella Maris | Instituto Adolfo Lutz, Interdiciplinary Procedures Center, Strategic Laboratory | Claudia Regina Gonçalves; Claudio Tavares Sacchi; Erica Valessa Ramos Gomes; Karoline Rodrigues Campos |
| EPI_ISL_693235 | Casmi Centro Atendimento Saude da Mulher e Infancia | Instituto Adolfo Lutz, Interdiciplinary Procedures Center, Strategic Laboratory | Claudia Regina Gonçalves; Claudio Tavares Sacchi; Erica Valessa Ramos Gomes; Karoline Rodrigues Campos |
| EPI_ISL_2105383 | Cebu TB Reference Laboratory | Philippine Genome Center | Alethea R. de Guzman; Anna Ong-Lim; Arianne A. Zamora; Asia Louisa U. Chong; Benedict A. Maralit; Candice Francheska B. Tambaoan; Carlo M. Lapid; Celia Carlos; Devon Ray Pacial; Edsel Maurice Salvaña; El King D. Morado; Eva Maria Cutiongco-de la Paz; Francis A. Tablizo; Irish Coleen A. Asin; Jaime C. Montoya; Jan Michael C. Yap; Jo-Hannah S. Llames; John Q. Wong; Joshua Gregor A. Dizon; Juan Antonio R. Magalang; Karol Sophia Agape R. Padilla; Kenneth M. Kim; Kris P. Punayan; Marc Edsel C. Ayes; Marc Jerrone R. Castro; Maria Rosario Singh-Vergeire and Cynthia P. Saloma; Maria Sofia L. Yangzon; Marissa Alejandria; Razel Nikka M. Hao; Rianna Patricia S. Cruz; Sheila Mae M. Araiza |
| EPI_ISL_522491 | Center for Laboratory Control of Infectious Diseases, Korea Centers for Diseases Control and Prevention | Center for Laboratory Control of Infectious Diseases, Korea Centers for Diseases Control and Prevention | Ae Kyung Park; Eunkyung Shin; Heui Man Kim; Jeong-Min Kim; Jin Sun No; Junyoung Kim; Myung Guk Han; Yoon-Seok Chung |
| EPI_ISL_940924 | Centers for Disease Control and Prevention, Dengue Branch | Centers for Disease Control and Prevention, Dengue Branch | Betzabel Flores; Gabriela Paz-Bailey; Gilberto A. Santiago; Glenda Gonzalez; Jorge L. Munoz-Jordan; Keyla Charriez |
| EPI_ISL_930857, EPI_ISL_942898, EPI_ISL_943581, EPI_ISL_943584, EPI_ISL_943606 | Central Laboratory of Public Health of Rio Grande do Sul (Lacen-RS) | State Center for Health Surveillance of the Health Department of the State of Rio Grande do Sul (CEVS/SES-RS) | ; Aline Campos; Amanda da Silva; Anelise Schaurich; Barcellos R; Campos A; Claudia Dornelles; Crescente L; Cynthia Molina; Da Silva A; Dornelles C; Fernanda Godinho; Fonseca V; Garay L; Godinho F; Gonzalez A; Gregianini T; Lara Crescente; Leticia Garay; Molina C; Regina Barcellos; Richard Salvato; Salvato R; Schaurich A; Tatiana Gregianini; Vagner Fonseca |
| EPI_ISL_978498, EPI_ISL_978506, EPI_ISL_978515, EPI_ISL_978517, EPI_ISL_978518, EPI_ISL_978525, EPI_ISL_978529, EPI_ISL_1068319, EPI_ISL_1068363, EPI_ISL_1068364, EPI_ISL_1068369, EPI_ISL_1068371, EPI_ISL_1068373, EPI_ISL_1068376, EPI_ISL_1068377, EPI_ISL_1068378, EPI_ISL_1068380, EPI_ISL_1068394, EPI_ISL_1583642, EPI_ISL_1583643, EPI_ISL_1583648, EPI_ISL_1583649, EPI_ISL_1583651, EPI_ISL_1583657, EPI_ISL_1583658, EPI_ISL_1583659, EPI_ISL_1583660, EPI_ISL_1583668, EPI_ISL_1583671, EPI_ISL_1583686, EPI_ISL_1583727, EPI_ISL_3255056, EPI_ISL_3255082, EPI_ISL_3255084, EPI_ISL_3266080, EPI_ISL_3266082, EPI_ISL_3266083, EPI_ISL_3266085, EPI_ISL_3266086, EPI_ISL_3266096, EPI_ISL_3266097, EPI_ISL_3266121, EPI_ISL_3506972, EPI_ISL_3506975, EPI_ISL_3506976, EPI_ISL_3506981, EPI_ISL_3506982, EPI_ISL_3506983, EPI_ISL_3506986, EPI_ISL_3631208 | | | |
| see above | Central Public Health Laboratory - LACEN -Bahia, Salvador, Brazil | Central Public Health Laboratory - LACEN - Bahia, Salvador, Brazil | Arabela Leal; Breno Dominguez; Felicidade Pereira; Jaqueline Gomes; Luciana Oliveira; Luiz Alcantara; Marcela Gómez; Marta Giovanetti; Patrícia Cajado; Stephane Tosta; Vagner Fonseca; Vanessa Nardy |
| EPI_ISL_515547 | Centro Medico da Policia Militar do Estado de Sao Paulo | Instituto Adolfo Lutz, Interdiciplinary Procedures Center, Strategic Laboratory | Claudia Regina Gonçalves; Claudio Tavares Sacchi; Erica Valessa Ramos Gomes |
| EPI_ISL_693244 | Centro Médico da Polícia Militar do Estado de Sao Paulo | Instituto Adolfo Lutz, Interdiciplinary Procedures Center, Strategic Laboratory | Claudia Regina Gonçalves; Claudio Tavares Sacchi; Erica Valessa Ramos Gomes; Karoline Rodrigues Campos |
| EPI_ISL_2612324, EPI_ISL_2612328, EPI_ISL_2612348, EPI_ISL_2612349, EPI_ISL_2612353, EPI_ISL_2612355, EPI_ISL_2612361, EPI_ISL_2612364, EPI_ISL_2612393, EPI_ISL_2612394, EPI_ISL_2612395 | | | |
| see above | Centro de Infectologia Charles Mérieux/ Laboratório Rodolphe Mérieux, FUNDHACRE | Bioinformatics Laboratory / LNCC | Alessandra P Lamarca; Alexandra L Gerber; Ana Paula de C Guimarães; Ana Tereza R Vasconcelos; Andreas Stocker; Cirley Maria de Oliveira Lobato; Douglas Terra Machado; Luiz Fellype Alves de Souza; Luiz G P de Almeida; Ronaldo da Silva F Jr |
| EPI_ISL_2491693, EPI_ISL_2491756, EPI_ISL_2491757, EPI_ISL_2491758, EPI_ISL_2491760, EPI_ISL_2491762, EPI_ISL_2491772, EPI_ISL_2491780 | | | |
| see above | Centro de Pesquisa Gonçalo Moniz (CPqGM - FIOCRUZ/BA) | Laboratory of Respiratory Viruses and Measles, Oswaldo Cruz Institute, FIOCRUZ | Alice Sampaio Rocha; Ana Carolina Mendonca; Anna Carolina Paixao; Camila I. de Oliveira; Elisa Cavalcante Pereira; Fernando Motta; Luciana Appolinario; Marilda Siqueira on behalf of the Fiocruz COVID-19 Genomic Surveillance Network; Paola Resende; Renata Serrano Lopes; Ricardo Khouri; Taina Venas |
| EPI_ISL_583500 | Centro de Saude I Tacito Leite de Carvalho e Silva | Instituto Adolfo Lutz, Interdiciplinary Procedures Center, Strategic Laboratory | Claudia Regina Gonçalves; Claudio Tavares Sacchi; Erica Valessa Ramos Gomes; Karoline Rodrigues Campos |
| EPI_ISL_1493591 | Centro de Saude II Dr Alcides Facundo Arroyo | Instituto Adolfo Lutz, Interdiciplinary Procedures Center, Strategic Laboratory | Caio Vinicius Dias Lopes; Claudia Regina Gonçalves; Claudio Tavares Sacchi; Erica Valessa Ramos Gomes; Karoline Rodrigues Campos |
| EPI_ISL_1520134, EPI_ISL_1520135 | Centro de Saude II Dr Jose Paione Mococa | Instituto Adolfo Lutz, Interdiciplinary Procedures Center, Strategic Laboratory | Caio Vinicius Dias Lopes; Claudia Regina Gonçalves; Claudio Tavares Sacchi; Erica Valessa Ramos Gomes; Karoline Rodrigues Campos |
| EPI_ISL_1533691 | Centro de Saude II Dr. Jose Paione Mococa | Instituto Adolfo Lutz, Interdiciplinary Procedures Center, Strategic Laboratory | Caio Vinicius Dias Lopes; Claudia Regina Gonçalves; Claudio Tavares Sacchi; Erica Valessa Ramos Gomes; Karoline Rodrigues Campos; Leonardo Jose Tadeu de Araujo |
| EPI_ISL_1396459 | Centro de Tecnología en Salud Pública de la Universidad Nacional de Rosario | Laboratorio Mixto de Biotecnología Acuática (LMBA) on behalf of 'Proyecto Argentino Interinstitucional de genomica de SARS-CoV-2' (PAIS Consortium) | Adriana Giri; Agustina Cerri; Ana Cavatorta; Ana Paletta; Diego Chouhy; Elisa Bolatti; Elizabeth Tapia (argenTAG); Federico Remes Lenicov; Flavio Spetale; Gastón Viarengo; Ignacio García Labari; Javier Murillo; Joaquín Ezpeleta; Julian Acosta; Laura Angelone; Leandro Ciappina; María Re; Pablo Casal; Pilar Bulacio; Silvana Spinelli; Silvia Arranz; Sofía Lavista Llanos; Vanina Villanova; Victoria Posner |
| EPI_ISL_882659 | Centro de Triagem Covid19 | Instituto Adolfo Lutz, Interdiciplinary Procedures Center, Strategic Laboratory | Claudia Regina Gonçalves; Claudio Tavares Sacchi; Erica Valessa Ramos Gomes; Karoline Rodrigues Campos |
| EPI_ISL_468305, EPI_ISL_468307, EPI_ISL_735411, EPI_ISL_735424, EPI_ISL_735426 | Centro de Vigilancia a Saude de Diadema | Instituto Adolfo Lutz, Interdiciplinary Procedures Center, Strategic Laboratory | Claudia Regina Gonçalves; Claudio Tavares Sacchi; Erica Valessa Ramos Gomes; Karoline Rodrigues Campos |
| EPI_ISL_693240, | Centro de Vigilância | Instituto Adolfo Lutz, | Claudia Regina Gonçalves; Claudio Tavares Sacchi; Erica Valessa Ramos Gomes; Karoline Rodrigues Campos |

| | | | |
|---|---|---|---|
| EPI_ISL_693242 | a Saude de Diadema | Interdiciplinary Procedures Center, Strategic Laboratory | |
| EPI_ISL_1164590 | Charité Universitätsmedizin Berlin, Institut für Virologie/Labor Berlin | Charité Universitätsmedizin Berlin, Institut für Virologie | Barbara Mühlemann; Christian Drosten; Julia Schneider; Jörn Beheim-Schwarzbach; Talitha Veith; Terry Jones; Victor M Corman |
| EPI_ISL_581703 | Clinical Virology | Clinical Bacteriology | Adrian Egli; Alexander Gensch; Alfredo Mari; Christian Nickel; Hans Hirsch; Hans Pargger; Helena MB Seth-Smith; Julia Bielicki; Karoline Leuzinger; Kirstine K. Soegaard; Madlen Stange; Manuel Battegay; Martin Siegemund; Michael Osthoff; Michael Schweitzer; Myrta Brunner; Rita Schneider-SliFemalea; Roland Bingisser; Sarah Tschudin-Sutter; Simon Fuchs; Stefano Bassetti; Tim Roloff |
| EPI_ISL_2277174, EPI_ISL_2309973 | Colorado Department of Public Health and Environment | Colorado Department of Public Health and Environment | Alexandria Rossheim; Diana Ir; Emily A. Travanty; Laura Bankers; Molly C. Hetherington-Rauth; Sarah Elizabeth Totten; Shannon R. Matzinger |
| EPI_ISL_583497 | Complexo Hospitalar Ouro Verde de Campinas | Instituto Adolfo Lutz, Interdiciplinary Procedures Center, Strategic Laboratory | Claudia Regina Gonçalves; Claudio Tavares Sacchi; Erica Valessa Ramos Gomes; Karoline Rodrigues Campos |
| EPI_ISL_1121323 | Complexo Hospitalar Padre Bentode Guarulhos | Instituto Adolfo Lutz, Interdiciplinary Procedures Center, Strategic Laboratory | Caio Vinicius Dias Lopes; Claudia Regina Gonçalves; Claudio Tavares Sacchi; Erica Valessa Ramos Gomes; Karoline Rodrigues Campos |
| EPI_ISL_523970 | Conjunto Hospitalar do Mandaqui | Instituto Adolfo Lutz, Interdiciplinary Procedures Center, Strategic Laboratory | Claudia Regina Gonçalves; Claudio Tavares Sacchi; Erica Valessa Ramos Gomes |
| EPI_ISL_2105543, EPI_ISL_2156282, EPI_ISL_2156299, EPI_ISL_2171229, EPI_ISL_2188677, EPI_ISL_2188728, EPI_ISL_2188775 | | | |
| see above | Cotabato Regional and Medical Center | Philippine Genome Center | Alethea R. de Guzman; Anna Ong-Lim; Arianne A. Zamora; Asia Louisa U. Chong; Benedict A. Maralit; Candice Francheska B. Tambaoan; Carlo M. Lapid; Celia Carlos; Devon Ray Pacial; Edsel Maurice Salvaña; El King D. Morado; Elcid Aaron R. Pangilinan; Eva Maria Cutiongco-de la Paz; Francis A. Tablizo; Irish Coleen A. Asin; Jaime C. Montoya; Jan Michael C. Yap; Jo-Hannah S. Llames; John Q. Wong; Joshua Gregor A. Dizon; Juan Antonio R. Magalang; Karol Sophia Agape R. Padilla; Kenneth M. Kim; Kris P. Punayan; Marc Edsel C. Ayes; Marc Jerrone R. Castro; Maria Rosario Singh-Vergeire and Cynthia P. Saloma; Maria Sofia L. Yangzon; Marissa Alejandria; Razel Nikka M. Hao; Renato Jacinto Q. Mantaring; Rianna Patricia S. Cruz; Sheila Mae M. Araiza |
| EPI_ISL_693207 | Cs II Doutor Antonio Vicoso Moreira de Rezende | Instituto Adolfo Lutz, Interdiciplinary Procedures Center, Strategic Laboratory | Claudia Regina Gonçalves; Claudio Tavares Sacchi; Erica Valessa Ramos Gomes; Karoline Rodrigues Campos |
| EPI_ISL_833168 | DB Diagnosticos do Brasil | Instituto Adolfo Lutz, Interdiciplinary Procedures Center, Strategic Laboratory | Claudia Regina Gonçalves; Claudio Tavares Sacchi; Erica Valessa Ramos Gomes; Karoline Rodrigues Campos |
| EPI_ISL_476282 | DB Diagnósticos do Brasil | Instituto de Medicina Tropical da Univesidade de São Paulo | Camila Alves Maia da Silva; Darlan da Silva Candido; Erika Regina Manuli; Ester Sabino; Flavia Cristina da Silva Sales; Giula Magalhaes Ferreira; Jaqueline Goes de Jesus; Julien Theze; Mariana Severo Ramundo; Nuno Faria; Samples: Nelson Gaburo Jr; Sequencing: Ingra Morales Claro; Thais de Moura Coletti |
| EPI_ISL_1469579, EPI_ISL_1469633, EPI_ISL_1469637, EPI_ISL_1469638, EPI_ISL_1469675, EPI_ISL_1469677, EPI_ISL_1469684, EPI_ISL_1469692, EPI_ISL_1469731, EPI_ISL_1469781, EPI_ISL_1479121 | | | |
| see above | DIRETORIA DE VIGILANCIA EM SAUDE | Epiclin | Ana Paula Muterle; Carolina Comerlato; Eliana Márcia Da Ros Wendland; Fernando Hayashi Sant'Anna; Janira Prichula; Juliana Comerlato |
| EPI_ISL_2344691 | DIRETORIA MUNICIPAL DE SAUDE DE ENGENHEIRO COELHO | Instituto Butantan / FZEA-USP-Pirassununga | Antonio Jorge Martins; Claudia Renata dos Santos Barros; David Schlesinger; Debora Botequio Moretti; Dimas Tadeu Covas; Elaine Cristina Marqueze; Elaine Vieira Santos; Evandra Strazza Rodrigues; Heidge Fukumasu; Jayme Augusto de Souza-Neto; José Salvatore Leister Patané; Luiz Alcantara; Luiz Lehmann Coutinho; Maria Carolina Elias; Maurício Lacerda Nogueira; Rafael dos Santos Bezerra; Raul Machado Neto; Rejane Maria Tommasini Grotto; Ricardo Haddad; Sandra Coccuzzo Sampaio Vessoni; Simone Kashima; Svetoslav Nanev Slavov; Vincent Louis Viala |
| EPI_ISL_857239 | DOHMH Corona | New York City Public Health Laboratory | Jade Wang; et al. |
| EPI_ISL_937224 | DOHMH Jamaica | New York City Public Health Laboratory | Jade Wang; et al. |
| EPI_ISL_2105331, EPI_ISL_2156371, EPI_ISL_2156403, EPI_ISL_2156405, EPI_ISL_2156406 | Davao One World Diagnostic Center Incorporated | Philippine Genome Center | Alethea R. de Guzman; Anna Ong-Lim; Arianne A. Zamora; Asia Louisa U. Chong; Benedict A. Maralit; Candice Francheska B. Tambaoan; Carlo M. Lapid; Celia Carlos; Devon Ray Pacial; Edsel Maurice Salvaña; El King D. Morado; Elcid Aaron R. Pangilinan; Eva Maria Cutiongco-de la Paz; Francis A. Tablizo; Irish Coleen A. Asin; Jaime C. Montoya; Jan Michael C. Yap; Jo-Hannah S. Llames; John Q. Wong; Joshua Gregor A. Dizon; Juan Antonio R. Magalang; Karol Sophia Agape R. Padilla; Kenneth M. Kim; Kris P. Punayan; Marc Edsel C. Ayes; Marc Jerrone R. Castro; Maria Rosario Singh-Vergeire and Cynthia P. Saloma; Maria Sofia L. Yangzon; Marissa Alejandria; Razel Nikka M. Hao; Rianna Patricia S. Cruz; Sheila Mae M. Araiza |
| EPI_ISL_906065 | Day Hospital de Ermelino Matarazzo | Instituto Adolfo Lutz, Interdiciplinary Procedures Center, Strategic Laboratory | Claudia Regina Gonçalves; Claudio Tavares Sacchi; Erica Valessa Ramos Gomes; Karoline Rodrigues Campos |
| EPI_ISL_1939017 | Departamento de Microbiología, CDB, Hospital Clínic, Barcelona | SeqCOVID-SPAIN consortium/IBV(CSIC) | Aida Peiró and SeqCOVID-SPAIN consortium; Andrea Vergara; Elisa Rubio; Jéssica Navero; Mikel Martínez |
| EPI_ISL_603034 | Departamento de Vigilância à Saúde | Instituto Adolfo Lutz, Interdiciplinary Procedures Center, Strategic Laboratory | Claudia Regina Gonçalves; Claudio Tavares Sacchi; Erica Valessa Ramos Gomes; Karoline Rodrigues Campos |
| EPI_ISL_1340751, EPI_ISL_1340754, EPI_ISL_1340756, EPI_ISL_1340757, EPI_ISL_1340764 | Departamento de Virologia, Laboratorio Central de Salud Pública, Avenida Venezuela y Teniente Escurra, Asunción, Paraguay | Laboratory of Respiratory Viruses and Measles, Oswaldo Cruz Institute, FIOCRUZ | Alice Sampaio Rocha; Ana Carolina Mendonca; Anna Carolina Paixao; Cynthia Vazquez; Fernando Motta; Luciana Appolinario; Marilda Siqueira on behalf of the Fiocruz COVID-19 Genomic Surveillance Network; Paola Resende; Renata Serrano Lopes |
| EPI_ISL_1667472 | Department of Laboratory Medicine, National Taiwan University Hospital | Microbial Genomics Core Lab, National Taiwan University Centers of Genomic and Precision Medicine | Chiao-Ling Li; Pei-Jer Chen; Shan-Chwen Chang; Shiou-Hwei Yeh; Sui-Yuan Chang; Ya-Yun Lai; You-Yu Lin |
| EPI_ISL_1034353 | Department of Medical Microbiology, St. Olavs hospital | Norwegian Institute of Public Health, Department of Virology | Atiya R Ali; Hilde Elshaug; Hilde Vollan; Ignacio Garcia Llorente; Kamilla Heddeland Instefjord; Karoline Bragstad; Kathrine Stene-Johansen; Marie Paulsen Madsen; Olav Hungnes; Rasmus Riis Kopperud; Serina B Engebretsen |
| EPI_ISL_619577 | Department of Virus and Microbiological Special Diagnostics, Statens Serum Institut, Denmark | Albertsen lab, Department of Chemistry and Bioscience, Aalborg University, Denmark | Danish Covid-19 Genome Consortia |
| EPI_ISL_1092725 | Diagnosticos da America - DASA | Instituto Adolfo Lutz, Interdiciplinary Procedures Center, Strategic Laboratory | Claudia Regina Gonçalves; Claudio Tavares Sacchi; Erica Valessa Ramos Gomes; Karoline Rodrigues Campos |
| EPI_ISL_1533724 | Diretoria Municipal de Saude | Instituto Adolfo Lutz, Interdiciplinary | Caio Vinicius Dias Lopes; Claudia Regina Gonçalves; Claudio Tavares Sacchi; Erica Valessa Ramos Gomes; Karoline Rodrigues Campos; Leonardo Jose Tadeu de Araujo |

| | | | |
|---|---|---|---|
| | | Procedures Center, Strategic Laboratory | |
| EPI_ISL_1469560, EPI_ISL_1469609, EPI_ISL_1469610, EPI_ISL_1469616, EPI_ISL_1469623, EPI_ISL_1469625, EPI_ISL_1469627, EPI_ISL_1469656, EPI_ISL_1469669, EPI_ISL_1469708, EPI_ISL_1469748, EPI_ISL_1469783, EPI_ISL_1469792, EPI_ISL_1469796, EPI_ISL_1469802, EPI_ISL_1469808, EPI_ISL_1469823, EPI_ISL_1469835 | | | |
| see above | Diretoria de Vigilância em Saúde | Epiclin | Ana Paula Muterle; Carolina Comerlato; Eliana Márcia Da Ros Wendland; Fernando Hayashi Sant'Anna; Janira Prichula; Juliana Comerlato |
| EPI_ISL_583499 | Distrito Sanitario Sul Campinas | Instituto Adolfo Lutz, Interdiciplinary Procedures Center, Strategic Laboratory | Claudia Regina Gonçalves; Claudio Tavares Sacchi; Erica Valessa Ramos Gomes; Karoline Rodrigues Campos |
| EPI_ISL_747337, EPI_ISL_850198, EPI_ISL_2161051, EPI_ISL_2161155 | Division of Emerging Infectious Diseases, Bureau of Infectious Diseases Diagnosis Control, Korea Disease Control and Prevention Agency | Division of Emerging Infectious Diseases, Bureau of Infectious Diseases Diagnosis Control, Korea Disease Control and Prevention Agency | Ae Kyung Park; Chae Young Lee; Chaeyoung Lee; Eun-Jin Kim; Heui Man Kim; Il-Hwan Kim; Jeong-Ah Kim; Jeong-Min Kim; Jin Sun No; Namjoo Lee; Sang Hee Woo |
| EPI_ISL_2105306, EPI_ISL_2105360, EPI_ISL_2105365, EPI_ISL_2105433, EPI_ISL_2105541, EPI_ISL_2188278 | Dr. Jorge P. Royeca Hospital | Philippine Genome Center | Alethea R. de Guzman; Anna Ong-Lim; Arianne A. Zamora; Asia Louisa U. Chong; Benedict A. Maralit; Candice Francheska B. Tambaoan; Carlo M. Lapid; Celia Carlos; Devon Ray Pacial; Edsel Maurice Salvaña; El King D. Morado; Elcid Aaron R. Pangilinan; Eva Maria Cutiongco-de la Paz; Francis A. Tablizo; Irish Coleen A. Asin; Jaime C. Montoya; Jan Michael C. Yap; Jo-Hannah S. Llames; John Q. Wong; Joshua Gregor A. Dizon; Juan Antonio R. Magalang; Karol Sophia Agape R. Padilla; Kenneth M. Kim; Kris P. Punayan; Marc Edsel C. Ayes; Marc Jerrone R. Castro; Maria Rosario Singh-Vergeire and Cynthia P. Saloma; Maria Sofia L. Yangzon; Marissa Alejandria; Razel Nikka M. Hao; Rianna Patricia S. Cruz; Sheila Mae M. Araiza |
| EPI_ISL_1063789 | Evandro Chagas Institute | Evandro Chagas Institute Virology | A.M.; Barbagelata; E.C.; E.M.A.; Ferreira; J.A.; Junior; K.C.; L.C.; L.S.; M.C.; P.S.; Pinheiro; Santos; Silva; Sousa; Sousa Junior; W.D.C.; da Silva |
| EPI_ISL_1469555, EPI_ISL_1469572, EPI_ISL_1469620, EPI_ISL_1469661 | FUNDACAO DE SAUDE PUBLICA DE NOVO HAMBURGO FSNH | Epiclin | Ana Paula Muterle; Carolina Comerlato; Eliana Márcia Da Ros Wendland; Fernando Hayashi Sant'Anna; Janira Prichula; Juliana Comerlato |
| EPI_ISL_1469704, EPI_ISL_1469723 | FUNDACAO DE SAUDE PUBLICA SAO CAMILO DE ESTEIO | Epiclin | Ana Paula Muterle; Carolina Comerlato; Eliana Márcia Da Ros Wendland; Fernando Hayashi Sant'Anna; Janira Prichula; Juliana Comerlato |
| EPI_ISL_1469586 | FUNDACAO HOSPITALAR SAO JOSE | Epiclin | Ana Paula Muterle; Carolina Comerlato; Eliana Márcia Da Ros Wendland; Fernando Hayashi Sant'Anna; Janira Prichula; Juliana Comerlato |
| EPI_ISL_2105553 | First Aide Diagnostic Center | Philippine Genome Center | Alethea R. de Guzman; Anna Ong-Lim; Arianne A. Zamora; Asia Louisa U. Chong; Benedict A. Maralit; Candice Francheska B. Tambaoan; Carlo M. Lapid; Celia Carlos; Devon Ray Pacial; Edsel Maurice Salvaña; El King D. Morado; Eva Maria Cutiongco-de la Paz; Francis A. Tablizo; Irish Coleen A. Asin; Jaime C. Montoya; Jan Michael C. Yap; Jo-Hannah S. Llames; John Q. Wong; Joshua Gregor A. Dizon; Juan Antonio R. Magalang; Karol Sophia Agape R. Padilla; Kenneth M. Kim; Kris P. Punayan; Marc Edsel C. Ayes; Marc Jerrone R. Castro; Maria Rosario Singh-Vergeire and Cynthia P. Saloma; Maria Sofia L. Yangzon; Marissa Alejandria; Razel Nikka M. Hao; Rianna Patricia S. Cruz; Sheila Mae M. Araiza |
| EPI_ISL_431180, EPI_ISL_431240 | Fujian Center for Disease Control and Prevention | Fujian Center for Disease Control and Prevention | Huang Zhimiao; Lin Qi; Weng Yuwei; Zhang Yanhua |
| EPI_ISL_1182550, EPI_ISL_1182587, EPI_ISL_1182601, EPI_ISL_1182602, EPI_ISL_1182609, EPI_ISL_1182612 | Fundação Ezequiel Dias (FUNED) | Coordenação Geral de Laboratórios de Saúde Pública (CGLAB/DAEVS/SVS/MS) | Vagner Fonseca; et al. |
| EPI_ISL_1469713, EPI_ISL_1469845 | Fundação Hospitalar de Sapucaia do Sul | Epiclin | Ana Paula Muterle; Carolina Comerlato; Eliana Márcia Da Ros Wendland; Fernando Hayashi Sant'Anna; Janira Prichula; Juliana Comerlato |
| EPI_ISL_1469561 | Fundação de Saúde Pública São Camilo de Esteio | Epiclin | Ana Paula Muterle; Carolina Comerlato; Eliana Márcia Da Ros Wendland; Fernando Hayashi Sant'Anna; Janira Prichula; Juliana Comerlato |
| EPI_ISL_3320805 | Furst Medical Laboratory | Norwegian Institute of Public Health, Department of Virology | Atiya R Ali; Debech Nadia; Engebretsen Serina Beate; Garcia Llorente Ignacio; Hilde Elshaug; Hilde Vollan; Jon Bråte; Kamilla Heddeland Instefjord; Karoline Bragstad; Kathrine Stene-Johansen; Line Victoria Moen; Marie Paulsen Madsen; Olav Hungnes; Pedersen Benedikte Nevjen; Rasmus Riis Kopperud |
| EPI_ISL_746686, EPI_ISL_1167743, EPI_ISL_1321466 | Genetica Molecular and Subdepartamento de Virologia ISP Chile | Instituto de Salud Publica de Chile | Andres Castillo; Barbara Parra; Gisselle Barra; Jaime Lagos; Javier Tognarelli; Jorge Fernandez; Karen Orostica; Loredana Arata; Patricia Bustos; Rodrigo Fasce |
| EPI_ISL_861914 | Genomika Einstein | LATE - Laboratório de Técnicas Especiais - Hospital Israelita Albert Einstein | Ana Paula Moreira Salles; Deyvid Amgarten; Fernanda de Mello Malta; João Bosco Oliveira Filho; João Renato Rebello Pinho; Pedro Henrique Sebe Rodrigues; Raquel Riyuzo |
| EPI_ISL_960776 | Germano de sousa | Instituto Gulbenkian de Ciencia | Cathy Paulino; João Costa; João Sobral; Maria Costa; Ricardo Leite; Susana Ladeiro |
| EPI_ISL_745827, EPI_ISL_745855 | Ginkgo Bioworks Clinical Laboratory | Utah Public Health Laboratory | Alex Plocik; Becky Schilling; Birgitte Simen; David R. Hillyard; E. Susan Slechta; Erin L. Young; James McGann; Jeffrey B. Stevenson; Jim Griffin; Keith Robison; Kelly Oakeson; Malaika McKenzie-Bennett; Martha Pierson; Melanie A. Mallory; Michael T. Pyne; Michelle Spencer; Rebecca Littlefield; Salika M. Shakir; Tara Gallagher |
| EPI_ISL_1503139 | Gorgas Memorial Laboratory of Health Studies | Gorgas Memorial Laboratory of Health Studies | Castillo Jorge; Franco Danilo; Gonzalez Claudia; Jessica Gondola; Leyda Abrego; Lopez-Verges Sandra; Marlenne Castillo; Martinez Alexander; Moreno Ambar; Moreno Brechla; Oris Chavarria; Ortiz Alma |
| EPI_ISL_508266 | Government Medical College | National Institute of Biomedical Genomics | Arindam Maitra; Dhaval Khatri; Jyoti Iravane; Maitrik Dave; Saumitra Das |
| EPI_ISL_2105471, EPI_ISL_2105483, EPI_ISL_2105502, EPI_ISL_2105555 | Governor Celestino Gallares Memorial Hospital | Philippine Genome Center | Alethea R. de Guzman; Anna Ong-Lim; Arianne A. Zamora; Asia Louisa U. Chong; Benedict A. Maralit; Candice Francheska B. Tambaoan; Carlo M. Lapid; Celia Carlos; Devon Ray Pacial; Edsel Maurice Salvaña; El King D. Morado; Eva Maria Cutiongco-de la Paz; Francis A. Tablizo; Irish Coleen A. Asin; Jaime C. Montoya; Jan Michael C. Yap; Jo-Hannah S. Llames; John Q. Wong; Joshua Gregor A. Dizon; Juan Antonio R. Magalang; Karol Sophia Agape R. Padilla; Kenneth M. Kim; Kris P. Punayan; Marc Edsel C. Ayes; Marc Jerrone R. Castro; Maria Rosario Singh-Vergeire and Cynthia P. Saloma; Maria Sofia L. Yangzon; Marissa Alejandria; Razel Nikka M. Hao; Rianna Patricia S. Cruz; Sheila Mae M. Araiza |
| EPI_ISL_1040815 | Groote Schuur Hospital wc GSH | NHLS/UCT | Arash Iranzadeh; Bruna Galvao; Carolyn Williamson; Deelan Doolabh; Diana Hardie; Innocent Mudau; Kruger Marais; Lynn Tyers; Marvin Hsiao; Stephen Korsman |
| EPI_ISL_3127913 | Guam Public Health Laboratory | Centers for Disease Control and Prevention Division of Viral Diseases, Pathogen Discovery | Alex Burgin; Ben L. Rambo-Martin; Clinton R. Paden; Dakota Howard; Dave Wentworth; Dhwani Batra; Jasmine Padilla; Justin Lee; Krista Queen; Kristen Knipe; Kristine Lacek; Mark Burroughs; Matthew Schmerer; Meghan Bentz; Mili Sheth; Peter Cook; Sam Shepard; Sarah Nobles; Suxiang Tong; Vivien Dugan; Yvette Unoarumhi |
| EPI_ISL_732044, EPI_ISL_732052 | H Beatriz Angelo | Instituto Nacional de Saude (INSA) | Borges et al |
| EPI_ISL_1117216 | H Guimaraes | Instituto Nacional de Saude (INSA) | Borges et al |
| EPI_ISL_1172014 | HC_FMUSP | Laboratório de Parasitologia Médica - Instituto de Medicina Tropical - Universidade de São Paulo | Brazil-UK Centre for Arbovirus Discovery Diagnosis Genomics and Epidemiology (CADDE) Genomic Network - Instituto de Medicina Tropical |
| EPI_ISL_3912417 | HGF HOSPITAL GERAL DE | Analytical Competence Molecular Epidemiology | Cleber Furtado Aksenen; Fabio Miyajima; Fernando Braga Stehling; Francisco Eder de Moura Lopes; Jamille Maria Mendes Bezerra; Joaquim Cesar do Nascimento Sousa Junior; Pedro Miguel Carneiro Jeronimo; Suzana Porto Almeida & Lucas Delerino on behalf of COVID-19 FIOCRUZ Genomic Network; Thais Ferreira de Oliveira; Thais de Oliveira Costa; Ticiane Cavalcante de Souza; Veridiana Pessoa Miyajima |

| | | | |
|---|---|---|---|
| | FORTALEZA | Lab/ACME, Oswaldo Cruz Foundation, Ceara (FIOCRUZ CE) | |
| EPI_ISL_2017243, EPI_ISL_2017251, EPI_ISL_2017253, EPI_ISL_2017258, EPI_ISL_2017259, EPI_ISL_2017264, EPI_ISL_2017266, EPI_ISL_2017271, EPI_ISL_2017275, EPI_ISL_2017276, EPI_ISL_2017309, EPI_ISL_2017457, EPI_ISL_2102517, EPI_ISL_2187714, EPI_ISL_2187726, EPI_ISL_2187746, EPI_ISL_2187747, EPI_ISL_2187783, EPI_ISL_2227560, EPI_ISL_2348593, EPI_ISL_2497434, EPI_ISL_2617626, EPI_ISL_3259351 | | | |
| see above | HLAGYN - Laboratorio de Imunologia de Transplantes de Goias | HLAGYN - Laboratorio de Imunologia de Transplantes de Goias | Alessandro Leonardo Alvares Magalhaes; Daniel Ferreira de Sousa; Danielle de Paiva Rezende; Erika Lopes Rocha Batista; Fernando Antonio Vinhal dos Santos; Frederico Rodrigues Vinhal; Lucas Carlos Gomes Pereira; Paola Cristina Resende Silva; Raphael Bessa Parmigiane; Sabrina Sara Moreira Duarte |
| EPI_ISL_1912832, EPI_ISL_1912892 | HMH | New York Genome Center | Andre Corvelo; Barry Kreiswirth; David Perlin; Dayna M. Oschwald; Jose Mediavilla; Kaelea Composto; Kar Chow; Liang Chen; Marcus Cunningham; Michael Zody; Samantha Fennessey; Tom Maniatis |
| EPI_ISL_1684086 | HOSPITAL CLINIC | Instituto de Salud Carlos III | A. Monzón; F. Casas; I. Jiménez; I.VILA ESTAPE; Iglesias-Caballero; JORDI; M. Sandonís; P. Zaballos; S. Camarero; S. Cuesta; S. Pozo; S. Varona; V. Vázquez-Morón |
| EPI_ISL_3912420 | HOSPITAL DE CAMPANHA COVID 19 DR EUDASIO BARROSO HMEB | Analytical Competence Molecular Epidemiology Lab/ACME, Oswaldo Cruz Foundation, Ceara (FIOCRUZ CE) | Cleber Furtado Aksenen; Fabio Miyajima; Fernando Braga Stehling; Francisco Eder de Moura Lopes; Jamille Maria Mendes Bezerra; Joaquim Cesar do Nascimento Sousa Junior; Pedro Miguel Carneiro Jeronimo; Suzana Porto Almeida & Lucas Delerino on behalf of COVID-19 FIOCRUZ Genomic Network; Thais Ferreira de Oliveira; Thais de Oliveira Costa; Ticiane Cavalcante de Souza; Veridiana Pessoa Miyajima |
| EPI_ISL_3912421 | HOSPITAL E MATERNIDADE NOSSA SENHORA DOS MILAGRES | Analytical Competence Molecular Epidemiology Lab/ACME, Oswaldo Cruz Foundation, Ceara (FIOCRUZ CE) | Cleber Furtado Aksenen; Fabio Miyajima; Fernando Braga Stehling; Francisco Eder de Moura Lopes; Jamille Maria Mendes Bezerra; Joaquim Cesar do Nascimento Sousa Junior; Pedro Miguel Carneiro Jeronimo; Suzana Porto Almeida & Lucas Delerino on behalf of COVID-19 FIOCRUZ Genomic Network; Thais Ferreira de Oliveira; Thais de Oliveira Costa; Ticiane Cavalcante de Souza; Veridiana Pessoa Miyajima |
| EPI_ISL_1469746 | HOSPITAL MONTENEGRO | Epiclin | Ana Paula Muterle; Carolina Comerlato; Eliana Márcia Da Ros Wendland; Fernando Hayashi Sant'Anna; Janira Prichula; Juliana Comerlato |
| EPI_ISL_3912424 | HOSPITAL MUNICIPAL DE CAMPOS SALES | Analytical Competence Molecular Epidemiology Lab/ACME, Oswaldo Cruz Foundation, Ceara (FIOCRUZ CE) | Cleber Furtado Aksenen; Fabio Miyajima; Fernando Braga Stehling; Francisco Eder de Moura Lopes; Jamille Maria Mendes Bezerra; Joaquim Cesar do Nascimento Sousa Junior; Pedro Miguel Carneiro Jeronimo; Suzana Porto Almeida & Lucas Delerino on behalf of COVID-19 FIOCRUZ Genomic Network; Thais Ferreira de Oliveira; Thais de Oliveira Costa; Ticiane Cavalcante de Souza; Veridiana Pessoa Miyajima |
| EPI_ISL_2345448 | HOSPITAL MUNICIPAL DE IBIUNA IBIUNA SP | Instituto Butantan / ESALQ-Piracicaba | Antonio Jorge Martins; Claudia Renata dos Santos Barros; David Schlesinger; Debora Botequio Moretti; Dimas Tadeu Covas; Elaine Cristina Marqueze; Elaine Vieira Santos; Evandra Strazza Rodrigues; Heidge Fukumasu; Jayme Augusto de Souza-Neto; José Salvatore Leister Patané; Luiz Alcantara; Luiz Lehmann Coutinho; Maria Carolina Elias; Maurício Lacerda Nogueira; Rafael dos Santos Bezerra; Raul Machado Neto; Rejane Maria Tommasini Grotto; Ricardo Haddad; Sandra Coccuzzo Sampaio Vessoni; Simone Kashima; Svetoslav Nanev Slavov; Vincent Louis Viala |
| EPI_ISL_1966922 | HOSPITAL MUNICIPAL DE ITABERA | Instituto Butantan / ESALQ-USP (Piracicaba) | Antonio Jorge Martins; Bianca Cechetto Carlos. Mendelics: Bibiana Santos; Claudia Renata dos Santos Barros; Cíntia Bittar; David Schlesinger. Hemocentro Ribeirão Preto: Simone Kashima; Debora Botequio Moretti; Elaine Cristina Marqueze; Elaine Vieira dos Santos; Elisangela Chicaroni Mattos; Erika Freitas; Evandra Strazza Rodrigues; Felipe Allan da Silva da Costa; Flavia Aburjaile; Fábio Sossai Possebon; Guilherme Campos; Guilherme Targino Valente; Heidge Fukumasu. USP-Botucatu: Rejane Maria Tommasini Grotto; Helena Lage Ferreira; Instituto Butantan: Dimas Tadeu Covas; Jardelina de Souza Todao Bernardino; Jayme A. Souza-Neto; Jessika Cristina Chagas Lesbon; Jorge A. Petroli Marchesi; José Salvatore Leister Patané; João Paulo Kitajima; João Pessoa Araújo Jr.; Leila Sabrina Ullmann; Loyze Paola Oliveira de Lima; Luiz Aurelio de Campos Crispin. Centro de Genômica Funcional da ESALQ: Luiz Lehmann Coutinho; Luiz Carlos Junior de Alcantara; Lívia Sacchetto; Maisa C. Pereira Parra; Maria Carolina Elias; Marta Giovanetti; Marília Moraes; Maurício Lacerda Nogueira. Prefeitura de Sao Paulo: Melissa Palmieri.; Patricia Akemi Assato; Paula Rahal; Paulo Inacio da Costa; Rafael dos Santos Bezerra; Raquel de Lello Rocha Campos Cassano. NGS Soluções Genômicas: Pilar Drummond Sampaio Corrêa Mariani. FZEA-USP Pirassununga: Mirele Daiana Poleti; Raul Machado Neto; Ricardo Augusto Brassaloti; Ricardo Haddad; Rodrigo Tocantins Calado. FAMERP-SJRP: Cecília Artico Banho; Sandra Coccuzzo Sampaio; Svetoslav Nanev Slavov; Vagner Fonseca; Vincent Louis Viala |
| EPI_ISL_1469624, EPI_ISL_1469631 | HOSPITAL MUNICIPAL GETULIO VARGAS | Epiclin | Ana Paula Muterle; Carolina Comerlato; Eliana Márcia Da Ros Wendland; Fernando Hayashi Sant'Anna; Janira Prichula; Juliana Comerlato |
| EPI_ISL_1510080, EPI_ISL_1510081 | HOSPITAL SAN PEDRO DE ALCANTARA - C.D. 05 | Instituto de Salud Carlos III | A. Monzón; F. Casas; GUADALUPE; I. Jiménez; I.RODRIGUEZ RODRIGUEZ; Iglesias-Caballero; M. Sandonís; P. Zaballos; S. Camarero; S. Cuesta; S. Pozo; S. Varona; V. Vázquez-Morón |
| EPI_ISL_2758675, EPI_ISL_2758699, EPI_ISL_2758700, EPI_ISL_2758702, EPI_ISL_2758703, EPI_ISL_2758704, EPI_ISL_2758708, EPI_ISL_2758712, EPI_ISL_2758767, EPI_ISL_2758768, EPI_ISL_2758769, EPI_ISL_2758770, EPI_ISL_2758774, EPI_ISL_2758775 | | | |
| see above | HUEM/IBMP | IPEC Guarapuava | NAPI-Genômica (Novos Arranjo de Pesquisa e Inovação em Genômica): Ademar Dantas da Cunha Júnior Adriano Ferrasa Adriano Mondini Aldo Przybysz Alessandra Lourenço Cecchini Armani Alex Sandro Jorge Alexandra Ivo de Medeiros Alexandre Maller Aline Cristina Batista Rodrigues Johann Ana Lucia Ferreira Ana Marisa Fusco Almeida Anderson Joel Martino Andrade André Luís Laforga Vanzela Andrea Duarte Doetzer Andrea Name Colado Simao Andressa Pereira de Souza Anelisa Ramão Angelica Beate Winter Boldt Anna Hermínia Castro Gomes de Amorim Anna Silvia Penteado Setti da Rocha Antonio Camilo da Silva Filho Antonio Stabelini Neto Arthur Hirata Bertachi Barbara Mendes Paz Chao Betty Cristiane Kuhn Bruno Ambrozio Galindo Bruno Ribeiro Cruz Camilla Reginatto De Pierri Carla Fredrichsen Moya Araujo Carla Fredrichsen Moya Araujo Carlos Alberto Oliveira de Biagi Junior Carlos Augusto Nassar Carlos Eduardo Buss Carlos Gilberto Carlotti Junior Carlos Henrique Schneider Carolina Panis Carolina Weigert Galvão Caroline de Jesus Coelho Donha Caroline Guisantes de Salvo Toni Caryna Eurich Mazur Catiuscie Cabreira da Silva Tortorella Celso F. D. Doliveira Cesar Luiz Boguszewski Christiane Pienna Soares Chung Man Chin Claudia Moro Cleverson Busso Cristiane Cominetti Daiane Priscila Simão-Silva Dalila Luciola Zanette Daniel de Paula Daniel de Paula Daniel Rech Daniela Fiori Gradia Daniela Pretti da Cunha Tirapelli Daniela Viganó Zanoti Jeronymo Daniele Ukan Danielle Malheiros Ferreira Danielle Venturini Deborah Catharine de Assis Leite Deivid Calebe de Souza Dennis Armando Bertolini Edenir Inez Pamero Edna Maria Vissoci Reiche Edson Roberto Arpini Miguel Eduardo José de Almeida Araújo Eliana Carolina Vespero Eliandro Reis Tavares Elza Kimura Grimshaw Emanuel Maltempi de Souza Emanuele Cristina Gustani Buss Émerson Carraro Emiliana Cristina Melo ENILze Maria de Souza Fonseca Ribeiro Enilze Maria de Souza Fonseca Ribeiro Erika Izumi Erika Seki Kioshima Cotica Evani Marques Pereira Fabio Negretti Fábio Rodrigues Ferreira Seiva Felipe Dunin dos Santos Felipe Tuon Fernanda Andreia Rosa Fernanda Cestaro Prado Cortez Fernanda Ivanski Fernanda Maris Peria Flavia Regina Oliveira de Barros Franciele Aní Caovilla Follador Franciele Mara Lucca Zanardo Bohm Francinete Ramos Campos Fulviana Silva Nishiyama GABRIEL RIBEIRO CORDEIRO Gabriela Datsch Bennemann Gisele Santos de Oliveira Glaucio Valdameri Glauco Akelinghton Freire Vitiello Glauco Vieira Miranda Glaura Scantamburlo ALves Fernandes Guilherme Ferreira Silveira Gustavo Bianchini Porfírio Gustavo Lenci Marques Hélito Volpato Hildebrando Masshiro Nagai Huei Diana Lee Ilce Mara de Syllos Cólus Iris Rabinovich Israel Gomy Jackson Kawakami Jacques Duílio Brancher Jaime Luís Lopes Rocha Jaqueline Carvalho de Oliveira Jean Henrique da Silva Rodrigues Jean Leandro dos Santos Jeane Eliete Laguila Visentainer João Paulo Bianchi Ximenez Joaquim Manoel da Silva Jociani Ascari Joel Donazzolo Jorge Luis Maria Ruiz Jose Knopfholz José Luis da Conceição Silva José Sebastião dos Santos Joseane Carla Schabarum Juliana Cheleski Wiggers Juliana Mara Serpeloni Juliana Morini Küpper Cardoso Perseguini Karen Brajão de Oliveira Karin Braun Prado Karine Aparecida de Lima Katiany Rizzieri Caleffi Ferracioli Katiuscia de Oliveira Francisco Gabriel Kelvinson Fernandes Viana Larissa Beatriz Cossalter Larissa Danielle Bahls Pinto Laurival Antonio Vilas Boas Léia Carolina Lucio Líbero Mezzadri Neto Ligia Carla Faccin Galhardi Lirane Elize Defante Ferreto Luciana Furlaneto Maia Luciana Oliveira de Fariña Luciana Reis Azevedo Alanis Luciane Regina Cavalli Lucy Megumi Yamauchi Lioni Luis Paulo Gomes Mascarenhas Luis Paulo Gomes Mascarenhas Luís Paulo Mascarenhas Lupe Furtado Alle Lyvia Regina Biagi Silva Bertachi Mara Antonia Ramos Costa Mara L. Cordeiro Marcela Maria Birolim Marcelo Ricardo Vicari Marcia Edilaine Lopes Consolaro Marcia Holsbach Beltrame Marcia Regina Eches Perugini Marcos Abdo Arbex Marcos Pileggi MARCOS TADEU GRZELCZAK Marcus Peikriszwili Tartaruga Maria Angelica Ehara Watanabe Maria Antonia Ramos Costa Maria Claudia Gross Maria José Soares Mendes Giannini Maria Leandra Terencio Maria Lúcia Bonfleur Maria Luiza Guimarães de Oliveira Maria Luiza Petzl-Erler Mariana Abe Vicente Cavagnari Marina Kimiko Kadowaki Marise Fonseca dos Santos Marla Karine Amarante Maurício Turkiewicz Mauro Antonio Alves Castro Michel Rodrigo Zambrano Passarini Michele Potrich Michelle Orane Schemberger Milena Massumi Kozonoe Mônica Degraf Cavallin Monica Tereza Suldofski Mucio Luiz de Assis Cirino Nadia Graciele Krohn Najeh Maissar Khalil Nédia de Castilhos Ghisi Neide Tomimura Costa Neiva Leite Neyva Maria Lopes Romeiro Patrícia Amâncio da Rosa Patricia Dayane Carvalho Schaker Patricia Oehlmeyer Nassar Patrícia Savio de Araújo-Souza Patricia Silva Lucio Paulo Henrique Couto Souza Paulo Roberto Donadio Percy Nohama Quirino Alves de Lima Neto Rafael Deminice Rafael dos Santos Bezerra Raquel Alves dos Santos Renan Manozzo Galante Renata Ernlund Freitas de Macedo Rita de Cássia Garcia Simão Roberta Losi Guembarovski Roberto H. Herai Roberto Rosati Rodrigo Ferreira Rodrigo Rodrigues Matiello Rogério Neri Shinsato Rogério Pincela Mateus Rosane Aparecida Ribeiro Rosilene Fressatti Cardoso Rosilene Fressatti Cardoso Sandra Mara Guse Scós Venske Selene Elifio Esposito Sérgio Ossamu Ioshii Silvana Giuliatti Silvia Mara de Souza Halick Silvio Henrique Maia de Almeida Simone Neumann Wendt Spencer Luiz Marques Payão Stefan Wolanski Negrão Stephane Janaina de Moura Escobar Sueli Fumie Yamada Ogatta SUELI PERCIO QUINAIA Taciane Finatto Tatiana Mayumi Veiga Iriyoda Tayza Katelline Danilau Ostroski Tony Alexander Hild Valeria Valente Vanessa Nascimento Kozak Vanessa Santos Sotomaior Victor Breno Pedrosa Victoria Zeghbi Cochenski Borba Vivian Rotuno Moure Valdameri Wander Rogerio Pavanelli Weber Cláudio Francisco Nunes da Silva Willian Augusto de Melo Yohandra Reyes Torres |
| EPI_ISL_2758771 | HUEM/LACEN | IPEC Guarapuava | NAPI-Genômica (Novos Arranjo de Pesquisa e Inovação em Genômica): Ademar Dantas da Cunha Júnior Adriano Ferrasa Adriano Mondini Aldo Przybysz Alessandra Lourenço Cecchini Armani Alex Sandro Jorge Alexandra Ivo de Medeiros Alexandre Maller Aline Cristina Batista Rodrigues Johann Ana Lucia Ferreira Ana Marisa Fusco Almeida Anderson Joel Martino Andrade André Luís Laforga Vanzela Andrea Duarte Doetzer Andrea Name Colado Simao Andressa Pereira de Souza Anelisa Ramão Angelica Beate Winter Boldt Anna Hermínia Castro Gomes de Amorim Anna Silvia Penteado Setti da Rocha Antonio Camilo da Silva Filho Antonio Stabelini Neto Arthur Hirata Bertachi Barbara Mendes Paz Chao Betty Cristiane Kuhn Bruno Ambrozio Galindo Bruno Ribeiro Cruz Camilla Reginatto De Pierri Carla Fredrichsen Moya Araujo Carla Fredrichsen Moya Araujo Carlos Alberto Oliveira de Biagi Junior Carlos Augusto Nassar Carlos Eduardo Buss Carlos Gilberto Carlotti Junior Carlos Henrique Schneider Carolina Panis Carolina Weigert Galvão Caroline de Jesus Coelho Donha Caroline Guisantes de Salvo Toni Caryna Eurich Mazur Catiuscie Cabreira da Silva Tortorella Celso F. D. Doliveira Cesar Luiz Boguszewski Christiane Pienna Soares Chung Man Chin Claudia Moro Cleverson Busso Cristiane Cominetti Daiane Priscila Simão-Silva Dalila Luciola Zanette Daniel de Paula Daniel de Paula Daniel Rech Daniela Fiori Gradia Daniela Pretti da Cunha Tirapelli Daniela Viganó Zanoti Jeronymo Daniele Ukan Danielle Malheiros Ferreira Danielle Venturini Deborah Catharine de Assis Leite Deivid Calebe de Souza Dennis Armando Bertolini Edenir Inez Pamero Edna Maria Vissoci Reiche Edson Roberto Arpini Miguel Eduardo José de Almeida Araújo Eliana Carolina Vespero Eliandro Reis Tavares Elza Kimura Grimshaw Emanuel Maltempi de Souza Emanuele Cristina Gustani Buss Émerson Carraro Emiliana Cristina Melo ENILze Maria de Souza Fonseca Ribeiro Enilze Maria de Souza Fonseca Ribeiro Erika Izumi Erika Seki Kioshima Cotica Evani Marques Pereira Fabio Negretti Fábio Rodrigues Ferreira Seiva Felipe Dunin dos Santos Felipe Tuon Fernanda Andreia Rosa Fernanda Cestaro Prado Cortez Fernanda Ivanski Fernanda Maris Peria Flavia Regina Oliveira de Barros Franciele Aní Caovilla Follador Franciele Mara Lucca Zanardo Bohm Francinete Ramos Campos Fulviana Silva Nishiyama GABRIEL RIBEIRO CORDEIRO Gabriela Datsch Bennemann Gisele Santos de Oliveira Glaucio Valdameri Glauco Akelinghton Freire Vitiello Glauco Vieira Miranda Glaura Scantamburlo ALves Fernandes Guilherme Ferreira Silveira Gustavo Bianchini Porfírio Gustavo Lenci Marques Hélito Volpato Hildebrando Masshiro Nagai Huei Diana Lee Ilce Mara de Syllos Cólus Iris Rabinovich Israel Gomy Jackson Kawakami Jacques Duílio Brancher Jaime Luís Lopes Rocha Jaqueline Carvalho de Oliveira Jean Henrique da Silva Rodrigues Jean Leandro dos Santos Jeane Eliete Laguila Visentainer João Paulo Bianchi Ximenez Joaquim Manoel da Silva Jociani Ascari Joel Donazzolo Jorge Luis Maria Ruiz Jose Knopfholz José Luis da Conceição Silva José Sebastião dos Santos Joseane Carla Schabarum Juliana Cheleski Wiggers Juliana Mara Serpeloni Juliana Morini Küpper Cardoso Perseguini Karen Brajão de Oliveira Karin Braun Prado Karine Aparecida de Lima Katiany Rizzieri Caleffi Ferracioli Katiuscia de Oliveira Francisco Gabriel Kelvinson Fernandes Viana Larissa Beatriz Cossalter Larissa Danielle Bahls Pinto Laurival Antonio Vilas Boas Léia Carolina Lucio Líbero Mezzadri Neto Ligia Carla Faccin Galhardi Lirane Elize Defante Ferreto Luciana Furlaneto Maia Luciana Oliveira de Fariña Luciana Reis Azevedo Alanis Luciane Regina Cavalli Lucy Megumi Yamauchi Lioni Luis Paulo Gomes Mascarenhas Luis Paulo Gomes Mascarenhas Luís Paulo Mascarenhas Lupe Furtado Alle Lyvia Regina Biagi Silva Bertachi Mara Antonia Ramos Costa Mara L. Cordeiro Marcela Maria Birolim Marcelo Ricardo Vicari Marcia Edilaine Lopes Consolaro Marcia Holsbach Beltrame Marcia Regina Eches Perugini Marcos Abdo Arbex Marcos Pileggi MARCOS TADEU GRZELCZAK Marcus Peikriszwili Tartaruga Maria Angelica Ehara Watanabe Maria Antonia Ramos Costa Maria Claudia Gross Maria José Soares Mendes Giannini Maria Leandra Terencio Maria Lúcia Bonfleur Maria Luiza Guimarães de Oliveira Maria Luiza Petzl-Erler Mariana Abe Vicente Cavagnari Marina Kimiko Kadowaki Marise Fonseca dos Santos Marla Karine Amarante Maurício Turkiewicz Mauro Antonio Alves Castro Michel Rodrigo Zambrano Passarini Michele Potrich Michelle Orane Schemberger Milena Massumi Kozonoe Mônica Degraf Cavallin Monica Tereza Suldofski Mucio Luiz de Assis Cirino Nadia Graciele Krohn Najeh Maissar Khalil Nédia de Castilhos Ghisi Neide Tomimura Costa Neiva Leite Neyva Maria Lopes Romeiro Patrícia Amâncio da Rosa Patricia Dayane Carvalho Schaker Patricia Oehlmeyer Nassar Patrícia Savio de Araújo-Souza Patricia Silva Lucio Paulo Henrique Couto Souza Paulo Roberto Donadio Percy Nohama Quirino Alves de Lima Neto Rafael Deminice Rafael dos Santos Bezerra Raquel Alves dos Santos Renan Manozzo Galante Renata Ernlund Freitas de Macedo Rita de Cássia Garcia Simão Roberta Losi Guembarovski Roberto H. Herai Roberto Rosati Rodrigo Ferreira Rodrigo Rodrigues Matiello Rogério Neri Shinsato Rogério Pincela Mateus Rosane Aparecida Ribeiro Rosilene Fressatti Cardoso Rosilene Fressatti Cardoso Sandra Mara Guse Scós Venske Selene Elifio Esposito Sérgio Ossamu Ioshii Silvana Giuliatti Silvia Mara de Souza Halick Silvio Henrique Maia de Almeida Simone Neumann Wendt Spencer Luiz Marques Payão Stefan Wolanski Negrão Stephane Janaina de Moura Escobar Sueli Fumie Yamada Ogatta SUELI PERCIO QUINAIA Taciane Finatto Tatiana Mayumi Veiga Iriyoda Tayza Katelline Danilau Ostroski Tony Alexander Hild Valeria Valente Vanessa Nascimento Kozak Vanessa Santos Sotomaior Victor Breno Pedrosa Victoria Zeghbi Cochenski Borba Vivian Rotuno Moure Valdameri Wander Rogerio Pavanelli Weber Cláudio Francisco Nunes da Silva Willian Augusto de Melo Yohandra Reyes Torres |
| EPI_ISL_1268114, EPI_ISL_1269948, EPI_ISL_1270371, EPI_ISL_1297937, EPI_ISL_2320148 | Helix/Illumina | Centers for Disease Control and Prevention Division of Viral Diseases, Pathogen Discovery | Adrian Paskey; Alexandre Bolze; Ary Ascencio; Ben L. Rambo-Martin; Benjamin Rambo-Martin; Brad Sickler; Charlotte Rivera-Garcia; Christine Tran; Christopher Gulvick; Clinton R. Paden; Dakota Howard; Darlene Wagner; David Becker; Dhwani Batra; Duncan MacCannell; Efren Sandoval; Eileen de Feo; Elizabeth Cirulli; Eric Allen; Geraint Levan; James Lu; Jan Antico; Jason Caravas; Jason Nguyen; Jimmy Ramirez; Jingtao Liu; Kara Moser; Kelly Schiabor Barrett; Kim Gietzen; Magnus Isaksson; Marc Laurent; Matthew Schmerer; Matthew Tolentino; Nicole L. Washington; Peter W. Cook; Phil Febbo; Ryan Cho; Scott Sammons; Shannon Wickline; Shatavia Morrison; Sherry Wang; Simon White; Summer Galloway; Suxiang Tong; Tyler Cassens; William Lee; Yvette Unoarumhi |
| EPI_ISL_471562, EPI_ISL_471581, EPI_ISL_515564, EPI_ISL_523977 | Hosp. Municipal Prof. Dr. Alípio Corrêa Netto | Instituto Adolfo Lutz, Interdiciplinary Procedures Center, Strategic Laboratory | Claudia Regina Gonçalves; Claudio Tavares Sacchi; Erica Valessa Ramos Gomes |
| EPI_ISL_693229 | Hospital 8 de Maio | Instituto Adolfo Lutz, Interdiciplinary Procedures Center, Strategic Laboratory | Claudia Regina Gonçalves; Claudio Tavares Sacchi; Erica Valessa Ramos Gomes; Karoline Rodrigues Campos |
| EPI_ISL_574580 | Hospital Cidade Tiradentes Carmen Prudente | Instituto Adolfo Lutz, Interdiciplinary Procedures Center, Strategic Laboratory | Claudia Regina Gonçalves; Claudio Tavares Sacchi; Erica Valessa Ramos Gomes; Karoline Rodrigues Campos |

| | | | |
|---|---|---|---|
| EPI_ISL_574591, EPI_ISL_574592, EPI_ISL_603030 | Hospital Domingos Leonardo Ceravolo Presidente Prudente | Instituto Adolfo Lutz, Interdiciplinary Procedures Center, Strategic Laboratory | Claudia Regina Gonçalves; Claudio Tavares Sacchi; Erica Valessa Ramos Gomes; Karoline Rodrigues Campos |
| EPI_ISL_574594 | Hospital Escola da Universidade de Taubate | Instituto Adolfo Lutz, Interdiciplinary Procedures Center, Strategic Laboratory | Claudia Regina Gonçalves; Claudio Tavares Sacchi; Erica Valessa Ramos Gomes; Karoline Rodrigues Campos |
| EPI_ISL_574588, EPI_ISL_583490 | Hospital Estadual Sumare | Instituto Adolfo Lutz, Interdiciplinary Procedures Center, Strategic Laboratory | Claudia Regina Gonçalves; Claudio Tavares Sacchi; Erica Valessa Ramos Gomes; Karoline Rodrigues Campos |
| EPI_ISL_583501 | Hospital Estadual de CampanhaCOVID 19 Barradas | Instituto Adolfo Lutz, Interdiciplinary Procedures Center, Strategic Laboratory | Claudia Regina Gonçalves; Claudio Tavares Sacchi; Erica Valessa Ramos Gomes; Karoline Rodrigues Campos |
| EPI_ISL_1647843, EPI_ISL_1647850 | Hospital General Universitario Gregorio Marañón | Hospital General Universitario Gregorio Marañón | Cristina Rodriguez-Grande; Darío García de Viedma; Laura Pérez-Lago; Patricia Muñoz; Pedro Sola Campoy; Pilar Catalán; Sergio Buenestado Serrano |
| EPI_ISL_852976 | Hospital General Universitario Gregorio Marañón | SeqCOVID-SPAIN consortium/IBV(CSIC) | Darío García de Viedma; Julia Suárez; Laura Pérez-Lago; Marta Herranz; Patricia Muñoz and SeqCOVID-SPAIN consortium; Pedro J Sola-Campoy; Pilar Catalán; Sergio Buenestado-Serrano; Victor Manuel de la Cueva |
| EPI_ISL_471541, EPI_ISL_523971 | Hospital Geral Santa Marcelina | Instituto Adolfo Lutz, Interdiciplinary Procedures Center, Strategic Laboratory | Claudia Regina Gonçalves; Claudio Tavares Sacchi; Erica Valessa Ramos Gomes |
| EPI_ISL_534317 | Hospital Geral de Itapevi | Instituto Adolfo Lutz, Interdiciplinary Procedures Center, Strategic Laboratory | Claudia Regina Gonçalves; Claudio Tavares Sacchi; Erica Valessa Ramos Gomes |
| EPI_ISL_603037 | Hospital Geral de Pedreira | Instituto Adolfo Lutz, Interdiciplinary Procedures Center, Strategic Laboratory | Claudia Regina Gonçalves; Claudio Tavares Sacchi; Erica Valessa Ramos Gomes; Karoline Rodrigues Campos |
| EPI_ISL_861636 | Hospital Geral de Sao Mateus São Paulo | Instituto Adolfo Lutz, Interdiciplinary Procedures Center, Strategic Laboratory | Claudia Regina Gonçalves; Claudio Tavares Sacchi; Erica Valessa Ramos Gomes; Karoline Rodrigues Campos |
| EPI_ISL_515555 | Hospital Geral de Vila Nova Cachoeirinha | Instituto Adolfo Lutz, Interdiciplinary Procedures Center, Strategic Laboratory | Claudia Regina Gonçalves; Claudio Tavares Sacchi; Erica Valessa Ramos Gomes |
| EPI_ISL_574595 | Hospital Geral de Vila Penteado Dr. Jose Pamgella | Instituto Adolfo Lutz, Interdiciplinary Procedures Center, Strategic Laboratory | Claudia Regina Gonçalves; Claudio Tavares Sacchi; Erica Valessa Ramos Gomes; Karoline Rodrigues Campos |
| EPI_ISL_523957 | Hospital Itamaraty | Instituto Adolfo Lutz, Interdiciplinary Procedures Center, Strategic Laboratory | Claudia Regina Gonçalves; Claudio Tavares Sacchi; Erica Valessa Ramos Gomes |
| EPI_ISL_524462 | Hospital Metropolitano | Instituto Adolfo Lutz, Interdiciplinary Procedures Center, Strategic Laboratory | Claudia Regina Gonçalves; Claudio Tavares Sacchi; Erica Valessa Ramos Gomes |
| EPI_ISL_3031336 | Hospital Metropolitano Dr. Célio de Castro | Instituto René Rachou / Fiocruz Minas | Anna Salim; Cristina Fonseca; Gabriel Fernandes; Mariana Melo; Núbia Fernandes; Pedro Alves; Rosiane Pereira; Rubens do Monte Neto; Sandra Gava; Thaís Santos; Thaís Silva; Wilma Patrícia Bernardes |
| EPI_ISL_515541, EPI_ISL_515561 | Hospital Montemagno | Instituto Adolfo Lutz, Interdiciplinary Procedures Center, Strategic Laboratory | Claudia Regina Gonçalves; Claudio Tavares Sacchi; Erica Valessa Ramos Gomes |
| EPI_ISL_534318, EPI_ISL_693208, EPI_ISL_693209 | Hospital Municipal Antonio Giglio | Instituto Adolfo Lutz, Interdiciplinary Procedures Center, Strategic Laboratory | Claudia Regina Gonçalves; Claudio Tavares Sacchi; Erica Valessa Ramos Gomes; Karoline Rodrigues Campos |
| EPI_ISL_861658 | Hospital Municipal Antônio Giglio | Instituto Adolfo Lutz, Interdiciplinary Procedures Center, Strategic Laboratory | Claudia Regina Gonçalves; Claudio Tavares Sacchi; Erica Valessa Ramos Gomes; Karoline Rodrigues Campos |
| EPI_ISL_471549 | Hospital Municipal Carmen Prudente | Instituto Adolfo Lutz, Interdiciplinary Procedures Center, Strategic Laboratory | Claudia Regina Gonçalves; Claudio Tavares Sacchi; Erica Valessa Ramos Gomes |
| EPI_ISL_1303544 | Hospital Municipal Cidade Tiradentes Carmen Prudente | Instituto Adolfo Lutz, Interdiciplinary Procedures Center, Strategic Laboratory | Caio Vinicius Dias Lopes; Claudia Regina Gonçalves; Claudio Tavares Sacchi; Erica Valessa Ramos Gomes; Karoline Rodrigues Campos |
| EPI_ISL_515562 | Hospital Municipal Doutor Alexandre Zaio | Instituto Adolfo Lutz, Interdiciplinary Procedures Center, Strategic Laboratory | Claudia Regina Gonçalves; Claudio Tavares Sacchi; Erica Valessa Ramos Gomes |
| EPI_ISL_693203 | Hospital Municipal Doutor Arthur Ribeiro de Saboya | Instituto Adolfo Lutz, Interdiciplinary Procedures Center, Strategic Laboratory | Claudia Regina Gonçalves; Claudio Tavares Sacchi; Erica Valessa Ramos Gomes; Karoline Rodrigues Campos |
| EPI_ISL_468311, EPI_ISL_468312 | Hospital Municipal Dr Ignacio Proenca de Gouvea | Instituto Adolfo Lutz, Interdiciplinary Procedures Center, Strategic Laboratory | Claudia Regina Gonçalves; Claudio Tavares Sacchi; Erica Valessa Ramos Gomes |
| EPI_ISL_523985 | Hospital Municipal Dr. Benedicto Montenegro | Instituto Adolfo Lutz, Interdiciplinary Procedures Center, Strategic Laboratory | Claudia Regina Gonçalves; Claudio Tavares Sacchi; Erica Valessa Ramos Gomes |
| EPI_ISL_882672 | Hospital Municipal Dr. Guido Guida | Instituto Adolfo Lutz, Interdiciplinary Procedures Center, Strategic Laboratory | Claudia Regina Gonçalves; Claudio Tavares Sacchi; Erica Valessa Ramos Gomes; Karoline Rodrigues Campos |
| EPI_ISL_515553, | Hospital Municipal | Instituto Adolfo Lutz, | Claudia Regina Gonçalves; Claudio Tavares Sacchi; Erica Valessa Ramos Gomes; Karoline Rodrigues Campos |

| | | | |
|---|---|---|---|
| EPI_ISL_574577, EPI_ISL_574579 | Dr. Ignacio Proença de Gouvea | Interdiciplinary Procedures Center, Strategic Laboratory | |
| EPI_ISL_515548, EPI_ISL_515563, EPI_ISL_574589 | Hospital Municipal Dr. Jose Soares Hungria | Instituto Adolfo Lutz, Interdiciplinary Procedures Center, Strategic Laboratory | Claudia Regina Gonçalves; Claudio Tavares Sacchi; Erica Valessa Ramos Gomes; Karoline Rodrigues Campos |
| EPI_ISL_861640, EPI_ISL_861641 | Hospital Municipal Dr. Moyses Deutsch | Instituto Adolfo Lutz, Interdiciplinary Procedures Center, Strategic Laboratory | Claudia Regina Gonçalves; Claudio Tavares Sacchi; Erica Valessa Ramos Gomes; Karoline Rodrigues Campos |
| EPI_ISL_583498 | Hospital Municipal Dr. Waldemar Tebaldi | Instituto Adolfo Lutz, Interdiciplinary Procedures Center, Strategic Laboratory | Claudia Regina Gonçalves; Claudio Tavares Sacchi; Erica Valessa Ramos Gomes; Karoline Rodrigues Campos |
| EPI_ISL_1469754, EPI_ISL_1469778 | Hospital Municipal Getúlio Vargas | Epiclin | Ana Paula Muterle; Carolina Comerlato; Eliana Márcia Da Ros Wendland; Fernando Hayashi Sant'Anna; Janira Prichula; Juliana Comerlato |
| EPI_ISL_693206 | Hospital Municipal Mario Gatti | Instituto Adolfo Lutz, Interdiciplinary Procedures Center, Strategic Laboratory | Claudia Regina Gonçalves; Claudio Tavares Sacchi; Erica Valessa Ramos Gomes; Karoline Rodrigues Campos |
| EPI_ISL_527870, EPI_ISL_574578 | Hospital Municipal Mário Gatti | Instituto Adolfo Lutz, Interdiciplinary Procedures Center, Strategic Laboratory | Claudia Regina Gonçalves; Claudio Tavares Sacchi; Erica Valessa Ramos Gomes; Karoline Rodrigues Campos |
| EPI_ISL_861671, EPI_ISL_882660 | Hospital Municipal Prefeito Waldemar Costa Filho | Instituto Adolfo Lutz, Interdiciplinary Procedures Center, Strategic Laboratory | Claudia Regina Gonçalves; Claudio Tavares Sacchi; Erica Valessa Ramos Gomes; Karoline Rodrigues Campos |
| EPI_ISL_527856 | Hospital Municipal Prof. Waldomiro de Paula | Instituto Adolfo Lutz, Interdiciplinary Procedures Center, Strategic Laboratory | Claudia Regina Gonçalves; Claudio Tavares Sacchi; Erica Valessa Ramos Gomes |
| EPI_ISL_1520110 | Hospital Municipal Reynaldo Guerra Cajati | Instituto Adolfo Lutz, Interdiciplinary Procedures Center, Strategic Laboratory | Caio Vinicius Dias Lopes; Claudia Regina Gonçalves; Claudio Tavares Sacchi; Erica Valessa Ramos Gomes; Karoline Rodrigues Campos |
| EPI_ISL_603028 | Hospital Municipal Santa Ana | Instituto Adolfo Lutz, Interdiciplinary Procedures Center, Strategic Laboratory | Claudia Regina Gonçalves; Claudio Tavares Sacchi; Erica Valessa Ramos Gomes; Karoline Rodrigues Campos |
| EPI_ISL_524468, EPI_ISL_527859 | Hospital Municipal Vereador Jose Storopolli | Instituto Adolfo Lutz, Interdiciplinary Procedures Center, Strategic Laboratory | Claudia Regina Gonçalves; Claudio Tavares Sacchi; Erica Valessa Ramos Gomes |
| EPI_ISL_471647 | Hospital Municipal de Barueri Dr. Francisco Moran | Instituto Adolfo Lutz, Interdiciplinary Procedures Center, Strategic Laboratory | Claudia Regina Gonçalves; Claudio Tavares Sacchi; Erica Valessa Ramos Gomes |
| EPI_ISL_527860 | Hospital Municipal de Parelheiros Josanias Castanha Braga | Instituto Adolfo Lutz, Interdiciplinary Procedures Center, Strategic Laboratory | Claudia Regina Gonçalves; Claudio Tavares Sacchi; Erica Valessa Ramos Gomes |
| EPI_ISL_527862 | Hospital Municipal de Urgência | Instituto Adolfo Lutz, Interdiciplinary Procedures Center, Strategic Laboratory | Claudia Regina Gonçalves; Claudio Tavares Sacchi; Erica Valessa Ramos Gomes |
| EPI_ISL_468308, EPI_ISL_468315, EPI_ISL_515520, EPI_ISL_515546, EPI_ISL_515552, EPI_ISL_523955, EPI_ISL_523974, EPI_ISL_527863 | | | |
| see above | Hospital Municipal do Tatuape Carmino Caricchio | Instituto Adolfo Lutz, Interdiciplinary Procedures Center, Strategic Laboratory | Claudia Regina Gonçalves; Claudio Tavares Sacchi; Erica Valessa Ramos Gomes |
| EPI_ISL_735399 | Hospital Municiplal Dr Ignacio de gouvea | Instituto Adolfo Lutz, Interdiciplinary Procedures Center, Strategic Laboratory | Claudia Regina Gonçalves; Claudio Tavares Sacchi; Erica Valessa Ramos Gomes; Karoline Rodrigues Campos |
| EPI_ISL_735428, EPI_ISL_735429, EPI_ISL_735431, EPI_ISL_906066 | Hospital Nipo Brasileiro | Instituto Adolfo Lutz, Interdiciplinary Procedures Center, Strategic Laboratory | Claudia Regina Gonçalves; Claudio Tavares Sacchi; Erica Valessa Ramos Gomes; Karoline Rodrigues Campos |
| EPI_ISL_527857 | Hospital Regional Vale do Ribeira | Instituto Adolfo Lutz, Interdiciplinary Procedures Center, Strategic Laboratory | Claudia Regina Gonçalves; Claudio Tavares Sacchi; Erica Valessa Ramos Gomes |
| EPI_ISL_524463 | Hospital Regional de Cotia | Instituto Adolfo Lutz, Interdiciplinary Procedures Center, Strategic Laboratory | Claudia Regina Gonçalves; Claudio Tavares Sacchi; Erica Valessa Ramos Gomes |
| EPI_ISL_735418 | Hospital Regional do Vale do Paraiba | Instituto Adolfo Lutz, Interdiciplinary Procedures Center, Strategic Laboratory | Claudia Regina Gonçalves; Claudio Tavares Sacchi; Erica Valessa Ramos Gomes; Karoline Rodrigues Campos |
| EPI_ISL_861649 | Hospital Renascença Campinas | Instituto Adolfo Lutz, Interdiciplinary Procedures Center, Strategic Laboratory | Claudia Regina Gonçalves; Claudio Tavares Sacchi; Erica Valessa Ramos Gomes; Karoline Rodrigues Campos |
| EPI_ISL_833165 | Hospital Samaritano | Instituto Adolfo Lutz, Interdiciplinary Procedures Center, Strategic Laboratory | Claudia Regina Gonçalves; Claudio Tavares Sacchi; Erica Valessa Ramos Gomes; Karoline Rodrigues Campos |
| EPI_ISL_471552 | Hospital Sancta Maggiore | Instituto Adolfo Lutz, Interdiciplinary Procedures Center, Strategic Laboratory | Claudia Regina Gonçalves; Claudio Tavares Sacchi; Erica Valessa Ramos Gomes |
| EPI_ISL_603036 | Hospital Santa Ana | Instituto Adolfo Lutz, Interdiciplinary Procedures Center, Strategic Laboratory | Claudia Regina Gonçalves; Claudio Tavares Sacchi; Erica Valessa Ramos Gomes; Karoline Rodrigues Campos |
| EPI_ISL_693196 | Hospital Santa | Instituto Adolfo Lutz, | Claudia Regina Gonçalves; Claudio Tavares Sacchi; Erica Valessa Ramos Gomes; Karoline Rodrigues Campos |

| | | | |
|---|---|---|---|
| | Clara | Interdiciplinary Procedures Center, Strategic Laboratory | |
| EPI_ISL_693233 | Hospital Santa Cruz | Instituto Adolfo Lutz, Interdiciplinary Procedures Center, Strategic Laboratory | Claudia Regina Gonçalves; Claudio Tavares Sacchi; Erica Valessa Ramos Gomes; Karoline Rodrigues Campos |
| EPI_ISL_693236, EPI_ISL_861646, EPI_ISL_861647, EPI_ISL_861650, EPI_ISL_861654, EPI_ISL_861655 | Hospital Santa Marcelina Sao Paulo | Instituto Adolfo Lutz, Interdiciplinary Procedures Center, Strategic Laboratory | Claudia Regina Gonçalves; Claudio Tavares Sacchi; Erica Valessa Ramos Gomes; Karoline Rodrigues Campos |
| EPI_ISL_861644 | Hospital Santa Virginia | Instituto Adolfo Lutz, Interdiciplinary Procedures Center, Strategic Laboratory | Claudia Regina Gonçalves; Claudio Tavares Sacchi; Erica Valessa Ramos Gomes; Karoline Rodrigues Campos |
| EPI_ISL_1493590 | Hospital Sao Marcos da Samamorro Agudo | Instituto Adolfo Lutz, Interdiciplinary Procedures Center, Strategic Laboratory | Caio Vinicius Dias Lopes; Claudia Regina Gonçalves; Claudio Tavares Sacchi; Erica Valessa Ramos Gomes; Karoline Rodrigues Campos |
| EPI_ISL_471545, EPI_ISL_515545, EPI_ISL_515559, EPI_ISL_515560, EPI_ISL_523969, EPI_ISL_523981, EPI_ISL_523988, EPI_ISL_693201, EPI_ISL_861639 | | | |
| see above | Hospital Sao Paulo de Ensino da Unifesp | Instituto Adolfo Lutz, Interdiciplinary Procedures Center, Strategic Laboratory | Claudia Regina Gonçalves; Claudio Tavares Sacchi; Erica Valessa Ramos Gomes; Karoline Rodrigues Campos |
| EPI_ISL_1469593 | Hospital Sapiranga | Epiclin | Ana Paula Muterle; Carolina Comerlato; Eliana Márcia Da Ros Wendland; Fernando Hayashi Sant’Anna; Janira Prichula; Juliana Comerlato |
| EPI_ISL_2352015, EPI_ISL_2516467, EPI_ISL_2516544 | Hospital Universitari Bellvitge | Microbiology Department | Aida Gonzalez-Diaz; Carmen Ardanuy; Jordi Camara; Jordi Niubó; Laura Calatayud; M Angeles Domínguez; Miguel Fernandez-Huerta; Sara Marti |
| EPI_ISL_468318, EPI_ISL_468321 | Hospital Universitario da USP | Instituto Adolfo Lutz, Interdiciplinary Procedures Center, Strategic Laboratory | Claudia Regina Gonçalves; Claudio Tavares Sacchi; Erica Valessa Ramos Gomes |
| EPI_ISL_471539 | Hospital Universitario da USP Sao Paulo | Instituto Adolfo Lutz, Interdiciplinary Procedures Center, Strategic Laboratory | Claudia Regina Gonçalves; Claudio Tavares Sacchi; Erica Valessa Ramos Gomes |
| EPI_ISL_534314 | Hospital Universitario da USP de SP | Instituto Adolfo Lutz, Interdiciplinary Procedures Center, Strategic Laboratory | Claudia Regina Gonçalves; Claudio Tavares Sacchi; Erica Valessa Ramos Gomes |
| EPI_ISL_1469570 | Hospital Universitário | Epiclin | Ana Paula Muterle; Carolina Comerlato; Eliana Márcia Da Ros Wendland; Fernando Hayashi Sant’Anna; Janira Prichula; Juliana Comerlato |
| EPI_ISL_1469774 | Hospital Universitário de Canoas | Epiclin | Ana Paula Muterle; Carolina Comerlato; Eliana Márcia Da Ros Wendland; Fernando Hayashi Sant’Anna; Janira Prichula; Juliana Comerlato |
| EPI_ISL_3031311 | Hospital da Baleia | Instituto René Rachou / Fiocruz Minas | Alana Oliveira; Anna Salim; Camila Corsini; Daniel Miranda; Gabriel Fernandes; Mozar de Castro; Nathalie Almeida; Pedro Alves; Priscilla Filgueiras; Rafaella Fortini; Raphael Silva; Raquel Vilela; Rubens do Monte Neto; Sarah Gomes; Thaís Silva; Wander Jeremias |
| EPI_ISL_476373, EPI_ISL_476445, EPI_ISL_476446, EPI_ISL_476469 | Hospital da Clínicas da Faculdade de Medicina da Universidade de São Paulo | Instituto de Medicina Tropical da Univesidade de São Paulo | Camila Alves Maia da Silva; Carolina S. Lazar; Cecilia Salete Alencar; Darlan da Silva Candido; Erika Regina Manuli; Ester Sabino; Flavia Cristina da Silva Sales; Giula Magalhaes Ferreira; Jaqueline Goes de Jesus; Julien Theze; Mariana Severo Ramundo; Nuno Faria; Samples: Ingra Morales Claro; Sequencing: Ingra Morales Claro; Silvia F. Costa; Thais de Moura Coletti |
| EPI_ISL_3031319, EPI_ISL_3031335 | Hospital das Clínicas da UFMG | Instituto René Rachou / Fiocruz Minas | Anna Salim; Cristina Fonseca; Gabriel Fernandes; Matheus Westin; Núbia Fernandes; Pedro Alves; Rosiane Pereira; Rubens do Monte Neto; Sandra Gava; Thaís Santos; Thaís Silva; Wilma Patrícia Bernardes |
| EPI_ISL_693205 | Hospital de Campanha Covid-19 Assis | Instituto Adolfo Lutz, Interdiciplinary Procedures Center, Strategic Laboratory | Claudia Regina Gonçalves; Claudio Tavares Sacchi; Erica Valessa Ramos Gomes; Karoline Rodrigues Campos |
| EPI_ISL_735396 | Hospital de Camplanha COVID 19 SER | Instituto Adolfo Lutz, Interdiciplinary Procedures Center, Strategic Laboratory | Claudia Regina Gonçalves; Claudio Tavares Sacchi; Erica Valessa Ramos Gomes; Karoline Rodrigues Campos |
| EPI_ISL_882661, EPI_ISL_882662 | Hospital de Santa Barbara de Goias | Instituto Adolfo Lutz, Interdiciplinary Procedures Center, Strategic Laboratory | Claudia Regina Gonçalves; Claudio Tavares Sacchi; Erica Valessa Ramos Gomes; Karoline Rodrigues Campos |
| EPI_ISL_534319, EPI_ISL_534320 | Hospital do Serv Pub ESTAFCO Morato de Oliveira | Instituto Adolfo Lutz, Interdiciplinary Procedures Center, Strategic Laboratory | Claudia Regina Gonçalves; Claudio Tavares Sacchi; Erica Valessa Ramos Gomes |
| EPI_ISL_693199 | Hospital do Servidor Publico Estadual Francisco Morato de Oliveira | Instituto Adolfo Lutz, Interdiciplinary Procedures Center, Strategic Laboratory | Claudia Regina Gonçalves; Claudio Tavares Sacchi; Erica Valessa Ramos Gomes; Karoline Rodrigues Campos |
| EPI_ISL_471548, EPI_ISL_515565, EPI_ISL_523965, EPI_ISL_523978, EPI_ISL_523982 | Hospital do Servidor Público Estadual Francisco Morato de Oliveira | Instituto Adolfo Lutz, Interdiciplinary Procedures Center, Strategic Laboratory | Claudia Regina Gonçalves; Claudio Tavares Sacchi; Erica Valessa Ramos Gomes |
| EPI_ISL_527861 | Hospital e Maternidade Celso Pierro | Instituto Adolfo Lutz, Interdiciplinary Procedures Center, Strategic Laboratory | 01246-1301; 355 - Brazil; Av. Dr. Arnaldo; Cerqueira Cesar; São Paulo - SP |
| EPI_ISL_693200 | Hospital e Maternidade Mairipora | Instituto Adolfo Lutz, Interdiciplinary Procedures Center, Strategic Laboratory | Claudia Regina Gonçalves; Claudio Tavares Sacchi; Erica Valessa Ramos Gomes; Karoline Rodrigues Campos |
| EPI_ISL_693241 | Hospital e Maternidade Sao Lucas | Instituto Adolfo Lutz, Interdiciplinary Procedures Center, Strategic Laboratory | Claudia Regina Gonçalves; Claudio Tavares Sacchi; Erica Valessa Ramos Gomes; Karoline Rodrigues Campos |
| EPI_ISL_861657 | Hospital e Maternidade Sino Brasileiro | Instituto Adolfo Lutz, Interdiciplinary Procedures Center, Strategic Laboratory | Claudia Regina Gonçalves; Claudio Tavares Sacchi; Erica Valessa Ramos Gomes; Karoline Rodrigues Campos |

| | | | |
|---|---|---|---|
| EPI_ISL_527865 | Hospital e Maternidade São Cristóvão | Instituto Adolfo Lutz, Interdiciplinary Procedures Center, Strategic Laboratory | Claudia Regina Gonçalves; Claudio Tavares Sacchi; Erica Valessa Ramos Gomes |
| EPI_ISL_527868 | Hospital e Maternidade do Braz | Instituto Adolfo Lutz, Interdiciplinary Procedures Center, Strategic Laboratory | Claudia Regina Gonçalves; Claudio Tavares Sacchi; Erica Valessa Ramos Gomes |
| EPI_ISL_861645 | Hospital e Pronto Socorro Comunitario Vila Iolanda | Instituto Adolfo Lutz, Interdiciplinary Procedures Center, Strategic Laboratory | Claudia Regina Gonçalves; Claudio Tavares Sacchi; Erica Valessa Ramos Gomes; Karoline Rodrigues Campos |
| EPI_ISL_693195, EPI_ISL_693230, EPI_ISL_693232, EPI_ISL_735412, EPI_ISL_861648, EPI_ISL_861680 | Hospital e Pronto Socorro Portinari | Instituto Adolfo Lutz, Interdiciplinary Procedures Center, Strategic Laboratory | Claudia Regina Gonçalves; Claudio Tavares Sacchi; Erica Valessa Ramos Gomes; Karoline Rodrigues Campos |
| EPI_ISL_790234, EPI_ISL_1074736, EPI_ISL_1075791, EPI_ISL_1076528, EPI_ISL_1079280, EPI_ISL_1304728, EPI_ISL_2191321, EPI_ISL_2223909 | | | |
| see above | Houston Methodist Hospital | Houston Methodist Hospital | David W. Bernard; Heather Hendrickson; Ilya J. Finkelstein; James J. Davis; Jessica Cambric; Jimmy Gollihar; Kristina Reppond; Layne Pruitt; Madison N. Shyer; Marcus Nguyen; Matthew Ojeda Saavedra; Maulik Shukla; Paul A. Christensen; Prasanti Yerramilli; Randall J. Olsen; Robert Olson; Ryan Gadd; S. Wesley Long; Sishir Subedi; and James M. Musser |
| EPI_ISL_984263, EPI_ISL_1078981, EPI_ISL_1078983, EPI_ISL_1078984, EPI_ISL_1078991, EPI_ISL_1078996, EPI_ISL_1079003, EPI_ISL_1079006, EPI_ISL_1079158, EPI_ISL_1079163, EPI_ISL_1079166, EPI_ISL_1086051, EPI_ISL_1086056, EPI_ISL_1121326 | | | |
| see above | IAL Regional de Bauru | Instituto Adolfo Lutz, Interdiciplinary Procedures Center, Strategic Laboratory | Caio Vinicius Dias Lopes; Claudia Regina Gonçalves; Claudio Tavares Sacchi; Erica Valessa Ramos Gomes; Karoline Rodrigues Campos |
| EPI_ISL_984248, EPI_ISL_984254, EPI_ISL_1171646, EPI_ISL_1171647, EPI_ISL_1196297, EPI_ISL_1196298 | IAL Regional de Marilia | Instituto Adolfo Lutz, Interdiciplinary Procedures Center, Strategic Laboratory | Caio Vinicius Dias Lopes; Claudia Regina Gonçalves; Claudio Tavares Sacchi; Erica Valessa Ramos Gomes; Karoline Rodrigues Campos |
| EPI_ISL_1171664, EPI_ISL_1171668, EPI_ISL_1171669, EPI_ISL_1171671 | IAL Regional de Presidente Prudente | Instituto Adolfo Lutz, Interdiciplinary Procedures Center, Strategic Laboratory | Caio Vinicius Dias Lopes; Claudia Regina Gonçalves; Claudio Tavares Sacchi; Erica Valessa Ramos Gomes; Karoline Rodrigues Campos |
| EPI_ISL_1358296, EPI_ISL_1358297, EPI_ISL_1381044, EPI_ISL_1381046, EPI_ISL_1381049, EPI_ISL_1381064 | IAL Regional de Santo Andre | Instituto Adolfo Lutz, Interdiciplinary Procedures Center, Strategic Laboratory | Caio Vinicius Dias Lopes; Claudia Regina Gonçalves; Claudio Tavares Sacchi; Erica Valessa Ramos Gomes; Karoline Rodrigues Campos |
| EPI_ISL_1123374, EPI_ISL_1171623, EPI_ISL_1171625, EPI_ISL_1171627, EPI_ISL_1171631, EPI_ISL_1171633, EPI_ISL_1171635, EPI_ISL_1171636 | | | |
| see above | IAL Regional de Santos | Instituto Adolfo Lutz, Interdiciplinary Procedures Center, Strategic Laboratory | Caio Vinicius Dias Lopes; Claudia Regina Gonçalves; Claudio Tavares Sacchi; Erica Valessa Ramos Gomes; Karoline Rodrigues Campos |
| EPI_ISL_1293056, EPI_ISL_1293057, EPI_ISL_1293064, EPI_ISL_1293072, EPI_ISL_1293081 | IAL Regional de Sorocaba | Instituto Adolfo Lutz, Interdiciplinary Procedures Center, Strategic Laboratory | Caio Vinicius Dias Lopes; Claudia Regina Gonçalves; Claudio Tavares Sacchi; Erica Valessa Ramos Gomes; Karoline Rodrigues Campos |
| EPI_ISL_1303527, EPI_ISL_1303528 | IAL Regional de São Jose do Rio Preto | Instituto Adolfo Lutz, Interdiciplinary Procedures Center, Strategic Laboratory | Caio Vinicius Dias Lopes; Claudia Regina Gonçalves; Claudio Tavares Sacchi; Erica Valessa Ramos Gomes; Karoline Rodrigues Campos |
| EPI_ISL_2234880, EPI_ISL_2234881, EPI_ISL_2234884, EPI_ISL_2234887, EPI_ISL_2234888, EPI_ISL_2234892, EPI_ISL_2234894, EPI_ISL_2234896, EPI_ISL_2234898, EPI_ISL_2444785, EPI_ISL_2444788, EPI_ISL_2444796, EPI_ISL_2444839, EPI_ISL_4071901 | | | |
| see above | IICS-UNA | IICS-UNA | Adriana Valenzuela; Alejandra Rojas; Chyntia Diaz; Eva Nara; Fatima Cardozo; Florencia del Puerto; Joel Ortiz; Jonas Fernandez; Laura Franco; Laura Mendoza; Leticia Rojas; Magaly Martinez; Maria Eugenia Galeano. |
| EPI_ISL_1213317 | IMT-UFRN/RN | Bioinformatics Laboratory / LNCC | Alessandra P Lamarca; Alexandra L Gerber; Ana Paula Melo Mariano; Ana Paula de C Guimarães; Ana Tereza R Vasconcelos; Angela Maria Guimarães Santos; Bianca Mendes Maciel; Danielle Angst Secco; Eduardo Sérgio Soares Sousa; Eloiza Helena Campana; Francisco Paulo Freire Neto; George Rego Albuquerque; Kátia Castanho Scortecci; Lucymara Fassarella Agnez Lima; Luiz G P de Almeida; Luís Cristóvão Porto; Otavio J. Brustolini; Paulo Ricardo Nascimento; Ronaldo da Silva Francisco Jr; Sandra Rocha Gadelha; Selma Maria Bezerra Jeronimo; Vinícius Pietta Perez |
| EPI_ISL_732116 | INSA | Instituto Nacional de Saude (INSA) | Borges et al |
| EPI_ISL_2758668, EPI_ISL_2758669, EPI_ISL_2758671, EPI_ISL_2758672, EPI_ISL_2758806 | IPEC Guarapuava | IPEC Guarapuava | NAPI-Genômica (Novos Arranjo de Pesquisa e Inovação em Genômica): Ademar Dantas da Cunha Júnior Adriano Ferrasa Adriano Mondini Aldo Przybysz Alessandra Lourenço Cecchini Armani Alex Sandro Jorge Alexandra Ivo de Medeiros Alexandre Maller Aline Cristina Batista Rodrigues Johann Ana Lucia Ferreira Ana Marisa Fusco Almeida Anderson Joel Martino Andrade André Luís Laforga Vanzela Andrea Duarte Doetzer Andrea Name Colado Simao Andressa Pereira de Souza Anelisa Ramão Angelica Beate Winter Boldt Anna Hermínia Castro Gomes de Amorim Anna Silvia Penteado Setti da Rocha Antonio Camilo da Silva Filho Antonio Stabelini Neto Arthur Hirata Bertachi Barbara Mendes Paz Chao Betty Cristiane Kuhn Bruno Ambrozio Galindo Bruno Ribeiro Cruz Camilla Reginatto De Pierri Carla Fredrichsen Moya Araujo Carla Fredrichsen Moya Araujo Carlos Alberto Oliveira de Biagi Junior Carlos Augusto Nassar Carlos Eduardo Buss Carlos Gilberto Carlotti Junior Carlos Henrique Schneider Carolina Panis Carolina Weigert Galvão Caroline de Jesus Coelho Donha Caroline Guisantes de Salvo Toni Caryna Eurich Mazur Catiuscie Cabreira da Silva Tortorella Celso F. D. Doliveira Cesar Luiz Boguszewski Christiane Pienna Soares Chung Man Chin Claudia Moro Cleverson Busso Cristiane Cominetti Daiane Priscila Simão-Silva Dalila Luciola Zanette Daniel de Paula Daniel de Paula Daniel Rech Daniela Fiori Gradia Daniela Pretti da Cunha Tirapelli Daniela Viganó Zanoti Jeronymo Daniele Ukan Danielle Malheiros Ferreira Danielle Venturini Deborah Catharine de Assis Leite Deivid Calebe de Souza Dennis Armando Bertolini Edenir Inez Pamero Edna Maria Vissoci Reiche Edson Roberto Arpini Miguel Eduardo José de Almeida Araújo Eliana Carolina Vespero Eliandro Reis Tavares Elza Kimura Grimshaw Emanuel Maltempi de Souza Emanuele Cristina Gustani Buss Émerson Carraro Emiliana Cristina Melo ENILze Maria de Souza Fonseca Ribeiro Enilze Maria de Souza Fonseca Ribeiro Erika Izumi Erika Seki Kioshima Cotica Evani Marques Pereira Fabio Negretti Fábio Rodrigues Ferreira Seiva Felipe Dunin dos Santos Felipe Tuon Fernanda Andreia Rosa Fernanda Cestaro Prado Cortez Fernanda Ivanski Fernanda Maris Peria Flavia Regina Oliveira de Barros Franciele Aní Caovilla Follador Franciele Mara Lucca Zanardo Bohm Francinete Ramos Campos Fulviana Silva Nishiyama GABRIEL RIBEIRO CORDEIRO Gabriela Datsch Bennemann Gisele Santos de Oliveira Glaucio Valdameri Glauco Akelinghton Freire Vitiello Glauco Vieira Miranda Glaura Scantamburlo ALves Fernandes Guilherme Ferreira Silveira Gustavo Bianchini Porfírio Gustavo Lenci Marques Hélito Volpato Hildebrando Masshiro Nagai Huei Diana Lee Ilce Mara de Syllos Cólus Iris Rabinovich Israel Gomy Jackson Kawakami Jacques Duílio Brancher Jaime Luís Lopes Rocha Jaqueline Carvalho de Oliveira Jean Henrique da Silva Rodrigues Jean Leandro dos Santos Jeane Eliete Laguila Visentainer João Paulo Bianchi Ximenez Joaquim Manoel da Silva Jociani Ascari Joel Donazzolo Jorge Luis Maria Ruiz Jose Knopfholz José Luis da Conceição Silva José Sebastião dos Santos Joseane Carla Schabarum Juliana Cheleski Wiggers Juliana Mara Serpeloni Juliana Morini Küpper Cardoso Perseguini Karen Brajão de Oliveira Karin Braun Prado Karine Aparecida de Lima Katiany Rizzieri Caleffi Ferracioli Katiuscia de Oliveira Francisco Gabriel Kelvinson Fernandes Viana Larissa Beatriz Cossalter Larissa Danielle Bahls Pinto Laurival Antonio Vilas Boas Léia Carolina Lucio Líbero Mezzadri Neto Ligia Carla Faccin Galhardi Lirane Elize Defante Ferreto Luciana Furlaneto Maia Luciana Oliveira de Fariña Luciana Reis Azevedo Alanis Luciane Regina Cavalli Lucy Megumi Yamauchi Lioni Luis Paulo Gomes Mascarenhas Luis Paulo Gomes Mascarenhas Luís Paulo Mascarenhas Lupe Furtado Alle Lyvia Regina Biagi Silva Bertachi Mara Antonia Ramos Costa Mara L. Cordeiro Marcela Maria Birolim Marcelo Ricardo Vicari Marcia Edilaine Lopes Consolaro Marcia Holsbach Beltrame Marcia Regina Eches Perugini Marcos Abdo Arbex Marcos Pileggi MARCOS TADEU GRZELCZAK Marcus Peikriszwili Tartaruga Maria Angelica Ehara Watanabe Maria Antonia Ramos Costa Maria Claudia Gross Maria José Soares Mendes Giannini Maria Leandra Terencio Maria Lúcia Bonfleur Maria Luiza Guimarães de Oliveira Maria Luiza Petzl-Erler Mariana Abe Vicente Cavagnari Marina Kimiko Kadowaki Marise Fonseca dos Santos Marla Karine Amarante Maurício Turkiewicz Mauro Antonio Alves Castro Michel Rodrigo Zambrano Passarini Michele Potrich Michelle Orane Schemberger Milena Massumi Kozonoe Mônica Degraf Cavallin Monica Tereza Suldofski Mucio Luiz de Assis Cirino Nadia Graciele Krohn Najeh Maissar Khalil Nédia de Castilhos Ghisi Neide Tomimura Costa Neiva Leite Neyva Maria Lopes Romeiro Patrícia Amâncio da Rosa Patricia Dayane Carvalho Schaker Patricia Oehlmeyer Nassar Patrícia Savio de Araújo-Souza Patricia Silva Lucio Paulo Henrique Couto Souza Paulo Roberto Donadio Percy Nohama Quirino Alves de Lima Neto Rafael Deminice Rafael dos Santos Bezerra Raquel Alves dos Santos Renan Manozzo Galante Renata Ernlund Freitas de Macedo Rita de Cássia Garcia Simão Roberta Losi Guembarovski Roberto H. Herai Roberto Rosati Rodrigo Ferreira Rodrigo Rodrigues Matiello Rogério Neri Shinsato Rogério Pincela Mateus Rosane Aparecida Ribeiro Rosilene Fressatti Cardoso Rosilene Fressatti Cardoso Sandra Mara Guse Scós Venske Selene Elifio Esposito Sérgio Ossamu Ioshii Silvana Giuliatti Silvia Mara de Souza Halick Silvio Henrique Maia de Almeida Simone Neumann Wendt Spencer Luiz Marques Payão Stefan Wolanski Negrão Stephane Janaina de Moura Escobar Sueli Fumie Yamada Ogatta SUELI PERCIO QUINAIA Taciane Finatto Tatiana Mayumi Veiga Iriyoda Tayza Katelline Danilau Ostroski Tony Alexander Hild Valeria Valente Vanessa Nascimento Kozak Vanessa Santos Sotomaior Victor Breno Pedrosa Victoria Zeghbi Cochenski Borba Vivian Rotuno Moure Valdameri Wander Rogerio Pavanelli Weber Cláudio Francisco Nunes da Silva Willian Augusto de Melo Yohandra Reyes Torres |
| EPI_ISL_2464654, EPI_ISL_2465722 | IdISSC/Hospital Clínico San Carlos de Madrid | SeqCOVID-SPAIN consortium/IBV(CSIC) | Alberto Delgado-Iribarren; Esther Culebras López; Jorge Matias-Guiu; Luis Ortega Medina; Silvia Sánchez Ramón; Ulises Gómez-Pinedo and SeqCOVID-SPAIN consortium; Vicente Estrada Pérez |
| EPI_ISL_1827855 | Institute for Health Research, Epidemiological Surveillance and Training (IRESSEF) | Abbott Laboratories | Adbou Padane; Ambroise Ahouidi; Aminata Dia; Aminata Mboup; Ana Olivo; Anna julienne selbe Ndiaye; Barbara Harris; Cyrille Diedhiou; Gavin Cloherty; Mary Rodgers; Moustapha Mbow; Nafissatou Leye; Ndeye Diabou Diagne; Papa Alassane Diaw; Souleymane Mboup; Todd Meyer |
| EPI_ISL_672705, EPI_ISL_672711, EPI_ISL_672719, EPI_ISL_672720, EPI_ISL_672748 | Institute of Tropical Medicine at the University of São Paulo (IMT-USP) | Laboratório de Parasitologia Médica - Instituto de Medicina Tropical - Universidade de São Paulo | Brazil-UK Centre for Arbovirus Discovery Diagnosis Genomics and Epidemiology (CADDE) Genomic Network - Instituto de Medicina Tropical |
| EPI_ISL_2245187 | Instituto Adolfo Lutz | Coordenação Geral de Laboratórios de Saúde Pública (CGLAB/DAEVS/SVS/MS) | Vagner Fonseca; et al. |
| EPI_ISL_755640, EPI_ISL_755643, EPI_ISL_755654, EPI_ISL_776750, EPI_ISL_776752, EPI_ISL_776753, EPI_ISL_776755, EPI_ISL_776756, EPI_ISL_776761, EPI_ISL_792101, EPI_ISL_792104, EPI_ISL_792106, EPI_ISL_792107, EPI_ISL_792108, EPI_ISL_792109, EPI_ISL_792110, EPI_ISL_792111, EPI_ISL_792112, EPI_ISL_792113, EPI_ISL_792114, EPI_ISL_833152, EPI_ISL_833153, EPI_ISL_833154, EPI_ISL_861625, EPI_ISL_861626, EPI_ISL_861627, EPI_ISL_861629, EPI_ISL_861630, EPI_ISL_861631, EPI_ISL_861632, EPI_ISL_861633, EPI_ISL_861634, EPI_ISL_861643, EPI_ISL_861663 | | | |

| | | | |
|---|---|---|---|
| see above | Instituto Adolfo Lutz - Central | Instituto Adolfo Lutz, Interdiciplinary Procedures Center, Strategic Laboratory | Claudia Regina Gonçalves; Claudio Tavares Sacchi; Erica Valessa Ramos Gomes; Karoline Rodrigues Campos |
| EPI_ISL_776768, EPI_ISL_1039702 | Instituto Adolfo Lutz - Regional de Aracatuba | Instituto Adolfo Lutz, Interdiciplinary Procedures Center, Strategic Laboratory | Claudia Regina Gonçalves; Claudio Tavares Sacchi; Erica Valessa Ramos Gomes; Karoline Rodrigues Campos |
| EPI_ISL_2919249 | Instituto Adolfo Lutz - Regional de Bauru | Instituto Adolfo Lutz, Interdiciplinary Procedures Center, Strategic Laboratory | Caio Vinicius Dias Lopes; Claudia Regina Gonçalves; Claudio Tavares Sacchi; Erica Valessa Ramos Gomes; Karoline Rodrigues Campos |
| EPI_ISL_755655 | Instituto Adolfo Lutz - Regional de Campinas | Instituto Adolfo Lutz, Interdiciplinary Procedures Center, Strategic Laboratory | Claudia Regina Gonçalves; Claudio Tavares Sacchi; Erica Valessa Ramos Gomes; Karoline Rodrigues Campos |
| EPI_ISL_776757, EPI_ISL_776758, EPI_ISL_776767, EPI_ISL_984243, EPI_ISL_1821229 | Instituto Adolfo Lutz - Regional de Marilia | Instituto Adolfo Lutz, Interdiciplinary Procedures Center, Strategic Laboratory | Caio Vinicius Dias Lopes; Claudia Regina Gonçalves; Claudio Tavares Sacchi; Erica Valessa Ramos Gomes; Karoline Rodrigues Campos; Leonardo Jose Tadeu de Araujo |
| EPI_ISL_977471, EPI_ISL_977475, EPI_ISL_977478, EPI_ISL_977480, EPI_ISL_977481, EPI_ISL_977485, EPI_ISL_977488, EPI_ISL_985170, EPI_ISL_1039696 | | | |
| see above | Instituto Adolfo Lutz - Regional de Presidente Prudente | Instituto Adolfo Lutz, Interdiciplinary Procedures Center, Strategic Laboratory | Claudia Regina Gonçalves; Claudio Tavares Sacchi; Erica Valessa Ramos Gomes; Karoline Rodrigues Campos |
| EPI_ISL_735401, EPI_ISL_735402, EPI_ISL_735403, EPI_ISL_735404, EPI_ISL_861667, EPI_ISL_1625985, EPI_ISL_1625996, EPI_ISL_1626008 | | | |
| see above | Instituto Adolfo Lutz - Regional de Rio Claro | Instituto Adolfo Lutz, Interdiciplinary Procedures Center, Strategic Laboratory | Caio Vinicius Dias Lopes; Claudia Regina Gonçalves; Claudio Tavares Sacchi; Erica Valessa Ramos Gomes; Karoline Rodrigues Campos; Katia Correa de Oliveira Santos; Leonardo Jose Tadeu de Araujo |
| EPI_ISL_755647, EPI_ISL_776765, EPI_ISL_776769, EPI_ISL_792103, EPI_ISL_833157 | Instituto Adolfo Lutz - Regional de Santo Andre | Instituto Adolfo Lutz, Interdiciplinary Procedures Center, Strategic Laboratory | Claudia Regina Gonçalves; Claudio Tavares Sacchi; Erica Valessa Ramos Gomes; Karoline Rodrigues Campos |
| EPI_ISL_735400 | Instituto Adolfo Lutz - Regional de Santos | Instituto Adolfo Lutz, Interdiciplinary Procedures Center, Strategic Laboratory | Claudia Regina Gonçalves; Claudio Tavares Sacchi; Erica Valessa Ramos Gomes; Karoline Rodrigues Campos |
| EPI_ISL_833156 | Instituto Adolfo Lutz - Regional de Sorocaba | Instituto Adolfo Lutz, Interdiciplinary Procedures Center, Strategic Laboratory | Claudia Regina Gonçalves; Claudio Tavares Sacchi; Erica Valessa Ramos Gomes; Karoline Rodrigues Campos |
| EPI_ISL_792115, EPI_ISL_792116, EPI_ISL_985171, EPI_ISL_985172, EPI_ISL_985173, EPI_ISL_1039699, EPI_ISL_1039703 | | | |
| see above | Instituto Adolfo Lutz - Regional de Taubate | Instituto Adolfo Lutz, Interdiciplinary Procedures Center, Strategic Laboratory | Claudia Regina Gonçalves; Claudio Tavares Sacchi; Erica Valessa Ramos Gomes; Karoline Rodrigues Campos |
| EPI_ISL_977473, EPI_ISL_977474, EPI_ISL_977476, EPI_ISL_977477, EPI_ISL_977483, EPI_ISL_977484, EPI_ISL_977487, EPI_ISL_984242, EPI_ISL_984246, EPI_ISL_985175, EPI_ISL_1039697, EPI_ISL_1039705, EPI_ISL_1039707, EPI_ISL_1039708, EPI_ISL_1039709, EPI_ISL_1039710, EPI_ISL_1171620, EPI_ISL_1731593, EPI_ISL_2003160, EPI_ISL_2614514, EPI_ISL_2614516, EPI_ISL_2614517, EPI_ISL_2756442, EPI_ISL_2756452 | | | |
| see above | Instituto Adolfo Lutz Central | Instituto Adolfo Lutz, Interdiciplinary Procedures Center, Strategic Laboratory | Caio Vinicius Dias Lopes; Claudia Regina Gonçalves; Claudio Tavares Sacchi; Erica Valessa Ramos Gomes; Karoline Rodrigues Campos; Katia Correa de Oliveira Santos; Leonardo Jose Tadeu de Araujo |
| EPI_ISL_4104676, EPI_ISL_4104692, EPI_ISL_4104697 | Instituto Butantan | Instituto Butantan | Antonio Jorge Martins; Claudia Renata dos Santos Barros; David Schlesinger; Debora Botequio Moretti; Dimas Tadeu Covas; Elaine Cristina Marqueze; Elaine Vieira Santos; Evandra Strazza Rodrigues; Heidge Fukumasu; Jayme Augusto de Souza-Neto; José Salvatore Leister Patané; Luiz Alcantara; Luiz Lehmann Coutinho; Maria Carolina Elias; Maurício Lacerda Nogueira; Rafael dos Santos Bezerra; Raul Machado Neto; Rejane Maria Tommasini Grotto; Ricardo Haddad; Sandra Coccuzzo Sampaio Vessoni; Simone Kashima; Svetoslav Nanev Slavov; Vincent Louis Viala |
| EPI_ISL_2344421, EPI_ISL_2344422, EPI_ISL_2344426, EPI_ISL_2344429, EPI_ISL_2344430, EPI_ISL_2344431, EPI_ISL_2344435, EPI_ISL_2344438, EPI_ISL_2344439, EPI_ISL_2344445, EPI_ISL_2344448, EPI_ISL_2344450, EPI_ISL_2344453, EPI_ISL_2344454, EPI_ISL_2344457, EPI_ISL_2344459, EPI_ISL_2425442 | | | |
| see above | Instituto Butantan | Instituto de Medicina Tropical de Sao Paulo | Brazil-UK Centre for Arbovirus Discovery Diagnosis Genomics and Epidemiology (CADDE) Genomic Network - Instituto de Medicina Tropical |
| EPI_ISL_3478866 | Instituto Nacional de Enfermedades Virales Humanas Dr. Julio I. Maiztegui | Laboratorio Mixto de Biotecnología Acuática (LMBA) on behalf of 'Proyecto Argentino Interinstitucional de genomica de SARS-CoV-2' (PAIS Consortium) | Ada Nazar; Adriana Giri; Agustina Cerri; Agustina Pacual; Anabel Sinchi; COFECyT SF-11; Camila Gonzalez; Carina Bonacalza; Carina Sen; Carlos Figueroa; Cintia Fabbri; Diego Chouhy; Elisa Bolatti; Elizabeth Tapia; Flavio Spetale; Florencia Mascali; Focem COF 03/11 COVID-19); Gastón Viarengo; Germán R. Perez; Ignacio García Labari; Javier Murillo; Joaquín Ezpeleta; Julia Brignone; Leandro Ciappina; Maria Laura Casela; Mariana Viegas (Financiamiento: argenTAG; Mariel Feroci; María Alejandra Morales; María Laura Martin; María Re; María de los Angeles Conti; Matías Abalo; Pablo Casal; Pilar Bulacio; Proyecto IP COVID-19 N°08; Silvana Spinelli; Silvia Arranz; Sofía Lavista Llanos; Sylvia Garcia; Vanina Villanova; Victoria Luppo; Victoria Posner |
| EPI_ISL_955192, EPI_ISL_1262637, EPI_ISL_1298462 | Instituto Nacional de Medicina Genomica | Instituto Nacional de Medicina Genomica | Alcaraz N; Cedro-Tanda A; Cisneros-Villanueva M; Gonzalez-Barrera D; Herrera-Montalvo LA; Hidalgo-Miranda A; Mendoza-Vargas A; Peñaloza-Figueroa F; Ramirez-Vega O; Rangel-DeLeon D; Reyes-Grajeda JP |
| EPI_ISL_492036 | Instituto de Biologia do Exército | Laboratório Metabolismo Macromolecular FirminoTorres de Castro, Instituto de Biofísica Carlos Chagas Filho, Universidade Federal do Rio de Janeiro | Aline Rosa Vianna de Souza; Bianca Catarina Azevedo Cabral; Caleb GM Santos; Clarissa Damaso; Elizabeth Valentin; Marcio da Costa Cipitelli; Marcos Dornelas-Ribeiro; Nádia Vaez Gonçalves da Cruz; Rodrigo Soares de Moura Neto; Rosane Silva; Tatiana LS Nogueira; Virginia Sara Grancieri do Amaral |
| EPI_ISL_861242, EPI_ISL_875540, EPI_ISL_875541, EPI_ISL_875542, EPI_ISL_875543, EPI_ISL_875544, EPI_ISL_875545, EPI_ISL_875546, EPI_ISL_875547, EPI_ISL_875548, EPI_ISL_875549, EPI_ISL_875550, EPI_ISL_888671, EPI_ISL_888672, EPI_ISL_1000670 | | | |
| see above | Instituto de Biotecnologia - UNESP-Botucatu-SP | Instituto de Biotecnologia - UNESP-Botucatu-SP | Camila Dantas Malossi; Fábio Sossai Possebon; João Pessoa Araújo Jr.; Leila Sabrina Ullmann; Paula Rahal; Paulo Inacio da Costa |
| EPI_ISL_836978 | Irmandade da Santa Casa de Misericordia de Lorena | Instituto Adolfo Lutz, Interdiciplinary Procedures Center, Strategic Laboratory | Claudia Regina Gonçalves; Claudio Tavares Sacchi; Erica Valessa Ramos Gomes; Karoline Rodrigues Campos |
| EPI_ISL_1287622 | Istituto Zooprofilattico Sperimentale del Mezzogiorno | TIGEM | Antonio Grimaldi Patrizia Annunziata Francesco Panariello Biancamaria Pierri Claudia Tiberio Valentina Bouche Chiara Colantuono Maria Concetta Cuomo Denise Di Concilio Lucio Di Filippo Anna Manfredi Marcello Salvi Antonio Limone Luigi Atripaldi Pellegrino Cerino Andrea Ballabio Davide Cacchiarelli |
| EPI_ISL_887535, EPI_ISL_981183 | Johns Hopkins Hospital Department of Pathology | Johns Hopkins Hospital Department of Pathology | Adannaya Amadi; C. Paul Morris; Chun Huai Luo; Heba H. Mostafa; Matthew Schwartz; Nicholas Gallagher |

| | | | |
|---|---|---|---|
| EPI_ISL_890322 | KU Leuven, Rega Institute, Clinical and Epidemiological Virology | KU Leuven, Rega Institute, Clinical and Epidemiological Virology | Bert Vanmechelen; Joan Marti-Carerras; Piet Maes; Tony Wawina-Bokalanga |
| EPI_ISL_690818 | Kanagawa Prefectural Institute of Public Health | Pathogen Genomics Center, National Institute of Infectious Diseases | Kentaro Itokawa; Makoto Kuroda; Masanori Hashino; Rina Tanaka; Tsuyoshi Sekizuka |
| EPI_ISL_766625, EPI_ISL_1008415 | Klinisk mikrobiologi | The Public Health Agency of Sweden | Anna Risberg; Anna-Malin Linde; Carlo Berg; Department of Microbiology; Karin Tegmark-Wisell; Maria Lind Karlberg; Mattias Haukland; Mia Brytting; Noura Walai; Oskar Karlsson Lindsjo; Petra Edquist; Petra Holmstrom; Reza Advani; Sofia Stamouli; The Public Health Agency of Sweden |
| EPI_ISL_1795418, EPI_ISL_1795420, EPI_ISL_1795421, EPI_ISL_1795422, EPI_ISL_2345552, EPI_ISL_2345573, EPI_ISL_2345574, EPI_ISL_2345580 | | | |
| see above | LABORATORIO DE FRANCA | Instituto Butantan / ESALQ-Piracicaba | Antonio Jorge Martins; Bianca Cechetto Carlos. Mendelics: Bibiana Santos; Claudia Renata dos Santos Barros; David Schlesinger; David Schlesinger. Hemocentro Ribeirão Preto: Simone Kashima; Debora Botequio Moretti; Debora Botequio Moretti. Centro de Genômica Funcional da ESALQ: Luiz Lehmann Coutinho; Dimas Tadeu Covas; Elaine Cristina Marqueze; Elaine Vieira Santos; Elaine Vieira dos Santos; Elisangela Chicaroni Mattos; Erika Freitas; Evandra Strazza Rodrigues; Felipe Allan da Silva da Costa; Flavia Aburjaile; Guilherme Targino Valente; Heidge Fukumasu; Heidge Fukumasu. USP-Botucatu: Rejane Maria Tommasini Grotto; Instituto Butantan: Alexander Roberto Precioso; Jayme A. Souza-Neto; Jayme Augusto de Souza-Neto; Jessika Cristina Chagas Lesbon; José Salvatore Leister Patané; João Paulo Kitajima; Luiz Alcantara; Luiz Carlos Junior de Alcantara; Luiz Lehmann Coutinho; Maria Carolina Elias; Marta Giovanetti; Maurício Lacerda Nogueira; Patricia Akemi Assato; Rafael dos Santos Bezerra; Raquel de Lello Rocha Campos Cassano. NGS Soluções Genômicas: Pilar Drummond Sampaio Corrêa Mariani. FZEA-USP Pirassununga: Mirele Daiana Poleti; Raul Machado Neto; Rejane Maria Tommasini Grotto; Ricardo Augusto Brassaloti; Ricardo Haddad; Rodrigo Tocantins Calado.; Sandra Coccuzzo Sampaio; Sandra Coccuzzo Sampaio Vessoni; Simone Kashima; Svetoslav Nanev Slavov; Vagner Fonseca; Vincent Louis Viala |
| EPI_ISL_1261698 | LACEN - Laboratório Central de Saúde Pública de Pernambuco | Evandro Chagas Institute | A.M.; Barbagelata; E.C.; E.M.A.; Ferreira; J.A.; Junior; K.C.; L.C.; L.S.; M.C.; P.S.; Pinheiro; Santos; Silva; Sousa; Sousa Junior; W.D.C.; da Silva |
| EPI_ISL_925916, EPI_ISL_926446 | LACEN - Laboratório Central de Saúde Pública do Amazonas | Evandro Chagas Institute Virology | A.M.; Barbagelata; E.C.; E.M.A.; Ferreira; J.A.; Junior; K.C.; L.C.; L.S.; M.C.; P.S.; Pinheiro; Santos; Silva; Sousa; Sousa Junior; W.D.C.; da Silva |
| EPI_ISL_918512 | LACEN - Laboratório Central de Saúde Pública do Amazonas | Evandro Chagas Institute | A.M.; Barbagelata; E.C.; E.M.A.; Ferreira; J.A.; Junior; K.C.; L.C.; L.S.; M.C.; P.S.; Pinheiro; Santos; Silva; Sousa; Sousa Junior; W.D.C.; da Silva |
| EPI_ISL_1164995, EPI_ISL_2488770 | LACEN - Laboratório Central de Saúde Pública do Ceará | Evandro Chagas Institute | A.M.; Barbagelata; E.C.; E.M.A.; Ferreira; J.A.; Junior; K.C.; L.C.; L.S.; M.C.; P.S.; Pinheiro; Santos; Silva; Sousa; Sousa Junior; W.D.C.; da Silva |
| EPI_ISL_2488773, EPI_ISL_2488810 | LACEN - Laboratório Central de Saúde Pública do Maranhao | Evandro Chagas Institute | A.M.; Barbagelata; E.C.; E.M.A.; Ferreira; J.A.; Junior; K.C.; L.C.; L.S.; M.C.; P.S.; Pinheiro; Santos; Silva; Sousa; Sousa Junior; W.D.C.; da Silva |
| EPI_ISL_918515, EPI_ISL_918550 | LACEN - Laboratório Central de Saúde Pública do Para | Evandro Chagas Institute | A.M.; Barbagelata; E.C.; E.M.A.; Ferreira; J.A.; Junior; K.C.; L.C.; L.S.; M.C.; P.S.; Pinheiro; Santos; Silva; Sousa; Sousa Junior; W.D.C.; da Silva |
| EPI_ISL_1086377, EPI_ISL_1164994 | LACEN - Laboratório Central de Saúde Pública do Paraiba | Evandro Chagas Institute | A.M.; Barbagelata; E.C.; E.M.A.; Ferreira; J.A.; Junior; K.C.; L.C.; L.S.; M.C.; P.S.; Pinheiro; Santos; Silva; Sousa; Sousa Junior; W.D.C.; da Silva |
| EPI_ISL_1086376 | LACEN - Laboratório Central de Saúde Pública do Rio Grande do Norte | Evandro Chagas Institute | A.M.; Barbagelata; E.C.; E.M.A.; Ferreira; J.A.; Junior; K.C.; L.C.; L.S.; M.C.; P.S.; Pinheiro; Santos; Silva; Sousa; Sousa Junior; W.D.C.; da Silva |
| EPI_ISL_3912433 | LACEN LABORATORIO CENTRAL DE SAUDE PUBLICA | Analytical Competence Molecular Epidemiology Lab/ACME, Oswaldo Cruz Foundation, Ceara (FIOCRUZ CE) | Cleber Furtado Aksenen; Fabio Miyajima; Fernando Braga Stehling; Francisco Eder de Moura Lopes; Jamille Maria Mendes Bezerra; Joaquim Cesar do Nascimento Sousa Junior; Pedro Miguel Carneiro Jeronimo; Suzana Porto Almeida & Lucas Delerino on behalf of COVID-19 FIOCRUZ Genomic Network; Thais Ferreira de Oliveira; Thais de Oliveira Costa; Ticiane Cavalcante de Souza; Veridiana Pessoa Miyajima |
| EPI_ISL_1293052, EPI_ISL_1303499 | LACEN de Rondonia | Instituto Adolfo Lutz, Interdiciplinary Procedures Center, Strategic Laboratory | Caio Vinicius Dias Lopes; Claudia Regina Gonçalves; Claudio Tavares Sacchi; Erica Valessa Ramos Gomes; Karoline Rodrigues Campos |
| EPI_ISL_1196287, EPI_ISL_1196288, EPI_ISL_1196291, EPI_ISL_1196293 | LACEN do Distrito Federal | Instituto Adolfo Lutz, Interdiciplinary Procedures Center, Strategic Laboratory | Caio Vinicius Dias Lopes; Claudia Regina Gonçalves; Claudio Tavares Sacchi; Erica Valessa Ramos Gomes; Karoline Rodrigues Campos |
| EPI_ISL_943988 | LACEN do Estado de Goias | Instituto Adolfo Lutz, Interdiciplinary Procedures Center, Strategic Laboratory | Claudia Regina Gonçalves; Claudio Tavares Sacchi; Erica Valessa Ramos Gomes; Karoline Rodrigues Campos |
| EPI_ISL_3316170, EPI_ISL_3316222 | LACEN do Estado de Mato Grosso | Instituto Adolfo Lutz, Interdiciplinary Procedures Center, Strategic Laboratory | Caio Vinicius Dias Lopes; Claudia Regina Gonçalves; Claudio Tavares Sacchi; Karoline Rodrigues Campos; Leonardo Tadeu de Araujo; Marlon Benedito Nascimento Santos |
| EPI_ISL_943974, EPI_ISL_943975, EPI_ISL_943976, EPI_ISL_943977, EPI_ISL_943978, EPI_ISL_943979, EPI_ISL_943981, EPI_ISL_943983, EPI_ISL_943985, EPI_ISL_943991, EPI_ISL_2919221 | | | |
| see above | LACEN do Estado de Tocantins | Instituto Adolfo Lutz, Interdiciplinary Procedures Center, Strategic Laboratory | Caio Vinicius Dias Lopes; Claudia Regina Gonçalves; Claudio Tavares Sacchi; Erica Valessa Ramos Gomes; Karoline Rodrigues Campos |
| EPI_ISL_1040825, EPI_ISL_1040826, EPI_ISL_1040827, EPI_ISL_1040828, EPI_ISL_1040830, EPI_ISL_1040832, EPI_ISL_1040834, EPI_ISL_1040838, EPI_ISL_1040841, EPI_ISL_1040846, EPI_ISL_1040847, EPI_ISL_1040849, EPI_ISL_1040850, EPI_ISL_1121329, EPI_ISL_1139052, EPI_ISL_1139054, EPI_ISL_1139056, EPI_ISL_1139057, EPI_ISL_1139060, EPI_ISL_1139067, EPI_ISL_1171621, EPI_ISL_1358305, EPI_ISL_1358308, EPI_ISL_1358311, EPI_ISL_1358314, EPI_ISL_1468437 | | | |
| see above | LACEN do Mato Grosso do Sul | Instituto Adolfo Lutz, Interdiciplinary Procedures Center, Strategic Laboratory | Caio Vinicius Dias Lopes; Claudia Regina Gonçalves; Claudio Tavares Sacchi; Erica Valessa Ramos Gomes; Karoline Rodrigues Campos |
| EPI_ISL_502875 | LACEN/PE | LABBE, Federal University of Pernambuco | ANTONIO CARLOS DE FREITAS; BRUNO SAMPAIO; HEIDI LACERDA ALVES DA CRUZ; MAIRA GALDINO DA ROCHA PITTA; MARCOS ANTONIO DE MORAIS JUNIOR; MARCOS DA SILVEIRA REGUEIRA NETO; MICHELLY CRISTINY PEREIRA; REGINALDO GONCALVES DE LIMA NETO; SERGIO DE SA LEITAO PAIVA JUNIOR; VALDIR DE QUEIROZ BALBINO.; WILSON JOSE DA SILVA JUNIOR; ZILDENE DE SOUSA SILVEIRA |
| EPI_ISL_2821285, EPI_ISL_2821287, EPI_ISL_2821291, EPI_ISL_2821293, EPI_ISL_2821298, EPI_ISL_2821305, EPI_ISL_2821311, EPI_ISL_2821316, EPI_ISL_2821318, EPI_ISL_2821320, EPI_ISL_2821321, EPI_ISL_2821323, EPI_ISL_2821324, EPI_ISL_2821325, EPI_ISL_3046171, EPI_ISL_3060274, EPI_ISL_3704467, EPI_ISL_3704474, EPI_ISL_3704487, EPI_ISL_3704496, EPI_ISL_3704499, EPI_ISL_3704504, EPI_ISL_3704511, EPI_ISL_3704514, EPI_ISL_3704518, EPI_ISL_3704530, EPI_ISL_3704534, EPI_ISL_3704536, EPI_ISL_3704539, EPI_ISL_3704543, EPI_ISL_3704546, EPI_ISL_3704549, EPI_ISL_3704553, EPI_ISL_3704582, EPI_ISL_3704585, EPI_ISL_3704590, EPI_ISL_3704597, EPI_ISL_3704614, EPI_ISL_3704622, EPI_ISL_3704631, EPI_ISL_3704640, EPI_ISL_3704647, EPI_ISL_3704651, EPI_ISL_3704654, EPI_ISL_3704658, EPI_ISL_3704662, EPI_ISL_3704666, EPI_ISL_3704669, EPI_ISL_3704673, EPI_ISL_3704676, EPI_ISL_3704680, EPI_ISL_3704687, EPI_ISL_3704691, EPI_ISL_3704694, EPI_ISL_3704698, EPI_ISL_3704706, EPI_ISL_3704709, EPI_ISL_3704746, EPI_ISL_3704749, EPI_ISL_3704753, EPI_ISL_3704756, EPI_ISL_3704764, EPI_ISL_3704771, EPI_ISL_3704775, EPI_ISL_3704778, EPI_ISL_3704785, EPI_ISL_3704807, EPI_ISL_3704814, EPI_ISL_3704818, EPI_ISL_3704821, EPI_ISL_3704833, EPI_ISL_3704848, EPI_ISL_3704880, EPI_ISL_3704884, EPI_ISL_3704917, EPI_ISL_3704922, EPI_ISL_3704937, EPI_ISL_3704952, EPI_ISL_3704963, EPI_ISL_3705014, EPI_ISL_3705017, EPI_ISL_3705032, EPI_ISL_3705077, EPI_ISL_3705083, EPI_ISL_3705090, EPI_ISL_3705092, EPI_ISL_3705095, EPI_ISL_3705127, EPI_ISL_3835305, EPI_ISL_3835306, EPI_ISL_3835307, EPI_ISL_3835309, EPI_ISL_3835311, EPI_ISL_3835313, EPI_ISL_3835315, EPI_ISL_3835316, EPI_ISL_3835317, EPI_ISL_3835318, EPI_ISL_3835319, EPI_ISL_3835320, EPI_ISL_3835321, EPI_ISL_3835324, EPI_ISL_3835325, EPI_ISL_3835326, EPI_ISL_3835327, EPI_ISL_3835328, EPI_ISL_3835329, EPI_ISL_3835331, EPI_ISL_3835332, EPI_ISL_3835335, EPI_ISL_3835336, EPI_ISL_3835337, EPI_ISL_3835338, EPI_ISL_3835339, EPI_ISL_3835340 | | | |
| see above | LACEN/PE | WallauLab on behalf of Fiocruz COVID-19 Genomic Surveillance Network | Alexandre Freitas da Silva; Antonio Mauro Rezende; Cassia Docena; Constância Flávia Junqueira Ayres; Cássia Docena; Duschinka Ribeiro Duarte Guedes; Elisama Helvecio; Filipe Zimmer Dezordi; Gabriel Luz Wallau; Gustavo Barbosa de Lima; Lais Ceschini Machado; Larissa Krokovsky; Laís Ceschini Machado; Lilian Caroliny Amorim Silva; Marcelo Henrique dos Santos Paiva; Matheus Filgueira Bezerra; Sinval Pinto Brandão Filho |
| EPI_ISL_1213220, EPI_ISL_1213226, EPI_ISL_1213269, EPI_ISL_1213294, EPI_ISL_1213401 | LAFEM/UESC | Bioinformatics Laboratory / LNCC | Alessandra P Lamarca; Alexandra L Gerber; Ana Paula Melo Mariano; Ana Paula de C Guimarães; Ana Tereza R Vasconcelos; Angela Maria Guimarães Santos; Bianca Mendes Maciel; Danielle Angst Secco; Eduardo Sérgio Soares Sousa; Eloiza Helena Campana; Francisco Paulo Freire Neto; George Rego Albuquerque; Kátia Castanho Scortecci; Lucymara Fassarella Agnez Lima; Luiz G P de Almeida; Luís Cristóvão Porto; Otavio J. Brustolini; Paulo Ricardo Nascimento; Ronaldo da Silva Francisco Jr; Sandra Rocha Gadelha; Selma Maria Bezerra Jeronimo; Vinícius Pietta Perez |
| EPI_ISL_861867, EPI_ISL_861868, EPI_ISL_861875, EPI_ISL_861876, EPI_ISL_861879, EPI_ISL_861885, EPI_ISL_861890, EPI_ISL_861892, EPI_ISL_861894, EPI_ISL_861895, EPI_ISL_861896, EPI_ISL_861900, EPI_ISL_861902, EPI_ISL_861903, EPI_ISL_861905, EPI_ISL_861906, EPI_ISL_861909, EPI_ISL_861912, EPI_ISL_861913, EPI_ISL_3457423 | | | |
| see above | LATE - Laboratório de Técnicas | LATE - Laboratório de Técnicas Especiais - | Alexandre Hideaki Takara; Ana Paula Moreira Salles; Anelisie da Silva Santos; Deyvid Amgarten; Erick Gustavo Dorlass; Fernanda de Mello Malta; João Renato Rebello Pinho; Marcio Anunciacao Menezes; Pedro Henrique Sebe Rodrigues; Raquel Riyuzo |

| | | | |
|---|---|---|---|
| | Especiais - Hospital Israelita Albert Einstein | Hospital Israelita Albert Einstein | |
| EPI_ISL_1213453, EPI_ISL_1213454, EPI_ISL_1213458, EPI_ISL_1213459, EPI_ISL_1213461 | LBM/UFPB | Bioinformatics Laboratory / LNCC | Alessandra P Lamarca; Alexandra L Gerber; Ana Paula Melo Mariano; Ana Paula de C Guimarães; Ana Tereza R Vasconcelos; Angela Maria Guimarães Santos; Bianca Mendes Maciel; Danielle Angst Secco; Eduardo Sérgio Soares Sousa; Eloiza Helena Campana; Francisco Paulo Freire Neto; George Rego Albuquerque; Kátia Castanho Scortecci; Lucymara Fassarella Agnez Lima; Luiz G P de Almeida; Luís Cristóvão Porto; Otavio J. Brustolini; Paulo Ricardo Nascimento; Ronaldo da Silva Francisco Jr; Sandra Rocha Gadelha; Selma Maria Bezerra Jeronimo; Vinícius Pietta Perez |
| EPI_ISL_755644, EPI_ISL_833162, EPI_ISL_861669 | Lab LOC - Itapecerica da Serra | Instituto Adolfo Lutz, Interdiciplinary Procedures Center, Strategic Laboratory | Claudia Regina Gonçalves; Claudio Tavares Sacchi; Erica Valessa Ramos Gomes; Karoline Rodrigues Campos |
| EPI_ISL_985178, EPI_ISL_1039698, EPI_ISL_1039704 | Lab Loc - Itapecerica da Serra | Instituto Adolfo Lutz, Interdiciplinary Procedures Center, Strategic Laboratory | Claudia Regina Gonçalves; Claudio Tavares Sacchi; Erica Valessa Ramos Gomes; Karoline Rodrigues Campos |
| EPI_ISL_511103 | Labeto - CAB - Leiria | Instituto Nacional de Saude (INSA) | Borges et al |
| EPI_ISL_2557340, EPI_ISL_2557341, EPI_ISL_2557342, EPI_ISL_2557346, EPI_ISL_2557350, EPI_ISL_2557353, EPI_ISL_2557358 | | | |
| see above | Laboratorio Central de Saude Publica do Estado de Minas Gerais (LACEN/MG) | Laboratory of Respiratory Viruses and Measles, Oswaldo Cruz Institute, FIOCRUZ | Alice Sampaio Rocha; Ana Carolina Mendonca; Andre Felipe Leal Bernardes; Anna Carolina Paixao; Elisa Cavalcante Pereira; Fernando Motta; Luciana Appolinario; Marilda Siqueira on behalf of the Fiocruz COVID-19 Genomic Surveillance Network; Paola Resende; Renata Serrano Lopes; Taina Venas |
| EPI_ISL_2645855, EPI_ISL_2645860, EPI_ISL_2645861, EPI_ISL_2645904 | Laboratorio Central de Saude Publica do Estado do Para (LACEN/PA) | Laboratory of Respiratory Viruses and Measles, Oswaldo Cruz Institute, FIOCRUZ | Alice Sampaio Rocha; Ana Carolina Mendonca; Anna Carolina Paixao; Elisa Cavalcante Pereira; Fernando Motta; Luciana Appolinario; Marilda Siqueira on behalf of the Fiocruz COVID-19 Genomic Surveillance Network; Paola Resende; Renata Serrano Lopes; Taina Venas; Valnete Andrade |
| EPI_ISL_1416434 | Laboratorio Central de Epidemiología (LCE) | Instituto de Biotecnología de la UNAM | Alejandro Sanchez-Flores; Alfredo Herrera-Estrella; Alicia Ocaña-Mondragón; Angel Gustavo Salas-Lais; Bernardo Martínez-Miguel; Blanca Taboada; Brenda Irasema Maldonado-Meza; Carla Ivón Herrera-Najera; Carlos F. Arias; Celia Boukadida; Clara Esperanza Santacruz-Tinoco; Concepción Grajales-Muñiz; Consorcio Mexicano de Vigilancia Genómica (CoViGen-Mex). Authors (in alphabetical order): Julio Elias Alvarado-Yaah; Fernando Fontove-Herrera; Francisco Pulido; Gloria Elena Espinoza-Ayala; Gloria María Molina-Salinas; Gloria Vazquez; Hector Esteban Paz-Juárez; Hector Montoya-Fuentes; Helen Haydee Fernanda Ramírez-Plascencia; Jorge Ivan Salinal-Nevarez; José Antonio Enciso-Moreno; José Esteban Muñoz-Medina; José de Jesús Nuñez-Contreras; Juan Bautista Chale-Dzul; Luis Alberto Ochoa-Carrera; Margarita Matías-Florentino; María Guadalupe Santiago-Mauricio; María Guadalupe de Jesús Mireles-Rivera; Nelly Sélem-Mojica; Pavel Isa; Ricardo Grande; Santiago Ávila-Ríos; Víctor Hugo Borja-Aburto |
| EPI_ISL_2491721, EPI_ISL_2491723, EPI_ISL_2491724, EPI_ISL_2491725, EPI_ISL_2491730, EPI_ISL_2491731, EPI_ISL_2491751 | | | |
| see above | Laboratorio Central de Saude Publica do Estado da Bahia (LACEN/BA) | Laboratory of Respiratory Viruses and Measles, Oswaldo Cruz Institute, FIOCRUZ | Alice Sampaio Rocha; Ana Carolina Mendonca; Anna Carolina Paixao; Elisa Cavalcante Pereira; Felicidade Pereira; Fernando Motta; Luciana Appolinario; Marilda Siqueira on behalf of the Fiocruz COVID-19 Genomic Surveillance Network; Paola Resende; Renata Serrano Lopes; Taina Venas |
| EPI_ISL_2157550, EPI_ISL_2536352 | Laboratorio Central de Saude Publica do Estado da Paraiba (LACEN-PB) | Laboratory of Respiratory Viruses and Measles, Oswaldo Cruz Institute, FIOCRUZ | Alice Sampaio Rocha; Ana Carolina Mendonca; Anna Carolina Paixao; Dalane Loudal Florentino Teixeira; Elisa Cavalcante Pereira; Fernando Motta; Joao Felipe Bezerra; Luciana Appolinario; Marilda Siqueira on behalf of the Fiocruz COVID-19 Genomic Surveillance Network; Paola Resende; Renata Serrano Lopes; Taina Venas |
| EPI_ISL_2645637, EPI_ISL_2645638, EPI_ISL_2645656, EPI_ISL_2645685, EPI_ISL_2645688, EPI_ISL_3434817 | Laboratorio Central de Saude Publica do Estado de Alagoas (LACEN/AL) | Laboratory of Respiratory Viruses and Measles, Oswaldo Cruz Institute, FIOCRUZ | Agatha Soares; Alice Sampaio Rocha; Ana Carolina Mendonca; Anderson Brandao Leite; Anna Carolina Paixao; Elisa Cavalcante Pereira; Fernando Motta; Ighor Arantes; Luciana Appolinario; Marilda Siqueira on behalf of the Fiocruz COVID-19 Genomic Surveillance Network; Paola Resende; Renata Serrano Lopes; Taina Venas |
| EPI_ISL_2645420 | Laboratorio Central de Saude Publica do Estado de Minas Gerais (LACEN-MG) | Laboratory of Respiratory Viruses and Measles, Oswaldo Cruz Institute, FIOCRUZ | Alice Sampaio Rocha; Ana Carolina Mendonca; Andre Felipe Leal Bernardes; Anna Carolina Paixao; Elisa Cavalcante Pereira; Fernando Motta; Luciana Appolinario; Marilda Siqueira on behalf of the Fiocruz COVID-19 Genomic Surveillance Network; Paola Resende; Renata Serrano Lopes; Taina Venas |
| EPI_ISL_2660459, EPI_ISL_2660473, EPI_ISL_2660476, EPI_ISL_2660493, EPI_ISL_2660496, EPI_ISL_2660498, EPI_ISL_2660500, EPI_ISL_2660502, EPI_ISL_2660504 | | | |
| see above | Laboratorio Central de Saude Publica do Estado de Minas Gerais (LACEN/MG) | Laboratory of Respiratory Viruses and Measles, Oswaldo Cruz Institute, FIOCRUZ | Alice Sampaio Rocha; Ana Carolina Mendonca; Andre Felipe Leal Bernardes; Anna Carolina Paixao; Elisa Cavalcante Pereira; Fernando Motta; Luciana Appolinario; Marilda Siqueira on behalf of the Fiocruz COVID-19 Genomic Surveillance Network; Paola Resende; Renata Serrano Lopes; Taina Venas |
| EPI_ISL_2157587, EPI_ISL_2196238, EPI_ISL_2660596, EPI_ISL_2660600, EPI_ISL_2660663, EPI_ISL_2660664, EPI_ISL_2660670, EPI_ISL_2660678, EPI_ISL_2660689, EPI_ISL_2660690 | | | |
| see above | Laboratorio Central de Saude Publica do Estado de Sergipe (LACEN/SE) | Laboratory of Respiratory Viruses and Measles, Oswaldo Cruz Institute, FIOCRUZ | Alice Sampaio Rocha; Ana Carolina Mendonca; Anna Carolina Paixao; Cliomar Alves dos Santos; Elisa Cavalcante Pereira; Fernando Motta; Luciana Appolinario; Marilda Siqueira on behalf of the Fiocruz COVID-19 Genomic Surveillance Network; Paola Resende; Renata Serrano Lopes; Tainá Moreira Martins Venas |
| EPI_ISL_2645546, EPI_ISL_2645549, EPI_ISL_2645551, EPI_ISL_2645554, EPI_ISL_2645561, EPI_ISL_2645564, EPI_ISL_2645565, EPI_ISL_2645569, EPI_ISL_2645574, EPI_ISL_2645578, EPI_ISL_2645579, EPI_ISL_2645586, EPI_ISL_2645588, EPI_ISL_2645599, EPI_ISL_2645606, EPI_ISL_2645613, EPI_ISL_2645614, EPI_ISL_2645626, EPI_ISL_2645627, EPI_ISL_2645633 | | | |
| see above | Laboratorio Central de Saude Publica do Estado do Espirito Santo (LACEN/ES) | Laboratory of Respiratory Viruses and Measles, Oswaldo Cruz Institute, FIOCRUZ | Alice Sampaio Rocha; Ana Carolina Mendonca; Anna Carolina Paixao; Eilisa Cavalcante Pereira; Fernando Motta; Luciana Appolinario; Marilda Siqueira on behalf of the Fiocruz COVID-19 Genomic Surveillance Network; Paola Resende; Renata Serrano Lopes; Rodrigo Ribeiro Rodrigues; Taina Venas |
| EPI_ISL_1465225, EPI_ISL_2983247, EPI_ISL_2983248, EPI_ISL_2983249, EPI_ISL_2983255, EPI_ISL_2983316, EPI_ISL_2983318, EPI_ISL_2983472 | | | |
| see above | Laboratorio Central de Saude Publica do Estado do Maranhao (LACEN-MA) | Laboratory of Respiratory Viruses and Measles, Oswaldo Cruz Institute, FIOCRUZ | Agatha Cristinne Prudencio; Alice Sampaio Rocha; Ana Carolina Mendonca; Anna Carolina Paixao; Elisa Cavalcante Pereira; Fernando Motta; Ighor Leonardo Arantes Gomes; Lidio Gonçalves Lima Neto; Luciana Appolinario; Marilda Siqueira on behalf of the Fiocruz COVID-19 Genomic Surveillance Network; Paola Resende; Renata Serrano Lopes; Taina Moreira Venas; Tainá Venas |
| EPI_ISL_2775424, EPI_ISL_2775425, EPI_ISL_2775437, EPI_ISL_2775444, EPI_ISL_2775452, EPI_ISL_2775454, EPI_ISL_2775455, EPI_ISL_2775456, EPI_ISL_2775458, EPI_ISL_2775463, EPI_ISL_2775466, EPI_ISL_2775469, EPI_ISL_2775470, EPI_ISL_2775472, EPI_ISL_2775473, EPI_ISL_2775474, EPI_ISL_2775476, EPI_ISL_2775477, EPI_ISL_2775478, EPI_ISL_2775479, EPI_ISL_2775480, EPI_ISL_2775481 | | | |
| see above | Laboratorio Central de Saude Publica do Estado do Parana (Instituto de Biologia Molecular do Paraná (LAC)EN-PR) | Instituto Carlos Chagas - Fiocruz | Alessandra De Melo Aguiar; Andreia Akemi Suzukawa; Andréa Rodrigues Ávila; Bruno Dallagiovanna; Dalila Zanette; Eduardo Balsanelli; Emanuel Maltempi de Souza; Fabio Passetti; Fabricio Klerynton Marchini; Fábio de Oliveira Pedrosa; Guilherme Becker; Helisson Faoro; Hellen Geremias dos Santos; Irina Nastassja Riediger; Letusa Albrecht; Lucas Blanes; Luis Gustavo Morello; Lysangela Ronalte Alves; Maria do Carmo Debur; Mauro de Medeiros Oliveira; Michelle Orane Schemberger; Paola Cristina Resende; Sheila Cristina Nardeli; Tiago Gräf; Valter Antônio de Baura |
| EPI_ISL_2731459, EPI_ISL_2731461, EPI_ISL_2731467, EPI_ISL_3061893, EPI_ISL_3243095, EPI_ISL_3243097, EPI_ISL_3243098, EPI_ISL_3243099, EPI_ISL_3243101, EPI_ISL_3243102, EPI_ISL_3243104, EPI_ISL_3243105, EPI_ISL_3243106, EPI_ISL_3243107, EPI_ISL_3243108, EPI_ISL_3243109, EPI_ISL_3243110, EPI_ISL_3243111, EPI_ISL_3243112 | | | |
| see above | Laboratorio Central de Saude Publica do Estado do Parana (LACEN/PR) | Laboratory of Respiratory Viruses and Measles, Oswaldo Cruz Institute, FIOCRUZ | Agatha Soares; Alice Sampaio Rocha; Ana Carolina Mendonca; Anna Carolina Paixao; Elisa Cavalcante Pereira; Fernando Motta; Ighor Leonardo Arantes; Irina Riediger; Luciana Appolinario; Marilda Siqueira on behalf of the Fiocruz COVID-19 Genomic Surveillance Network; Paola Resende; Renata Serrano Lopes; Taina Venas |
| EPI_ISL_2603526, EPI_ISL_2661786, EPI_ISL_2661795, EPI_ISL_2661797, EPI_ISL_2661798, EPI_ISL_2661799, EPI_ISL_2661801, EPI_ISL_2661819, EPI_ISL_2661827, EPI_ISL_2661865 | | | |
| see above | Laboratorio Central de Saude Publica do Estado do Rio Grande do Sul (LACEN-RS) | Laboratory of Respiratory Viruses and Measles, Oswaldo Cruz Institute, FIOCRUZ | Alice Sampaio Rocha; Ana Carolina Mendonca; Anderson Brandao Leite; Anna Carolina Paixao; Elisa Cavalcante Pereira; Fernando Motta; Luciana Appolinario; Marilda Siqueira on behalf of the Fiocruz COVID-19 Genomic Surveillance Network; Paola Resende; Renata Serrano Lopes; Richard Salvato; Taina Venas; Tatiana Schaffer Gregianini |
| EPI_ISL_3048754, EPI_ISL_3048755, EPI_ISL_3048772, EPI_ISL_3048797, EPI_ISL_3048810, EPI_ISL_3048813, EPI_ISL_3048815, EPI_ISL_3048820, EPI_ISL_3048821, EPI_ISL_3048824, EPI_ISL_3048826, EPI_ISL_3048827, EPI_ISL_3048831, EPI_ISL_3048834 | | | |
| see above | Laboratorio Central de Saude Publica do Estado do Rio Grande do Sul (LACEN-RS) | Laboratório de Biologia Molecular da Universidade Federal de Ciências da Saúde de Porto Alegre | Adriana Seixas; Ana B. G. Veiga; Ana Paula Muterle Varela; Fabiana Quoos Mayer; Fernando Hayashi Sant'Anna; Janira Prichula; Letícia Garay Martins; Richard Steiner Salvato; Tatiana Schäffer Gregianini |
| EPI_ISL_2731471 | Laboratorio Central | Laboratory of | Alice Sampaio Rocha; Ana Carolina Mendonca; Andrea Cony Cavalcanti; Anna Carolina Paixao; Elisa Cavalcante Pereira; Fernando Motta; Luciana Appolinario; Marilda Siqueira on behalf of the Fiocruz COVID-19 Genomic Surveillance Network; Paola Resende; Renata Serrano Lopes; Taina Venas |

| | | | |
|---|---|---|---|
| | de Saude Publica do Estado do Rio de Janeiro (LACEN/RJ) | Respiratory Viruses and Measles, Oswaldo Cruz Institute, FIOCRUZ | |
| EPI_ISL_979352 | Laboratorio Estatal de Salud Pública de Nuevo León | Laboratorio de Infectología Molecular, Departamento de Bioquímica y Medicina Molecular,Facultad de Medicina - Universidad Autónoma de Nuevo León | Ana M. Rivas-Estilla; Consuelo Treviño-Garza; Daniel Arellanos-Soto; Else del Carmen García-García; Gloria A. Jasso-de-la-Peña; Kame A. Galán-Huerta; Manuel E. de-la-O-Cavazos; María F. Herrera-Saldivar; Natalia Martínez-Acuña; Roberto Montes-de-Oca; Samuel Buentello-Wong; Sonia A. Lozano-Sepúlveda |
| EPI_ISL_735398 | Laboratorio Fleury | Instituto Adolfo Lutz, Interdiciplinary Procedures Center, Strategic Laboratory | Claudia Regina Gonçalves; Claudio Tavares Sacchi; Erica Valessa Ramos Gomes; Karoline Rodrigues Campos |
| EPI_ISL_861628 | Laboratorio Municipal de Guarulhos | Instituto Adolfo Lutz, Interdiciplinary Procedures Center, Strategic Laboratory | Claudia Regina Gonçalves; Claudio Tavares Sacchi; Erica Valessa Ramos Gomes; Karoline Rodrigues Campos |
| EPI_ISL_801386, EPI_ISL_801387, EPI_ISL_801388, EPI_ISL_801389, EPI_ISL_801390, EPI_ISL_801391, EPI_ISL_801392, EPI_ISL_801393, EPI_ISL_801394, EPI_ISL_801395, EPI_ISL_801396, EPI_ISL_833131, EPI_ISL_1068082, EPI_ISL_1068089, EPI_ISL_1068090, EPI_ISL_1068093, EPI_ISL_1068095, EPI_ISL_1068096, EPI_ISL_1068101, EPI_ISL_1068102, EPI_ISL_1068107, EPI_ISL_1068127, EPI_ISL_1068129, EPI_ISL_1068130, EPI_ISL_1068132, EPI_ISL_1068133, EPI_ISL_1068134, EPI_ISL_1068135, EPI_ISL_1068137, EPI_ISL_1068146, EPI_ISL_1068148, EPI_ISL_1068152, EPI_ISL_1068161, EPI_ISL_1068168, EPI_ISL_1068172, EPI_ISL_1068175, EPI_ISL_1068182, EPI_ISL_1068190, EPI_ISL_1068192, EPI_ISL_1068197, EPI_ISL_1068205, EPI_ISL_1068206, EPI_ISL_1068208, EPI_ISL_1068209, EPI_ISL_1068210, EPI_ISL_1068211, EPI_ISL_1068213, EPI_ISL_1068214, EPI_ISL_1068217, EPI_ISL_1068218, EPI_ISL_1068242, EPI_ISL_1068244, EPI_ISL_1068245, EPI_ISL_1068246, EPI_ISL_1068247, EPI_ISL_1068250, EPI_ISL_1068253, EPI_ISL_1068254, EPI_ISL_1068257, EPI_ISL_1661251, EPI_ISL_2777360, EPI_ISL_2777371, EPI_ISL_2777387, EPI_ISL_2777390, EPI_ISL_2777391, EPI_ISL_2777392, EPI_ISL_2777394 | | | |
| see above | Laboratorio de Ecologia de Doencas Transmissiveis na Amazonia, Instituto Leonidas e Maria Deane - Fiocruz Amazonia | Laboratorio de Ecologia de Doencas Transmissiveis na Amazonia, Instituto Leonidas e Maria Deane - Fiocruz Amazonia | André Corado; Debora Duarte; Felipe Naveca; Felipe Naveca on behalf of the Fiocruz COVID-19 Genomic Surveillance Network; Fernanda Nascimento; George Silva; Karina Pessoa; Luciana Gonçalves; Maria Júlia Brandão; Matilde Mejía; Michele Jesus; Valdinete Nascimento; Victor Souza; Ágatha Costa |
| EPI_ISL_1511641 | Laboratorio de Patologia Clinica - UNICAMP | Laboratorio de Estudos de Virus Emergentes | Alessandro S. Farias; Aline Vieira; André S. Vieira; Angelica Schreiber; Antonio C. G. Carlos Jr; Barbara F. N. Carvalho; Camila L. Simeoni; Daniel A. Toledo-Teixeira; Emerson S.S. França; Ester C. Sabino; Fabiana Granja; Fernando R. Spilki; Gisele A. Pedroso; Ingra M. Claro; José Luiz Proenca-Modena; Julia Forato; Kamila C. S. Krywacz; Karina Bispo-dos-Santos; Luciana S. Mofatto; Luís Felipe Bachur; Luís G. O. Cardoso; Magnun N. N. Santos; Marcelo A. Mori; Maria H. P. Pavan; Maria L. Moretti; Mariene R. Amorim; Natalia S. Brunetti; Nuno R. Faria; Patricia A. F. Leme; Pierina L. Parise; Rodrigo Angerami; Tania R. Zaccariotto; William M. Souza |
| EPI_ISL_2008937, EPI_ISL_2008939, EPI_ISL_2107304, EPI_ISL_2544838, EPI_ISL_2544848, EPI_ISL_2544849, EPI_ISL_2544851, EPI_ISL_2544857, EPI_ISL_2544858, EPI_ISL_2544862, EPI_ISL_2544873, EPI_ISL_2544879, EPI_ISL_2544880, EPI_ISL_2544894, EPI_ISL_2544895, EPI_ISL_3048949, EPI_ISL_3048950 | | | |
| see above | Laboratorio de Pesquisa em Virologia, FAMERP, SJRP | Laboratorio de Pesquisa em Virologia, FAMERP, SJRP | Cecília Artico Banho; Cíntia Bittar; Fábio Sossai Possebon; Guilherme Campos; Helena Lage Ferreira; Jorge A. Petroli Marchesi; João Pessoa Araújo Jr.; Leila Sabrina Ullmann; Lívia Sacchetto; Maisa C. Pereira Parra; Marília Moraes; Maurício L. Nogueira.; Paula Rahal; Paulo Inacio da Costa |
| EPI_ISL_1534655 | Laboratorio de Referencia Nacional de Virus Respiratorio. Instituto Nacional de Salud Perú | Laboratorio de Referencia Nacional de Biotecnología y Biología Molecular. Instituto Nacional de Salud Perú | Carlos Padilla Rojas; Henri Bailon Calderon; Johanna Balbuena Torrez; Karolyn Vega Chozo; Luis Barcena; Marco Galarza Perez; Maribel Huaringa Nuñez; Nancy Rojas Serrano; Omar Caceres Rey; Priscila Lope Pari |
| EPI_ISL_1111146 | Laboratorio de Referencia Nacional de Virus Respiratorio. Instituto Nacional de Salud Perú | Laboratorio de Referencia Nacional de Enteropatógenos. Instituto Nacional de Salud del Perú | Fiorella Orellana Peralta; Iris Silva Molina; Junior Caro Castro; Ronnie Gavilan Chavez; Veronica Hurtado Vela; Willi Quino Sifuentes |
| EPI_ISL_623130, EPI_ISL_717807, EPI_ISL_717808, EPI_ISL_717810, EPI_ISL_717811, EPI_ISL_717812, EPI_ISL_717813, EPI_ISL_717814, EPI_ISL_717815, EPI_ISL_717818, EPI_ISL_717819, EPI_ISL_717820, EPI_ISL_717821, EPI_ISL_717822, EPI_ISL_717823, EPI_ISL_717824, EPI_ISL_717825, EPI_ISL_717826, EPI_ISL_717827, EPI_ISL_717828, EPI_ISL_717829, EPI_ISL_717830 | | | |
| see above | Laboratorio de Virologia Molecular / UFRJ | Bioinformatics Laboratory / LNCC | Alexandra L Gerber; Amilcar Tanuri; Ana Paula de C Guimarães; Ana Tereza R de Vasconcelos; Andréa Cony Cavalcanti; Carolina M Voloch; Claudia dos Santos Rodrigues; Covid19-UFRJ Workgroup; Cynthia C Cardoso; Diana Mariani; Luiz G P de Almeida; Luís Cristóvão Pôrto; Orlando C. Ferreira; Otavio Bustrolini; Otavio J. Brustolini; Renato S Aguiar; Ronaldo S Francisco Jr; Ronaldo da Silva F Jr; Terezinha M P P Castiñeira; Terezinha M P P Castiñeiras |
| EPI_ISL_3090038, EPI_ISL_3090039 | Laboratorio de Virología Molecular, Centro Universitario Regional del Litoral Norte. Universidad de la República, Salto, Uruguay | Centro de Innovación en Vigilancia Epidemiológica (CiVE), Institut Pasteur Montevideo, Uruguay | Alicia Costábile; Alvaro Fajardo; Ana Moller; Andrés Lizasoain; Belén González; Bernardina Rivera; Cecilia Alonso; Cecilia Salazar; Gonzalo Bello; Gonzalo Moratorio; Gregorio Iraola; Henry Albornoz; Ignacio Ferrés; Javier Hurtado; Juan Zanetti; Julio Medina; Luciana Griffero; Lucía Spangenberg; Ma Noel Bentancor; Ma Pía Techera; Mailen Arleo; Martina Alonso; Matías Maidana; Mauricio Méndez; Melissa Duquía; Mercedes Paz; Natalia Rego; Natalia Reyes; Nicolas Nin; Odhille Chappos; Paula Perbolianachis; Pilar Moreno; Rodney Colina; Rodrigo Arce; Tamara Fernández-Calero; Tania Possi; Veronica Noya; Viviana Bortagaray |
| EPI_ISL_1798980, EPI_ISL_1799147, EPI_ISL_1928690 | Laboratory Corporation of America | Centers for Disease Control and Prevention Division of Viral Diseases, Pathogen Discovery | Adrian Paskey; Amanda Douglas; Amanda Suchanek; Andrea Throop; Ayla Burns; Benjamin Rambo-Martin; Bobbi Croy; Brian Krueger; Brian Norvell; Christopher Gulvick; Christos Petropoulos; Clinton R. Paden; Craig Lukasik; Dakota Howard; Darlene Wagner; Debbie Boles; Dhwani Batra; Duncan MacCannell; Eyad Almasri; Goran Stevovic; Howard Engler; Hrushikesh Deshmukh; Jake Humphrey; Jana Schroth; Jason Caravas; Joe Voshell; John Pruitt; Jonathan Meltzer; Jonathan Williams; Kara Moser; Kimberly Wagner; Lax Iyer; Lyndon Tilson; Manoj Jain; Marcia Eisenberg; Mary Ann Cristobal; Mary Williamson; Matthew Schmerer; Michael Levandoski; Mike Sapeta; Mindy Nye; Minoo Agarwal; Mohan Kolli; Nuthawin Charoensri; Oren Cohen; Peter W. Cook; Prashant Gupta; Qian Zeng; Rama Ghatti; Scott Parker; Scott Ryan; Scott Sammons; Shatavia Morrison; Stanley Letovsky; Steven Ragan; Suresh Babu Selvaraju; Susan Countryman; Susan Hicks; Suzanne Dale; Thomas Urban; Tim Kuphal; Tricia Zwiefelhofer; Vincent Drouillon; Yvette Unoarumhi |
| EPI_ISL_1021947 | Laboratory Corporation of America | Respiratory Viruses Branch, Division of Viral Diseases, Centers for Disease Control and Prevention | Ben L. Rambo-Martin; Clinton R. Paden; Dakota Howard; Dhwani Batra; Duncan MacCannell; Peter W. Cook; Suxiang Tong |
| EPI_ISL_754913 | Laboratory Diagnostics and Clinical Immunology of Developmental Age, Medical University of Warsaw | genXone SA, Research & Development Laboratory; The Faculty of Mathematics, Informatics and Mechanics of the University of Warsaw | Anna Gambin; Grzegorz Nowicki; Jakub Grabowski; Maciej Sykulski; Michał Kaszuba; Monika Mańkowska-Woźniak; Natalia Drwęska-Matelska; Urszula Demkow; Łukasz Krych |
| EPI_ISL_2614072 | Laboratory of Molecular Virology, Federal University of Rio de Janeito, UFRJ | Laboratory of Respiratory Viruses and Measles, Oswaldo Cruz Institute, FIOCRUZ | Alice Sampaio Rocha; Amilcar Tanuri; Ana Carolina Mendonca; Anna Carolina Paixao; Elisa Cavalcante Pereira; Fernando Motta; Luciana Appolinario; Marilda Siqueira on behalf of the Fiocruz COVID-19 Genomic Surveillance Network; Paola Resende; Renata Serrano Lopes; Taina Venas |
| EPI_ISL_467356, EPI_ISL_467359, EPI_ISL_467366, EPI_ISL_541354, EPI_ISL_541355, EPI_ISL_541359, EPI_ISL_2443595, EPI_ISL_2443635, EPI_ISL_2443636, EPI_ISL_2557388, EPI_ISL_2557400, EPI_ISL_2603477, EPI_ISL_2603487, EPI_ISL_2603499, EPI_ISL_2603501, EPI_ISL_2603505, EPI_ISL_2603512, EPI_ISL_2603516, EPI_ISL_2603517, EPI_ISL_2603520, EPI_ISL_2614095, EPI_ISL_2614103, EPI_ISL_2614104, EPI_ISL_2614144, EPI_ISL_2614156, EPI_ISL_2614160, EPI_ISL_2614169, EPI_ISL_2614181, EPI_ISL_2677218, EPI_ISL_2731462, EPI_ISL_2731463, EPI_ISL_2731464, EPI_ISL_2731465, EPI_ISL_2731477, EPI_ISL_2731479, EPI_ISL_2731484, EPI_ISL_2731485, EPI_ISL_2731489, EPI_ISL_2731490, EPI_ISL_2731496, EPI_ISL_2731497, EPI_ISL_2731500, EPI_ISL_2731502, EPI_ISL_2731503, EPI_ISL_2731505, EPI_ISL_3190277, EPI_ISL_3190280, EPI_ISL_3190284, EPI_ISL_3243093, EPI_ISL_3434944, EPI_ISL_3434945 | | | |
| see above | Laboratory of Respiratory Viruses and Measles, Oswaldo Cruz Institute, FIOCRUZ | Laboratory of Respiratory Viruses and Measles, Oswaldo Cruz Institute, FIOCRUZ | Agatha Cristinne Prudencio; Alice Sampaio Rocha; Aline Mattos; Ana Carolina Mendonca; Ana Carolina Mendonça; Anna Carolina Paixao; Anna Carolina Paixão; Braulia Caetano; Cinthia Avila; Cristiana Garcia; Elisa Cavalcante Pereira; Fernando Motta; Ighor Leonardo Arantes Gomes; Jonathan Lopes; Luciana Appolinario; Maria Ogrzewalska; Marilda Siqueira on behalf of the Fiocruz COVID-19 Genomic Surveillance Network; Milene Miranda; Paola Resende; Renata Serrano Lopes; Roxana Loayza; Taina Venas |
| EPI_ISL_456088 | Laboratório Central de Saúde Pública Noel Nutels (LACEN-RJ) | Laboratory of Respiratory Viruses and Measles, Oswaldo Cruz Institute, FIOCRUZ | Aline Mattos; Braulia Caetano; Cristiana Garcia; Fernando Motta; Jonathan Lopes; Luciana Appolinario; Maria Ogrzewalska; Marilda Siqueira on behalf of the Fiocruz COVID-19 Genomic Surveillance Network; Milene Miranda; Paola Resende |
| EPI_ISL_2241528 | Laboratório Central de Saúde Pública da Bahia | Coordenação Geral de Laboratórios de Saúde Pública (CGLAB/DAEVS/SVS/MS) | Vagner Fonseca; et al. |
| EPI_ISL_2241498, EPI_ISL_2241529, | Laboratório Central de Saúde Pública da | Coordenação Geral de Laboratórios de Saúde | Vagner Fonseca; et al. |

| | | | |
|---|---|---|---|
| EPI_ISL_2241566, EPI_ISL_2241572, EPI_ISL_2241596 | Paraiba | Pública (CGLAB/DAEVS/SVS/MS) | |
| EPI_ISL_2308426, EPI_ISL_2308452, EPI_ISL_2308469 | Laboratório Central de Saúde Pública de Alagoas | Coordenação Geral de Laboratórios de Saúde Pública (CGLAB/DAEVS/SVS/MS) | Vagner Fonseca; et al. |
| EPI_ISL_2298748 | Laboratório Central de Saúde Pública de Roraima | Coordenação Geral de Laboratórios de Saúde Pública (CGLAB/DAEVS/SVS/MS) | Vagner Fonseca; et al. |
| EPI_ISL_2293009 | Laboratório Central de Saúde Pública de Santa Catarina | Coordenação Geral de Laboratórios de Saúde Pública (CGLAB/DAEVS/SVS/MS) | Vagner Fonseca; et al. |
| EPI_ISL_2241557, EPI_ISL_2241609 | Laboratório Central de Saúde Pública de Sergipe | Coordenação Geral de Laboratórios de Saúde Pública (CGLAB/DAEVS/SVS/MS) | Vagner Fonseca; et al. |
| EPI_ISL_2245069 | Laboratório Central de Saúde Pública do Amapá | Coordenação Geral de Laboratórios de Saúde Pública (CGLAB/DAEVS/SVS/MS) | Vagner Fonseca; et al. |
| EPI_ISL_2298865 | Laboratório Central de Saúde Pública do Amazonas | Coordenação Geral de Laboratórios de Saúde Pública (CGLAB/DAEVS/SVS/MS) | Vagner Fonseca; et al. |
| EPI_ISL_1239117 | Laboratório Central de Saúde Pública do Espírito Santo | Coordenação Geral de Laboratórios de Saúde Pública (CGLAB) | ; Vagner Fonseca et al |
| EPI_ISL_792605 | Laboratório Central de Saúde Pública do Estado da Paraíba (LACEN-PB) | Laboratory of Respiratory Viruses and Measles, Oswaldo Cruz Institute, FIOCRUZ | Ana Carolina Mendonca; Anna Carolina Paixao; Dalane Loudal Florentino Teixeira; Fernando Motta; João Felipe Bezerra; Luciana Appolinario; Marilda Siqueira on behalf of the Fiocruz COVID-19 Genomic Surveillance Network; Paola Resende; Romero Henrique Teixeira de Vasconcelos; Thiago Franco de Oliveira Carneiro |
| EPI_ISL_427292, EPI_ISL_792643 | Laboratório Central de Saúde Pública do Estado de Alagoas (LACEN-AL) | Laboratory of Respiratory Viruses and Measles, Oswaldo Cruz Institute, FIOCRUZ | Aline Mattos; Ana Carolina Mendonca; Anderson Brandao Leite; Anna Carolina Paixao; Braulia Caetano; Cristiana Garcia; Fernando Motta; Jonathan Lopes; Luciana Appolinario; Maria Ogrzewalska; Marilda Siqueira on behalf of the Fiocruz COVID-19 Genomic Surveillance Network; Milene Miranda; Paola Resende; Priscila Born; Sunando Roy |
| EPI_ISL_500483, EPI_ISL_572371 | Laboratório Central de Saúde Pública do Estado de Pernambuco (LACEN-PE) | WallauLab, Aggeu Magalhaes Institute | Alexandre Freitas da Silva; Antonio Mauro Rezende; Armando de Menezes Neto; Bruna Santos Lima Figueiredo de Sá; Caroline Targino Alves da Silva; Claudio Eduardo Cavalcanti; Constância Flávia Junqueira Ayres; Cássia Docena; Derciliano Lopes da Cruz; Duschinka Ribeiro Duarte Guedes; Elisama Helvecio; Filipe Zimmer Dezordi; Gabriel Luz Wallau on behalf of the Fiocruz COVID-19 Genomic Surveillance Network; Gonzalo Bello; Kamila Gaudêncio da Silva Sales; Larissa Krokovsky; Laís Ceschini Machado; Luciane Caroline Albuquerque Bezerra; Luydson Richardson Silva Vasconcelos; Marcelo Henrique Santos Paiva; Maria Almerice Lopes da Silva; Matheus Filgueira Bezerra; Michelle da Silva Barros; Paola Cristina Resende; Renata Pessôa Germano Mendes; Rodrigo Moraes Loyo Arcoverde; Severino Jefferson Ribeiro da Silva; Sinval Pinto Brandão Filho; Tiago Gräf; Wheverton Ricardo Correia do Nascimento |
| EPI_ISL_541372, EPI_ISL_541386 | Laboratório Central de Saúde Pública do Estado de Sergipe (LACEN-SE) | Laboratory of Respiratory Viruses and Measles, Oswaldo Cruz Institute, FIOCRUZ | Ana Carolina Mendonça; Anna Carolina Paixão; Clioma Santos; Fernando Motta; Jonathan Lopes; Luciana Appolinario; Marilda Siqueira on behalf of the Fiocruz COVID-19 Genomic Surveillance Network; Paola Resende |
| EPI_ISL_801397, EPI_ISL_801398, EPI_ISL_801399, EPI_ISL_801400, EPI_ISL_801401, EPI_ISL_801402, EPI_ISL_801403 | | | |
| see above | Laboratório Central de Saúde Pública do Estado do Amazonas (LACEN-AM) | Laboratorio de Ecologia de Doencas Transmissiveis na Amazonia, Instituto Leonidas e Maria Deane - Fiocruz Amazonia | André Corado; Debora Duarte; Felipe Naveca on behalf of the Fiocruz COVID-19 Genomic Surveillance Network; Fernanda Nascimento; George Silva; Luciana Gonçalves; Maria Júlia Brandão; Michele Jesus; Valdinete Nascimento; Victor Souza; Ágatha Costa |
| EPI_ISL_541343, EPI_ISL_541344, EPI_ISL_792647, EPI_ISL_792649, EPI_ISL_792653, EPI_ISL_792654 | Laboratório Central de Saúde Pública do Estado do Paraná (LACEN-PR) | Laboratory of Respiratory Viruses and Measles, Oswaldo Cruz Institute, FIOCRUZ | Ana Carolina Mendonca; Ana Carolina Mendonça; Anna Carolina Paixao; Anna Carolina Paixão; Fernando Motta; Irina Nastassja Riediger; Irina Riediger; Jonathan Lopes; Luciana Appolinario; Maria do Carmo Debur; Marilda Siqueira on behalf of the Fiocruz COVID-19 Genomic Surveillance Network; Paola Resende |
| EPI_ISL_729801, EPI_ISL_729803, EPI_ISL_729805, EPI_ISL_729806, EPI_ISL_729808, EPI_ISL_729813, EPI_ISL_729840, EPI_ISL_729845, EPI_ISL_729852, EPI_ISL_729853, EPI_ISL_729854, EPI_ISL_729856, EPI_ISL_729861 | | | |
| see above | Laboratório Central de Saúde Pública do Estado do Rio Grande do Sul (LACEN-RS) | Laboratory of Respiratory Viruses and Measles, Oswaldo Cruz Institute, FIOCRUZ | Ana Carolina Mendonça; Anna Carolina Paixão; Fernando Motta; Luciana Appolinario; Marilda Siqueira on behalf of the Fiocruz COVID-19 Genomic Surveillance Network; Marilda Tereza Mar da Rosa; Paola Resende; Tatiana Schaffer Gregianini |
| EPI_ISL_2248770, EPI_ISL_2298750 | Laboratório Central de Saúde Pública do Maranhão | Coordenação Geral de Laboratórios de Saúde Pública (CGLAB/DAEVS/SVS/MS) | Vagner Fonseca; et al. |
| EPI_ISL_2241593, EPI_ISL_2241607, EPI_ISL_2246287 | Laboratório Central de Saúde Pública do Piaui | Coordenação Geral de Laboratórios de Saúde Pública (CGLAB/DAEVS/SVS/MS) | Vagner Fonseca; et al. |
| EPI_ISL_2241508 | Laboratório Central de Saúde Pública do Rio Grande do Norte | Coordenação Geral de Laboratórios de Saúde Pública (CGLAB/DAEVS/SVS/MS) | Vagner Fonseca; et al. |
| EPI_ISL_1182547, EPI_ISL_1182610, EPI_ISL_1182621, EPI_ISL_1182623, EPI_ISL_2249348, EPI_ISL_2249352, EPI_ISL_2249353, EPI_ISL_2249355, EPI_ISL_2249379, EPI_ISL_2249382 | | | |
| see above | Laboratório Central de Saúde Pública do Rio Grande do Sul | Coordenação Geral de Laboratórios de Saúde Pública (CGLAB/DAEVS/SVS/MS) | Vagner Fonseca; et al. |
| EPI_ISL_2249404, EPI_ISL_2249405, EPI_ISL_2249407, EPI_ISL_2249409, EPI_ISL_2249426 | Laboratório Central de Saúde Pública do Rio de Janeiro | Coordenação Geral de Laboratórios de Saúde Pública (CGLAB/DAEVS/SVS/MS) | Vagner Fonseca; et al. |
| EPI_ISL_1182562, EPI_ISL_1182567, EPI_ISL_1182584, EPI_ISL_1182613, EPI_ISL_1182614 | Laboratório Central do Estado do Paraná | Coordenação Geral de Laboratórios de Saúde Pública (CGLAB/DAEVS/SVS/MS) | Vagner Fonseca; et al. |
| EPI_ISL_1182554 | Laboratório Central do Estado do Rio de Janeiro | Coordenação Geral de Laboratórios de Saúde Pública (CGLAB/DAEVS/SVS/MS) | Vagner Fonseca; et al. |
| EPI_ISL_1213309, | Laboratório | Bioinformatics | Alessandra P Lamarca; Alexandra L Gerber; Ana Paula Melo Mariano; Ana Paula de C Guimarães; Ana Tereza R Vasconcelos; Angela Maria Guimarães Santos; Bianca Mendes Maciel; Danielle Angst Secco; Eduardo Sérgio Soares Sousa; Eloiza Helena Campana; Francisco Paulo Freire Neto; George Rego Albuquerque; Kátia Castanho |

| | | | |
|---|---|---|---|
| EPI_ISL_1213397 | HLA/UERJ | Laboratory / LNCC | Scortecci; Lucymara Fassarella Agnez Lima; Luiz G P de Almeida; Luís Cristóvão Porto; Otavio J. Brustolini; Paulo Ricardo Nascimento; Ronaldo da Silva Francisco Jr; Sandra Rocha Gadelha; Selma Maria Bezerra Jeronimo; Vinícius Pietta Perez |
| EPI_ISL_693220, EPI_ISL_693223, EPI_ISL_693224, EPI_ISL_693243 | Laboratório Municipal de Piracicaba | Instituto Adolfo Lutz, Interdiciplinary Procedures Center, Strategic Laboratory | Claudia Regina Gonçalves; Claudio Tavares Sacchi; Erica Valessa Ramos Gomes; Karoline Rodrigues Campos |
| EPI_ISL_940608 | Laboratório Sao Lucas | Instituto Adolfo Lutz, Interdiciplinary Procedures Center, Strategic Laboratory | Claudia Regina Gonçalves; Claudio Tavares Sacchi; Erica Valessa Ramos Gomes; Karoline Rodrigues Campos |
| EPI_ISL_1494970, EPI_ISL_1495004, EPI_ISL_1497548 | Laboratório de Biologia Integrativa | Laboratório de Biologia Integrativa | Alessandro Clayton de Souza Ferreira; Aline Brito de Lima; Carolina Moreira Voloch; Daniel Costa Queiroz; Danielle Alves Gomes Zauli; Diego Menezes Bonfim; Filipe Romero Rebello Moreira; Frederico Scott Varella Malta; Joice do Prado Silva; Lucyene Miguita Luiz; Nuno Rodrigues Faria; Paula Luize Camargos Fonseca; Rafael Marques de Souza; Renan Pedra de Souza; Renato Santana Aguiar; Rennan Garcias Moreira; Victor Cavalcanti Pardini; Victor Emmanuel Viana Geddes |
| EPI_ISL_2466150, EPI_ISL_2466153, EPI_ISL_2466157, EPI_ISL_2466161, EPI_ISL_2466172, EPI_ISL_2466176, EPI_ISL_2466186, EPI_ISL_2466188, EPI_ISL_2466189, EPI_ISL_2466229 | | | |
| see above | Laboratório de Biologia Molecular de Doenças Infecciosas e do Câncer (LADIC - UFRN) | Laboratory of Respiratory Viruses and Measles, Oswaldo Cruz Institute, FIOCRUZ | Alice Sampaio Rocha; Ana Carolina Mendonca; Anna Carolina Paixao; Elisa Cavalcante Pereira; Fernando Motta; Josélio Araújo; Luciana Appolinario; Marilda Siqueira on behalf of the Fiocruz COVID-19 Genomic Surveillance Network; Paola Resende; Renata Serrano Lopes; Taina Venas |
| EPI_ISL_770558, EPI_ISL_770562, EPI_ISL_770569, EPI_ISL_770572, EPI_ISL_770576, EPI_ISL_770577, EPI_ISL_770582, EPI_ISL_770585, EPI_ISL_770586, EPI_ISL_770588, EPI_ISL_770590, EPI_ISL_770614, EPI_ISL_770623, EPI_ISL_770626, EPI_ISL_770627, EPI_ISL_770629, EPI_ISL_779156, EPI_ISL_779160, EPI_ISL_779161, EPI_ISL_779163, EPI_ISL_779165, EPI_ISL_779166, EPI_ISL_779167, EPI_ISL_779168 | | | |
| see above | Laboratório de Microbiologia Molecular - Universidade FEEVALE | Bioinformatics Laboratory / LNCC | Alana Witt Hansen; Alessandra Pavan Lamarca da Silva; Alexandra L Gerber; Ana Karolina Eisen Antunes; Ana Luiza Ziulkoski; Ana Paula de C Guimarães; Ana Tereza R de Vasconcelos; Bruna Hermann; Fagner Henrique Heldt; Felipe Benites; Fernando Rosado Spilki; Juliana Schons; Juliane Deise Fleck; Karoline Schallenberger; Larissa Mallmann; Luiz G P de Almeida; Matheus Nunes Weber; Meriane Demoliner; Paula Rodrigues de Almeida; Ronaldo da Silva F Jr; Vyctoria Goes |
| EPI_ISL_2918996, EPI_ISL_2918997, EPI_ISL_2918998, EPI_ISL_2918999, EPI_ISL_2919001, EPI_ISL_2919002, EPI_ISL_2919003, EPI_ISL_2919004, EPI_ISL_2919005, EPI_ISL_2919006, EPI_ISL_2919007, EPI_ISL_2919008, EPI_ISL_2919009, EPI_ISL_2919010, EPI_ISL_2919011, EPI_ISL_2919015, EPI_ISL_2919016, EPI_ISL_2925736, EPI_ISL_2928507, EPI_ISL_2928508 | | | |
| see above | Laboratório de Microbiologia Molecular - Universidade FEEVALE | Laboratório de Microbiologia Molecular - Universidade FEEVALE | Alana Witt Hansen; Fernando Rosado Spilki; Flávio Silveira; Fágner Henrique Heldt; Juliana Schons Gulart; Juliana Schons Gularte; Juliane Deise Fleck; Mariana Soares da Silva; Matheus Nunes Weber; Meriane Demoliner; Michele Filippi.; Micheli Filippi.; Paula Rodrigues de Almeida |
| EPI_ISL_1799499, EPI_ISL_1799505, EPI_ISL_2431431, EPI_ISL_2928344 | Laboratório de Microbiologia Molecular - Universidade FEEVALE | Molecular Microbiology Laboratory | Alana Witt Hansen; Fernando Rosado Spilki; Flávio Silveira; Fágner Henrique Heldt; Juliana Schons Gulart; Juliana Schons Gularte; Juliane Deise Fleck; Mariana Soares da Silva; Matheus Nunes Weber; Meriane Demoliner; Michele Filippi.; Micheli Filippi.; Paula Rodrigues de Almeida |
| EPI_ISL_831645, EPI_ISL_831660, EPI_ISL_831688, EPI_ISL_831689, EPI_ISL_831938, EPI_ISL_832009, EPI_ISL_832011 | | | |
| see above | Laboratório de Microbiologia Molecular - Universidade FEEVALE | Universidade Federal de Ciências da Saúde de Porto Alegre | Amanda de Menezes Mayer; Carla Andretta Moreira Neves; Claudia Elizabeth Thompson; Fernando Rosado Spilki; Gabriel Dickin Caldana; Gabriela Bettella Cybis; Lívia Kmetzsch; Patrícia Aline Gröhs Ferrareze; Ricardo Ariel Zimerman; Vinícius Bonetti Franceschi |
| EPI_ISL_476341, EPI_ISL_476395, EPI_ISL_476398 | Laboratório de Patologia Clínica - UNICAMP | Laboratório de Estudos de Vírus Emergentes - UNICAMP | Angelica Schreiber; Camila Simeoni; Darlan da Silva Candido; Jaqueline Goes Jesus e William Marciel de Souza; José Luiz Proença-Modena; Julia Forato; Julien Theze; Luiz Gonzaga; Magnun Nueldo Nunes dos Santos; Marcilio Jorge Fumagalli; Mariene Ribeiro Amorim; Nuno Rodrigues Faria |
| EPI_ISL_1785610, EPI_ISL_1785612 | Laboratório de Pesquisa em Virologia, FAMERP, SJRP | Laboratório de Pesquisa em Virologia, FAMERP, SJRP | Cecília Artico Banho; Cíntia Bittar; Fábio Sossai Possebon; Guilherme Campos; Helena Lage Ferreira; Jorge A. Petroli Marchesi; João Pessoa Araújo Jr.; Leila Sabrina Ullmann; Lívia Sacchetto; Maisa C. Pereira Parra; Marília Moraes; Maurício L. Nogueira; Paula Rahal; Paulo Inacio da Costa |
| EPI_ISL_1464675, EPI_ISL_1464677 | Laboratório de Virologia - UNIFESP | Laboratory of Respiratory Viruses and Measles, Oswaldo Cruz Institute, FIOCRUZ | Alice Sampaio Rocha; Ana Carolina Mendonca; Anna Carolina Paixao; Fernando Motta; Luciana Appolinario; Marilda Siqueira on behalf of the Fiocruz COVID-19 Genomic Surveillance Network; Nancy Belei; Paola Resende; Renata Serrano Lopes |
| EPI_ISL_2629608, EPI_ISL_2629624, EPI_ISL_2629634 | Laboratório de Virologia Molecular - Universidade Federal do Rio de Janeiro | Laboratório de Virologia Molecular - Universidade Federal do Rio de Janeiro | ; Alice Laschuk Herlinger; Amilcar Tanuri; André Felipe Andrade dos Santos; Carolina Moreira Voloch; Cássia Cristina Alves Gonçalves; Diana Mariani; Débora Souza Faffe; Filipe Romero Rebello Moreira; Francine Bittencourt Schiffler; Isabela de Carvalho Leitão; Marcelo Calado de Paula Tôrres; Matheus Augusto Calvano Cosentino; Mirela D'arc; Orlando da Costa Ferreira Junior; Rafael Mello Galliez; Raíssa Mirella dos Santos Cunha da Costa; Renato Santana de Aguiar; Terezinha Marta Pereira Pinto Castineiras; Thamiris dos Santos Miranda; Átila Duque Rossi |
| EPI_ISL_2196357, EPI_ISL_2196360, EPI_ISL_2677096, EPI_ISL_2677099, EPI_ISL_2677126, EPI_ISL_2677234, EPI_ISL_2677244, EPI_ISL_2677289, EPI_ISL_2677291, EPI_ISL_2677292, EPI_ISL_2677299, EPI_ISL_2677300, EPI_ISL_2677307, EPI_ISL_2677308, EPI_ISL_3061901 | | | |
| see above | Labortorio Central de Saude Publica do Estado de Santa Catarina (LACEN/SC) | Laboratory of Respiratory Viruses and Measles, Oswaldo Cruz Institute, FIOCRUZ | Alice Sampaio Rocha; Ana Carolina Mendonca; Anna Carolina Paixao; Darcita Buerger Rovaris; Elisa Cavalcante Pereira; Fernando Motta; Luciana Appolinario; Marilda Siqueira on behalf of the Fiocruz COVID-19 Genomic Surveillance Network; Paola Resende; Renata Serrano Lopes; Sandra Bianchini Fernandes; Taina Venas |
| EPI_ISL_2614351 | Labortorio Central de Saude Publica do Estado do Rio de Janeiro (LACEN/RJ) | Laboratory of Respiratory Viruses and Measles, Oswaldo Cruz Institute, FIOCRUZ | Alice Sampaio Rocha; Ana Carolina Mendonca; Andrea Cony Cavalcanti; Anna Carolina Paixao; Elisa Cavalcante Pereira; Fernando Motta; Luciana Appolinario; Marilda Siqueira on behalf of the Fiocruz COVID-19 Genomic Surveillance Network; Paola Resende; Renata Serrano Lopes; Taina Venas |
| EPI_ISL_1358302 | Lacen de Tocantins | Instituto Adolfo Lutz, Interdiciplinary Procedures Center, Strategic Laboratory | Caio Vinicius Dias Lopes; Claudia Regina Gonçalves; Claudio Tavares Sacchi; Erica Valessa Ramos Gomes; Karoline Rodrigues Campos |
| EPI_ISL_2157343, EPI_ISL_2196362 | Lboratorio Central de Saude Publica do Estado do Parana (LACEN/PR) | Laboratory of Respiratory Viruses and Measles, Oswaldo Cruz Institute, FIOCRUZ | Alice Sampaio Rocha; Ana Carolina Mendonca; Anna Carolina Paixao; Elisa Cavalcante Pereira; Fernando Motta; Irina Riediger; Luciana Appolinario; Marilda Siqueira on behalf of the Fiocruz COVID-19 Genomic Surveillance Network; Paola Resende; Renata Serrano Lopes; Taina Venas |
| EPI_ISL_703236, EPI_ISL_730661 | Lighthouse Lab in Alderley Park | Wellcome Sanger Institute for the COVID-19 Genomics UK (COG-UK) Consortium | Cordelia Langford; David K. Jackson; Dominic Kwiatkowski; Ewan Harrison; Ian Johnston; Jacquelyn Wynn; John Sillitoe on behalf of the Wellcome Sanger Institute COVID-19 Surveillance Team; Mairead Hyland; Roberto Amato; Sonia Goncalves; The Lighthouse Lab in Alderley Park and Alex Alderton |
| EPI_ISL_551467 | Lighthouse Lab in Alderley Park | Wellcome Sanger Institute for the COVID-19 Genomics UK (COG-UK) consortium | Cordelia Langford; David K. Jackson; Dominic Kwiatkowski; Ewan Harrison; Ian Johnston; John Sillitoe on behalf of the Wellcome Sanger Institute COVID-19 Surveillance Team (http://www.sanger.ac.uk/covid-team); Roberto Amato; Sonia Goncalves; The Lighthouse Lab in Alderley Park and Alex Alderton |
| EPI_ISL_673528, EPI_ISL_857679 | Lighthouse Lab in Cambridge | Wellcome Sanger Institute for the COVID-19 Genomics UK (COG-UK) Consortium | Cordelia Langford; David K. Jackson; Dominic Kwiatkowski; Ewan Harrison; Ian Johnston; John Sillitoe on behalf of the Wellcome Sanger Institute COVID-19 Surveillance Team; Rob Howes; Roberto Amato; Sonia Goncalves; The Lighthouse Lab in Cambridge and Alex Alderton |
| EPI_ISL_590506 | Lighthouse Lab in Glasgow | Wellcome Sanger Institute for the COVID-19 Genomics UK (COG-UK) consortium | Anna Dominiczak and Alex Alderton; Carol Clugston; Cordelia Langford; David Gray; David K. Jackson; Dominic Kwiatkowski; Ewan Harrison; Harper VanSteenhouse; Ian Johnston; John Sillitoe on behalf of the Wellcome Sanger Institute COVID-19 Surveillance Team (http://www.sanger.ac.uk/covid-team); Roberto Amato; Sonia Goncalves; Yumi Kasai |
| EPI_ISL_760881, EPI_ISL_760963, EPI_ISL_3954939 | Lighthouse Lab in Milton Keynes | Wellcome Sanger Institute for the COVID-19 Genomics UK (COG-UK) Consortium | Cordelia Langford; David K. Jackson; Dominic Kwiatkowski; Ewan Harrison; Ian Johnston; Jeffrey Barrett; John Sillitoe on behalf of the Wellcome Sanger Institute COVID-19 Surveillance Team; Roberto Amato; Sonia Goncalves; The Lighthouse Lab in Milton Keynes and Alex Alderton |
| EPI_ISL_549833, EPI_ISL_629164, | Lighthouse Lab in Milton Keynes | Wellcome Sanger Institute for the COVID- | Cordelia Langford; David K. Jackson; Dominic Kwiatkowski; Ewan Harrison; Ian Johnston; John Sillitoe on behalf of the Wellcome Sanger Institute COVID-19 Surveillance Team; John Sillitoe on behalf of the Wellcome Sanger Institute COVID-19 Surveillance Team (http://www.sanger.ac.uk/covid-team); Roberto Amato; Sonia Goncalves; The Lighthouse Lab in Milton Keynes and Alex Alderton |

| | | | |
|---|---|---|---|
| EPI_ISL_630998, EPI_ISL_631036 | | 19 Genomics UK (COG-UK) consortium | |
| EPI_ISL_1227371, EPI_ISL_1227396, EPI_ISL_1227400, EPI_ISL_1227427, EPI_ISL_1227445, EPI_ISL_1227446, EPI_ISL_1227933, EPI_ISL_1228176, EPI_ISL_1228572, EPI_ISL_1397621, EPI_ISL_1623976, EPI_ISL_2278731 | | | |
| see above | MONTEFIORE MEDICAL CENTER LABORATORIES | Wadsworth Center, New York State Department of Health | Alexis Russel; Alexis Russell; Catharine Prussing; Daryl M. Lamson; Erasmus Schneider; Erica Lasek-Nesselquist; John Kelly; Jonathan Plitnick; Kirsten St. George; Matthew Shudt; Melissa A Leisner; Navjot Singh |
| EPI_ISL_802105, EPI_ISL_802106, EPI_ISL_1300937, EPI_ISL_1301233, EPI_ISL_1301238, EPI_ISL_1709347, EPI_ISL_1709917 | | | |
| see above | MSHS Clinical Microbiology Laboratories | MSHS Pathogen Surveillance Program | Adolfo García-Sastre; Adriana van de Guchte; Ajay Obla; Alberto Paniz-Mondolfi; Ana S. Gonzalez-Reiche; Andrew Kasarskis; Angela Amoako; Ashley S. Salimbangon; Ashley Salimbangon; Betsaida Salom Melo; Bremy Alburquerque; Brianne Ciferri; Charles Gleason; Daniel Floda; Deena R. Altman; Denise Jurczyszak; Elena Hirsch; Emilia Mia Sordillo; Emily Ferreri; Gintaras Deikus; Giulio Kleiner; Gopi Patel; Hala Alshammary; Harm van Bakel; Irina Oussenko; Jayeeta Dutta; Juan Soto; Julia Matthews; Katherine Beach; Kathryn Twyman; Kayla Russo; Komal Srivastava; Levy Sominsky; Mahmoud Awawda; Marta Luksza; Matthew M. Hernandez; Melissa Gitman; Michael D. Nowak; Mitchell J. Sullivan; Nancy Francoeur; Rachel Chernet; Robert Sebra; Sarah Schaefer; Shelcie Fabre; Shwetha Hara Sridhar; Viviana Simon; Ying-Chih Wang; Zenab Khan |
| EPI_ISL_2691009 | MSK Microbiology Lab | MSK Microbiology Lab | Esther Babady; Krupa Jani; Tracy McMillen |
| EPI_ISL_1152591 | MVZ Medizinisches Labor Hannover GmbH | Robert Koch Institute | |
| EPI_ISL_1719809 | Microbiology Department, Laboratori Clínic Metropolitana Nord. Hospital Universitari Germans Trias i Pujol. | Can Ruti SARS-CoV-2 Sequencing Hub (HUGTiP/IrsiCaixa/IGTP) | Alba Sánchez; Anna Not; Antoni E Bordoy; Bonaventura Clotet; Cristina Casañ; Cristina Esteban; Francesc Catala-Moll; Gemma Clara; Ignacio Blanco; Marc Noguera-Julian; Maria Casadellà; Mariona Parera; Mercedes Guerrero; Montserrat Giménez; Pere-Joan Cardona; Pilar Armengol; Roger Paredes; Verónica Saludes; and Elisa Martró on behalf of the Can Ruti SARS-CoV-2 Sequencing Hub. |
| EPI_ISL_547433, EPI_ISL_547434, EPI_ISL_547435, EPI_ISL_547437 | Microbiology, Department of Pathology, St. Bernard's Hospital, Gibraltar Health Authority | Respiratory Virus Unit, Microbiology Services Colindale, Public Health England | Charlotte Gillborn-Jones (Gibraltar); Dr Nicholas Cortes (Gibraltar); PHE Covid Sequencing Team |
| EPI_ISL_735413 | Militello Centro de Diagnosticos e Biopesquisa Clinica | Instituto Adolfo Lutz, Interdiciplinary Procedures Center, Strategic Laboratory | Claudia Regina Gonçalves; Claudio Tavares Sacchi; Erica Valessa Ramos Gomes; Karoline Rodrigues Campos |
| EPI_ISL_1534512, EPI_ISL_1911798, EPI_ISL_3256939, EPI_ISL_3406510, EPI_ISL_3406753, EPI_ISL_3414403, EPI_ISL_3414677 | | | |
| see above | Ministry of Health Turkey | Ministry of Health Turkey | Fatma Bayrakdar; Gulay Korukluoglu; Gülay Korukluoğlu; Suleyman Yalcin; Süleyman Yalcin; Yasemin Cosgun; Yasemin Coşgun |
| EPI_ISL_1714420, EPI_ISL_1714447, EPI_ISL_1714459, EPI_ISL_1714483, EPI_ISL_1714533, EPI_ISL_1714591, EPI_ISL_1714616, EPI_ISL_1714620 | | | |
| see above | Ministry of Public Health / Hamad Medical Corporation | Weill Cornell Medical College - Qatar (WCM-Q), Genomics Core Laboratory / Qatar Genome Project (QGP) | Chadi Saad MOPH and HMC: Abdullatif Al-Khal; Dina Elgakhlab; Einas A. E. Al-Kuwari; Hamad E. Al-Romaihi; Hamda Alromaihi; Joel A Malek. QGP: Fatima H. Al-Kuwari; Laith Abu-Raddad; Mashael A. Al-Bader; Meryem Bensaad; Mohammed Al-Thani; Muna A. S. Al-Maslamani; Peter V. Coyle; Reham A. El-Kahlout. QBB: Tasneem Al-Hamad; Roberto Bertollini; Salih Al-Marri; Shameem Younuskunju; WCMQ: Ayeda A. Ahmed; Yasmin Mohamoud |
| EPI_ISL_1597713, EPI_ISL_1597714 | Montefiore Medical Center | Abbott | Amy Fox; Ana Olivo; Ana Vallari; Barbara Harris; Gavin Cloherty; Mary Rodgers; Todd Meyer; Yitz Goldstein |
| EPI_ISL_1017531 | Murphy Medical Associates | Grubaugh Lab - Yale School of Public Health | Annie Watkins; Caleb Neal; Chantal Vogels; Joseph Fauver; Mallery Breban; Mary Petrone; Nathan Grubaugh; Steven Murphy; Tara Alpert |
| EPI_ISL_1966563 | NUCLEO DE SAUDE VILA FALCAO DE BAURU | Instituto Butantan / Mendelics | Antonio Jorge Martins; Bianca Cechetto Carlos. Mendelics: Bibiana Santos; Claudia Renata dos Santos Barros; Cíntia Bittar; David Schlesinger. Hemocentro Ribeirão Preto: Simone Kashima; Debora Botequio Moretti; Elaine Cristina Marqueze; Elaine Vieira dos Santos; Elisangela Chicaroni Mattos; Erika Freitas; Evandra Strazza Rodrigues; Felipe Allan da Silva da Costa; Flavia Aburjaile; Fábio Sossai Possebon; Guilherme Campos; Guilherme Targino Valente; Heidge Fukumasu. USP-Botucatu: Rejane Maria Tommasini Grotto; Helena Lage Ferreira; Instituto Butantan: Dimas Tadeu Covas; Jardelina de Souza Todao Bernardino; Jayme A. Souza-Neto; Jessika Cristina Chagas Lesbon; Jorge A. Petroli Marchesi; José Salvatore Leister Patané; João Paulo Kitajima; João Pessoa Araújo Jr.; Leila Sabrina Ullmann; Loyze Paola Oliveira de Lima; Luiz Aurelio de Campos Crispin. Centro de Genômica Funcional da ESALQ: Luiz Lehmann Coutinho; Luiz Carlos Junior de Alcantara; Lívia Sacchetto; Maisa C. Pereira Parra; Maria Carolina Elias; Marta Giovanetti; Marília Moraes; Maurício Lacerda Nogueira. Prefeitura de Sao Paulo: Melissa Palmieri.; Patricia Akemi Assato; Paula Rahal; Paulo Inacio da Costa; Rafael dos Santos Bezerra; Raquel de Lello Rocha Campos Cassano. NGS Soluções Genômicas: Pilar Drummond Sampaio Corrêa Mariani. FZEA-USP Pirassununga: Mirele Daiana Poleti; Raul Machado Neto; Ricardo Augusto Brassaloti; Ricardo Haddad; Rodrigo Tocantins Calado. FAMERP-SJRP: Cecília Artico Banho; Sandra Coccuzzo Sampaio; Svetoslav Nanev Slavov; Vagner Fonseca; Vincent Louis Viala |
| EPI_ISL_3046174, EPI_ISL_3046188, EPI_ISL_3046190, EPI_ISL_3046193, EPI_ISL_3046196, EPI_ISL_3046199, EPI_ISL_3046200, EPI_ISL_3046201, EPI_ISL_3046204, EPI_ISL_3046207, EPI_ISL_3046214, EPI_ISL_3046216, EPI_ISL_3046219, EPI_ISL_3046228, EPI_ISL_3046230, EPI_ISL_3046232, EPI_ISL_3046238, EPI_ISL_3046239, EPI_ISL_3046244, EPI_ISL_3046245, EPI_ISL_3046260, EPI_ISL_3046262, EPI_ISL_3060269, EPI_ISL_3060271, EPI_ISL_3060272, EPI_ISL_3060273, EPI_ISL_3134727, EPI_ISL_3134738, EPI_ISL_3134741, EPI_ISL_3134742, EPI_ISL_3134743, EPI_ISL_3134744, EPI_ISL_3134748, EPI_ISL_3134751, EPI_ISL_3134819, EPI_ISL_3134821, EPI_ISL_3134822, EPI_ISL_3134827, EPI_ISL_3134828, EPI_ISL_3134829, EPI_ISL_3703673 | | | |
| see above | NUPIT/UFPE | WallauLab on behalf of Fiocruz COVID-19 Genomic Surveillance Network | Alexandre Freitas da Silva; Cassia Docena; Constância Flávia Junqueira Ayres; Filipe Zimmer Dezordi; Gabriel Luz Wallau; Gustavo Barbosa de Lima; Lais Ceschini Machado; Lilian Caroliny Amorim Silva; Maira Galdino da Rocha Pitta; Marcelo Henrique dos Santos Paiva; Matheus Filgueira Bezerra; Michelly Cristiny Pereira; Rômulo Pessoa e Silva; Sinval Pinto Brandão Filho |
| EPI_ISL_1227698 | NYC Pandemic Response Lab | Wadsworth Center, New York State Department of Health | Alexis Russel; Daryl M. Lamson; Erasmus Schneider; Erica Lasek-Nesselquist; John Kelly; Jonathan Plitnick; Kirsten St. George; Matthew Shudt; Melissa A Leisner; Navjot Singh |
| EPI_ISL_1379204 | NYP-WCM | New York Genome Center | Andre Corvelo; Arryn Craney; Chris Mason; Dayna M. Oschwald; Hanna Rennert; Lars F Westblade; Margaret Elizabeth Ross; Melissa Cushing; Michael Zody; Olivier Elemento; Priya Velu; Samantha Fennessey; Tom Maniatis |
| EPI_ISL_1415196 | National Centre For Cell Science | National Centre For Cell Science – INSACOG | Ajay Pillai; Dhiraj Paul; INSACOG Consortium team; Manoj Kumar Bhat; Mitali Inamdar; Mohak P Gujare; Shivang P. Bhanushali; Sonal Manik Chavan; Yogesh Shouche. |
| EPI_ISL_416036 | National Influenza Center - Instituto Adolfo Lutz | Instituto Adolfo Lutz, Interdiciplinary Procedures Center, Strategic Laboratory | Adriana Bugno; Adriano Abbud; Carlos Henrique Camargo; Claudia Regina Gonçalves; Claudio Tavares Sacchi; Daniela Bernardes Borges da Silva; Erica Valessa Ramos Gomes; Fabiana Cristina Pereira dos Santos; Maria do Carmo Sampaio Tavares Timenetsky; Simone Guadagnucci Morillo; Terezinha Maria de Paiva |
| EPI_ISL_1447440 | National Institute of Infectious Diseases (NIID) | National Institute of Infectious Diseases (NIID) | Hideka Miura; Kentaro Itokawa; Kiyoko Okamoto; Kumiko Araki; Makoto Kuroda; Masanori Hashino; Rina Tanaka; Seiichiro Fujisaki; Shinichiro Hirai; Tsuyoshi Sekizuka |
| EPI_ISL_728187 | National Public Health Laboratory, National Centre for Infectious Diseases | National Public Health Laboratory, National Centre for Infectious Diseases | Lin Cui; Raymond Tzer Pin Lin; Sophie Octavia; Tze Minn Mak; Zhenyang Zhou |
| EPI_ISL_875351 | National Virus Reference Laboratory | National Virus Reference Laboratory | Cillian F De Gascun; Gabriel Gonzalez; Jonathan Dean; Michael Carr |
| EPI_ISL_2105404, EPI_ISL_2105410, EPI_ISL_2105412, EPI_ISL_2105416, EPI_ISL_2105419, EPI_ISL_2105422, EPI_ISL_2105423, EPI_ISL_2105427, EPI_ISL_2105431, EPI_ISL_2105435, EPI_ISL_2105439, EPI_ISL_2105444, EPI_ISL_2105450, EPI_ISL_2105451, EPI_ISL_2105452, EPI_ISL_2105460 | | | |
| see above | Negros Oriental Provincial Hospital | Philippine Genome Center | Alethea R. de Guzman; Anna Ong-Lim; Arianne A. Zamora; Asia Louisa U. Chong; Benedict A. Maralit; Candice Francheska B. Tambaoan; Carlo M. Lapid; Celia Carlos; Devon Ray Pacial; Edsel Maurice Salvaña; El King D. Morado; Eva Maria Cutiongco-de la Paz; Francis A. Tablizo; Irish Coleen A. Asin; Jaime C. Montoya; Jan Michael C. Yap; Jo-Hannah S. Llames; John Q. Wong; Joshua Gregor A. Dizon; Juan Antonio R. Magalang; Karol Sophia Agape R. Padilla; Kenneth M. Kim; Kris P. Punayan; Marc Edsel C. Ayes; Marc Jerrone R. Castro; Maria Rosario Singh-Vergeire and Cynthia P. Saloma; Maria Sofia L. Yangzon; Marissa Alejandria; Razel Nikka M. Hao; Rianna Patricia S. Cruz; Sheila Mae M. Araiza |
| EPI_ISL_896351, EPI_ISL_1016306, EPI_ISL_1397496, EPI_ISL_1397501, EPI_ISL_1624013, EPI_ISL_1624022, EPI_ISL_1624056, EPI_ISL_1624076, EPI_ISL_1624081 | | | |
| see above | New York Presbyterian Hospital | Wadsworth Center, New York State Department of Health | Alexis Russel; Alexis Russell; Catharine Prussing; Daryl M. Lamson; Erasmus Schneider; Erica Lasek-Nesselquist; John Kelly; Jonathan Plitnick; Kirsten St. George; Matthew Shudt; Melissa A Leisner; Navjot Singh |
| EPI_ISL_2105401, EPI_ISL_2105556, EPI_ISL_2156485, EPI_ISL_2156584, EPI_ISL_2156591, EPI_ISL_2156592, EPI_ISL_2156593, EPI_ISL_2189279 | | | |
| see above | Northern Mindanao TB Regional Center | Philippine Genome Center | Alethea R. de Guzman; Anna Ong-Lim; Arianne A. Zamora; Asia Louisa U. Chong; Benedict A. Maralit; Candice Francheska B. Tambaoan; Carlo M. Lapid; Celia Carlos; Devon Ray Pacial; Edsel Maurice Salvaña; El King D. Morado; Elcid Aaron R. Pangilinan; Eva Maria Cutiongco-de la Paz; Francis A. Tablizo; Irish Coleen A. Asin; Jaime C. Montoya; Jan Michael C. Yap; Jo-Hannah S. Llames; John Q. Wong; Joshua Gregor A. Dizon; Juan Antonio R. Magalang; Karol Sophia Agape R. Padilla; Kenneth M. Kim; Kris P. Punayan; Marc Edsel C. Ayes; Marc Jerrone R. Castro; Maria Rosario Singh-Vergeire and Cynthia P. Saloma; Maria Sofia L. Yangzon; Marissa Alejandria; Razel Nikka M. Hao; Renato Jacinto Q. Mantaring; Rianna Patricia S. Cruz; Sheila Mae M. Araiza |
| EPI_ISL_534326 | Notre Dame Intermedica Saude AS | Instituto Adolfo Lutz, Interdiciplinary Procedures Center, Strategic Laboratory | Claudia Regina Gonçalves; Claudio Tavares Sacchi; Erica Valessa Ramos Gomes |
| EPI_ISL_1117384, EPI_ISL_1117388, | Nucleo de Pesquisa em Inovacao | LABBE, Federal University of | Bruno Sampaio; Heidi Lacerda Alves da Cruz; Maira Galdino da Rocha Pitta; Marco Katzenberger; Marcos da Silveira Regueira Neto; Michelly Cristiny Pereira; Reginaldo Goncalves de Lima Neto; Valdir de Queiroz Balbino; Wilson Jose da Silva Junior |

| | | | |
|---|---|---|---|
| EPI_ISL_1117408, EPI_ISL_1117429 | Terapeutica - UFPE | Pernambuco | |
| EPI_ISL_534316 | OS Mun Santana Lauro Ribas Braga | Instituto Adolfo Lutz, Interdiciplinary Procedures Center, Strategic Laboratory | Claudia Regina Gonçalves; Claudio Tavares Sacchi; Erica Valessa Ramos Gomes |
| EPI_ISL_1131143, EPI_ISL_1430700 | Osaka Institute of Public Health, Morinomiya Center | Pathogen Genomics Center, National Institute of Infectious Diseases | Kentaro Itokawa; Makoto Kuroda; Masanori Hashino; Rina Tanaka; Tsuyoshi Sekizuka |
| EPI_ISL_2171170 | Ospital ng ParaÃ±aque II | Philippine Genome Center | Alethea R. de Guzman; Anna Ong-Lim; Arianne A. Zamora; Asia Louisa U. Chong; Benedict A. Maralit; Candice Francheska B. Tambaoan; Carlo M. Lapid; Celia Carlos; Devon Ray Pacial; Edsel Maurice Salvaña; El King D. Morado; Elcid Aaron R. Pangilinan; Eva Maria Cutiongco-de la Paz; Francis A. Tablizo; Irish Coleen A. Asin; Jaime C. Montoya; Jan Michael C. Yap; Jo-Hannah S. Llames; John Q. Wong; Joshua Gregor A. Dizon; Juan Antonio R. Magalang; Karol Sophia Agape R. Padilla; Kenneth M. Kim; Kris P. Punayan; Marc Edsel C. Ayes; Marc Jerrone R. Castro; Maria Rosario Singh-Vergeire and Cynthia P. Saloma; Maria Sofia L. Yangzon; Marissa Alejandria; Razel Nikka M. Hao; Rianna Patricia S. Cruz; Sheila Mae M. Araiza |
| EPI_ISL_2661879, EPI_ISL_2661903 | Oswaldo Cruz Institute, FIOCRUZ/CE | Analytical Competence Molecular Epidemiology Lab/ACME, Oswaldo Cruz Foundation, Ceara (FIOCRUZ CE) | Alice Sampaio Rocha; Ana Carolina Mendonca; Ana Carolina Paixao; Elisa Cavalcante Pereira; Fabio Miyajima; Fernando Motta; Luciana Appolinario; Marilda Siqueira on behalf of the Fiocruz COVID-19 Genomic Surveillance Network; Paola Resende; Renata Serrano Lopes; Taina Venas |
| EPI_ISL_638782, EPI_ISL_651545, EPI_ISL_652142 | Oxford Viromics, NDM, University of Oxford; Oxford University Hospitals; Basingstoke and North Hampshire Hospital | COVID-19 Genomics UK (COG-UK) Consortium | Alex Mobbs; Amy Trebes; Anita Justice; Catrin Moore; Christophe Fraser; David Bonsall; David Buck; Emma Wise; George Macintyre; Jessica Lynch; John Todd; Mariateresa de Cesare; Matilde Mori; Monique Andersson; Nathan Moore; Nick Cortes; Robert Shaw; Stephen Kidd; Tanya Golubchik; Timothy Peto |
| EPI_ISL_861673 | PA Novo Osasco | Instituto Adolfo Lutz, Interdiciplinary Procedures Center, Strategic Laboratory | Claudia Regina Gonçalves; Claudio Tavares Sacchi; Erica Valessa Ramos Gomes; Karoline Rodrigues Campos |
| EPI_ISL_2156328 | PHILIPPINE AIRPORT DIAGNOSTIC LABORATORY | Philippine Genome Center | Alethea R. de Guzman; Anna Ong-Lim; Arianne A. Zamora; Asia Louisa U. Chong; Benedict A. Maralit; Candice Francheska B. Tambaoan; Carlo M. Lapid; Celia Carlos; Devon Ray Pacial; Edsel Maurice Salvaña; El King D. Morado; Elcid Aaron R. Pangilinan; Eva Maria Cutiongco-de la Paz; Francis A. Tablizo; Irish Coleen A. Asin; Jaime C. Montoya; Jan Michael C. Yap; Jo-Hannah S. Llames; John Q. Wong; Joshua Gregor A. Dizon; Juan Antonio R. Magalang; Karol Sophia Agape R. Padilla; Kenneth M. Kim; Kris P. Punayan; Marc Edsel C. Ayes; Marc Jerrone R. Castro; Maria Rosario Singh-Vergeire and Cynthia P. Saloma; Maria Sofia L. Yangzon; Marissa Alejandria; Razel Nikka M. Hao; Rianna Patricia S. Cruz; Sheila Mae M. Araiza |
| EPI_ISL_1445090, EPI_ISL_1966104 | POLICLINICA COVID 19 ITAPETININGA | Instituto Butantan / Mendelics | Antonio Jorge Martins; Bianca Cechetto Carlos. Mendelics: Bibiana Santos; Bibiana Santos; Claudia Renata dos Santos Barros; Cíntia Bittar; David Schlesinger; David Schlesinger. Hemocentro Ribeirão Preto: Simone Kashima; Debora Botequio Moretti; Dimas Tadeu Covas; Elaine Cristina Marqueze; Elaine Vieira dos Santos; Elisangela Chicaroni Mattos; Erika Freitas; Evandra Strazza Rodrigues; Felipe Allan da Silva da Costa; Flavia Aburjaile; Fábio Sossai Possebon; Guilherme Campos; Guilherme Targino Valente; Heidge Fukumasu. USP-Botucatu: Rejane Maria Tommasini Grotto; Helena Lage Ferreira; Instituto Butantan: Dimas Tadeu Covas; Jardelina de Souza Todao Bernardino; Jayme A. Souza-Neto; Jessika Cristina Chagas Lesbon; Jorge A. Petroli Marchesi; José Salvatore Leister Patané; João Paulo Kitajima; João Pessoa Araújo Jr.; Leila Sabrina Ullmann; Loyze Paola Oliveira de Lima; Luiz Aurelio de Campos Crispin. Centro de Genômica Funcional da ESALQ: Luiz Lehmann Coutinho; Luiz Carlos Junior de Alcantara; Lívia Sacchetto; Maisa C. Pereira Parra; Maria Carolina Elias; Marta Giovanetti; Marília Moraes; Maurício Lacerda Nogueira. Prefeitura de Sao Paulo: Melissa Palmieri.; Patricia Akemi Assato; Paula Rahal; Paulo Inacio da Costa; Rafael dos Santos Bezerra; Raquel de Lello Rocha Campos Cassano. NGS Soluções Genômicas: Pilar Drummond Sampaio Corrêa Mariani. FZEA-USP Pirassununga: Mirele Daiana Poleti; Raul Machado Neto; Ricardo Augusto Brassaloti; Ricardo Haddad; Rodrigo Tocantins Calado.; Rodrigo Tocantins Calado. FAMERP-SJRP: Cecília Artico Banho; Sandra Coccuzzo Sampaio; Simone Kashima; Svetoslav Nanev Slavov; Vagner Fonseca; Vincent Louis Viala |
| EPI_ISL_1795399, EPI_ISL_2345349 | POLICLINICA HORTOLANDIA | Instituto Butantan / ESALQ-Piracicaba | Antonio Jorge Martins; Bianca Cechetto Carlos. Mendelics: Bibiana Santos; Claudia Renata dos Santos Barros; David Schlesinger; David Schlesinger. Hemocentro Ribeirão Preto: Simone Kashima; Debora Botequio Moretti; Debora Botequio Moretti. Centro de Genômica Funcional da ESALQ: Luiz Lehmann Coutinho; Dimas Tadeu Covas; Elaine Cristina Marqueze; Elaine Vieira Santos; Elaine Vieira dos Santos; Elisangela Chicaroni Mattos; Erika Freitas; Evandra Strazza Rodrigues; Felipe Allan da Silva da Costa; Flavia Aburjaile; Guilherme Targino Valente; Heidge Fukumasu; Heidge Fukumasu. USP-Botucatu: Rejane Maria Tommasini Grotto; Instituto Butantan: Alexander Roberto Precioso; Jayme A. Souza-Neto; Jayme Augusto de Souza-Neto; Jessika Cristina Chagas Lesbon; José Salvatore Leister Patané; João Paulo Kitajima; Luiz Alcantara; Luiz Carlos Junior de Alcantara; Luiz Lehmann Coutinho; Maria Carolina Elias; Marta Giovanetti; Maurício Lacerda Nogueira; Patricia Akemi Assato; Rafael dos Santos Bezerra; Raquel de Lello Rocha Campos Cassano. NGS Soluções Genômicas: Pilar Drummond Sampaio Corrêa Mariani. FZEA-USP Pirassununga: Mirele Daiana Poleti; Raul Machado Neto; Rejane Maria Tommasini Grotto; Ricardo Augusto Brassaloti; Ricardo Haddad; Rodrigo Tocantins Calado.; Sandra Coccuzzo Sampaio; Sandra Coccuzzo Sampaio Vessoni; Simone Kashima; Svetoslav Nanev Slavov; Vagner Fonseca; Vincent Louis Viala |
| EPI_ISL_1966691 | PRONTO ATENDIMENTO MUNICIPAL DE JACUPIRANGA | Instituto Butantan / Mendelics | Antonio Jorge Martins; Bianca Cechetto Carlos. Mendelics: Bibiana Santos; Claudia Renata dos Santos Barros; Cíntia Bittar; David Schlesinger. Hemocentro Ribeirão Preto: Simone Kashima; Debora Botequio Moretti; Elaine Cristina Marqueze; Elaine Vieira dos Santos; Elisangela Chicaroni Mattos; Erika Freitas; Evandra Strazza Rodrigues; Felipe Allan da Silva da Costa; Flavia Aburjaile; Fábio Sossai Possebon; Guilherme Campos; Guilherme Targino Valente; Heidge Fukumasu. USP-Botucatu: Rejane Maria Tommasini Grotto; Helena Lage Ferreira; Instituto Butantan: Dimas Tadeu Covas; Jardelina de Souza Todao Bernardino; Jayme A. Souza-Neto; Jessika Cristina Chagas Lesbon; Jorge A. Petroli Marchesi; José Salvatore Leister Patané; João Paulo Kitajima; João Pessoa Araújo Jr.; Leila Sabrina Ullmann; Loyze Paola Oliveira de Lima; Luiz Aurelio de Campos Crispin. Centro de Genômica Funcional da ESALQ: Luiz Lehmann Coutinho; Luiz Carlos Junior de Alcantara; Lívia Sacchetto; Maisa C. Pereira Parra; Maria Carolina Elias; Marta Giovanetti; Marília Moraes; Maurício Lacerda Nogueira. Prefeitura de Sao Paulo: Melissa Palmieri.; Patricia Akemi Assato; Paula Rahal; Paulo Inacio da Costa; Rafael dos Santos Bezerra; Raquel de Lello Rocha Campos Cassano. NGS Soluções Genômicas: Pilar Drummond Sampaio Corrêa Mariani. FZEA-USP Pirassununga: Mirele Daiana Poleti; Raul Machado Neto; Ricardo Augusto Brassaloti; Ricardo Haddad; Rodrigo Tocantins Calado. FAMERP-SJRP: Cecília Artico Banho; Sandra Coccuzzo Sampaio; Svetoslav Nanev Slavov; Vagner Fonseca; Vincent Louis Viala |
| EPI_ISL_2209413 | PRONTO ATENDIMENTO MUNICIPAL ITALO SANTUCCI | Instituto Butantan | Antonio Jorge Martins; Claudia Renata dos Santos Barros; David Schlesinger; Debora Botequio Moretti; Dimas Tadeu Covas; Elaine Cristina Marqueze; Elaine Vieira Santos; Evandra Strazza Rodrigues; Heidge Fukumasu; Jayme Augusto de Souza-Neto; José Salvatore Leister Patané; Luiz Alcantara; Luiz Lehmann Coutinho; Maria Carolina Elias; Maurício Lacerda Nogueira; Rafael dos Santos Bezerra; Raul Machado Neto; Rejane Maria Tommasini Grotto; Ricardo Haddad; Sandra Coccuzzo Sampaio Vessoni; Simone Kashima; Svetoslav Nanev Slavov; Vincent Louis Viala. |
| EPI_ISL_2344592 | PRONTO ATENDIMENTO MUNICIPAL ITALO SANTUCCI | Instituto Butantan / UNESP-Botucatu | Antonio Jorge Martins; Claudia Renata dos Santos Barros; David Schlesinger; Debora Botequio Moretti; Dimas Tadeu Covas; Elaine Cristina Marqueze; Elaine Vieira Santos; Evandra Strazza Rodrigues; Heidge Fukumasu; Jayme Augusto de Souza-Neto; José Salvatore Leister Patané; Luiz Alcantara; Luiz Lehmann Coutinho; Maria Carolina Elias; Maurício Lacerda Nogueira; Rafael dos Santos Bezerra; Raul Machado Neto; Rejane Maria Tommasini Grotto; Ricardo Haddad; Sandra Coccuzzo Sampaio Vessoni; Simone Kashima; Svetoslav Nanev Slavov; Vincent Louis Viala |
| EPI_ISL_2209422 | PRONTO SOCORRO MUNICIPAL DE TAUBATE | Instituto Butantan | Antonio Jorge Martins; Claudia Renata dos Santos Barros; David Schlesinger; Debora Botequio Moretti; Dimas Tadeu Covas; Elaine Cristina Marqueze; Elaine Vieira Santos; Evandra Strazza Rodrigues; Heidge Fukumasu; Jayme Augusto de Souza-Neto; José Salvatore Leister Patané; Luiz Alcantara; Luiz Lehmann Coutinho; Maria Carolina Elias; Maurício Lacerda Nogueira; Rafael dos Santos Bezerra; Raul Machado Neto; Rejane Maria Tommasini Grotto; Ricardo Haddad; Sandra Coccuzzo Sampaio Vessoni; Simone Kashima; Svetoslav Nanev Slavov; Vincent Louis Viala. |
| EPI_ISL_2344686 | PRONTO SOCORRO MUNICIPAL DE TAUBATE | Instituto Butantan / UNESP-Botucatu | Antonio Jorge Martins; Claudia Renata dos Santos Barros; David Schlesinger; Debora Botequio Moretti; Dimas Tadeu Covas; Elaine Cristina Marqueze; Elaine Vieira Santos; Evandra Strazza Rodrigues; Heidge Fukumasu; Jayme Augusto de Souza-Neto; José Salvatore Leister Patané; Luiz Alcantara; Luiz Lehmann Coutinho; Maria Carolina Elias; Maurício Lacerda Nogueira; Rafael dos Santos Bezerra; Raul Machado Neto; Rejane Maria Tommasini Grotto; Ricardo Haddad; Sandra Coccuzzo Sampaio Vessoni; Simone Kashima; Svetoslav Nanev Slavov; Vincent Louis Viala |
| EPI_ISL_2105557 | PROVIDERS MULTI-PURPOSE COOPERATIVE MEDICAL CENTER | Philippine Genome Center | Alethea R. de Guzman; Anna Ong-Lim; Arianne A. Zamora; Asia Louisa U. Chong; Benedict A. Maralit; Candice Francheska B. Tambaoan; Carlo M. Lapid; Celia Carlos; Devon Ray Pacial; Edsel Maurice Salvaña; El King D. Morado; Eva Maria Cutiongco-de la Paz; Francis A. Tablizo; Irish Coleen A. Asin; Jaime C. Montoya; Jan Michael C. Yap; Jo-Hannah S. Llames; John Q. Wong; Joshua Gregor A. Dizon; Juan Antonio R. Magalang; Karol Sophia Agape R. Padilla; Kenneth M. Kim; Kris P. Punayan; Marc Edsel C. Ayes; Marc Jerrone R. Castro; Maria Rosario Singh-Vergeire and Cynthia P. Saloma; Maria Sofia L. Yangzon; Marissa Alejandria; Razel Nikka M. Hao; Rianna Patricia S. Cruz; Sheila Mae M. Araiza |
| EPI_ISL_534322 | PS Mun Julio Tupy | Instituto Adolfo Lutz, Interdiciplinary Procedures Center, Strategic Laboratory | Claudia Regina Gonçalves; Claudio Tavares Sacchi; Erica Valessa Ramos Gomes |
| EPI_ISL_515524, EPI_ISL_515566, EPI_ISL_524466, EPI_ISL_527866 | PS Municipal Dr Lauro Ribas Braga | Instituto Adolfo Lutz, Interdiciplinary Procedures Center, Strategic Laboratory | 01246-1301; 355 - Brazil; Av. Dr. Arnaldo; Cerqueira Cesar; Claudia Regina Gonçalves; Claudio Tavares Sacchi; Erica Valessa Ramos Gomes; São Paulo - SP |
| EPI_ISL_524465 | PS Municipal Dr. Caetano Virgílio Neto | Instituto Adolfo Lutz, Interdiciplinary Procedures Center, Strategic Laboratory | Claudia Regina Gonçalves; Claudio Tavares Sacchi; Erica Valessa Ramos Gomes |
| EPI_ISL_534321, EPI_ISL_861659, EPI_ISL_861660, EPI_ISL_861661, EPI_ISL_906067 | PS e Maternidade Nair Fonseca Leitao Arantes | Instituto Adolfo Lutz, Interdiciplinary Procedures Center, Strategic Laboratory | Claudia Regina Gonçalves; Claudio Tavares Sacchi; Erica Valessa Ramos Gomes; Karoline Rodrigues Campos |
| EPI_ISL_861666 | PSF Dr. Antonio Pires de Almeida | Instituto Adolfo Lutz, Interdiciplinary Procedures Center, Strategic Laboratory | Claudia Regina Gonçalves; Claudio Tavares Sacchi; Erica Valessa Ramos Gomes; Karoline Rodrigues Campos |
| EPI_ISL_984932, EPI_ISL_984938, EPI_ISL_1041373, EPI_ISL_1097913, EPI_ISL_1098162, EPI_ISL_1258936 | Pandemic Response Lab - NYC | Pandemic Response Lab, R&D | Cybill del Castillo; Haiping Hao; Henry Lee; Jon Laurent; Melissa Hopkins; Michael Hammerling; Pradeep Bugga; Shinyoung Clair Kang; William Ward |
| EPI_ISL_2156364 | Pasig City Children's Hospital - Child's Hope | Philippine Genome Center | Alethea R. de Guzman; Anna Ong-Lim; Arianne A. Zamora; Asia Louisa U. Chong; Benedict A. Maralit; Candice Francheska B. Tambaoan; Carlo M. Lapid; Celia Carlos; Devon Ray Pacial; Edsel Maurice Salvaña; El King D. Morado; Elcid Aaron R. Pangilinan; Eva Maria Cutiongco-de la Paz; Francis A. Tablizo; Irish Coleen A. Asin; Jaime C. Montoya; Jan Michael C. Yap; Jo-Hannah S. Llames; John Q. Wong; Joshua Gregor A. Dizon; Juan Antonio R. Magalang; Karol Sophia Agape R. Padilla; Kenneth M. Kim; Kris P. Punayan; Marc Edsel C. Ayes; Marc Jerrone R. Castro; Maria Rosario Singh-Vergeire and Cynthia P. Saloma; Maria Sofia L. Yangzon; Marissa Alejandria; Razel Nikka M. Hao; Rianna Patricia S. Cruz; Sheila Mae M. Araiza |
| EPI_ISL_591352, EPI_ISL_591372, EPI_ISL_591402, EPI_ISL_591411, EPI_ISL_591449, EPI_ISL_591450, EPI_ISL_591537, EPI_ISL_591538, EPI_ISL_667561, EPI_ISL_667578, EPI_ISL_667626, EPI_ISL_667633, EPI_ISL_667634, EPI_ISL_667639, EPI_ISL_667649, EPI_ISL_667650, EPI_ISL_667659, EPI_ISL_667663, EPI_ISL_685539, EPI_ISL_686307, EPI_ISL_690635, EPI_ISL_736895, EPI_ISL_768652, EPI_ISL_768709, EPI_ISL_779209, EPI_ISL_779216, EPI_ISL_779245, EPI_ISL_779246, EPI_ISL_901583, EPI_ISL_901605, EPI_ISL_1127124, EPI_ISL_1127125 | | | |
| see above | Pathogen Genomics | Pathogen Genomics | Kentaro Itokawa; Makoto Kuroda; Masanori Hashino; Rina Tanaka; Tsuyoshi Sekizuka |

| | | | |
|---|---|---|---|
| | Center, National Institute of Infectious Diseases | Center, National Institute of Infectious Diseases | |
| EPI_ISL_2105405, EPI_ISL_2105420, EPI_ISL_2105432, EPI_ISL_2105446, EPI_ISL_2105459 | Philippine Red Cross - Misamis Oriental | Philippine Genome Center | Alethea R. de Guzman; Anna Ong-Lim; Arianne A. Zamora; Asia Louisa U. Chong; Benedict A. Maralit; Candice Francheska B. Tambaoan; Carlo M. Lapid; Celia Carlos; Devon Ray Pacial; Edsel Maurice Salvaña; El King D. Morado; Eva Maria Cutiongco-de la Paz; Francis A. Tablizo; Irish Coleen A. Asin; Jaime C. Montoya; Jan Michael C. Yap; Jo-Hannah S. Llames; John Q. Wong; Joshua Gregor A. Dizon; Juan Antonio R. Magalang; Karol Sophia Agape R. Padilla; Kenneth M. Kim; Kris P. Punayan; Marc Edsel C. Ayes; Marc Jerrone R. Castro; Maria Rosario Singh-Vergeire and Cynthia P. Saloma; Maria Sofia L. Yangzon; Marissa Alejandria; Razel Nikka M. Hao; Rianna Patricia S. Cruz; Sheila Mae M. Araiza |
| EPI_ISL_2171200, EPI_ISL_2189213 | Philippine Red Cross - Port Area | Philippine Genome Center | Alethea R. de Guzman; Anna Ong-Lim; Arianne A. Zamora; Asia Louisa U. Chong; Benedict A. Maralit; Candice Francheska B. Tambaoan; Carlo M. Lapid; Celia Carlos; Devon Ray Pacial; Edsel Maurice Salvaña; El King D. Morado; Elcid Aaron R. Pangilinan; Eva Maria Cutiongco-de la Paz; Francis A. Tablizo; Irish Coleen A. Asin; Jaime C. Montoya; Jan Michael C. Yap; Jo-Hannah S. Llames; John Q. Wong; Joshua Gregor A. Dizon; Juan Antonio R. Magalang; Karol Sophia Agape R. Padilla; Kenneth M. Kim; Kris P. Punayan; Marc Edsel C. Ayes; Marc Jerrone R. Castro; Maria Rosario Singh-Vergeire and Cynthia P. Saloma; Maria Sofia L. Yangzon; Marissa Alejandria; Razel Nikka M. Hao; Renato Jacinto Q. Mantaring; Rianna Patricia S. Cruz; Sheila Mae M. Araiza |
| EPI_ISL_2105381 | Philippine Red Cross Logistics and Multipurpose Center | Philippine Genome Center | Alethea R. de Guzman; Anna Ong-Lim; Arianne A. Zamora; Asia Louisa U. Chong; Benedict A. Maralit; Candice Francheska B. Tambaoan; Carlo M. Lapid; Celia Carlos; Devon Ray Pacial; Edsel Maurice Salvaña; El King D. Morado; Eva Maria Cutiongco-de la Paz; Francis A. Tablizo; Irish Coleen A. Asin; Jaime C. Montoya; Jan Michael C. Yap; Jo-Hannah S. Llames; John Q. Wong; Joshua Gregor A. Dizon; Juan Antonio R. Magalang; Karol Sophia Agape R. Padilla; Kenneth M. Kim; Kris P. Punayan; Marc Edsel C. Ayes; Marc Jerrone R. Castro; Maria Rosario Singh-Vergeire and Cynthia P. Saloma; Maria Sofia L. Yangzon; Marissa Alejandria; Razel Nikka M. Hao; Rianna Patricia S. Cruz; Sheila Mae M. Araiza |
| EPI_ISL_2663256 | Plataforma de Vigilancia Molecular (PVM) - FIOCRUZ/BA | Plataforma de Vigilancia Molecular (PVM) - FIOCRUZ/BA | Bruno Bezerril Andrade; Camila I. de Oliveira on behalf of the Fiocruz COVID-19 Genomic Surveillance Network.; Clarissa Araújo Gurgel; Leonardo Paiva Farias; Marina Cucco; Ricardo Khouri; Tiago Graf |
| EPI_ISL_513514, EPI_ISL_513532, EPI_ISL_513546, EPI_ISL_513557, EPI_ISL_513578 | Programa de Oncovirologia, Instituto Nacional de Câncer | Programa de Oncovirologia, Instituto Nacional de Câncer | Andreia C. de Melo; Brunna M. Alves; Claudia Cicala; James Arthos; João P.B. Viola; Juliana D. Siqueira; Livia R. Goes; Marcelo A. Soares |
| EPI_ISL_1469657 | Pronto Atendimento Campo Bom | Epiclin | Ana Paula Muterle; Carolina Comerlato; Eliana Márcia Da Ros Wendland; Fernando Hayashi Sant'Anna; Janira Prichula; Juliana Comerlato |
| EPI_ISL_1469721 | Pronto Atendimento Cruzeiro do Sul | Epiclin | Ana Paula Muterle; Carolina Comerlato; Eliana Márcia Da Ros Wendland; Fernando Hayashi Sant'Anna; Janira Prichula; Juliana Comerlato |
| EPI_ISL_603021, EPI_ISL_693204 | Pronto Socorro Dr. Conrado Cesarino Nuvolini | Instituto Adolfo Lutz, Interdiciplinary Procedures Center, Strategic Laboratory | Claudia Regina Gonçalves; Claudio Tavares Sacchi; Erica Valessa Ramos Gomes; Karoline Rodrigues Campos |
| EPI_ISL_471556 | Pronto Socorro Jose Ibrahin | Instituto Adolfo Lutz, Interdiciplinary Procedures Center, Strategic Laboratory | Claudia Regina Gonçalves; Claudio Tavares Sacchi; Erica Valessa Ramos Gomes |
| EPI_ISL_515529 | Pronto Socorro Municipal Julio Tupy | Instituto Adolfo Lutz, Interdiciplinary Procedures Center, Strategic Laboratory | Claudia Regina Gonçalves; Claudio Tavares Sacchi; Erica Valessa Ramos Gomes |
| EPI_ISL_693202 | Pronto Socorro Municipal Prof. Joao Catarin Mezomo | Instituto Adolfo Lutz, Interdiciplinary Procedures Center, Strategic Laboratory | Claudia Regina Gonçalves; Claudio Tavares Sacchi; Erica Valessa Ramos Gomes; Karoline Rodrigues Campos |
| EPI_ISL_515554, EPI_ISL_523958 | Pronto Socorro Municipal de Perus | Instituto Adolfo Lutz, Interdiciplinary Procedures Center, Strategic Laboratory | Claudia Regina Gonçalves; Claudio Tavares Sacchi; Erica Valessa Ramos Gomes |
| EPI_ISL_693231 | Pronto Socorro Municipal de Santa Branca | Instituto Adolfo Lutz, Interdiciplinary Procedures Center, Strategic Laboratory | Claudia Regina Gonçalves; Claudio Tavares Sacchi; Erica Valessa Ramos Gomes; Karoline Rodrigues Campos |
| EPI_ISL_693210 | Pronto-Socorro Dr. Osmar Mesquita | Instituto Adolfo Lutz, Interdiciplinary Procedures Center, Strategic Laboratory | Claudia Regina Gonçalves; Claudio Tavares Sacchi; Erica Valessa Ramos Gomes; Karoline Rodrigues Campos |
| EPI_ISL_693257, EPI_ISL_693278 | Queensland Health Forensic and Scientific Services | Queensland Health Forensic and Scientific Services | Son Nguyen et al |
| EPI_ISL_854921 | Quest Diagnostics | Quest Diagnostics | Anderson, B.; Bernstein; D.F.; Gerasimova, A.; Hua, M.; I.A.; K.E.; Kagan; L.E.; Lacbawan, F.; Liu Y.; Livingston; Owen, R.; Perez, A.; R.M.; Rosenthal; S.H.; Shalhout; Shlyakhter; Tanpaiboon, P. |
| EPI_ISL_1266775 | Quest Diagnostics Incorporated | Centers for Disease Control and Prevention Division of Viral Diseases, Pathogen Discovery | A. Gerasimova; A. Perez; B. Anderson; Ben L. Rambo-Martin; Clinton R. Paden; Dakota Howard; Dhwani Batra; Duncan MacCannell; F. Lacbawan; I. A. Shlyakhter; K.E. Livingston; L.E. Bernstein; M. Hua; P. Tanpaiboon; Peter W. Cook; R. M. Kagan; R. Owen; R. V. Rolando; S. H. Rosenthal; Suxiang Tong; Y. Liu |
| EPI_ISL_1086554, EPI_ISL_1086960, EPI_ISL_1086980, EPI_ISL_1086982, EPI_ISL_1087267, EPI_ISL_1088169 | Quest Diagnostics Incorporated | Respiratory Viruses Branch, Division of Viral Diseases, Centers for Disease Control and Prevention | A. Gerasimova; A. Perez; B. Anderson; Ben L. Rambo-Martin; Clinton R. Paden; Dakota Howard; Dhwani Batra; Duncan MacCannell; F. Lacbawan; I. A. Shlyakhter; K.E. Livingston; L.E. Bernstein; M. Hua; P. Tanpaiboon; Peter W. Cook; R. M. Kagan; R. Owen; R. V. Rolando; S. H. Rosenthal; Suxiang Tong; Y. Liu |
| EPI_ISL_2105310 | Quezon City General Hospital (PGC) | Philippine Genome Center | Alethea R. de Guzman; Anna Ong-Lim; Arianne A. Zamora; Asia Louisa U. Chong; Benedict A. Maralit; Candice Francheska B. Tambaoan; Carlo M. Lapid; Celia Carlos; Devon Ray Pacial; Edsel Maurice Salvaña; El King D. Morado; Eva Maria Cutiongco-de la Paz; Francis A. Tablizo; Irish Coleen A. Asin; Jaime C. Montoya; Jan Michael C. Yap; Jo-Hannah S. Llames; John Q. Wong; Joshua Gregor A. Dizon; Juan Antonio R. Magalang; Karol Sophia Agape R. Padilla; Kenneth M. Kim; Kris P. Punayan; Marc Edsel C. Ayes; Marc Jerrone R. Castro; Maria Rosario Singh-Vergeire and Cynthia P. Saloma; Maria Sofia L. Yangzon; Marissa Alejandria; Razel Nikka M. Hao; Rianna Patricia S. Cruz; Sheila Mae M. Araiza |
| EPI_ISL_2101297 | RSUD Mangusada | Eijkman Institute for Molecular Biology, National Agency for Research and Innovation; Molecular Biology Laboratory, Faculty Medicine and Health Sciences, Warmadewa University | Amin Soebandrio; Edison Johar; Erly Sintya; Frilasita A Yudhaputri; Hidayat Trimarsanto; Ida Ayu Wayan Mahayani; Iskandar Adnan; Khin Saw Myint; Lidwina Priliani; Lydia V. Panggalo; Muhammad Rezki Rasyak; Safarina G Malik; Sri Masyeni; Sukma Oktavianthi; Willy Agustine |
| EPI_ISL_920984 | Regional Virus Laboratory, Belfast Health and Social Care Trust | COVID-19 Genomics UK (COG-UK) Consortium | Alison Watt; Ciara Cox; Conall McCaughey; David Simpson; Derek Fairley; James McKenna; Mairead Connor; Susan Feeney; Tanya Curran; Zoltan Molnar |
| EPI_ISL_2105289, EPI_ISL_2105337, EPI_ISL_2105418, EPI_ISL_2105461, EPI_ISL_2156351 | Research Institute for Tropical Medicine, Inc. (RITM) | Philippine Genome Center | Alethea R. de Guzman; Anna Ong-Lim; Arianne A. Zamora; Asia Louisa U. Chong; Benedict A. Maralit; Candice Francheska B. Tambaoan; Carlo M. Lapid; Celia Carlos; Devon Ray Pacial; Edsel Maurice Salvaña; El King D. Morado; Elcid Aaron R. Pangilinan; Eva Maria Cutiongco-de la Paz; Francis A. Tablizo; Irish Coleen A. Asin; Jaime C. Montoya; Jan Michael C. Yap; Jo-Hannah S. Llames; John Q. Wong; Joshua Gregor A. Dizon; Juan Antonio R. Magalang; Karol Sophia Agape R. Padilla; Kenneth M. Kim; Kris P. Punayan; Marc Edsel C. Ayes; Marc Jerrone R. Castro; Maria Rosario Singh-Vergeire and Cynthia P. Saloma; Maria Sofia L. Yangzon; Marissa Alejandria; Razel Nikka M. Hao; Rianna Patricia S. Cruz; Sheila Mae M. Araiza |
| EPI_ISL_1046774, EPI_ISL_1046782, EPI_ISL_1425749, EPI_ISL_1427816, EPI_ISL_1927843, EPI_ISL_2131797 | SARS-CoV-2 testing team, National Institute of Infectious Diseases | Pathogen Genomics Center, National Institute of Infectious Diseases | ; Chikako Shimokawa; Daisuke Kobayashi; Eunsil Park; Hazuka Y Furihata; Ken Miyazawa; Kentaro Itokawa; Kento Fukano; Makoto Kuroda; Masanori Hashino; Masumichi Saito; Minoru Nagi; Naomi Nojiri; Nozomu Hanaoka; Rina Tanaka; Sana Uchikoba; Takanobu Kato; Takashi Sakudoh; Tsuguto Fujimoto; Tsuyoshi Sekizuka; Yasutaka Hoshino; Yasuyuki Morishima; Yoshihiro Kaku |
| EPI_ISL_1445177, EPI_ISL_1966112 | SAUDE COLETIVA CAPAO BONITO | Instituto Butantan / Mendelics | Antonio Jorge Martins; Bianca Cechetto Carlos. Mendelics: Bibiana Santos; Bibiana Santos; Claudia Renata dos Santos Barros; Cíntia Bittar; David Schlesinger; David Schlesinger. Hemocentro Ribeirão Preto: Simone Kashima; Debora Botequio Moretti; Dimas Tadeu Covas; Elaine Cristina Marqueze; Elaine Vieira dos Santos; Elisangela Chicaroni Mattos; Erika Freitas; Evandra Strazza Rodrigues; Felipe Allan da Silva da Costa; Flavia Aburjaile; Fábio Sossai Possebon; Guilherme Campos; Guilherme Targino Valente; Heidge Fukumasu. USP-Botucatu: Rejane Maria Tommasini Grotto; Helena Lage Ferreira; Instituto Butantan: Dimas Tadeu Covas; Jardelina de Souza Todao Bernardino; Jayme A. Souza-Neto; Jessika Cristina Chagas Lesbon; Jorge A. Petroli Marchesi; José Salvatore Leister Patané; João Paulo Kitajima; João Pessoa Araújo Jr.; Leila Sabrina Ullmann; Loyze Paola Oliveira de Lima; Luiz Aurelio de Campos Crispin. Centro de Genômica Funcional da ESALQ: Luiz Lehmann Coutinho; Luiz Carlos Junior de Alcantara; Lívia Sacchetto; Maisa C. Pereira Parra; Maria Carolina Elias; Marta Giovanetti; Marília Moraes; Maurício Lacerda Nogueira. Prefeitura de Sao Paulo: Melissa Palmieri.; Patricia Akemi Assato; Paula Rahal; Paulo Inacio da Costa; Rafael dos Santos Bezerra; Raquel de Lello Rocha Campos Cassano. NGS Soluções Genômicas: Pilar |

| | | | |
|---|---|---|---|
| | | | Drummond Sampaio Corrêa Mariani. FZEA-USP Pirassununga: Mirele Daiana Poleti; Raul Machado Neto; Ricardo Augusto Brassaloti; Ricardo Haddad; Rodrigo Tocantins Calado.; Rodrigo Tocantins Calado. FAMERP-SJRP: Cecília Artico Banho; Sandra Coccuzzo Sampaio; Simone Kashima; Svetoslav Nanev Slavov; Vagner Fonseca; Vincent Louis Viala |
| EPI_ISL_1445168, EPI_ISL_1445242, EPI_ISL_1445244, EPI_ISL_1966073, EPI_ISL_1966194, EPI_ISL_1966488 | SECAO CENTRO DE DIAGNOSTICO SECEDI | Instituto Butantan / Mendelics | Antonio Jorge Martins; Bianca Cechetto Carlos. Mendelics: Bibiana Santos; Bibiana Santos; Claudia Renata dos Santos Barros; Cíntia Bittar; David Schlesinger; David Schlesinger. Hemocentro Ribeirão Preto: Simone Kashima; Debora Botequio Moretti; Dimas Tadeu Covas; Elaine Cristina Marqueze; Elaine Vieira dos Santos; Elisangela Chicaroni Mattos; Erika Freitas; Evandra Strazza Rodrigues; Felipe Allan da Silva da Costa; Flavia Aburjaile; Fábio Sossai Possebon; Guilherme Campos; Guilherme Targino Valente; Heidge Fukumasu. USP-Botucatu: Rejane Maria Tommasini Grotto; Helena Lage Ferreira; Instituto Butantan: Dimas Tadeu Covas; Jardelina de Souza Todao Bernardino; Jayme A. Souza-Neto; Jessika Cristina Chagas Lesbon; Jorge A. Petroli Marchesi; José Salvatore Leister Patané; João Paulo Kitajima; João Pessoa Araújo Jr.; Leila Sabrina Ullmann; Loyze Paola Oliveira de Lima; Luiz Aurelio de Campos Crispin. Centro de Genômica Funcional da ESALQ: Luiz Lehmann Coutinho; Luiz Carlos Junior de Alcantara; Lívia Sacchetto; Maisa C. Pereira Parra; Maria Carolina Elias; Marta Giovanetti; Marília Moraes; Maurício Lacerda Nogueira. Prefeitura de Sao Paulo: Melissa Palmieri.; Patricia Akemi Assato; Paula Rahal; Paulo Inacio da Costa; Rafael dos Santos Bezerra; Raquel de Lello Rocha Campos Cassano. NGS Soluções Genômicas: Pilar Drummond Sampaio Corrêa Mariani. FZEA-USP Pirassununga: Mirele Daiana Poleti; Raul Machado Neto; Ricardo Augusto Brassaloti; Ricardo Haddad; Rodrigo Tocantins Calado.; Rodrigo Tocantins Calado. FAMERP-SJRP: Cecília Artico Banho; Sandra Coccuzzo Sampaio; Simone Kashima; Svetoslav Nanev Slavov; Vagner Fonseca; Vincent Louis Viala |
| EPI_ISL_2344674 | SECRETARIA MUNICIPAL DE SAUDE DE CORDEIROPOLIS | Instituto Butantan / FZEA-USP-Pirassununga | Antonio Jorge Martins; Claudia Renata dos Santos Barros; David Schlesinger; Debora Botequio Moretti; Dimas Tadeu Covas; Elaine Cristina Marqueze; Elaine Vieira Santos; Evandra Strazza Rodrigues; Heidge Fukumasu; Jayme Augusto de Souza-Neto; José Salvatore Leister Patané; Luiz Alcantara; Luiz Lehmann Coutinho; Maria Carolina Elias; Maurício Lacerda Nogueira; Rafael dos Santos Bezerra; Raul Machado Neto; Rejane Maria Tommasini Grotto; Ricardo Haddad; Sandra Coccuzzo Sampaio Vessoni; Simone Kashima; Svetoslav Nanev Slavov; Vincent Louis Viala |
| EPI_ISL_1469683, EPI_ISL_1469687 | SECRETARIA MUNICIPAL DE SAUDE DE TAQUARA | Epiclin | Ana Paula Muterle; Carolina Comerlato; Eliana Márcia Da Ros Wendland; Fernando Hayashi Sant'Anna; Janira Prichula; Juliana Comerlato |
| EPI_ISL_1469740 | SECRETARIA MUNICIPAL DE SAUDE DE TRES COROAS | Epiclin | Ana Paula Muterle; Carolina Comerlato; Eliana Márcia Da Ros Wendland; Fernando Hayashi Sant'Anna; Janira Prichula; Juliana Comerlato |
| EPI_ISL_1445134, EPI_ISL_1966178 | SECRETARIA MUNICIPAL DE SAUDE SOROCABA | Instituto Butantan / Mendelics | Antonio Jorge Martins; Bianca Cechetto Carlos. Mendelics: Bibiana Santos; Bibiana Santos; Claudia Renata dos Santos Barros; Cíntia Bittar; David Schlesinger; David Schlesinger. Hemocentro Ribeirão Preto: Simone Kashima; Debora Botequio Moretti; Dimas Tadeu Covas; Elaine Cristina Marqueze; Elaine Vieira dos Santos; Elisangela Chicaroni Mattos; Erika Freitas; Evandra Strazza Rodrigues; Felipe Allan da Silva da Costa; Flavia Aburjaile; Fábio Sossai Possebon; Guilherme Campos; Guilherme Targino Valente; Heidge Fukumasu. USP-Botucatu: Rejane Maria Tommasini Grotto; Helena Lage Ferreira; Instituto Butantan: Dimas Tadeu Covas; Jardelina de Souza Todao Bernardino; Jayme A. Souza-Neto; Jessika Cristina Chagas Lesbon; Jorge A. Petroli Marchesi; José Salvatore Leister Patané; João Paulo Kitajima; João Pessoa Araújo Jr.; Leila Sabrina Ullmann; Loyze Paola Oliveira de Lima; Luiz Aurelio de Campos Crispin. Centro de Genômica Funcional da ESALQ: Luiz Lehmann Coutinho; Luiz Carlos Junior de Alcantara; Lívia Sacchetto; Maisa C. Pereira Parra; Maria Carolina Elias; Marta Giovanetti; Marília Moraes; Maurício Lacerda Nogueira. Prefeitura de Sao Paulo: Melissa Palmieri.; Patricia Akemi Assato; Paula Rahal; Paulo Inacio da Costa; Rafael dos Santos Bezerra; Raquel de Lello Rocha Campos Cassano. NGS Soluções Genômicas: Pilar Drummond Sampaio Corrêa Mariani. FZEA-USP Pirassununga: Mirele Daiana Poleti; Raul Machado Neto; Ricardo Augusto Brassaloti; Ricardo Haddad; Rodrigo Tocantins Calado.; Rodrigo Tocantins Calado. FAMERP-SJRP: Cecília Artico Banho; Sandra Coccuzzo Sampaio; Simone Kashima; Svetoslav Nanev Slavov; Vagner Fonseca; Vincent Louis Viala |
| EPI_ISL_2346082 | SERRANA | Instituto Butantan / Mendelics | Antonio Jorge Martins; Claudia Renata dos Santos Barros; David Schlesinger; Debora Botequio Moretti; Dimas Tadeu Covas; Elaine Cristina Marqueze; Elaine Vieira Santos; Evandra Strazza Rodrigues; Heidge Fukumasu; Jayme Augusto de Souza-Neto; José Salvatore Leister Patané; Luiz Alcantara; Luiz Lehmann Coutinho; Maria Carolina Elias; Maurício Lacerda Nogueira; Rafael dos Santos Bezerra; Raul Machado Neto; Rejane Maria Tommasini Grotto; Ricardo Haddad; Sandra Coccuzzo Sampaio Vessoni; Simone Kashima; Svetoslav Nanev Slavov; Vincent Louis Viala |
| EPI_ISL_1795398, EPI_ISL_2345454 | SMS SECRETARIA MUNICIPAL DE SAUDE DE BOITUVA | Instituto Butantan / ESALQ-Piracicaba | Antonio Jorge Martins; Bianca Cechetto Carlos. Mendelics: Bibiana Santos; Claudia Renata dos Santos Barros; David Schlesinger; David Schlesinger. Hemocentro Ribeirão Preto: Simone Kashima; Debora Botequio Moretti; Debora Botequio Moretti. Centro de Genômica Funcional da ESALQ: Luiz Lehmann Coutinho; Dimas Tadeu Covas; Elaine Cristina Marqueze; Elaine Vieira Santos; Elaine Vieira dos Santos; Elisangela Chicaroni Mattos; Erika Freitas; Evandra Strazza Rodrigues; Felipe Allan da Silva da Costa; Flavia Aburjaile; Guilherme Targino Valente; Heidge Fukumasu; Heidge Fukumasu. USP-Botucatu: Rejane Maria Tommasini Grotto; Instituto Butantan: Alexander Roberto Precioso; Jayme A. Souza-Neto; Jayme Augusto de Souza-Neto; Jessika Cristina Chagas Lesbon; José Salvatore Leister Patané; João Paulo Kitajima; Luiz Alcantara; Luiz Carlos Junior de Alcantara; Luiz Lehmann Coutinho; Maria Carolina Elias; Marta Giovanetti; Maurício Lacerda Nogueira; Patricia Akemi Assato; Rafael dos Santos Bezerra; Raquel de Lello Rocha Campos Cassano. NGS Soluções Genômicas: Pilar Drummond Sampaio Corrêa Mariani. FZEA-USP Pirassununga: Mirele Daiana Poleti; Raul Machado Neto; Rejane Maria Tommasini Grotto; Ricardo Augusto Brassaloti; Ricardo Haddad; Rodrigo Tocantins Calado.; Sandra Coccuzzo Sampaio; Sandra Coccuzzo Sampaio Vessoni; Simone Kashima; Svetoslav Nanev Slavov; Vagner Fonseca; Vincent Louis Viala |
| EPI_ISL_1966261 | SMS SECRETARIA MUNICIPAL DE SAUDE DE BOITUVA | Instituto Butantan / Mendelics | Antonio Jorge Martins; Bianca Cechetto Carlos. Mendelics: Bibiana Santos; Claudia Renata dos Santos Barros; Cíntia Bittar; David Schlesinger. Hemocentro Ribeirão Preto: Simone Kashima; Debora Botequio Moretti; Elaine Cristina Marqueze; Elaine Vieira dos Santos; Elisangela Chicaroni Mattos; Erika Freitas; Evandra Strazza Rodrigues; Felipe Allan da Silva da Costa; Flavia Aburjaile; Fábio Sossai Possebon; Guilherme Campos; Guilherme Targino Valente; Heidge Fukumasu. USP-Botucatu: Rejane Maria Tommasini Grotto; Helena Lage Ferreira; Instituto Butantan: Dimas Tadeu Covas; Jardelina de Souza Todao Bernardino; Jayme A. Souza-Neto; Jessika Cristina Chagas Lesbon; Jorge A. Petroli Marchesi; José Salvatore Leister Patané; João Paulo Kitajima; João Pessoa Araújo Jr.; Leila Sabrina Ullmann; Loyze Paola Oliveira de Lima; Luiz Aurelio de Campos Crispin. Centro de Genômica Funcional da ESALQ: Luiz Lehmann Coutinho; Luiz Carlos Junior de Alcantara; Lívia Sacchetto; Maisa C. Pereira Parra; Maria Carolina Elias; Marta Giovanetti; Marília Moraes; Maurício Lacerda Nogueira. Prefeitura de Sao Paulo: Melissa Palmieri.; Patricia Akemi Assato; Paula Rahal; Paulo Inacio da Costa; Rafael dos Santos Bezerra; Raquel de Lello Rocha Campos Cassano. NGS Soluções Genômicas: Pilar Drummond Sampaio Corrêa Mariani. FZEA-USP Pirassununga: Mirele Daiana Poleti; Raul Machado Neto; Ricardo Augusto Brassaloti; Ricardo Haddad; Rodrigo Tocantins Calado. FAMERP-SJRP: Cecília Artico Banho; Sandra Coccuzzo Sampaio; Svetoslav Nanev Slavov; Vagner Fonseca; Vincent Louis Viala |
| EPI_ISL_547575 | SVO Jundiaí | Instituto Adolfo Lutz, Interdiciplinary Procedures Center, Strategic Laboratory | Claudia Regina Gonçalves; Claudio Tavares Sacchi; Erica Valessa Ramos Gomes; Karoline Rodrigues Campos |
| EPI_ISL_1854406 | SYNLAB | Instituto Nacional de Saude (INSA) | Borges et al |
| EPI_ISL_2105340, EPI_ISL_2105394 | Safeguard DNA Diagnostic, Inc. | Philippine Genome Center | Alethea R. de Guzman; Anna Ong-Lim; Arianne A. Zamora; Asia Louisa U. Chong; Benedict A. Maralit; Candice Francheska B. Tambaoan; Carlo M. Lapid; Celia Carlos; Devon Ray Pacial; Edsel Maurice Salvaña; El King D. Morado; Eva Maria Cutiongco-de la Paz; Francis A. Tablizo; Irish Coleen A. Asin; Jaime C. Montoya; Jan Michael C. Yap; Jo-Hannah S. Llames; John Q. Wong; Joshua Gregor A. Dizon; Juan Antonio R. Magalang; Karol Sophia Agape R. Padilla; Kenneth M. Kim; Kris P. Punayan; Marc Edsel C. Ayes; Marc Jerrone R. Castro; Maria Rosario Singh-Vergeire and Cynthia P. Saloma; Maria Sofia L. Yangzon; Marissa Alejandria; Razel Nikka M. Hao; Rianna Patricia S. Cruz; Sheila Mae M. Araiza |
| EPI_ISL_1426592, EPI_ISL_1426593, EPI_ISL_1427064 | Sakai City Institute of Public Health | Pathogen Genomics Center, National Institute of Infectious Diseases | Kentaro Itokawa; Makoto Kuroda; Masanori Hashino; Rina Tanaka; Tsuyoshi Sekizuka |
| EPI_ISL_2350041, EPI_ISL_2350066, EPI_ISL_2603719, EPI_ISL_2603730 | Salud Digna | Instituto Nacional de Medicina Genomica | Abraham Campos-Romero; Cedro-Tanda A; Escobar-Arrazola; Gonzalez-Barrera D; Herrera-Montalvo LA.; Hidalgo-Miranda A; Luna-Ruiz Marco; M; Mendoza-Vargas A; Moreno-Camacho José Luis; Munguia-Garza P; Ramirez-Vega O; Rangel-DeLeon D; Reyes-Grajeda JP; Rodriguez-Gallegos Jorge |
| EPI_ISL_2105287 | San Antonio City of Ilagan Hospital | Philippine Genome Center | Alethea R. de Guzman; Anna Ong-Lim; Arianne A. Zamora; Asia Louisa U. Chong; Benedict A. Maralit; Candice Francheska B. Tambaoan; Carlo M. Lapid; Celia Carlos; Devon Ray Pacial; Edsel Maurice Salvaña; El King D. Morado; Eva Maria Cutiongco-de la Paz; Francis A. Tablizo; Irish Coleen A. Asin; Jaime C. Montoya; Jan Michael C. Yap; Jo-Hannah S. Llames; John Q. Wong; Joshua Gregor A. Dizon; Juan Antonio R. Magalang; Karol Sophia Agape R. Padilla; Kenneth M. Kim; Kris P. Punayan; Marc Edsel C. Ayes; Marc Jerrone R. Castro; Maria Rosario Singh-Vergeire and Cynthia P. Saloma; Maria Sofia L. Yangzon; Marissa Alejandria; Razel Nikka M. Hao; Rianna Patricia S. Cruz; Sheila Mae M. Araiza |
| EPI_ISL_750177 | Sanatorio Americano | Institut Pasteur de Montevideo | Ana Carolina Mendonça; Andrés Lizasoain; Camila Simoes; Cecilia Alonso; Cecilia Salazar; Daiana Mir; Fernando López-Tort; Fernando Motta; Gonzalo Bello; Ighor Arantes; Ignacio Ferrés; Jose Sotelo; Leticia Maya; Letícia Garay Martins; Luciana Appolinario; Lucía Spangenberg; Mailen Arleo; Mariana Brandes; Marilda Mendonça Siqueira; Marilda Tereza Mar da Rosa; María José Benitez-Galeano; Martín Graña; Matías Castells; Matías Victoria; Matías Salvo; Natalia Rego; Natalia Reyes; Pablo Smircich; Paola Cristina Resende; Rodney Colina; Tamara Fernandez-Calero; Tania Possi; Tatiana Schäffer Gregianini; Verónica Noya; Yasser Vega |
| EPI_ISL_583492 | Santa Casa Anna Cintra | Instituto Adolfo Lutz, Interdiciplinary Procedures Center, Strategic Laboratory | Claudia Regina Gonçalves; Claudio Tavares Sacchi; Erica Valessa Ramos Gomes; Karoline Rodrigues Campos |
| EPI_ISL_1468454 | Santa Casa de Andradina | Instituto Adolfo Lutz, Interdiciplinary Procedures Center, Strategic Laboratory | Caio Vinicius Dias Lopes; Claudia Regina Gonçalves; Claudio Tavares Sacchi; Erica Valessa Ramos Gomes; Karoline Rodrigues Campos |
| EPI_ISL_1493592 | Santa Casa de Guaira | Instituto Adolfo Lutz, Interdiciplinary Procedures Center, Strategic Laboratory | Caio Vinicius Dias Lopes; Claudia Regina Gonçalves; Claudio Tavares Sacchi; Erica Valessa Ramos Gomes; Karoline Rodrigues Campos |
| EPI_ISL_603024, EPI_ISL_603038 | Santa Casa de Misericordia de Araçatuba | Instituto Adolfo Lutz, Interdiciplinary Procedures Center, Strategic Laboratory | Claudia Regina Gonçalves; Claudio Tavares Sacchi; Erica Valessa Ramos Gomes; Karoline Rodrigues Campos |
| EPI_ISL_693212 | Santa Casa de Misericordia de Braganca Paulista | Instituto Adolfo Lutz, Interdiciplinary Procedures Center, Strategic Laboratory | Claudia Regina Gonçalves; Claudio Tavares Sacchi; Erica Valessa Ramos Gomes; Karoline Rodrigues Campos |
| EPI_ISL_1468453 | Santa Casa de Misericordia de Pereira Barreto | Instituto Adolfo Lutz, Interdiciplinary Procedures Center, Strategic Laboratory | Caio Vinicius Dias Lopes; Claudia Regina Gonçalves; Claudio Tavares Sacchi; Erica Valessa Ramos Gomes; Karoline Rodrigues Campos |
| EPI_ISL_524469 | Santa Casa de Misericordia de Sao Paulo | Instituto Adolfo Lutz, Interdiciplinary Procedures Center, Strategic Laboratory | Claudia Regina Gonçalves; Claudio Tavares Sacchi; Erica Valessa Ramos Gomes |
| EPI_ISL_693198 | Santa Casa de Misericordia de Sao Paulo - Hospital Central | Instituto Adolfo Lutz, Interdiciplinary Procedures Center, Strategic Laboratory | Claudia Regina Gonçalves; Claudio Tavares Sacchi; Erica Valessa Ramos Gomes; Karoline Rodrigues Campos |
| EPI_ISL_693211 | Santa Casa de Misericordia e Maternidade | Instituto Adolfo Lutz, Interdiciplinary Procedures Center, Strategic Laboratory | Claudia Regina Gonçalves; Claudio Tavares Sacchi; Erica Valessa Ramos Gomes; Karoline Rodrigues Campos |
| EPI_ISL_547579 | Santa Casa de Misericórdia de Araçatuba | Instituto Adolfo Lutz, Interdiciplinary Procedures Center, | Claudia Regina Gonçalves; Claudio Tavares Sacchi; Erica Valessa Ramos Gomes; Karoline Rodrigues Campos |

| | | | |
|---|---|---|---|
| | | Strategic Laboratory | |
| EPI_ISL_524464, EPI_ISL_1121322 | Santa Casa de Santa Isabel | Instituto Adolfo Lutz, Interdiciplinary Procedures Center, Strategic Laboratory | Caio Vinicius Dias Lopes; Claudia Regina Gonçalves; Claudio Tavares Sacchi; Erica Valessa Ramos Gomes; Karoline Rodrigues Campos |
| EPI_ISL_1468466 | Santa Casa de Sao Carlos | Instituto Adolfo Lutz, Interdiciplinary Procedures Center, Strategic Laboratory | Caio Vinicius Dias Lopes; Claudia Regina Gonçalves; Claudio Tavares Sacchi; Erica Valessa Ramos Gomes; Karoline Rodrigues Campos |
| EPI_ISL_1533692 | Santa Casa de Sao Paulo | Instituto Adolfo Lutz, Interdiciplinary Procedures Center, Strategic Laboratory | Caio Vinicius Dias Lopes; Claudia Regina Gonçalves; Claudio Tavares Sacchi; Erica Valessa Ramos Gomes; Karoline Rodrigues Campos; Leonardo Jose Tadeu de Araujo |
| EPI_ISL_693238, EPI_ISL_693239 | Secao Centro de Diagnostico Secedi | Instituto Adolfo Lutz, Interdiciplinary Procedures Center, Strategic Laboratory | Claudia Regina Gonçalves; Claudio Tavares Sacchi; Erica Valessa Ramos Gomes; Karoline Rodrigues Campos |
| EPI_ISL_735405 | Secretaria Minucipal de Saude de Birigui | Instituto Adolfo Lutz, Interdiciplinary Procedures Center, Strategic Laboratory | Claudia Regina Gonçalves; Claudio Tavares Sacchi; Erica Valessa Ramos Gomes; Karoline Rodrigues Campos |
| EPI_ISL_2919013, EPI_ISL_2919014, EPI_ISL_2928510 | Secretaria Municipal da Saúde-Porto Alegre | Laboratório de Microbiologia Molecular - Universidade FEEVALE | Alana Witt Hansen; Fernando Rosado Spilki; Flávio Silveira; Fágner Henrique Heldt; Juliana Schons Gulart; Juliana Schons Gularte; Juliane Deise Fleck; Mariana Soares da Silva; Matheus Nunes Weber; Meriane Demoliner; Michele Filippi.; Micheli Filippi.; Paula Rodrigues de Almeida |
| EPI_ISL_882658 | Secretaria Municipal de Saude | Instituto Adolfo Lutz, Interdiciplinary Procedures Center, Strategic Laboratory | Claudia Regina Gonçalves; Claudio Tavares Sacchi; Erica Valessa Ramos Gomes; Karoline Rodrigues Campos |
| EPI_ISL_708530 | Secretaria Municipal de Saude de Fernandópolis | Instituto Adolfo Lutz, Interdiciplinary Procedures Center, Strategic Laboratory | Carlos Henrique Camargo; Claudia Regina Gonçalves; Claudio Tavares Sacchi; Erica Valessa Ramos Gomes; Fernanda Modesto Tolentino Binhardi; Janaina Other Martins Montanha; Karoline Rodrigues Campos; Marcia Maria Costa Nunes Soares; Maricelia Navarro Pinheiro Flores |
| EPI_ISL_574583 | Secretaria Municipal de Saude de Jandira | Instituto Adolfo Lutz, Interdiciplinary Procedures Center, Strategic Laboratory | Claudia Regina Gonçalves; Claudio Tavares Sacchi; Erica Valessa Ramos Gomes; Karoline Rodrigues Campos |
| EPI_ISL_574597 | Secretaria Municipal de Saude de Jarinu | Instituto Adolfo Lutz, Interdiciplinary Procedures Center, Strategic Laboratory | Claudia Regina Gonçalves; Claudio Tavares Sacchi; Erica Valessa Ramos Gomes; Karoline Rodrigues Campos |
| EPI_ISL_1040823 | Secretaria Municipal de Saude de Piracaia | Instituto Adolfo Lutz, Interdiciplinary Procedures Center, Strategic Laboratory | Claudia Regina Gonçalves; Claudio Tavares Sacchi; Erica Valessa Ramos Gomes; Karoline Rodrigues Campos |
| EPI_ISL_833164 | Secretaria Municipal de Saude de Santa Barbara d'oeste | Instituto Adolfo Lutz, Interdiciplinary Procedures Center, Strategic Laboratory | Claudia Regina Gonçalves; Claudio Tavares Sacchi; Erica Valessa Ramos Gomes; Karoline Rodrigues Campos |
| EPI_ISL_547576, EPI_ISL_603035 | Secretaria Municipal de Saúde | Instituto Adolfo Lutz, Interdiciplinary Procedures Center, Strategic Laboratory | Claudia Regina Gonçalves; Claudio Tavares Sacchi; Erica Valessa Ramos Gomes; Karoline Rodrigues Campos |
| EPI_ISL_693221 | Secretaria Municipal de Saúde de Birigui | Instituto Adolfo Lutz, Interdiciplinary Procedures Center, Strategic Laboratory | Claudia Regina Gonçalves; Claudio Tavares Sacchi; Erica Valessa Ramos Gomes; Karoline Rodrigues Campos |
| EPI_ISL_693215 | Secretaria Municipal de Saúde de Iracemapolis | Instituto Adolfo Lutz, Interdiciplinary Procedures Center, Strategic Laboratory | Claudia Regina Gonçalves; Claudio Tavares Sacchi; Erica Valessa Ramos Gomes; Karoline Rodrigues Campos |
| EPI_ISL_1469588, EPI_ISL_1469647 | Secretaria Municipal de Saúde de Taquara | Epiclin | Ana Paula Muterle; Carolina Comerlato; Eliana Márcia Da Ros Wendland; Fernando Hayashi Sant'Anna; Janira Prichula; Juliana Comerlato |
| EPI_ISL_1469803 | Secretaria Municipal de Saúde de Três Coroas | Epiclin | Ana Paula Muterle; Carolina Comerlato; Eliana Márcia Da Ros Wendland; Fernando Hayashi Sant'Anna; Janira Prichula; Juliana Comerlato |
| EPI_ISL_693228 | Secretaria Municipal de Sorocaba | Instituto Adolfo Lutz, Interdiciplinary Procedures Center, Strategic Laboratory | Claudia Regina Gonçalves; Claudio Tavares Sacchi; Erica Valessa Ramos Gomes; Karoline Rodrigues Campos |
| EPI_ISL_471542 | Secretaria de Saude de Mogi das Cruzes | Instituto Adolfo Lutz, Interdiciplinary Procedures Center, Strategic Laboratory | Claudia Regina Gonçalves; Claudio Tavares Sacchi; Erica Valessa Ramos Gomes |
| EPI_ISL_583502 | Serv de Vig Sanitaria Epidemio e CTRL de Zoonoses Guaruja | Instituto Adolfo Lutz, Interdiciplinary Procedures Center, Strategic Laboratory | Claudia Regina Gonçalves; Claudio Tavares Sacchi; Erica Valessa Ramos Gomes; Karoline Rodrigues Campos |
| EPI_ISL_2135301, EPI_ISL_2135303 | Servicio Virosis Respiratorias-Departamento Virología-INEI | Instituto Nacional Enfermedades Infecciosas C.G.Malbran | Avaro M.; Baumeister E.; Benedetti E.; Campos J.; Cisterna D.; Dattero ME; Lorenzo F.; Molina V.; Perandones C.; Poklepovich T.; Pontoriero A.; Russo M.; Tuduri E. |
| EPI_ISL_574598 | Servico de Verificacao de Obito SVO | Instituto Adolfo Lutz, Interdiciplinary Procedures Center, Strategic Laboratory | Claudia Regina Gonçalves; Claudio Tavares Sacchi; Erica Valessa Ramos Gomes; Karoline Rodrigues Campos |
| EPI_ISL_1165070 | Siti Khodijah Hospital | Institute of Tropical Disease, Universitas Airlangga | Aldise M Nastri; Gatot Soegiarto; Jezzy R Dewantari; Kazufumi Shimizu; Krisnoadi Rahardjo; Laksmi Wulandari; Maria I Lusida; Muhammad Hamdan; Resti Yudhawati; Rima R Prasetya; Soetjipto; Yasuko Mori |
| EPI_ISL_1154442 | Sonic - Labor Dr. von Froreich GmbH | Robert Koch Institute | |
| EPI_ISL_490026, EPI_ISL_593687, EPI_ISL_593698, EPI_ISL_593711 | South Eastern Area Laboratory Services (SEALS) | NSW Health Pathology - Institute of Clinical Pathology and Medical Research; Westmead Hospital; University of Sydney | CIDM-PH et al. |
| EPI_ISL_2105232 | South Super | Philippine Genome | Alethea R. de Guzman; Anna Ong-Lim; Arianne A. Zamora; Asia Louisa U. Chong; Benedict A. Maralit; Candice Francheska B. Tambaoan; Carlo M. Lapid; Celia Carlos; Devon Ray Pacial; Edsel Maurice Salvaña; El King D. Morado; Eva Maria Cutiongco-de la Paz; Francis A. Tablizo; Irish Coleen A. Asin; Jaime C. Montoya; Jan Michael C. Yap; Jo- |

| | | | |
|---|---|---|---|
| | Highway Molecular Diagnostic Laboratory | Center | Hannah S. Llames; John Q. Wong; Joshua Gregor A. Dizon; Juan Antonio R. Magalang; Karol Sophia Agape R. Padilla; Kenneth M. Kim; Kris P. Punayan; Marc Edsel C. Ayes; Marc Jerrone R. Castro; Maria Rosario Singh-Vergeire and Cynthia P. Saloma; Maria Sofia L. Yangzon; Marissa Alejandria; Razel Nikka M. Hao; Rianna Patricia S. Cruz; Sheila Mae M. Araiza |
| EPI_ISL_2105264, EPI_ISL_2105266, EPI_ISL_2105272 | Southern Philippines Medical Center (SPMC) | Philippine Genome Center | Alethea R. de Guzman; Anna Ong-Lim; Arianne A. Zamora; Asia Louisa U. Chong; Benedict A. Maralit; Candice Francheska B. Tambaoan; Carlo M. Lapid; Celia Carlos; Devon Ray Pacial; Edsel Maurice Salvaña; El King D. Morado; Eva Maria Cutiongco-de la Paz; Francis A. Tablizo; Irish Coleen A. Asin; Jaime C. Montoya; Jan Michael C. Yap; Jo-Hannah S. Llames; John Q. Wong; Joshua Gregor A. Dizon; Juan Antonio R. Magalang; Karol Sophia Agape R. Padilla; Kenneth M. Kim; Kris P. Punayan; Marc Edsel C. Ayes; Marc Jerrone R. Castro; Maria Rosario Singh-Vergeire and Cynthia P. Saloma; Maria Sofia L. Yangzon; Marissa Alejandria; Razel Nikka M. Hao; Rianna Patricia S. Cruz; Sheila Mae M. Araiza |
| EPI_ISL_2105293, EPI_ISL_2105294, EPI_ISL_2105300, EPI_ISL_2105301, EPI_ISL_2105302, EPI_ISL_2105303, EPI_ISL_2105305, EPI_ISL_2105308, EPI_ISL_2105309, EPI_ISL_2105314, EPI_ISL_2105316, EPI_ISL_2105319, EPI_ISL_2105321, EPI_ISL_2105324, EPI_ISL_2105330, EPI_ISL_2105334, EPI_ISL_2105336, EPI_ISL_2105338, EPI_ISL_2105342, EPI_ISL_2105343, EPI_ISL_2105344, EPI_ISL_2105345, EPI_ISL_2105347, EPI_ISL_2105348, EPI_ISL_2105349, EPI_ISL_2105350, EPI_ISL_2105351, EPI_ISL_2105354, EPI_ISL_2105355, EPI_ISL_2105357, EPI_ISL_2105358, EPI_ISL_2105362, EPI_ISL_2105368, EPI_ISL_2105372, EPI_ISL_2105374, EPI_ISL_2105376, EPI_ISL_2105384, EPI_ISL_2105385, EPI_ISL_2105386, EPI_ISL_2105387, EPI_ISL_2105389, EPI_ISL_2105425 | | | |
| see above | Southern Philippines Medical Center (SPMC-SNL-MBL) | Philippine Genome Center | Alethea R. de Guzman; Anna Ong-Lim; Arianne A. Zamora; Asia Louisa U. Chong; Benedict A. Maralit; Candice Francheska B. Tambaoan; Carlo M. Lapid; Celia Carlos; Devon Ray Pacial; Edsel Maurice Salvaña; El King D. Morado; Eva Maria Cutiongco-de la Paz; Francis A. Tablizo; Irish Coleen A. Asin; Jaime C. Montoya; Jan Michael C. Yap; Jo-Hannah S. Llames; John Q. Wong; Joshua Gregor A. Dizon; Juan Antonio R. Magalang; Karol Sophia Agape R. Padilla; Kenneth M. Kim; Kris P. Punayan; Marc Edsel C. Ayes; Marc Jerrone R. Castro; Maria Rosario Singh-Vergeire and Cynthia P. Saloma; Maria Sofia L. Yangzon; Marissa Alejandria; Razel Nikka M. Hao; Rianna Patricia S. Cruz; Sheila Mae M. Araiza |
| EPI_ISL_1364498 | Stanford Health Care | Stanford University School of Medicine, Clinical Virology Laboratory | ChunHong Huang; Daniel Solis; Fumiko Yamamoto; James Zehnder; Malaya K. Sahoo; Mamdouh Sibai; Michelle Verghese; and Benjamin A. Pinsky |
| EPI_ISL_1760559, EPI_ISL_2280076 | TXDSHS | TXDSHS | Anita Pokharel; Bonnie Oh; Chun Wang; Grace Kubin; Jenny Zhang; Karen Bobier; Lorraine Rodriguez; Maliha Rahman; Mayela Pedrueza; Myong Koag; Rachel Lee; Rashmi Tuladhar |
| EPI_ISL_1660606, EPI_ISL_1660607, EPI_ISL_1660610, EPI_ISL_1660613, EPI_ISL_1660614, EPI_ISL_1760108 | Texas Department of State Health Services (TXDSHS) | Texas Department of State Health Services (TXDSHS) | Anita Pokharel; Bonnie Oh; Chun Wang; Grace Kubin; Jenny Zhang; Lorraine Rodriguez; Maliha Rahman; Mayela Pedrueza; Myong Koag; Rachel Lee; Rashmi Tuladhar |
| EPI_ISL_672205 | The Ashley Laboratory, Stanford University | Chan-Zuckerberg Biohub | CZB Cliahub Consortium |
| EPI_ISL_2105280, EPI_ISL_2105546, EPI_ISL_2188275 | The Lord's Grace Medical and Industrial Clinic | Philippine Genome Center | Alethea R. de Guzman; Anna Ong-Lim; Arianne A. Zamora; Asia Louisa U. Chong; Benedict A. Maralit; Candice Francheska B. Tambaoan; Carlo M. Lapid; Celia Carlos; Devon Ray Pacial; Edsel Maurice Salvaña; El King D. Morado; Elcid Aaron R. Pangilinan; Eva Maria Cutiongco-de la Paz; Francis A. Tablizo; Irish Coleen A. Asin; Jaime C. Montoya; Jan Michael C. Yap; Jo-Hannah S. Llames; John Q. Wong; Joshua Gregor A. Dizon; Juan Antonio R. Magalang; Karol Sophia Agape R. Padilla; Kenneth M. Kim; Kris P. Punayan; Marc Edsel C. Ayes; Marc Jerrone R. Castro; Maria Rosario Singh-Vergeire and Cynthia P. Saloma; Maria Sofia L. Yangzon; Marissa Alejandria; Razel Nikka M. Hao; Rianna Patricia S. Cruz; Sheila Mae M. Araiza |
| EPI_ISL_2105292, EPI_ISL_2105335, EPI_ISL_2105369, EPI_ISL_2105411 | The Medical City -Â Ortigas | Philippine Genome Center | Alethea R. de Guzman; Anna Ong-Lim; Arianne A. Zamora; Asia Louisa U. Chong; Benedict A. Maralit; Candice Francheska B. Tambaoan; Carlo M. Lapid; Celia Carlos; Devon Ray Pacial; Edsel Maurice Salvaña; El King D. Morado; Eva Maria Cutiongco-de la Paz; Francis A. Tablizo; Irish Coleen A. Asin; Jaime C. Montoya; Jan Michael C. Yap; Jo-Hannah S. Llames; John Q. Wong; Joshua Gregor A. Dizon; Juan Antonio R. Magalang; Karol Sophia Agape R. Padilla; Kenneth M. Kim; Kris P. Punayan; Marc Edsel C. Ayes; Marc Jerrone R. Castro; Maria Rosario Singh-Vergeire and Cynthia P. Saloma; Maria Sofia L. Yangzon; Marissa Alejandria; Razel Nikka M. Hao; Rianna Patricia S. Cruz; Sheila Mae M. Araiza |
| EPI_ISL_3912447 | UAPS LUIS COSTA | Analytical Competence Molecular Epidemiology Lab/ACME, Oswaldo Cruz Foundation, Ceara (FIOCRUZ CE) | Cleber Furtado Aksenen; Fabio Miyajima; Fernando Braga Stehling; Francisco Eder de Moura Lopes; Jamille Maria Mendes Bezerra; Joaquim Cesar do Nascimento Sousa Junior; Pedro Miguel Carneiro Jeronimo; Suzana Porto Almeida & Lucas Delerino on behalf of COVID-19 FIOCRUZ Genomic Network; Thais Ferreira de Oliveira; Thais de Oliveira Costa; Ticiane Cavalcante de Souza; Veridiana Pessoa Miyajima |
| EPI_ISL_3912450 | UAPS PEDRO CELESTINO | Analytical Competence Molecular Epidemiology Lab/ACME, Oswaldo Cruz Foundation, Ceara (FIOCRUZ CE) | Cleber Furtado Aksenen; Fabio Miyajima; Fernando Braga Stehling; Francisco Eder de Moura Lopes; Jamille Maria Mendes Bezerra; Joaquim Cesar do Nascimento Sousa Junior; Pedro Miguel Carneiro Jeronimo; Suzana Porto Almeida & Lucas Delerino on behalf of COVID-19 FIOCRUZ Genomic Network; Thais Ferreira de Oliveira; Thais de Oliveira Costa; Ticiane Cavalcante de Souza; Veridiana Pessoa Miyajima |
| EPI_ISL_735419 | UBS Alvarenga | Instituto Adolfo Lutz, Interdiciplinary Procedures Center, Strategic Laboratory | Claudia Regina Gonçalves; Claudio Tavares Sacchi; Erica Valessa Ramos Gomes; Karoline Rodrigues Campos |
| EPI_ISL_837053 | UBS Darcy Alves e Robalinho | Instituto Adolfo Lutz, Interdiciplinary Procedures Center, Strategic Laboratory | Claudia Regina Gonçalves; Claudio Tavares Sacchi; Erica Valessa Ramos Gomes; Karoline Rodrigues Campos |
| EPI_ISL_735422 | UBS Dematchi | Instituto Adolfo Lutz, Interdiciplinary Procedures Center, Strategic Laboratory | Claudia Regina Gonçalves; Claudio Tavares Sacchi; Erica Valessa Ramos Gomes; Karoline Rodrigues Campos |
| EPI_ISL_1966553 | UBS II DE TANABI MILTON MARTINS PERCHES | Instituto Butantan / FZEA-USP (Pirassununga) | Antonio Jorge Martins; Bianca Cechetto Carlos. Mendelics: Bibiana Santos; Claudia Renata dos Santos Barros; Cíntia Bittar; David Schlesinger. Hemocentro Ribeirão Preto: Simone Kashima; Debora Botequio Moretti; Elaine Cristina Marqueze; Elaine Vieira dos Santos; Elisangela Chicaroni Mattos; Erika Freitas; Evandra Strazza Rodrigues; Felipe Allan da Silva da Costa; Flavia Aburjaile; Fábio Sossai Possebon; Guilherme Campos; Guilherme Targino Valente; Heidge Fukumasu. USP-Botucatu: Rejane Maria Tommasini Grotto; Helena Lage Ferreira; Instituto Butantan: Dimas Tadeu Covas; Jardelina de Souza Todao Bernardino; Jayme A. Souza-Neto; Jessika Cristina Chagas Lesbon; Jorge A. Petroli Marchesi; José Salvatore Leister Patané; João Paulo Kitajima; João Pessoa Araújo Jr.; Leila Sabrina Ullmann; Loyze Paola Oliveira de Lima; Luiz Aurelio de Campos Crispin. Centro de Genômica Funcional da ESALQ: Luiz Lehmann Coutinho; Luiz Carlos Junior de Alcantara; Lívia Sacchetto; Maisa C. Pereira Parra; Maria Carolina Elias; Marta Giovanetti; Marília Moraes; Maurício Lacerda Nogueira. Prefeitura de Sao Paulo: Melissa Palmieri.; Patricia Akemi Assato; Paula Rahal; Paulo Inacio da Costa; Rafael dos Santos Bezerra; Raquel de Lello Rocha Campos Cassano. NGS Soluções Genômicas: Pilar Drummond Sampaio Corrêa Mariani. FZEA-USP Pirassununga: Mirele Daiana Poleti; Raul Machado Neto; Ricardo Augusto Brassaloti; Ricardo Haddad; Rodrigo Tocantins Calado. FAMERP-SJRP: Cecília Artico Banho; Sandra Coccuzzo Sampaio; Svetoslav Nanev Slavov; Vagner Fonseca; Vincent Louis Viala |
| EPI_ISL_837054 | UBS Jose Sabino Ferreira | Instituto Adolfo Lutz, Interdiciplinary Procedures Center, Strategic Laboratory | Claudia Regina Gonçalves; Claudio Tavares Sacchi; Erica Valessa Ramos Gomes; Karoline Rodrigues Campos |
| EPI_ISL_735421 | UBS Sta Terezinha | Instituto Adolfo Lutz, Interdiciplinary Procedures Center, Strategic Laboratory | Claudia Regina Gonçalves; Claudio Tavares Sacchi; Erica Valessa Ramos Gomes; Karoline Rodrigues Campos |
| EPI_ISL_471648 | UBS e Pronto Socorro Jd. Jacira | Instituto Adolfo Lutz, Interdiciplinary Procedures Center, Strategic Laboratory | Claudia Regina Gonçalves; Claudio Tavares Sacchi; Erica Valessa Ramos Gomes |
| EPI_ISL_2758653, EPI_ISL_2758655, EPI_ISL_2758659, EPI_ISL_2758660, EPI_ISL_2758663, EPI_ISL_2758664, EPI_ISL_2758665, EPI_ISL_2758666, EPI_ISL_2758680, EPI_ISL_2758689, EPI_ISL_2758690, EPI_ISL_2758691, EPI_ISL_2758697, EPI_ISL_2758721, EPI_ISL_2758751, EPI_ISL_2758753, EPI_ISL_2758755, EPI_ISL_2758766, EPI_ISL_2758777, EPI_ISL_2758779, EPI_ISL_2758783, EPI_ISL_2758784, EPI_ISL_2758785, EPI_ISL_2758788, EPI_ISL_2758792 | | | |
| see above | UEL | IPEC Guarapuava | NAPI-Genômica (Novos Arranjo de Pesquisa e Inovação em Genômica): Ademar Dantas da Cunha Júnior Adriano Ferrasa Adriano Mondini Aldo Przybysz Alessandra Lourenço Cecchini Armani Alex Sandro Jorge Alexandra Ivo de Medeiros Alexandre Maller Aline Cristina Batista Rodrigues Johann Ana Lucia Ferreira Ana Marisa Fusco Almeida Anderson Joel Martino Andrade André Luís Laforga Vanzela Andrea Duarte Doetzer Andrea Name Colado Simao Andressa Pereira de Souza Anelisa Ramão Angelica Beate Winter Boldt Anna Hermínia Castro Gomes de Amorim Anna Silvia Penteado Setti da Rocha Antonio Camilo da Silva Filho Antonio Stabelini Neto Arthur Hirata Bertachi Barbara Mendes Paz Chao Betty Cristiane Kuhn Bruno Ambrozio Galindo Bruno Ribeiro Cruz Camilla Reginatto De Pierri Carla Fredrichsen Moya Araujo Carla Fredrichsen Moya Araujo Carlos Alberto Oliveira de Biagi Junior Carlos Augusto Nassar Carlos Eduardo Buss Carlos Gilberto Carlotti Junior Carlos Henrique Schneider Carolina Panis Carolina Weigert Galvão Caroline de Jesus Coelho Donha Caroline Guisantes de Salvo Toni Caryna Eurich Mazur Catiuscie Cabreira da Silva Tortorella Celso F. D. Doliveira Cesar Luiz Boguszewski Christiane Pienna Soares Chung Man Chin Claudia Moro Cleverson Busso Cristiane Cominetti Daiane Priscila Simão-Silva Dalila Luciola Zanette Daniel de Paula Daniel de Paula Daniel Rech Daniela Fiori Gradia Daniela Pretti da Cunha Tirapelli Daniela Viganó Zanoti Jeronymo Daniele Ukan Danielle Malheiros Ferreira Danielle Venturini Deborah Catharine de Assis Leite Deivid Calebe de Souza Dennis Armando Bertolini Edenir Inez Pamero Edna Maria Vissoci Reiche Edson Roberto Arpini Miguel Eduardo José de Almeida Araújo Eliana Carolina Vespero Eliandro Reis Tavares Elza Kimura Grimshaw Emanuel Maltempi de Souza Emanuele Cristina Gustani Buss Émerson Carraro Emiliana Cristina Melo ENILze Maria de Souza Fonseca Ribeiro Enilze Maria de Souza Fonseca Ribeiro Erika Izumi Erika Seki Kioshima Cotica Evani Marques Pereira Fabio Negretti Fábio Rodrigues Ferreira Seiva Felipe Dunin dos Santos Felipe Tuon Fernanda Andreia Rosa Fernanda Cestaro Prado Cortez Fernanda Ivanski Fernanda Maris Peria Flavia Regina Oliveira de Barros Franciele Aní Caovilla Follador Franciele Mara Lucca Zanardo Bohm Francinete Ramos Campos Fulviana Silva Nishiyama GABRIEL RIBEIRO CORDEIRO Gabriela Datsch Bennemann Gisele Santos de Oliveira Glaucio Valdameri Glauco Akelinghton Freire Vitiello Glauco Vieira Miranda Glaura Scantamburlo ALves Fernandes Guilherme Ferreira Silveira Gustavo Bianchini Porfírio Gustavo Lenci Marques Hélito Volpato Hildebrando Masshiro Nagai Huei Diana Lee Ilce Mara de Syllos Cólus Iris Rabinovich Israel Gomy Jackson Kawakami Jacques Duílio Brancher Jaime Luís Lopes Rocha Jaqueline Carvalho de Oliveira Jean Henrique da Silva Rodrigues Jean Leandro dos Santos Jeane Eliete Laguila Visentainer João Paulo Bianchi Ximenez Joaquim Manoel da Silva Jociani Ascari Joel Donazzolo Jorge Luis Maria Ruiz Jose Knopfholz José Luis da Conceição Silva José Sebastião dos Santos Joseane Carla Schabarum Juliana Cheleski Wiggers Juliana Mara Serpeloni Juliana Morini Küpper Cardoso Perseguini Karen Brajão de Oliveira Karin Braun Prado Karine Aparecida de Lima Katiany Rizzieri Caleffi Ferracioli Katiuscia de Oliveira Francisco Gabriel Kelvinson Fernandes Viana Larissa Beatriz Cossalter Larissa Danielle Bahls Pinto Laurival Antonio Vilas Boas Léia Carolina Lucio Líbero Mezzadri Neto Ligia Carla Faccin Galhardi Lirane Elize Defante Ferreto Luciana Furlaneto Maia Luciana Oliveira de Fariña Luciana Reis Azevedo Alanis Luciane Regina Cavalli Lucy Megumi Yamauchi Lioni Luis Paulo Gomes Mascarenhas Luis Paulo Gomes Mascarenhas Luís Paulo Mascarenhas Lupe Furtado Alle Lyvia Regina Biagi Silva Bertachi Mara Antonia Ramos Costa Mara L. Cordeiro Marcela Maria Birolim Marcelo Ricardo Vicari Marcia Edilaine Lopes Consolaro Marcia Holsbach Beltrame Marcia Regina Eches Perugini Marcos Abdo Arbex Marcos Pileggi MARCOS TADEU GRZELCZAK Marcus Peikriszwili Tartaruga Maria Angelica Ehara Watanabe Maria Antonia Ramos Costa Maria Claudia Gross Maria José Soares Mendes Giannini Maria Leandra Terencio Maria Lúcia Bonfleur Maria Luiza Guimarães de Oliveira Maria Luiza Petzl-Erler Mariana Abe Vicente Cavagnari Marina Kimiko Kadowaki Marise Fonseca dos Santos Marla Karine Amarante Maurício Turkiewicz Mauro Antonio Alves Castro Michel Rodrigo Zambrano Passarini Michele Potrich Michelle Orane Schemberger Milena Massumi Kozonoe Mônica Degraf Cavallin Monica Tereza Suldofski Mucio Luiz de Assis Cirino Nadia Graciele Krohn Najeh Maissar Khalil Nédia de Castilhos Ghisi Neide Tomimura Costa Neiva Leite Neyva Maria Lopes Romeiro Patrícia Amâncio da Rosa Patricia Dayane Carvalho Schaker Patricia Oehlmeyer Nassar Patrícia Savio de Araújo-Souza Patricia Silva Lucio Paulo Henrique Couto Souza Paulo Roberto Donadio Percy Nohama Quirino Alves de Lima Neto Rafael Deminice Rafael dos Santos Bezerra Raquel Alves dos Santos Renan Manozzo Galante Renata Ernlund Freitas de Macedo Rita de Cássia Garcia Simão Roberta Losi Guembarovski Roberto H. Herai Roberto Rosati Rodrigo Ferreira Rodrigo Rodrigues Matiello Rogério Neri Shinsato Rogério Pincela Mateus Rosane Aparecida Ribeiro Rosilene Fressatti Cardoso Rosilene Fressatti Cardoso Sandra Mara Guse Scós Venske Selene Elifio Esposito Sérgio Ossamu Ioshii Silvana Giuliatti Silvia Mara de Souza Halick Silvio Henrique Maia de Almeida Simone Neumann Wendt Spencer Luiz Marques Payão Stefan Wolanski Negrão Stephane Janaina de Moura Escobar Sueli Fumie Yamada Ogatta SUELI PERCIO QUINAIA Taciane Finatto Tatiana Mayumi Veiga Iriyoda Tayza Katelline Danilau Ostroski Tony Alexander Hild Valeria Valente Vanessa Nascimento Kozak Vanessa Santos Sotomaior Victor Breno Pedrosa Victoria Zeghbi Cochenski Borba Vivian Rotuno Moure Valdameri Wander Rogerio Pavanelli Weber Cláudio Francisco Nunes da Silva Willian Augusto de Melo Yohandra Reyes Torres |
| EPI_ISL_941550, EPI_ISL_941552 | ULS Guarda | Instituto Nacional de Saude (INSA) | Borges et al |
| EPI_ISL_732179, EPI_ISL_732180, EPI_ISL_732181, EPI_ISL_732183, EPI_ISL_732184 | ULS Guarda | Instituto Nacional de Saude (INSA) and Instituto Gulbenkian de Ciencia (IGC) | Borges et al |

| | | | |
|---|---|---|---|
| EPI_ISL_941583 | ULS Litoral Alentejano | Instituto Nacional de Saude (INSA) | Borges et al |
| EPI_ISL_941896 | ULSNE - Braganca | Instituto Nacional de Saude (INSA) and Instituto Gulbenkian de Ciencia (IGC) | Borges et al |
| EPI_ISL_1469719, EPI_ISL_1469733 | UNIDADE DE PRONTO ATENDIMENTO DE SAPUCAIA DO SUL UPA | Epiclin | Ana Paula Muterle; Carolina Comerlato; Eliana Márcia Da Ros Wendland; Fernando Hayashi Sant'Anna; Janira Prichula; Juliana Comerlato |
| EPI_ISL_1469720, EPI_ISL_1469722 | UNIDADE SANITARIA DE IGREJINHA | Epiclin | Ana Paula Muterle; Carolina Comerlato; Eliana Márcia Da Ros Wendland; Fernando Hayashi Sant'Anna; Janira Prichula; Juliana Comerlato |
| EPI_ISL_2209934 | UNIDADE SENTINELA COVID19 | Instituto Butantan | Antonio Jorge Martins; Claudia Renata dos Santos Barros; David Schlesinger; Debora Botequio Moretti; Dimas Tadeu Covas; Elaine Cristina Marqueze; Elaine Vieira Santos; Evandra Strazza Rodrigues; Heidge Fukumasu; Jayme Augusto de Souza-Neto; José Salvatore Leister Patané; Luiz Alcantara; Luiz Lehmann Coutinho; Maria Carolina Elias; Maurício Lacerda Nogueira; Rafael dos Santos Bezerra; Raul Machado Neto; Rejane Maria Tommasini Grotto; Ricardo Haddad; Sandra Coccuzzo Sampaio Vessoni; Simone Kashima; Svetoslav Nanev Slavov; Vincent Louis Viala. |
| EPI_ISL_1966752 | UNIDADE SENTINELA COVID19 | Instituto Butantan / Mendelics | Antonio Jorge Martins; Bianca Cechetto Carlos. Mendelics: Bibiana Santos; Claudia Renata dos Santos Barros; Cíntia Bittar; David Schlesinger. Hemocentro Ribeirão Preto: Simone Kashima; Debora Botequio Moretti; Elaine Cristina Marqueze; Elaine Vieira dos Santos; Elisangela Chicaroni Mattos; Erika Freitas; Evandra Strazza Rodrigues; Felipe Allan da Silva da Costa; Flavia Aburjaile; Fábio Sossai Possebon; Guilherme Campos; Guilherme Targino Valente; Heidge Fukumasu. USP-Botucatu: Rejane Maria Tommasini Grotto; Helena Lage Ferreira; Instituto Butantan: Dimas Tadeu Covas; Jardelina de Souza Todao Bernardino; Jayme A. Souza-Neto; Jessika Cristina Chagas Lesbon; Jorge A. Petroli Marchesi; José Salvatore Leister Patané; João Paulo Kitajima; João Pessoa Araújo Jr.; Leila Sabrina Ullmann; Loyze Paola Oliveira de Lima; Luiz Aurelio de Campos Crispin. Centro de Genômica Funcional da ESALQ: Luiz Lehmann Coutinho; Luiz Carlos Junior de Alcantara; Lívia Sacchetto; Maisa C. Pereira Parra; Maria Carolina Elias; Marta Giovanetti; Marília Moraes; Maurício Lacerda Nogueira. Prefeitura de Sao Paulo: Melissa Palmieri.; Patricia Akemi Assato; Paula Rahal; Paulo Inacio da Costa; Rafael dos Santos Bezerra; Raquel de Lello Rocha Campos Cassano. NGS Soluções Genômicas: Pilar Drummond Sampaio Corrêa Mariani. FZEA-USP Pirassununga: Mirele Daiana Poleti; Raul Machado Neto; Ricardo Augusto Brassaloti; Ricardo Haddad; Rodrigo Tocantins Calado. FAMERP-SJRP: Cecília Artico Banho; Sandra Coccuzzo Sampaio; Svetoslav Nanev Slavov; Vagner Fonseca; Vincent Louis Viala |
| EPI_ISL_2344658 | UNIDADE SENTINELA COVID19 | Instituto Butantan / UNESP-Botucatu | Antonio Jorge Martins; Claudia Renata dos Santos Barros; David Schlesinger; Debora Botequio Moretti; Dimas Tadeu Covas; Elaine Cristina Marqueze; Elaine Vieira Santos; Evandra Strazza Rodrigues; Heidge Fukumasu; Jayme Augusto de Souza-Neto; José Salvatore Leister Patané; Luiz Alcantara; Luiz Lehmann Coutinho; Maria Carolina Elias; Maurício Lacerda Nogueira; Rafael dos Santos Bezerra; Raul Machado Neto; Rejane Maria Tommasini Grotto; Ricardo Haddad; Sandra Coccuzzo Sampaio Vessoni; Simone Kashima; Svetoslav Nanev Slavov; Vincent Louis Viala |
| EPI_ISL_2105325 | UP Health Services (PGC) | Philippine Genome Center | Alethea R. de Guzman; Anna Ong-Lim; Arianne A. Zamora; Asia Louisa U. Chong; Benedict A. Maralit; Candice Francheska B. Tambaoan; Carlo M. Lapid; Celia Carlos; Devon Ray Pacial; Edsel Maurice Salvaña; El King D. Morado; Eva Maria Cutiongco-de la Paz; Francis A. Tablizo; Irish Coleen A. Asin; Jaime C. Montoya; Jan Michael C. Yap; Jo-Hannah S. Llames; John Q. Wong; Joshua Gregor A. Dizon; Juan Antonio R. Magalang; Karol Sophia Agape R. Padilla; Kenneth M. Kim; Kris P. Punayan; Marc Edsel C. Ayes; Marc Jerrone R. Castro; Maria Rosario Singh-Vergeire and Cynthia P. Saloma; Maria Sofia L. Yangzon; Marissa Alejandria; Razel Nikka M. Hao; Rianna Patricia S. Cruz; Sheila Mae M. Araiza |
| EPI_ISL_523983 | UPA Campo Limpo | Instituto Adolfo Lutz, Interdiciplinary Procedures Center, Strategic Laboratory | Claudia Regina Gonçalves; Claudio Tavares Sacchi; Erica Valessa Ramos Gomes |
| EPI_ISL_2346035 | UPA DR FRANCO DA ROCHA | Instituto Butantan | Antonio Jorge Martins; Claudia Renata dos Santos Barros; David Schlesinger; Debora Botequio Moretti; Dimas Tadeu Covas; Elaine Cristina Marqueze; Elaine Vieira Santos; Evandra Strazza Rodrigues; Heidge Fukumasu; Jayme Augusto de Souza-Neto; José Salvatore Leister Patané; Luiz Alcantara; Luiz Lehmann Coutinho; Maria Carolina Elias; Maurício Lacerda Nogueira; Rafael dos Santos Bezerra; Raul Machado Neto; Rejane Maria Tommasini Grotto; Ricardo Haddad; Sandra Coccuzzo Sampaio Vessoni; Simone Kashima; Svetoslav Nanev Slavov; Vincent Louis Viala |
| EPI_ISL_1123372 | UPA I Santa Isabel | Instituto Adolfo Lutz, Interdiciplinary Procedures Center, Strategic Laboratory | Caio Vinicius Dias Lopes; Claudia Regina Gonçalves; Claudio Tavares Sacchi; Erica Valessa Ramos Gomes; Karoline Rodrigues Campos |
| EPI_ISL_534311 | UPA III 26 de Agosto | Instituto Adolfo Lutz, Interdiciplinary Procedures Center, Strategic Laboratory | Claudia Regina Gonçalves; Claudio Tavares Sacchi; Erica Valessa Ramos Gomes |
| EPI_ISL_583496 | UPA Jandira | Instituto Adolfo Lutz, Interdiciplinary Procedures Center, Strategic Laboratory | Claudia Regina Gonçalves; Claudio Tavares Sacchi; Erica Valessa Ramos Gomes; Karoline Rodrigues Campos |
| EPI_ISL_693237, EPI_ISL_693245 | UPA Santa Isabel | Instituto Adolfo Lutz, Interdiciplinary Procedures Center, Strategic Laboratory | Claudia Regina Gonçalves; Claudio Tavares Sacchi; Erica Valessa Ramos Gomes; Karoline Rodrigues Campos |
| EPI_ISL_523975, EPI_ISL_523980 | UPA Tito Lopes | Instituto Adolfo Lutz, Interdiciplinary Procedures Center, Strategic Laboratory | Claudia Regina Gonçalves; Claudio Tavares Sacchi; Erica Valessa Ramos Gomes |
| EPI_ISL_468316 | UPA Vila Assis | Instituto Adolfo Lutz, Interdiciplinary Procedures Center, Strategic Laboratory | Claudia Regina Gonçalves; Claudio Tavares Sacchi; Erica Valessa Ramos Gomes |
| EPI_ISL_861682 | UPA Vila Santa Catarina | Instituto Adolfo Lutz, Interdiciplinary Procedures Center, Strategic Laboratory | Claudia Regina Gonçalves; Claudio Tavares Sacchi; Erica Valessa Ramos Gomes; Karoline Rodrigues Campos |
| EPI_ISL_861656 | UPA de Jandira | Instituto Adolfo Lutz, Interdiciplinary Procedures Center, Strategic Laboratory | Claudia Regina Gonçalves; Claudio Tavares Sacchi; Erica Valessa Ramos Gomes; Karoline Rodrigues Campos |
| EPI_ISL_1445201, EPI_ISL_1966234 | USAFA TUDE BASTOS | Instituto Butantan / Mendelics | Antonio Jorge Martins; Bianca Cechetto Carlos. Mendelics: Bibiana Santos; Bibiana Santos; Claudia Renata dos Santos Barros; Cíntia Bittar; David Schlesinger; David Schlesinger. Hemocentro Ribeirão Preto: Simone Kashima; Debora Botequio Moretti; Dimas Tadeu Covas; Elaine Cristina Marqueze; Elaine Vieira dos Santos; Elisangela Chicaroni Mattos; Erika Freitas; Evandra Strazza Rodrigues; Felipe Allan da Silva da Costa; Flavia Aburjaile; Fábio Sossai Possebon; Guilherme Campos; Guilherme Targino Valente; Heidge Fukumasu. USP-Botucatu: Rejane Maria Tommasini Grotto; Helena Lage Ferreira; Instituto Butantan: Dimas Tadeu Covas; Jardelina de Souza Todao Bernardino; Jayme A. Souza-Neto; Jessika Cristina Chagas Lesbon; Jorge A. Petroli Marchesi; José Salvatore Leister Patané; João Paulo Kitajima; João Pessoa Araújo Jr.; Leila Sabrina Ullmann; Loyze Paola Oliveira de Lima; Luiz Aurelio de Campos Crispin. Centro de Genômica Funcional da ESALQ: Luiz Lehmann Coutinho; Luiz Carlos Junior de Alcantara; Lívia Sacchetto; Maisa C. Pereira Parra; Maria Carolina Elias; Marta Giovanetti; Marília Moraes; Maurício Lacerda Nogueira. Prefeitura de Sao Paulo: Melissa Palmieri.; Patricia Akemi Assato; Paula Rahal; Paulo Inacio da Costa; Rafael dos Santos Bezerra; Raquel de Lello Rocha Campos Cassano. NGS Soluções Genômicas: Pilar Drummond Sampaio Corrêa Mariani. FZEA-USP Pirassununga: Mirele Daiana Poleti; Raul Machado Neto; Ricardo Augusto Brassaloti; Ricardo Haddad; Rodrigo Tocantins Calado.; Rodrigo Tocantins Calado. FAMERP-SJRP: Cecília Artico Banho; Sandra Coccuzzo Sampaio; Simone Kashima; Svetoslav Nanev Slavov; Vagner Fonseca; Vincent Louis Viala |
| EPI_ISL_1324137, EPI_ISL_1324140 | UW Virology Lab | UW Virology Lab | Alexander Greninger; Hong Xie; Keith R Jerome; Lasata Shrestha; Margaret Mills; Meei-Li Huang; Michelle Lin; Noah Baker; Pavitra Roychoudhury; Saraswathi Sathees; Sean Ellis; Shah Mohamed Bakhash |
| EPI_ISL_734865 | UZ Leuven, National Reference Laboratory for Coronaviruses, Laboratory Medicine, Leuven, Belgium | KU Leuven, Rega Institute, Clinical and Epidemiological Virology | Bert Vanmechelen; Joan Marti-Carerras; Piet Maes; Tony Wawina-Bokalanga |
| EPI_ISL_693225 | Ubs Vila Rosa - Olímpia Gomes De Almeida | Instituto Adolfo Lutz, Interdiciplinary Procedures Center, Strategic Laboratory | Claudia Regina Gonçalves; Claudio Tavares Sacchi; Erica Valessa Ramos Gomes; Karoline Rodrigues Campos |
| EPI_ISL_735397 | Unidade Respiratória Nova Hortolandia | Instituto Adolfo Lutz, Interdiciplinary Procedures Center, Strategic Laboratory | Claudia Regina Gonçalves; Claudio Tavares Sacchi; Erica Valessa Ramos Gomes; Karoline Rodrigues Campos |
| EPI_ISL_1469576 | Unidade Sanitária de Igrejinha | Epiclin | Ana Paula Muterle; Carolina Comerlato; Eliana Márcia Da Ros Wendland; Fernando Hayashi Sant'Anna; Janira Prichula; Juliana Comerlato |
| EPI_ISL_1469664, EPI_ISL_1469714 | Unidade de Atendimento DST AIDS TB e Han | Epiclin | Ana Paula Muterle; Carolina Comerlato; Eliana Márcia Da Ros Wendland; Fernando Hayashi Sant'Anna; Janira Prichula; Juliana Comerlato |
| EPI_ISL_574590 | Unidade de Printo Atendimento UPA I Santa Isabel | Instituto Adolfo Lutz, Interdiciplinary Procedures Center, Strategic Laboratory | Claudia Regina Gonçalves; Claudio Tavares Sacchi; Erica Valessa Ramos Gomes; Karoline Rodrigues Campos |

| | | | |
|---|---|---|---|
| EPI_ISL_735409 | Unidade de Pronto Atendimento Carlos Lourenco | Instituto Adolfo Lutz, Interdiciplinary Procedures Center, Strategic Laboratory | Claudia Regina Gonçalves; Claudio Tavares Sacchi; Erica Valessa Ramos Gomes; Karoline Rodrigues Campos |
| EPI_ISL_693214 | Unidade de Pronto Atendimento Central de Caraguatatuba | Instituto Adolfo Lutz, Interdiciplinary Procedures Center, Strategic Laboratory | Claudia Regina Gonçalves; Claudio Tavares Sacchi; Erica Valessa Ramos Gomes; Karoline Rodrigues Campos |
| EPI_ISL_882665 | Unidade de Pronto Atendimento Dra Zilda Arns | Instituto Adolfo Lutz, Interdiciplinary Procedures Center, Strategic Laboratory | Claudia Regina Gonçalves; Claudio Tavares Sacchi; Erica Valessa Ramos Gomes; Karoline Rodrigues Campos |
| EPI_ISL_693226 | Unidade de Pronto Atendimento Sao José | Instituto Adolfo Lutz, Interdiciplinary Procedures Center, Strategic Laboratory | Claudia Regina Gonçalves; Claudio Tavares Sacchi; Erica Valessa Ramos Gomes; Karoline Rodrigues Campos |
| EPI_ISL_735406 | Unidade de Pronto Atendimento UPA I Sta Isabel | Instituto Adolfo Lutz, Interdiciplinary Procedures Center, Strategic Laboratory | Claudia Regina Gonçalves; Claudio Tavares Sacchi; Erica Valessa Ramos Gomes; Karoline Rodrigues Campos |
| EPI_ISL_735417 | Unidade de Pronto Atendimento de Agenor de Campos | Instituto Adolfo Lutz, Interdiciplinary Procedures Center, Strategic Laboratory | Claudia Regina Gonçalves; Claudio Tavares Sacchi; Erica Valessa Ramos Gomes; Karoline Rodrigues Campos |
| EPI_ISL_1469580 | Unidade de Pronto Atendimento de Sapucaia do Sul | Epiclin | Ana Paula Muterle; Carolina Comerlato; Eliana Márcia Da Ros Wendland; Fernando Hayashi Sant'Anna; Janira Prichula; Juliana Comerlato |
| EPI_ISL_693216, EPI_ISL_693217 | Unidade de Vigilância Epiudemiológica de Araras | Instituto Adolfo Lutz, Interdiciplinary Procedures Center, Strategic Laboratory | Claudia Regina Gonçalves; Claudio Tavares Sacchi; Erica Valessa Ramos Gomes; Karoline Rodrigues Campos |
| EPI_ISL_2491722 | Universidade Federal do Sul da Bahia (UFSB) | Laboratory of Respiratory Viruses and Measles, Oswaldo Cruz Institute, FIOCRUZ | Alice Sampaio Rocha; Ana Carolina Mendonca; Anna Carolina Paixao; Elisa Cavalcante Pereira; Felicidade Pereira; Fernando Motta; Luciana Appolinario; Marilda Siqueira on behalf of the Fiocruz COVID-19 Genomic Surveillance Network; Paola Resende; Renata Serrano Lopes; Taina Venas; Thiago Mafra |
| EPI_ISL_873097 | University of Michigan Clinical Microbiology Laboratory | Lauring Lab, University of Michigan, Department of Microbiology and Immunology | Valesano |
| EPI_ISL_2105391 | University of the Philippines National Institutes of Health (UP NIH) | Philippine Genome Center | Alethea R. de Guzman; Anna Ong-Lim; Arianne A. Zamora; Asia Louisa U. Chong; Benedict A. Maralit; Candice Francheska B. Tambaoan; Carlo M. Lapid; Celia Carlos; Devon Ray Pacial; Edsel Maurice Salvaña; El King D. Morado; Eva Maria Cutiongco-de la Paz; Francis A. Tablizo; Irish Coleen A. Asin; Jaime C. Montoya; Jan Michael C. Yap; Jo-Hannah S. Llames; John Q. Wong; Joshua Gregor A. Dizon; Juan Antonio R. Magalang; Karol Sophia Agape R. Padilla; Kenneth M. Kim; Kris P. Punayan; Marc Edsel C. Ayes; Marc Jerrone R. Castro; Maria Rosario Singh-Vergeire and Cynthia P. Saloma; Maria Sofia L. Yangzon; Marissa Alejandria; Razel Nikka M. Hao; Rianna Patricia S. Cruz; Sheila Mae M. Araiza |
| EPI_ISL_1286592, EPI_ISL_1286611 | Universitätsklinikum Heidelberg | Robert Koch Institute | |
| EPI_ISL_693234 | Upa Vereador Jose Da Rocha Goncalves | Instituto Adolfo Lutz, Interdiciplinary Procedures Center, Strategic Laboratory | Claudia Regina Gonçalves; Claudio Tavares Sacchi; Erica Valessa Ramos Gomes; Karoline Rodrigues Campos |
| EPI_ISL_2284160 | VA Connecticut Healthcare System | Yale Center for Genomic Analysis | Brooke Sullivan; Curt Scharfe; Irina Tikhonova; Kaya Bilguvar; Shrikant Mane |
| EPI_ISL_1469573 | VIGILANCIA EM SAUDE NH | Epiclin | Ana Paula Muterle; Carolina Comerlato; Eliana Márcia Da Ros Wendland; Fernando Hayashi Sant'Anna; Janira Prichula; Juliana Comerlato |
| EPI_ISL_1445229, EPI_ISL_1445232, EPI_ISL_1445235, EPI_ISL_1445249, EPI_ISL_1445251, EPI_ISL_1445270, EPI_ISL_1966059, EPI_ISL_1966062, EPI_ISL_1966067, EPI_ISL_1966091, EPI_ISL_2170898 | | | |
| see above | VIGILANCIA EPIDEMIOLOGICA | Instituto Butantan / Mendelics | Antonio Jorge Martins; Bianca Cechetto Carlos. Mendelics: Bibiana Santos; Bibiana Santos; Claudia Renata dos Santos Barros; Cíntia Bittar; David Schlesinger; David Schlesinger. Hemocentro Ribeirão Preto: Simone Kashima; Debora Botequio Moretti; Dimas Tadeu Covas; Elaine Cristina Marqueze; Elaine Vieira dos Santos; Elisangela Chicaroni Mattos; Erika Freitas; Evandra Strazza Rodrigues; Felipe Allan da Silva da Costa; Flavia Aburjaile; Fábio Sossai Possebon; Guilherme Campos; Guilherme Targino Valente; Heidge Fukumasu. USP-Botucatu: Rejane Maria Tommasini Grotto; Helena Lage Ferreira; Instituto Butantan: Dimas Tadeu Covas; Jardelina de Souza Todao Bernardino; Jayme A. Souza-Neto; Jessika Cristina Chagas Lesbon; Jorge A. Petroli Marchesi; José Salvatore Leister Patané; João Paulo Kitajima; João Pessoa Araújo Jr.; Leila Sabrina Ullmann; Loyze Paola Oliveira de Lima; Luiz Aurelio de Campos Crispin. Centro de Genômica Funcional da ESALQ: Luiz Lehmann Coutinho; Luiz Carlos Junior de Alcantara; Lívia Sacchetto; Maisa C. Pereira Parra; Maria Carolina Elias; Marta Giovanetti; Marília Moraes; Maurício Lacerda Nogueira. Prefeitura de Sao Paulo: Melissa Palmieri.; Patricia Akemi Assato; Paula Rahal; Paulo Inacio da Costa; Rafael dos Santos Bezerra; Raquel de Lello Rocha Campos Cassano. NGS Soluções Genômicas: Pilar Drummond Sampaio Corrêa Mariani. FZEA-USP Pirassununga: Mirele Daiana Poleti; Raul Machado Neto; Ricardo Augusto Brassaloti; Ricardo Haddad; Rodrigo Tocantins Calado.; Rodrigo Tocantins Calado. FAMERP-SJRP: Cecília Artico Banho; Sandra Coccuzzo Sampaio; Simone Kashima; Svetoslav Nanev Slavov; Vagner Fonseca; Vincent Louis Viala |
| EPI_ISL_2345734 | VIGILANCIA EPIDEMIOLOGICA JARDINOPOLIS SP | Instituto Butantan / Mendelics | Antonio Jorge Martins; Claudia Renata dos Santos Barros; David Schlesinger; Debora Botequio Moretti; Dimas Tadeu Covas; Elaine Cristina Marqueze; Elaine Vieira Santos; Evandra Strazza Rodrigues; Heidge Fukumasu; Jayme Augusto de Souza-Neto; José Salvatore Leister Patané; Luiz Alcantara; Luiz Lehmann Coutinho; Maria Carolina Elias; Maurício Lacerda Nogueira; Rafael dos Santos Bezerra; Raul Machado Neto; Rejane Maria Tommasini Grotto; Ricardo Haddad; Sandra Coccuzzo Sampaio Vessoni; Simone Kashima; Svetoslav Nanev Slavov; Vincent Louis Viala |
| EPI_ISL_2105341 | Valenzuela Hope Molecular Laboratory | Philippine Genome Center | Alethea R. de Guzman; Anna Ong-Lim; Arianne A. Zamora; Asia Louisa U. Chong; Benedict A. Maralit; Candice Francheska B. Tambaoan; Carlo M. Lapid; Celia Carlos; Devon Ray Pacial; Edsel Maurice Salvaña; El King D. Morado; Eva Maria Cutiongco-de la Paz; Francis A. Tablizo; Irish Coleen A. Asin; Jaime C. Montoya; Jan Michael C. Yap; Jo-Hannah S. Llames; John Q. Wong; Joshua Gregor A. Dizon; Juan Antonio R. Magalang; Karol Sophia Agape R. Padilla; Kenneth M. Kim; Kris P. Punayan; Marc Edsel C. Ayes; Marc Jerrone R. Castro; Maria Rosario Singh-Vergeire and Cynthia P. Saloma; Maria Sofia L. Yangzon; Marissa Alejandria; Razel Nikka M. Hao; Rianna Patricia S. Cruz; Sheila Mae M. Araiza |
| EPI_ISL_468313, EPI_ISL_468319, EPI_ISL_603033 | Vigilancia Epidemiologica de São Bernardo do Campo | Instituto Adolfo Lutz, Interdiciplinary Procedures Center, Strategic Laboratory | Claudia Regina Gonçalves; Claudio Tavares Sacchi; Erica Valessa Ramos Gomes; Karoline Rodrigues Campos |
| EPI_ISL_515542 | Vigilância Epidemiológica de Leme | Instituto Adolfo Lutz, Interdiciplinary Procedures Center, Strategic Laboratory | Claudia Regina Gonçalves; Claudio Tavares Sacchi; Erica Valessa Ramos Gomes |
| EPI_ISL_603023 | Vigilância em Saúde Visa Sul | Instituto Adolfo Lutz, Interdiciplinary Procedures Center, Strategic Laboratory | Claudia Regina Gonçalves; Claudio Tavares Sacchi; Erica Valessa Ramos Gomes; Karoline Rodrigues Campos |
| EPI_ISL_547573 | Vigilância em Saúde de Cajamar | Instituto Adolfo Lutz, Interdiciplinary Procedures Center, Strategic Laboratory | Claudia Regina Gonçalves; Claudio Tavares Sacchi; Erica Valessa Ramos Gomes; Karoline Rodrigues Campos |
| EPI_ISL_1469696 | Vigilância em Saúde de Sapucaia do Sul | Epiclin | Ana Paula Muterle; Carolina Comerlato; Eliana Márcia Da Ros Wendland; Fernando Hayashi Sant'Anna; Janira Prichula; Juliana Comerlato |
| EPI_ISL_1004317 | Viollier AG | Department of Biosystems Science and Engineering, ETH Zürich | Andrea Patrignani; Andreia Cabral de Gouvea; Catharine Aquino; Chaoran Chen; Christiane Beckmann; Christoph Noppen; David Dreifuss; Doris Popovic; Griffin White; Ivan Topolsky; Jay Tracy; Katharina Jahn; Lara Fuhrmann; Laura Neff; Lennart Opitz; Maria Domenica Moccia; Maurice Redondo; Niko Beerenwinkel; Noemie Santamaria de Souza; Olivier Kobel; Philipp Jablonski; Ralph Schlapbach; Sarah Nadeau; Simon Grüter; Sophie Seidel; Tanja Stadler; Timothy Sykes |
| EPI_ISL_478249 | Virology Department, Royal Infirmary of Edinburgh, NHS Lothian / School of Biological Sciences, University of Edinburgh / Institute of Genetics and Molecular | COVID-19 Genomics UK (COG-UK) Consortium | Balcaza C; Colquhoun R; Dewar R; Gallagher M; Hill V; Jackson B; McCrone JT; McHugh M; O'Toole Á; Rambaut A; Rooke S; Scher E; Templeton K; Williams TC; Yu X |

| | | | |
|---|---|---|---|
| | Medicine, University of Edinburgh | | |
| EPI_ISL_1968604, EPI_ISL_2273882, EPI_ISL_2273938, EPI_ISL_2283150, EPI_ISL_2283162, EPI_ISL_2283229, EPI_ISL_2402835, EPI_ISL_2402881, EPI_ISL_2450673, EPI_ISL_2450676 | | | |
| see above | Weill Cornell Medicine | New York Genome Center | Andre Corvelo; Arryn Craney; Chris Mason; Dayna M. Oschwald; Hanna Rennert; Lars F Westblade; Margaret Elizabeth Ross; Melissa Cushing; Michael Zody; Olivier Elemento; Priya Velu; Samantha Fennessey; Tom Maniatis |
| EPI_ISL_461606, EPI_ISL_461678, EPI_ISL_473651, EPI_ISL_478094, EPI_ISL_493851, EPI_ISL_493868 | West of Scotland Specialist Virology Centre, NHSGGC / MRC-University of Glasgow Centre for Virus Research | COVID-19 Genomics UK (COG-UK) Consortium | Alasdair MacLean; Alice Broos; Ana da Silva Filipe; Antonia Ho; Daniel Mair; David L Robertson; Elihu Aranday-Cortes; Emma Thomson; James Shepherd; Jenna Nichols; Joseph Hughes; Kathy Li; Kathy Smollett; Kirstyn Brunker; Kyriaki Nomikou; Lily Tong; Marc Niebel; Natasha Jesudason; Natasha Johnson; Patawee Asamaphan; Rajiv Shah; Richard Orton; Rory Gunson; Sarah McDonald; Sreenu Vattipally; Stephen Carmichael; Yasmin Parr |
| EPI_ISL_803346 | Wisconsin State Laboratory of Hygiene Communicable Disease Division | Wisconsin State Laboratory of Hygiene Communicable Disease Division | Abigail C. Shockey; Kelsey R. Florek |
| EPI_ISL_486429 | unknown | Clinical Laboratory, Hospital Israelita Albert Einstein | Amgarten, D.; C.L. and Pinho; F.G.; Guedes; J.R.; Malta, F.; Mangueira; R.A.; R.L.; Santana; de Menezes |
| EPI_ISL_458140, EPI_ISL_458141, EPI_ISL_458146, EPI_ISL_458147, EPI_ISL_524783, EPI_ISL_524785, EPI_ISL_524786, EPI_ISL_524787, EPI_ISL_848562, EPI_ISL_848563, EPI_ISL_848565, EPI_ISL_848566, EPI_ISL_848571, EPI_ISL_848582, EPI_ISL_848583, EPI_ISL_848585, EPI_ISL_848587, EPI_ISL_848588, EPI_ISL_848589, EPI_ISL_848590, EPI_ISL_848592, EPI_ISL_848593, EPI_ISL_848595, EPI_ISL_848611, EPI_ISL_848615, EPI_ISL_848617, EPI_ISL_848618, EPI_ISL_848619, EPI_ISL_848620, EPI_ISL_848621, EPI_ISL_848622, EPI_ISL_848623, EPI_ISL_848624, EPI_ISL_848628, EPI_ISL_918518 | | | |
| see above | Evandro Chagas Institute | Evandro Chagas Institute | A.M.; Barbagelata; E.C.; E.M.A.; Ferreira; G.M.R; H.R; J.A.; Junior; K.C.; L.C.; L.S.; M.C.; Martins; P.S.; Pinheiro; Resque; Santos; Silva; Sousa; Sousa Junior; Viana; W.D.C.; da Silva |

We gratefully acknowledge the following Authors from the Originating laboratories responsible for obtaining the specimens, as well as the Submitting laboratories where the genome data were generated and shared via GISAID, on which this research is based.

All Submitters of data may be contacted directly via www.gisaid.org

Authors are sorted alphabetically.

| Accession ID | Originating Laboratory | Submitting Laboratory | Authors |
|---|---|---|---|
| EPI_ISL_4060359 | "OK Public Health Laboratory, Oklahoma State DOH" | Centers for Disease Control and Prevention Division of Viral Diseases, Pathogen Discovery | Alex Burgin; Ben Rambo-Martin; Clinton Paden; Dakota Howard; Dave Wentworth; Dhwani Batra; Jasmine Padilla; Justin Lee; Krista Queen; Kristen Knipe; Kristine Lacek; Mark Burroughs; Matthew Schmerer; Meghan Bentz; Mili Sheth; Peter Cook; Sam Shepard; Sarah Nobles; Suxiang Tong; Vivien Dugan; Yvette Unoarumhi |
| EPI_ISL_3813952 | "PA Department of Health, Bureau of Laboratories" | Centers for Disease Control and Prevention Division of Viral Diseases, Pathogen Discovery | Alex Burgin; Ben Rambo-Martin; Clinton Paden; Dakota Howard; Dave Wentworth; Dhwani Batra; Jasmine Padilla; Justin Lee; Krista Queen; Kristen Knipe; Kristine Lacek; Mark Burroughs; Matthew Schmerer; Meghan Bentz; Mili Sheth; Peter Cook; Sam Shepard; Sarah Nobles; Suxiang Tong; Vivien Dugan; Yvette Unoarumhi |
| EPI_ISL_3062381 | A.S.L. ROMA 2 - SISP | Department of General Diagnostics; Department of Virology; Istituto Zooprofilattico Sperimentale del Lazio e della Toscana (IZSLT) | Alessia Franco; Antonella Cersini; Antonio Battisti.; Elena L. Diaconu; Fabiola Feltrin; Giuseppe Manna; Patricia Alba; Raffaela Conti; Teresa Scicluna; Virginia Carfora |
| EPI_ISL_3638943 | AREA DE SALUD ACOSTA | Inciensa, Instituto Costarricense de Investigación y Enseñanza en Nutrición y Salud | Adriana Godínez; Claudio Soto-Garita; Estela Cordero; Francisco Duarte; Hebleen Porras; José Luis Vargas; Mariela Gutiérrez & Joselyn Prado; Melany Calderón |
| EPI_ISL_3638752 | AREA DE SALUD ALAJUELITA | Inciensa, Instituto Costarricense de Investigación y Enseñanza en Nutrición y Salud | Adriana Godínez; Claudio Soto-Garita; Estela Cordero; Francisco Duarte; Hebleen Porras; José Luis Vargas; Mariela Gutiérrez & Joselyn Prado; Melany Calderón |
| EPI_ISL_3638775, EPI_ISL_3948510 | AREA DE SALUD ASERRI | Inciensa, Instituto Costarricense de Investigación y Enseñanza en Nutrición y Salud | Adriana Godínez; Claudio Soto-Garita; Estela Cordero; Francisco Duarte; Hebleen Porras; José Luis Vargas; Mariela Gutiérrez & Joselyn Prado; Melany Calderón |
| EPI_ISL_3639056 | AREA DE SALUD BAGACES | Inciensa, Instituto Costarricense de Investigación y Enseñanza en Nutrición y Salud | Adriana Godínez; Claudio Soto-Garita; Estela Cordero; Francisco Duarte; Hebleen Porras; Joselyn Prado & Adriana Bermúdez; José Luis Vargas; Mariela Gutiérrez; Melany Calderón |
| EPI_ISL_3298361, EPI_ISL_3948509 | AREA DE SALUD CARTAGO | Inciensa, Instituto Costarricense de Investigación y Enseñanza en Nutrición y Salud | Adriana Godínez; Claudio Soto-Garita; Estela Cordero; Francisco Duarte; Hebleen Porras; Joselyn Prado & Monserrat Segura; Joselyn Prado & Moserrat Segura; José Luis Vargas; Mariela Gutiérrez; Melany Calderón |
| EPI_ISL_3948523 | AREA DE SALUD CAÑAS | Inciensa, Instituto Costarricense de Investigación y Enseñanza en Nutrición y Salud | Adriana Godínez; Claudio Soto-Garita; Estela Cordero; Francisco Duarte; Hebleen Porras; Joselyn Prado & Adriana Bermúdez; José Luis Vargas; Mariela Gutiérrez; Melany Calderón |
| EPI_ISL_3638917 | AREA DE SALUD GOICOECHEA 2 - CLINICA DR. JIMENEZ NUÑEZ | Inciensa, Instituto Costarricense de Investigación y Enseñanza en Nutrición y Salud | Adriana Godínez; Claudio Soto-Garita; Estela Cordero; Francisco Duarte; Hebleen Porras; Joselyn Prado & Juan Carlos Cartes; José Luis Vargas; Mariela Gutiérrez; Melany Calderón |
| EPI_ISL_3638970 | AREA DE SALUD GOLFITO | Inciensa, Instituto Costarricense de Investigación y Enseñanza en Nutrición y Salud | Adriana Godínez; Claudio Soto-Garita; Estela Cordero; Francisco Duarte; Hebleen Porras; Joselyn Prado & Eduardo Blanco; José Luis Vargas; Mariela Gutiérrez; Melany Calderón |
| EPI_ISL_3639111 | AREA DE SALUD GUATUSO | Inciensa, Instituto Costarricense de Investigación y Enseñanza en Nutrición y Salud | Adriana Godínez; Claudio Soto-Garita; Estela Cordero; Francisco Duarte; Hebleen Porras; Joselyn Prado & Francisco Chacón; José Luis Vargas; Mariela Gutiérrez; Melany Calderón |
| EPI_ISL_3948491 | AREA DE SALUD HATILLO - CLINICA DR. SOLON NUÑEZ | Inciensa, Instituto Costarricense de Investigación y Enseñanza en Nutrición y Salud | Adriana Godínez; Claudio Soto-Garita; Estela Cordero; Francisco Duarte; Hebleen Porras; Joselyn Prado & Roberto Brilla; José Luis Vargas; Mariela Gutiérrez; Melany Calderón |
| EPI_ISL_3948531 | AREA DE SALUD HEREDIA-VIRILLA | Inciensa, Instituto Costarricense de Investigación y Enseñanza en Nutrición y Salud | Adriana Godínez; Claudio Soto-Garita; Estela Cordero; Francisco Duarte; Hebleen Porras; José Luis Vargas; Mariela Gutiérrez & Joselyn Prado; Melany Calderón |
| EPI_ISL_3639114 | AREA DE SALUD LA UNION | Inciensa, Instituto Costarricense de Investigación y Enseñanza en Nutrición y Salud | Adriana Godínez; Claudio Soto-Garita; Estela Cordero; Francisco Duarte; Hebleen Porras; Joselyn Prado & Mónica Charpentier; José Luis Vargas; Mariela Gutiérrez; Melany Calderón |
| EPI_ISL_3464549, EPI_ISL_3948486 | AREA DE SALUD MATA REDONDA-HOSPITAL - CLINICA DR. MORENO CAÑAS | Inciensa, Instituto Costarricense de Investigación y Enseñanza en Nutrición y Salud | Adriana Godínez; Claudio Soto-Garita; Estela Cordero; Francisco Duarte; Hebleen Porras; Joselyn Prado & Ricardo González; Joselyn Prado & Ricardo González Cascante; José Luis Vargas; Mariela Gutiérrez; Melany Calderón |
| EPI_ISL_3638959 | AREA DE SALUD MONTES DE ORO | Inciensa, Instituto Costarricense de Investigación y Enseñanza en Nutrición y Salud | Adriana Godínez; Claudio Soto-Garita; Estela Cordero; Francisco Duarte; Hebleen Porras; Joselyn Prado & María José Gómez; José Luis Vargas; Mariela Gutiérrez; Melany Calderón |
| EPI_ISL_3639026, EPI_ISL_3948496, EPI_ISL_3948506 | AREA DE SALUD OREAMUNO-PACAYAS-TIERRA BLANCA | Inciensa, Instituto Costarricense de Investigación y Enseñanza en Nutrición y Salud | Adriana Godínez; Claudio Soto-Garita; Estela Cordero; Francisco Duarte; Hebleen Porras; Joselyn Prado & Carolina Loría; José Luis Vargas; Mariela Gutiérrez; Melany Calderón |
| EPI_ISL_3638947 | AREA DE SALUD PAQUERA | Inciensa, Instituto Costarricense de Investigación y Enseñanza en Nutrición y Salud | Adriana Godínez; Claudio Soto-Garita; Estela Cordero; Francisco Duarte; Hebleen Porras; Joselyn Prado & Ronald Núñez; José Luis Vargas; Mariela Gutiérrez; Melany Calderón |
| EPI_ISL_3639075 | AREA DE SALUD PAVAS (COOPESALUD) | Inciensa, Instituto Costarricense de Investigación y Enseñanza en Nutrición y Salud | Adriana Godínez; Claudio Soto-Garita; Estela Cordero; Francisco Duarte; Hebleen Porras; José Luis Vargas; Mariela Gutiérrez & Joselyn Prado; Melany Calderón |
| EPI_ISL_3639166 | AREA DE SALUD SANTA ANA (COOPESANA) | Inciensa, Instituto Costarricense de Investigación y Enseñanza en Nutrición y Salud | Adriana Godínez; Claudio Soto-Garita; Estela Cordero; Francisco Duarte; Hebleen Porras; José Luis Vargas; Mariela Gutiérrez & Joselyn Prado; Melany Calderón |
| EPI_ISL_3638874, EPI_ISL_3948483 | AREA DE SALUD SIQUIRRES | Inciensa, Instituto Costarricense de Investigación y Enseñanza en Nutrición y Salud | Adriana Godínez; Claudio Soto-Garita; Estela Cordero; Francisco Duarte; Hebleen Porras; José Luis Vargas; Mariela Gutiérrez & Joselyn Prado; Melany Calderón |
| EPI_ISL_3464548 | AREA DE SALUD TURRIALBA-JIMENEZ | Inciensa, Instituto Costarricense de Investigación y Enseñanza en Nutrición y Salud | Adriana Godínez; Claudio Soto-Garita; Estela Cordero; Francisco Duarte; Hebleen Porras; Joselyn Prado & Mónica Charpentier Artavia; José Luis Vargas; Mariela Gutiérrez; Melany Calderón |
| EPI_ISL_3126848 | ASST FBF Sacco | Laboratory of Clinical Microbiology, Virology and Bioemergencies, ASST Fatebenefratelli Sacco - Sacco Hospital | Valeria Micheli |
| EPI_ISL_3905332, EPI_ISL_3905334, EPI_ISL_3905349, EPI_ISL_3905355, EPI_ISL_3905359 | AUSTRAL-omics, UACh | AUSTRAL-omics, UACh | Andrea Silva; Carolina Encina; Cristian Molina; Daniela Plaza; Luis Guzmán; Suany Quesada |

EPI_ISL_2965605, EPI_ISL_2965612, EPI_ISL_2965614, EPI_ISL_2965618, EPI_ISL_2965622, EPI_ISL_2965629, EPI_ISL_3047420, EPI_ISL_3047481, EPI_ISL_3120265, EPI_ISL_3241830, EPI_ISL_3254986, EPI_ISL_3254988

| | | | |
|---|---|---|---|
| see above | AZDelta | AZ Delta Medical Laboratories in Roeselare, Belgium | Dieter De Smet; Frederik Van Hoecke; Geert Martens; Merijn Vanhee; on behalf of AZ Delta COVID-19 Genomics core (member of Genomic surveillance of SARS-CoV-2 in Belgium network) |

EPI_ISL_3218448, EPI_ISL_3219960, EPI_ISL_3220023, EPI_ISL_3220055, EPI_ISL_3220252, EPI_ISL_3220296, EPI_ISL_3305499, EPI_ISL_3321321, EPI_ISL_3321430, EPI_ISL_3321538, EPI_ISL_3321571, EPI_ISL_3321615, EPI_ISL_3321698, EPI_ISL_3321960, EPI_ISL_3321977, EPI_ISL_3322059, EPI_ISL_3322142, EPI_ISL_3322214, EPI_ISL_3322410, EPI_ISL_3322447, EPI_ISL_3322542, EPI_ISL_3322669, EPI_ISL_3322823, EPI_ISL_3322950, EPI_ISL_3323038, EPI_ISL_3323175, EPI_ISL_3323222, EPI_ISL_3323407, EPI_ISL_3323495, EPI_ISL_3323715, EPI_ISL_3323716, EPI_ISL_3323724, EPI_ISL_3323743, EPI_ISL_3323817, EPI_ISL_3323927, EPI_ISL_3324046, EPI_ISL_3324127, EPI_ISL_3324215, EPI_ISL_3324410, EPI_ISL_3324544, EPI_ISL_3324821, EPI_ISL_3324885, EPI_ISL_3325057, EPI_ISL_3325227,

| | | | |
|---|---|---|---|
| EPI_ISL_3430033, EPI_ISL_3430185, EPI_ISL_3430191, EPI_ISL_3430233, EPI_ISL_3430365, EPI_ISL_3430438, EPI_ISL_3430545, EPI_ISL_3430701, EPI_ISL_3430703, EPI_ISL_3812948, EPI_ISL_3813212, EPI_ISL_3813278, EPI_ISL_3813308, EPI_ISL_3813456, EPI_ISL_3813472, EPI_ISL_3814408, EPI_ISL_3815183, EPI_ISL_3815189, EPI_ISL_3815200, EPI_ISL_3815203, EPI_ISL_3815484, EPI_ISL_3816025, EPI_ISL_3816060, EPI_ISL_3816564, EPI_ISL_3816690, EPI_ISL_3816702, EPI_ISL_3816733, EPI_ISL_3818302, EPI_ISL_3818609, EPI_ISL_3818636, EPI_ISL_3818660, EPI_ISL_3818819, EPI_ISL_3818994, EPI_ISL_3819082, EPI_ISL_3819561, EPI_ISL_3819995, EPI_ISL_3821296, EPI_ISL_3821558, EPI_ISL_3821669, EPI_ISL_3822499, EPI_ISL_3822715, EPI_ISL_3822867, EPI_ISL_3842668, EPI_ISL_3842790, EPI_ISL_3843072, EPI_ISL_3843074, EPI_ISL_3843076, EPI_ISL_3843079, EPI_ISL_3843081, EPI_ISL_3843083, EPI_ISL_3843088, EPI_ISL_3844428, EPI_ISL_3844584, EPI_ISL_3845171, EPI_ISL_3845238, EPI_ISL_3845370, EPI_ISL_3845396, EPI_ISL_3845852, EPI_ISL_3846130, EPI_ISL_3846340, EPI_ISL_3846638, EPI_ISL_3847406, EPI_ISL_3847734, EPI_ISL_3848009, EPI_ISL_3848251, EPI_ISL_3849901, EPI_ISL_3849909, EPI_ISL_3851079, EPI_ISL_3851199, EPI_ISL_3851863, EPI_ISL_3852806, EPI_ISL_3853010, EPI_ISL_3853618, EPI_ISL_3853911, EPI_ISL_3855192, EPI_ISL_3855436, EPI_ISL_3856611, EPI_ISL_3856616, EPI_ISL_3856672, EPI_ISL_3858368, EPI_ISL_3858383, EPI_ISL_3859123, EPI_ISL_3859582, EPI_ISL_3861401, EPI_ISL_3862132, EPI_ISL_3862573, EPI_ISL_3864154, EPI_ISL_4057062, EPI_ISL_4059095, EPI_ISL_4059273 | | | |
| see above | Aegis Sciences Corporation | Centers for Disease Control and Prevention Division of Viral Diseases, Pathogen Discovery | Adrian Paskey; Alec Vest; Benjamin Rambo-Martin; Christopher Gulvick; Clinton Paden; Clinton R. Paden; Cyndi Clark; Dakota Howard; Darlene Wagner; Dhwani Batra; Dillon Nall; Duncan MacCannell; Ethan Sanders; Holly Houdeshell; Jason Caravas; Kara Moser; Matthew Hardison; Matthew Schmerer; Ola Kvalvaag; Patrick Campbell; Peter Cook; Peter W. Cook; Rob Case; Scott Sammons; Shatavia Morrison; Shaun Westlund; Vikramsinha Ghorpade; Yvette Unoarumhi |
| EPI_ISL_3388728 | Alverno Labs | RIPHL at Rush University Medical Center | Edith Perez; Felix Araujo Perez; Kevin Kunstman; Laura Furtado; Marieta Hyde; Max Kolton; Melissa Mace; Stefan Green |
| EPI_ISL_3089191, EPI_ISL_3089196, EPI_ISL_3089199, EPI_ISL_3089200, EPI_ISL_3089201, EPI_ISL_3089202, EPI_ISL_3089203, EPI_ISL_3090065, EPI_ISL_3216770, EPI_ISL_3217106, EPI_ISL_3217130, EPI_ISL_3217144, EPI_ISL_3262733, EPI_ISL_3262749, EPI_ISL_3262760, EPI_ISL_3319874 | | | |
| see above | Arizona State Public Health Laboratory | Arizona State Public Health Laboratory | Jessica Escobar; Katherine Fullerton; Linda Getsinger; Matthew Contursi; Nobuko Fukushima; Stacy White; Trung Huynh; Victor Waddell |
| EPI_ISL_3260474, EPI_ISL_3493438 | Arizona State University | Arizona State University | Ajeet Bains; Efrem S. Lim; Joshua LaBaer; LaRinda A. Holland; Matthew F. Smith; Nathaniel Johnson; Nicholas J. Mellor; Peter T. Skidmore; Rabia Maqsood; Vel Murugan |
| EPI_ISL_3553476 | Arkansas Children's Hospital | Center for Global Health, University of New Mexico Health Sciences Center | Ashton Ingold; Bobby Boyanton; Darrell Dinwiddie; Daryl Domman; Joshua L. Kennedy; Kurt Schwalm; Valerie Morley |
| EPI_ISL_3099255 | Arkansas Public Health Laboratory, Arkansas Department of Health | University of Minnesota Genomics Center | Corbin Dirkx; Daryl M. Gohl; Jaquelyn Kuriger-Laber; John Garbe |
| EPI_ISL_3307684, EPI_ISL_3797192 | Austrian Agency for Health and Food Safety (AGES) | Bergthaler laboratory, CeMM Research Center for Molecular Medicine of the Austrian Academy of Sciences | Andreas Bergthaler; Anna Schedl; Bekir Erguner; Benedikt Agerer; Christoph Bock; Fabian Amman; Jan Laine; Lukas Endler; Maelle Le Moing; Martin Senekowitsch; Matthew Thornton; Michael Schuster; Petr Triska; Thomas Penz |
| EPI_ISL_3912132 | BANCO DE OLHOS DO CEARA | Analytical Competence Molecular Epidemiology Lab/ACME, Oswaldo Cruz Foundation, Ceara (FIOCRUZ CE) | Cleber Furtado Aksenen; Fabio Miyajima; Fernando Braga Stehling; Francisco Eder de Moura Lopes; Jamille Maria Mendes Bezerra; Joaquim Cesar do Nascimento Sousa Junior; Pedro Miguel Carneiro Jeronimo; Suzana Porto Almeida & Lucas Delerino on behalf of COVID-19 FIOCRUZ Genomic Network; Thais Ferreira de Oliveira; Thais de Oliveira Costa; Ticiane Cavalcante de Souza; Veridiana Pessoa Miyajima |
| EPI_ISL_3579326 | BIOFAST | Instituto Butantan | Antonio Jorge Martins; Claudia Renata dos Santos Barros; David Schlesinger; Debora Botequio Moretti; Dimas Tadeu Covas; Elaine Cristina Marqueze; Elaine Vieira Santos; Evandra Strazza Rodrigues; Heidge Fukumasu; Jayme Augusto de Souza-Neto; José Salvatore Leister Patané; Luiz Alcantara; Luiz Lehmann Coutinho; Maria Carolina Elias; Maurício Lacerda Nogueira; Rafael dos Santos Bezerra; Raul Machado Neto; Rejane Maria Tommasini Grotto; Ricardo Haddad; Sandra Coccuzzo Sampaio Vessoni; Simone Kashima; Svetoslav Nanev Slavov; Vincent Louis Viala |
| EPI_ISL_3835033 | BOSTON HEART DIAGNOSTICS CORP | Wadsworth Center, New York State Department of Health | Alexis Russell; Catharine Prussing; Daryl M. Lamson; Erasmus Schneider; Erica Lasek-Nesselquist; John Kelly; Jonathan Plitnick; Kirsten St. George; Matthew Shudt; Melissa A Leisner; Navjot Singh |
| EPI_ISL_3087039, EPI_ISL_3087088, EPI_ISL_3214808 | Baylor Scott & White-Temple | Baylor Scott & White-Temple | Ari Rao; Kimberly Walker; Linden Morales; Marcus Volz; Shelby Johnson |
| EPI_ISL_3980440, EPI_ISL_3981426 | Biomédicos de Mérida | Biomédicos de Mérida | CANCHE-PECH JOSE REYES; CAUICH-VERA KRISTIAN; CEBALLOS-LOPEZ ADRIAN; CHIN-HAU LETICIA; GARCIA-GONZALEZ IGRID; LOPEZ-NOVELO MARIA E; MENDOZA-OLIVEROS TAHALI; SAURI-MEDINA EMILY |
| EPI_ISL_3084623, EPI_ISL_3084663, EPI_ISL_3084906, EPI_ISL_3155849, EPI_ISL_3407759, EPI_ISL_3407760, EPI_ISL_3408337, EPI_ISL_3431226 | | | |
| see above | Broad Institute Clinical Research Sequencing Platform | Infectious Disease Program, Broad Institute of Harvard and MIT | Adams, G.; B.L.; B.W.; Bauer, M.; Birren; Blumenstiel, B.; Brown, C.; Carter, A.; Chaluvadi, S.; D.J.; DeFelice, M.; DeRuff, K.; Dodge, S.; Gabriel, S.; Gallagher, G.; Gladden-Young, A.; Granger, B.; J.E.; K.J.; Lagerborg, K.; Larkin, K.; Lee, M.; Lemieux; Lennon, N.; Loreth, C.; Madoff, L.; McGovern, S.; Meldrim, J.; Normandin, E.; P.C.; Park; Pearlman, L.; Reilly, S.; Rudy, M.; Sabeti; Siddle; Smole, S.; Tomkins-Tinch, C.; Vicente, G.; and MacInnis |
| EPI_ISL_3723154 | CDPH VBL | California Department of Public Health | Emily Smith on behalf of CDPH-COVIDNet and UCI Genome Sciences Center/GHTF |
| EPI_ISL_3536165 | CENTRO DE ATENCAO ESPECIALIZADA | Analytical Competence Molecular Epidemiology Lab/ACME, Oswaldo Cruz Foundation, Ceara (FIOCRUZ CE) | Cleber Furtado Aksenen; Fabio Miyajima; Fernando Braga Stehling; Francisco Eder de Moura Lopes; Jamille Maria Mendes Bezerra; Joaquim César do Nascimento Sousa Junior; Pedro Miguel Carneiro Jeronimo; Suzana Porto Almeida e Lucas Delerino; Thais Ferreira de Oliveira; Thais de Oliveira Costa; Ticiane Cavalcante de Souza; Veridiana Pessoa Miyajima |
| EPI_ISL_3536353, EPI_ISL_3912134, EPI_ISL_3912135, EPI_ISL_3912136, EPI_ISL_3912137 | CENTRO DE ATENDIMENTO PARA ENFRENTAMENTO AO COVID 19 | Analytical Competence Molecular Epidemiology Lab/ACME, Oswaldo Cruz Foundation, Ceara (FIOCRUZ CE) | Cleber Furtado Aksenen; Fabio Miyajima; Fernando Braga Stehling; Francisco Eder de Moura Lopes; Jamille Maria Mendes Bezerra; Joaquim Cesar do Nascimento Sousa Junior; Joaquim César do Nascimento Sousa Junior; Pedro Miguel Carneiro Jeronimo; Suzana Porto Almeida & Lucas Delerino on behalf of COVID-19 FIOCRUZ Genomic Network; Suzana Porto Almeida e Lucas Delerino; Thais Ferreira de Oliveira; Thais de Oliveira Costa; Ticiane Cavalcante de Souza; Veridiana Pessoa Miyajima |
| EPI_ISL_3696640, EPI_ISL_3696644, EPI_ISL_3696647, EPI_ISL_3696659, EPI_ISL_3696668, EPI_ISL_3696752, EPI_ISL_3696769, EPI_ISL_3696788, EPI_ISL_3696808 | | | |
| see above | CENTRO DE ENFRENTAMENTO COVID | Instituto Butantan | Antonio Jorge Martins; Claudia Renata dos Santos Barros; David Schlesinger; Debora Botequio Moretti; Dimas Tadeu Covas; Elaine Cristina Marqueze; Elaine Vieira Santos; Evandra Strazza Rodrigues; Heidge Fukumasu; Jayme Augusto de Souza-Neto; José Salvatore Leister Patané; Luiz Alcantara; Luiz Lehmann Coutinho; Maria Carolina Elias; Maurício Lacerda Nogueira; Rafael dos Santos Bezerra; Raul Machado Neto; Rejane Maria Tommasini Grotto; Ricardo Haddad; Sandra Coccuzzo Sampaio Vessoni; Simone Kashima; Svetoslav Nanev Slavov; Vincent Louis Viala |
| EPI_ISL_3922299 | CENTRO DE SAUDE II DR GABRIEL MESQUITA VARGEM GDE DO SUL | Instituto Butantan | Antonio Jorge Martins; Claudia Renata dos Santos Barros; David Schlesinger; Debora Botequio Moretti; Dimas Tadeu Covas; Elaine Cristina Marqueze; Elaine Vieira Santos; Evandra Strazza Rodrigues; Heidge Fukumasu; Jayme Augusto de Souza-Neto; José Salvatore Leister Patané; Luiz Alcantara; Luiz Lehmann Coutinho; Maria Carolina Elias; Maurício Lacerda Nogueira; Rafael dos Santos Bezerra; Raul Machado Neto; Rejane Maria Tommasini Grotto; Ricardo Haddad; Sandra Coccuzzo Sampaio Vessoni; Simone Kashima; Svetoslav Nanev Slavov; Vincent Louis Viala |
| EPI_ISL_3131368 | CERBALLIANCE MIDI-PYRENEES | CNR Virus des Infections Respiratoires - France SUD | Antonin Bal; Bruno Lina; Gregory Destras; Gwendolyne Burfin; Hadrien Regue; Laurence Josset; Martine Valette; Quentin Semanas |
| EPI_ISL_3269615, EPI_ISL_3269616, EPI_ISL_3269617, EPI_ISL_3269618, EPI_ISL_3398100, EPI_ISL_3547308, EPI_ISL_3547309, EPI_ISL_3547311 | | | |
| see above | CH de l'Ouest Guyanais Frank Joly | Institut Pasteur de la Guyane | Anne Lavergne; Antoine Enfissi; Arielle Salmier; Dominique Rousset |
| EPI_ISL_3190118, EPI_ISL_3190121, EPI_ISL_3269602, EPI_ISL_3269603, EPI_ISL_3269604, EPI_ISL_3269605, EPI_ISL_3269612, EPI_ISL_3269613, EPI_ISL_3277483, EPI_ISL_3370958, EPI_ISL_3370970, EPI_ISL_3370973, EPI_ISL_3398115, EPI_ISL_3398116, EPI_ISL_3501660 | | | |
| see above | CHC Andrée Rosemon | Institut Pasteur de la Guyane | Anne Lavergne; Antoine Enfissi; Arielle Salmier; Dominique Rousset |
| EPI_ISL_3758167, EPI_ISL_3758168, EPI_ISL_3758169, EPI_ISL_3758171, EPI_ISL_3758178, EPI_ISL_3758179, EPI_ISL_3758181, EPI_ISL_3758184, EPI_ISL_3758185, EPI_ISL_3758188, EPI_ISL_3758189, EPI_ISL_3758191, EPI_ISL_3758192, EPI_ISL_3758194, EPI_ISL_3825455, EPI_ISL_3825456, EPI_ISL_3825458, EPI_ISL_3825460, EPI_ISL_3825462, EPI_ISL_3825469, EPI_ISL_4061622, EPI_ISL_4061624, EPI_ISL_4061634, EPI_ISL_4061641, EPI_ISL_4061644, EPI_ISL_4061645 | | | |
| see above | CHECK-UP - Medicina e Diagnóstico - Matriz | HLAGYN - Laboratorio de Imunologia de Transplantes de Goias | Elaize Maria Gomes de Paula; Fernando Antonio Vinhal dos Santos; Frederico Rodrigues Vinhal; Gladstone Rodrigues da Cunha Filho; Kamila Oliveira Reis De Freitas; Lucas Carlos Gomes Pereira; Nubia Silva Araújo; Sabrina Sara Moreira Duarte |
| EPI_ISL_3804696, EPI_ISL_3838197, EPI_ISL_3866870, EPI_ISL_3866877 | CHU NIMES | CHU NIMES | Agathe Boudet; Marie-Josée Carles; Sophie Bravo; Stephan Robin |
| EPI_ISL_3234582, EPI_ISL_3471631 | CHU Purpan - Laboratoire de Virologie - Institut Fédératif de Biologie | CHU Purpan - Laboratoire de Virologie - Institut Fédératif de Biologie | Bulach T.; Donnadieu C.; Izopet J.; Latour J.; Milhes M.; Nicot F.; Ranger N.; Salin G.; Tremeaux P. |
| EPI_ISL_3131351 | CHU TOULOUSE | CNR Virus des Infections Respiratoires - France SUD | Antonin Bal; Bruno Lina; Gregory Destras; Gwendolyne Burfin; Hadrien Regue; Laurence Josset; Martine Valette; Quentin Semanas |
| EPI_ISL_3267802, EPI_ISL_3267825 | CHUV | Laboratory of genomics and metagenomics | Claire Bertelli; Damien Jacot; Gilbert Greub; Sébastien Aeby; Trestan Pillonel |
| EPI_ISL_3387077 | CLILAB | Microbiology Department | Aida Gonzalez-Diaz; Carmen Ardanuy; Jordi Camara; Jordi Niubó; Laura Calatayud; M Angeles Domínguez; Sara Marti; Yolanda Hernandez |
| EPI_ISL_3129963, EPI_ISL_3129965, EPI_ISL_3190132, EPI_ISL_3190133, EPI_ISL_3269608, EPI_ISL_3277486, EPI_ISL_3277487, EPI_ISL_3277488, EPI_ISL_3277489, EPI_ISL_3277492, EPI_ISL_3277493, EPI_ISL_3277494, EPI_ISL_3277496, EPI_ISL_3370962, EPI_ISL_3398104, EPI_ISL_3398107, EPI_ISL_3547310, EPI_ISL_3547312 | | | |
| see above | CNR Institut Pasteur de la Guyane | Institut Pasteur de la Guyane | Anne Lavergne; Antoine Enfissi; Arielle Salmier; Dominique Rousset |
| EPI_ISL_3131720 | CNR Virus des Infections Respiratoires - France SUD | CNR Virus des Infections Respiratoires - France SUD | Antonin Bal; Bruno Lina; Gregory Destras; Gwendolyne Burfin; Hadrien Regue; Laurence Josset; Martine Valette; Quentin Semanas |
| EPI_ISL_3570212, EPI_ISL_3570218 | Center for Laboratory Medicine | Center for Laboratory Medicine | Yannick Gerth |
| EPI_ISL_3505620, EPI_ISL_3505630 | Center for Research on Health in Latin America | Institute of Microbiology, Universidad San Francisco de Quito | Belén Prado-Vivar; Bernardo Gutiérrez; Erika Muñoz; Fernanda Zurita; Gabriel Trueba; Jaime Costales; Juan José Guadalupe; Michelle Grunauer; Monica Becerra-Wong; Patricio Rojas-Silva; Paúl Cárdenas; Sully Márquez; Verónica Barragán |

| | | | |
|---|---|---|---|
| EPI_ISL_3539226, EPI_ISL_3539227, EPI_ISL_3539228, EPI_ISL_3802961 | Central Laboratory of Public Health of Bahia State (LACEN/BA) | Laboratory of Respiratory Viruses and Measles, Oswaldo Cruz Institute, FIOCRUZ | Agatha Soares; Alice Sampaio Rocha; Ana Carolina Mendonca; Anna Carolina Paixao; Elisa Cavalcante Pereira; Felicidade Pereira; Fernando Motta; Ighor Arantes; Luciana Appolinario; Marilda Siqueira on behalf of the Fiocruz COVID-19 Genomic Surveillance Network; Paola Resende; Renata Serrano Lopes; Taina Venas |
| EPI_ISL_3462621, EPI_ISL_3462696, EPI_ISL_3462697, EPI_ISL_3462698, EPI_ISL_3462699, EPI_ISL_3462700, EPI_ISL_3462701, EPI_ISL_3462702, EPI_ISL_3462703, EPI_ISL_3462704, EPI_ISL_3462705, EPI_ISL_3462706, EPI_ISL_3462707, EPI_ISL_3462708, EPI_ISL_3462709, EPI_ISL_3462710, EPI_ISL_3462711, EPI_ISL_3462712, EPI_ISL_3462713, EPI_ISL_3462714, EPI_ISL_3462715, EPI_ISL_3462716, EPI_ISL_3462717, EPI_ISL_3462718, EPI_ISL_3462719, EPI_ISL_3462720, EPI_ISL_3462721, EPI_ISL_3462722, EPI_ISL_3462723, EPI_ISL_3462724, EPI_ISL_3462725, EPI_ISL_3462726, EPI_ISL_3462729, EPI_ISL_3462730, EPI_ISL_3462731, EPI_ISL_3464734, EPI_ISL_4003116, EPI_ISL_4003120, EPI_ISL_4003121, EPI_ISL_4003122, EPI_ISL_4003123, EPI_ISL_4003125, EPI_ISL_4003126, EPI_ISL_4003127, EPI_ISL_4003129, EPI_ISL_4003132, EPI_ISL_4003135, EPI_ISL_4003137, EPI_ISL_4003141, EPI_ISL_4003142, EPI_ISL_4003143, EPI_ISL_4003144, EPI_ISL_4003145, EPI_ISL_4003146, EPI_ISL_4003147, EPI_ISL_4003148, EPI_ISL_4003151, EPI_ISL_4003152, EPI_ISL_4003153, EPI_ISL_4003155, EPI_ISL_4003156 | | | |
| see above | Central Laboratory, Bureau of Public Health (BOG) and Academic Hospital Paramaribo | Erasmus Medical Center | Bas B Oude Munnink; Cherise Beek; Consuella Partowidjojo; Dion Gajadin; Ed PF IJzerman; Emmanuelle Munger; Gary Gummels; Ingrid SK Krishnadath; Lycke Woittiez; Marion PG Koopmans; Mireille Van de Veer; Phyllis Pinas; Princes Wongsowidjojo; Radjesh Ori; Ranisha Doerbalie; Rohma Banwari; Soeradj Harkisoen; Stephen Vreden; Tilotmadebie Ramlal; Verne Nanhoe |
| EPI_ISL_3635659, EPI_ISL_3635660, EPI_ISL_3635661, EPI_ISL_3635662, EPI_ISL_3635663, EPI_ISL_3635664, EPI_ISL_3635665, EPI_ISL_3635666, EPI_ISL_3635667, EPI_ISL_3635668, EPI_ISL_3635669, EPI_ISL_3635670, EPI_ISL_3635671, EPI_ISL_3635672, EPI_ISL_3635673, EPI_ISL_3635674, EPI_ISL_3944572, EPI_ISL_3944574, EPI_ISL_3944577, EPI_ISL_3944578, EPI_ISL_3944579, EPI_ISL_3944580, EPI_ISL_3944581, EPI_ISL_3944582, EPI_ISL_3944583, EPI_ISL_3944585, EPI_ISL_3944586, EPI_ISL_3944587, EPI_ISL_3944588, EPI_ISL_3944589, EPI_ISL_3944590, EPI_ISL_3944591, EPI_ISL_3944594, EPI_ISL_3944595, EPI_ISL_3944596, EPI_ISL_3944597, EPI_ISL_3944598, EPI_ISL_3944599, EPI_ISL_3944600, EPI_ISL_3944601, EPI_ISL_3944602, EPI_ISL_3944603, EPI_ISL_3944604, EPI_ISL_3944605, EPI_ISL_3944606, EPI_ISL_3944607, EPI_ISL_3944608, EPI_ISL_3944609, EPI_ISL_3944610, EPI_ISL_3944611, EPI_ISL_3944612, EPI_ISL_3944613, EPI_ISL_3944614, EPI_ISL_3944616, EPI_ISL_3944617, EPI_ISL_3944618, EPI_ISL_3944619, EPI_ISL_3944620, EPI_ISL_3944621, EPI_ISL_3944622, EPI_ISL_3944623, EPI_ISL_3944624, EPI_ISL_3944625, EPI_ISL_3944627 | | | |
| see above | Central Public Health Laboratory - LACEN - Bahia, Salvador, Brazil | Central Public Health Laboratory - LACEN -Bahia, Salvador, Brazil | Arabela Leal; Felicidade Pereira; Gabriela Menezes; Jaqueline Gomes; Lenisa Dandara; Luciana Oliveira; Luiz Alcantara; Marcela Gómez; Marta Giovanetti; Stephane Tosta; Vagner Fonseca; Vanessa Nardy |
| EPI_ISL_3129967, EPI_ISL_3129969, EPI_ISL_3129970 | Centre Hospitalier de l'Ouest Guyanais | Institut Pasteur de la Guyane | Anne Lavergne; Antoine Enfissi; Arielle Salmier; Dominique Rousset |
| EPI_ISL_3805729 | Centro de Investigacion Biomedica del Noreste (CIBIN) | Unidad de Genomica Avanzada | ; Alejandra Garcia-Gasca; Alejandra Hernandez-Teran; Alejandro Sanchez-Flores; Alfredo Herrera-Estrella; Alicia Ocaña-Mondragon; Andreu Comas-Garcia; Angel Gustavo Salas-Lais; Antonio Loza Roman; Bernardo Martinez-Miguel; Blanca Taboada; Brenda Irasema Maldonado-Meza; Bruno Gomez-Gil; Carla Ivon Herrera-Najera; Carlos F. Arias; Celia Boukadida; Celida Duque Molina; Celida Martinez- Rodriguez; Clara Esperanza Santacruz-Tinoco; Concepcion Grajales-Muñiz; Consorcio Mexicano de Vigilancia Genomica (CoViGen-Mex). Authors (in alphabetical order): Julio Elias Alvarado-Yaah; Cristobal Chaidez-Quiroz; Daniel Fregoso-Rueda; Daniel Lira Morales; Eduardo Becerril-Vargas; Fernando Fontove-Herrera; Fidencio Mejia-Nepomuceno; Francisco Pulido; Gloria Elena Espinosa-Ayala; Gloria Maria Molina-Salinas; Gloria Vazquez; Hector Esteban Paz-Juarez; Hector Montoya-Fuentes; Helen Haydee Fernanda Ramirez-Plascencia; Irvin Gonzalez-Lopez; Jean Pierre Gonzalez; Jesus Hernandez; Joel Armando Vazquez-Perez.; Jorge Salas-Hernandez; Jose Antonio Enciso-Moreno; Jose Arturo Martinez-Orozco; Jose Esteban Muñoz-Medina; Jose de Jesus Nuñez-Contreras; Juan Bautista Chale-Dzul; Julissa Enciso-Ibarra; Luis Alberto Ochoa-Carrera; Margarita Matias-Florentino; Maria Guadalupe Santiago-Mauricio; Maria Guadalupe de Jesus Mireles-Rivera; Mario Mujica-Sanchez; Marissa Perez-Garcia; Nelly Selem-Mojica; Pavel Isa; Ricardo Ciria Merce; Ricardo Grande; Rosa Maria Gutierrez Rios; Santiago avila-Rios; Selene Zarate; Susana Lopez; Veronica Mata-Haro; Victor Eduardo Garcia-Arias; Victor Hugo Borja-Aburto |
| EPI_ISL_3557185, EPI_ISL_3557225 | Centro de Investigación Biomédica de Occidente (CIBO) | Centro de Investigación en Enfermedades Infecciosas (CIENI), Instituto Nacional de Enfermedades Respiratorias (INER) | Alejandra García-Gasca; Alejandra Hernández-Terán; Alejandro Sánchez-Flores; Alfredo Herrera-Estrella; Alicia Ocaña-Mondragón; Andreu Comas-García; Angel Gustavo Salas-Lais; Antonio Loza Román; Bernardo Martínez-Miguel; Blanca Taboada; Brenda Irasema Maldonado-Meza; Bruno Gómez-Gil; Carla Ivón Herrera-Najera; Carlos F. Arias; Celia Boukadida; Clara Esperanza Santacruz-Tinoco; Concepción Grajales-Muñiz; Consorcio Mexicano de Vigilancia Genómica (CoViGen-Mex). Authors (in alphabetical order): Julio Elias Alvarado-Yaah; Cristóbal Cháidez-Quiróz; Célida Duque Molina; Célida Martínez- Rodríguez; Daniel Fregoso-Rueda; Daniel Lira Morales; Eduardo Becerril-Vargas; Fernando Fontove-Herrera; Fidencio Mejía-Nepomuceno; Francisco Pulido; Gloria Elena Espinosa-Ayala; Gloria María Molina-Salinas; Gloria Vazquez; Hector Esteban Paz-Juárez; Hector Montoya-Fuentes; Helen Haydee Fernanda Ramírez-Plascencia; Irvin González-López; Jean Pierre González; Jesús Hernández; Joel Armando Vázquez-Pérez.; Jorge Salas-Hernández; José Antonio Enciso-Moreno; José Arturo Martínez-Orozco; José Esteban Muñoz-Medina; José de Jesús Nuñez-Contreras; Juan Bautista Chale-Dzul; Julissa Enciso-Ibarra; Kathia Elizabeth Tapia-Diaz; Luis Alberto Ochoa-Carrera; Margarita Matías-Florentino; Mario Mújica-Sánchez; Marissa Perez-Garcia; María Guadalupe Santiago-Mauricio; María Guadalupe de Jesús Mireles-Rivera; Nelly Sélem-Mojica; Pavel Isa; Ricardo Ciria Merce; Ricardo Grande; Rosa María Gutiérrez Rios; Santiago Ávila-Ríos; Selene Zárate; Susana Lopez; Verónica Mata-Haro; Victor Eduardo García-Arias; Víctor Hugo Borja-Aburto |
| EPI_ISL_3347554 | Centro de Investigación Biomédica de Occidente (CIBO) | Instituto de Biotecnología de la UNAM | ; Alejandra García-Gasca; Alejandra Hernández-Terán; Alejandro Sánchez-Flores; Alfredo Herrera-Estrella; Alicia Ocaña-Mondragón; Andreu Comas-García; Angel Gustavo Salas-Lais; Antonio Loza Román; Bernardo Martínez-Miguel; Blanca Taboada; Brenda Irasema Maldonado-Meza; Bruno Gómez-Gil; Carla Ivón Herrera-Najera; Carlos F. Arias; Celia Boukadida; Clara Esperanza Santacruz-Tinoco; Concepción Grajales-Muñiz; Consorcio Mexicano de Vigilancia Genómica (CoViGen-Mex). Authors (in alphabetical order): Julio Elias Alvarado-Yaah; Cristóbal Cháidez-Quiróz; Célida Duque Molina; Célida Martínez- Rodríguez; Daniel Fregoso-Rueda; Daniel Lira Morales; Eduardo Becerril-Vargas; Fernando Fontove-Herrera; Fidencio Mejía-Nepomuceno; Francisco Pulido; Gloria Elena Espinosa-Ayala; Gloria María Molina-Salinas; Gloria Vazquez; Hector Esteban Paz-Juárez; Hector Montoya-Fuentes; Helen Haydee Fernanda Ramírez-Plascencia; Irvin González-López; Jean Pierre González; Jesús Hernández; Joel Armando Vázquez-Pérez).; Jorge Salas-Hernández; José Antonio Enciso-Moreno; José Arturo Martínez-Orozco; José Esteban Muñoz-Medina; José de Jesús Nuñez-Contreras; Juan Bautista Chale-Dzul; Julissa Enciso-Ibarra; Kathia Elizabeth Tapia-Diaz; Luis Alberto Ochoa-Carrera; Margarita Matías-Florentino; Mario Mújica-Sánchez; Marissa Perez-Garcia; María Guadalupe Santiago-Mauricio; María Guadalupe de Jesús Mireles-Rivera; Nelly Sélem-Mojica; Pavel Isa; Ricardo Ciria Merce; Ricardo Grande; Rosa María Gutiérrez Rios; Santiago Ávila-Ríos; Selene Zárate; Susana Lopez; Verónica Mata-Haro; Victor Eduardo García-Arias; Víctor Hugo Borja-Aburto |
| EPI_ISL_3556954 | Centro de Investigación Biomédica del Noreste (CIBIN) | Centro de Investigación en Enfermedades Infecciosas (CIENI), Instituto Nacional de Enfermedades Respiratorias (INER) | Alejandra García-Gasca; Alejandra Hernández-Terán; Alejandro Sánchez-Flores; Alfredo Herrera-Estrella; Alicia Ocaña-Mondragón; Andreu Comas-García; Angel Gustavo Salas-Lais; Antonio Loza Román; Bernardo Martínez-Miguel; Blanca Taboada; Brenda Irasema Maldonado-Meza; Bruno Gómez-Gil; Carla Ivón Herrera-Najera; Carlos F. Arias; Celia Boukadida; Clara Esperanza Santacruz-Tinoco; Concepción Grajales-Muñiz; Consorcio Mexicano de Vigilancia Genómica (CoViGen-Mex). Authors (in alphabetical order): Julio Elias Alvarado-Yaah; Cristóbal Cháidez-Quiróz; Célida Duque Molina; Célida Martínez- Rodríguez; Daniel Fregoso-Rueda; Daniel Lira Morales; Eduardo Becerril-Vargas; Fernando Fontove-Herrera; Fidencio Mejía-Nepomuceno; Francisco Pulido; Gloria Elena Espinosa-Ayala; Gloria María Molina-Salinas; Gloria Vazquez; Hector Esteban Paz-Juárez; Hector Montoya-Fuentes; Helen Haydee Fernanda Ramírez-Plascencia; Irvin González-López; Jean Pierre González; Jesús Hernández; Joel Armando Vázquez-Pérez.; Jorge Salas-Hernández; José Antonio Enciso-Moreno; José Arturo Martínez-Orozco; José Esteban Muñoz-Medina; José de Jesús Nuñez-Contreras; Juan Bautista Chale-Dzul; Julissa Enciso-Ibarra; Kathia Elizabeth Tapia-Diaz; Luis Alberto Ochoa-Carrera; Margarita Matías-Florentino; Mario Mújica-Sánchez; Marissa Perez-Garcia; María Guadalupe Santiago-Mauricio; María Guadalupe de Jesús Mireles-Rivera; Nelly Sélem-Mojica; Pavel Isa; Ricardo Ciria Merce; Ricardo Grande; Rosa María Gutiérrez Rios; Santiago Ávila-Ríos; Selene Zárate; Susana Lopez; Verónica Mata-Haro; Victor Eduardo García-Arias; Víctor Hugo Borja-Aburto |
| EPI_ISL_3347532 | Centro de Investigación Biomédica del Noreste (CIBIN) | Instituto de Biotecnología de la UNAM | ; Alejandra García-Gasca; Alejandra Hernández-Terán; Alejandro Sánchez-Flores; Alfredo Herrera-Estrella; Alicia Ocaña-Mondragón; Andreu Comas-García; Angel Gustavo Salas-Lais; Antonio Loza Román; Bernardo Martínez-Miguel; Blanca Taboada; Brenda Irasema Maldonado-Meza; Bruno Gómez-Gil; Carla Ivón Herrera-Najera; Carlos F. Arias; Celia Boukadida; Clara Esperanza Santacruz-Tinoco; Concepción Grajales-Muñiz; Consorcio Mexicano de Vigilancia Genómica (CoViGen-Mex). Authors (in alphabetical order): Julio Elias Alvarado-Yaah; Cristóbal Cháidez-Quiróz; Célida Duque Molina; Célida Martínez- Rodríguez; Daniel Fregoso-Rueda; Daniel Lira Morales; Eduardo Becerril-Vargas; Fernando Fontove-Herrera; Fidencio Mejía-Nepomuceno; Francisco Pulido; Gloria Elena Espinosa-Ayala; Gloria María Molina-Salinas; Gloria Vazquez; Hector Esteban Paz-Juárez; Hector Montoya-Fuentes; Helen Haydee Fernanda Ramírez-Plascencia; Irvin González-López; Jean Pierre González; Jesús Hernández; Joel Armando Vázquez-Pérez).; Jorge Salas-Hernández; José Antonio Enciso-Moreno; José Arturo Martínez-Orozco; José Esteban Muñoz-Medina; José de Jesús Nuñez-Contreras; Juan Bautista Chale-Dzul; Julissa Enciso-Ibarra; Kathia Elizabeth Tapia-Diaz; Luis Alberto Ochoa-Carrera; Margarita Matías-Florentino; Mario Mújica-Sánchez; Marissa Perez-Garcia; María Guadalupe Santiago-Mauricio; María Guadalupe de Jesús Mireles-Rivera; Nelly Sélem-Mojica; Pavel Isa; Ricardo Ciria Merce; Ricardo Grande; Rosa María Gutiérrez Rios; Santiago Ávila-Ríos; Selene Zárate; Susana Lopez; Verónica Mata-Haro; Victor Eduardo García-Arias; Víctor Hugo Borja-Aburto |
| EPI_ISL_3983576 | City of Milwaukee Health Department Laboratory | City of Milwaukee Health Department Laboratory | Amy Bauer; Manjeet Khubbar; Nandu Balakrishnan; Samantha Scott; Sanjib Bhattacharyya |
| EPI_ISL_3076828, EPI_ISL_3076837 | Clinical Microbiology, Infection Prevention and Control | Section for Molecular Diagnostics | Björn Hallström; Jonas Björkman |
| EPI_ISL_3092377 | Cotugno | TIGEM | Antonio Grimaldi Patrizia Annunziata Francesco Panariello Biancamaria Pierri Claudia Tiberio Teresa Giuliano Valentina Bouche Chiara Colantuono Maria Concetta Cuomo Denise Di Concilio Lucio Di Filippo Anna Manfredi Marcello Salvi Antonio Limone Luigi Atripaldi Pellegrino Cerino Andrea Ballabio Davide Cacchiarelli |
| EPI_ISL_3920461 | DC Public Health Lab/ Dept. of Forensic Sciences | DC Public Health Lab/ Dept. of Forensic Sciences | Brittany Hamilton; Connie Maza; Elizabeth Zelaya; Janis Doss; Jocelyn Hauser; Monica Mann; Nathan Bruns; Sarah Scott; Scott Nguyen |
| EPI_ISL_3841265, EPI_ISL_3841266, EPI_ISL_3841267, EPI_ISL_3841268, EPI_ISL_3841269, EPI_ISL_3841279, EPI_ISL_3841300, EPI_ISL_3841301, EPI_ISL_3841302, EPI_ISL_3841303, EPI_ISL_3841304, EPI_ISL_3841309, EPI_ISL_3841310 | | | |
| see above | DNAGYN | LGBio (Laboratorio de Genetica & Biodiversidade) | Alex Honda Bernardes; Amanda Alves de Melo; Aparecido Divino da Cruz; Cintia Pelegrineti Targueta de Azevedo Brito; Daniela de Melo e Silva; Elisangela de Paula Silveira Lacerda; Juliana Santana de Curcio; Luiz Augusto Pereira; Marc Alexandre Duarte Gigonzac; Mariana Pires de Campos Telles; Ramilla dos Santos Braga; Renata de Oliveira Dias; Rhewter Nunes; Thais Cidália Vieira Gigonzac; Thais Guimarães Castro; Thays Millena Alves Pedroso |
| EPI_ISL_3692711 | DPHL | Delaware Public Health Lab | Rebecca Savage |
| EPI_ISL_3644513, EPI_ISL_4093606, EPI_ISL_4093614 | Department of Virology and Immunology, University of Helsinki and Helsinki University Hospital, Huslab Finland | Department of Virology, Faculty of Medicine, University of Helsinki, Helsinki, Finland | Essi Korhonen; Hanna Jarva; Hanna Liimatainen; Hannimari Kallio-Kokko; Harri Kangas; Hussein Alburkat; Jenni Virtanen; Maija Lappalainen; Maija Suvanto; Olli Vapalahti; Pekka Ellonen; Phuoc Truong; Ravi Kant; Sari Hannula; Satu Kurkela; Teemu Smura |
| EPI_ISL_3160431, EPI_ISL_3404737 | Dianalabs SA | Genesupport | Geraldine Jost; Katia Jaton; Nadia Liassine; Tanguy ARAUD |
| EPI_ISL_3372586, EPI_ISL_3372600, EPI_ISL_3372604 | Dirección regional de salud del Callao (DIRESA-CALLAO) | Centro de Investigaciones Tecnológicas, Biomédicas y Medioambientales (CITBM) | B; Huaman; J. Alarcon; M. Cuellar; M. Ramirez; M. Sovero |
| EPI_ISL_3369223, EPI_ISL_3369226, EPI_ISL_3369229 | Division of Emerging Infectious Diseases, Bureau of Infectious Diseases Diagnosis Control, Korea Disease Control and Prevention Agency | Division of Emerging Infectious Diseases, Bureau of Infectious Diseases Diagnosis Control, Korea Disease Control and Prevention Agency | Ae Kyung Park; Chae Young Lee; Eun-Jin Kim; Heui Man Kim; Il-Hwan Kim; Jeong-Ah Kim |
| EPI_ISL_4076118, EPI_ISL_4076448, EPI_ISL_4077052 | Dutch COVID-19 response team | National Institute for Public Health and the Environment (RIVM) | Adam Meijer; AnneMarie van den Brandt; Annelies Kroneman; Bas van der Veer; Chantal Reusken; Dennis Schmitz; Dirk Eggink; Euníce Then; Florian Zwagemaker; Harry Vennema; Ivo van Walle; Jeroen Cremer; Karim Hajji; Kim Freriks; Lisa Wijsman; Lynn Aarts; Ryanne Jaarsma; Sanne Bos; Sharon van den Brink; Stijn van Rossum; on behalf of the national COVID-19 response team |
| EPI_ISL_3459623 | EL CAMINO HOSPITAL | Santa Clara County Public Health Laboratory | Santa Clara County Public Health Department |
| EPI_ISL_3692162, EPI_ISL_3692329, EPI_ISL_3937038, EPI_ISL_3937241 | EXCITE Lab | Andersen lab at Scripps Research | Abigail Schnapper; Angela Scioscia; Art Mendoza; Cathy Woerle; Chip Schooley; David Pride; Helena Tubb; Jacquelyn Berumen; Liam McGinnis; Natasha Martin Cheryl Anderson; Omid Bakhtar; SEARCH Alliance San Diego with Aaron Harding; Sawyer Farmer; Sharon Reed; Tommy Valles + SEARCH |
| EPI_ISL_4054617 | Emory Molecular Diagnostics Laboratory, Emory Healthcare | Piantadosi Lab, Emory Department of Pathology | Ahmed Babiker; Anne Piantadosi; Ludy Registre Carmola |
| EPI_ISL_3902824, EPI_ISL_3903286, EPI_ISL_3903287, EPI_ISL_3903289, EPI_ISL_3920890, EPI_ISL_3920891 | Florida Bureau of Public Health Laboratories | Florida Bureau of Public Health Laboratories | Jason Blanton; Jiaqi Li; Namratha Tarigopula; Sarah Schmedes |

| | | | |
|---|---|---|---|
| EPI_ISL_3064782, EPI_ISL_3064893, EPI_ISL_3064897, EPI_ISL_3064963, EPI_ISL_3208534, EPI_ISL_3209081, EPI_ISL_3209621, EPI_ISL_3210043, EPI_ISL_3211689, EPI_ISL_3211976, EPI_ISL_3212343, EPI_ISL_3301008, EPI_ISL_3301513, EPI_ISL_3301592, EPI_ISL_3302738, EPI_ISL_3303032, EPI_ISL_3303035, EPI_ISL_3303516, EPI_ISL_3348010, EPI_ISL_3349409, EPI_ISL_3349460, EPI_ISL_3350552, EPI_ISL_3397129, EPI_ISL_3457436, EPI_ISL_3457484, EPI_ISL_3520910, EPI_ISL_3521421, EPI_ISL_3522426, EPI_ISL_3523079, EPI_ISL_3608006, EPI_ISL_3608193, EPI_ISL_3608291, EPI_ISL_3659923, EPI_ISL_3662599, EPI_ISL_3719656, EPI_ISL_4022689, EPI_ISL_4024741, EPI_ISL_4032778, EPI_ISL_4034302, EPI_ISL_4034554 | | | |
| see above | Fulgent Genetics | Centers for Disease Control and Prevention Division of Viral Diseases, Pathogen Discovery | Adrian Paskey; Becky Tsai; Benafsh Sapra; Benjamin Rambo-Martin; Christopher Gulvick; Clinton Paden; Clinton R. Paden; Dakota Howard; Darlene Wagner; Dhwani Batra; Doreen Ng; Duncan MacCannell; Harry Gao; James Xie; Jason Caravas; John Gao; Joseph Fierro; Kara Moser; Matthew Schmerer; Mickey Li; Peter Cook; Peter W. Cook; Scott Sammons; Shatavia Morrison; Yan Meng; Yvette Unoarumhi |
| EPI_ISL_3231764, EPI_ISL_3435077, EPI_ISL_3435078 | GA Department of Public Health | GA Department of Public Health | Aliyah Fields; Cynthia Dixey; Jonathan Edwards; Sharmila Talekar; Stacy Reeves; Taylor Smith; Tonia Parrott |
| EPI_ISL_3476495 | GH A.CHENEVIER-H.MONDOR | Department of Virology, Henri Mondor University Hospital, Assistance Publique Hôpitaux de Paris, Université Paris-Est Créteil, INSERM U955 | Alexandre Soulier; Christophe Rodriguez; Elisabeth Trawinski; Guillaume Gricourt; Jean-Michel Pawlotsky; Melissa N'Debi; Slim Fourati; Vanessa Demontant |
| EPI_ISL_3185657, EPI_ISL_3185663, EPI_ISL_3185669, EPI_ISL_3185670, EPI_ISL_3185671, EPI_ISL_3185680, EPI_ISL_3185682, EPI_ISL_3185684, EPI_ISL_3185697, EPI_ISL_3185716, EPI_ISL_3185723, EPI_ISL_3185724, EPI_ISL_3185736, EPI_ISL_3185740, EPI_ISL_3185743, EPI_ISL_3185754, EPI_ISL_3185755, EPI_ISL_3185761, EPI_ISL_3185767, EPI_ISL_3185772, EPI_ISL_3185777, EPI_ISL_3185785, EPI_ISL_3185796, EPI_ISL_3185797, EPI_ISL_3185799, EPI_ISL_3185800, EPI_ISL_3185801, EPI_ISL_3185805, EPI_ISL_3185808, EPI_ISL_3262663, EPI_ISL_3262666, EPI_ISL_3262669, EPI_ISL_3262674, EPI_ISL_3262677, EPI_ISL_3262678, EPI_ISL_3262679, EPI_ISL_3262687, EPI_ISL_3262701, EPI_ISL_3262704, EPI_ISL_3262705, EPI_ISL_3262706, EPI_ISL_3262712, EPI_ISL_3262718, EPI_ISL_3341868, EPI_ISL_3342040, EPI_ISL_3342042, EPI_ISL_3342043, EPI_ISL_3342044, EPI_ISL_3342045, EPI_ISL_3358542, EPI_ISL_3358544, EPI_ISL_3358546, EPI_ISL_3358548, EPI_ISL_3363548, EPI_ISL_3363569, EPI_ISL_3363582, EPI_ISL_3363618, EPI_ISL_3363627, EPI_ISL_3363631, EPI_ISL_3363635, EPI_ISL_3363646, EPI_ISL_3363649, EPI_ISL_3363653, EPI_ISL_3363656, EPI_ISL_3363659, EPI_ISL_3363674, EPI_ISL_3364915, EPI_ISL_3364923, EPI_ISL_3364926, EPI_ISL_3364929, EPI_ISL_3364936, EPI_ISL_3364954, EPI_ISL_3364958, EPI_ISL_3364967, EPI_ISL_3364971, EPI_ISL_3364987, EPI_ISL_3364991, EPI_ISL_3365018, EPI_ISL_3365022, EPI_ISL_3365026, EPI_ISL_3368409, EPI_ISL_3368430, EPI_ISL_3368439, EPI_ISL_3368453, EPI_ISL_3368454, EPI_ISL_3368464, EPI_ISL_3368777, EPI_ISL_3368778, EPI_ISL_3368785, EPI_ISL_3368786, EPI_ISL_3368788, EPI_ISL_3368794, EPI_ISL_3368795, EPI_ISL_3368796, EPI_ISL_3368797, EPI_ISL_3368799, EPI_ISL_3368803, EPI_ISL_3368805, EPI_ISL_3368809, EPI_ISL_3368851, EPI_ISL_3368852, EPI_ISL_3368856, EPI_ISL_3368864, EPI_ISL_3368866, EPI_ISL_3368875, EPI_ISL_3368876, EPI_ISL_3368878, EPI_ISL_3368882, EPI_ISL_3369291, EPI_ISL_3369293, EPI_ISL_3369295, EPI_ISL_3369300, EPI_ISL_3369304, EPI_ISL_3369341, EPI_ISL_3369342, EPI_ISL_3369343, EPI_ISL_3369344, EPI_ISL_3369345, EPI_ISL_3369346, EPI_ISL_3369350, EPI_ISL_3369351, EPI_ISL_3369356, EPI_ISL_3369376, EPI_ISL_3369378, EPI_ISL_3369379, EPI_ISL_3369380, EPI_ISL_3369395, EPI_ISL_3369396, EPI_ISL_3369399, EPI_ISL_3369401, EPI_ISL_3369405, EPI_ISL_3369406, EPI_ISL_3369407, EPI_ISL_3536006, EPI_ISL_3536017, EPI_ISL_3536022, EPI_ISL_3536028, EPI_ISL_3536095, EPI_ISL_3536096, EPI_ISL_3536101, EPI_ISL_3536113, EPI_ISL_3536114, EPI_ISL_3536119, EPI_ISL_3536120, EPI_ISL_3536131, EPI_ISL_3536132, EPI_ISL_3536133, EPI_ISL_3536135, EPI_ISL_3536136, EPI_ISL_3536140, EPI_ISL_3536144, EPI_ISL_3537148, EPI_ISL_3537152, EPI_ISL_3537154, EPI_ISL_3537160, EPI_ISL_3537165, EPI_ISL_3537166, EPI_ISL_3537169, EPI_ISL_3537170, EPI_ISL_3537176, EPI_ISL_3537186, EPI_ISL_3537188, EPI_ISL_3537189, EPI_ISL_3537311, EPI_ISL_3537312, EPI_ISL_3537313, EPI_ISL_3537314, EPI_ISL_3537319, EPI_ISL_3537320, EPI_ISL_3537329, EPI_ISL_3537332, EPI_ISL_3537333, EPI_ISL_3537334, EPI_ISL_3537336, EPI_ISL_3546372, EPI_ISL_3546373, EPI_ISL_3546379, EPI_ISL_3981960, EPI_ISL_3981961, EPI_ISL_3981963, EPI_ISL_3981969, EPI_ISL_3981970, EPI_ISL_3981971, EPI_ISL_3981974, EPI_ISL_3981975, EPI_ISL_3981977, EPI_ISL_3981979, EPI_ISL_3981999, EPI_ISL_3982399, EPI_ISL_3982401, EPI_ISL_3982422, EPI_ISL_3982428, EPI_ISL_3982623, EPI_ISL_3982630, EPI_ISL_3982631, EPI_ISL_3982636, EPI_ISL_3982637, EPI_ISL_3982643, EPI_ISL_3982644, EPI_ISL_3982649, EPI_ISL_3982650, EPI_ISL_3982651, EPI_ISL_3982653, EPI_ISL_3982654, EPI_ISL_3982655, EPI_ISL_3982656, EPI_ISL_3982657, EPI_ISL_3982658, EPI_ISL_3982664, EPI_ISL_3987648, EPI_ISL_3987649, EPI_ISL_3987651, EPI_ISL_3987661, EPI_ISL_3987662, EPI_ISL_3987663, EPI_ISL_3987665, EPI_ISL_3987868, EPI_ISL_3987875, EPI_ISL_3987894, EPI_ISL_3987895, EPI_ISL_3987920, EPI_ISL_3988092, EPI_ISL_3988093, EPI_ISL_3988094, EPI_ISL_3988095, EPI_ISL_3988096, EPI_ISL_3988102, EPI_ISL_3988111, EPI_ISL_3988112, EPI_ISL_3988113, EPI_ISL_3988117, EPI_ISL_3988118, EPI_ISL_3988119, EPI_ISL_3988124, EPI_ISL_3988128, EPI_ISL_3988143, EPI_ISL_3988144, EPI_ISL_3988149, EPI_ISL_3988150, EPI_ISL_3988153, EPI_ISL_3988216, EPI_ISL_3988233, EPI_ISL_3988234, EPI_ISL_3988235, EPI_ISL_3988239, EPI_ISL_3988252, EPI_ISL_3988255, EPI_ISL_3988264, EPI_ISL_3988268, EPI_ISL_3988269, EPI_ISL_3988289, EPI_ISL_3988299, EPI_ISL_4005019, EPI_ISL_4005021, EPI_ISL_4005023, EPI_ISL_4005024, EPI_ISL_4005025, EPI_ISL_4005027, EPI_ISL_4005031, EPI_ISL_4005044, EPI_ISL_4005045, EPI_ISL_4005046, EPI_ISL_4005080, EPI_ISL_4005081, EPI_ISL_4005084, EPI_ISL_4005085, EPI_ISL_4005087, EPI_ISL_4005089, EPI_ISL_4005091, EPI_ISL_4005092, EPI_ISL_4005093, EPI_ISL_4005094, EPI_ISL_4005095, EPI_ISL_4005096, EPI_ISL_4005097, EPI_ISL_4005098, EPI_ISL_4005101, EPI_ISL_4005102, EPI_ISL_4005103, EPI_ISL_4005104, EPI_ISL_4005105, EPI_ISL_4005106, EPI_ISL_4005107, EPI_ISL_4005108, EPI_ISL_4005109, EPI_ISL_4005133, EPI_ISL_4005153, EPI_ISL_4005155, EPI_ISL_4005157, EPI_ISL_4005159, EPI_ISL_4005160, EPI_ISL_4005161, EPI_ISL_4005162, EPI_ISL_4005171, EPI_ISL_4005172, EPI_ISL_4005173, EPI_ISL_4005177, EPI_ISL_4005181, EPI_ISL_4005182, EPI_ISL_4005183, EPI_ISL_4005184, EPI_ISL_4005185, EPI_ISL_4005186, EPI_ISL_4005188, EPI_ISL_4005221, EPI_ISL_4005222, EPI_ISL_4005224, EPI_ISL_4005228, EPI_ISL_4005230, EPI_ISL_4005232, EPI_ISL_4005233 | | | |
| see above | Genetica Molecular and Subdepartamento de Virologia ISP Chile | Instituto de Salud Publica de Chile | Andres Castillo; Barbara Parra; Constanza Campano; Gisselle Barra; Javier Tognarelli; Jorge Fernandez; Karen Orostica; Loredana Arata; Patricia Bustos; Rodrigo Fasce; Soledad Ulloa |
| EPI_ISL_3624115 | Gravity Diagnostics, LLC | Gravity Diagnostics, LLC | Gravity Diagnostics |
| EPI_ISL_3636676 | H Evora | Instituto Nacional de Saude (INSA) | Borges et al |
| EPI_ISL_3204354 | H Garcia de Orta | Instituto Nacional de Saude (INSA) | Borges et al |
| EPI_ISL_3536208, EPI_ISL_3536209, EPI_ISL_3536211, EPI_ISL_3536212, EPI_ISL_3912161 | HEMOCE CENTRO DE HEMATOLOGIA E HEMOTERAPIA DO CEARA | Analytical Competence Molecular Epidemiology Lab/ACME, Oswaldo Cruz Foundation, Ceara (FIOCRUZ CE) | Cleber Furtado Aksenen; Fabio Miyajima; Fernando Braga Stehling; Francisco Eder de Moura Lopes; Jamille Maria Mendes Bezerra; Joaquim Cesar do Nascimento Sousa Junior; Joaquim César do Nascimento Sousa Junior; Pedro Miguel Carneiro Jeronimo; Suzana Porto Almeida & Lucas Delerino on behalf of COVID-19 FIOCRUZ Genomic Network; Suzana Porto Almeida e Lucas Delerino; Thais Ferreira de Oliveira; Thais de Oliveira Costa; Ticiane Cavalcante de Souza; Veridiana Pessoa Miyajima |
| EPI_ISL_3912168, EPI_ISL_3912169 | HIAS HOSPITAL INFANTIL ALBERT SABIN | Analytical Competence Molecular Epidemiology Lab/ACME, Oswaldo Cruz Foundation, Ceara (FIOCRUZ CE) | Cleber Furtado Aksenen; Fabio Miyajima; Fernando Braga Stehling; Francisco Eder de Moura Lopes; Jamille Maria Mendes Bezerra; Joaquim Cesar do Nascimento Sousa Junior; Pedro Miguel Carneiro Jeronimo; Suzana Porto Almeida & Lucas Delerino on behalf of COVID-19 FIOCRUZ Genomic Network; Thais Ferreira de Oliveira; Thais de Oliveira Costa; Ticiane Cavalcante de Souza; Veridiana Pessoa Miyajima |
| EPI_ISL_3259358, EPI_ISL_3259359, EPI_ISL_3259363, EPI_ISL_3259364, EPI_ISL_3259365, EPI_ISL_3259366, EPI_ISL_3259368, EPI_ISL_3259371, EPI_ISL_3259376, EPI_ISL_3259377, EPI_ISL_3259378, EPI_ISL_3259379, EPI_ISL_3259380, EPI_ISL_3274748, EPI_ISL_3274749, EPI_ISL_3274753, EPI_ISL_3274755, EPI_ISL_3274757, EPI_ISL_3274762, EPI_ISL_3274763, EPI_ISL_3274767, EPI_ISL_3274769, EPI_ISL_3274770, EPI_ISL_3274774, EPI_ISL_3274777, EPI_ISL_3274783, EPI_ISL_3274784, EPI_ISL_3274786, EPI_ISL_3274788, EPI_ISL_3299430, EPI_ISL_3386132, EPI_ISL_3386137, EPI_ISL_3386141, EPI_ISL_3386149, EPI_ISL_3386152, EPI_ISL_3536439, EPI_ISL_3536441, EPI_ISL_3536443, EPI_ISL_3536446, EPI_ISL_3536452, EPI_ISL_3536453, EPI_ISL_3536458, EPI_ISL_3536459, EPI_ISL_3536466, EPI_ISL_3536470, EPI_ISL_3536472, EPI_ISL_3536474, EPI_ISL_3536478, EPI_ISL_3536479, EPI_ISL_3536480, EPI_ISL_3536482, EPI_ISL_3536485, EPI_ISL_3536489, EPI_ISL_3536490, EPI_ISL_3536494, EPI_ISL_3536496, EPI_ISL_3536498, EPI_ISL_3536507, EPI_ISL_3536508, EPI_ISL_3536511, EPI_ISL_3536516, EPI_ISL_3536518, EPI_ISL_3536520, EPI_ISL_3536521, EPI_ISL_3536534, EPI_ISL_3536537, EPI_ISL_3758060, EPI_ISL_3758065, EPI_ISL_3758074, EPI_ISL_3758080, EPI_ISL_3758086, EPI_ISL_3758087, EPI_ISL_3758092, EPI_ISL_3758093, EPI_ISL_3758097, EPI_ISL_3758100, EPI_ISL_3758109, EPI_ISL_3758111, EPI_ISL_3758127, EPI_ISL_3758128, EPI_ISL_3758130, EPI_ISL_3758133, EPI_ISL_3758135, EPI_ISL_3758141, EPI_ISL_3758142, EPI_ISL_3758145, EPI_ISL_3758146, EPI_ISL_3758149, EPI_ISL_3758154, EPI_ISL_3758156, EPI_ISL_3922206, EPI_ISL_3922210, EPI_ISL_3922212, EPI_ISL_3922213, EPI_ISL_3922215, EPI_ISL_3922223, EPI_ISL_3922229, EPI_ISL_3922230, EPI_ISL_3922232, EPI_ISL_3922236, EPI_ISL_3922238, EPI_ISL_3922250, EPI_ISL_3922254, EPI_ISL_3922256, EPI_ISL_3922257, EPI_ISL_3997118, EPI_ISL_3997120, EPI_ISL_3997123, EPI_ISL_3997129, EPI_ISL_3997135, EPI_ISL_3997144, EPI_ISL_3997153, EPI_ISL_3997160, EPI_ISL_3997163, EPI_ISL_3997164, EPI_ISL_3997172, EPI_ISL_3997180, EPI_ISL_3997183, EPI_ISL_3997185, EPI_ISL_3997190, EPI_ISL_3997195, EPI_ISL_3997209, EPI_ISL_3997210, EPI_ISL_3997211, EPI_ISL_4061629, EPI_ISL_4061663, EPI_ISL_4061664, EPI_ISL_4061667, EPI_ISL_4061670 | | | |
| see above | HLAGYN - Laboratorio de Imunologia de Transplantes de Goias | HLAGYN - Laboratorio de Imunologia de Transplantes de Goias | Alessandro Leonardo Alvares Magalhaes; Erika Lopes Rocha Batista; Fernando Antonio Vinhal dos Santos; Frederico Rodrigues Vinhal; Kamila Oliveira Reis De Freitas.; Lucas Carlos Gomes Pereira; Sabrina Sara Moreira Duarte |
| EPI_ISL_3536229, EPI_ISL_3536235, EPI_ISL_3536244, EPI_ISL_3536250, EPI_ISL_3536260, EPI_ISL_3536261, EPI_ISL_3536262, EPI_ISL_3912170, EPI_ISL_3912171, EPI_ISL_3912172 | | | |
| see above | HM HOSPITAL DE MESSEJANA DR CARLOS ALBERTO STUDART GOMES | Analytical Competence Molecular Epidemiology Lab/ACME, Oswaldo Cruz Foundation, Ceara (FIOCRUZ CE) | Cleber Furtado Aksenen; Fabio Miyajima; Fernando Braga Stehling; Francisco Eder de Moura Lopes; Jamille Maria Mendes Bezerra; Joaquim Cesar do Nascimento Sousa Junior; Joaquim César do Nascimento Sousa Junior; Pedro Miguel Carneiro Jeronimo; Suzana Porto Almeida & Lucas Delerino on behalf of COVID-19 FIOCRUZ Genomic Network; Suzana Porto Almeida e Lucas Delerino; Thais Ferreira de Oliveira; Thais de Oliveira Costa; Ticiane Cavalcante de Souza; Veridiana Pessoa Miyajima |
| EPI_ISL_3997818 | HOSPITAL DE BELLVITGE | Microbiology Department | Aida Gonzalez-Diaz; Carmen Ardanuy; Jordi Camara; Jordi Niubó; Laura Calatayud; M Angeles Domínguez; Sara Marti; Yolanda Hernandez |
| EPI_ISL_3639085 | HOSPITAL DR. RAFAEL A. CALDERON GUARDIA | Inciensa, Instituto Costarricense de Investigación y Enseñanza en Nutrición y Salud | Adriana Godínez; Claudio Soto-Garita; Estela Cordero; Francisco Duarte; Hebleen Porras; Joselyn Prado & Fabian Salas; José Luis Vargas; Mariela Gutiérrez; Melany Calderón |
| EPI_ISL_3912194, EPI_ISL_3912195 | HOSPITAL E MATERNIDADE DRA ZILDA ARNS NEUMANN | Analytical Competence Molecular Epidemiology Lab/ACME, Oswaldo Cruz Foundation, Ceara (FIOCRUZ CE) | Cleber Furtado Aksenen; Fabio Miyajima; Fernando Braga Stehling; Francisco Eder de Moura Lopes; Jamille Maria Mendes Bezerra; Joaquim Cesar do Nascimento Sousa Junior; Pedro Miguel Carneiro Jeronimo; Suzana Porto Almeida & Lucas Delerino on behalf of COVID-19 FIOCRUZ Genomic Network; Thais Ferreira de Oliveira; Thais de Oliveira Costa; Ticiane Cavalcante de Souza; Veridiana Pessoa Miyajima |
| EPI_ISL_3536163, EPI_ISL_3536164, EPI_ISL_3536171, EPI_ISL_3536172, EPI_ISL_3536268, EPI_ISL_3536276, EPI_ISL_3536333, EPI_ISL_3536396, EPI_ISL_3912199, EPI_ISL_3912202, EPI_ISL_3912203, EPI_ISL_3912205 | | | |
| see above | HOSPITAL ESTADUAL LEONARDO DA VINCI | Analytical Competence Molecular Epidemiology Lab/ACME, Oswaldo Cruz Foundation, Ceara (FIOCRUZ CE) | Cleber Furtado Aksenen; Fabio Miyajima; Fernando Braga Stehling; Francisco Eder de Moura Lopes; Jamille Maria Mendes Bezerra; Joaquim Cesar do Nascimento Sousa Junior; Joaquim César do Nascimento Sousa Junior; Pedro Miguel Carneiro Jeronimo; Suzana Porto Almeida & Lucas Delerino on behalf of COVID-19 FIOCRUZ Genomic Network; Suzana Porto Almeida e Lucas Delerino; Thais Ferreira de Oliveira; Thais de Oliveira Costa; Ticiane Cavalcante de Souza; Veridiana Pessoa Miyajima |
| EPI_ISL_3948522 | HOSPITAL LA ANEXION | Inciensa, Instituto Costarricense de Investigación y Enseñanza en Nutrición y Salud | Adriana Godínez; Claudio Soto-Garita; Estela Cordero; Francisco Duarte; Hebleen Porras; Joselyn Prado & Adriana Bermúdez; José Luis Vargas; Mariela Gutiérrez; Melany Calderón |
| EPI_ISL_3639148 | HOSPITAL MONSEÑOR SANABRIA | Inciensa, Instituto Costarricense de Investigación y Enseñanza en Nutrición y Salud | Adriana Godínez; Claudio Soto-Garita; Estela Cordero; Francisco Duarte; Hebleen Porras; Joselyn Prado & Andrea Moreno; José Luis Vargas; Mariela Gutiérrez; Melany Calderón |
| EPI_ISL_3922309 | HOSPITAL MUNICIPAL DE ITAPIRA | Instituto Butantan | Antonio Jorge Martins; Claudia Renata dos Santos Barros; David Schlesinger; Debora Botequio Moretti; Dimas Tadeu Covas; Elaine Cristina Marqueze; Elaine Vieira Santos; Evandra Strazza Rodrigues; Heidge Fukumasu; Jayme Augusto de Souza-Neto; José Salvatore Leister Patané; Luiz Alcantara; Luiz Lehmann Coutinho; Maria Carolina Elias; Maurício Lacerda Nogueira; Rafael dos Santos Bezerra; Raul Machado Neto; Rejane Maria Tommasini Grotto; Ricardo Haddad; Sandra Coccuzzo Sampaio Vessoni; Simone Kashima; Svetoslav Nanev Slavov; Vincent Louis Viala |
| EPI_ISL_3912214, EPI_ISL_3912215 | HOSPITAL MUNICIPAL NOSSA SENHORA DOS MILAGRES | Analytical Competence Molecular Epidemiology Lab/ACME, Oswaldo Cruz Foundation, Ceara (FIOCRUZ CE) | Cleber Furtado Aksenen; Fabio Miyajima; Fernando Braga Stehling; Francisco Eder de Moura Lopes; Jamille Maria Mendes Bezerra; Joaquim Cesar do Nascimento Sousa Junior; Pedro Miguel Carneiro Jeronimo; Suzana Porto Almeida & Lucas Delerino on behalf of COVID-19 FIOCRUZ Genomic Network; Thais Ferreira de Oliveira; Thais de Oliveira Costa; Ticiane Cavalcante de Souza; Veridiana Pessoa Miyajima |
| EPI_ISL_3761771, EPI_ISL_3761792, EPI_ISL_3761800 | HOSPITAL NACIONAL DE NIÑOS | Inciensa, Instituto Costarricense de Investigación y Enseñanza en Nutrición y Salud | Cristian Pérez-Corrales & Valeria Peralta-Barquero |
| EPI_ISL_3912223 | HOSPITAL REGIONAL DO SERTAO CENTRAL | Analytical Competence Molecular Epidemiology Lab/ACME, Oswaldo Cruz Foundation, Ceara (FIOCRUZ CE) | Cleber Furtado Aksenen; Fabio Miyajima; Fernando Braga Stehling; Francisco Eder de Moura Lopes; Jamille Maria Mendes Bezerra; Joaquim Cesar do Nascimento Sousa Junior; Pedro Miguel Carneiro Jeronimo; Suzana Porto Almeida & Lucas Delerino on behalf of COVID-19 FIOCRUZ Genomic Network; Thais Ferreira de Oliveira; Thais de Oliveira Costa; Ticiane Cavalcante de Souza; Veridiana Pessoa Miyajima |
| EPI_ISL_3639017 | HOSPITAL SAN FRANCISCO DE ASIS | Inciensa, Instituto Costarricense de Investigación y Enseñanza en Nutrición y Salud | Adriana Godínez; Claudio Soto-Garita; Estela Cordero; Francisco Duarte; Hebleen Porras; Joselyn Prado & Adrián Fallas Mora; José Luis Vargas; Mariela Gutiérrez; Melany Calderón |
| EPI_ISL_3948490 | HOSPITAL SAN JUAN DE DIOS | Inciensa, Instituto Costarricense de Investigación y Enseñanza en Nutrición y Salud | Adriana Godínez; Claudio Soto-Garita; Estela Cordero; Francisco Duarte; Hebleen Porras; Joselyn Prado & Juliana Mora; José Luis Vargas; Mariela Gutiérrez; Melany Calderón |
| EPI_ISL_3638824 | HOSPITAL SAN RAFAEL DE ALAJUELA | Inciensa, Instituto Costarricense de Investigación y Enseñanza en Nutrición y Salud | Adriana Godínez; Claudio Soto-Garita; Estela Cordero; Francisco Duarte; Hebleen Porras; Joselyn Prado & Christian Sanchez; José Luis Vargas; Mariela Gutiérrez; Melany Calderón |

| | | | |
|---|---|---|---|
| EPI_ISL_3639143 | HOSPITAL SAN VICENTE DE PAUL | Inciensa, Instituto Costarricense de Investigación y Enseñanza en Nutrición y Salud | Adriana Godínez; Claudio Soto-Garita; Estela Cordero; Francisco Duarte; Hebleen Porras; Joselyn Prado & Silvia Sáenz; José Luis Vargas; Mariela Gutiérrez; Melany Calderón |
| EPI_ISL_3298356, EPI_ISL_3948541 | HOSPITAL SAN VITO DE COTO BRUS | Inciensa, Instituto Costarricense de Investigación y Enseñanza en Nutrición y Salud | Adriana Godínez; Claudio Soto-Garita; Estela Cordero; Francisco Duarte; Hebleen Porras; Joselyn Prado & Maria Fernanda Matamoros; José Luis Vargas; Mariela Gutiérrez; Mariela Gutiérrez & Joselyn Prado; Melany Calderón |
| EPI_ISL_3912228 | HOSPITAL SAO JOSE DE DOENCAS INFECCIOSAS | Analytical Competence Molecular Epidemiology Lab/ACME, Oswaldo Cruz Foundation, Ceara (FIOCRUZ CE) | Cleber Furtado Aksenen; Fabio Miyajima; Fernando Braga Stehling; Francisco Eder de Moura Lopes; Jamille Maria Mendes Bezerra; Joaquim Cesar do Nascimento Sousa Junior; Pedro Miguel Carneiro Jeronimo; Suzana Porto Almeida & Lucas Delerino on behalf of COVID-19 FIOCRUZ Genomic Network; Thais Ferreira de Oliveira; Thais de Oliveira Costa; Ticiane Cavalcante de Souza; Veridiana Pessoa Miyajima |
| EPI_ISL_3188634 | Hospital General Universitario Gregorio Marañón | Hospital General Universitario Gregorio Marañón | Cristina Rodriguez-Grande; Darío García de Viedma; Julia Suárez; Laura Pérez-Lago; Marta Herranz Martin; Patricia Muñoz; Pedro Sola Campoy; Pilar Catalán; Sergio Buenestado Serrano; Victor Manuel de la Cueva |
| EPI_ISL_3260112 | Hospital Jaime Ferre - SAMCO Rafaela | Grupo de Genómica y Bioinformática del Instituto de Investigación de la Cadena Láctea CONICET-INTA on behalf of 'Proyecto Argentino Interinstitucional de genomica de SARS-CoV-2' (PAIS Consortium) | AF; Aliprandi D; Amadio; C; Eberhardt; Irazoqui; Isaia; JF; JM; MF; Pandolfi; Quaranta; Soratti R; V |
| EPI_ISL_4005953, EPI_ISL_4005957 | Hospital Sharp | Microbial Genomics Laboratory | ; Alejandra García-Gasca; Alejandra Hernández-Terán; Alejandro Sánchez-Flores; Alfredo Herrera-Estrella; Alicia Ocaña-Mondragón; Andreu Comas-García; Angel Gustavo Salas-Lais; Antonio Loza Román; Bernardo Martínez-Miguel; Blanca Taboada; Brenda Irasema Maldonado-Meza; Bruno Gómez-Gil; Carla Ivón Herrera-Najera; Carlos F. Arias; Celia Boukadida; Clara Esperanza Santacruz-Tinoco; Concepción Grajales-Muñiz; Consorcio Mexicano de Vigilancia Genómica (CoViGen-Mex). Authors (in alphabetical order): Julio Elias Alvarado-Yaah; Cristóbal Cháidez-Quiróz; Célida Duque Molina; Célida Martínez- Rodríguez; Daniel Fregoso-Rueda; Daniel Lira Morales; Eduardo Becerril-Vargas; Fernando Fontove-Herrera; Fidencio Mejía-Nepomuceno; Francisco Pulido; Gloria Elena Espinosa-Ayala; Gloria María Molina-Salinas; Gloria Vazquez; Hector Esteban Paz-Juárez; Hector Montoya-Fuentes; Helen Haydee Fernanda Ramírez-Plascencia; Irvin González-López; Jean Pierre González; Jesús Hernández; Joel Armando Vázquez-Pérez.; Jorge Salas-Hernández; José Antonio Enciso-Moreno; José Arturo Martínez-Orozco; José Esteban Muñoz-Medina; José de Jesús Nuñez-Contreras; Juan Bautista Chale-Dzul; Julissa Enciso-Ibarra; Luis Alberto Ochoa-Carrera; Margarita Matías-Florentino; Mario Mújica-Sánchez; Marissa Perez-Garcia; María Guadalupe Santiago-Mauricio; María Guadalupe de Jesús Mireles-Rivera; Nelly Sélem-Mojica; Pavel Isa; Ricardo Ciria Merce; Ricardo Grande; Rosa María Gutiérrez Rios; Santiago Ávila-Ríos; Selene Zárate; Susana Lopez; Verónica Mata-Haro; Victor Eduardo García-Arias; Víctor Hugo Borja-Aburto |
| EPI_ISL_3696641, EPI_ISL_3696682, EPI_ISL_3696726 | Hospital de Campanha de Guaratinguetá | Instituto Butantan | Antonio Jorge Martins; Claudia Renata dos Santos Barros; David Schlesinger; Debora Botequio Moretti; Dimas Tadeu Covas; Elaine Cristina Marqueze; Elaine Vieira Santos; Evandra Strazza Rodrigues; Heidge Fukumasu; Jayme Augusto de Souza-Neto; José Salvatore Leister Patané; Luiz Alcantara; Luiz Lehmann Coutinho; Maria Carolina Elias; Maurício Lacerda Nogueira; Rafael dos Santos Bezerra; Raul Machado Neto; Rejane Maria Tommasini Grotto; Ricardo Haddad; Sandra Coccuzzo Sampaio Vessoni; Simone Kashima; Svetoslav Nanev Slavov; Vincent Louis Viala |
| EPI_ISL_3398956, EPI_ISL_4004142, EPI_ISL_4004281 | Houston Health Dept. | Houston Health Dept. | Adolpho Lara; Pamela Brown; Ryker Penn; Yanlai Lai |
| EPI_ISL_3303826, EPI_ISL_3369475, EPI_ISL_3936902 | IN State Department of Health Laboratory Services | IN State Department of Health Laboratory Services | Brian Pope; Cassandra Campion; Jamie Yeadon; Kyle Brownlee; Lixia Liu; Mark Glazier; Melissa Hindenlang |
| EPI_ISL_3068188, EPI_ISL_3068202, EPI_ISL_3068241, EPI_ISL_3274395, EPI_ISL_3274430, EPI_ISL_3274458, EPI_ISL_3274460, EPI_ISL_3274480, EPI_ISL_3274497, EPI_ISL_3274502, EPI_ISL_3274520, EPI_ISL_3274525, EPI_ISL_3274530, EPI_ISL_3274569, EPI_ISL_3274608, EPI_ISL_3274615 | | | |
| see above | INSPI-CRN DE INFLUENZA Y OTROS VIRUS RESPIRATORIOS | NIC-INSPI | Alfredo Bruno; Daniel Ramos; Domenica de Mora.; Jimmy Garcés; Johanna Laines; Lizbeth Patiño; Manuel Gonzalez; Maria Angelica Becerra; Maritza Olmedo; Michelle Páez |
| EPI_ISL_3088327 | INTERLAB | Omics Sciences Laboratory | Darlyn Amaya; Derly Andrade Molina; Gabriel Morey León; Juan Carlos Fernández Cadena; Rubén Armas González |
| EPI_ISL_3068295, EPI_ISL_3068586, EPI_ISL_3161423, EPI_ISL_3161554, EPI_ISL_3161569, EPI_ISL_3419716, EPI_ISL_3419948 | | | |
| see above | IRCCS San Gallicano Dermatological Institute | IRCCS Regina Elena National Cancer Institute | Aldo Morrone; Alice Massacci; Andrea Cazzani; Eleonora Sperandio; Elisabetta Trento; Fabrizio Ensoli; Francesca De Nicola; Francesca Maione; Francesca Sivori; Frauke Goeman; Fulvia Pimpinelli; Gennaro Ciliberto; Giovanna D'agosto; Giovanni Blandino; Giulia Orlandi; Ilaria Cavallo; Ludovica Ciuffreda; Matteo Pallocca; Maurizio Fanciulli; Sabrina Strano; Sara Donzelli; Sara Petrolo |
| EPI_ISL_3547122, EPI_ISL_3547148 | IVY3 Central Lab, Vanderbilt University Medical Center | Lauring Lab, University of Michigan, Department of Microbiology and Immunology | Gilbert |
| EPI_ISL_3156477, EPI_ISL_3272419, EPI_ISL_3640996, EPI_ISL_3664324 | Idaho Bureau of Laboratories | Boise VA Medical Center, PALMS | Aimee Ceniseros; Cheri Lamb McFarlane; Christian Loera; Dr. Christopher Ball; Emily Bartlett; Emily Bartlette; James Razor; Matthew Burns; Ying Pei |
| EPI_ISL_3642917, EPI_ISL_3722427, EPI_ISL_4084693 | Idaho Bureau of Laboratories | Center for Global Health, University of New Mexico Health Sciences Center | Abigail Cortez; Aimee Ceniseros; Cassidy Vanwarmerdam; Christian Loera; Christopher Ball; Darrell Dinwiddie; Daryl Domman; Josie Santos; Kurt Schwalm; Matthew Burns; Robert Voermans; Valerie Morley |
| EPI_ISL_3232011 | Illinois Department of Public Health | Illinois Department of Public Health - Chicago Lab | Ira Heimler; Joel Price; Vineet K. Dhiman |
| EPI_ISL_3347020 | Incyte Diagnostics Spokane | Seattle Flu Study | Amanda Adler; Barry R. Lutz; Benjamin Pelle; Brian Hiatt; Caitlin R. Wolf; Chris D. Frazar; Deborah A. Nickerson; Elisabeth Brandstetter; Erica Ryke; Geoff Melly; Helen Y. Chu; Janet A. Englund; Jay Shendure; Jover Lee; Kairsten Fay; Kirsten Lacombe; Lea M. Starita; Mark J. Rieder; Matthew Richardson; Matthew Thompson; Melissa Truong; Michael Boeckh; Michael Famulare; Misja Ilcisin; Peter D. Han; Philip Dykema; Romesh Gautom; Scott Lindquist; Thomas R. Sibley; Trevor Bedford |
| EPI_ISL_3299775 | Infinity Biologix | Centers for Disease Control and Prevention Division of Viral Diseases, Pathogen Discovery | Adrian Paskey; Benjamin Rambo-Martin; Chirayu Goswami; Christian Bixby; Christopher Gulvick; Clinton R. Paden; Dakota Howard; Darlene Wagner; Dhwani Batra; Duncan MacCannell; Jason Caravas; Jonathan Schultz; Kara Moser; Matthew Schmerer; Peter W. Cook; Robin Grimwood; Russ Hager; Scott Sammons; Shatavia Morrison; Yihe Wang; Yvette Unoarumhi |
| EPI_ISL_3505610, EPI_ISL_3505611, EPI_ISL_3824606 | Institute of Microbiology, Universidad San Francisco de Quito | Institute of Microbiology, Universidad San Francisco de Quito | Alexandra Gonzalez; Belén Prado-Vivar; Bernardo Gutiérrez; Betty Angulo; Erika B. Muñoz; Fernanda Zurita; Francisco Guerra; Gabriel Trueba; Hugo Vergara; Jeaninna Peña; Juan José Guadalupe; Juan Pablo Román; Liu Yuqian; Luis Fuenmayor; Luz-Angelica Castillo; Marco Viteri Yanez; Mateo Carvajal; Michelle Grunauer; Michelle Jacome; Monica Becerra-Wong; Nancy Flores Lastra; Natalia Parra; Patricio Rojas-Silva; Paúl Cárdenas; Ronny Javier Pibaque; Stephanie Espín-Arroba; Sully Márquez; Valeria Armijos; Verónica Barragán |
| EPI_ISL_3545781, EPI_ISL_3545787, EPI_ISL_3864599 | Instituto Adolfo Lutz - Regional de Marilia | Instituto Adolfo Lutz, Interdiciplinary Procedures Center, Strategic Laboratory | Caio Vinicius Dias Lopes; Claudia Regina Gonçalves; Claudio Tavares Sacchi; Karoline Rodrigues Campos; Leonardo Tadeu de Araujo; Marlon Benedito Nascomento Santos |
| EPI_ISL_3545770, EPI_ISL_3545771, EPI_ISL_3545772, EPI_ISL_3545780, EPI_ISL_4081145, EPI_ISL_4081147, EPI_ISL_4081148, EPI_ISL_4081149, EPI_ISL_4081150, EPI_ISL_4081151, EPI_ISL_4081152, EPI_ISL_4081153 | | | |
| see above | Instituto Adolfo Lutz - Regional de Sao Jose do Rio Preto | Instituto Adolfo Lutz, Interdiciplinary Procedures Center, Strategic Laboratory | Caio Vinicius Dias Lopes; Claudia Regina Gonçalves; Claudio Tavares Sacchi; Karoline Rodrigues Campos; Leonardo Tadeu de Araujo; Marlon Benedito Nascomento Santos; Marlon Benedito Nascomento Santos |
| EPI_ISL_3545774, EPI_ISL_3545775, EPI_ISL_3545785 | Instituto Adolfo Lutz - Regional de Sorocaba | Instituto Adolfo Lutz, Interdiciplinary Procedures Center, Strategic Laboratory | Caio Vinicius Dias Lopes; Claudia Regina Gonçalves; Claudio Tavares Sacchi; Karoline Rodrigues Campos; Leonardo Tadeu de Araujo; Marlon Benedito Nascomento Santos |
| EPI_ISL_3691688, EPI_ISL_3691689, EPI_ISL_3691690, EPI_ISL_3691694, EPI_ISL_3691698, EPI_ISL_3691699, EPI_ISL_3864540, EPI_ISL_3864542, EPI_ISL_3864544, EPI_ISL_3864551, EPI_ISL_3864556, EPI_ISL_3864558, EPI_ISL_3864560, EPI_ISL_3864562, EPI_ISL_3864564, EPI_ISL_3864568, EPI_ISL_3864572, EPI_ISL_3864574, EPI_ISL_3864576, EPI_ISL_3864579, EPI_ISL_3864585, EPI_ISL_3864589, EPI_ISL_3864590, EPI_ISL_3864592, EPI_ISL_3864598, EPI_ISL_4081132, EPI_ISL_4081134, EPI_ISL_4081137, EPI_ISL_4081140, EPI_ISL_4081141, EPI_ISL_4081142, EPI_ISL_4081144 | | | |
| see above | Instituto Adolfo Lutz Central | Instituto Adolfo Lutz, Interdiciplinary Procedures Center, Strategic Laboratory | Caio Vinicius Dias Lopes; Claudia Regina Gonçalves; Claudio Tavares Sacchi; Karoline Rodrigues Campos; Leonardo Tadeu de Araujo; Marlon Benedito Nascomento Santos; Marlon Benedito Nascomento Santos |
| EPI_ISL_3982817 | Instituto Nacional de Enfermedades Respiratorias (INER) | Centro de Investigación en Enfermedades Infecciosas (CIENI), Instituto Nacional de Enfermedades Respiratorias (INER) | Alejandra García-Gasca; Alejandra Hernández-Terán; Alejandro Sánchez-Flores; Alfredo Herrera-Estrella; Alicia Ocaña-Mondragón; Andreu Comas-García; Angel Gustavo Salas-Lais; Antonio Loza Román; Bernardo Martínez-Miguel; Blanca Taboada; Brenda Irasema Maldonado-Meza; Bruno Gómez-Gil; Carla Ivón Herrera-Najera; Carlos F. Arias; Celia Boukadida; Clara Esperanza Santacruz-Tinoco; Concepción Grajales-Muñiz; Consorcio Mexicano de Vigilancia Genómica (CoViGen-Mex). Authors (in alphabetical order): Julio Elias Alvarado-Yaah; Cristóbal Cháidez-Quiróz; Célida Duque Molina; Célida Martínez-Rodríguez; Daniel Fregoso-Rueda; Daniel Lira Morales; Eduardo Becerril-Vargas; Eduardo Rivera-Martínez; Fernando Fontove-Herrera; Fidencio Mejía-Nepomuceno; Francisco Pulido; Gabriel Chavira-Trujillo; Gloria Elena Espinosa-Ayala; Gloria María Molina-Salinas; Gloria Vazquez; Hector Montoya-Fuentes; Helen Haydee Fernanda Ramírez-Plascencia; Irvin González-López; Jean Pierre González; Jesús Hernández; Joel Armando Vázquez-Pérez).; Jorge Salas-Hernández; José Antonio Enciso-Moreno; José Arturo Martínez-Orozco; José Esteban Muñoz-Medina; José de Jesús Nuñez-Contreras; Juan Bautista Chale-Dzul; Julissa Enciso-Ibarra; Kathia Elizabeth Tapia-Diaz; Luis Alberto Ochoa-Carrera; Margarita Matías-Florentino; Mario Mújica-Sánchez; Marissa Perez-Garcia; María Eugenia Jiménez-Corona; María Guadalupe Santiago-Mauricio; María Guadalupe de Jesús Mireles-Rivera; Nelly Sélem-Mojica; Pavel Isa; Ricardo Ciria Merce; Ricardo Grande; Rosa María Gutiérrez Rios; Rosario Vazquez-Larios; Santiago Ávila-Ríos; Selene Zárate; Susana Lopez; Verónica Mata-Haro; Victor Eduardo García-Arias; Víctor Hugo Borja-Aburto |
| EPI_ISL_3118933, EPI_ISL_3824319, EPI_ISL_3824320, EPI_ISL_3824321, EPI_ISL_3824322, EPI_ISL_3824323, EPI_ISL_3824326, EPI_ISL_3824330, EPI_ISL_3824332, EPI_ISL_3824333, EPI_ISL_3824336, EPI_ISL_3824337, EPI_ISL_3824338, EPI_ISL_3824339, EPI_ISL_3824340, EPI_ISL_3824341, EPI_ISL_3824350 | | | |
| see above | Instituto de Biotecnologia - UNESP-Botucatu-SP | Instituto de Biotecnologia - UNESP-Botucatu-SP | Cecília Artico Banho; Cíntia Bittar; Fábio Sossai Possebon; Guilherme Campos; Helena Lage Ferreira; Jorge A. Petroli Marchesi; João Pessoa Araújo Jr.; Leila Sabrina Ullmann; Lívia Sacchetto; Maisa C. Pereira Parra; Marília Moraes; Maurício L. Nogueira; Paula Rahal; Paulo Inacio da Costa |
| EPI_ISL_3262656 | Kansas Health and Environmental Lab | Kansas Health and Environmental Lab | Amanda Bradley; Jonathan Barnell; Mike Grose; and Phil Adam |
| EPI_ISL_3471630 | LABO BIO DOC | CHU Purpan - Laboratoire de Virologie - Institut Fédératif de Biologie | Bulach T.; Donnadieu C.; Izopet J.; Latour J.; Milhes M.; Nicot F.; Ranger N.; Salin G.; Tremeaux P. |
| EPI_ISL_3379109 | LABORATOIRE ACCOLAB SUD OUEST | CNR Virus des Infections Respiratoires - France SUD | Antonin Bal; Bruno Lina; Gregory Destras; Gwendolyne Burfin; Hadrien Regue; Laurence Josset; Martine Valette; Quentin Semanas |

| | | | |
|---|---|---|---|
| EPI_ISL_3131258 | LABORATOIRE CBM DE MURET | CNR Virus des Infections Respiratoires - France SUD | Antonin Bal; Bruno Lina; Gregory Destras; Gwendolyne Burfin; Hadrien Regue; Laurence Josset; Martine Valette; Quentin Semanas |
| EPI_ISL_3841255, EPI_ISL_3841280, EPI_ISL_3841281, EPI_ISL_3841282, EPI_ISL_3841283, EPI_ISL_3841284, EPI_ISL_3841313, EPI_ISL_3841314 | | | |
| see above | LACEN | LGBio (Laboratorio de Genetica & Biodiversidade) | Alex Honda Bernardes; Amanda Alves de Melo; Aparecido Divino da Cruz; Cintia Pelegrineti Targueta de Azevedo Brito; Daniela de Melo e Silva; Elisangela de Paula Silveira Lacerda; Juliana Santana de Curcio; Luiz Augusto Pereira; Marc Alexandre Duarte Gigonzac; Mariana Pires de Campos Telles; Ramilla dos Santos Braga; Renata de Oliveira Dias; Rhewter Nunes; Thais Cidália Vieira Gigonzac; Thais Guimarães Castro; Thays Millena Alves Pedroso |
| EPI_ISL_3447459, EPI_ISL_3447460, EPI_ISL_3447461, EPI_ISL_3447462, EPI_ISL_3447465, EPI_ISL_3447466, EPI_ISL_3447468, EPI_ISL_3447471, EPI_ISL_3447473, EPI_ISL_3447476, EPI_ISL_3447480, EPI_ISL_3447481, EPI_ISL_3447482, EPI_ISL_3447483, EPI_ISL_3447484, EPI_ISL_3447485, EPI_ISL_3447489, EPI_ISL_3447569, EPI_ISL_3447570, EPI_ISL_3447571, EPI_ISL_3447573, EPI_ISL_3447574, EPI_ISL_3447575, EPI_ISL_3447576, EPI_ISL_3447577, EPI_ISL_3447578, EPI_ISL_3447579, EPI_ISL_3447580, EPI_ISL_3447582, EPI_ISL_3447583, EPI_ISL_3447584, EPI_ISL_3447585, EPI_ISL_3447587, EPI_ISL_3447592, EPI_ISL_3447598, EPI_ISL_3703726, EPI_ISL_3703733, EPI_ISL_3703737, EPI_ISL_3703755, EPI_ISL_3703772, EPI_ISL_3703778, EPI_ISL_3703779, EPI_ISL_3703780, EPI_ISL_3835240, EPI_ISL_3835242, EPI_ISL_3835246, EPI_ISL_3835248, EPI_ISL_3835249, EPI_ISL_3835251, EPI_ISL_3835255, EPI_ISL_3835257, EPI_ISL_3835258, EPI_ISL_3835264, EPI_ISL_3835265, EPI_ISL_3835276, EPI_ISL_3835277, EPI_ISL_3835282, EPI_ISL_3835283, EPI_ISL_3835284, EPI_ISL_3835286, EPI_ISL_3835290, EPI_ISL_3835295, EPI_ISL_3835299, EPI_ISL_3835303, EPI_ISL_3835304, EPI_ISL_3835354, EPI_ISL_3835355, EPI_ISL_3835357 | | | |
| see above | LACEN/PE | WallauLab on behalf of Fiocruz COVID-19 Genomic Surveillance Network | Alexandre Freitas da Silva; Cassia Docena; Constância Flávia Junqueira Ayres; Filipe Zimmer Dezordi; Gabriel Luz Wallau; Gustavo Barbosa de Lima; Lais Ceschini Machado; Lilian Caroliny Amorim Silva; Marcelo Henrique dos Santos Paiva; Matheus Filgueira Bezerra; Sinval Pinto Brandão Filho |
| EPI_ISL_3912232, EPI_ISL_3912233, EPI_ISL_3912234 | LACEN_CENTRO DE TESTAGEM PARA VIAJANTE | Analytical Competence Molecular Epidemiology Lab/ACME, Oswaldo Cruz Foundation, Ceara (FIOCRUZ CE) | Cleber Furtado Aksenen; Fabio Miyajima; Fernando Braga Stehling; Francisco Eder de Moura Lopes; Jamille Maria Mendes Bezerra; Joaquim Cesar do Nascimento Sousa Junior; Pedro Miguel Carneiro Jeronimo; Suzana Porto Almeida & Lucas Delerino on behalf of COVID-19 FIOCRUZ Genomic Network; Thais Ferreira de Oliveira; Thais de Oliveira Costa; Ticiane Cavalcante de Souza; Veridiana Pessoa Miyajima |
| EPI_ISL_3047717, EPI_ISL_3047718, EPI_ISL_3047732, EPI_ISL_3047737, EPI_ISL_3047739, EPI_ISL_3047740, EPI_ISL_3047746, EPI_ISL_3134923, EPI_ISL_3134924, EPI_ISL_3134925, EPI_ISL_3134931, EPI_ISL_3134936, EPI_ISL_3134938, EPI_ISL_3134940, EPI_ISL_3134941, EPI_ISL_3134942, EPI_ISL_3134944, EPI_ISL_3134945, EPI_ISL_3134948, EPI_ISL_3134952, EPI_ISL_3134953, EPI_ISL_3134956, EPI_ISL_3134961, EPI_ISL_3134966, EPI_ISL_3134968, EPI_ISL_3134969, EPI_ISL_3134970, EPI_ISL_3134971, EPI_ISL_3134972, EPI_ISL_3134974, EPI_ISL_3134975, EPI_ISL_3134976, EPI_ISL_3149366, EPI_ISL_3246152, EPI_ISL_3246153, EPI_ISL_3246155, EPI_ISL_3246158, EPI_ISL_3246159, EPI_ISL_3246160, EPI_ISL_3246161, EPI_ISL_3246162, EPI_ISL_3246170, EPI_ISL_3246172, EPI_ISL_3246174, EPI_ISL_3246177, EPI_ISL_3246179, EPI_ISL_3246181, EPI_ISL_3246183, EPI_ISL_3246185, EPI_ISL_3246186, EPI_ISL_3246187, EPI_ISL_3246188, EPI_ISL_3246191, EPI_ISL_3246193, EPI_ISL_3246196, EPI_ISL_3385140, EPI_ISL_3385141, EPI_ISL_3385142, EPI_ISL_3385143, EPI_ISL_3385144, EPI_ISL_3385145, EPI_ISL_3385146, EPI_ISL_3385147, EPI_ISL_3385149, EPI_ISL_3385150, EPI_ISL_3385151, EPI_ISL_3385152, EPI_ISL_3385153, EPI_ISL_3385154, EPI_ISL_3505191, EPI_ISL_3505193, EPI_ISL_3505195, EPI_ISL_3505196, EPI_ISL_3505197, EPI_ISL_3505204, EPI_ISL_3505206, EPI_ISL_3804720, EPI_ISL_3804723, EPI_ISL_3804724, EPI_ISL_3804725, EPI_ISL_3804727, EPI_ISL_3804728, EPI_ISL_3804729, EPI_ISL_3804730, EPI_ISL_3804733, EPI_ISL_3804735, EPI_ISL_3804736, EPI_ISL_3804738, EPI_ISL_3804744, EPI_ISL_3941511, EPI_ISL_3941512, EPI_ISL_3941513, EPI_ISL_3941515, EPI_ISL_3941518, EPI_ISL_4081415, EPI_ISL_4081417 | | | |
| see above | LATE - Laboratório de Técnicas Especiais - Hospital Israelita Albert Einstein | LATE - Laboratório de Técnicas Especiais - Hospital Israelita Albert Einstein | Alexandre Hideaki Takara; Ana Paula Moreira Salles; Anelisie da Silva Santos; Deyvid Amgarten; Erick Gustavo Dorlass; Fernanda de Mello Malta; João Renato Rebello Pinho; Marcio Anunciacao Menezes; Pedro Henrique Sebe Rodrigues; Raquel Riyuzo |
| EPI_ISL_3383156 | LBM SEALAB DARRASSE ET ASSOCIES | CNR Virus des Infections Respiratoires - France SUD | Antonin Bal; Bruno Lina; Gregory Destras; Gwendolyne Burfin; Hadrien Regue; Laurence Josset; Martine Valette; Quentin Semanas |
| EPI_ISL_3206149 | LBM UNIBIO VALENTIN | CNR Virus des Infections Respiratoires - France SUD | Antonin Bal; Bruno Lina; Gregory Destras; Gwendolyne Burfin; Hadrien Regue; Laurence Josset; Martine Valette; Quentin Semanas |
| EPI_ISL_3459386, EPI_ISL_3459387 | LDSP CAQUETA | Instituto Nacional de Salud | Carlos Franco-Muñoz; Carmen Osorio; Diana Malo; Diego A. Álvarez-Díaz; Diego Andrés Prada; Gerardo Santamaría; Hector Alejandro Ruiz-Moreno; Jhonnatan Reales-González; Jorge Rivera; Juan Camilo Martinez; Julian Naizaque; Katherine Laiton-Donato; Lisseth Pardo; Magdalena Wiesner; Marcela Mercado-Reyes; Maria T. Herrera-Sepúlveda; Marta Lopez Blanco; Martha Lucia Ospina Martinez; Paola Rojas; Sergio Gomez; Sheryll Corchuelo; Ángela Alarcon Cruz |
| EPI_ISL_3923743 | LESP Chiapas | Instituto de Diagnostico y Referencia Epidemiologicos (INDRE) | Abril Rodriguez-Maldonado; Ariadna Medina-Benitez; Claudia Wong-Arambula; Ernesto Ramirez-Gonzalez.; Gisela Barrera-Badillo; Irma Lopez-Martinez; Joaquin Quiroz-Mercado; Lucia Hernandez-Rivas; Maribel Gonzalez-Villa; Natividad Cruz-Ortiz; Sergio Rangel-Guerrero; Tatiana Nunez-Garcia; Vanessa Rivero-Arredondo |
| EPI_ISL_3459941, EPI_ISL_3459942 | LESP Queretaro | Instituto de Diagnostico y Referencia Epidemiologicos (INDRE) | Abril Rodriguez-Maldonado; Ariadna Medina-Benitez; Claudia Wong-Arambula; Ernesto Ramirez-Gonzalez.; Gisela Barrera-Badillo; Irma Lopez-Martinez; Joaquin Quiroz-Mercado; Lucia Hernandez-Rivas; Maribel Gonzalez-Villa; Natividad Cruz-Ortiz; Sergio Rangel-Guerrero; Tatiana Nunez-Garcia; Vanessa Rivero-Arredondo |
| EPI_ISL_3459945, EPI_ISL_3923753, EPI_ISL_3923754 | LESP Sonora | Instituto de Diagnostico y Referencia Epidemiologicos (INDRE) | Abril Rodriguez-Maldonado; Ariadna Medina-Benitez; Claudia Wong-Arambula; Ernesto Ramirez-Gonzalez.; Gisela Barrera-Badillo; Irma Lopez-Martinez; Joaquin Quiroz-Mercado; Lucia Hernandez-Rivas; Maribel Gonzalez-Villa; Natividad Cruz-Ortiz; Sergio Rangel-Guerrero; Tatiana Nunez-Garcia; Vanessa Rivero-Arredondo |
| EPI_ISL_3459955, EPI_ISL_3769141, EPI_ISL_3923744 | LESP Tabasco | Instituto de Diagnostico y Referencia Epidemiologicos (INDRE) | Abril Rodriguez-Maldonado; Ariadna Medina-Benitez; Claudia Wong-Arambula; Ernesto Ramirez-Gonzalez.; Gisela Barrera-Badillo; Irma Lopez-Martinez; Joaquin Quiroz-Mercado; Lucia Hernandez-Rivas; Maribel Gonzalez-Villa; Natividad Cruz-Ortiz; Sergio Rangel-Guerrero; Tatiana Nunez-Garcia; Vanessa Rivero-Arredondo |
| EPI_ISL_3508198 | LESPH | T4 OLIGO | CONNIE ALEJANDRA HERRERA-GOMEZ; HECTOR ALBERTO VILLAFUENTES-TELLEZ; LAURA SOFIA CASTILLO-ORTEGA; MELITO GARCIA-REYES; OCTAVIO GARCIA-GONZALEZ; YAJAIRA SULIM VIGUERAS-MORALES |
| EPI_ISL_3868205, EPI_ISL_3868313 | Lab voor klinische biologie | Lab voor klinische biologie | Bruno Verhasselt; Hannelore Hamerlinck; Marija Janevska |
| EPI_ISL_3801471 | Labo Analyses Med | National Reference Center for Viruses of Respiratory Infections, Institut Pasteur, Paris | Angela Brisebarre; Camille Capel; Christophe Malabat; Corinne Maufrais; Etienne Simon-Lorière; Frédéric Lemoine; Hub de Bioinformatique et Biostatistique; Louise Lefrançois; Marion Barbet; Maud Vanpeene; Méline Bizard; Sylvie Behillil; Sylvie Van der Werf; Vincent Enouf; Vincent Vieillefond |
| EPI_ISL_3239261 | Labor Becker & Kollegen (Standort München) | Robert Koch Institute | |
| EPI_ISL_3121513 | Laborarztpraxis Osnabrück | Robert Koch Institute | |
| EPI_ISL_3129957, EPI_ISL_3129960, EPI_ISL_3129961, EPI_ISL_3129962, EPI_ISL_3277498, EPI_ISL_3277499, EPI_ISL_3277500, EPI_ISL_3501652, EPI_ISL_3501654, EPI_ISL_3501656, EPI_ISL_3501657, EPI_ISL_3501659, EPI_ISL_3903682, EPI_ISL_3903683, EPI_ISL_3903684, EPI_ISL_3903685, EPI_ISL_3903686 | | | |
| see above | Laboratoire Carage | Institut Pasteur de la Guyane | Anne Lavergne; Antoine Enfissi; Arielle Salmier; Dominique Rousset |
| EPI_ISL_4028969, EPI_ISL_4028985, EPI_ISL_4028988, EPI_ISL_4028992, EPI_ISL_4028999, EPI_ISL_4029003 | Laboratoire National de Santé Publique – LNSP | Genomics and Proteomics Departament, Gorgas Memorial Institute For Health Studies | Alexander A Martinez; Ambar Moreno; Claudia Estrada; Claudia Gonzalez V; César Roberto Conde Pereira; Jessica Gondola; Leyda Abrego; Marlene Castillo; Melissa Gaitan; Oris Chavarria |
| EPI_ISL_3800194 | Laboratori de Referencia de Catalunya | Laboratori de Referencia de Catalunya | Bellosillo B.; Canal M.; Hernandez JJ.; Padilla E.; Ramirez A.; Vilas A. |
| EPI_ISL_3539231, EPI_ISL_3539235, EPI_ISL_3539236, EPI_ISL_3539237, EPI_ISL_4080819, EPI_ISL_4080821, EPI_ISL_4080822, EPI_ISL_4080823, EPI_ISL_4080828, EPI_ISL_4080829, EPI_ISL_4080830, EPI_ISL_4080831, EPI_ISL_4080833, EPI_ISL_4080835, EPI_ISL_4080836, EPI_ISL_4080837, EPI_ISL_4080838, EPI_ISL_4080840, EPI_ISL_4080842, EPI_ISL_4080844, EPI_ISL_4080845, EPI_ISL_4080846, EPI_ISL_4080853, EPI_ISL_4080854, EPI_ISL_4080859, EPI_ISL_4080860, EPI_ISL_4080863, EPI_ISL_4080867, EPI_ISL_4080868, EPI_ISL_4080872, EPI_ISL_4080875, EPI_ISL_4080876, EPI_ISL_4080877, EPI_ISL_4080878, EPI_ISL_4080879, EPI_ISL_4080880, EPI_ISL_4080887, EPI_ISL_4080889, EPI_ISL_4080891, EPI_ISL_4080892, EPI_ISL_4080895, EPI_ISL_4081028, EPI_ISL_4081068 | | | |
| see above | Laboratorio Central de Saude Publica do Estado de Goias (LACEN/GO) | Laboratory of Respiratory Viruses and Measles, Oswaldo Cruz Institute, FIOCRUZ | Agatha Cristinne Prudencio; Alice Sampaio Rocha; Ana Carolina Mendonca; Ana Flavia Mendonça; Anna Carolina Paixao; Carmen Helena Ramos; Cassiane Casanova; Elisa Cavalcante Pereira; Fernando Motta; Flavia Pereira Amorim da Silva; Ighor Leonardo Arantes Gomes; Luciana Appolinario; Luiz Augusto Pereira; Marilda Siqueira on behalf of the Fiocruz COVID-19 Genomic Surveillance Network; Paola Resende; Rafael Souza Guedes; Renata Serrano Lopes; Taina Venas; Vinicius Lemes da Silva |
| EPI_ISL_3539239, EPI_ISL_3539240, EPI_ISL_3802966, EPI_ISL_4061289, EPI_ISL_4061290, EPI_ISL_4061291, EPI_ISL_4061293, EPI_ISL_4061295 | | | |
| see above | Laboratorio Central de Saude Publica do Estado do Para (LACEN/PA) | Laboratory of Respiratory Viruses and Measles, Oswaldo Cruz Institute, FIOCRUZ | Agatha Soares; Alice Sampaio Rocha; Ana Carolina Mendonca; Anna Carolina Paixao; Elisa Cavalcante Pereira; Fernando Motta; Ighor Arantes; Luciana Appolinario; Marilda Siqueira on behalf of the Fiocruz COVID-19 Genomic Surveillance Network; Paola Resende; Renata Serrano Lopes; Taina Venas; Valnete Andrade |
| EPI_ISL_3245245, EPI_ISL_3245246, EPI_ISL_3245247, EPI_ISL_3245248, EPI_ISL_3245257, EPI_ISL_3245258, EPI_ISL_3245259, EPI_ISL_3245260, EPI_ISL_3245262, EPI_ISL_3245267, EPI_ISL_3245270, EPI_ISL_3245271, EPI_ISL_3245274, EPI_ISL_3245275, EPI_ISL_3245277, EPI_ISL_3245278, EPI_ISL_3245279, EPI_ISL_3245282, EPI_ISL_3245283, EPI_ISL_3245284, EPI_ISL_3245285, EPI_ISL_3245289, EPI_ISL_3245290, EPI_ISL_3245291, EPI_ISL_3245293, EPI_ISL_3245298, EPI_ISL_3245301, EPI_ISL_3245302, EPI_ISL_3245304, EPI_ISL_3245306, EPI_ISL_3245307, EPI_ISL_3245308, EPI_ISL_3245309, EPI_ISL_3245310, EPI_ISL_3245311, EPI_ISL_3245312, EPI_ISL_3245323, EPI_ISL_3245324, EPI_ISL_3245326, EPI_ISL_3245327, EPI_ISL_3245328, EPI_ISL_3245329, EPI_ISL_3245330, EPI_ISL_3245333, EPI_ISL_3245336, EPI_ISL_3615150, EPI_ISL_3615151, EPI_ISL_3615152, EPI_ISL_3615153, EPI_ISL_3615154, EPI_ISL_3615155, EPI_ISL_3615156, EPI_ISL_3615157, EPI_ISL_3615158, EPI_ISL_3615159, EPI_ISL_3615160, EPI_ISL_3615166, EPI_ISL_3615167, EPI_ISL_3615168, EPI_ISL_3615169, EPI_ISL_3615170, EPI_ISL_3615171, EPI_ISL_3615172, EPI_ISL_3615173, EPI_ISL_3615174, EPI_ISL_3615175, EPI_ISL_3615176, EPI_ISL_3615178, EPI_ISL_3615179, EPI_ISL_3615180, EPI_ISL_3615181, EPI_ISL_3615182, EPI_ISL_3615183, EPI_ISL_3615184, EPI_ISL_3615185, EPI_ISL_3615186, EPI_ISL_3615187, EPI_ISL_3615188, EPI_ISL_3615189, EPI_ISL_3615191, EPI_ISL_3615192, EPI_ISL_3615193, EPI_ISL_3615194, EPI_ISL_3615195, EPI_ISL_3615196, EPI_ISL_3615197, EPI_ISL_3615199, EPI_ISL_3615200, EPI_ISL_3615201, EPI_ISL_3615202, EPI_ISL_3615235, EPI_ISL_3615237, EPI_ISL_3615238, EPI_ISL_3920871, EPI_ISL_3920895, EPI_ISL_3920952, EPI_ISL_3920967, EPI_ISL_3920976, EPI_ISL_3920982, EPI_ISL_3920987, EPI_ISL_3921008, EPI_ISL_3921026, EPI_ISL_3921114, EPI_ISL_3921120, EPI_ISL_3921158, EPI_ISL_3921178, EPI_ISL_3921217, EPI_ISL_3921253 | | | |
| see above | Laboratorio Central Noel Nutels | Bioinformatics Laboratory / LNCC | Alessandra P Lamarca; Alexandra L Gerber; Amilcar Tanuri; Ana Paula de C Guimaraes; Ana Tereza R Vasconcelos; Andrea Cony Cavalcanti; Caio Luiz Pereira Ribeiro; Cintia Policarpo; Claudia Maria Braga de Mello; Cristiane Gomes da Silva; Douglas Terra Machado; Erica Ramos dos Santos Nascimento; Fernanda Leitao dos Santos; Flavio Dias da Silva; Gleidson da Silva de Oliveira; Leandro Magalhaes de Souza; Liliane Cavalcante; Luiz G P de Almeida; Marcio Henrique de Oliveira Garcia; Mario Sergio Ribeiro; Ricardo Jose Barbosa Salviano; Ronaldo da Silva F Jr; Silvia Carvalho |
| EPI_ISL_3805733, EPI_ISL_3805734 | Laboratorio Central de Epidemiologia (LCE) | Unidad de Genomica Avanzada | ; Alejandra Garcia-Gasca; Alejandra Hernandez-Teran; Alejandro Sanchez-Flores; Alfredo Herrera-Estrella; Alicia Ocaña-Mondragon; Andreu Comas-Garcia; Angel Gustavo Salas-Lais; Antonio Loza Roman; Bernardo Martinez-Miguel; Blanca Taboada; Brenda Irasema Maldonado-Meza; Bruno Gomez-Gil; Carla Ivon Herrera-Najera; Carlos F. Arias; Celia Boukadida; Celida Duque Molina; Celida Martinez- Rodriguez; Clara Esperanza Santacruz-Tinoco; Concepcion Grajales-Muñiz; Consorcio Mexicano de Vigilancia Genomica (CoViGen-Mex). Authors (in alphabetical order): Julio Elias Alvarado-Yaah; Cristobal Chaidez-Quiroz; Daniel Fregoso-Rueda; Daniel Lira Morales; Eduardo Becerril-Vargas; Fernando Fontove-Herrera; Fidencio Mejia-Nepomuceno; Francisco Pulido; Gloria Elena Espinosa-Ayala; Gloria Maria Molina-Salinas; Gloria Vazquez; Hector Esteban Paz-Juarez; Hector Montoya-Fuentes; Helen Haydee Fernanda Ramirez-Plascencia; Irvin Gonzalez-Lopez; Jean Pierre Gonzalez; Jesus Hernandez; Joel Armando Vazquez-Perez.; Jorge Salas-Hernandez; Jose Antonio Enciso-Moreno; Jose Arturo Martinez-Orozco; Jose Esteban Muñoz-Medina; Jose de Jesus Nuñez-Contreras; Juan Bautista Chale-Dzul; Julissa Enciso-Ibarra; Luis Alberto Ochoa-Carrera; Margarita Matias-Florentino; Maria Guadalupe Santiago-Mauricio; Maria Guadalupe de Jesus Mireles-Rivera; Mario Mujica-Sanchez; Marissa Perez-Garcia; Nelly Selem-Mojica; Pavel Isa; Ricardo Ciria Merce; Ricardo Grande; Rosa Maria Gutierrez Rios; Santiago avila-Rios; Selene Zarate; Susana Lopez; Veronica Mata-Haro; Victor Eduardo Garcia-Arias; Victor Hugo Borja-Aburto |
| EPI_ISL_3557087 | Laboratorio Central de Epidemiología (LCE) | Centro de Investigación en Enfermedades Infecciosas (CIENI), Instituto Nacional de Enfermedades Respiratorias (INER) | Alejandra García-Gasca; Alejandra Hernández-Terán; Alejandro Sánchez-Flores; Alfredo Herrera-Estrella; Alicia Ocaña-Mondragón; Andreu Comas-García; Angel Gustavo Salas-Lais; Antonio Loza Román; Bernardo Martínez-Miguel; Blanca Taboada; Brenda Irasema Maldonado-Meza; Bruno Gómez-Gil; Carla Ivón Herrera-Najera; Carlos F. Arias; Celia Boukadida; Clara Esperanza Santacruz-Tinoco; Concepción Grajales-Muñiz; Consorcio Mexicano de Vigilancia Genómica (CoViGen-Mex). Authors (in alphabetical order): Julio Elias Alvarado-Yaah; Cristóbal Cháidez-Quiróz; Célida Duque Molina; Célida Martínez- Rodríguez; Daniel Fregoso-Rueda; Daniel Lira Morales; Eduardo Becerril-Vargas; Fernando Fontove-Herrera; Fidencio Mejía-Nepomuceno; Francisco Pulido; Gloria Elena Espinosa-Ayala; Gloria María Molina-Salinas; Gloria Vazquez; Hector Esteban Paz-Juárez; Hector Montoya-Fuentes; Helen Haydee Fernanda Ramírez-Plascencia; Irvin González-López; Jean Pierre González; Jesús Hernández; Joel Armando Vázquez-Pérez.; Jorge Salas-Hernández; José Antonio Enciso-Moreno; José Arturo Martínez-Orozco; José Esteban Muñoz-Medina; José de Jesús Nuñez-Contreras; Juan Bautista Chale-Dzul; Julissa Enciso-Ibarra; Kathia Elizabeth Tapia-Diaz; Luis Alberto Ochoa-Carrera; Margarita Matías-Florentino; Mario Mújica-Sánchez; Marissa Perez-Garcia; María Guadalupe Santiago-Mauricio; María Guadalupe de Jesús Mireles-Rivera; Nelly Sélem-Mojica; Pavel Isa; Ricardo Ciria Merce; Ricardo Grande; Rosa María Gutiérrez Rios; Santiago Ávila-Ríos; Selene Zárate; Susana Lopez; Verónica Mata-Haro; Victor Eduardo García-Arias; Víctor Hugo Borja-Aburto |
| EPI_ISL_3347549, EPI_ISL_3347559 | Laboratorio Central de Epidemiología (LCE) | Instituto de Biotecnología de la UNAM | ; Alejandra García-Gasca; Alejandra Hernández-Terán; Alejandro Sánchez-Flores; Alfredo Herrera-Estrella; Alicia Ocaña-Mondragón; Andreu Comas-García; Angel Gustavo Salas-Lais; Antonio Loza Román; Bernardo Martínez-Miguel; Blanca Taboada; Brenda Irasema Maldonado-Meza; Bruno Gómez-Gil; Carla Ivón Herrera-Najera; Carlos F. Arias; Celia Boukadida; Clara Esperanza Santacruz-Tinoco; Concepción Grajales-Muñiz; Consorcio Mexicano de Vigilancia Genómica (CoViGen-Mex). Authors (in alphabetical order): Julio Elias Alvarado-Yaah; Cristóbal Cháidez-Quiróz; Célida Duque Molina; Célida Martínez- Rodríguez; Daniel Fregoso-Rueda; Daniel Lira Morales; Eduardo Becerril-Vargas; Fernando Fontove-Herrera; Fidencio Mejía-Nepomuceno; Francisco Pulido; Gloria Elena Espinosa-Ayala; Gloria María Molina-Salinas; Gloria Vazquez; Hector Esteban Paz-Juárez; Hector Montoya-Fuentes; Helen Haydee Fernanda Ramírez-Plascencia; Irvin González-López; Jean Pierre González; Jesús Hernández; Joel Armando Vázquez-Pérez).; Jorge Salas-Hernández; José Antonio Enciso-Moreno; José Arturo Martínez-Orozco; José Esteban Muñoz-Medina; José de Jesús Nuñez-Contreras; Juan Bautista Chale-Dzul; Julissa Enciso-Ibarra; Kathia Elizabeth Tapia-Diaz; Luis Alberto Ochoa-Carrera; Margarita Matías-Florentino; Mario Mújica-Sánchez; Marissa Perez-Garcia; María Guadalupe Santiago-Mauricio; María Guadalupe de Jesús Mireles-Rivera; Nelly Sélem-Mojica; Pavel Isa; Ricardo Ciria Merce; Ricardo Grande; Rosa María Gutiérrez Rios; Santiago Ávila-Ríos; Selene Zárate; Susana Lopez; Verónica Mata-Haro; Victor Eduardo García-Arias; Víctor Hugo Borja-Aburto |

| | | | |
|---|---|---|---|
| EPI_ISL_3825480 | Laboratorio Central de Salud Publica | Laboratorio de Biología Molecular, Instituto de Medicina Regional on behalf of 'Proyecto Argentino Interinstitucional de genomica de SARS-CoV-2' (PAIS Consortium) | Andrea Ayala; Bettina Brusés; Erica Struss; Esteban Paredes; Griselda Oria; Horacio Lucero.; Javier Mussin; Laura Formichelli; Melina Lorenzini Campos; Raúl Maximiliano Acevedo; Verónica Gómez; Victoria Femenías |
| EPI_ISL_3235235, EPI_ISL_3235236, EPI_ISL_3235237, EPI_ISL_4061421, EPI_ISL_4061422, EPI_ISL_4061423 | Laboratorio Central de Saude Publica do Estado da Bahia (LACEN/BA) | Laboratory of Respiratory Viruses and Measles, Oswaldo Cruz Institute, FIOCRUZ | Agatha Soares; Alice Sampaio Rocha; Ana Carolina Mendonca; Anna Carolina Paixao; Elisa Cavalcante Pereira; Felicidade Pereira; Fernando Motta; Ighor Arantes; Luciana Appolinario; Marilda Siqueira on behalf of the Fiocruz COVID-19 Genomic Surveillance Network; Paola Resende; Renata Serrano Lopes; Taina Venas |
| EPI_ISL_3235244, EPI_ISL_3434759, EPI_ISL_3434760, EPI_ISL_3434761, EPI_ISL_3434766, EPI_ISL_3434769, EPI_ISL_3434770, EPI_ISL_3434779, EPI_ISL_3434780, EPI_ISL_3434781, EPI_ISL_3434782, EPI_ISL_3434789, EPI_ISL_3832397, EPI_ISL_4081069, EPI_ISL_4081070, EPI_ISL_4081071, EPI_ISL_4081072, EPI_ISL_4081073, EPI_ISL_4081074, EPI_ISL_4081075, EPI_ISL_4081076, EPI_ISL_4081077, EPI_ISL_4081078, EPI_ISL_4081079, EPI_ISL_4081080, EPI_ISL_4081081, EPI_ISL_4081082, EPI_ISL_4081083, EPI_ISL_4081084, EPI_ISL_4081086, EPI_ISL_4081088, EPI_ISL_4081089 | | | |
| see above | Laboratorio Central de Saude Publica do Estado da Paraiba (LACEN/PB) | Laboratory of Respiratory Viruses and Measles, Oswaldo Cruz Institute, FIOCRUZ | Alice Sampaio Rocha; Ana Carolina Mendonca; Anna Carolina Paixao; Dalane Loudal Florentino Teixeira; Elisa Cavalcante Pereira; Fernando Motta; Joao Felipe Bezerra; Luciana Appolinario; Marilda Siqueira on behalf of the Fiocruz COVID-19 Genomic Surveillance Network; Paola Resende; Renata Serrano Lopes; Taina Venas |
| EPI_ISL_3434820, EPI_ISL_3434844, EPI_ISL_3434845, EPI_ISL_3434846, EPI_ISL_3434847, EPI_ISL_3434848, EPI_ISL_3434894, EPI_ISL_3434896, EPI_ISL_3434943, EPI_ISL_3802904, EPI_ISL_3802905, EPI_ISL_3802906, EPI_ISL_3802907, EPI_ISL_3802908, EPI_ISL_3802909, EPI_ISL_3802910, EPI_ISL_3802911, EPI_ISL_3802912, EPI_ISL_3802913, EPI_ISL_3802914, EPI_ISL_3802915, EPI_ISL_3802916, EPI_ISL_3802917, EPI_ISL_3802918, EPI_ISL_3802919, EPI_ISL_3802921, EPI_ISL_3802922, EPI_ISL_3802923, EPI_ISL_3802924, EPI_ISL_3802925, EPI_ISL_3802926, EPI_ISL_3802927, EPI_ISL_3802929, EPI_ISL_3802930, EPI_ISL_3802931, EPI_ISL_3802932, EPI_ISL_4061393, EPI_ISL_4061394, EPI_ISL_4061395, EPI_ISL_4061396, EPI_ISL_4061397, EPI_ISL_4061398, EPI_ISL_4061399, EPI_ISL_4061400, EPI_ISL_4061401, EPI_ISL_4061402, EPI_ISL_4061403, EPI_ISL_4061404, EPI_ISL_4061405, EPI_ISL_4061407, EPI_ISL_4061409, EPI_ISL_4061410, EPI_ISL_4061411, EPI_ISL_4061412, EPI_ISL_4061413, EPI_ISL_4061415, EPI_ISL_4061417, EPI_ISL_4061418, EPI_ISL_4061419, EPI_ISL_4061420 | | | |
| see above | Laboratorio Central de Saude Publica do Estado de Alagoas (LACEN/AL) | Laboratory of Respiratory Viruses and Measles, Oswaldo Cruz Institute, FIOCRUZ | Agatha Soares; Alice Sampaio Rocha; Ana Carolina Mendonca; Anderson Brandao Leite; Anna Carolina Paixao; Elisa Cavalcante Pereira; Fernando Motta; Ighor Arantes; Luciana Appolinario; Marilda Siqueira on behalf of the Fiocruz COVID-19 Genomic Surveillance Network; Paola Resende; Renata Serrano Lopes; Rubens Pasa; Taina Venas |
| EPI_ISL_3434792, EPI_ISL_3434793, EPI_ISL_3434794, EPI_ISL_3434796, EPI_ISL_3434797, EPI_ISL_3434800, EPI_ISL_3434918, EPI_ISL_3434919, EPI_ISL_3434920, EPI_ISL_3434921, EPI_ISL_3434922, EPI_ISL_3434923, EPI_ISL_3434924, EPI_ISL_3435054, EPI_ISL_3435055, EPI_ISL_3435056, EPI_ISL_3435057, EPI_ISL_3435058, EPI_ISL_3435061, EPI_ISL_3435062, EPI_ISL_3435063, EPI_ISL_3435064, EPI_ISL_3435066, EPI_ISL_3539245, EPI_ISL_3539772, EPI_ISL_3539773, EPI_ISL_3827990, EPI_ISL_3828012, EPI_ISL_3828016 | | | |
| see above | Laboratorio Central de Saude Publica do Estado de Santa Catarina (LACEN/SC) | Laboratory of Respiratory Viruses and Measles, Oswaldo Cruz Institute, FIOCRUZ | Alice Sampaio Rocha; Ana Carolina Mendonca; Anna Carolina Paixao; Darcita Buerger Rovaris; Elisa Cavalcante Pereira; Fernando Motta; Luciana Appolinario; Marilda Siqueira on behalf of the Fiocruz COVID-19 Genomic Surveillance Network; Paola Resende; Renata Serrano Lopes; Sandra Bianchini Fernandes; Taina Venas |
| EPI_ISL_4061449, EPI_ISL_4061450 | Laboratorio Central de Saude Publica do Estado de Sergipe (LACEN/SE) | Laboratory of Respiratory Viruses and Measles, Oswaldo Cruz Institute, FIOCRUZ | Agatha Soares; Alice Sampaio Rocha; Ana Carolina Mendonca; Anna Carolina Paixao; Cliomar Alves dos Santos; Elisa Cavalcante Pereira; Fernando Motta; Ighor Arantes; Luciana Appolinario; Marilda Siqueira on behalf of the Fiocruz COVID-19 Genomic Surveillance Network; Paola Resende; Renata Serrano Lopes; Tainá Moreira Martins Venas |
| EPI_ISL_3539795, EPI_ISL_3539796, EPI_ISL_3539797, EPI_ISL_3539798, EPI_ISL_3539799, EPI_ISL_3539800, EPI_ISL_3539801, EPI_ISL_3539802, EPI_ISL_3539803, EPI_ISL_3539804, EPI_ISL_3539805, EPI_ISL_3539806, EPI_ISL_3539807, EPI_ISL_3539808, EPI_ISL_3539809, EPI_ISL_3539811, EPI_ISL_3539813, EPI_ISL_3539814, EPI_ISL_3539815, EPI_ISL_3539816, EPI_ISL_3539817, EPI_ISL_3539818, EPI_ISL_3539819, EPI_ISL_3539820, EPI_ISL_3539821, EPI_ISL_3539822, EPI_ISL_3539823, EPI_ISL_3539824, EPI_ISL_3539825, EPI_ISL_3539826, EPI_ISL_3539828, EPI_ISL_4081029, EPI_ISL_4081030, EPI_ISL_4081031, EPI_ISL_4081033, EPI_ISL_4081034, EPI_ISL_4081035, EPI_ISL_4081036, EPI_ISL_4081037, EPI_ISL_4081038, EPI_ISL_4081039, EPI_ISL_4081040, EPI_ISL_4081041, EPI_ISL_4081042, EPI_ISL_4081044, EPI_ISL_4081046, EPI_ISL_4081047, EPI_ISL_4081049, EPI_ISL_4081050, EPI_ISL_4081052, EPI_ISL_4081053, EPI_ISL_4081054, EPI_ISL_4081056, EPI_ISL_4081057, EPI_ISL_4081058, EPI_ISL_4081059, EPI_ISL_4081060, EPI_ISL_4081061, EPI_ISL_4081062, EPI_ISL_4081063, EPI_ISL_4081065, EPI_ISL_4081066 | | | |
| see above | Laboratorio Central de Saude Publica do Estado do Amapa (LACEN/AP) | Laboratory of Respiratory Viruses and Measles, Oswaldo Cruz Institute, FIOCRUZ | Agatha Cristinne Prudencio; Alice Sampaio Rocha; Ana Carolina Mendonca; Andreia Santos Costa; Anna Carolina Paixao; Anne Caroline da Silva Soledade; Elisa Cavalcante Pereira; Fernando Motta; Ighor Leonardo Arantes Gomes; Lindomar dos Anjos Silva; Luciana Appolinario; Marcia Socorro Pereira Cavalcante; Marilda Siqueira on behalf of the Fiocruz COVID-19 Genomic Surveillance Network; Paola Resende; Renata Serrano Lopes; Taina Venas |
| EPI_ISL_3434721, EPI_ISL_3434729, EPI_ISL_3434730, EPI_ISL_3434732, EPI_ISL_3434733, EPI_ISL_3434734, EPI_ISL_3434735, EPI_ISL_3434736, EPI_ISL_3434737, EPI_ISL_3434738, EPI_ISL_3434739, EPI_ISL_3434740, EPI_ISL_3434741, EPI_ISL_3434744, EPI_ISL_3434745, EPI_ISL_3434747, EPI_ISL_3434748, EPI_ISL_3434951, EPI_ISL_3434989, EPI_ISL_3434990, EPI_ISL_3434992, EPI_ISL_3434993, EPI_ISL_3434994, EPI_ISL_3434999, EPI_ISL_3435000, EPI_ISL_3435001, EPI_ISL_3435002, EPI_ISL_3435003, EPI_ISL_3435004, EPI_ISL_3435006, EPI_ISL_3435007, EPI_ISL_3435009, EPI_ISL_3435010, EPI_ISL_3435012, EPI_ISL_3435017, EPI_ISL_3435018, EPI_ISL_3435020, EPI_ISL_3435021, EPI_ISL_3435022, EPI_ISL_3435023, EPI_ISL_3435026, EPI_ISL_3435027, EPI_ISL_3435028, EPI_ISL_3435033, EPI_ISL_3435034, EPI_ISL_3435036, EPI_ISL_3435040, EPI_ISL_3435041, EPI_ISL_3435068, EPI_ISL_3539933, EPI_ISL_3539934, EPI_ISL_3827993, EPI_ISL_3828001, EPI_ISL_3828009, EPI_ISL_3828014, EPI_ISL_4080765 | | | |
| see above | Laboratorio Central de Saude Publica do Estado do Espirito Santo (LACEN/ES) | Laboratory of Respiratory Viruses and Measles, Oswaldo Cruz Institute, FIOCRUZ | Alice Sampaio Rocha; Ana Carolina Mendonca; Anna Carolina Paixao; Eilisa Cavalcante Pereira; Elisa Cavalcante Pereira; Fernando Motta; Luciana Appolinario; Marilda Siqueira on behalf of the Fiocruz COVID-19 Genomic Surveillance Network; Paola Resende; Renata Serrano Lopes; Rodrigo Ribeiro Rodrigues; Taina Venas |
| EPI_ISL_3914061, EPI_ISL_3914069, EPI_ISL_3914078, EPI_ISL_3914099, EPI_ISL_3914116, EPI_ISL_3914138, EPI_ISL_3914150, EPI_ISL_3914202, EPI_ISL_3914219, EPI_ISL_3914249, EPI_ISL_3914254, EPI_ISL_3914291, EPI_ISL_3914331, EPI_ISL_3914334, EPI_ISL_3914339 | | | |
| see above | Laboratorio Central de Saude Publica do Estado do Parana (Instituto de Biologia Molecular do Paraná (LACEN-PR) | Instituto Carlos Chagas - Fiocruz | Alessandra De Melo Aguiar; Andreia Akemi Suzukawa; Andréa Rodrigues Ávila; Bruno Dallagiovanna; Dalila Zanette; Eduardo Balsanelli; Emanuel Maltempi de Souza; Fabio Passetti; Fabricio Klerynton Marchini; Fábio de Oliveira Pedrosa; Guilherme Becker; Helisson Faoro; Hellen Geremias dos Santos; Irina Nastassja Riediger; Letusa Albrecht; Lucas Blanes; Luis Gustavo Morello; Lysangela Ronalte Alves; Maria do Carmo Debur; Mauro de Medeiros Oliveira; Michelle Orane Schemberger; Paola Cristina Resende; Sheila Cristina Nardeli; Tiago Gräf; Valter Antônio de Baura |
| EPI_ISL_4061330, EPI_ISL_4061424 | Laboratorio Central de Saude Publica do Estado do Parana (LACEN/PR) | Laboratory of Respiratory Viruses and Measles, Oswaldo Cruz Institute, FIOCRUZ | Agatha Soares; Alice Sampaio Rocha; Ana Carolina Mendonca; Anna Carolina Paixao; Elisa Cavalcante Pereira; Fernando Motta; Ighor Leonardo Arantes; Irina Riediger; Luciana Appolinario; Marilda Siqueira on behalf of the Fiocruz COVID-19 Genomic Surveillance Network; Paola Resende; Renata Serrano Lopes; Taina Venas |
| EPI_ISL_3235268, EPI_ISL_3235270, EPI_ISL_3235271, EPI_ISL_3539831, EPI_ISL_3539832, EPI_ISL_3539834, EPI_ISL_3539837, EPI_ISL_3539838, EPI_ISL_3539839, EPI_ISL_3832401, EPI_ISL_3832403, EPI_ISL_3832404, EPI_ISL_3832405, EPI_ISL_3832406, EPI_ISL_3832407, EPI_ISL_3832408, EPI_ISL_3832409 | | | |
| see above | Laboratorio Central de Saude Publica do Estado do Rio Grande do Norte (LACEN/RN) | Laboratory of Respiratory Viruses and Measles, Oswaldo Cruz Institute, FIOCRUZ | Agatha Soares; Alice Sampaio Rocha; Ana Carolina Mendonca; Ana Paula Ferreira Costa; Anna Carolina Paixao; Antonnyo Palmielly Diogenes Lima; Aurélio de Oliveira Bento; Elisa Cavalcante Pereira; Fernando Motta; Gessika Brenna Costa Alves; Heglayne Pereira Vital da Silva; Iago de Souza Gomes; Ighor Arantes; Isabelle Cristina Clemente dos Santos; Janaina Sonale Cavalcante Nogueira de Oliveira; Jayra Juliana Paiva Alves Abrantes; Jonas José da Silva; Luciana Appolinario; Marilda Siqueira on behalf of the Fiocruz COVID-19 Genomic Surveillance Network; Paola Resende; Renata Serrano Lopes; Taina Venas; Themis Rocha de Souza; Vítor Gabriel Saldanha Fernandes |
| EPI_ISL_3190298, EPI_ISL_3190300, EPI_ISL_3190301, EPI_ISL_3190306, EPI_ISL_3190307, EPI_ISL_3190308, EPI_ISL_3235231, EPI_ISL_3235232, EPI_ISL_3235233, EPI_ISL_3235234, EPI_ISL_3235276, EPI_ISL_3235277, EPI_ISL_3235279, EPI_ISL_3235284, EPI_ISL_3235285, EPI_ISL_3235286, EPI_ISL_3235287, EPI_ISL_3235288, EPI_ISL_3539922, EPI_ISL_3832417, EPI_ISL_3832418, EPI_ISL_3832419, EPI_ISL_3832420, EPI_ISL_3832421, EPI_ISL_3832422, EPI_ISL_3832423, EPI_ISL_3832424, EPI_ISL_3832425, EPI_ISL_3832426, EPI_ISL_3832428, EPI_ISL_3832429, EPI_ISL_3832431, EPI_ISL_3832432, EPI_ISL_3832433, EPI_ISL_3832434, EPI_ISL_3832435, EPI_ISL_3832436, EPI_ISL_3832437, EPI_ISL_3832438 | | | |
| see above | Laboratorio Central de Saude Publica do Estado do Rio Grande do Sul (LACEN-RS) | Laboratory of Respiratory Viruses and Measles, Oswaldo Cruz Institute, FIOCRUZ | Agatha Cristinne Soares; Alice Sampaio Rocha; Ana Carolina Mendonca; Anderson Brandao Leite; Anna Carolina Paixao; Elisa Cavalcante Pereira; Fernando Motta; Ighor Leonardo Arantes; Luciana Appolinario; Marilda Siqueira on behalf of the Fiocruz COVID-19 Genomic Surveillance Network; Paola Resende; Renata Serrano Lopes; Richard Salvato; Taina Venas; Tatiana Schaffer Gregianini |
| EPI_ISL_3235260, EPI_ISL_3235261, EPI_ISL_3235263, EPI_ISL_3235266, EPI_ISL_3434790, EPI_ISL_3434941, EPI_ISL_3539766, EPI_ISL_4061336, EPI_ISL_4080788 | | | |
| see above | Laboratorio Central de Saude Publica do Estado do Rio de Janeiro (LACEN/RJ) | Laboratory of Respiratory Viruses and Measles, Oswaldo Cruz Institute, FIOCRUZ | Agatha Cristinne Prudencio Soares; Agatha Soares; Alice Sampaio Rocha; Ana Carolina Mendonca; Andrea Cony Cavalcanti; Anna Carolina Paixao; Elisa Cavalcante Pereira; Fernando Motta; Ighor Arantes; Ighor Leonardo Arantes Gomes; Luciana Appolinario; Marilda Siqueira on behalf of the Fiocruz COVID-19 Genomic Surveillance Network; Paola Resende; Renata Serrano Lopes; Taina Venas |
| EPI_ISL_4080800, EPI_ISL_4080801 | Laboratorio Central de Saude Publica do Estado do Tocantins (LACEN/TO) | Laboratory of Respiratory Viruses and Measles, Oswaldo Cruz Institute, FIOCRUZ | Agatha Soares; Alice Sampaio Rocha; Ana Carolina Mendonca; Anna Carolina Paixao; Elisa Cavalcante Pereira; Fernando Motta; Ighor Arantes; Jucimaria Dantas Galvao; Luciana Appolinario; Marilda Siqueira on behalf of the Fiocruz COVID-19 Genomic Surveillance Network; Paola Resende; Renata Serrano Lopes; Taina Venas |
| EPI_ISL_4003464 | Laboratorio Central, Ministerio de Salud Córdoba | Instituto de Patologia Vegetal (CIAP-INTA) on behalf of 'Proyecto Argentino Interinstitucional de genomica de SARS-CoV-2' (PAIS Consortium) | Barbas, G.; Castro, G.; Debat, HJ.; FD; Fernández; M; M.B.; Marquez, N.; Pisano; Re, V. |
| EPI_ISL_3527817, EPI_ISL_3527818, EPI_ISL_4051092 | Laboratorio Nacional de Salud | Asociación de Salud Integral / Clínica Familiar "Luis Ángel García" | Ana S. Gonzalez-Reiche; Claudia Rangel; Danicela Mercado; Eduardo Arathon; Hilda Ruiz; Luis Aguirre; Luis Rivas; Narda Medina; Oscar Bonilla; Osmar Gamboa. |
| EPI_ISL_4029327 | Laboratorio Nacional de Salud, Ministerio de Salud Publica y Asistencia Social | Genomics and Proteomics Department, Gorgas Memorial Institute For Health Studies | Alexander A Martinez; Ambar Moreno; Claudia Estrada; Claudia Gonzalez V; César Roberto Conde Pereira; Jessica Gondola; Leyda Abrego; Marlene Castillo; Melissa Gaitan; Oris Chavarria |
| EPI_ISL_3707440 | Laboratorio de Genómica Microbiana, Universidad Peruana Cayetano Heredia | cov0937 | Alejandra Dávila-Barclay; Diego Cuicapuza; Guillermo Salvatierra; Janet Huancachoque; Luis González; Pablo Tsukayama; Pedro E. Romero; Pool Marcos |
| EPI_ISL_3535744, EPI_ISL_3535754, EPI_ISL_3642589, EPI_ISL_3642591, EPI_ISL_3758927, EPI_ISL_3758932, EPI_ISL_3758949, EPI_ISL_3758953, EPI_ISL_3758955, EPI_ISL_3944414 | | | |
| see above | Laboratorio de Infectologia y Virologia Molecular | Laboratory of Molecular Virology, School of Medicine, Pontificia Universidad Catolica de Chile | Ana Maria Contreras; Andres E. Munoz-Marcos; Carlos Palma; Catalina Pardo-Roa; Constanza Maldonado; Constanza Martinez-Valdevenito; Eileen Serrano; Erick Salinas; Estefany Poblete; Francisco Melo; Jennifer Angulo; Jorge Levican; Leonardo I. Almonacid; M. Belen Leyton; Marcela Ferres; Maria Jose Avendano; Rafael A. Medina; Tamara Garcia-Salum |
| EPI_ISL_3134280, EPI_ISL_3134281 | Laboratorio de Medicina Molecular, Universidad de Magallanes | Centro Asistencial Docente y de Investigacion, Universidad de Magallanes | Carolina Pérez; Constanza Ceroni; Diego Alvarez; Inés Cid; Jacqueline Aldridge; Jorge González; Juan Ríos; Marcelo Navarrete |

| | | | |
|---|---|---|---|
| EPI_ISL_3761574, EPI_ISL_3761579, EPI_ISL_3761580, EPI_ISL_3761581, EPI_ISL_3761582, EPI_ISL_3761585, EPI_ISL_3761591, EPI_ISL_3761592, EPI_ISL_3761594, EPI_ISL_3761595, EPI_ISL_3761596, EPI_ISL_3761598, EPI_ISL_3761601, EPI_ISL_3761606, EPI_ISL_3761609, EPI_ISL_3761610, EPI_ISL_3761611, EPI_ISL_3761612, EPI_ISL_3761620, EPI_ISL_3761622, EPI_ISL_3761624, EPI_ISL_3761627, EPI_ISL_3761628, EPI_ISL_3761630, EPI_ISL_3761632, EPI_ISL_3761704, EPI_ISL_3761706, EPI_ISL_3761707, EPI_ISL_3761708, EPI_ISL_3761709, EPI_ISL_3761710, EPI_ISL_3761711, EPI_ISL_3761712, EPI_ISL_3761713, EPI_ISL_3761714, EPI_ISL_3761715, EPI_ISL_3761717, EPI_ISL_3761718, EPI_ISL_3761719, EPI_ISL_3761720, EPI_ISL_3761721, EPI_ISL_3761722, EPI_ISL_3761723, EPI_ISL_3761725, EPI_ISL_3761726, EPI_ISL_3761727, EPI_ISL_3761728, EPI_ISL_3761729, EPI_ISL_3761730, EPI_ISL_3761731, EPI_ISL_3761732, EPI_ISL_3761733, EPI_ISL_3761734, EPI_ISL_3761735, EPI_ISL_3761737, EPI_ISL_3761738, EPI_ISL_3761739, EPI_ISL_3761741, EPI_ISL_3761742, EPI_ISL_3761743, EPI_ISL_3761744, EPI_ISL_3761745, EPI_ISL_3761746, EPI_ISL_3761747, EPI_ISL_3761748, EPI_ISL_3761764 | | | |
| see above | Laboratorio de Pesquisa em Virologia, FAMERP, SJRP | Laboratorio de Pesquisa em Virologia, FAMERP, SJRP | Beatriz de Carvalho Marques; Cecília Artico Banho; Cíntia Bittar; Fábio Sossai Possebon; Guilherme Campos; Helena Lage Ferreira; Jorge A. Petroli Marchesi; João Pessoa Araújo Jr.; Leila Sabrina Ullmann; Lívia Sacchetto; Maisa C. Pereira Parra; Marília Moraes; Maurício L. Nogueira.; Paula Rahal; Paulo Inacio da Costa |
| EPI_ISL_3128480, EPI_ISL_3128489, EPI_ISL_3128514 | Laboratorio di Riferimento Regionale della Sicilia Occidentale per l'Emergenza COVID-19 | Laboratorio di Riferimento Regionale della Sicilia Occidentale per l'Emergenza COVID-19 | Carmelo Massimo Maida; Claudio Costantino; Daniela Di Naro; Fabio Tramuto; Francesco Vitale; Giorgio Graziano; Giulia Randazzo; Vincenzo Restivo; Walter Mazzucco |
| EPI_ISL_3112892, EPI_ISL_3112926, EPI_ISL_3112966, EPI_ISL_3326658, EPI_ISL_3327091, EPI_ISL_3327106, EPI_ISL_3327216, EPI_ISL_3327292, EPI_ISL_3327316, EPI_ISL_3327427, EPI_ISL_3327588, EPI_ISL_3327645, EPI_ISL_3327692, EPI_ISL_3327808, EPI_ISL_3327818, EPI_ISL_3327821, EPI_ISL_3328088, EPI_ISL_3328830, EPI_ISL_3328910, EPI_ISL_3329030, EPI_ISL_3329416, EPI_ISL_3329419, EPI_ISL_3329810, EPI_ISL_3330061, EPI_ISL_3330610, EPI_ISL_3430984, EPI_ISL_3431004, EPI_ISL_3509873, EPI_ISL_3511002, EPI_ISL_3511592, EPI_ISL_3511728, EPI_ISL_3512236, EPI_ISL_3512804, EPI_ISL_3512852, EPI_ISL_3513703, EPI_ISL_3514248, EPI_ISL_3514551, EPI_ISL_3514681, EPI_ISL_3514751, EPI_ISL_3515016, EPI_ISL_3515472, EPI_ISL_3516400, EPI_ISL_3516594, EPI_ISL_3517195, EPI_ISL_3517622, EPI_ISL_3517635, EPI_ISL_3517743, EPI_ISL_3517752, EPI_ISL_3517886, EPI_ISL_3517888, EPI_ISL_3517889, EPI_ISL_3517894, EPI_ISL_3518681, EPI_ISL_3518713, EPI_ISL_3518737, EPI_ISL_3518906, EPI_ISL_3518952, EPI_ISL_3519272, EPI_ISL_3519538, EPI_ISL_3519674, EPI_ISL_3519678, EPI_ISL_3519729, EPI_ISL_3519755, EPI_ISL_3520352, EPI_ISL_3606078, EPI_ISL_3606095, EPI_ISL_3606481, EPI_ISL_3607175, EPI_ISL_3607407, EPI_ISL_3677210, EPI_ISL_3677950, EPI_ISL_3678937, EPI_ISL_3679475, EPI_ISL_3680607, EPI_ISL_3682881, EPI_ISL_3685600, EPI_ISL_3685808, EPI_ISL_3686172, EPI_ISL_3686239, EPI_ISL_3686347, EPI_ISL_3686369, EPI_ISL_3686758, EPI_ISL_3687475, EPI_ISL_3687570, EPI_ISL_3688090, EPI_ISL_3740926, EPI_ISL_3740930, EPI_ISL_3740967, EPI_ISL_3744016, EPI_ISL_3744061, EPI_ISL_3744510, EPI_ISL_3746227, EPI_ISL_3746785, EPI_ISL_3748522, EPI_ISL_3749211, EPI_ISL_3749538, EPI_ISL_3749614, EPI_ISL_3750255, EPI_ISL_3750679, EPI_ISL_3750743, EPI_ISL_3751992, EPI_ISL_3752013, EPI_ISL_3752144 | | | |
| see above | Laboratory Corporation of America | Centers for Disease Control and Prevention Division of Viral Diseases, Pathogen Discovery | Adrian Paskey; Amanda Douglas; Amanda Suchanek; Andrea Throop; Ayla Burns; Benjamin Rambo-Martin; Bobbi Croy; Brian Krueger; Brian Norvell; Christopher Gulvick; Christos Petropoulos; Clinton Paden; Clinton R. Paden; Craig Lukasik; Dakota Howard; Darlene Wagner; Debbie Boles; Dhwani Batra; Duncan MacCannell; Eyad Almasri; Goran Stevovic; Howard Engler; Hrushikesh Deshmukh; Jake Humphrey; Jana Schroth; Jason Caravas; Joe Voshell; John Pruitt; Jonathan Meltzer; Jonathan Williams; Kara Moser; Kimberly Wagner; Lax Iyer; Lisa Pfefferle; Lyndon Tilson; Manoj Jain; Marcia Eisenberg; Mary Ann Cristobal; Mary Cristobal; Mary Williamson; Matthew Robinson; Matthew Schmerer; Michael Levandoski; Mike Sapeta; Mindy Nye; Minoo Agarwal; Mohan Kolli; Nuthawin Charoensri; Oren Cohen; Peter Cook; Peter W. Cook; Prashant Gupta; Qian Zeng; Rama Ghatti; Scott Parker; Scott Ryan; Scott Sammons; Shatavia Morrison; Stanley Letovsky; Steven Ragan; Suresh Babu Selvaraju; Suresh Selvaraju; Susan Countryman; Susan Hicks; Suzanne Dale; Thomas Urban; Tim Kuphal; Tricia Zwiefelhofer; Vincent Drouillon; Yvette Unoarumhi |
| EPI_ISL_3664280 | Laboratory for Molecular Diagnostics,IPHMN | Eurofins Genomics Europe Sequencing GmbH | Danijela Vujošević |
| EPI_ISL_3832400 | Laboratory of Respiratory Viruses and Measles, Oswaldo Cruz Institute, FIOCRUZ | Laboratory of Respiratory Viruses and Measles, Oswaldo Cruz Institute, FIOCRUZ | Agatha Soares; Alice Sampaio Rocha; Ana Carolina Mendonca; Anna Carolina Paixao; Elisa Cavalcante Pereira; Fernando Motta; Ighor Arantes; Luciana Appolinario; Marilda Siqueira on behalf of the Fiocruz COVID-19 Genomic Surveillance Network; Paola Resende; Renata Serrano Lopes; Taina Venas |
| EPI_ISL_3460255, EPI_ISL_3460258, EPI_ISL_3461056, EPI_ISL_3461057, EPI_ISL_3461061, EPI_ISL_3461062, EPI_ISL_3461063, EPI_ISL_3461064, EPI_ISL_3461066, EPI_ISL_3461067, EPI_ISL_3461097, EPI_ISL_3461098, EPI_ISL_3461100, EPI_ISL_3461102, EPI_ISL_3461103, EPI_ISL_3461105, EPI_ISL_3461108, EPI_ISL_3492485, EPI_ISL_3492486, EPI_ISL_3492487, EPI_ISL_3492488, EPI_ISL_3492489, EPI_ISL_3492490, EPI_ISL_3492491, EPI_ISL_3492500, EPI_ISL_3492505, EPI_ISL_3492508, EPI_ISL_3492514, EPI_ISL_3756575, EPI_ISL_3806774 | | | |
| see above | Laboratório de Microbiologia Molecular - Universidade FEEVALE | Molecular Microbiology Laboratory | Alana Witt Hansen; Fernando Rosado Spilki; Fágner Henrique Heldt; Juliana Schons Gulart; Juliana Schons Gularte; Juliane Deise Fleck; Mariana Soares da Silva; Matheus Nunes Weber; Meriane Demoliner; Michele Filippi.; Micheli Filippi; Paula Rodrigues de Almeida; Vyctoria Malayhka de Abreu Góes Pereira. |
| EPI_ISL_3529997 | Lighthouse Lab in Glasgow | Wellcome Sanger Institute for the COVID-19 Genomics UK (COG-UK) Consortium | Anna Dominiczak and Alex Alderton; Carol Clugston; Cordelia Langford; David Gray; David K. Jackson; Dominic Kwiatkowski; Ewan Harrison; Harper VanSteenhouse; Ian Johnston; Jeffrey Barrett; John Sillitoe on behalf of the Wellcome Sanger Institute COVID-19 Surveillance Team; Roberto Amato; Sonia Goncalves; Yumi Kasai |
| EPI_ISL_3221445 | Lighthouse Lab in Milton Keynes | Wellcome Sanger Institute for the COVID-19 Genomics UK (COG-UK) Consortium | Cordelia Langford; David K. Jackson; Dominic Kwiatkowski; Ewan Harrison; Ian Johnston; Jeffrey Barrett; John Sillitoe on behalf of the Wellcome Sanger Institute COVID-19 Surveillance Team; Roberto Amato; Sonia Goncalves; The Lighthouse Lab in Milton Keynes and Alex Alderton |
| EPI_ISL_3254285 | Lincoln Diagnostics | New York City Public Health Laboratory | Jade Wang; et al. |
| EPI_ISL_3536368, EPI_ISL_3912236 | MATERNIDADE ESCOLA ASSIS CHATEAUBRIAND | Analytical Competence Molecular Epidemiology Lab/ACME, Oswaldo Cruz Foundation, Ceara (FIOCRUZ CE) | Cleber Furtado Aksenen; Fabio Miyajima; Fernando Braga Stehling; Francisco Eder de Moura Lopes; Jamille Maria Mendes Bezerra; Joaquim Cesar do Nascimento Sousa Junior; Joaquim César do Nascimento Sousa Junior; Pedro Miguel Carneiro Jeronimo; Suzana Porto Almeida & Lucas Delerino on behalf of COVID-19 FIOCRUZ Genomic Network; Suzana Porto Almeida e Lucas Delerino; Thais Ferreira de Oliveira; Thais de Oliveira Costa; Ticiane Cavalcante de Souza; Veridiana Pessoa Miyajima |
| EPI_ISL_3491530, EPI_ISL_3491531 | MD DOH Laboratories Administration | Centers for Disease Control and Prevention Division of Viral Diseases, Pathogen Discovery | Alex Burgin; Ben Rambo-Martin; Clinton Paden; Dakota Howard; Dave Wentworth; Dhwani Batra; Jasmine Padilla; Justin Lee; Krista Queen; Kristen Knipe; Kristine Lacek; Mark Burroughs; Matthew Schmerer; Meghan Bentz; Mili Sheth; Peter Cook; Sam Shepard; Sarah Nobles; Suxiang Tong; Vivien Dugan; Yvette Unoarumhi |
| EPI_ISL_3241905 | MD PHL | MD PHL | Ami Patel; Eric N. Keller; Jillian Loomis; Kwang Low; Terence L. Moore; and Robert Myers |
| EPI_ISL_3152291, EPI_ISL_3235641 | MD PHL | Maryland Department of Health Laboratories Administration | Ami Patel; Eric N. Keller; Jillian Loomis; Kwang Low; Terence L. Moore; and Robert Myers |
| EPI_ISL_3432935, EPI_ISL_3432936, EPI_ISL_3432937, EPI_ISL_3432938, EPI_ISL_3432939, EPI_ISL_3432940, EPI_ISL_3432941, EPI_ISL_3432942, EPI_ISL_3432943, EPI_ISL_3432944, EPI_ISL_3432945, EPI_ISL_3432946, EPI_ISL_3432947, EPI_ISL_3432948, EPI_ISL_3432949, EPI_ISL_3432950, EPI_ISL_3432951, EPI_ISL_3803222 | | | |
| see above | MEPHI, Aix Marseille University | MEPHI, Aix Marseille University | Anthony LEVASSEUR |
| EPI_ISL_3494663 | MVZ Labor Dr. Fenner und Kollegen (Standort Hamburg) | Robert Koch Institute | |
| EPI_ISL_3298934 | Maine Health and Environmental Testing Laboratory | Tewhey Lab, The Jackson Laboratory | Barter, M.; Dewey, H.; H. and Tewhey, R.; Iosue, F.; Lynch, R.; Matluk, N.; Munger |
| EPI_ISL_3427182, EPI_ISL_3427279, EPI_ISL_3427338, EPI_ISL_3427340, EPI_ISL_3427581, EPI_ISL_3427603, EPI_ISL_3427658, EPI_ISL_3427913, EPI_ISL_3428594, EPI_ISL_3603040, EPI_ISL_3603806, EPI_ISL_3604040, EPI_ISL_3604274, EPI_ISL_3604495, EPI_ISL_3865103, EPI_ISL_3865218, EPI_ISL_3865230 | | | |
| see above | Mako Medical | Centers for Disease Control and Prevention Division of Viral Diseases, Pathogen Discovery | Adrian Paskey; Benjamin Rambo-Martin; Christopher Gulvick; Clinton Paden; Clinton R. Paden; Dakota Howard; Darlene Wagner; Dhwani Batra; Duncan MacCannell; Jason Caravas; Kara Moser; Lauren Moon; Matthew Schmerer; Matthew Tugwell; Peter Cook; Peter W. Cook; Scott Sammons; Shatavia Morrison; Yvette Unoarumhi |
| EPI_ISL_3549791 | Maryland Genomics, Institute for Genome Sciences, University of Maryland School of Medicine | Maryland Genomics, Institute for Genome Sciences, University of Maryland School of Medicine | Claire M; Fraser; Hazen; Holly; Humphrys; Ivette; Jacques; Jonathan; Kranthi; Lim; Lisa D; Luke J; Mike; Ott; Ravel; Roussey; Sadzewicz; Sandra; Santana-Cruz; Tallon; Tracy; Vavikolanu |
| EPI_ISL_3877468 | Medizinisch-Diagnostisches Labor Kempten allgäulab | Robert Koch Institute | |
| EPI_ISL_3713461 | Medizinische Laboratorien Düsseldorf | Robert Koch Institute | |
| EPI_ISL_3156056 | Microbiology Department, Laboratori Clínic Metropolitana Nord. Hospital Universitari Germans Trias i Pujol | Can Ruti SARS-CoV-2 Sequencing Hub (HUGTiP/IrsiCaixa/IGTP) | Alba Sánchez; Alexia París; Anna Not; Antoni E Bordoy; Bonaventura Clotet; Cristina Casañ; David Panisello; Francesc Catala-Moll; Gemma Clara; Ignacio Blanco; Laia Soler; Lauro Sumoy; Marc Noguera-Julian; Maria Casadellà; Mariona Parera; Mercedes Guerrero; Montserrat Giménez; Pere-Joan Cardona; Pilar Armengol; Roger Paredes; Verónica Saludes; and Elisa Martró on behalf of the Can Ruti SARS-CoV-2 Sequencing Hub |
| EPI_ISL_3875815 | Microbiology Department. Complexo Hospitalario Universitario de Vigo | Microbiology Department. Complexo Hospitalario Universitario de Vigo | Alfaya N; Alonso I; Alvarez M; Cabrera JJ; Carballo R; Cores O; Cortizo S; Davina C; Martinez L; Mediero G; Perez S; Potel C; Regueiro B; Rey S; Vasallo FJ; del-Campo V |
| EPI_ISL_3133570 | Microvida | Microvida | Jaco J. Verweij; Joep J. J. M. Stohr; Suzan D. Pas |
| EPI_ISL_3256532, EPI_ISL_3256534, EPI_ISL_3256539, EPI_ISL_3256543, EPI_ISL_3256552, EPI_ISL_3256583, EPI_ISL_3256891, EPI_ISL_3257132, EPI_ISL_3257153, EPI_ISL_3306730, EPI_ISL_3315243, EPI_ISL_3315245, EPI_ISL_3315250, EPI_ISL_3315258, EPI_ISL_3315259, EPI_ISL_3315260, EPI_ISL_3315291, EPI_ISL_3315442, EPI_ISL_3315443, EPI_ISL_3315444, EPI_ISL_3315445, EPI_ISL_3315512, EPI_ISL_3315614, EPI_ISL_3315615, EPI_ISL_3315616, EPI_ISL_3315617, EPI_ISL_3315618, EPI_ISL_3315630, EPI_ISL_3315652, EPI_ISL_3315757, EPI_ISL_3315759, EPI_ISL_3315764, EPI_ISL_3315772, EPI_ISL_3315773, EPI_ISL_3315774, EPI_ISL_3402376, EPI_ISL_3402377, EPI_ISL_3402378, EPI_ISL_3402379, EPI_ISL_3402380, EPI_ISL_3402381, EPI_ISL_3402382, EPI_ISL_3402383, EPI_ISL_3402384, EPI_ISL_3402385, EPI_ISL_3402403, EPI_ISL_3402408, EPI_ISL_3402429, EPI_ISL_3402430, EPI_ISL_3402431, EPI_ISL_3402432, EPI_ISL_3402477, EPI_ISL_3402482, EPI_ISL_3402503, EPI_ISL_3402504, EPI_ISL_3402505, EPI_ISL_3402506, EPI_ISL_3403233, EPI_ISL_3403294, EPI_ISL_3405064, EPI_ISL_3405211, EPI_ISL_3405361, EPI_ISL_3405511, EPI_ISL_3405607, EPI_ISL_3405629, EPI_ISL_3405707, EPI_ISL_3405729, EPI_ISL_3405807, EPI_ISL_3405829, EPI_ISL_3405907, EPI_ISL_3405929, EPI_ISL_3406462, EPI_ISL_3406705, EPI_ISL_3406948, EPI_ISL_3412436, EPI_ISL_3412446, EPI_ISL_3412468, EPI_ISL_3412492, EPI_ISL_3412501, EPI_ISL_3412525, EPI_ISL_3414355, EPI_ISL_3414629, EPI_ISL_3414795, EPI_ISL_3414917, EPI_ISL_3415132, EPI_ISL_3415254, EPI_ISL_3415431, EPI_ISL_3415440, EPI_ISL_3415461, EPI_ISL_3415485, EPI_ISL_3415494, EPI_ISL_3415518, EPI_ISL_3533151, EPI_ISL_3533871, EPI_ISL_3533872, EPI_ISL_3534125 | | | |
| see above | Ministry of Health Turkey | Ministry of Health Turkey | Fatma Bayrakdar; Gulay Korukluoglu; Gülay Korukluoğlu; Suleyman Yalcin; Süleyman Yalcin; Yasemin Cosgun; Yasemin Coşgun |
| EPI_ISL_2988441, EPI_ISL_3133154, EPI_ISL_3133159, EPI_ISL_3268226 | Montana Public Health Laboratory | Montana Public Health Laboratory | Carrie Biskupiak; Deborah Gibson; Joy Ritter; Michael Dills; Michelle Mozer |
| EPI_ISL_3900727 | NJDOH, Public Health and Environmental Laboratories | NJ PHEL | Allison Roder; Byeong Jeong; Chelsea San Filippo; Dana Woell; Jacquelyn Deverell; Lindsey Bodnar; Ryan Pachucki; Shiv K. Verma |

| | | | |
|---|---|---|---|
| EPI_ISL_3151733, EPI_ISL_3232172, EPI_ISL_3668028 | NJDOH, Public Health and Environmental Laboratories | NJ_PHEL | Allison Roder; Byeong Jeong; Chelsea San Filippo; Dana Woell; Jacquelyn Deverell; Lindsey Bodnar; Ryan Pachucki; Shiv K. Verma |
| EPI_ISL_3704078, EPI_ISL_3704099, EPI_ISL_3704109, EPI_ISL_3704118, EPI_ISL_3704147, EPI_ISL_3704151, EPI_ISL_3704154, EPI_ISL_3704158, EPI_ISL_3704170, EPI_ISL_3704181, EPI_ISL_3704202, EPI_ISL_3704223, EPI_ISL_3704242, EPI_ISL_3704252, EPI_ISL_3704261, EPI_ISL_3704270, EPI_ISL_3704279, EPI_ISL_3704282, EPI_ISL_3704286, EPI_ISL_3704290, EPI_ISL_3704299, EPI_ISL_3704309, EPI_ISL_3704318, EPI_ISL_3704330, EPI_ISL_3704334, EPI_ISL_3704337, EPI_ISL_3704342, EPI_ISL_3704345, EPI_ISL_3704353, EPI_ISL_3704364, EPI_ISL_3704374, EPI_ISL_3704389, EPI_ISL_3704390, EPI_ISL_3704393, EPI_ISL_3704394, EPI_ISL_3704395, EPI_ISL_3704396, EPI_ISL_3704397, EPI_ISL_3704398, EPI_ISL_3704399, EPI_ISL_3704400, EPI_ISL_3704401, EPI_ISL_3704402, EPI_ISL_3704403, EPI_ISL_3704404, EPI_ISL_3704405, EPI_ISL_3704407, EPI_ISL_3704408, EPI_ISL_3704409, EPI_ISL_3704411, EPI_ISL_3704412, EPI_ISL_3704413, EPI_ISL_3704414, EPI_ISL_3704418, EPI_ISL_3704421, EPI_ISL_3704424, EPI_ISL_3704428, EPI_ISL_3704431, EPI_ISL_3704435, EPI_ISL_3704438, EPI_ISL_3704442, EPI_ISL_3704445, EPI_ISL_3704449, EPI_ISL_3704458, EPI_ISL_3835341, EPI_ISL_3835342, EPI_ISL_3835343, EPI_ISL_3835344, EPI_ISL_3835345, EPI_ISL_3835346, EPI_ISL_3835347, EPI_ISL_3835348, EPI_ISL_3835350 | | | |
| see above | NUPIT/UFPE | WallauLab on behalf of Fiocruz COVID-19 Genomic Surveillance Network | Alexandre Freitas da Silva; Cassia Docena; Constância Flávia Junqueira Ayres; Filipe Zimmer Dezordi; Gabriel Luz Wallau; Gustavo Barbosa de Lima; Lais Ceschini Machado; Lilian Caroliny Amorim Silva; Maira Galdino da Rocha Pitta; Marcelo Henrique dos Santos Paiva; Matheus Filgueira Bezerra; Michelly Cristiny Pereira; Rômulo Pessoa e Silva; Sinval Pinto Brandão Filho |
| EPI_ISL_3102715, EPI_ISL_3102716 | Nevada State Public Health Laboratory | Nevada State Public Health Laboratory | Andrew Gorzalski; Mark Pandori |
| EPI_ISL_3329246, EPI_ISL_3812700, EPI_ISL_3819196, EPI_ISL_3999557 | New Mexico Department of Health Scientific Laboratory | New Mexico Department of Health Scientific Laboratory | Anastacia Griego-Fisher; D'eldra Malone; Ellie Johnson; Jennifer Benoit; Linda Salazar; Ratheesh Rajan; T. Mark Willmon |
| EPI_ISL_3185890 | Northwestern Memorial Hospital | Northwestern University - Ozer Lab | Chad J. Achenbach; Chao Qi; Egon A. Ozer; Judd F. Hultquist; Lacy M. Simons; Lawrence J. Jennings; Michael G. Ison; Ramon Lorenzo-Redondo; Taylor J. Dean |
| EPI_ISL_3547380 | OHSU-MM Lab | OHSU MM Lab | Yun Wu |
| EPI_ISL_3100170, EPI_ISL_3233062, EPI_ISL_3304459 | OSPEDALE SAN SALVATORE - MEDICINA DI LABORATORIO | Istituto Zooprofilattico Sperimentale dell'Abruzzo e Molise "G. Caporale" | Ancora M; Calistri P; Cammà C; Caporale M; Curini V; Delli Compagni E; Di Domenico M; Di Lollo Valeria; Di Pasquale A; Lorusso A; Mangone I; Marcacci M; Puglia I; Rinaldi A; Savini G; Scialabba S |
| EPI_ISL_3538946, EPI_ISL_3657512, EPI_ISL_4020772 | Ohio Department of Health Laboratory | Ohio Department of Health Laboratory | Allison Black; Brent Lee; Caitlin McDonnell; Eric Brandt; Erica Leasure; Glen McGillivary; Heather Blankenship; Holmes; Jade Mowery; Jennifer; Kelsey Florek; Keoni Omura; Kirtana Ramadugu; Quanta Brown; Stephanie Mccracken; Tyler Payne; and Tammy Bannerman |
| EPI_ISL_3088308 | Omics Sciences Laboratory | Omics Sciences Laboratory | Darlyn Amaya; Derly Andrade Molina; Gabriel Morey León; Juan Carlos Fernández Cadena; Rubén Armas González |
| EPI_ISL_3067395, EPI_ISL_3568685, EPI_ISL_3592930, EPI_ISL_3760834, EPI_ISL_4004814 | Oregon State Public Health Laboratory | Oregon State Public Health Laboratory | Eugene Yeboah; John Fontana and Shane Sevey; Laura Tsaknaridis; Rafia Razzaque; Vanda Makris |
| EPI_ISL_3013976 | Osp. S.Pertini | National Institute for Infectious Diseases (INMI) L. Spallanzani I.R.C.C.S | A Di Caro; B Bartolini; CEM Gruber; E Giombini; F Messina; F Santini; G Bonfiglio; M Rueca; MR Capobianchi; O Butera |
| EPI_ISL_3219524, EPI_ISL_3219525, EPI_ISL_3219526, EPI_ISL_3219527, EPI_ISL_3219528, EPI_ISL_3219529, EPI_ISL_3219530, EPI_ISL_3219531, EPI_ISL_3219532, EPI_ISL_3219533, EPI_ISL_3392632, EPI_ISL_3392633, EPI_ISL_3392634, EPI_ISL_3392635, EPI_ISL_3392636, EPI_ISL_3392637, EPI_ISL_3392638, EPI_ISL_3392642, EPI_ISL_3801546, EPI_ISL_3804146, EPI_ISL_3804147, EPI_ISL_3804148, EPI_ISL_3804149, EPI_ISL_3804156, EPI_ISL_3804157, EPI_ISL_3804158 | | | |
| see above | Outre mer | National Reference Center for Viruses of Respiratory Infections, Institut Pasteur, Paris | Angela Brisebarre; Camille Capel; Christophe Malabat; Corinne Maufrais; Dominique Rousset; Etienne Simon-Lorière; Frédéric Lemoine; Hub de Bioinformatique et Biostatistique; Hub de bioinformatique et Biostatistique; Louise Lefrançois; Marion Barbet; Maud Vanpeene; Méline Bizard; Sylvie Behillil; Sylvie Van der Werf; Vincent Enouf |
| EPI_ISL_3922300 | PRONTO SOCORRO MUNICIPAL TAMBAU | Instituto Butantan | Antonio Jorge Martins; Claudia Renata dos Santos Barros; David Schlesinger; Debora Botequio Moretti; Dimas Tadeu Covas; Elaine Cristina Marqueze; Elaine Vieira Santos; Evandra Strazza Rodrigues; Heidge Fukumasu; Jayme Augusto de Souza-Neto; José Salvatore Leister Patané; Luiz Alcantara; Luiz Lehmann Coutinho; Maria Carolina Elias; Maurício Lacerda Nogueira; Rafael dos Santos Bezerra; Raul Machado Neto; Rejane Maria Tommasini Grotto; Ricardo Haddad; Sandra Coccuzzo Sampaio Vessoni; Simone Kashima; Svetoslav Nanev Slavov; Vincent Louis Viala |
| EPI_ISL_3010693, EPI_ISL_3010747, EPI_ISL_3188491, EPI_ISL_3331609, EPI_ISL_3394309, EPI_ISL_3569307, EPI_ISL_3778434 | | | |
| see above | Pandemic Response Lab - NYC | Pandemic Response Lab, R&D | Alex Carpio; Cybill del Castillo; Dylan Law; Haiping Hao; Henry Lee; Isabel Fernandez Escapa; Jon Laurent; Melissa Hopkins; Michael Hammerling; Pradeep Bugga; Shinyoung Clair Kang; Sol Rey; William Ward |
| EPI_ISL_3707328 | Pathology, UAB | Pathology, UAB | E.J.; Hendrickson; Leal; Lefkowitz; Moates, D.; R.C.; S.M.; Van Der Pol; W.J. |
| EPI_ISL_3451831 | Pennsylvania Department of Health Bureau of Laboratories | Pennsylvania Department of Health Bureau of Laboratories | Dongxiang Xia |
| EPI_ISL_3474945, EPI_ISL_3476662, EPI_ISL_3478418 | Plateau technique Biomer Metz Verriers | Department of Virology, Henri Mondor University Hospital, Assistance Publique Hôpitaux de Paris, Université Paris-Est Créteil, INSERM U955 | Alexandre Soulier; Christophe Rodriguez; Elisabeth Trawinski; Guillaume Gricourt; Jean-Michel Pawlotsky; Melissa N'Debi; Slim Fourati; Vanessa Demontant |
| EPI_ISL_3943628 | Pomona Valley Hospital Medical Center | Los Angeles County Public Health Laboratories | P. Hemarajata et al. |
| EPI_ISL_3303908, EPI_ISL_3303911, EPI_ISL_3303921, EPI_ISL_3303925, EPI_ISL_3395239, EPI_ISL_3395300, EPI_ISL_3395551, EPI_ISL_3395650, EPI_ISL_3395680, EPI_ISL_3395681, EPI_ISL_3395786, EPI_ISL_3396761, EPI_ISL_3396821, EPI_ISL_3431067, EPI_ISL_3431076, EPI_ISL_3456275, EPI_ISL_3456505, EPI_ISL_3456512, EPI_ISL_3460809, EPI_ISL_3460811, EPI_ISL_3460820, EPI_ISL_3460821, EPI_ISL_3460830, EPI_ISL_3460839, EPI_ISL_3460840, EPI_ISL_3823483, EPI_ISL_3823527, EPI_ISL_3823856, EPI_ISL_3824012 | | | |
| see above | Quest Diagnostics Incorporated | Centers for Disease Control and Prevention Division of Viral Diseases, Pathogen Discovery | A. Gerasimova; A. Perez; Adrian Paskey; B. Anderson; Benjamin Rambo-Martin; Christopher Gulvick; Clinton Paden; Clinton R. Paden; Dakota Howard; Darlene Wagner; Dhwani Batra; Duncan MacCannell; F. Lacbawan; I. A. Shlyakhter; I. Shlyakhter; Jason Caravas; K. Livingston; K.E. Livingston; Kara Moser; L. Bernstein; L.E. Bernstein; M. Hua; Matthew Schmerer; P. Tanpaiboon; Peter Cook; Peter W. Cook; R. Kagan; R. M. Kagan; R. Owen; R. Rolando; R. V. Rolando; S. H. Rosenthal; S. Rosenthal; Sammons; Scott; Scott Sammons; Shatavia Morrison; Y. Liu; Yvette Unoarumhi |
| EPI_ISL_3315103 | RM2/Osp. S.Pertini | National Institute for Infectious Diseases (INMI) L. Spallanzani I.R.C.C.S | A Di Caro; B Bartolini; CEM Gruber; E Giombini; F Messina; F Santini; G Bonfiglio; M Rueca; MR Capobianchi; O Butera |
| EPI_ISL_3446918 | RM3/A.O. S.Camillo | National Institute for Infectious Diseases (INMI) L. Spallanzani I.R.C.C.S | A Di Caro; B Bartolini; CEM Gruber; E Giombini; F Messina; F Santini; G Bonfiglio; M Rueca; MR Capobianchi; O Butera |
| EPI_ISL_3080974, EPI_ISL_3179251, EPI_ISL_3291465, EPI_ISL_3291851, EPI_ISL_3573591, EPI_ISL_3574505, EPI_ISL_3959078, EPI_ISL_3959944 | | | |
| see above | Respiratory Virus Unit, Microbiology Services Colindale, Public Health England | COVID-19 Genomics UK (COG-UK) Consortium | PHE Covid Sequencing Team |
| EPI_ISL_3398872, EPI_ISL_3720972 | Rhode Island State Health Laboratory | Rhode Island State Health Laboratory | Ewa King; Kristin Carpenter-Azevedo; Richard C. Huard |
| EPI_ISL_3982687, EPI_ISL_3982721, EPI_ISL_3982725 | Ribeirao Preto | Laboratorio de Biologia Molecular Hemocentro de Ribeirao Preto | Antonio Jorge Martins; Claudia Renata dos Santos Barros; David Schlesinger; Debora Botequio Moretti; Dimas Tadeu Covas; Elaine Cristina Marqueze; Elaine Vieira Santos; Evandra Strazza Rodrigues; Heidge Fukumasu; Jayme Augusto de Souza-Neto; José Salvatore Leister Patané; Luiz Alcantara; Luiz Lehmann Coutinho; Maria Carolina Elias; Maurício Lacerda Nogueira; Rafael dos Santos Bezerra; Raul Machado Neto; Rejane Maria Tommasini Grotto; Ricardo Haddad; Sandra Coccuzzo Sampaio Vessoni; Simone Kashima; Svetoslav Nanev Slavov; Vincent Louis Viala |
| EPI_ISL_3982690, EPI_ISL_3982707, EPI_ISL_3982714, EPI_ISL_3982720, EPI_ISL_3982724, EPI_ISL_3982727 | Ribeirao Preto SP | Laboratorio de Biologia Molecular Hemocentro de Ribeirao Preto | Antonio Jorge Martins; Claudia Renata dos Santos Barros; David Schlesinger; Debora Botequio Moretti; Dimas Tadeu Covas; Elaine Cristina Marqueze; Elaine Vieira Santos; Evandra Strazza Rodrigues; Heidge Fukumasu; Jayme Augusto de Souza-Neto; José Salvatore Leister Patané; Luiz Alcantara; Luiz Lehmann Coutinho; Maria Carolina Elias; Maurício Lacerda Nogueira; Rafael dos Santos Bezerra; Raul Machado Neto; Rejane Maria Tommasini Grotto; Ricardo Haddad; Sandra Coccuzzo Sampaio Vessoni; Simone Kashima; Svetoslav Nanev Slavov; Vincent Louis Viala |
| EPI_ISL_3388720 | Rush University Medical Center | RIPHL at Rush University Medical Center | Diane Springer; Edith Perez; Felix Araujo Perez; Joyce Houlihan; Kevin Kunstman; Laura Furtado; Marieta Hyde; Mary Hayden; Max Kolton; Stefan Green |
| EPI_ISL_3909190 | SARS-CoV-2 Sequencing Castilla y Leon-Spain Consortium | SARS-CoV-2 Sequencing Castilla y Leon-Spain Consortium | Antonio Orduña-Domingo; Carlos Fuster Foz; Carmen Aldea-Mansilla; Carmen Gimeno Crespo; David Abad; Gabriel March Rosello; Gregoria Megías Lobón; Jose María Eiros Bouza; M. Isabel Fernandez-Natal; Marta Dominguez-Gil; Marta Hernandez; María Antonia García Castro; Mª Fe Brezmes-Valdivieso; Noelia Arenal Andrés; Silvia Rojo; Sonsoles Garcinuño Pérez |
| EPI_ISL_3076694, EPI_ISL_3543154, EPI_ISL_3983486 | SARS-CoV-2 testing team, National Institute of Infectious Diseases | Pathogen Genomics Center, National Institute of Infectious Diseases | Hazuka Y Furihata; Hiromizu Takahashi; Kentaro Itokawa; Makoto Kuroda; Masanori Hashino; Masumichi Saito; Naomi Nojiri; Nozomu Hanaoka; Rina Tanaka; Tsuguto Fujimoto; Tsuyoshi Sekizuka |
| EPI_ISL_3579325 | SECRETARIA MUNICIPAL DE SAUDE DE SANTA BARBARA D OESTE | Instituto Butantan | Antonio Jorge Martins; Claudia Renata dos Santos Barros; David Schlesinger; Debora Botequio Moretti; Dimas Tadeu Covas; Elaine Cristina Marqueze; Elaine Vieira Santos; Evandra Strazza Rodrigues; Heidge Fukumasu; Jayme Augusto de Souza-Neto; José Salvatore Leister Patané; Luiz Alcantara; Luiz Lehmann Coutinho; Maria Carolina Elias; Maurício Lacerda Nogueira; Rafael dos Santos Bezerra; Raul Machado Neto; Rejane Maria Tommasini Grotto; Ricardo Haddad; Sandra Coccuzzo Sampaio Vessoni; Simone Kashima; Svetoslav Nanev Slavov; Vincent Louis Viala |
| EPI_ISL_3912239 | SECRETARIA MUNICIPAL | Analytical Competence Molecular | Cleber Furtado Aksenen; Fabio Miyajima; Fernando Braga Stehling; Francisco Eder de Moura Lopes; Jamille Maria Mendes Bezerra; Joaquim Cesar do Nascimento Sousa Junior; Pedro Miguel Carneiro Jeronimo; Suzana Porto Almeida & Lucas Delerino on behalf of COVID-19 FIOCRUZ Genomic Network; Thais Ferreira de |

| | | | |
|---|---|---|---|
| | DE SAUDE DE TIANGUA | Epidemiology Lab/ACME, Oswaldo Cruz Foundation, Ceara (FIOCRUZ CE) | Oliveira; Thais de Oliveira Costa; Ticiane Cavalcante de Souza; Veridiana Pessoa Miyajima |
| EPI_ISL_3391909 | SELAS MEDILYS | Department of Virology, Henri Mondor University Hospital, Assistance Publique Hôpitaux de Paris, Université Paris-Est Créteil, INSERM U955 | Alexandre Soulier; Christophe Rodriguez; Elisabeth Trawinski; Guillaume Gricourt; Jean-Michel Pawlotsky; Melissa N'Debi; Slim Fourati; Vanessa Demontant |
| EPI_ISL_3204579 | SESARAM | Instituto Nacional de Saude (INSA) | Borges et al |
| EPI_ISL_3503634, EPI_ISL_3548258, EPI_ISL_3721736 | Salud Digna | Instituto Nacional de Medicina Genomica | Abraham Campos-Romero; Cedro-Tanda A; Escobar-Arrazola MA; Herrera-Montalvo LA.; Hidalgo-Miranda A; Luna-Ruiz Marco; Mendoza-Vargas A; Moreno-Camacho José Luis; Munguia-Garza P; Ramirez-Vega O; Rangel-DeLeon D; Reyes-Grajeda JP; Rodriguez-Gallegos Jorge; Yair Alfaro-Mora |
| EPI_ISL_3912477, EPI_ISL_3912498, EPI_ISL_3912518 | Salud Digna, A.C | Andersen lab at Scripps Research | Abraham Garcia Gil; Jose Luis Moreno Camacho; Marco Antonio Luna Ruiz-Esparza; Miguel A. Fernandez Rojas; SEARCH Alliance with Abraham Campos Romero |
| EPI_ISL_3912892, EPI_ISL_3912900, EPI_ISL_3912970, EPI_ISL_3912980 | San Diego County Public Health Laboratory | Andersen lab at Scripps Research | SEARCH Alliance San Diego with Ashleigh Murphy and Brett Austin |
| EPI_ISL_3708859, EPI_ISL_3842031 | San Diego County Public Health Laboratory | San Diego County Public Health Laboratory | Ashleigh Murphy Schafer; Brett Austin |
| EPI_ISL_3948575 | Sanquin B.V. | Sanquin B.V. | Anton van Weert; Jalenka van Wijk; Karin van Leeuwen; Lianne Koets; Maaike Derlagen; Marco Koppelman; Nadia Keijzer |
| EPI_ISL_3219274, EPI_ISL_3333990, EPI_ISL_3334041 | Santa Clara Valley Medical Center | Santa Clara County Public Health Laboratory | Santa Clara County Public Health Department |
| EPI_ISL_3261657, EPI_ISL_3757311 | Scripps Medical Laboratory | Andersen lab at Scripps Research | Ellen Stefanski; Ian Mchardy; SEARCH Alliance San Diego with Michael Quigley |
| EPI_ISL_4083362, EPI_ISL_4083435, EPI_ISL_4083447, EPI_ISL_4083509 | Servicio Virosis Respiratorias-Departamento Virología-INEI | Instituto Nacional Enfermedades Infecciosas C.G.Malbran | Avaro M.; Baumeister E.; Benedetti E.; Campos J.; Cisterna D.; Dattero ME; De Belder D.; Haim MS.; Lorenzo F.; Molina V.; Perandones C.; Poklepovich T.; Pontoriero A.; Russo M.; Sanchez Loria J.; Tuduri E. |
| EPI_ISL_3265888, EPI_ISL_3266010 | Servicio de Microbiología Clínica (Complejo Hospitalario de Navarra, Pamplona) | Centro de Secuenciación NASERTIC | Ana Miqueleiz; Ana Navascués; Carmen Ezpeleta Baquedano |
| EPI_ISL_3261327, EPI_ISL_3373926, EPI_ISL_3373973, EPI_ISL_3544107, EPI_ISL_3912619, EPI_ISL_3912881 | Sharp HealthCare Laboratory | Andersen lab at Scripps Research | Art Mendoza; Cathy Woerle; Jacquelyn Berumen; Liam McGinnis; Omid Bakhtar; SEARCH Alliance San Diego with Aaron Harding |
| EPI_ISL_3538746, EPI_ISL_3916315 | Stanford Health Care | Stanford University School of Medicine, Clinical Virology Laboratory | Becky Jiang; Bernadette Troung; James Zehnder; Malaya K. Sahoo; Nathan Hammond; Selamawit Bihon; and Benjamin A. Pinsky |
| EPI_ISL_3925268, EPI_ISL_3925285 | StarMed Healthcare | UNC Charlotte Environmental Monitoring Laboratory | Adam France; Cynthia Gibas; Erin Stiers; Jannatul Ferdous; Jessica Schlueter; Kevin Lambirth; Sam Kunkleman |
| EPI_ISL_3374243 | State Hygienic Laboratory at the University of Iowa | State Hygienic Laboratory at the University of Iowa | Alankar Kampoowale; Anna Yakos; Davis Rieckenberg; Erik Twait; Jeff Benfer; Kris Eveland; Kristen Zanon; Mariah Knutson; Valerie Reeb; Wes Hottel |
| EPI_ISL_3189705 | State Laboratories Division, Hawaii State Department of Health | State Laboratories Division, Hawaii State Department of Health | Ayana Garnet; Daniel Strange; Drew Kuwazaki; Edward Desmond; Pamela O'Brien; Razvan Sultana |
| EPI_ISL_3516374 | Supera | Laboratório de Biologia Molecular - Hemocentro de Ribeirão Preto | Sao Paulo State Network for Pandemic Alert of Emerging SARS-CoV-2 variants |
| EPI_ISL_3157680, EPI_ISL_3157776, EPI_ISL_3157825 | Swedish national genomic surveillance program of SARS-CoV-2 | The Public Health Agency of Sweden | Alma Brolund; Maria Lind Karlberg; Maximilian Riess; Swedish national genomic surveillance program of SARS-CoV-2 |
| EPI_ISL_3325819, EPI_ISL_3325849, EPI_ISL_3326334, EPI_ISL_3326700, EPI_ISL_3826189 | TGen North | TGen North | Brett Van Tassel; Chris French; Darrin Lemmer; Dave Engelthaler; Hayley Yaglom; Heather Centner; Jolene Bowers |
| EPI_ISL_3385370, EPI_ISL_3455613 | TXDSHS | TXDSHS | Anita Pokharel; Bonnie Oh; Chun Wang; Grace Kubin; Jenny Zhang; Karen Bobier; Lorraine Rodriguez; Maliha Rahman; Mayela Pedrueza; Myong Koag; Rachel Lee; Rashmi Tuladhar |
| EPI_ISL_3048178 | Tampa General Hospital Esoteric Lab | Tampa General Hospital Esoteric Research & Development Lab | Amorce Lima; Deanna Becker; Dominic Uy; Elaine Vendrone; Jon Faughn; Jorge Mecias-Frias; Marci O'Driscoll; Suzane Silbert; Vicki Healer |
| EPI_ISL_3922303 | UBS JARDIM FORTALEZA | Instituto Butantan | Antonio Jorge Martins; Claudia Renata dos Santos Barros; David Schlesinger; Debora Botequio Moretti; Dimas Tadeu Covas; Elaine Cristina Marqueze; Elaine Vieira Santos; Evandra Strazza Rodrigues; Heidge Fukumasu; Jayme Augusto de Souza-Neto; José Salvatore Leister Patané; Luiz Alcantara; Luiz Lehmann Coutinho; Maria Carolina Elias; Maurício Lacerda Nogueira; Rafael dos Santos Bezerra; Raul Machado Neto; Rejane Maria Tommasini Grotto; Ricardo Haddad; Sandra Coccuzzo Sampaio Vessoni; Simone Kashima; Svetoslav Nanev Slavov; Vincent Louis Viala |
| EPI_ISL_3922301, EPI_ISL_3922302 | UBS SANTA LIDIA | Instituto Butantan | Antonio Jorge Martins; Claudia Renata dos Santos Barros; David Schlesinger; Debora Botequio Moretti; Dimas Tadeu Covas; Elaine Cristina Marqueze; Elaine Vieira Santos; Evandra Strazza Rodrigues; Heidge Fukumasu; Jayme Augusto de Souza-Neto; José Salvatore Leister Patané; Luiz Alcantara; Luiz Lehmann Coutinho; Maria Carolina Elias; Maurício Lacerda Nogueira; Rafael dos Santos Bezerra; Raul Machado Neto; Rejane Maria Tommasini Grotto; Ricardo Haddad; Sandra Coccuzzo Sampaio Vessoni; Simone Kashima; Svetoslav Nanev Slavov; Vincent Louis Viala |
| EPI_ISL_3370434 | UC-Christus Clinical Hospital Laboratory | Laboratory of Molecular Virology, School of Medicine, Pontificia Universidad Catolica de Chile | Ana Maira Guzman; Andres E. Munoz-Marcos; Catalina Pardo-Roa; Eileen Serrano; Erick Salinas; Estefany Poblete; Francisco Melo; Jorge Levican; Leonardo I. Almonacid; Maria Jose Avendano; Maria Patricia Vega; Rafael A. Medina; Ricardo Enrique de la Barra; Tamara Garcia-Salum |
| EPI_ISL_3104703 | UCC (Universidad Central del Caribe) | Grubaugh Lab - Yale School of Public Health | Alejandaro Vallejo Degaudenzi; Anderson Brito; Annie Watkins; Chaney Kalinich; Chantal Vogels; Elisa Contreras; Esperanza Mendoza; Isabel Ott; Jessica Rothman; Joseph Fauver; Kendall Billig; Mallery Breban; Mary Petrone; Nathan Grubaugh; Robert Paulino-Ramirez; Tara Alpert; Tobias Koch; Victor Virgilio Calderon |
| EPI_ISL_3370910 | UCSC Genomics Institute | UCSC Genomics Institute | A. Marm Kilpatrick; Angie Hinrichs; Beth Shapiro; Bryan Thornlow; Ciara Wanket; David Haussler; Eric Beraut; Hugh Olsen; Ikenna Anigbogu; Isabel Bjork; Jakob McBroome; Jeremy Sanford; Joshua Kapp; Mark Akeson; Maximilian Haeussler; Michael Stone; Miten Jain; Molly Cassatt-Johnstone; Namrita Dhillon; Russell Corbett-Detig; Terren Chang; Yatish Turakhia |
| EPI_ISL_3912091 | UNIDADE SENTINELA DE JUAZEIRO DO NORTE | Analytical Competence Molecular Epidemiology Lab/ACME, Oswaldo Cruz Foundation, Ceara (FIOCRUZ CE) | Cleber Furtado Aksenen; Fabio Miyajima; Fernando Braga Stehling; Francisco Eder de Moura Lopes; Jamille Maria Mendes Bezerra; Joaquim Cesar do Nascimento Sousa Junior; Pedro Miguel Carneiro Jeronimo; Suzana Porto Almeida & Lucas Delerino on behalf of COVID-19 FIOCRUZ Genomic Network; Thais Ferreira de Oliveira; Thais de Oliveira Costa; Ticiane Cavalcante de Souza; Veridiana Pessoa Miyajima |
| EPI_ISL_3922304 | UPA SAO JOAO LAVRAS | Instituto Butantan | Antonio Jorge Martins; Claudia Renata dos Santos Barros; David Schlesinger; Debora Botequio Moretti; Dimas Tadeu Covas; Elaine Cristina Marqueze; Elaine Vieira Santos; Evandra Strazza Rodrigues; Heidge Fukumasu; Jayme Augusto de Souza-Neto; José Salvatore Leister Patané; Luiz Alcantara; Luiz Lehmann Coutinho; Maria Carolina Elias; Maurício Lacerda Nogueira; Rafael dos Santos Bezerra; Raul Machado Neto; Rejane Maria Tommasini Grotto; Ricardo Haddad; Sandra Coccuzzo Sampaio Vessoni; Simone Kashima; Svetoslav Nanev Slavov; Vincent Louis Viala |
| EPI_ISL_3939961 | URMC LABS | URMC LABS | Andrew Cameron; Dwight Hardy; Ghinwa Dumyati; Kelly DeLary; Stepan Voljehn |
| EPI_ISL_3942473, EPI_ISL_4001693, EPI_ISL_4079165 | US Air Force School of Aerospace Medicine | US Air Force School of Aerospace Medicine | Amanda Javorina; Anthony Fries; Carol Garrett; Clarise Starr; Cole Anderson; Deanna Muehleman; Elizabeth Macias; Fritz Castillo; Jennifer Black; Jennifer Meyer; Kelsey Lanter; Sarah Purves; William Gruner |
| EPI_ISL_3149580, EPI_ISL_3149665, EPI_ISL_3149692, EPI_ISL_3161179, EPI_ISL_3236592, EPI_ISL_3244868, EPI_ISL_3369571, EPI_ISL_3369588, EPI_ISL_3508018, EPI_ISL_3508031, EPI_ISL_3552266, EPI_ISL_3666624, EPI_ISL_3759154, EPI_ISL_3759507, EPI_ISL_3759541, EPI_ISL_3759553, EPI_ISL_3825874, EPI_ISL_3825976, EPI_ISL_3826062 | | | |
| see above | UW Virology Lab | UW Virology Lab | Alexander Greninger; Hong Xie; Keith R Jerome; Maria Lukes; Meei-Li Huang; Nathan Breit; Patrick Mathias; Pavitra Roychoudhury; Ricardo Perez; Robert J. Livingston; Savanna S. Carmack; Sean Ellis; Shah Mohamed Bakhash; Tien V. Nguyen |
| EPI_ISL_3557255 | Unidad de Investigación Biomédica de Zacatecas (UIBZ) | Centro de Investigación en Enfermedades Infecciosas (CIENI), Instituto Nacional de Enfermedades Respiratorias (INER) | Alejandra García-Gasca; Alejandra Hernández-Terán; Alejandro Sánchez-Flores; Alfredo Herrera-Estrella; Alicia Ocaña-Mondragón; Andreu Comas-García; Angel Gustavo Salas-Lais; Antonio Loza Román; Bernardo Martínez-Miguel; Blanca Taboada; Brenda Irasema Maldonado-Meza; Bruno Gómez-Gil; Carla Ivón Herrera-Najera; Carlos F. Arias; Celia Boukadida; Clara Esperanza Santacruz-Tinoco; Concepción Grajales-Muñiz; Consorcio Mexicano de Vigilancia Genómica (CoViGen-Mex). Authors (in alphabetical order): Julio Elias Alvarado-Yaah; Cristóbal Cháidez-Quiróz; Célida Duque Molina; Célida Martínez- Rodríguez; Daniel Fregoso-Rueda; Daniel Lira Morales; Eduardo Becerril-Vargas; Fernando Fontove-Herrera; Fidencio Mejía-Nepomuceno; Francisco Pulido; Gloria Elena Espinosa-Ayala; Gloria María Molina-Salinas; Gloria Vazquez; Hector Esteban Paz-Juárez; Hector Montoya-Fuentes; Helen Haydee Fernanda Ramírez-Plascencia; Irvin González-López; Jean Pierre González; Jesús Hernández; Joel Armando Vázquez-Pérez.; Jorge Salas-Hernández; José Antonio Enciso-Moreno; José Arturo Martínez-Orozco; José Esteban Muñoz-Medina; José de Jesús Nuñez-Contreras; Juan Bautista Chale-Dzul; Julissa Enciso-Ibarra; Kathia Elizabeth Tapia-Diaz; Luis Alberto Ochoa-Carrera; Margarita Matías-Florentino; Mario Mújica-Sánchez; Marissa Perez-Garcia; María Guadalupe Santiago-Mauricio; María Guadalupe de Jesús Mireles-Rivera; Nelly Sélem-Mojica; Pavel Isa; Ricardo Ciria Merce; Ricardo Grande; Rosa María Gutiérrez Rios; Santiago Ávila-Ríos; Selene Zárate; Susana Lopez; Verónica Mata-Haro; Victor Eduardo García-Arias; Víctor Hugo Borja-Aburto |

| | | | |
|---|---|---|---|
| EPI_ISL_4006331 | Unidad de Investigación Médica de Yucatán (UIMY) | Instituto de Biotecnología de la UNAM | ; Alejandra García-Gasca; Alejandra Hernández-Terán; Alejandro Sánchez-Flores; Alfredo Herrera-Estrella; Alicia Ocaña-Mondragón; Andreu Comas-García; Angel Gustavo Salas-Lais; Antonio Loza Román; Bernardo Martínez-Miguel; Blanca Taboada; Brenda Irasema Maldonado-Meza; Bruno Gómez-Gil; Carla Ivón Herrera-Najera; Carlos F. Arias; Celia Boukadida; Clara Esperanza Santacruz-Tinoco; Concepción Grajales-Muñiz; Consorcio Mexicano de Vigilancia Genómica (CoViGen-Mex). Authors (in alphabetical order): Julio Elias Alvarado-Yaah; Cristóbal Cháidez-Quiróz; Célida Duque Molina; Célida Martínez- Rodríguez; Daniel Fregoso-Rueda; Daniel Lira Morales; Eduardo Becerril-Vargas; Fernando Fontove-Herrera; Fidencio Mejía-Nepomuceno; Francisco Pulido; Gloria Elena Espinosa-Ayala; Gloria María Molina-Salinas; Gloria Vazquez; Hector Esteban Paz-Juárez; Hector Montoya-Fuentes; Helen Haydee Fernanda Ramírez-Plascencia; Irvin González-López; Jean Pierre González; Jesús Hernández; Joel Armando Vázquez-Pérez).; Jorge Salas-Hernández; José Antonio Enciso-Moreno; José Arturo Martínez-Orozco; José Esteban Muñoz-Medina; José de Jesús Nuñez-Contreras; Juan Bautista Chale-Dzul; Julissa Enciso-Ibarra; Kathia Elizabeth Tapia-Diaz; Luis Alberto Ochoa-Carrera; Margarita Matías-Florentino; Mario Müjica-Sánchez; Marissa Perez-Garcia; María Guadalupe Santiago-Mauricio; María Guadalupe de Jesús Mireles-Rivera; Nelly Sélem-Mojica; Pavel Isa; Ricardo Ciria Merce; Ricardo Grande; Rosa María Gutiérrez Rios; Santiago Ávila-Ríos; Selene Zárate; Susana Lopez; Verónica Mata-Haro; Victor Eduardo García-Arias; Víctor Hugo Borja-Aburto |
| EPI_ISL_3696701, EPI_ISL_3696711, EPI_ISL_3696758, EPI_ISL_3696767 | Unidade Básica de Saúde – Dr. José Francisco de Almeida | Instituto Butantan | Antonio Jorge Martins; Claudia Renata dos Santos Barros; David Schlesinger; Debora Botequio Moretti; Dimas Tadeu Covas; Elaine Cristina Marqueze; Elaine Vieira Santos; Evandra Strazza Rodrigues; Heidge Fukumasu; Jayme Augusto de Souza-Neto; José Salvatore Leister Patané; Luiz Alcantara; Luiz Lehmann Coutinho; Maria Carolina Elias; Maurício Lacerda Nogueira; Rafael dos Santos Bezerra; Raul Machado Neto; Rejane Maria Tommasini Grotto; Ricardo Haddad; Sandra Coccuzzo Sampaio Vessoni; Simone Kashima; Svetoslav Nanev Slavov; Vincent Louis Viala |
| EPI_ISL_3696811 | Unidade Mista de Saúde de Roseira | Instituto Butantan | Antonio Jorge Martins; Claudia Renata dos Santos Barros; David Schlesinger; Debora Botequio Moretti; Dimas Tadeu Covas; Elaine Cristina Marqueze; Elaine Vieira Santos; Evandra Strazza Rodrigues; Heidge Fukumasu; Jayme Augusto de Souza-Neto; José Salvatore Leister Patané; Luiz Alcantara; Luiz Lehmann Coutinho; Maria Carolina Elias; Maurício Lacerda Nogueira; Rafael dos Santos Bezerra; Raul Machado Neto; Rejane Maria Tommasini Grotto; Ricardo Haddad; Sandra Coccuzzo Sampaio Vessoni; Simone Kashima; Svetoslav Nanev Slavov; Vincent Louis Viala |
| EPI_ISL_3245351, EPI_ISL_3245353, EPI_ISL_3245361, EPI_ISL_3245377, EPI_ISL_3245382, EPI_ISL_3245388, EPI_ISL_3245391, EPI_ISL_3245393, EPI_ISL_3245402, EPI_ISL_3245403, EPI_ISL_3245404, EPI_ISL_3245405, EPI_ISL_3245407, EPI_ISL_3245413, EPI_ISL_3245414, EPI_ISL_3245415, EPI_ISL_3245418, EPI_ISL_3245420, EPI_ISL_3245421, EPI_ISL_3245423, EPI_ISL_3245425, EPI_ISL_3245437, EPI_ISL_3245439, EPI_ISL_3245449, EPI_ISL_3245452, EPI_ISL_3245453, EPI_ISL_3245456, EPI_ISL_3245459, EPI_ISL_3615203, EPI_ISL_3615204, EPI_ISL_3615205, EPI_ISL_3615206, EPI_ISL_3615207, EPI_ISL_3615208, EPI_ISL_3615210, EPI_ISL_3615211, EPI_ISL_3615212, EPI_ISL_3615213, EPI_ISL_3615214, EPI_ISL_3615215, EPI_ISL_3615217, EPI_ISL_3921467 | | | |
| see above | Unidade de apoio ao diagnostico da COVID - UNADIG | Bioinformatics Laboratory / LNCC | Alessandra P Lamarca; Alexandra L Gerber; Amilcar Tanuri; Ana Paula de C Guimaraes; Ana Tereza R Vasconcelos; Andrea Cony Cavalcanti; Caio Luiz Pereira Ribeiro; Cintia Policarpo; Claudia Maria Braga de Mello; Cristiane Gomes da Silva; Douglas Terra Machado; Erica Ramos dos Santos Nascimento; Fernanda Leitao dos Santos; Flavio Dias da Silva; Gleidson da Silva de Oliveira; Leandro Magalhaes de Souza; Liliane Cavalcante; Luiz G P de Almeida; Marcio Henrique de Oliveira Garcia; Mario Sergio Ribeiro; Ricardo Jose Barbosa Salviano; Ronaldo da Silva F Jr; Silvia Carvalho |
| EPI_ISL_3230063 | University Campus Bio-Medico of Rome (UCBM) | University Campus Bio-Medico of Rome (UCBM) | Angeletti S.; De Florio L.; Fogolari M.; Francesconi M.; Lintas C.; Riva E.; Veralli R. |
| EPI_ISL_3048434, EPI_ISL_3151472 | University Hospitals of Geneva, Laboratory of Virology | HUG, Laboratory of Virology and the Health2030 Genome Center | Aline Mamin; Ana Rita Goncalves; Deborah Penet; Emmanouil Dermitzakis; Francisco Perez; Henri Pegeot; Ioannis Xenarios; Keith Harshman; Laurent Kaiser; Lorenzo Cerutti; Melyssa Elies; Samuel Cordey |
| EPI_ISL_3893656 | University of Oregon COVID-19 MAP Laboratory | University of Oregon Genomics and Cell Charactericzation Core Facility (GC3F) | Ariana White; Demi Glidden; Douglas Turnbull; Jason Carriere; Jason Sydes; Jeff Bishop; Megan Criss; Peter Batzel |
| EPI_ISL_3231193, EPI_ISL_3231194, EPI_ISL_3231195 | Università degli Studi di Perugia | Istituto Zooprofilattico Sperimentale dell'Abruzzo e Molise "G. Caporale" | Ancora M; Biagetti M; Calistri P; Camilloni B; Cammà C; Curini V; Delli Compagni E; Di Domenico M; Di Pasquale A; Giammarioli M; Lorusso A; Mangone I; Marcacci M; Mencacci A; Puglia I; Rinaldi A; Savini G; Scialabba S |
| EPI_ISL_3144684, EPI_ISL_3263282, EPI_ISL_3263628, EPI_ISL_3449971, EPI_ISL_3745395, EPI_ISL_3833303 | Utah Public Health Laboratory | Utah Public Health Laboratory | Erin L. Young; Kelly F. Oakeson; Olinto Linares-Perdomo; Pooja Gupta |
| EPI_ISL_3710522 | VA Palo Alto Health Care System | Genomics and Discovery, Respiratory Viruses Branch, Division of Viral Diseases, Centers for Disease Control and Prevention | Adam Retchless; Anna Kelleher; Anna Uehara; Brian Lynch; Clinton R. Paden; Dhwani Batra; Haibin Wang; Han Jia Justin Ng; Jasmine Padilla; Jing Zhang; Justin Lee; Mark Burroughs; Mili Sheth; Morgan Davis; Peter Cook; Rachel Marine; Sarah Nobles; Suxiang Tong; Tara Coalter; Yan Li; Ying Tao |
| EPI_ISL_3259937 | VIDYMED LAUSANNE | Laboratory of genomics and metagenomics | Claire Bertelli; Damien Jacot; Gilbert Greub; Sébastien Aeby; Trestan Pillonel |
| EPI_ISL_3667098 | Valais Hospital, Central Institute | Valais Hospital, Central Institute | Alexis Dumoulin; Deborah Penet; Emmanouil Dermitzakis; Henri Pegeot; Ioannis Xenarios; Keith Harshman; Lorenzo Cerutti; Melyssa Elies |
| EPI_ISL_3853848 | Viollier AG | Department of Biosystems Science and Engineering, ETH Zürich | Andrea Patrizia Salzmann; Chaoran Chen; Christiane Beckmann; Christoph Noppen; Henriette Kurth; Ivan Topolsky; Kim Philipp Jablonski; Lara Fuhrmann; Louis du Plessis; Maurice Redondo; Niko Beerenwinkel; Olivier Kobel; Sarah Nadeau; Tanja Stadler |
| EPI_ISL_3048106, EPI_ISL_3117587, EPI_ISL_3318753, EPI_ISL_3402336, EPI_ISL_3667861, EPI_ISL_3667930, EPI_ISL_3693367, EPI_ISL_4000336 | | | |
| see above | Virginia Division of Consolidated Laboratory Services | Virginia Division of Consolidated Laboratory Services | Virginia Division of Consolidated Laboratory Services |
| EPI_ISL_3217059, EPI_ISL_3451010, EPI_ISL_3451033, EPI_ISL_3656901, EPI_ISL_3921836, EPI_ISL_3921837, EPI_ISL_3921838, EPI_ISL_3921839, EPI_ISL_3921840, EPI_ISL_3921841, EPI_ISL_3921842, EPI_ISL_3921843 | | | |
| see above | Washington State Department of Health Public Health Laboratories | Washington State Department of Health Public Health Laboratories | Avi Singh; Darren Lucas; Denny Russell; Drew MacKellar; Geoff Melly; Hannah Gray; Joenice Gonzalez; JohnAric Peterson; Philip Dykema; Rebecca Cao; Vanessa De Los Santos |
| EPI_ISL_3132091, EPI_ISL_3556239, EPI_ISL_4072757 | Wisconsin State Laboratory of Hygiene Communicable Disease Division | Wisconsin State Laboratory of Hygiene Communicable Disease Division | Abigail C. Shockey; Alicia J. Mooney; Erika M. Hanson; Kelsey R. Florek; Richard Griesser; Sara Wagner; Tonya Danz |
| EPI_ISL_3236383, EPI_ISL_3841133, EPI_ISL_4006008 | Yale Clinical Virology Lab | Grubaugh Lab - Yale School of Public Health | Anderson Brito; Annie Watkins; Chaney Kalinich; Chantal Vogels; Isabel Ott; Jessica Rothman; Joseph Fauver; Kendall Billig; Mallery Breban; Marie L. Landry; Mary Petrone; Nathan Grubaugh; Tara Alpert; Tobias Koch |

We gratefully acknowledge the following Authors from the Originating laboratories responsible for obtaining the specimens, as well as the Submitting laboratories where the genome data were generated and shared via GISAID, on which this research is based.

All Submitters of data may be contacted directly via www.gisaid.org

Authors are sorted alphabetically.

| Accession ID | Originating Laboratory | Submitting Laboratory | Authors |
|---|---|---|---|
| EPI_ISL_3128131, EPI_ISL_3128132, EPI_ISL_3128136 | "AZ SPHL, Arizona Department of Health Services" | Centers for Disease Control and Prevention Division of Viral Diseases, Pathogen Discovery | Alex Burgin; Ben L. Rambo-Martin; Clinton R. Paden; Dakota Howard; Dave Wentworth; Dhwani Batra; Jasmine Padilla; Justin Lee; Krista Queen; Kristen Knipe; Kristine Lacek; Mark Burroughs; Matthew Schmerer; Meghan Bentz; Mili Sheth; Peter Cook; Sam Shepard; Sarah Nobles; Suxiang Tong; Vivien Dugan; Yvette Unoarumhi |
| EPI_ISL_2987104, EPI_ISL_2987111, EPI_ISL_2987114 | "CDPH, Viral and Rickettsial Disease Laboratory" | Centers for Disease Control and Prevention Division of Viral Diseases, Pathogen Discovery | Alison Laufer Halpin; Ben L. Rambo-Martin; Clinton R. Paden; Dakota Howard; Darlene Wagner; Dave Wentworth; Dhwani Batra; Jasmine Padilla; Justin Lee; Katie Dillon; Krista Queen; Kristen Knipe; Kristine Lacek; Mark Burroughs; Matthew Schmerer; Mili Sheth; Peter Cook; Sam Shepard; Sarah Nobles; Shoshona Le; Suxiang Tong; Vivien Dugan; Yvette Unoarumhi |
| EPI_ISL_2787711 | "HI Dept. of Health, State Laboratories Division" | Centers for Disease Control and Prevention Division of Viral Diseases, Pathogen Discovery | Alison Laufer Halpin; Ben L. Rambo-Martin; Clinton R. Paden; Dakota Howard; Darlene Wagner; Dave Wentworth; Dhwani Batra; Jasmine Padilla; Justin Lee; Katie Dillon; Krista Queen; Kristen Knipe; Kristine Lacek; Mark Burroughs; Matthew Schmerer; Mili Sheth; Peter Cook; Sam Shepard; Sarah Nobles; Shoshona Le; Suxiang Tong; Vivien Dugan; Yvette Unoarumhi |
| EPI_ISL_2690045, EPI_ISL_3355284 | "NM Dept. Health, Scientific Laboratory Division " | Centers for Disease Control and Prevention Division of Viral Diseases, Pathogen Discovery | Alex Burgin; Alison Laufer Halpin; Ben L. Rambo-Martin; Ben Rambo-Martin; Clinton Paden; Clinton R. Paden; Dakota Howard; Darlene Wagner; Dave Wentworth; Dhwani Batra; Jasmine Padilla; Justin Lee; Katie Dillon; Krista Queen; Kristen Knipe; Kristine Lacek; Mark Burroughs; Matthew Schmerer; Meghan Bentz; Mili Sheth; Peter Cook; Sam Shepard; Sarah Nobles; Shoshona Le; Suxiang Tong; Vivien Dugan; Yvette Unoarumhi |
| EPI_ISL_3355430 | "NYSDOH Wadsworth Center, Virology Lab" | Centers for Disease Control and Prevention Division of Viral Diseases, Pathogen Discovery | Alex Burgin; Ben Rambo-Martin; Clinton Paden; Dakota Howard; Dave Wentworth; Dhwani Batra; Jasmine Padilla; Justin Lee; Krista Queen; Kristen Knipe; Kristine Lacek; Mark Burroughs; Matthew Schmerer; Meghan Bentz; Mili Sheth; Peter Cook; Sam Shepard; Sarah Nobles; Suxiang Tong; Vivien Dugan; Yvette Unoarumhi |
| EPI_ISL_3064753, EPI_ISL_4060386 | "OK Public Health Laboratory, Oklahoma State DOH" | Centers for Disease Control and Prevention Division of Viral Diseases, Pathogen Discovery | Alex Burgin; Ben L. Rambo-Martin; Ben Rambo-Martin; Clinton Paden; Clinton R. Paden; Dakota Howard; Dave Wentworth; Dhwani Batra; Jasmine Padilla; Justin Lee; Krista Queen; Kristen Knipe; Kristine Lacek; Mark Burroughs; Matthew Schmerer; Meghan Bentz; Mili Sheth; Peter Cook; Sam Shepard; Sarah Nobles; Suxiang Tong; Vivien Dugan; Yvette Unoarumhi |
| EPI_ISL_2494052, EPI_ISL_2494053, EPI_ISL_2494054, EPI_ISL_2494055, EPI_ISL_2494056, EPI_ISL_2494057 | AFIP LESTE | Instituto Butantan | Antonio Jorge Martins; Claudia Renata dos Santos Barros; David Schlesinger; Debora Botequio Moretti; Dimas Tadeu Covas; Elaine Cristina Marqueze; Elaine Vieira Santos; Evandra Strazza Rodrigues; Heidge Fukumasu; Jayme Augusto de Souza-Neto; José Salvatore Leister Patané; Luiz Alcantara; Luiz Lehmann Coutinho; Maria Carolina Elias; Maurício Lacerda Nogueira; Rafael dos Santos Bezerra; Raul Machado Neto; Rejane Maria Tommasini Grotto; Ricardo Haddad; Sandra Coccuzzo Sampaio Vessoni; Simone Kashima; Svetoslav Nanev Slavov; Vincent Louis Viala |
| EPI_ISL_2493269, EPI_ISL_2494058, EPI_ISL_2494059, EPI_ISL_2494060, EPI_ISL_2494062, EPI_ISL_2494063, EPI_ISL_2494064, EPI_ISL_2494066 | | | |
| see above | AFIP SUDESTE | Instituto Butantan | Antonio Jorge Martins; Claudia Renata dos Santos Barros; David Schlesinger; Debora Botequio Moretti; Dimas Tadeu Covas; Elaine Cristina Marqueze; Elaine Vieira Santos; Evandra Strazza Rodrigues; Heidge Fukumasu; Jayme Augusto de Souza-Neto; José Salvatore Leister Patané; Luiz Alcantara; Luiz Lehmann Coutinho; Maria Carolina Elias; Maurício Lacerda Nogueira; Rafael dos Santos Bezerra; Raul Machado Neto; Rejane Maria Tommasini Grotto; Ricardo Haddad; Sandra Coccuzzo Sampaio Vessoni; Simone Kashima; Svetoslav Nanev Slavov; Vincent Louis Viala |
| EPI_ISL_2493287, EPI_ISL_2494067, EPI_ISL_2494069, EPI_ISL_2494070, EPI_ISL_2494071, EPI_ISL_2494072, EPI_ISL_2494073, EPI_ISL_2494074, EPI_ISL_2494075, EPI_ISL_2494077, EPI_ISL_2494078, EPI_ISL_2494080, EPI_ISL_2494081, EPI_ISL_2494084, EPI_ISL_2494085, EPI_ISL_2494086, EPI_ISL_2494087, EPI_ISL_2494090, EPI_ISL_2494092, EPI_ISL_2494095, EPI_ISL_2494098, EPI_ISL_2494311, EPI_ISL_2494312, EPI_ISL_2494330 | | | |
| see above | AFIP SUL | Instituto Butantan | Antonio Jorge Martins; Claudia Renata dos Santos Barros; David Schlesinger; Debora Botequio Moretti; Dimas Tadeu Covas; Elaine Cristina Marqueze; Elaine Vieira Santos; Evandra Strazza Rodrigues; Heidge Fukumasu; Jayme Augusto de Souza-Neto; José Salvatore Leister Patané; Luiz Alcantara; Luiz Lehmann Coutinho; Maria Carolina Elias; Maurício Lacerda Nogueira; Rafael dos Santos Bezerra; Raul Machado Neto; Rejane Maria Tommasini Grotto; Ricardo Haddad; Sandra Coccuzzo Sampaio Vessoni; Simone Kashima; Svetoslav Nanev Slavov; Vincent Louis Viala |
| EPI_ISL_3031195 | ALBANY MEDICAL CENTER | Wadsworth Center, New York State Department of Health | Alexis Russell; Catharine Prussing; Daryl M. Lamson; Erasmus Schneider; Erica Lasek-Nesselquist; John Kelly; Jonathan Plitnick; Kirsten St. George; Matthew Shudt; Melissa A Leisner; Navjot Singh |
| EPI_ISL_3388726 | ALVERNO PROFESSIONAL CLINICAL LABORATORIES LLC | RIPHL at Rush University Medical Center | Edith Perez; Felix Araujo Perez; Julia Baranski; Kevin Kunstman; Laura Furtado; Marieta Hyde; Max Kolton; Stefan Green |
| EPI_ISL_2462327, EPI_ISL_2494134, EPI_ISL_2494137, EPI_ISL_2494138, EPI_ISL_2494331 | AMBULATORIO DE ESPECIALIDADE V E MOGI MIRIM | Instituto Butantan | Antonio Jorge Martins; Claudia Renata dos Santos Barros; David Schlesinger; Debora Botequio Moretti; Dimas Tadeu Covas; Elaine Cristina Marqueze; Elaine Vieira Santos; Evandra Strazza Rodrigues; Heidge Fukumasu; Jayme Augusto de Souza-Neto; José Salvatore Leister Patané; Luiz Alcantara; Luiz Lehmann Coutinho; Maria Carolina Elias; Maurício Lacerda Nogueira; Rafael dos Santos Bezerra; Raul Machado Neto; Rejane Maria Tommasini Grotto; Ricardo Haddad; Sandra Coccuzzo Sampaio Vessoni; Simone Kashima; Svetoslav Nanev Slavov; Vincent Louis Viala |
| EPI_ISL_2827997, EPI_ISL_2827998 | AREA DE SALUD BAGACES | Inciensa, Instituto Costarricense de Investigación y Enseñanza en Nutrición y Salud | Adriana Godínez; Claudio Soto-Garita; Estela Cordero; Francisco Duarte; Hebleen Porras; Joselyn Prado & Karol Matamoros; José Luis Vargas; Mariela Gutiérrez; Melany Calderón |
| EPI_ISL_3037817 | AREA DE SALUD BELEN-FLORES - CLINICA DR. JORGE VOLIO | Inciensa, Instituto Costarricense de Investigación y Enseñanza en Nutrición y Salud | Adriana Godínez; Caterina Guzmán; Claudio Soto-Garita; Estela Cordero; Francisco Duarte; Hebleen Porras; José Luis Vargas; Mariela Gutiérrez Joselyn Prado; Melany Calderón; Nazareth Ruiz & Sandra Mora Leitón |
| EPI_ISL_2827996, EPI_ISL_3298336 | AREA DE SALUD CARTAGO | Inciensa, Instituto Costarricense de Investigación y Enseñanza en Nutrición y Salud | Adriana Godínez; Claudio Soto-Garita; Estela Cordero; Francisco Duarte; Hebleen Porras; Joselyn Prado & Monserrat Segura; José Luis Vargas; Mariela Gutiérrez; Melany Calderón |
| EPI_ISL_3037814 | AREA DE SALUD CHACARITA - CLINICA DR. FRANCISCO QUINTANA | Inciensa, Instituto Costarricense de Investigación y Enseñanza en Nutrición y Salud | Adriana Godínez; Caterina Guzmán; Claudio Soto-Garita; Estela Cordero; Francisco Duarte; Hebleen Porras; José Luis Vargas; Mariela Gutiérrez Joselyn Prado; Melany Calderón; Nazareth Ruiz & María José Gómez |
| EPI_ISL_2827988, EPI_ISL_2827989, EPI_ISL_2827990 | AREA DE SALUD CIUDAD QUESADA | Inciensa, Instituto Costarricense de Investigación y Enseñanza en Nutrición y Salud | Adriana Godínez; Claudio Soto-Garita; Estela Cordero; Francisco Duarte; Hebleen Porras; Joselyn Prado & Juan Carlos Villalobos; José Luis Vargas; Mariela Gutiérrez; Melany Calderón |
| EPI_ISL_3298363 | AREA DE SALUD COBANO | Inciensa, Instituto Costarricense de Investigación y Enseñanza en Nutrición y Salud | Adriana Godínez; Claudio Soto-Garita; Estela Cordero; Francisco Duarte; Hebleen Porras; Joselyn Prado & María José Gómez; José Luis Vargas; Mariela Gutiérrez; Melany Calderón |
| EPI_ISL_3037821 | AREA DE SALUD COTO BRUS | Inciensa, Instituto Costarricense de Investigación y Enseñanza en Nutrición y Salud | Adriana Godínez; Caterina Guzmán; Claudio Soto-Garita; Estela Cordero; Francisco Duarte; Hebleen Porras; José Luis Vargas; Mariela Gutiérrez Joselyn Prado; Melany Calderón; Nazareth Ruiz & María Fernanda Matamoros |
| EPI_ISL_2827999 | AREA DE SALUD DESAMPARADOS 2 (COOPESALUD) | Inciensa, Instituto Costarricense de Investigación y Enseñanza en Nutrición y Salud | Adriana Godínez; Claudio Soto-Garita; Estela Cordero; Francisco Duarte; Hebleen Porras; Joselyn Prado & Alberto Quirós; José Luis Vargas; Mariela Gutiérrez; Melany Calderón |
| EPI_ISL_3298316 | AREA DE SALUD EL GUARCO | Inciensa, Instituto Costarricense de Investigación y Enseñanza en Nutrición y Salud | Adriana Godínez; Claudio Soto-Garita; Estela Cordero; Francisco Duarte; Hebleen Porras; Joselyn Prado & Mónica Charpentier; José Luis Vargas; Mariela Gutiérrez; Melany Calderón |
| EPI_ISL_2828006 | AREA DE SALUD ESCAZU (COOPESANA) | Inciensa, Instituto Costarricense de Investigación y Enseñanza en Nutrición y Salud | Adriana Godínez; Claudio Soto-Garita; Estela Cordero; Francisco Duarte; Hebleen Porras; Joselyn Prado & Juliana Mora; José Luis Vargas; Mariela Gutiérrez; Melany Calderón |
| EPI_ISL_2827985 | AREA DE SALUD ESPARZA | Inciensa, Instituto Costarricense de Investigación y Enseñanza en Nutrición y Salud | Adriana Godínez; Claudio Soto-Garita; Estela Cordero; Francisco Duarte; Hebleen Porras; Joselyn Prado & María José Gómez; José Luis Vargas; Mariela Gutiérrez; Melany Calderón |
| EPI_ISL_2828002, EPI_ISL_3037801 | AREA DE SALUD GOICOECHEA 2 - CLINICA DR. JIMENEZ NUÑEZ | Inciensa, Instituto Costarricense de Investigación y Enseñanza en Nutrición y Salud | & Nazareth Ruiz; Adriana Godínez; Caterina Guzmán; Claudio Soto-Garita; Estela Cordero; Francisco Duarte; Hebleen Porras; Joselyn Prado & Juan Carlos Cartes; José Luis Vargas; Mariela Gutiérrez; Mariela Gutiérrez Joselyn Prado; Melany Calderón |
| EPI_ISL_2827970 | AREA DE SALUD GUATUSO | Inciensa, Instituto Costarricense de Investigación y Enseñanza en Nutrición y Salud | Adriana Godínez; Claudio Soto-Garita; Estela Cordero; Francisco Duarte; Hebleen Porras; Joselyn Prado & Francisco Chacón; José Luis Vargas; Mariela Gutiérrez; Melany Calderón |
| EPI_ISL_2827961 | AREA DE SALUD HATILLO - CLINICA DR. SOLON NUÑEZ | Inciensa, Instituto Costarricense de Investigación y Enseñanza en Nutrición y Salud | Adriana Godínez; Claudio Soto-Garita; Estela Cordero; Francisco Duarte; Hebleen Porras; Joselyn Prado & Roberto Brilla; José Luis Vargas; Mariela Gutiérrez; Melany Calderón |
| EPI_ISL_3298313, EPI_ISL_3298314 | AREA DE SALUD MATA REDONDA-HOSPITAL - CLINICA DR. MORENO CAÑAS | Inciensa, Instituto Costarricense de Investigación y Enseñanza en Nutrición y Salud | Adriana Godínez; Claudio Soto-Garita; Estela Cordero; Francisco Duarte; Hebleen Porras; Joselyn Prado & Ricardo González; José Luis Vargas; Mariela Gutiérrez; Melany Calderón |
| EPI_ISL_3037804 | AREA DE SALUD OSA | Inciensa, Instituto Costarricense de | Adriana Godínez; Caterina Guzmán; Claudio Soto-Garita; Estela Cordero; Francisco Duarte; Hebleen Porras; José Luis Vargas; Mariela Gutiérrez Joselyn Prado; Melany Calderón; Nazareth Ruiz & Mariamilia Cob |

| | | | |
|---|---|---|---|
| | | Investigación y Enseñanza en Nutrición y Salud | |
| EPI_ISL_3037822 | AREA DE SALUD PARAISO-CERVANTES | Inciensa, Instituto Costarricense de Investigación y Enseñanza en Nutrición y Salud | Adriana Godínez; Caterina Guzmán; Claudio Soto-Garita; Estela Cordero; Francisco Duarte; Hebleen Porras; José Luis Vargas; Mariela Gutiérrez Joselyn Prado; Melany Calderón; Nazareth Ruiz & Mónica Charpentier |
| EPI_ISL_3037810 | AREA DE SALUD SIQUIRRES | Inciensa, Instituto Costarricense de Investigación y Enseñanza en Nutrición y Salud | Adriana Godínez; Caterina Guzmán; Claudio Soto-Garita; Estela Cordero; Francisco Duarte; Hebleen Porras; José Luis Vargas; Mariela Gutiérrez Joselyn Prado; Melany Calderón; Nazareth Ruiz & Ileana Chavez Peraza |
| EPI_ISL_3298305 | AREA DE SALUD ZAPOTE-CATEDRAL - CLINICA DR. CARLOS DURAN | Inciensa, Instituto Costarricense de Investigación y Enseñanza en Nutrición y Salud | Adriana Godínez; Claudio Soto-Garita; Estela Cordero; Francisco Duarte; Hebleen Porras; Joselyn Prado & Juliana Mora; José Luis Vargas; Mariela Gutiérrez; Melany Calderón |
| EPI_ISL_2695447 | ARS Algarve - Laboratorio Laura Ayres | Instituto Nacional de Saude (INSA) | Borges et al |
| EPI_ISL_2638106, EPI_ISL_3028238 | AULSS 2 Marca Trevigiana | Istituto Zooprofilattico Sperimentale delle Venezie | Adelaide Milani; Alessia Schivo; Alice Fusaro; Ambra Pastori; Annalisa Salviato; Antonia Ricci; Calogero Terregino; Edoardo Giussani; Elisa Palumbo; Erika Giorgia Quaranta; Isabella Monne; Luca Tassoni |
| EPI_ISL_2963463 | AULSS 5 Polesana | Istituto Zooprofilattico Sperimentale delle Venezie | Adelaide Milani; Alessia Schivo; Alice Fusaro; Ambra Pastori; Annalisa Salviato; Antonia Ricci; Calogero Terregino; Edoardo Giussani; Elisa Palumbo; Erika Giorgia Quaranta; Isabella Monne; Luca Tassoni |
| EPI_ISL_3006782 | AULSS 9 Scaligera | Istituto Zooprofilattico Sperimentale delle Venezie | Adelaide Milani; Alessia Schivo; Alice Fusaro; Ambra Pastori; Annalisa Salviato; Antonia Ricci; Calogero Terregino; Edoardo Giussani; Elisa Palumbo; Erika Giorgia Quaranta; Isabella Monne; Luca Tassoni |
| EPI_ISL_3039626 | AULSS 9 Scaligera - Legnago | Istituto Zooprofilattico Sperimentale delle Venezie | Adelaide Milani; Alessia Schivo; Alice Fusaro; Ambra Pastori; Annalisa Salviato; Antonia Ricci; Calogero Terregino; Edoardo Giussani; Elisa Palumbo; Erika Giorgia Quaranta; Isabella Monne; Luca Tassoni |
| EPI_ISL_2790130 | AZDelta | AZ Delta Medical Laboratories in Roeselare, Belgium | Dieter De Smet; Geert Martens; Merijn Vanhee; on behalf of AZ Delta COVID-19 Genomics core (member of Genomic surveillance of SARS-CoV-2 in Belgium network) |
| EPI_ISL_2370627, EPI_ISL_2370713, EPI_ISL_2422043, EPI_ISL_2422115, EPI_ISL_2422117, EPI_ISL_2442336, EPI_ISL_2489126, EPI_ISL_2489172, EPI_ISL_2489186, EPI_ISL_2489189, EPI_ISL_2489191, EPI_ISL_2489195, EPI_ISL_2489202, EPI_ISL_2489213, EPI_ISL_2489282, EPI_ISL_2489306, EPI_ISL_2489308, EPI_ISL_2489320, EPI_ISL_2489332, EPI_ISL_2489342, EPI_ISL_2489345, EPI_ISL_2489367, EPI_ISL_2489369, EPI_ISL_2489382, EPI_ISL_2489405, EPI_ISL_2489438, EPI_ISL_2489457, EPI_ISL_2489459, EPI_ISL_2489479, EPI_ISL_2489530, EPI_ISL_2489533, EPI_ISL_2489535, EPI_ISL_2489541, EPI_ISL_2489587, EPI_ISL_2489589, EPI_ISL_2489605, EPI_ISL_2489615, EPI_ISL_2489631, EPI_ISL_2489694, EPI_ISL_2489696, EPI_ISL_2489702, EPI_ISL_2489728, EPI_ISL_2489732, EPI_ISL_2489758, EPI_ISL_2489759, EPI_ISL_2489762, EPI_ISL_2489802, EPI_ISL_2527882, EPI_ISL_2527954, EPI_ISL_2527961, EPI_ISL_2527974, EPI_ISL_2528019, EPI_ISL_2528052, EPI_ISL_2528062, EPI_ISL_2528074, EPI_ISL_2528076, EPI_ISL_2528087, EPI_ISL_2528104, EPI_ISL_2528132, EPI_ISL_2528139, EPI_ISL_2528163, EPI_ISL_2528218, EPI_ISL_2528233, EPI_ISL_2528235, EPI_ISL_2528237, EPI_ISL_2528283, EPI_ISL_2528299, EPI_ISL_2528303, EPI_ISL_2528318, EPI_ISL_2528372, EPI_ISL_2528376, EPI_ISL_2528394, EPI_ISL_2528404, EPI_ISL_2528425, EPI_ISL_2528458, EPI_ISL_2528504, EPI_ISL_2528519, EPI_ISL_2528541, EPI_ISL_2528546, EPI_ISL_2528550, EPI_ISL_2528556, EPI_ISL_2528582, EPI_ISL_2528642, EPI_ISL_2528654, EPI_ISL_2528739, EPI_ISL_2528747, EPI_ISL_2528768, EPI_ISL_2528779, EPI_ISL_2528783, EPI_ISL_2528845, EPI_ISL_2528862, EPI_ISL_2528875, EPI_ISL_2528976, EPI_ISL_2528980, EPI_ISL_2528987, EPI_ISL_2529005, EPI_ISL_2530970, EPI_ISL_2531006, EPI_ISL_2531014, EPI_ISL_2531040, EPI_ISL_2531070, EPI_ISL_2534552, EPI_ISL_2686906, EPI_ISL_2686908, EPI_ISL_2686917, EPI_ISL_2686919, EPI_ISL_2686920, EPI_ISL_2686981, EPI_ISL_2686985, EPI_ISL_2686989, EPI_ISL_2686997, EPI_ISL_2687015, EPI_ISL_2687021, EPI_ISL_2687046, EPI_ISL_2687047, EPI_ISL_2687057, EPI_ISL_2687095, EPI_ISL_2687096, EPI_ISL_2687098, EPI_ISL_2687106, EPI_ISL_2687120, EPI_ISL_2687121, EPI_ISL_2687165, EPI_ISL_2687179, EPI_ISL_2687186, EPI_ISL_2687196, EPI_ISL_2687202, EPI_ISL_2687205, EPI_ISL_2687213, EPI_ISL_2687242, EPI_ISL_2687243, EPI_ISL_2687248, EPI_ISL_2687276, EPI_ISL_2687287, EPI_ISL_2687292, EPI_ISL_2687301, EPI_ISL_2687303, EPI_ISL_2687308, EPI_ISL_2687326, EPI_ISL_2687338, EPI_ISL_2687356, EPI_ISL_2687407, EPI_ISL_2687408, EPI_ISL_2687413, EPI_ISL_2687416, EPI_ISL_2687419, EPI_ISL_2687444, EPI_ISL_2687455, EPI_ISL_2687488, EPI_ISL_2687489, EPI_ISL_2687499, EPI_ISL_2687503, EPI_ISL_2687551, EPI_ISL_2687554, EPI_ISL_2687558, EPI_ISL_2687559, EPI_ISL_2709498, EPI_ISL_2709541, EPI_ISL_2709543, EPI_ISL_2784748, EPI_ISL_2784763, EPI_ISL_2784764, EPI_ISL_2784785, EPI_ISL_2784806, EPI_ISL_2784827, EPI_ISL_2784828, EPI_ISL_2784851, EPI_ISL_2784852, EPI_ISL_2784860, EPI_ISL_2784897, EPI_ISL_2784917, EPI_ISL_2784918, EPI_ISL_2784931, EPI_ISL_2784933, EPI_ISL_2784951, EPI_ISL_2784953, EPI_ISL_2784956, EPI_ISL_2784974, EPI_ISL_2784975, EPI_ISL_2784980, EPI_ISL_2784981, EPI_ISL_2784985, EPI_ISL_2784996, EPI_ISL_2784997, EPI_ISL_2785002, EPI_ISL_2785011, EPI_ISL_2785012, EPI_ISL_2785032, EPI_ISL_2785046, EPI_ISL_2785065, EPI_ISL_2785084, EPI_ISL_2785112, EPI_ISL_2785146, EPI_ISL_2785180, EPI_ISL_2785181, EPI_ISL_2785199, EPI_ISL_2785218, EPI_ISL_2785233, EPI_ISL_2785234, EPI_ISL_2785238, EPI_ISL_2785257, EPI_ISL_2785311, EPI_ISL_2785313, EPI_ISL_2785319, EPI_ISL_2785330, EPI_ISL_2785331, EPI_ISL_2785355, EPI_ISL_2785361, EPI_ISL_2785368, EPI_ISL_2785376, EPI_ISL_2785379, EPI_ISL_2785384, EPI_ISL_2785429, EPI_ISL_2785439, EPI_ISL_2785478, EPI_ISL_2785484, EPI_ISL_2785485, EPI_ISL_2785487, EPI_ISL_2785495, EPI_ISL_2785496, EPI_ISL_2832520, EPI_ISL_2832522, EPI_ISL_2832535, EPI_ISL_2875541, EPI_ISL_2875542, EPI_ISL_2875576, EPI_ISL_2875612, EPI_ISL_2875615, EPI_ISL_2875624, EPI_ISL_2875659, EPI_ISL_2875667, EPI_ISL_2875677, EPI_ISL_2875678, EPI_ISL_2875683, EPI_ISL_2875697, EPI_ISL_2875698, EPI_ISL_2875832, EPI_ISL_2875835, EPI_ISL_2875837, EPI_ISL_2875840, EPI_ISL_2875872, EPI_ISL_2875893, EPI_ISL_2875894, EPI_ISL_2877655, EPI_ISL_2877692, EPI_ISL_2877694, EPI_ISL_3217299, EPI_ISL_3217365, EPI_ISL_3217383, EPI_ISL_3217395, EPI_ISL_3217615, EPI_ISL_3217626, EPI_ISL_3217639, EPI_ISL_3217760, EPI_ISL_3217792, EPI_ISL_3217875, EPI_ISL_3218007, EPI_ISL_3218066, EPI_ISL_3218101, EPI_ISL_3218202, EPI_ISL_3218298, EPI_ISL_3218511, EPI_ISL_3218512, EPI_ISL_3218513, EPI_ISL_3218514, EPI_ISL_3218549, EPI_ISL_3218615, EPI_ISL_3218723, EPI_ISL_3220024, EPI_ISL_3220328, EPI_ISL_3220336, EPI_ISL_3220364, EPI_ISL_3220501, EPI_ISL_3220513, EPI_ISL_3220536, EPI_ISL_3220578, EPI_ISL_3220582, EPI_ISL_3220589, EPI_ISL_3220667, EPI_ISL_3220672, EPI_ISL_3220675, EPI_ISL_3220708, EPI_ISL_3220969, EPI_ISL_3220989, EPI_ISL_3221005, EPI_ISL_3221007, EPI_ISL_3221008, EPI_ISL_3221048, EPI_ISL_3305299, EPI_ISL_3305372, EPI_ISL_3305444, EPI_ISL_3305461, EPI_ISL_3321358, EPI_ISL_3321441, EPI_ISL_3321445, EPI_ISL_3321480, EPI_ISL_3321481, EPI_ISL_3323016 | | | |
| see above | Aegis Sciences Corporation | Centers for Disease Control and Prevention Division of Viral Diseases, Pathogen Discovery | Adrian Paskey; Alec Vest; Benjamin Rambo-Martin; Christopher Gulvick; Clinton R. Paden; Cyndi Clark; Dakota Howard; Darlene Wagner; Dhwani Batra; Dillon Nall; Duncan MacCannell; Ethan Sanders; Holly Houdeshell; Jason Caravas; Kara Moser; Matthew Hardison; Matthew Schmerer; Ola Kvalvaag; Patrick Campbell; Peter W. Cook; Rob Case; Scott Sammons; Shatavia Morrison; Shaun Westlund; Vikramsinha Ghorpade; Yvette Unoarumhi |
| EPI_ISL_2445953 | Affidea | Instituto Nacional de Saude (INSA) | Borges et al |
| EPI_ISL_2504004, EPI_ISL_2626722, EPI_ISL_2626723, EPI_ISL_2626727, EPI_ISL_2626735, EPI_ISL_2626737, EPI_ISL_2626738, EPI_ISL_2626743, EPI_ISL_2626745, EPI_ISL_2790766, EPI_ISL_2790772, EPI_ISL_2791464, EPI_ISL_2893332, EPI_ISL_2893333, EPI_ISL_2893334, EPI_ISL_2893375, EPI_ISL_2966419, EPI_ISL_2966439, EPI_ISL_2987162, EPI_ISL_3025339 | | | |
| see above | Alaska State Virology Laboratory | Alaska State Virology Laboratory | Elva House; Jack Chen; Jacob Zidek; Lisa Smith; Ph.D.; Stephanie DeRonde |
| EPI_ISL_3388711, EPI_ISL_3388803 | Alverno Labs | RIPHL at Rush University Medical Center | Edith Perez; Felix Araujo Perez; Kevin Kunstman; Laura Furtado; Marieta Hyde; Max Kolton; Melissa Mace; Stefan Green |
| EPI_ISL_2493286, EPI_ISL_2494039, EPI_ISL_2494040, EPI_ISL_2494041, EPI_ISL_2494042, EPI_ISL_2494043, EPI_ISL_2494044, EPI_ISL_2494045, EPI_ISL_2494046, EPI_ISL_2494047, EPI_ISL_2494048, EPI_ISL_2494049, EPI_ISL_2494050, EPI_ISL_2494051, EPI_ISL_2494329, EPI_ISL_2500985 | | | |
| see above | Ambulatório IB | Instituto Butantan | Antonio Jorge Martins; Claudia Renata dos Santos Barros; David Schlesinger; Debora Botequio Moretti; Dimas Tadeu Covas; Elaine Cristina Marqueze; Elaine Vieira Santos; Evandra Strazza Rodrigues; Heidge Fukumasu; Jayme Augusto de Souza-Neto; José Salvatore Leister Patané; Luiz Alcantara; Luiz Lehmann Coutinho; Maria Carolina Elias; Maurício Lacerda Nogueira; Rafael dos Santos Bezerra; Raul Machado Neto; Rejane Maria Tommasini Grotto; Ricardo Haddad; Sandra Coccuzzo Sampaio Vessoni; Simone Kashima; Svetoslav Nanev Slavov; Vincent Louis Viala |
| EPI_ISL_2650135, EPI_ISL_2650136, EPI_ISL_2650137, EPI_ISL_2650141, EPI_ISL_2650142, EPI_ISL_2650148, EPI_ISL_2650151, EPI_ISL_2716405, EPI_ISL_2716406, EPI_ISL_2716407, EPI_ISL_2716411, EPI_ISL_2754301, EPI_ISL_2754305, EPI_ISL_2854198, EPI_ISL_2854199, EPI_ISL_2854200, EPI_ISL_2854201, EPI_ISL_2919071, EPI_ISL_2919072, EPI_ISL_2919073, EPI_ISL_2919076, EPI_ISL_2919078, EPI_ISL_2919079, EPI_ISL_2919081, EPI_ISL_2919082, EPI_ISL_2919083, EPI_ISL_2919084, EPI_ISL_2919093, EPI_ISL_2919097, EPI_ISL_2919101, EPI_ISL_2943052, EPI_ISL_2943060, EPI_ISL_2981919, EPI_ISL_3089183, EPI_ISL_3090069, EPI_ISL_3217104 | | | |
| see above | Arizona State Public Health Laboratory | Arizona State Public Health Laboratory | Jessica Escobar; Katherine Fullerton; Linda Getsinger; Matthew Contursi; Nobuko Fukushima; Stacy White; Trung Huynh; Victor Waddell |
| EPI_ISL_2484113, EPI_ISL_2690351, EPI_ISL_2690380, EPI_ISL_2690385, EPI_ISL_2839100, EPI_ISL_2839101, EPI_ISL_2839102, EPI_ISL_2839103, EPI_ISL_2839104, EPI_ISL_2839105, EPI_ISL_2839106, EPI_ISL_2839107, EPI_ISL_2839108, EPI_ISL_2863926 | | | |
| see above | Arizona State University | Arizona State University | Efrem S. Lim; Joshua LaBaer; Joy M. Blain; LaRinda A. Holland; Matthew F. Smith; Nicholas J. Mellor; Peter T. Skidmore; Rabia Maqsood; Valerie Harris; Vel Murugan |
| EPI_ISL_3504828, EPI_ISL_3553424, EPI_ISL_3553485 | Arkansas Children's Hospital | Center for Global Health, University of New Mexico Health Sciences Center | Ashton Ingold; Bobby Boyanton; Darrell Dinwiddie; Daryl Domman; Joshua L. Kennedy; Kurt Schwalm; Valerie Morley |
| EPI_ISL_2715073, EPI_ISL_2715182, EPI_ISL_3015772, EPI_ISL_3015774 | Atlas Genomics | Seattle Flu Study | Amanda Adler; Barry R. Lutz; Benjamin Pelle; Brian Hiatt; Caitlin R. Wolf; Chris D. Frazar; Deborah A. Nickerson; Elisabeth Brandstetter; Erica Ryke; Geoff Melly; Helen Y. Chu; Janet A. Englund; Jay Shendure; Jover Lee; Kairsten Fay; Kirsten Lacombe; Lea M. Starita; Mark J. Rieder; Matthew Richardson; Matthew Thompson; Melissa Truong; Michael Boeckh; Michael Famulare; Misja Ilcisin; Peter D. Han; Philip Dykema; Romesh Gautom; Scott Lindquist; Thomas R. Sibley; Trevor Bedford |
| EPI_ISL_2758010, EPI_ISL_2758012 | Austrian Agency for Health and Food Safety (AGES) | Bergthaler laboratory, CeMM Research Center for Molecular Medicine of the Austrian Academy of Sciences | Andreas Bergthaler; Anna Schedl; Bekir Erguner; Benedikt Agerer; Christoph Bock; Fabian Amman; Jan Laine; Lukas Endler; Maelle Le Moing; Martin Senekowitsch; Matthew Thornton; Michael Schuster; Petr Triska; Thomas Penz |
| EPI_ISL_2493261, EPI_ISL_2493262, EPI_ISL_2494220, EPI_ISL_2494222, EPI_ISL_2494223 | BIOFAST CENTRO | Instituto Butantan | Antonio Jorge Martins; Claudia Renata dos Santos Barros; David Schlesinger; Debora Botequio Moretti; Dimas Tadeu Covas; Elaine Cristina Marqueze; Elaine Vieira Santos; Evandra Strazza Rodrigues; Heidge Fukumasu; Jayme Augusto de Souza-Neto; José Salvatore Leister Patané; Luiz Alcantara; Luiz Lehmann Coutinho; Maria Carolina Elias; Maurício Lacerda Nogueira; Rafael dos Santos Bezerra; Raul Machado Neto; Rejane Maria Tommasini Grotto; Ricardo Haddad; Sandra Coccuzzo Sampaio Vessoni; Simone Kashima; Svetoslav Nanev Slavov; Vincent Louis Viala |
| EPI_ISL_2493248, EPI_ISL_2494227, EPI_ISL_2494228, EPI_ISL_2494229, EPI_ISL_2494231, EPI_ISL_2494232, EPI_ISL_2494233, EPI_ISL_2494234, EPI_ISL_2494235, EPI_ISL_2494236, EPI_ISL_2494237, EPI_ISL_2494238, EPI_ISL_2494239, EPI_ISL_2494240, EPI_ISL_2494241, EPI_ISL_2494242, EPI_ISL_2494243, EPI_ISL_2494244, EPI_ISL_2494246, EPI_ISL_2494248, EPI_ISL_2494249, EPI_ISL_2494250, EPI_ISL_2494251, EPI_ISL_2494253, EPI_ISL_2494255, EPI_ISL_2494256, EPI_ISL_2494257, EPI_ISL_2494258, EPI_ISL_2494259, EPI_ISL_2494261, EPI_ISL_2494262, EPI_ISL_2494263, EPI_ISL_2494264, EPI_ISL_2494265, EPI_ISL_2494301 | | | |
| see above | BIOFAST LESTE | Instituto Butantan | Antonio Jorge Martins; Claudia Renata dos Santos Barros; David Schlesinger; Debora Botequio Moretti; Dimas Tadeu Covas; Elaine Cristina Marqueze; Elaine Vieira Santos; Evandra Strazza Rodrigues; Heidge Fukumasu; Jayme Augusto de Souza-Neto; José Salvatore Leister Patané; Luiz Alcantara; Luiz Lehmann Coutinho; Maria Carolina Elias; Maurício Lacerda Nogueira; Rafael dos Santos Bezerra; Raul Machado Neto; Rejane Maria Tommasini Grotto; Ricardo Haddad; Sandra Coccuzzo Sampaio Vessoni; Simone Kashima; Svetoslav Nanev Slavov; Vincent Louis Viala |
| EPI_ISL_2493270, EPI_ISL_2494214, EPI_ISL_2494215, EPI_ISL_2494216, EPI_ISL_2494217, EPI_ISL_2494333 | BIOFAST NORTE | Instituto Butantan | Antonio Jorge Martins; Claudia Renata dos Santos Barros; David Schlesinger; Debora Botequio Moretti; Dimas Tadeu Covas; Elaine Cristina Marqueze; Elaine Vieira Santos; Evandra Strazza Rodrigues; Heidge Fukumasu; Jayme Augusto de Souza-Neto; José Salvatore Leister Patané; Luiz Alcantara; Luiz Lehmann Coutinho; Maria Carolina Elias; Maurício Lacerda Nogueira; Rafael dos Santos Bezerra; Raul Machado Neto; Rejane Maria Tommasini Grotto; Ricardo Haddad; Sandra Coccuzzo Sampaio Vessoni; Simone Kashima; Svetoslav Nanev Slavov; Vincent Louis Viala |
| EPI_ISL_2493260, EPI_ISL_2493267, EPI_ISL_2494201, EPI_ISL_2494202, EPI_ISL_2494203, EPI_ISL_2494204, EPI_ISL_2494205, EPI_ISL_2494206, EPI_ISL_2494207, EPI_ISL_2494209, EPI_ISL_2494211, EPI_ISL_2494212, EPI_ISL_2494313 | | | |
| see above | BIOFAST SUL | Instituto Butantan | Antonio Jorge Martins; Claudia Renata dos Santos Barros; David Schlesinger; Debora Botequio Moretti; Dimas Tadeu Covas; Elaine Cristina Marqueze; Elaine Vieira Santos; Evandra Strazza Rodrigues; Heidge Fukumasu; Jayme Augusto de Souza-Neto; José Salvatore Leister Patané; Luiz Alcantara; Luiz Lehmann Coutinho; Maria Carolina Elias; Maurício Lacerda Nogueira; Rafael dos Santos Bezerra; Raul Machado Neto; Rejane Maria Tommasini Grotto; Ricardo Haddad; Sandra Coccuzzo Sampaio Vessoni; Simone Kashima; Svetoslav Nanev Slavov; Vincent Louis Viala |
| EPI_ISL_2933352, EPI_ISL_2933354, EPI_ISL_2933355, EPI_ISL_2933356, EPI_ISL_2933357, EPI_ISL_2933359, EPI_ISL_2933360, EPI_ISL_2933361, EPI_ISL_2933362, EPI_ISL_2933363, EPI_ISL_2933364, EPI_ISL_2933365, EPI_ISL_2933366, EPI_ISL_2933367, EPI_ISL_2933368, EPI_ISL_2933369, EPI_ISL_2933370, EPI_ISL_2933371, EPI_ISL_2933372, EPI_ISL_2933373, EPI_ISL_2933374, EPI_ISL_2933375, EPI_ISL_2933376, EPI_ISL_2933378, EPI_ISL_2933379, EPI_ISL_2933381, EPI_ISL_2933382, EPI_ISL_2933383, EPI_ISL_2933384, EPI_ISL_2933385, EPI_ISL_2933386, EPI_ISL_2933387, EPI_ISL_2933388, EPI_ISL_2933389, EPI_ISL_2933390, EPI_ISL_2933391, EPI_ISL_2933392, EPI_ISL_2933393, EPI_ISL_2933395, EPI_ISL_2933396, EPI_ISL_2933397, EPI_ISL_2933398, EPI_ISL_2933399, EPI_ISL_2933400, EPI_ISL_2933401, EPI_ISL_2933402, EPI_ISL_2933403, EPI_ISL_2933405, EPI_ISL_2933406, EPI_ISL_2933407, EPI_ISL_2933408, EPI_ISL_2933410, EPI_ISL_2933411, EPI_ISL_2933413, EPI_ISL_2933414, EPI_ISL_2933415, EPI_ISL_2933416, EPI_ISL_2933417, EPI_ISL_2933418, EPI_ISL_2933419, EPI_ISL_2933420, EPI_ISL_2933421, EPI_ISL_2933422, EPI_ISL_2933423, EPI_ISL_2933424, EPI_ISL_2933425, EPI_ISL_2933426, EPI_ISL_2933427, EPI_ISL_2933428, EPI_ISL_2933429, EPI_ISL_2933430, EPI_ISL_2933431, EPI_ISL_2933432, EPI_ISL_2933433, EPI_ISL_2933434, EPI_ISL_2933435, EPI_ISL_2933436, EPI_ISL_2933437, EPI_ISL_2933438, EPI_ISL_2933439, EPI_ISL_2933440, EPI_ISL_2933441, EPI_ISL_2933442, EPI_ISL_3130100 | | | |
| see above | BIOMNIS LYON | CNR Virus des Infections Respiratoires - France SUD | Antonin Bal; Bruno Lina; Gregory Destras; Gwendolyne Burfin; Hadrien Regue; Laurence Josset; Martine Valette; Quentin Semanas |
| EPI_ISL_3504577, EPI_ISL_3504653 | Baptist Health Medical Center | Center for Global Health, University of New Mexico Health Sciences Center | Amanda Novack; Darrell Dinwiddie; Daryl Domman; Dirk Haselow; Joshua L. Kennedy; Kurt Schwalm; Valerie Morley |
| EPI_ISL_2658950 | Basurto University Hospital: Clinical Microbiology Laboratory | Biocruces Bizkaia | Ana de la Hoz; Estibaliz Ugalde Zarraga; José Luis Díaz de Tuesta del Arco; Mikel Gallego Rodrigo; Mikel Urrutikoetxea-Gutiérrez; Mª Carmen Nieto Toboso |
| EPI_ISL_2626816, | Baylor Scott & White-Temple | Baylor Scott & White-Temple | Ari Rao; Kimberly Walker; Linden Morales; Marcus Volz; Shelby Johnson |

| | | | |
|---|---|---|---|
| EPI_ISL_2626831, EPI_ISL_2774303, EPI_ISL_2834162, EPI_ISL_2928247, EPI_ISL_2928257 | | | |
| EPI_ISL_2758372, EPI_ISL_2758373 | Berkeley Medical Center | WVU and Marshall University Combined Genomics Core Facilities | James Denvir; Peter Perrotta; Peter Stoilov; Ryan Percifield; Wesley Kimble |
| EPI_ISL_2464480 | Biogroup Bio Lam-LCD Saint-Denis | Department of Virology, Henri Mondor University Hospital, Assistance Publique Hôpitaux de Paris, Université Paris-Est Créteil, INSERM U955 | Alexandre Soulier; Christophe Rodriguez; Elisabeth Trawinski; Guillaume Gricourt; Jean-Michel Pawlotsky; Melissa N'Debi; Slim Fourati; Vanessa Demontant |
| EPI_ISL_3145907, EPI_ISL_3145908 | BioneXt Lab | Laboratoire national de sante, Microbiology, Microbial Genomics Platform | Anke Wienecke-Baldacchino; Catherine Ragimbeau; Elodie Solarino; Fatu Djabi; Jessica Tapp; Lise Pignon; Raoul Salmon; Tamir Abdelrahman; Thibault Ferrandon; Virginie Jover |
| EPI_ISL_2762051 | Bioscientia Labor Wermsdorf | Robert Koch Institute | |
| EPI_ISL_2633914 | Bioscientia MVZ Labor Karlsruhe GmbH | Robert Koch Institute | |
| EPI_ISL_3601715, EPI_ISL_3602355, EPI_ISL_3602558, EPI_ISL_3602851 | British Columbia Centre For Disease Control | BCCDC Public Health Laboratory | Ana Pacagnella; Corrinne Ng; Dan Fornika; John Tyson; Kim Macdonald; Kimia Kamelian; Linda Hoang; Loretta Janz; Mel Krajden; Prystajecky Natalie; Robert Azana; Shannon Russell |
| EPI_ISL_2991515, EPI_ISL_2991536, EPI_ISL_2991562, EPI_ISL_2991713, EPI_ISL_2991774, EPI_ISL_2991856, EPI_ISL_2991908 | | | |
| see above | Broad Institute Clinical Research Sequencing Platform | Infectious Disease Program, Broad Institute of Harvard and MIT | A.E.; Adams, G.; Anahtar, M.; B.L.; B.W.; Bauer, M.; Birren; Branda, J.; Carter, A.; Cerrato, F.; Chaluvadi, S.; Chapman; Cusick, C.; D.J.; DeRuff, K.; Flowers, K.; Gallagher, G.; Gladden-Young, A.; Gnirke, A.; Harris, J.; J.E.; K.J.; LaRocque, R.; Lagerborg, K.; Lemieux; Lin; Loreth, C.; MacInnis; Neumann, A.; Normandin, E.; P.C.; Park; Pierce, V.; Reilly, S.; Rosenberg, E.; Rudy, M.; Ryan, E.; S.B.; Sabeti; Shaw, B.; Siddle; Slater, D.; Smole, S.; Tomkins-Tinch, C.; Turbett, S. |
| EPI_ISL_2787323 | CA-Los Angeles County Public Health Laboratory | Centers for Disease Control and Prevention Division of Viral Diseases, Pathogen Discovery | Alison Laufer Halpin; Ben L. Rambo-Martin; Clinton R. Paden; Dakota Howard; Darlene Wagner; Dave Wentworth; Dhwani Batra; Jasmine Padilla; Justin Lee; Katie Dillon; Krista Queen; Kristen Knipe; Kristine Lacek; Mark Burroughs; Matthew Schmerer; Mili Sheth; Peter Cook; Sam Shepard; Sarah Nobles; Shoshona Le; Suxiang Tong; Vivien Dugan; Yvette Unoarumhi |
| EPI_ISL_2891968, EPI_ISL_2892078, EPI_ISL_2892202, EPI_ISL_2892208, EPI_ISL_2892277, EPI_ISL_2892318, EPI_ISL_2922118, EPI_ISL_2923361, EPI_ISL_2923388, EPI_ISL_2923436, EPI_ISL_2923562, EPI_ISL_2923659, EPI_ISL_2923663, EPI_ISL_2923741, EPI_ISL_2923742, EPI_ISL_3244455, EPI_ISL_3244465, EPI_ISL_3244470, EPI_ISL_3244476, EPI_ISL_3244477, EPI_ISL_3244492, EPI_ISL_3244501, EPI_ISL_3244519, EPI_ISL_3669509, EPI_ISL_3669565, EPI_ISL_3669568, EPI_ISL_3669569, EPI_ISL_3669570, EPI_ISL_3669574, EPI_ISL_3669575, EPI_ISL_3669578, EPI_ISL_3669579, EPI_ISL_3669580, EPI_ISL_3669582, EPI_ISL_3669583, EPI_ISL_3669588, EPI_ISL_3669589, EPI_ISL_3669590, EPI_ISL_3669591, EPI_ISL_3669592, EPI_ISL_3669594, EPI_ISL_3669595, EPI_ISL_3669596, EPI_ISL_3669597, EPI_ISL_3671253, EPI_ISL_3671254, EPI_ISL_3671256, EPI_ISL_3671257, EPI_ISL_3671258, EPI_ISL_3671260, EPI_ISL_3671262, EPI_ISL_3671263, EPI_ISL_3671264, EPI_ISL_3671265, EPI_ISL_3671266, EPI_ISL_3671268, EPI_ISL_3671270, EPI_ISL_3671271, EPI_ISL_3671272, EPI_ISL_3671275, EPI_ISL_3671277, EPI_ISL_3671279, EPI_ISL_3671280, EPI_ISL_3672280, EPI_ISL_3695229, EPI_ISL_3695265, EPI_ISL_3695284, EPI_ISL_3695292, EPI_ISL_3695305, EPI_ISL_3695355, EPI_ISL_3695366, EPI_ISL_3695411, EPI_ISL_3695521, EPI_ISL_3695589, EPI_ISL_3695610, EPI_ISL_3695767, EPI_ISL_3695799, EPI_ISL_3695801, EPI_ISL_3717096 | | | |
| see above | CDPH VBL | California Department of Public Health | CDPH-COVIDNet; Emily Smith on behalf of CDPH-COVIDNet and UCI Genome Sciences Center/GHTF; Emily Smith on behalf of CDPH-COVIDNet and UCSD EXCITE lab |
| EPI_ISL_2933353 | CEDIBIO-UNILABS-TOUNIS | CNR Virus des Infections Respiratoires - France SUD | Antonin Bal; Bruno Lina; Gregory Destras; Gwendolyne Burfin; Hadrien Regue; Laurence Josset; Martine Valette; Quentin Semanas |
| EPI_ISL_2761738, EPI_ISL_2761751, EPI_ISL_2761779, EPI_ISL_2761840, EPI_ISL_2761867 | CENTOGENE Frankfurt Laboratory: Niederlassung Industriepark Höchst | Robert Koch Institute | |
| EPI_ISL_2801364, EPI_ISL_3102261, EPI_ISL_3102274, EPI_ISL_3102278, EPI_ISL_3102288, EPI_ISL_3102305, EPI_ISL_3102495, EPI_ISL_3102496, EPI_ISL_3102497, EPI_ISL_3102498, EPI_ISL_3102499, EPI_ISL_3102501, EPI_ISL_3102502, EPI_ISL_3102503, EPI_ISL_3102504, EPI_ISL_3102507, EPI_ISL_3102524, EPI_ISL_3102525, EPI_ISL_3536352, EPI_ISL_3536354, EPI_ISL_3536355, EPI_ISL_3912138, EPI_ISL_3912139, EPI_ISL_3912140, EPI_ISL_3912141, EPI_ISL_3912142, EPI_ISL_3912143, EPI_ISL_3912144 | | | |
| see above | CENTRO DE ATENDIMENTO PARA ENFRENTAMENTO AO COVID 19 | Analytical Competence Molecular Epidemiology Lab/ACME, Oswaldo Cruz Foundation, Ceara (FIOCRUZ CE) | Cleber Furtado Aksenen; Cleber Furtado Aksenen e Suzana Porto Almeida; Fabio Miyajima; Fernando Braga Stehling; Francisco Eder de Moura Lopes; Jamille Maria Mendes Bezerra; Joaquim Cesar do Nascimento Sousa Junior; Joaquim César do Nascimento Sousa Junior; Pedro Miguel Carneiro Jeronimo; Suzana Porto Almeida & Lucas Delerino on behalf of COVID-19 FIOCRUZ Genomic Network; Suzana Porto Almeida e Lucas Delerino; Thais Ferreira de Oliveira; Thais de Oliveira Costa; Ticiane Cavalcante de Souza; Veridiana Pessoa Miyajima |
| EPI_ISL_2493790, EPI_ISL_2493791 | CENTRO DE GENOMAS | Instituto Butantan | Antonio Jorge Martins; Claudia Renata dos Santos Barros; David Schlesinger; Debora Botequio Moretti; Dimas Tadeu Covas; Elaine Cristina Marqueze; Elaine Vieira Santos; Evandra Strazza Rodrigues; Heidge Fukumasu; Jayme Augusto de Souza-Neto; José Salvatore Leister Patané; Luiz Alcantara; Luiz Lehmann Coutinho; Maria Carolina Elias; Maurício Lacerda Nogueira; Rafael dos Santos Bezerra; Raul Machado Neto; Rejane Maria Tommasini Grotto; Ricardo Haddad; Sandra Coccuzzo Sampaio Vessoni; Simone Kashima; Svetoslav Nanev Slavov; Vincent Louis Viala |
| EPI_ISL_2493824, EPI_ISL_2493826, EPI_ISL_2494127, EPI_ISL_2494128 | CENTRO DE REFERENCIA DO IDOSO DR HUMBERTO MENDES DE CARVALHO | Instituto Butantan | Antonio Jorge Martins; Claudia Renata dos Santos Barros; David Schlesinger; Debora Botequio Moretti; Dimas Tadeu Covas; Elaine Cristina Marqueze; Elaine Vieira Santos; Evandra Strazza Rodrigues; Heidge Fukumasu; Jayme Augusto de Souza-Neto; José Salvatore Leister Patané; Luiz Alcantara; Luiz Lehmann Coutinho; Maria Carolina Elias; Maurício Lacerda Nogueira; Rafael dos Santos Bezerra; Raul Machado Neto; Rejane Maria Tommasini Grotto; Ricardo Haddad; Sandra Coccuzzo Sampaio Vessoni; Simone Kashima; Svetoslav Nanev Slavov; Vincent Louis Viala |
| EPI_ISL_2494309 | CENTRO DE REFERENCIA E TREINAMENTO DSTAIDS SAO PAULO | Instituto Butantan | Antonio Jorge Martins; Claudia Renata dos Santos Barros; David Schlesinger; Debora Botequio Moretti; Dimas Tadeu Covas; Elaine Cristina Marqueze; Elaine Vieira Santos; Evandra Strazza Rodrigues; Heidge Fukumasu; Jayme Augusto de Souza-Neto; José Salvatore Leister Patané; Luiz Alcantara; Luiz Lehmann Coutinho; Maria Carolina Elias; Maurício Lacerda Nogueira; Rafael dos Santos Bezerra; Raul Machado Neto; Rejane Maria Tommasini Grotto; Ricardo Haddad; Sandra Coccuzzo Sampaio Vessoni; Simone Kashima; Svetoslav Nanev Slavov; Vincent Louis Viala |
| EPI_ISL_2494179 | CENTRO DE SAUDE DE ANALANDIA | Instituto Butantan | Antonio Jorge Martins; Claudia Renata dos Santos Barros; David Schlesinger; Debora Botequio Moretti; Dimas Tadeu Covas; Elaine Cristina Marqueze; Elaine Vieira Santos; Evandra Strazza Rodrigues; Heidge Fukumasu; Jayme Augusto de Souza-Neto; José Salvatore Leister Patané; Luiz Alcantara; Luiz Lehmann Coutinho; Maria Carolina Elias; Maurício Lacerda Nogueira; Rafael dos Santos Bezerra; Raul Machado Neto; Rejane Maria Tommasini Grotto; Ricardo Haddad; Sandra Coccuzzo Sampaio Vessoni; Simone Kashima; Svetoslav Nanev Slavov; Vincent Louis Viala |
| EPI_ISL_2493792, EPI_ISL_2493793, EPI_ISL_2493794, EPI_ISL_2493795, EPI_ISL_2493796 | CENTRO DE SAUDE DE PIRAJUI | Instituto Butantan | Antonio Jorge Martins; Claudia Renata dos Santos Barros; David Schlesinger; Debora Botequio Moretti; Dimas Tadeu Covas; Elaine Cristina Marqueze; Elaine Vieira Santos; Evandra Strazza Rodrigues; Heidge Fukumasu; Jayme Augusto de Souza-Neto; José Salvatore Leister Patané; Luiz Alcantara; Luiz Lehmann Coutinho; Maria Carolina Elias; Maurício Lacerda Nogueira; Rafael dos Santos Bezerra; Raul Machado Neto; Rejane Maria Tommasini Grotto; Ricardo Haddad; Sandra Coccuzzo Sampaio Vessoni; Simone Kashima; Svetoslav Nanev Slavov; Vincent Louis Viala |
| EPI_ISL_3536339, EPI_ISL_3536340 | CENTRO DE SAUDE DONA TEREZA HOLANDA DE OLIVEIRA | Analytical Competence Molecular Epidemiology Lab/ACME, Oswaldo Cruz Foundation, Ceara (FIOCRUZ CE) | Cleber Furtado Aksenen; Fabio Miyajima; Fernando Braga Stehling; Francisco Eder de Moura Lopes; Jamille Maria Mendes Bezerra; Joaquim César do Nascimento Sousa Junior; Pedro Miguel Carneiro Jeronimo; Suzana Porto Almeida e Lucas Delerino; Thais Ferreira de Oliveira; Thais de Oliveira Costa; Ticiane Cavalcante de Souza; Veridiana Pessoa Miyajima |
| EPI_ISL_2494195 | CENTRO DE SAUDE II DR JOSE DE FELIPE E S PINHAL SP | Instituto Butantan | Antonio Jorge Martins; Claudia Renata dos Santos Barros; David Schlesinger; Debora Botequio Moretti; Dimas Tadeu Covas; Elaine Cristina Marqueze; Elaine Vieira Santos; Evandra Strazza Rodrigues; Heidge Fukumasu; Jayme Augusto de Souza-Neto; José Salvatore Leister Patané; Luiz Alcantara; Luiz Lehmann Coutinho; Maria Carolina Elias; Maurício Lacerda Nogueira; Rafael dos Santos Bezerra; Raul Machado Neto; Rejane Maria Tommasini Grotto; Ricardo Haddad; Sandra Coccuzzo Sampaio Vessoni; Simone Kashima; Svetoslav Nanev Slavov; Vincent Louis Viala |
| EPI_ISL_2494165, EPI_ISL_2494186 | CENTRO DE SAUDE III DE DIVINOLANDIA | Instituto Butantan | Antonio Jorge Martins; Claudia Renata dos Santos Barros; David Schlesinger; Debora Botequio Moretti; Dimas Tadeu Covas; Elaine Cristina Marqueze; Elaine Vieira Santos; Evandra Strazza Rodrigues; Heidge Fukumasu; Jayme Augusto de Souza-Neto; José Salvatore Leister Patané; Luiz Alcantara; Luiz Lehmann Coutinho; Maria Carolina Elias; Maurício Lacerda Nogueira; Rafael dos Santos Bezerra; Raul Machado Neto; Rejane Maria Tommasini Grotto; Ricardo Haddad; Sandra Coccuzzo Sampaio Vessoni; Simone Kashima; Svetoslav Nanev Slavov; Vincent Louis Viala |
| EPI_ISL_2493822 | CENTRO DE SAUDE III SANTA LUCIA | Instituto Butantan | Antonio Jorge Martins; Claudia Renata dos Santos Barros; David Schlesinger; Debora Botequio Moretti; Dimas Tadeu Covas; Elaine Cristina Marqueze; Elaine Vieira Santos; Evandra Strazza Rodrigues; Heidge Fukumasu; Jayme Augusto de Souza-Neto; José Salvatore Leister Patané; Luiz Alcantara; Luiz Lehmann Coutinho; Maria Carolina Elias; Maurício Lacerda Nogueira; Rafael dos Santos Bezerra; Raul Machado Neto; Rejane Maria Tommasini Grotto; Ricardo Haddad; Sandra Coccuzzo Sampaio Vessoni; Simone Kashima; Svetoslav Nanev Slavov; Vincent Louis Viala |
| EPI_ISL_2494109, EPI_ISL_2494115, EPI_ISL_2494123 | CENTRO DE SAUDE III TABATINGA | Instituto Butantan | Antonio Jorge Martins; Claudia Renata dos Santos Barros; David Schlesinger; Debora Botequio Moretti; Dimas Tadeu Covas; Elaine Cristina Marqueze; Elaine Vieira Santos; Evandra Strazza Rodrigues; Heidge Fukumasu; Jayme Augusto de Souza-Neto; José Salvatore Leister Patané; Luiz Alcantara; Luiz Lehmann Coutinho; Maria Carolina Elias; Maurício Lacerda Nogueira; Rafael dos Santos Bezerra; Raul Machado Neto; Rejane Maria Tommasini Grotto; Ricardo Haddad; Sandra Coccuzzo Sampaio Vessoni; Simone Kashima; Svetoslav Nanev Slavov; Vincent Louis Viala |
| EPI_ISL_3536360, EPI_ISL_3536361 | CENTRO DE SAUDE IRACEMA BRAGA SANDERS | Analytical Competence Molecular Epidemiology Lab/ACME, Oswaldo Cruz Foundation, Ceara (FIOCRUZ CE) | Cleber Furtado Aksenen; Fabio Miyajima; Fernando Braga Stehling; Francisco Eder de Moura Lopes; Jamille Maria Mendes Bezerra; Joaquim César do Nascimento Sousa Junior; Pedro Miguel Carneiro Jeronimo; Suzana Porto Almeida e Lucas Delerino; Thais Ferreira de Oliveira; Thais de Oliveira Costa; Ticiane Cavalcante de Souza; Veridiana Pessoa Miyajima |
| EPI_ISL_2493950, EPI_ISL_2493951 | CENTRO DE SAUDE SAO ROQUE DR JOSE CARVALHO BRITO | Instituto Butantan | Antonio Jorge Martins; Claudia Renata dos Santos Barros; David Schlesinger; Debora Botequio Moretti; Dimas Tadeu Covas; Elaine Cristina Marqueze; Elaine Vieira Santos; Evandra Strazza Rodrigues; Heidge Fukumasu; Jayme Augusto de Souza-Neto; José Salvatore Leister Patané; Luiz Alcantara; Luiz Lehmann Coutinho; Maria Carolina Elias; Maurício Lacerda Nogueira; Rafael dos Santos Bezerra; Raul Machado Neto; Rejane Maria Tommasini Grotto; Ricardo Haddad; Sandra Coccuzzo Sampaio Vessoni; Simone Kashima; Svetoslav Nanev Slavov; Vincent Louis Viala |
| EPI_ISL_2943231 | CENTRO MEDICO NAVAL SEMAR LAB BIOLOGIA MOLECULAR | CENTRO MEDICO NAVAL SEMAR LAB PATOLOGIA MOLECULAR | MEIXUEIRO CALDERON CLAUDIA. MORALES RAMIREZ OMAR. BALTIERRA URIBE SHANTAL. ZUNO CORONADO TEUTLI. GARCIA ALEGRIA IRMA.ALVAREZ JIMENEZ VIOLETA DEJANIRA. LOPEZ BUSTOS ABRAHAM. CABRERA GONZALEZ MOISES. ROMERO ROMAN EDGAR. LIEVANO ESQUINCA JAZMIN. FIGUEROA ANTONIO VICTOR. DIAZ PONCE MEDRANO JUAN ALBERTO. |
| EPI_ISL_2462325, EPI_ISL_2494130 | CENTRO MUNICIPAL DE SAUDE RUY DE CAMARGO | Instituto Butantan | Antonio Jorge Martins; Claudia Renata dos Santos Barros; David Schlesinger; Debora Botequio Moretti; Dimas Tadeu Covas; Elaine Cristina Marqueze; Elaine Vieira Santos; Evandra Strazza Rodrigues; Heidge Fukumasu; Jayme Augusto de Souza-Neto; José Salvatore Leister Patané; Luiz Alcantara; Luiz Lehmann Coutinho; Maria Carolina Elias; Maurício Lacerda Nogueira; Rafael dos Santos Bezerra; Raul Machado Neto; Rejane Maria Tommasini Grotto; Ricardo Haddad; Sandra Coccuzzo Sampaio Vessoni; Simone Kashima; Svetoslav Nanev Slavov; Vincent Louis Viala |
| EPI_ISL_2494159, EPI_ISL_2494188 | CENTRO SAUDE III DR JOSE TADEU MOURAO | Instituto Butantan | Antonio Jorge Martins; Claudia Renata dos Santos Barros; David Schlesinger; Debora Botequio Moretti; Dimas Tadeu Covas; Elaine Cristina Marqueze; Elaine Vieira Santos; Evandra Strazza Rodrigues; Heidge Fukumasu; Jayme Augusto de Souza-Neto; José Salvatore Leister Patané; Luiz Alcantara; Luiz Lehmann Coutinho; Maria Carolina Elias; Maurício Lacerda Nogueira; Rafael dos Santos Bezerra; Raul Machado Neto; Rejane Maria Tommasini Grotto; Ricardo Haddad; Sandra Coccuzzo Sampaio Vessoni; Simone Kashima; Svetoslav Nanev Slavov; Vincent Louis Viala |
| EPI_ISL_2933380 | CERBALLIANCE MIDI-PYRENEES | CNR Virus des Infections Respiratoires - France SUD | Antonin Bal; Bruno Lina; Gregory Destras; Gwendolyne Burfin; Hadrien Regue; Laurence Josset; Martine Valette; Quentin Semanas |
| EPI_ISL_2637834 | CERBALLIANCE Valmy | Department of Virology, Henri Mondor University Hospital, Assistance Publique Hôpitaux de Paris, Université Paris-Est Créteil, INSERM U955 | Alexandre Soulier; Christophe Rodriguez; Elisabeth Trawinski; Guillaume Gricourt; Jean-Michel Pawlotsky; Melissa N'Debi; Slim Fourati; Vanessa Demontant |
| EPI_ISL_2887729 | CH Valenciennes | CHU Lille - Laboratoire de Virologie | AIT YAHYA Emilie; ALIDJINOU Enagnon Kazali; BOCKET Laurence; CREPIN Michel; DEMAY Christophe; ENGELMANN Ilka; GEFFROY Sandrine; GUIGON Aurélie; LAMBERT Valérie; LAZREK Mouna; NOBILLIAUX Florian; PREVOST Brigitte; TCHANTCHOU NJOSSE YANICK; THUILLIER Caroline; TINEZ Claire |
| EPI_ISL_2426982, EPI_ISL_2627848, EPI_ISL_2627853, EPI_ISL_2627854, EPI_ISL_2627855, EPI_ISL_2627856, EPI_ISL_2627857, EPI_ISL_2627858, EPI_ISL_2627859, EPI_ISL_2629224, EPI_ISL_2629232, EPI_ISL_2629233, EPI_ISL_2629234, EPI_ISL_2835977 | | | |

| | | | |
|---|---|---|---|
| see above | CHC Andrée Rosemon | Institut Pasteur de la Guyane | Anne Lavergne; Antoine Enfissi; Arielle Salmier; Dominique Rousset |
| EPI_ISL_2695339, EPI_ISL_2695345, EPI_ISL_2895275 | CHEDV | Instituto Nacional de Saude (INSA) | Borges et al |
| EPI_ISL_3234556 | CHU Purpan - Laboratoire de Virologie - Institut Fédératif de Biologie | CHU Purpan - Laboratoire de Virologie - Institut Fédératif de Biologie | Bulach T.; Donnadieu C.; Izopet J.; Latour J.; Milhes M.; Nicot F.; Ranger N.; Salin G.; Tremeaux P. |
| EPI_ISL_2695098 | CHULN - H Santa Maria | Instituto Nacional de Saude (INSA) | Borges et al |
| EPI_ISL_2637260, EPI_ISL_2957044, EPI_ISL_3030542, EPI_ISL_3030557 | CLILAB | Microbiology Department | Aida Gonzalez-Diaz; Carmen Ardanuy; Jordi Camara; Jordi Niubó; Laura Calatayud; M Angeles Domínguez; Miguel Fernandez-Huerta; Sara Marti |
| EPI_ISL_3298334 | CLINICA BIBLICA | Inciensa, Instituto Costarricense de Investigación y Enseñanza en Nutrición y Salud | Adriana Godínez; Claudio Soto-Garita; Estela Cordero; Francisco Duarte; Hebleen Porras; Joselyn Prado & Karla Gutiérrez; José Luis Vargas; Mariela Gutiérrez; Melany Calderón |
| EPI_ISL_2426983, EPI_ISL_2426984, EPI_ISL_2627860, EPI_ISL_2627861, EPI_ISL_2627864, EPI_ISL_2627865, EPI_ISL_2627866, EPI_ISL_2629235, EPI_ISL_2629236, EPI_ISL_2629237, EPI_ISL_2835978, EPI_ISL_2835979, EPI_ISL_2835980, EPI_ISL_2835981, EPI_ISL_2835982, EPI_ISL_2835984, EPI_ISL_2983480 | | | |
| see above | CNR Institut Pasteur de la Guyane | Institut Pasteur de la Guyane | Anne Lavergne; Antoine Enfissi; Arielle Salmier; Dominique Rousset |
| EPI_ISL_2671834 | COVID LAB | Cayman Islands Forensic Science Lab | Allen Grolla; Brittany Balcewich; Jonathan Smellie; Karla Montes |
| EPI_ISL_2494183 | CS III DR ANTONIO ANADAO SSGRAMA | Instituto Butantan | Antonio Jorge Martins; Claudia Renata dos Santos Barros; David Schlesinger; Debora Botequio Moretti; Dimas Tadeu Covas; Elaine Cristina Marqueze; Elaine Vieira Santos; Evandra Strazza Rodrigues; Heidge Fukumasu; Jayme Augusto de Souza-Neto; José Salvatore Leister Patané; Luiz Alcantara; Luiz Lehmann Coutinho; Maria Carolina Elias; Maurício Lacerda Nogueira; Rafael dos Santos Bezerra; Raul Machado Neto; Rejane Maria Tommasini Grotto; Ricardo Haddad; Sandra Coccuzzo Sampaio Vessoni; Simone Kashima; Svetoslav Nanev Slavov; Vincent Louis Viala |
| EPI_ISL_3912151, EPI_ISL_3912152 | CSF ARACATIACU | Analytical Competence Molecular Epidemiology Lab/ACME, Oswaldo Cruz Foundation, Ceara (FIOCRUZ CE) | Cleber Furtado Aksenen; Fabio Miyajima; Fernando Braga Stehling; Francisco Eder de Moura Lopes; Jamille Maria Mendes Bezerra; Joaquim Cesar do Nascimento Sousa Junior; Pedro Miguel Carneiro Jeronimo; Suzana Porto Almeida & Lucas Delerino on behalf of COVID-19 FIOCRUZ Genomic Network; Thais Ferreira de Oliveira; Thais de Oliveira Costa; Ticiane Cavalcante de Souza; Veridiana Pessoa Miyajima |
| EPI_ISL_3912153 | CSF COELCE | Analytical Competence Molecular Epidemiology Lab/ACME, Oswaldo Cruz Foundation, Ceara (FIOCRUZ CE) | Cleber Furtado Aksenen; Fabio Miyajima; Fernando Braga Stehling; Francisco Eder de Moura Lopes; Jamille Maria Mendes Bezerra; Joaquim Cesar do Nascimento Sousa Junior; Pedro Miguel Carneiro Jeronimo; Suzana Porto Almeida & Lucas Delerino on behalf of COVID-19 FIOCRUZ Genomic Network; Thais Ferreira de Oliveira; Thais de Oliveira Costa; Ticiane Cavalcante de Souza; Veridiana Pessoa Miyajima |
| EPI_ISL_3912155 | CSF JUNCO | Analytical Competence Molecular Epidemiology Lab/ACME, Oswaldo Cruz Foundation, Ceara (FIOCRUZ CE) | Cleber Furtado Aksenen; Fabio Miyajima; Fernando Braga Stehling; Francisco Eder de Moura Lopes; Jamille Maria Mendes Bezerra; Joaquim Cesar do Nascimento Sousa Junior; Pedro Miguel Carneiro Jeronimo; Suzana Porto Almeida & Lucas Delerino on behalf of COVID-19 FIOCRUZ Genomic Network; Thais Ferreira de Oliveira; Thais de Oliveira Costa; Ticiane Cavalcante de Souza; Veridiana Pessoa Miyajima |
| EPI_ISL_3912158 | CSF TERRENOS NOVOS 1 | Analytical Competence Molecular Epidemiology Lab/ACME, Oswaldo Cruz Foundation, Ceara (FIOCRUZ CE) | Cleber Furtado Aksenen; Fabio Miyajima; Fernando Braga Stehling; Francisco Eder de Moura Lopes; Jamille Maria Mendes Bezerra; Joaquim Cesar do Nascimento Sousa Junior; Pedro Miguel Carneiro Jeronimo; Suzana Porto Almeida & Lucas Delerino on behalf of COVID-19 FIOCRUZ Genomic Network; Thais Ferreira de Oliveira; Thais de Oliveira Costa; Ticiane Cavalcante de Souza; Veridiana Pessoa Miyajima |
| EPI_ISL_2493830, EPI_ISL_2494321 | CSII DR WASHINGTON LUIS M RODRIGUES DA SILVA PITANGUEIRAS | Instituto Butantan | Antonio Jorge Martins; Claudia Renata dos Santos Barros; David Schlesinger; Debora Botequio Moretti; Dimas Tadeu Covas; Elaine Cristina Marqueze; Elaine Vieira Santos; Evandra Strazza Rodrigues; Heidge Fukumasu; Jayme Augusto de Souza-Neto; José Salvatore Leister Patané; Luiz Alcantara; Luiz Lehmann Coutinho; Maria Carolina Elias; Maurício Lacerda Nogueira; Rafael dos Santos Bezerra; Raul Machado Neto; Rejane Maria Tommasini Grotto; Ricardo Haddad; Sandra Coccuzzo Sampaio Vessoni; Simone Kashima; Svetoslav Nanev Slavov; Vincent Louis Viala |
| EPI_ISL_2892470, EPI_ISL_2892517, EPI_ISL_3536935, EPI_ISL_4005556, EPI_ISL_4005559, EPI_ISL_4005560, EPI_ISL_4005561, EPI_ISL_4005562, EPI_ISL_4005564, EPI_ISL_4005578 | | | |
| see above | California Department of Public Health | California Department of Public Health | CDPH IDLB COVIDNet et al |
| EPI_ISL_3134074, EPI_ISL_3134105, EPI_ISL_3134106, EPI_ISL_3134109, EPI_ISL_3134116, EPI_ISL_3134118, EPI_ISL_3134121, EPI_ISL_3134124, EPI_ISL_3134128, EPI_ISL_3134129, EPI_ISL_3134147, EPI_ISL_3134158, EPI_ISL_3134162 | | | |
| see above | California Department of Public Health Valencia Branch Laboratory | California Department of Public Health | CDPH-COVIDNet; UCSF Center for Advanced Technology |
| EPI_ISL_2774013, EPI_ISL_3911439, EPI_ISL_3911440, EPI_ISL_3911441, EPI_ISL_3911466, EPI_ISL_3911488 | Cantacuzino National Military-Medical Institute, Viral Respiratory Infections Laboratory | Cantacuzino Institute Virology | Carmen Cherciu; Luiza Ustea; Mihaela Lazar; Mihaela Oprea; Nicoleta Paraschiv; Sorin Dinu |
| EPI_ISL_2502507, EPI_ISL_3462615, EPI_ISL_3462616, EPI_ISL_3462617, EPI_ISL_3462618, EPI_ISL_3462619, EPI_ISL_3462620, EPI_ISL_3462627, EPI_ISL_3462628, EPI_ISL_3462629, EPI_ISL_3462630, EPI_ISL_3462631, EPI_ISL_3462632, EPI_ISL_3462633, EPI_ISL_3462634, EPI_ISL_3462635, EPI_ISL_3462636, EPI_ISL_3462637, EPI_ISL_3462638, EPI_ISL_3462639, EPI_ISL_3462640, EPI_ISL_3462641, EPI_ISL_3462642, EPI_ISL_3462643, EPI_ISL_3462644, EPI_ISL_3462645, EPI_ISL_3462646, EPI_ISL_3462647, EPI_ISL_3462648, EPI_ISL_3462649, EPI_ISL_3462650, EPI_ISL_3462651, EPI_ISL_3462652, EPI_ISL_3462653, EPI_ISL_3462654, EPI_ISL_3462655, EPI_ISL_3462656, EPI_ISL_3462657, EPI_ISL_3462658, EPI_ISL_3462659, EPI_ISL_3462660, EPI_ISL_3462661, EPI_ISL_3462662, EPI_ISL_3462663, EPI_ISL_3462664, EPI_ISL_3462665, EPI_ISL_3462666, EPI_ISL_3462667, EPI_ISL_3462668, EPI_ISL_3462669, EPI_ISL_3462670, EPI_ISL_3462671, EPI_ISL_3462672, EPI_ISL_3462673, EPI_ISL_3462674, EPI_ISL_3462675, EPI_ISL_3462676, EPI_ISL_3462677, EPI_ISL_3462678, EPI_ISL_3462679, EPI_ISL_3462680, EPI_ISL_3462681, EPI_ISL_3462682, EPI_ISL_3462683, EPI_ISL_3462684, EPI_ISL_3462685, EPI_ISL_3462686, EPI_ISL_3462687, EPI_ISL_3462688, EPI_ISL_3462689, EPI_ISL_3462690, EPI_ISL_3462691, EPI_ISL_3462692, EPI_ISL_3462693, EPI_ISL_3462694, EPI_ISL_3462695, EPI_ISL_3462727, EPI_ISL_3462728, EPI_ISL_3464729, EPI_ISL_3464730, EPI_ISL_3464731, EPI_ISL_3464732, EPI_ISL_3464733, EPI_ISL_3464735, EPI_ISL_3464736 | | | |
| see above | Central Laboratory, Bureau of Public Health (BOG) and Academic Hospital Paramaribo | Erasmus Medical Center | Bas B Oude Munnink; Cherise Beek; Consuella Partowidjojo; Dion Gajadin; Ed PF IJzerman; Emmanuelle Munger; Gary Gummels; Ingrid SK Krishnadath; Lycke Woittiez; Marion PG Koopmans; Mireille Van de Veer; Phyllis Pinas; Princes Wongsowidjojo; Radjesh Ori; Ranisha Doerbalie; Rohma Banwari; Soeradj Harkisoen; Stephen Vreden; Tilotmadebie Ramlal; Verne Nanhoe |
| EPI_ISL_3255135, EPI_ISL_3255137, EPI_ISL_3255138, EPI_ISL_3255140, EPI_ISL_3255144, EPI_ISL_3255145, EPI_ISL_3255146, EPI_ISL_3255148, EPI_ISL_3255149, EPI_ISL_3255151, EPI_ISL_3255154, EPI_ISL_3255155, EPI_ISL_3255156, EPI_ISL_3255157, EPI_ISL_3255159, EPI_ISL_3255160, EPI_ISL_3255163, EPI_ISL_3255164, EPI_ISL_3255166, EPI_ISL_3255168, EPI_ISL_3255171, EPI_ISL_3255174, EPI_ISL_3255176, EPI_ISL_3255180, EPI_ISL_3255181, EPI_ISL_3255182, EPI_ISL_3255183, EPI_ISL_3255184, EPI_ISL_3255188, EPI_ISL_3255189, EPI_ISL_3255190, EPI_ISL_3255196, EPI_ISL_3255203, EPI_ISL_3255204, EPI_ISL_3255210, EPI_ISL_3255211, EPI_ISL_3255213, EPI_ISL_3255215, EPI_ISL_3255217, EPI_ISL_3255218, EPI_ISL_3255220, EPI_ISL_3255222, EPI_ISL_3255224, EPI_ISL_3255225, EPI_ISL_3255228, EPI_ISL_3255229, EPI_ISL_3255231, EPI_ISL_3255234, EPI_ISL_3255235, EPI_ISL_3255236, EPI_ISL_3255238, EPI_ISL_3255239, EPI_ISL_3255241, EPI_ISL_3255242, EPI_ISL_3255243, EPI_ISL_3255245, EPI_ISL_3255247, EPI_ISL_3255249, EPI_ISL_3255250, EPI_ISL_3255252, EPI_ISL_3255254, EPI_ISL_3255257, EPI_ISL_3255258, EPI_ISL_3255261, EPI_ISL_3255262, EPI_ISL_3255263, EPI_ISL_3255264, EPI_ISL_3255266, EPI_ISL_3255269, EPI_ISL_3255270, EPI_ISL_3255271, EPI_ISL_3255272, EPI_ISL_3255273, EPI_ISL_3255274, EPI_ISL_3255275, EPI_ISL_3255276, EPI_ISL_3255277, EPI_ISL_3255278, EPI_ISL_3255279, EPI_ISL_3255280, EPI_ISL_3255281 | | | |
| see above | Central Public Health Laboratory - LACEN - Bahia, Salvador, Brazil | Central Public Health Laboratory - LACEN -Bahia, Salvador, Brazil | Arabela Leal; Breno Dominguez; Felicidade Pereira; Jaqueline Gomes; Luciana Oliveira; Luiz Alcantara; Marcela Gómez; Marta Giovanetti; Patrícia Cajado; Stephane Tosta; Vagner Fonseca; Vanessa Nardy |
| EPI_ISL_3832900 | Central Washington Hospital | Seattle Flu Study | Amanda Adler; Barry R. Lutz; Benjamin Pelle; Brian Hiatt; Caitlin R. Wolf; Chris D. Frazar; Deborah A. Nickerson; Elisabeth Brandstetter; Erica Ryke; Geoff Melly; Helen Y. Chu; Janet A. Englund; Jay Shendure; Jover Lee; Kairsten Fay; Kirsten Lacombe; Lea M. Starita; Mark J. Rieder; Matthew Richardson; Matthew Thompson; Melissa Truong; Michael Boeckh; Michael Famulare; Misja Ilcisin; Peter D. Han; Philip Dykema; Romesh Gautom; Scott Lindquist; Thomas R. Sibley; Trevor Bedford |
| EPI_ISL_2491661 | Centre Hospitalier Universitaire Clermont-Ferrand | CHU Clermont-Ferrand, service de virologie | Bisseux Maxime; Combes Patricia; Henquell Cécile; Mirand Audrey |
| EPI_ISL_2671574, EPI_ISL_2671575, EPI_ISL_2671576, EPI_ISL_2671582, EPI_ISL_2671585, EPI_ISL_2671586, EPI_ISL_2671703, EPI_ISL_2942410, EPI_ISL_2942411, EPI_ISL_2942418, EPI_ISL_2942426, EPI_ISL_2942428, EPI_ISL_2942434, EPI_ISL_2942438, EPI_ISL_2942440, EPI_ISL_2942460, EPI_ISL_2942732, EPI_ISL_2942737 | | | |
| see above | Centro de Investigacion Biomedica de Occidente (CIBO) | Unidad de Genomica Avanzada | ; Alejandra Garcia-Gasca; Alejandra Hernandez-Teran; Alejandro Sanchez-Flores; Alfredo Herrera-Estrella; Alicia Ocaña-Mondragon; Andreu Comas-Garcia; Angel Gustavo Salas-Lais; Antonio Loza Roman; Bernardo Martinez-Miguel; Blanca Taboada; Brenda Irasema Maldonado-Meza; Bruno Gomez-Gil; Carla Ivon Herrera-Najera; Carlos F. Arias; Celia Boukadida; Celida Duque Molina; Celida Martinez- Rodriguez; Clara Esperanza Santacruz-Tinoco; Concepcion Grajales-Muñiz; Consorcio Mexicano de Vigilancia Genomica (CoViGen-Mex). Authors (in alphabetical order): Julio Elias Alvarado-Yaah; Cristobal Chaidez-Quiroz; Daniel Fregoso-Rueda; Daniel Lira Morales; Eduardo Becerril-Vargas; Fernando Fontove-Herrera; Fidencio Mejia-Nepomuceno; Francisco Pulido; Gloria Elena Espinosa-Ayala; Gloria Maria Molina-Salinas; Gloria Vazquez; Hector Esteban Paz-Juarez; Hector Montoya-Fuentes; Helen Haydee Fernanda Ramirez-Plascencia; Irvin Gonzalez-Lopez; Jean Pierre Gonzalez; Jesus Hernandez; Joel Armando Vazquez-Perez.; Jorge Salas-Hernandez; Jose Antonio Enciso-Moreno; Jose Arturo Martinez-Orozco; Jose Esteban Muñoz-Medina; Jose de Jesus Nuñez-Contreras; Juan Bautista Chale-Dzul; Julissa Enciso-Ibarra; Luis Alberto Ochoa-Carrera; Margarita Matias-Florentino; Maria Guadalupe Santiago-Mauricio; Maria Guadalupe de Jesus Mireles-Rivera; Mario Mujica-Sanchez; Marissa Perez-Garcia; Nelly Selem-Mojica; Pavel Isa; Ricardo Ciria Merce; Ricardo Grande; Rosa Maria Gutierrez Rios; Santiago avila-Rios; Selene Zarate; Susana Lopez; Veronica Mata-Haro; Victor Eduardo Garcia-Arias; Victor Hugo Borja-Aburto |
| EPI_ISL_2671560, EPI_ISL_2942484, EPI_ISL_2942755, EPI_ISL_2942756, EPI_ISL_2942770 | Centro de Investigacion Biomedica del Noreste (CIBIN) | Unidad de Genomica Avanzada | ; Alejandra Garcia-Gasca; Alejandra Hernandez-Teran; Alejandro Sanchez-Flores; Alfredo Herrera-Estrella; Alicia Ocaña-Mondragon; Andreu Comas-Garcia; Angel Gustavo Salas-Lais; Antonio Loza Roman; Bernardo Martinez-Miguel; Blanca Taboada; Brenda Irasema Maldonado-Meza; Bruno Gomez-Gil; Carla Ivon Herrera-Najera; Carlos F. Arias; Celia Boukadida; Celida Duque Molina; Celida Martinez- Rodriguez; Clara Esperanza Santacruz-Tinoco; Concepcion Grajales-Muñiz; Consorcio Mexicano de Vigilancia Genomica (CoViGen-Mex). Authors (in alphabetical order): Julio Elias Alvarado-Yaah; Cristobal Chaidez-Quiroz; Daniel Fregoso-Rueda; Daniel Lira Morales; Eduardo Becerril-Vargas; Fernando Fontove-Herrera; Fidencio Mejia-Nepomuceno; Francisco Pulido; Gloria Elena Espinosa-Ayala; Gloria Maria Molina-Salinas; Gloria Vazquez; Hector Esteban Paz-Juarez; Hector Montoya-Fuentes; Helen Haydee Fernanda Ramirez-Plascencia; Irvin Gonzalez-Lopez; Jean Pierre Gonzalez; Jesus Hernandez; Joel Armando Vazquez-Perez.; Jorge Salas-Hernandez; Jose Antonio Enciso-Moreno; Jose Arturo Martinez-Orozco; Jose Esteban Muñoz-Medina; Jose de Jesus Nuñez-Contreras; Juan Bautista Chale-Dzul; Julissa Enciso-Ibarra; Luis Alberto Ochoa-Carrera; Margarita Matias-Florentino; Maria Guadalupe Santiago-Mauricio; Maria Guadalupe de Jesus Mireles-Rivera; Mario Mujica-Sanchez; Marissa Perez-Garcia; Nelly Selem-Mojica; Pavel Isa; Ricardo Ciria Merce; Ricardo Grande; Rosa Maria Gutierrez Rios; Santiago avila-Rios; Selene Zarate; Susana Lopez; Veronica Mata-Haro; Victor Eduardo Garcia-Arias; Victor Hugo Borja-Aburto |
| EPI_ISL_3019538, EPI_ISL_3129919 | Centro de Investigación Biomédica de La Rioja - Hospital San Pedro Logroño | SeqCOVID-SPAIN consortium/IBV(CSIC) | José Manuel Azcona Gutiérrez; María Pilar Bea Escudero; María de Toro; Miriam Blasco Alberdi and SeqCOVID-SPAIN consortium |
| EPI_ISL_3155465, EPI_ISL_3155471, EPI_ISL_3155472, EPI_ISL_3155489, EPI_ISL_3155492, EPI_ISL_3155494, EPI_ISL_3155500 | | | |
| see above | Centro de Investigación Biomédica de Occidente (CIBO) | Centro de Investigación en Enfermedades Infecciosas (CIENI), Instituto Nacional de Enfermedades Respiratorias (INER) | ; Alejandra García-Gasca; Alejandra Hernández-Terán; Alejandro Sánchez-Flores; Alfredo Herrera-Estrella; Alicia Ocaña-Mondragón; Andreu Comas-García; Angel Gustavo Salas-Lais; Antonio Loza Román; Bernardo Martínez-Miguel; Blanca Taboada; Brenda Irasema Maldonado-Meza; Bruno Gómez-Gil; Carla Ivón Herrera-Najera; Carlos F. Arias; Celia Boukadida; Clara Esperanza Santacruz-Tinoco; Concepción Grajales-Muñiz; Consorcio Mexicano de Vigilancia Genómica (CoViGen-Mex). Authors (in alphabetical order): Julio Elias Alvarado-Yaah; Cristóbal Cháidez-Quiróz; Célida Duque Molina; Célida Martínez- Rodríguez; Daniel Fregoso-Rueda; Daniel Lira Morales; Eduardo Becerril-Vargas; Fernando Fontove-Herrera; Fidencio Mejía-Nepomuceno; Francisco Pulido; Gloria Elena Espinosa-Ayala; Gloria María Molina-Salinas; Gloria Vazquez; Hector Esteban Paz-Juárez; Hector Montoya-Fuentes; Helen Haydee Fernanda Ramírez-Plascencia; Irvin González-López; Jean Pierre González; Jesús Hernández; Joel Armando Vázquez-Pérez.; Jorge Salas-Hernández; José Antonio Enciso-Moreno; José Arturo Martínez-Orozco; José Esteban Muñoz-Medina; José de Jesús Nuñez-Contreras; Juan Bautista Chale-Dzul; Julissa Enciso-Ibarra; Kathia Elizabeth Tapia-Diaz; Luis Alberto Ochoa-Carrera; Margarita Matías-Florentino; Mario Mújica-Sánchez; Marissa Perez-Garcia; María Guadalupe Santiago-Mauricio; María Guadalupe de Jesús Mireles-Rivera; Nelly Sélem-Mojica; Pavel Isa; Ricardo Ciria Merce; Ricardo Grande; Rosa María Gutiérrez Rios; Santiago Ávila-Ríos; Selene Zárate; Susana Lopez; Verónica Mata-Haro; Victor Eduardo García-Arias; Víctor Hugo Borja-Aburto |
| EPI_ISL_2801721, EPI_ISL_2801726, EPI_ISL_2801729, EPI_ISL_2801731, EPI_ISL_2801733, EPI_ISL_2801736, EPI_ISL_2801742, EPI_ISL_2801746, EPI_ISL_2801750 | | | |
| see above | Centro de Investigación Biomédica de Occidente (CIBO) | Instituto de Biotecnología de la UNAM | ; Alejandra García-Gasca; Alejandra Hernández-Terán; Alejandro Sánchez-Flores; Alfredo Herrera-Estrella; Alicia Ocaña-Mondragón; Andreu Comas-García; Angel Gustavo Salas-Lais; Antonio Loza Román; Bernardo Martínez-Miguel; Blanca Taboada; Brenda Irasema Maldonado-Meza; Bruno Gómez-Gil; Carla Ivón Herrera-Najera; Carlos F. Arias; Celia Boukadida; Clara Esperanza Santacruz-Tinoco; Concepción Grajales-Muñiz; Consorcio Mexicano de Vigilancia Genómica (CoViGen-Mex). Authors (in alphabetical order): Julio Elias Alvarado-Yaah; Cristóbal Cháidez-Quiróz; Célida Duque Molina; Célida Martínez- Rodríguez; Daniel Fregoso-Rueda; Daniel Lira Morales; Eduardo Becerril-Vargas; Fernando Fontove-Herrera; Fidencio Mejía-Nepomuceno; Francisco Pulido; Gloria Elena Espinosa-Ayala; Gloria María Molina-Salinas; Gloria Vazquez; Hector Esteban Paz-Juárez; Hector Montoya-Fuentes; Helen Haydee Fernanda Ramírez-Plascencia; Irvin González-López; Jean Pierre González; Jesús Hernández; Joel Armando Vázquez-Pérez.; Jorge Salas-Hernández; José Antonio Enciso-Moreno; José Arturo Martínez-Orozco; José Esteban Muñoz-Medina; José de Jesús Nuñez-Contreras; Juan Bautista Chale-Dzul; |

| | | | |
|---|---|---|---|
| | | | Julissa Enciso-Ibarra; Luis Alberto Ochoa-Carrera; Margarita Matías-Florentino; Mario Mújica-Sánchez; Marissa Perez-Garcia; María Guadalupe Santiago-Mauricio; María Guadalupe de Jesús Mireles-Rivera; Nelly Sélem-Mojica; Pavel Isa; Ricardo Ciria Merce; Ricardo Grande; Rosa María Gutiérrez Rios; Santiago Ávila-Ríos; Selene Zárate; Susana Lopez; Verónica Mata-Haro; Víctor Eduardo García-Arias; Víctor Hugo Borja-Aburto |
| EPI_ISL_2681227, EPI_ISL_2801658, EPI_ISL_2801669, EPI_ISL_2801674, EPI_ISL_2801680, EPI_ISL_2801681 | Centro de Investigación Biomédica del Noreste (CIBIN) | Instituto de Biotecnología de la UNAM | ; Alejandra García-Gasca; Alejandra Hernández-Terán; Alejandro Sánchez-Flores; Alfredo Herrera-Estrella; Alicia Ocaña-Mondragón; Andreu Comas-García; Angel Gustavo Salas-Lais; Antonio Loza Román; Bernardo Martínez-Miguel; Blanca Taboada; Brenda Irasema Maldonado-Meza; Bruno Gómez-Gil; Carla Ivón Herrera-Najera; Carlos F. Arias; Celia Boukadida; Clara Esperanza Santacruz-Tinoco; Concepción Grajales-Muñiz; Consorcio Mexicano de Vigilancia Genómica (CoViGen-Mex). Authors (in alphabetical order): Julio Elias Alvarado-Yaah; Cristóbal Cháidez-Quiróz; Célida Duque Molina; Célida Martínez- Rodríguez; Daniel Fregoso-Rueda; Daniel Lira Morales; Eduardo Becerril-Vargas; Fernando Fontove-Herrera; Fidencio Mejía-Nepomuceno; Francisco Pulido; Gloria Elena Espinosa-Ayala; Gloria María Molina-Salinas; Gloria Vazquez; Hector Esteban Paz-Juárez; Hector Montoya-Fuentes; Helen Haydee Fernanda Ramírez-Plascencia; Irvin González-López; Jean Pierre González; Jesús Hernández; Joel Armando Vázquez-Pérez.; Jorge Salas-Hernández; José Antonio Enciso-Moreno; José Arturo Martínez-Orozco; José Esteban Muñoz-Medina; José de Jesús Nuñez-Contreras; Juan Bautista Chale-Dzul; Julissa Enciso-Ibarra; Luis Alberto Ochoa-Carrera; Margarita Matías-Florentino; Mario Mújica-Sánchez; Marissa Perez-Garcia; María Guadalupe Santiago-Mauricio; María Guadalupe de Jesús Mireles-Rivera; Nelly Sélem-Mojica; Pavel Isa; Ricardo Ciria Merce; Ricardo Grande; Rosa María Gutiérrez Rios; Santiago Ávila-Ríos; Selene Zárate; Susana Lopez; Verónica Mata-Haro; Víctor Eduardo García-Arias; Víctor Hugo Borja-Aburto |
| EPI_ISL_3242687, EPI_ISL_3242691, EPI_ISL_3242693, EPI_ISL_3242696, EPI_ISL_3242700, EPI_ISL_3242706, EPI_ISL_3242707, EPI_ISL_3242708, EPI_ISL_3242709, EPI_ISL_3242710, EPI_ISL_3242712, EPI_ISL_3242714, EPI_ISL_3242716, EPI_ISL_3242718, EPI_ISL_3242726, EPI_ISL_3242743 | | | |
| see above | Clinica INDISA | "Facultad de Ciencias de la Vida, UNAB" | "Claudio Meneses; Ariel Orellana"; Claudio Olmos; Daniel Leon; Dayan Sanhueza; Eduardo Castro; Gonzalo Campaña; Macarena Bastias; Paola Pidal; Ricardo Yusta; Sebastian Wolter; Susana Saez; Víctor Monreal; Waldo Diaz |
| EPI_ISL_2988680, EPI_ISL_3025679, EPI_ISL_3025685 | Clinical Microbiology, Infection Prevention and Control | Section for Molecular Diagnostics | Björn Hallström; Jonas Björkman |
| EPI_ISL_2466606, EPI_ISL_2610956 | Clinical Virology | Clinical Bacteriology | Adrian Egli; Alfredo Mari; Fanny Wegner; Hans Hirsch; Helena MB Seth-Smith; Julia Bielicki; Karoline Leuzinger; Manuel Battegay; Tim Roloff |
| EPI_ISL_2550357, EPI_ISL_2790623, EPI_ISL_2790625, EPI_ISL_2790645, EPI_ISL_2790667, EPI_ISL_2941636, EPI_ISL_3160140, EPI_ISL_3404119, EPI_ISL_3404121, EPI_ISL_3696528, EPI_ISL_3842262 | | | |
| see above | Colorado Department of Public Health and Environment | Colorado Department of Public Health and Environment | Alexandria Rossheim; Diana Ir; Emily A. Travanty; Laura Bankers; Mandy Waters; Michael A. Martin; Michael Martin; Molly C. Hetherington-Rauth; Sarah Elizabeth Totten; Shannon R. Matzinger |
| EPI_ISL_2930338 | Cotugno | TIGEM | Antonio Grimaldi Patrizia Annunziata Francesco Panariello Biancamaria Pierri Claudia Tiberio Teresa Giuliano Valentina Bouche Chiara Colantuono Maria Concetta Cuomo Denise Di Concilio Lucio Di Filippo Anna Manfredi Marcello Salvi Antonio Limone Luigi Atripaldi Pellegrino Cerino Andrea Ballabio Davide Cacchiarelli |
| EPI_ISL_2658982, EPI_ISL_2659116 | County of San Luis Obispo Public Health Laboratory | County of San Luis Obispo Public Health Laboratory | Frances Sidhu; Katrina Erwin |
| EPI_ISL_3273362 | Curative Labs | Curative Labs | Elias L. Salfati; Eugenia Khorosheva; George Way; J.Cesar Ignacio-Espinoza; Janet Chen; Mikhail Hanewich-Hollatz; Nabjot Sandhu; Sophia Quasem; Vladimir Slepnev; Zhiyi Xie |
| EPI_ISL_2941667, EPI_ISL_2941681, EPI_ISL_2941683, EPI_ISL_2941687 | Curative Labs | New Mexico Department of Health Scientific Laboratory | ; Anastacia Griego-Fisher; D'eldra Malone; Ellie Johnson; Jennifer Benoit; Keila Gutierrez; Linda Salazar; Ratheesh Rajan |
| EPI_ISL_3664275 | DASA | DASA | Adriano Bonaldi; Angelica Hristov; Annelise Lopes; Bianca Cota; Cristina Oliveira; Jose Levi; Lidia Yamamoto; Paulo Pierry; Rodrigo Guarischi; Rodrigo Salazar |
| EPI_ISL_2612553, EPI_ISL_2612554, EPI_ISL_2612555 | DOHMH Corona | New York City Public Health Laboratory | Jade Wang; et al. |
| EPI_ISL_2662745, EPI_ISL_2662750, EPI_ISL_2662763, EPI_ISL_2662776, EPI_ISL_3692598, EPI_ISL_3692608, EPI_ISL_3692632, EPI_ISL_3692660, EPI_ISL_3692719 | | | |
| see above | DPHL | Delaware Public Health Lab | Rebecca Savage |
| EPI_ISL_2429493, EPI_ISL_2431024, EPI_ISL_2665775, EPI_ISL_2666225, EPI_ISL_2667603, EPI_ISL_2669007, EPI_ISL_2669330, EPI_ISL_2669458, EPI_ISL_2671258, EPI_ISL_2675490, EPI_ISL_2747517, EPI_ISL_2748057, EPI_ISL_2748372, EPI_ISL_2784618, EPI_ISL_2819380 | | | |
| see above | Department of Bacteria, Parasites and Fungi, Statens Serum Institut, Copenhagen, Denmark | Statens Serum Institut Bioinformatics and Microbial Genomics | Danish Covid-19 Genome Consortium |
| EPI_ISL_3135153, EPI_ISL_3135891 | Department of Virology and Immunology, University of Helsinki and Helsinki University Hospital, Huslab Finland | Department of Virology, Faculty of Medicine, University of Helsinki, Helsinki, Finland | Essi Korhonen; Hanna Jarva; Hanna Liimatainen; Hannimari Kallio-Kokko; Harri Kangas; Hussein Alburkat; Jenni Virtanen; Maija Lappalainen; Maija Suvanto; Olli Vapalahti; Pekka Ellonen; Phuoc Truong; Ravi Kant; Sari Hannula; Satu Kurkela; Teemu Smura |
| EPI_ISL_2503963 | Diagnostyka | 1. Academic Center for Pathomorphological and Genetic-Molecular Diagnostics ltd, Bialystok, Poland 2. National Institute of Public Health - National Institute of Hygiene, Warsaw, Poland | Anetta Sulewska; Jacek Nikliński; Janusz Dzięcioł; Joanna Kiśluk; Joanna Zacharczuk; Konrad Raczkowski; Magdalena Nowakowska; Małgorzata Sadkowska-Todys; Piotr Karabowicz; Piotr Majewski; Przemysław Biecek. Joanna Reszeć; Radosław Charkiewicz; Tomasz Wołkowicz |
| EPI_ISL_3050671, EPI_ISL_3050674, EPI_ISL_3050700, EPI_ISL_3050701, EPI_ISL_3050704, EPI_ISL_3050720, EPI_ISL_3050723, EPI_ISL_3050725, EPI_ISL_3372572, EPI_ISL_3372573, EPI_ISL_3372577, EPI_ISL_3372580, EPI_ISL_3372584 | | | |
| see above | Dirección regional de salud del Callao (DIRESA-CALLAO) | Centro de Investigaciones Tecnológicas, Biomédicas y Medioambientales (CITBM) | B; Huaman; J. Alarcon; M. Cuellar; M. Ramirez; M. Sovero |
| EPI_ISL_2967363, EPI_ISL_2967364, EPI_ISL_2967365, EPI_ISL_2967403, EPI_ISL_2967544, EPI_ISL_2967545, EPI_ISL_3933507 | | | |
| see above | Division of Emerging Infectious Diseases, Bureau of Infectious Diseases Diagnosis Control, Korea Disease Control and Prevention Agency | Division of Emerging Infectious Diseases, Bureau of Infectious Diseases Diagnosis Control, Korea Disease Control and Prevention Agency | Ae Kyung Park; Chae Young Lee; Eun-Jin Kim; Heui Man Kim; Il-Hwan Kim; Jeong-Ah Kim; Jeong-Min Kim |
| EPI_ISL_2405572, EPI_ISL_2405606, EPI_ISL_2406331, EPI_ISL_2475680, EPI_ISL_2475694, EPI_ISL_2475695, EPI_ISL_2475701, EPI_ISL_2610234, EPI_ISL_2610296, EPI_ISL_2610674, EPI_ISL_2610700, EPI_ISL_2610733, EPI_ISL_2610743, EPI_ISL_2672423, EPI_ISL_2672546, EPI_ISL_2672729, EPI_ISL_2673170, EPI_ISL_2673225, EPI_ISL_2787886, EPI_ISL_2787900, EPI_ISL_2787902, EPI_ISL_2787905, EPI_ISL_2787907, EPI_ISL_2862759, EPI_ISL_2862761, EPI_ISL_2862765, EPI_ISL_2862769, EPI_ISL_2981755, EPI_ISL_2981895, EPI_ISL_3136605, EPI_ISL_3136615, EPI_ISL_3137106 | | | |
| see above | Dutch COVID-19 response team | National Institute for Public Health and the Environment (RIVM) | Adam Meijer; AnneMarie van den Brandt; Annelies Kroneman; Bas van der Veer; Chantal Reusken; Dennis Schmitz; Dirk Eggink; Euníce Then; Florian Zwagemaker; Harry Vennema; James Groot; Jeroen Cremer; Karim Hajji; Kim Freriks; Linda van de Nes; Lisa Wijsman; Lynn Aarts; Melissa van Tuil; Ryanne Jaarsma; Sanne Bos; Sharon van den Brink; Sjoerd Kuiling; on behalf of the national COVID-19 response team |
| EPI_ISL_4059813, EPI_ISL_4059814, EPI_ISL_4059815, EPI_ISL_4059816, EPI_ISL_4059817, EPI_ISL_4059818, EPI_ISL_4059819, EPI_ISL_4059820, EPI_ISL_4059821, EPI_ISL_4059822, EPI_ISL_4059823, EPI_ISL_4059824, EPI_ISL_4060090, EPI_ISL_4060091, EPI_ISL_4060093, EPI_ISL_4060094, EPI_ISL_4060095, EPI_ISL_4060103, EPI_ISL_4060106, EPI_ISL_4060107, EPI_ISL_4060109, EPI_ISL_4060110 | | | |
| see above | Dynacare Manitoba | Manitoba Cadham Provincial Laboratory | David Alexander; Hilary Racher; Hongbin Li; Melissa Desaulnier; Tintu Abraham |
| EPI_ISL_3215320, EPI_ISL_3215323, EPI_ISL_3215815, EPI_ISL_3475847, EPI_ISL_3475851 | EXCITE Lab | Andersen lab at Scripps Research | Abigail Schnapper; Angela Scioscia; Araz Majnoonian; Ashkan Hassani; Chip Schooley; David Pride; Fitri Carrissa Wijaya; Greg Humphrey; Helena Tubb; Laura Kohn; Marlene Flores; Megan Nguyen; Natasha Martin Cheryl Anderson; Patricia Gonzalez-Zuniga; Rebecca Fielding-Miller; Rodolfo Salido; Sawyer Farmer; Sharon Reed; Smruthi Karthikeyan; Tommy Valles + SEARCH; Victor Cantu; Vinton Omaleki |
| EPI_ISL_3718929 | Emory Molecular Diagnostics Laboratory, Emory Healthcare | Piantadosi Lab, Emory Department of Pathology | Ahmed Babiker; Anne Piantadosi; Ludy Registre Carmola |
| EPI_ISL_3246302 | Eurofins Diatherix | Hudsonalpha Genome Sequencing Center | Ada Stewart; Jane Grimwood; Jenell Webber; Jennifer Cart; John Lovell; Joshua Stough; Leslie Malone; Lori H. Handley; Melissa Williams; Stefan Brzezinski; Teresa Jones; and Jeremy Schmutz |
| EPI_ISL_2634510, EPI_ISL_2634515, EPI_ISL_2634522, EPI_ISL_2634526, EPI_ISL_2634534, EPI_ISL_2634536, EPI_ISL_2634551, EPI_ISL_2634563, EPI_ISL_2634567, EPI_ISL_2634585, EPI_ISL_2634605, EPI_ISL_2634610, EPI_ISL_2634612, EPI_ISL_2634619, EPI_ISL_2634652, EPI_ISL_2634667, EPI_ISL_2634669, EPI_ISL_2634687 | | | |
| see above | Eurofins LifeCodexx GmbH | Robert Koch Institute | |
| EPI_ISL_2494267, EPI_ISL_2494269, EPI_ISL_2494271, EPI_ISL_2494272, EPI_ISL_2494302 | FACULDADE DE MEDICINA ABC | Instituto Butantan | Antonio Jorge Martins; Claudia Renata dos Santos Barros; David Schlesinger; Debora Botequio Moretti; Dimas Tadeu Covas; Elaine Cristina Marqueze; Elaine Vieira Santos; Evandra Strazza Rodrigues; Heidge Fukumasu; Jayme Augusto de Souza-Neto; José Salvatore Leister Patané; Luiz Alcantara; Luiz Lehmann Coutinho; Maria Carolina Elias; Maurício Lacerda Nogueira; Rafael dos Santos Bezerra; Raul Machado Neto; Rejane Maria Tommasini Grotto; Ricardo Haddad; Sandra Coccuzzo Sampaio Vessoni; Simone Kashima; Svetoslav Nanev Slavov; Vincent Louis Viala |
| EPI_ISL_2840301 | FL Bur. of Public Health Laboratories-Jacksonville | Centers for Disease Control and Prevention Division of Viral Diseases, Pathogen Discovery | Alison Laufer Halpin; Ben L. Rambo-Martin; Clinton R. Paden; Dakota Howard; Darlene Wagner; Dave Wentworth; Dhwani Batra; Jasmine Padilla; Justin Lee; Katie Dillon; Krista Queen; Kristen Knipe; Kristine Lacek; Mark Burroughs; Matthew Schmerer; Mili Sheth; Peter Cook; Sam Shepard; Sarah Nobles; Shoshona Le; Suxiang Tong; Vivien Dugan; Yvette Unoarumhi |
| EPI_ISL_2840425 | FL Bureau of Public Health Laboratories-Miami | Centers for Disease Control and Prevention Division of Viral Diseases, Pathogen Discovery | Alison Laufer Halpin; Ben L. Rambo-Martin; Clinton R. Paden; Dakota Howard; Darlene Wagner; Dave Wentworth; Dhwani Batra; Jasmine Padilla; Justin Lee; Katie Dillon; Krista Queen; Kristen Knipe; Kristine Lacek; Mark Burroughs; Matthew Schmerer; Mili Sheth; Peter Cook; Sam Shepard; Sarah Nobles; Shoshona Le; Suxiang Tong; Vivien Dugan; Yvette Unoarumhi |
| EPI_ISL_2787209, EPI_ISL_3355380 | FL Bureau of Public Health Laboratories-Tampa | Centers for Disease Control and Prevention Division of Viral Diseases, Pathogen Discovery | Alex Burgin; Alison Laufer Halpin; Ben L. Rambo-Martin; Ben Rambo-Martin; Clinton Paden; Clinton R. Paden; Dakota Howard; Darlene Wagner; Dave Wentworth; Dhwani Batra; Jasmine Padilla; Justin Lee; Katie Dillon; Krista Queen; Kristen Knipe; Kristine Lacek; Mark Burroughs; Matthew Schmerer; Meghan Bentz; Mili Sheth; Peter Cook; Sam Shepard; Sarah Nobles; Shoshona Le; Suxiang Tong; Vivien Dugan; Yvette Unoarumhi |
| EPI_ISL_2919148, EPI_ISL_2919158 | Fakultní nemocnice Hradec Králové | Fakultní nemocnice Hradec Králové | Helena Kovarikova; Marketa Gancarcikova |
| EPI_ISL_3230593 | Fimlab Laboratoriot Oy Tampere | Expert Microbiology, National | Carita Savolainen-Kopra; Erika Lindh; Haider al-Hello; Jani Halkilahti; Kirsi Liitsola; Niina Ikonen; Olli Vapalahti; Pekka Ellonen; Phuoc Truong; Päivi Laurila; Ravi Kant; Sari Hannula; Soile Blomqvist; Teemu Smura |

| | | | |
|---|---|---|---|
| | | Institute for Health and Welfare | |
| EPI_ISL_2567245, EPI_ISL_2567246, EPI_ISL_2567247, EPI_ISL_2836332, EPI_ISL_2836414, EPI_ISL_2836488, EPI_ISL_2836489, EPI_ISL_2836498, EPI_ISL_2836505, EPI_ISL_2836511, EPI_ISL_2836551, EPI_ISL_2836643, EPI_ISL_2836736, EPI_ISL_2836738, EPI_ISL_2836800, EPI_ISL_2836841, EPI_ISL_2836844, EPI_ISL_2920458, EPI_ISL_2920461, EPI_ISL_2920464, EPI_ISL_2920466, EPI_ISL_2920491, EPI_ISL_2920531, EPI_ISL_2920544, EPI_ISL_2920550, EPI_ISL_2920560, EPI_ISL_2920573, EPI_ISL_2920575, EPI_ISL_2920590, EPI_ISL_2920611, EPI_ISL_2920613, EPI_ISL_2920620, EPI_ISL_2920623, EPI_ISL_2920624, EPI_ISL_2920625, EPI_ISL_2920627, EPI_ISL_2920630, EPI_ISL_2920676, EPI_ISL_3100753, EPI_ISL_3100765, EPI_ISL_3100776, EPI_ISL_3100829, EPI_ISL_3100840, EPI_ISL_3100847, EPI_ISL_3100889, EPI_ISL_3100902, EPI_ISL_3100952, EPI_ISL_3100959, EPI_ISL_3100969, EPI_ISL_3100971, EPI_ISL_3903285 | | | |
| see above | Florida Bureau of Public Health Laboratories | Florida Bureau of Public Health Laboratories | Jason Blanton; Jiaqi Li; Namratha Tarigopula; Sarah Schmedes |
| EPI_ISL_2663252 | Fondazione IRCCS Ca' Granda Ospedale Maggiore Policlinico | Fondazione IRCCS Ca' Granda Ospedale Maggiore Policlinico | Ferruccio Ceriotti; Sara Uceda Renteria |
| EPI_ISL_2371530, EPI_ISL_2371531, EPI_ISL_2371548, EPI_ISL_2441851, EPI_ISL_2441852, EPI_ISL_2441875, EPI_ISL_2441877, EPI_ISL_2441886, EPI_ISL_2441904, EPI_ISL_2441907, EPI_ISL_2441935, EPI_ISL_2479997, EPI_ISL_2479998, EPI_ISL_2480036, EPI_ISL_2480099, EPI_ISL_2480108, EPI_ISL_2480143, EPI_ISL_2525669, EPI_ISL_2525727, EPI_ISL_2525729, EPI_ISL_2527565, EPI_ISL_2527735, EPI_ISL_2527753, EPI_ISL_2527816, EPI_ISL_2598704, EPI_ISL_2598705, EPI_ISL_2598726, EPI_ISL_2598733, EPI_ISL_2598791, EPI_ISL_2599922, EPI_ISL_2686408, EPI_ISL_2686431, EPI_ISL_2686449, EPI_ISL_2686452, EPI_ISL_2686486, EPI_ISL_2686496, EPI_ISL_2686554, EPI_ISL_2686555, EPI_ISL_2686561, EPI_ISL_2686573, EPI_ISL_2686609, EPI_ISL_2686623, EPI_ISL_2686651, EPI_ISL_2686717, EPI_ISL_2686720, EPI_ISL_2686732, EPI_ISL_2785548, EPI_ISL_2785551, EPI_ISL_2785592, EPI_ISL_2785601, EPI_ISL_2785636, EPI_ISL_2785644, EPI_ISL_2785648, EPI_ISL_2785680, EPI_ISL_2785681, EPI_ISL_2785737, EPI_ISL_2785858, EPI_ISL_2785861, EPI_ISL_2785872, EPI_ISL_2785912, EPI_ISL_2785940, EPI_ISL_2873401, EPI_ISL_2873586, EPI_ISL_2873631, EPI_ISL_2873692, EPI_ISL_2873762, EPI_ISL_2873765, EPI_ISL_2873796, EPI_ISL_2873808, EPI_ISL_2928558, EPI_ISL_2928640, EPI_ISL_2928678, EPI_ISL_2928681, EPI_ISL_2929136, EPI_ISL_2929138, EPI_ISL_2929152, EPI_ISL_2929153, EPI_ISL_2929158, EPI_ISL_3016511, EPI_ISL_3016654, EPI_ISL_3065030, EPI_ISL_3208204, EPI_ISL_3208320, EPI_ISL_3208332, EPI_ISL_3208819, EPI_ISL_3299864, EPI_ISL_3299867, EPI_ISL_3299871, EPI_ISL_3299967, EPI_ISL_3300021 | | | |
| see above | Fulgent Genetics | Centers for Disease Control and Prevention Division of Viral Diseases, Pathogen Discovery | Adrian Paskey; Becky Tsai; Benafsh Sapra; Benjamin Rambo-Martin; Christopher Gulvick; Clinton R. Paden; Dakota Howard; Darlene Wagner; Dhwani Batra; Doreen Ng; Duncan MacCannell; Harry Gao; James Xie; Jason Caravas; John Gao; Joseph Fierro; Kara Moser; Matthew Schmerer; Mickey Li; Peter W. Cook; Scott Sammons; Shatavia Morrison; Yan Meng; Yvette Unoarumhi |
| EPI_ISL_2644693 | Furst Medical Laboratory | Norwegian Institute of Public Health, Department of Virology | Atiya R Ali; Debech Nadia; Engebretsen Serina Beate; Garcia Llorente Ignacio; Hilde Elshaug; Hilde Vollan; Jon Bråte; Kamilla Heddeland Instefjord; Karoline Bragstad; Kathrine Stene-Johansen; Line Victoria Moen; Marie Paulsen Madsen; Olav Hungnes; Pedersen Benedikte Nevjen; Rasmus Riis Kopperud |
| EPI_ISL_2645769 | GA Department of Public Health | GA Department of Public Health | Aliyah Fields; Cynthia Dixey; Jonathan Edwards; Stacy Reeves; Taylor Smith; Tonia Parrott |
| EPI_ISL_2690029, EPI_ISL_3128097, EPI_ISL_3800440, EPI_ISL_3800483 | GA Department of Public Health Laboratory | Centers for Disease Control and Prevention Division of Viral Diseases, Pathogen Discovery | Alex Burgin; Alison Laufer Halpin; Ben L. Rambo-Martin; Ben Rambo-Martin; Clinton Paden; Clinton R. Paden; Dakota Howard; Darlene Wagner; Dave Wentworth; Dhwani Batra; Jasmine Padilla; Justin Lee; Katie Dillon; Krista Queen; Kristen Knipe; Kristine Lacek; Mark Burroughs; Matthew Schmerer; Meghan Bentz; Mili Sheth; Peter Cook; Sam Shepard; Sarah Nobles; Shoshona Le; Suxiang Tong; Vivien Dugan; Yvette Unoarumhi |
| EPI_ISL_2689598, EPI_ISL_2754920 | GA Department of Public Health Laboratory | Genomics and Discovery, Respiratory Viruses Branch, Division of Viral Diseases, Centers for Disease Control and Prevention | Adam Retchless; Anna Kelleher; Anna Uehara; Brian Lynch; Clinton R. Paden; Dhwani Batra; Haibin Wang; Han Jia Justin Ng; Jasmine Padilla; Jing Zhang; Justin Lee; Krista Queen; Mark Burroughs; Mili Sheth; Morgan Davis; Peter Cook; Rachel Marine; Sarah Nobles; Suxiang Tong; Tara Coalter; Yan Li; Ying Tao |
| EPI_ISL_3031058 | GLENS FALLS HOSPITAL LABORATORY | Wadsworth Center, New York State Department of Health | Alexis Russell; Catharine Prussing; Daryl M. Lamson; Erasmus Schneider; Erica Lasek-Nesselquist; John Kelly; Jonathan Plitnick; Kirsten St. George; Matthew Shudt; Melissa A Leisner; Navjot Singh |
| EPI_ISL_2493943 | GRUPO TECNICO DE VIGILANCIA SANITARIA E EPIDEMIOLOGICA | Instituto Butantan | Antonio Jorge Martins; Claudia Renata dos Santos Barros; David Schlesinger; Debora Botequio Moretti; Dimas Tadeu Covas; Elaine Cristina Marqueze; Elaine Vieira Santos; Evandra Strazza Rodrigues; Heidge Fukumasu; Jayme Augusto de Souza-Neto; José Salvatore Leister Patané; Luiz Alcantara; Luiz Lehmann Coutinho; Maria Carolina Elias; Maurício Lacerda Nogueira; Rafael dos Santos Bezerra; Raul Machado Neto; Rejane Maria Tommasini Grotto; Ricardo Haddad; Sandra Coccuzzo Sampaio Vessoni; Simone Kashima; Svetoslav Nanev Slavov; Vincent Louis Viala |
| EPI_ISL_2543582, EPI_ISL_2543616, EPI_ISL_2543627, EPI_ISL_2543630, EPI_ISL_2543631, EPI_ISL_2543632, EPI_ISL_2543633, EPI_ISL_2543634, EPI_ISL_2543635, EPI_ISL_2543636, EPI_ISL_2543638, EPI_ISL_2543642, EPI_ISL_2543643, EPI_ISL_2543644, EPI_ISL_2543645, EPI_ISL_2543646, EPI_ISL_2543647, EPI_ISL_2543649, EPI_ISL_2543661, EPI_ISL_2543665, EPI_ISL_2557255, EPI_ISL_2557256, EPI_ISL_2557258, EPI_ISL_2557262, EPI_ISL_2557269, EPI_ISL_2557279, EPI_ISL_2557283, EPI_ISL_2557284, EPI_ISL_2557294, EPI_ISL_2557300, EPI_ISL_2659117, EPI_ISL_2659122, EPI_ISL_2659126, EPI_ISL_2659128, EPI_ISL_2659132, EPI_ISL_2659136, EPI_ISL_2659157, EPI_ISL_2659167, EPI_ISL_2659171, EPI_ISL_2659178, EPI_ISL_2659179, EPI_ISL_2659189, EPI_ISL_2659191, EPI_ISL_2659192, EPI_ISL_2659193, EPI_ISL_2663430, EPI_ISL_2663442, EPI_ISL_2663444, EPI_ISL_2663449, EPI_ISL_2663452, EPI_ISL_2663560, EPI_ISL_2663562, EPI_ISL_2663563, EPI_ISL_2663565, EPI_ISL_2663568, EPI_ISL_2663574, EPI_ISL_2663576, EPI_ISL_2663580, EPI_ISL_2663582, EPI_ISL_2663583, EPI_ISL_2663587, EPI_ISL_2663592, EPI_ISL_2663601, EPI_ISL_2663607, EPI_ISL_2663616, EPI_ISL_2756060, EPI_ISL_2756061, EPI_ISL_2756066, EPI_ISL_2756069, EPI_ISL_2756094, EPI_ISL_2756095, EPI_ISL_2756097, EPI_ISL_2756108, EPI_ISL_2756109, EPI_ISL_2756112, EPI_ISL_2756114, EPI_ISL_2756116, EPI_ISL_2756122, EPI_ISL_2756124, EPI_ISL_2756126, EPI_ISL_2756128, EPI_ISL_2756130, EPI_ISL_2756141, EPI_ISL_2756144, EPI_ISL_2756148, EPI_ISL_2756149, EPI_ISL_2756150, EPI_ISL_2756157, EPI_ISL_2756166, EPI_ISL_2756178, EPI_ISL_2756179, EPI_ISL_2756183, EPI_ISL_2756184, EPI_ISL_2756187, EPI_ISL_2756188, EPI_ISL_2756189, EPI_ISL_2756190, EPI_ISL_2756195, EPI_ISL_2756230, EPI_ISL_2756234, EPI_ISL_2756240, EPI_ISL_2756241, EPI_ISL_2756245, EPI_ISL_2756253, EPI_ISL_2756269, EPI_ISL_2756274, EPI_ISL_2756276, EPI_ISL_2756282, EPI_ISL_2756283, EPI_ISL_2756287, EPI_ISL_2756295, EPI_ISL_2756296, EPI_ISL_2756297, EPI_ISL_2756309, EPI_ISL_2756311, EPI_ISL_2756317, EPI_ISL_2756327, EPI_ISL_2756331, EPI_ISL_2756335, EPI_ISL_2756338, EPI_ISL_2892596, EPI_ISL_2892598, EPI_ISL_2892609, EPI_ISL_2892610, EPI_ISL_2892612, EPI_ISL_2892627, EPI_ISL_2892630, EPI_ISL_2892631, EPI_ISL_2892632, EPI_ISL_2892636, EPI_ISL_2892638, EPI_ISL_2892639, EPI_ISL_2892642, EPI_ISL_2892643, EPI_ISL_2892645, EPI_ISL_2892646, EPI_ISL_2892652, EPI_ISL_2892655, EPI_ISL_2892660, EPI_ISL_2892664, EPI_ISL_2892667, EPI_ISL_2892668, EPI_ISL_2892673, EPI_ISL_2892675, EPI_ISL_2892678, EPI_ISL_2892692, EPI_ISL_2892727, EPI_ISL_2892729, EPI_ISL_2892731, EPI_ISL_2892741, EPI_ISL_2892746, EPI_ISL_2892747, EPI_ISL_2892755, EPI_ISL_2892757, EPI_ISL_2892761, EPI_ISL_2892762, EPI_ISL_2892777, EPI_ISL_2892780, EPI_ISL_2892792, EPI_ISL_2892821, EPI_ISL_2892823, EPI_ISL_2892825, EPI_ISL_2892828, EPI_ISL_2892830, EPI_ISL_2892831, EPI_ISL_2892851, EPI_ISL_2892854, EPI_ISL_2892856, EPI_ISL_2892857, EPI_ISL_2892858, EPI_ISL_2892860, EPI_ISL_2892862, EPI_ISL_2892865, EPI_ISL_2892866, EPI_ISL_2892868, EPI_ISL_2892870, EPI_ISL_2892871, EPI_ISL_2892878, EPI_ISL_2892879, EPI_ISL_2892882, EPI_ISL_2892885, EPI_ISL_2892891, EPI_ISL_2892894, EPI_ISL_2892898, EPI_ISL_2892900, EPI_ISL_2892901, EPI_ISL_2892908, EPI_ISL_2892921, EPI_ISL_2892922, EPI_ISL_2892923, EPI_ISL_2892924, EPI_ISL_2892925, EPI_ISL_2892930, EPI_ISL_2892931, EPI_ISL_2892936, EPI_ISL_2892946, EPI_ISL_2892951, EPI_ISL_2892959, EPI_ISL_2892961, EPI_ISL_2892964, EPI_ISL_2892965, EPI_ISL_2892967, EPI_ISL_2892968, EPI_ISL_2892969, EPI_ISL_2892970, EPI_ISL_2892971, EPI_ISL_2892972, EPI_ISL_2892973, EPI_ISL_2892974, EPI_ISL_2892975, EPI_ISL_2892976, EPI_ISL_2892978, EPI_ISL_2892980, EPI_ISL_2892982, EPI_ISL_2892993, EPI_ISL_2892995, EPI_ISL_2893001, EPI_ISL_2893003, EPI_ISL_2893004, EPI_ISL_2893006, EPI_ISL_2893009, EPI_ISL_2893010, EPI_ISL_2893013, EPI_ISL_2893014, EPI_ISL_2893015, EPI_ISL_2893016, EPI_ISL_2893017, EPI_ISL_2893018, EPI_ISL_2894896, EPI_ISL_2894897, EPI_ISL_2894898, EPI_ISL_2894899, EPI_ISL_2894905, EPI_ISL_2894906, EPI_ISL_2894916, EPI_ISL_2894917, EPI_ISL_2894918, EPI_ISL_2894919, EPI_ISL_2894924, EPI_ISL_2894928, EPI_ISL_2894929, EPI_ISL_2894930, EPI_ISL_2894931, EPI_ISL_2894938, EPI_ISL_2894947, EPI_ISL_2894952, EPI_ISL_2894960, EPI_ISL_2894963, EPI_ISL_2894967, EPI_ISL_2894970, EPI_ISL_2896150, EPI_ISL_2896151, EPI_ISL_2896153, EPI_ISL_2896154, EPI_ISL_2896160, EPI_ISL_2896162, EPI_ISL_2896165, EPI_ISL_2896169, EPI_ISL_2896170, EPI_ISL_2896172, EPI_ISL_2896173, EPI_ISL_2896174, EPI_ISL_2896175, EPI_ISL_2896176, EPI_ISL_2896179, EPI_ISL_2896181, EPI_ISL_2930489, EPI_ISL_2930492, EPI_ISL_2930495, EPI_ISL_2930501, EPI_ISL_2930502, EPI_ISL_2930504, EPI_ISL_2930512, EPI_ISL_2930513, EPI_ISL_2930514, EPI_ISL_2930515, EPI_ISL_2930516, EPI_ISL_2930517, EPI_ISL_2930525, EPI_ISL_2930526, EPI_ISL_2930527, EPI_ISL_2930529, EPI_ISL_2930531, EPI_ISL_2930532, EPI_ISL_2930533, EPI_ISL_2930536, EPI_ISL_2930550, EPI_ISL_2930552, EPI_ISL_3132174, EPI_ISL_3132175, EPI_ISL_3132177, EPI_ISL_3132188, EPI_ISL_3132197, EPI_ISL_3132199, EPI_ISL_3132200, EPI_ISL_3132211, EPI_ISL_3132212, EPI_ISL_3132213, EPI_ISL_3132216, EPI_ISL_3132238, EPI_ISL_3132240, EPI_ISL_3132241, EPI_ISL_3132242, EPI_ISL_3132266, EPI_ISL_3132268, EPI_ISL_3132270, EPI_ISL_3132642, EPI_ISL_3132643, EPI_ISL_3132645, EPI_ISL_3132646, EPI_ISL_3132647, EPI_ISL_3132650, EPI_ISL_3132659, EPI_ISL_3132661, EPI_ISL_3132679, EPI_ISL_3132680, EPI_ISL_3132681, EPI_ISL_3132682, EPI_ISL_3132684, EPI_ISL_3132685, EPI_ISL_3132691, EPI_ISL_3132692, EPI_ISL_3132695, EPI_ISL_3132696, EPI_ISL_3144905, EPI_ISL_3144907, EPI_ISL_3144909, EPI_ISL_3144912, EPI_ISL_3144916, EPI_ISL_3144923, EPI_ISL_3144927, EPI_ISL_3144931, EPI_ISL_3144932, EPI_ISL_3149082, EPI_ISL_3149084, EPI_ISL_3149087, EPI_ISL_3149090, EPI_ISL_3149091, EPI_ISL_3149094, EPI_ISL_3149095, EPI_ISL_3149098, EPI_ISL_3149099, EPI_ISL_3149101, EPI_ISL_3149102, EPI_ISL_3149118, EPI_ISL_3149120, EPI_ISL_3149124, EPI_ISL_3149126, EPI_ISL_3185388, EPI_ISL_3185389, EPI_ISL_3185391, EPI_ISL_3185394, EPI_ISL_3185400, EPI_ISL_3185402, EPI_ISL_3185409, EPI_ISL_3185410, EPI_ISL_3185411, EPI_ISL_3185417, EPI_ISL_3185419, EPI_ISL_3185421, EPI_ISL_3185425, EPI_ISL_3185426, EPI_ISL_3185428, EPI_ISL_3185429, EPI_ISL_3185432, EPI_ISL_3185433, EPI_ISL_3185434, EPI_ISL_3185435, EPI_ISL_3185436, EPI_ISL_3185453, EPI_ISL_3185458, EPI_ISL_3185464, EPI_ISL_3185469, EPI_ISL_3185472, EPI_ISL_3185483, EPI_ISL_3185484, EPI_ISL_3185486, EPI_ISL_3185487, EPI_ISL_3185489, EPI_ISL_3185492, EPI_ISL_3185494, EPI_ISL_3185495, EPI_ISL_3185496, EPI_ISL_3185502, EPI_ISL_3185503, EPI_ISL_3185507, EPI_ISL_3185508, EPI_ISL_3185510, EPI_ISL_3185513, EPI_ISL_3185522, EPI_ISL_3185528, EPI_ISL_3185539, EPI_ISL_3185542, EPI_ISL_3185545, EPI_ISL_3185546, EPI_ISL_3185547, EPI_ISL_3185548, EPI_ISL_3185549, EPI_ISL_3185550, EPI_ISL_3185551, EPI_ISL_3185555, EPI_ISL_3185557, EPI_ISL_3185566, EPI_ISL_3185569, EPI_ISL_3185572, EPI_ISL_3185583, EPI_ISL_3185587, EPI_ISL_3185591, EPI_ISL_3185593, EPI_ISL_3185601, EPI_ISL_3185602, EPI_ISL_3185610, EPI_ISL_3185612, EPI_ISL_3185613, EPI_ISL_3185614, EPI_ISL_3185619, EPI_ISL_3185621, EPI_ISL_3185624, EPI_ISL_3185625, EPI_ISL_3185640, EPI_ISL_3185641, EPI_ISL_3185672, EPI_ISL_3185678, EPI_ISL_3185699, EPI_ISL_3185705, EPI_ISL_3185706, EPI_ISL_3185708, EPI_ISL_3185717, EPI_ISL_3185719, EPI_ISL_3185720, EPI_ISL_3185730, EPI_ISL_3185770, EPI_ISL_3220038, EPI_ISL_3220043, EPI_ISL_3220045, EPI_ISL_3220047, EPI_ISL_3220058, EPI_ISL_3220065, EPI_ISL_3220068, EPI_ISL_3220074, EPI_ISL_3220075, EPI_ISL_3220077, EPI_ISL_3220087, EPI_ISL_3220090, EPI_ISL_3220098, EPI_ISL_3220101, EPI_ISL_3220129, EPI_ISL_3220142, EPI_ISL_3220143, EPI_ISL_3358536, EPI_ISL_3358538, EPI_ISL_3358540, EPI_ISL_3358562, EPI_ISL_3358566, EPI_ISL_3358571, EPI_ISL_3358572, EPI_ISL_3368885, EPI_ISL_3368887, EPI_ISL_3368888, EPI_ISL_3368889, EPI_ISL_3368891, EPI_ISL_3368896, EPI_ISL_3368897, EPI_ISL_3369339, EPI_ISL_3369340, EPI_ISL_3369386, EPI_ISL_3981972, EPI_ISL_3981976 | | | |
| see above | Genetica Molecular and Subdepartamento de Virologia ISP Chile | Instituto de Salud Publica de Chile | Andres Castillo; Barbara Parra; Constanza Campano; Gisselle Barra; Javier Tognarelli; Jorge Fernandez; Karen Orostica; Loredana Arata; Patricia Bustos; Rodrigo Fasce; Soledad Ulloa |
| EPI_ISL_2621554 | Genomics, Poplar Healthcare | Genomics, Poplar Healthcare | Truong, M. |
| EPI_ISL_4054613, EPI_ISL_4054615 | Grady Memorial Hospital, Clinical Microbiology Laboratory | Piantadosi Lab, Emory Department of Pathology | Anne Piantadosi; Ludy Registre Carmola; Yun F (Wayne) Wang |
| EPI_ISL_2537141, EPI_ISL_2537146, EPI_ISL_2537202, EPI_ISL_2537203, EPI_ISL_2537207, EPI_ISL_2537217, EPI_ISL_2537219, EPI_ISL_2537233, EPI_ISL_2854941, EPI_ISL_2854951, EPI_ISL_2854955, EPI_ISL_2854961, EPI_ISL_2985641, EPI_ISL_2985642, EPI_ISL_2985664, EPI_ISL_2985665, EPI_ISL_2985668, EPI_ISL_2985714, EPI_ISL_2985761 | | | |
| see above | Gravity Diagnostics, LLC | Gravity Diagnostics, LLC | Gravity Diagnostics |
| EPI_ISL_2376254, EPI_ISL_2376255, EPI_ISL_2501001 | Gundersen Clinical Microbiology Laboratory | Kabara Cancer Research Institute | Craig S. Richmond; Paraic A. Kenny |
| EPI_ISL_2695275 | H Braga | Instituto Nacional de Saude (INSA) | Borges et al |
| EPI_ISL_3102422, EPI_ISL_3536150, EPI_ISL_3536363 | H J M A HOSPITAL JOSE MARTINIANO DE ALENCAR | Analytical Competence Molecular Epidemiology Lab/ACME, Oswaldo Cruz Foundation, Ceara (FIOCRUZ CE) | Cleber Furtado Aksenen; Fabio Miyajima; Fernando Braga Stehling; Francisco Eder de Moura Lopes; Jamille Maria Mendes Bezerra; Joaquim César do Nascimento Sousa Junior; Pedro Miguel Carneiro Jeronimo; Suzana Porto Almeida e Lucas Delerino; Thais Ferreira de Oliveira; Thais de Oliveira Costa; Ticiane Cavalcante de Souza; Veridiana Pessoa Miyajima |
| EPI_ISL_3102348, EPI_ISL_3102356, EPI_ISL_3102359, EPI_ISL_3102427, EPI_ISL_3536210, EPI_ISL_3536213 | HEMOCE CENTRO DE HEMATOLOGIA E HEMOTERAPIA DO CEARA | Analytical Competence Molecular Epidemiology Lab/ACME, Oswaldo Cruz Foundation, Ceara (FIOCRUZ CE) | Cleber Furtado Aksenen; Fabio Miyajima; Fernando Braga Stehling; Francisco Eder de Moura Lopes; Jamille Maria Mendes Bezerra; Joaquim César do Nascimento Sousa Junior; Pedro Miguel Carneiro Jeronimo; Suzana Porto Almeida e Lucas Delerino; Thais Ferreira de Oliveira; Thais de Oliveira Costa; Ticiane Cavalcante de Souza; Veridiana Pessoa Miyajima |
| EPI_ISL_3912162 | HEMOCE DE SOBRAL | Analytical Competence Molecular Epidemiology Lab/ACME, Oswaldo Cruz Foundation, Ceara (FIOCRUZ CE) | Cleber Furtado Aksenen; Fabio Miyajima; Fernando Braga Stehling; Francisco Eder de Moura Lopes; Jamille Maria Mendes Bezerra; Joaquim Cesar do Nascimento Sousa Junior; Pedro Miguel Carneiro Jeronimo; Suzana Porto Almeida & Lucas Delerino on behalf of COVID-19 FIOCRUZ Genomic Network; Thais Ferreira de Oliveira; Thais de Oliveira Costa; Ticiane Cavalcante de Souza; Veridiana Pessoa Miyajima |
| EPI_ISL_3550432, EPI_ISL_3550434, EPI_ISL_3550440 | HG LAB TRUCK GmbH | Institute of Virology Department of Hygiene, Microbiology and Public Health at Innsbruck Medical University | Andre Volland; Annika Roessler; Barbara Falkensammer; David Bante; Dorothee von Laer; Heribert Stoiber; Janine Kimpel; Lisa Pipperger; Lydia Riepler; Wegene Borena |
| EPI_ISL_3102339, EPI_ISL_3102345, EPI_ISL_3102352, EPI_ISL_3102381, EPI_ISL_3102392, EPI_ISL_3102397, EPI_ISL_3536166, EPI_ISL_3536214 | | | |
| see above | HGCC HOSPITAL GERAL DR CESAR CALS | Analytical Competence Molecular Epidemiology Lab/ACME, Oswaldo Cruz Foundation, Ceara (FIOCRUZ CE) | Cleber Furtado Aksenen; Fabio Miyajima; Fernando Braga Stehling; Francisco Eder de Moura Lopes; Jamille Maria Mendes Bezerra; Joaquim César do Nascimento Sousa Junior; Pedro Miguel Carneiro Jeronimo; Suzana Porto Almeida e Lucas Delerino; Thais Ferreira de Oliveira; Thais de Oliveira Costa; Ticiane Cavalcante de Souza; Veridiana Pessoa Miyajima |
| EPI_ISL_3102337, EPI_ISL_3102340, EPI_ISL_3102347, EPI_ISL_3102357, EPI_ISL_3102361, EPI_ISL_3102363, EPI_ISL_3102378, EPI_ISL_3102398, EPI_ISL_3102401, EPI_ISL_3536206, EPI_ISL_3536216, EPI_ISL_3536218, EPI_ISL_3536219, EPI_ISL_3536220, EPI_ISL_3536221 | | | |
| see above | HIAS HOSPITAL INFANTIL ALBERT SABIN | Analytical Competence Molecular Epidemiology Lab/ACME, Oswaldo Cruz Foundation, Ceara (FIOCRUZ CE) | Cleber Furtado Aksenen; Fabio Miyajima; Fernando Braga Stehling; Francisco Eder de Moura Lopes; Jamille Maria Mendes Bezerra; Joaquim César do Nascimento Sousa Junior; Pedro Miguel Carneiro Jeronimo; Suzana Porto Almeida e Lucas Delerino; Thais Ferreira de Oliveira; Thais de Oliveira Costa; Ticiane Cavalcante de Souza; Veridiana Pessoa Miyajima |
| EPI_ISL_2497443, EPI_ISL_2497444, EPI_ISL_2497445, EPI_ISL_2497446, EPI_ISL_2497447, EPI_ISL_2497449, EPI_ISL_2497450, EPI_ISL_2497451, EPI_ISL_2497452, EPI_ISL_2497453, EPI_ISL_2497455, EPI_ISL_2497457, EPI_ISL_2497458, EPI_ISL_2497461, EPI_ISL_2497462, EPI_ISL_2497463, EPI_ISL_2497464, EPI_ISL_2497467, EPI_ISL_2497469, EPI_ISL_2497470, EPI_ISL_2617589, EPI_ISL_2617592, EPI_ISL_2617593, EPI_ISL_2617594, EPI_ISL_2617595, EPI_ISL_2617609, EPI_ISL_2617610, EPI_ISL_2617612, EPI_ISL_2617627, EPI_ISL_2680895, EPI_ISL_2680896, EPI_ISL_2680897, EPI_ISL_2680898, EPI_ISL_2680899, EPI_ISL_2680900, EPI_ISL_2680901, EPI_ISL_2680902, EPI_ISL_2680903, EPI_ISL_2680904, EPI_ISL_2680905, EPI_ISL_2680906, EPI_ISL_2680907, EPI_ISL_2680909, EPI_ISL_2680910, EPI_ISL_2680912, EPI_ISL_2680914, EPI_ISL_2680915, EPI_ISL_2921535, EPI_ISL_2921539, EPI_ISL_2921540, EPI_ISL_2921541, EPI_ISL_2921542, EPI_ISL_2921543, EPI_ISL_2921544, EPI_ISL_2921550, EPI_ISL_2921552, EPI_ISL_2921555, EPI_ISL_2921556, EPI_ISL_2921557, EPI_ISL_2921559, EPI_ISL_2921560, EPI_ISL_2921564, EPI_ISL_2921565, EPI_ISL_2921566, EPI_ISL_2921567, EPI_ISL_2921568, EPI_ISL_2921569, EPI_ISL_2921571, EPI_ISL_2921574, EPI_ISL_2921576, EPI_ISL_2921577, EPI_ISL_2921578, EPI_ISL_2921580, EPI_ISL_2921583, EPI_ISL_2921586, EPI_ISL_2921587, EPI_ISL_2921588, EPI_ISL_2921590, EPI_ISL_2921591, EPI_ISL_2921592, EPI_ISL_2921595, EPI_ISL_2921599, EPI_ISL_2921602, EPI_ISL_2921604, EPI_ISL_2921606, EPI_ISL_2921609, EPI_ISL_2928511, EPI_ISL_3254319, EPI_ISL_3254320, EPI_ISL_3254322, EPI_ISL_3254323, EPI_ISL_3254325, EPI_ISL_3254326, EPI_ISL_3254327, EPI_ISL_3254328, EPI_ISL_3254330, EPI_ISL_3254332, EPI_ISL_3254333, EPI_ISL_3254349, EPI_ISL_3254350, EPI_ISL_3254352, EPI_ISL_3254353, EPI_ISL_3254354, EPI_ISL_3254356, EPI_ISL_3254357, EPI_ISL_3254358, EPI_ISL_3254359, EPI_ISL_3254360, EPI_ISL_3254361, | | | |

| | | | |
|---|---|---|---|
| EPI_ISL_3254362, EPI_ISL_3254363, EPI_ISL_3254364, EPI_ISL_3254365, EPI_ISL_3254366, EPI_ISL_3254368, EPI_ISL_3254369, EPI_ISL_3254370, EPI_ISL_3254373, EPI_ISL_3254374, EPI_ISL_3254380, EPI_ISL_3254381, EPI_ISL_3254383, EPI_ISL_3254384, EPI_ISL_3254386, EPI_ISL_3254388, EPI_ISL_3254390, EPI_ISL_3254391, EPI_ISL_3254392, EPI_ISL_3254393, EPI_ISL_3254395, EPI_ISL_3254396, EPI_ISL_3254397, EPI_ISL_3254398, EPI_ISL_3254400, EPI_ISL_3254402, EPI_ISL_3254404, EPI_ISL_3254405, EPI_ISL_3254406, EPI_ISL_3254407, EPI_ISL_3254409, EPI_ISL_3254411, EPI_ISL_3254412, EPI_ISL_3254413, EPI_ISL_3254414, EPI_ISL_3254416, EPI_ISL_3254418, EPI_ISL_3254419, EPI_ISL_3254422, EPI_ISL_3254423, EPI_ISL_3254424, EPI_ISL_3254425, EPI_ISL_3254426, EPI_ISL_3254427, EPI_ISL_3254428, EPI_ISL_3254429, EPI_ISL_3254431, EPI_ISL_3254433, EPI_ISL_3254434, EPI_ISL_3254435, EPI_ISL_3254438, EPI_ISL_3254439, EPI_ISL_3254440, EPI_ISL_3254442, EPI_ISL_3254443, EPI_ISL_3254445, EPI_ISL_3254447, EPI_ISL_3254448, EPI_ISL_3254449, EPI_ISL_3254450, EPI_ISL_3254451, EPI_ISL_3254452, EPI_ISL_3254453, EPI_ISL_3254454, EPI_ISL_3254455, EPI_ISL_3254458, EPI_ISL_3254459, EPI_ISL_3254460, EPI_ISL_3254461, EPI_ISL_3254462, EPI_ISL_3254463, EPI_ISL_3254464, EPI_ISL_3254466, EPI_ISL_3254467, EPI_ISL_3254468, EPI_ISL_3254469, EPI_ISL_3254470, EPI_ISL_3254471, EPI_ISL_3254472, EPI_ISL_3254473, EPI_ISL_3254475, EPI_ISL_3254476, EPI_ISL_3254477, EPI_ISL_3254479, EPI_ISL_3254480, EPI_ISL_3254481, EPI_ISL_3254482, EPI_ISL_3254485, EPI_ISL_3254486, EPI_ISL_3254487, EPI_ISL_3254489, EPI_ISL_3254491, EPI_ISL_3254492, EPI_ISL_3254493, EPI_ISL_3254494, EPI_ISL_3254495, EPI_ISL_3254496, EPI_ISL_3254498, EPI_ISL_3254499, EPI_ISL_3254501, EPI_ISL_3254502, EPI_ISL_3254505, EPI_ISL_3254506, EPI_ISL_3254507, EPI_ISL_3254508, EPI_ISL_3254509, EPI_ISL_3254510, EPI_ISL_3254511, EPI_ISL_3254512, EPI_ISL_3254513, EPI_ISL_3254514, EPI_ISL_3254515, EPI_ISL_3254516, EPI_ISL_3254517, EPI_ISL_3254518, EPI_ISL_3254519, EPI_ISL_3254520, EPI_ISL_3254521, EPI_ISL_3254522, EPI_ISL_3254523, EPI_ISL_3254526, EPI_ISL_3254528, EPI_ISL_3254530, EPI_ISL_3254531, EPI_ISL_3254533, EPI_ISL_3254535, EPI_ISL_3254536, EPI_ISL_3254538, EPI_ISL_3254539, EPI_ISL_3254541, EPI_ISL_3254542, EPI_ISL_3254543, EPI_ISL_3254544, EPI_ISL_3254546, EPI_ISL_3254547, EPI_ISL_3254548, EPI_ISL_3254549, EPI_ISL_3254551, EPI_ISL_3254553, EPI_ISL_3254554, EPI_ISL_3254556, EPI_ISL_3254558, EPI_ISL_3254560, EPI_ISL_3254562, EPI_ISL_3254563, EPI_ISL_3254564, EPI_ISL_3254565, EPI_ISL_3254568, EPI_ISL_3254569, EPI_ISL_3254571, EPI_ISL_3254572, EPI_ISL_3254573, EPI_ISL_3254574, EPI_ISL_3254575, EPI_ISL_3254576, EPI_ISL_3254578, EPI_ISL_3254579, EPI_ISL_3254580, EPI_ISL_3254581, EPI_ISL_3254582, EPI_ISL_3254583, EPI_ISL_3254584, EPI_ISL_3254585, EPI_ISL_3254586, EPI_ISL_3254587, EPI_ISL_3254589, EPI_ISL_3254590, EPI_ISL_3254594, EPI_ISL_3254595, EPI_ISL_3254596, EPI_ISL_3254597, EPI_ISL_3254598, EPI_ISL_3254601, EPI_ISL_3254602, EPI_ISL_3254604, EPI_ISL_3254605, EPI_ISL_3254606, EPI_ISL_3254610, EPI_ISL_3254611, EPI_ISL_3254612, EPI_ISL_3254613, EPI_ISL_3254614, EPI_ISL_3254615, EPI_ISL_3254618, EPI_ISL_3254619, EPI_ISL_3254620, EPI_ISL_3254621, EPI_ISL_3254623, EPI_ISL_3254624, EPI_ISL_3254625, EPI_ISL_3254626, EPI_ISL_3254627, EPI_ISL_3254628, EPI_ISL_3254629, EPI_ISL_3254630, EPI_ISL_3254633, EPI_ISL_3254634, EPI_ISL_3254635, EPI_ISL_3254636, EPI_ISL_3254637, EPI_ISL_3254640, EPI_ISL_3254641, EPI_ISL_3254642, EPI_ISL_3254643, EPI_ISL_3254645, EPI_ISL_3254647, EPI_ISL_3254651, EPI_ISL_3254652, EPI_ISL_3254653, EPI_ISL_3254657, EPI_ISL_3254659, EPI_ISL_3254660, EPI_ISL_3254664, EPI_ISL_3254665, EPI_ISL_3254672, EPI_ISL_3254673, EPI_ISL_3254676, EPI_ISL_3254677, EPI_ISL_3254684, EPI_ISL_3259354, EPI_ISL_3299426, EPI_ISL_3299428, EPI_ISL_3299429, EPI_ISL_3386151, EPI_ISL_3386153, EPI_ISL_3386154, EPI_ISL_4061630, EPI_ISL_4061647, EPI_ISL_4061648, EPI_ISL_4061651, EPI_ISL_4061652, EPI_ISL_4061655, EPI_ISL_4061657, EPI_ISL_4061659, EPI_ISL_4061661 | | | |
| see above | HLAGYN - Laboratorio de Imunologia de Transplantes de Goias | HLAGYN - Laboratorio de Imunologia de Transplantes de Goias | Alessandro Leonardo Alvares Magalhaes; Daniel Ferreira de Sousa; Danielle de Paiva Rezende; Erika Lopes Rocha Batista; Fernando Antonio Vinhal dos Santos; Frederico Rodrigues Vinhal; Kamila Oliveira Reis De Freitas.; Lucas Carlos Gomes Pereira; Sabrina Sara Moreira Duarte |
| EPI_ISL_3102408, EPI_ISL_3102419, EPI_ISL_3102426, EPI_ISL_3102430, EPI_ISL_3102431, EPI_ISL_3102432, EPI_ISL_3102434, EPI_ISL_3102436, EPI_ISL_3102440, EPI_ISL_3102444, EPI_ISL_3102448, EPI_ISL_3102453, EPI_ISL_3102457, EPI_ISL_3102461, EPI_ISL_3102462, EPI_ISL_3102464, EPI_ISL_3102469, EPI_ISL_3536205, EPI_ISL_3536222, EPI_ISL_3536223, EPI_ISL_3536224, EPI_ISL_3536225, EPI_ISL_3536227, EPI_ISL_3536228, EPI_ISL_3536230, EPI_ISL_3536232, EPI_ISL_3536233, EPI_ISL_3536236, EPI_ISL_3536237, EPI_ISL_3536238, EPI_ISL_3536239, EPI_ISL_3536240, EPI_ISL_3536241, EPI_ISL_3536242, EPI_ISL_3536243, EPI_ISL_3536245, EPI_ISL_3536247, EPI_ISL_3536248, EPI_ISL_3536249, EPI_ISL_3536251, EPI_ISL_3536252, EPI_ISL_3536254, EPI_ISL_3536257, EPI_ISL_3536258, EPI_ISL_3536259, EPI_ISL_3912173 | | | |
| see above | HM HOSPITAL DE MESSEJANA DR CARLOS ALBERTO STUDART GOMES | Analytical Competence Molecular Epidemiology Lab/ACME, Oswaldo Cruz Foundation, Ceara (FIOCRUZ CE) | Cleber Furtado Aksenen; Fabio Miyajima; Fernando Braga Stehling; Francisco Eder de Moura Lopes; Jamille Maria Mendes Bezerra; Joaquim Cesar do Nascimento Sousa Junior; Joaquim César do Nascimento Sousa Junior; Pedro Miguel Carneiro Jeronimo; Suzana Porto Almeida & Lucas Delerino on behalf of COVID-19 FIOCRUZ Genomic Network; Suzana Porto Almeida e Lucas Delerino; Thais Ferreira de Oliveira; Thais de Oliveira Costa; Ticiane Cavalcante de Souza; Veridiana Pessoa Miyajima |
| EPI_ISL_3536177, EPI_ISL_3912188 | HOSP DR THADEU DE PAULO BRITO | Analytical Competence Molecular Epidemiology Lab/ACME, Oswaldo Cruz Foundation, Ceara (FIOCRUZ CE) | Cleber Furtado Aksenen; Fabio Miyajima; Fernando Braga Stehling; Francisco Eder de Moura Lopes; Jamille Maria Mendes Bezerra; Joaquim Cesar do Nascimento Sousa Junior; Joaquim César do Nascimento Sousa Junior; Pedro Miguel Carneiro Jeronimo; Suzana Porto Almeida & Lucas Delerino on behalf of COVID-19 FIOCRUZ Genomic Network; Suzana Porto Almeida e Lucas Delerino; Thais Ferreira de Oliveira; Thais de Oliveira Costa; Ticiane Cavalcante de Souza; Veridiana Pessoa Miyajima |
| EPI_ISL_3536181, EPI_ISL_3536182, EPI_ISL_3536183, EPI_ISL_3912191 | HOSP MUN ABELARDO GADELHA DA ROCHA | Analytical Competence Molecular Epidemiology Lab/ACME, Oswaldo Cruz Foundation, Ceara (FIOCRUZ CE) | Cleber Furtado Aksenen; Fabio Miyajima; Fernando Braga Stehling; Francisco Eder de Moura Lopes; Jamille Maria Mendes Bezerra; Joaquim Cesar do Nascimento Sousa Junior; Joaquim César do Nascimento Sousa Junior; Pedro Miguel Carneiro Jeronimo; Suzana Porto Almeida & Lucas Delerino on behalf of COVID-19 FIOCRUZ Genomic Network; Suzana Porto Almeida e Lucas Delerino; Thais Ferreira de Oliveira; Thais de Oliveira Costa; Ticiane Cavalcante de Souza; Veridiana Pessoa Miyajima |
| EPI_ISL_2493850, EPI_ISL_2493851, EPI_ISL_2493853, EPI_ISL_2493854, EPI_ISL_2493855, EPI_ISL_2493856, EPI_ISL_2493857, EPI_ISL_2493859, EPI_ISL_2493860, EPI_ISL_2493905, EPI_ISL_2493918, EPI_ISL_2493920, EPI_ISL_2493921, EPI_ISL_2493923, EPI_ISL_2493924, EPI_ISL_2493925, EPI_ISL_2493926, EPI_ISL_2493927, EPI_ISL_2493929, EPI_ISL_2493937, EPI_ISL_2493938, EPI_ISL_2493954 | | | |
| see above | HOSP MUN DE MOGI DAS CRUZES PREF WALDEMAR COSTA FILHO | Instituto Butantan | Antonio Jorge Martins; Claudia Renata dos Santos Barros; David Schlesinger; Debora Botequio Moretti; Dimas Tadeu Covas; Elaine Cristina Marqueze; Elaine Vieira Santos; Evandra Strazza Rodrigues; Heidge Fukumasu; Jayme Augusto de Souza-Neto; José Salvatore Leister Patané; Luiz Alcantara; Luiz Lehmann Coutinho; Maria Carolina Elias; Maurício Lacerda Nogueira; Rafael dos Santos Bezerra; Raul Machado Neto; Rejane Maria Tommasini Grotto; Ricardo Haddad; Sandra Coccuzzo Sampaio Vessoni; Simone Kashima; Svetoslav Nanev Slavov; Vincent Louis Viala |
| EPI_ISL_2493941 | HOSP MUN JOSANIAS CASTANHA BRAGA | Instituto Butantan | Antonio Jorge Martins; Claudia Renata dos Santos Barros; David Schlesinger; Debora Botequio Moretti; Dimas Tadeu Covas; Elaine Cristina Marqueze; Elaine Vieira Santos; Evandra Strazza Rodrigues; Heidge Fukumasu; Jayme Augusto de Souza-Neto; José Salvatore Leister Patané; Luiz Alcantara; Luiz Lehmann Coutinho; Maria Carolina Elias; Maurício Lacerda Nogueira; Rafael dos Santos Bezerra; Raul Machado Neto; Rejane Maria Tommasini Grotto; Ricardo Haddad; Sandra Coccuzzo Sampaio Vessoni; Simone Kashima; Svetoslav Nanev Slavov; Vincent Louis Viala |
| EPI_ISL_2812743 | HOSPITAL CAN MISSES | HOSPITAL UNIVERSITARIO SON ESPASES | Hospital Universitario Son Espases; Servicio de Microbiología |
| EPI_ISL_3102285 | HOSPITAL CURA DARS | Analytical Competence Molecular Epidemiology Lab/ACME, Oswaldo Cruz Foundation, Ceara (FIOCRUZ CE) | Cleber Furtado Aksenen; Fabio Miyajima; Fernando Braga Stehling; Francisco Eder de Moura Lopes; Jamille Maria Mendes Bezerra; Joaquim César do Nascimento Sousa Junior; Pedro Miguel Carneiro Jeronimo; Suzana Porto Almeida e Lucas Delerino; Thais Ferreira de Oliveira; Thais de Oliveira Costa; Ticiane Cavalcante de Souza; Veridiana Pessoa Miyajima |
| EPI_ISL_2493956, EPI_ISL_2493959 | HOSPITAL DE CAMPANHA COVID 19 DE MAIRIPORA | Instituto Butantan | Antonio Jorge Martins; Claudia Renata dos Santos Barros; David Schlesinger; Debora Botequio Moretti; Dimas Tadeu Covas; Elaine Cristina Marqueze; Elaine Vieira Santos; Evandra Strazza Rodrigues; Heidge Fukumasu; Jayme Augusto de Souza-Neto; José Salvatore Leister Patané; Luiz Alcantara; Luiz Lehmann Coutinho; Maria Carolina Elias; Maurício Lacerda Nogueira; Rafael dos Santos Bezerra; Raul Machado Neto; Rejane Maria Tommasini Grotto; Ricardo Haddad; Sandra Coccuzzo Sampaio Vessoni; Simone Kashima; Svetoslav Nanev Slavov; Vincent Louis Viala |
| EPI_ISL_2493873, EPI_ISL_2493876, EPI_ISL_2493877 | HOSPITAL DE CAMPANHA COVID 19 MUNICIPIO DE TAUBATE | Instituto Butantan | Antonio Jorge Martins; Claudia Renata dos Santos Barros; David Schlesinger; Debora Botequio Moretti; Dimas Tadeu Covas; Elaine Cristina Marqueze; Elaine Vieira Santos; Evandra Strazza Rodrigues; Heidge Fukumasu; Jayme Augusto de Souza-Neto; José Salvatore Leister Patané; Luiz Alcantara; Luiz Lehmann Coutinho; Maria Carolina Elias; Maurício Lacerda Nogueira; Rafael dos Santos Bezerra; Raul Machado Neto; Rejane Maria Tommasini Grotto; Ricardo Haddad; Sandra Coccuzzo Sampaio Vessoni; Simone Kashima; Svetoslav Nanev Slavov; Vincent Louis Viala |
| EPI_ISL_3037826 | HOSPITAL DE NIÑOS DR. CARLOS SAENZ HERRERA | Inciensa, Instituto Costarricense de Investigación y Enseñanza en Nutrición y Salud | Adriana Godínez; Caterina Guzmán; Claudio Soto-Garita; Estela Cordero; Francisco Duarte; Hebleen Porras; José Luis Vargas; Mariela Gutiérrez Joselyn Prado; Melany Calderón; Nazareth Ruiz & Cristian Pérez |
| EPI_ISL_3459388 | HOSPITAL DEPARTAMENTAL DE VILLAVICENCIO | Instituto Nacional de Salud | Carlos Franco-Muñoz; Carmen Osorio; Diana Malo; Diego A. Álvarez-Díaz; Diego Andrés Prada; Gerardo Santamaría; Hector Alejandro Ruiz-Moreno; Jhonnatan Reales-González; Jorge Rivera; Juan Camilo Martinez; Julian Naizaque; Katherine Laiton-Donato; Lisseth Pardo; Magdalena Wiesner; Marcela Mercado-Reyes; Maria T. Herrera-Sepúlveda; Marta Lopez Blanco; Martha Lucia Ospina Martinez; Paola Rojas; Sergio Gomez; Sheryll Corchuelo; Ángela Alarcon Cruz |
| EPI_ISL_2827976, EPI_ISL_2827977 | HOSPITAL DR. ENRIQUE BALTODANO BRICEÑO | Inciensa, Instituto Costarricense de Investigación y Enseñanza en Nutrición y Salud | Adriana Godínez; Claudio Soto-Garita; Estela Cordero; Francisco Duarte; Hebleen Porras; Joselyn Prado & Adriana Bermúdez; José Luis Vargas; Mariela Gutiérrez; Melany Calderón |
| EPI_ISL_3464519 | HOSPITAL DR. FERNANDO ESCALANTE PRADILLA | Inciensa, Instituto Costarricense de Investigación y Enseñanza en Nutrición y Salud | Adriana Godínez; Claudio Soto-Garita; Estela Cordero; Francisco Duarte; Hebleen Porras; Joselyn Prado & Yendri Ramírez Alpízar; José Luis Vargas; Mariela Gutiérrez; Melany Calderón |
| EPI_ISL_2827967, EPI_ISL_2828010, EPI_ISL_3037815 | HOSPITAL DR. RAFAEL A. CALDERON GUARDIA | Inciensa, Instituto Costarricense de Investigación y Enseñanza en Nutrición y Salud | Adriana Godínez; Caterina Guzmán; Claudio Soto-Garita; Estela Cordero; Francisco Duarte; Hebleen Porras; Joselyn Prado & Fabian Salas; José Luis Vargas; Mariela Gutiérrez; Mariela Gutiérrez Joselyn Prado; Melany Calderón; Nazareth Ruiz & Fabián Salas |
| EPI_ISL_2827963, EPI_ISL_2827964, EPI_ISL_2827965 | HOSPITAL DR. WILLIAM ALLEN | Inciensa, Instituto Costarricense de Investigación y Enseñanza en Nutrición y Salud | Adriana Godínez; Claudio Soto-Garita; Estela Cordero; Francisco Duarte; Hebleen Porras; José Luis Vargas; Mariela Gutiérrez & Joselyn Prado; Melany Calderón |
| EPI_ISL_3102412, EPI_ISL_3102472, EPI_ISL_3536263, EPI_ISL_3536264, EPI_ISL_3536265 | HOSPITAL E MATERNIDADE DRA ZILDA ARNS NEUMANN | Analytical Competence Molecular Epidemiology Lab/ACME, Oswaldo Cruz Foundation, Ceara (FIOCRUZ CE) | Cleber Furtado Aksenen; Fabio Miyajima; Fernando Braga Stehling; Francisco Eder de Moura Lopes; Jamille Maria Mendes Bezerra; Joaquim César do Nascimento Sousa Junior; Pedro Miguel Carneiro Jeronimo; Suzana Porto Almeida e Lucas Delerino; Thais Ferreira de Oliveira; Thais de Oliveira Costa; Ticiane Cavalcante de Souza; Veridiana Pessoa Miyajima |
| EPI_ISL_2493257, EPI_ISL_2493258, EPI_ISL_2493863, EPI_ISL_2493865, EPI_ISL_2493870, EPI_ISL_2493871, EPI_ISL_2500986 | | | |
| see above | HOSPITAL E MATERNIDADE NOSSA SENHORA DA AJUDA | Instituto Butantan | Antonio Jorge Martins; Claudia Renata dos Santos Barros; David Schlesinger; Debora Botequio Moretti; Dimas Tadeu Covas; Elaine Cristina Marqueze; Elaine Vieira Santos; Evandra Strazza Rodrigues; Heidge Fukumasu; Jayme Augusto de Souza-Neto; José Salvatore Leister Patané; Luiz Alcantara; Luiz Lehmann Coutinho; Maria Carolina Elias; Maurício Lacerda Nogueira; Rafael dos Santos Bezerra; Raul Machado Neto; Rejane Maria Tommasini Grotto; Ricardo Haddad; Sandra Coccuzzo Sampaio Vessoni; Simone Kashima; Svetoslav Nanev Slavov; Vincent Louis Viala |
| EPI_ISL_3102275, EPI_ISL_3102334, EPI_ISL_3102336, EPI_ISL_3102341, EPI_ISL_3102349, EPI_ISL_3102351, EPI_ISL_3102354, EPI_ISL_3102360, EPI_ISL_3102364, EPI_ISL_3102372, EPI_ISL_3102374, EPI_ISL_3102375, EPI_ISL_3102376, EPI_ISL_3102383, EPI_ISL_3102384, EPI_ISL_3102386, EPI_ISL_3102387, EPI_ISL_3102393, EPI_ISL_3102394, EPI_ISL_3102395, EPI_ISL_3102396, EPI_ISL_3102399, EPI_ISL_3102403, EPI_ISL_3102405, EPI_ISL_3102506, EPI_ISL_3536149, EPI_ISL_3536154, EPI_ISL_3536155, EPI_ISL_3536169, EPI_ISL_3536170, EPI_ISL_3536174, EPI_ISL_3536175, EPI_ISL_3536176, EPI_ISL_3536184, EPI_ISL_3536186, EPI_ISL_3536187, EPI_ISL_3536188, EPI_ISL_3536193, EPI_ISL_3536266, EPI_ISL_3536267, EPI_ISL_3536270, EPI_ISL_3536271, EPI_ISL_3536272, EPI_ISL_3536273, EPI_ISL_3536274, EPI_ISL_3536275, EPI_ISL_3536277, EPI_ISL_3536278, EPI_ISL_3536279, EPI_ISL_3536280, EPI_ISL_3536281, EPI_ISL_3536282, EPI_ISL_3536283, EPI_ISL_3536284, EPI_ISL_3536285, EPI_ISL_3536286, EPI_ISL_3536287, EPI_ISL_3536288, EPI_ISL_3536312, EPI_ISL_3536313, EPI_ISL_3536323, EPI_ISL_3536324, EPI_ISL_3536332, EPI_ISL_3536334, EPI_ISL_3536341, EPI_ISL_3536342, EPI_ISL_3536347, EPI_ISL_3536357, EPI_ISL_3536359, EPI_ISL_3536364, EPI_ISL_3536372, EPI_ISL_3536376, EPI_ISL_3536377, EPI_ISL_3536393, EPI_ISL_3536395, EPI_ISL_3536398, EPI_ISL_3912206 | | | |
| see above | HOSPITAL ESTADUAL LEONARDO DA VINCI | Analytical Competence Molecular Epidemiology Lab/ACME, Oswaldo Cruz Foundation, Ceara (FIOCRUZ CE) | Cleber Furtado Aksenen; Fabio Miyajima; Fernando Braga Stehling; Francisco Eder de Moura Lopes; Jamille Maria Mendes Bezerra; Joaquim Cesar do Nascimento Sousa Junior; Joaquim César do Nascimento Sousa Junior; Pedro Miguel Carneiro Jeronimo; Suzana Porto Almeida & Lucas Delerino on behalf of COVID-19 FIOCRUZ Genomic Network; Suzana Porto Almeida e Lucas Delerino; Thais Ferreira de Oliveira; Thais de Oliveira Costa; Ticiane Cavalcante de Souza; Veridiana Pessoa Miyajima |
| EPI_ISL_2493861, EPI_ISL_2493957 | HOSPITAL HELIOPOLIS UNIDADE DE GESTAO ASSISTENCIAL I SP | Instituto Butantan | Antonio Jorge Martins; Claudia Renata dos Santos Barros; David Schlesinger; Debora Botequio Moretti; Dimas Tadeu Covas; Elaine Cristina Marqueze; Elaine Vieira Santos; Evandra Strazza Rodrigues; Heidge Fukumasu; Jayme Augusto de Souza-Neto; José Salvatore Leister Patané; Luiz Alcantara; Luiz Lehmann Coutinho; Maria Carolina Elias; Maurício Lacerda Nogueira; Rafael dos Santos Bezerra; Raul Machado Neto; Rejane Maria Tommasini Grotto; Ricardo Haddad; Sandra Coccuzzo Sampaio Vessoni; Simone Kashima; Svetoslav Nanev Slavov; Vincent Louis Viala |
| EPI_ISL_2626156 | HOSPITAL MANACOR | HOSPITAL UNIVERSITARIO SON ESPASES | Hospital Universitario Son Espases; Servicio de Microbiología |
| EPI_ISL_3102330 | HOSPITAL MATERNIDADE AGENOR ARAUJO | Analytical Competence Molecular Epidemiology Lab/ACME, Oswaldo Cruz Foundation, Ceara (FIOCRUZ CE) | Cleber Furtado Aksenen; Fabio Miyajima; Fernando Braga Stehling; Francisco Eder de Moura Lopes; Jamille Maria Mendes Bezerra; Joaquim César do Nascimento Sousa Junior; Pedro Miguel Carneiro Jeronimo; Suzana Porto Almeida e Lucas Delerino; Thais Ferreira de Oliveira; Thais de Oliveira Costa; Ticiane Cavalcante de Souza; Veridiana Pessoa Miyajima |
| EPI_ISL_2827991, EPI_ISL_2827992, EPI_ISL_3298328 | HOSPITAL MEXICO | Inciensa, Instituto Costarricense de Investigación y Enseñanza en Nutrición y Salud | Adriana Godínez; Claudio Soto-Garita; Estela Cordero; Francisco Duarte; Hebleen Porras; Joselyn Prado & Juan Carlos Villalobos; José Luis Vargas; Mariela Gutiérrez; Melany Calderón |
| EPI_ISL_2827981, EPI_ISL_2828014 | HOSPITAL MONSEÑOR SANABRIA | Inciensa, Instituto Costarricense de Investigación y Enseñanza en Nutrición y Salud | Adriana Godínez; Claudio Soto-Garita; Estela Cordero; Francisco Duarte; Hebleen Porras; Joselyn Prado & María José Gómez; José Luis Vargas; Mariela Gutiérrez; Melany Calderón |
| EPI_ISL_2493882 | HOSPITAL MUN DR GUIDO GUIDA | Instituto Butantan | Antonio Jorge Martins; Claudia Renata dos Santos Barros; David Schlesinger; Debora Botequio Moretti; Dimas Tadeu Covas; Elaine Cristina Marqueze; Elaine Vieira Santos; Evandra Strazza Rodrigues; Heidge Fukumasu; Jayme Augusto de Souza-Neto; José Salvatore Leister Patané; Luiz Alcantara; Luiz Lehmann Coutinho; Maria Carolina Elias; Maurício Lacerda Nogueira; Rafael dos Santos Bezerra; Raul Machado Neto; Rejane Maria Tommasini Grotto; Ricardo Haddad; Sandra Coccuzzo Sampaio Vessoni; Simone Kashima; Svetoslav Nanev Slavov; Vincent Louis Viala |
| EPI_ISL_2493955 | HOSPITAL MUNICIPAL DE IBIUNA IBIUNA SP | Instituto Butantan | Antonio Jorge Martins; Claudia Renata dos Santos Barros; David Schlesinger; Debora Botequio Moretti; Dimas Tadeu Covas; Elaine Cristina Marqueze; Elaine Vieira Santos; Evandra Strazza Rodrigues; Heidge Fukumasu; Jayme Augusto de Souza-Neto; José Salvatore Leister Patané; Luiz Alcantara; Luiz Lehmann Coutinho; Maria Carolina Elias; Maurício Lacerda Nogueira; Rafael dos Santos Bezerra; Raul Machado Neto; Rejane Maria Tommasini Grotto; Ricardo Haddad; Sandra Coccuzzo Sampaio Vessoni; Simone Kashima; Svetoslav Nanev Slavov; Vincent Louis Viala |

| | | | |
|---|---|---|---|
| EPI_ISL_2462326, EPI_ISL_2494132, EPI_ISL_2494135, EPI_ISL_2494136, EPI_ISL_2494139, EPI_ISL_2494140 | HOSPITAL MUNICIPAL DE ITAPIRA | Instituto Butantan | Antonio Jorge Martins; Claudia Renata dos Santos Barros; David Schlesinger; Debora Botequio Moretti; Dimas Tadeu Covas; Elaine Cristina Marqueze; Elaine Vieira Santos; Evandra Strazza Rodrigues; Heidge Fukumasu; Jayme Augusto de Souza-Neto; José Salvatore Leister Patané; Luiz Alcantara; Luiz Lehmann Coutinho; Maria Carolina Elias; Maurício Lacerda Nogueira; Rafael dos Santos Bezerra; Raul Machado Neto; Rejane Maria Tommasini Grotto; Ricardo Haddad; Sandra Coccuzzo Sampaio Vessoni; Simone Kashima; Svetoslav Nanev Slavov; Vincent Louis Viala |
| EPI_ISL_2801347 | HOSPITAL MUNICIPAL DR EDUARDO DIAS | Analytical Competence Molecular Epidemiology Lab/ACME, Oswaldo Cruz Foundation, Ceara (FIOCRUZ CE) | Cleber Furtado Aksenen e Suzana Porto Almeida; Fabio Miyajima; Fernando Braga Stehling; Francisco Eder de Moura Lopes; Jamille Maria Mendes Bezerra; Joaquim César do Nascimento Sousa Junior; Pedro Miguel Carneiro Jeronimo; Thais Ferreira de Oliveira; Thais de Oliveira Costa; Ticiane Cavalcante de Souza; Veridiana Pessoa Miyajima |
| EPI_ISL_2462315, EPI_ISL_2493821, EPI_ISL_2493827 | HOSPITAL MUNICIPAL DR MARIO GATTI CAMPINAS | Instituto Butantan | Antonio Jorge Martins; Claudia Renata dos Santos Barros; David Schlesinger; Debora Botequio Moretti; Dimas Tadeu Covas; Elaine Cristina Marqueze; Elaine Vieira Santos; Evandra Strazza Rodrigues; Heidge Fukumasu; Jayme Augusto de Souza-Neto; José Salvatore Leister Patané; Luiz Alcantara; Luiz Lehmann Coutinho; Maria Carolina Elias; Maurício Lacerda Nogueira; Rafael dos Santos Bezerra; Raul Machado Neto; Rejane Maria Tommasini Grotto; Ricardo Haddad; Sandra Coccuzzo Sampaio Vessoni; Simone Kashima; Svetoslav Nanev Slavov; Vincent Louis Viala |
| EPI_ISL_2494164 | HOSPITAL MUNICIPAL JORGE CURY DE SANTA MARIA DA SERRA | Instituto Butantan | Antonio Jorge Martins; Claudia Renata dos Santos Barros; David Schlesinger; Debora Botequio Moretti; Dimas Tadeu Covas; Elaine Cristina Marqueze; Elaine Vieira Santos; Evandra Strazza Rodrigues; Heidge Fukumasu; Jayme Augusto de Souza-Neto; José Salvatore Leister Patané; Luiz Alcantara; Luiz Lehmann Coutinho; Maria Carolina Elias; Maurício Lacerda Nogueira; Rafael dos Santos Bezerra; Raul Machado Neto; Rejane Maria Tommasini Grotto; Ricardo Haddad; Sandra Coccuzzo Sampaio Vessoni; Simone Kashima; Svetoslav Nanev Slavov; Vincent Louis Viala |
| EPI_ISL_2801348, EPI_ISL_2801349, EPI_ISL_2801355, EPI_ISL_2801370, EPI_ISL_3102265, EPI_ISL_3102299, EPI_ISL_3102508, EPI_ISL_3536345, EPI_ISL_3912216, EPI_ISL_3912217, EPI_ISL_3912218, EPI_ISL_3912219, EPI_ISL_3912220 | | | |
| see above | HOSPITAL MUNICIPAL NOSSA SENHORA DOS MILAGRES | Analytical Competence Molecular Epidemiology Lab/ACME, Oswaldo Cruz Foundation, Ceara (FIOCRUZ CE) | Cleber Furtado Aksenen; Cleber Furtado Aksenen e Suzana Porto Almeida; Fabio Miyajima; Fernando Braga Stehling; Francisco Eder de Moura Lopes; Jamille Maria Mendes Bezerra; Joaquim Cesar do Nascimento Sousa Junior; Joaquim César do Nascimento Sousa Junior; Pedro Miguel Carneiro Jeronimo; Suzana Porto Almeida & Lucas Delerino on behalf of COVID-19 FIOCRUZ Genomic Network; Suzana Porto Almeida e Lucas Delerino; Thais Ferreira de Oliveira; Thais de Oliveira Costa; Ticiane Cavalcante de Souza; Veridiana Pessoa Miyajima |
| EPI_ISL_3761773, EPI_ISL_3761782 | HOSPITAL NACIONAL DE NIÑOS | Inciensa, Instituto Costarricense de Investigación y Enseñanza en Nutrición y Salud | Cristian Pérez-Corrales & Valeria Peralta-Barquero |
| EPI_ISL_3102505 | HOSPITAL REGIONAL DE IGUATU | Analytical Competence Molecular Epidemiology Lab/ACME, Oswaldo Cruz Foundation, Ceara (FIOCRUZ CE) | Cleber Furtado Aksenen; Fabio Miyajima; Fernando Braga Stehling; Francisco Eder de Moura Lopes; Jamille Maria Mendes Bezerra; Joaquim César do Nascimento Sousa Junior; Pedro Miguel Carneiro Jeronimo; Suzana Porto Almeida e Lucas Delerino; Thais Ferreira de Oliveira; Thais de Oliveira Costa; Ticiane Cavalcante de Souza; Veridiana Pessoa Miyajima |
| EPI_ISL_3102304 | HOSPITAL REGIONAL DO SERTAO CENTRAL | Analytical Competence Molecular Epidemiology Lab/ACME, Oswaldo Cruz Foundation, Ceara (FIOCRUZ CE) | Cleber Furtado Aksenen; Fabio Miyajima; Fernando Braga Stehling; Francisco Eder de Moura Lopes; Jamille Maria Mendes Bezerra; Joaquim César do Nascimento Sousa Junior; Pedro Miguel Carneiro Jeronimo; Suzana Porto Almeida e Lucas Delerino; Thais Ferreira de Oliveira; Thais de Oliveira Costa; Ticiane Cavalcante de Souza; Veridiana Pessoa Miyajima |
| EPI_ISL_3912225, EPI_ISL_3912226 | HOSPITAL REGIONAL UNIMED | Analytical Competence Molecular Epidemiology Lab/ACME, Oswaldo Cruz Foundation, Ceara (FIOCRUZ CE) | Cleber Furtado Aksenen; Fabio Miyajima; Fernando Braga Stehling; Francisco Eder de Moura Lopes; Jamille Maria Mendes Bezerra; Joaquim Cesar do Nascimento Sousa Junior; Pedro Miguel Carneiro Jeronimo; Suzana Porto Almeida & Lucas Delerino on behalf of COVID-19 FIOCRUZ Genomic Network; Thais Ferreira de Oliveira; Thais de Oliveira Costa; Ticiane Cavalcante de Souza; Veridiana Pessoa Miyajima |
| EPI_ISL_2827973, EPI_ISL_3037808, EPI_ISL_3037811, EPI_ISL_3298308 | HOSPITAL SAN JUAN DE DIOS | Inciensa, Instituto Costarricense de Investigación y Enseñanza en Nutrición y Salud | Adriana Godínez; Caterina Guzmán; Claudio Soto-Garita; Estela Cordero; Francisco Duarte; Hebleen Porras; Joselyn Prado & Juliana Mora; Joselyn Prado & Paola Sánchez; José Luis Vargas; Mariela Gutiérrez; Mariela Gutiérrez Joselyn Prado; Melany Calderón; Nazareth Ruiz & Juliana Mora |
| EPI_ISL_2827986 | HOSPITAL SAN VICENTE DE PAUL | Inciensa, Instituto Costarricense de Investigación y Enseñanza en Nutrición y Salud | Adriana Godínez; Claudio Soto-Garita; Estela Cordero; Francisco Duarte; Hebleen Porras; Joselyn Prado & Josiany Arce; José Luis Vargas; Mariela Gutiérrez; Melany Calderón |
| EPI_ISL_3102428, EPI_ISL_3102449, EPI_ISL_3102451, EPI_ISL_3536153, EPI_ISL_3536289, EPI_ISL_3536290 | HOSPITAL SAO JOSE DE DOENCAS INFECCIOSAS | Analytical Competence Molecular Epidemiology Lab/ACME, Oswaldo Cruz Foundation, Ceara (FIOCRUZ CE) | Cleber Furtado Aksenen; Fabio Miyajima; Fernando Braga Stehling; Francisco Eder de Moura Lopes; Jamille Maria Mendes Bezerra; Joaquim César do Nascimento Sousa Junior; Pedro Miguel Carneiro Jeronimo; Suzana Porto Almeida e Lucas Delerino; Thais Ferreira de Oliveira; Thais de Oliveira Costa; Ticiane Cavalcante de Souza; Veridiana Pessoa Miyajima |
| EPI_ISL_2494168, EPI_ISL_2494172 | HOSPITAL SAO JOSE ITIRAPINA | Instituto Butantan | Antonio Jorge Martins; Claudia Renata dos Santos Barros; David Schlesinger; Debora Botequio Moretti; Dimas Tadeu Covas; Elaine Cristina Marqueze; Elaine Vieira Santos; Evandra Strazza Rodrigues; Heidge Fukumasu; Jayme Augusto de Souza-Neto; José Salvatore Leister Patané; Luiz Alcantara; Luiz Lehmann Coutinho; Maria Carolina Elias; Maurício Lacerda Nogueira; Rafael dos Santos Bezerra; Raul Machado Neto; Rejane Maria Tommasini Grotto; Ricardo Haddad; Sandra Coccuzzo Sampaio Vessoni; Simone Kashima; Svetoslav Nanev Slavov; Vincent Louis Viala |
| EPI_ISL_3536335 | HOSPITAL SAO MATEUS | Analytical Competence Molecular Epidemiology Lab/ACME, Oswaldo Cruz Foundation, Ceara (FIOCRUZ CE) | Cleber Furtado Aksenen; Fabio Miyajima; Fernando Braga Stehling; Francisco Eder de Moura Lopes; Jamille Maria Mendes Bezerra; Joaquim César do Nascimento Sousa Junior; Pedro Miguel Carneiro Jeronimo; Suzana Porto Almeida e Lucas Delerino; Thais Ferreira de Oliveira; Thais de Oliveira Costa; Ticiane Cavalcante de Souza; Veridiana Pessoa Miyajima |
| EPI_ISL_3532488 | HOSPITAL UNIVERSITARIO 12 DE OCTUBRE | HOSPITAL UNIVERSITARIO 12 DE OCTUBRE | Carmen Martín-Higuera; Esther Viedma; Irene Múñoz-Gallego; M.ª Dolores Folgueira; Mar Aguilera; Noelia Moral; Rafael Delgado; Sagrario Zurita |
| EPI_ISL_3102435, EPI_ISL_3102454, EPI_ISL_3536326 | HOSPITAL UNIVERSITARIO WALTER CANTIDIO | Analytical Competence Molecular Epidemiology Lab/ACME, Oswaldo Cruz Foundation, Ceara (FIOCRUZ CE) | Cleber Furtado Aksenen; Fabio Miyajima; Fernando Braga Stehling; Francisco Eder de Moura Lopes; Jamille Maria Mendes Bezerra; Joaquim César do Nascimento Sousa Junior; Pedro Miguel Carneiro Jeronimo; Suzana Porto Almeida e Lucas Delerino; Thais Ferreira de Oliveira; Thais de Oliveira Costa; Ticiane Cavalcante de Souza; Veridiana Pessoa Miyajima |
| EPI_ISL_3235332, EPI_ISL_3235349, EPI_ISL_3235350, EPI_ISL_3235351, EPI_ISL_3235357 | Hackensack Medical Center | New York Genome Center | Amy Baldwin; Andre Corvelo; Barry Kreiswirth; David Perlin; Dayna M. Oschwald; Jose Mediavilla; Kaelea Composto; Kar Chow; Liang Chen; Marcus Cunningham; Michael Zody; Samantha Fennessey; Tom Maniatis |
| EPI_ISL_2663225, EPI_ISL_2727868, EPI_ISL_2727869, EPI_ISL_3118528 | HealthPartners Central Lab | Minnesota Department of Health, Public Health Laboratory | Alexandra Lorentz; Jacob Garfin; Matt Plumb; and Xiong Wang |
| EPI_ISL_3149731 | Hosp. Pediatrico Dr. Avelino Castelán | Laboratorio de Biología Molecular, Instituto de Medicina Regional on behalf of 'Proyecto Argentino Interinstitucional de genomica de SARS-CoV-2' (PAIS Consortium) | Bettina Brusés; Gioia Lucia Marino; Griselda Oria; Horacio Lucero.; Javier Mussin; Julieta Ronchi; Laura Formichelli; María Andrea Gili; María Cecilia López; Melina Lorenzini Campos; Raúl Maximiliano Acevedo |
| EPI_ISL_2923267 | Hospital | National Reference Center for Viruses of Respiratory Infections, Institut Pasteur, Paris | Angela Brisebarre; Camille Capel; Christophe Malabat; Corinne Maufrais; Etienne Simon-Lorière; FARFOUR Eric; Frédéric Lemoine; Gael Millot; Louise Lefrançois; Marion Barbet; Maud Vanpeene; Méline Bizard; Sylvie Behillil; Sylvie Van der Werf; Vincent Enouf |
| EPI_ISL_3092579 | Hospital Alvarez Buylla | Laboratorio de Virología HUCA | Abreu F; Alvarez-Arguelles ME; Boga JA; Castelló C; Costales I; Coto E; Gómez de Oña J; Martín-Rodríguez G; Melón S; Perez-Martínez Z; Rojo S; Sandoval M |
| EPI_ISL_2812647, EPI_ISL_3076926 | Hospital Carmen y Severo Ochoa | Laboratorio de Virología HUCA | Abreu F; Alvarez-Arguelles ME; Boga JA; Castelló C; Costales I; Coto E; Gómez de Oña J; Martín-Rodríguez G; Melón S; Perez-Martínez Z; Rojo S; Sandoval M |
| EPI_ISL_3147475 | Hospital Center Emile Mayrisch | Laboratoire national de sante, Microbiology, Microbial Genomics Platform | Anke Wienecke-Baldacchino; Catherine Ragimbeau; Cynthia Oxacelay; Elodie Solarino; Fatu Djabi; Jessica Tapp; Lise Pignon; Raoul Salmon; Tamir Abdelrahman; Virginie Jover |
| EPI_ISL_3147487, EPI_ISL_3147545 | Hospital Center Luxembourg | Laboratoire national de sante, Microbiology, Microbial Genomics Platform | Anke Wienecke-Baldacchino; Catherine Ragimbeau; Elodie Solarino; Fatu Djabi; Jean-Hugues Francois; Jessica Tapp; Lise Pignon; Michel Kohnen; Raoul Salmon; Tamir Abdelrahman; Virginie Jover |
| EPI_ISL_2402499, EPI_ISL_2484017, EPI_ISL_2484018, EPI_ISL_2601701, EPI_ISL_2601714, EPI_ISL_2675302, EPI_ISL_2782321, EPI_ISL_2782332, EPI_ISL_2782348, EPI_ISL_2782352, EPI_ISL_2782356, EPI_ISL_3167156, EPI_ISL_3505984, EPI_ISL_3505988 | | | |
| see above | Hospital General Universitario Gregorio Marañón | Hospital General Universitario Gregorio Marañón | Cristina Rodriguez-Grande; Darío García de Viedma; Julia Suárez; Laura Pérez-Lago; Marta Herranz Martin; Patricia Muñoz; Pedro Sola Campoy; Pilar Catalán; Sergio Buenestado Serrano; Victor Manuel de la Cueva |
| EPI_ISL_3216392 | Hospital General Universitario de Alicante - Instituto de Investigación Sanitaria y Biomédica de Alicante | SeqCOVID-SPAIN consortium/IBV(CSIC) | Carmen Molina Pardines and SeqCOVID-SPAIN consortium; Maripaz Ventero Martín |
| EPI_ISL_3033082, EPI_ISL_3033084, EPI_ISL_3033086, EPI_ISL_3260092, EPI_ISL_3260093, EPI_ISL_3260094, EPI_ISL_3260108 | | | |
| see above | Hospital Jaime Ferre - SAMCO Rafaela | Grupo de Genómica y Bioinformática del Instituto de Investigación de la Cadena Láctea CONICET-INTA on behalf of | AF; Aliprandi D; Amadio; C; Eberhardt; Irazoqui; Isaia; JF; JM; MF; Pandolfi; Quaranta; Soratti R; V |

| | | | |
|---|---|---|---|
| | | 'Proyecto Argentino Interinstitucional de genomica de SARS-CoV-2' (PAIS Consortium) | |
| EPI_ISL_2894556, EPI_ISL_2894559, EPI_ISL_2987697, EPI_ISL_2987700, EPI_ISL_2987707 | Hospital Margarita Maza de Juárez | Microbial Genomics Laboratory | ; Alejandra García-Gasca; Alejandra Hernández-Terán; Alejandro Sánchez-Flores; Alfredo Herrera-Estrella; Alicia Ocaña-Mondragón; Andreu Comas-García; Angel Gustavo Salas-Lais; Antonio Loza Román; Bernardo Martínez-Miguel; Blanca Taboada; Brenda Irasema Maldonado-Meza; Bruno Gómez-Gil; Carla Ivón Herrera-Najera; Carlos F. Arias; Celia Boukadida; Clara Esperanza Santacruz-Tinoco; Concepción Grajales-Muñiz; Consorcio Mexicano de Vigilancia Genómica (CoViGen-Mex). Authors (in alphabetical order): Julio Elias Alvarado-Yaah; Cristóbal Cháidez-Quiróz; Célida Duque Molina; Célida Martínez- Rodríguez; Daniel Fregoso-Rueda; Daniel Lira Morales; Eduardo Becerril-Vargas; Fernando Fontove-Herrera; Fidencio Mejía-Nepomuceno; Francisco Pulido; Gloria Elena Espinosa-Ayala; Gloria María Molina-Salinas; Gloria Vazquez; Hector Esteban Paz-Juárez; Hector Montoya-Fuentes; Helen Haydee Fernanda Ramírez-Plascencia; Irvin González-López; Jean Pierre González; Jesús Hernández; Joel Armando Vázquez-Pérez.; Jorge Salas-Hernández; José Antonio Enciso-Moreno; José Arturo Martínez-Orozco; José Esteban Muñoz-Medina; José de Jesús Nuñez-Contreras; Juan Bautista Chale-Dzul; Julissa Enciso-Ibarra; Luis Alberto Ochoa-Carrera; Margarita Matías-Florentino; Mario Mújica-Sánchez; Marissa Perez-Garcia; María Guadalupe Santiago-Mauricio; María Guadalupe de Jesús Mireles-Rivera; Nelly Sélem-Mojica; Pavel Isa; Ricardo Ciria Merce; Ricardo Grande; Rosa María Gutiérrez Rios; Santiago Ávila-Ríos; Selene Zárate; Susana Lopez; Verónica Mata-Haro; Victor Eduardo García-Arias; Víctor Hugo Borja-Aburto |
| EPI_ISL_2524776, EPI_ISL_2628167, EPI_ISL_2628170, EPI_ISL_2628171, EPI_ISL_2828226, EPI_ISL_2828227, EPI_ISL_2828233 | | | |
| see above | Hospital Universitari Arnau de Vilanova | Hospital Universitari Vall d'Hebron - Vall d'Hebron Institut de Recerca | Alejandra González-Sánchez; Andrés Antón; Ariadna Rando; Carla Castillo; Cristina Andrés; Damir Garcia-Cehic; Josep Quer; Juliana Esperalba; Maria Carmen Martin; Maria Gema Codina; Maria Piñana; Tomàs Pumarola |
| EPI_ISL_2637324, EPI_ISL_2861766 | Hospital Universitari Bellvitge | Microbiology Department | Aida Gonzalez-Diaz; Carmen Ardanuy; Jordi Camara; Jordi Niubó; Laura Calatayud; M Angeles Domínguez; Miguel Fernandez-Huerta; Sara Marti |
| EPI_ISL_2566592, EPI_ISL_2566593, EPI_ISL_2566594, EPI_ISL_2566595, EPI_ISL_2566596, EPI_ISL_2652058, EPI_ISL_2652059, EPI_ISL_2652064, EPI_ISL_2657359, EPI_ISL_2657366 | | | |
| see above | Hospital Universitari Joan XXIII | Hospital Universitari Joan XXIII | Carla Martín; Clara Benavent; Cristina Gutiérrez; Ester Picó; Gemma Recio; Margarida Terrón; Natalia Bastón |
| EPI_ISL_2712852, EPI_ISL_2712870, EPI_ISL_2828111, EPI_ISL_2828116, EPI_ISL_2828151, EPI_ISL_2828171, EPI_ISL_2828215, EPI_ISL_2828217, EPI_ISL_2885302, EPI_ISL_2982331, EPI_ISL_2982387, EPI_ISL_3009983, EPI_ISL_3984105 | | | |
| see above | Hospital Universitari Vall d'Hebron - Vall d'Hebron Institut de Recerca | Hospital Universitari Vall d'Hebron - Vall d'Hebron Institut de Recerca | Alejandra González-Sánchez; Andrés Antón; Ariadna Rando; Carla Castillo; Cristina Andrés; Damir Garcia-Cehic; Josep Quer; Juliana Esperalba; Karen García; Maria Carmen Martin; Maria Gema Codina; Maria Piñana; Rodrigo Vásquez; Tomàs Pumarola |
| EPI_ISL_2600854, EPI_ISL_2775934, EPI_ISL_2775946, EPI_ISL_2775959, EPI_ISL_2775966, EPI_ISL_2775970, EPI_ISL_2820672, EPI_ISL_2931444, EPI_ISL_2931450, EPI_ISL_3087929, EPI_ISL_3087940 | | | |
| see above | Hospital Universitari i Politècnic La Fe de València | SeqCOVID-SPAIN consortium/IBV(CSIC) | Ana Gil Brusola; Eva González Barberá; José Luis López Hontangas and SeqCOVID-SPAIN consortium; María Dolores Gómez Ruiz; Salvador Giner Almaraz |
| EPI_ISL_2545330, EPI_ISL_2620963, EPI_ISL_2620966, EPI_ISL_2773856 | Hospital Universitario Virgen de las Nieves de Granada-SAS | SeqCOVID-SPAIN consortium/IBV(CSIC) | Irene Pedrosa Corral; José M. Navarro-Marí and SeqCOVID-SPAIN consortium; Sara Sanbonmatsu Gámez |
| EPI_ISL_2925674, EPI_ISL_2925675, EPI_ISL_2925676, EPI_ISL_2925677 | Hospital de Cabueñes | Laboratorio de Virología HUCA | Abreu F; Alvarez-Arguelles ME; Boga JA; Castelló C; Costales I; Coto E; Gómez de Oña J; Martín-Rodríguez G; Melón; Melón S; Perez-Martínez Z; Rojo S; Sandoval M |
| EPI_ISL_2788571 | Hospital of Southern Norway - Kristiansand, Department of Medical Microbiology | Norwegian Institute of Public Health, Department of Virology | Atiya R Ali; Debech Nadia; Engebretsen Serina Beate; Garcia Llorente Ignacio; Hilde Elshaug; Hilde Vollan; Jon Bråte; Kamilla Heddeland Instefjord; Karoline Bragstad; Kathrine Stene-Johansen; Line Victoria Moen; Marie Paulsen Madsen; Olav Hungnes; Pedersen Benedikte Nevjen; Rasmus Riis Kopperud |
| EPI_ISL_3411959 | Hospital of the University of Pennsylvania Molecular Pathology Lab | Bushman | Andrew D. Marques; Aoife M. Roche; Arupa Ganguly; Ayannah S. Fitzgerald; Brendan Kelly; Jevon Graham-Wooten; John K. Everett; Kyle Rodino; Layla A. Khatib; Mike Feldman; Ronald G. Collman and Frederic Bushan; Samantha A. Whiteside; Scott Sherrill-Mix; Shantan Reddy; Young Hwang |
| EPI_ISL_2597697, EPI_ISL_3023264, EPI_ISL_3023266, EPI_ISL_3023276, EPI_ISL_3023329, EPI_ISL_3023330, EPI_ISL_3023373 | | | |
| see above | Hospital of the University of Pennsylvania Molecular Pathology Lab | Bushman Lab - University of Pennsylvania | Aoife M. Roche; Arupa Ganguly; Ayannah S. Fitzgerald; Brendan Kelly; Jevon Graham-Wooten; John K. Everett; Kyle Rodino; Layla A. Khatib; Mike Feldman; Ronald G. Collman and Frederic Bushan; Samantha A. Whiteside; Scott Sherrill-Mix; Shantan Reddy; Young Hwang |
| EPI_ISL_2616771, EPI_ISL_2968598, EPI_ISL_2968613, EPI_ISL_2968620, EPI_ISL_2968636, EPI_ISL_2968664, EPI_ISL_2968665, EPI_ISL_3398946 | | | |
| see above | Houston Health Dept. | Houston Health Dept. | Adolpho Lara; Pamela Brown; Ryker Penn; Yanlai Lai |
| EPI_ISL_2689890, EPI_ISL_2787361, EPI_ISL_2787367, EPI_ISL_2787369, EPI_ISL_2787380, EPI_ISL_2787389, EPI_ISL_2787392, EPI_ISL_3355076 | | | |
| see above | IA State Hygienic Laboratory | Centers for Disease Control and Prevention Division of Viral Diseases, Pathogen Discovery | Alex Burgin; Alison Laufer Halpin; Ben L. Rambo-Martin; Ben Rambo-Martin; Clinton Paden; Clinton R. Paden; Dakota Howard; Darlene Wagner; Dave Wentworth; Dhwani Batra; Jasmine Padilla; Justin Lee; Katie Dillon; Krista Queen; Kristen Knipe; Kristine Lacek; Mark Burroughs; Matthew Schmerer; Meghan Bentz; Mili Sheth; Peter Cook; Sam Shepard; Sarah Nobles; Shoshona Le; Suxiang Tong; Vivien Dugan; Yvette Unoarumhi |
| EPI_ISL_2690063, EPI_ISL_2690064, EPI_ISL_2840317, EPI_ISL_2840320, EPI_ISL_2987054, EPI_ISL_3128023, EPI_ISL_3128024, EPI_ISL_3128028, EPI_ISL_3355243, EPI_ISL_3355266, EPI_ISL_3355267, EPI_ISL_3355326 | | | |
| see above | ID Bureau of Laboratories | Centers for Disease Control and Prevention Division of Viral Diseases, Pathogen Discovery | Alex Burgin; Alison Laufer Halpin; Ben L. Rambo-Martin; Ben Rambo-Martin; Clinton Paden; Clinton R. Paden; Dakota Howard; Darlene Wagner; Dave Wentworth; Dhwani Batra; Jasmine Padilla; Justin Lee; Katie Dillon; Krista Queen; Kristen Knipe; Kristine Lacek; Mark Burroughs; Matthew Schmerer; Meghan Bentz; Mili Sheth; Peter Cook; Sam Shepard; Sarah Nobles; Shoshona Le; Suxiang Tong; Vivien Dugan; Yvette Unoarumhi |
| EPI_ISL_2812810 | IFO Regina Elena | National Institute for Infectious Diseases (INMI) L. Spallanzani I.R.C.C.S | A Di Caro; B Bartolini; CEM Gruber; E Giombini; F Messina; F Santini; G Bonfiglio; M Rueca; MR Capobianchi; O Butera |
| EPI_ISL_2788130, EPI_ISL_2788133, EPI_ISL_2788138, EPI_ISL_2788144, EPI_ISL_2788147, EPI_ISL_2840290 | IL Dept. of Public Health Springfield Laboratory | Centers for Disease Control and Prevention Division of Viral Diseases, Pathogen Discovery | Alison Laufer Halpin; Ben L. Rambo-Martin; Clinton R. Paden; Dakota Howard; Darlene Wagner; Dave Wentworth; Dhwani Batra; Jasmine Padilla; Justin Lee; Katie Dillon; Krista Queen; Kristen Knipe; Kristine Lacek; Mark Burroughs; Matthew Schmerer; Mili Sheth; Peter Cook; Sam Shepard; Sarah Nobles; Shoshona Le; Suxiang Tong; Vivien Dugan; Yvette Unoarumhi |
| EPI_ISL_2544640, EPI_ISL_2544660, EPI_ISL_2557242, EPI_ISL_2725041, EPI_ISL_2860400, EPI_ISL_3030076 | IN State Department of Health Laboratory Services | IN State Department of Health Laboratory Services | Brian Pope; Cassandra Campion; Jamie Yeadon; Kyle Brownlee; Lixia Liu; Mark Glazier; Melissa Hindenlang |
| EPI_ISL_2493987, EPI_ISL_2493994, EPI_ISL_2494001, EPI_ISL_2494327 | INSIDE CRSN CACHOEIRINHA/CASA VERDE | Instituto Butantan | Antonio Jorge Martins; Claudia Renata dos Santos Barros; David Schlesinger; Debora Botequio Moretti; Dimas Tadeu Covas; Elaine Cristina Marqueze; Elaine Vieira Santos; Evandra Strazza Rodrigues; Heidge Fukumasu; Jayme Augusto de Souza-Neto; José Salvatore Leister Patané; Luiz Alcantara; Luiz Lehmann Coutinho; Maria Carolina Elias; Maurício Lacerda Nogueira; Rafael dos Santos Bezerra; Raul Machado Neto; Rejane Maria Tommasini Grotto; Ricardo Haddad; Sandra Coccuzzo Sampaio Vessoni; Simone Kashima; Svetoslav Nanev Slavov; Vincent Louis Viala |
| EPI_ISL_2493973, EPI_ISL_2493982, EPI_ISL_2493988, EPI_ISL_2493992, EPI_ISL_2493998, EPI_ISL_2494024, EPI_ISL_2494035 | | | |
| see above | INSIDE CRSN FREGUESIA/BRASILÂNDIA | Instituto Butantan | Antonio Jorge Martins; Claudia Renata dos Santos Barros; David Schlesinger; Debora Botequio Moretti; Dimas Tadeu Covas; Elaine Cristina Marqueze; Elaine Vieira Santos; Evandra Strazza Rodrigues; Heidge Fukumasu; Jayme Augusto de Souza-Neto; José Salvatore Leister Patané; Luiz Alcantara; Luiz Lehmann Coutinho; Maria Carolina Elias; Maurício Lacerda Nogueira; Rafael dos Santos Bezerra; Raul Machado Neto; Rejane Maria Tommasini Grotto; Ricardo Haddad; Sandra Coccuzzo Sampaio Vessoni; Simone Kashima; Svetoslav Nanev Slavov; Vincent Louis Viala |
| EPI_ISL_2493979, EPI_ISL_2494026 | INSIDE CRSN PERUS | Instituto Butantan | Antonio Jorge Martins; Claudia Renata dos Santos Barros; David Schlesinger; Debora Botequio Moretti; Dimas Tadeu Covas; Elaine Cristina Marqueze; Elaine Vieira Santos; Evandra Strazza Rodrigues; Heidge Fukumasu; Jayme Augusto de Souza-Neto; José Salvatore Leister Patané; Luiz Alcantara; Luiz Lehmann Coutinho; Maria Carolina Elias; Maurício Lacerda Nogueira; Rafael dos Santos Bezerra; Raul Machado Neto; Rejane Maria Tommasini Grotto; Ricardo Haddad; Sandra Coccuzzo Sampaio Vessoni; Simone Kashima; Svetoslav Nanev Slavov; Vincent Louis Viala |
| EPI_ISL_2493201, EPI_ISL_2493967, EPI_ISL_2493971, EPI_ISL_2493993, EPI_ISL_2494003, EPI_ISL_2494027 | INSIDE CRSN PIRITUBA | Instituto Butantan | Antonio Jorge Martins; Claudia Renata dos Santos Barros; David Schlesinger; Debora Botequio Moretti; Dimas Tadeu Covas; Elaine Cristina Marqueze; Elaine Vieira Santos; Evandra Strazza Rodrigues; Heidge Fukumasu; Jayme Augusto de Souza-Neto; José Salvatore Leister Patané; Luiz Alcantara; Luiz Lehmann Coutinho; Maria Carolina Elias; Maurício Lacerda Nogueira; Rafael dos Santos Bezerra; Raul Machado Neto; Rejane Maria Tommasini Grotto; Ricardo Haddad; Sandra Coccuzzo Sampaio Vessoni; Simone Kashima; Svetoslav Nanev Slavov; Vincent Louis Viala |
| EPI_ISL_2493259, EPI_ISL_2493984, EPI_ISL_2494006, EPI_ISL_2494007 | INSIDE CRSN VILA MARIA/VILA GUILHERME | Instituto Butantan | Antonio Jorge Martins; Claudia Renata dos Santos Barros; David Schlesinger; Debora Botequio Moretti; Dimas Tadeu Covas; Elaine Cristina Marqueze; Elaine Vieira Santos; Evandra Strazza Rodrigues; Heidge Fukumasu; Jayme Augusto de Souza-Neto; José Salvatore Leister Patané; Luiz Alcantara; Luiz Lehmann Coutinho; Maria Carolina Elias; Maurício Lacerda Nogueira; Rafael dos Santos Bezerra; Raul Machado Neto; Rejane Maria Tommasini Grotto; Ricardo Haddad; Sandra Coccuzzo Sampaio Vessoni; Simone Kashima; Svetoslav Nanev Slavov; Vincent Louis Viala |
| EPI_ISL_2493265, EPI_ISL_2493968, EPI_ISL_2494000, EPI_ISL_2494030, EPI_ISL_2494033 | INSIDE CRSO BUTANTÃ | Instituto Butantan | Antonio Jorge Martins; Claudia Renata dos Santos Barros; David Schlesinger; Debora Botequio Moretti; Dimas Tadeu Covas; Elaine Cristina Marqueze; Elaine Vieira Santos; Evandra Strazza Rodrigues; Heidge Fukumasu; Jayme Augusto de Souza-Neto; José Salvatore Leister Patané; Luiz Alcantara; Luiz Lehmann Coutinho; Maria Carolina Elias; Maurício Lacerda Nogueira; Rafael dos Santos Bezerra; Raul Machado Neto; Rejane Maria Tommasini Grotto; Ricardo Haddad; Sandra Coccuzzo Sampaio Vessoni; Simone Kashima; Svetoslav Nanev Slavov; Vincent Louis Viala |
| EPI_ISL_2493200 | INSIDE CRSO LAPA/PINHEIROS | Instituto Butantan | Antonio Jorge Martins; Claudia Renata dos Santos Barros; David Schlesinger; Debora Botequio Moretti; Dimas Tadeu Covas; Elaine Cristina Marqueze; Elaine Vieira Santos; Evandra Strazza Rodrigues; Heidge Fukumasu; Jayme Augusto de Souza-Neto; José Salvatore Leister Patané; Luiz Alcantara; Luiz Lehmann Coutinho; Maria Carolina Elias; Maurício Lacerda Nogueira; Rafael dos Santos Bezerra; Raul Machado Neto; Rejane Maria Tommasini Grotto; Ricardo Haddad; Sandra Coccuzzo Sampaio Vessoni; Simone Kashima; Svetoslav Nanev Slavov; Vincent Louis Viala |
| EPI_ISL_2493964, EPI_ISL_2493972, EPI_ISL_2493976, EPI_ISL_2493978, EPI_ISL_2493995, EPI_ISL_2494012, EPI_ISL_2494016, EPI_ISL_2494017, EPI_ISL_2494018, EPI_ISL_2494029, EPI_ISL_2494034, EPI_ISL_2494038 | | | |
| see above | INSIDE CRSS CAPELA DO SOCORRO | Instituto Butantan | Antonio Jorge Martins; Claudia Renata dos Santos Barros; David Schlesinger; Debora Botequio Moretti; Dimas Tadeu Covas; Elaine Cristina Marqueze; Elaine Vieira Santos; Evandra Strazza Rodrigues; Heidge Fukumasu; Jayme Augusto de Souza-Neto; José Salvatore Leister Patané; Luiz Alcantara; Luiz Lehmann Coutinho; Maria Carolina Elias; Maurício Lacerda Nogueira; Rafael dos Santos Bezerra; Raul Machado Neto; Rejane Maria Tommasini Grotto; Ricardo Haddad; Sandra Coccuzzo Sampaio Vessoni; Simone Kashima; Svetoslav Nanev Slavov; Vincent Louis Viala |
| EPI_ISL_2493960, EPI_ISL_2493961, EPI_ISL_2493977, EPI_ISL_2493989, EPI_ISL_2493990, EPI_ISL_2494005, EPI_ISL_2494036 | | | |

| | | | |
|---|---|---|---|
| see above | INSIDE CRSS PARELHEIROS | Instituto Butantan | Antonio Jorge Martins; Claudia Renata dos Santos Barros; David Schlesinger; Debora Botequio Moretti; Dimas Tadeu Covas; Elaine Cristina Marqueze; Elaine Vieira Santos; Evandra Strazza Rodrigues; Heidge Fukumasu; Jayme Augusto de Souza-Neto; José Salvatore Leister Patané; Luiz Alcantara; Luiz Lehmann Coutinho; Maria Carolina Elias; Maurício Lacerda Nogueira; Rafael dos Santos Bezerra; Raul Machado Neto; Rejane Maria Tommasini Grotto; Ricardo Haddad; Sandra Coccuzzo Sampaio Vessoni; Simone Kashima; Svetoslav Nanev Slavov; Vincent Louis Viala |
| EPI_ISL_2493274, EPI_ISL_2493996, EPI_ISL_2494316 | INSIDE CRSSU IPIRANGA | Instituto Butantan | Antonio Jorge Martins; Claudia Renata dos Santos Barros; David Schlesinger; Debora Botequio Moretti; Dimas Tadeu Covas; Elaine Cristina Marqueze; Elaine Vieira Santos; Evandra Strazza Rodrigues; Heidge Fukumasu; Jayme Augusto de Souza-Neto; José Salvatore Leister Patané; Luiz Alcantara; Luiz Lehmann Coutinho; Maria Carolina Elias; Maurício Lacerda Nogueira; Rafael dos Santos Bezerra; Raul Machado Neto; Rejane Maria Tommasini Grotto; Ricardo Haddad; Sandra Coccuzzo Sampaio Vessoni; Simone Kashima; Svetoslav Nanev Slavov; Vincent Louis Viala |
| EPI_ISL_2493970 | INSIDE CRSSU JABAQUARA/VILA MARIANA | Instituto Butantan | Antonio Jorge Martins; Claudia Renata dos Santos Barros; David Schlesinger; Debora Botequio Moretti; Dimas Tadeu Covas; Elaine Cristina Marqueze; Elaine Vieira Santos; Evandra Strazza Rodrigues; Heidge Fukumasu; Jayme Augusto de Souza-Neto; José Salvatore Leister Patané; Luiz Alcantara; Luiz Lehmann Coutinho; Maria Carolina Elias; Maurício Lacerda Nogueira; Rafael dos Santos Bezerra; Raul Machado Neto; Rejane Maria Tommasini Grotto; Ricardo Haddad; Sandra Coccuzzo Sampaio Vessoni; Simone Kashima; Svetoslav Nanev Slavov; Vincent Louis Viala |
| EPI_ISL_2494004, EPI_ISL_2494028, EPI_ISL_2494031, EPI_ISL_2494310 | INSIDE CRSSU MOOCA/ARICANDUVA | Instituto Butantan | Antonio Jorge Martins; Claudia Renata dos Santos Barros; David Schlesinger; Debora Botequio Moretti; Dimas Tadeu Covas; Elaine Cristina Marqueze; Elaine Vieira Santos; Evandra Strazza Rodrigues; Heidge Fukumasu; Jayme Augusto de Souza-Neto; José Salvatore Leister Patané; Luiz Alcantara; Luiz Lehmann Coutinho; Maria Carolina Elias; Maurício Lacerda Nogueira; Rafael dos Santos Bezerra; Raul Machado Neto; Rejane Maria Tommasini Grotto; Ricardo Haddad; Sandra Coccuzzo Sampaio Vessoni; Simone Kashima; Svetoslav Nanev Slavov; Vincent Louis Viala |
| EPI_ISL_2493962, EPI_ISL_2493965, EPI_ISL_2493983, EPI_ISL_2493999, EPI_ISL_2494009, EPI_ISL_2494019, EPI_ISL_2494020 | | | |
| see above | INSIDE CRSSU SAPOPEMBA/VILA PRUDENTE | Instituto Butantan | Antonio Jorge Martins; Claudia Renata dos Santos Barros; David Schlesinger; Debora Botequio Moretti; Dimas Tadeu Covas; Elaine Cristina Marqueze; Elaine Vieira Santos; Evandra Strazza Rodrigues; Heidge Fukumasu; Jayme Augusto de Souza-Neto; José Salvatore Leister Patané; Luiz Alcantara; Luiz Lehmann Coutinho; Maria Carolina Elias; Maurício Lacerda Nogueira; Rafael dos Santos Bezerra; Raul Machado Neto; Rejane Maria Tommasini Grotto; Ricardo Haddad; Sandra Coccuzzo Sampaio Vessoni; Simone Kashima; Svetoslav Nanev Slavov; Vincent Louis Viala |
| EPI_ISL_2893596, EPI_ISL_2893597, EPI_ISL_2893598, EPI_ISL_2893599, EPI_ISL_2893600, EPI_ISL_2893601, EPI_ISL_2893602, EPI_ISL_2893604, EPI_ISL_2893605, EPI_ISL_2897544, EPI_ISL_2988448, EPI_ISL_2988462, EPI_ISL_2988463, EPI_ISL_2988466, EPI_ISL_2988479, EPI_ISL_2988486, EPI_ISL_2988501, EPI_ISL_2988502, EPI_ISL_2988506, EPI_ISL_2988519, EPI_ISL_2988528, EPI_ISL_2988529, EPI_ISL_2988531, EPI_ISL_2988552, EPI_ISL_2988556, EPI_ISL_2988560, EPI_ISL_2988561, EPI_ISL_2988568, EPI_ISL_2988573, EPI_ISL_2988577, EPI_ISL_2988581, EPI_ISL_3010437, EPI_ISL_3068144, EPI_ISL_3068159, EPI_ISL_3068166, EPI_ISL_3068194, EPI_ISL_3068213, EPI_ISL_3068218, EPI_ISL_3068223, EPI_ISL_3068228, EPI_ISL_3068231, EPI_ISL_3133207, EPI_ISL_3274398, EPI_ISL_3274462, EPI_ISL_3274503, EPI_ISL_3274512, EPI_ISL_3274524, EPI_ISL_3274539, EPI_ISL_3274540, EPI_ISL_3274593 | | | |
| see above | INSPI-CRN DE INFLUENZA Y OTROS VIRUS RESPIRATORIOS | NIC-INSPI | Alfredo Bruno; Daniel Ramos; Domenica de Mora.; Jimmy Garcés; Johanna Laines; Lizbeth Patiño; Manuel Gonzalez; Marcela Mejía; Maria Angelica Becerra; Maritza Olmedo; Mayra Wilca; Michelle Páez; Milton Medina |
| EPI_ISL_2604875, EPI_ISL_2604876, EPI_ISL_2604877, EPI_ISL_3088317, EPI_ISL_3088319 | INTERLAB | Omics Sciences Laboratory | Darlyn Amaya; Derly Andrade Molina; Gabriel Morey León; Juan Carlos Fernández Cadena; Rubén Armas González |
| EPI_ISL_2923003, EPI_ISL_2923039, EPI_ISL_2923047, EPI_ISL_2985121, EPI_ISL_2985167, EPI_ISL_2985302, EPI_ISL_3068250 | | | |
| see above | IRCCS San Gallicano Dermatological Institute | IRCCS Regina Elena National Cancer Institute | Aldo Morrone; Alice Massacci; Andrea Cazzani; Eleonora Sperandio; Elisabetta Trento; Fabrizio Ensoli; Francesca De Nicola; Frauke Goeman; Fulvia Pimpinelli; Gennaro Ciliberto; Giovanni Blandino; Giulia Orlandi; Ilaria Cavallo; Ludovica Ciuffreda; Martina Pontone; Matteo Pallocca; Maurizio Fanciulli; Sara Donzelli; Sara Petrolo; Serena Salvo; di Domenico Enea Gino |
| EPI_ISL_2726937, EPI_ISL_2726940, EPI_ISL_2757934, EPI_ISL_2757935, EPI_ISL_2757937 | ISDH | Quantigen Biosciences | Paul J. Childress |
| EPI_ISL_2628209, EPI_ISL_2628300, EPI_ISL_2628305 | IVIC | Laboratorio de Virología Molecular | Carmen L Loureiro; CoViVen Group; Domingo J Garzaro; Esmeralda Vizzi; Flor H Pujol; Héctor R Rangel; José Luis Zambrano; Lieska Rodríguez; Mariana Hidalgo; Pierina D´Angelo; Rossana C Jaspe; Víctor Alarcón; Yoneira Sulbaran; Zoila Moros |
| EPI_ISL_2789980, EPI_ISL_2801469, EPI_ISL_3010167, EPI_ISL_3281530 | IVY3 Central Lab, Vanderbilt University Medical Center | Lauring Lab, University of Michigan, Department of Microbiology and Immunology | Gilbert; Valesano |
| EPI_ISL_3090836, EPI_ISL_3090838, EPI_ISL_3090841, EPI_ISL_3100649, EPI_ISL_3100653 | Idaho Bureau of Laboratories | Boise VA Medical Center, PALMS | Aimee Ceniseros; Cheri Lamb McFarlane; Christian Loera; Dr. Christopher Ball; Emily Bartlette; James Razor; Matthew Burns; Ying Pei |
| EPI_ISL_3066972, EPI_ISL_3066973 | Idaho Bureau of Laboratories | Center for Global Health, University of New Mexico Health Sciences Center | Christopher Ball; Darrell Dinwiddie; Daryl Domman; Kurt Schwalm; Matthew Burns; Robert Voermans; Valerie Morley |
| EPI_ISL_2801140, EPI_ISL_2801141, EPI_ISL_3254817 | Illinois Department of Public Health | Gagnon Lab, Southern Illinois University | Keith Gagnon |
| EPI_ISL_2800846, EPI_ISL_2800849, EPI_ISL_2800850, EPI_ISL_2800851, EPI_ISL_2800852, EPI_ISL_2800853, EPI_ISL_2800857, EPI_ISL_2800859, EPI_ISL_2800861, EPI_ISL_2800862, EPI_ISL_2800863, EPI_ISL_2800865, EPI_ISL_3232014, EPI_ISL_3232015, EPI_ISL_3232018, EPI_ISL_3232020, EPI_ISL_3232021 | | | |
| see above | Illinois Department of Public Health | Illinois Department of Public Health - Chicago Lab | Ira Heimler; Joel Price; Vineet K. Dhiman |
| EPI_ISL_2403009, EPI_ISL_2403012, EPI_ISL_2403053, EPI_ISL_2403055, EPI_ISL_2403074, EPI_ISL_2403083, EPI_ISL_2403148, EPI_ISL_2403158, EPI_ISL_2403165, EPI_ISL_2403168, EPI_ISL_2403173, EPI_ISL_2403175, EPI_ISL_2403176, EPI_ISL_2403177, EPI_ISL_2403179, EPI_ISL_2403180, EPI_ISL_2403183, EPI_ISL_2598169, EPI_ISL_2598176, EPI_ISL_2598186, EPI_ISL_2598202, EPI_ISL_2598206, EPI_ISL_2598207, EPI_ISL_2598244, EPI_ISL_2598245, EPI_ISL_2598246, EPI_ISL_2774312, EPI_ISL_2774316, EPI_ISL_2774320, EPI_ISL_2774330, EPI_ISL_2774331, EPI_ISL_2774332, EPI_ISL_2774333, EPI_ISL_2774336, EPI_ISL_2774339, EPI_ISL_2774346, EPI_ISL_2774353, EPI_ISL_2774365, EPI_ISL_2774367, EPI_ISL_2774368, EPI_ISL_2774370, EPI_ISL_2774371, EPI_ISL_2774380, EPI_ISL_2774382, EPI_ISL_2774383, EPI_ISL_2774384, EPI_ISL_2774385, EPI_ISL_2774389, EPI_ISL_2774391, EPI_ISL_2774392, EPI_ISL_2885392, EPI_ISL_2885410, EPI_ISL_2885459, EPI_ISL_2885461 | | | |
| see above | Illinois Department of Public Health - Springfield Lab | Illinois Department of Public Health - Springfield Lab | Bryan Sim; Gordon McCall |
| EPI_ISL_2715220, EPI_ISL_2735971, EPI_ISL_3015619, EPI_ISL_3015628, EPI_ISL_3015629, EPI_ISL_3105046, EPI_ISL_3105047, EPI_ISL_3105050, EPI_ISL_3105051, EPI_ISL_3105052, EPI_ISL_3347000 | | | |
| see above | Incyte Diagnostics Spokane | Seattle Flu Study | Amanda Adler; Barry R. Lutz; Benjamin Pelle; Brian Hiatt; Caitlin R. Wolf; Chris D. Frazar; Deborah A. Nickerson; Elisabeth Brandstetter; Erica Ryke; Geoff Melly; Helen Y. Chu; Janet A. Englund; Jay Shendure; Jover Lee; Kairsten Fay; Kirsten Lacombe; Lea M. Starita; Mark J. Rieder; Matthew Richardson; Matthew Thompson; Melissa Truong; Michael Boeckh; Michael Famulare; Misja Ilcisin; Peter D. Han; Philip Dykema; Romesh Gautom; Scott Lindquist; Thomas R. Sibley; Trevor Bedford |
| EPI_ISL_2660401, EPI_ISL_2660405 | Infectious Disease Program, Broad Institute of Harvard and MIT | Infectious Disease Program, Broad Institute of Harvard and MIT | A.E.; Adams, G.; Anahtar, M.; B.L.; B.W.; Bauer, M.; Birren; Branda, J.; Carter, A.; Cerrato, F.; Chaluvadi, S.; Chapman; Cusick, C.; D.J.; DeRuff, K.; Flowers, K.; Gallagher, G.; Gladden-Young, A.; Gnirke, A.; Harris, J.; J.E.; K.J.; LaRocque, R.; Lagerborg, K.; Lemieux; Lin; Loreth, C.; MacInnis; Neumann, A.; Normandin, E.; P.C.; Park; Pierce, V.; Reilly, S.; Rosenberg, E.; Rudy, M.; Ryan, E.; S.B.; Sabeti; Shaw, B.; Siddle; Slater, D.; Smole, S.; Tomkins-Tinch, C.; Turbett, S. |
| EPI_ISL_2557441, EPI_ISL_2557445, EPI_ISL_2557465, EPI_ISL_2557518, EPI_ISL_2557527, EPI_ISL_2557538, EPI_ISL_2557541, EPI_ISL_2557574, EPI_ISL_2557601, EPI_ISL_2557614, EPI_ISL_2557626, EPI_ISL_2652607, EPI_ISL_2653014, EPI_ISL_2653015, EPI_ISL_2653035, EPI_ISL_2653056, EPI_ISL_2653059, EPI_ISL_2686857, EPI_ISL_2686883, EPI_ISL_2686893, EPI_ISL_2786681, EPI_ISL_2786699, EPI_ISL_2876051, EPI_ISL_3020027, EPI_ISL_3111088 | | | |
| see above | Infinity Biologix | Centers for Disease Control and Prevention Division of Viral Diseases, Pathogen Discovery | Adrian Paskey; Benjamin Rambo-Martin; Chirayu Goswami; Christian Bixby; Christopher Gulvick; Clinton R. Paden; Dakota Howard; Darlene Wagner; Dhwani Batra; Duncan MacCannell; Jason Caravas; Jonathan Schultz; Kara Moser; Matthew Schmerer; Peter W. Cook; Robin Grimwood; Russ Hager; Scott Sammons; Shatavia Morrison; Yihe Wang; Yvette Unoarumhi |
| EPI_ISL_3031143 | Innlandet Hospital Trust, Division Lillehammer, Department for Medical Microbiology | Norwegian Institute of Public Health, Department of Virology | Atiya R Ali; Debech Nadia; Engebretsen Serina Beate; Garcia Llorente Ignacio; Hilde Elshaug; Hilde Vollan; Jon Bråte; Kamilla Heddeland Instefjord; Karoline Bragstad; Kathrine Stene-Johansen; Line Victoria Moen; Marie Paulsen Madsen; Olav Hungnes; Pedersen Benedikte Nevjen; Rasmus Riis Kopperud |
| EPI_ISL_2761729, EPI_ISL_2761732, EPI_ISL_2956071, EPI_ISL_2956076 | Institut für Labormedizin Mikrobiologie und Hygiene | Robert Koch Institute | |
| EPI_ISL_3426469, EPI_ISL_3453310, EPI_ISL_3504024 | Institute of Microbiology and Immunology, Faculty of Medicine, University of Ljubljana | Institute of Microbiology and Immunology, Faculty of Medicine, University of Ljubljana | Alen Suljič; Andraž Celar; Dominika Šturm; Doroteja Vlaj; Mario Poljak; Matic Brvar; Miša Korva; Patricija Pozvek; Samo Zakotnik; Tatjana Avšič – Županc; Tomaž Mark Zorec; Špela Pleh |
| EPI_ISL_2492351 | Institute of Microbiology, Universidad San Francisco de Quito | Institute of Microbiology, Universidad San Francisco de Quito | Belén Prado-Vivar; Bernardo Gutiérrez; Fernanda Zurita; Gabriel Trueba; Juan José Guadalupe; Lofrein Vera; Michelle Grunauer; Monica Becerra-Wong; Patricio Rojas-Silva; Paúl Cárdenas; Sully Márquez; Verónica Barragán |
| EPI_ISL_2919255, EPI_ISL_3316158, EPI_ISL_3316159, EPI_ISL_3316181, EPI_ISL_3316193, EPI_ISL_3316202, EPI_ISL_3316225 | | | |
| see above | Instituto Adolfo Lutz - Regional de Bauru | Instituto Adolfo Lutz, Interdiciplinary Procedures Center, Strategic Laboratory | Caio Vinicius Dias Lopes; Claudia Regina Gonçalves; Claudio Tavares Sacchi; Erica Valessa Ramos Gomes; Karoline Rodrigues Campos; Leonardo Tadeu de Araujo; Marlon Benedito Nascomento Santos |
| EPI_ISL_2828684, EPI_ISL_2828685, EPI_ISL_2919263, EPI_ISL_3259692, EPI_ISL_3259694, EPI_ISL_3259696, EPI_ISL_3259697, EPI_ISL_3259698, EPI_ISL_3259699, EPI_ISL_3259700, EPI_ISL_3259701, EPI_ISL_3259702, EPI_ISL_3259705, EPI_ISL_3259706, EPI_ISL_3259708, EPI_ISL_3259709, EPI_ISL_3259710, EPI_ISL_3259712, EPI_ISL_3259713, EPI_ISL_3259716, EPI_ISL_3259717, EPI_ISL_3545799 | | | |
| see above | Instituto Adolfo Lutz - Regional de Campinas | Instituto Adolfo Lutz, Interdiciplinary Procedures Center, Strategic Laboratory | Caio Vinicius Dias Lopes; Claudia Regina Gonçalves; Claudio Tavares Sacchi; Erica Valessa Ramos Gomes; Karoline Rodrigues Campos; Leonardo Tadeu de Araujo; Marlon Benedito Nascomento Santos |
| EPI_ISL_3545788, EPI_ISL_3545789, EPI_ISL_3545796, EPI_ISL_3545800, EPI_ISL_3545801, EPI_ISL_3545802, EPI_ISL_3717840, EPI_ISL_3717841, EPI_ISL_3717842, EPI_ISL_3717843, EPI_ISL_3717844, EPI_ISL_3717845, EPI_ISL_3717846, EPI_ISL_3717847, EPI_ISL_3717848, EPI_ISL_3717849, EPI_ISL_3717850, EPI_ISL_3717851, EPI_ISL_3717853, EPI_ISL_3717854, EPI_ISL_3717855, EPI_ISL_3717857, EPI_ISL_3717858, EPI_ISL_3717859, EPI_ISL_3717860, EPI_ISL_3717861, EPI_ISL_3717864, EPI_ISL_3717866, EPI_ISL_3717868, EPI_ISL_3717869, EPI_ISL_3717870, EPI_ISL_3717871, EPI_ISL_3717872, EPI_ISL_3717873, EPI_ISL_3717874, EPI_ISL_3717875, EPI_ISL_3717887, EPI_ISL_3717889, EPI_ISL_3946613, EPI_ISL_3946614 | | | |
| see above | Instituto Adolfo Lutz - Regional de Marilia | Instituto Adolfo Lutz, Interdiciplinary Procedures Center, Strategic Laboratory | Caio Vinicius Dias Lopes; Claudia Regina Gonçalves; Claudio Tavares Sacchi; Karoline Rodrigues Campos; Leonardo Tadeu de Araujo; Marlon Benedito Nascimento Santos; Marlon Benedito Nascomento Santos |
| EPI_ISL_2614617, | Instituto Adolfo Lutz - Regional de | Instituto Adolfo Lutz, | Caio Vinicius Dias Lopes; Claudia Regina Gonçalves; Claudio Tavares Sacchi; Erica Valessa Ramos Gomes; Karoline Rodrigues Campos; Leonardo Jose Tadeu de Araujo; Leonardo Tadeu de Araujo; Marlon Benedito Nascomento Santos |

| | | | |
|---|---|---|---|
| EPI_ISL_3671894 | Presidente Prudente | Interdiciplinary Procedures Center, Strategic Laboratory | |
| EPI_ISL_3671866, EPI_ISL_3671867, EPI_ISL_3671868, EPI_ISL_3671869, EPI_ISL_3671870, EPI_ISL_3671872, EPI_ISL_3671873, EPI_ISL_3671874, EPI_ISL_3671875, EPI_ISL_3671876 | | | |
| see above | Instituto Adolfo Lutz - Regional de Presidente Prudente | Instituto Adolfo Lutz, Interdisciplinary Procedures Center, Strategic Laboratory | Caio Vinicius Dias Lopes; Claudia Regina Gonçalves; Claudio Tavares Sacchi; Karoline Rodrigues Campos; Leonardo Tadeu de Araujo; Marlon Benedito Nascomento Santos |
| EPI_ISL_3316164, EPI_ISL_3316167, EPI_ISL_3316171, EPI_ISL_3316178, EPI_ISL_3316183, EPI_ISL_3316192, EPI_ISL_3316240, EPI_ISL_3316243, EPI_ISL_3316301, EPI_ISL_3316307, EPI_ISL_3545793 | | | |
| see above | Instituto Adolfo Lutz - Regional de Ribeirao Preto | Instituto Adolfo Lutz, Interdisciplinary Procedures Center, Strategic Laboratory | Caio Vinicius Dias Lopes; Claudia Regina Gonçalves; Claudio Tavares Sacchi; Karoline Rodrigues Campos; Leonardo Tadeu de Araujo; Marlon Benedito Nascomento Santos |
| EPI_ISL_3671877 | Instituto Adolfo Lutz - Regional de Ribeirao Preto | Instituto Adolfo Lutz, Interdisciplinary Procedures Center, Strategic Laboratory | Caio Vinicius Dias Lopes; Claudia Regina Gonçalves; Claudio Tavares Sacchi; Karoline Rodrigues Campos; Leonardo Tadeu de Araujo; Marlon Benedito Nascomento Santos |
| EPI_ISL_3316293, EPI_ISL_3316294, EPI_ISL_3316295, EPI_ISL_3316296, EPI_ISL_3316298, EPI_ISL_3316299 | Instituto Adolfo Lutz - Regional de Rio Claro | Instituto Adolfo Lutz, Interdiciplinary Procedures Center, Strategic Laboratory | Caio Vinicius Dias Lopes; Claudia Regina Gonçalves; Claudio Tavares Sacchi; Karoline Rodrigues Campos; Leonardo Tadeu de Araujo; Marlon Benedito Nascomento Santos |
| EPI_ISL_2828669, EPI_ISL_2828670, EPI_ISL_2828671, EPI_ISL_2828672, EPI_ISL_2828673, EPI_ISL_2828674, EPI_ISL_2828675, EPI_ISL_2828677, EPI_ISL_2828678, EPI_ISL_2828679, EPI_ISL_2828680, EPI_ISL_2828681, EPI_ISL_2828682, EPI_ISL_3717876, EPI_ISL_3717877, EPI_ISL_3717879, EPI_ISL_3717880, EPI_ISL_3717882, EPI_ISL_3717883, EPI_ISL_3717884, EPI_ISL_3717886, EPI_ISL_3717888, EPI_ISL_3946615, EPI_ISL_4056259, EPI_ISL_4056260, EPI_ISL_4056262, EPI_ISL_4056263, EPI_ISL_4056264, EPI_ISL_4056267, EPI_ISL_4056268, EPI_ISL_4056269, EPI_ISL_4056270, EPI_ISL_4056271, EPI_ISL_4056272, EPI_ISL_4056273, EPI_ISL_4056275, EPI_ISL_4056276, EPI_ISL_4056277, EPI_ISL_4056278, EPI_ISL_4056279, EPI_ISL_4056280, EPI_ISL_4056281, EPI_ISL_4056282, EPI_ISL_4056283, EPI_ISL_4056284, EPI_ISL_4056285, EPI_ISL_4056286, EPI_ISL_4056287, EPI_ISL_4056289, EPI_ISL_4056290, EPI_ISL_4056291, EPI_ISL_4056292, EPI_ISL_4056294, EPI_ISL_4056295 | | | |
| see above | Instituto Adolfo Lutz - Regional de Santo Andre | Instituto Adolfo Lutz, Interdiciplinary Procedures Center, Strategic Laboratory | Caio Vinicius Dias Lopes; Claudia Regina Gonçalves; Claudio Tavares Sacchi; Erica Valessa Ramos Gomes; Karoline Rodrigues Campos; Leonardo Tadeu de Araujo; Marlon Benedito Nascimento Santos |
| EPI_ISL_3671878 | Instituto Adolfo Lutz - Regional de Santo Andre | Instituto Adolfo Lutz, Interdisciplinary Procedures Center, Strategic Laboratory | Caio Vinicius Dias Lopes; Claudia Regina Gonçalves; Claudio Tavares Sacchi; Karoline Rodrigues Campos; Leonardo Tadeu de Araujo; Marlon Benedito Nascomento Santos |
| EPI_ISL_3671879, EPI_ISL_3671880, EPI_ISL_3671881, EPI_ISL_3671882, EPI_ISL_3671883, EPI_ISL_3671884, EPI_ISL_3671886, EPI_ISL_3671887, EPI_ISL_3671888, EPI_ISL_3671890, EPI_ISL_3671891 | | | |
| see above | Instituto Adolfo Lutz - Regional de Santos | Instituto Adolfo Lutz, Interdisciplinary Procedures Center, Strategic Laboratory | Caio Vinicius Dias Lopes; Claudia Regina Gonçalves; Claudio Tavares Sacchi; Karoline Rodrigues Campos; Leonardo Tadeu de Araujo; Marlon Benedito Nascomento Santos |
| EPI_ISL_2919265, EPI_ISL_2919266, EPI_ISL_2919267, EPI_ISL_3259720, EPI_ISL_3259721, EPI_ISL_3259722, EPI_ISL_3259723, EPI_ISL_3259724, EPI_ISL_3259725, EPI_ISL_3259726, EPI_ISL_3259728, EPI_ISL_3316244 | | | |
| see above | Instituto Adolfo Lutz - Regional de Sao Jose do Rio Preto | Instituto Adolfo Lutz, Interdisciplinary Procedures Center, Strategic Laboratory | Caio Vinicius Dias Lopes; Claudia Regina Gonçalves; Claudio Tavares Sacchi; Erica Valessa Ramos Gomes; Karoline Rodrigues Campos; Leonardo Tadeu de Araujo; Marlon Benedito Nascomento Santos |
| EPI_ISL_3671892 | Instituto Adolfo Lutz - Regional de Sao Jose do Rio Preto | Instituto Adolfo Lutz, Interdisciplinary Procedures Center, Strategic Laboratory | Caio Vinicius Dias Lopes; Claudia Regina Gonçalves; Claudio Tavares Sacchi; Karoline Rodrigues Campos; Leonardo Tadeu de Araujo; Marlon Benedito Nascomento Santos |
| EPI_ISL_3259678, EPI_ISL_3259680, EPI_ISL_3259682, EPI_ISL_3259683, EPI_ISL_3259685, EPI_ISL_3259686, EPI_ISL_3259688, EPI_ISL_3259690, EPI_ISL_3316161, EPI_ISL_3316165, EPI_ISL_3316184, EPI_ISL_3316201, EPI_ISL_3316237, EPI_ISL_3545790, EPI_ISL_3545791, EPI_ISL_3545797, EPI_ISL_3946602, EPI_ISL_3946603, EPI_ISL_3946605, EPI_ISL_3946607, EPI_ISL_3946611, EPI_ISL_3946612 | | | |
| see above | Instituto Adolfo Lutz - Regional de Sorocaba | Instituto Adolfo Lutz, Interdisciplinary Procedures Center, Strategic Laboratory | Caio Vinicius Dias Lopes; Claudia Regina Gonçalves; Claudio Tavares Sacchi; Erica Valessa Ramos Gomes; Karoline Rodrigues Campos; Leonardo Tadeu de Araujo; Marlon Benedito Nascomento Santos |
| EPI_ISL_2919233, EPI_ISL_2919234, EPI_ISL_2919268, EPI_ISL_2919269, EPI_ISL_2919270, EPI_ISL_2919271, EPI_ISL_2919272, EPI_ISL_2919273, EPI_ISL_2919274, EPI_ISL_2919275, EPI_ISL_2919278, EPI_ISL_2919280, EPI_ISL_2919281, EPI_ISL_2919285, EPI_ISL_2919286, EPI_ISL_2919287, EPI_ISL_2919288, EPI_ISL_2919289, EPI_ISL_2919290, EPI_ISL_2919291, EPI_ISL_2919292, EPI_ISL_2919294, EPI_ISL_3259730, EPI_ISL_3259731, EPI_ISL_3259732, EPI_ISL_3259733, EPI_ISL_3259734, EPI_ISL_3259735, EPI_ISL_3259736, EPI_ISL_3259738, EPI_ISL_3259739, EPI_ISL_3259740, EPI_ISL_3259741, EPI_ISL_3259742, EPI_ISL_3259743, EPI_ISL_3259744, EPI_ISL_3259745, EPI_ISL_3259746, EPI_ISL_3259747, EPI_ISL_3259748, EPI_ISL_3259750, EPI_ISL_3259751, EPI_ISL_3259752, EPI_ISL_3259754, EPI_ISL_3259755, EPI_ISL_3259756, EPI_ISL_3259757, EPI_ISL_3259759, EPI_ISL_3259760, EPI_ISL_3259762, EPI_ISL_3259763, EPI_ISL_3259764, EPI_ISL_3316245, EPI_ISL_3316246, EPI_ISL_3316247, EPI_ISL_3316248, EPI_ISL_3316250, EPI_ISL_3316251, EPI_ISL_3316252, EPI_ISL_3316253, EPI_ISL_3316254, EPI_ISL_3316255, EPI_ISL_3316258, EPI_ISL_3316259, EPI_ISL_3316260, EPI_ISL_3316261, EPI_ISL_3316262, EPI_ISL_3316266, EPI_ISL_3316267, EPI_ISL_3316268, EPI_ISL_3316269, EPI_ISL_3316271, EPI_ISL_3316273, EPI_ISL_3316274, EPI_ISL_3316277, EPI_ISL_3316278, EPI_ISL_3316279, EPI_ISL_3316280, EPI_ISL_3316281, EPI_ISL_3316282, EPI_ISL_3316283, EPI_ISL_3316287, EPI_ISL_3316288, EPI_ISL_3316289, EPI_ISL_3316290, EPI_ISL_3316291, EPI_ISL_3316292, EPI_ISL_3545792, EPI_ISL_3545798, EPI_ISL_3545803, EPI_ISL_3671895, EPI_ISL_3671896, EPI_ISL_3671897, EPI_ISL_3671898, EPI_ISL_3671899, EPI_ISL_3671900, EPI_ISL_3671904, EPI_ISL_3671905, EPI_ISL_3691691, EPI_ISL_3691692, EPI_ISL_3691693, EPI_ISL_3691700, EPI_ISL_3710516, EPI_ISL_3717835, EPI_ISL_3717836, EPI_ISL_3717838 | | | |
| see above | Instituto Adolfo Lutz Central | Instituto Adolfo Lutz, Interdisciplinary Procedures Center, Strategic Laboratory | Caio Vinicius Dias Lopes; Claudia Regina Gonçalves; Claudio Tavares Sacchi; Erica Valessa Ramos Gomes; Karoline Rodrigues Campos; Leonardo Tadeu de Araujo; Marlon Benedito Nascimento Santos; Marlon Benedito Nascomento Santos |
| EPI_ISL_3671893 | Instituto Adolfo Lutz Central | Instituto Adolfo Lutz, Interdisciplinary Procedures Center, Strategic Laboratory | Caio Vinicius Dias Lopes; Claudia Regina Gonçalves; Claudio Tavares Sacchi; Karoline Rodrigues Campos; Leonardo Tadeu de Araujo; Marlon Benedito Nascomento Santos |
| EPI_ISL_2924058 | Instituto Adolfo Lutz Central | Instituto Adolfo Lutz, Rapid Response Center, Strategic Laboratory | Caio Vinicius Dias Lopes; Claudia Regina Gonçalves; Claudio Tavares Sacchi; Erica Valessa Ramos Gomes; Karoline Rodrigues Campos; Leonardo Jose Tadeu de Araujo |
| EPI_ISL_2777215, EPI_ISL_2777223 | Instituto Nacional de Cancerología | Centro de Investigación en Enfermedades Infecciosas (CIENI), Instituto Nacional de Enfermedades Respiratorias (INER) | Alejandra Garcia-Gasca; Alejandra Hernández-Terán; Alejandro Sanchez-Flores; Alfredo Herrera-Estrella; Alicia Ocaña-Mondragón; Andreu Comas-García; Angel Gustavo Salas-Lais; Antonio Loza Román; Bernardo Martínez-Miguel; Blanca Taboada; Brenda Irasema Maldonado-Meza; Bruno Gomez-Gil; Carla Ivón Herrera-Najera; Carlos F. Arias; Celia Boukadida; Clara Esperanza Santacruz-Tinoco; Concepción Grajales-Muñiz; Consorcio Mexicano de Vigilancia Genómica (CoViGen-Mex). Authors (in alphabetical order): Julio Elias Alvarado-Yaah; Cristóbal Cháidez-Quiróz; Célida Duque Molina; Célida Martínez-Rodríguez; Daniel Fregoso-Rueda; Daniel Lira Morales; Eduardo Becerril-Vargas; Fernando Fontove-Herrera; Fidencio Mejía-Nepomuceno; Francisco Pulido; Gloria Elena Espinosa-Ayala; Gloria María Molina-Salinas; Gloria Vazquez; Hector Esteban Paz-Juárez; Hector Montoya-Fuentes; Helen Haydee Fernanda Ramírez-Plascencia; Irvin González-López; Jean Pierre González; Joel Armando Vázquez-Pérez.; Jorge Salas-Hernández; José Antonio Enciso-Moreno; José Arturo Martínez-Orozco; José Esteban Muñoz-Medina; José de Jesús Nuñez-Contreras; Juan Bautista Chale-Dzul; Julissa Enciso-Ibarra; Kathia Elizabeth Tapia-Diaz; Luis Alberto Ochoa-Carrera; Margarita Matías-Florentino; Mario Mújica-Sánchez; Marissa Pérez-García; María Guadalupe Santiago-Mauricio; María Guadalupe de Jesús Mireles-Rivera; Nelly Sélem-Mojica; Pavel Isa; Ricardo Ciria Merce; Ricardo Grande; Rosa María Gutierrez Rios; Santiago Ávila-Ríos; Selene Zárate; Susana Lopez; Victor Eduardo Garcia-Arias; Víctor Hugo Borja-Aburto |
| EPI_ISL_2648116, EPI_ISL_2648117, EPI_ISL_2777212, EPI_ISL_3268374, EPI_ISL_3268376, EPI_ISL_3268377 | Instituto Nacional de Enfermedades Respiratorias (INER) | Centro de Investigación en Enfermedades Infecciosas (CIENI), Instituto Nacional de Enfermedades Respiratorias (INER) | ; Alejandra Garcia-Gasca; Alejandra Hernández-Terán; Alejandro Sanchez-Flores; Alfredo Herrera-Estrella; Alicia Ocaña-Mondragón; Andreu Comas-García; Angel Gustavo Salas-Lais; Antonio Loza Román; Bernardo Martínez-Miguel; Blanca Taboada; Brenda Irasema Maldonado-Meza; Bruno Gomez-Gil; Carla Ivón Herrera-Najera; Carlos F. Arias; Celia Boukadida; Clara Esperanza Santacruz-Tinoco; Concepción Grajales-Muñiz; Consorcio Mexicano de Vigilancia Genómica (CoViGen-Mex). Authors (in alphabetical order): Julio Elias Alvarado-Yaah; Cristóbal Cháidez-Quiróz; Célida Duque Molina; Célida Martínez- Rodríguez; Daniel Fregoso-Rueda; Daniel Lira Morales; Eduardo Becerril-Vargas; Fernando Fontove-Herrera; Fidencio Mejía-Nepomuceno; Francisco Pulido; Gloria Elena Espinosa-Ayala; Gloria María Molina-Salinas; Gloria Vazquez; Hector Esteban Paz-Juárez; Hector Montoya-Fuentes; Helen Haydee Fernanda Ramírez-Plascencia; Irvin González-López; Jean Pierre González; Joel Armando Vázquez-Pérez.; Jorge Salas-Hernández; José Antonio Enciso-Moreno; José Arturo Martínez-Orozco; José Esteban Muñoz-Medina; José de Jesús Nuñez-Contreras; Juan Bautista Chale-Dzul; Julissa Enciso-Ibarra; Kathia Elizabeth Tapia-Diaz; Luis Alberto Ochoa-Carrera; Margarita Matías-Florentino; Mario Mújica-Sánchez; Marissa Pérez-García; María Guadalupe Santiago-Mauricio; María Guadalupe de Jesús Mireles-Rivera; Nelly Sélem-Mojica; Pavel Isa; Ricardo Ciria Merce; Ricardo Grande; Rosa María Gutierrez Rios; Santiago Ávila-Ríos; Selene Zárate; Susana Lopez; Victor Eduardo Garcia-Arias; Víctor Hugo Borja-Aburto |
| EPI_ISL_2810063, EPI_ISL_2894311 | Instituto Nacional de Medicina Genomica | Instituto Nacional de Medicina Genomica | Cedro-Tanda A; Cisneros- Villanueva M; Escobar-Arrazola; Gonzalez-Barrera D; Herrera-Montalvo LA.; Hidalgo-Miranda A; M; Mendoza-Vargas A; Munguia-Garza P; Ramirez-Vega O; Rangel-DeLeon D; Reyes-Grajeda JP |
| EPI_ISL_2843904, EPI_ISL_2843907, EPI_ISL_2843909, EPI_ISL_2843910, EPI_ISL_2843911, EPI_ISL_2843916, EPI_ISL_2843917, EPI_ISL_2843919, EPI_ISL_2843921, EPI_ISL_2843923, EPI_ISL_2843927, EPI_ISL_2843930, EPI_ISL_2843931, EPI_ISL_2843934, EPI_ISL_2843937, EPI_ISL_2843938, EPI_ISL_2843943, EPI_ISL_2843945, EPI_ISL_2843946, EPI_ISL_2843948, EPI_ISL_2843951, EPI_ISL_2843955, EPI_ISL_2843956, EPI_ISL_2843957, EPI_ISL_2843958, EPI_ISL_2843959, EPI_ISL_2843962, EPI_ISL_2843963, EPI_ISL_2843964, EPI_ISL_2843968, EPI_ISL_2843969, EPI_ISL_2843971, EPI_ISL_2843972, EPI_ISL_2843974, EPI_ISL_2843977, EPI_ISL_2843978, EPI_ISL_2843979, EPI_ISL_2843981, EPI_ISL_2843983, EPI_ISL_2843984, EPI_ISL_2843985, EPI_ISL_2843987, EPI_ISL_2843988, EPI_ISL_2843989, EPI_ISL_2843991, EPI_ISL_2843992, EPI_ISL_2843993, EPI_ISL_2843994, EPI_ISL_2843996, EPI_ISL_2843997, EPI_ISL_2843998, EPI_ISL_2843999, EPI_ISL_2844002, EPI_ISL_2844003, EPI_ISL_2844004, EPI_ISL_2844005, EPI_ISL_2844006, EPI_ISL_2844007, EPI_ISL_2844008, EPI_ISL_2844009, EPI_ISL_2844010, EPI_ISL_2844011, EPI_ISL_2844012, EPI_ISL_2844014, EPI_ISL_2844016, EPI_ISL_2844017, EPI_ISL_2844018, EPI_ISL_2844019, EPI_ISL_2844020, EPI_ISL_2844022, EPI_ISL_2844024, EPI_ISL_2844025, EPI_ISL_2844026, EPI_ISL_2844027, EPI_ISL_2844030, EPI_ISL_2844032, EPI_ISL_2844035, EPI_ISL_2844036, EPI_ISL_2844037, EPI_ISL_2844040, EPI_ISL_2844041, EPI_ISL_2844043, EPI_ISL_2844045, EPI_ISL_2844047, EPI_ISL_2844048, EPI_ISL_2844050, EPI_ISL_2844051, EPI_ISL_2844053, EPI_ISL_2844057, EPI_ISL_2844058, EPI_ISL_2844061, EPI_ISL_2844063, EPI_ISL_2844065, EPI_ISL_2844067, EPI_ISL_2844072, EPI_ISL_2844073, EPI_ISL_2844077, EPI_ISL_2844078, EPI_ISL_2844081, EPI_ISL_2844084, EPI_ISL_2844086, EPI_ISL_2844087, EPI_ISL_2844090, EPI_ISL_2844091, EPI_ISL_2844097, EPI_ISL_2844099, EPI_ISL_2844103, EPI_ISL_2844105, EPI_ISL_2844106, EPI_ISL_2844107, EPI_ISL_2844108, EPI_ISL_2844111, EPI_ISL_2844112, EPI_ISL_2844113, EPI_ISL_2844114, EPI_ISL_2844115, EPI_ISL_2844117, EPI_ISL_2844118, EPI_ISL_2844119, EPI_ISL_2844126, EPI_ISL_2844127, EPI_ISL_2844128, EPI_ISL_2844133, EPI_ISL_2844137, EPI_ISL_2844140, EPI_ISL_2844142, EPI_ISL_2844144, EPI_ISL_2844147, EPI_ISL_2844148, EPI_ISL_2844149, EPI_ISL_2844150, EPI_ISL_2844155, EPI_ISL_2844156, EPI_ISL_2844161, EPI_ISL_2844162, EPI_ISL_2844165, EPI_ISL_2844170, EPI_ISL_2844172, EPI_ISL_2844179, EPI_ISL_2844181, EPI_ISL_2844183, EPI_ISL_2844188, EPI_ISL_2844189, EPI_ISL_2844190, EPI_ISL_2844191, EPI_ISL_2844193, EPI_ISL_2844195, EPI_ISL_3030615, EPI_ISL_3118852, EPI_ISL_3118857, EPI_ISL_3118859, EPI_ISL_3118860, EPI_ISL_3118865, EPI_ISL_3118868, EPI_ISL_3118875, EPI_ISL_3118878, EPI_ISL_3118879, EPI_ISL_3118883, EPI_ISL_3118884, EPI_ISL_3118886, EPI_ISL_3118894, EPI_ISL_3118899, EPI_ISL_3118901, EPI_ISL_3118902, EPI_ISL_3118908, EPI_ISL_3118909, EPI_ISL_3118910, EPI_ISL_3118914, EPI_ISL_3118916, EPI_ISL_3118923, EPI_ISL_3118924, EPI_ISL_3118925, EPI_ISL_3118928, EPI_ISL_3118932, EPI_ISL_3118936, EPI_ISL_3118938, EPI_ISL_3118939, EPI_ISL_3118941, EPI_ISL_3118943, EPI_ISL_3118945, EPI_ISL_3244532, EPI_ISL_3244533, EPI_ISL_3244534, EPI_ISL_3244536, EPI_ISL_3244537, EPI_ISL_3244539, EPI_ISL_3244540, EPI_ISL_3244541, EPI_ISL_3244542, EPI_ISL_3244544, EPI_ISL_3244552, EPI_ISL_3244555, EPI_ISL_3244557, EPI_ISL_3244561, EPI_ISL_3244562, EPI_ISL_3244566, EPI_ISL_3244572, EPI_ISL_3244573, EPI_ISL_3244574, EPI_ISL_3244575, EPI_ISL_3244579, EPI_ISL_3244584, EPI_ISL_3244588, EPI_ISL_3244592, EPI_ISL_3244594 | | | |
| see above | Instituto de Biotecnologia - UNESP-Botucatu-SP | Instituto de Biotecnologia - UNESP-Botucatu-SP | Cecília Artico Banho; Cíntia Bittar; Fábio Sossai Possebon; Guilherme Campos; Helena Lage Ferreira; Jorge A. Petroli Marchesi; João Pessoa Araújo Jr.; Leila Sabrina Ullmann; Lívia Sacchetto; Maisa C. Pereira Parra; Marília Moraes; Maurício L. Nogueira; Paula Rahal; Paulo Inacio da Costa |
| EPI_ISL_3236442, EPI_ISL_3236443 | Instituto de Medicina Tropical & Salud Global Universidad Iberoamericana | Grubaugh Lab - Yale School of Public Health | Alejandaro Vallejo Degaudenzi; Anderson Brito; Annie Watkins; Chaney Kalinich; Chantal Vogels; Elisa Contreras; Esperanza Mendoza; Isabel Ott; Jessica Rothman; Joseph Fauver; Kendall Billig; Mallery Breban; Mary Petrone; Nathan Grubaugh; Robert Paulino-Ramirez; Tara Alpert; Tobias Koch; Victor Virgilio Calderon |
| EPI_ISL_3020244, EPI_ISL_3020245 | Instituto de Medicina Tropical Alexander Von Humboldt, Sede Cusco | Laboratorio de Genómica Microbiana, Universidad Peruana Cayetano Heredia | Alejandra Dávila-Barclay; Diego Cuicapuza; Guillermo Salvatierra; Janet Huancachoque; Luis González; Pablo Tsukayama; Pedro E. Romero; Pool Marcos |
| EPI_ISL_3105054, EPI_ISL_3347068, EPI_ISL_3347084, EPI_ISL_3347092 | Interpath Laboratory | Seattle Flu Study | Amanda Adler; Barry R. Lutz; Benjamin Pelle; Brian Hiatt; Caitlin R. Wolf; Chris D. Frazar; Deborah A. Nickerson; Elisabeth Brandstetter; Erica Ryke; Geoff Melly; Helen Y. Chu; Janet A. Englund; Jay Shendure; Jover Lee; Kairsten Fay; Kirsten Lacombe; Lea M. Starita; Mark J. Rieder; Matthew Richardson; Matthew Thompson; Melissa Truong; Michael Boeckh; Michael Famulare; Misja Ilcisin; Peter D. Han; Philip Dykema; Romesh Gautom; Scott Lindquist; Thomas R. Sibley; Trevor Bedford |
| EPI_ISL_3230430 | Islab, Etelä-Savon aluelaboratorio | Expert Microbiology, National Institute for Health and Welfare | Carita Savolainen-Kopra; Erika Lindh; Haider al-Hello; Jani Halkilahti; Kirsi Liitsola; Niina Ikonen; Olli Vapalahti; Pekka Ellonen; Phuoc Truong; Päivi Laurila; Ravi Kant; Sari Hannula; Soile Blomqvist; Teemu Smura |
| EPI_ISL_2645182 | Islab, Pohjois-Karjalan aluelaboratorio | Expert Microbiology, National Institute for Health and Welfare | Carita Savolainen-Kopra; Erika Lindh; Haider al-Hello; Jani Halkilahti; Kirsi Liitsola; Niina Ikonen; Olli Vapalahti; Pekka Ellonen; Phuoc Truong; Päivi Laurila; Ravi Kant; Sari Hannula; Soile Blomqvist; Teemu Smura |
| EPI_ISL_3230614, | Islab, Pohjois-Savon aluelaboratorio | Expert Microbiology, National | Carita Savolainen-Kopra; Erika Lindh; Haider al-Hello; Jani Halkilahti; Kirsi Liitsola; Niina Ikonen; Olli Vapalahti; Pekka Ellonen; Phuoc Truong; Päivi Laurila; Ravi Kant; Sari Hannula; Soile Blomqvist; Teemu Smura |

| | | | |
|---|---|---|---|
| EPI_ISL_3230615 | | Institute for Health and Welfare | |
| EPI_ISL_3279359 | Israel Central Virology laboratory | Israel National Consortium for SARS-CoV-2 sequencing | Dana Bar-Ilan; Danit Sofer; Efrat Dahan Bucris; Efrat Glick-Saar; Ella Mendelson; Gideon Rechavi; Michal Mandelboim; Miranda Geva; Neta Zuckerman; Omer Asraf; Omri Nayshool; Oran Erster; Orna Mor |
| EPI_ISL_2692081, EPI_ISL_2692084 | John H Stroger Jr Hospital of Cook County | RIPHL at Rush University Medical Center | Ah-Young Sung; Felix Araujo Perez; Kevin Kunstman; Laura Furtado; Marieta Hyde; Max Kolton; Sharon Welbel; Stefan Green |
| EPI_ISL_2690054 | KS Health and Environmental Laboratories | Centers for Disease Control and Prevention Division of Viral Diseases, Pathogen Discovery | Alison Laufer Halpin; Ben L. Rambo-Martin; Clinton R. Paden; Dakota Howard; Darlene Wagner; Dave Wentworth; Dhwani Batra; Jasmine Padilla; Justin Lee; Katie Dillon; Krista Queen; Kristen Knipe; Kristine Lacek; Mark Burroughs; Matthew Schmerer; Mili Sheth; Peter Cook; Sam Shepard; Sarah Nobles; Shoshona Le; Suxiang Tong; Vivien Dugan; Yvette Unoarumhi |
| EPI_ISL_3561409, EPI_ISL_3561424, EPI_ISL_3561441, EPI_ISL_3561827, EPI_ISL_3825803, EPI_ISL_4102639, EPI_ISL_4102687 | | | |
| see above | Kaiser Permanente NW Reginal Lab | OHSU MM Lab | Amber Halse; Jeannine Lama; Xuan Qin; Yun Wu |
| EPI_ISL_2443131, EPI_ISL_2455455, EPI_ISL_2558702, EPI_ISL_2833925 | Kansas Health and Environmental Lab | Kansas Health and Environmental Lab | Ben Olsen; Jonathan Barnell; Katherine Wiggins; Mike Grose; and Phil Adam |
| EPI_ISL_2898394, EPI_ISL_2899349 | Karolinska University Hospital Solna | Karolinska University Hospital | Annelie Bjerkner; Isak Sylvin; Jan Albert; Karolina Ininbergs; Lina Guerra Blomqvist; Lynda Eneh; Martin Ekman; Martina Wahlund; Robert Dyrdak; Sandra Broddesson; Tanja Normark; Tobias Allander; Valtteri Wirta; Zhibing Yun |
| EPI_ISL_4080592 | Kentucky State Public Health Lab | Kentucky State Public Health Lab | Anjayya Tummala; Chitralipi Kattunga; Joshua Tobias; Karim George; Matthew Johnson; Rachel Zinner; Rhonda Lucas; Vaneet Arora; William Grooms |
| EPI_ISL_2483832 | Klinisch Laboratorium ZNA | Klinisch Laboratorium ZNA | Verstrepen et al. |
| EPI_ISL_3130541 | LABORATOIRE CERBALLIANCE PLT VILLON | CNR Virus des Infections Respiratoires - France SUD | Antonin Bal; Bruno Lina; Gregory Destras; Gwendolyne Burfin; Hadrien Regue; Laurence Josset; Martine Valette; Quentin Semanas |
| EPI_ISL_2494141, EPI_ISL_2494142, EPI_ISL_2494143 | LABORATORIO DE FRANCA | Instituto Butantan | Antonio Jorge Martins; Claudia Renata dos Santos Barros; David Schlesinger; Debora Botequio Moretti; Dimas Tadeu Covas; Elaine Cristina Marqueze; Elaine Vieira Santos; Evandra Strazza Rodrigues; Heidge Fukumasu; Jayme Augusto de Souza-Neto; José Salvatore Leister Patané; Luiz Alcantara; Luiz Lehmann Coutinho; Maria Carolina Elias; Maurício Lacerda Nogueira; Rafael dos Santos Bezerra; Raul Machado Neto; Rejane Maria Tommasini Grotto; Ricardo Haddad; Sandra Coccuzzo Sampaio Vessoni; Simone Kashima; Svetoslav Nanev Slavov; Vincent Louis Viala |
| EPI_ISL_2494166, EPI_ISL_2494176, EPI_ISL_2494184 | LABORATORIO MUNICIPAL DE ANALISES CLINICAS DE RIO CLARO | Instituto Butantan | Antonio Jorge Martins; Claudia Renata dos Santos Barros; David Schlesinger; Debora Botequio Moretti; Dimas Tadeu Covas; Elaine Cristina Marqueze; Elaine Vieira Santos; Evandra Strazza Rodrigues; Heidge Fukumasu; Jayme Augusto de Souza-Neto; José Salvatore Leister Patané; Luiz Alcantara; Luiz Lehmann Coutinho; Maria Carolina Elias; Maurício Lacerda Nogueira; Rafael dos Santos Bezerra; Raul Machado Neto; Rejane Maria Tommasini Grotto; Ricardo Haddad; Sandra Coccuzzo Sampaio Vessoni; Simone Kashima; Svetoslav Nanev Slavov; Vincent Louis Viala |
| EPI_ISL_2494163, EPI_ISL_2494182, EPI_ISL_2494200, EPI_ISL_2494338 | LABORATORIO MUNICIPAL DE PIRACICABA | Instituto Butantan | Antonio Jorge Martins; Claudia Renata dos Santos Barros; David Schlesinger; Debora Botequio Moretti; Dimas Tadeu Covas; Elaine Cristina Marqueze; Elaine Vieira Santos; Evandra Strazza Rodrigues; Heidge Fukumasu; Jayme Augusto de Souza-Neto; José Salvatore Leister Patané; Luiz Alcantara; Luiz Lehmann Coutinho; Maria Carolina Elias; Maurício Lacerda Nogueira; Rafael dos Santos Bezerra; Raul Machado Neto; Rejane Maria Tommasini Grotto; Ricardo Haddad; Sandra Coccuzzo Sampaio Vessoni; Simone Kashima; Svetoslav Nanev Slavov; Vincent Louis Viala |
| EPI_ISL_2493881, EPI_ISL_2493884, EPI_ISL_2493885, EPI_ISL_2493886, EPI_ISL_2493887, EPI_ISL_2493906, EPI_ISL_2493908, EPI_ISL_2493948, EPI_ISL_2500987 | | | |
| see above | LABORATORIO MUNICIPAL DE SUZANO | Instituto Butantan | Antonio Jorge Martins; Claudia Renata dos Santos Barros; David Schlesinger; Debora Botequio Moretti; Dimas Tadeu Covas; Elaine Cristina Marqueze; Elaine Vieira Santos; Evandra Strazza Rodrigues; Heidge Fukumasu; Jayme Augusto de Souza-Neto; José Salvatore Leister Patané; Luiz Alcantara; Luiz Lehmann Coutinho; Maria Carolina Elias; Maurício Lacerda Nogueira; Rafael dos Santos Bezerra; Raul Machado Neto; Rejane Maria Tommasini Grotto; Ricardo Haddad; Sandra Coccuzzo Sampaio Vessoni; Simone Kashima; Svetoslav Nanev Slavov; Vincent Louis Viala |
| EPI_ISL_2895184 | LAC UCoimbra | Instituto Nacional de Saude (INSA) | Borges et al |
| EPI_ISL_3404852, EPI_ISL_3404856 | LACEN | LGBio (Laboratorio de Genetica & Biodiversidade) | Alex Honda Bernardes; Amanda Alves de Melo; Aparecido Divino da Cruz; Cintia Pelegrineti Targueta de Azevedo Brito; Daniela de Melo e Silva; Elisangela de Paula Silveira Lacerda; Juliana Santana de Curcio; Luiz Augusto Pereira; Marc Alexandre Duarte Gigonzac; Mariana Pires de Campos Telles; Ramilla dos Santos Braga; Renata de Oliveira Dias; Rhewter Nunes; Thais Cidália Vieira Gigonzac; Thais Guimarães Castro; Thays Millena Alves Pedroso |
| EPI_ISL_3671907, EPI_ISL_3946592, EPI_ISL_3946593, EPI_ISL_3946594 | LACEN do Distrito Federal | Instituto Adolfo Lutz, Interdiciplinary Procedures Center, Strategic Laboratory | Caio Vinicius Dias Lopes; Claudia Regina Gonçalves; Claudio Tavares Sacchi; Karoline Rodrigues Campos; Leonardo Tadeu de Araujo; Marlon Benedito Nascimento Santos |
| EPI_ISL_2894548, EPI_ISL_2896234, EPI_ISL_2896235, EPI_ISL_2896236, EPI_ISL_2896238, EPI_ISL_2896239, EPI_ISL_2896240, EPI_ISL_2896241 | | | |
| see above | LACEN do Estado de Mato Grosso | Instituto Adolfo Lutz, Interdiciplinary Procedures Center, Strategic Laboratory | Caio Vinicius Dias Lopes; Claudia Regina Gonçalves; Claudio Tavares Sacchi; Karoline Rodrigues Campos |
| EPI_ISL_3545804, EPI_ISL_3545805, EPI_ISL_3691360, EPI_ISL_3691362 | LACEN do Estado do Mato Grosso do Sul | Instituto Adolfo Lutz, Interdiciplinary Procedures Center, Strategic Laboratory | Caio Vinicius Dias Lopes; Claudia Regina Gonçalves; Claudio Tavares Sacchi; Karoline Rodrigues Campos; Leonardo Tadeu de Araujo; Marlon Benedito Nascimento Santos |
| EPI_ISL_3835195, EPI_ISL_3835196, EPI_ISL_3835197, EPI_ISL_3835198, EPI_ISL_3835200, EPI_ISL_3835201, EPI_ISL_3835202, EPI_ISL_3835204, EPI_ISL_3835205, EPI_ISL_3835209, EPI_ISL_3835210, EPI_ISL_3835211, EPI_ISL_3835212, EPI_ISL_3835213, EPI_ISL_3835215, EPI_ISL_3835216, EPI_ISL_3835217, EPI_ISL_3835218, EPI_ISL_3835219, EPI_ISL_3835220, EPI_ISL_3835222, EPI_ISL_3835224, EPI_ISL_3835225, EPI_ISL_3835229, EPI_ISL_3835230, EPI_ISL_3835231, EPI_ISL_3835233, EPI_ISL_3835234, EPI_ISL_3835235, EPI_ISL_3835237, EPI_ISL_3835238, EPI_ISL_3835239 | | | |
| see above | LACEN/PB | WallauLab on behalf of Fiocruz COVID-19 Genomic Surveillance Network | Alexandre Freitas da Silva; Cassia Docena; Constância Flávia Junqueira Ayres; Filipe Zimmer Dezordi; Gabriel Luz Wallau; Gustavo Barbosa de Lima; Lais Ceschini Machado; Lilian Caroliny Amorim Silva; Maira Galdino da Rocha Pitta; Marcelo Henrique dos Santos Paiva; Matheus Filgueira Bezerra; Michelly Cristiny Pereira; Rômulo Pessoa e Silva; Sinval Pinto Brandão Filho |
| EPI_ISL_3046268, EPI_ISL_3046270, EPI_ISL_3046272, EPI_ISL_3046273, EPI_ISL_3046275, EPI_ISL_3046277, EPI_ISL_3046278, EPI_ISL_3046280, EPI_ISL_3046281, EPI_ISL_3046282, EPI_ISL_3046283, EPI_ISL_3046284, EPI_ISL_3046285, EPI_ISL_3046286, EPI_ISL_3046287, EPI_ISL_3046288, EPI_ISL_3046290, EPI_ISL_3046291, EPI_ISL_3046292, EPI_ISL_3046293, EPI_ISL_3046294, EPI_ISL_3046295, EPI_ISL_3046296, EPI_ISL_3046297, EPI_ISL_3046299, EPI_ISL_3046300, EPI_ISL_3046301, EPI_ISL_3046305, EPI_ISL_3046306, EPI_ISL_3046307, EPI_ISL_3046308, EPI_ISL_3046309, EPI_ISL_3046313, EPI_ISL_3046314, EPI_ISL_3046315, EPI_ISL_3046316, EPI_ISL_3046317, EPI_ISL_3046318, EPI_ISL_3046320, EPI_ISL_3046322, EPI_ISL_3046323, EPI_ISL_3046325, EPI_ISL_3046326, EPI_ISL_3046332, EPI_ISL_3046338, EPI_ISL_3046339, EPI_ISL_3046344, EPI_ISL_3046345, EPI_ISL_3134665, EPI_ISL_3134667, EPI_ISL_3134669, EPI_ISL_3134672, EPI_ISL_3134676, EPI_ISL_3134680, EPI_ISL_3134681, EPI_ISL_3134682, EPI_ISL_3134683, EPI_ISL_3134684, EPI_ISL_3134685, EPI_ISL_3134687, EPI_ISL_3134688, EPI_ISL_3134689, EPI_ISL_3134690, EPI_ISL_3134691, EPI_ISL_3134692, EPI_ISL_3134693, EPI_ISL_3134695, EPI_ISL_3134696, EPI_ISL_3134698, EPI_ISL_3134701, EPI_ISL_3134702, EPI_ISL_3134703, EPI_ISL_3134704, EPI_ISL_3134705, EPI_ISL_3134706, EPI_ISL_3134707, EPI_ISL_3134708, EPI_ISL_3134709, EPI_ISL_3134710, EPI_ISL_3134715, EPI_ISL_3134716, EPI_ISL_3134717, EPI_ISL_3134718, EPI_ISL_3134719, EPI_ISL_3134720, EPI_ISL_3134721, EPI_ISL_3134834, EPI_ISL_3134858, EPI_ISL_3447469, EPI_ISL_3447470, EPI_ISL_3447472, EPI_ISL_3447475, EPI_ISL_3447478, EPI_ISL_3447479, EPI_ISL_3447487, EPI_ISL_3447488, EPI_ISL_3447600 | | | |
| see above | LACEN/PE | WallauLab on behalf of Fiocruz COVID-19 Genomic Surveillance Network | Alexandre Freitas da Silva; Cassia Docena; Constância Flávia Junqueira Ayres; Filipe Zimmer Dezordi; Gabriel Luz Wallau; Gustavo Barbosa de Lima; Lais Ceschini Machado; Lilian Caroliny Amorim Silva; Marcelo Henrique dos Santos Paiva; Matheus Filgueira Bezerra; Sinval Pinto Brandão Filho |
| EPI_ISL_3149318, EPI_ISL_3149319, EPI_ISL_3149320, EPI_ISL_3149321 | LANIIA | LANIIA-Nayarit | ; Alejandra García-Gasca; Alejandra Hernández-Terán; Alejandro Sánchez-Flores; Alfredo Herrera-Estrella; Alicia Ocaña-Mondragón; Andreu Comas-García; Angel Gustavo Salas-Lais; Antonio Loza Román; Bernardo Martínez-Miguel; Blanca Taboada; Brenda Irasema Maldonado-Meza; Bruno Gómez-Gil; Carla Ivón Herrera-Najera; Carlos Eduardo Covantes-Rosales; Carlos F. Arias; Celia Boukadida; Clara Esperanza Santacruz-Tinoco; Concepción Grajales-Muñiz; Consorcio Mexicano de Vigilancia Genómica (CoViGen-Mex). Authors (in alphabetical order): Julio Elias Alvarado-Yaah; Cristóbal Cháidez-Quiróz; Célida Duque Molina; Célida Martínez- Rodríguez; Daniel Alberto Girón Pérez; Daniel Fregoso-Rueda; Daniel Lira Morales; Eduardo Becerril-Vargas; Fernando Fontove-Herrera; Fidencio Mejía-Nepomuceno; Francisco Pulido; Gloria Elena Espinosa-Ayala; Gloria María Molina-Salinas; Gloria Vazquez; Hector Esteban Paz-Juárez; Hector Montoya-Fuentes; Helen Haydee Fernanda Ramírez-Plascencia; Irvin González-López; Jean Pierre González; Jesús Hernández; Joel Armando Vázquez-Pérez. LANNIA Nayarit: Manuel Ivan Girón Pérez; Jorge Salas-Hernández; José Antonio Enciso-Moreno; José Arturo Martínez-Orozco; José Esteban Muñoz-Medina; José de Jesús Nuñez-Contreras; Juan Bautista Chale-Dzul; Julissa Enciso-Ibarra; Luis Alberto Ochoa-Carrera; Margarita Matías-Florentino; Mario Mújica-Sánchez; Marissa Perez-Garcia; María Guadalupe Santiago-Mauricio; María Guadalupe de Jesús Mireles-Rivera; Nelly Sélem-Mojica; Pavel Isa; Ricardo Ciria Merce; Ricardo Grande; Rosa María Gutiérrez Rios; Santiago Ávila-Ríos; Selene Zárate; Susana Lopez; Ulises Mercado.; Verónica Mata-Haro; Victor Eduardo García-Arias; Víctor Hugo Borja-Aburto |
| EPI_ISL_3010759, EPI_ISL_3010760, EPI_ISL_3010764, EPI_ISL_3010768, EPI_ISL_3010776, EPI_ISL_3010777, EPI_ISL_3010781, EPI_ISL_3010783, EPI_ISL_3010784, EPI_ISL_3010791, EPI_ISL_3010793, EPI_ISL_3010794, EPI_ISL_3010798, EPI_ISL_3010800, EPI_ISL_3010801, EPI_ISL_3010802, EPI_ISL_3010803, EPI_ISL_3010805, EPI_ISL_3010806, EPI_ISL_3010807, EPI_ISL_3010808, EPI_ISL_3010809, EPI_ISL_3010816, EPI_ISL_3010817, EPI_ISL_3010818, EPI_ISL_3010819, EPI_ISL_3010823, EPI_ISL_3010824, EPI_ISL_3010825, EPI_ISL_3010826, EPI_ISL_3010827, EPI_ISL_3010831, EPI_ISL_3010832, EPI_ISL_3010833, EPI_ISL_3010835, EPI_ISL_3010836, EPI_ISL_3010841, EPI_ISL_3010842, EPI_ISL_3010843, EPI_ISL_3010845, EPI_ISL_3010846, EPI_ISL_3010847, EPI_ISL_3010848, EPI_ISL_3010853, EPI_ISL_3010856, EPI_ISL_3010859, EPI_ISL_3010860, EPI_ISL_3010861, EPI_ISL_3010863, EPI_ISL_3010864, EPI_ISL_3010866, EPI_ISL_3010870, EPI_ISL_3010871, EPI_ISL_3010874, EPI_ISL_3010875, EPI_ISL_3010876, EPI_ISL_3010877, EPI_ISL_3010879, EPI_ISL_3010881, EPI_ISL_3010886, EPI_ISL_3010892, EPI_ISL_3010893, EPI_ISL_3010894, EPI_ISL_3010895, EPI_ISL_3010896, EPI_ISL_3010898, EPI_ISL_3010900, EPI_ISL_3010901, EPI_ISL_3010902, EPI_ISL_3010905, EPI_ISL_3010907, EPI_ISL_3010910, EPI_ISL_3010911, EPI_ISL_3010912, EPI_ISL_3010914, EPI_ISL_3010917, EPI_ISL_3010918, EPI_ISL_3010920, EPI_ISL_3010921, EPI_ISL_3010923, EPI_ISL_3010924, EPI_ISL_3010926, EPI_ISL_3010934, EPI_ISL_3047701, EPI_ISL_3047702, EPI_ISL_3047704, EPI_ISL_3047706, EPI_ISL_3047707, EPI_ISL_3047708, EPI_ISL_3047709, EPI_ISL_3047711, EPI_ISL_3047712, EPI_ISL_3047713, EPI_ISL_3047715, EPI_ISL_3047716, EPI_ISL_3047719, EPI_ISL_3047720, EPI_ISL_3047721, EPI_ISL_3047726, EPI_ISL_3047727, EPI_ISL_3047728, EPI_ISL_3047729, EPI_ISL_3047730, EPI_ISL_3047731, EPI_ISL_3047733, EPI_ISL_3047734, EPI_ISL_3047735, EPI_ISL_3047738, EPI_ISL_3047741, EPI_ISL_3047742, EPI_ISL_3047743, EPI_ISL_3047744, EPI_ISL_3047745, EPI_ISL_3134921, EPI_ISL_3134922, EPI_ISL_3134930, EPI_ISL_3134934, EPI_ISL_3134939, EPI_ISL_3134950, EPI_ISL_3134955, EPI_ISL_3134960, EPI_ISL_3134965, EPI_ISL_3134967, EPI_ISL_3134977, EPI_ISL_3246189, EPI_ISL_3261062, EPI_ISL_3261063, EPI_ISL_3261064, EPI_ISL_3261065, EPI_ISL_3261067, EPI_ISL_3261068, EPI_ISL_3261071, EPI_ISL_3261072, EPI_ISL_3261073, EPI_ISL_3261074, EPI_ISL_3261075, EPI_ISL_3261076, EPI_ISL_3261077, EPI_ISL_3261078, EPI_ISL_3261079, EPI_ISL_3261080, EPI_ISL_3261081, EPI_ISL_3261083, EPI_ISL_3261084, EPI_ISL_3261086, EPI_ISL_3261087, EPI_ISL_3261090, EPI_ISL_3261091, EPI_ISL_3261092, EPI_ISL_3261093, EPI_ISL_3261094, EPI_ISL_3261095, EPI_ISL_3261096, EPI_ISL_3261097, EPI_ISL_3261099, EPI_ISL_3261100, EPI_ISL_3261101, EPI_ISL_3261102, EPI_ISL_3261104, EPI_ISL_3261105, EPI_ISL_3261106, EPI_ISL_3261109, EPI_ISL_3261110, EPI_ISL_3261113, EPI_ISL_3261115, EPI_ISL_3267907, EPI_ISL_3267908, EPI_ISL_3267909, EPI_ISL_3267910, EPI_ISL_3267911, EPI_ISL_3267912, EPI_ISL_3267917, EPI_ISL_3267918, EPI_ISL_3267919, EPI_ISL_3267921, EPI_ISL_3267922, EPI_ISL_3267923, EPI_ISL_3267924, EPI_ISL_3267928, EPI_ISL_3267930, EPI_ISL_3267933, EPI_ISL_3267934, EPI_ISL_3267935, EPI_ISL_3267936, EPI_ISL_3267940, EPI_ISL_3267942, EPI_ISL_3267943, EPI_ISL_3267945, EPI_ISL_3267947, EPI_ISL_3267948, EPI_ISL_3267949, EPI_ISL_3267950, EPI_ISL_3267951, EPI_ISL_3267953, EPI_ISL_3267954, EPI_ISL_3267955, EPI_ISL_3267957, EPI_ISL_3267958, EPI_ISL_3267959, EPI_ISL_3267960, EPI_ISL_3267961, EPI_ISL_3267962, EPI_ISL_3267963, EPI_ISL_3267965, EPI_ISL_3267967, EPI_ISL_3267969, EPI_ISL_3267970, EPI_ISL_3267972, EPI_ISL_3267973, EPI_ISL_3267975, EPI_ISL_3267976, EPI_ISL_3267978, EPI_ISL_3267980, EPI_ISL_3267982, EPI_ISL_3267983, EPI_ISL_3267985, EPI_ISL_3267987, EPI_ISL_3267988, EPI_ISL_3267989, EPI_ISL_3267990, EPI_ISL_3267991, EPI_ISL_3267993, EPI_ISL_3267994, EPI_ISL_3267995, EPI_ISL_3267996, EPI_ISL_3267997, EPI_ISL_3267999, EPI_ISL_3268001, EPI_ISL_3268002, EPI_ISL_3268003, EPI_ISL_3268004, EPI_ISL_3268005, EPI_ISL_3268006, EPI_ISL_3268007, EPI_ISL_3268010, EPI_ISL_3268011, EPI_ISL_3268012, EPI_ISL_3268014, EPI_ISL_3268016, EPI_ISL_3268017, EPI_ISL_3268018, EPI_ISL_3268019, EPI_ISL_3268020, EPI_ISL_3268023, EPI_ISL_3304422 | | | |
| see above | LATE - Laboratório de Técnicas Especiais - Hospital Israelita Albert Einstein | LATE - Laboratório de Técnicas Especiais - Hospital Israelita Albert Einstein | Alexandre Hideaki Takara; Ana Paula Moreira Salles; Anelisie da Silva Santos; Deyvid Amgarten; Erick Gustavo Dorlass; Fernanda de Mello Malta; João Renato Rebello Pinho; Marcio Anunciacao Menezes; Pedro Henrique Sebe Rodrigues; Raquel Riyuzo |
| EPI_ISL_2841778, EPI_ISL_2841782, EPI_ISL_2841785, EPI_ISL_2841787 | LBM ALPHABIO, Marseille | LBM ALPHABIO, Marseille | Vincent GARCIA |
| EPI_ISL_2827783 | LDSP - CAQUETA HOSPITAL MALVINAS | Instituto Nacional de Salud-Dirección de Investigación en Salud Pública | Carlos Franco-Muñoz; Carmen Osorio; Diana Malo; Diego A. Álvarez-Díaz; Diego Andrés Prada; Gerardo Santamaría; Hector Alejandro Ruiz-Moreno; Jhonnatan Reales-González; Jorge Rivera; Juan Camilo Martinez; Julian Naizaque; Katherine Laiton-Donato; Lisseth Pardo; Magdalena Wiesner; Marcela Mercado-Reyes; Maria T. Herrera-Sepúlveda; Marta Lopez Blanco; Martha Lucia Ospina Martinez; Paola Rojas; Sergio Gomez; Sheryll Corchuelo; Ángela Alarcon Cruz |
| EPI_ISL_2827779 | LDSP - CAQUETA HOSPITAL MARIA INMACULADA | Instituto Nacional de Salud-Dirección de Investigación en Salud Pública | Carlos Franco-Muñoz; Carmen Osorio; Diana Malo; Diego A. Álvarez-Díaz; Diego Andrés Prada; Gerardo Santamaría; Hector Alejandro Ruiz-Moreno; Jhonnatan Reales-González; Jorge Rivera; Juan Camilo Martinez; Julian Naizaque; Katherine Laiton-Donato; Lisseth Pardo; Magdalena Wiesner; Marcela Mercado-Reyes; Maria T. Herrera-Sepúlveda; Marta Lopez Blanco; Martha Lucia Ospina Martinez; Paola Rojas; Sergio Gomez; Sheryll Corchuelo; Ángela Alarcon Cruz |
| EPI_ISL_3007486 | LDSP VILLAVICENCIO - CLINICA PRIMAVERA DE VILLAVICENCIO | Instituto Nacional de Salud-Dirección de Investigación en Salud Pública | Carlos Franco-Muñoz; Carmen Osorio; Diana Malo; Diego A. Álvarez-Díaz; Diego Andrés Prada; Gerardo Santamaría; Hector Alejandro Ruiz-Moreno; Jhonnatan Reales-González; Jorge Rivera; Juan Camilo Martinez; Julian Naizaque; Katherine Laiton-Donato; Lisseth Pardo; Magdalena Wiesner; Marcela Mercado-Reyes; Maria T. Herrera-Sepúlveda; Marta Lopez Blanco; Martha Lucia Ospina Martinez; Paola Rojas; Sergio Gomez; Sheryll Corchuelo; Ángela Alarcon Cruz |
| EPI_ISL_2835071, | LESP Aguascalientes | Instituto de Diagnostico y | Abril Rodriguez-Maldonado; Ariadna Medina-Benitez; Claudia Wong-Arambula; Ernesto Ramirez-Gonzalez.; Gisela Barrera-Badillo; Irma Lopez-Martinez; Joaquin Quiroz-Mercado; Lucia Hernandez-Rivas; Maribel Gonzalez-Villa; Natividad Cruz-Ortiz; Sergio Rangel-Guerrero; Tatiana Nunez-Garcia; Vanessa |

| | | | |
|---|---|---|---|
| EPI_ISL_3265588 | | Referencia Epidemiologicos (INDRE) | Rivero-Arredondo |
| EPI_ISL_3033578 | LESP Baja California Sur | Instituto de Diagnostico y Referencia Epidemiologicos (INDRE) | Abril Rodriguez-Maldonado; Ariadna Medina-Benitez; Claudia Wong-Arambula; Ernesto Ramirez-Gonzalez.; Gisela Barrera-Badillo; Irma Lopez-Martinez; Joaquin Quiroz-Mercado; Lucia Hernandez-Rivas; Maribel Gonzalez-Villa; Natividad Cruz-Ortiz; Sergio Rangel-Guerrero; Tatiana Nunez-Garcia; Vanessa Rivero-Arredondo |
| EPI_ISL_3265618 | LESP Campeche | Instituto de Diagnostico y Referencia Epidemiologicos (INDRE) | Abril Rodriguez-Maldonado; Ariadna Medina-Benitez; Claudia Wong-Arambula; Ernesto Ramirez-Gonzalez.; Gisela Barrera-Badillo; Irma Lopez-Martinez; Joaquin Quiroz-Mercado; Lucia Hernandez-Rivas; Maribel Gonzalez-Villa; Natividad Cruz-Ortiz; Sergio Rangel-Guerrero; Tatiana Nunez-Garcia; Vanessa Rivero-Arredondo |
| EPI_ISL_2859010, EPI_ISL_2859011 | LESP Chiapas | Instituto de Diagnostico y Referencia Epidemiologicos (INDRE) | Abril Rodriguez-Maldonado; Ariadna Medina-Benitez; Claudia Wong-Arambula; Ernesto Ramirez-Gonzalez.; Gisela Barrera-Badillo; Irma Lopez-Martinez; Joaquin Quiroz-Mercado; Lucia Hernandez-Rivas; Maribel Gonzalez-Villa; Natividad Cruz-Ortiz; Sergio Rangel-Guerrero; Tatiana Nunez-Garcia; Vanessa Rivero-Arredondo |
| EPI_ISL_2859062, EPI_ISL_2937769, EPI_ISL_2937770, EPI_ISL_2937804, EPI_ISL_2937805, EPI_ISL_3033593, EPI_ISL_3033594 | | | |
| see above | LESP Ciudad de Mexico | Instituto de Diagnostico y Referencia Epidemiologicos (INDRE) | Abril Rodriguez-Maldonado; Ariadna Medina-Benitez; Claudia Wong-Arambula; Ernesto Ramirez-Gonzalez.; Gisela Barrera-Badillo; Irma Lopez-Martinez; Joaquin Quiroz-Mercado; Lucia Hernandez-Rivas; Maribel Gonzalez-Villa; Natividad Cruz-Ortiz; Sergio Rangel-Guerrero; Tatiana Nunez-Garcia; Vanessa Rivero-Arredondo |
| EPI_ISL_2674642 | LESP Durango | Instituto de Diagnostico y Referencia Epidemiologicos (INDRE) | Abril Rodriguez-Maldonado; Ariadna Medina-Benitez; Claudia Wong-Arambula; Ernesto Ramirez-Gonzalez.; Gisela Barrera-Badillo; Irma Lopez-Martinez; Joaquin Quiroz-Mercado; Lucia Hernandez-Rivas; Natividad Cruz-Ortiz; Sergio Rangel-Guerrero; Tatiana Nunez-Garcia; Vanessa Rivero-Arredondo |
| EPI_ISL_2674664 | LESP Estado de Mexico | Instituto de Diagnostico y Referencia Epidemiologicos (INDRE) | Abril Rodriguez-Maldonado; Ariadna Medina-Benitez; Claudia Wong-Arambula; Ernesto Ramirez-Gonzalez.; Gisela Barrera-Badillo; Irma Lopez-Martinez; Joaquin Quiroz-Mercado; Lucia Hernandez-Rivas; Natividad Cruz-Ortiz; Sergio Rangel-Guerrero; Tatiana Nunez-Garcia; Vanessa Rivero-Arredondo |
| EPI_ISL_2674695, EPI_ISL_2779056, EPI_ISL_2779057, EPI_ISL_2779058, EPI_ISL_2779120, EPI_ISL_2779186, EPI_ISL_2779187, EPI_ISL_2779188, EPI_ISL_2779190, EPI_ISL_2779191, EPI_ISL_2779192, EPI_ISL_2800917, EPI_ISL_2800924, EPI_ISL_2858991, EPI_ISL_2858992, EPI_ISL_2858993, EPI_ISL_2937772 | | | |
| see above | LESP Guanajuato | Instituto de Diagnostico y Referencia Epidemiologicos (INDRE) | Abril Rodriguez-Maldonado; Ariadna Medina-Benitez; Claudia Wong-Arambula; Ernesto Ramirez-Gonzalez.; Gisela Barrera-Badillo; Irma Lopez-Martinez; Joaquin Quiroz-Mercado; Lucia Hernandez-Rivas; Maribel Gonzalez-Villa; Natividad Cruz-Ortiz; Sergio Rangel-Guerrero; Tatiana Nunez-Garcia; Vanessa Rivero-Arredondo |
| EPI_ISL_2674665, EPI_ISL_2674677, EPI_ISL_2859012, EPI_ISL_3033562, EPI_ISL_3033566, EPI_ISL_3265597, EPI_ISL_3278284 | | | |
| see above | LESP Hidalgo | Instituto de Diagnostico y Referencia Epidemiologicos (INDRE) | Abril Rodriguez-Maldonado; Ariadna Medina-Benitez; Claudia Wong-Arambula; Ernesto Ramirez-Gonzalez.; Gisela Barrera-Badillo; Irma Lopez-Martinez; Joaquin Quiroz-Mercado; Lucia Hernandez-Rivas; Maribel Gonzalez-Villa; Natividad Cruz-Ortiz; Sergio Rangel-Guerrero; Tatiana Nunez-Garcia; Vanessa Rivero-Arredondo |
| EPI_ISL_2779175, EPI_ISL_2858995, EPI_ISL_2937757, EPI_ISL_2937801, EPI_ISL_2937815, EPI_ISL_3459946, EPI_ISL_3459947 | | | |
| see above | LESP Jalisco | Instituto de Diagnostico y Referencia Epidemiologicos (INDRE) | Abril Rodriguez-Maldonado; Ariadna Medina-Benitez; Claudia Wong-Arambula; Ernesto Ramirez-Gonzalez.; Gisela Barrera-Badillo; Irma Lopez-Martinez; Joaquin Quiroz-Mercado; Lucia Hernandez-Rivas; Maribel Gonzalez-Villa; Natividad Cruz-Ortiz; Sergio Rangel-Guerrero; Tatiana Nunez-Garcia; Vanessa Rivero-Arredondo |
| EPI_ISL_3033575, EPI_ISL_3033592 | LESP Michoacan | Instituto de Diagnostico y Referencia Epidemiologicos (INDRE) | Abril Rodriguez-Maldonado; Ariadna Medina-Benitez; Claudia Wong-Arambula; Ernesto Ramirez-Gonzalez.; Gisela Barrera-Badillo; Irma Lopez-Martinez; Joaquin Quiroz-Mercado; Lucia Hernandez-Rivas; Maribel Gonzalez-Villa; Natividad Cruz-Ortiz; Sergio Rangel-Guerrero; Tatiana Nunez-Garcia; Vanessa Rivero-Arredondo |
| EPI_ISL_2937774, EPI_ISL_3265605 | LESP Nayarit | Instituto de Diagnostico y Referencia Epidemiologicos (INDRE) | Abril Rodriguez-Maldonado; Ariadna Medina-Benitez; Claudia Wong-Arambula; Ernesto Ramirez-Gonzalez.; Gisela Barrera-Badillo; Irma Lopez-Martinez; Joaquin Quiroz-Mercado; Lucia Hernandez-Rivas; Maribel Gonzalez-Villa; Natividad Cruz-Ortiz; Sergio Rangel-Guerrero; Tatiana Nunez-Garcia; Vanessa Rivero-Arredondo |
| EPI_ISL_2859054 | LESP Nuevo Leon | Instituto de Diagnostico y Referencia Epidemiologicos (INDRE) | Abril Rodriguez-Maldonado; Ariadna Medina-Benitez; Claudia Wong-Arambula; Ernesto Ramirez-Gonzalez.; Gisela Barrera-Badillo; Irma Lopez-Martinez; Joaquin Quiroz-Mercado; Lucia Hernandez-Rivas; Maribel Gonzalez-Villa; Natividad Cruz-Ortiz; Sergio Rangel-Guerrero; Tatiana Nunez-Garcia; Vanessa Rivero-Arredondo |
| EPI_ISL_2674700, EPI_ISL_3033596, EPI_ISL_3033597 | LESP Puebla | Instituto de Diagnostico y Referencia Epidemiologicos (INDRE) | Abril Rodriguez-Maldonado; Ariadna Medina-Benitez; Claudia Wong-Arambula; Ernesto Ramirez-Gonzalez.; Gisela Barrera-Badillo; Irma Lopez-Martinez; Joaquin Quiroz-Mercado; Lucia Hernandez-Rivas; Maribel Gonzalez-Villa; Natividad Cruz-Ortiz; Sergio Rangel-Guerrero; Tatiana Nunez-Garcia; Vanessa Rivero-Arredondo |
| EPI_ISL_2545693, EPI_ISL_2545694, EPI_ISL_2937758, EPI_ISL_2937798 | LESP Quintana Roo | Instituto de Diagnostico y Referencia Epidemiologicos (INDRE) | Abril Rodriguez-Maldonado; Ariadna Medina-Benitez; Claudia Wong-Arambula; Ernesto Ramirez-Gonzalez.; Gisela Barrera-Badillo; Irma Lopez-Martinez; Joaquin Quiroz-Mercado; Lucia Hernandez-Rivas; Maribel Gonzalez-Villa; Natividad Cruz-Ortiz; Sergio Rangel-Guerrero; Tatiana Nunez-Garcia; Vanessa Rivero-Arredondo |
| EPI_ISL_2779159, EPI_ISL_2859059, EPI_ISL_2859060, EPI_ISL_2937773 | LESP San Luis Potosi | Instituto de Diagnostico y Referencia Epidemiologicos (INDRE) | Abril Rodriguez-Maldonado; Ariadna Medina-Benitez; Claudia Wong-Arambula; Ernesto Ramirez-Gonzalez.; Gisela Barrera-Badillo; Irma Lopez-Martinez; Joaquin Quiroz-Mercado; Lucia Hernandez-Rivas; Maribel Gonzalez-Villa; Natividad Cruz-Ortiz; Sergio Rangel-Guerrero; Tatiana Nunez-Garcia; Vanessa Rivero-Arredondo |
| EPI_ISL_2545692, EPI_ISL_2674634, EPI_ISL_2859013, EPI_ISL_2859015, EPI_ISL_3033574 | LESP Sinaloa | Instituto de Diagnostico y Referencia Epidemiologicos (INDRE) | Abril Rodriguez-Maldonado; Ariadna Medina-Benitez; Claudia Wong-Arambula; Ernesto Ramirez-Gonzalez.; Gisela Barrera-Badillo; Irma Lopez-Martinez; Joaquin Quiroz-Mercado; Lucia Hernandez-Rivas; Maribel Gonzalez-Villa; Natividad Cruz-Ortiz; Sergio Rangel-Guerrero; Tatiana Nunez-Garcia; Vanessa Rivero-Arredondo |
| EPI_ISL_2674648, EPI_ISL_2674649, EPI_ISL_2779055, EPI_ISL_2779185, EPI_ISL_2859051, EPI_ISL_2937767, EPI_ISL_2937795, EPI_ISL_2937797, EPI_ISL_3033580, EPI_ISL_3265593, EPI_ISL_3265594, EPI_ISL_3459940 | | | |
| see above | LESP Sonora | Instituto de Diagnostico y Referencia Epidemiologicos (INDRE) | Abril Rodriguez-Maldonado; Ariadna Medina-Benitez; Claudia Wong-Arambula; Ernesto Ramirez-Gonzalez.; Gisela Barrera-Badillo; Irma Lopez-Martinez; Joaquin Quiroz-Mercado; Lucia Hernandez-Rivas; Maribel Gonzalez-Villa; Natividad Cruz-Ortiz; Sergio Rangel-Guerrero; Tatiana Nunez-Garcia; Vanessa Rivero-Arredondo |
| EPI_ISL_2779160, EPI_ISL_2779169, EPI_ISL_2859016, EPI_ISL_2937811, EPI_ISL_2937812, EPI_ISL_2937813, EPI_ISL_3033583 | | | |
| see above | LESP Tabasco | Instituto de Diagnostico y Referencia Epidemiologicos (INDRE) | Abril Rodriguez-Maldonado; Ariadna Medina-Benitez; Claudia Wong-Arambula; Ernesto Ramirez-Gonzalez.; Gisela Barrera-Badillo; Irma Lopez-Martinez; Joaquin Quiroz-Mercado; Lucia Hernandez-Rivas; Maribel Gonzalez-Villa; Natividad Cruz-Ortiz; Sergio Rangel-Guerrero; Tatiana Nunez-Garcia; Vanessa Rivero-Arredondo |
| EPI_ISL_2779060, EPI_ISL_2779061, EPI_ISL_2779062, EPI_ISL_2779063, EPI_ISL_2779064, EPI_ISL_2779065, EPI_ISL_2858996, EPI_ISL_2858997, EPI_ISL_2858998, EPI_ISL_2858999, EPI_ISL_2859030, EPI_ISL_2859031, EPI_ISL_2859032, EPI_ISL_2937775, EPI_ISL_2937776, EPI_ISL_2937777, EPI_ISL_2937828, EPI_ISL_2937829, EPI_ISL_2937830, EPI_ISL_3033465, EPI_ISL_3033466, EPI_ISL_3033469, EPI_ISL_3265606 | | | |
| see above | LESP Tamaulipas | Instituto de Diagnostico y Referencia Epidemiologicos (INDRE) | Abril Rodriguez-Maldonado; Ariadna Medina-Benitez; Claudia Wong-Arambula; Ernesto Ramirez-Gonzalez.; Gisela Barrera-Badillo; Irma Lopez-Martinez; Joaquin Quiroz-Mercado; Lucia Hernandez-Rivas; Maribel Gonzalez-Villa; Natividad Cruz-Ortiz; Sergio Rangel-Guerrero; Tatiana Nunez-Garcia; Vanessa Rivero-Arredondo |
| EPI_ISL_3265602 | LESP Veracruz | Instituto de Diagnostico y Referencia Epidemiologicos (INDRE) | Abril Rodriguez-Maldonado; Ariadna Medina-Benitez; Claudia Wong-Arambula; Ernesto Ramirez-Gonzalez.; Gisela Barrera-Badillo; Irma Lopez-Martinez; Joaquin Quiroz-Mercado; Lucia Hernandez-Rivas; Maribel Gonzalez-Villa; Natividad Cruz-Ortiz; Sergio Rangel-Guerrero; Tatiana Nunez-Garcia; Vanessa Rivero-Arredondo |
| EPI_ISL_2545649 | LHUB-ULB | Labo Klinische Biologie, UZA | Basil Britto Xavier; Christine Lammens; Herman Goossens; Jasmine Coppens; Marie Le Mercier; Veerle Matheeussen |
| EPI_ISL_2873838 | LUM Laboratorio Unico Metropolitano - AUSL BOLOGNA | U.O. Microbiologia, Laboratorio Unico Centro Servizi - AUSL della Romagna | Giorgio Dirani |
| EPI_ISL_2695394 | Lab La Salete Robles - VN Famalicao | Instituto Nacional de Saude (INSA) | Borges et al |
| EPI_ISL_2861479, EPI_ISL_3868312 | Lab voor klinische biologie | Lab voor klinische biologie | Bruno Verhasselt; Hannelore Hamerlinck; Marija Janevska |
| EPI_ISL_2725223 | Lab. Microbiologia e Virologia Cotugno A.O. dei Colli - Istituto Zooprofilattico Sperimentale del Mezzogiorno | Telethon Institute of Genetics and Medicine (TIGEM) | Antonio Grimaldi Patrizia Annunziata Francesco Panariello Biancamaria Pierri Claudia Tiberio Teresa Giuliano Valentina Bouche Chiara Colantuono Maria Concetta Cuomo Denise Di Concilio Lucio Di Filippo Anna Manfredi Marcello Salvi Antonio Limone Luigi Atripaldi Pellegrino Cerino Andrea Ballabio Davide Cacchiarelli |
| EPI_ISL_2650016 | LabPLUS | Institute of Environmental Science and Research (ESR) | Anja Werno; Antje van der Linden; Arlo Upton; Chris Mansell; David Hammer; Dragana Drinkovic; Erasmus Smit; Gary McAuliffe; Hana Sofia Andersson; Hermes Perez; James Ussher; Jill Sherwood; Jing Wang; Joep de Ligt; Josh Freeman; Julia Howard; Juliet Elvy; Lauren Jelly; Mary DeAlmeida; Matt Blakiston; Matt Storey; Matthew Rogers; Max Bloomfield; Michael Addidle; Michelle Balm; Muhammad Faisal; Nikki Freed; Olin Silander; Olivia Stroeven; Rachel Boyle; Sally Roberts; SallyAnn Harbison; Sarah Jefferies; Sharmini Muttaiyah; Susan Morpeth; Susan Taylor; Timothy Blackmore; Vani Sathyendran; Veronica Playle; Virginia Hope; Xiaoyun Ren |
| EPI_ISL_2535296, EPI_ISL_2790872, EPI_ISL_2790971, EPI_ISL_2790978, EPI_ISL_2835744, EPI_ISL_2835851, EPI_ISL_2835852, EPI_ISL_2835853, EPI_ISL_2835854, EPI_ISL_2835856, EPI_ISL_2923134, EPI_ISL_2923135, EPI_ISL_2923176, EPI_ISL_2923179, EPI_ISL_2923181, EPI_ISL_2958763, EPI_ISL_3058210, EPI_ISL_3245972 | | | |
| see above | Labo Analyses Med | National Reference Center for Viruses of Respiratory Infections, Institut Pasteur, Paris | Amaury Vaysse; Angela Brisebarre; BéNéDicte Roquebert; Camille Capel; Christophe Malabat; Corinne Maufrais; Damien Mornico; Emmanuelle Permal; Etienne Simon-Lorière; Eve Haguenoer; Frédéric Lemoine; Gael Millot; Hub Bioinformatique Biostatistiques; Hub de bioinformatique et biostatistique; Louise Lefrançois; Marion Barbet; Maud Vanpeene; Méline Bizard; OphéLie Said-Delattre; POTIRON Grégoire; Sabine Trombert; Sabine Trombert; Sylvie Behillil; Sylvie Van der Werf; TROMBERT Sabine; Victoire Baillet; Vincent Enouf |
| EPI_ISL_2504046, EPI_ISL_3113202 | Labor Berlin Charité Vivantes GmbH / Institut für Virologie | Charité Universitätsmedizin Berlin, Institut für Virologie/Labor Berlin | Barbara Mühlemann; Christian Drosten; Christine Stephan; Peter Menzel; Rolf Schwarzer; Terry Jones; Victor M Corman |
| EPI_ISL_2689539 | Labor Dr. Fenner und Kollegen | Heinrich Pette Institute, Leibniz | Adam Grundhoff; Alexis Robitaille; Johannes Knobloch; Martin Aepfelbacher; Nicole Fischer; Thomas Günther |

| | | | |
|---|---|---|---|
| EPI_ISL_2878457 | Laboratoire Ana-L | Institute for Experimental Virology<br>Department of Virology, Henri Mondor University Hospital, Assistance Publique Hôpitaux de Paris, Université Paris-Est Créteil, INSERM U955 | Alexandre Soulier; Christophe Rodriguez; Elisabeth Trawinski; Guillaume Gricourt; Jean-Michel Pawlotsky; Melissa N'Debi; Slim Fourati; Vanessa Demontant |
| EPI_ISL_2426981, EPI_ISL_2627849, EPI_ISL_2629225, EPI_ISL_2629226, EPI_ISL_2629228, EPI_ISL_2629229, EPI_ISL_2835986, EPI_ISL_2835987, EPI_ISL_2835988, EPI_ISL_2835989, EPI_ISL_2835990 | | | |
| see above | Laboratoire Carage | Institut Pasteur de la Guyane | Anne Lavergne; Antoine Enfissi; Arielle Salmier; Dominique Rousset |
| EPI_ISL_3458779, EPI_ISL_3458806, EPI_ISL_3458809, EPI_ISL_3458826, EPI_ISL_3459046, EPI_ISL_3459085 | Laboratoire de santé publique du Québec | Laboratoire de santé publique du Québec | Guillaume Bourque; Ioannis Ragoussis; Jesse Shapiro; Mark Lathrop and Michel Roger on behalf of the CoVSeQ research group; Sandrine Moreira |
| EPI_ISL_3906483 | Laboratori de Referència de Catalunya | SeqCOVID-SPAIN consortium/IBV(CSIC) | Aida Esperanza Ramírez Marinero; Alba Vilas Basil; Eduardo Padilla León; Mireia Canal Aranda and SeqCOVID-SPAIN consortium |
| EPI_ISL_2983182, EPI_ISL_2983185, EPI_ISL_2983186, EPI_ISL_2983231, EPI_ISL_2983233, EPI_ISL_2983234, EPI_ISL_3190351, EPI_ISL_3190352, EPI_ISL_3190354, EPI_ISL_3190355 | | | |
| see above | Laboratorio Central de Saude Publica do Estado de Goias (LACEN/GO) | Laboratory of Respiratory Viruses and Measles, Oswaldo Cruz Institute, FIOCRUZ | Agatha Cristinne Prudencio; Alice Sampaio Rocha; Ana Carolina Mendonca; Ana Flavia Mendonça; Anna Carolina Paixao; Carmen Helena Ramos; Cassiane Casanova; Elisa Cavalcante Pereira; Fernando Motta; Flavia Pereira Amorim da Silva; Ighor Leonardo Arantes Gomes; Luciana Appolinario; Luiz Augusto Pereira; Marilda Siqueira on behalf of the Fiocruz COVID-19 Genomic Surveillance Network; Paola Resende; Rafael Souza Guedes; Renata Serrano Lopes; Taina Venas; Vinicius Lemes da Silva |
| EPI_ISL_3539750, EPI_ISL_3539751, EPI_ISL_3539752, EPI_ISL_3539753, EPI_ISL_3539755, EPI_ISL_3539756, EPI_ISL_3539757, EPI_ISL_3539758, EPI_ISL_3539759, EPI_ISL_3539760, EPI_ISL_3539761, EPI_ISL_3539762, EPI_ISL_3539763, EPI_ISL_3539764, EPI_ISL_3539765 | | | |
| see above | Laboratorio Central de Saude Publica do Estado de Minas Gerais (LACEN/MG) | Laboratory of Respiratory Viruses and Measles, Oswaldo Cruz Institute, FIOCRUZ | Alice Sampaio Rocha; Ana Carolina Mendonca; Anna Carolina Paixao; Elisa Cavalcante Pereira; Fernando Motta; Luciana Appolinario; Marilda Siqueira on behalf of the Fiocruz COVID-19 Genomic Surveillance Network; Paola Resende; Renata Serrano Lopes; Rubens Pasa; Taina Venas |
| EPI_ISL_2645434, EPI_ISL_2645435, EPI_ISL_2731453, EPI_ISL_2731454, EPI_ISL_2731455, EPI_ISL_2731456, EPI_ISL_2863578, EPI_ISL_2863580, EPI_ISL_2863582, EPI_ISL_2863706, EPI_ISL_2863707, EPI_ISL_2863709, EPI_ISL_2863711, EPI_ISL_2863801, EPI_ISL_2863802, EPI_ISL_2863803, EPI_ISL_2863804, EPI_ISL_2863805, EPI_ISL_2863806, EPI_ISL_2863807, EPI_ISL_2863808, EPI_ISL_2863809, EPI_ISL_2863810, EPI_ISL_2863811, EPI_ISL_2863812, EPI_ISL_2863813, EPI_ISL_2863814, EPI_ISL_2863815, EPI_ISL_2863816, EPI_ISL_2863817, EPI_ISL_2863819, EPI_ISL_2863820, EPI_ISL_2863821, EPI_ISL_2863822, EPI_ISL_2863823, EPI_ISL_2863824, EPI_ISL_2863825, EPI_ISL_2863826, EPI_ISL_2863827, EPI_ISL_2863828, EPI_ISL_2863829, EPI_ISL_2863830, EPI_ISL_2863831, EPI_ISL_2863832, EPI_ISL_2863833, EPI_ISL_2863834, EPI_ISL_2863835, EPI_ISL_2863837, EPI_ISL_2863838, EPI_ISL_2863839, EPI_ISL_2863840, EPI_ISL_2863841, EPI_ISL_2863842, EPI_ISL_2863843, EPI_ISL_2863844, EPI_ISL_2863845, EPI_ISL_2863846, EPI_ISL_2863847, EPI_ISL_2863848, EPI_ISL_2863849, EPI_ISL_2863850, EPI_ISL_2863851, EPI_ISL_2863852, EPI_ISL_2863853, EPI_ISL_2863854, EPI_ISL_2863894, EPI_ISL_4080766, EPI_ISL_4080769, EPI_ISL_4080770 | | | |
| see above | Laboratorio Central de Saude Publica do Estado do Para (LACEN/PA) | Laboratory of Respiratory Viruses and Measles, Oswaldo Cruz Institute, FIOCRUZ | Agatha Cristinne Prudencio; Agatha Cristinne Prudencio Soares; Agatha Soares; Alice Sampaio Rocha; Ana Carolina Mendonca; Anna Carolina Paixao; Elisa Cavalcante Pereira; Fernando Motta; Ighor Arantes; Ighor Leonardo Arantes Gomes; Luciana Appolinario; Marilda Siqueira on behalf of the Fiocruz COVID-19 Genomic Surveillance Network; Paola Resende; Renata Serrano Lopes; Taina Venas; Valnete Andrade |
| EPI_ISL_2535078, EPI_ISL_2535087, EPI_ISL_2535088, EPI_ISL_2535108, EPI_ISL_2535109, EPI_ISL_2535110, EPI_ISL_2691315, EPI_ISL_2691316, EPI_ISL_2691317, EPI_ISL_2691318, EPI_ISL_2691319, EPI_ISL_2691321, EPI_ISL_2691322, EPI_ISL_2691323, EPI_ISL_2691324, EPI_ISL_2691325, EPI_ISL_2691326, EPI_ISL_2691327, EPI_ISL_2691328, EPI_ISL_2691330, EPI_ISL_2691331, EPI_ISL_2691332, EPI_ISL_2691333, EPI_ISL_2691334, EPI_ISL_2691336, EPI_ISL_2691337, EPI_ISL_2691338, EPI_ISL_2691339, EPI_ISL_2691340, EPI_ISL_2691341, EPI_ISL_2691343, EPI_ISL_2691344, EPI_ISL_2691345, EPI_ISL_2691347, EPI_ISL_2691348, EPI_ISL_2691349, EPI_ISL_2691350, EPI_ISL_2691351, EPI_ISL_2691352, EPI_ISL_2691357, EPI_ISL_2691359, EPI_ISL_2691360, EPI_ISL_2691362, EPI_ISL_2691364, EPI_ISL_2691366, EPI_ISL_2691367, EPI_ISL_2691368, EPI_ISL_2691369, EPI_ISL_2691370, EPI_ISL_2691371, EPI_ISL_2691372, EPI_ISL_2691373, EPI_ISL_2691374, EPI_ISL_2691375, EPI_ISL_2691376, EPI_ISL_2691377, EPI_ISL_2691378, EPI_ISL_2691380, EPI_ISL_2691381, EPI_ISL_2691383, EPI_ISL_2691384, EPI_ISL_2691385, EPI_ISL_2691387, EPI_ISL_2691388, EPI_ISL_2691389, EPI_ISL_2691390, EPI_ISL_2691391, EPI_ISL_2691392, EPI_ISL_2691393, EPI_ISL_2691394, EPI_ISL_2691395, EPI_ISL_2691396, EPI_ISL_2691397, EPI_ISL_2691398, EPI_ISL_2691399, EPI_ISL_2691400, EPI_ISL_2691401, EPI_ISL_2691402, EPI_ISL_2691403, EPI_ISL_2691404, EPI_ISL_2691409, EPI_ISL_2691410, EPI_ISL_2691411, EPI_ISL_2691412, EPI_ISL_2691414, EPI_ISL_2691415, EPI_ISL_2691416, EPI_ISL_2691417, EPI_ISL_2691419, EPI_ISL_2691420, EPI_ISL_2691424, EPI_ISL_2691425, EPI_ISL_2691426, EPI_ISL_2691427, EPI_ISL_2691428, EPI_ISL_2691430, EPI_ISL_2691431, EPI_ISL_2691432, EPI_ISL_2691433, EPI_ISL_2691434, EPI_ISL_2691435, EPI_ISL_2691436, EPI_ISL_2691437, EPI_ISL_2691438, EPI_ISL_2691439, EPI_ISL_2691441, EPI_ISL_2691442, EPI_ISL_2691443, EPI_ISL_2691444, EPI_ISL_2691445, EPI_ISL_2691446, EPI_ISL_2691447, EPI_ISL_2691448, EPI_ISL_2691449, EPI_ISL_2691451, EPI_ISL_2691453, EPI_ISL_2691454, EPI_ISL_2691455, EPI_ISL_2691456, EPI_ISL_2691457, EPI_ISL_2691458, EPI_ISL_2691459, EPI_ISL_2691460, EPI_ISL_2691461, EPI_ISL_2691462, EPI_ISL_2691463, EPI_ISL_2691464, EPI_ISL_2691465, EPI_ISL_2691466, EPI_ISL_2691467, EPI_ISL_2691468, EPI_ISL_2691469, EPI_ISL_2691470, EPI_ISL_2691471, EPI_ISL_2691472, EPI_ISL_2691473, EPI_ISL_2691474, EPI_ISL_2691476, EPI_ISL_2691477, EPI_ISL_2691478, EPI_ISL_2691479, EPI_ISL_2691480, EPI_ISL_2691481, EPI_ISL_2691482, EPI_ISL_2691483, EPI_ISL_2691484, EPI_ISL_2691485, EPI_ISL_2691486, EPI_ISL_2691487, EPI_ISL_2691488, EPI_ISL_2691489, EPI_ISL_2691490, EPI_ISL_2691491, EPI_ISL_2691492, EPI_ISL_2691493, EPI_ISL_2691494, EPI_ISL_2691495, EPI_ISL_2691496, EPI_ISL_2691497, EPI_ISL_2691499, EPI_ISL_2691500, EPI_ISL_2691501, EPI_ISL_2691502, EPI_ISL_2691503, EPI_ISL_2691504, EPI_ISL_2691505, EPI_ISL_2691506, EPI_ISL_2691507, EPI_ISL_2691508, EPI_ISL_2691509, EPI_ISL_2691510, EPI_ISL_2691511, EPI_ISL_2691512, EPI_ISL_2691513, EPI_ISL_2691515, EPI_ISL_2691517, EPI_ISL_2691518, EPI_ISL_2691519, EPI_ISL_2691520, EPI_ISL_2691521, EPI_ISL_2691522, EPI_ISL_2691525, EPI_ISL_2691527, EPI_ISL_2691528, EPI_ISL_2691529, EPI_ISL_2691530, EPI_ISL_2691531, EPI_ISL_2691533, EPI_ISL_2691534, EPI_ISL_2691535, EPI_ISL_2691537, EPI_ISL_2691538, EPI_ISL_2691539, EPI_ISL_2691540, EPI_ISL_2691541, EPI_ISL_2691542, EPI_ISL_2691543, EPI_ISL_2691545, EPI_ISL_2691546, EPI_ISL_2691547, EPI_ISL_2691548, EPI_ISL_2691549, EPI_ISL_2691550, EPI_ISL_2691551, EPI_ISL_2691552, EPI_ISL_2691553, EPI_ISL_2691554, EPI_ISL_2691555, EPI_ISL_2691556, EPI_ISL_2691557, EPI_ISL_2691558, EPI_ISL_2691560, EPI_ISL_2691561, EPI_ISL_2691563, EPI_ISL_2691564, EPI_ISL_2691565, EPI_ISL_2691566, EPI_ISL_2691567, EPI_ISL_2691568, EPI_ISL_2836977, EPI_ISL_2836978, EPI_ISL_2836979, EPI_ISL_2836981, EPI_ISL_2836982, EPI_ISL_2836983, EPI_ISL_2836984, EPI_ISL_2836985, EPI_ISL_2836988, EPI_ISL_2836989, EPI_ISL_2836990, EPI_ISL_2836992, EPI_ISL_2836996, EPI_ISL_2837001, EPI_ISL_2837002, EPI_ISL_2837004, EPI_ISL_2837005, EPI_ISL_2837006, EPI_ISL_2837007, EPI_ISL_2837008, EPI_ISL_2837009, EPI_ISL_2837010, EPI_ISL_2837011, EPI_ISL_2837012, EPI_ISL_2837013, EPI_ISL_2837014, EPI_ISL_2837015, EPI_ISL_2837017, EPI_ISL_2837019, EPI_ISL_2837020, EPI_ISL_2837021, EPI_ISL_2837022, EPI_ISL_2837026, EPI_ISL_2837027, EPI_ISL_2837028, EPI_ISL_2837029, EPI_ISL_2837030, EPI_ISL_2837031, EPI_ISL_2837032, EPI_ISL_2837033, EPI_ISL_2837034, EPI_ISL_2837035, EPI_ISL_2837036, EPI_ISL_2837037, EPI_ISL_2837038, EPI_ISL_2837040, EPI_ISL_2837043, EPI_ISL_2837045, EPI_ISL_2837046, EPI_ISL_2837048, EPI_ISL_2837049, EPI_ISL_2837050, EPI_ISL_2837051, EPI_ISL_2837052, EPI_ISL_2837053, EPI_ISL_2837054, EPI_ISL_2837055, EPI_ISL_2837056, EPI_ISL_2837060, EPI_ISL_2837063, EPI_ISL_2837064, EPI_ISL_2837065, EPI_ISL_2837066, EPI_ISL_2837067, EPI_ISL_2837068, EPI_ISL_2837069, EPI_ISL_2837071, EPI_ISL_2837072, EPI_ISL_2837074, EPI_ISL_2837075, EPI_ISL_2837076, EPI_ISL_2837077, EPI_ISL_2837078, EPI_ISL_2837079, EPI_ISL_2837080, EPI_ISL_2837081, EPI_ISL_2837082, EPI_ISL_2837083, EPI_ISL_2837084, EPI_ISL_2837087, EPI_ISL_2837088, EPI_ISL_2837089, EPI_ISL_2837091, EPI_ISL_2837094, EPI_ISL_2837095, EPI_ISL_2837098, EPI_ISL_2837099, EPI_ISL_2837100, EPI_ISL_2837101, EPI_ISL_2837102, EPI_ISL_2837103, EPI_ISL_2837105, EPI_ISL_2837106, EPI_ISL_2837107, EPI_ISL_2837108, EPI_ISL_2837109, EPI_ISL_2837110, EPI_ISL_2837111, EPI_ISL_2837113, EPI_ISL_2837114, EPI_ISL_2837115, EPI_ISL_2837118, EPI_ISL_2837119, EPI_ISL_2837120, EPI_ISL_2837122, EPI_ISL_2837123, EPI_ISL_2837124, EPI_ISL_2837125, EPI_ISL_2837126, EPI_ISL_2837130, EPI_ISL_2837132, EPI_ISL_2837133, EPI_ISL_2837134, EPI_ISL_2837135, EPI_ISL_2837136, EPI_ISL_2837137, EPI_ISL_2837138, EPI_ISL_2837139, EPI_ISL_2837140, EPI_ISL_2837141, EPI_ISL_2837142, EPI_ISL_2837143, EPI_ISL_2837144, EPI_ISL_2837146, EPI_ISL_2837147, EPI_ISL_2837148, EPI_ISL_2837149, EPI_ISL_2837150, EPI_ISL_2837151, EPI_ISL_2837152, EPI_ISL_2837153, EPI_ISL_2837154, EPI_ISL_2837155, EPI_ISL_2837156, EPI_ISL_2837157, EPI_ISL_2837159, EPI_ISL_2837160, EPI_ISL_2837162, EPI_ISL_2837163, EPI_ISL_2837165, EPI_ISL_2837167, EPI_ISL_2837168, EPI_ISL_2837169, EPI_ISL_2837170, EPI_ISL_2837171, EPI_ISL_2837172, EPI_ISL_2837179, EPI_ISL_2837180, EPI_ISL_2837181, EPI_ISL_2837182, EPI_ISL_2837183, EPI_ISL_2837184, EPI_ISL_2837186, EPI_ISL_2837187, EPI_ISL_2837189, EPI_ISL_2837190, EPI_ISL_2837191, EPI_ISL_2837192, EPI_ISL_2837193, EPI_ISL_2837194, EPI_ISL_2837195, EPI_ISL_2837196, EPI_ISL_2837197, EPI_ISL_2837198, EPI_ISL_2837199, EPI_ISL_2837200, EPI_ISL_2837202, EPI_ISL_2837203, EPI_ISL_2837204, EPI_ISL_2837205, EPI_ISL_2837206, EPI_ISL_2837207, EPI_ISL_3245102, EPI_ISL_3245105, EPI_ISL_3245106, EPI_ISL_3245108, EPI_ISL_3245112, EPI_ISL_3245113, EPI_ISL_3245119, EPI_ISL_3245122, EPI_ISL_3245123, EPI_ISL_3245124, EPI_ISL_3245129, EPI_ISL_3245130, EPI_ISL_3245131, EPI_ISL_3245134, EPI_ISL_3245136, EPI_ISL_3245137, EPI_ISL_3245138, EPI_ISL_3245139, EPI_ISL_3245141, EPI_ISL_3245142, EPI_ISL_3245144, EPI_ISL_3245145, EPI_ISL_3245146, EPI_ISL_3245147, EPI_ISL_3245149, EPI_ISL_3245151, EPI_ISL_3245152, EPI_ISL_3245153, EPI_ISL_3245155, EPI_ISL_3245156, EPI_ISL_3245157, EPI_ISL_3245160, EPI_ISL_3245161, EPI_ISL_3245162, EPI_ISL_3245164, EPI_ISL_3245165, EPI_ISL_3245167, EPI_ISL_3245168, EPI_ISL_3245171, EPI_ISL_3245172, EPI_ISL_3245175, EPI_ISL_3245176, EPI_ISL_3245177, EPI_ISL_3245179, EPI_ISL_3245180, EPI_ISL_3245183, EPI_ISL_3245184, EPI_ISL_3245185, EPI_ISL_3245189, EPI_ISL_3245191, EPI_ISL_3245193, EPI_ISL_3245194, EPI_ISL_3245197, EPI_ISL_3245198, EPI_ISL_3245200, EPI_ISL_3245201, EPI_ISL_3245203, EPI_ISL_3245204, EPI_ISL_3245206, EPI_ISL_3245207, EPI_ISL_3245209, EPI_ISL_3245210, EPI_ISL_3245211, EPI_ISL_3245212, EPI_ISL_3245213, EPI_ISL_3245214, EPI_ISL_3245216, EPI_ISL_3245217, EPI_ISL_3245218, EPI_ISL_3245220, EPI_ISL_3245221, EPI_ISL_3245222, EPI_ISL_3245223, EPI_ISL_3245224, EPI_ISL_3245230, EPI_ISL_3245232, EPI_ISL_3245234, EPI_ISL_3245235, EPI_ISL_3245237, EPI_ISL_3245239, EPI_ISL_3245300, EPI_ISL_3245340 | | | |
| see above | Laboratorio Central Noel Nutels | Bioinformatics Laboratory / LNCC | Alessandra P Lamarca; Alexandra L Gerber; Amilcar Tanuri; Ana Paula de C Guimaraes; Ana Tereza R Vasconcelos; Andrea Cony Cavalcanti; Caio Luiz Pereira Ribeiro; Cassia Alves; Cintia Policarpo; Claudia Maria Braga de Mello; Cristiane Gomes da Silva; Diana Mariani; Douglas Terra Machado; Erica Ramos dos Santos Nascimento; Fernanda Leitao dos Santos; Flavio Dias da Silva; Gleidson da Silva de Oliveira; Leandro Magalhaes de Souza; Liliane Cavalcante; Luiz G P de Almeida; Marcio Henrique de Oliveira Garcia; Mario Sergio Ribeiro; Ricardo Jose Barbosa Salviano; Ronaldo da Silva F Jr; Silvia Carvalho |
| EPI_ISL_2942513, EPI_ISL_2942520, EPI_ISL_2942527, EPI_ISL_2942547, EPI_ISL_2942569, EPI_ISL_2942573, EPI_ISL_2942581, EPI_ISL_2942606, EPI_ISL_2942837, EPI_ISL_2942893, EPI_ISL_2942903, EPI_ISL_2942904, EPI_ISL_2942908, EPI_ISL_2942924 | | | |
| see above | Laboratorio Central de Epidemiologia (LCE) | Unidad de Genomica Avanzada | ; Alejandra Garcia-Gasca; Alejandra Hernandez-Teran; Alejandro Sanchez-Flores; Alfredo Herrera-Estrella; Alicia Ocaña-Mondragon; Andreu Comas-Garcia; Angel Gustavo Salas-Lais; Antonio Loza Roman; Bernardo Martinez-Miguel; Blanca Taboada; Brenda Irasema Maldonado-Meza; Bruno Gomez-Gil; Carla Ivon Herrera-Najera; Carlos F. Arias; Celia Boukadida; Celida Duque Molina; Celida Martinez- Rodriguez; Clara Esperanza Santacruz-Tinoco; Concepcion Grajales-Muñiz; Consorcio Mexicano de Vigilancia Genomica (CoViGen-Mex). Authors (in alphabetical order): Julio Elias Alvarado-Yaah; Cristobal Chaidez-Quiroz; Daniel Fregoso-Rueda; Daniel Lira Morales; Eduardo Becerril-Vargas; Fernando Fontove-Herrera; Fidencio Mejia-Nepomuceno; Francisco Pulido; Gloria Elena Espinosa-Ayala; Gloria Maria Molina-Salinas; Gloria Vazquez; Hector Esteban Paz-Juarez; Hector Montoya-Fuentes; Helen Haydee Fernanda Ramirez-Plascencia; Irvin Gonzalez-Lopez; Jean Pierre Gonzalez; Jesus Hernandez; Joel Armando Vazquez-Perez.; Jorge Salas-Hernandez; Jose Antonio Enciso-Moreno; Jose Arturo Martinez-Orozco; Jose Esteban Muñoz-Medina; Jose de Jesus Nuñez-Contreras; Juan Bautista Chale-Dzul; Julissa Enciso-Ibarra; Luis Alberto Ochoa-Carrera; Margarita Matias-Florentino; Maria Guadalupe Santiago-Mauricio; Maria Guadalupe de Jesus Mireles-Rivera; Mario Mujica-Sanchez; Marissa Perez-Garcia; Nelly Selem-Mojica; Pavel Isa; Ricardo Ciria Merce; Ricardo Grande; Rosa Maria Gutierrez Rios; Santiago avila-Rios; Selene Zarate; Susana Lopez; Veronica Mata-Haro; Victor Eduardo Garcia-Arias; Victor Hugo Borja-Aburto |
| EPI_ISL_3155578, EPI_ISL_3155611, EPI_ISL_3155623, EPI_ISL_3155625 | Laboratorio Central de Epidemiología (LCE) | Centro de Investigación en Enfermedades Infecciosas (CIENI), Instituto Nacional de Enfermedades Respiratorias (INER) | ; Alejandra García-Gasca; Alejandra Hernández-Terán; Alejandro Sánchez-Flores; Alfredo Herrera-Estrella; Alicia Ocaña-Mondragón; Andreu Comas-García; Angel Gustavo Salas-Lais; Antonio Loza Román; Bernardo Martínez-Miguel; Blanca Taboada; Brenda Irasema Maldonado-Meza; Bruno Gómez-Gil; Carla Ivón Herrera-Najera; Carlos F. Arias; Celia Boukadida; Clara Esperanza Santacruz-Tinoco; Concepción Grajales-Muñiz; Consorcio Mexicano de Vigilancia Genómica (CoViGen-Mex). Authors (in alphabetical order): Julio Elias Alvarado-Yaah; Cristóbal Cháidez-Quiróz; Célida Duque Molina; Célida Martínez- Rodríguez; Daniel Fregoso-Rueda; Daniel Lira Morales; Eduardo Becerril-Vargas; Fernando Fontove-Herrera; Fidencio Mejía-Nepomuceno; Francisco Pulido; Gloria Elena Espinosa-Ayala; Gloria María Molina-Salinas; Gloria Vazquez; Hector Esteban Paz-Juárez; Hector Montoya-Fuentes; Helen Haydee Fernanda Ramírez-Plascencia; Irvin González-López; Jean Pierre González; Jesús Hernández; Joel Armando Vázquez-Pérez.; Jorge Salas-Hernández; José Antonio Enciso-Moreno; José Arturo Martínez-Orozco; José Esteban Muñoz-Medina; José de Jesús Nuñez-Contreras; Juan Bautista Chale-Dzul; Julissa Enciso-Ibarra; Kathia Elizabeth Tapia-Diaz; Luis Alberto Ochoa-Carrera; Margarita Matías-Florentino; Mario Mújica-Sánchez; Marissa Pérez-Garcia; María Guadalupe Santiago-Mauricio; María Guadalupe de Jesús Mireles-Rivera; Nelly Sélem-Mojica; Pavel Isa; Ricardo Ciria Merce; Ricardo Grande; Rosa María Gutiérrez Rios; Santiago Ávila-Ríos; Selene Zárate; Susana Lopez; Verónica Mata-Haro; Victor Eduardo García-Arias; Víctor Hugo Borja-Aburto |
| EPI_ISL_2681076, EPI_ISL_2681112, EPI_ISL_2681158, EPI_ISL_2681163, EPI_ISL_2681179, EPI_ISL_2681182, EPI_ISL_2681225, EPI_ISL_2801768, EPI_ISL_2801769, EPI_ISL_2801796, EPI_ISL_2801834, EPI_ISL_2801846, EPI_ISL_2801860, EPI_ISL_2801867, EPI_ISL_2801883, EPI_ISL_2801884 | | | |
| see above | Laboratorio Central de Epidemiología (LCE) | Instituto de Biotecnología de la UNAM | ; Alejandra García-Gasca; Alejandra Hernández-Terán; Alejandro Sánchez-Flores; Alfredo Herrera-Estrella; Alicia Ocaña-Mondragón; Andreu Comas-García; Angel Gustavo Salas-Lais; Antonio Loza Román; Bernardo Martínez-Miguel; Blanca Taboada; Brenda Irasema Maldonado-Meza; Bruno Gómez-Gil; Carla Ivón Herrera-Najera; Carlos F. Arias; Celia Boukadida; Clara Esperanza Santacruz-Tinoco; Concepción Grajales-Muñiz; Consorcio Mexicano de Vigilancia Genómica (CoViGen-Mex). Authors (in alphabetical order): Julio Elias Alvarado-Yaah; Cristóbal Cháidez-Quiróz; Célida Duque Molina; Célida Martínez- Rodríguez; Daniel Fregoso-Rueda; Daniel Lira Morales; Eduardo Becerril-Vargas; Fernando Fontove-Herrera; Fidencio Mejía-Nepomuceno; Francisco Pulido; Gloria Elena Espinosa-Ayala; Gloria María Molina-Salinas; Gloria Vazquez; Hector Esteban Paz-Juárez; Hector Montoya-Fuentes; Helen Haydee Fernanda Ramírez-Plascencia; Irvin González-López; Jean Pierre González; Jesús Hernández; Joel Armando Vázquez-Pérez.; Jorge Salas-Hernández; José Antonio Enciso-Moreno; José Arturo Martínez-Orozco; José Esteban Muñoz-Medina; José de Jesús Nuñez-Contreras; Juan Bautista Chale-Dzul; Julissa Enciso-Ibarra; Luis Alberto Ochoa-Carrera; Margarita Matías-Florentino; Mario Mújica-Sánchez; Marissa Pérez-Garcia; María Guadalupe Santiago-Mauricio; María Guadalupe de Jesús Mireles-Rivera; Nelly Sélem-Mojica; Pavel Isa; Ricardo Ciria Merce; Ricardo Grande; Rosa María Gutiérrez Rios; Santiago Ávila-Ríos; Selene Zárate; Susana Lopez; Verónica Mata-Haro; Victor Eduardo García-Arias; Víctor Hugo Borja-Aburto |
| EPI_ISL_3434751, EPI_ISL_3434752, EPI_ISL_3434755, EPI_ISL_3434758 | Laboratorio Central de Saude Publica do Estado Maranhao (LACEN-MA) | Laboratory of Respiratory Viruses and Measles, Oswaldo Cruz Institute, FIOCRUZ | Alice Sampaio Rocha; Ana Carolina Mendonca; Anna Carolina Paixao; Elisa Cavalcante Pereira; Fernando Motta; Lidio Gonçalves Lima Neto; Luciana Appolinario; Marilda Siqueira on behalf of the Fiocruz COVID-19 Genomic Surveillance Network; Paola Resende; Renata Serrano Lopes; Taina Venas |
| EPI_ISL_2645408, EPI_ISL_2645409, EPI_ISL_2645410, EPI_ISL_2863589, EPI_ISL_2863591, EPI_ISL_2863593, EPI_ISL_2863695, EPI_ISL_2863714, EPI_ISL_2863716, EPI_ISL_2983374, EPI_ISL_2983375, EPI_ISL_2983376, EPI_ISL_3045424, EPI_ISL_3045425, EPI_ISL_3190169, EPI_ISL_3190170, EPI_ISL_3190171 | | | |
| see above | Laboratorio Central de Saude Publica do Estado da Bahia (LACEN/BA) | Laboratory of Respiratory Viruses and Measles, Oswaldo Cruz Institute, FIOCRUZ | Alice Sampaio Rocha; Ana Carolina Mendonca; Anna Carolina Paixao; Elisa Cavalcante Pereira; Felicidade Pereira; Fernando Motta; Luciana Appolinario; Marilda Siqueira on behalf of the Fiocruz COVID-19 Genomic Surveillance Network; Paola Resende; Renata Serrano Lopes; Taina Venas |
| EPI_ISL_3190174, EPI_ISL_3190175, EPI_ISL_3190176, EPI_ISL_3190178, EPI_ISL_3190179, EPI_ISL_3190180, EPI_ISL_3190181, EPI_ISL_3190182, EPI_ISL_3190183, EPI_ISL_3190184, EPI_ISL_3190185, EPI_ISL_3190186, EPI_ISL_3190187, EPI_ISL_3190188, EPI_ISL_3235238, EPI_ISL_3235239, EPI_ISL_3235240, EPI_ISL_3235241, EPI_ISL_3235242, EPI_ISL_3235243, EPI_ISL_3235245, EPI_ISL_3235246, EPI_ISL_3235247, EPI_ISL_3235248, EPI_ISL_3235249, EPI_ISL_3235250, EPI_ISL_3235251, EPI_ISL_3235252, EPI_ISL_3235253, EPI_ISL_3235254, EPI_ISL_3235255, EPI_ISL_3434726, EPI_ISL_3434762, EPI_ISL_3434763, EPI_ISL_3434764, EPI_ISL_3434765, EPI_ISL_3434767, EPI_ISL_3434768, EPI_ISL_3434772, EPI_ISL_3434773, EPI_ISL_3434774, EPI_ISL_3434775, EPI_ISL_3434776, EPI_ISL_3434777, EPI_ISL_3434778, EPI_ISL_3434783, EPI_ISL_3434784, EPI_ISL_3434785, EPI_ISL_3434786, EPI_ISL_3434787, EPI_ISL_3434788, EPI_ISL_3434802, EPI_ISL_3434905, EPI_ISL_3539936 | | | |
| see above | Laboratorio Central de Saude Publica do Estado da Paraiba (LACEN/PB) | Laboratory of Respiratory Viruses and Measles, Oswaldo Cruz Institute, FIOCRUZ | Alice Sampaio Rocha; Ana Carolina Mendonca; Anna Carolina Paixao; Dalane Loudal Florentino Teixeira; Elisa Cavalcante Pereira; Fernando Motta; Joao Felipe Bezerra; Luciana Appolinario; Marilda Siqueira on behalf of the Fiocruz COVID-19 Genomic Surveillance Network; Paola Resende; Renata Serrano Lopes; Taina Venas |
| EPI_ISL_2645386, EPI_ISL_2645400, EPI_ISL_2645401, EPI_ISL_2645403, EPI_ISL_2645404, EPI_ISL_2983064, EPI_ISL_2983067, EPI_ISL_2983068, EPI_ISL_2983069, EPI_ISL_2983070, EPI_ISL_2983071, EPI_ISL_2983072, EPI_ISL_2983073, EPI_ISL_2983074, EPI_ISL_2983075, EPI_ISL_2983076, EPI_ISL_2983077, EPI_ISL_2983078, EPI_ISL_2983079, EPI_ISL_2983080, EPI_ISL_2983081, EPI_ISL_2983082, EPI_ISL_2983083, EPI_ISL_2983084, EPI_ISL_2983085, EPI_ISL_2983338, EPI_ISL_2983478, EPI_ISL_3190227, EPI_ISL_3190228, EPI_ISL_3190229, EPI_ISL_3190230, EPI_ISL_3190231, EPI_ISL_3190232, EPI_ISL_3190233, EPI_ISL_3190234, EPI_ISL_3190235, EPI_ISL_3190236, EPI_ISL_3190310, EPI_ISL_3190312, EPI_ISL_3190313, EPI_ISL_3190314, EPI_ISL_3190315, EPI_ISL_3190316, EPI_ISL_3190317, EPI_ISL_3190318, EPI_ISL_3190319, EPI_ISL_3190320, EPI_ISL_3190321, EPI_ISL_3190322, EPI_ISL_3190323, EPI_ISL_3190325, EPI_ISL_3190327, EPI_ISL_3190328, EPI_ISL_3190329, EPI_ISL_3190330, EPI_ISL_3190331, EPI_ISL_3190332, EPI_ISL_3190335, EPI_ISL_3190339, EPI_ISL_3190340, EPI_ISL_3190341, EPI_ISL_3190342, EPI_ISL_3190343, EPI_ISL_3190344, EPI_ISL_3190345, EPI_ISL_3190346, EPI_ISL_3190347, EPI_ISL_3190348, EPI_ISL_3190349, EPI_ISL_3434825, EPI_ISL_3434826, EPI_ISL_3434827, EPI_ISL_3434828, EPI_ISL_3434829, EPI_ISL_3434830, EPI_ISL_3434831, EPI_ISL_3434832, EPI_ISL_3434833, EPI_ISL_3434834, EPI_ISL_3434835, EPI_ISL_3434836, EPI_ISL_3434837, EPI_ISL_3434838, EPI_ISL_3434839, EPI_ISL_3434840, EPI_ISL_3434841, EPI_ISL_3434842, EPI_ISL_3434843, EPI_ISL_3434849, EPI_ISL_3434850, EPI_ISL_3434851, EPI_ISL_3434852, EPI_ISL_3434853, EPI_ISL_3434854, EPI_ISL_3434855, EPI_ISL_3434856, EPI_ISL_3434857, EPI_ISL_3434858, EPI_ISL_3434859, EPI_ISL_3434860, EPI_ISL_3434861, EPI_ISL_3434862, EPI_ISL_3434863, EPI_ISL_3434864, EPI_ISL_3434865, EPI_ISL_3434866, EPI_ISL_3434867, EPI_ISL_3434868, EPI_ISL_3434869, EPI_ISL_3434870, EPI_ISL_3434871, EPI_ISL_3434872, EPI_ISL_3434873, EPI_ISL_3434874, EPI_ISL_3434875, EPI_ISL_3434876, EPI_ISL_3434877, EPI_ISL_3434878, EPI_ISL_3434879, EPI_ISL_3434880, EPI_ISL_3434881, EPI_ISL_3434882, EPI_ISL_3434883, EPI_ISL_3434884, EPI_ISL_3434885, EPI_ISL_3434886, EPI_ISL_3434887, EPI_ISL_3434888, EPI_ISL_3434889, EPI_ISL_3434890, EPI_ISL_3434891, EPI_ISL_3434892, EPI_ISL_3434893, EPI_ISL_3434895, EPI_ISL_3434897, EPI_ISL_3434939, EPI_ISL_3434942, EPI_ISL_3539925, EPI_ISL_3539926, EPI_ISL_3539927, EPI_ISL_3539928 | | | |
| see above | Laboratorio Central de Saude Publica do Estado de Alagoas (LACEN/AL) | Laboratory of Respiratory Viruses and Measles, Oswaldo Cruz | Agatha Cristinne Prudencio; Agatha Soares; Alice Sampaio Rocha; Ana Carolina Mendonca; Anderson Brandao Leite; Anna Carolina Paixao; Elisa Cavalcante Pereira; Fernando Motta; Ighor Arantes; Ighor Leonardo Arantes Gomes; Luciana Appolinario; Marilda Siqueira on behalf of the Fiocruz COVID-19 Genomic Surveillance Network; Paola Resende; Renata Serrano Lopes; Taina Venas |

| | | | |
|---|---|---|---|
| | | Institute, FIOCRUZ | |
| EPI_ISL_2863647, EPI_ISL_2863648, EPI_ISL_2863651, EPI_ISL_2863653, EPI_ISL_2863655, EPI_ISL_2863656, EPI_ISL_2863658, EPI_ISL_2863660, EPI_ISL_2863662, EPI_ISL_2863664, EPI_ISL_2863666, EPI_ISL_2863712, EPI_ISL_2983218, EPI_ISL_2983339, EPI_ISL_2983340, EPI_ISL_2983342, EPI_ISL_2983343, EPI_ISL_2983344, EPI_ISL_2983345, EPI_ISL_2983346, EPI_ISL_2983347, EPI_ISL_2983348, EPI_ISL_2983349, EPI_ISL_2983350, EPI_ISL_2983351, EPI_ISL_2983352, EPI_ISL_2983353, EPI_ISL_2983354, EPI_ISL_2983355, EPI_ISL_2983356, EPI_ISL_2983357, EPI_ISL_2983359, EPI_ISL_2983360, EPI_ISL_2983361, EPI_ISL_2983362, EPI_ISL_2983363, EPI_ISL_2983364, EPI_ISL_2983365, EPI_ISL_2983366, EPI_ISL_2983367, EPI_ISL_2983368, EPI_ISL_2983369, EPI_ISL_2983370, EPI_ISL_2983371, EPI_ISL_2983372 | | | |
| see above | Laboratorio Central de Saude Publica do Estado de Santa Catarina (LACEN-SC) | Laboratory of Respiratory Viruses and Measles, Oswaldo Cruz Institute, FIOCRUZ | Agatha Cristinne Prudencio; Alice Sampaio Rocha; Ana Carolina Mendonca; Anna Carolina Paixao; Darcita Buerger Rovaris; Elisa Cavalcante Pereira; Fernando Motta; Ighor Leonardo Arantes Gomes; Luciana Appolinario; Marilda Siqueira on behalf of the Fiocruz COVID-19 Genomic Surveillance Network; Paola Resende; Renata Serrano Lopes; Sandra Bianchini Fernandes; Taina Venas |
| EPI_ISL_2983383, EPI_ISL_2983384, EPI_ISL_2983385, EPI_ISL_2983387, EPI_ISL_2983388, EPI_ISL_2983389, EPI_ISL_2983391, EPI_ISL_2983392, EPI_ISL_2983394, EPI_ISL_2983395, EPI_ISL_2983397, EPI_ISL_2983398, EPI_ISL_2983400, EPI_ISL_2983402, EPI_ISL_2983403, EPI_ISL_2983404, EPI_ISL_2983405, EPI_ISL_2983406, EPI_ISL_2983407, EPI_ISL_2983409, EPI_ISL_2983410, EPI_ISL_2983411, EPI_ISL_2983412, EPI_ISL_2983414, EPI_ISL_2983415, EPI_ISL_3190167, EPI_ISL_3190219, EPI_ISL_3190221, EPI_ISL_3190222, EPI_ISL_3190223, EPI_ISL_3190224, EPI_ISL_3190225, EPI_ISL_3235294, EPI_ISL_3235296, EPI_ISL_3235297, EPI_ISL_3235298, EPI_ISL_3235300, EPI_ISL_3235301, EPI_ISL_3235303, EPI_ISL_3434908, EPI_ISL_3434910, EPI_ISL_3434911, EPI_ISL_3434912, EPI_ISL_3434913, EPI_ISL_3434914, EPI_ISL_3434915, EPI_ISL_3434925, EPI_ISL_3434926, EPI_ISL_3434927, EPI_ISL_3434928, EPI_ISL_3434929, EPI_ISL_3434930, EPI_ISL_3434931, EPI_ISL_3434932, EPI_ISL_3434933, EPI_ISL_3434934, EPI_ISL_3434935, EPI_ISL_3434936, EPI_ISL_3434937, EPI_ISL_3434940, EPI_ISL_3539246, EPI_ISL_3539247, EPI_ISL_3539248, EPI_ISL_3539249, EPI_ISL_3539770, EPI_ISL_3539771 | | | |
| see above | Laboratorio Central de Saude Publica do Estado de Santa Catarina (LACEN/SC) | Laboratory of Respiratory Viruses and Measles, Oswaldo Cruz Institute, FIOCRUZ | Alice Sampaio Rocha; Ana Carolina Mendonca; Anna Carolina Paixao; Darcita Buerger Rovaris; Elisa Cavalcante Pereira; Fernando Motta; Luciana Appolinario; Marilda Siqueira on behalf of the Fiocruz COVID-19 Genomic Surveillance Network; Paola Resende; Renata Serrano Lopes; Sandra Bianchini Fernandes; Taina Venas |
| EPI_ISL_2863573, EPI_ISL_2863575, EPI_ISL_2863621, EPI_ISL_2863623, EPI_ISL_2863626, EPI_ISL_2863630, EPI_ISL_2863637, EPI_ISL_2863639, EPI_ISL_2863641, EPI_ISL_2863643, EPI_ISL_2863645, EPI_ISL_2863719, EPI_ISL_3803004, EPI_ISL_3803006, EPI_ISL_3803007, EPI_ISL_3803008, EPI_ISL_3803009, EPI_ISL_3803010, EPI_ISL_3803012, EPI_ISL_3803014, EPI_ISL_3803015, EPI_ISL_4061451, EPI_ISL_4061452, EPI_ISL_4061453 | | | |
| see above | Laboratorio Central de Saude Publica do Estado de Sergipe (LACEN/SE) | Laboratory of Respiratory Viruses and Measles, Oswaldo Cruz Institute, FIOCRUZ | Agatha Soares; Alice Sampaio Rocha; Ana Carolina Mendonca; Anna Carolina Paixao; Cliomar Alves dos Santos; Elisa Cavalcante Pereira; Fernando Motta; Ighor Arantes; Luciana Appolinario; Marilda Siqueira on behalf of the Fiocruz COVID-19 Genomic Surveillance Network; Paola Resende; Renata Serrano Lopes; Tainá Moreira Martins Venas |
| EPI_ISL_3434898, EPI_ISL_3434899, EPI_ISL_3434900, EPI_ISL_3434901, EPI_ISL_3434902, EPI_ISL_3434903, EPI_ISL_3434904, EPI_ISL_3434956, EPI_ISL_3434957, EPI_ISL_3434958, EPI_ISL_3434959, EPI_ISL_3434960, EPI_ISL_3434961, EPI_ISL_3434962, EPI_ISL_3434964, EPI_ISL_3434965, EPI_ISL_3434966, EPI_ISL_3434967, EPI_ISL_3434968, EPI_ISL_3434969, EPI_ISL_3434970, EPI_ISL_3434971, EPI_ISL_3434972, EPI_ISL_3434973, EPI_ISL_3434974, EPI_ISL_3434975, EPI_ISL_3434976, EPI_ISL_3434977, EPI_ISL_3434978, EPI_ISL_3434979, EPI_ISL_3434980, EPI_ISL_3434981, EPI_ISL_3435067, EPI_ISL_3539742, EPI_ISL_3539743, EPI_ISL_3539744, EPI_ISL_3539745, EPI_ISL_3539746, EPI_ISL_3539747, EPI_ISL_3539748, EPI_ISL_3539931 | | | |
| see above | Laboratorio Central de Saude Publica do Estado do Amapa (LACEN/AP) | Laboratory of Respiratory Viruses and Measles, Oswaldo Cruz Institute, FIOCRUZ | Agatha Cristinne Prudencio; Alice Sampaio Rocha; Ana Carolina Mendonca; Andreia Santos Costa; Anna Carolina Paixao; Anne Caroline da Silva Soledade; Elisa Cavalcante Pereira; Fernando Motta; Ighor Leonardo Arantes Gomes; Lindomar dos Anjos Silva; Luciana Appolinario; Marcia Socorro Pereira Cavalcante; Marilda Siqueira on behalf of the Fiocruz COVID-19 Genomic Surveillance Network; Paola Resende; Renata Serrano Lopes; Taina Venas |
| EPI_ISL_2375477, EPI_ISL_3045370, EPI_ISL_3045463, EPI_ISL_3045466, EPI_ISL_3045467, EPI_ISL_3045556, EPI_ISL_3190244, EPI_ISL_3190245, EPI_ISL_3190246, EPI_ISL_3190247, EPI_ISL_3190248, EPI_ISL_3190249, EPI_ISL_3190250, EPI_ISL_3190251, EPI_ISL_3190252, EPI_ISL_3190253, EPI_ISL_3434746, EPI_ISL_3434987, EPI_ISL_3434988, EPI_ISL_3434995, EPI_ISL_3434996, EPI_ISL_3434997, EPI_ISL_3434998, EPI_ISL_3435008, EPI_ISL_3435011, EPI_ISL_3435013, EPI_ISL_3435014, EPI_ISL_3435015, EPI_ISL_3435016, EPI_ISL_3435019, EPI_ISL_3435024, EPI_ISL_3435025, EPI_ISL_3435029, EPI_ISL_3435030, EPI_ISL_3435031, EPI_ISL_3435035, EPI_ISL_3435037, EPI_ISL_3435038, EPI_ISL_3435039, EPI_ISL_3827985, EPI_ISL_3827994, EPI_ISL_3827997, EPI_ISL_3828000, EPI_ISL_3828003, EPI_ISL_3828004, EPI_ISL_3828006, EPI_ISL_3828008, EPI_ISL_3828010, EPI_ISL_3828011 | | | |
| see above | Laboratorio Central de Saude Publica do Estado do Espirito Santo (LACEN/ES) | Laboratory of Respiratory Viruses and Measles, Oswaldo Cruz Institute, FIOCRUZ | Alice Sampaio Rocha; Ana Carolina Mendonca; Anna Carolina Paixao; Eilisa Cavalcante Pereira; Elisa Cavalcante Pereira; Fernando Motta; Luciana Appolinario; Marilda Siqueira on behalf of the Fiocruz COVID-19 Genomic Surveillance Network; Paola Resende; Renata Serrano Lopes; Rodrigo Ribeiro Rodrigues; Taina Venas |
| EPI_ISL_3190261 | Laboratorio Central de Saude Publica do Estado do Maranhao (LACEN-MA) | Laboratory of Respiratory Viruses and Measles, Oswaldo Cruz Institute, FIOCRUZ | Agatha Soares; Alice Sampaio Rocha; Ana Carolina Mendonca; Anna Carolina Paixao; Elisa Cavalcante Pereira; Fernando Motta; Ighor Arantes; Lidio Gonçalves Lima Neto; Luciana Appolinario; Marilda Siqueira on behalf of the Fiocruz COVID-19 Genomic Surveillance Network; Paola Resende; Renata Serrano Lopes; Tainá Venas |
| EPI_ISL_3913651, EPI_ISL_3913656, EPI_ISL_3913660, EPI_ISL_3913664, EPI_ISL_3913669, EPI_ISL_3913676, EPI_ISL_3913681, EPI_ISL_3913685, EPI_ISL_3913690, EPI_ISL_3913694, EPI_ISL_3913698, EPI_ISL_3913703, EPI_ISL_3913707, EPI_ISL_3913712, EPI_ISL_3913715, EPI_ISL_3913720, EPI_ISL_3913724, EPI_ISL_3913729, EPI_ISL_3913733, EPI_ISL_3913738, EPI_ISL_3913741, EPI_ISL_3913746, EPI_ISL_3913751, EPI_ISL_3913755, EPI_ISL_3913759, EPI_ISL_3913764, EPI_ISL_3913772, EPI_ISL_3913775, EPI_ISL_3913780, EPI_ISL_3913784, EPI_ISL_3913792, EPI_ISL_3913797, EPI_ISL_3913801, EPI_ISL_3913809, EPI_ISL_3913818, EPI_ISL_3913823, EPI_ISL_3913827, EPI_ISL_3913832, EPI_ISL_3913841, EPI_ISL_3913845, EPI_ISL_3913849, EPI_ISL_3913854, EPI_ISL_3913862, EPI_ISL_3913874, EPI_ISL_3913879, EPI_ISL_3913882, EPI_ISL_3913891, EPI_ISL_3913896, EPI_ISL_3913900, EPI_ISL_3913905, EPI_ISL_3913909, EPI_ISL_3913913, EPI_ISL_3913917, EPI_ISL_3913921, EPI_ISL_3913930, EPI_ISL_3913934, EPI_ISL_3913938, EPI_ISL_3913943, EPI_ISL_3913947, EPI_ISL_3913955, EPI_ISL_3913960, EPI_ISL_3913964, EPI_ISL_3913969, EPI_ISL_3913977, EPI_ISL_3913981, EPI_ISL_3913985, EPI_ISL_3913989, EPI_ISL_3913994, EPI_ISL_3914002, EPI_ISL_3914007, EPI_ISL_3914010, EPI_ISL_3914014, EPI_ISL_3914019, EPI_ISL_3914023, EPI_ISL_3914027, EPI_ISL_3914032, EPI_ISL_3914040, EPI_ISL_3914045, EPI_ISL_3914053, EPI_ISL_3914064, EPI_ISL_3914073, EPI_ISL_3914082, EPI_ISL_3914090, EPI_ISL_3914095, EPI_ISL_3914103, EPI_ISL_3914107, EPI_ISL_3914111, EPI_ISL_3914119, EPI_ISL_3914124, EPI_ISL_3914129, EPI_ISL_3914134, EPI_ISL_3914142, EPI_ISL_3914147, EPI_ISL_3914155, EPI_ISL_3914158, EPI_ISL_3914166, EPI_ISL_3914171, EPI_ISL_3914175, EPI_ISL_3914193, EPI_ISL_3914198, EPI_ISL_3914206, EPI_ISL_3914223, EPI_ISL_3914236, EPI_ISL_3914240, EPI_ISL_3914244, EPI_ISL_3914258, EPI_ISL_3914271, EPI_ISL_3914279, EPI_ISL_3914287, EPI_ISL_3914296, EPI_ISL_3914300, EPI_ISL_3914304, EPI_ISL_3914308, EPI_ISL_3914317, EPI_ISL_3914326, EPI_ISL_3914342, EPI_ISL_3914347, EPI_ISL_3914351, EPI_ISL_3914355, EPI_ISL_3914359, EPI_ISL_3914363, EPI_ISL_3914368, EPI_ISL_3914372, EPI_ISL_3914381, EPI_ISL_3914384, EPI_ISL_3914389, EPI_ISL_3914394, EPI_ISL_3914397, EPI_ISL_3914402, EPI_ISL_3914407, EPI_ISL_3914415, EPI_ISL_3914418, EPI_ISL_3914423, EPI_ISL_3914427, EPI_ISL_3914430, EPI_ISL_3914434 | | | |
| see above | Laboratorio Central de Saude Publica do Estado do Parana (Instituto de Biologia Molecular do Paraná (LACEN-PR) | Instituto Carlos Chagas - Fiocruz | Alessandra De Melo Aguiar; Andreia Akemi Suzukawa; Andréa Rodrigues Ávila; Bruno Dallagiovanna; Dalila Zanette; Eduardo Balsanelli; Emanuel Maltempi de Souza; Fabio Passetti; Fabricio Klerynton Marchini; Fábio de Oliveira Pedrosa; Guilherme Becker; Helisson Faoro; Hellen Geremias dos Santos; Irina Nastassja Riediger; Letusa Albrecht; Lucas Blanes; Luis Gustavo Morello; Lysangela Ronalte Alves; Maria do Carmo Debur; Mauro de Medeiros Oliveira; Michelle Orane Schemberger; Paola Cristina Resende; Sheila Cristina Nardeli; Tiago Gräf; Valter Antônio de Baura |
| EPI_ISL_2645445, EPI_ISL_2645446, EPI_ISL_2645448, EPI_ISL_2645449, EPI_ISL_2645450, EPI_ISL_2645451, EPI_ISL_2645452, EPI_ISL_2645453, EPI_ISL_2645454, EPI_ISL_2645455, EPI_ISL_2645456, EPI_ISL_2645457, EPI_ISL_2645459, EPI_ISL_2645460, EPI_ISL_2645462, EPI_ISL_2645464, EPI_ISL_2645467, EPI_ISL_2645468, EPI_ISL_2645469, EPI_ISL_2645473, EPI_ISL_2645475, EPI_ISL_2645476, EPI_ISL_2645477, EPI_ISL_2645478, EPI_ISL_2645479, EPI_ISL_2645481, EPI_ISL_2645483, EPI_ISL_2645486, EPI_ISL_2645493, EPI_ISL_2645496, EPI_ISL_2645502, EPI_ISL_2983426, EPI_ISL_2983427, EPI_ISL_2983428, EPI_ISL_2983430, EPI_ISL_2983431, EPI_ISL_2983433, EPI_ISL_2983434, EPI_ISL_2983435, EPI_ISL_2983438, EPI_ISL_2983439, EPI_ISL_2983440, EPI_ISL_2983441, EPI_ISL_2983442, EPI_ISL_2983443, EPI_ISL_2983444, EPI_ISL_2983445, EPI_ISL_2983446, EPI_ISL_2983449, EPI_ISL_2983451, EPI_ISL_3190190, EPI_ISL_3190191, EPI_ISL_3190192, EPI_ISL_3190266, EPI_ISL_3190267, EPI_ISL_3190268, EPI_ISL_3190360, EPI_ISL_3190361, EPI_ISL_3190362, EPI_ISL_3190363, EPI_ISL_3190364, EPI_ISL_3190365, EPI_ISL_3190366, EPI_ISL_3190367, EPI_ISL_3190369, EPI_ISL_3190371, EPI_ISL_3190372, EPI_ISL_3190373, EPI_ISL_3190374, EPI_ISL_3190375, EPI_ISL_3190376, EPI_ISL_3190377, EPI_ISL_3190378, EPI_ISL_3190379, EPI_ISL_3190380, EPI_ISL_3190381, EPI_ISL_3190382, EPI_ISL_3190383, EPI_ISL_3190384, EPI_ISL_3190385, EPI_ISL_3190387, EPI_ISL_3190388, EPI_ISL_3190389, EPI_ISL_3190390, EPI_ISL_3190391, EPI_ISL_3435048 | | | |
| see above | Laboratorio Central de Saude Publica do Estado do Parana (LACEN/PR) | Laboratory of Respiratory Viruses and Measles, Oswaldo Cruz Institute, FIOCRUZ | Agatha Soares; Alice Sampaio Rocha; Ana Carolina Mendonca; Anna Carolina Paixao; Elisa Cavalcante Pereira; Fernando Motta; Ighor Arantes; Ighor Leonardo Arantes; Irina Riediger; Luciana Appolinario; Marilda Siqueira on behalf of the Fiocruz COVID-19 Genomic Surveillance Network; Paola Resende; Renata Serrano Lopes; Taina Venas |
| EPI_ISL_3190392, EPI_ISL_3190393, EPI_ISL_3190394, EPI_ISL_3190395, EPI_ISL_3190396, EPI_ISL_3190397, EPI_ISL_3190398, EPI_ISL_3190399, EPI_ISL_3190400, EPI_ISL_3190401, EPI_ISL_3190404, EPI_ISL_3235269, EPI_ISL_3235272, EPI_ISL_3235273, EPI_ISL_3235274, EPI_ISL_3235275 | | | |
| see above | Laboratorio Central de Saude Publica do Estado do Rio Grande do Norte (LACEN/RN) | Laboratory of Respiratory Viruses and Measles, Oswaldo Cruz Institute, FIOCRUZ | Agatha Soares; Alice Sampaio Rocha; Ana Carolina Mendonca; Ana Paula Ferreira Costa; Anna Carolina Paixao; Antonnyo Palmielly Diogenes Lima; Aurélio de Oliveira Bento; Elisa Cavalcante Pereira; Fernando Motta; Gessika Brenna Costa Alves; Heglayne Pereira Vital da Silva; Iago de Souza Gomes; Ighor Arantes; Isabelle Cristina Clemente dos Santos; Janaina Sonale Cavalcante Nogueira de Oliveira; Jayra Juliana Paiva Alves Abrantes; Jonas José da Silva; Luciana Appolinario; Marilda Siqueira on behalf of the Fiocruz COVID-19 Genomic Surveillance Network; Paola Resende; Renata Serrano Lopes; Taina Venas; Themis Rocha de Souza; Vítor Gabriel Saldanha Fernandes |
| EPI_ISL_2603441, EPI_ISL_2603529, EPI_ISL_2603530, EPI_ISL_2603531, EPI_ISL_2603532, EPI_ISL_2603533, EPI_ISL_2603534, EPI_ISL_2603535, EPI_ISL_2982762, EPI_ISL_2982763, EPI_ISL_2982764, EPI_ISL_2982765, EPI_ISL_2982766, EPI_ISL_2982767, EPI_ISL_2982768, EPI_ISL_2982775, EPI_ISL_2982776, EPI_ISL_2982777, EPI_ISL_2982781, EPI_ISL_2982784, EPI_ISL_2982788, EPI_ISL_2982792, EPI_ISL_2982795, EPI_ISL_2982796, EPI_ISL_2982797, EPI_ISL_2982798, EPI_ISL_2982800, EPI_ISL_2982801, EPI_ISL_2982802, EPI_ISL_2982803, EPI_ISL_2982807, EPI_ISL_2982808, EPI_ISL_2982811, EPI_ISL_2982813, EPI_ISL_2982814, EPI_ISL_2982815, EPI_ISL_2982816, EPI_ISL_2982818, EPI_ISL_2982819, EPI_ISL_2982820, EPI_ISL_2982821, EPI_ISL_2982824, EPI_ISL_2982826, EPI_ISL_2982827, EPI_ISL_2982828, EPI_ISL_2982829, EPI_ISL_2982830, EPI_ISL_3190303, EPI_ISL_3190305, EPI_ISL_3190406, EPI_ISL_3190407, EPI_ISL_3235278, EPI_ISL_3235280, EPI_ISL_3235281, EPI_ISL_3235282, EPI_ISL_3235283, EPI_ISL_3235289, EPI_ISL_3235290, EPI_ISL_3434907, EPI_ISL_3539241, EPI_ISL_3539242, EPI_ISL_3539243, EPI_ISL_3539244, EPI_ISL_3539767, EPI_ISL_3539768, EPI_ISL_3539769, EPI_ISL_3539840, EPI_ISL_3539841, EPI_ISL_3539842, EPI_ISL_3539843, EPI_ISL_3539845, EPI_ISL_3539846, EPI_ISL_3539847, EPI_ISL_3539848, EPI_ISL_3539849, EPI_ISL_3539850, EPI_ISL_3539851, EPI_ISL_3539852, EPI_ISL_3539853, EPI_ISL_3539855, EPI_ISL_3539856, EPI_ISL_3539858, EPI_ISL_3539859, EPI_ISL_3539861, EPI_ISL_3539863, EPI_ISL_3539864, EPI_ISL_3539865, EPI_ISL_3539867, EPI_ISL_3539868, EPI_ISL_3539869, EPI_ISL_3539870, EPI_ISL_3539871, EPI_ISL_3539872, EPI_ISL_3539873, EPI_ISL_3539874, EPI_ISL_3539875, EPI_ISL_3539876, EPI_ISL_3539877, EPI_ISL_3539878, EPI_ISL_3539880, EPI_ISL_3539881, EPI_ISL_3539882, EPI_ISL_3539883, EPI_ISL_3539884, EPI_ISL_3539885, EPI_ISL_3539886, EPI_ISL_3539887, EPI_ISL_3539888, EPI_ISL_3539891, EPI_ISL_3539892, EPI_ISL_3539893, EPI_ISL_3539894, EPI_ISL_3539895, EPI_ISL_3539896, EPI_ISL_3539897, EPI_ISL_3539898, EPI_ISL_3539901, EPI_ISL_3539902, EPI_ISL_3539903, EPI_ISL_3539904, EPI_ISL_3539905, EPI_ISL_3539938, EPI_ISL_3539939, EPI_ISL_3539940, EPI_ISL_3539941, EPI_ISL_3539943, EPI_ISL_3539944, EPI_ISL_3539945, EPI_ISL_3539946, EPI_ISL_3539947, EPI_ISL_3539948, EPI_ISL_3539949, EPI_ISL_3539950, EPI_ISL_3539951, EPI_ISL_3539952, EPI_ISL_3539953, EPI_ISL_3539954, EPI_ISL_3539956, EPI_ISL_3539957, EPI_ISL_3539958, EPI_ISL_3539961, EPI_ISL_3539962, EPI_ISL_3539963, EPI_ISL_3539964, EPI_ISL_3539965, EPI_ISL_3539966, EPI_ISL_3539967, EPI_ISL_3539968, EPI_ISL_3539970, EPI_ISL_3539971, EPI_ISL_3539972, EPI_ISL_3539973, EPI_ISL_3539974, EPI_ISL_3539975, EPI_ISL_3539976, EPI_ISL_3539977, EPI_ISL_3539978, EPI_ISL_3539979, EPI_ISL_3539980, EPI_ISL_3539985, EPI_ISL_3539986, EPI_ISL_3539988, EPI_ISL_3539989, EPI_ISL_3539990, EPI_ISL_3539991, EPI_ISL_3539992, EPI_ISL_3539993, EPI_ISL_3539994, EPI_ISL_3539995, EPI_ISL_3539996, EPI_ISL_3539997, EPI_ISL_3832410, EPI_ISL_3832411, EPI_ISL_3832412, EPI_ISL_3832413, EPI_ISL_3832414, EPI_ISL_3832415, EPI_ISL_3832416, EPI_ISL_4061391 | | | |
| see above | Laboratorio Central de Saude Publica do Estado do Rio Grande do Sul (LACEN-RS) | Laboratory of Respiratory Viruses and Measles, Oswaldo Cruz Institute, FIOCRUZ | Agatha Cristinne Soares; Alice Sampaio Rocha; Ana Carolina Mendonca; Anderson Brandao Leite; Anna Carolina Paixao; Elisa Cavalcante Pereira; Fernando Motta; Ighor Leonardo Arantes; Luciana Appolinario; Marilda Siqueira on behalf of the Fiocruz COVID-19 Genomic Surveillance Network; Paola Resende; Renata Serrano Lopes; Richard Salvato; Taina Venas; Tatiana Schaffer Gregianini |
| EPI_ISL_2863584, EPI_ISL_2863585, EPI_ISL_2863586, EPI_ISL_2863587, EPI_ISL_2863588, EPI_ISL_2863602, EPI_ISL_2863604, EPI_ISL_2863667, EPI_ISL_2863669, EPI_ISL_2863671, EPI_ISL_2863673, EPI_ISL_2863675, EPI_ISL_2863677, EPI_ISL_2863679, EPI_ISL_2863680, EPI_ISL_2863698, EPI_ISL_2863700, EPI_ISL_2863702, EPI_ISL_2863704, EPI_ISL_2863717, EPI_ISL_2863721, EPI_ISL_2863723, EPI_ISL_2863727, EPI_ISL_2982742, EPI_ISL_2982744, EPI_ISL_2982745, EPI_ISL_2982746, EPI_ISL_2982747, EPI_ISL_2982748, EPI_ISL_2982749, EPI_ISL_2982751, EPI_ISL_2982752, EPI_ISL_2982754, EPI_ISL_2982756, EPI_ISL_2983214, EPI_ISL_2983215, EPI_ISL_2983216 | | | |
| see above | Laboratorio Central de Saude Publica do Estado do Rio de Janeiro (LACEN-RJ) | Laboratory of Respiratory Viruses and Measles, Oswaldo Cruz Institute, FIOCRUZ | Agatha Cristinne Prudencio; Alice Sampaio Rocha; Ana Carolina Mendonca; Andrea Cony Cavalcanti; Anna Carolina Paixao; Elisa Cavalcante Pereira; Fernando Motta; Ighor Leonardo Arantes Gomes; Luciana Appolinario; Marilda Siqueira on behalf of the Fiocruz COVID-19 Genomic Surveillance Network; Paola Resende; Renata Serrano Lopes; Taina Venas |
| EPI_ISL_3434906 | Laboratorio Central de Saude Publica do Estado do Rio de Janeiro (LACEN/RJ) | Laboratory of Respiratory Viruses and Measles, Oswaldo Cruz Institute, FIOCRUZ | Agatha Cristinne Prudencio Soares; Alice Sampaio Rocha; Ana Carolina Mendonca; Andrea Cony Cavalcanti; Anna Carolina Paixao; Elisa Cavalcante Pereira; Fernando Motta; Ighor Leonardo Arantes Gomes; Luciana Appolinario; Marilda Siqueira on behalf of the Fiocruz COVID-19 Genomic Surveillance Network; Paola Resende; Renata Serrano Lopes; Taina Venas |
| EPI_ISL_3235304, EPI_ISL_3235305, EPI_ISL_3235306, EPI_ISL_3235307, EPI_ISL_3235308, EPI_ISL_3235313 | Laboratorio Central de Saude Publica do Estado do Tocantins (LACEN/TO) | Laboratory of Respiratory Viruses and Measles, Oswaldo Cruz Institute, FIOCRUZ | Agatha Soares; Alice Sampaio Rocha; Ana Carolina Mendonca; Anna Carolina Paixao; Elisa Cavalcante Pereira; Fernando Motta; Ighor Arantes; Jucimaria Dantas Galvao; Luciana Appolinario; Marilda Siqueira on behalf of the Fiocruz COVID-19 Genomic Surveillance Network; Paola Resende; Renata Serrano Lopes; Taina Venas |
| EPI_ISL_3033064, EPI_ISL_3033065, EPI_ISL_3033068, EPI_ISL_3033071, EPI_ISL_3033072, EPI_ISL_3033074, EPI_ISL_3033078, EPI_ISL_3033079, EPI_ISL_3260096, EPI_ISL_3260097, EPI_ISL_3260098, EPI_ISL_3260099, EPI_ISL_3260100, EPI_ISL_3260104, EPI_ISL_3260105 | | | |
| see above | Laboratorio Central de la Ciudad de Santa Fe | Grupo de Genómica y Bioinformática del Instituto de Investigación de la Cadena Láctea CONICET-INTA on behalf of 'Proyecto Argentino Interinstitucional de genomica de SARS-CoV-2' (PAIS Consortium) | AF; Amadio; C; Eberhardt; G; Irazoqui; JM; MF; Mugna; Ojeda; Pastor; Rompato; V |
| EPI_ISL_2940274, EPI_ISL_2940275, EPI_ISL_2940285, EPI_ISL_2940290 | Laboratorio Central, Ministerio de Salud Córdoba | Instituto de Patologia Vegetal (CIAP-INTA) on behalf of 'Proyecto Argentino Interinstitucional de genomica de SARS-CoV-2' (PAIS Consortium) | Barbas, G.; Castro, G.; Debat, HJ.; FD; Fernández; M; M.B.; Marquez, N.; Pisano; Re, V. |
| EPI_ISL_2662459 | Laboratorio HUB -Azienda Ospedaliero Universitaria - AOU – Cagliari | Laboratorio SPOKE Biologia Molecolare -Azienda Ospedaliero Universitaria - AOU – Cagliari | Alessandra Scano; Ferdinando Coghe; Germano Orrù; Miriam Loddo; Riccardo Cappai; Sara Fais; Valentina Medda |
| EPI_ISL_3527820, EPI_ISL_3527822, EPI_ISL_3835365, | Laboratorio Nacional de Salud | Asociación de Salud Integral / Clínica Familiar "Luis Ángel García" | Ana S. Gonzalez-Reiche; Claudia Rangel; Danicela Mercado; Eduardo Arathon; Hilda Ruiz; Luis Aguirre; Luis Rivas; Narda Medina; Oscar Bonilla; Osmar Gamboa. |

| | | | |
|---|---|---|---|
| EPI_ISL_3835378, EPI_ISL_3835379, EPI_ISL_3835385 | | | |
| EPI_ISL_3275241, EPI_ISL_3275242, EPI_ISL_4029319 | Laboratorio Nacional de Salud, Ministerio de Salud Publica y Asistencia Social | Genomics and Proteomics Departament, Gorgas Memorial Institute For Health Studies | Alexander A Martinez; Ambar Moreno; Claudia Estrada; Claudia Gonzalez V; César Roberto Conde Pereira; Jessica Gondola; Leyda Abrego; Marlene Castillo; Melissa Gaitan; Oris Chavarria |
| EPI_ISL_2777453, EPI_ISL_3050307, EPI_ISL_4030314, EPI_ISL_4030315, EPI_ISL_4030316, EPI_ISL_4030330, EPI_ISL_4030332, EPI_ISL_4030338, EPI_ISL_4030344, EPI_ISL_4030345, EPI_ISL_4030356, EPI_ISL_4030359, EPI_ISL_4030365 | | | |
| see above | Laboratorio de Ecologia de Doencas Transmissiveis na Amazonia, Instituto Leonidas e Maria Deane - Fiocruz Amazonia | Laboratorio de Ecologia de Doencas Transmissiveis na Amazonia, Instituto Leonidas e Maria Deane - Fiocruz Amazonia | Alex Martins; André Corado; Debora Duarte; Fabiola Mendonça da Silva Chui; Felipe Naveca; Fernanda Nascimento; Fernando Fonseca de Almeida Val; George Silva; Gisely Cardoso de Melo; Karina Pessoa; Luciana Gonçalves; Marcus Vinicius Guimarães de Lacerda; Maria Júlia Brandão; Maria Paula Gomes Mourão; Mariana Xavier Simão; Matilde Mejía; Michele Jesus; Valdinete Nascimento; Vanderson de Souza Sampaio; Victor Souza; Ágatha Costa |
| EPI_ISL_3133693, EPI_ISL_3370436, EPI_ISL_3370440, EPI_ISL_3370456, EPI_ISL_3642594 | Laboratorio de Infectologia y Virologia Molecular | Laboratory of Molecular Virology, School of Medicine, Pontificia Universidad Catolica de Chile | Alejandro Bhrun; Ana Maria Contreras; Andres E. Munoz-Marcos; Carlos Palma; Catalina Pardo-Roa; Constanza Maldonado; Constanza Martinez-Valdevenito; Eileen Serrano; Erick Salinas; Estefany Poblete; Francisco Melo; Jennifer Angulo; Jorge Levican; Leonardo I. Almonacid; M. Belen Leyton; Magdalena Vera; Marcela Ferres; Maria Jose Avendano; Rafael A. Medina; Tamara Garcia-Salum |
| EPI_ISL_2854154, EPI_ISL_3134255 | Laboratorio de Medicina Molecular, Universidad de Magallanes | Centro Asistencial Docente y de Investigacion, Universidad de Magallanes | Carolina Pérez; Constanza Ceroni; Diego Alvarez; Inés Cid; Jacqueline Aldridge; Jorge González; Juan Ríos; Marcelo Navarrete; Roberto Uribe-Paredes |
| EPI_ISL_3048979, EPI_ISL_3048980, EPI_ISL_3048981, EPI_ISL_3048982, EPI_ISL_3048983, EPI_ISL_3048984, EPI_ISL_3048985, EPI_ISL_3048986, EPI_ISL_3048987, EPI_ISL_3048988, EPI_ISL_3048989, EPI_ISL_3048990, EPI_ISL_3048991, EPI_ISL_3048992, EPI_ISL_3048993, EPI_ISL_3048994, EPI_ISL_3048995, EPI_ISL_3048996, EPI_ISL_3048997, EPI_ISL_3048998, EPI_ISL_3049000, EPI_ISL_3049001, EPI_ISL_3049002, EPI_ISL_3049003, EPI_ISL_3049004, EPI_ISL_3049005, EPI_ISL_3049007, EPI_ISL_3049008, EPI_ISL_3049010, EPI_ISL_3049012, EPI_ISL_3049016, EPI_ISL_3049017, EPI_ISL_3049018, EPI_ISL_3049019, EPI_ISL_3049020, EPI_ISL_3049023, EPI_ISL_3049025, EPI_ISL_3049027, EPI_ISL_3049029, EPI_ISL_3055529, EPI_ISL_3055530, EPI_ISL_3055532, EPI_ISL_3761573, EPI_ISL_3761586, EPI_ISL_3761587, EPI_ISL_3761588, EPI_ISL_3761589, EPI_ISL_3761597, EPI_ISL_3761599, EPI_ISL_3761605, EPI_ISL_3761617, EPI_ISL_3761637, EPI_ISL_3761638, EPI_ISL_3761639, EPI_ISL_3761640, EPI_ISL_3761641, EPI_ISL_3761642, EPI_ISL_3761643, EPI_ISL_3761644, EPI_ISL_3761647, EPI_ISL_3761648, EPI_ISL_3761649, EPI_ISL_3761650, EPI_ISL_3761651, EPI_ISL_3761652, EPI_ISL_3761653, EPI_ISL_3761654, EPI_ISL_3761655, EPI_ISL_3761656, EPI_ISL_3761657, EPI_ISL_3761659, EPI_ISL_3761661, EPI_ISL_3761662, EPI_ISL_3761663, EPI_ISL_3761664, EPI_ISL_3761665, EPI_ISL_3761666, EPI_ISL_3761667, EPI_ISL_3761668, EPI_ISL_3761669, EPI_ISL_3761670, EPI_ISL_3761671, EPI_ISL_3761672, EPI_ISL_3761673, EPI_ISL_3761675, EPI_ISL_3761676, EPI_ISL_3761677, EPI_ISL_3761678, EPI_ISL_3761679, EPI_ISL_3761680, EPI_ISL_3761681, EPI_ISL_3761684, EPI_ISL_3761685, EPI_ISL_3761686, EPI_ISL_3761687, EPI_ISL_3761688, EPI_ISL_3761689, EPI_ISL_3761690, EPI_ISL_3761693, EPI_ISL_3761694, EPI_ISL_3761695, EPI_ISL_3761696, EPI_ISL_3761697, EPI_ISL_3761699, EPI_ISL_3761754, EPI_ISL_3761755, EPI_ISL_3761756, EPI_ISL_3761757, EPI_ISL_3761758, EPI_ISL_3761759, EPI_ISL_3761761, EPI_ISL_3761767 | | | |
| see above | Laboratorio de Pesquisa em Virologia, FAMERP, SJRP | Laboratorio de Pesquisa em Virologia, FAMERP, SJRP | Beatriz de Carvalho Marques; Cecília Artico Banho; Cíntia Bittar; Fábio Sossai Possebon; Guilherme Campos; Helena Lage Ferreira; Jorge A. Petroli Marchesi; João Pessoa Araújo Jr.; Leila Sabrina Ullmann; Lívia Sacchetto; Maisa C. Pereira Parra; Marília Moraes; Maurício L. Nogueira.; Paula Rahal; Paulo Inacio da Costa |
| EPI_ISL_3672424, EPI_ISL_3672425, EPI_ISL_3672427, EPI_ISL_3672429, EPI_ISL_3672430, EPI_ISL_3672431, EPI_ISL_3672432, EPI_ISL_3672433, EPI_ISL_3672435, EPI_ISL_3672436, EPI_ISL_3672439, EPI_ISL_3826386, EPI_ISL_3826387, EPI_ISL_3826390, EPI_ISL_3826391, EPI_ISL_3826416, EPI_ISL_3826418, EPI_ISL_3826419, EPI_ISL_3826421, EPI_ISL_3826422, EPI_ISL_3826423, EPI_ISL_3826432, EPI_ISL_3826435, EPI_ISL_3826438, EPI_ISL_3826439, EPI_ISL_3826443, EPI_ISL_3826444, EPI_ISL_3991565, EPI_ISL_4052705, EPI_ISL_4052707, EPI_ISL_4052708, EPI_ISL_4052709, EPI_ISL_4052710, EPI_ISL_4052727, EPI_ISL_4052731, EPI_ISL_4080441, EPI_ISL_4080442, EPI_ISL_4080443, EPI_ISL_4080444, EPI_ISL_4080445, EPI_ISL_4080446, EPI_ISL_4080447, EPI_ISL_4080448, EPI_ISL_4080450, EPI_ISL_4080456, EPI_ISL_4080457, EPI_ISL_4080458, EPI_ISL_4080459, EPI_ISL_4080460, EPI_ISL_4080461, EPI_ISL_4080462, EPI_ISL_4080464, EPI_ISL_4080466, EPI_ISL_4080467, EPI_ISL_4080468, EPI_ISL_4080469, EPI_ISL_4080470, EPI_ISL_4080472, EPI_ISL_4080473, EPI_ISL_4080476, EPI_ISL_4080477, EPI_ISL_4080478, EPI_ISL_4080479, EPI_ISL_4080482, EPI_ISL_4080485, EPI_ISL_4080486, EPI_ISL_4080487, EPI_ISL_4080488, EPI_ISL_4080491, EPI_ISL_4080492, EPI_ISL_4080493, EPI_ISL_4080495, EPI_ISL_4080502, EPI_ISL_4080503, EPI_ISL_4080506, EPI_ISL_4080511, EPI_ISL_4080512, EPI_ISL_4080514, EPI_ISL_4080515, EPI_ISL_4080516, EPI_ISL_4080518, EPI_ISL_4080521, EPI_ISL_4080522, EPI_ISL_4080523, EPI_ISL_4080524, EPI_ISL_4080525, EPI_ISL_4080526, EPI_ISL_4080528, EPI_ISL_4080529, EPI_ISL_4080530, EPI_ISL_4080532, EPI_ISL_4080536, EPI_ISL_4080538, EPI_ISL_4080539, EPI_ISL_4080540, EPI_ISL_4080541, EPI_ISL_4080542, EPI_ISL_4080543, EPI_ISL_4080544, EPI_ISL_4080545, EPI_ISL_4080550, EPI_ISL_4080552, EPI_ISL_4080554, EPI_ISL_4080555 | | | |
| see above | Laboratorio de Referencia Nacional de Virus Respiratorios. Centro Nacional de Salud Publica. Instituto Nacional de Salud Peru. | Laboratorio de Referencia Nacional de Virus Respiratorios. Centro Nacional de Salud Publica. Instituto Nacional de Salud Peru. | Carlos Padilla Rojas; Henri Bailon Calderon; Iris Silva Molina; Joseph Huayra Niquen; Lely Solari Zerpa; Luis Barcena Flores; Marco Galarza Perez; Nancy Rojas Serrano; Nieves Sevilla Castañeda; Omar Caceres Rey; Orson Mestanza Millones; Princesa Medrano Alhuay; Priscila Lope Pari; Sandra Morales Ruiz; Sara Gordillo Vilchez; Steve Acedo Lazo; Veronica Hurtado Vela; Victor Jimenez Vasquez; Wendy Lizarraga Olivares |
| EPI_ISL_2608291, EPI_ISL_2812650, EPI_ISL_2812707, EPI_ISL_2812709, EPI_ISL_3076938, EPI_ISL_3076940 | Laboratorio de Virología HUCA | Laboratorio de Virología HUCA | Abreu F; Alvarez-Arguelles ME; Boga JA; Castelló C; Costales I; Coto E; Gómez de Oña J; Martín-Rodríguez G; Melón S; Perez-Martínez Z; Rojo S; Sandoval M |
| EPI_ISL_2544205, EPI_ISL_2708965, EPI_ISL_2708968, EPI_ISL_2918882 | Laboratorio di Microbiologia | Laboratorio di Microbiologia | Martinetti Lucchini Gladys; Valeria Spina |
| EPI_ISL_2765565 | Laboratorio di Riferimento Regionale della Sicilia Occidentale per l'Emergenza COVID-19 | Laboratorio di Riferimento Regionale della Sicilia Occidentale per l'Emergenza COVID-19 | Carmelo Massimo Maida; Claudio Costantino; Daniela Di Naro; Fabio Tramuto; Francesco Vitale; Giorgio Graziano; Giulia Randazzo; Vincenzo Restivo; Walter Mazzucco |
| EPI_ISL_2680918, EPI_ISL_2680925, EPI_ISL_2987688, EPI_ISL_2987689, EPI_ISL_2987691, EPI_ISL_2987692 | Laboratorios Delia Barraza | Laboratorio nacional-LANIIA-CIAD | ; Alejandra García-Gasca; Alejandra Hernández-Terán; Alejandro Sánchez-Flores; Alfredo Herrera-Estrella; Alicia Ocaña-Mondragón; Andreu Comas-García; Angel Gustavo Salas-Lais; Antonio Loza Román; Bernardo Martínez-Miguel; Blanca Taboada; Brenda Irasema Maldonado-Meza; Bruno Gómez-Gil; Carla Ivón Herrera-Najera; Carlos F. Arias; Celia Boukadida; Clara Esperanza Santacruz-Tinoco; Claudia Soto Félix; Concepción Grajales-Muñiz; Consorcio Mexicano de Vigilancia Genómica (CoViGen-Mex). Authors (in alphabetical order): Julio Elias Alvarado-Yaah; Cristóbal Cháidez-Quiróz; Célida Duque Molina; Célida Martínez- Rodríguez; Daniel Fregoso-Rueda; Daniel Lira Morales; Eduardo Becerril-Vargas; Fernando Fontove-Herrera; Fidencio Mejía-Nepomuceno; Francisco Pulido; Gloria Elena Espinosa-Ayala; Gloria María Molina-Salinas; Gloria Vazquez; Hector Esteban Paz-Juárez; Hector Montoya-Fuentes; Helen Haydee Fernanda Ramírez-Plascencia; Irvin González-López; Jean Pierre González; Jesús Hernández; Joel Armando Vázquez-Pérez. Laboratorios Delia Barraza: María Delia Barraza Sámano; Jorge Salas-Hernández; José Antonio Enciso-Moreno; José Arturo Martínez-Orozco; José Esteban Muñoz-Medina; José de Jesús Nuñez-Contreras; Juan Bautista Chale-Dzul; Julissa Enciso-Ibarra; Luis Alberto Ochoa-Carrera; Margarita Matías-Florentino; Mario Mújica-Sánchez; Marissa Perez-Garcia; María Guadalupe Santiago-Mauricio; María Guadalupe de Jesús Mireles-Rivera; Melissa García Angulo.; Nelly Sélem-Mojica; Pavel Isa; Ricardo Ciria Merce; Ricardo Grande; Rosa María Gutiérrez Rios; Santiago Ávila-Ríos; Selene Zárate; Susana Lopez; Verónica Mata-Haro; Victor Eduardo García-Arias; Víctor Hugo Borja-Aburto |
| EPI_ISL_2399380, EPI_ISL_2399399, EPI_ISL_2399403, EPI_ISL_2480207, EPI_ISL_2480218, EPI_ISL_2480220, EPI_ISL_2480232, EPI_ISL_2480233, EPI_ISL_2480272, EPI_ISL_2480312, EPI_ISL_2480368, EPI_ISL_2480378, EPI_ISL_2480380, EPI_ISL_2480382, EPI_ISL_2480402, EPI_ISL_2480404, EPI_ISL_2480418, EPI_ISL_2480509, EPI_ISL_2480522, EPI_ISL_2480565, EPI_ISL_2480818, EPI_ISL_2480858, EPI_ISL_2480859, EPI_ISL_2480860, EPI_ISL_2480861, EPI_ISL_2480922, EPI_ISL_2480923, EPI_ISL_2480924, EPI_ISL_2480944, EPI_ISL_2480976, EPI_ISL_2481145, EPI_ISL_2481220, EPI_ISL_2481221, EPI_ISL_2481224, EPI_ISL_2481225, EPI_ISL_2481252, EPI_ISL_2481293, EPI_ISL_2481301, EPI_ISL_2481445, EPI_ISL_2481448, EPI_ISL_2481449, EPI_ISL_2481472, EPI_ISL_2481485, EPI_ISL_2481554, EPI_ISL_2481565, EPI_ISL_2481580, EPI_ISL_2481612, EPI_ISL_2481647, EPI_ISL_2481651, EPI_ISL_2481652, EPI_ISL_2481658, EPI_ISL_2481698, EPI_ISL_2481706, EPI_ISL_2481710, EPI_ISL_2481736, EPI_ISL_2481748, EPI_ISL_2481761, EPI_ISL_2481764, EPI_ISL_2481838, EPI_ISL_2481847, EPI_ISL_2481856, EPI_ISL_2481879, EPI_ISL_2481939, EPI_ISL_2481961, EPI_ISL_2481962, EPI_ISL_2481964, EPI_ISL_2481965, EPI_ISL_2481967, EPI_ISL_2481968, EPI_ISL_2481969, EPI_ISL_2481970, EPI_ISL_2482020, EPI_ISL_2482027, EPI_ISL_2482037, EPI_ISL_2482038, EPI_ISL_2482039, EPI_ISL_2482040, EPI_ISL_2482045, EPI_ISL_2482048, EPI_ISL_2482050, EPI_ISL_2482055, EPI_ISL_2482057, EPI_ISL_2482078, EPI_ISL_2482118, EPI_ISL_2482127, EPI_ISL_2482130, EPI_ISL_2482138, EPI_ISL_2482175, EPI_ISL_2482185, EPI_ISL_2482192, EPI_ISL_2482215, EPI_ISL_2482216, EPI_ISL_2482222, EPI_ISL_2482252, EPI_ISL_2482253, EPI_ISL_2482255, EPI_ISL_2611122, EPI_ISL_2611124, EPI_ISL_2611135, EPI_ISL_2611136, EPI_ISL_2611147, EPI_ISL_2611151, EPI_ISL_2611183, EPI_ISL_2611187, EPI_ISL_2611190, EPI_ISL_2611194, EPI_ISL_2611220, EPI_ISL_2611229, EPI_ISL_2611232, EPI_ISL_2611233, EPI_ISL_2611238, EPI_ISL_2611239, EPI_ISL_2611240, EPI_ISL_2611242, EPI_ISL_2611281, EPI_ISL_2611301, EPI_ISL_2611304, EPI_ISL_2611313, EPI_ISL_2611321, EPI_ISL_2611324, EPI_ISL_2611332, EPI_ISL_2611334, EPI_ISL_2611344, EPI_ISL_2611349, EPI_ISL_2611364, EPI_ISL_2611372, EPI_ISL_2611384, EPI_ISL_2611392, EPI_ISL_2611397, EPI_ISL_2611398, EPI_ISL_2611418, EPI_ISL_2611428, EPI_ISL_2611447, EPI_ISL_2611482, EPI_ISL_2611488, EPI_ISL_2611512, EPI_ISL_2611516, EPI_ISL_2611539, EPI_ISL_2611552, EPI_ISL_2611555, EPI_ISL_2611558, EPI_ISL_2611561, EPI_ISL_2611592, EPI_ISL_2611593, EPI_ISL_2611607, EPI_ISL_2611626, EPI_ISL_2611627, EPI_ISL_2611631, EPI_ISL_2611645, EPI_ISL_2611646, EPI_ISL_2611656, EPI_ISL_2611767, EPI_ISL_2611812, EPI_ISL_2611823, EPI_ISL_2872170, EPI_ISL_2872173, EPI_ISL_2872187, EPI_ISL_2872208, EPI_ISL_2872299, EPI_ISL_2872305, EPI_ISL_2872334, EPI_ISL_2872430, EPI_ISL_2872473, EPI_ISL_2872474, EPI_ISL_2872620, EPI_ISL_2872677, EPI_ISL_2872710, EPI_ISL_2872735, EPI_ISL_2872736, EPI_ISL_2872785, EPI_ISL_2872809, EPI_ISL_2872810, EPI_ISL_2872831, EPI_ISL_2872841, EPI_ISL_2872842, EPI_ISL_2872844, EPI_ISL_2872873, EPI_ISL_2872874, EPI_ISL_2872881, EPI_ISL_2872927, EPI_ISL_2872939, EPI_ISL_2872941, EPI_ISL_2872997, EPI_ISL_2873078, EPI_ISL_2873088, EPI_ISL_2873122, EPI_ISL_2873124, EPI_ISL_2873165, EPI_ISL_2873167, EPI_ISL_2873189, EPI_ISL_2873190, EPI_ISL_2873196, EPI_ISL_2873199, EPI_ISL_2873200, EPI_ISL_2873201, EPI_ISL_2873221, EPI_ISL_2873222, EPI_ISL_2873227, EPI_ISL_2873232, EPI_ISL_2873239, EPI_ISL_2873244, EPI_ISL_2873254, EPI_ISL_2873255, EPI_ISL_2873279, EPI_ISL_2873282, EPI_ISL_2873284, EPI_ISL_2873320, EPI_ISL_2926208, EPI_ISL_2926213, EPI_ISL_2926255, EPI_ISL_2926257, EPI_ISL_2926272, EPI_ISL_2926339, EPI_ISL_2926360, EPI_ISL_2926369, EPI_ISL_2926376, EPI_ISL_2926396, EPI_ISL_2926410, EPI_ISL_2926415, EPI_ISL_2926429, EPI_ISL_2926451, EPI_ISL_2926500, EPI_ISL_2926514, EPI_ISL_2926523, EPI_ISL_2926530, EPI_ISL_2926531, EPI_ISL_2926532, EPI_ISL_2926537, EPI_ISL_2926676, EPI_ISL_2926745, EPI_ISL_2926778, EPI_ISL_2926847, EPI_ISL_2926903, EPI_ISL_2926963, EPI_ISL_2926966, EPI_ISL_2926978, EPI_ISL_2927001, EPI_ISL_2927084, EPI_ISL_2927114, EPI_ISL_2927115, EPI_ISL_2927132, EPI_ISL_2927159, EPI_ISL_2927167, EPI_ISL_2927240, EPI_ISL_2927242, EPI_ISL_2927265, EPI_ISL_2927307, EPI_ISL_2927312, EPI_ISL_2927320, EPI_ISL_2927321, EPI_ISL_2927327, EPI_ISL_2927332, EPI_ISL_2927340, EPI_ISL_2927360, EPI_ISL_2927367, EPI_ISL_2927392, EPI_ISL_2927393, EPI_ISL_2927402, EPI_ISL_2927486, EPI_ISL_2927489, EPI_ISL_2927548, EPI_ISL_2927588, EPI_ISL_2927626, EPI_ISL_2927646, EPI_ISL_2927676, EPI_ISL_2927691, EPI_ISL_2927694, EPI_ISL_2927695, EPI_ISL_2927715, EPI_ISL_3016915, EPI_ISL_3016930, EPI_ISL_3016967, EPI_ISL_3016975, EPI_ISL_3017014, EPI_ISL_3017017, EPI_ISL_3017019, EPI_ISL_3017026, EPI_ISL_3017174, EPI_ISL_3017176, EPI_ISL_3017221, EPI_ISL_3017242, EPI_ISL_3017279, EPI_ISL_3017327, EPI_ISL_3017348, EPI_ISL_3017349, EPI_ISL_3017519, EPI_ISL_3017520, EPI_ISL_3017521, EPI_ISL_3017548, EPI_ISL_3017579, EPI_ISL_3017588, EPI_ISL_3017589, EPI_ISL_3017598, EPI_ISL_3017614, EPI_ISL_3017615, EPI_ISL_3017654, EPI_ISL_3017665, EPI_ISL_3017697, EPI_ISL_3017717, EPI_ISL_3017719, EPI_ISL_3017731, EPI_ISL_3017743, EPI_ISL_3017744, EPI_ISL_3017776, EPI_ISL_3017801, EPI_ISL_3017802, EPI_ISL_3017812, EPI_ISL_3017833, EPI_ISL_3017855, EPI_ISL_3017887, EPI_ISL_3017905, EPI_ISL_3017911, EPI_ISL_3017950, EPI_ISL_3017969, EPI_ISL_3017992, EPI_ISL_3017994, EPI_ISL_3018005, EPI_ISL_3018006, EPI_ISL_3018095, EPI_ISL_3018106, EPI_ISL_3018151, EPI_ISL_3020069, EPI_ISL_3111330, EPI_ISL_3111339, EPI_ISL_3111358, EPI_ISL_3111457, EPI_ISL_3111471, EPI_ISL_3111489, EPI_ISL_3111491, EPI_ISL_3111492, EPI_ISL_3111576, EPI_ISL_3111677, EPI_ISL_3111779, EPI_ISL_3111811, EPI_ISL_3111814, EPI_ISL_3111848, EPI_ISL_3112180, EPI_ISL_3112302, EPI_ISL_3112304, EPI_ISL_3112416, EPI_ISL_3112459, EPI_ISL_3112478, EPI_ISL_3112540, EPI_ISL_3112569, EPI_ISL_3112652, EPI_ISL_3112764, EPI_ISL_3112995, EPI_ISL_3113038, EPI_ISL_3113219, EPI_ISL_3113280, EPI_ISL_3113287, EPI_ISL_3113321, EPI_ISL_3113359, EPI_ISL_3113497, EPI_ISL_3113502, EPI_ISL_3113622, EPI_ISL_3325548, EPI_ISL_3325577, EPI_ISL_3325732, EPI_ISL_3325734, EPI_ISL_3326343, EPI_ISL_3326376, EPI_ISL_3326546, EPI_ISL_3326562, EPI_ISL_3326572, EPI_ISL_3326649, EPI_ISL_3326911, EPI_ISL_3328025, EPI_ISL_3328026, EPI_ISL_3328443, EPI_ISL_3328455, EPI_ISL_3328467, EPI_ISL_3328481, EPI_ISL_3328507, EPI_ISL_3328514, EPI_ISL_3328532, EPI_ISL_3328657, EPI_ISL_3328756, EPI_ISL_3330637, EPI_ISL_3330657, EPI_ISL_3330704 | | | |
| see above | Laboratory Corporation of America | Centers for Disease Control and Prevention Division of Viral Diseases, Pathogen Discovery | ; Adrian Paskey; Amanda Douglas; Amanda Suchanek; Andrea Throop; Ayla Burns; Benjamin Rambo-Martin; Bobbi Croy; Brian Krueger; Brian Norvell; Christopher Gulvick; Christos Petropoulos; Clinton R. Paden; Craig Lukasik; Dakota Howard; Darlene Wagner; Debbie Boles; Dhwani Batra; Duncan MacCannell; Eyad Almasri; Goran Stevovic; Howard Engler; Hrushikesh Deshmukh; Jake Humphrey; Jana Schroth; Jason Caravas; Joe Voshell; John Pruitt; Jonathan Meltzer; Jonathan Williams; Kara Moser; Kimberly Wagner; Lax Iyer; Lisa Pfefferle; Lyndon Tilson; Manoj Jain; Marcia Eisenberg; Mary Ann Cristobal; Mary Williamson; Matthew Robinson; Matthew Schmerer; Michael Levandoski; Mike Sapeta; Mindy Nye; Minoo Agarwal; Mohan Kolli; Nuthawin Charoensri; Oren Cohen; Peter W. Cook; Prashant Gupta; Qian Zeng; Rama Ghatti; Scott Parker; Scott Ryan; Scott Sammons; Shatavia Morrison; Stanley Letovsky; Steven Ragan; Suresh Babu Selvaraju; Susan Countryman; Susan Hicks; Suzanne Dale; Thomas Urban; Tim Kuphal; Tricia Zwiefelhofer; Vincent Drouillon; Yvette Unoarumhi |
| EPI_ISL_2811913 | Laboratory Medicine, University of Washington | University of Washington/Fred Hutch | Castor; J.L. |
| EPI_ISL_2557405, EPI_ISL_2557407, EPI_ISL_2557409, EPI_ISL_2557410, EPI_ISL_2645506, EPI_ISL_2863605, EPI_ISL_2863607, EPI_ISL_2863609, EPI_ISL_2863611, EPI_ISL_2863613, EPI_ISL_2863614, EPI_ISL_2863616, EPI_ISL_2863689, EPI_ISL_2863691, EPI_ISL_2863693, EPI_ISL_2863797, EPI_ISL_2863798, EPI_ISL_2863799, EPI_ISL_2863800, EPI_ISL_2863897, EPI_ISL_2982717, EPI_ISL_2982718, EPI_ISL_2982719, EPI_ISL_2982720, EPI_ISL_2982721, EPI_ISL_2982722, EPI_ISL_2982724, EPI_ISL_2982725, EPI_ISL_2982726, EPI_ISL_2982727, EPI_ISL_2982729, EPI_ISL_2982730, EPI_ISL_2982731, EPI_ISL_2982732, EPI_ISL_2982733, EPI_ISL_2982736, EPI_ISL_2982737, EPI_ISL_2982738, EPI_ISL_2982740, EPI_ISL_2982865, EPI_ISL_2982866, EPI_ISL_2983187, EPI_ISL_2983236, EPI_ISL_2983237, EPI_ISL_2983379, EPI_ISL_2983380, EPI_ISL_2983381, EPI_ISL_2983382, EPI_ISL_3045374, EPI_ISL_3045375, EPI_ISL_3045376, EPI_ISL_3045377, EPI_ISL_3045382, EPI_ISL_3045383, EPI_ISL_3045423, EPI_ISL_3045470, EPI_ISL_3045472, EPI_ISL_3045473, EPI_ISL_3045474, EPI_ISL_3045475, EPI_ISL_3045476, EPI_ISL_3045477, EPI_ISL_3045478, EPI_ISL_3045479, EPI_ISL_3045480, EPI_ISL_3045481, EPI_ISL_3045482, EPI_ISL_3045485, EPI_ISL_3045488, EPI_ISL_3045489, EPI_ISL_3045490, EPI_ISL_3045491, EPI_ISL_3045492, EPI_ISL_3045494, EPI_ISL_3045495, EPI_ISL_3045496, EPI_ISL_3045497, EPI_ISL_3045498, EPI_ISL_3045499, EPI_ISL_3045500, EPI_ISL_3045501, EPI_ISL_3045502, EPI_ISL_3045503, EPI_ISL_3045505, EPI_ISL_3045506, EPI_ISL_3045507, EPI_ISL_3045511, EPI_ISL_3045512, EPI_ISL_3045513, EPI_ISL_3045515, EPI_ISL_3045518, EPI_ISL_3045519, EPI_ISL_3045521, EPI_ISL_3045522, EPI_ISL_3045523, EPI_ISL_3045524, EPI_ISL_3045525, EPI_ISL_3045526, EPI_ISL_3045527, EPI_ISL_3045528, EPI_ISL_3045529, EPI_ISL_3045531, EPI_ISL_3045532, EPI_ISL_3045533, EPI_ISL_3045537, EPI_ISL_3045538, EPI_ISL_3045539, EPI_ISL_3045540, EPI_ISL_3045544, EPI_ISL_3045545, EPI_ISL_3045546, EPI_ISL_3045547, EPI_ISL_3045548, EPI_ISL_3045549, EPI_ISL_3045551, EPI_ISL_3045552, EPI_ISL_3045553, EPI_ISL_3190168, EPI_ISL_3435049, EPI_ISL_3435050, EPI_ISL_3435051, EPI_ISL_3435052, EPI_ISL_3539998, EPI_ISL_3539999 | | | |
| see above | Laboratory of Respiratory Viruses and Measles, Oswaldo Cruz Institute, FIOCRUZ | Laboratory of Respiratory Viruses and Measles, Oswaldo Cruz Institute, FIOCRUZ | Agatha Cristinne Prudencio; Agatha Cristinne Prudencio Soares; Alice Sampaio Rocha; Ana Carolina Mendonca; Anna Carolina Paixao; Elisa Cavalcante Pereira; Fernando Motta; Ighor Leonardo Arantes Gomes; Luciana Appolinario; Marilda Siqueira on behalf of the Fiocruz COVID-19 Genomic Surveillance Network; Paola Resende; Renata Serrano Lopes; Taina Venas |
| EPI_ISL_3025606, EPI_ISL_3025615, EPI_ISL_3025620, EPI_ISL_3025626, EPI_ISL_3461216, EPI_ISL_3461217, EPI_ISL_3461239, EPI_ISL_3461240, EPI_ISL_3461243, EPI_ISL_3461285, EPI_ISL_3461328 | | | |
| see above | Laboratory, Gravity Diagnostics | Laboratory, Gravity Diagnostics | Diagnostics, G. |
| EPI_ISL_2777293, EPI_ISL_2777512, EPI_ISL_2777564, EPI_ISL_2777616, EPI_ISL_2777631, EPI_ISL_2777651, EPI_ISL_2777658, EPI_ISL_2777669, EPI_ISL_2777690, EPI_ISL_2777692, EPI_ISL_2777806, EPI_ISL_2777863, EPI_ISL_2777896, EPI_ISL_2777897, EPI_ISL_2777898, EPI_ISL_3050298, EPI_ISL_3050302, EPI_ISL_3050303, EPI_ISL_3050308, EPI_ISL_3050309, EPI_ISL_3050312, EPI_ISL_3050315, EPI_ISL_3050319, EPI_ISL_3050322, EPI_ISL_3050332, EPI_ISL_3050337, EPI_ISL_3050344, EPI_ISL_3050354, EPI_ISL_3050363, EPI_ISL_3050367, EPI_ISL_3050375, EPI_ISL_3050376, EPI_ISL_3050384, EPI_ISL_3050387, EPI_ISL_3050391, EPI_ISL_3050394, EPI_ISL_3050401, EPI_ISL_3050403, EPI_ISL_3050406, EPI_ISL_3050407, EPI_ISL_3050415, EPI_ISL_3050431, EPI_ISL_3050434, EPI_ISL_3050436, EPI_ISL_3050464, EPI_ISL_3050470, EPI_ISL_3050471, EPI_ISL_3050490, EPI_ISL_3050501, EPI_ISL_3050502, EPI_ISL_3050503, EPI_ISL_3050506, EPI_ISL_3050519, EPI_ISL_3050529, EPI_ISL_3050538, EPI_ISL_3050551, EPI_ISL_3050558, EPI_ISL_3050565, EPI_ISL_3050566, EPI_ISL_3050570, EPI_ISL_3050593, EPI_ISL_3050605, EPI_ISL_3050624, EPI_ISL_3050627, EPI_ISL_3050636 | | | |
| see above | Laboratório Central de Saúde Pública do Amazonas - LACEN-AM | Laboratorio de Ecologia de Doencas Transmissiveis na Amazonia, Instituto Leonidas e Maria Deane - Fiocruz Amazonia | André Corado; Debora Duarte; Felipe Naveca; Fernanda Nascimento; George Silva; Karina Pessoa; Luciana Gonçalves; Maria Júlia Brandão; Matilde Mejía; Michele Jesus; Valdinete Nascimento; Victor Souza; Ágatha Costa |
| EPI_ISL_3426438, EPI_ISL_3584949, EPI_ISL_3585952, EPI_ISL_3587490, EPI_ISL_3602957, EPI_ISL_3602958, EPI_ISL_3602960, EPI_ISL_3602964, EPI_ISL_3610353, EPI_ISL_3610354, EPI_ISL_3610355, EPI_ISL_3610356, EPI_ISL_3610358, EPI_ISL_3610359 | | | |
| see above | Laboratório Central de Saúde Pública do Estado do Tocantins (LACEN/TO) | Bioinformatics and Biotechnology Laboratory (Labinftec/Federal | Bergmann Morais Ribeiro; Chácaras 69/72; Fabrício Souza Campos; Fernando Lucas Melo; Gurupi/TO Ueric José Borges de Souza; Jucimária Dantas Galvão; Laboratório de Bioinformática e Biotecnologia; Lote 7; Raíssa Nunes dos Santos; Rua Badejós; Ueric José Borges de Souza; Zona Rural \| 77.402-970\| UFT Campus de Gurupi |

| | | | |
|---|---|---|---|
| | | University of Tocantins, Campus of Gurupi) | |
| EPI_ISL_2835216, EPI_ISL_2835220, EPI_ISL_2835221, EPI_ISL_2835222, EPI_ISL_2835223, EPI_ISL_2983455, EPI_ISL_2983457, EPI_ISL_2983459, EPI_ISL_2983460, EPI_ISL_2983462, EPI_ISL_2983463, EPI_ISL_2983464, EPI_ISL_2983466, EPI_ISL_2983467, EPI_ISL_2983468, EPI_ISL_3539775, EPI_ISL_3539776, EPI_ISL_3539777, EPI_ISL_3539778, EPI_ISL_3539779, EPI_ISL_3539780, EPI_ISL_3539781, EPI_ISL_3539782, EPI_ISL_3539783, EPI_ISL_3539784, EPI_ISL_3539785, EPI_ISL_3539786, EPI_ISL_3539787, EPI_ISL_3539789, EPI_ISL_3539790 | | | |
| see above | Laboratório de Biotecnologia Aplicada (LBA) - Laboratório de Biologia Molecular - Hospital das Clínicas, Faculdade de Medicina de Botucatu, Departamento de Bioprocessos e Biotecnologia - Faculdade de Ciências Agronômicas, UNESP – Botucatu/SP | Laboratory of Respiratory Viruses and Measles, Oswaldo Cruz Institute, FIOCRUZ | Alice Sampaio Rocha; Ana Carolina Mendonca; Anna Carolina Paixao; Elisa Cavalcante Pereira; Felipe Allan da Silva Costa; Fernando Motta; Jayme Augusto de Souza Neto; Leonardo Nazario de Moraes; Luciana Appolinario; Marilda Siqueira on behalf of the Fiocruz COVID-19 Genomic Surveillance Network; Paola Resende; Patricia Akemi Assato; Rejane Maria Tommasini; Renata Serrano Lopes; Taina Venas |
| EPI_ISL_3456068, EPI_ISL_3456069, EPI_ISL_3873632, EPI_ISL_3873661, EPI_ISL_3873663, EPI_ISL_3873665, EPI_ISL_3873666 | | | |
| see above | Laboratório de Microbiologia Molecular - Universidade FEEVALE | Molecular Microbiology Laboratory | Alana Witt Hansen; Fernando Rosado Spilki; Fágner Henrique Heldt; Juliana Schons Gulart; Juliana Schons Gularte; Juliane Deise Fleck; Mariana Soares da Silva; Matheus Nunes Weber; Meriane Demoliner; Michele Filippi.; Micheli Filippi; Paula Rodrigues de Almeida; Vyctoria Malayhka de Abreu Góes Pereira. |
| EPI_ISL_2557412, EPI_ISL_2557413 | Labortorio Central de Saude Publica do Estado de Santa Catarina (LACEN/SC) | Laboratory of Respiratory Viruses and Measles, Oswaldo Cruz Institute, FIOCRUZ | Alice Sampaio Rocha; Ana Carolina Mendonca; Anna Carolina Paixao; Darcita Buerger Rovaris; Elisa Cavalcante Pereira; Fernando Motta; Luciana Appolinario; Marilda Siqueira on behalf of the Fiocruz COVID-19 Genomic Surveillance Network; Paola Resende; Renata Serrano Lopes; Sandra Bianchini Fernandes; Taina Venas |
| EPI_ISL_2645505, EPI_ISL_2863891, EPI_ISL_3045378, EPI_ISL_3045379, EPI_ISL_3045380, EPI_ISL_3190193, EPI_ISL_3190194, EPI_ISL_3190195, EPI_ISL_3190196, EPI_ISL_3190197, EPI_ISL_3190198, EPI_ISL_3190199, EPI_ISL_3190200, EPI_ISL_3190203, EPI_ISL_3190204, EPI_ISL_3190205, EPI_ISL_3190207, EPI_ISL_3190209, EPI_ISL_3190210, EPI_ISL_3190214, EPI_ISL_3190216, EPI_ISL_3190296 | | | |
| see above | Labortorio Central de Saude Publica do Estado do Rio de Janeiro (LACEN/RJ) | Laboratory of Respiratory Viruses and Measles, Oswaldo Cruz Institute, FIOCRUZ | Agatha Cristinne Prudencio Soares; Alice Sampaio Rocha; Ana Carolina Mendonca; Andrea Cony Cavalcanti; Anna Carolina Paixao; Elisa Cavalcante Pereira; Fernando Motta; Ighor Leonardo Arantes Gomes; Luciana Appolinario; Marilda Siqueira on behalf of the Fiocruz COVID-19 Genomic Surveillance Network; Paola Resende; Renata Serrano Lopes; Taina Venas |
| EPI_ISL_2645507, EPI_ISL_2645512, EPI_ISL_2645513, EPI_ISL_2863600, EPI_ISL_2983418, EPI_ISL_2983419, EPI_ISL_2983420, EPI_ISL_2983422, EPI_ISL_2983423 | | | |
| see above | Labortorio Central de Saude Publica do Estado do Tocantins (LACEN/TO) | Laboratory of Respiratory Viruses and Measles, Oswaldo Cruz Institute, FIOCRUZ | Alice Sampaio Rocha; Ana Carolina Mendonca; Anna Carolina Paixao; Elisa Cavalcante Pereira; Fernando Motta; Jucimaria Dantas Galvao; Luciana Appolinario; Marilda Siqueira on behalf of the Fiocruz COVID-19 Genomic Surveillance Network; Paola Resende; Renata Serrano Lopes; Taina Venas |
| EPI_ISL_2613809, EPI_ISL_2613810, EPI_ISL_2613811, EPI_ISL_2613812, EPI_ISL_2613813, EPI_ISL_2613814, EPI_ISL_2613815 | | | |
| see above | Lboratorio Central de Saude Publica do Estado do Parana (LACEN/PR) | Laboratory of Respiratory Viruses and Measles, Oswaldo Cruz Institute, FIOCRUZ | Alice Sampaio Rocha; Ana Carolina Mendonca; Anna Carolina Paixao; Elisa Cavalcante Pereira; Fernando Motta; Irina Riediger; Luciana Appolinario; Marilda Siqueira on behalf of the Fiocruz COVID-19 Genomic Surveillance Network; Paola Resende; Renata Serrano Lopes; Taina Venas |
| EPI_ISL_2392333, EPI_ISL_2395953, EPI_ISL_2437005, EPI_ISL_2446442 | Lighthouse Lab in Milton Keynes | Wellcome Sanger Institute for the COVID-19 Genomics UK (COG-UK) Consortium | Cordelia Langford; David K. Jackson; Dominic Kwiatkowski; Ewan Harrison; Ian Johnston; Jeffrey Barrett; John Sillitoe on behalf of the Wellcome Sanger Institute COVID-19 Surveillance Team; Roberto Amato; Sonia Goncalves; The Lighthouse Lab in Milton Keynes and Alex Alderton |
| EPI_ISL_2390202 | Limbach - MVZ Labor Ravensburg Labor Dr. Gärtner | Robert Koch Institute | |
| EPI_ISL_2842581, EPI_ISL_2842582, EPI_ISL_2842583, EPI_ISL_2842587, EPI_ISL_2842594 | Lurie Children's Hospital of Chicago | Northwestern University - Ozer Lab | Egon A. Ozer; Judd F. Hultquist; Lacy M. Simons; Larry K. Kociolek; Michael G. Ison; Ramon Lorenzo-Redondo; Taylor J. Dean; William J. Muller; Xiaotian; Zheng |
| EPI_ISL_3102429, EPI_ISL_3102455 | MATERNIDADE ESCOLA ASSIS CHATEAUBRIAND | Analytical Competence Molecular Epidemiology Lab/ACME, Oswaldo Cruz Foundation, Ceara (FIOCRUZ CE) | Cleber Furtado Aksenen; Fabio Miyajima; Fernando Braga Stehling; Francisco Eder de Moura Lopes; Jamille Maria Mendes Bezerra; Joaquim César do Nascimento Sousa Junior; Pedro Miguel Carneiro Jeronimo; Suzana Porto Almeida e Lucas Delerino; Thais Ferreira de Oliveira; Thais de Oliveira Costa; Ticiane Cavalcante de Souza; Veridiana Pessoa Miyajima |
| EPI_ISL_2598083, EPI_ISL_2709816, EPI_ISL_2790050, EPI_ISL_2861207, EPI_ISL_2862024, EPI_ISL_2955557, EPI_ISL_2955566, EPI_ISL_2955569, EPI_ISL_2980037, EPI_ISL_3020114, EPI_ISL_3045923, EPI_ISL_3099901 | | | |
| see above | MD PHL | MD PHL | Maryland Department of Health Laboratories Administration |
| EPI_ISL_2957399 | MDI Limbach Berlin GmbH; MVZ Labor Berlin | Robert Koch Institute | |
| EPI_ISL_3230761 | MEDILAB | CHU Poitiers | Agnes BEBY-DEFAUX; Birama N'DIAYE; Caroline MICHAUD; Magali GARCIA; Manon PRAT; Maxime PICHON; Nicolas LEVEQUE; Valentin BON-BARET |
| EPI_ISL_2659675, EPI_ISL_2659754 | MEPHI, Aix Marseille University | MEPHI, Aix Marseille University | Anthony LEVASSEUR |
| EPI_ISL_2626996 | MONTEFIORE MEDICAL CENTER LABORATORIES | Wadsworth Center, New York State Department of Health | Alexis Russell; Catharine Prussing; Daryl M. Lamson; Erasmus Schneider; Erica Lasek-Nesselquist; John Kelly; Jonathan Plitnick; Kirsten St. George; Matthew Shudt; Melissa A Leisner; Navjot Singh |
| EPI_ISL_2787554 | MS Public Health Laboratory | Centers for Disease Control and Prevention Division of Viral Diseases, Pathogen Discovery | Alison Laufer Halpin; Ben L. Rambo-Martin; Clinton R. Paden; Dakota Howard; Darlene Wagner; Dave Wentworth; Dhwani Batra; Jasmine Padilla; Justin Lee; Katie Dillon; Krista Queen; Kristen Knipe; Kristine Lacek; Mark Burroughs; Matthew Schmerer; Mili Sheth; Peter Cook; Sam Shepard; Sarah Nobles; Shoshona Le; Suxiang Tong; Vivien Dugan; Yvette Unoarumhi |
| EPI_ISL_2646337, EPI_ISL_2646340, EPI_ISL_2858795, EPI_ISL_2858801 | MS Public Health Laboratory | University of Mississippi Medical Center, Molecular and Genomics Core Facility | Ashley C. Johnson; D. Ashley Robinson; Ithiel J. Frame; Krishna K. Ayyalasomayajula; Michael R. Garrett; Wenjie Wu |
| EPI_ISL_2690127 | MT Public Health Laboratory | Centers for Disease Control and Prevention Division of Viral Diseases, Pathogen Discovery | Alison Laufer Halpin; Ben L. Rambo-Martin; Clinton R. Paden; Dakota Howard; Darlene Wagner; Dave Wentworth; Dhwani Batra; Jasmine Padilla; Justin Lee; Katie Dillon; Krista Queen; Kristen Knipe; Kristine Lacek; Mark Burroughs; Matthew Schmerer; Mili Sheth; Peter Cook; Sam Shepard; Sarah Nobles; Shoshona Le; Suxiang Tong; Vivien Dugan; Yvette Unoarumhi |
| EPI_ISL_2472447, EPI_ISL_2472482, EPI_ISL_2632831, EPI_ISL_2635874, EPI_ISL_2635888, EPI_ISL_2762510 | MVZ Labor Dr. Limbach & Kollegen GbR | Robert Koch Institute | |
| EPI_ISL_2647377, EPI_ISL_2647466, EPI_ISL_2647483, EPI_ISL_2647487, EPI_ISL_2647528, EPI_ISL_2647529, EPI_ISL_2647588, EPI_ISL_2647651, EPI_ISL_2647655, EPI_ISL_2647694, EPI_ISL_2647714, EPI_ISL_2647716, EPI_ISL_2647845, EPI_ISL_2647854, EPI_ISL_2647866, EPI_ISL_2647869, EPI_ISL_2647889, EPI_ISL_2647890, EPI_ISL_2647936, EPI_ISL_2647956, EPI_ISL_2786002, EPI_ISL_2786004, EPI_ISL_2786086, EPI_ISL_2786093, EPI_ISL_2786115, EPI_ISL_2786129, EPI_ISL_2786191, EPI_ISL_2786251, EPI_ISL_2786283, EPI_ISL_2786329, EPI_ISL_2786442, EPI_ISL_2786460, EPI_ISL_2786470, EPI_ISL_2786508, EPI_ISL_2786527, EPI_ISL_2786580, EPI_ISL_2875242, EPI_ISL_2875281, EPI_ISL_2875282, EPI_ISL_2875290, EPI_ISL_2875351, EPI_ISL_2875368, EPI_ISL_2875397, EPI_ISL_2875402, EPI_ISL_2875438, EPI_ISL_2875477, EPI_ISL_2875483, EPI_ISL_2875503, EPI_ISL_3065124, EPI_ISL_3065150, EPI_ISL_3065172, EPI_ISL_3065282, EPI_ISL_3065479, EPI_ISL_3065499, EPI_ISL_3110617, EPI_ISL_3110717, EPI_ISL_3110748, EPI_ISL_3110749, EPI_ISL_3110831, EPI_ISL_3110860, EPI_ISL_3110893, EPI_ISL_3110919 | | | |
| see above | Mako Medical | Centers for Disease Control and Prevention Division of Viral Diseases, Pathogen Discovery | Adrian Paskey; Benjamin Rambo-Martin; Christopher Gulvick; Clinton R. Paden; Dakota Howard; Darlene Wagner; Dhwani Batra; Duncan MacCannell; Jason Caravas; Kara Moser; Lauren Moon; Matthew Schmerer; Matthew Tugwell; Peter W. Cook; Scott Sammons; Shatavia Morrison; Yvette Unoarumhi |
| EPI_ISL_3009153, EPI_ISL_3009168, EPI_ISL_3435356, EPI_ISL_3435357, EPI_ISL_3435359 | Maryland Genomics, Institute for Genome Sciences, University of Maryland School of Medicine | Maryland Genomics, Institute for Genome Sciences, University of Maryland School of Medicine | Claire M; Fraser; Hazen; Holly; Humphrys; Ivette; Jacques; Jonathan; Kranthi; Lim; Lisa D; Luke J; Mike; Ott; Ravel; Roussey; Sadzewicz; Sandra; Santana-Cruz; Tallon; Tracy; Vavikolanu |
| EPI_ISL_2550770 | Maryland Public Health Laboratory (MD PHL) | Maryland Public Health Laboratory (MD PHL) | Maryland Department of Health Laboratories Administration |
| EPI_ISL_2484936, EPI_ISL_2688808, EPI_ISL_2688819, EPI_ISL_2920258, EPI_ISL_2920266, EPI_ISL_2920271 | Massachusetts State Public Health Laboratory | Massachusetts State Public Health Laboratory | Andrew Lang; Glen Gallagher; Sandra Smole; Timelia Fink |
| EPI_ISL_2958947 | Mayo Clinic & Mayo Clinic Laboratories | Minnesota Department of Health, Public Health Laboratory | Alexandra Lorentz; Jacob Garfin; Matt Plumb; and Xiong Wang |
| EPI_ISL_2544257 | Medica | Institute of Medical Virology | Alexandra Trkola; Annette Audigé; Cyril Shah; Gabriela Ziltener; Guido Bloemberg; Jon Huder; Jürg Böni; Kevin Steiner; Maria Grünberg; Maryam Zaheri; Michael Huber; Riccarda Capaul; Stefan Schmutz; Verena Kufner |
| EPI_ISL_2636815 | Medizinisch-Diagnostisches Labor Kempten allgäulab | Robert Koch Institute | |
| EPI_ISL_2433970, EPI_ISL_2433982 | Microbiologia CATLAB | Can Ruti SARS-CoV-2 Sequencing Hub (HUGTiP/IrsiCaixa/IGTP) | Alba Sánchez; Anna Not; Antoni E Bordoy; Bonaventura Clotet; Cristina Casañ; Cristina Esteban; Francesc Catala-Moll; Gemma Clara; Ignacio Blanco; Marc Noguera-Julian; Maria Casadellà; Mariona Parera; Mercedes Guerrero; Montserrat Giménez; Pere-Joan Cardona; Pilar Armengol; Roger Paredes; Verónica Saludes; and Elisa Martró on behalf of the Can Ruti SARS-CoV-2 Sequencing Hub. |

| | | | |
|---|---|---|---|
| EPI_ISL_2448064, EPI_ISL_2650271, EPI_ISL_2650293, EPI_ISL_2650328, EPI_ISL_2832110, EPI_ISL_2832172 | Microbiologia CATLAB | Can Ruti SARS-CoV-2 Sequencing Hub (HUGTiP/IrsiCaixa/IGTP) | Alba Snchez; Alba Sánchez; Alexia París; Anna Not; Antoni E Bordoy; Bonaventura Clotet; Cristina Casa; Cristina Casañ; Cristina Esteban; David Panisello; Francesc Catala-Moll; Gemma Clara; Ignacio Blanco; Laia Soler; Lauro Sumoy; Marc Noguera-Julian; Maria Casadell; Maria Casadellà; Mariona Parera; Mercedes Guerrero; Montserrat Gimnez; Montserrat Giménez; Pere-Joan Cardona; Pilar Armengol; Roger Paredes; Vernica Saludes; Verónica Saludes; and Elisa Martr on behalf of the Can Ruti SARS-CoV-2 Sequencing Hub.; and Elisa Martró on behalf of the Can Ruti SARS-CoV-2 Sequencing Hub |
| EPI_ISL_2448054, EPI_ISL_2448086, EPI_ISL_2671448, EPI_ISL_2812471, EPI_ISL_2812485, EPI_ISL_2832166, EPI_ISL_2832188, EPI_ISL_2897003, EPI_ISL_2897006, EPI_ISL_2926161, EPI_ISL_2926172, EPI_ISL_3027975, EPI_ISL_3163990, EPI_ISL_3320201, EPI_ISL_3320217, EPI_ISL_3455752 | | | |
| see above | Microbiology Department, Laboratori Clínic Metropolitana Nord. Hospital Universitari Germans Trias i Pujol | Can Ruti SARS-CoV-2 Sequencing Hub (HUGTiP/IrsiCaixa/IGTP) | Alba Snchez; Alba Sánchez; Alexia París; Anna Not; Antoni E Bordoy; Bonaventura Clotet; Cristina Casa; Cristina Casañ; Cristina Esteban; David Panisello; Francesc Catala-Moll; Gemma Clara; Ignacio Blanco; Laia Soler; Lauro Sumoy; Marc Noguera-Julian; Maria Casadell; Maria Casadellà; Mariona Parera; Mercedes Guerrero; Montserrat Gimnez; Montserrat Giménez; Pere-Joan Cardona; Pilar Armengol; Roger Paredes; Vernica Saludes; Verónica Saludes; and Elisa Martr on behalf of the Can Ruti SARS-CoV-2 Sequencing Hub.; and Elisa Martró on behalf of the Can Ruti SARS-CoV-2 Sequencing Hub |
| EPI_ISL_2756396, EPI_ISL_2774104 | Microbiology Department, University Hospital Donostia | Microbiology Department, University Hospital Donostia | Cilla G.; Gomez M; Marimon JM; Martin-Peñaranda T; Montes M; Piñeiro L; Sorarrain A |
| EPI_ISL_2501367, EPI_ISL_2501368, EPI_ISL_2861303, EPI_ISL_2967893 | Microbiology Department. Complexo Hospitalario Universitario de Vigo | Microbiology Department. Complexo Hospitalario Universitario de Vigo | Alfaya N; Alonso I; Alvarez M; Cabrera JJ; Carballo R; Cores O; Cortizo S; Davina C; Martinez L; Mediero G; Perez S; Potel C; Regueiro B; Rey S; Vasallo FJ; del-Campo V |
| EPI_ISL_3026893 | Microbiology Division, SC DHEC | Microbiology Division, SC DHEC | Freeman; J.S. |
| EPI_ISL_2936682, EPI_ISL_2958605, EPI_ISL_2984697, EPI_ISL_2984741, EPI_ISL_2987761, EPI_ISL_2987789, EPI_ISL_2987790, EPI_ISL_2987793, EPI_ISL_2987795, EPI_ISL_2987796, EPI_ISL_2987810, EPI_ISL_2987811, EPI_ISL_2987812, EPI_ISL_2987813, EPI_ISL_2987815, EPI_ISL_2987816, EPI_ISL_2987817, EPI_ISL_2987818, EPI_ISL_2987819, EPI_ISL_2987923, EPI_ISL_2987973, EPI_ISL_2988015, EPI_ISL_2988020, EPI_ISL_3032541, EPI_ISL_3032559, EPI_ISL_3032569, EPI_ISL_3032574, EPI_ISL_3032580, EPI_ISL_3032586, EPI_ISL_3032587, EPI_ISL_3032633, EPI_ISL_3032636, EPI_ISL_3032638, EPI_ISL_3032644, EPI_ISL_3032654, EPI_ISL_3032706, EPI_ISL_3032817, EPI_ISL_3063340, EPI_ISL_3063345, EPI_ISL_3063349, EPI_ISL_3063351, EPI_ISL_3063361, EPI_ISL_3063365, EPI_ISL_3066700, EPI_ISL_3066701, EPI_ISL_3066703, EPI_ISL_3066705, EPI_ISL_3066706, EPI_ISL_3066707, EPI_ISL_3066709, EPI_ISL_3066710, EPI_ISL_3066726, EPI_ISL_3066727, EPI_ISL_3066729, EPI_ISL_3066730, EPI_ISL_3066744, EPI_ISL_3066745, EPI_ISL_3404496, EPI_ISL_3404498, EPI_ISL_3404499, EPI_ISL_3404541, EPI_ISL_3906776, EPI_ISL_3906902, EPI_ISL_3907002, EPI_ISL_3907003 | | | |
| see above | Ministry of Health Turkey | Ministry of Health Turkey | Fatma Bayrakdar; Gulay Korukluoglu; Gülay Korukluoğlu; Suleyman Yalcin; Süleyman Yalcin; Yasemin Cosgun; Yasemin Coşgun |
| EPI_ISL_2425383, EPI_ISL_2501152, EPI_ISL_2565864, EPI_ISL_2658515, EPI_ISL_2727839, EPI_ISL_2727855, EPI_ISL_2958944, EPI_ISL_3099434, EPI_ISL_3099435, EPI_ISL_3099436 | | | |
| see above | Minnesota Department of Health, Public Health Laboratory | Minnesota Department of Health, Public Health Laboratory | Alexandra Lorentz; Jacob Garfin; Matt Plumb; and Xiong Wang |
| EPI_ISL_2635085, EPI_ISL_2753648, EPI_ISL_3011018 | Missouri State Public Health Laboratory | Missouri State Public Health Laboratory | Ashley New; Joshua Barry; Matthew Sinn |
| EPI_ISL_3142934, EPI_ISL_3142935 | Molecular & Genomic Pathology Laboratory, Thomas Jefferson University Hospital | Molecular & Genomic Pathology Laboratory, Thomas Jefferson University Hospital | Nitika Badjatia; Run Jin; Shannon Mahoney; Zi-xuan Wang |
| EPI_ISL_2692159, EPI_ISL_2692168, EPI_ISL_2710337, EPI_ISL_2731437, EPI_ISL_2941725, EPI_ISL_2941763, EPI_ISL_2943174, EPI_ISL_2943199, EPI_ISL_2943208, EPI_ISL_2943219, EPI_ISL_2943220, EPI_ISL_2982696, EPI_ISL_2982701, EPI_ISL_2982709 | | | |
| see above | Montana Public Health Laboratory | Montana Public Health Laboratory | Carrie Biskupiak; Deborah Gibson; Joy Ritter; Michael Dills; Michelle Mozer |
| EPI_ISL_4060426 | NC State Laboratory of Public Health | Centers for Disease Control and Prevention Division of Viral Diseases, Pathogen Discovery | Alex Burgin; Ben Rambo-Martin; Clinton Paden; Dakota Howard; Dave Wentworth; Dhwani Batra; Jasmine Padilla; Justin Lee; Krista Queen; Kristen Knipe; Kristine Lacek; Mark Burroughs; Matthew Schmerer; Meghan Bentz; Mili Sheth; Peter Cook; Sam Shepard; Sarah Nobles; Suxiang Tong; Vivien Dugan; Yvette Unoarumhi |
| EPI_ISL_3242077 | ND Dept. of Health Laboratory Services-Microbiology | Centers for Disease Control and Prevention Division of Viral Diseases, Pathogen Discovery | Alex Burgin; Ben L. Rambo-Martin; Clinton R. Paden; Dakota Howard; Dave Wentworth; Dhwani Batra; Jasmine Padilla; Justin Lee; Krista Queen; Kristen Knipe; Kristine Lacek; Mark Burroughs; Matthew Schmerer; Meghan Bentz; Mili Sheth; Peter Cook; Sam Shepard; Sarah Nobles; Suxiang Tong; Vivien Dugan; Yvette Unoarumhi |
| EPI_ISL_2689917, EPI_ISL_2689927, EPI_ISL_2689930, EPI_ISL_2987007 | NE Public Health Laboratory | Centers for Disease Control and Prevention Division of Viral Diseases, Pathogen Discovery | Alison Laufer Halpin; Ben L. Rambo-Martin; Clinton R. Paden; Dakota Howard; Darlene Wagner; Dave Wentworth; Dhwani Batra; Jasmine Padilla; Justin Lee; Katie Dillon; Krista Queen; Kristen Knipe; Kristine Lacek; Mark Burroughs; Matthew Schmerer; Mili Sheth; Peter Cook; Sam Shepard; Sarah Nobles; Shoshona Le; Suxiang Tong; Vivien Dugan; Yvette Unoarumhi |
| EPI_ISL_2689938, EPI_ISL_2787857, EPI_ISL_2787928, EPI_ISL_3800472 | NJ Public Health and Environmental Laboratories | Centers for Disease Control and Prevention Division of Viral Diseases, Pathogen Discovery | Alex Burgin; Alison Laufer Halpin; Ben L. Rambo-Martin; Ben Rambo-Martin; Clinton Paden; Clinton R. Paden; Dakota Howard; Darlene Wagner; Dave Wentworth; Dhwani Batra; Jasmine Padilla; Justin Lee; Katie Dillon; Krista Queen; Kristen Knipe; Kristine Lacek; Mark Burroughs; Matthew Schmerer; Meghan Bentz; Mili Sheth; Peter Cook; Sam Shepard; Sarah Nobles; Shoshona Le; Suxiang Tong; Vivien Dugan; Yvette Unoarumhi |
| EPI_ISL_2443124, EPI_ISL_2557117, EPI_ISL_2557131, EPI_ISL_2557134, EPI_ISL_2557148, EPI_ISL_2557157, EPI_ISL_2649830, EPI_ISL_2649831, EPI_ISL_2842649, EPI_ISL_2842660, EPI_ISL_2842684, EPI_ISL_3023754, EPI_ISL_3129992 | | | |
| see above | NJDOH, Public Health and Environmental Laboratories | NJ_PHEL | Allison Roder; Byeong Jeong; Chelsea San Filippo; Dana Woell; Jacquelyn Deverell; Lindsey Bodnar; Ryan Pachucki; Shiv K. Verma |
| EPI_ISL_2627155, EPI_ISL_2627177, EPI_ISL_2627179, EPI_ISL_3031060, EPI_ISL_3031066, EPI_ISL_3031067 | NORTHWELL HEALTH LABORATORIES | Wadsworth Center, New York State Department of Health | Alexis Russell; Catharine Prussing; Daryl M. Lamson; Erasmus Schneider; Erica Lasek-Nesselquist; John Kelly; Jonathan Plitnick; Kirsten St. George; Matthew Shudt; Melissa A Leisner; Navjot Singh |
| EPI_ISL_2821280, EPI_ISL_2821281, EPI_ISL_2821282, EPI_ISL_2821283, EPI_ISL_2821284, EPI_ISL_3134752, EPI_ISL_3134754, EPI_ISL_3134755, EPI_ISL_3134759, EPI_ISL_3134760, EPI_ISL_3134763, EPI_ISL_3134765, EPI_ISL_3134768, EPI_ISL_3134769, EPI_ISL_3134770, EPI_ISL_3134771, EPI_ISL_3134772, EPI_ISL_3134774, EPI_ISL_3134775, EPI_ISL_3134776, EPI_ISL_3134778, EPI_ISL_3134779, EPI_ISL_3134780, EPI_ISL_3134781, EPI_ISL_3134783, EPI_ISL_3134784, EPI_ISL_3134785, EPI_ISL_3134786, EPI_ISL_3134789, EPI_ISL_3134790, EPI_ISL_3134792, EPI_ISL_3134793, EPI_ISL_3134794, EPI_ISL_3134795, EPI_ISL_3134796, EPI_ISL_3134797, EPI_ISL_3134798, EPI_ISL_3134799, EPI_ISL_3134800, EPI_ISL_3134801, EPI_ISL_3134803, EPI_ISL_3134804, EPI_ISL_3134805, EPI_ISL_3134806, EPI_ISL_3134808, EPI_ISL_3134813, EPI_ISL_3447458, EPI_ISL_3447490, EPI_ISL_3447491, EPI_ISL_3447492, EPI_ISL_3447493, EPI_ISL_3447494, EPI_ISL_3447495, EPI_ISL_3447496, EPI_ISL_3447498, EPI_ISL_3447500, EPI_ISL_3447501, EPI_ISL_3447502, EPI_ISL_3447503, EPI_ISL_3447504, EPI_ISL_3447505, EPI_ISL_3447506, EPI_ISL_3447507, EPI_ISL_3447508, EPI_ISL_3447510, EPI_ISL_3447511, EPI_ISL_3447512, EPI_ISL_3447513, EPI_ISL_3447514, EPI_ISL_3447515, EPI_ISL_3447516, EPI_ISL_3447517, EPI_ISL_3447518, EPI_ISL_3447519, EPI_ISL_3447521, EPI_ISL_3447522, EPI_ISL_3447524, EPI_ISL_3447525, EPI_ISL_3447526, EPI_ISL_3447527, EPI_ISL_3447528, EPI_ISL_3447529, EPI_ISL_3447530, EPI_ISL_3447531, EPI_ISL_3447533, EPI_ISL_3447534, EPI_ISL_3447535, EPI_ISL_3447536, EPI_ISL_3447537, EPI_ISL_3447538, EPI_ISL_3447539, EPI_ISL_3447540, EPI_ISL_3447541, EPI_ISL_3447542, EPI_ISL_3447543, EPI_ISL_3447545, EPI_ISL_3447546, EPI_ISL_3447547, EPI_ISL_3447548, EPI_ISL_3447549, EPI_ISL_3447550, EPI_ISL_3447551, EPI_ISL_3447552, EPI_ISL_3447553, EPI_ISL_3447554, EPI_ISL_3447556, EPI_ISL_3447557, EPI_ISL_3447558, EPI_ISL_3447559, EPI_ISL_3447560, EPI_ISL_3447561, EPI_ISL_3447562, EPI_ISL_3447563, EPI_ISL_3447564, EPI_ISL_3447565, EPI_ISL_3447566, EPI_ISL_3447567, EPI_ISL_3447568, EPI_ISL_3447594, EPI_ISL_3447595, EPI_ISL_3447596, EPI_ISL_3447597, EPI_ISL_3703782, EPI_ISL_3703783, EPI_ISL_3703784, EPI_ISL_3703785, EPI_ISL_3703787, EPI_ISL_3703788, EPI_ISL_3703789, EPI_ISL_3703792, EPI_ISL_3703794, EPI_ISL_3703796, EPI_ISL_3703797, EPI_ISL_3703798, EPI_ISL_3703799, EPI_ISL_3703801, EPI_ISL_3703813, EPI_ISL_3703816, EPI_ISL_3703823, EPI_ISL_3703827, EPI_ISL_3703831, EPI_ISL_3703834, EPI_ISL_3703846, EPI_ISL_3703849, EPI_ISL_3703853, EPI_ISL_3703868, EPI_ISL_3703876, EPI_ISL_3703886, EPI_ISL_3703905, EPI_ISL_3703909, EPI_ISL_3703912, EPI_ISL_3703917, EPI_ISL_3703928, EPI_ISL_3703938, EPI_ISL_3703948, EPI_ISL_3703958, EPI_ISL_3703962, EPI_ISL_3703969, EPI_ISL_3703972, EPI_ISL_3703980, EPI_ISL_3703991, EPI_ISL_3704002, EPI_ISL_3704012, EPI_ISL_3704016, EPI_ISL_3704024, EPI_ISL_3704035, EPI_ISL_3704043, EPI_ISL_3704066, EPI_ISL_3704070, EPI_ISL_3704074 | | | |
| see above | NUPIT/UFPE | WallauLab on behalf of Fiocruz COVID-19 Genomic Surveillance Network | Alexandre Freitas da Silva; Antonio Mauro Rezende; Cassia Docena; Constância Flávia Junqueira Ayres; Cássia Docena; Duschinka Ribeiro Duarte Guedes; Elisama Helvecio; Filipe Zimmer Dezordi; Gabriel Luz Wallau; Gustavo Barbosa de Lima; Lais Ceschini Machado; Larissa Krokovsky; Laís Ceschini Machado; Lilian Caroliny Amorim Silva; Maira Galdino da Rocha Pitta; Marcelo Henrique dos Santos Paiva; Matheus Filgueira Bezerra; Michelly Cristiny Pereira; Rômulo Pessoa e Silva; Sinval Pinto Brandão Filho |
| EPI_ISL_2663535, EPI_ISL_2753941 | NYU Langone Health | Departments of Pathology and Medicine, New York University School of Medicine | Adriana Heguy; Christian Marier; Dacia Dimartino; Emily Guzman; Gael Westby; Guiqing Wang; Paolo Cotzia; Paul Zappile; Peter Meyn; Sitharam Ramaswami; Yutong Zhang |
| EPI_ISL_3063113 | National Institute of Public Health | National Institute of Public Health | Alexander Nagy; Dusan Trnka; Helena Jirincova; Jaromira Vecerova; Timotej Suri |
| EPI_ISL_2621962, EPI_ISL_2621963 | National Public Health Laboratory, National Centre for Infectious Diseases | National Public Health Laboratory, National Centre for Infectious Diseases | Lin Cui; Raymond Tzer Pin Lin; Royce Ang; Tze Minn Mak; Zhenyang Zhou |
| EPI_ISL_2308536, EPI_ISL_2382430, EPI_ISL_2466750, EPI_ISL_2597931, EPI_ISL_2689123 | Nebraska Public Health Laboratory | NPHL COVID-19 Response Team | NPHL COVID-19 Response Team |
| EPI_ISL_2612507 | Nevada State Public Health Laboratory | Nevada State Public Health Laboratory | Andrew Gorzalski; Mark Pandori |
| EPI_ISL_2652076, EPI_ISL_2941647, EPI_ISL_3086158 | New Mexico Department of Health Scientific Laboratory | New Mexico Department of Health Scientific Laboratory | ; Anastacia Griego-Fisher; D'eldra Malone; Ellie Johnson; Jennifer Benoit; Keila Gutierrez; Linda Salazar; Ratheesh Rajan |
| EPI_ISL_3230464 | NordLab Kajaani | Expert Microbiology, National Institute for Health and Welfare | Carita Savolainen-Kopra; Erika Lindh; Haider al-Hello; Jani Halkilahti; Kirsi Liitsola; Niina Ikonen; Olli Vapalahti; Pekka Ellonen; Phuoc Truong; Päivi Laurila; Ravi Kant; Sari Hannula; Soile Blomqvist; Teemu Smura |
| EPI_ISL_2736189, EPI_ISL_2736201 | Northwest Laboratory | Seattle Flu Study | Amanda Adler; Barry R. Lutz; Benjamin Pelle; Brian Hiatt; Caitlin R. Wolf; Chris D. Frazar; Deborah A. Nickerson; Elisabeth Brandstetter; Erica Ryke; Geoff Melly; Helen Y. Chu; Janet A. Englund; Jay Shendure; Jover Lee; Kairsten Fay; Kirsten Lacombe; Lea M. Starita; Mark J. Rieder; Matthew Richardson; Matthew Thompson; Melissa Truong; Michael Boeckh; Michael Famulare; Misja Ilcisin; Peter D. Han; Philip Dykema; Romesh Gautom; Scott Lindquist; Thomas R. Sibley; Trevor Bedford |
| EPI_ISL_2373049, EPI_ISL_2842596, EPI_ISL_2842597, EPI_ISL_2842605, EPI_ISL_2842608, EPI_ISL_2842612, EPI_ISL_2842613 | | | |
| see above | Northwestern Memorial Hospital | Northwestern University - Ozer Lab | Chad J. Achenbach; Chao Qi; Egon A. Ozer; Judd F. Hultquist; Lacy M. Simons; Lawrence J. Jennings; Michael G. Ison; Ramon Lorenzo-Redondo; Taylor J. Dean |
| EPI_ISL_3388710 | Northwestern Memorial Hospital | RIPHL at Rush University Medical Center | Amber Kimble; Chao Qi; Edith Perez; Felix Araujo Perez; Kevin Kunstman; Laura Furtado; Marieta Hyde; Max Kolton; Stefan Green |
| EPI_ISL_2838856, EPI_ISL_2838857, EPI_ISL_2838858, EPI_ISL_2838859, EPI_ISL_2838860, EPI_ISL_2838861, EPI_ISL_2838862, EPI_ISL_2838863, EPI_ISL_2838864, EPI_ISL_2838865, EPI_ISL_2838866, EPI_ISL_2838867, EPI_ISL_2838869, EPI_ISL_2838870, EPI_ISL_2838871, EPI_ISL_2838872, EPI_ISL_2838873, EPI_ISL_2838874, EPI_ISL_2838875, EPI_ISL_2838876, EPI_ISL_2838877, EPI_ISL_2838878, EPI_ISL_2838879, EPI_ISL_2838880, EPI_ISL_2838881, EPI_ISL_2838882, EPI_ISL_2838883 | | | |
| see above | OHSU Lab Services Molecular Microbiology | Oregon SARS-CoV-2 Genome | '[UsersoroakBox SyncPHI_SARS-COV-2_Sample_Sheet_infoGISAID_Submission_Infometa_master_2021.xlsx]release20_final'!D:AC; 26; FALSE); VLOOKUP(C466; VLOOKUP(C468; VLOOKUP(C474; VLOOKUP(C477; VLOOKUP(C483; VLOOKUP(C488; VLOOKUP(C502; VLOOKUP(C503; VLOOKUP(C504; |

| | Lab | Sequencing Center | VLOOKUP(C505; VLOOKUP(C508; VLOOKUP(C510; VLOOKUP(C516; VLOOKUP(C517; VLOOKUP(C518; VLOOKUP(C519; VLOOKUP(C521; VLOOKUP(C522; VLOOKUP(C525; VLOOKUP(C526; VLOOKUP(C528; VLOOKUP(C529; VLOOKUP(C530; VLOOKUP(C533; VLOOKUP(C534; VLOOKUP(C539; VLOOKUP(C547 |
|---|---|---|---|
| EPI_ISL_2689979, EPI_ISL_2689981, EPI_ISL_2689982, EPI_ISL_2689984, EPI_ISL_2787421, EPI_ISL_2787436, EPI_ISL_2787447, EPI_ISL_2787450, EPI_ISL_2787453, EPI_ISL_2787459, EPI_ISL_2840415, EPI_ISL_2987015, EPI_ISL_3355222 | | | |
| see above | OR State PHL-Virology/Immunology Section | Centers for Disease Control and Prevention Division of Viral Diseases, Pathogen Discovery | Alex Burgin; Alison Laufer Halpin; Ben L. Rambo-Martin; Ben Rambo-Martin; Clinton Paden; Clinton R. Paden; Dakota Howard; Darlene Wagner; Dave Wentworth; Dhwani Batra; Jasmine Padilla; Justin Lee; Katie Dillon; Krista Queen; Kristen Knipe; Kristine Lacek; Mark Burroughs; Matthew Schmerer; Meghan Bentz; Mili Sheth; Peter Cook; Sam Shepard; Sarah Nobles; Shoshona Le; Suxiang Tong; Vivien Dugan; Yvette Unoarumhi |
| EPI_ISL_2966615 | Ochsner Health | Bioinfoexperts, LLC | Amy Feehan; David J. Nolan; Judy Crabtree; Julia-Garcia-Diaz; Lucio Miele; Rebecca Rose; Susanna L. Lamers; Tessa LaFleur |
| EPI_ISL_2621529 | Omega Diagnostics at Mounes | Omega Diagnostics at Mounes | Ashleigh McGrail; Cynthia Corley; Latira Haynes-Jacob; MD; Vivek Khare |
| EPI_ISL_2423535, EPI_ISL_2604871 | Omics Sciences Laboratory | Omics Sciences Laboratory | Darlyn Amaya; Derly Andrade Molina; Gabriel Morey León; Juan Carlos Fernández Cadena; Rubén Armas González |
| EPI_ISL_2928154, EPI_ISL_2928156, EPI_ISL_2928158, EPI_ISL_2928161, EPI_ISL_2928167, EPI_ISL_2928169, EPI_ISL_2928172, EPI_ISL_2928173, EPI_ISL_2928176, EPI_ISL_2928183, EPI_ISL_2928193, EPI_ISL_2928195, EPI_ISL_2928196, EPI_ISL_3010309, EPI_ISL_3099977 | | | |
| see above | Orange County Public Health Laboratory | Orange County Public Health Laboratory | Angelica Torres; Julia Wolfe |
| EPI_ISL_2424249, EPI_ISL_2424250, EPI_ISL_2473664, EPI_ISL_2473665, EPI_ISL_2473666, EPI_ISL_2473667, EPI_ISL_2557922, EPI_ISL_2557923, EPI_ISL_2557926, EPI_ISL_2604198, EPI_ISL_2604205, EPI_ISL_2604209, EPI_ISL_2612540, EPI_ISL_2612541, EPI_ISL_2612545, EPI_ISL_2631151, EPI_ISL_2631155, EPI_ISL_2631187, EPI_ISL_2727464, EPI_ISL_2727476, EPI_ISL_2727487, EPI_ISL_2790735, EPI_ISL_2790737, EPI_ISL_2790739, EPI_ISL_2790740, EPI_ISL_2790741, EPI_ISL_2790742, EPI_ISL_2790744, EPI_ISL_2790745, EPI_ISL_2790746, EPI_ISL_2893422, EPI_ISL_2893423, EPI_ISL_2893424, EPI_ISL_2893425, EPI_ISL_2893426, EPI_ISL_2893427, EPI_ISL_2893428, EPI_ISL_2893429, EPI_ISL_2928905, EPI_ISL_2933949, EPI_ISL_2955407, EPI_ISL_3010589, EPI_ISL_3010594, EPI_ISL_3048118, EPI_ISL_3048120, EPI_ISL_3067393, EPI_ISL_3090726 | | | |
| see above | Oregon State Public Health Laboratory | Oregon State Public Health Laboratory | Eugene Yeboah; John Fontana and Shane Sevey; Laura Tsaknaridis; Rafia Razzaque; Vanda Makris |
| EPI_ISL_2925501 | Ospedale di Genzano | National Institute for Infectious Diseases (INMI) L. Spallanzani I.R.C.C.S | A Di Caro; B Bartolini; CEM Gruber; E Giombini; F Messina; F Santini; G Bonfiglio; M Rueca; MR Capobianchi; O Butera |
| EPI_ISL_3219518, EPI_ISL_3219519, EPI_ISL_3219520, EPI_ISL_3219521, EPI_ISL_3219522, EPI_ISL_3219523 | Outre mer | National Reference Center for Viruses of Respiratory Infections, Institut Pasteur, Paris | Angela Brisebarre; Camille Capel; Christophe Malabat; Corinne Maufrais; Dominique Rousset; Etienne Simon-Lorière; Frédéric Lemoine; Hub de Bioinformatique et Biostatistique; Louise Lefrançois; Marion Barbet; Maud Vanpeene; Méline Bizard; Sylvie Behillil; Sylvie Van der Werf; Vincent Enouf |
| EPI_ISL_3105045, EPI_ISL_3105048, EPI_ISL_3105049 | Overlake Hospital | Seattle Flu Study | Amanda Adler; Barry R. Lutz; Benjamin Pelle; Brian Hiatt; Caitlin R. Wolf; Chris D. Frazar; Deborah A. Nickerson; Elisabeth Brandstetter; Erica Ryke; Geoff Melly; Helen Y. Chu; Janet A. Englund; Jay Shendure; Jover Lee; Kairsten Fay; Kirsten Lacombe; Lea M. Starita; Mark J. Rieder; Matthew Richardson; Matthew Thompson; Melissa Truong; Michael Boeckh; Michael Famulare; Misja Ilcisin; Peter D. Han; Philip Dykema; Romesh Gautom; Scott Lindquist; Thomas R. Sibley; Trevor Bedford |
| EPI_ISL_2903092 | Oxford Viromics, NDM, University of Oxford; Oxford University Hospitals; Basingstoke and North Hampshire Hospital | COVID-19 Genomics UK (COG-UK) Consortium | Alex Mobbs; Amy Trebes; Anita Justice; Catrin Moore; Christophe Fraser; David Bonsall; David Buck; Emma Wise; George Macintyre; Jessica Lynch; John Todd; Mariateresa de Cesare; Matilde Mori; Monique Andersson; Nathan Moore; Nick Cortes; Robert Shaw; Stephen Kidd; Tanya Golubchik; Timothy Peto |
| EPI_ISL_2493913 | POLICLINICA COVID 19 ITAPETININGA | Instituto Butantan | Antonio Jorge Martins; Claudia Renata dos Santos Barros; David Schlesinger; Debora Botequio Moretti; Dimas Tadeu Covas; Elaine Cristina Marqueze; Elaine Vieira Santos; Evandra Strazza Rodrigues; Heidge Fukumasu; Jayme Augusto de Souza-Neto; José Salvatore Leister Patané; Luiz Alcantara; Luiz Lehmann Coutinho; Maria Carolina Elias; Maurício Lacerda Nogueira; Rafael dos Santos Bezerra; Raul Machado Neto; Rejane Maria Tommasini Grotto; Ricardo Haddad; Sandra Coccuzzo Sampaio Vessoni; Simone Kashima; Svetoslav Nanev Slavov; Vincent Louis Viala |
| EPI_ISL_3536291 | POLO BASE DO TRILHO | Analytical Competence Molecular Epidemiology Lab/ACME, Oswaldo Cruz Foundation, Ceara (FIOCRUZ CE) | Cleber Furtado Aksenen; Fabio Miyajima; Fernando Braga Stehling; Francisco Eder de Moura Lopes; Jamille Maria Mendes Bezerra; Joaquim César do Nascimento Sousa Junior; Pedro Miguel Carneiro Jeronimo; Suzana Porto Almeida e Lucas Delerino; Thais Ferreira de Oliveira; Thais de Oliveira Costa; Ticiane Cavalcante de Souza; Veridiana Pessoa Miyajima |
| EPI_ISL_2493883, EPI_ISL_2493896, EPI_ISL_2493902, EPI_ISL_2493904, EPI_ISL_2493911, EPI_ISL_2493915, EPI_ISL_2493932, EPI_ISL_2493935, EPI_ISL_2493936, EPI_ISL_2494325, EPI_ISL_2494326 | | | |
| see above | PRONTO ATENDIMENTO DE VARGEM GRANDE PAULISTA | Instituto Butantan | Antonio Jorge Martins; Claudia Renata dos Santos Barros; David Schlesinger; Debora Botequio Moretti; Dimas Tadeu Covas; Elaine Cristina Marqueze; Elaine Vieira Santos; Evandra Strazza Rodrigues; Heidge Fukumasu; Jayme Augusto de Souza-Neto; José Salvatore Leister Patané; Luiz Alcantara; Luiz Lehmann Coutinho; Maria Carolina Elias; Maurício Lacerda Nogueira; Rafael dos Santos Bezerra; Raul Machado Neto; Rejane Maria Tommasini Grotto; Ricardo Haddad; Sandra Coccuzzo Sampaio Vessoni; Simone Kashima; Svetoslav Nanev Slavov; Vincent Louis Viala |
| EPI_ISL_2493846 | PS E AMBULATORIO ENGENHEIRO CARDOSO | Instituto Butantan | Antonio Jorge Martins; Claudia Renata dos Santos Barros; David Schlesinger; Debora Botequio Moretti; Dimas Tadeu Covas; Elaine Cristina Marqueze; Elaine Vieira Santos; Evandra Strazza Rodrigues; Heidge Fukumasu; Jayme Augusto de Souza-Neto; José Salvatore Leister Patané; Luiz Alcantara; Luiz Lehmann Coutinho; Maria Carolina Elias; Maurício Lacerda Nogueira; Rafael dos Santos Bezerra; Raul Machado Neto; Rejane Maria Tommasini Grotto; Ricardo Haddad; Sandra Coccuzzo Sampaio Vessoni; Simone Kashima; Svetoslav Nanev Slavov; Vincent Louis Viala |
| EPI_ISL_2493900 | PSF DR ANTONIO PIRES DE ALMEIDA PORTO FELIZ | Instituto Butantan | Antonio Jorge Martins; Claudia Renata dos Santos Barros; David Schlesinger; Debora Botequio Moretti; Dimas Tadeu Covas; Elaine Cristina Marqueze; Elaine Vieira Santos; Evandra Strazza Rodrigues; Heidge Fukumasu; Jayme Augusto de Souza-Neto; José Salvatore Leister Patané; Luiz Alcantara; Luiz Lehmann Coutinho; Maria Carolina Elias; Maurício Lacerda Nogueira; Rafael dos Santos Bezerra; Raul Machado Neto; Rejane Maria Tommasini Grotto; Ricardo Haddad; Sandra Coccuzzo Sampaio Vessoni; Simone Kashima; Svetoslav Nanev Slavov; Vincent Louis Viala |
| EPI_ISL_2349631, EPI_ISL_2349675, EPI_ISL_2446041, EPI_ISL_2446051, EPI_ISL_2446088, EPI_ISL_2446091, EPI_ISL_2446094, EPI_ISL_2631339, EPI_ISL_2631340, EPI_ISL_2631349, EPI_ISL_2631350, EPI_ISL_2631354, EPI_ISL_2631359, EPI_ISL_2631360, EPI_ISL_2631366, EPI_ISL_2631386, EPI_ISL_2631397, EPI_ISL_2631407, EPI_ISL_2634916, EPI_ISL_2634943, EPI_ISL_2827487, EPI_ISL_2827488, EPI_ISL_2827489, EPI_ISL_2827500, EPI_ISL_2923915, EPI_ISL_2923945, EPI_ISL_3010646, EPI_ISL_3010649, EPI_ISL_3010653, EPI_ISL_3010670, EPI_ISL_3010678, EPI_ISL_3010754, EPI_ISL_3010757 | | | |
| see above | Pandemic Response Lab - NYC | Pandemic Response Lab, R&D | Cybill del Castillo; Dylan Law; Haiping Hao; Henry Lee; Isabel Fernandez Escapa; Jon Laurent; Katharine Nelson; Melissa Hopkins; Michael Hammerling; Pradeep Bugga; Shinyoung Clair Kang; Sol Rey; William Ward |
| EPI_ISL_2832117, EPI_ISL_2832124, EPI_ISL_2832127, EPI_ISL_2897033, EPI_ISL_2897056 | Parc Taulí Hospital Universitari | Can Ruti SARS-CoV-2 Sequencing Hub (HUGTiP/IrsiCaixa/IGTP) | Alba Sánchez; Alexia París; Anna Not; Antoni E Bordoy; Bonaventura Clotet; Cristina Casañ; David Panisello; Francesc Catala-Moll; Gemma Clara; Ignacio Blanco; Laia Soler; Lauro Sumoy; Marc Noguera-Julian; Maria Casadellà; Mariona Parera; Mercedes Guerrero; Montserrat Giménez; Pere-Joan Cardona; Pilar Armengol; Roger Paredes; Verónica Saludes; and Elisa Martró on behalf of the Can Ruti SARS-CoV-2 Sequencing Hub |
| EPI_ISL_3050138, EPI_ISL_3050162, EPI_ISL_3398772 | Pathology, UAB | Pathology, UAB | E.J.; Hendrickson; Hernandez, J.; Leal; Lefkowitz; Moates, D.; R.C.; S.M. |
| EPI_ISL_3127929 | Philadelphia Department of Public Health | Centers for Disease Control and Prevention Division of Viral Diseases, Pathogen Discovery | Alex Burgin; Ben L. Rambo-Martin; Clinton R. Paden; Dakota Howard; Dave Wentworth; Dhwani Batra; Jasmine Padilla; Justin Lee; Krista Queen; Kristen Knipe; Kristine Lacek; Mark Burroughs; Matthew Schmerer; Meghan Bentz; Mili Sheth; Peter Cook; Sam Shepard; Sarah Nobles; Suxiang Tong; Vivien Dugan; Yvette Unoarumhi |
| EPI_ISL_2861583 | Platform BIS UZA/UAntwerpen | Labo Klinische Biologie, UZA | Basil Britto Xavier; Christine Lammens; Herman Goossens; Ines Verbesselt; Jasmine Coppens; Kathleen Holemans; Marie Le Mercier; Veerle Matheeussen |
| EPI_ISL_3120103 | Policlinico San Donato | Laboratory of Clinical Microbiology, Virology and Bioemergencies, ASST Fatebenefratelli Sacco - Sacco Hospital | Valeria Micheli |
| EPI_ISL_2925488 | Policlinico Tor Vergata | National Institute for Infectious Diseases (INMI) L. Spallanzani I.R.C.C.S | A Di Caro; B Bartolini; CEM Gruber; E Giombini; F Messina; F Santini; G Bonfiglio; M Rueca; MR Capobianchi; O Butera |
| EPI_ISL_3276769 | Providence Saint John's Health Center | Los Angeles County Public Health Laboratories | P. Hemarajata et al. |
| EPI_ISL_2699447, EPI_ISL_2709231 | Public Health Laboratory, Minnesota Department of Health | University of Minnesota Genomics Center | Corbin Dirkx; Daryl M. Gohl; Jaquelyn Kuriger-Laber; John Garbe; and Sean Wang |
| EPI_ISL_3132189 | Puerto Rico Department of Health | Centers for Disease Control and Prevention, Dengue Branch | Betzabel Flores; Gabriela Paz-Bailey; Gilberto A. Santiago; Glenda Gonzalez; Jorge L. Munoz-Jordan; Keyla Charriez |
| EPI_ISL_2615917, EPI_ISL_2615920 | QLabs | WVU and Marshall University Combined Genomics Core Facilities | James Denvir; Peter Perrotta; Peter Stoilov; Ryan Percifield; Wesley Kimble |
| EPI_ISL_2451428, EPI_ISL_2451463, EPI_ISL_2451473, EPI_ISL_2451509, EPI_ISL_2451518, EPI_ISL_2451524, EPI_ISL_2451538, EPI_ISL_2451549, EPI_ISL_2451555, EPI_ISL_2451559, EPI_ISL_2451565, EPI_ISL_2451566, EPI_ISL_2463008, EPI_ISL_2463013, EPI_ISL_2463014, EPI_ISL_2463015, EPI_ISL_2463017, EPI_ISL_2488822, EPI_ISL_2488823, EPI_ISL_2488824, EPI_ISL_2488825, EPI_ISL_2488827, EPI_ISL_2488829, EPI_ISL_2488830, EPI_ISL_2488831, EPI_ISL_2488832, EPI_ISL_2488833, EPI_ISL_2488834, EPI_ISL_2488835, EPI_ISL_2488837, EPI_ISL_2488840, EPI_ISL_2488841, EPI_ISL_2488844, EPI_ISL_2488850, EPI_ISL_2488851, EPI_ISL_2488853, EPI_ISL_2488854, EPI_ISL_2488855, EPI_ISL_2488856, EPI_ISL_2488857, EPI_ISL_2488860, EPI_ISL_2534519, EPI_ISL_2534521, EPI_ISL_2534522, EPI_ISL_2534524, EPI_ISL_2534525, EPI_ISL_2534526, EPI_ISL_2534527, EPI_ISL_2534528, EPI_ISL_2534529, EPI_ISL_2534530, EPI_ISL_2534531, EPI_ISL_2534532, EPI_ISL_2534553, EPI_ISL_2534554, EPI_ISL_2534555, EPI_ISL_2534556, EPI_ISL_2534557, EPI_ISL_2534558, EPI_ISL_2534560, EPI_ISL_2534565, EPI_ISL_2649948, EPI_ISL_2649950, EPI_ISL_2649951, EPI_ISL_2649952, EPI_ISL_2649953, EPI_ISL_2649955, EPI_ISL_2649956, EPI_ISL_2649957, EPI_ISL_2649958, EPI_ISL_2649959, EPI_ISL_2649960, EPI_ISL_2649962, EPI_ISL_2649963, EPI_ISL_2649964, EPI_ISL_2649966, EPI_ISL_2649967, EPI_ISL_2649968, EPI_ISL_2649969, EPI_ISL_2649979, EPI_ISL_2649980, EPI_ISL_2649981, EPI_ISL_2649982, EPI_ISL_2649983, EPI_ISL_2649984, EPI_ISL_2649987, EPI_ISL_2686360, EPI_ISL_2686361, EPI_ISL_2686362, EPI_ISL_2686364, EPI_ISL_2686365, EPI_ISL_2686366, EPI_ISL_2686367, EPI_ISL_2686370, EPI_ISL_2686373, EPI_ISL_2686374, EPI_ISL_2686375, EPI_ISL_2686376, EPI_ISL_2686377, EPI_ISL_2686380, EPI_ISL_2686381, EPI_ISL_2686386, EPI_ISL_2686387, EPI_ISL_2686388, EPI_ISL_2872074, EPI_ISL_2876105, EPI_ISL_2876106, EPI_ISL_2876107, EPI_ISL_2876109, EPI_ISL_2876110, EPI_ISL_2876113, EPI_ISL_2876114, EPI_ISL_2876115, EPI_ISL_2876116, EPI_ISL_2876119, EPI_ISL_2876120, EPI_ISL_2876121, EPI_ISL_2876122, EPI_ISL_2876123, EPI_ISL_2876124, EPI_ISL_2876125, EPI_ISL_2876126, EPI_ISL_2876127, EPI_ISL_2876128, EPI_ISL_2876129, EPI_ISL_2876132, EPI_ISL_2876134, EPI_ISL_2876135, EPI_ISL_2876137, EPI_ISL_2876142, EPI_ISL_2876145, EPI_ISL_2876146, EPI_ISL_2876148, EPI_ISL_2876149, EPI_ISL_2876153, EPI_ISL_2876154, EPI_ISL_2876155, EPI_ISL_2876160, EPI_ISL_2876162, EPI_ISL_2876163, EPI_ISL_2876166, EPI_ISL_2876167, EPI_ISL_2876616, EPI_ISL_2876617, EPI_ISL_2876619, EPI_ISL_2876620, EPI_ISL_2876621, EPI_ISL_2876624, EPI_ISL_2876626, EPI_ISL_2876628, EPI_ISL_2876629, EPI_ISL_2876630, EPI_ISL_2876631, EPI_ISL_2876632, EPI_ISL_2876633, EPI_ISL_2876636, EPI_ISL_2876638, EPI_ISL_2876639, EPI_ISL_2876640, EPI_ISL_2876644, EPI_ISL_2876645, EPI_ISL_2876646, EPI_ISL_2876647, EPI_ISL_2876649, EPI_ISL_2876650, EPI_ISL_2876651, EPI_ISL_2876652, EPI_ISL_2876653, EPI_ISL_2876654, EPI_ISL_2876656, EPI_ISL_2876660, EPI_ISL_2876661, EPI_ISL_2876662, EPI_ISL_2876663, EPI_ISL_2876665, EPI_ISL_2876666, EPI_ISL_2876667, EPI_ISL_2876668, EPI_ISL_2876672, EPI_ISL_2876673, EPI_ISL_2876675, EPI_ISL_2876676, EPI_ISL_2876677, EPI_ISL_2876734, EPI_ISL_2876735, EPI_ISL_2876737, EPI_ISL_2876741, EPI_ISL_2876742, EPI_ISL_2876743, EPI_ISL_2876744, EPI_ISL_2876745, EPI_ISL_2876746, EPI_ISL_2876748, EPI_ISL_2876749, EPI_ISL_2876750, EPI_ISL_2876752, EPI_ISL_2876753, EPI_ISL_2876755, EPI_ISL_2876756, EPI_ISL_2876757, EPI_ISL_2876758, EPI_ISL_2876760, EPI_ISL_2876762, EPI_ISL_2876763, EPI_ISL_2876764, EPI_ISL_2876765, EPI_ISL_2876766, EPI_ISL_2876768, EPI_ISL_2876770, EPI_ISL_2876771, EPI_ISL_2876772, EPI_ISL_2876774, EPI_ISL_2876775, EPI_ISL_2876776, EPI_ISL_2876777, EPI_ISL_2876778, EPI_ISL_2876781, EPI_ISL_2876782, EPI_ISL_2876783, EPI_ISL_2876785, EPI_ISL_2876788, EPI_ISL_2876789, EPI_ISL_2876791, EPI_ISL_2876793, EPI_ISL_2876794, EPI_ISL_2876795, EPI_ISL_2876798, EPI_ISL_2876799, EPI_ISL_2876800, EPI_ISL_2876801, EPI_ISL_2876803, EPI_ISL_2876804, EPI_ISL_2876806, EPI_ISL_2876807, EPI_ISL_2876809, EPI_ISL_2876810, EPI_ISL_2876813, EPI_ISL_2876814, EPI_ISL_2876818, EPI_ISL_2876824, EPI_ISL_2876825, EPI_ISL_2876827, EPI_ISL_2876830, EPI_ISL_2876831, EPI_ISL_2876832, EPI_ISL_2876834, EPI_ISL_2876837, EPI_ISL_2876838, EPI_ISL_2876839, EPI_ISL_2876840, EPI_ISL_2876841, EPI_ISL_2876842, EPI_ISL_2876843, EPI_ISL_2876844, EPI_ISL_2876845, EPI_ISL_2876846, EPI_ISL_2876849, EPI_ISL_2876852, EPI_ISL_2876853, EPI_ISL_2876854, EPI_ISL_2876855, EPI_ISL_2876858, EPI_ISL_2876859, EPI_ISL_2876860, EPI_ISL_2876861, EPI_ISL_2876862, EPI_ISL_2876866, EPI_ISL_2876868, EPI_ISL_2876869, EPI_ISL_2876870, EPI_ISL_2876874, EPI_ISL_2876875, EPI_ISL_2876877, EPI_ISL_2876879, EPI_ISL_2876881, EPI_ISL_2876882, EPI_ISL_2876884, EPI_ISL_2876885, EPI_ISL_2876886, EPI_ISL_2876888, EPI_ISL_2876892, EPI_ISL_2876893, EPI_ISL_2876896, EPI_ISL_2876897, EPI_ISL_2876898, EPI_ISL_2876899, EPI_ISL_2876901, EPI_ISL_2876902, EPI_ISL_2876904, EPI_ISL_2876906, EPI_ISL_2876908, EPI_ISL_2876909, EPI_ISL_2876914, EPI_ISL_2876916, EPI_ISL_2876917, EPI_ISL_2876918, EPI_ISL_2876919, EPI_ISL_2876920, EPI_ISL_2876922, EPI_ISL_2876924, EPI_ISL_2876925, EPI_ISL_2876926, EPI_ISL_2876927, EPI_ISL_2876928, EPI_ISL_2876930, EPI_ISL_2876933, EPI_ISL_2876934, EPI_ISL_3085829, EPI_ISL_3085830, EPI_ISL_3085831, EPI_ISL_3085836, EPI_ISL_3085840, EPI_ISL_3085841, EPI_ISL_3085843, EPI_ISL_3085848, EPI_ISL_3085849, EPI_ISL_3085850, EPI_ISL_3085851, EPI_ISL_3085852, EPI_ISL_3085854, EPI_ISL_3085855, EPI_ISL_3085856, EPI_ISL_3085858, EPI_ISL_3085862, EPI_ISL_3085864, EPI_ISL_3085866, EPI_ISL_3303840, EPI_ISL_3303843, EPI_ISL_3303845, EPI_ISL_3303847, EPI_ISL_3303850, EPI_ISL_3303852, EPI_ISL_3303853, EPI_ISL_3303855, EPI_ISL_3303856, EPI_ISL_3303857, EPI_ISL_3303859, EPI_ISL_3303860, EPI_ISL_3303865, EPI_ISL_3303867, EPI_ISL_3303868, EPI_ISL_3303872, EPI_ISL_3303875, EPI_ISL_3303876, EPI_ISL_3303879, EPI_ISL_3303881, EPI_ISL_3303884, EPI_ISL_3303886, EPI_ISL_3303887, EPI_ISL_3303890, EPI_ISL_3303891, EPI_ISL_3303893, EPI_ISL_3303897, EPI_ISL_3303898, EPI_ISL_3303900, EPI_ISL_3303901, EPI_ISL_3303904, EPI_ISL_3303906, EPI_ISL_3303909, EPI_ISL_3303912, EPI_ISL_3303916, EPI_ISL_3303917, EPI_ISL_3303918, EPI_ISL_3304999, EPI_ISL_3305007, EPI_ISL_3305011, EPI_ISL_3305013, EPI_ISL_3305015, EPI_ISL_3305027, EPI_ISL_3305033, EPI_ISL_3305039, EPI_ISL_3305040, EPI_ISL_3305045, EPI_ISL_3305056, EPI_ISL_3305065, EPI_ISL_3305070, EPI_ISL_3305072, EPI_ISL_3305078, EPI_ISL_3305080, EPI_ISL_3305093, EPI_ISL_3305098, EPI_ISL_3305099, EPI_ISL_3305100, EPI_ISL_3305105, EPI_ISL_3305111, EPI_ISL_3305122, EPI_ISL_3305124, EPI_ISL_3305130, EPI_ISL_3305133, EPI_ISL_3305135, EPI_ISL_3305140, EPI_ISL_3305143, EPI_ISL_3305148, EPI_ISL_3305154, EPI_ISL_3305155, EPI_ISL_3305156, EPI_ISL_3305165, EPI_ISL_3305170, EPI_ISL_3305171, EPI_ISL_3305183 | | | |
| see above | Quest Diagnostics Incorporated | Centers for Disease Control and | A. Gerasimova; A. Perez; Adrian Paskey; B. Anderson; Benjamin Rambo-Martin; Christopher Gulvick; Clinton R. Paden; Dakota Howard; Darlene Wagner; Dhwani Batra; Duncan MacCannell; F. Lacbawan; I. A. Shlyakhter; Jason Caravas; K.E. Livingston; Kara Moser; L.E. Bernstein; M. Hua; Matthew |

| | | | |
|---|---|---|---|
| | | Prevention Division of Viral Diseases, Pathogen Discovery | Schmerer; P. Tanpaiboon; Peter W. Cook; R. M. Kagan; R. Owen; R. V. Rolando; S. H. Rosenthal; Scott Sammons; Shatavia Morrison; Y. Liu; Yvette Unoarumhi |
| EPI_ISL_2645357, EPI_ISL_2787984, EPI_ISL_2787987, EPI_ISL_2840374, EPI_ISL_3355373 | RI State Health Laboratories | Centers for Disease Control and Prevention Division of Viral Diseases, Pathogen Discovery | Alex Burgin; Alison Laufer Halpin; Ben L. Rambo-Martin; Ben Rambo-Martin; Clinton Paden; Clinton R. Paden; Dakota Howard; Darlene Wagner; Dave Wentworth; Dhwani Batra; Jasmine Padilla; Justin Lee; Katie Dillon; Krista Queen; Kristen Knipe; Kristine Lacek; Mark Burroughs; Matthew Schmerer; Meghan Bentz; Mili Sheth; Peter Cook; Sam Shepard; Sarah Nobles; Shoshona Le; Suxiang Tong; Vivien Dugan; Yvette Unoarumhi |
| EPI_ISL_3091694 | Respiratory Viruses Branch, Centers for Disease Control and Prevention | Respiratory Viruses Branch, Centers for Disease Control and Prevention | Anderson, B.; Batra, D.; Bernstein, L.; C.R.; Caravas, J.; Cook; Gerasimova, A.; Gulvick, C.; Howard, D.; Hua, M.; I.A.; Kagan; Lacbawan, F.; Liu, Y.; Livingston, K.; MacCannell, D.; Morrison, S.; Moser, K.; Owen, R.; P.W.; Paden; Paskey, A.; Perez, A.; R.M.; R.V.; Rambo-Martin, B.; Rolando; Rosenthal; S.H.; Sammons, S.; Schmerer, M.; Shlyakhter; Tanpaiboon, P.; Unoarumhi, Y.; Wagner, D. |
| EPI_ISL_3982730 | Ribeirao Preto | Ribeirao Preto | Antonio Jorge Martins; Claudia Renata dos Santos Barros; David Schlesinger; Debora Botequio Moretti; Dimas Tadeu Covas; Elaine Cristina Marqueze; Elaine Vieira Santos; Evandra Strazza Rodrigues; Heidge Fukumasu; Jayme Augusto de Souza-Neto; José Salvatore Leister Patané; Luiz Alcantara; Luiz Lehmann Coutinho; Maria Carolina Elias; Maurício Lacerda Nogueira; Rafael dos Santos Bezerra; Raul Machado Neto; Rejane Maria Tommasini Grotto; Ricardo Haddad; Sandra Coccuzzo Sampaio Vessoni; Simone Kashima; Svetoslav Nanev Slavov; Vincent Louis Viala |
| EPI_ISL_3982693, EPI_ISL_3982697, EPI_ISL_3982701, EPI_ISL_3982711 | Ribeirao Preto SP | Laboratorio de Biologia Molecular Hemocentro de Ribeirao Preto | Antonio Jorge Martins; Claudia Renata dos Santos Barros; David Schlesinger; Debora Botequio Moretti; Dimas Tadeu Covas; Elaine Cristina Marqueze; Elaine Vieira Santos; Evandra Strazza Rodrigues; Heidge Fukumasu; Jayme Augusto de Souza-Neto; José Salvatore Leister Patané; Luiz Alcantara; Luiz Lehmann Coutinho; Maria Carolina Elias; Maurício Lacerda Nogueira; Rafael dos Santos Bezerra; Raul Machado Neto; Rejane Maria Tommasini Grotto; Ricardo Haddad; Sandra Coccuzzo Sampaio Vessoni; Simone Kashima; Svetoslav Nanev Slavov; Vincent Louis Viala |
| EPI_ISL_2691942, EPI_ISL_2931090, EPI_ISL_2931092, EPI_ISL_2931093, EPI_ISL_2931094, EPI_ISL_2931100, EPI_ISL_2931102, EPI_ISL_3388714, EPI_ISL_3388715, EPI_ISL_3388808 | | | |
| see above | Rush University Medical Center | RIPHL at Rush University Medical Center | Diane Springer; Edith Perez; Felix Araujo Perez; Joyce Houlihan; Kevin Kunstman; Laura Furtado; Marieta Hyde; Mary Hayden; Max Kolton; Stefan Green |
| EPI_ISL_2925707 | S. Camillo | National Institute for Infectious Diseases (INMI) L. Spallanzani I.R.C.C.S | A Di Caro; B Bartolini; CEM Gruber; E Giombini; F Messina; F Santini; G Bonfiglio; M Rueca; MR Capobianchi; O Butera |
| EPI_ISL_2840624, EPI_ISL_2840626, EPI_ISL_2840628 | S.C. Laboratorio Analisi, ASL 3 Liguria | U.O. Igiene, Ospedale Policlinico San Martino | Bruzzone Bianca; Caligiuri Patrizia; De Pace Vanessa; Domnich Alexander; Icardi Giancarlo on behalf of SARS-CoV-2 ITALIAN RESEARCH ENTERPRISE-(SCIRE) Collaborative Group; Orsi Andrea; Ricucci Valentina; Spitaleri Antonino |
| EPI_ISL_2493909 | SANTA CASA SALTO DE PIRAPORA | Instituto Butantan | Antonio Jorge Martins; Claudia Renata dos Santos Barros; David Schlesinger; Debora Botequio Moretti; Dimas Tadeu Covas; Elaine Cristina Marqueze; Elaine Vieira Santos; Evandra Strazza Rodrigues; Heidge Fukumasu; Jayme Augusto de Souza-Neto; José Salvatore Leister Patané; Luiz Alcantara; Luiz Lehmann Coutinho; Maria Carolina Elias; Maurício Lacerda Nogueira; Rafael dos Santos Bezerra; Raul Machado Neto; Rejane Maria Tommasini Grotto; Ricardo Haddad; Sandra Coccuzzo Sampaio Vessoni; Simone Kashima; Svetoslav Nanev Slavov; Vincent Louis Viala |
| EPI_ISL_2494169, EPI_ISL_2494177, EPI_ISL_2494189, EPI_ISL_2494196 | SANTA CASA SANTA RITA DO PASSA QUATRO | Instituto Butantan | Antonio Jorge Martins; Claudia Renata dos Santos Barros; David Schlesinger; Debora Botequio Moretti; Dimas Tadeu Covas; Elaine Cristina Marqueze; Elaine Vieira Santos; Evandra Strazza Rodrigues; Heidge Fukumasu; Jayme Augusto de Souza-Neto; José Salvatore Leister Patané; Luiz Alcantara; Luiz Lehmann Coutinho; Maria Carolina Elias; Maurício Lacerda Nogueira; Rafael dos Santos Bezerra; Raul Machado Neto; Rejane Maria Tommasini Grotto; Ricardo Haddad; Sandra Coccuzzo Sampaio Vessoni; Simone Kashima; Svetoslav Nanev Slavov; Vincent Louis Viala |
| EPI_ISL_2957883, EPI_ISL_2958106, EPI_ISL_2986110, EPI_ISL_2986642, EPI_ISL_2986643, EPI_ISL_2986644, EPI_ISL_3587525 | | | |
| see above | SARS-CoV-2 Sequencing Castilla y Leon-Spain Consortium | SARS-CoV-2 Sequencing Castilla y Leon-Spain Consortium | Antonio Orduña-Domingo; Carlos Fuster Foz; Carmen Aldea-Mansilla; Carmen Gimeno Crespo; David Abad; Gabriel March Rosello; Gabriel March Rossello; Gregoria Megías Lobón; Jose María Eiros Bouza; Laura Sánchez de Prada; M. Isabel Fernandez-Natal; Marta Dominguez-Gil; Marta Hernandez; María Antonia García Castro; Mª Fe Brezmes-Valdivieso; Noelia Arenal Andrés; Silvia Rojo; Sonsoles Garcinuño Pérez |
| EPI_ISL_2621771, EPI_ISL_2621772, EPI_ISL_2828304, EPI_ISL_2930565 | SARS-CoV-2 testing team, National Institute of Infectious Diseases | Pathogen Genomics Center, National Institute of Infectious Diseases | Hazuka Y Furihata; Hiromizu Takahashi; Kentaro Itokawa; Makoto Kuroda; Masanori Hashino; Masumichi Saito; Naomi Nojiri; Nozomu Hanaoka; Rina Tanaka; Sana Uchikoba; Tsuguto Fujimoto; Tsuyoshi Sekizuka |
| EPI_ISL_2828387, EPI_ISL_2828388 | SC (UCO) Igiene e Sanità Pubblica, ASUGI, Trieste | ARGO Laboratorio Genomica ed Epigenomica | Barbone F; Breda C; Busetti M; D'Agaro P; Dal Monego S; Degasperi M; Fontana F; Licastro D; Marcello A; Piscianz E; Segat L |
| EPI_ISL_2828404 | SC (UCO) Igiene e Sanità Pubblica, ASUGI, Trieste and SC Microbiologia e Virologia, ASFO, Pordenone | ARGO Laboratorio Genomica ed Epigenomica | Barbone F; Breda C; D'Agaro P; Dal Monego S; De Rosa R; Degasperi M; Licastro D; Marcello A; Segat L |
| EPI_ISL_2494100 | SECAO CENTRO DE DIAGNOSTICO SECEDI | Instituto Butantan | Antonio Jorge Martins; Claudia Renata dos Santos Barros; David Schlesinger; Debora Botequio Moretti; Dimas Tadeu Covas; Elaine Cristina Marqueze; Elaine Vieira Santos; Evandra Strazza Rodrigues; Heidge Fukumasu; Jayme Augusto de Souza-Neto; José Salvatore Leister Patané; Luiz Alcantara; Luiz Lehmann Coutinho; Maria Carolina Elias; Maurício Lacerda Nogueira; Rafael dos Santos Bezerra; Raul Machado Neto; Rejane Maria Tommasini Grotto; Ricardo Haddad; Sandra Coccuzzo Sampaio Vessoni; Simone Kashima; Svetoslav Nanev Slavov; Vincent Louis Viala |
| EPI_ISL_2494161, EPI_ISL_2494170, EPI_ISL_2494178 | SECRETARIA DE SAUDE DE SANTA GERTRUDES | Instituto Butantan | Antonio Jorge Martins; Claudia Renata dos Santos Barros; David Schlesinger; Debora Botequio Moretti; Dimas Tadeu Covas; Elaine Cristina Marqueze; Elaine Vieira Santos; Evandra Strazza Rodrigues; Heidge Fukumasu; Jayme Augusto de Souza-Neto; José Salvatore Leister Patané; Luiz Alcantara; Luiz Lehmann Coutinho; Maria Carolina Elias; Maurício Lacerda Nogueira; Rafael dos Santos Bezerra; Raul Machado Neto; Rejane Maria Tommasini Grotto; Ricardo Haddad; Sandra Coccuzzo Sampaio Vessoni; Simone Kashima; Svetoslav Nanev Slavov; Vincent Louis Viala |
| EPI_ISL_2494167, EPI_ISL_2494190 | SECRETARIA DE SAUDE DE SAO PEDRO | Instituto Butantan | Antonio Jorge Martins; Claudia Renata dos Santos Barros; David Schlesinger; Debora Botequio Moretti; Dimas Tadeu Covas; Elaine Cristina Marqueze; Elaine Vieira Santos; Evandra Strazza Rodrigues; Heidge Fukumasu; Jayme Augusto de Souza-Neto; José Salvatore Leister Patané; Luiz Alcantara; Luiz Lehmann Coutinho; Maria Carolina Elias; Maurício Lacerda Nogueira; Rafael dos Santos Bezerra; Raul Machado Neto; Rejane Maria Tommasini Grotto; Ricardo Haddad; Sandra Coccuzzo Sampaio Vessoni; Simone Kashima; Svetoslav Nanev Slavov; Vincent Louis Viala |
| EPI_ISL_2493903 | SECRETARIA MUNICIPAL DA SAUDE DE ITARARE | Instituto Butantan | Antonio Jorge Martins; Claudia Renata dos Santos Barros; David Schlesinger; Debora Botequio Moretti; Dimas Tadeu Covas; Elaine Cristina Marqueze; Elaine Vieira Santos; Evandra Strazza Rodrigues; Heidge Fukumasu; Jayme Augusto de Souza-Neto; José Salvatore Leister Patané; Luiz Alcantara; Luiz Lehmann Coutinho; Maria Carolina Elias; Maurício Lacerda Nogueira; Rafael dos Santos Bezerra; Raul Machado Neto; Rejane Maria Tommasini Grotto; Ricardo Haddad; Sandra Coccuzzo Sampaio Vessoni; Simone Kashima; Svetoslav Nanev Slavov; Vincent Louis Viala |
| EPI_ISL_2493275, EPI_ISL_2493276, EPI_ISL_2494110 | SECRETARIA MUNICIPAL DE SAUDE | Instituto Butantan | Antonio Jorge Martins; Claudia Renata dos Santos Barros; David Schlesinger; Debora Botequio Moretti; Dimas Tadeu Covas; Elaine Cristina Marqueze; Elaine Vieira Santos; Evandra Strazza Rodrigues; Heidge Fukumasu; Jayme Augusto de Souza-Neto; José Salvatore Leister Patané; Luiz Alcantara; Luiz Lehmann Coutinho; Maria Carolina Elias; Maurício Lacerda Nogueira; Rafael dos Santos Bezerra; Raul Machado Neto; Rejane Maria Tommasini Grotto; Ricardo Haddad; Sandra Coccuzzo Sampaio Vessoni; Simone Kashima; Svetoslav Nanev Slavov; Vincent Louis Viala |
| EPI_ISL_2494185 | SECRETARIA MUNICIPAL DE SAUDE DE CORDEIROPOLIS | Instituto Butantan | Antonio Jorge Martins; Claudia Renata dos Santos Barros; David Schlesinger; Debora Botequio Moretti; Dimas Tadeu Covas; Elaine Cristina Marqueze; Elaine Vieira Santos; Evandra Strazza Rodrigues; Heidge Fukumasu; Jayme Augusto de Souza-Neto; José Salvatore Leister Patané; Luiz Alcantara; Luiz Lehmann Coutinho; Maria Carolina Elias; Maurício Lacerda Nogueira; Rafael dos Santos Bezerra; Raul Machado Neto; Rejane Maria Tommasini Grotto; Ricardo Haddad; Sandra Coccuzzo Sampaio Vessoni; Simone Kashima; Svetoslav Nanev Slavov; Vincent Louis Viala |
| EPI_ISL_2493825 | SECRETARIA MUNICIPAL DE SAUDE DE MORUNGABA | Instituto Butantan | Antonio Jorge Martins; Claudia Renata dos Santos Barros; David Schlesinger; Debora Botequio Moretti; Dimas Tadeu Covas; Elaine Cristina Marqueze; Elaine Vieira Santos; Evandra Strazza Rodrigues; Heidge Fukumasu; Jayme Augusto de Souza-Neto; José Salvatore Leister Patané; Luiz Alcantara; Luiz Lehmann Coutinho; Maria Carolina Elias; Maurício Lacerda Nogueira; Rafael dos Santos Bezerra; Raul Machado Neto; Rejane Maria Tommasini Grotto; Ricardo Haddad; Sandra Coccuzzo Sampaio Vessoni; Simone Kashima; Svetoslav Nanev Slavov; Vincent Louis Viala |
| EPI_ISL_2493802, EPI_ISL_2494306, EPI_ISL_2494307, EPI_ISL_2494308 | SECRETARIA MUNICIPAL DE SAUDE DE PEDERNEIRAS | Instituto Butantan | Antonio Jorge Martins; Claudia Renata dos Santos Barros; David Schlesinger; Debora Botequio Moretti; Dimas Tadeu Covas; Elaine Cristina Marqueze; Elaine Vieira Santos; Evandra Strazza Rodrigues; Heidge Fukumasu; Jayme Augusto de Souza-Neto; José Salvatore Leister Patané; Luiz Alcantara; Luiz Lehmann Coutinho; Maria Carolina Elias; Maurício Lacerda Nogueira; Rafael dos Santos Bezerra; Raul Machado Neto; Rejane Maria Tommasini Grotto; Ricardo Haddad; Sandra Coccuzzo Sampaio Vessoni; Simone Kashima; Svetoslav Nanev Slavov; Vincent Louis Viala |
| EPI_ISL_2462329, EPI_ISL_2494146 | SECRETARIA MUNICIPAL DE SAUDE DE PRES BERNARDES SP | Instituto Butantan | Antonio Jorge Martins; Claudia Renata dos Santos Barros; David Schlesinger; Debora Botequio Moretti; Dimas Tadeu Covas; Elaine Cristina Marqueze; Elaine Vieira Santos; Evandra Strazza Rodrigues; Heidge Fukumasu; Jayme Augusto de Souza-Neto; José Salvatore Leister Patané; Luiz Alcantara; Luiz Lehmann Coutinho; Maria Carolina Elias; Maurício Lacerda Nogueira; Rafael dos Santos Bezerra; Raul Machado Neto; Rejane Maria Tommasini Grotto; Ricardo Haddad; Sandra Coccuzzo Sampaio Vessoni; Simone Kashima; Svetoslav Nanev Slavov; Vincent Louis Viala |
| EPI_ISL_2801350, EPI_ISL_2801359, EPI_ISL_2801366, EPI_ISL_3102500, EPI_ISL_3536383, EPI_ISL_3536384, EPI_ISL_3536386, EPI_ISL_3536387, EPI_ISL_3536388, EPI_ISL_3536389, EPI_ISL_3536390, EPI_ISL_3536391, EPI_ISL_3912241, EPI_ISL_3912244, EPI_ISL_3912245, EPI_ISL_3912387 | | | |
| see above | SECRETARIA MUNICIPAL DE SAUDE DE TIANGUA | Analytical Competence Molecular Epidemiology Lab/ACME, Oswaldo Cruz Foundation, Ceara (FIOCRUZ CE) | Cleber Furtado Aksenen; Cleber Furtado Aksenen e Suzana Porto Almeida; Fabio Miyajima; Fernando Braga Stehling; Francisco Eder de Moura Lopes; Jamille Maria Mendes Bezerra; Joaquim Cesar do Nascimento Sousa Junior; Joaquim César do Nascimento Sousa Junior; Pedro Miguel Carneiro Jeronimo; Suzana Porto Almeida & Lucas Delerino on behalf of COVID-19 FIOCRUZ Genomic Network; Suzana Porto Almeida e Lucas Delerino; Thais Ferreira de Oliveira; Thais de Oliveira Costa; Ticiane Cavalcante de Souza; Veridiana Pessoa Miyajima |
| EPI_ISL_2895352, EPI_ISL_2895353, EPI_ISL_2895354 | SEEBMO | Instituto Nacional de Saude (INSA) | Borges et al |
| EPI_ISL_2795758 | SELAS MEDILYS | Department of Virology, Henri Mondor University Hospital, Assistance Publique Hôpitaux de Paris, Université Paris-Est Créteil, INSERM U955 | Alexandre Soulier; Christophe Rodriguez; Elisabeth Trawinski; Guillaume Gricourt; Jean-Michel Pawlotsky; Melissa N'Debi; Slim Fourati; Vanessa Demontant |
| EPI_ISL_2493832, EPI_ISL_2493833, EPI_ISL_2493837, EPI_ISL_2493838, EPI_ISL_2493839, EPI_ISL_2493840, EPI_ISL_2493842, EPI_ISL_2493890, EPI_ISL_2493892, EPI_ISL_2493894, EPI_ISL_2494294, EPI_ISL_2494322 | | | |
| see above | SERV DE VIG SANITARIA EPIDEMIO E CTRL DE ZOONOZES GUARUJA | Instituto Butantan | Antonio Jorge Martins; Claudia Renata dos Santos Barros; David Schlesinger; Debora Botequio Moretti; Dimas Tadeu Covas; Elaine Cristina Marqueze; Elaine Vieira Santos; Evandra Strazza Rodrigues; Heidge Fukumasu; Jayme Augusto de Souza-Neto; José Salvatore Leister Patané; Luiz Alcantara; Luiz Lehmann Coutinho; Maria Carolina Elias; Maurício Lacerda Nogueira; Rafael dos Santos Bezerra; Raul Machado Neto; Rejane Maria Tommasini Grotto; Ricardo Haddad; Sandra Coccuzzo Sampaio Vessoni; Simone Kashima; Svetoslav Nanev Slavov; Vincent Louis Viala |
| EPI_ISL_2895384, EPI_ISL_2895389 | SESARAM | Instituto Nacional de Saude (INSA) | Borges et al |
| EPI_ISL_2462317, EPI_ISL_2493872, EPI_ISL_2493874, EPI_ISL_2493942 | SMS ARUJA | Instituto Butantan | Antonio Jorge Martins; Claudia Renata dos Santos Barros; David Schlesinger; Debora Botequio Moretti; Dimas Tadeu Covas; Elaine Cristina Marqueze; Elaine Vieira Santos; Evandra Strazza Rodrigues; Heidge Fukumasu; Jayme Augusto de Souza-Neto; José Salvatore Leister Patané; Luiz Alcantara; Luiz Lehmann Coutinho; Maria Carolina Elias; Maurício Lacerda Nogueira; Rafael dos Santos Bezerra; Raul Machado Neto; Rejane Maria Tommasini Grotto; Ricardo Haddad; Sandra Coccuzzo Sampaio Vessoni; Simone Kashima; Svetoslav Nanev Slavov; Vincent Louis Viala |
| EPI_ISL_3536156, EPI_ISL_3536157, EPI_ISL_3536158, EPI_ISL_3536159, EPI_ISL_3536161 | SOCIEDADE HOSPITALAR PE DIONISIO | Analytical Competence Molecular Epidemiology Lab/ACME, Oswaldo Cruz Foundation, Ceara (FIOCRUZ CE) | Cleber Furtado Aksenen; Fabio Miyajima; Fernando Braga Stehling; Francisco Eder de Moura Lopes; Jamille Maria Mendes Bezerra; Joaquim César do Nascimento Sousa Junior; Pedro Miguel Carneiro Jeronimo; Suzana Porto Almeida e Lucas Delerino; Thais Ferreira de Oliveira; Thais de Oliveira Costa; Ticiane Cavalcante de Souza; Veridiana Pessoa Miyajima |
| EPI_ISL_2627152, EPI_ISL_2868528 | STONY BROOK UNIVERSITY HOSPITAL | Wadsworth Center, New York State Department of Health | Alexis Russell; Catharine Prussing; Daryl M. Lamson; Erasmus Schneider; Erica Lasek-Nesselquist; John Kelly; Jonathan Plitnick; Kirsten St. George; Matthew Shudt; Melissa A Leisner; Navjot Singh |

| | | | |
|---|---|---|---|
| EPI_ISL_2868260, EPI_ISL_2868261 | SUNY UPSTATE MEDICAL UNIVERSITY | Wadsworth Center, New York State Department of Health | Alexis Russell; Catharine Prussing; Daryl M. Lamson; Erasmus Schneider; Erica Lasek-Nesselquist; John Kelly; Jonathan Plitnick; Kirsten St. George; Matthew Shudt; Melissa A Leisner; Navjot Singh |
| EPI_ISL_2933358 | SYNLAB GASCOGNE | CNR Virus des Infections Respiratoires - France SUD | Antonin Bal; Bruno Lina; Gregory Destras; Gwendolyne Burfin; Hadrien Regue; Laurence Josset; Martine Valette; Quentin Semanas |
| EPI_ISL_3044006 | SYNLAB Labor München Zentrum LMZ | Robert Koch Institute | |
| EPI_ISL_2472365 | SYNLAB MVZ Berlin | Robert Koch Institute | |
| EPI_ISL_2389554, EPI_ISL_2389772, EPI_ISL_2633059, EPI_ISL_2635424, EPI_ISL_2844917 | SYNLAB MVZ Heidelberg | Robert Koch Institute | |
| EPI_ISL_2635200, EPI_ISL_2759897 | SYNLAB MVZ Leverkusen | Robert Koch Institute | |
| EPI_ISL_2469537, EPI_ISL_2633330 | SYNLAB MVZ Trier | Robert Koch Institute | |
| EPI_ISL_2469483, EPI_ISL_2472105 | SYNLAB MVZ Weiden | Robert Koch Institute | |
| EPI_ISL_2603678, EPI_ISL_2603701, EPI_ISL_2603702, EPI_ISL_2603710, EPI_ISL_2603733, EPI_ISL_2603770, EPI_ISL_2603771, EPI_ISL_2617007, EPI_ISL_2617014, EPI_ISL_2692580, EPI_ISL_2692655, EPI_ISL_2692656, EPI_ISL_2692762, EPI_ISL_2692792, EPI_ISL_2692797, EPI_ISL_2810033, EPI_ISL_2810080, EPI_ISL_2810086, EPI_ISL_2810096, EPI_ISL_2810103, EPI_ISL_2810169, EPI_ISL_2810256, EPI_ISL_2810270, EPI_ISL_2894265, EPI_ISL_2894378, EPI_ISL_2894410, EPI_ISL_2978363, EPI_ISL_2978442, EPI_ISL_2978444, EPI_ISL_2978449, EPI_ISL_2978489, EPI_ISL_2978499, EPI_ISL_2978517, EPI_ISL_3067926, EPI_ISL_3067935, EPI_ISL_3067936, EPI_ISL_3067937, EPI_ISL_3721739 | | | |
| see above | Salud Digna | Instituto Nacional de Medicina Genomica | Abraham Campos-Romero; Cedro-Tanda A; Escobar-Arrazola; Escobar-Arrazola MA; Gonzalez-Barrera D; Herrera-Montalvo LA.; Hidalgo-Miranda A; Luna-Ruiz Marco; M; Mendoza-Vargas A; Moreno-Camacho José Luis; Munguia-Garza P; Ramirez-Vega O; Rangel-DeLeon D; Reyes-Grajeda JP; Rodriguez-Gallegos Jorge; Yair Alfaro-Mora |
| EPI_ISL_3373977, EPI_ISL_3373979, EPI_ISL_3373982, EPI_ISL_3373994, EPI_ISL_3374014, EPI_ISL_3374016, EPI_ISL_3374019, EPI_ISL_3374020, EPI_ISL_3374023, EPI_ISL_3374024, EPI_ISL_3374025, EPI_ISL_3374026, EPI_ISL_3374030, EPI_ISL_3374031, EPI_ISL_3374036, EPI_ISL_3374038, EPI_ISL_3374039, EPI_ISL_3912490, EPI_ISL_3912512 | | | |
| see above | Salud Digna, A.C | Andersen lab at Scripps Research | Abraham Garcia Gil; Jose Luis Moreno Camacho; Marco Antonio Luna Ruiz-Esparza; Miguel A. Fernandez Rojas; SEARCH Alliance with Abraham Campos Romero |
| EPI_ISL_3374049, EPI_ISL_3374052, EPI_ISL_3374055, EPI_ISL_3374057, EPI_ISL_3374060, EPI_ISL_3374068, EPI_ISL_3374069, EPI_ISL_3374071, EPI_ISL_3374113, EPI_ISL_3374114, EPI_ISL_3374121 | | | |
| see above | San Diego County Public Health Laboratory | Andersen lab at Scripps Research | Brett Austin; Jovan Shephard; SEARCH Alliance San Diego with Ashleigh Murphy |
| EPI_ISL_2840862, EPI_ISL_2840896, EPI_ISL_2841534, EPI_ISL_2841536, EPI_ISL_2841614, EPI_ISL_2842703, EPI_ISL_2842704, EPI_ISL_2842705, EPI_ISL_2873837, EPI_ISL_3275048, EPI_ISL_3346596, EPI_ISL_3346597 | | | |
| see above | San Diego County Public Health Laboratory | San Diego County Public Health Laboratory | Ashleigh Murphy Schafer; Brett Austin; San Diego County Public Health Laboratory |
| EPI_ISL_2650574, EPI_ISL_2691314, EPI_ISL_2934069, EPI_ISL_2967940 | Santa Clara Valley Medical Center | Santa Clara County Public Health Laboratory | Santa Clara County Public Health Department |
| EPI_ISL_2627004 | Saratoga Hospital Laboratory | Wadsworth Center, New York State Department of Health | Alexis Russell; Catharine Prussing; Daryl M. Lamson; Erasmus Schneider; Erica Lasek-Nesselquist; John Kelly; Jonathan Plitnick; Kirsten St. George; Matthew Shudt; Melissa A Leisner; Navjot Singh |
| EPI_ISL_2735928, EPI_ISL_2735931 | Seattle Flu Study | Seattle Flu Study | Amanda Adler; Barry R. Lutz; Benjamin Pelle; Caitlin R. Wolf; Chris D. Frazar; Deborah A. Nickerson; Elisabeth Brandstetter; Erica Ryke; Helen Y. Chu; Janet A. Englund; Jay Shendure; Jeff Duchin; Jover Lee; Kairsten Fay; Karen Cowgill; Kirsten Lacombe; Lea M. Starita; Mark J. Rieder; Matthew Richardson; Matthew Thompson; Melissa Truong; Michael Boeckh; Michael Famulare; Misja Ilcisin; Peter D. Han; Stephanie Schrag; Thomas R. Sibley; Trevor Bedford |
| EPI_ISL_2680930 | Servicio Diagnóstico Clínico | Laboratorio nacional-LANIIA-CIAD | ; Alejandra García-Gasca; Alejandra Hernández-Terán; Alejandro Sánchez-Flores; Alfredo Herrera-Estrella; Alicia Ocaña-Mondragón; Andreu Comas-García; Angel Gustavo Salas-Lais; Antonio Loza Román; Bernardo Martínez-Miguel; Blanca Taboada; Brenda Irasema Maldonado-Meza; Bruno Gómez-Gil; Carla Ivón Herrera-Najera; Carlos F. Arias; Celia Boukadida; Clara Esperanza Santacruz-Tinoco; Concepción Grajales-Muñiz; Consorcio Mexicano de Vigilancia Genómica (CoViGen-Mex). Authors (in alphabetical order): Julio Elias Alvarado-Yaah; Cristóbal Cháidez-Quiróz; Célida Duque Molina; Célida Martínez- Rodríguez; Daniel Fregoso-Rueda; Daniel Lira Morales; Eduardo Becerril-Vargas; Fernando Fontove-Herrera; Fidencio Mejía-Nepomuceno; Francisco Pulido; Gloria Elena Espinosa-Ayala; Gloria María Molina-Salinas; Gloria Vazquez; Hector Esteban Paz-Juárez; Hector Montoya-Fuentes; Helen Haydee Fernanda Ramírez-Plascencia; Irvin González-López; Jean Pierre González; Jesús Hernández; Joel Armando Vázquez-Pérez.; Jorge Salas-Hernández; José Antonio Enciso-Moreno; José Arturo Martínez-Orozco; José Esteban Muñoz-Medina; José de Jesús Nuñez-Contreras; Juan Bautista Chale-Dzul; Julissa Enciso-Ibarra; Luis Alberto Ochoa-Carrera; Margarita Matías-Florentino; Mario Mújica-Sánchez; Marissa Perez-Garcia; María Guadalupe Santiago-Mauricio; María Guadalupe de Jesús Mireles-Rivera; Nelly Sélem-Mojica; Pavel Isa; Ricardo Ciria Merce; Ricardo Grande; Rosa María Gutiérrez Rios; Santiago Ávila-Ríos; Selene Zárate; Susana Lopez; Verónica Mata-Haro; Victor Eduardo García-Arias; Víctor Hugo Borja-Aburto |
| EPI_ISL_4082378, EPI_ISL_4082415, EPI_ISL_4082433, EPI_ISL_4082434, EPI_ISL_4082437, EPI_ISL_4082445, EPI_ISL_4082479, EPI_ISL_4082507, EPI_ISL_4082509, EPI_ISL_4082510, EPI_ISL_4082521, EPI_ISL_4082522, EPI_ISL_4082524, EPI_ISL_4082530, EPI_ISL_4082536, EPI_ISL_4082544, EPI_ISL_4082545, EPI_ISL_4082572, EPI_ISL_4082576, EPI_ISL_4082594, EPI_ISL_4082602, EPI_ISL_4082609, EPI_ISL_4082611, EPI_ISL_4082629, EPI_ISL_4082634, EPI_ISL_4082638, EPI_ISL_4082652, EPI_ISL_4082664, EPI_ISL_4082665, EPI_ISL_4082713, EPI_ISL_4082715, EPI_ISL_4082717, EPI_ISL_4082718, EPI_ISL_4082722, EPI_ISL_4082728, EPI_ISL_4082747, EPI_ISL_4082777, EPI_ISL_4082778, EPI_ISL_4082805, EPI_ISL_4082810, EPI_ISL_4082833, EPI_ISL_4082834, EPI_ISL_4082837, EPI_ISL_4082841, EPI_ISL_4082843, EPI_ISL_4082845, EPI_ISL_4082847, EPI_ISL_4082848, EPI_ISL_4082849, EPI_ISL_4082852, EPI_ISL_4082878, EPI_ISL_4082887, EPI_ISL_4082893, EPI_ISL_4082909, EPI_ISL_4082915, EPI_ISL_4082922, EPI_ISL_4082932, EPI_ISL_4082941, EPI_ISL_4082964, EPI_ISL_4082991, EPI_ISL_4082997, EPI_ISL_4082998, EPI_ISL_4083001, EPI_ISL_4083004, EPI_ISL_4083008, EPI_ISL_4083009, EPI_ISL_4083010, EPI_ISL_4083018, EPI_ISL_4083019, EPI_ISL_4083021, EPI_ISL_4083022, EPI_ISL_4083023, EPI_ISL_4083024, EPI_ISL_4083026, EPI_ISL_4083028, EPI_ISL_4083030, EPI_ISL_4083031, EPI_ISL_4083033, EPI_ISL_4083034, EPI_ISL_4083035, EPI_ISL_4083037, EPI_ISL_4083046, EPI_ISL_4083063, EPI_ISL_4083074, EPI_ISL_4083084, EPI_ISL_4083097, EPI_ISL_4083100, EPI_ISL_4083101, EPI_ISL_4083117, EPI_ISL_4083138, EPI_ISL_4083152, EPI_ISL_4083162, EPI_ISL_4083170, EPI_ISL_4083171, EPI_ISL_4083178, EPI_ISL_4083224, EPI_ISL_4083226, EPI_ISL_4083237, EPI_ISL_4083244, EPI_ISL_4083248, EPI_ISL_4083249, EPI_ISL_4083250, EPI_ISL_4083255, EPI_ISL_4083257, EPI_ISL_4083264, EPI_ISL_4083266, EPI_ISL_4083268, EPI_ISL_4083269, EPI_ISL_4083285, EPI_ISL_4083296, EPI_ISL_4083298, EPI_ISL_4083317, EPI_ISL_4083335, EPI_ISL_4083358, EPI_ISL_4083360, EPI_ISL_4083396, EPI_ISL_4083426, EPI_ISL_4083429, EPI_ISL_4083432, EPI_ISL_4083434 | | | |
| see above | Servicio Virosis Respiratorias-Departamento Virología-INEI | Instituto Nacional Enfermedades Infecciosas C.G.Malbran | Avaro M.; Baumeister E.; Benedetti E.; Campos J.; Cisterna D.; Dattero ME; De Belder D.; Haim MS.; Lorenzo F.; Molina V.; Perandones C.; Poklepovich T.; Pontoriero A.; Russo M.; Sanchez Loria J.; Tuduri E. |
| EPI_ISL_3149734, EPI_ISL_3388829 | Servicio de Inmunología del Hospital Julio Perrando | Laboratorio de Biología Molecular, Instituto de Medicina Regional on behalf of 'Proyecto Argentino Interinstitucional de genomica de SARS-CoV-2' (PAIS Consortium) | Bettina Brusés; Florencia Vallejos Schulze; Griselda Oria; Horacio Lucero.; Javier Mussin; Laura Formichelli; María Delia Foussal; Melina Lorenzini Campos; Raúl Maximiliano Acevedo |
| EPI_ISL_2558044, EPI_ISL_2758565, EPI_ISL_2758594, EPI_ISL_3219110, EPI_ISL_3219120 | Servicio de Microbiologia Hospital Ramon y Cajal | Servicio de Microbiologia Hospital Ramon y Cajal | Galan JC; Martinez L. Abreu M; Ponce M; y Gonzalez-Alba JM |
| EPI_ISL_2516600, EPI_ISL_2516673, EPI_ISL_2516674, EPI_ISL_2516693, EPI_ISL_2516694, EPI_ISL_2516695, EPI_ISL_2516705, EPI_ISL_2516706, EPI_ISL_2516707, EPI_ISL_2516708, EPI_ISL_2516716, EPI_ISL_2516725, EPI_ISL_2516752, EPI_ISL_2516755, EPI_ISL_2516859, EPI_ISL_2934624 | | | |
| see above | Servicio de Microbiología Clínica (Complejo Hospitalario de Navarra, Pamplona) | Centro de Secuenciación NASERTIC | Ana Miqueleiz; Ana Navascués; Carmen Ezpeleta Baquedano |
| EPI_ISL_2658202, EPI_ISL_2658207, EPI_ISL_2658212, EPI_ISL_2658215, EPI_ISL_2658235, EPI_ISL_2658236, EPI_ISL_2658243, EPI_ISL_3060990 | | | |
| see above | Servicio de Microbiología. Consorcio Hospital General Universitario de Valencia | SeqCOVID-SPAIN consortium/IBV(CSIC) | Begoña Fuster Escrivá; Carme Salvador García; Concepción Gimeno Cardona and SeqCOVID-SPAIN consortium; María Dolores Ocete; Rafael Medina González |
| EPI_ISL_2600911, EPI_ISL_2600917, EPI_ISL_2931406 | Servicio de Microbiología. Hospital Clínico Universitario de Valencia | SeqCOVID-SPAIN consortium/IBV(CSIC) | David Navarro Ortega; Eliseo Albert Vicent; Ignacio Torres and SeqCOVID-SPAIN consortium |
| EPI_ISL_3008587 | Servicio de Microbiología. Hospital General Universitario de Castellón | SeqCOVID-SPAIN consortium/IBV(CSIC) | María Dolores Tirado Balaguer and SeqCOVID-SPAIN consortium; Rosario Moreno Muñoz |
| EPI_ISL_3063787, EPI_ISL_3907299 | Servicio de Microbiología. Hospital Universitario Doctor Peset | SeqCOVID-SPAIN consortium/IBV(CSIC) | José Miguel Nogueira Coito and SeqCOVID-SPAIN consortium; Juan Alberola Enguídanos; Juan José Camarena Miñana; Rosa González Pellicer |
| EPI_ISL_2790433 | Servizo de Microbioloxía. Complexo Hospitalario de Santiago de Compostela | Servizo de Microbioloxía. Complexo Hospitalario de Santiago de Compostela | Amparo Coira; Antonio Aguilera; Carlos García_Riestra; Daniel Navarro; Gema Barbeito; Iria Roca; José Llovo; Laura Millán; Manuela Hernández; María Luisa Pérez_del_Molino; Mercedes Treviño; Rocío trastoy; Teresa Lopez_Valiño; Xosé Costa |
| EPI_ISL_3024497 | Shamir Medical Center (Asaf Harofe) | Shamir Medical Center (Asaf Harofe) | Abu Hamad Ramzia; Adina Bar Chaim; Anna Vishnevsky; Chen Weiner; Nir Rainy; Patricia Benveniste-Lekovitz; Reut Sorek Abramovich; Yevgeni Yegorov |
| EPI_ISL_2712542, EPI_ISL_2920435, EPI_ISL_3373825 | Sharp HealthCare Laboratory | Andersen lab at Scripps Research | Art Mendoza; Cathy Woerle; Jacquelyn Berumen; Liam McGinnis; Omid Bakhtar; SEARCH Alliance San Diego with Aaron Harding |
| EPI_ISL_3254818, EPI_ISL_3254819, EPI_ISL_3552166 | Simple Laboratories | Gagnon Lab, Southern Illinois University | Keith Gagnon |
| EPI_ISL_2484889 | Southern Nevada Public Health Laboratory | Southern Nevada Public Health Laboratory | Michael Picker |
| EPI_ISL_2918821 | Spital Limmattal | Institute of Medical Virology | Alexandra Trkola; Annette Audigé; Catharine Aquino; Cyril Shah; Daniel Ehrsam; Gabriela Ziltener; Guido Bloemberg; Hubert Rehrauer; Isabel Stürmer; Joel Wirz; Jon Huder; Jürg Böni; Kevin Steiner; Maria Grünberg; Maryam Zaheri; Michael Huber; Riccarda Capaul; Stefan Schmutz; Verena Kufner; Weihong Qi |
| EPI_ISL_3050186, EPI_ISL_3050210, | Stanford Health Care | Stanford University School of Medicine, Clinical Virology | Becky Jiang; Bernadette Troung; Daniel Solis; Hannah Wang; James Zehnder; Malaya K. Sahoo; Mamdouh Sibai; Nathan Hammond; Selamawit Bihon; and Benjamin A. Pinsky |

| | | | |
|---|---|---|---|
| EPI_ISL_4072051, EPI_ISL_4072052 | | Laboratory | |
| EPI_ISL_3419023 | StarMed Healthcare | UNC Charlotte Environmental Monitoring Laboratory | Adam France; Cynthia Gibas; Erin Stiers; Jannatul Ferdous; Jessica Schlueter; Kevin Lambirth; Sam Kunkleman |
| EPI_ISL_2724749, EPI_ISL_2724788, EPI_ISL_2724799, EPI_ISL_2724825, EPI_ISL_2724828, EPI_ISL_2724830, EPI_ISL_2724833, EPI_ISL_2724834, EPI_ISL_2724842, EPI_ISL_2724844, EPI_ISL_2724849, EPI_ISL_2724873, EPI_ISL_2724880, EPI_ISL_2724900, EPI_ISL_2724909, EPI_ISL_3033098, EPI_ISL_3033104, EPI_ISL_3033105, EPI_ISL_3033106, EPI_ISL_3033120, EPI_ISL_3033163, EPI_ISL_3033187, EPI_ISL_3033188, EPI_ISL_3033266, EPI_ISL_3307957 | | | |
| see above | State Laboratories Division, Hawaii State Department of Health | State Laboratories Division, Hawaii State Department of Health | Ayana Garnet; Daniel Strange; Drew Kuwazaki; Edward Desmond; Pamela O'Brien; Razvan Sultana |
| EPI_ISL_2800567, EPI_ISL_2800569, EPI_ISL_2800572, EPI_ISL_2800574, EPI_ISL_2800575, EPI_ISL_2800576, EPI_ISL_2800580, EPI_ISL_2800590, EPI_ISL_2800591, EPI_ISL_2800592, EPI_ISL_3049994, EPI_ISL_3050004, EPI_ISL_3050008, EPI_ISL_3050050, EPI_ISL_3050131 | | | |
| see above | State Testing Facility | Altius Institute for Biomedical Research | Alex Isner; Alex Nguyen; Amanda Gale; Audra Johnson; Clem Green; Daniel Bates; Eric Thorland; Jacob Rodriguez; Jean Robinston; Jemma Nelson; Jessica Kunder; John Stamatoyannopoulos; Joshua Richards; Julia Wald; Kneshay Harper; Lauren Mitchell; Mark Frerker; Matt Hartman; Michael Buckley; Muhammad Halimun; Rebecca Bruders; Sadie Patraw; Sofia Olsson; Tobias Ragoczy |
| EPI_ISL_2581162, EPI_ISL_2581166, EPI_ISL_2581180, EPI_ISL_2618952, EPI_ISL_2618969, EPI_ISL_2619010, EPI_ISL_2619021, EPI_ISL_2619038, EPI_ISL_2792714, EPI_ISL_2792769, EPI_ISL_2792804, EPI_ISL_2792874, EPI_ISL_2792929, EPI_ISL_2792931, EPI_ISL_2792935, EPI_ISL_2793103, EPI_ISL_2793165, EPI_ISL_2888088, EPI_ISL_2888091, EPI_ISL_2888115, EPI_ISL_2980940, EPI_ISL_2980941, EPI_ISL_3157564, EPI_ISL_3795575, EPI_ISL_3998556, EPI_ISL_3998558, EPI_ISL_3998560, EPI_ISL_3998561, EPI_ISL_3998562, EPI_ISL_3998563, EPI_ISL_3998569, EPI_ISL_3998570, EPI_ISL_3998572, EPI_ISL_3998578, EPI_ISL_3998581, EPI_ISL_3998584, EPI_ISL_3998869, EPI_ISL_3998892 | | | |
| see above | Swedish national genomic surveillance program of SARS-CoV-2 | The Public Health Agency of Sweden | Alma Brolund; Maria Lind Karlberg; Maximilian Riess; Swedish national genomic surveillance program of SARS-CoV-2 |
| EPI_ISL_2769146 | Swiss Analysis AG | Institute of Medical Virology | Alexandra Trkola; Annette Audigé; Cyril Shah; Gabriela Ziltener; Guido Bloemberg; Jon Huder; Jürg Böni; Kevin Steiner; Maria Grünberg; Maryam Zaheri; Michael Huber; Riccarda Capaul; Stefan Schmutz; Verena Kufner |
| EPI_ISL_3030108 | Synlab Suisse SA | Clinical Bacteriology | Adrian Egli; Alfredo Mari; Fanny Wegner; Hans Hirsch; Helena MB Seth-Smith; Julia Bielicki; Karoline Leuzinger; Manuel Battegay; Tim Roloff |
| EPI_ISL_2626957, EPI_ISL_2626963, EPI_ISL_2868602, EPI_ISL_2868604, EPI_ISL_2868605, EPI_ISL_2868607 | TEMPUS LABS INC | Wadsworth Center, New York State Department of Health | Alexis Russell; Catharine Prussing; Daryl M. Lamson; Erasmus Schneider; Erica Lasek-Nesselquist; John Kelly; Jonathan Plitnick; Kirsten St. George; Matthew Shudt; Melissa A Leisner; Navjot Singh |
| EPI_ISL_2769191, EPI_ISL_2769204, EPI_ISL_2769277, EPI_ISL_3088803, EPI_ISL_3325976, EPI_ISL_3325992, EPI_ISL_3326351, EPI_ISL_3326437, EPI_ISL_3326706, EPI_ISL_3460398, EPI_ISL_3460423, EPI_ISL_3460429, EPI_ISL_3460463, EPI_ISL_3460472, EPI_ISL_3460490, EPI_ISL_3460511, EPI_ISL_3460515, EPI_ISL_3460516, EPI_ISL_3460532, EPI_ISL_3460614, EPI_ISL_3661970, EPI_ISL_3826274 | | | |
| see above | TGen North | TGen North | "Jolene Bowers; Brett Van Tassel; Chris French; Darrin Lemmer; Dave Engelthaler; Dave Engelthaler"; Hayley Yaglom; Heather Centner; Jolene Bowers |
| EPI_ISL_3242037 | TX-Houston Health Department Laboratory | Centers for Disease Control and Prevention Division of Viral Diseases, Pathogen Discovery | Alex Burgin; Ben L. Rambo-Martin; Clinton R. Paden; Dakota Howard; Dave Wentworth; Dhwani Batra; Jasmine Padilla; Justin Lee; Krista Queen; Kristen Knipe; Kristine Lacek; Mark Burroughs; Matthew Schmerer; Meghan Bentz; Mili Sheth; Peter Cook; Sam Shepard; Sarah Nobles; Suxiang Tong; Vivien Dugan; Yvette Unoarumhi |
| EPI_ISL_3267887 | TXDSHS | TXDSHS | Anita Pokharel; Bonnie Oh; Chun Wang; Grace Kubin; Jenny Zhang; Karen Bobier; Lorraine Rodriguez; Maliha Rahman; Mayela Pedrueza; Myong Koag; Rachel Lee; Rashmi Tuladhar |
| EPI_ISL_2790535, EPI_ISL_2800904, EPI_ISL_2800905, EPI_ISL_2833809, EPI_ISL_2966844, EPI_ISL_2966850 | Tampa General Hospital Esoteric Lab | Tampa General Hospital Esoteric Research & Development Lab | Amorce Lima; Deanna Becker; Dominic Uy; Elaine Vendrone; Jon Faughn; Jorge Mecias-Frias; Suzane Silbert; Vicki Healer |
| EPI_ISL_3925995, EPI_ISL_3926065, EPI_ISL_3930809 | The National University Hospital of Iceland | deCODE genetics | Agnar Helgason; Alma Moller; Arna B Agustsdottir; Arnaldur Gylfason; Asgeir Sigurdsson; Aslaug Jonasdottir; Berglind Eiriksdottir; Bjarni Thorbjornsson; Brynjar O Jensson; Daniel F Gudbjartsson; Droplaug N Magnusdottir; Elisabet E Gardarsdottir; Emil A Thorarensen; Gardar Sveinbjornsson; Gisli Masson; Gudmundur Georgsson; Gudmundur L Norddahl; Gudrun Sigmundsdottir; Hakon Jonsson; Hannes Eggertsson; Hilma Holm; Ingileif Jonsdottir; Jona Saemundsdottir; Kamilla S Josefsdottir; Kari Stefansson; Karl G Kristinsson; Kjartan R Gudmundsson; Kristin E Sveinsdottir; Kristjan E Hjorleifsson; Louise le Roux; Maney Sveinsdottir; Olafia S Gretarsdottir; Olafur T Magnusson; Pall Melsted; Patrick Sulem; Run Fridriksdottir; Solvi Rognvaldsson; Thora R Gunnarsdottir; Thordur Kristjansson; Thorolfur Gudnason; Unnur Thorsteinsdottir |
| EPI_ISL_2762132 | Thüringer Landesamtes für Verbraucherschutz | Robert Koch Institute | |
| EPI_ISL_2774572 | Torrance Memorial Medical Center | Los Angeles County Public Health Laboratories | P. Hemarajata et al. |
| EPI_ISL_3152417 | TriCore Reference Laboratories | Center for Global Health, University of New Mexico Health Sciences Center | Cecilia Thompson; Darrell Dinwiddie; Daryl Domman; Karissa Culbreath; Kendra Pesko; Kurt Schwalm; Valerie Morley |
| EPI_ISL_3049350 | Tulare County Public Health Laboratory | Tulare County Public Health Laboratory | Denise L Lopez; Joshua Kootstra |
| EPI_ISL_2840625 | U.O. Igiene, Ospedale Policlinico San Martino IRCCS Genova | U.O. Igiene, Ospedale Policlinico San Martino | Bruzzone Bianca; Caligiuri Patrizia; De Pace Vanessa; Domnich Alexander; Icardi Giancarlo on behalf of SARS-CoV-2 ITALIAN RESEARCH ENTERPRISE-(SCIRE) Collaborative Group; Orsi Andrea; Ricucci Valentina |
| EPI_ISL_3266323 | U.O. Microbiologia Laboratorio Unico Centro Servizi - AUSL della Romagna | U.O. Microbiologia, Laboratorio Unico Centro Servizi - AUSL della Romagna | Giorgio Dirani |
| EPI_ISL_3912250 | UAPS FERNANDO FACANHA | Analytical Competence Molecular Epidemiology Lab/ACME, Oswaldo Cruz Foundation, Ceara (FIOCRUZ CE) | Cleber Furtado Aksenen; Fabio Miyajima; Fernando Braga Stehling; Francisco Eder de Moura Lopes; Jamille Maria Mendes Bezerra; Joaquim Cesar do Nascimento Sousa Junior; Pedro Miguel Carneiro Jeronimo; Suzana Porto Almeida & Lucas Delerino on behalf of COVID-19 FIOCRUZ Genomic Network; Thais Ferreira de Oliveira; Thais de Oliveira Costa; Ticiane Cavalcante de Souza; Veridiana Pessoa Miyajima |
| EPI_ISL_3912251 | UAPS FLORESTA | Analytical Competence Molecular Epidemiology Lab/ACME, Oswaldo Cruz Foundation, Ceara (FIOCRUZ CE) | Cleber Furtado Aksenen; Fabio Miyajima; Fernando Braga Stehling; Francisco Eder de Moura Lopes; Jamille Maria Mendes Bezerra; Joaquim Cesar do Nascimento Sousa Junior; Pedro Miguel Carneiro Jeronimo; Suzana Porto Almeida & Lucas Delerino on behalf of COVID-19 FIOCRUZ Genomic Network; Thais Ferreira de Oliveira; Thais de Oliveira Costa; Ticiane Cavalcante de Souza; Veridiana Pessoa Miyajima |
| EPI_ISL_3912264 | UBASF AMADOR | Analytical Competence Molecular Epidemiology Lab/ACME, Oswaldo Cruz Foundation, Ceara (FIOCRUZ CE) | Cleber Furtado Aksenen; Fabio Miyajima; Fernando Braga Stehling; Francisco Eder de Moura Lopes; Jamille Maria Mendes Bezerra; Joaquim Cesar do Nascimento Sousa Junior; Pedro Miguel Carneiro Jeronimo; Suzana Porto Almeida & Lucas Delerino on behalf of COVID-19 FIOCRUZ Genomic Network; Thais Ferreira de Oliveira; Thais de Oliveira Costa; Ticiane Cavalcante de Souza; Veridiana Pessoa Miyajima |
| EPI_ISL_3912265 | UBASF DE GUARIBAS | Analytical Competence Molecular Epidemiology Lab/ACME, Oswaldo Cruz Foundation, Ceara (FIOCRUZ CE) | Cleber Furtado Aksenen; Fabio Miyajima; Fernando Braga Stehling; Francisco Eder de Moura Lopes; Jamille Maria Mendes Bezerra; Joaquim Cesar do Nascimento Sousa Junior; Pedro Miguel Carneiro Jeronimo; Suzana Porto Almeida & Lucas Delerino on behalf of COVID-19 FIOCRUZ Genomic Network; Thais Ferreira de Oliveira; Thais de Oliveira Costa; Ticiane Cavalcante de Souza; Veridiana Pessoa Miyajima |
| EPI_ISL_3912268 | UBS ACUDE DO MATO | Analytical Competence Molecular Epidemiology Lab/ACME, Oswaldo Cruz Foundation, Ceara (FIOCRUZ CE) | Cleber Furtado Aksenen; Fabio Miyajima; Fernando Braga Stehling; Francisco Eder de Moura Lopes; Jamille Maria Mendes Bezerra; Joaquim Cesar do Nascimento Sousa Junior; Pedro Miguel Carneiro Jeronimo; Suzana Porto Almeida & Lucas Delerino on behalf of COVID-19 FIOCRUZ Genomic Network; Thais Ferreira de Oliveira; Thais de Oliveira Costa; Ticiane Cavalcante de Souza; Veridiana Pessoa Miyajima |
| EPI_ISL_3912270, EPI_ISL_3912272, EPI_ISL_3912273, EPI_ISL_3912275, EPI_ISL_3912276, EPI_ISL_3912277 | UBS CAIC II | Analytical Competence Molecular Epidemiology Lab/ACME, Oswaldo Cruz Foundation, Ceara (FIOCRUZ CE) | Cleber Furtado Aksenen; Fabio Miyajima; Fernando Braga Stehling; Francisco Eder de Moura Lopes; Jamille Maria Mendes Bezerra; Joaquim Cesar do Nascimento Sousa Junior; Pedro Miguel Carneiro Jeronimo; Suzana Porto Almeida & Lucas Delerino on behalf of COVID-19 FIOCRUZ Genomic Network; Thais Ferreira de Oliveira; Thais de Oliveira Costa; Ticiane Cavalcante de Souza; Veridiana Pessoa Miyajima |
| EPI_ISL_2494144 | UBS CELESTINO CARNIELO | Instituto Butantan | Antonio Jorge Martins; Claudia Renata dos Santos Barros; David Schlesinger; Debora Botequio Moretti; Dimas Tadeu Covas; Elaine Cristina Marqueze; Elaine Vieira Santos; Evandra Strazza Rodrigues; Heidge Fukumasu; Jayme Augusto de Souza-Neto; José Salvatore Leister Patané; Luiz Alcantara; Luiz Lehmann Coutinho; Maria Carolina Elias; Maurício Lacerda Nogueira; Rafael dos Santos Bezerra; Raul Machado Neto; Rejane Maria Tommasini Grotto; Ricardo Haddad; Sandra Coccuzzo Sampaio Vessoni; Simone Kashima; Svetoslav Nanev Slavov; Vincent Louis Viala |
| EPI_ISL_3912278 | UBS CENTRO | Analytical Competence Molecular Epidemiology Lab/ACME, Oswaldo Cruz Foundation, Ceara (FIOCRUZ CE) | Cleber Furtado Aksenen; Fabio Miyajima; Fernando Braga Stehling; Francisco Eder de Moura Lopes; Jamille Maria Mendes Bezerra; Joaquim Cesar do Nascimento Sousa Junior; Pedro Miguel Carneiro Jeronimo; Suzana Porto Almeida & Lucas Delerino on behalf of COVID-19 FIOCRUZ Genomic Network; Thais Ferreira de Oliveira; Thais de Oliveira Costa; Ticiane Cavalcante de Souza; Veridiana Pessoa Miyajima |
| EPI_ISL_2493866, EPI_ISL_2493868, EPI_ISL_2493869 | UBS DE JAMBEIRO JOCELINA MEIRELES | Instituto Butantan | Antonio Jorge Martins; Claudia Renata dos Santos Barros; David Schlesinger; Debora Botequio Moretti; Dimas Tadeu Covas; Elaine Cristina Marqueze; Elaine Vieira Santos; Evandra Strazza Rodrigues; Heidge Fukumasu; Jayme Augusto de Souza-Neto; José Salvatore Leister Patané; Luiz Alcantara; Luiz Lehmann Coutinho; Maria Carolina Elias; Maurício Lacerda Nogueira; Rafael dos Santos Bezerra; Raul Machado Neto; Rejane Maria Tommasini Grotto; Ricardo Haddad; Sandra Coccuzzo Sampaio Vessoni; Simone Kashima; Svetoslav Nanev Slavov; Vincent Louis Viala |
| EPI_ISL_3536374 | UBS DE MUFUMBAL | Analytical Competence Molecular Epidemiology Lab/ACME, Oswaldo Cruz Foundation, Ceara (FIOCRUZ CE) | Cleber Furtado Aksenen; Fabio Miyajima; Fernando Braga Stehling; Francisco Eder de Moura Lopes; Jamille Maria Mendes Bezerra; Joaquim César do Nascimento Sousa Junior; Pedro Miguel Carneiro Jeronimo; Suzana Porto Almeida e Lucas Delerino; Thais Ferreira de Oliveira; Thais de Oliveira Costa; Ticiane Cavalcante de Souza; Veridiana Pessoa Miyajima |
| EPI_ISL_3912280, EPI_ISL_3912281, EPI_ISL_3912282, EPI_ISL_3912283, EPI_ISL_3912284, EPI_ISL_3912285, EPI_ISL_3912287 | | | |
| see above | UBS DO BAIRRO SAO JOSE | Analytical Competence Molecular Epidemiology Lab/ACME, Oswaldo Cruz Foundation, Ceara (FIOCRUZ | Cleber Furtado Aksenen; Fabio Miyajima; Fernando Braga Stehling; Francisco Eder de Moura Lopes; Jamille Maria Mendes Bezerra; Joaquim Cesar do Nascimento Sousa Junior; Pedro Miguel Carneiro Jeronimo; Suzana Porto Almeida & Lucas Delerino on behalf of COVID-19 FIOCRUZ Genomic Network; Thais Ferreira de Oliveira; Thais de Oliveira Costa; Ticiane Cavalcante de Souza; Veridiana Pessoa Miyajima |

| | | | |
|---|---|---|---|
| | | CE) | |
| EPI_ISL_3536375 | UBS DO CAMPO LINDO | Analytical Competence Molecular Epidemiology Lab/ACME, Oswaldo Cruz Foundation, Ceara (FIOCRUZ CE) | Cleber Furtado Aksenen; Fabio Miyajima; Fernando Braga Stehling; Francisco Eder de Moura Lopes; Jamille Maria Mendes Bezerra; Joaquim César do Nascimento Sousa Junior; Pedro Miguel Carneiro Jeronimo; Suzana Porto Almeida e Lucas Delerino; Thais Ferreira de Oliveira; Thais de Oliveira Costa; Ticiane Cavalcante de Souza; Veridiana Pessoa Miyajima |
| EPI_ISL_2494114, EPI_ISL_2494118, EPI_ISL_2494119, EPI_ISL_2494120, EPI_ISL_2494121, EPI_ISL_2494122, EPI_ISL_2494126, EPI_ISL_2494129 | | | |
| see above | UBS DR ALFREDO DANTAS DE SOUZA UMUARAMA | Instituto Butantan | Antonio Jorge Martins; Claudia Renata dos Santos Barros; David Schlesinger; Debora Botequio Moretti; Dimas Tadeu Covas; Elaine Cristina Marqueze; Elaine Vieira Santos; Evandra Strazza Rodrigues; Heidge Fukumasu; Jayme Augusto de Souza-Neto; José Salvatore Leister Patané; Luiz Alcantara; Luiz Lehmann Coutinho; Maria Carolina Elias; Maurício Lacerda Nogueira; Rafael dos Santos Bezerra; Raul Machado Neto; Rejane Maria Tommasini Grotto; Ricardo Haddad; Sandra Coccuzzo Sampaio Vessoni; Simone Kashima; Svetoslav Nanev Slavov; Vincent Louis Viala |
| EPI_ISL_2494320 | UBS JARDIM CAIUBY | Instituto Butantan | Antonio Jorge Martins; Claudia Renata dos Santos Barros; David Schlesinger; Debora Botequio Moretti; Dimas Tadeu Covas; Elaine Cristina Marqueze; Elaine Vieira Santos; Evandra Strazza Rodrigues; Heidge Fukumasu; Jayme Augusto de Souza-Neto; José Salvatore Leister Patané; Luiz Alcantara; Luiz Lehmann Coutinho; Maria Carolina Elias; Maurício Lacerda Nogueira; Rafael dos Santos Bezerra; Raul Machado Neto; Rejane Maria Tommasini Grotto; Ricardo Haddad; Sandra Coccuzzo Sampaio Vessoni; Simone Kashima; Svetoslav Nanev Slavov; Vincent Louis Viala |
| EPI_ISL_2493849 | UBS JARDIM ODETE | Instituto Butantan | Antonio Jorge Martins; Claudia Renata dos Santos Barros; David Schlesinger; Debora Botequio Moretti; Dimas Tadeu Covas; Elaine Cristina Marqueze; Elaine Vieira Santos; Evandra Strazza Rodrigues; Heidge Fukumasu; Jayme Augusto de Souza-Neto; José Salvatore Leister Patané; Luiz Alcantara; Luiz Lehmann Coutinho; Maria Carolina Elias; Maurício Lacerda Nogueira; Rafael dos Santos Bezerra; Raul Machado Neto; Rejane Maria Tommasini Grotto; Ricardo Haddad; Sandra Coccuzzo Sampaio Vessoni; Simone Kashima; Svetoslav Nanev Slavov; Vincent Louis Viala |
| EPI_ISL_3912288 | UBS MARATOAN | Analytical Competence Molecular Epidemiology Lab/ACME, Oswaldo Cruz Foundation, Ceara (FIOCRUZ CE) | Cleber Furtado Aksenen; Fabio Miyajima; Fernando Braga Stehling; Francisco Eder de Moura Lopes; Jamille Maria Mendes Bezerra; Joaquim Cesar do Nascimento Sousa Junior; Pedro Miguel Carneiro Jeronimo; Suzana Porto Almeida & Lucas Delerino on behalf of COVID-19 FIOCRUZ Genomic Network; Thais Ferreira de Oliveira; Thais de Oliveira Costa; Ticiane Cavalcante de Souza; Veridiana Pessoa Miyajima |
| EPI_ISL_3419663, EPI_ISL_3419683, EPI_ISL_3419684, EPI_ISL_3419690 | UC Davis Genome Center and Healthy Davis Together | UC Davis Genome Center | UC Davis |
| EPI_ISL_3067001, EPI_ISL_3067009, EPI_ISL_3067011, EPI_ISL_3067012, EPI_ISL_3067050, EPI_ISL_3370432, EPI_ISL_3370433, EPI_ISL_3370457 | | | |
| see above | UC-Christus Clinical Hospital Laboratory | Laboratory of Molecular Virology, School of Medicine, Pontificia Universidad Catolica de Chile | Ana Maira Guzman; Andres E. Munoz-Marcos; Catalina Pardo-Roa; Eileen Serrano; Erick Salinas; Estefany Poblete; Francisco Melo; Jorge Levican; Leonardo I. Almonacid; Maria Jose Avendano; Maria Patricia Vega; Rafael A. Medina; Ricardo Enrique de la Barra; Tamara Garcia-Salum |
| EPI_ISL_2860319, EPI_ISL_2860326 | UCC | Grubaugh Lab - Yale School of Public Health | Anderson Brito; Andrea Firpo; Annie Watkins; Chaney Kalinich; Chantal Vogels; David Rivera Aponte; Ernesto Vazquez; Fabiola Cruz-Lopez; Fabiola Fontanet; Isabel Ott; Jessica Rothman; Joseph Fauver; Karolane Gonzalez; Kendall Billig; Mallery Breban; Mary Petrone; Natalie Machargo; Nathan Grubaugh; Patricia Serrano; Tania Mitwalli; Tara Alpert; Tobias Koch; Vianca Aponte |
| EPI_ISL_2960091, EPI_ISL_2960100, EPI_ISL_2960102, EPI_ISL_3025377, EPI_ISL_3025378, EPI_ISL_3025382, EPI_ISL_3025383, EPI_ISL_3025385, EPI_ISL_3025388, EPI_ISL_3104709, EPI_ISL_3236525 | | | |
| see above | UCC (Universidad Central del Caribe) | Grubaugh Lab - Yale School of Public Health | Alejandaro Vallejo Degaudenzi; Anderson Brito; Andrea Firpo; Annie Watkins; Chaney Kalinich; Chantal Vogels; David Rivera Aponte; Elisa Contreras; Ernesto Vazquez; Esperanza Mendoza; Fabiola Cruz-Lopez; Fabiola Fontanet; Isabel Ott; Jessica Rothman; Joseph Fauver; Karolane Gonzalez; Kendall Billig; Mallery Breban; Mary Petrone; Natalie Machargo; Nathan Grubaugh; Patricia Serrano; Robert Paulino-Ramirez; Tania Mitwalli; Tara Alpert; Tobias Koch; Vianca Aponte; Victor Virgilio Calderon |
| EPI_ISL_3506323, EPI_ISL_3710096 | UCLA Clinical Micro Lab | Los Angeles County Public Health Laboratories | P. Hemarajata et al. |
| EPI_ISL_2547293, EPI_ISL_2547300, EPI_ISL_2547301, EPI_ISL_2547302, EPI_ISL_2547313, EPI_ISL_2547319, EPI_ISL_2547320, EPI_ISL_2547321, EPI_ISL_2661538, EPI_ISL_2661544, EPI_ISL_2661545, EPI_ISL_2661546, EPI_ISL_2661547, EPI_ISL_2661548, EPI_ISL_2661549, EPI_ISL_3462085, EPI_ISL_3462086, EPI_ISL_3462088, EPI_ISL_3462089, EPI_ISL_3462090 | | | |
| see above | UHTL, University Hospitals | UHTL, University Hospitals | Alouani, D.; Sadri, N.; Song, X. |
| EPI_ISL_2695325, EPI_ISL_2695326 | ULSM - Matosinhos | Instituto Nacional de Saude (INSA) | Borges et al |
| EPI_ISL_3061561 | UNC-CH COVID Surveillance Lab | Jeremy Wang | Amir Barzin; Amy James Loftis; Corbin Jones; Jeremy Wang; Melissa Miller; Susan Fiscus |
| EPI_ISL_3066764 | UNIDAD DE DIAGNOSTICO HEMATO ONCOLOGICA | Instituto Nacional de Salud | Carlos Franco-Muñoz; Carmen Osorio; Diana Malo; Diego A. Álvarez-Díaz; Diego Andrés Prada; Gerardo Santamaría; Hector Alejandro Ruiz-Moreno; Jhonnatan Reales-González; Jorge Rivera; Juan Camilo Martinez; Julian Naizaque; Katherine Laiton-Donato; Lisseth Pardo; Magdalena Wiesner; Marcela Mercado-Reyes; Maria T.Herrera-Sepúlveda; Marta Lopez Blanco; Martha Lucia Ospina Martinez; Paola Rojas; Sergio Gomez; Sheryll Corchuelo; Ángela Alarcon Cruz |
| EPI_ISL_3385787 | UNIDAD DE DIAGNOSTICO HEMATO ONCOLOGICA | Instituto Nacional de Salud-Dirección de Investigación en Salud Pública | Carlos Franco-Muñoz; Carmen Osorio; Diana Malo; Diego A. Álvarez-Díaz; Diego Andrés Prada; Gerardo Santamaría; Hector Alejandro Ruiz-Moreno; Jhonnatan Reales-González; Jorge Rivera; Juan Camilo Martinez; Julian Naizaque; Katherine Laiton-Donato; Lisseth Pardo; Magdalena Wiesner; Marcela Mercado-Reyes; Maria T. Herrera-Sepúlveda; Marta Lopez Blanco; Martha Lucia Ospina Martinez; Paola Rojas; Sergio Gomez; Sheryll Corchuelo; Ángela Alarcon Cruz |
| EPI_ISL_3536379 | UNIDADE BASICA DE SAUDE ALTO BRILHANTE | Analytical Competence Molecular Epidemiology Lab/ACME, Oswaldo Cruz Foundation, Ceara (FIOCRUZ CE) | Cleber Furtado Aksenen; Fabio Miyajima; Fernando Braga Stehling; Francisco Eder de Moura Lopes; Jamille Maria Mendes Bezerra; Joaquim César do Nascimento Sousa Junior; Pedro Miguel Carneiro Jeronimo; Suzana Porto Almeida e Lucas Delerino; Thais Ferreira de Oliveira; Thais de Oliveira Costa; Ticiane Cavalcante de Souza; Veridiana Pessoa Miyajima |
| EPI_ISL_3536380 | UNIDADE BASICA DE SAUDE COLIBRIS | Analytical Competence Molecular Epidemiology Lab/ACME, Oswaldo Cruz Foundation, Ceara (FIOCRUZ CE) | Cleber Furtado Aksenen; Fabio Miyajima; Fernando Braga Stehling; Francisco Eder de Moura Lopes; Jamille Maria Mendes Bezerra; Joaquim César do Nascimento Sousa Junior; Pedro Miguel Carneiro Jeronimo; Suzana Porto Almeida e Lucas Delerino; Thais Ferreira de Oliveira; Thais de Oliveira Costa; Ticiane Cavalcante de Souza; Veridiana Pessoa Miyajima |
| EPI_ISL_3912293 | UNIDADE BASICA DE SAUDE DA GIRILANDIA | Analytical Competence Molecular Epidemiology Lab/ACME, Oswaldo Cruz Foundation, Ceara (FIOCRUZ CE) | Cleber Furtado Aksenen; Fabio Miyajima; Fernando Braga Stehling; Francisco Eder de Moura Lopes; Jamille Maria Mendes Bezerra; Joaquim Cesar do Nascimento Sousa Junior; Pedro Miguel Carneiro Jeronimo; Suzana Porto Almeida & Lucas Delerino on behalf of COVID-19 FIOCRUZ Genomic Network; Thais Ferreira de Oliveira; Thais de Oliveira Costa; Ticiane Cavalcante de Souza; Veridiana Pessoa Miyajima |
| EPI_ISL_3912294, EPI_ISL_3912295 | UNIDADE BASICA DE SAUDE DA SEDE | Analytical Competence Molecular Epidemiology Lab/ACME, Oswaldo Cruz Foundation, Ceara (FIOCRUZ CE) | Cleber Furtado Aksenen; Fabio Miyajima; Fernando Braga Stehling; Francisco Eder de Moura Lopes; Jamille Maria Mendes Bezerra; Joaquim Cesar do Nascimento Sousa Junior; Pedro Miguel Carneiro Jeronimo; Suzana Porto Almeida & Lucas Delerino on behalf of COVID-19 FIOCRUZ Genomic Network; Thais Ferreira de Oliveira; Thais de Oliveira Costa; Ticiane Cavalcante de Souza; Veridiana Pessoa Miyajima |
| EPI_ISL_3912296 | UNIDADE BASICA DE SAUDE DE BOA AGUA | Analytical Competence Molecular Epidemiology Lab/ACME, Oswaldo Cruz Foundation, Ceara (FIOCRUZ CE) | Cleber Furtado Aksenen; Fabio Miyajima; Fernando Braga Stehling; Francisco Eder de Moura Lopes; Jamille Maria Mendes Bezerra; Joaquim Cesar do Nascimento Sousa Junior; Pedro Miguel Carneiro Jeronimo; Suzana Porto Almeida & Lucas Delerino on behalf of COVID-19 FIOCRUZ Genomic Network; Thais Ferreira de Oliveira; Thais de Oliveira Costa; Ticiane Cavalcante de Souza; Veridiana Pessoa Miyajima |
| EPI_ISL_3912297 | UNIDADE BASICA DE SAUDE DE ROLDAO | Analytical Competence Molecular Epidemiology Lab/ACME, Oswaldo Cruz Foundation, Ceara (FIOCRUZ CE) | Cleber Furtado Aksenen; Fabio Miyajima; Fernando Braga Stehling; Francisco Eder de Moura Lopes; Jamille Maria Mendes Bezerra; Joaquim Cesar do Nascimento Sousa Junior; Pedro Miguel Carneiro Jeronimo; Suzana Porto Almeida & Lucas Delerino on behalf of COVID-19 FIOCRUZ Genomic Network; Thais Ferreira de Oliveira; Thais de Oliveira Costa; Ticiane Cavalcante de Souza; Veridiana Pessoa Miyajima |
| EPI_ISL_3536344 | UNIDADE BASICA DE SAUDE DE SAO VICENTE | Analytical Competence Molecular Epidemiology Lab/ACME, Oswaldo Cruz Foundation, Ceara (FIOCRUZ CE) | Cleber Furtado Aksenen; Fabio Miyajima; Fernando Braga Stehling; Francisco Eder de Moura Lopes; Jamille Maria Mendes Bezerra; Joaquim César do Nascimento Sousa Junior; Pedro Miguel Carneiro Jeronimo; Suzana Porto Almeida e Lucas Delerino; Thais Ferreira de Oliveira; Thais de Oliveira Costa; Ticiane Cavalcante de Souza; Veridiana Pessoa Miyajima |
| EPI_ISL_3912298 | UNIDADE BASICA DE SAUDE DIVINO ESPIRITO SANTO | Analytical Competence Molecular Epidemiology Lab/ACME, Oswaldo Cruz Foundation, Ceara (FIOCRUZ CE) | Cleber Furtado Aksenen; Fabio Miyajima; Fernando Braga Stehling; Francisco Eder de Moura Lopes; Jamille Maria Mendes Bezerra; Joaquim Cesar do Nascimento Sousa Junior; Pedro Miguel Carneiro Jeronimo; Suzana Porto Almeida & Lucas Delerino on behalf of COVID-19 FIOCRUZ Genomic Network; Thais Ferreira de Oliveira; Thais de Oliveira Costa; Ticiane Cavalcante de Souza; Veridiana Pessoa Miyajima |
| EPI_ISL_3912299, EPI_ISL_3912300 | UNIDADE BASICA DE SAUDE DO CAMPO DE AVIACAO | Analytical Competence Molecular Epidemiology Lab/ACME, Oswaldo Cruz Foundation, Ceara (FIOCRUZ CE) | Cleber Furtado Aksenen; Fabio Miyajima; Fernando Braga Stehling; Francisco Eder de Moura Lopes; Jamille Maria Mendes Bezerra; Joaquim Cesar do Nascimento Sousa Junior; Pedro Miguel Carneiro Jeronimo; Suzana Porto Almeida & Lucas Delerino on behalf of COVID-19 FIOCRUZ Genomic Network; Thais Ferreira de Oliveira; Thais de Oliveira Costa; Ticiane Cavalcante de Souza; Veridiana Pessoa Miyajima |
| EPI_ISL_3912301 | UNIDADE BASICA DE SAUDE DO SAO FRANCISCO | Analytical Competence Molecular Epidemiology Lab/ACME, Oswaldo Cruz Foundation, Ceara (FIOCRUZ CE) | Cleber Furtado Aksenen; Fabio Miyajima; Fernando Braga Stehling; Francisco Eder de Moura Lopes; Jamille Maria Mendes Bezerra; Joaquim Cesar do Nascimento Sousa Junior; Pedro Miguel Carneiro Jeronimo; Suzana Porto Almeida & Lucas Delerino on behalf of COVID-19 FIOCRUZ Genomic Network; Thais Ferreira de Oliveira; Thais de Oliveira Costa; Ticiane Cavalcante de Souza; Veridiana Pessoa Miyajima |
| EPI_ISL_2494175 | UNIDADE BASICA DE SAUDE DR MATHEUS GABRIEL BONASSA | Instituto Butantan | Antonio Jorge Martins; Claudia Renata dos Santos Barros; David Schlesinger; Debora Botequio Moretti; Dimas Tadeu Covas; Elaine Cristina Marqueze; Elaine Vieira Santos; Evandra Strazza Rodrigues; Heidge Fukumasu; Jayme Augusto de Souza-Neto; José Salvatore Leister Patané; Luiz Alcantara; Luiz Lehmann Coutinho; Maria Carolina Elias; Maurício Lacerda Nogueira; Rafael dos Santos Bezerra; Raul Machado Neto; Rejane Maria Tommasini Grotto; Ricardo Haddad; Sandra Coccuzzo Sampaio Vessoni; Simone Kashima; Svetoslav Nanev Slavov; Vincent Louis Viala |
| EPI_ISL_3536329 | UNIDADE BASICA DE SAUDE FAZENDINHA | Analytical Competence Molecular Epidemiology Lab/ACME, Oswaldo Cruz Foundation, Ceara (FIOCRUZ CE) | Cleber Furtado Aksenen; Fabio Miyajima; Fernando Braga Stehling; Francisco Eder de Moura Lopes; Jamille Maria Mendes Bezerra; Joaquim César do Nascimento Sousa Junior; Pedro Miguel Carneiro Jeronimo; Suzana Porto Almeida e Lucas Delerino; Thais Ferreira de Oliveira; Thais de Oliveira Costa; Ticiane Cavalcante de Souza; Veridiana Pessoa Miyajima |
| EPI_ISL_3536178, EPI_ISL_3536179, EPI_ISL_3536180 | UNIDADE BASICA DE SAUDE HORACIO FONTENELE DE SAMPAIO | Analytical Competence Molecular Epidemiology Lab/ACME, Oswaldo Cruz Foundation, Ceara (FIOCRUZ CE) | Cleber Furtado Aksenen; Fabio Miyajima; Fernando Braga Stehling; Francisco Eder de Moura Lopes; Jamille Maria Mendes Bezerra; Joaquim César do Nascimento Sousa Junior; Pedro Miguel Carneiro Jeronimo; Suzana Porto Almeida e Lucas Delerino; Thais Ferreira de Oliveira; Thais de Oliveira Costa; Ticiane Cavalcante de Souza; Veridiana Pessoa Miyajima |
| EPI_ISL_3536330 | UNIDADE BASICA DE SAUDE PICOS | Analytical Competence Molecular Epidemiology Lab/ACME, Oswaldo Cruz Foundation, Ceara (FIOCRUZ CE) | Cleber Furtado Aksenen; Fabio Miyajima; Fernando Braga Stehling; Francisco Eder de Moura Lopes; Jamille Maria Mendes Bezerra; Joaquim César do Nascimento Sousa Junior; Pedro Miguel Carneiro Jeronimo; Suzana Porto Almeida e Lucas Delerino; Thais Ferreira de Oliveira; Thais de Oliveira Costa; Ticiane Cavalcante de Souza; Veridiana Pessoa Miyajima |

| | | | |
|---|---|---|---|
| EPI_ISL_3536381 | UNIDADE BASICA DE SAUDE SANTA TEREZA | Analytical Competence Molecular Epidemiology Lab/ACME, Oswaldo Cruz Foundation, Ceara (FIOCRUZ CE) | Cleber Furtado Aksenen; Fabio Miyajima; Fernando Braga Stehling; Francisco Eder de Moura Lopes; Jamille Maria Mendes Bezerra; Joaquim César do Nascimento Sousa Junior; Pedro Miguel Carneiro Jeronimo; Suzana Porto Almeida e Lucas Delerino; Thais Ferreira de Oliveira; Thais de Oliveira Costa; Ticiane Cavalcante de Souza; Veridiana Pessoa Miyajima |
| EPI_ISL_3536382 | UNIDADE BASICA DE SAUDE SANTANA | Analytical Competence Molecular Epidemiology Lab/ACME, Oswaldo Cruz Foundation, Ceara (FIOCRUZ CE) | Cleber Furtado Aksenen; Fabio Miyajima; Fernando Braga Stehling; Francisco Eder de Moura Lopes; Jamille Maria Mendes Bezerra; Joaquim César do Nascimento Sousa Junior; Pedro Miguel Carneiro Jeronimo; Suzana Porto Almeida e Lucas Delerino; Thais Ferreira de Oliveira; Thais de Oliveira Costa; Ticiane Cavalcante de Souza; Veridiana Pessoa Miyajima |
| EPI_ISL_3536365, EPI_ISL_3912303 | UNIDADE BASICA DE SAUDE SEDE I | Analytical Competence Molecular Epidemiology Lab/ACME, Oswaldo Cruz Foundation, Ceara (FIOCRUZ CE) | Cleber Furtado Aksenen; Fabio Miyajima; Fernando Braga Stehling; Francisco Eder de Moura Lopes; Jamille Maria Mendes Bezerra; Joaquim Cesar do Nascimento Sousa Junior; Joaquim César do Nascimento Sousa Junior; Pedro Miguel Carneiro Jeronimo; Suzana Porto Almeida & Lucas Delerino on behalf of COVID-19 FIOCRUZ Genomic Network; Suzana Porto Almeida e Lucas Delerino; Thais Ferreira de Oliveira; Thais de Oliveira Costa; Ticiane Cavalcante de Souza; Veridiana Pessoa Miyajima |
| EPI_ISL_3536338 | UNIDADE BASICA DE SAUDE VALMIR XAVIER | Analytical Competence Molecular Epidemiology Lab/ACME, Oswaldo Cruz Foundation, Ceara (FIOCRUZ CE) | Cleber Furtado Aksenen; Fabio Miyajima; Fernando Braga Stehling; Francisco Eder de Moura Lopes; Jamille Maria Mendes Bezerra; Joaquim César do Nascimento Sousa Junior; Pedro Miguel Carneiro Jeronimo; Suzana Porto Almeida e Lucas Delerino; Thais Ferreira de Oliveira; Thais de Oliveira Costa; Ticiane Cavalcante de Souza; Veridiana Pessoa Miyajima |
| EPI_ISL_3536148 | UNIDADE BASICA DE SAUDE VILA COBAL | Analytical Competence Molecular Epidemiology Lab/ACME, Oswaldo Cruz Foundation, Ceara (FIOCRUZ CE) | Cleber Furtado Aksenen; Fabio Miyajima; Fernando Braga Stehling; Francisco Eder de Moura Lopes; Jamille Maria Mendes Bezerra; Joaquim César do Nascimento Sousa Junior; Pedro Miguel Carneiro Jeronimo; Suzana Porto Almeida e Lucas Delerino; Thais Ferreira de Oliveira; Thais de Oliveira Costa; Ticiane Cavalcante de Souza; Veridiana Pessoa Miyajima |
| EPI_ISL_3912307, EPI_ISL_3912308, EPI_ISL_3912309 | UNIDADE DE PRONTO ATENDIMENTO ARACATI UPA ARACATI | Analytical Competence Molecular Epidemiology Lab/ACME, Oswaldo Cruz Foundation, Ceara (FIOCRUZ CE) | Cleber Furtado Aksenen; Fabio Miyajima; Fernando Braga Stehling; Francisco Eder de Moura Lopes; Jamille Maria Mendes Bezerra; Joaquim Cesar do Nascimento Sousa Junior; Pedro Miguel Carneiro Jeronimo; Suzana Porto Almeida & Lucas Delerino on behalf of COVID-19 FIOCRUZ Genomic Network; Thais Ferreira de Oliveira; Thais de Oliveira Costa; Ticiane Cavalcante de Souza; Veridiana Pessoa Miyajima |
| EPI_ISL_3912311 | UNIDADE DE PRONTO ATENDIMENTO DA JUREMA | Analytical Competence Molecular Epidemiology Lab/ACME, Oswaldo Cruz Foundation, Ceara (FIOCRUZ CE) | Cleber Furtado Aksenen; Fabio Miyajima; Fernando Braga Stehling; Francisco Eder de Moura Lopes; Jamille Maria Mendes Bezerra; Joaquim Cesar do Nascimento Sousa Junior; Pedro Miguel Carneiro Jeronimo; Suzana Porto Almeida & Lucas Delerino on behalf of COVID-19 FIOCRUZ Genomic Network; Thais Ferreira de Oliveira; Thais de Oliveira Costa; Ticiane Cavalcante de Souza; Veridiana Pessoa Miyajima |
| EPI_ISL_2462319, EPI_ISL_2494102, EPI_ISL_2494104, EPI_ISL_2494105, EPI_ISL_2494106, EPI_ISL_2494334 | UNIDADE DE PRONTO ATENDIMENTO DE AGENOR DE CAMPOS | Instituto Butantan | Antonio Jorge Martins; Claudia Renata dos Santos Barros; David Schlesinger; Debora Botequio Moretti; Dimas Tadeu Covas; Elaine Cristina Marqueze; Elaine Vieira Santos; Evandra Strazza Rodrigues; Heidge Fukumasu; Jayme Augusto de Souza-Neto; José Salvatore Leister Patané; Luiz Alcantara; Luiz Lehmann Coutinho; Maria Carolina Elias; Maurício Lacerda Nogueira; Rafael dos Santos Bezerra; Raul Machado Neto; Rejane Maria Tommasini Grotto; Ricardo Haddad; Sandra Coccuzzo Sampaio Vessoni; Simone Kashima; Svetoslav Nanev Slavov; Vincent Louis Viala |
| EPI_ISL_3912314, EPI_ISL_3912315, EPI_ISL_3912316, EPI_ISL_3912317 | UNIDADE DE PRONTO ATENDIMENTO DE EUSEBIO | Analytical Competence Molecular Epidemiology Lab/ACME, Oswaldo Cruz Foundation, Ceara (FIOCRUZ CE) | Cleber Furtado Aksenen; Fabio Miyajima; Fernando Braga Stehling; Francisco Eder de Moura Lopes; Jamille Maria Mendes Bezerra; Joaquim Cesar do Nascimento Sousa Junior; Pedro Miguel Carneiro Jeronimo; Suzana Porto Almeida & Lucas Delerino on behalf of COVID-19 FIOCRUZ Genomic Network; Thais Ferreira de Oliveira; Thais de Oliveira Costa; Ticiane Cavalcante de Souza; Veridiana Pessoa Miyajima |
| EPI_ISL_2493835 | UNIDADE DE PRONTO ATENDIMENTO DE PITANGUEIRAS | Instituto Butantan | Antonio Jorge Martins; Claudia Renata dos Santos Barros; David Schlesinger; Debora Botequio Moretti; Dimas Tadeu Covas; Elaine Cristina Marqueze; Elaine Vieira Santos; Evandra Strazza Rodrigues; Heidge Fukumasu; Jayme Augusto de Souza-Neto; José Salvatore Leister Patané; Luiz Alcantara; Luiz Lehmann Coutinho; Maria Carolina Elias; Maurício Lacerda Nogueira; Rafael dos Santos Bezerra; Raul Machado Neto; Rejane Maria Tommasini Grotto; Ricardo Haddad; Sandra Coccuzzo Sampaio Vessoni; Simone Kashima; Svetoslav Nanev Slavov; Vincent Louis Viala |
| EPI_ISL_2493844 | UNIDADE DE SAUDE DA FAMILIA JARDIM AMERICA ITAQUAQUECETUBA | Instituto Butantan | Antonio Jorge Martins; Claudia Renata dos Santos Barros; David Schlesinger; Debora Botequio Moretti; Dimas Tadeu Covas; Elaine Cristina Marqueze; Elaine Vieira Santos; Evandra Strazza Rodrigues; Heidge Fukumasu; Jayme Augusto de Souza-Neto; José Salvatore Leister Patané; Luiz Alcantara; Luiz Lehmann Coutinho; Maria Carolina Elias; Maurício Lacerda Nogueira; Rafael dos Santos Bezerra; Raul Machado Neto; Rejane Maria Tommasini Grotto; Ricardo Haddad; Sandra Coccuzzo Sampaio Vessoni; Simone Kashima; Svetoslav Nanev Slavov; Vincent Louis Viala |
| EPI_ISL_2494181 | UNIDADE DE VIGILANCIA EPIDEMIOLOGICA | Instituto Butantan | Antonio Jorge Martins; Claudia Renata dos Santos Barros; David Schlesinger; Debora Botequio Moretti; Dimas Tadeu Covas; Elaine Cristina Marqueze; Elaine Vieira Santos; Evandra Strazza Rodrigues; Heidge Fukumasu; Jayme Augusto de Souza-Neto; José Salvatore Leister Patané; Luiz Alcantara; Luiz Lehmann Coutinho; Maria Carolina Elias; Maurício Lacerda Nogueira; Rafael dos Santos Bezerra; Raul Machado Neto; Rejane Maria Tommasini Grotto; Ricardo Haddad; Sandra Coccuzzo Sampaio Vessoni; Simone Kashima; Svetoslav Nanev Slavov; Vincent Louis Viala |
| EPI_ISL_2493947 | UNIDADE MISTA ARACOIABA DA SERRA | Instituto Butantan | Antonio Jorge Martins; Claudia Renata dos Santos Barros; David Schlesinger; Debora Botequio Moretti; Dimas Tadeu Covas; Elaine Cristina Marqueze; Elaine Vieira Santos; Evandra Strazza Rodrigues; Heidge Fukumasu; Jayme Augusto de Souza-Neto; José Salvatore Leister Patané; Luiz Alcantara; Luiz Lehmann Coutinho; Maria Carolina Elias; Maurício Lacerda Nogueira; Rafael dos Santos Bezerra; Raul Machado Neto; Rejane Maria Tommasini Grotto; Ricardo Haddad; Sandra Coccuzzo Sampaio Vessoni; Simone Kashima; Svetoslav Nanev Slavov; Vincent Louis Viala |
| EPI_ISL_3912320 | UNIDADE PRONTO ATENDIMENTO AUTRAN NUNES | Analytical Competence Molecular Epidemiology Lab/ACME, Oswaldo Cruz Foundation, Ceara (FIOCRUZ CE) | Cleber Furtado Aksenen; Fabio Miyajima; Fernando Braga Stehling; Francisco Eder de Moura Lopes; Jamille Maria Mendes Bezerra; Joaquim Cesar do Nascimento Sousa Junior; Pedro Miguel Carneiro Jeronimo; Suzana Porto Almeida & Lucas Delerino on behalf of COVID-19 FIOCRUZ Genomic Network; Thais Ferreira de Oliveira; Thais de Oliveira Costa; Ticiane Cavalcante de Souza; Veridiana Pessoa Miyajima |
| EPI_ISL_3102331 | UNIDADE PRONTO ATENDIMENTO PRAIA DO FUTURO | Analytical Competence Molecular Epidemiology Lab/ACME, Oswaldo Cruz Foundation, Ceara (FIOCRUZ CE) | Cleber Furtado Aksenen; Fabio Miyajima; Fernando Braga Stehling; Francisco Eder de Moura Lopes; Jamille Maria Mendes Bezerra; Joaquim César do Nascimento Sousa Junior; Pedro Miguel Carneiro Jeronimo; Suzana Porto Almeida e Lucas Delerino; Thais Ferreira de Oliveira; Thais de Oliveira Costa; Ticiane Cavalcante de Souza; Veridiana Pessoa Miyajima |
| EPI_ISL_3102298, EPI_ISL_3536194, EPI_ISL_3536195, EPI_ISL_3536196, EPI_ISL_3536197, EPI_ISL_3536198, EPI_ISL_3536201, EPI_ISL_3912087, EPI_ISL_3912088, EPI_ISL_3912094, EPI_ISL_3912337, EPI_ISL_3912338, EPI_ISL_3912339, EPI_ISL_3912341, EPI_ISL_3912342, EPI_ISL_3912343, EPI_ISL_3912344, EPI_ISL_3912345, EPI_ISL_3912348, EPI_ISL_3912355, EPI_ISL_3912358, EPI_ISL_3912359, EPI_ISL_3912362, EPI_ISL_3912363, EPI_ISL_3912364, EPI_ISL_3912365, EPI_ISL_3912366, EPI_ISL_3912367, EPI_ISL_3912369, EPI_ISL_3912370, EPI_ISL_3912371, EPI_ISL_3912372, EPI_ISL_3912373, EPI_ISL_3912377, EPI_ISL_3912378, EPI_ISL_3912379 | | | |
| see above | UNIDADE REFERENCIA COVID 19 | Analytical Competence Molecular Epidemiology Lab/ACME, Oswaldo Cruz Foundation, Ceara (FIOCRUZ CE) | Cleber Furtado Aksenen; Fabio Miyajima; Fernando Braga Stehling; Francisco Eder de Moura Lopes; Jamille Maria Mendes Bezerra; Joaquim Cesar do Nascimento Sousa Junior; Joaquim César do Nascimento Sousa Junior; Pedro Miguel Carneiro Jeronimo; Suzana Porto Almeida & Lucas Delerino on behalf of COVID-19 FIOCRUZ Genomic Network; Suzana Porto Almeida e Lucas Delerino; Thais Ferreira de Oliveira; Thais de Oliveira Costa; Ticiane Cavalcante de Souza; Veridiana Pessoa Miyajima |
| EPI_ISL_2493897, EPI_ISL_2493912 | UNIDADE SENTINELA COVID19 | Instituto Butantan | Antonio Jorge Martins; Claudia Renata dos Santos Barros; David Schlesinger; Debora Botequio Moretti; Dimas Tadeu Covas; Elaine Cristina Marqueze; Elaine Vieira Santos; Evandra Strazza Rodrigues; Heidge Fukumasu; Jayme Augusto de Souza-Neto; José Salvatore Leister Patané; Luiz Alcantara; Luiz Lehmann Coutinho; Maria Carolina Elias; Maurício Lacerda Nogueira; Rafael dos Santos Bezerra; Raul Machado Neto; Rejane Maria Tommasini Grotto; Ricardo Haddad; Sandra Coccuzzo Sampaio Vessoni; Simone Kashima; Svetoslav Nanev Slavov; Vincent Louis Viala |
| EPI_ISL_2493907 | UNIDADE SENTINELA VILA APARECIDA | Instituto Butantan | Antonio Jorge Martins; Claudia Renata dos Santos Barros; David Schlesinger; Debora Botequio Moretti; Dimas Tadeu Covas; Elaine Cristina Marqueze; Elaine Vieira Santos; Evandra Strazza Rodrigues; Heidge Fukumasu; Jayme Augusto de Souza-Neto; José Salvatore Leister Patané; Luiz Alcantara; Luiz Lehmann Coutinho; Maria Carolina Elias; Maurício Lacerda Nogueira; Rafael dos Santos Bezerra; Raul Machado Neto; Rejane Maria Tommasini Grotto; Ricardo Haddad; Sandra Coccuzzo Sampaio Vessoni; Simone Kashima; Svetoslav Nanev Slavov; Vincent Louis Viala |
| EPI_ISL_2628717, EPI_ISL_2628825, EPI_ISL_2628962, EPI_ISL_2645941, EPI_ISL_2645995, EPI_ISL_2646099, EPI_ISL_2796397 | | | |
| see above | UNILABS | Instituto Nacional de Saude (INSA) | Borges et al |
| EPI_ISL_2796048 | UNILABS | Instituto Nacional de Saude (INSA) and Instituto Gulbenkian de Ciencia (IGC) | Borges et al |
| EPI_ISL_2820961 | UOC Microbiologia e Virologia, Azienda Ospedaliera Universitaria Senese, Siena, Italy | Dipartimento di Biotecnologie Mediche | Claudia Gandolfo; David Pinzauti; Francesco Santoro; Gabriele Anichini; Gianni Gori Savellini; Gianni Pozzi; Maria Grazia Cusi |
| EPI_ISL_2493816, EPI_ISL_2493848, EPI_ISL_2493852, EPI_ISL_2493858 | UPA SADAKO SEDOGUTI | Instituto Butantan | Antonio Jorge Martins; Claudia Renata dos Santos Barros; David Schlesinger; Debora Botequio Moretti; Dimas Tadeu Covas; Elaine Cristina Marqueze; Elaine Vieira Santos; Evandra Strazza Rodrigues; Heidge Fukumasu; Jayme Augusto de Souza-Neto; José Salvatore Leister Patané; Luiz Alcantara; Luiz Lehmann Coutinho; Maria Carolina Elias; Maurício Lacerda Nogueira; Rafael dos Santos Bezerra; Raul Machado Neto; Rejane Maria Tommasini Grotto; Ricardo Haddad; Sandra Coccuzzo Sampaio Vessoni; Simone Kashima; Svetoslav Nanev Slavov; Vincent Louis Viala |
| EPI_ISL_2920237 | URMC LABS | URMC LABS | Andrew Cameron; Dwight Hardy; Kelly DeLary; Stepan Voljehn |
| EPI_ISL_2868325, EPI_ISL_2868527, EPI_ISL_3031239 | URMC LABS | Wadsworth Center, New York State Department of Health | Alexis Russell; Catharine Prussing; Daryl M. Lamson; Erasmus Schneider; Erica Lasek-Nesselquist; John Kelly; Jonathan Plitnick; Kirsten St. George; Matthew Shudt; Melissa A Leisner; Navjot Singh |
| EPI_ISL_3048152, EPI_ISL_3048160 | US Air Force School of Aerospace Medicine | US Air Force School of Aerospace Medicine | Amanda Javorina; Anthony Fries; Carol Garrett; Clarise Starr; Elizabeth Macias; Jennifer Meyer; Sarah Purves; William Gruner |
| EPI_ISL_2493879 | USAFA MARACANA | Instituto Butantan | Antonio Jorge Martins; Claudia Renata dos Santos Barros; David Schlesinger; Debora Botequio Moretti; Dimas Tadeu Covas; Elaine Cristina Marqueze; Elaine Vieira Santos; Evandra Strazza Rodrigues; Heidge Fukumasu; Jayme Augusto de Souza-Neto; José Salvatore Leister Patané; Luiz Alcantara; Luiz Lehmann Coutinho; Maria Carolina Elias; Maurício Lacerda Nogueira; Rafael dos Santos Bezerra; Raul Machado Neto; Rejane Maria Tommasini Grotto; Ricardo Haddad; Sandra Coccuzzo Sampaio Vessoni; Simone Kashima; Svetoslav Nanev Slavov; Vincent Louis Viala |
| EPI_ISL_2493895 | USAFA MIRIM | Instituto Butantan | Antonio Jorge Martins; Claudia Renata dos Santos Barros; David Schlesinger; Debora Botequio Moretti; Dimas Tadeu Covas; Elaine Cristina Marqueze; Elaine Vieira Santos; Evandra Strazza Rodrigues; Heidge Fukumasu; Jayme Augusto de Souza-Neto; José Salvatore Leister Patané; Luiz Alcantara; Luiz Lehmann Coutinho; Maria Carolina Elias; Maurício Lacerda Nogueira; Rafael dos Santos Bezerra; Raul Machado Neto; Rejane Maria Tommasini Grotto; Ricardo Haddad; Sandra Coccuzzo Sampaio Vessoni; Simone Kashima; Svetoslav Nanev Slavov; Vincent Louis Viala |
| EPI_ISL_2493831 | USAFA SAO JORGE | Instituto Butantan | Antonio Jorge Martins; Claudia Renata dos Santos Barros; David Schlesinger; Debora Botequio Moretti; Dimas Tadeu Covas; Elaine Cristina Marqueze; Elaine Vieira Santos; Evandra Strazza Rodrigues; Heidge Fukumasu; Jayme Augusto de Souza-Neto; José Salvatore Leister Patané; Luiz Alcantara; Luiz Lehmann Coutinho; Maria Carolina Elias; Maurício Lacerda Nogueira; Rafael dos Santos Bezerra; Raul Machado Neto; Rejane Maria Tommasini Grotto; Ricardo Haddad; Sandra Coccuzzo Sampaio Vessoni; Simone Kashima; Svetoslav Nanev Slavov; Vincent Louis Viala |
| EPI_ISL_2493801 | USF JARDIM AMERICA FIGUEIRA | Instituto Butantan | Antonio Jorge Martins; Claudia Renata dos Santos Barros; David Schlesinger; Debora Botequio Moretti; Dimas Tadeu Covas; Elaine Cristina Marqueze; Elaine Vieira Santos; Evandra Strazza Rodrigues; Heidge Fukumasu; Jayme Augusto de Souza-Neto; José Salvatore Leister Patané; Luiz Alcantara; Luiz Lehmann Coutinho; Maria Carolina Elias; Maurício Lacerda Nogueira; Rafael dos Santos Bezerra; Raul Machado Neto; Rejane Maria Tommasini Grotto; Ricardo Haddad; Sandra Coccuzzo Sampaio Vessoni; Simone Kashima; Svetoslav Nanev Slavov; Vincent Louis Viala |
| EPI_ISL_2493811, EPI_ISL_2493814, EPI_ISL_2493817 | USF JARDIM CAMANDUCAIA | Instituto Butantan | Antonio Jorge Martins; Claudia Renata dos Santos Barros; David Schlesinger; Debora Botequio Moretti; Dimas Tadeu Covas; Elaine Cristina Marqueze; Elaine Vieira Santos; Evandra Strazza Rodrigues; Heidge Fukumasu; Jayme Augusto de Souza-Neto; José Salvatore Leister Patané; Luiz Alcantara; Luiz Lehmann Coutinho; Maria Carolina Elias; Maurício Lacerda Nogueira; Rafael dos Santos Bezerra; Raul Machado Neto; Rejane Maria Tommasini Grotto; Ricardo Haddad; Sandra Coccuzzo Sampaio Vessoni; Simone Kashima; Svetoslav Nanev Slavov; Vincent Louis Viala |
| EPI_ISL_2493799, EPI_ISL_2493810, | USF JARDIM DAS AVES MOREIRINHA | Instituto Butantan | Antonio Jorge Martins; Claudia Renata dos Santos Barros; David Schlesinger; Debora Botequio Moretti; Dimas Tadeu Covas; Elaine Cristina Marqueze; Elaine Vieira Santos; Evandra Strazza Rodrigues; Heidge Fukumasu; Jayme Augusto de Souza-Neto; José Salvatore Leister Patané; Luiz Alcantara; Luiz Lehmann Coutinho; Maria Carolina Elias; Maurício Lacerda Nogueira; Rafael dos Santos Bezerra; Raul Machado Neto; Rejane Maria Tommasini Grotto; Ricardo Haddad; Sandra Coccuzzo Sampaio Vessoni; Simone Kashima; Svetoslav Nanev Slavov; Vincent Louis Viala |

| | | | |
|---|---|---|---|
| EPI_ISL_2493812, EPI_ISL_2493813 | | | |
| EPI_ISL_2493819 | USF JARDIM JOSELY | Instituto Butantan | Antonio Jorge Martins; Claudia Renata dos Santos Barros; David Schlesinger; Debora Botequio Moretti; Dimas Tadeu Covas; Elaine Cristina Marqueze; Elaine Vieira Santos; Evandra Strazza Rodrigues; Heidge Fukumasu; Jayme Augusto de Souza-Neto; José Salvatore Leister Patané; Luiz Alcantara; Luiz Lehmann Coutinho; Maria Carolina Elias; Maurício Lacerda Nogueira; Rafael dos Santos Bezerra; Raul Machado Neto; Rejane Maria Tommasini Grotto; Ricardo Haddad; Sandra Coccuzzo Sampaio Vessoni; Simone Kashima; Svetoslav Nanev Slavov; Vincent Louis Viala |
| EPI_ISL_2493798, EPI_ISL_2493809 | USF JARDIM SAO DIMAS | Instituto Butantan | Antonio Jorge Martins; Claudia Renata dos Santos Barros; David Schlesinger; Debora Botequio Moretti; Dimas Tadeu Covas; Elaine Cristina Marqueze; Elaine Vieira Santos; Evandra Strazza Rodrigues; Heidge Fukumasu; Jayme Augusto de Souza-Neto; José Salvatore Leister Patané; Luiz Alcantara; Luiz Lehmann Coutinho; Maria Carolina Elias; Maurício Lacerda Nogueira; Rafael dos Santos Bezerra; Raul Machado Neto; Rejane Maria Tommasini Grotto; Ricardo Haddad; Sandra Coccuzzo Sampaio Vessoni; Simone Kashima; Svetoslav Nanev Slavov; Vincent Louis Viala |
| EPI_ISL_2493815 | USF JARDIM SILVESTRE II | Instituto Butantan | Antonio Jorge Martins; Claudia Renata dos Santos Barros; David Schlesinger; Debora Botequio Moretti; Dimas Tadeu Covas; Elaine Cristina Marqueze; Elaine Vieira Santos; Evandra Strazza Rodrigues; Heidge Fukumasu; Jayme Augusto de Souza-Neto; José Salvatore Leister Patané; Luiz Alcantara; Luiz Lehmann Coutinho; Maria Carolina Elias; Maurício Lacerda Nogueira; Rafael dos Santos Bezerra; Raul Machado Neto; Rejane Maria Tommasini Grotto; Ricardo Haddad; Sandra Coccuzzo Sampaio Vessoni; Simone Kashima; Svetoslav Nanev Slavov; Vincent Louis Viala |
| EPI_ISL_2493818 | USF SANTA MARIA DO AMPARO | Instituto Butantan | Antonio Jorge Martins; Claudia Renata dos Santos Barros; David Schlesinger; Debora Botequio Moretti; Dimas Tadeu Covas; Elaine Cristina Marqueze; Elaine Vieira Santos; Evandra Strazza Rodrigues; Heidge Fukumasu; Jayme Augusto de Souza-Neto; José Salvatore Leister Patané; Luiz Alcantara; Luiz Lehmann Coutinho; Maria Carolina Elias; Maurício Lacerda Nogueira; Rafael dos Santos Bezerra; Raul Machado Neto; Rejane Maria Tommasini Grotto; Ricardo Haddad; Sandra Coccuzzo Sampaio Vessoni; Simone Kashima; Svetoslav Nanev Slavov; Vincent Louis Viala |
| EPI_ISL_2494145 | USF SERROTE REGISTRO | Instituto Butantan | Antonio Jorge Martins; Claudia Renata dos Santos Barros; David Schlesinger; Debora Botequio Moretti; Dimas Tadeu Covas; Elaine Cristina Marqueze; Elaine Vieira Santos; Evandra Strazza Rodrigues; Heidge Fukumasu; Jayme Augusto de Souza-Neto; José Salvatore Leister Patané; Luiz Alcantara; Luiz Lehmann Coutinho; Maria Carolina Elias; Maurício Lacerda Nogueira; Rafael dos Santos Bezerra; Raul Machado Neto; Rejane Maria Tommasini Grotto; Ricardo Haddad; Sandra Coccuzzo Sampaio Vessoni; Simone Kashima; Svetoslav Nanev Slavov; Vincent Louis Viala |
| EPI_ISL_2493800 | USF TRES PONTES | Instituto Butantan | Antonio Jorge Martins; Claudia Renata dos Santos Barros; David Schlesinger; Debora Botequio Moretti; Dimas Tadeu Covas; Elaine Cristina Marqueze; Elaine Vieira Santos; Evandra Strazza Rodrigues; Heidge Fukumasu; Jayme Augusto de Souza-Neto; José Salvatore Leister Patané; Luiz Alcantara; Luiz Lehmann Coutinho; Maria Carolina Elias; Maurício Lacerda Nogueira; Rafael dos Santos Bezerra; Raul Machado Neto; Rejane Maria Tommasini Grotto; Ricardo Haddad; Sandra Coccuzzo Sampaio Vessoni; Simone Kashima; Svetoslav Nanev Slavov; Vincent Louis Viala |
| EPI_ISL_2493914 | USF YEN WANN SHENG | Instituto Butantan | Antonio Jorge Martins; Claudia Renata dos Santos Barros; David Schlesinger; Debora Botequio Moretti; Dimas Tadeu Covas; Elaine Cristina Marqueze; Elaine Vieira Santos; Evandra Strazza Rodrigues; Heidge Fukumasu; Jayme Augusto de Souza-Neto; José Salvatore Leister Patané; Luiz Alcantara; Luiz Lehmann Coutinho; Maria Carolina Elias; Maurício Lacerda Nogueira; Rafael dos Santos Bezerra; Raul Machado Neto; Rejane Maria Tommasini Grotto; Ricardo Haddad; Sandra Coccuzzo Sampaio Vessoni; Simone Kashima; Svetoslav Nanev Slavov; Vincent Louis Viala |
| EPI_ISL_2432602, EPI_ISL_2432619, EPI_ISL_2497213, EPI_ISL_2497214, EPI_ISL_2497215, EPI_ISL_2497216, EPI_ISL_2497221, EPI_ISL_2497232, EPI_ISL_2497247, EPI_ISL_2497263, EPI_ISL_2497268, EPI_ISL_2497287, EPI_ISL_2497292, EPI_ISL_2497299, EPI_ISL_2497303, EPI_ISL_2497310, EPI_ISL_2497325, EPI_ISL_2497338, EPI_ISL_2497356, EPI_ISL_2497365, EPI_ISL_2497383, EPI_ISL_2497392, EPI_ISL_2497396, EPI_ISL_2497932, EPI_ISL_2497933, EPI_ISL_2497934, EPI_ISL_2497935, EPI_ISL_2497936, EPI_ISL_2497937, EPI_ISL_2497938, EPI_ISL_2497939, EPI_ISL_2497940, EPI_ISL_2501473, EPI_ISL_2501477, EPI_ISL_2501481, EPI_ISL_2501482, EPI_ISL_2501483, EPI_ISL_2501487, EPI_ISL_2501489, EPI_ISL_2501490, EPI_ISL_2501491, EPI_ISL_2501492, EPI_ISL_2501493, EPI_ISL_2501494, EPI_ISL_2545020, EPI_ISL_2545075, EPI_ISL_2545107, EPI_ISL_2545119, EPI_ISL_2545123, EPI_ISL_2545153, EPI_ISL_2545154, EPI_ISL_2545155, EPI_ISL_2545157, EPI_ISL_2545159, EPI_ISL_2545160, EPI_ISL_2545161, EPI_ISL_2545162, EPI_ISL_2545163, EPI_ISL_2545165, EPI_ISL_2545166, EPI_ISL_2545167, EPI_ISL_2545168, EPI_ISL_2545169, EPI_ISL_2545170, EPI_ISL_2545171, EPI_ISL_2545172, EPI_ISL_2545173, EPI_ISL_2545174, EPI_ISL_2545175, EPI_ISL_2545176, EPI_ISL_2545177, EPI_ISL_2549343, EPI_ISL_2549346, EPI_ISL_2549354, EPI_ISL_2549382, EPI_ISL_2549399, EPI_ISL_2549432, EPI_ISL_2549447, EPI_ISL_2549458, EPI_ISL_2549471, EPI_ISL_2549478, EPI_ISL_2549495, EPI_ISL_2549496, EPI_ISL_2549504, EPI_ISL_2549519, EPI_ISL_2549520, EPI_ISL_2549523, EPI_ISL_2549532, EPI_ISL_2549540, EPI_ISL_2549543, EPI_ISL_2549548, EPI_ISL_2549553, EPI_ISL_2549555, EPI_ISL_2549556, EPI_ISL_2549557, EPI_ISL_2549558, EPI_ISL_2549559, EPI_ISL_2617628, EPI_ISL_2617970, EPI_ISL_2617975, EPI_ISL_2617987, EPI_ISL_2617992, EPI_ISL_2618002, EPI_ISL_2618005, EPI_ISL_2618023, EPI_ISL_2618038, EPI_ISL_2618049, EPI_ISL_2618053, EPI_ISL_2618056, EPI_ISL_2618088, EPI_ISL_2693116, EPI_ISL_2693129, EPI_ISL_2693180, EPI_ISL_2693253, EPI_ISL_2693260, EPI_ISL_2693278, EPI_ISL_2693299, EPI_ISL_2693684, EPI_ISL_2693746, EPI_ISL_2693781, EPI_ISL_2693801, EPI_ISL_2693813, EPI_ISL_2716479, EPI_ISL_2716488, EPI_ISL_2716734, EPI_ISL_2802896, EPI_ISL_2802923, EPI_ISL_2802937, EPI_ISL_2802944, EPI_ISL_2802955, EPI_ISL_2802959, EPI_ISL_2802990, EPI_ISL_2802995, EPI_ISL_2802998, EPI_ISL_2803005, EPI_ISL_2803309, EPI_ISL_2864531, EPI_ISL_2864533, EPI_ISL_2864534, EPI_ISL_2864537, EPI_ISL_2895012, EPI_ISL_2937933, EPI_ISL_2967614, EPI_ISL_2967625, EPI_ISL_2967658, EPI_ISL_2967691, EPI_ISL_2967692, EPI_ISL_2967707, EPI_ISL_2967725, EPI_ISL_3025927, EPI_ISL_3025931, EPI_ISL_3025955, EPI_ISL_3025960, EPI_ISL_3032167, EPI_ISL_3063560, EPI_ISL_3063563, EPI_ISL_3063607, EPI_ISL_3063609, EPI_ISL_3063657, EPI_ISL_3063684 | | | |
| see above | UW Virology Lab | UW Virology Lab | Alexander Greninger; Hong Xie; Keith R Jerome; Lasata Shrestha; Maria Lukes; Meei-Li Huang; Michelle Lin; Nathan Breit; Noah R. Baker; Patrick Mathias; Pavitra Roychoudhury; Ricardo Perez; Robert J. Livingston; Sean Ellis; Shah Mohamed Bakhash; Tien V. Nguyen |
| EPI_ISL_2942330 | Unidad de Investigacion Medica de Yucatan (UIMY) | Unidad de Genomica Avanzada | ; Alejandra Garcia-Gasca; Alejandra Hernandez-Teran; Alejandro Sanchez-Flores; Alfredo Herrera-Estrella; Alicia Ocaña-Mondragon; Andreu Comas-Garcia; Angel Gustavo Salas-Lais; Antonio Loza Roman; Bernardo Martinez-Miguel; Blanca Taboada; Brenda Irasema Maldonado-Meza; Bruno Gomez-Gil; Carla Ivon Herrera-Najera; Carlos F. Arias; Celia Boukadida; Celida Duque Molina; Celida Martinez- Rodriguez; Clara Esperanza Santacruz-Tinoco; Concepcion Grajales-Muñiz; Consorcio Mexicano de Vigilancia Genomica (CoViGen-Mex). Authors (in alphabetical order): Julio Elias Alvarado-Yaah; Cristobal Chaidez-Quiroz; Daniel Fregoso-Rueda; Daniel Lira Morales; Eduardo Becerril-Vargas; Fernando Fontove-Herrera; Fidencio Mejia-Nepomuceno; Francisco Pulido; Gloria Elena Espinosa-Ayala; Gloria Maria Molina-Salinas; Gloria Vazquez; Hector Esteban Paz-Juarez; Hector Montoya-Fuentes; Helen Haydee Fernanda Ramirez-Plascencia; Irvin Gonzalez-Lopez; Jean Pierre Gonzalez; Jesus Hernandez; Joel Armando Vazquez-Perez.; Jorge Salas-Hernandez; Jose Antonio Enciso-Moreno; Jose Arturo Martinez-Orozco; Jose Esteban Muñoz-Medina; Jose de Jesus Nuñez-Contreras; Juan Bautista Chale-Dzul; Julissa Enciso-Ibarra; Luis Alberto Ochoa-Carrera; Margarita Matias-Florentino; Maria Guadalupe Santiago-Mauricio; Maria Guadalupe de Jesus Mireles-Rivera; Mario Mujica-Sanchez; Marissa Perez-Garcia; Nelly Selem-Mojica; Pavel Isa; Ricardo Ciria Merce; Ricardo Grande; Rosa Maria Gutierrez Rios; Santiago avila-Rios; Selene Zarate; Susana Lopez; Veronica Mata-Haro; Victor Eduardo Garcia-Arias; Victor Hugo Borja-Aburto |
| EPI_ISL_2681209 | Unidad de Investigación Biomédica de Zacatecas (UIBZ) | Instituto de Biotecnología de la UNAM | ; Alejandra García-Gasca; Alejandra Hernández-Terán; Alejandro Sánchez-Flores; Alfredo Herrera-Estrella; Alicia Ocaña-Mondragón; Andreu Comas-García; Angel Gustavo Salas-Lais; Antonio Loza Román; Bernardo Martínez-Miguel; Blanca Taboada; Brenda Irasema Maldonado-Meza; Bruno Gómez-Gil; Carla Ivón Herrera-Najera; Carlos F. Arias; Celia Boukadida; Clara Esperanza Santacruz-Tinoco; Concepción Grajales-Muñiz; Consorcio Mexicano de Vigilancia Genómica (CoViGen-Mex). Authors (in alphabetical order): Julio Elias Alvarado-Yaah; Cristóbal Cháidez-Quiróz; Célida Duque Molina; Célida Martínez- Rodríguez; Daniel Fregoso-Rueda; Daniel Lira Morales; Eduardo Becerril-Vargas; Fernando Fontove-Herrera; Fidencio Mejía-Nepomuceno; Francisco Pulido; Gloria Elena Espinosa-Ayala; Gloria María Molina-Salinas; Gloria Vazquez; Hector Esteban Paz-Juárez; Hector Montoya-Fuentes; Helen Haydee Fernanda Ramírez-Plascencia; Irvin González-López; Jean Pierre González; Jesús Hernández; Joel Armando Vázquez-Pérez.; Jorge Salas-Hernández; José Antonio Enciso-Moreno; José Arturo Martínez-Orozco; José Esteban Muñoz-Medina; José de Jesús Nuñez-Contreras; Juan Bautista Chale-Dzul; Julissa Enciso-Ibarra; Luis Alberto Ochoa-Carrera; Margarita Matías-Florentino; Mario Mújica-Sánchez; Marissa Perez-Garcia; María Guadalupe Santiago-Mauricio; María Guadalupe de Jesús Mireles-Rivera; Nelly Sélem-Mojica; Pavel Isa; Ricardo Ciria Merce; Ricardo Grande; Rosa María Gutiérrez Rios; Santiago Ávila-Ríos; Selene Zárate; Susana Lopez; Verónica Mata-Haro; Victor Eduardo García-Arias; Víctor Hugo Borja-Aburto |
| EPI_ISL_2681055, EPI_ISL_2681090, EPI_ISL_2681141, EPI_ISL_2681244, EPI_ISL_2801588, EPI_ISL_2801617, EPI_ISL_2801642, EPI_ISL_2801656, EPI_ISL_2801657 | | | |
| see above | Unidad de Investigación Médica de Yucatán (UIMY) | Instituto de Biotecnología de la UNAM | ; Alejandra García-Gasca; Alejandra Hernández-Terán; Alejandro Sánchez-Flores; Alfredo Herrera-Estrella; Alicia Ocaña-Mondragón; Andreu Comas-García; Angel Gustavo Salas-Lais; Antonio Loza Román; Bernardo Martínez-Miguel; Blanca Taboada; Brenda Irasema Maldonado-Meza; Bruno Gómez-Gil; Carla Ivón Herrera-Najera; Carlos F. Arias; Celia Boukadida; Clara Esperanza Santacruz-Tinoco; Concepción Grajales-Muñiz; Consorcio Mexicano de Vigilancia Genómica (CoViGen-Mex). Authors (in alphabetical order): Julio Elias Alvarado-Yaah; Cristóbal Cháidez-Quiróz; Célida Duque Molina; Célida Martínez- Rodríguez; Daniel Fregoso-Rueda; Daniel Lira Morales; Eduardo Becerril-Vargas; Fernando Fontove-Herrera; Fidencio Mejía-Nepomuceno; Francisco Pulido; Gloria Elena Espinosa-Ayala; Gloria María Molina-Salinas; Gloria Vazquez; Hector Esteban Paz-Juárez; Hector Montoya-Fuentes; Helen Haydee Fernanda Ramírez-Plascencia; Irvin González-López; Jean Pierre González; Jesús Hernández; Joel Armando Vázquez-Pérez.; Jorge Salas-Hernández; José Antonio Enciso-Moreno; José Arturo Martínez-Orozco; José Esteban Muñoz-Medina; José de Jesús Nuñez-Contreras; Juan Bautista Chale-Dzul; Julissa Enciso-Ibarra; Luis Alberto Ochoa-Carrera; Margarita Matías-Florentino; Mario Mújica-Sánchez; Marissa Perez-Garcia; María Guadalupe Santiago-Mauricio; María Guadalupe de Jesús Mireles-Rivera; Nelly Sélem-Mojica; Pavel Isa; Ricardo Ciria Merce; Ricardo Grande; Rosa María Gutiérrez Rios; Santiago Ávila-Ríos; Selene Zárate; Susana Lopez; Verónica Mata-Haro; Victor Eduardo García-Arias; Víctor Hugo Borja-Aburto |
| EPI_ISL_2691569, EPI_ISL_2691570, EPI_ISL_2691571, EPI_ISL_2691572, EPI_ISL_2691574, EPI_ISL_2691575, EPI_ISL_2691576, EPI_ISL_2691577, EPI_ISL_2691578, EPI_ISL_2691579, EPI_ISL_2691581, EPI_ISL_2691582, EPI_ISL_2691583, EPI_ISL_2691584, EPI_ISL_2691586, EPI_ISL_2691587, EPI_ISL_2691590, EPI_ISL_2691591, EPI_ISL_2691592, EPI_ISL_2691593, EPI_ISL_2691594, EPI_ISL_2691595, EPI_ISL_2691596, EPI_ISL_2691598, EPI_ISL_2691599, EPI_ISL_2691600, EPI_ISL_2691601, EPI_ISL_2691602, EPI_ISL_2691603, EPI_ISL_2691604, EPI_ISL_2691605, EPI_ISL_2691606, EPI_ISL_2691607, EPI_ISL_2691608, EPI_ISL_2691609, EPI_ISL_2691610, EPI_ISL_2691611, EPI_ISL_2691612, EPI_ISL_2691613, EPI_ISL_2691614, EPI_ISL_2691615, EPI_ISL_2691616, EPI_ISL_2691617, EPI_ISL_2691618, EPI_ISL_2691619, EPI_ISL_2691620, EPI_ISL_2691621, EPI_ISL_2691622, EPI_ISL_2691628, EPI_ISL_2691629, EPI_ISL_2691630, EPI_ISL_2691631, EPI_ISL_2691633, EPI_ISL_2691635, EPI_ISL_2691636, EPI_ISL_2691638, EPI_ISL_2691639, EPI_ISL_2691640, EPI_ISL_2691642, EPI_ISL_2691643, EPI_ISL_2691644, EPI_ISL_2691647, EPI_ISL_2691648, EPI_ISL_2691649, EPI_ISL_2691650, EPI_ISL_2691651, EPI_ISL_2691652, EPI_ISL_2691653, EPI_ISL_2691655, EPI_ISL_2691657, EPI_ISL_2691658, EPI_ISL_2691659, EPI_ISL_2691660, EPI_ISL_2691661, EPI_ISL_2691662, EPI_ISL_2691664, EPI_ISL_2691665, EPI_ISL_2691666, EPI_ISL_2691667, EPI_ISL_2691668, EPI_ISL_2691669, EPI_ISL_2691670, EPI_ISL_2691671, EPI_ISL_2691672, EPI_ISL_2691673, EPI_ISL_2691674, EPI_ISL_2691677, EPI_ISL_2691678, EPI_ISL_2691679, EPI_ISL_2691680, EPI_ISL_2691681, EPI_ISL_2691682, EPI_ISL_2691683, EPI_ISL_2691684, EPI_ISL_2837208, EPI_ISL_2837209, EPI_ISL_2837210, EPI_ISL_2837211, EPI_ISL_2837213, EPI_ISL_2837214, EPI_ISL_2837215, EPI_ISL_2837216, EPI_ISL_2837217, EPI_ISL_2837218, EPI_ISL_2837219, EPI_ISL_2837221, EPI_ISL_2837222, EPI_ISL_2837223, EPI_ISL_2837224, EPI_ISL_2837225, EPI_ISL_2837226, EPI_ISL_2837227, EPI_ISL_2837228, EPI_ISL_2837229, EPI_ISL_2837231, EPI_ISL_2837232, EPI_ISL_2837233, EPI_ISL_2837234, EPI_ISL_2837235, EPI_ISL_2837236, EPI_ISL_2837238, EPI_ISL_2837239, EPI_ISL_2837240, EPI_ISL_2837241, EPI_ISL_2837242, EPI_ISL_2837244, EPI_ISL_2837245, EPI_ISL_2837249, EPI_ISL_2837250, EPI_ISL_2837252, EPI_ISL_2837253, EPI_ISL_2837254, EPI_ISL_2837255, EPI_ISL_2837256, EPI_ISL_2837257, EPI_ISL_2837258, EPI_ISL_2837259, EPI_ISL_2837260, EPI_ISL_2837261, EPI_ISL_2837262, EPI_ISL_2837264, EPI_ISL_2837265, EPI_ISL_2837267, EPI_ISL_2837268, EPI_ISL_2837269, EPI_ISL_2837270, EPI_ISL_2837272, EPI_ISL_2837273, EPI_ISL_2837274, EPI_ISL_2837275, EPI_ISL_2837276, EPI_ISL_2837277, EPI_ISL_2837279, EPI_ISL_2837280, EPI_ISL_2837282, EPI_ISL_2837283, EPI_ISL_2837284, EPI_ISL_2837285, EPI_ISL_2837287, EPI_ISL_2837288, EPI_ISL_2837289, EPI_ISL_2837290, EPI_ISL_2837291, EPI_ISL_2837292, EPI_ISL_2837295, EPI_ISL_2837298, EPI_ISL_2837300, EPI_ISL_2837301, EPI_ISL_2837302, EPI_ISL_2837303, EPI_ISL_2837304, EPI_ISL_2837305, EPI_ISL_2837306, EPI_ISL_2837307, EPI_ISL_2837308, EPI_ISL_2837309, EPI_ISL_2837310, EPI_ISL_2837312, EPI_ISL_2837313, EPI_ISL_2837314, EPI_ISL_3072149, EPI_ISL_3072151, EPI_ISL_3072152, EPI_ISL_3072153, EPI_ISL_3072155, EPI_ISL_3072158, EPI_ISL_3072159, EPI_ISL_3072160, EPI_ISL_3072161, EPI_ISL_3072163, EPI_ISL_3072166, EPI_ISL_3072167, EPI_ISL_3072168, EPI_ISL_3072170, EPI_ISL_3072171, EPI_ISL_3072172, EPI_ISL_3072173, EPI_ISL_3072174, EPI_ISL_3072177, EPI_ISL_3072178, EPI_ISL_3072179, EPI_ISL_3072180, EPI_ISL_3072181, EPI_ISL_3072185, EPI_ISL_3072186, EPI_ISL_3072187, EPI_ISL_3072188, EPI_ISL_3072189, EPI_ISL_3072191, EPI_ISL_3072192, EPI_ISL_3072193, EPI_ISL_3072194, EPI_ISL_3072196, EPI_ISL_3072198, EPI_ISL_3072199, EPI_ISL_3072200, EPI_ISL_3072202, EPI_ISL_3072203, EPI_ISL_3072204, EPI_ISL_3072205, EPI_ISL_3072206, EPI_ISL_3072207, EPI_ISL_3072208, EPI_ISL_3072209, EPI_ISL_3072210, EPI_ISL_3072214, EPI_ISL_3072221, EPI_ISL_3072223, EPI_ISL_3072224, EPI_ISL_3072225, EPI_ISL_3072226, EPI_ISL_3072227, EPI_ISL_3072228, EPI_ISL_3072229, EPI_ISL_3072230, EPI_ISL_3072232, EPI_ISL_3072233, EPI_ISL_3072234, EPI_ISL_3072235, EPI_ISL_3072236, EPI_ISL_3072237, EPI_ISL_3072238, EPI_ISL_3072239, EPI_ISL_3072240, EPI_ISL_3072241, EPI_ISL_3072242, EPI_ISL_3072244, EPI_ISL_3072245, EPI_ISL_3072246, EPI_ISL_3072247, EPI_ISL_3072248, EPI_ISL_3072249, EPI_ISL_3072251, EPI_ISL_3072253, EPI_ISL_3072254, EPI_ISL_3072255, EPI_ISL_3072257, EPI_ISL_3072258, EPI_ISL_3072261, EPI_ISL_3072263, EPI_ISL_3072265, EPI_ISL_3072266, EPI_ISL_3072267, EPI_ISL_3072268, EPI_ISL_3072270, EPI_ISL_3072271, EPI_ISL_3072272, EPI_ISL_3072273, EPI_ISL_3072274, EPI_ISL_3072276, EPI_ISL_3072277, EPI_ISL_3072278, EPI_ISL_3072281, EPI_ISL_3072282, EPI_ISL_3072283, EPI_ISL_3072285, EPI_ISL_3072286, EPI_ISL_3072288, EPI_ISL_3072289, EPI_ISL_3072290, EPI_ISL_3072292, EPI_ISL_3072294, EPI_ISL_3072297, EPI_ISL_3072299, EPI_ISL_3072300, EPI_ISL_3072302, EPI_ISL_3072306, EPI_ISL_3072308, EPI_ISL_3072309, EPI_ISL_3072311, EPI_ISL_3072312, EPI_ISL_3072313, EPI_ISL_3072314, EPI_ISL_3072315, EPI_ISL_3072316, EPI_ISL_3072317, EPI_ISL_3072318, EPI_ISL_3072319, EPI_ISL_3072320, EPI_ISL_3072322, EPI_ISL_3072323, EPI_ISL_3072324, EPI_ISL_3072327, EPI_ISL_3072329, EPI_ISL_3072331, EPI_ISL_3072333, EPI_ISL_3072335, EPI_ISL_3072337, EPI_ISL_3072341, EPI_ISL_3072343, EPI_ISL_3072347, EPI_ISL_3072359, EPI_ISL_3072366, EPI_ISL_3072370, EPI_ISL_3072374, EPI_ISL_3072378, EPI_ISL_3072394, EPI_ISL_3072396, EPI_ISL_3072399, EPI_ISL_3072401, EPI_ISL_3072403, EPI_ISL_3072409, EPI_ISL_3072411, EPI_ISL_3072415, EPI_ISL_3072417, EPI_ISL_3072423, EPI_ISL_3072425, EPI_ISL_3072427, EPI_ISL_3072431, EPI_ISL_3072434, EPI_ISL_3072436, EPI_ISL_3072438, EPI_ISL_3072440, EPI_ISL_3072444, EPI_ISL_3072448, EPI_ISL_3072450, EPI_ISL_3072452, EPI_ISL_3072454, EPI_ISL_3072456, EPI_ISL_3072458, EPI_ISL_3072462, EPI_ISL_3072463, EPI_ISL_3072465, EPI_ISL_3072467, EPI_ISL_3072469, EPI_ISL_3072475, EPI_ISL_3072477, EPI_ISL_3072479, EPI_ISL_3072481, EPI_ISL_3072483, EPI_ISL_3072485, EPI_ISL_3072487, EPI_ISL_3072491, EPI_ISL_3072494, EPI_ISL_3072497, EPI_ISL_3072501, EPI_ISL_3072507, EPI_ISL_3072509, EPI_ISL_3072513, EPI_ISL_3072519, EPI_ISL_3072521, EPI_ISL_3072525, EPI_ISL_3072531, EPI_ISL_3072535, EPI_ISL_3072543, EPI_ISL_3072547, EPI_ISL_3072551, EPI_ISL_3072555, EPI_ISL_3072559, EPI_ISL_3072563, EPI_ISL_3072565, EPI_ISL_3072571, EPI_ISL_3072575, EPI_ISL_3072577, EPI_ISL_3072579, EPI_ISL_3072581, EPI_ISL_3072584, EPI_ISL_3072586, EPI_ISL_3072590, EPI_ISL_3072592, EPI_ISL_3072595, EPI_ISL_3072597, EPI_ISL_3072598, EPI_ISL_3072604, EPI_ISL_3072606, EPI_ISL_3072612, EPI_ISL_3072614, EPI_ISL_3072622, EPI_ISL_3072626, EPI_ISL_3245362, EPI_ISL_3245383, EPI_ISL_3245395, EPI_ISL_3245396, EPI_ISL_3245409, EPI_ISL_3245411 | | | |
| see above | Unidade de apoio ao diagnostico da COVID - UNADIG | Bioinformatics Laboratory / LNCC | Alessandra P Lamarca; Alexandra L Gerber; Amilcar Tanuri; Ana Paula de C Guimaraes; Ana Tereza R Vasconcelos; Andrea Cony Cavalcanti; Caio Luiz Pereira Ribeiro; Cassia Alves; Cintia Policarpo; Claudia Maria Braga de Mello; Cristiane Gomes da Silva; Diana Mariani; Douglas Terra Machado; Erica Ramos dos Santos Nascimento; Fernanda Leitao dos Santos; Flavio Dias da Silva; Gleidson da Silva de Oliveira; Leandro Magalhaes de Souza; Liliane Cavalcante; Luiz G P de Almeida; Marcio Henrique de Oliveira Garcia; Mario Sergio Ribeiro; Ricardo Jose Barbosa Salviano; Ronaldo da Silva F Jr; Silvia Carvalho |
| EPI_ISL_2544253 | Unilabs | Institute of Medical Virology | Alexandra Trkola; Annette Audigé; Cyril Shah; Gabriela Ziltener; Guido Bloemberg; Jon Huder; Jürg Böni; Kevin Steiner; Maria Grünberg; Maryam Zaheri; Michael Huber; Riccarda Capaul; Stefan Schmutz; Verena Kufner |
| EPI_ISL_2840091, EPI_ISL_2840156, EPI_ISL_2897535, EPI_ISL_2899164, EPI_ISL_2899544 | Unilabs | Karolinska University Hospital | Annelie Bjerkner; Isak Sylvin; Jan Albert; Karolina Ininbergs; Lina Guerra Blomqvist; Lynda Eneh; Martin Ekman; Martina Wahlund; Robert Dyrdak; Sandra Broddesson; Tanja Normark; Tobias Allander; Valtteri Wirta; Zhibing Yun |
| EPI_ISL_2776218 | Universidad Central del Caribe (UCC) | Grubaugh Lab - Yale School of Public Health | Anderson Brito; Andrea Firpo; Annie Watkins; Chaney Kalinich; Chantal Vogels; David Rivera Aponte; Ernesto Vazquez; Fabiola Cruz-Lopez; Fabiola Fontanet; Isabel Ott; Jessica Rothman; Joseph Fauver; Kendall Billig; Mallery Breban; Mary Petrone; Natalie Machargo; Nathan Grubaugh; Patricia Serrano; Tania Mitwalli; Tara Alpert; Tobias Koch; Vianca Aponte |
| EPI_ISL_3477567 | Universidad del Valle, LDAB-Laboratorio de Diagnostico de Agentes Biologicos | Universidad del Valle, TAO-Lab, VIREM, NEAS Network | Andres Castillo; Beatriz Parra & COVID-19 Team Univalle; Diana López-Alvarez; Nelson Rivera Franco |
| EPI_ISL_3076939 | University Clinical Hospital of Mostar, Department of Microbiology and Molecular Diagnostics | University of Sarajevo, Veterinary Faculty, Laboratory for Molecular Diagnostic and Research Laboratory | Goletic S; Goletic T; Hodzic A.; Jazic A.; Nicevic M.; Ostojic M; Sabic E.; Softic A; Terzic I. |
| EPI_ISL_2405336, EPI_ISL_2681457 | University Hospitals of Geneva, Laboratory of Virology | HUG, Laboratory of Virology and the Health2030 Genome Center | Ana Rita Goncalves; Deborah Penet; Emmanouil Dermitzakis; Henri Pegeot; Ioannis Xenarios; Keith Harshman; Laurent Kaiser; Lorenzo Cerutti; Melyssa Elies; Samuel Cordey |
| EPI_ISL_2692019, EPI_ISL_2692022, EPI_ISL_2692023, EPI_ISL_3388801 | University of Chicago Medicine | RIPHL at Rush University Medical Center | Edith Perez; Felix Araujo Perez; Kevin Kunstman; Laura Furtado; Lauren Megger; Marieta Hyde; Max Kolton; Stefan Green |
| EPI_ISL_2692112, EPI_ISL_2692121, EPI_ISL_3388712 | University of Illinois at Chicago | RIPHL at Rush University Medical Center | Edith Perez; Felix Araujo Perez; Iuliana Bentea; Julie Bentea; Kevin Kunstman; Laura Furtado; Marieta Hyde; Max Kolton; Omar Perez; Stefan Green |
| EPI_ISL_3537377, EPI_ISL_3537381 | University of Iowa Hospitals and Clinics | State Hygienic Laboratory at the University of Iowa | Alankar Kampoowale; Anna Yakos; Erik Twait; Kris Eveland; Mariah Knutson; Valerie Reeb; Wes Hottel |
| EPI_ISL_2690251 | University of Mississippi Medical Center, Department of Pathology | University of Mississippi Medical Center, Molecular and Genomics Core Facility | Ashley C. Johnson; D. Ashley Robinson; Ithiel J. Frame; Krishna K. Ayyalasomayajula; Michael R. Garrett; Wenjie Wu |
| EPI_ISL_2650508, EPI_ISL_2650510, EPI_ISL_2650512, EPI_ISL_2650515 | University of Oregon COVID-19 MAP Laboratory | University of Oregon Genomics and Cell Characterization Core Facility (GC3F) | Ariana White; Douglas Turnbull; Jason Carriere; Jason Sydes; Jeff Bishop; Peter Batzel |

| | | | |
|---|---|---|---|
| EPI_ISL_2484682, EPI_ISL_2918786 | UniversitätsSpital Zürich | Institute of Medical Virology | Alexandra Trkola; Annette Audigé; Catharine Aquino; Cyril Shah; Daniel Ehrsam; Gabriela Ziltener; Guido Bloemberg; Hubert Rehrauer; Isabel Stürmer; Joel Wirz; Jon Huder; Jürg Böni; Kevin Steiner; Maria Grünberg; Maryam Zaheri; Michael Huber; Riccarda Capaul; Stefan Schmutz; Verena Kufner; Weihong Qi |
| EPI_ISL_2544269 | UniversitätsSpital Zürich 009 | Institute of Medical Virology | Alexandra Trkola; Annette Audigé; Cyril Shah; Gabriela Ziltener; Guido Bloemberg; Jon Huder; Jürg Böni; Kevin Steiner; Maria Grünberg; Maryam Zaheri; Michael Huber; Riccarda Capaul; Stefan Schmutz; Verena Kufner |
| EPI_ISL_3506141 | Usansolo-Galdakao University Hospital | Cruces University Hospital | Ana Belén de la Hz; Ana Gual-de-Torrella; Izaskun Alejo-Cancho; Mikel Gallego |
| EPI_ISL_2678949, EPI_ISL_2678954, EPI_ISL_2678965, EPI_ISL_2679234, EPI_ISL_2679344, EPI_ISL_2856304, EPI_ISL_2856315, EPI_ISL_2856371, EPI_ISL_2856376, EPI_ISL_2856512, EPI_ISL_2856556, EPI_ISL_2856563, EPI_ISL_2856580, EPI_ISL_2856711, EPI_ISL_2856721, EPI_ISL_2856739, EPI_ISL_2856811, EPI_ISL_2856814, EPI_ISL_2856853, EPI_ISL_2856854, EPI_ISL_2856888, EPI_ISL_2856909, EPI_ISL_2856931, EPI_ISL_2856966, EPI_ISL_2857044, EPI_ISL_2857077, EPI_ISL_2857164, EPI_ISL_2857232, EPI_ISL_2857257, EPI_ISL_2857336, EPI_ISL_2857347, EPI_ISL_2857582, EPI_ISL_2857585, EPI_ISL_2857608, EPI_ISL_2857704, EPI_ISL_2857706, EPI_ISL_2857746, EPI_ISL_2857747, EPI_ISL_2857767, EPI_ISL_3132731, EPI_ISL_3132736, EPI_ISL_3132778, EPI_ISL_3132958, EPI_ISL_3132965, EPI_ISL_3133017, EPI_ISL_3144682, EPI_ISL_3268526, EPI_ISL_3268532, EPI_ISL_3268537, EPI_ISL_3268559, EPI_ISL_3268601, EPI_ISL_3268609, EPI_ISL_3268615, EPI_ISL_3268701, EPI_ISL_3268786, EPI_ISL_3268835, EPI_ISL_3268861, EPI_ISL_3268907, EPI_ISL_3268916, EPI_ISL_3268945, EPI_ISL_3268952, EPI_ISL_3269897, EPI_ISL_3745387, EPI_ISL_3745388, EPI_ISL_3745390, EPI_ISL_3745392, EPI_ISL_3745393, EPI_ISL_3745397, EPI_ISL_3745398, EPI_ISL_3745400, EPI_ISL_3756245, EPI_ISL_3756246, EPI_ISL_3756247, EPI_ISL_3756249, EPI_ISL_3756250, EPI_ISL_3756251, EPI_ISL_3756252, EPI_ISL_3756253, EPI_ISL_3756254, EPI_ISL_3756255, EPI_ISL_3756256, EPI_ISL_3756257, EPI_ISL_3756258, EPI_ISL_3756259, EPI_ISL_3756260 | | | |
| see above | Utah Public Health Laboratory | Utah Public Health Laboratory | Erin L. Young; Kelly F. Oakeson; Olinto Linares-Perdomo; Pooja Gupta; Tara Gallagher |
| EPI_ISL_3355143, EPI_ISL_3355159, EPI_ISL_3355209 | VA-Division of Consolidated Laboratory Services | Centers for Disease Control and Prevention Division of Viral Diseases, Pathogen Discovery | Alex Burgin; Ben Rambo-Martin; Clinton Paden; Dakota Howard; Dave Wentworth; Dhwani Batra; Jasmine Padilla; Justin Lee; Krista Queen; Kristen Knipe; Kristine Lacek; Mark Burroughs; Matthew Schmerer; Meghan Bentz; Mili Sheth; Peter Cook; Sam Shepard; Sarah Nobles; Suxiang Tong; Vivien Dugan; Yvette Unoarumhi |
| EPI_ISL_2462316, EPI_ISL_2493841 | VIGILANCIA EM SAUDE DE FCO MORATO | Instituto Butantan | Antonio Jorge Martins; Claudia Renata dos Santos Barros; David Schlesinger; Debora Botequio Moretti; Dimas Tadeu Covas; Elaine Cristina Marqueze; Elaine Vieira Santos; Evandra Strazza Rodrigues; Heidge Fukumasu; Jayme Augusto de Souza-Neto; José Salvatore Leister Patané; Luiz Alcantara; Luiz Lehmann Coutinho; Maria Carolina Elias; Maurício Lacerda Nogueira; Rafael dos Santos Bezerra; Raul Machado Neto; Rejane Maria Tommasini Grotto; Ricardo Haddad; Sandra Coccuzzo Sampaio Vessoni; Simone Kashima; Svetoslav Nanev Slavov; Vincent Louis Viala |
| EPI_ISL_2494111, EPI_ISL_2494112, EPI_ISL_2494113 | VIGILANCIA EM SAUDE SAE VIGILANCIA EPIDEMIOLOGICA | Instituto Butantan | Antonio Jorge Martins; Claudia Renata dos Santos Barros; David Schlesinger; Debora Botequio Moretti; Dimas Tadeu Covas; Elaine Cristina Marqueze; Elaine Vieira Santos; Evandra Strazza Rodrigues; Heidge Fukumasu; Jayme Augusto de Souza-Neto; José Salvatore Leister Patané; Luiz Alcantara; Luiz Lehmann Coutinho; Maria Carolina Elias; Maurício Lacerda Nogueira; Rafael dos Santos Bezerra; Raul Machado Neto; Rejane Maria Tommasini Grotto; Ricardo Haddad; Sandra Coccuzzo Sampaio Vessoni; Simone Kashima; Svetoslav Nanev Slavov; Vincent Louis Viala |
| EPI_ISL_2493916 | VIGILANCIA EPIDEMIOLOGICA DE FERRAZ DE VASCONCELOS | Instituto Butantan | Antonio Jorge Martins; Claudia Renata dos Santos Barros; David Schlesinger; Debora Botequio Moretti; Dimas Tadeu Covas; Elaine Cristina Marqueze; Elaine Vieira Santos; Evandra Strazza Rodrigues; Heidge Fukumasu; Jayme Augusto de Souza-Neto; José Salvatore Leister Patané; Luiz Alcantara; Luiz Lehmann Coutinho; Maria Carolina Elias; Maurício Lacerda Nogueira; Rafael dos Santos Bezerra; Raul Machado Neto; Rejane Maria Tommasini Grotto; Ricardo Haddad; Sandra Coccuzzo Sampaio Vessoni; Simone Kashima; Svetoslav Nanev Slavov; Vincent Louis Viala |
| EPI_ISL_2494162, EPI_ISL_2494173, EPI_ISL_2494336 | VIGILANCIA EPIDEMIOLOGICA DE LEME | Instituto Butantan | Antonio Jorge Martins; Claudia Renata dos Santos Barros; David Schlesinger; Debora Botequio Moretti; Dimas Tadeu Covas; Elaine Cristina Marqueze; Elaine Vieira Santos; Evandra Strazza Rodrigues; Heidge Fukumasu; Jayme Augusto de Souza-Neto; José Salvatore Leister Patané; Luiz Alcantara; Luiz Lehmann Coutinho; Maria Carolina Elias; Maurício Lacerda Nogueira; Rafael dos Santos Bezerra; Raul Machado Neto; Rejane Maria Tommasini Grotto; Ricardo Haddad; Sandra Coccuzzo Sampaio Vessoni; Simone Kashima; Svetoslav Nanev Slavov; Vincent Louis Viala |
| EPI_ISL_2494171, EPI_ISL_2494191, EPI_ISL_2494197, EPI_ISL_2494198 | VIGILANCIA EPIDEMIOLOGICA E CONTROLE DE VETORES PIRASSUNUN | Instituto Butantan | Antonio Jorge Martins; Claudia Renata dos Santos Barros; David Schlesinger; Debora Botequio Moretti; Dimas Tadeu Covas; Elaine Cristina Marqueze; Elaine Vieira Santos; Evandra Strazza Rodrigues; Heidge Fukumasu; Jayme Augusto de Souza-Neto; José Salvatore Leister Patané; Luiz Alcantara; Luiz Lehmann Coutinho; Maria Carolina Elias; Maurício Lacerda Nogueira; Rafael dos Santos Bezerra; Raul Machado Neto; Rejane Maria Tommasini Grotto; Ricardo Haddad; Sandra Coccuzzo Sampaio Vessoni; Simone Kashima; Svetoslav Nanev Slavov; Vincent Louis Viala |
| EPI_ISL_2493944, EPI_ISL_2493945, EPI_ISL_2493949, EPI_ISL_2493953, EPI_ISL_2493958 | VIGILANCIA SANITARIA E VIG EPIDEMIOLOGICA DE ITAPEVI | Instituto Butantan | Antonio Jorge Martins; Claudia Renata dos Santos Barros; David Schlesinger; Debora Botequio Moretti; Dimas Tadeu Covas; Elaine Cristina Marqueze; Elaine Vieira Santos; Evandra Strazza Rodrigues; Heidge Fukumasu; Jayme Augusto de Souza-Neto; José Salvatore Leister Patané; Luiz Alcantara; Luiz Lehmann Coutinho; Maria Carolina Elias; Maurício Lacerda Nogueira; Rafael dos Santos Bezerra; Raul Machado Neto; Rejane Maria Tommasini Grotto; Ricardo Haddad; Sandra Coccuzzo Sampaio Vessoni; Simone Kashima; Svetoslav Nanev Slavov; Vincent Louis Viala |
| EPI_ISL_2544492, EPI_ISL_2662372, EPI_ISL_2662403, EPI_ISL_2662445, EPI_ISL_2662446, EPI_ISL_2724466, EPI_ISL_2938392, EPI_ISL_2938458, EPI_ISL_3007545, EPI_ISL_3007610, EPI_ISL_3007621 | | | |
| see above | Viollier AG | Department of Biosystems Science and Engineering, ETH Zürich | Andrea Patrignani; Andrea Patrizia Salzmann; Andreia Cabral de Gouvea; Catharine Aquino; Chaoran Chen; Christian Beisel; Christiane Beckmann; Christoph Noppen; Daniel Ehrsam; Doris Popovic; Elodie Burcklen; Griffin White; Henriette Kurth; Ina Nissen; Isabel Stürmer; Ivan Topolsky; Jay Tracy; Kim Philipp Jablonski; Lara Fuhrmann; Laura Neff; Lennart Opitz; Maria Domenica Moccia; Maurice Redondo; Mirjam Feldkamp; Natascha Santacroce; Niko Beerenwinkel; Olivier Kobel; Ralph Schlapbach; Rebecca Denes; Sarah Nadeau; Simon Grüter; Tanja Stadler; Timothy Sykes |
| EPI_ISL_2420698, EPI_ISL_2420707 | Viollier AG | Viollier AG | Andrea Patrizia Salzmann; Christiane Beckmann; Christoph Noppen; Henriette Kurth; Maurice Redondo; Olivier Kobel |
| EPI_ISL_2603346, EPI_ISL_2603359, EPI_ISL_2689853, EPI_ISL_2689854, EPI_ISL_2776029, EPI_ISL_2776030, EPI_ISL_2776043, EPI_ISL_2776050, EPI_ISL_2776054, EPI_ISL_2920793, EPI_ISL_2982646, EPI_ISL_2982657, EPI_ISL_3025879, EPI_ISL_3048097, EPI_ISL_3123430 | | | |
| see above | Virginia Division of Consolidated Laboratory Services | Virginia Division of Consolidated Laboratory Services | Virginia DCLS; Virginia Division of Consolidated Laboratory Services |
| EPI_ISL_3230510 | Vita Laboratoriot Oy | Expert Microbiology, National Institute for Health and Welfare | Carita Savolainen-Kopra; Erika Lindh; Haider al-Hello; Jani Halkilahti; Kirsi Liitsola; Niina Ikonen; Olli Vapalahti; Pekka Ellonen; Phuoc Truong; Päivi Laurila; Ravi Kant; Sari Hannula; Soile Blomqvist; Teemu Smura |
| EPI_ISL_2690098, EPI_ISL_2690108, EPI_ISL_2690110, EPI_ISL_2788021, EPI_ISL_2788026 | WA State Department of Health | Centers for Disease Control and Prevention Division of Viral Diseases, Pathogen Discovery | Alison Laufer Halpin; Ben L. Rambo-Martin; Clinton R. Paden; Dakota Howard; Darlene Wagner; Dave Wentworth; Dhwani Batra; Jasmine Padilla; Justin Lee; Katie Dillon; Krista Queen; Kristen Knipe; Kristine Lacek; Mark Burroughs; Matthew Schmerer; Mili Sheth; Peter Cook; Sam Shepard; Sarah Nobles; Shoshona Le; Suxiang Tong; Vivien Dugan; Yvette Unoarumhi |
| EPI_ISL_3031272, EPI_ISL_3031273, EPI_ISL_3031274 | WESTCHESTER MEDICAL CENTER | Wadsworth Center, New York State Department of Health | Alexis Russell; Catharine Prussing; Daryl M. Lamson; Erasmus Schneider; Erica Lasek-Nesselquist; John Kelly; Jonathan Plitnick; Kirsten St. George; Matthew Shudt; Melissa A Leisner; Navjot Singh |
| EPI_ISL_2787585 | WI State Laboratory of Hygiene | Centers for Disease Control and Prevention Division of Viral Diseases, Pathogen Discovery | Alison Laufer Halpin; Ben L. Rambo-Martin; Clinton R. Paden; Dakota Howard; Darlene Wagner; Dave Wentworth; Dhwani Batra; Jasmine Padilla; Justin Lee; Katie Dillon; Krista Queen; Kristen Knipe; Kristine Lacek; Mark Burroughs; Matthew Schmerer; Mili Sheth; Peter Cook; Sam Shepard; Sarah Nobles; Shoshona Le; Suxiang Tong; Vivien Dugan; Yvette Unoarumhi |
| EPI_ISL_2597899 | WSSE w Gdańsku | 1. Tricity SARS-CoV-2 sequencing consortium: University of Gdansk, Medical University of Gdansk, Vaxican Ltd., Invicta Ltd. 2. National Institute of Public Health - National Institute of Hygiene, Warsaw, Poland | Celina Cybulska; Karolina Gackowska; Katarzyna Groth; Katarzyna Zacharczuk; Krystyna Bienkowska Szewczyk; Lukasz Rabalski; Maciej Grzybek; Maciej Kosinski; Magdalena Nowakowska; Marcin Lubocki; Małgorzata Sadkowska-Todys; Tomasz Wołkowicz |
| EPI_ISL_3009183 | WSSE w Katowicach | 1. Academic Center for Pathomorphological and Genetic-Molecular Diagnostics ltd, Bialystok, Poland 2. National Institute of Public Health - National Institute of Hygiene, Warsaw, Poland | Anetta Sulewska; Jacek Nikliński; Janusz Dzięcioł; Joanna Kiśluk; Katarzyna Zacharczuk; Konrad Raczkowski; Magdalena Nowakowska; Małgorzata Sadkowska-Todys; Piotr Karabowicz; Piotr Majewski; Przemysław Biecek. Joanna Reszeć; Radosław Charkiewicz; Tomasz Wołkowicz |
| EPI_ISL_2557057, EPI_ISL_2557063, EPI_ISL_2557066, EPI_ISL_2557067, EPI_ISL_2557071, EPI_ISL_2557075, EPI_ISL_2557079, EPI_ISL_2651042, EPI_ISL_2651054, EPI_ISL_2651062, EPI_ISL_2651064, EPI_ISL_2651077, EPI_ISL_2651083, EPI_ISL_2651084, EPI_ISL_2651086, EPI_ISL_2651090, EPI_ISL_2651097, EPI_ISL_2651105, EPI_ISL_2716463, EPI_ISL_2716464, EPI_ISL_2716466, EPI_ISL_2716469, EPI_ISL_2716470, EPI_ISL_2759880, EPI_ISL_2834708, EPI_ISL_3016356, EPI_ISL_3016360, EPI_ISL_3016387, EPI_ISL_3216979 | | | |
| see above | Washington State Department of Health Public Health Laboratories | Washington State Department of Health Public Health Laboratories | Avi Singh; Darren Lucas; Denny Russell; Drew MacKellar; Geoff Melly; Hannah Gray; Joenice Gonzalez; JohnAric Peterson; Philip Dykema; Rebecca Cao; Vanessa De Los Santos |
| EPI_ISL_2774524, EPI_ISL_3235364, EPI_ISL_3235365, EPI_ISL_3246657 | Weill Cornell Medicine | New York Genome Center | Amy Baldwin; Andre Corvelo; Arryn Craney; Chris Mason; Dayna M. Oschwald; Hanna Rennert; Lars F Westblade; Margaret Elizabeth Ross; Melissa Cushing; Michael Zody; Olivier Elemento; Priya Velu; Samantha Fennessey; Tom Maniatis |
| EPI_ISL_2876102 | Wexner Medical Center | The Ohio State University College of Medicine | Koenig, S.; Seminetta, J. |
| EPI_ISL_2400234, EPI_ISL_2499942, EPI_ISL_2534256, EPI_ISL_2612191, EPI_ISL_2612195, EPI_ISL_2627938, EPI_ISL_2627939, EPI_ISL_2675197, EPI_ISL_2677640, EPI_ISL_2756526, EPI_ISL_2756532 | | | |
| see above | Wichita State University - Molecular Diagnostics Lab | Kansas Health and Environmental Lab | Ben Olsen; Jonathan Barnell; Katherine Wiggins; Mike Grose; and Phil Adam |
| EPI_ISL_2833947 | Wichita State University- Molecular Diagnostic Lab | Kansas Health and Environmental Lab | Ben Olsen; Jonathan Barnell; Katherine Wiggins; Mike Grose; and Phil Adam |
| EPI_ISL_2382562, EPI_ISL_2382563, EPI_ISL_2383017, EPI_ISL_2383018 | Wichita State University- Molecular Diagonstic lab | Kansas Health and Environmental Lab | Ben Olsen; Jonathan Barnell; Katherine Wiggins; Mike Grose; and Phil Adam |
| EPI_ISL_2383180, EPI_ISL_2561917, EPI_ISL_2561926 | Willis-Knighton Medical Center Hospital Laboratory | LSUHS Emerging Viral Threat Laboratory | Alexander Mijalis; Andrew D. Yurochko; April N. Johnson; Christopher G. Kevil; Gregory L. Ware; Jennifer L. Carroll; Jeremy P. Kamil; John A. Vanchiere; Joseph A. Bocchini; Lorie M. Atkins; Maarten Van Diest; Rona S. Scott |
| EPI_ISL_2558601, EPI_ISL_2558668, EPI_ISL_2558669, EPI_ISL_2727681, EPI_ISL_2727687, EPI_ISL_2727705, EPI_ISL_2727708, EPI_ISL_2727712, EPI_ISL_2727766, EPI_ISL_2727775, EPI_ISL_2727782, EPI_ISL_2727792, EPI_ISL_2727816, EPI_ISL_2727817, EPI_ISL_2727826, EPI_ISL_3131920, EPI_ISL_3131936, EPI_ISL_3131949, EPI_ISL_3131960, EPI_ISL_3234718, EPI_ISL_3234720 | | | |
| see above | Wisconsin State Laboratory of Hygiene Communicable Disease Division | Wisconsin State Laboratory of Hygiene Communicable Disease Division | Abigail C. Shockey; Alicia J. Mooney; Erika M. Hanson; Kelsey R. Florek; Richard Griesser; Sara Wagner; Tonya Danz |
| EPI_ISL_2712919, EPI_ISL_2712921, EPI_ISL_2726819, EPI_ISL_2754064, EPI_ISL_2801550, EPI_ISL_2833845, EPI_ISL_2834976, EPI_ISL_2960293, EPI_ISL_3038893, EPI_ISL_3062475, EPI_ISL_3160600, EPI_ISL_3160630, EPI_ISL_3207429 | | | |
| see above | Wyoming Public Health Laboratory | Wyoming Public Health Laboratory | Ashley Norberg; Brian Dominguez; Cari Sloma; Channing Weber; Chayse Rowley; Elliot Thomasson; Jim Mildenberger; Marley Goetz; Robert Petit; Sam Britz; Taylor Fearing; and Rob Christensen |

| | | | |
|---|---|---|---|
| EPI_ISL_2716232, EPI_ISL_2716245, EPI_ISL_2860312, EPI_ISL_3025425, EPI_ISL_3104693, EPI_ISL_3104694 | Yale Clinical Virology Lab | Grubaugh Lab - Yale School of Public Health | Anderson Brito; Annie Watkins; Chaney Kalinich; Chantal Vogels; Isabel Ott; Jessica Rothman; Joseph Fauver; Kendall Billig; Mallery Breban; Marie L. Landry; Mary Petrone; Nathan Grubaugh; Tara Alpert; Tobias Koch |

We gratefully acknowledge the following Authors from the Originating laboratories responsible for obtaining the specimens, as well as the Submitting laboratories where the genome data were generated and shared via GISAID, on which this research is based.

All Submitters of data may be contacted directly via www.gisaid.org

Authors are sorted alphabetically.

| Accession ID | Originating Laboratory | Submitting Laboratory | Authors |
|---|---|---|---|
| EPI_ISL_2689871, EPI_ISL_2689873 | "AK State Public Health Lab, State Health Department" | Centers for Disease Control and Prevention Division of Viral Diseases, Pathogen Discovery | Alison Laufer Halpin; Ben L. Rambo-Martin; Clinton R. Paden; Dakota Howard; Darlene Wagner; Dave Wentworth; Dhwani Batra; Jasmine Padilla; Justin Lee; Katie Dillon; Krista Queen; Kristen Knipe; Kristine Lacek; Mark Burroughs; Matthew Schmerer; Mili Sheth; Peter Cook; Sam Shepard; Sarah Nobles; Shoshona Le; Suxiang Tong; Vivien Dugan; Yvette Unoarumhi |
| EPI_ISL_3354922 | "AR Dept. of Health-PHL, Molecular Diagnostics" | Centers for Disease Control and Prevention Division of Viral Diseases, Pathogen Discovery | Alex Burgin; Ben Rambo-Martin; Clinton Paden; Dakota Howard; Dave Wentworth; Dhwani Batra; Jasmine Padilla; Justin Lee; Krista Queen; Kristen Knipe; Kristine Lacek; Mark Burroughs; Matthew Schmerer; Meghan Bentz; Mili Sheth; Peter Cook; Sam Shepard; Sarah Nobles; Suxiang Tong; Vivien Dugan; Yvette Unoarumhi |
| EPI_ISL_2463003 | "TX DSHS, Lab Services Section MC 1947" | Centers for Disease Control and Prevention Division of Viral Diseases, Pathogen Discovery | Alison Laufer Halpin; Ben L. Rambo-Martin; Clinton R. Paden; Dakota Howard; Darlene Wagner; Dave Wentworth; Dhwani Batra; Jasmine Padilla; Justin Lee; Katie Dillon; Krista Queen; Kristen Knipe; Kristine Lacek; Mark Burroughs; Matthew Schmerer; Mili Sheth; Peter Cook; Sam Shepard; Sarah Nobles; Shoshona Le; Suxiang Tong; Vivien Dugan; Yvette Unoarumhi |
| EPI_ISL_2650703 | A.S.L. NOVARA - ISTITUTO S.S. TRINITA' - BORGOMANERO - S.S. LABORATORIO ANALISI CHIMICO CLINICHE - | Fondazione del Piemonte per l'Oncologia IRCCS | Antonino Sottile; Giorgio Giardina; Paola Marino; Silvia Brossa |
| EPI_ISL_3347194, EPI_ISL_3347195, EPI_ISL_3347196, EPI_ISL_3347197, EPI_ISL_3347198, EPI_ISL_3347199, EPI_ISL_3347200, EPI_ISL_3347201, EPI_ISL_3347202 | | | |
| see above | ADILAB | Laboratorio Departamental de Salud Publica de Antioquia | Andres F. Cardona-Rios; Gloria Isabel Escobar; Idabely Betancur Ortiz; Juan P. Hernandez-Ortiz; Maria Stella López |
| EPI_ISL_2493256, EPI_ISL_2493285, EPI_ISL_2493739, EPI_ISL_2493740, EPI_ISL_2493741, EPI_ISL_2493742, EPI_ISL_2493744, EPI_ISL_2493745, EPI_ISL_2493749, EPI_ISL_2493751, EPI_ISL_2493753, EPI_ISL_2493754, EPI_ISL_2493755, EPI_ISL_2493756, EPI_ISL_2493758, EPI_ISL_2493759, EPI_ISL_2493761, EPI_ISL_2493762, EPI_ISL_2494293 | | | |
| see above | AFIP | Instituto Butantan | Antonio Jorge Martins; Claudia Renata dos Santos Barros; David Schlesinger; Debora Botequio Moretti; Dimas Tadeu Covas; Elaine Cristina Marqueze; Elaine Vieira Santos; Evandra Strazza Rodrigues; Heidge Fukumasu; Jayme Augusto de Souza-Neto; José Salvatore Leister Patané; Luiz Alcantara; Luiz Lehmann Coutinho; Maria Carolina Elias; Maurício Lacerda Nogueira; Rafael dos Santos Bezerra; Raul Machado Neto; Rejane Maria Tommasini Grotto; Ricardo Haddad; Sandra Coccuzzo Sampaio Vessoni; Simone Kashima; Svetoslav Nanev Slavov; Vincent Louis Viala |
| EPI_ISL_2229280, EPI_ISL_2229281, EPI_ISL_2229282, EPI_ISL_2383983 | AL Dept. of Public Health Bureau of Clinical Laboratories | Centers for Disease Control and Prevention Division of Viral Diseases, Pathogen Discovery | Alison Laufer Halpin; Ben L. Rambo-Martin; Clinton R. Paden; Dakota Howard; Darlene Wagner; Dave Wentworth; Dhwani Batra; Jasmine Padilla; Justin Lee; Katie Dillon; Krista Queen; Kristen Knipe; Kristine Lacek; Mark Burroughs; Matthew Schmerer; Mili Sheth; Peter Cook; Sam Shepard; Sarah Nobles; Shoshona Le; Suxiang Tong; Vivien Dugan; Yvette Unoarumhi |
| EPI_ISL_2098500, EPI_ISL_2279505, EPI_ISL_2627157 | ALBANY MEDICAL CENTER | Wadsworth Center, New York State Department of Health | Alexis Russell; Catharine Prussing; Daryl M. Lamson; Erasmus Schneider; Erica Lasek-Nesselquist; John Kelly; Jonathan Plitnick; Kirsten St. George; Matthew Shudt; Melissa A Leisner; Navjot Singh |
| EPI_ISL_2208874 | AMB MEDICO DR FRANCISCO FELIX DE MENDONCA SAUDADE | Instituto Butantan / ESALQ-Piracicaba | Antonio Jorge Martins; Claudia Renata dos Santos Barros; David Schlesinger; Debora Botequio Moretti; Dimas Tadeu Covas; Elaine Cristina Marqueze; Elaine Vieira Santos; Evandra Strazza Rodrigues; Heidge Fukumasu; Jayme Augusto de Souza-Neto; José Salvatore Leister Patané; Luiz Alcantara; Luiz Lehmann Coutinho; Maria Carolina Elias; Maurício Lacerda Nogueira; Rafael dos Santos Bezerra; Raul Machado Neto; Rejane Maria Tommasini Grotto; Ricardo Haddad; Sandra Coccuzzo Sampaio Vessoni; Simone Kashima; Svetoslav Nanev Slavov; Vincent Louis Viala |
| EPI_ISL_2493553 | AMBULATORIO AVIACAO | Instituto Butantan | Antonio Jorge Martins; Claudia Renata dos Santos Barros; David Schlesinger; Debora Botequio Moretti; Dimas Tadeu Covas; Elaine Cristina Marqueze; Elaine Vieira Santos; Evandra Strazza Rodrigues; Heidge Fukumasu; Jayme Augusto de Souza-Neto; José Salvatore Leister Patané; Luiz Alcantara; Luiz Lehmann Coutinho; Maria Carolina Elias; Maurício Lacerda Nogueira; Rafael dos Santos Bezerra; Raul Machado Neto; Rejane Maria Tommasini Grotto; Ricardo Haddad; Sandra Coccuzzo Sampaio Vessoni; Simone Kashima; Svetoslav Nanev Slavov; Vincent Louis Viala |
| EPI_ISL_2493549, EPI_ISL_2493554 | AMBULATORIO BOQUEIRAO | Instituto Butantan | Antonio Jorge Martins; Claudia Renata dos Santos Barros; David Schlesinger; Debora Botequio Moretti; Dimas Tadeu Covas; Elaine Cristina Marqueze; Elaine Vieira Santos; Evandra Strazza Rodrigues; Heidge Fukumasu; Jayme Augusto de Souza-Neto; José Salvatore Leister Patané; Luiz Alcantara; Luiz Lehmann Coutinho; Maria Carolina Elias; Maurício Lacerda Nogueira; Rafael dos Santos Bezerra; Raul Machado Neto; Rejane Maria Tommasini Grotto; Ricardo Haddad; Sandra Coccuzzo Sampaio Vessoni; Simone Kashima; Svetoslav Nanev Slavov; Vincent Louis Viala |
| EPI_ISL_2493608, EPI_ISL_2493615 | AMBULATORIO COVID 19 | Instituto Butantan | Antonio Jorge Martins; Claudia Renata dos Santos Barros; David Schlesinger; Debora Botequio Moretti; Dimas Tadeu Covas; Elaine Cristina Marqueze; Elaine Vieira Santos; Evandra Strazza Rodrigues; Heidge Fukumasu; Jayme Augusto de Souza-Neto; José Salvatore Leister Patané; Luiz Alcantara; Luiz Lehmann Coutinho; Maria Carolina Elias; Maurício Lacerda Nogueira; Rafael dos Santos Bezerra; Raul Machado Neto; Rejane Maria Tommasini Grotto; Ricardo Haddad; Sandra Coccuzzo Sampaio Vessoni; Simone Kashima; Svetoslav Nanev Slavov; Vincent Louis Viala |
| EPI_ISL_2445420, EPI_ISL_2445422, EPI_ISL_2493255, EPI_ISL_2493529, EPI_ISL_2493530, EPI_ISL_2493532, EPI_ISL_2493533, EPI_ISL_2493534, EPI_ISL_2500980 | | | |
| see above | AMBULATORIO DE ESPECIALIDADE V E MOGI MIRIM | Instituto Butantan | Antonio Jorge Martins; Claudia Renata dos Santos Barros; David Schlesinger; Debora Botequio Moretti; Dimas Tadeu Covas; Elaine Cristina Marqueze; Elaine Vieira Santos; Evandra Strazza Rodrigues; Heidge Fukumasu; Jayme Augusto de Souza-Neto; José Salvatore Leister Patané; Luiz Alcantara; Luiz Lehmann Coutinho; Maria Carolina Elias; Maurício Lacerda Nogueira; Rafael dos Santos Bezerra; Raul Machado Neto; Rejane Maria Tommasini Grotto; Ricardo Haddad; Sandra Coccuzzo Sampaio Vessoni; Simone Kashima; Svetoslav Nanev Slavov; Vincent Louis Viala |
| EPI_ISL_2473762, EPI_ISL_2473763, EPI_ISL_2473770, EPI_ISL_2473772, EPI_ISL_2473774, EPI_ISL_2493364 | AMBULATÓRIO INSTITUTO BUTANTAN | Instituto Butantan | Antonio Jorge Martins; Claudia Renata dos Santos Barros; David Schlesinger; Debora Botequio Moretti; Dimas Tadeu Covas; Elaine Cristina Marqueze; Elaine Vieira Santos; Evandra Strazza Rodrigues; Heidge Fukumasu; Jayme Augusto de Souza-Neto; José Salvatore Leister Patané; Luiz Alcantara; Luiz Lehmann Coutinho; Maria Carolina Elias; Maurício Lacerda Nogueira; Rafael dos Santos Bezerra; Raul Machado Neto; Rejane Maria Tommasini Grotto; Ricardo Haddad; Sandra Coccuzzo Sampaio Vessoni; Simone Kashima; Svetoslav Nanev Slavov; Vincent Louis Viala |
| EPI_ISL_2451779 | AR Dept. of Health-PHL, Molecular Diagnostics | Centers for Disease Control and Prevention Division of Viral Diseases, Pathogen Discovery | Alison Laufer Halpin; Ben L. Rambo-Martin; Clinton R. Paden; Dakota Howard; Darlene Wagner; Dave Wentworth; Dhwani Batra; Jasmine Padilla; Justin Lee; Katie Dillon; Krista Queen; Kristen Knipe; Kristine Lacek; Mark Burroughs; Matthew Schmerer; Mili Sheth; Peter Cook; Sam Shepard; Sarah Nobles; Shoshona Le; Suxiang Tong; Vivien Dugan; Yvette Unoarumhi |
| EPI_ISL_2155825 | AR Dept. of Health-PHL, Molecular Diagnostics | Pathogen Discovery, Respiratory Viruses Branch, Division of Viral Diseases, Centers for Disease Control and Prevention | Adam Retchless; Anna Kelleher; Anna Uehara; Brian Lynch; Clinton R. Paden; Dhwani Batra; Haibin Wang; Han Jia Justin Ng; Jasmine Padilla; Jing Zhang; Justin Lee; Krista Queen; Mark Burroughs; Mili Sheth; Morgan Davis; Peter Cook; Rachel Marine; Sarah Nobles; Suxiang Tong; Tara Coalter; Yan Li; Ying Tao |
| EPI_ISL_2502728 | AREA DE SALUD CARRILLO | Inciensa, Instituto Costarricense de InvestigaciOn y Enseñanza en NutriciOn y Salud | Adriana Godínez; Claudio Soto-Garita; Estela Cordero; Francisco Duarte; Hebleen Porras; Jose Luis Vargas; Joselyn Prado & Ivanna Krize-Morún; Mariela Gutierrez; Melany CalderOn |
| EPI_ISL_2273001 | AREA DE SALUD CORONADO | Inciensa, Instituto Costarricense de Investigación y Enseñanza en Nutrición y Salud | Adriana Godínez; Claudio Soto-Garita; Estela Cordero; Francisco Duarte; Hebleen Porras; Joselyn Prado & Mariel López; José Luis Vargas; Mariela Gutiérrez; Melany Calderón |
| EPI_ISL_2502725 | AREA DE SALUD CURRIDABAT 2 | Inciensa, Instituto Costarricense de InvestigaciOn y Enseñanza en NutriciOn y Salud | Adriana Godínez; Claudio Soto-Garita; Estela Cordero; Francisco Duarte; Hebleen Porras; Jose Luis Vargas; Joselyn Prado & Mariel LOpez; Mariela Gutierrez; Melany CalderOn |
| EPI_ISL_2272983 | AREA DE SALUD GOICOECHEA 2 - CLINICA DR. JIMENEZ NUÑEZ | Inciensa, Instituto Costarricense de Investigación y Enseñanza en Nutrición y Salud | Adriana Godínez; Claudio Soto-Garita; Estela Cordero; Francisco Duarte; Hebleen Porras; Joselyn Prado & Juan Carlos Cartes Parra; José Luis Vargas; Mariela Gutiérrez; Melany Calderón |
| EPI_ISL_2502747 | AREA DE SALUD PARAISO-CERVANTES | Inciensa, Instituto Costarricense de InvestigaciOn y Enseñanza en NutriciOn y Salud | Adriana Godínez; Claudio Soto-Garita; Estela Cordero; Francisco Duarte; Hebleen Porras; Jose Luis Vargas; Mariela Gutierrez & Joselyn Prado; Melany CalderOn |
| EPI_ISL_2273004 | AREA DE SALUD SAN JUAN-SAN DIEGO-CONCEPCION 2 | Inciensa, Instituto Costarricense de Investigación y Enseñanza en Nutrición y Salud | Adriana Godínez; Claudio Soto-Garita; Estela Cordero; Francisco Duarte; Hebleen Porras; Joselyn Prado & Mariel López; José Luis Vargas; Mariela Gutiérrez; Melany Calderón |
| EPI_ISL_2658269 | AREA DE SALUD ZAPOTE-CATEDRAL - CLINICA DR. CARLOS DURAN | Inciensa, Instituto Costarricense de Investigación y Enseñanza en Nutrición y Salud | Adriana Godínez; Claudio Soto-Garita; Estela Cordero; Francisco Duarte; Hebleen Porras; Joselyn Prado & Andony Cordero-Jiménez; José Luis Vargas; Mariela Gutiérrez; Melany Calderón |
| EPI_ISL_1999796, EPI_ISL_1999817, EPI_ISL_1999841, EPI_ISL_1999873, EPI_ISL_2000082, EPI_ISL_2000100, EPI_ISL_2000104, EPI_ISL_2000119, EPI_ISL_2039060, EPI_ISL_2039083, EPI_ISL_2039096, EPI_ISL_2039124, EPI_ISL_2039127, EPI_ISL_2039144, EPI_ISL_2039149, EPI_ISL_2039153, EPI_ISL_2039158, EPI_ISL_2039163, EPI_ISL_2039390, EPI_ISL_2039416, EPI_ISL_2039460, EPI_ISL_2039469, EPI_ISL_2039470, EPI_ISL_2039485, EPI_ISL_2039527, EPI_ISL_2039666, EPI_ISL_2039783, EPI_ISL_2039843, EPI_ISL_2039845, EPI_ISL_2040165, EPI_ISL_2040252, EPI_ISL_2040294, EPI_ISL_2040306, EPI_ISL_2040348, EPI_ISL_2040377, EPI_ISL_2041076, EPI_ISL_2041407, EPI_ISL_2041424, EPI_ISL_2041519, EPI_ISL_2041681, EPI_ISL_2041807, EPI_ISL_2041816, EPI_ISL_2041845, EPI_ISL_2041892, EPI_ISL_2041893, EPI_ISL_2041936, EPI_ISL_2041942, EPI_ISL_2042059, EPI_ISL_2042103, EPI_ISL_2042228, EPI_ISL_2042229, EPI_ISL_2042248, EPI_ISL_2042494, EPI_ISL_2042505, EPI_ISL_2042539, EPI_ISL_2042605, EPI_ISL_2042611, EPI_ISL_2042629, EPI_ISL_2042646, EPI_ISL_2042650, EPI_ISL_2042675, EPI_ISL_2042721, EPI_ISL_2042729, EPI_ISL_2042745, EPI_ISL_2042813, EPI_ISL_2042844, EPI_ISL_2042853, EPI_ISL_2042896, EPI_ISL_2042900, EPI_ISL_2042917, EPI_ISL_2042980, EPI_ISL_2043034, EPI_ISL_2043041, EPI_ISL_2043116, EPI_ISL_2043172, EPI_ISL_2043175, EPI_ISL_2043178, EPI_ISL_2043196, EPI_ISL_2043197, EPI_ISL_2043238, EPI_ISL_2043249, EPI_ISL_2043259, EPI_ISL_2043318, EPI_ISL_2043358, EPI_ISL_2043361, EPI_ISL_2043365, EPI_ISL_2043430, EPI_ISL_2043447, EPI_ISL_2043466, EPI_ISL_2043489, EPI_ISL_2043503, EPI_ISL_2043505, EPI_ISL_2043511, EPI_ISL_2043517, EPI_ISL_2043523, EPI_ISL_2043527, EPI_ISL_2043545, EPI_ISL_2043549, EPI_ISL_2089479, EPI_ISL_2089506, EPI_ISL_2089512, EPI_ISL_2089591, EPI_ISL_2089605, EPI_ISL_2089608, EPI_ISL_2089609, EPI_ISL_2089626, EPI_ISL_2089660, EPI_ISL_2089703, EPI_ISL_2089707, EPI_ISL_2089842, EPI_ISL_2089849, EPI_ISL_2089857, EPI_ISL_2089866, EPI_ISL_2089921, EPI_ISL_2089943, EPI_ISL_2089953, EPI_ISL_2089959, EPI_ISL_2089992, EPI_ISL_2089993, EPI_ISL_2089995, EPI_ISL_2090010, EPI_ISL_2090030, EPI_ISL_2090061, EPI_ISL_2090063, EPI_ISL_2090196, EPI_ISL_2090257, EPI_ISL_2090266, EPI_ISL_2090271, EPI_ISL_2090311, EPI_ISL_2090329, EPI_ISL_2090352, EPI_ISL_2090354, EPI_ISL_2090361, EPI_ISL_2090368, EPI_ISL_2090395, EPI_ISL_2090402, EPI_ISL_2090403, EPI_ISL_2090431, EPI_ISL_2090455, EPI_ISL_2090456, EPI_ISL_2090457, EPI_ISL_2132545, EPI_ISL_2145803, EPI_ISL_2145804, EPI_ISL_2145821, EPI_ISL_2145837, EPI_ISL_2145851, EPI_ISL_2145866, EPI_ISL_2145869, EPI_ISL_2145870, EPI_ISL_2145899, EPI_ISL_2145903, EPI_ISL_2145911, EPI_ISL_2145912, EPI_ISL_2145925, EPI_ISL_2145965, EPI_ISL_2145972, EPI_ISL_2146010, EPI_ISL_2146112, EPI_ISL_2146137, EPI_ISL_2146167, EPI_ISL_2146186, EPI_ISL_2146237, EPI_ISL_2146239, EPI_ISL_2146257, EPI_ISL_2146258, EPI_ISL_2146280, EPI_ISL_2146319, EPI_ISL_2146322, EPI_ISL_2146337, EPI_ISL_2146354, EPI_ISL_2146387, EPI_ISL_2146399, EPI_ISL_2146401, EPI_ISL_2146448, EPI_ISL_2146466, EPI_ISL_2146467, EPI_ISL_2146468, EPI_ISL_2146493, EPI_ISL_2146521, EPI_ISL_2146522, EPI_ISL_2146528, EPI_ISL_2146529, EPI_ISL_2146551, EPI_ISL_2146562, EPI_ISL_2146628, EPI_ISL_2146644, EPI_ISL_2146653, EPI_ISL_2146657, EPI_ISL_2146684, EPI_ISL_2146719, EPI_ISL_2146722, EPI_ISL_2146736, EPI_ISL_2146775, EPI_ISL_2146780, EPI_ISL_2146784, EPI_ISL_2146795, EPI_ISL_2146824, EPI_ISL_2146833, EPI_ISL_2146877, EPI_ISL_2146886, EPI_ISL_2146894, EPI_ISL_2146921, EPI_ISL_2146957, EPI_ISL_2146959, EPI_ISL_2147010, EPI_ISL_2147040, EPI_ISL_2147045, EPI_ISL_2147071, EPI_ISL_2147101, EPI_ISL_2147103, EPI_ISL_2147115, EPI_ISL_2147126, EPI_ISL_2147131, EPI_ISL_2147132, EPI_ISL_2147133, EPI_ISL_2147147, EPI_ISL_2147159, EPI_ISL_2147188, EPI_ISL_2147244, EPI_ISL_2147252, EPI_ISL_2147398, EPI_ISL_2147403, EPI_ISL_2147405, EPI_ISL_2147419, EPI_ISL_2147454, EPI_ISL_2147461, EPI_ISL_2147484, EPI_ISL_2147503, EPI_ISL_2147520, EPI_ISL_2147522, EPI_ISL_2147537, EPI_ISL_2147542, EPI_ISL_2147547, EPI_ISL_2147548, EPI_ISL_2147564, EPI_ISL_2147572, EPI_ISL_2147604, EPI_ISL_2147605, EPI_ISL_2147606, EPI_ISL_2147616, EPI_ISL_2147667, EPI_ISL_2147669, EPI_ISL_2147671, EPI_ISL_2147755, EPI_ISL_2147817, EPI_ISL_2147832, EPI_ISL_2147887, EPI_ISL_2147897, EPI_ISL_2147921, EPI_ISL_2147943, EPI_ISL_2147949, EPI_ISL_2147956, EPI_ISL_2147967, EPI_ISL_2147968, EPI_ISL_2148006, EPI_ISL_2148008, EPI_ISL_2148068, EPI_ISL_2148106, EPI_ISL_2148190, EPI_ISL_2148195, EPI_ISL_2148200, EPI_ISL_2148245, EPI_ISL_2148248, EPI_ISL_2148257, EPI_ISL_2148258, EPI_ISL_2148277, EPI_ISL_2148312, EPI_ISL_2148322, EPI_ISL_2148373, EPI_ISL_2148378, EPI_ISL_2148406, EPI_ISL_2148416, EPI_ISL_2148433, EPI_ISL_2148451, EPI_ISL_2148459, EPI_ISL_2148505, EPI_ISL_2148507, EPI_ISL_2148527, EPI_ISL_2148541, EPI_ISL_2148576, EPI_ISL_2148632, EPI_ISL_2148635, EPI_ISL_2148668, EPI_ISL_2148710, EPI_ISL_2148728, EPI_ISL_2148763, EPI_ISL_2148766, EPI_ISL_2148767, EPI_ISL_2148768, EPI_ISL_2148778, EPI_ISL_2148782, EPI_ISL_2148788, EPI_ISL_2148798, EPI_ISL_2148813, EPI_ISL_2148814, EPI_ISL_2148817, EPI_ISL_2148853, EPI_ISL_2148901, EPI_ISL_2148902, EPI_ISL_2148905, EPI_ISL_2148910, EPI_ISL_2148913, EPI_ISL_2148914, EPI_ISL_2148925, EPI_ISL_2148939, EPI_ISL_2148947, EPI_ISL_2148948, EPI_ISL_2149145, EPI_ISL_2149216, EPI_ISL_2149217, EPI_ISL_2149302, EPI_ISL_2149311, EPI_ISL_2149318, EPI_ISL_2149340, EPI_ISL_2149356, EPI_ISL_2149385, EPI_ISL_2149391, EPI_ISL_2149418, EPI_ISL_2149448, EPI_ISL_2149499, EPI_ISL_2149541, EPI_ISL_2149542, EPI_ISL_2149543, EPI_ISL_2149549, EPI_ISL_2149565, EPI_ISL_2149589, EPI_ISL_2149594, EPI_ISL_2149632, EPI_ISL_2149655, EPI_ISL_2149656, EPI_ISL_2149697, EPI_ISL_2149707, EPI_ISL_2149728, EPI_ISL_2149745, EPI_ISL_2149753, EPI_ISL_2149775, EPI_ISL_2149795, EPI_ISL_2149829, EPI_ISL_2149861, EPI_ISL_2149919, EPI_ISL_2149922, EPI_ISL_2149926, EPI_ISL_2150079, EPI_ISL_2150082, EPI_ISL_2150099, EPI_ISL_2150117, EPI_ISL_2150139, EPI_ISL_2150140, EPI_ISL_2150151, EPI_ISL_2150191, EPI_ISL_2150192, EPI_ISL_2150218, EPI_ISL_2150222, EPI_ISL_2150315, EPI_ISL_2150325, EPI_ISL_2150330, EPI_ISL_2150361, EPI_ISL_2150384, EPI_ISL_2150433, EPI_ISL_2150465, EPI_ISL_2150466, EPI_ISL_2150494, EPI_ISL_2150501, EPI_ISL_2150507, EPI_ISL_2150535, EPI_ISL_2150539, EPI_ISL_2150542, EPI_ISL_2150554, EPI_ISL_2150567, EPI_ISL_2150685, EPI_ISL_2150686, EPI_ISL_2150786, EPI_ISL_2150805, | | | |

| | | | |
|---|---|---|---|
| EPI_ISL_2150852, EPI_ISL_2150854, EPI_ISL_2150906, EPI_ISL_2150907, EPI_ISL_2150937, EPI_ISL_2150960, EPI_ISL_2150964, EPI_ISL_2158851, EPI_ISL_2158874, EPI_ISL_2158883, EPI_ISL_2158911, EPI_ISL_2158919, EPI_ISL_2158936, EPI_ISL_2158973, EPI_ISL_2158982, EPI_ISL_2158985, EPI_ISL_2159125, EPI_ISL_2159128, EPI_ISL_2159135, EPI_ISL_2159158, EPI_ISL_2159172, EPI_ISL_2159215, EPI_ISL_2159220, EPI_ISL_2159250, EPI_ISL_2159252, EPI_ISL_2159263, EPI_ISL_2159307, EPI_ISL_2159329, EPI_ISL_2159366, EPI_ISL_2159367, EPI_ISL_2159430, EPI_ISL_2159431, EPI_ISL_2159463, EPI_ISL_2180537, EPI_ISL_2180582, EPI_ISL_2180586, EPI_ISL_2180588, EPI_ISL_2180656, EPI_ISL_2180666, EPI_ISL_2180671, EPI_ISL_2180720, EPI_ISL_2180723, EPI_ISL_2180732, EPI_ISL_2180746, EPI_ISL_2180749, EPI_ISL_2180751, EPI_ISL_2180799, EPI_ISL_2180818, EPI_ISL_2180832, EPI_ISL_2180865, EPI_ISL_2180880, EPI_ISL_2180910, EPI_ISL_2180948, EPI_ISL_2181054, EPI_ISL_2181101, EPI_ISL_2181107, EPI_ISL_2181134, EPI_ISL_2181174, EPI_ISL_2181175, EPI_ISL_2181196, EPI_ISL_2181225, EPI_ISL_2181234, EPI_ISL_2181251, EPI_ISL_2181278, EPI_ISL_2181279, EPI_ISL_2181292, EPI_ISL_2181338, EPI_ISL_2181349, EPI_ISL_2181372, EPI_ISL_2181375, EPI_ISL_2181376, EPI_ISL_2181387, EPI_ISL_2181388, EPI_ISL_2181403, EPI_ISL_2181434, EPI_ISL_2181446, EPI_ISL_2181554, EPI_ISL_2181556, EPI_ISL_2181560, EPI_ISL_2181578, EPI_ISL_2181604, EPI_ISL_2181610, EPI_ISL_2181626, EPI_ISL_2181637, EPI_ISL_2181644, EPI_ISL_2187363, EPI_ISL_2187377, EPI_ISL_2187389, EPI_ISL_2187403, EPI_ISL_2187408, EPI_ISL_2187409, EPI_ISL_2187432, EPI_ISL_2187442, EPI_ISL_2187486, EPI_ISL_2187490, EPI_ISL_2187505, EPI_ISL_2187513, EPI_ISL_2187574, EPI_ISL_2187609, EPI_ISL_2187614, EPI_ISL_2202991, EPI_ISL_2203094, EPI_ISL_2203219, EPI_ISL_2203221, EPI_ISL_2203229, EPI_ISL_2204817, EPI_ISL_2204857, EPI_ISL_2241648, EPI_ISL_2241669, EPI_ISL_2241671, EPI_ISL_2241678, EPI_ISL_2280329, EPI_ISL_2280351, EPI_ISL_2280354, EPI_ISL_2280356, EPI_ISL_2280357, EPI_ISL_2280404, EPI_ISL_2280407, EPI_ISL_2280413, EPI_ISL_2280414, EPI_ISL_2280426, EPI_ISL_2280442, EPI_ISL_2280467, EPI_ISL_2280504, EPI_ISL_2280505, EPI_ISL_2280513, EPI_ISL_2280521, EPI_ISL_2280530, EPI_ISL_2280547, EPI_ISL_2280553, EPI_ISL_2280554, EPI_ISL_2280564, EPI_ISL_2280579, EPI_ISL_2280601, EPI_ISL_2280604, EPI_ISL_2280652, EPI_ISL_2280653, EPI_ISL_2280654, EPI_ISL_2280655, EPI_ISL_2280680, EPI_ISL_2280681, EPI_ISL_2280736, EPI_ISL_2280753, EPI_ISL_2280760, EPI_ISL_2280761, EPI_ISL_2280762, EPI_ISL_2280829, EPI_ISL_2280847, EPI_ISL_2280854, EPI_ISL_2280933, EPI_ISL_2280934, EPI_ISL_2280948, EPI_ISL_2280963, EPI_ISL_2280974, EPI_ISL_2280987, EPI_ISL_2281044, EPI_ISL_2281063, EPI_ISL_2281079, EPI_ISL_2281100, EPI_ISL_2281105, EPI_ISL_2281112, EPI_ISL_2281126, EPI_ISL_2281148, EPI_ISL_2281208, EPI_ISL_2283819, EPI_ISL_2283822, EPI_ISL_2283828, EPI_ISL_2283846, EPI_ISL_2283848, EPI_ISL_2283854, EPI_ISL_2283856, EPI_ISL_2283861, EPI_ISL_2283863, EPI_ISL_2283867, EPI_ISL_2283869, EPI_ISL_2283876, EPI_ISL_2283878, EPI_ISL_2321252, EPI_ISL_2321267, EPI_ISL_2321301, EPI_ISL_2321338, EPI_ISL_2321358, EPI_ISL_2367654, EPI_ISL_2367658, EPI_ISL_2367673, EPI_ISL_2368210, EPI_ISL_2368238, EPI_ISL_2368259, EPI_ISL_2368285, EPI_ISL_2368286, EPI_ISL_2368299, EPI_ISL_2368336, EPI_ISL_2368340, EPI_ISL_2368381, EPI_ISL_2368385, EPI_ISL_2368386, EPI_ISL_2368471, EPI_ISL_2368478, EPI_ISL_2368542, EPI_ISL_2368559, EPI_ISL_2368582, EPI_ISL_2368622, EPI_ISL_2368657, EPI_ISL_2368681, EPI_ISL_2368698, EPI_ISL_2368786, EPI_ISL_2368800, EPI_ISL_2368838, EPI_ISL_2368927, EPI_ISL_2368969, EPI_ISL_2369960, EPI_ISL_2370031, EPI_ISL_2370088, EPI_ISL_2370188, EPI_ISL_2370336, EPI_ISL_2370538, EPI_ISL_2370622, EPI_ISL_2372306, EPI_ISL_2372310, EPI_ISL_2372312, EPI_ISL_2372322, EPI_ISL_2372347, EPI_ISL_2372372, EPI_ISL_2372384, EPI_ISL_2372386, EPI_ISL_2372435, EPI_ISL_2372445, EPI_ISL_2372452, EPI_ISL_2421834, EPI_ISL_2421849, EPI_ISL_2421869, EPI_ISL_2421876, EPI_ISL_2421882, EPI_ISL_2421885, EPI_ISL_2421886, EPI_ISL_2421896, EPI_ISL_2421908, EPI_ISL_2421937, EPI_ISL_2421943, EPI_ISL_2421946, EPI_ISL_2421998, EPI_ISL_2422002, EPI_ISL_2422055, EPI_ISL_2422093, EPI_ISL_2422111, EPI_ISL_2442324, EPI_ISL_2489829, EPI_ISL_2489837, EPI_ISL_2489841, EPI_ISL_2489843, EPI_ISL_2489846, EPI_ISL_2489875, EPI_ISL_2489884, EPI_ISL_2489887, EPI_ISL_2600264, EPI_ISL_2600265, EPI_ISL_2600290 | | | |
| see above | Aegis Sciences Corporation | Centers for Disease Control and Prevention Division of Viral Diseases, Pathogen Discovery | Adrian Paskey; Alec Vest; Benjamin Rambo-Martin; Christopher Gulvick; Clinton R. Paden; Cyndi Clark; Dakota Howard; Darlene Wagner; Dhwani Batra; Dillon Nall; Duncan MacCannell; Ethan Sanders; Holly Houdeshell; Jason Caravas; Kara Moser; Matthew Hardison; Matthew Schmerer; Ola Kvalvaag; Patrick Campbell; Peter W. Cook; Rob Case; Scott Sammons; Shatavia Morrison; Shaun Westlund; Vikramsinha Ghorpade; Yvette Unoarumhi |
| EPI_ISL_2249137 | Affidea | Instituto Nacional de Saude (INSA) and Centro de Investigaçao em Biodiversidade e Recursos Geneticos (CIBIO), Universidade do Porto | Borges et al |
| EPI_ISL_2136442, EPI_ISL_2154069, EPI_ISL_2187648, EPI_ISL_2272531, EPI_ISL_2272538, EPI_ISL_2308647, EPI_ISL_2376008, EPI_ISL_2376069, EPI_ISL_2376089, EPI_ISL_2376098, EPI_ISL_2422100, EPI_ISL_2422101, EPI_ISL_2455425 | | | |
| see above | Alaska State Virology Laboratory | Alaska State Virology Laboratory | Elva House; Jack Chen; Jacob Zidek; Lisa Smith; Ph.D.; Stephanie DeRonde |
| EPI_ISL_3243025, EPI_ISL_3243026, EPI_ISL_3243028, EPI_ISL_3243029, EPI_ISL_3243030, EPI_ISL_3243032, EPI_ISL_3243033 | | | |
| see above | Applied DNA Sciences Inc | Applied DNA Sciences Inc | Huan Dong; Lucas Liu; Spencer William; Yuhua Sun |
| EPI_ISL_2227487, EPI_ISL_2227488, EPI_ISL_2227489, EPI_ISL_2325174 | Arizona State Public Health Laboratory | Arizona State Public Health Laboratory | Jessica Escobar; Katherine Fullerton; Linda Getsinger; Nobuko Fukushima; Stacy White; Trung Huynh; Victor Waddell |
| EPI_ISL_2007175, EPI_ISL_2007176, EPI_ISL_2007178, EPI_ISL_2015335, EPI_ISL_2015344, EPI_ISL_2153660, EPI_ISL_2153661, EPI_ISL_2153663, EPI_ISL_2153664, EPI_ISL_2250017, EPI_ISL_2250018, EPI_ISL_2250021, EPI_ISL_2250036, EPI_ISL_2250047, EPI_ISL_2250060, EPI_ISL_2250082, EPI_ISL_2250100, EPI_ISL_2250102, EPI_ISL_2250123, EPI_ISL_2250130, EPI_ISL_2250132, EPI_ISL_2250147 | | | |
| see above | Arizona State University | Arizona State University | Efrem S. Lim; Joshua LaBaer; Joy M. Blain; LaRinda A. Holland; Matthew F. Smith; Nicholas J. Mellor; Peter T. Skidmore; Rabia Maqsood; Valerie Harris; Vel Murugan |
| EPI_ISL_2484741, EPI_ISL_2484742, EPI_ISL_2931027, EPI_ISL_2931039, EPI_ISL_2931040 | Arkansas Children's Hospital | Center for Global Health, University of New Mexico Health Sciences Center | Amanda Novack; Darrell Dinwiddie; Daryl Domman; Dirk Haselow; Joshua L. Kennedy; Kurt Schwalm; Valerie Morley |
| EPI_ISL_2484740, EPI_ISL_2931024, EPI_ISL_2939042, EPI_ISL_2939077, EPI_ISL_2939081, EPI_ISL_2939084 | Arkansas Department of Health | Center for Global Health, University of New Mexico Health Sciences Center | Amanda Novack; Atul Kothari; Darrell Dinwiddie; Daryl Domman; Dirk Haselow; Joshua L. Kennedy; Kurt Schwalm; Valerie Morley |
| EPI_ISL_2692305, EPI_ISL_2692447, EPI_ISL_2715034 | Atlas Genomics | Seattle Flu Study | Amanda Adler; Barry R. Lutz; Benjamin Pelle; Brian Hiatt; Caitlin R. Wolf; Chris D. Frazar; Deborah A. Nickerson; Elisabeth Brandstetter; Erica Ryke; Geoff Melly; Helen Y. Chu; Janet A. Englund; Jay Shendure; Jover Lee; Kairsten Fay; Kirsten Lacombe; Lea M. Starita; Mark J. Rieder; Matthew Richardson; Matthew Thompson; Melissa Truong; Michael Boeckh; Michael Famulare; Misja Ilcisin; Peter D. Han; Philip Dykema; Romesh Gautom; Scott Lindquist; Thomas R. Sibley; Trevor Bedford |
| EPI_ISL_2485062, EPI_ISL_2485063, EPI_ISL_2485158 | Austrian Agency for Health and Food Safety (AGES) | Bergthaler laboratory, CeMM Research Center for Molecular Medicine of the Austrian Academy of Sciences | Andreas Bergthaler; Anna Schedl; Bekir Erguner; Benedikt Agerer; Christoph Bock; Fabian Amman; Jan Laine; Lukas Endler; Maelle Le Moing; Martin Senekowitsch; Michael Schuster; Petr Triska; Thomas Penz |
| EPI_ISL_2249139 | Avelab | Instituto Nacional de Saude (INSA) and Centro de Investigaçao em Biodiversidade e Recursos Geneticos (CIBIO), Universidade do Porto | Borges et al |
| EPI_ISL_1840731, EPI_ISL_1840732, EPI_ISL_1840750, EPI_ISL_1840758, EPI_ISL_1840770, EPI_ISL_1840781, EPI_ISL_2080258, EPI_ISL_2080274, EPI_ISL_2080282, EPI_ISL_2080294, EPI_ISL_2080295, EPI_ISL_2080297, EPI_ISL_2318906 | | | |
| see above | Azienda Sanitaria dell'Alto Adige Laboratorio Aziendale di Microbiologia e Virologia | Istituto di Genomica Applicata | Davide Scaglione; Eleonora Paparelli; Elisa Masi; Elisabetta Giacobazzi; Elisabetta Pagani; Gabriele Magris; Irena Jurman; Irene Bianconi; Michele Morgante; Stefanie Wieser; Vera Vendramin |
| EPI_ISL_2444173, EPI_ISL_2444202 | BADY BASSITT | Instituto Butantan / FAMERP | Antonio Jorge Martins; Claudia Renata dos Santos Barros; David Schlesinger; Debora Botequio Moretti; Dimas Tadeu Covas; Elaine Cristina Marqueze; Elaine Vieira Santos; Evandra Strazza Rodrigues; Heidge Fukumasu; Jayme Augusto de Souza-Neto; José Salvatore Leister Patané; Luiz Alcantara; Luiz Lehmann Coutinho; Maria Carolina Elias; Maurício Lacerda Nogueira; Rafael dos Santos Bezerra; Raul Machado Neto; Rejane Maria Tommasini Grotto; Ricardo Haddad; Sandra Coccuzzo Sampaio Vessoni; Simone Kashima; Svetoslav Nanev Slavov; Vincent Louis Viala |
| EPI_ISL_2444212 | BARRETOS | Instituto Butantan / FAMERP | Antonio Jorge Martins; Claudia Renata dos Santos Barros; David Schlesinger; Debora Botequio Moretti; Dimas Tadeu Covas; Elaine Cristina Marqueze; Elaine Vieira Santos; Evandra Strazza Rodrigues; Heidge Fukumasu; Jayme Augusto de Souza-Neto; José Salvatore Leister Patané; Luiz Alcantara; Luiz Lehmann Coutinho; Maria Carolina Elias; Maurício Lacerda Nogueira; Rafael dos Santos Bezerra; Raul Machado Neto; Rejane Maria Tommasini Grotto; Ricardo Haddad; Sandra Coccuzzo Sampaio Vessoni; Simone Kashima; Svetoslav Nanev Slavov; Vincent Louis Viala |
| EPI_ISL_2493199, EPI_ISL_2493763, EPI_ISL_2493765, EPI_ISL_2493768, EPI_ISL_2493769, EPI_ISL_2493771, EPI_ISL_2493772, EPI_ISL_2493773, EPI_ISL_2493774, EPI_ISL_2493776, EPI_ISL_2493777, EPI_ISL_2493778, EPI_ISL_2493780, EPI_ISL_2493782, EPI_ISL_2493784, EPI_ISL_2493786, EPI_ISL_2493787, EPI_ISL_2493788, EPI_ISL_2494314, EPI_ISL_2494315, EPI_ISL_2494319, EPI_ISL_2500974 | | | |
| see above | BIOFAST | Instituto Butantan | Antonio Jorge Martins; Claudia Renata dos Santos Barros; David Schlesinger; Debora Botequio Moretti; Dimas Tadeu Covas; Elaine Cristina Marqueze; Elaine Vieira Santos; Evandra Strazza Rodrigues; Heidge Fukumasu; Jayme Augusto de Souza-Neto; José Salvatore Leister Patané; Luiz Alcantara; Luiz Lehmann Coutinho; Maria Carolina Elias; Maurício Lacerda Nogueira; Rafael dos Santos Bezerra; Raul Machado Neto; Rejane Maria Tommasini Grotto; Ricardo Haddad; Sandra Coccuzzo Sampaio Vessoni; Simone Kashima; Svetoslav Nanev Slavov; Vincent Louis Viala |
| EPI_ISL_2445085 | BIOFAST CENTRO SANTA CECÍLIA | Instituto Butantan | Antonio Jorge Martins; Claudia Renata dos Santos Barros; David Schlesinger; Debora Botequio Moretti; Dimas Tadeu Covas; Elaine Cristina Marqueze; Elaine Vieira Santos; Evandra Strazza Rodrigues; Heidge Fukumasu; Jayme Augusto de Souza-Neto; José Salvatore Leister Patané; Luiz Alcantara; Luiz Lehmann Coutinho; Maria Carolina Elias; Maurício Lacerda Nogueira; Rafael dos Santos Bezerra; Raul Machado Neto; Rejane Maria Tommasini Grotto; Ricardo Haddad; Sandra Coccuzzo Sampaio Vessoni; Simone Kashima; Svetoslav Nanev Slavov; Vincent Louis Viala |
| EPI_ISL_2445086 | BIOFAST CENTRO SÉ | Instituto Butantan | Antonio Jorge Martins; Claudia Renata dos Santos Barros; David Schlesinger; Debora Botequio Moretti; Dimas Tadeu Covas; Elaine Cristina Marqueze; Elaine Vieira Santos; Evandra Strazza Rodrigues; Heidge Fukumasu; Jayme Augusto de Souza-Neto; José Salvatore Leister Patané; Luiz Alcantara; Luiz Lehmann Coutinho; Maria Carolina Elias; Maurício Lacerda Nogueira; Rafael dos Santos Bezerra; Raul Machado Neto; Rejane Maria Tommasini Grotto; Ricardo Haddad; Sandra Coccuzzo Sampaio Vessoni; Simone Kashima; Svetoslav Nanev Slavov; Vincent Louis Viala |
| EPI_ISL_2445094 | BIOFAST LESTE GUAIANASES | Instituto Butantan | Antonio Jorge Martins; Claudia Renata dos Santos Barros; David Schlesinger; Debora Botequio Moretti; Dimas Tadeu Covas; Elaine Cristina Marqueze; Elaine Vieira Santos; Evandra Strazza Rodrigues; Heidge Fukumasu; Jayme Augusto de Souza-Neto; José Salvatore Leister Patané; Luiz Alcantara; Luiz Lehmann Coutinho; Maria Carolina Elias; Maurício Lacerda Nogueira; Rafael dos Santos Bezerra; Raul Machado Neto; Rejane Maria Tommasini Grotto; Ricardo Haddad; Sandra Coccuzzo Sampaio Vessoni; Simone Kashima; Svetoslav Nanev Slavov; Vincent Louis Viala |
| EPI_ISL_2445099, EPI_ISL_2445102 | BIOFAST LESTE ITAIM PAULISTA | Instituto Butantan | Antonio Jorge Martins; Claudia Renata dos Santos Barros; David Schlesinger; Debora Botequio Moretti; Dimas Tadeu Covas; Elaine Cristina Marqueze; Elaine Vieira Santos; Evandra Strazza Rodrigues; Heidge Fukumasu; Jayme Augusto de Souza-Neto; José Salvatore Leister Patané; Luiz Alcantara; Luiz Lehmann Coutinho; Maria Carolina Elias; Maurício Lacerda Nogueira; Rafael dos Santos Bezerra; Raul Machado Neto; Rejane Maria Tommasini Grotto; Ricardo Haddad; Sandra Coccuzzo Sampaio Vessoni; Simone Kashima; Svetoslav Nanev Slavov; Vincent Louis Viala |
| EPI_ISL_2445088, EPI_ISL_2445090, EPI_ISL_2445092 | BIOFAST LESTE ITAQUERA | Instituto Butantan | Antonio Jorge Martins; Claudia Renata dos Santos Barros; David Schlesinger; Debora Botequio Moretti; Dimas Tadeu Covas; Elaine Cristina Marqueze; Elaine Vieira Santos; Evandra Strazza Rodrigues; Heidge Fukumasu; Jayme Augusto de Souza-Neto; José Salvatore Leister Patané; Luiz Alcantara; Luiz Lehmann Coutinho; Maria Carolina Elias; Maurício Lacerda Nogueira; Rafael dos Santos Bezerra; Raul Machado Neto; Rejane Maria Tommasini Grotto; Ricardo Haddad; Sandra Coccuzzo Sampaio Vessoni; Simone Kashima; Svetoslav Nanev Slavov; Vincent Louis Viala |
| EPI_ISL_2445097, EPI_ISL_2445098 | BIOFAST LESTE SÃO MIGUEL PAULISTA | Instituto Butantan | Antonio Jorge Martins; Claudia Renata dos Santos Barros; David Schlesinger; Debora Botequio Moretti; Dimas Tadeu Covas; Elaine Cristina Marqueze; Elaine Vieira Santos; Evandra Strazza Rodrigues; Heidge Fukumasu; Jayme Augusto de Souza-Neto; José Salvatore Leister Patané; Luiz Alcantara; Luiz Lehmann Coutinho; Maria Carolina Elias; Maurício Lacerda Nogueira; Rafael dos Santos Bezerra; Raul Machado Neto; Rejane Maria Tommasini Grotto; Ricardo Haddad; Sandra Coccuzzo Sampaio Vessoni; Simone Kashima; Svetoslav Nanev Slavov; Vincent Louis Viala |
| EPI_ISL_2445103 | BIOFAST LESTE TIRADENTES | Instituto Butantan | Antonio Jorge Martins; Claudia Renata dos Santos Barros; David Schlesinger; Debora Botequio Moretti; Dimas Tadeu Covas; Elaine Cristina Marqueze; Elaine Vieira Santos; Evandra Strazza Rodrigues; Heidge Fukumasu; Jayme Augusto de Souza-Neto; José Salvatore Leister Patané; Luiz Alcantara; Luiz Lehmann Coutinho; Maria Carolina Elias; Maurício Lacerda Nogueira; Rafael dos Santos Bezerra; Raul Machado Neto; Rejane Maria Tommasini Grotto; Ricardo Haddad; Sandra Coccuzzo Sampaio Vessoni; Simone Kashima; Svetoslav Nanev Slavov; Vincent Louis Viala |
| EPI_ISL_2445105, EPI_ISL_2445107, EPI_ISL_2473811 | BIOFAST NORTE SANTANA/JAÇANA/TUCURUVI | Instituto Butantan | Antonio Jorge Martins; Claudia Renata dos Santos Barros; David Schlesinger; Debora Botequio Moretti; Dimas Tadeu Covas; Elaine Cristina Marqueze; Elaine Vieira Santos; Evandra Strazza Rodrigues; Heidge Fukumasu; Jayme Augusto de Souza-Neto; José Salvatore Leister Patané; Luiz Alcantara; Luiz Lehmann Coutinho; Maria Carolina Elias; Maurício Lacerda Nogueira; Rafael dos Santos Bezerra; Raul Machado Neto; Rejane Maria Tommasini Grotto; Ricardo Haddad; Sandra Coccuzzo Sampaio Vessoni; Simone Kashima; Svetoslav Nanev Slavov; Vincent Louis Viala |
| EPI_ISL_2445108, EPI_ISL_2445109, EPI_ISL_2445112 | BIOFAST SUL SANTO AMARO/CIDADE ADEMAR | Instituto Butantan | Antonio Jorge Martins; Claudia Renata dos Santos Barros; David Schlesinger; Debora Botequio Moretti; Dimas Tadeu Covas; Elaine Cristina Marqueze; Elaine Vieira Santos; Evandra Strazza Rodrigues; Heidge Fukumasu; Jayme Augusto de Souza-Neto; José Salvatore Leister Patané; Luiz Alcantara; Luiz Lehmann Coutinho; Maria Carolina Elias; Maurício Lacerda Nogueira; Rafael dos Santos Bezerra; Raul Machado Neto; Rejane Maria Tommasini Grotto; Ricardo Haddad; Sandra Coccuzzo Sampaio Vessoni; Simone Kashima; Svetoslav Nanev Slavov; Vincent Louis Viala |
| EPI_ISL_2373844 | BIOMNIS LYON | CNR Virus des Infections Respiratoires - France SUD | Antonin Bal; Bruno Lina; Gregory Destras; Gwendolyne Burfin; Hadrien Regue; Laurence Josset; Martine Valette; Quentin Semanas |
| EPI_ISL_2421233 | BOSTON HEART DIAGNOSTICS CORP | Wadsworth Center, New York State Department of Health | Alexis Russell; Catharine Prussing; Daryl M. Lamson; Erasmus Schneider; Erica Lasek-Nesselquist; John Kelly; Jonathan Plitnick; Kirsten St. George; Matthew Shudt; Melissa A Leisner; Navjot Singh |
| EPI_ISL_2484739 | Baptist Health Medical Center | Center for Global Health, University of New Mexico Health Sciences Center | Amanda Novack; Darrell Dinwiddie; Daryl Domman; Dirk Haselow; Joshua L. Kennedy; Kurt Schwalm; Valerie Morley |
| EPI_ISL_2262754, EPI_ISL_2262815 | Bayerisches Landesamt für Gesundheit und Lebensmittelsicherheit (LGL) | Robert Koch Institute | |
| EPI_ISL_2083061, | Baylor Scott & White-Temple | Baylor Scott & White-Temple | Ari Rao; Kimberly Walker; Linden Morales; Marcus Volz; Shelby Hendrickson; Shelby Johnson |

| | | | |
|---|---|---|---|
| EPI_ISL_2083080, EPI_ISL_2193240, EPI_ISL_2193265, EPI_ISL_2443472, EPI_ISL_2443507 | | | |
| EPI_ISL_2204446 | Berkeley Medical Center | WVU and Marshall University Combined Genomics Core Facilities | James Denvir; Peter Perrotta; Peter Stoilov; Ryan Percifield; Wesley Kimble |
| EPI_ISL_2105670, EPI_ISL_2105671 | Biolab Diagnostic Laboratories | Biolab Diagnostic Laboratories | Ahmad Tibi; Amid Abdelnour; Badia Saddedin; Eiad Atwa; Issa Abu-Dayyeh; Lama Hussein; Shayma Ali |
| EPI_ISL_2698103, EPI_ISL_2698104 | Biology, UFLA, Universidade Federal de Lavras | Biology, UFLA | Barcante, J.; Cherem, J.; Fernandes, G.; Luciano, P.; Melo, D.; Pylro, V. |
| EPI_ISL_1917098, EPI_ISL_1917111, EPI_ISL_1917129, EPI_ISL_2401290, EPI_ISL_2401348, EPI_ISL_2401349, EPI_ISL_2401558, EPI_ISL_3145466, EPI_ISL_3145467, EPI_ISL_3145468, EPI_ISL_3145469, EPI_ISL_3145470, EPI_ISL_3145471, EPI_ISL_3145473, EPI_ISL_3145474 | | | |
| see above | BioneXt Lab | Laboratoire national de sante, Microbiology, Microbial Genomics Platform | Anke Wienecke-Baldacchino; Catherine Ragimbeau; Elodie Solarino; Fatu Djabi; Jessica Tapp; Lise Pignon; Raoul Salmon; Tamir Abdelrahman; Thibault Ferrandon; Virginie Jover |
| EPI_ISL_3494467, EPI_ISL_3494483 | Boston University CTL | Boston University/National Emerging Infectious Disease Laboratories | Catherine Klapperich; Jacquelyn Turcinovic; John H. Connor; Lena Landeverde; Lynn Doucette-Stamm |
| EPI_ISL_3484463, EPI_ISL_3484465, EPI_ISL_3485505, EPI_ISL_3485949, EPI_ISL_3486879 | British Columbia Centre For Disease Control | BCCDC Public Health Laboratory | Ana Pacagnella; Corrinne Ng; Dan Fornika; John Tyson; Kim Macdonald; Kimia Kamelian; Linda Hoang; Loretta Janz; Mel Krajden; Prystajecky Natalie; Robert Azana; Shannon Russell |
| EPI_ISL_2192739, EPI_ISL_2192747, EPI_ISL_2192775, EPI_ISL_2192804, EPI_ISL_2192833, EPI_ISL_2192842, EPI_ISL_2192857, EPI_ISL_2192880, EPI_ISL_2192882, EPI_ISL_2192928, EPI_ISL_2192931, EPI_ISL_2192947, EPI_ISL_2193017, EPI_ISL_2193033, EPI_ISL_2193040, EPI_ISL_2193055, EPI_ISL_2193060, EPI_ISL_2193067, EPI_ISL_2193107, EPI_ISL_2193152, EPI_ISL_2193179 | | | |
| see above | Broad Institute Clinical Research Sequencing Platform | Infectious Disease Program, Broad Institute of Harvard and MIT | Adams, G.; B.L.; B.W.; Bauer, M.; Birren; Blumenstiel, B.; Brown, C.; Carter, A.; Chaluvadi, S.; D.J.; DeFelice, M.; DeRuff, K.; Dodge, S.; Gabriel, S.; Gallagher, G.; Gladden-Young, A.; Granger, B.; J.E.; K.J.; Lagerborg, K.; Larkin, K.; Lee, M.; Lemieux; Lennon, N.; Loreth, C.; Madoff, L.; McGovern, S.; Meldrim, J.; Normandin, E.; P.C.; Park; Pearlman, L.; Reilly, S.; Rudy, M.; Sabeti; Siddle; Smole, S.; Tomkins-Tinch, C.; Vicente, G.; and MacInnis |
| EPI_ISL_2341513, EPI_ISL_2496898, EPI_ISL_2496899, EPI_ISL_2496900, EPI_ISL_2496901, EPI_ISL_2496902, EPI_ISL_2496905, EPI_ISL_2496906, EPI_ISL_2496907, EPI_ISL_2496909, EPI_ISL_2496913, EPI_ISL_2497648, EPI_ISL_2497649, EPI_ISL_2497650, EPI_ISL_2497651, EPI_ISL_2497652, EPI_ISL_2497655, EPI_ISL_2497665, EPI_ISL_2497668, EPI_ISL_2500044, EPI_ISL_2500102, EPI_ISL_2500120, EPI_ISL_2500149, EPI_ISL_2500164, EPI_ISL_2500170, EPI_ISL_2500186, EPI_ISL_2500210 | | | |
| see above | CA DPH Viral and Rickettsial Disease Laboratory | Chan-Zuckerberg Biohub | CZB Cliahub Consortium |
| EPI_ISL_2105135, EPI_ISL_2248730, EPI_ISL_2383987 | CA-Los Angeles County Public Health Laboratory | Centers for Disease Control and Prevention Division of Viral Diseases, Pathogen Discovery | Alison Laufer Halpin; Ben L. Rambo-Martin; Clinton R. Paden; Dakota Howard; Darlene Wagner; Dave Wentworth; Dhwani Batra; Jasmine Padilla; Justin Lee; Katie Dillon; Krista Queen; Kristen Knipe; Kristine Lacek; Mark Burroughs; Matthew Schmerer; Mili Sheth; Peter Cook; Sam Shepard; Sarah Nobles; Shoshona Le; Suxiang Tong; Vivien Dugan; Yvette Unoarumhi |
| EPI_ISL_2445514, EPI_ISL_2473723, EPI_ISL_2473788, EPI_ISL_2473818 | CADIP VILA REGINA FERNANDOPOLIS | Instituto Butantan | Antonio Jorge Martins; Claudia Renata dos Santos Barros; David Schlesinger; Debora Botequio Moretti; Dimas Tadeu Covas; Elaine Cristina Marqueze; Elaine Vieira Santos; Evandra Strazza Rodrigues; Heidge Fukumasu; Jayme Augusto de Souza-Neto; José Salvatore Leister Patané; Luiz Alcantara; Luiz Lehmann Coutinho; Maria Carolina Elias; Maurício Lacerda Nogueira; Rafael dos Santos Bezerra; Raul Machado Neto; Rejane Maria Tommasini Grotto; Ricardo Haddad; Sandra Coccuzzo Sampaio Vessoni; Simone Kashima; Svetoslav Nanev Slavov; Vincent Louis Viala |
| EPI_ISL_2825980, EPI_ISL_2825981, EPI_ISL_2826003, EPI_ISL_2826004, EPI_ISL_2882291, EPI_ISL_2890943, EPI_ISL_2891723, EPI_ISL_2891724, EPI_ISL_2891725, EPI_ISL_2891748, EPI_ISL_2891769, EPI_ISL_2891824, EPI_ISL_2891826, EPI_ISL_2891829, EPI_ISL_2891837, EPI_ISL_2891839, EPI_ISL_2891846, EPI_ISL_2891855, EPI_ISL_2891886, EPI_ISL_2891920, EPI_ISL_2891930, EPI_ISL_2891939, EPI_ISL_2891951, EPI_ISL_2891986, EPI_ISL_2892018, EPI_ISL_2892127, EPI_ISL_2892222, EPI_ISL_2892223, EPI_ISL_2892226, EPI_ISL_2892228, EPI_ISL_2892233, EPI_ISL_2892261, EPI_ISL_2892323, EPI_ISL_2892337, EPI_ISL_2892339, EPI_ISL_2892343, EPI_ISL_2892357, EPI_ISL_2892359, EPI_ISL_2921703, EPI_ISL_2921765, EPI_ISL_2921766, EPI_ISL_2921767, EPI_ISL_2921827, EPI_ISL_2921828, EPI_ISL_2921837, EPI_ISL_2921838, EPI_ISL_2921839, EPI_ISL_2921840, EPI_ISL_2921841, EPI_ISL_2921862, EPI_ISL_2921887, EPI_ISL_2921892, EPI_ISL_2921915, EPI_ISL_2921947, EPI_ISL_2921948, EPI_ISL_2921975, EPI_ISL_2921977, EPI_ISL_2921978, EPI_ISL_2921979, EPI_ISL_2921980, EPI_ISL_2922040, EPI_ISL_2922063, EPI_ISL_2922138, EPI_ISL_2922139, EPI_ISL_2922154, EPI_ISL_2922177, EPI_ISL_2922178, EPI_ISL_2922205, EPI_ISL_2922206, EPI_ISL_2922208, EPI_ISL_2922279, EPI_ISL_2923616, EPI_ISL_2976970, EPI_ISL_2976984, EPI_ISL_2976988, EPI_ISL_2977007, EPI_ISL_2977020, EPI_ISL_2977025, EPI_ISL_2977046, EPI_ISL_2977053, EPI_ISL_2977067, EPI_ISL_2977068, EPI_ISL_2977075, EPI_ISL_2977076, EPI_ISL_2977095, EPI_ISL_2977096, EPI_ISL_2977108, EPI_ISL_2977148, EPI_ISL_2977166, EPI_ISL_2977170, EPI_ISL_2977173, EPI_ISL_2977198, EPI_ISL_2977205, EPI_ISL_2977229, EPI_ISL_2977236, EPI_ISL_2977267, EPI_ISL_2977271, EPI_ISL_2977281, EPI_ISL_2977288, EPI_ISL_2977299, EPI_ISL_2977307, EPI_ISL_2977313, EPI_ISL_2977315, EPI_ISL_2977319, EPI_ISL_2977322, EPI_ISL_2977326, EPI_ISL_2977339, EPI_ISL_2977346, EPI_ISL_2977356, EPI_ISL_2977361, EPI_ISL_2977372, EPI_ISL_2977378, EPI_ISL_2977385, EPI_ISL_2977391, EPI_ISL_2977393, EPI_ISL_2978154, EPI_ISL_3243752, EPI_ISL_3243757, EPI_ISL_3243767, EPI_ISL_3243785, EPI_ISL_3243810, EPI_ISL_3243836, EPI_ISL_3243837, EPI_ISL_3243847, EPI_ISL_3243855, EPI_ISL_3243857, EPI_ISL_3243875, EPI_ISL_3243878, EPI_ISL_3243879, EPI_ISL_3243902, EPI_ISL_3243909, EPI_ISL_3243912, EPI_ISL_3243914, EPI_ISL_3243921, EPI_ISL_3243941, EPI_ISL_3243956, EPI_ISL_3243960, EPI_ISL_3244010, EPI_ISL_3244011, EPI_ISL_3244013, EPI_ISL_3244022, EPI_ISL_3244026, EPI_ISL_3244053, EPI_ISL_3244061, EPI_ISL_3244065, EPI_ISL_3244066, EPI_ISL_3244081, EPI_ISL_3244106, EPI_ISL_3244113, EPI_ISL_3244126, EPI_ISL_3244145, EPI_ISL_3244147, EPI_ISL_3244149, EPI_ISL_3244160, EPI_ISL_3244171, EPI_ISL_3244173, EPI_ISL_3244178, EPI_ISL_3244180, EPI_ISL_3244193, EPI_ISL_3244201, EPI_ISL_3244222, EPI_ISL_3244231, EPI_ISL_3244237, EPI_ISL_3244246, EPI_ISL_3244266, EPI_ISL_3244277, EPI_ISL_3244288, EPI_ISL_3244296, EPI_ISL_3244300, EPI_ISL_3244311, EPI_ISL_3244320, EPI_ISL_3244363, EPI_ISL_3244375, EPI_ISL_3244408, EPI_ISL_3244439, EPI_ISL_3244448 | | | |
| see above | CDPH VBL | California Department of Public Health | CDPH-COVIDNet; Emily Smith on behalf of CDPH-COVIDNet and UCSD EXCITE lab; UCLA Technology Center for Genomics & Bioinformatics; UCSD EXCITE lab |
| EPI_ISL_2097252 | CENTERS LAB NJ, LLC (DBA MEDLABS DIAGNOSTICS) | Wadsworth Center, New York State Department of Health | Alexis Russell; Catharine Prussing; Daryl M. Lamson; Erasmus Schneider; Erica Lasek-Nesselquist; John Kelly; Jonathan Plitnick; Kirsten St. George; Matthew Shudt; Melissa A Leisner; Navjot Singh |
| EPI_ISL_2344961, EPI_ISL_2344963, EPI_ISL_2344967, EPI_ISL_2344974, EPI_ISL_2344981, EPI_ISL_2344984, EPI_ISL_2344986, EPI_ISL_2344991, EPI_ISL_2344992, EPI_ISL_2344993, EPI_ISL_2344994, EPI_ISL_2344996, EPI_ISL_2344998 | | | |
| see above | CENTRO DE APOIO EPIDEMIOLOGICO | Instituto Butantan / ESALQ-Piracicaba | Antonio Jorge Martins; Claudia Renata dos Santos Barros; David Schlesinger; Debora Botequio Moretti; Dimas Tadeu Covas; Elaine Cristina Marqueze; Elaine Vieira Santos; Evandra Strazza Rodrigues; Heidge Fukumasu; Jayme Augusto de Souza-Neto; José Salvatore Leister Patané; Luiz Alcantara; Luiz Lehmann Coutinho; Maria Carolina Elias; Maurício Lacerda Nogueira; Rafael dos Santos Bezerra; Raul Machado Neto; Rejane Maria Tommasini Grotto; Ricardo Haddad; Sandra Coccuzzo Sampaio Vessoni; Simone Kashima; Svetoslav Nanev Slavov; Vincent Louis Viala |
| EPI_ISL_2209086, EPI_ISL_2209721, EPI_ISL_2345689, EPI_ISL_2345735, EPI_ISL_2345779, EPI_ISL_2345830, EPI_ISL_2345832, EPI_ISL_2345878 | | | |
| see above | CENTRO DE APOIO EPIDEMIOLOGICO | Instituto Butantan / Mendelics | Antonio Jorge Martins; Claudia Renata dos Santos Barros; David Schlesinger; Debora Botequio Moretti; Dimas Tadeu Covas; Elaine Cristina Marqueze; Elaine Vieira Santos; Evandra Strazza Rodrigues; Heidge Fukumasu; Jayme Augusto de Souza-Neto; José Salvatore Leister Patané; Luiz Alcantara; Luiz Lehmann Coutinho; Maria Carolina Elias; Maurício Lacerda Nogueira; Rafael dos Santos Bezerra; Raul Machado Neto; Rejane Maria Tommasini Grotto; Ricardo Haddad; Sandra Coccuzzo Sampaio Vessoni; Simone Kashima; Svetoslav Nanev Slavov; Vincent Louis Viala |
| EPI_ISL_3102309, EPI_ISL_3102329, EPI_ISL_3102511, EPI_ISL_3102512, EPI_ISL_3102515 | CENTRO DE ATENDIMENTO PARA ENFRENTAMENTO A COVID 19 | Analytical Competence Molecular Epidemiology Lab/ACME, Oswaldo Cruz Foundation, Ceara (FIOCRUZ CE) | Cleber Furtado Aksenen; Fabio Miyajima; Fernando Braga Stehling; Francisco Eder de Moura Lopes; Jamille Maria Mendes Bezerra; Joaquim César do Nascimento Sousa Junior; Pedro Miguel Carneiro Jeronimo; Suzana Porto Almeida e Lucas Delerino; Thais Ferreira de Oliveira; Thais de Oliveira Costa; Ticiane Cavalcante de Souza; Veridiana Pessoa Miyajima |
| EPI_ISL_2801337, EPI_ISL_2801341, EPI_ISL_2801343, EPI_ISL_2801344, EPI_ISL_2801346, EPI_ISL_2801363, EPI_ISL_3102270, EPI_ISL_3102277, EPI_ISL_3102519 | | | |
| see above | CENTRO DE ATENDIMENTO PARA ENFRENTAMENTO AO COVID 19 | Analytical Competence Molecular Epidemiology Lab/ACME, Oswaldo Cruz Foundation, Ceara (FIOCRUZ CE) | Cleber Furtado Aksenen; Cleber Furtado Aksenen e Suzana Porto Almeida; Fabio Miyajima; Fernando Braga Stehling; Francisco Eder de Moura Lopes; Jamille Maria Mendes Bezerra; Joaquim César do Nascimento Sousa Junior; Pedro Miguel Carneiro Jeronimo; Suzana Porto Almeida e Lucas Delerino; Thais Ferreira de Oliveira; Thais de Oliveira Costa; Ticiane Cavalcante de Souza; Veridiana Pessoa Miyajima |
| EPI_ISL_2493548 | CENTRO DE ESPECIALIDADES DE AGUAI AGUAI | Instituto Butantan | Antonio Jorge Martins; Claudia Renata dos Santos Barros; David Schlesinger; Debora Botequio Moretti; Dimas Tadeu Covas; Elaine Cristina Marqueze; Elaine Vieira Santos; Evandra Strazza Rodrigues; Heidge Fukumasu; Jayme Augusto de Souza-Neto; José Salvatore Leister Patané; Luiz Alcantara; Luiz Lehmann Coutinho; Maria Carolina Elias; Maurício Lacerda Nogueira; Rafael dos Santos Bezerra; Raul Machado Neto; Rejane Maria Tommasini Grotto; Ricardo Haddad; Sandra Coccuzzo Sampaio Vessoni; Simone Kashima; Svetoslav Nanev Slavov; Vincent Louis Viala |
| EPI_ISL_2445551 | CENTRO DE ESPECIALIDADES DE PRIMAVERA | Instituto Butantan | Antonio Jorge Martins; Claudia Renata dos Santos Barros; David Schlesinger; Debora Botequio Moretti; Dimas Tadeu Covas; Elaine Cristina Marqueze; Elaine Vieira Santos; Evandra Strazza Rodrigues; Heidge Fukumasu; Jayme Augusto de Souza-Neto; José Salvatore Leister Patané; Luiz Alcantara; Luiz Lehmann Coutinho; Maria Carolina Elias; Maurício Lacerda Nogueira; Rafael dos Santos Bezerra; Raul Machado Neto; Rejane Maria Tommasini Grotto; Ricardo Haddad; Sandra Coccuzzo Sampaio Vessoni; Simone Kashima; Svetoslav Nanev Slavov; Vincent Louis Viala |
| EPI_ISL_2210219 | CENTRO DE ESPECIALIDADES DE PRIMAVERA | Instituto Butantan / Mendelics | Antonio Jorge Martins; Claudia Renata dos Santos Barros; David Schlesinger; Debora Botequio Moretti; Dimas Tadeu Covas; Elaine Cristina Marqueze; Elaine Vieira Santos; Evandra Strazza Rodrigues; Heidge Fukumasu; Jayme Augusto de Souza-Neto; José Salvatore Leister Patané; Luiz Alcantara; Luiz Lehmann Coutinho; Maria Carolina Elias; Maurício Lacerda Nogueira; Rafael dos Santos Bezerra; Raul Machado Neto; Rejane Maria Tommasini Grotto; Ricardo Haddad; Sandra Coccuzzo Sampaio Vessoni; Simone Kashima; Svetoslav Nanev Slavov; Vincent Louis Viala |
| EPI_ISL_2445539 | CENTRO DE ESPECIALIDADES MARIAS DA GLORIA | Instituto Butantan | Antonio Jorge Martins; Claudia Renata dos Santos Barros; David Schlesinger; Debora Botequio Moretti; Dimas Tadeu Covas; Elaine Cristina Marqueze; Elaine Vieira Santos; Evandra Strazza Rodrigues; Heidge Fukumasu; Jayme Augusto de Souza-Neto; José Salvatore Leister Patané; Luiz Alcantara; Luiz Lehmann Coutinho; Maria Carolina Elias; Maurício Lacerda Nogueira; Rafael dos Santos Bezerra; Raul Machado Neto; Rejane Maria Tommasini Grotto; Ricardo Haddad; Sandra Coccuzzo Sampaio Vessoni; Simone Kashima; Svetoslav Nanev Slavov; Vincent Louis Viala |
| EPI_ISL_2493375 | CENTRO DE ESPECIALIDDS MEDICAS IRMA LEOPOLDINA PIRASSUNUNGA | Instituto Butantan | Antonio Jorge Martins; Claudia Renata dos Santos Barros; David Schlesinger; Debora Botequio Moretti; Dimas Tadeu Covas; Elaine Cristina Marqueze; Elaine Vieira Santos; Evandra Strazza Rodrigues; Heidge Fukumasu; Jayme Augusto de Souza-Neto; José Salvatore Leister Patané; Luiz Alcantara; Luiz Lehmann Coutinho; Maria Carolina Elias; Maurício Lacerda Nogueira; Rafael dos Santos Bezerra; Raul Machado Neto; Rejane Maria Tommasini Grotto; Ricardo Haddad; Sandra Coccuzzo Sampaio Vessoni; Simone Kashima; Svetoslav Nanev Slavov; Vincent Louis Viala |
| EPI_ISL_2493436, EPI_ISL_2493437, EPI_ISL_2493438 | CENTRO DE GENOMAS SUL | Instituto Butantan | Antonio Jorge Martins; Claudia Renata dos Santos Barros; David Schlesinger; Debora Botequio Moretti; Dimas Tadeu Covas; Elaine Cristina Marqueze; Elaine Vieira Santos; Evandra Strazza Rodrigues; Heidge Fukumasu; Jayme Augusto de Souza-Neto; José Salvatore Leister Patané; Luiz Alcantara; Luiz Lehmann Coutinho; Maria Carolina Elias; Maurício Lacerda Nogueira; Rafael dos Santos Bezerra; Raul Machado Neto; Rejane Maria Tommasini Grotto; Ricardo Haddad; Sandra Coccuzzo Sampaio Vessoni; Simone Kashima; Svetoslav Nanev Slavov; Vincent Louis Viala |
| EPI_ISL_2345815, EPI_ISL_2345860, EPI_ISL_2345865 | CENTRO DE REFERENCIA DO IDOSO DR HUMBERTO MENDES DE CARVALHO | Instituto Butantan / Mendelics | Antonio Jorge Martins; Claudia Renata dos Santos Barros; David Schlesinger; Debora Botequio Moretti; Dimas Tadeu Covas; Elaine Cristina Marqueze; Elaine Vieira Santos; Evandra Strazza Rodrigues; Heidge Fukumasu; Jayme Augusto de Souza-Neto; José Salvatore Leister Patané; Luiz Alcantara; Luiz Lehmann Coutinho; Maria Carolina Elias; Maurício Lacerda Nogueira; Rafael dos Santos Bezerra; Raul Machado Neto; Rejane Maria Tommasini Grotto; Ricardo Haddad; Sandra Coccuzzo Sampaio Vessoni; Simone Kashima; Svetoslav Nanev Slavov; Vincent Louis Viala |
| EPI_ISL_2493394 | CENTRO DE SAUDE | Instituto Butantan | Antonio Jorge Martins; Claudia Renata dos Santos Barros; David Schlesinger; Debora Botequio Moretti; Dimas Tadeu Covas; Elaine Cristina Marqueze; Elaine Vieira Santos; Evandra Strazza Rodrigues; Heidge Fukumasu; Jayme Augusto de Souza-Neto; José Salvatore Leister Patané; Luiz Alcantara; Luiz Lehmann Coutinho; Maria Carolina Elias; Maurício Lacerda Nogueira; Rafael dos Santos Bezerra; Raul Machado Neto; Rejane Maria Tommasini Grotto; Ricardo Haddad; Sandra Coccuzzo Sampaio Vessoni; Simone Kashima; Svetoslav Nanev Slavov; Vincent Louis Viala |
| EPI_ISL_2344852 | CENTRO DE SAUDE DE ANALANDIA | Instituto Butantan / ESALQ-Piracicaba | Antonio Jorge Martins; Claudia Renata dos Santos Barros; David Schlesinger; Debora Botequio Moretti; Dimas Tadeu Covas; Elaine Cristina Marqueze; Elaine Vieira Santos; Evandra Strazza Rodrigues; Heidge Fukumasu; Jayme Augusto de Souza-Neto; José Salvatore Leister Patané; Luiz Alcantara; Luiz Lehmann Coutinho; Maria Carolina Elias; Maurício Lacerda Nogueira; Rafael dos Santos Bezerra; Raul Machado Neto; Rejane Maria Tommasini Grotto; Ricardo Haddad; Sandra Coccuzzo Sampaio Vessoni; Simone Kashima; Svetoslav Nanev Slavov; Vincent Louis Viala |
| EPI_ISL_2345222 | CENTRO DE SAUDE DE AVAI | Instituto Butantan / ESALQ-Piracicaba | Antonio Jorge Martins; Claudia Renata dos Santos Barros; David Schlesinger; Debora Botequio Moretti; Dimas Tadeu Covas; Elaine Cristina Marqueze; Elaine Vieira Santos; Evandra Strazza Rodrigues; Heidge Fukumasu; Jayme Augusto de Souza-Neto; José Salvatore Leister Patané; Luiz Alcantara; Luiz Lehmann Coutinho; Maria Carolina Elias; Maurício Lacerda Nogueira; Rafael dos Santos Bezerra; Raul Machado Neto; Rejane Maria Tommasini Grotto; Ricardo Haddad; Sandra Coccuzzo Sampaio Vessoni; Simone Kashima; Svetoslav Nanev Slavov; Vincent Louis Viala |
| EPI_ISL_1967212 | CENTRO DE SAUDE DE BARBOSA | Instituto Butantan / Mendelics | Antonio Jorge Martins; Bianca Cechetto Carlos. Mendelics: Bibiana Santos; Claudia Renata dos Santos Barros; Cíntia Bittar; David Schlesinger. Hemocentro Ribeirão Preto: Simone Kashima; Debora Botequio Moretti; Elaine Cristina Marqueze; Elaine Vieira dos Santos; Elisangela Chicaroni Mattos; Erika Freitas; Evandra Strazza Rodrigues; Felipe Allan da Silva da Costa; Flavia Aburjaile; Fábio Sossai Possebon; Guilherme Campos; Guilherme Targino Valente; Heidge Fukumasu. USP-Botucatu: Rejane Maria Tommasini Grotto; Helena Lage Ferreira; Instituto Butantan: Dimas Tadeu Covas; Jardelina de Souza Todao Bernardino; Jayme A. Souza-Neto; Jessika Cristina Chagas Lesbon; Jorge A. Petroli Marchesi; José Salvatore Leister Patané; João Paulo Kitajima; João Pessoa Araújo Jr.; Leila Sabrina Ullmann; Loyze Paola Oliveira de Lima; Luiz Aurelio de Campos Crispin. Centro de Genômica Funcional da ESALQ: Luiz Lehmann Coutinho; Luiz Carlos Junior de Alcantara; Lívia Sacchetto; Maisa C. Pereira Parra; Maria Carolina Elias; Marta Giovanetti; Marília Moraes; Maurício Lacerda Nogueira. Prefeitura de Sao Paulo: Melissa Palmieri.; Patricia Akemi Assato; Paula Rahal; Paulo Inacio da Costa; Rafael dos Santos Bezerra; Raquel de Lello Rocha Campos Cassano. NGS Soluções Genômicas: Pilar Drummond Sampaio Corrêa Mariani. FZEA-USP Pirassununga: Mirele Daiana Poleti; Raul Machado Neto; Ricardo Augusto Brassaloti; Ricardo Haddad; Rodrigo Tocantins Calado. FAMERP-SJRP: Cecília Artico Banho; Sandra Coccuzzo Sampaio; Svetoslav Nanev Slavov; Vagner Fonseca; Vincent Louis Viala |

| | | | |
|---|---|---|---|
| EPI_ISL_2473820 | CENTRO DE SAUDE DE CANITAR | Instituto Butantan | Antonio Jorge Martins; Claudia Renata dos Santos Barros; David Schlesinger; Debora Botequio Moretti; Dimas Tadeu Covas; Elaine Cristina Marqueze; Elaine Vieira Santos; Evandra Strazza Rodrigues; Heidge Fukumasu; Jayme Augusto de Souza-Neto; José Salvatore Leister Patané; Luiz Alcantara; Luiz Lehmann Coutinho; Maria Carolina Elias; Maurício Lacerda Nogueira; Rafael dos Santos Bezerra; Raul Machado Neto; Rejane Maria Tommasini Grotto; Ricardo Haddad; Sandra Coccuzzo Sampaio Vessoni; Simone Kashima; Svetoslav Nanev Slavov; Vincent Louis Viala |
| EPI_ISL_2445197 | CENTRO DE SAUDE DE CRUZALIA | Instituto Butantan | Antonio Jorge Martins; Claudia Renata dos Santos Barros; David Schlesinger; Debora Botequio Moretti; Dimas Tadeu Covas; Elaine Cristina Marqueze; Elaine Vieira Santos; Evandra Strazza Rodrigues; Heidge Fukumasu; Jayme Augusto de Souza-Neto; José Salvatore Leister Patané; Luiz Alcantara; Luiz Lehmann Coutinho; Maria Carolina Elias; Maurício Lacerda Nogueira; Rafael dos Santos Bezerra; Raul Machado Neto; Rejane Maria Tommasini Grotto; Ricardo Haddad; Sandra Coccuzzo Sampaio Vessoni; Simone Kashima; Svetoslav Nanev Slavov; Vincent Louis Viala |
| EPI_ISL_2445153 | CENTRO DE SAUDE DE FLORIDA PAULISTA | Instituto Butantan | Antonio Jorge Martins; Claudia Renata dos Santos Barros; David Schlesinger; Debora Botequio Moretti; Dimas Tadeu Covas; Elaine Cristina Marqueze; Elaine Vieira Santos; Evandra Strazza Rodrigues; Heidge Fukumasu; Jayme Augusto de Souza-Neto; José Salvatore Leister Patané; Luiz Alcantara; Luiz Lehmann Coutinho; Maria Carolina Elias; Maurício Lacerda Nogueira; Rafael dos Santos Bezerra; Raul Machado Neto; Rejane Maria Tommasini Grotto; Ricardo Haddad; Sandra Coccuzzo Sampaio Vessoni; Simone Kashima; Svetoslav Nanev Slavov; Vincent Louis Viala |
| EPI_ISL_2473713 | CENTRO DE SAUDE DE IBIRAREMA | Instituto Butantan | Antonio Jorge Martins; Claudia Renata dos Santos Barros; David Schlesinger; Debora Botequio Moretti; Dimas Tadeu Covas; Elaine Cristina Marqueze; Elaine Vieira Santos; Evandra Strazza Rodrigues; Heidge Fukumasu; Jayme Augusto de Souza-Neto; José Salvatore Leister Patané; Luiz Alcantara; Luiz Lehmann Coutinho; Maria Carolina Elias; Maurício Lacerda Nogueira; Rafael dos Santos Bezerra; Raul Machado Neto; Rejane Maria Tommasini Grotto; Ricardo Haddad; Sandra Coccuzzo Sampaio Vessoni; Simone Kashima; Svetoslav Nanev Slavov; Vincent Louis Viala |
| EPI_ISL_2445428, EPI_ISL_2445429, EPI_ISL_2445430, EPI_ISL_2445431, EPI_ISL_2445433, EPI_ISL_2445434, EPI_ISL_2493586 | | | |
| see above | CENTRO DE SAUDE DE LUCELIA | Instituto Butantan | Antonio Jorge Martins; Claudia Renata dos Santos Barros; David Schlesinger; Debora Botequio Moretti; Dimas Tadeu Covas; Elaine Cristina Marqueze; Elaine Vieira Santos; Evandra Strazza Rodrigues; Heidge Fukumasu; Jayme Augusto de Souza-Neto; José Salvatore Leister Patané; Luiz Alcantara; Luiz Lehmann Coutinho; Maria Carolina Elias; Maurício Lacerda Nogueira; Rafael dos Santos Bezerra; Raul Machado Neto; Rejane Maria Tommasini Grotto; Ricardo Haddad; Sandra Coccuzzo Sampaio Vessoni; Simone Kashima; Svetoslav Nanev Slavov; Vincent Louis Viala |
| EPI_ISL_2209593, EPI_ISL_2345657, EPI_ISL_2345702, EPI_ISL_2345753 | CENTRO DE SAUDE DE MARACAI | Instituto Butantan / Mendelics | Antonio Jorge Martins; Claudia Renata dos Santos Barros; David Schlesinger; Debora Botequio Moretti; Dimas Tadeu Covas; Elaine Cristina Marqueze; Elaine Vieira Santos; Evandra Strazza Rodrigues; Heidge Fukumasu; Jayme Augusto de Souza-Neto; José Salvatore Leister Patané; Luiz Alcantara; Luiz Lehmann Coutinho; Maria Carolina Elias; Maurício Lacerda Nogueira; Rafael dos Santos Bezerra; Raul Machado Neto; Rejane Maria Tommasini Grotto; Ricardo Haddad; Sandra Coccuzzo Sampaio Vessoni; Simone Kashima; Svetoslav Nanev Slavov; Vincent Louis Viala |
| EPI_ISL_2345236 | CENTRO DE SAUDE DE OCAUCU | Instituto Butantan / ESALQ-Piracicaba | Antonio Jorge Martins; Claudia Renata dos Santos Barros; David Schlesinger; Debora Botequio Moretti; Dimas Tadeu Covas; Elaine Cristina Marqueze; Elaine Vieira Santos; Evandra Strazza Rodrigues; Heidge Fukumasu; Jayme Augusto de Souza-Neto; José Salvatore Leister Patané; Luiz Alcantara; Luiz Lehmann Coutinho; Maria Carolina Elias; Maurício Lacerda Nogueira; Rafael dos Santos Bezerra; Raul Machado Neto; Rejane Maria Tommasini Grotto; Ricardo Haddad; Sandra Coccuzzo Sampaio Vessoni; Simone Kashima; Svetoslav Nanev Slavov; Vincent Louis Viala |
| EPI_ISL_2345192, EPI_ISL_2345201 | CENTRO DE SAUDE DE OSVALDO CRUZ | Instituto Butantan / ESALQ-Piracicaba | Antonio Jorge Martins; Claudia Renata dos Santos Barros; David Schlesinger; Debora Botequio Moretti; Dimas Tadeu Covas; Elaine Cristina Marqueze; Elaine Vieira Santos; Evandra Strazza Rodrigues; Heidge Fukumasu; Jayme Augusto de Souza-Neto; José Salvatore Leister Patané; Luiz Alcantara; Luiz Lehmann Coutinho; Maria Carolina Elias; Maurício Lacerda Nogueira; Rafael dos Santos Bezerra; Raul Machado Neto; Rejane Maria Tommasini Grotto; Ricardo Haddad; Sandra Coccuzzo Sampaio Vessoni; Simone Kashima; Svetoslav Nanev Slavov; Vincent Louis Viala |
| EPI_ISL_2473744, EPI_ISL_2473745 | CENTRO DE SAUDE DE PIRAJUI | Instituto Butantan | Antonio Jorge Martins; Claudia Renata dos Santos Barros; David Schlesinger; Debora Botequio Moretti; Dimas Tadeu Covas; Elaine Cristina Marqueze; Elaine Vieira Santos; Evandra Strazza Rodrigues; Heidge Fukumasu; Jayme Augusto de Souza-Neto; José Salvatore Leister Patané; Luiz Alcantara; Luiz Lehmann Coutinho; Maria Carolina Elias; Maurício Lacerda Nogueira; Rafael dos Santos Bezerra; Raul Machado Neto; Rejane Maria Tommasini Grotto; Ricardo Haddad; Sandra Coccuzzo Sampaio Vessoni; Simone Kashima; Svetoslav Nanev Slavov; Vincent Louis Viala |
| EPI_ISL_2445193, EPI_ISL_2445194 | CENTRO DE SAUDE DE RIBEIRAO DO SUL | Instituto Butantan | Antonio Jorge Martins; Claudia Renata dos Santos Barros; David Schlesinger; Debora Botequio Moretti; Dimas Tadeu Covas; Elaine Cristina Marqueze; Elaine Vieira Santos; Evandra Strazza Rodrigues; Heidge Fukumasu; Jayme Augusto de Souza-Neto; José Salvatore Leister Patané; Luiz Alcantara; Luiz Lehmann Coutinho; Maria Carolina Elias; Maurício Lacerda Nogueira; Rafael dos Santos Bezerra; Raul Machado Neto; Rejane Maria Tommasini Grotto; Ricardo Haddad; Sandra Coccuzzo Sampaio Vessoni; Simone Kashima; Svetoslav Nanev Slavov; Vincent Louis Viala |
| EPI_ISL_2209288, EPI_ISL_2345730, EPI_ISL_2345775, EPI_ISL_2362246 | CENTRO DE SAUDE DR JOAO JOAQUIM FERREIRA CRISTAIS PAULISTA | Instituto Butantan / Mendelics | Antonio Jorge Martins; Claudia Renata dos Santos Barros; David Schlesinger; Debora Botequio Moretti; Dimas Tadeu Covas; Elaine Cristina Marqueze; Elaine Vieira Santos; Evandra Strazza Rodrigues; Heidge Fukumasu; Jayme Augusto de Souza-Neto; José Salvatore Leister Patané; Luiz Alcantara; Luiz Lehmann Coutinho; Maria Carolina Elias; Maurício Lacerda Nogueira; Rafael dos Santos Bezerra; Raul Machado Neto; Rejane Maria Tommasini Grotto; Ricardo Haddad; Sandra Coccuzzo Sampaio Vessoni; Simone Kashima; Svetoslav Nanev Slavov; Vincent Louis Viala |
| EPI_ISL_2493354 | CENTRO DE SAUDE DR JOSE AGOSTINHO NOGUEIRA PAULICEIA | Instituto Butantan | Antonio Jorge Martins; Claudia Renata dos Santos Barros; David Schlesinger; Debora Botequio Moretti; Dimas Tadeu Covas; Elaine Cristina Marqueze; Elaine Vieira Santos; Evandra Strazza Rodrigues; Heidge Fukumasu; Jayme Augusto de Souza-Neto; José Salvatore Leister Patané; Luiz Alcantara; Luiz Lehmann Coutinho; Maria Carolina Elias; Maurício Lacerda Nogueira; Rafael dos Santos Bezerra; Raul Machado Neto; Rejane Maria Tommasini Grotto; Ricardo Haddad; Sandra Coccuzzo Sampaio Vessoni; Simone Kashima; Svetoslav Nanev Slavov; Vincent Louis Viala |
| EPI_ISL_2493377, EPI_ISL_2493378, EPI_ISL_2494150 | CENTRO DE SAUDE DR MARIO DIAS DE AGUIAR CAPIVARI | Instituto Butantan | Antonio Jorge Martins; Claudia Renata dos Santos Barros; David Schlesinger; Debora Botequio Moretti; Dimas Tadeu Covas; Elaine Cristina Marqueze; Elaine Vieira Santos; Evandra Strazza Rodrigues; Heidge Fukumasu; Jayme Augusto de Souza-Neto; José Salvatore Leister Patané; Luiz Alcantara; Luiz Lehmann Coutinho; Maria Carolina Elias; Maurício Lacerda Nogueira; Rafael dos Santos Bezerra; Raul Machado Neto; Rejane Maria Tommasini Grotto; Ricardo Haddad; Sandra Coccuzzo Sampaio Vessoni; Simone Kashima; Svetoslav Nanev Slavov; Vincent Louis Viala |
| EPI_ISL_2445441, EPI_ISL_2445442, EPI_ISL_2445445, EPI_ISL_2445450, EPI_ISL_2445453 | CENTRO DE SAUDE DR NELSON DA CUNHA BASTOS | Instituto Butantan | Antonio Jorge Martins; Claudia Renata dos Santos Barros; David Schlesinger; Debora Botequio Moretti; Dimas Tadeu Covas; Elaine Cristina Marqueze; Elaine Vieira Santos; Evandra Strazza Rodrigues; Heidge Fukumasu; Jayme Augusto de Souza-Neto; José Salvatore Leister Patané; Luiz Alcantara; Luiz Lehmann Coutinho; Maria Carolina Elias; Maurício Lacerda Nogueira; Rafael dos Santos Bezerra; Raul Machado Neto; Rejane Maria Tommasini Grotto; Ricardo Haddad; Sandra Coccuzzo Sampaio Vessoni; Simone Kashima; Svetoslav Nanev Slavov; Vincent Louis Viala |
| EPI_ISL_2344832, EPI_ISL_2344873, EPI_ISL_2344904, EPI_ISL_2344905, EPI_ISL_2344907 | CENTRO DE SAUDE DR RENATO DE CARVALHO RIBEIRO ANGATUBA | Instituto Butantan / ESALQ-Piracicaba | Antonio Jorge Martins; Claudia Renata dos Santos Barros; David Schlesinger; Debora Botequio Moretti; Dimas Tadeu Covas; Elaine Cristina Marqueze; Elaine Vieira Santos; Evandra Strazza Rodrigues; Heidge Fukumasu; Jayme Augusto de Souza-Neto; José Salvatore Leister Patané; Luiz Alcantara; Luiz Lehmann Coutinho; Maria Carolina Elias; Maurício Lacerda Nogueira; Rafael dos Santos Bezerra; Raul Machado Neto; Rejane Maria Tommasini Grotto; Ricardo Haddad; Sandra Coccuzzo Sampaio Vessoni; Simone Kashima; Svetoslav Nanev Slavov; Vincent Louis Viala |
| EPI_ISL_2473787 | CENTRO DE SAUDE II CARDOSO | Instituto Butantan | Antonio Jorge Martins; Claudia Renata dos Santos Barros; David Schlesinger; Debora Botequio Moretti; Dimas Tadeu Covas; Elaine Cristina Marqueze; Elaine Vieira Santos; Evandra Strazza Rodrigues; Heidge Fukumasu; Jayme Augusto de Souza-Neto; José Salvatore Leister Patané; Luiz Alcantara; Luiz Lehmann Coutinho; Maria Carolina Elias; Maurício Lacerda Nogueira; Rafael dos Santos Bezerra; Raul Machado Neto; Rejane Maria Tommasini Grotto; Ricardo Haddad; Sandra Coccuzzo Sampaio Vessoni; Simone Kashima; Svetoslav Nanev Slavov; Vincent Louis Viala |
| EPI_ISL_2445529, EPI_ISL_2445531, EPI_ISL_2445533 | CENTRO DE SAUDE II DE POTIRENDABA CESARINO BENFATTI | Instituto Butantan | Antonio Jorge Martins; Claudia Renata dos Santos Barros; David Schlesinger; Debora Botequio Moretti; Dimas Tadeu Covas; Elaine Cristina Marqueze; Elaine Vieira Santos; Evandra Strazza Rodrigues; Heidge Fukumasu; Jayme Augusto de Souza-Neto; José Salvatore Leister Patané; Luiz Alcantara; Luiz Lehmann Coutinho; Maria Carolina Elias; Maurício Lacerda Nogueira; Rafael dos Santos Bezerra; Raul Machado Neto; Rejane Maria Tommasini Grotto; Ricardo Haddad; Sandra Coccuzzo Sampaio Vessoni; Simone Kashima; Svetoslav Nanev Slavov; Vincent Louis Viala |
| EPI_ISL_2493643 | CENTRO DE SAUDE II DR GABRIEL MESQUITA VARGEM GDE DO SUL | Instituto Butantan | Antonio Jorge Martins; Claudia Renata dos Santos Barros; David Schlesinger; Debora Botequio Moretti; Dimas Tadeu Covas; Elaine Cristina Marqueze; Elaine Vieira Santos; Evandra Strazza Rodrigues; Heidge Fukumasu; Jayme Augusto de Souza-Neto; José Salvatore Leister Patané; Luiz Alcantara; Luiz Lehmann Coutinho; Maria Carolina Elias; Maurício Lacerda Nogueira; Rafael dos Santos Bezerra; Raul Machado Neto; Rejane Maria Tommasini Grotto; Ricardo Haddad; Sandra Coccuzzo Sampaio Vessoni; Simone Kashima; Svetoslav Nanev Slavov; Vincent Louis Viala |
| EPI_ISL_2345650, EPI_ISL_2345695, EPI_ISL_2362262 | CENTRO DE SAUDE II DR GABRIEL MESQUITA VARGEM GDE DO SUL | Instituto Butantan / Mendelics | Antonio Jorge Martins; Claudia Renata dos Santos Barros; David Schlesinger; Debora Botequio Moretti; Dimas Tadeu Covas; Elaine Cristina Marqueze; Elaine Vieira Santos; Evandra Strazza Rodrigues; Heidge Fukumasu; Jayme Augusto de Souza-Neto; José Salvatore Leister Patané; Luiz Alcantara; Luiz Lehmann Coutinho; Maria Carolina Elias; Maurício Lacerda Nogueira; Rafael dos Santos Bezerra; Raul Machado Neto; Rejane Maria Tommasini Grotto; Ricardo Haddad; Sandra Coccuzzo Sampaio Vessoni; Simone Kashima; Svetoslav Nanev Slavov; Vincent Louis Viala |
| EPI_ISL_2494160 | CENTRO DE SAUDE II DR JOSE DE FELIPE E S PINHAL SP | Instituto Butantan | Antonio Jorge Martins; Claudia Renata dos Santos Barros; David Schlesinger; Debora Botequio Moretti; Dimas Tadeu Covas; Elaine Cristina Marqueze; Elaine Vieira Santos; Evandra Strazza Rodrigues; Heidge Fukumasu; Jayme Augusto de Souza-Neto; José Salvatore Leister Patané; Luiz Alcantara; Luiz Lehmann Coutinho; Maria Carolina Elias; Maurício Lacerda Nogueira; Rafael dos Santos Bezerra; Raul Machado Neto; Rejane Maria Tommasini Grotto; Ricardo Haddad; Sandra Coccuzzo Sampaio Vessoni; Simone Kashima; Svetoslav Nanev Slavov; Vincent Louis Viala |
| EPI_ISL_2445317, EPI_ISL_2493537, EPI_ISL_2493540, EPI_ISL_2493542, EPI_ISL_2493571, EPI_ISL_2493572, EPI_ISL_2493574, EPI_ISL_2493575, EPI_ISL_2493576, EPI_ISL_2493578, EPI_ISL_2493580, EPI_ISL_2493582, EPI_ISL_2493583, EPI_ISL_2493675, EPI_ISL_2493676, EPI_ISL_2493680, EPI_ISL_2493682, EPI_ISL_2493683 | | | |
| see above | CENTRO DE SAUDE II IBITINGA | Instituto Butantan | Antonio Jorge Martins; Claudia Renata dos Santos Barros; David Schlesinger; Debora Botequio Moretti; Dimas Tadeu Covas; Elaine Cristina Marqueze; Elaine Vieira Santos; Evandra Strazza Rodrigues; Heidge Fukumasu; Jayme Augusto de Souza-Neto; José Salvatore Leister Patané; Luiz Alcantara; Luiz Lehmann Coutinho; Maria Carolina Elias; Maurício Lacerda Nogueira; Rafael dos Santos Bezerra; Raul Machado Neto; Rejane Maria Tommasini Grotto; Ricardo Haddad; Sandra Coccuzzo Sampaio Vessoni; Simone Kashima; Svetoslav Nanev Slavov; Vincent Louis Viala |
| EPI_ISL_2209353, EPI_ISL_2209368, EPI_ISL_2209527, EPI_ISL_2345644, EPI_ISL_2345649, EPI_ISL_2345724, EPI_ISL_2345743, EPI_ISL_2345747, EPI_ISL_2345766, EPI_ISL_2345791, EPI_ISL_2345861 | | | |
| see above | CENTRO DE SAUDE II IBITINGA | Instituto Butantan / Mendelics | Antonio Jorge Martins; Claudia Renata dos Santos Barros; David Schlesinger; Debora Botequio Moretti; Dimas Tadeu Covas; Elaine Cristina Marqueze; Elaine Vieira Santos; Evandra Strazza Rodrigues; Heidge Fukumasu; Jayme Augusto de Souza-Neto; José Salvatore Leister Patané; Luiz Alcantara; Luiz Lehmann Coutinho; Maria Carolina Elias; Maurício Lacerda Nogueira; Rafael dos Santos Bezerra; Raul Machado Neto; Rejane Maria Tommasini Grotto; Ricardo Haddad; Sandra Coccuzzo Sampaio Vessoni; Simone Kashima; Svetoslav Nanev Slavov; Vincent Louis Viala |
| EPI_ISL_2493336, EPI_ISL_2493337, EPI_ISL_2493338, EPI_ISL_2493339, EPI_ISL_2493344, EPI_ISL_2493347 | CENTRO DE SAUDE II JUNQUEIROPOLIS | Instituto Butantan | Antonio Jorge Martins; Claudia Renata dos Santos Barros; David Schlesinger; Debora Botequio Moretti; Dimas Tadeu Covas; Elaine Cristina Marqueze; Elaine Vieira Santos; Evandra Strazza Rodrigues; Heidge Fukumasu; Jayme Augusto de Souza-Neto; José Salvatore Leister Patané; Luiz Alcantara; Luiz Lehmann Coutinho; Maria Carolina Elias; Maurício Lacerda Nogueira; Rafael dos Santos Bezerra; Raul Machado Neto; Rejane Maria Tommasini Grotto; Ricardo Haddad; Sandra Coccuzzo Sampaio Vessoni; Simone Kashima; Svetoslav Nanev Slavov; Vincent Louis Viala |
| EPI_ISL_2445603 | CENTRO DE SAUDE II SAO MIGUEL ARCANJO | Instituto Butantan | Antonio Jorge Martins; Claudia Renata dos Santos Barros; David Schlesinger; Debora Botequio Moretti; Dimas Tadeu Covas; Elaine Cristina Marqueze; Elaine Vieira Santos; Evandra Strazza Rodrigues; Heidge Fukumasu; Jayme Augusto de Souza-Neto; José Salvatore Leister Patané; Luiz Alcantara; Luiz Lehmann Coutinho; Maria Carolina Elias; Maurício Lacerda Nogueira; Rafael dos Santos Bezerra; Raul Machado Neto; Rejane Maria Tommasini Grotto; Ricardo Haddad; Sandra Coccuzzo Sampaio Vessoni; Simone Kashima; Svetoslav Nanev Slavov; Vincent Louis Viala |
| EPI_ISL_2493263 | CENTRO DE SAUDE III AFFONSO LUZZI SANTA CRUZ DAS PALMEIRAS | Instituto Butantan | Antonio Jorge Martins; Claudia Renata dos Santos Barros; David Schlesinger; Debora Botequio Moretti; Dimas Tadeu Covas; Elaine Cristina Marqueze; Elaine Vieira Santos; Evandra Strazza Rodrigues; Heidge Fukumasu; Jayme Augusto de Souza-Neto; José Salvatore Leister Patané; Luiz Alcantara; Luiz Lehmann Coutinho; Maria Carolina Elias; Maurício Lacerda Nogueira; Rafael dos Santos Bezerra; Raul Machado Neto; Rejane Maria Tommasini Grotto; Ricardo Haddad; Sandra Coccuzzo Sampaio Vessoni; Simone Kashima; Svetoslav Nanev Slavov; Vincent Louis Viala |
| EPI_ISL_2209307, EPI_ISL_2209312, EPI_ISL_2345792, EPI_ISL_2362260 | CENTRO DE SAUDE III AFFONSO LUZZI SANTA CRUZ DAS PALMEIRAS | Instituto Butantan / Mendelics | Antonio Jorge Martins; Claudia Renata dos Santos Barros; David Schlesinger; Debora Botequio Moretti; Dimas Tadeu Covas; Elaine Cristina Marqueze; Elaine Vieira Santos; Evandra Strazza Rodrigues; Heidge Fukumasu; Jayme Augusto de Souza-Neto; José Salvatore Leister Patané; Luiz Alcantara; Luiz Lehmann Coutinho; Maria Carolina Elias; Maurício Lacerda Nogueira; Rafael dos Santos Bezerra; Raul Machado Neto; Rejane Maria Tommasini Grotto; Ricardo Haddad; Sandra Coccuzzo Sampaio Vessoni; Simone Kashima; Svetoslav Nanev Slavov; Vincent Louis Viala |
| EPI_ISL_2345195 | CENTRO DE SAUDE III ALTO ALEGRE | Instituto Butantan / ESALQ-Piracicaba | Antonio Jorge Martins; Claudia Renata dos Santos Barros; David Schlesinger; Debora Botequio Moretti; Dimas Tadeu Covas; Elaine Cristina Marqueze; Elaine Vieira Santos; Evandra Strazza Rodrigues; Heidge Fukumasu; Jayme Augusto de Souza-Neto; José Salvatore Leister Patané; Luiz Alcantara; Luiz Lehmann Coutinho; Maria Carolina Elias; Maurício Lacerda Nogueira; Rafael dos Santos Bezerra; Raul Machado Neto; Rejane Maria Tommasini Grotto; Ricardo Haddad; Sandra Coccuzzo Sampaio Vessoni; Simone Kashima; Svetoslav Nanev Slavov; Vincent Louis Viala |
| EPI_ISL_2493539, EPI_ISL_2493544, EPI_ISL_2493547, EPI_ISL_2493570 | CENTRO DE SAUDE III BORBOREMA | Instituto Butantan | Antonio Jorge Martins; Claudia Renata dos Santos Barros; David Schlesinger; Debora Botequio Moretti; Dimas Tadeu Covas; Elaine Cristina Marqueze; Elaine Vieira Santos; Evandra Strazza Rodrigues; Heidge Fukumasu; Jayme Augusto de Souza-Neto; José Salvatore Leister Patané; Luiz Alcantara; Luiz Lehmann Coutinho; Maria Carolina Elias; Maurício Lacerda Nogueira; Rafael dos Santos Bezerra; Raul Machado Neto; Rejane Maria Tommasini Grotto; Ricardo Haddad; Sandra Coccuzzo Sampaio Vessoni; Simone Kashima; Svetoslav Nanev Slavov; Vincent Louis Viala |
| EPI_ISL_2494317, EPI_ISL_2494332 | CENTRO DE SAUDE III DE AGUAS DE SAO PEDRO | Instituto Butantan | Antonio Jorge Martins; Claudia Renata dos Santos Barros; David Schlesinger; Debora Botequio Moretti; Dimas Tadeu Covas; Elaine Cristina Marqueze; Elaine Vieira Santos; Evandra Strazza Rodrigues; Heidge Fukumasu; Jayme Augusto de Souza-Neto; José Salvatore Leister Patané; Luiz Alcantara; Luiz Lehmann Coutinho; Maria Carolina Elias; Maurício Lacerda Nogueira; Rafael dos Santos Bezerra; Raul Machado Neto; Rejane Maria Tommasini Grotto; Ricardo Haddad; Sandra Coccuzzo Sampaio Vessoni; Simone Kashima; Svetoslav Nanev Slavov; Vincent Louis Viala |
| EPI_ISL_2345188 | CENTRO DE SAUDE III DE BOCAINA | Instituto Butantan / ESALQ-Piracicaba | Antonio Jorge Martins; Claudia Renata dos Santos Barros; David Schlesinger; Debora Botequio Moretti; Dimas Tadeu Covas; Elaine Cristina Marqueze; Elaine Vieira Santos; Evandra Strazza Rodrigues; Heidge Fukumasu; Jayme Augusto de Souza-Neto; José Salvatore Leister Patané; Luiz Alcantara; Luiz Lehmann Coutinho; Maria Carolina Elias; Maurício Lacerda Nogueira; Rafael dos Santos Bezerra; Raul Machado Neto; Rejane Maria Tommasini Grotto; Ricardo Haddad; Sandra Coccuzzo Sampaio Vessoni; Simone Kashima; Svetoslav Nanev Slavov; Vincent Louis Viala |
| EPI_ISL_2493669 | CENTRO DE SAUDE III DOBRADA | Instituto Butantan | Antonio Jorge Martins; Claudia Renata dos Santos Barros; David Schlesinger; Debora Botequio Moretti; Dimas Tadeu Covas; Elaine Cristina Marqueze; Elaine Vieira Santos; Evandra Strazza Rodrigues; Heidge Fukumasu; Jayme Augusto de Souza-Neto; José Salvatore Leister Patané; Luiz Alcantara; Luiz Lehmann Coutinho; Maria Carolina Elias; Maurício Lacerda Nogueira; Rafael dos Santos Bezerra; Raul Machado Neto; Rejane Maria Tommasini Grotto; Ricardo Haddad; Sandra Coccuzzo Sampaio Vessoni; Simone Kashima; Svetoslav Nanev Slavov; Vincent Louis Viala |
| EPI_ISL_2209702 | CENTRO DE SAUDE III NELCIDIO DA SILVEIRA BASTOS | Instituto Butantan / Mendelics | Antonio Jorge Martins; Claudia Renata dos Santos Barros; David Schlesinger; Debora Botequio Moretti; Dimas Tadeu Covas; Elaine Cristina Marqueze; Elaine Vieira Santos; Evandra Strazza Rodrigues; Heidge Fukumasu; Jayme Augusto de Souza-Neto; José Salvatore Leister Patané; Luiz Alcantara; Luiz Lehmann Coutinho; Maria Carolina Elias; Maurício Lacerda Nogueira; Rafael dos Santos Bezerra; Raul Machado Neto; Rejane Maria Tommasini Grotto; Ricardo Haddad; Sandra Coccuzzo Sampaio Vessoni; Simone Kashima; Svetoslav Nanev Slavov; Vincent Louis Viala |
| EPI_ISL_2208938, EPI_ISL_2209968, EPI_ISL_2210119, EPI_ISL_2345697, EPI_ISL_2345842 | CENTRO DE SAUDE III SALES OLIVEIRA | Instituto Butantan / Mendelics | Antonio Jorge Martins; Claudia Renata dos Santos Barros; David Schlesinger; Debora Botequio Moretti; Dimas Tadeu Covas; Elaine Cristina Marqueze; Elaine Vieira Santos; Evandra Strazza Rodrigues; Heidge Fukumasu; Jayme Augusto de Souza-Neto; José Salvatore Leister Patané; Luiz Alcantara; Luiz Lehmann Coutinho; Maria Carolina Elias; Maurício Lacerda Nogueira; Rafael dos Santos Bezerra; Raul Machado Neto; Rejane Maria Tommasini Grotto; Ricardo Haddad; Sandra Coccuzzo Sampaio Vessoni; Simone Kashima; Svetoslav Nanev Slavov; Vincent Louis Viala |
| EPI_ISL_2493677 | CENTRO DE SAUDE III TABATINGA | Instituto Butantan | Antonio Jorge Martins; Claudia Renata dos Santos Barros; David Schlesinger; Debora Botequio Moretti; Dimas Tadeu Covas; Elaine Cristina Marqueze; Elaine Vieira Santos; Evandra Strazza Rodrigues; Heidge Fukumasu; Jayme Augusto de Souza-Neto; José Salvatore Leister Patané; Luiz Alcantara; Luiz Lehmann |

| | | | |
|---|---|---|---|
| | | | Coutinho; Maria Carolina Elias; Maurício Lacerda Nogueira; Rafael dos Santos Bezerra; Raul Machado Neto; Rejane Maria Tommasini Grotto; Ricardo Haddad; Sandra Coccuzzo Sampaio Vessoni; Simone Kashima; Svetoslav Nanev Slavov; Vincent Louis Viala |
| EPI_ISL_2445360, EPI_ISL_2445365, EPI_ISL_2445448, EPI_ISL_2445451, EPI_ISL_2445452 | CENTRO INTEGRADO DE SAUDE | Instituto Butantan | Antonio Jorge Martins; Claudia Renata dos Santos Barros; David Schlesinger; Debora Botequio Moretti; Dimas Tadeu Covas; Elaine Cristina Marqueze; Elaine Vieira Santos; Evandra Strazza Rodrigues; Heidge Fukumasu; Jayme Augusto de Souza-Neto; José Salvatore Leister Patané; Luiz Alcantara; Luiz Lehmann Coutinho; Maria Carolina Elias; Maurício Lacerda Nogueira; Rafael dos Santos Bezerra; Raul Machado Neto; Rejane Maria Tommasini Grotto; Ricardo Haddad; Sandra Coccuzzo Sampaio Vessoni; Simone Kashima; Svetoslav Nanev Slavov; Vincent Louis Viala |
| EPI_ISL_2493292, EPI_ISL_2493300 | CENTRO MEDICO PMESP | Instituto Butantan | Antonio Jorge Martins; Claudia Renata dos Santos Barros; David Schlesinger; Debora Botequio Moretti; Dimas Tadeu Covas; Elaine Cristina Marqueze; Elaine Vieira Santos; Evandra Strazza Rodrigues; Heidge Fukumasu; Jayme Augusto de Souza-Neto; José Salvatore Leister Patané; Luiz Alcantara; Luiz Lehmann Coutinho; Maria Carolina Elias; Maurício Lacerda Nogueira; Rafael dos Santos Bezerra; Raul Machado Neto; Rejane Maria Tommasini Grotto; Ricardo Haddad; Sandra Coccuzzo Sampaio Vessoni; Simone Kashima; Svetoslav Nanev Slavov; Vincent Louis Viala |
| EPI_ISL_2345129 | CENTRO MEDICO PMESP | Instituto Butantan / ESALQ-Piracicaba | Antonio Jorge Martins; Claudia Renata dos Santos Barros; David Schlesinger; Debora Botequio Moretti; Dimas Tadeu Covas; Elaine Cristina Marqueze; Elaine Vieira Santos; Evandra Strazza Rodrigues; Heidge Fukumasu; Jayme Augusto de Souza-Neto; José Salvatore Leister Patané; Luiz Alcantara; Luiz Lehmann Coutinho; Maria Carolina Elias; Maurício Lacerda Nogueira; Rafael dos Santos Bezerra; Raul Machado Neto; Rejane Maria Tommasini Grotto; Ricardo Haddad; Sandra Coccuzzo Sampaio Vessoni; Simone Kashima; Svetoslav Nanev Slavov; Vincent Louis Viala |
| EPI_ISL_2493597, EPI_ISL_2493600, EPI_ISL_2493606, EPI_ISL_2493625 | CENTRO MEDICO SOCIAL COMUNITARIO JANUARIO TEODORO DE SOUZA | Instituto Butantan | Antonio Jorge Martins; Claudia Renata dos Santos Barros; David Schlesinger; Debora Botequio Moretti; Dimas Tadeu Covas; Elaine Cristina Marqueze; Elaine Vieira Santos; Evandra Strazza Rodrigues; Heidge Fukumasu; Jayme Augusto de Souza-Neto; José Salvatore Leister Patané; Luiz Alcantara; Luiz Lehmann Coutinho; Maria Carolina Elias; Maurício Lacerda Nogueira; Rafael dos Santos Bezerra; Raul Machado Neto; Rejane Maria Tommasini Grotto; Ricardo Haddad; Sandra Coccuzzo Sampaio Vessoni; Simone Kashima; Svetoslav Nanev Slavov; Vincent Louis Viala |
| EPI_ISL_2493433, EPI_ISL_2493445 | CENTRO SAUDE III DR JOSE TADEU MOURAO | Instituto Butantan | Antonio Jorge Martins; Claudia Renata dos Santos Barros; David Schlesinger; Debora Botequio Moretti; Dimas Tadeu Covas; Elaine Cristina Marqueze; Elaine Vieira Santos; Evandra Strazza Rodrigues; Heidge Fukumasu; Jayme Augusto de Souza-Neto; José Salvatore Leister Patané; Luiz Alcantara; Luiz Lehmann Coutinho; Maria Carolina Elias; Maurício Lacerda Nogueira; Rafael dos Santos Bezerra; Raul Machado Neto; Rejane Maria Tommasini Grotto; Ricardo Haddad; Sandra Coccuzzo Sampaio Vessoni; Simone Kashima; Svetoslav Nanev Slavov; Vincent Louis Viala |
| EPI_ISL_2614611, EPI_ISL_2614612 | CETESB - Companhia Ambiental do Estado de Sao Paulo | Instituto Adolfo Lutz, Interdiciplinary Procedures Center, Strategic Laboratory | Caio Vinicius Dias Lopes; Claudia Regina Gonçalves; Claudio Tavares Sacchi; Erica Valessa Ramos Gomes; Karoline Rodrigues Campos; Leonardo Jose Tadeu de Araujo; Luisa Zanolli Moreno; Maria Ines Zanoli Sato; Mikaela R. F. Barbosa |
| EPI_ISL_2015908 | CH DE SAINTONGE | CNR Virus des Infections Respiratoires - France SUD | Antonin Bal; Bruno Lina; Gregory Destras; Gwendolyne Burfin; Hadrien Regue; Laurence Josset; Martine Valette; Quentin Semanas |
| EPI_ISL_2627846 | CHC Andrée Rosemon | Institut Pasteur de la Guyane | Anne Lavergne; Antoine Enfissi; Arielle Salmier; Dominique Rousset |
| EPI_ISL_2248789, EPI_ISL_2248792 | CHEDV | Instituto Nacional de Saude (INSA) | Borges et al |
| EPI_ISL_2004290, EPI_ISL_2248827, EPI_ISL_2362105, EPI_ISL_2362114 | CHLC | Instituto Nacional de Saude (INSA) | Borges et al |
| EPI_ISL_2510151 | CHUGA-IBP-laboratoire de Virologie | IBP-laboratoire de virologie | Alban Caporossi; Anne Signori -Schmuck; Anne-Karen Faure; Aurélie Truffot; Benjamin Nemoz; Hugo Jardin; Julien Andréani; Julien Lupo; Léa Ponderand; Pascal Poignard; Patrice Morand; Raphaële Germi; Sylvie Larrat |
| EPI_ISL_1941821 | CHUV | Laboratory of genomics and metagenomics | Claire Bertelli; Damien Jacot; Gilbert Greub; Sebastien Aeby; Trestan Pillonel |
| EPI_ISL_2533931 | CIAD-Hermosillo | Immunology Laboratory | ; Alejandra García-Gasca; Alejandra Hernández-Terán; Alejandro Sánchez-Flores; Alfredo Herrera-Estrella; Alicia Ocaña-Mondragón; Andreu Comas-García; Angel Gustavo Salas-Lais; Antonio Loza Román; Bernardo Martínez-Miguel; Blanca Taboada; Brenda Irasema Maldonado-Meza; Bruno Gómez-Gil; Carla Ivón Herrera-Najera; Carlos F. Arias; Celia Boukadida; Clara Esperanza Santacruz-Tinoco; Concepción Grajales-Muñiz; Consorcio Mexicano de Vigilancia Genómica (CoViGen-Mex). Authors (in alphabetical order): Julio Elias Alvarado-Yaah; Cristóbal Cháidez-Quiróz; Célida Duque Molina; Célida Martínez-Rodríguez; Daniel Fregoso-Rueda; Daniel Lira Morales; Eduardo Becerril-Vargas; Fernando Fontove-Herrera; Fidencio Mejía-Nepomuceno; Francisco Pulido; Gloria Elena Espinosa-Ayala; Gloria María Molina-Salinas; Gloria Vazquez; Hector Esteban Paz-Juárez; Hector Montoya-Fuentes; Helen Haydee Fernanda Ramírez-Plascencia; Irvin González-López; Jean Pierre González; Jesús Hernández; Joel Armando Vázquez-Pérez.; Jorge Salas-Hernández; José Antonio Enciso-Moreno; José Arturo Martínez-Orozco; José Esteban Muñoz-Medina; José de Jesús Nuñez-Contreras; Juan Bautista Chale-Dzul; Julissa Enciso-Ibarra; Luis Alberto Ochoa-Carrera; Margarita Matías-Florentino; Mario Mújica-Sánchez; Marissa Perez-Garcia; María Guadalupe Santiago-Mauricio; María Guadalupe de Jesús Mireles-Rivera; Nelly Sélem-Mojica; Pavel Isa; Ricardo Ciria Merce; Ricardo Grande; Rosa María Gutiérrez Rios; Santiago Ávila-Ríos; Selene Zárate; Susana Lopez; Verónica Mata-Haro; Victor Eduardo García-Arias; Víctor Hugo Borja-Aburto |
| EPI_ISL_2017661, EPI_ISL_2017665, EPI_ISL_2017668, EPI_ISL_2351958, EPI_ISL_2351959, EPI_ISL_2351960, EPI_ISL_2351964, EPI_ISL_2351968, EPI_ISL_2351969, EPI_ISL_2351970, EPI_ISL_2351978, EPI_ISL_2516497, EPI_ISL_2516531 | | | |
| see above | CLILAB | Microbiology Department | Aida Gonzalez-Diaz; Carmen Ardanuy; Jordi Camara; Jordi Niubó; Laura Calatayud; M Angeles Domínguez; Miguel Fernandez-Huerta; Sara Marti |
| EPI_ISL_3355515 | CLINICA PUTUMAYO | Corporacion CorpoGen-Universidad de los Andes-Universidad Central | Christian Romero; Jorge Duitama; Juan Manuel Anzola; Laura González; Maryam Chaib De Mares; María Mercedes Zambrano; Nelly Díaz; Patricia Del Portillo; Silvia Restrepo |
| EPI_ISL_2362548 | CNR Institut Pasteur de la Guyane | Institut Pasteur de la Guyane | Anne Lavergne; Antoine Enfissi; Arielle Salmier; Dominique Rousset |
| EPI_ISL_2373756 | CNR Virus des Infections Respiratoires - France SUD | CNR Virus des Infections Respiratoires - France SUD | Antonin Bal; Bruno Lina; Gregory Destras; Gwendolyne Burfin; Hadrien Regue; Laurence Josset; Martine Valette; Quentin Semanas |
| EPI_ISL_2493384, EPI_ISL_2493385, EPI_ISL_2493446, EPI_ISL_2494153 | COORDENADORIA MUNICIPAL DE SAUDE DE IRACEMAPOLIS | Instituto Butantan | Antonio Jorge Martins; Claudia Renata dos Santos Barros; David Schlesinger; Debora Botequio Moretti; Dimas Tadeu Covas; Elaine Cristina Marqueze; Elaine Vieira Santos; Evandra Strazza Rodrigues; Heidge Fukumasu; Jayme Augusto de Souza-Neto; José Salvatore Leister Patané; Luiz Alcantara; Luiz Lehmann Coutinho; Maria Carolina Elias; Maurício Lacerda Nogueira; Rafael dos Santos Bezerra; Raul Machado Neto; Rejane Maria Tommasini Grotto; Ricardo Haddad; Sandra Coccuzzo Sampaio Vessoni; Simone Kashima; Svetoslav Nanev Slavov; Vincent Louis Viala |
| EPI_ISL_2345111 | COVID 19 CENTRO DE COMBATE AO CORONAVIRUS CCC JANDIRA | Instituto Butantan / ESALQ-Piracicaba | Antonio Jorge Martins; Claudia Renata dos Santos Barros; David Schlesinger; Debora Botequio Moretti; Dimas Tadeu Covas; Elaine Cristina Marqueze; Elaine Vieira Santos; Evandra Strazza Rodrigues; Heidge Fukumasu; Jayme Augusto de Souza-Neto; José Salvatore Leister Patané; Luiz Alcantara; Luiz Lehmann Coutinho; Maria Carolina Elias; Maurício Lacerda Nogueira; Rafael dos Santos Bezerra; Raul Machado Neto; Rejane Maria Tommasini Grotto; Ricardo Haddad; Sandra Coccuzzo Sampaio Vessoni; Simone Kashima; Svetoslav Nanev Slavov; Vincent Louis Viala |
| EPI_ISL_2378754 | CS DE APARECIDA DOESTE | Instituto Butantan | Antonio Jorge Martins; Claudia Renata dos Santos Barros; David Schlesinger; Debora Botequio Moretti; Dimas Tadeu Covas; Elaine Cristina Marqueze; Elaine Vieira Santos; Evandra Strazza Rodrigues; Heidge Fukumasu; Jayme Augusto de Souza-Neto; José Salvatore Leister Patané; Luiz Alcantara; Luiz Lehmann Coutinho; Maria Carolina Elias; Maurício Lacerda Nogueira; Rafael dos Santos Bezerra; Raul Machado Neto; Rejane Maria Tommasini Grotto; Ricardo Haddad; Sandra Coccuzzo Sampaio Vessoni; Simone Kashima; Svetoslav Nanev Slavov; Vincent Louis Viala |
| EPI_ISL_2445209, EPI_ISL_2445210, EPI_ISL_2473720 | CS DE BALSAMO | Instituto Butantan | Antonio Jorge Martins; Claudia Renata dos Santos Barros; David Schlesinger; Debora Botequio Moretti; Dimas Tadeu Covas; Elaine Cristina Marqueze; Elaine Vieira Santos; Evandra Strazza Rodrigues; Heidge Fukumasu; Jayme Augusto de Souza-Neto; José Salvatore Leister Patané; Luiz Alcantara; Luiz Lehmann Coutinho; Maria Carolina Elias; Maurício Lacerda Nogueira; Rafael dos Santos Bezerra; Raul Machado Neto; Rejane Maria Tommasini Grotto; Ricardo Haddad; Sandra Coccuzzo Sampaio Vessoni; Simone Kashima; Svetoslav Nanev Slavov; Vincent Louis Viala |
| EPI_ISL_2344863 | CS DE BALSAMO | Instituto Butantan / ESALQ-Piracicaba | Antonio Jorge Martins; Claudia Renata dos Santos Barros; David Schlesinger; Debora Botequio Moretti; Dimas Tadeu Covas; Elaine Cristina Marqueze; Elaine Vieira Santos; Evandra Strazza Rodrigues; Heidge Fukumasu; Jayme Augusto de Souza-Neto; José Salvatore Leister Patané; Luiz Alcantara; Luiz Lehmann Coutinho; Maria Carolina Elias; Maurício Lacerda Nogueira; Rafael dos Santos Bezerra; Raul Machado Neto; Rejane Maria Tommasini Grotto; Ricardo Haddad; Sandra Coccuzzo Sampaio Vessoni; Simone Kashima; Svetoslav Nanev Slavov; Vincent Louis Viala |
| EPI_ISL_2344823, EPI_ISL_2344824, EPI_ISL_2344857, EPI_ISL_2344860 | CS DE CATIGUA | Instituto Butantan / ESALQ-Piracicaba | Antonio Jorge Martins; Claudia Renata dos Santos Barros; David Schlesinger; Debora Botequio Moretti; Dimas Tadeu Covas; Elaine Cristina Marqueze; Elaine Vieira Santos; Evandra Strazza Rodrigues; Heidge Fukumasu; Jayme Augusto de Souza-Neto; José Salvatore Leister Patané; Luiz Alcantara; Luiz Lehmann Coutinho; Maria Carolina Elias; Maurício Lacerda Nogueira; Rafael dos Santos Bezerra; Raul Machado Neto; Rejane Maria Tommasini Grotto; Ricardo Haddad; Sandra Coccuzzo Sampaio Vessoni; Simone Kashima; Svetoslav Nanev Slavov; Vincent Louis Viala |
| EPI_ISL_2445212, EPI_ISL_2494297 | CS DE COSMORAMA | Instituto Butantan | Antonio Jorge Martins; Claudia Renata dos Santos Barros; David Schlesinger; Debora Botequio Moretti; Dimas Tadeu Covas; Elaine Cristina Marqueze; Elaine Vieira Santos; Evandra Strazza Rodrigues; Heidge Fukumasu; Jayme Augusto de Souza-Neto; José Salvatore Leister Patané; Luiz Alcantara; Luiz Lehmann Coutinho; Maria Carolina Elias; Maurício Lacerda Nogueira; Rafael dos Santos Bezerra; Raul Machado Neto; Rejane Maria Tommasini Grotto; Ricardo Haddad; Sandra Coccuzzo Sampaio Vessoni; Simone Kashima; Svetoslav Nanev Slavov; Vincent Louis Viala |
| EPI_ISL_2344871, EPI_ISL_2344874 | CS DE COSMORAMA | Instituto Butantan / ESALQ-Piracicaba | Antonio Jorge Martins; Claudia Renata dos Santos Barros; David Schlesinger; Debora Botequio Moretti; Dimas Tadeu Covas; Elaine Cristina Marqueze; Elaine Vieira Santos; Evandra Strazza Rodrigues; Heidge Fukumasu; Jayme Augusto de Souza-Neto; José Salvatore Leister Patané; Luiz Alcantara; Luiz Lehmann Coutinho; Maria Carolina Elias; Maurício Lacerda Nogueira; Rafael dos Santos Bezerra; Raul Machado Neto; Rejane Maria Tommasini Grotto; Ricardo Haddad; Sandra Coccuzzo Sampaio Vessoni; Simone Kashima; Svetoslav Nanev Slavov; Vincent Louis Viala |
| EPI_ISL_2344882 | CS DE MARAPOAMA | Instituto Butantan / ESALQ-Piracicaba | Antonio Jorge Martins; Claudia Renata dos Santos Barros; David Schlesinger; Debora Botequio Moretti; Dimas Tadeu Covas; Elaine Cristina Marqueze; Elaine Vieira Santos; Evandra Strazza Rodrigues; Heidge Fukumasu; Jayme Augusto de Souza-Neto; José Salvatore Leister Patané; Luiz Alcantara; Luiz Lehmann Coutinho; Maria Carolina Elias; Maurício Lacerda Nogueira; Rafael dos Santos Bezerra; Raul Machado Neto; Rejane Maria Tommasini Grotto; Ricardo Haddad; Sandra Coccuzzo Sampaio Vessoni; Simone Kashima; Svetoslav Nanev Slavov; Vincent Louis Viala |
| EPI_ISL_2493424 | CS DE MONTE APRAZIVEL | Instituto Butantan | Antonio Jorge Martins; Claudia Renata dos Santos Barros; David Schlesinger; Debora Botequio Moretti; Dimas Tadeu Covas; Elaine Cristina Marqueze; Elaine Vieira Santos; Evandra Strazza Rodrigues; Heidge Fukumasu; Jayme Augusto de Souza-Neto; José Salvatore Leister Patané; Luiz Alcantara; Luiz Lehmann Coutinho; Maria Carolina Elias; Maurício Lacerda Nogueira; Rafael dos Santos Bezerra; Raul Machado Neto; Rejane Maria Tommasini Grotto; Ricardo Haddad; Sandra Coccuzzo Sampaio Vessoni; Simone Kashima; Svetoslav Nanev Slavov; Vincent Louis Viala |
| EPI_ISL_2493405, EPI_ISL_2493406, EPI_ISL_2493407, EPI_ISL_2493412, EPI_ISL_2493414, EPI_ISL_2493416 | CS DE NEVES PAULISTA | Instituto Butantan | Antonio Jorge Martins; Claudia Renata dos Santos Barros; David Schlesinger; Debora Botequio Moretti; Dimas Tadeu Covas; Elaine Cristina Marqueze; Elaine Vieira Santos; Evandra Strazza Rodrigues; Heidge Fukumasu; Jayme Augusto de Souza-Neto; José Salvatore Leister Patané; Luiz Alcantara; Luiz Lehmann Coutinho; Maria Carolina Elias; Maurício Lacerda Nogueira; Rafael dos Santos Bezerra; Raul Machado Neto; Rejane Maria Tommasini Grotto; Ricardo Haddad; Sandra Coccuzzo Sampaio Vessoni; Simone Kashima; Svetoslav Nanev Slavov; Vincent Louis Viala |
| EPI_ISL_2445213, EPI_ISL_2445214, EPI_ISL_2445215, EPI_ISL_2445217, EPI_ISL_2445218, EPI_ISL_2473721, EPI_ISL_2473722, EPI_ISL_2493661, EPI_ISL_2493662 | | | |
| see above | CS DE NOVO HORIZONTE | Instituto Butantan | Antonio Jorge Martins; Claudia Renata dos Santos Barros; David Schlesinger; Debora Botequio Moretti; Dimas Tadeu Covas; Elaine Cristina Marqueze; Elaine Vieira Santos; Evandra Strazza Rodrigues; Heidge Fukumasu; Jayme Augusto de Souza-Neto; José Salvatore Leister Patané; Luiz Alcantara; Luiz Lehmann Coutinho; Maria Carolina Elias; Maurício Lacerda Nogueira; Rafael dos Santos Bezerra; Raul Machado Neto; Rejane Maria Tommasini Grotto; Ricardo Haddad; Sandra Coccuzzo Sampaio Vessoni; Simone Kashima; Svetoslav Nanev Slavov; Vincent Louis Viala |
| EPI_ISL_2344872, EPI_ISL_2344875, EPI_ISL_2344887, EPI_ISL_2344889 | CS DE ONDA VERDE | Instituto Butantan / ESALQ-Piracicaba | Antonio Jorge Martins; Claudia Renata dos Santos Barros; David Schlesinger; Debora Botequio Moretti; Dimas Tadeu Covas; Elaine Cristina Marqueze; Elaine Vieira Santos; Evandra Strazza Rodrigues; Heidge Fukumasu; Jayme Augusto de Souza-Neto; José Salvatore Leister Patané; Luiz Alcantara; Luiz Lehmann Coutinho; Maria Carolina Elias; Maurício Lacerda Nogueira; Rafael dos Santos Bezerra; Raul Machado Neto; Rejane Maria Tommasini Grotto; Ricardo Haddad; Sandra Coccuzzo Sampaio Vessoni; Simone Kashima; Svetoslav Nanev Slavov; Vincent Louis Viala |
| EPI_ISL_2493411, EPI_ISL_2493419, EPI_ISL_2493429 | CS DE ORINDIUVA | Instituto Butantan | Antonio Jorge Martins; Claudia Renata dos Santos Barros; David Schlesinger; Debora Botequio Moretti; Dimas Tadeu Covas; Elaine Cristina Marqueze; Elaine Vieira Santos; Evandra Strazza Rodrigues; Heidge Fukumasu; Jayme Augusto de Souza-Neto; José Salvatore Leister Patané; Luiz Alcantara; Luiz Lehmann Coutinho; Maria Carolina Elias; Maurício Lacerda Nogueira; Rafael dos Santos Bezerra; Raul Machado Neto; Rejane Maria Tommasini Grotto; Ricardo Haddad; Sandra Coccuzzo Sampaio Vessoni; Simone Kashima; Svetoslav Nanev Slavov; Vincent Louis Viala |
| EPI_ISL_2493410, EPI_ISL_2493413, EPI_ISL_2493415, EPI_ISL_2493417, EPI_ISL_2493420 | CS DE PALMARES PAULISTA | Instituto Butantan | Antonio Jorge Martins; Claudia Renata dos Santos Barros; David Schlesinger; Debora Botequio Moretti; Dimas Tadeu Covas; Elaine Cristina Marqueze; Elaine Vieira Santos; Evandra Strazza Rodrigues; Heidge Fukumasu; Jayme Augusto de Souza-Neto; José Salvatore Leister Patané; Luiz Alcantara; Luiz Lehmann Coutinho; Maria Carolina Elias; Maurício Lacerda Nogueira; Rafael dos Santos Bezerra; Raul Machado Neto; Rejane Maria Tommasini Grotto; Ricardo Haddad; Sandra Coccuzzo Sampaio Vessoni; Simone Kashima; Svetoslav Nanev Slavov; Vincent Louis Viala |
| EPI_ISL_2209164, EPI_ISL_2344859 | CS DE PALMARES PAULISTA | Instituto Butantan / ESALQ-Piracicaba | Antonio Jorge Martins; Claudia Renata dos Santos Barros; David Schlesinger; Debora Botequio Moretti; Dimas Tadeu Covas; Elaine Cristina Marqueze; Elaine Vieira Santos; Evandra Strazza Rodrigues; Heidge Fukumasu; Jayme Augusto de Souza-Neto; José Salvatore Leister Patané; Luiz Alcantara; Luiz Lehmann Coutinho; Maria Carolina Elias; Maurício Lacerda Nogueira; Rafael dos Santos Bezerra; Raul Machado Neto; Rejane Maria Tommasini Grotto; Ricardo Haddad; Sandra Coccuzzo Sampaio Vessoni; Simone Kashima; Svetoslav Nanev Slavov; Vincent Louis Viala |
| EPI_ISL_2445211 | CS DE PLANALTO | Instituto Butantan | Antonio Jorge Martins; Claudia Renata dos Santos Barros; David Schlesinger; Debora Botequio Moretti; Dimas Tadeu Covas; Elaine Cristina Marqueze; Elaine Vieira Santos; Evandra Strazza Rodrigues; Heidge Fukumasu; Jayme Augusto de Souza-Neto; José Salvatore Leister Patané; Luiz Alcantara; Luiz Lehmann |

| | | | |
|---|---|---|---|
| | | | Coutinho; Maria Carolina Elias; Maurício Lacerda Nogueira; Rafael dos Santos Bezerra; Raul Machado Neto; Rejane Maria Tommasini Grotto; Ricardo Haddad; Sandra Coccuzzo Sampaio Vessoni; Simone Kashima; Svetoslav Nanev Slavov; Vincent Louis Viala |
| EPI_ISL_2493362 | CS DE ROSANA | Instituto Butantan | Antonio Jorge Martins; Claudia Renata dos Santos Barros; David Schlesinger; Debora Botequio Moretti; Dimas Tadeu Covas; Elaine Cristina Marqueze; Elaine Vieira Santos; Evandra Strazza Rodrigues; Heidge Fukumasu; Jayme Augusto de Souza-Neto; José Salvatore Leister Patané; Luiz Alcantara; Luiz Lehmann Coutinho; Maria Carolina Elias; Maurício Lacerda Nogueira; Rafael dos Santos Bezerra; Raul Machado Neto; Rejane Maria Tommasini Grotto; Ricardo Haddad; Sandra Coccuzzo Sampaio Vessoni; Simone Kashima; Svetoslav Nanev Slavov; Vincent Louis Viala |
| EPI_ISL_2493400 | CS DE SANTA ALBERTINA | Instituto Butantan | Antonio Jorge Martins; Claudia Renata dos Santos Barros; David Schlesinger; Debora Botequio Moretti; Dimas Tadeu Covas; Elaine Cristina Marqueze; Elaine Vieira Santos; Evandra Strazza Rodrigues; Heidge Fukumasu; Jayme Augusto de Souza-Neto; José Salvatore Leister Patané; Luiz Alcantara; Luiz Lehmann Coutinho; Maria Carolina Elias; Maurício Lacerda Nogueira; Rafael dos Santos Bezerra; Raul Machado Neto; Rejane Maria Tommasini Grotto; Ricardo Haddad; Sandra Coccuzzo Sampaio Vessoni; Simone Kashima; Svetoslav Nanev Slavov; Vincent Louis Viala |
| EPI_ISL_2344854, EPI_ISL_2344855, EPI_ISL_2344856 | CS DE SANTA ALBERTINA | Instituto Butantan / ESALQ-Piracicaba | Antonio Jorge Martins; Claudia Renata dos Santos Barros; David Schlesinger; Debora Botequio Moretti; Dimas Tadeu Covas; Elaine Cristina Marqueze; Elaine Vieira Santos; Evandra Strazza Rodrigues; Heidge Fukumasu; Jayme Augusto de Souza-Neto; José Salvatore Leister Patané; Luiz Alcantara; Luiz Lehmann Coutinho; Maria Carolina Elias; Maurício Lacerda Nogueira; Rafael dos Santos Bezerra; Raul Machado Neto; Rejane Maria Tommasini Grotto; Ricardo Haddad; Sandra Coccuzzo Sampaio Vessoni; Simone Kashima; Svetoslav Nanev Slavov; Vincent Louis Viala |
| EPI_ISL_2493421, EPI_ISL_2493422 | CS DE SANTA RITA DOESTE | Instituto Butantan | Antonio Jorge Martins; Claudia Renata dos Santos Barros; David Schlesinger; Debora Botequio Moretti; Dimas Tadeu Covas; Elaine Cristina Marqueze; Elaine Vieira Santos; Evandra Strazza Rodrigues; Heidge Fukumasu; Jayme Augusto de Souza-Neto; José Salvatore Leister Patané; Luiz Alcantara; Luiz Lehmann Coutinho; Maria Carolina Elias; Maurício Lacerda Nogueira; Rafael dos Santos Bezerra; Raul Machado Neto; Rejane Maria Tommasini Grotto; Ricardo Haddad; Sandra Coccuzzo Sampaio Vessoni; Simone Kashima; Svetoslav Nanev Slavov; Vincent Louis Viala |
| EPI_ISL_2493430, EPI_ISL_2493431 | CS DE VALENTIM GENTIL | Instituto Butantan | Antonio Jorge Martins; Claudia Renata dos Santos Barros; David Schlesinger; Debora Botequio Moretti; Dimas Tadeu Covas; Elaine Cristina Marqueze; Elaine Vieira Santos; Evandra Strazza Rodrigues; Heidge Fukumasu; Jayme Augusto de Souza-Neto; José Salvatore Leister Patané; Luiz Alcantara; Luiz Lehmann Coutinho; Maria Carolina Elias; Maurício Lacerda Nogueira; Rafael dos Santos Bezerra; Raul Machado Neto; Rejane Maria Tommasini Grotto; Ricardo Haddad; Sandra Coccuzzo Sampaio Vessoni; Simone Kashima; Svetoslav Nanev Slavov; Vincent Louis Viala |
| EPI_ISL_2493299 | CS II DE IGARAPAVA | Instituto Butantan | Antonio Jorge Martins; Claudia Renata dos Santos Barros; David Schlesinger; Debora Botequio Moretti; Dimas Tadeu Covas; Elaine Cristina Marqueze; Elaine Vieira Santos; Evandra Strazza Rodrigues; Heidge Fukumasu; Jayme Augusto de Souza-Neto; José Salvatore Leister Patané; Luiz Alcantara; Luiz Lehmann Coutinho; Maria Carolina Elias; Maurício Lacerda Nogueira; Rafael dos Santos Bezerra; Raul Machado Neto; Rejane Maria Tommasini Grotto; Ricardo Haddad; Sandra Coccuzzo Sampaio Vessoni; Simone Kashima; Svetoslav Nanev Slavov; Vincent Louis Viala |
| EPI_ISL_2445399, EPI_ISL_2445401 | CS II DR ANTONIO VICOSO MOREIRA DE REZENDE SUMARE | Instituto Butantan | Antonio Jorge Martins; Claudia Renata dos Santos Barros; David Schlesinger; Debora Botequio Moretti; Dimas Tadeu Covas; Elaine Cristina Marqueze; Elaine Vieira Santos; Evandra Strazza Rodrigues; Heidge Fukumasu; Jayme Augusto de Souza-Neto; José Salvatore Leister Patané; Luiz Alcantara; Luiz Lehmann Coutinho; Maria Carolina Elias; Maurício Lacerda Nogueira; Rafael dos Santos Bezerra; Raul Machado Neto; Rejane Maria Tommasini Grotto; Ricardo Haddad; Sandra Coccuzzo Sampaio Vessoni; Simone Kashima; Svetoslav Nanev Slavov; Vincent Louis Viala |
| EPI_ISL_2445455, EPI_ISL_2445458, EPI_ISL_2445459, EPI_ISL_2445461, EPI_ISL_2445462 | CS II DR JAHYR DE PAULA RIBEIRO GUARA | Instituto Butantan | Antonio Jorge Martins; Claudia Renata dos Santos Barros; David Schlesinger; Debora Botequio Moretti; Dimas Tadeu Covas; Elaine Cristina Marqueze; Elaine Vieira Santos; Evandra Strazza Rodrigues; Heidge Fukumasu; Jayme Augusto de Souza-Neto; José Salvatore Leister Patané; Luiz Alcantara; Luiz Lehmann Coutinho; Maria Carolina Elias; Maurício Lacerda Nogueira; Rafael dos Santos Bezerra; Raul Machado Neto; Rejane Maria Tommasini Grotto; Ricardo Haddad; Sandra Coccuzzo Sampaio Vessoni; Simone Kashima; Svetoslav Nanev Slavov; Vincent Louis Viala |
| EPI_ISL_2345262 | CS II DR JAHYR DE PAULA RIBEIRO GUARA | Instituto Butantan / ESALQ-Piracicaba | Antonio Jorge Martins; Claudia Renata dos Santos Barros; David Schlesinger; Debora Botequio Moretti; Dimas Tadeu Covas; Elaine Cristina Marqueze; Elaine Vieira Santos; Evandra Strazza Rodrigues; Heidge Fukumasu; Jayme Augusto de Souza-Neto; José Salvatore Leister Patané; Luiz Alcantara; Luiz Lehmann Coutinho; Maria Carolina Elias; Maurício Lacerda Nogueira; Rafael dos Santos Bezerra; Raul Machado Neto; Rejane Maria Tommasini Grotto; Ricardo Haddad; Sandra Coccuzzo Sampaio Vessoni; Simone Kashima; Svetoslav Nanev Slavov; Vincent Louis Viala |
| EPI_ISL_2445406 | CS II DR MIGUEL VITALIANO ORLANDIA | Instituto Butantan | Antonio Jorge Martins; Claudia Renata dos Santos Barros; David Schlesinger; Debora Botequio Moretti; Dimas Tadeu Covas; Elaine Cristina Marqueze; Elaine Vieira Santos; Evandra Strazza Rodrigues; Heidge Fukumasu; Jayme Augusto de Souza-Neto; José Salvatore Leister Patané; Luiz Alcantara; Luiz Lehmann Coutinho; Maria Carolina Elias; Maurício Lacerda Nogueira; Rafael dos Santos Bezerra; Raul Machado Neto; Rejane Maria Tommasini Grotto; Ricardo Haddad; Sandra Coccuzzo Sampaio Vessoni; Simone Kashima; Svetoslav Nanev Slavov; Vincent Louis Viala |
| EPI_ISL_2345078, EPI_ISL_2345194, EPI_ISL_2345198, EPI_ISL_2345200 | CS II DR MIGUEL VITALIANO ORLANDIA | Instituto Butantan / ESALQ-Piracicaba | Antonio Jorge Martins; Claudia Renata dos Santos Barros; David Schlesinger; Debora Botequio Moretti; Dimas Tadeu Covas; Elaine Cristina Marqueze; Elaine Vieira Santos; Evandra Strazza Rodrigues; Heidge Fukumasu; Jayme Augusto de Souza-Neto; José Salvatore Leister Patané; Luiz Alcantara; Luiz Lehmann Coutinho; Maria Carolina Elias; Maurício Lacerda Nogueira; Rafael dos Santos Bezerra; Raul Machado Neto; Rejane Maria Tommasini Grotto; Ricardo Haddad; Sandra Coccuzzo Sampaio Vessoni; Simone Kashima; Svetoslav Nanev Slavov; Vincent Louis Viala |
| EPI_ISL_2209850 | CS II DR SEBASTIAO RIBEIRO DO VALLE | Instituto Butantan / Mendelics | Antonio Jorge Martins; Claudia Renata dos Santos Barros; David Schlesinger; Debora Botequio Moretti; Dimas Tadeu Covas; Elaine Cristina Marqueze; Elaine Vieira Santos; Evandra Strazza Rodrigues; Heidge Fukumasu; Jayme Augusto de Souza-Neto; José Salvatore Leister Patané; Luiz Alcantara; Luiz Lehmann Coutinho; Maria Carolina Elias; Maurício Lacerda Nogueira; Rafael dos Santos Bezerra; Raul Machado Neto; Rejane Maria Tommasini Grotto; Ricardo Haddad; Sandra Coccuzzo Sampaio Vessoni; Simone Kashima; Svetoslav Nanev Slavov; Vincent Louis Viala |
| EPI_ISL_2493573 | CS III DR ANTONIO ANADAO SSGRAMA | Instituto Butantan | Antonio Jorge Martins; Claudia Renata dos Santos Barros; David Schlesinger; Debora Botequio Moretti; Dimas Tadeu Covas; Elaine Cristina Marqueze; Elaine Vieira Santos; Evandra Strazza Rodrigues; Heidge Fukumasu; Jayme Augusto de Souza-Neto; José Salvatore Leister Patané; Luiz Alcantara; Luiz Lehmann Coutinho; Maria Carolina Elias; Maurício Lacerda Nogueira; Rafael dos Santos Bezerra; Raul Machado Neto; Rejane Maria Tommasini Grotto; Ricardo Haddad; Sandra Coccuzzo Sampaio Vessoni; Simone Kashima; Svetoslav Nanev Slavov; Vincent Louis Viala |
| EPI_ISL_2345273, EPI_ISL_2345274, EPI_ISL_2345278 | CS IOLANDA FLEURY DE CAMPOS GUEDES NUPORANGA | Instituto Butantan / ESALQ-Piracicaba | Antonio Jorge Martins; Claudia Renata dos Santos Barros; David Schlesinger; Debora Botequio Moretti; Dimas Tadeu Covas; Elaine Cristina Marqueze; Elaine Vieira Santos; Evandra Strazza Rodrigues; Heidge Fukumasu; Jayme Augusto de Souza-Neto; José Salvatore Leister Patané; Luiz Alcantara; Luiz Lehmann Coutinho; Maria Carolina Elias; Maurício Lacerda Nogueira; Rafael dos Santos Bezerra; Raul Machado Neto; Rejane Maria Tommasini Grotto; Ricardo Haddad; Sandra Coccuzzo Sampaio Vessoni; Simone Kashima; Svetoslav Nanev Slavov; Vincent Louis Viala |
| EPI_ISL_2344831 | CSA COMPLEXO DE SAUDE ANHANGUERA LTDA | Instituto Butantan | Antonio Jorge Martins; Claudia Renata dos Santos Barros; David Schlesinger; Debora Botequio Moretti; Dimas Tadeu Covas; Elaine Cristina Marqueze; Elaine Vieira Santos; Evandra Strazza Rodrigues; Heidge Fukumasu; Jayme Augusto de Souza-Neto; José Salvatore Leister Patané; Luiz Alcantara; Luiz Lehmann Coutinho; Maria Carolina Elias; Maurício Lacerda Nogueira; Rafael dos Santos Bezerra; Raul Machado Neto; Rejane Maria Tommasini Grotto; Ricardo Haddad; Sandra Coccuzzo Sampaio Vessoni; Simone Kashima; Svetoslav Nanev Slavov; Vincent Louis Viala |
| EPI_ISL_2344906 | CSA COMPLEXO DE SAUDE ANHANGUERA TUCURUVI | Instituto Butantan | Antonio Jorge Martins; Claudia Renata dos Santos Barros; David Schlesinger; Debora Botequio Moretti; Dimas Tadeu Covas; Elaine Cristina Marqueze; Elaine Vieira Santos; Evandra Strazza Rodrigues; Heidge Fukumasu; Jayme Augusto de Souza-Neto; José Salvatore Leister Patané; Luiz Alcantara; Luiz Lehmann Coutinho; Maria Carolina Elias; Maurício Lacerda Nogueira; Rafael dos Santos Bezerra; Raul Machado Neto; Rejane Maria Tommasini Grotto; Ricardo Haddad; Sandra Coccuzzo Sampaio Vessoni; Simone Kashima; Svetoslav Nanev Slavov; Vincent Louis Viala |
| EPI_ISL_2493301 | CSII DR JOSE FERREIRA TELLES | Instituto Butantan | Antonio Jorge Martins; Claudia Renata dos Santos Barros; David Schlesinger; Debora Botequio Moretti; Dimas Tadeu Covas; Elaine Cristina Marqueze; Elaine Vieira Santos; Evandra Strazza Rodrigues; Heidge Fukumasu; Jayme Augusto de Souza-Neto; José Salvatore Leister Patané; Luiz Alcantara; Luiz Lehmann Coutinho; Maria Carolina Elias; Maurício Lacerda Nogueira; Rafael dos Santos Bezerra; Raul Machado Neto; Rejane Maria Tommasini Grotto; Ricardo Haddad; Sandra Coccuzzo Sampaio Vessoni; Simone Kashima; Svetoslav Nanev Slavov; Vincent Louis Viala |
| EPI_ISL_2345171, EPI_ISL_2345246 | CSII DR JOSE FERREIRA TELLES | Instituto Butantan / ESALQ-Piracicaba | Antonio Jorge Martins; Claudia Renata dos Santos Barros; David Schlesinger; Debora Botequio Moretti; Dimas Tadeu Covas; Elaine Cristina Marqueze; Elaine Vieira Santos; Evandra Strazza Rodrigues; Heidge Fukumasu; Jayme Augusto de Souza-Neto; José Salvatore Leister Patané; Luiz Alcantara; Luiz Lehmann Coutinho; Maria Carolina Elias; Maurício Lacerda Nogueira; Rafael dos Santos Bezerra; Raul Machado Neto; Rejane Maria Tommasini Grotto; Ricardo Haddad; Sandra Coccuzzo Sampaio Vessoni; Simone Kashima; Svetoslav Nanev Slavov; Vincent Louis Viala |
| EPI_ISL_2493622 | CSII DR WASHINGTON LUIS M RODRIGUES DA SILVA PITANGUEIRAS | Instituto Butantan | Antonio Jorge Martins; Claudia Renata dos Santos Barros; David Schlesinger; Debora Botequio Moretti; Dimas Tadeu Covas; Elaine Cristina Marqueze; Elaine Vieira Santos; Evandra Strazza Rodrigues; Heidge Fukumasu; Jayme Augusto de Souza-Neto; José Salvatore Leister Patané; Luiz Alcantara; Luiz Lehmann Coutinho; Maria Carolina Elias; Maurício Lacerda Nogueira; Rafael dos Santos Bezerra; Raul Machado Neto; Rejane Maria Tommasini Grotto; Ricardo Haddad; Sandra Coccuzzo Sampaio Vessoni; Simone Kashima; Svetoslav Nanev Slavov; Vincent Louis Viala |
| EPI_ISL_2208992, EPI_ISL_2345680, EPI_ISL_2345824, EPI_ISL_2345868, EPI_ISL_2362268 | CSII DR WASHINGTON LUIS M RODRIGUES DA SILVA PITANGUEIRAS | Instituto Butantan / Mendelics | Antonio Jorge Martins; Claudia Renata dos Santos Barros; David Schlesinger; Debora Botequio Moretti; Dimas Tadeu Covas; Elaine Cristina Marqueze; Elaine Vieira Santos; Evandra Strazza Rodrigues; Heidge Fukumasu; Jayme Augusto de Souza-Neto; José Salvatore Leister Patané; Luiz Alcantara; Luiz Lehmann Coutinho; Maria Carolina Elias; Maurício Lacerda Nogueira; Rafael dos Santos Bezerra; Raul Machado Neto; Rejane Maria Tommasini Grotto; Ricardo Haddad; Sandra Coccuzzo Sampaio Vessoni; Simone Kashima; Svetoslav Nanev Slavov; Vincent Louis Viala |
| EPI_ISL_2105892 | CSL, Maryland Department of Health | CSL, Maryland Department of Health | Keller, E.; Loomis, J.; Low, K. |
| EPI_ISL_2537469, EPI_ISL_2892480, EPI_ISL_3536926, EPI_ISL_3536928, EPI_ISL_3536930, EPI_ISL_3536933, EPI_ISL_4005512, EPI_ISL_4005520, EPI_ISL_4005522, EPI_ISL_4005534, EPI_ISL_4005535, EPI_ISL_4005537, EPI_ISL_4005543, EPI_ISL_4005549, EPI_ISL_4005551 | | | |
| see above | California Department of Public Health | California Department of Public Health | CDPH IDLB COVIDNet et al |
| EPI_ISL_3134058, EPI_ISL_3134076, EPI_ISL_3134087 | California Department of Public Health Valencia Branch Laboratory | California Department of Public Health | CDPH-COVIDNet; UCSF Center for Advanced Technology |
| EPI_ISL_2778847, EPI_ISL_2778870, EPI_ISL_2778895, EPI_ISL_2778935 | California Department of Public Health Valencia Branch Laboratory (CDPH VBL) | California Department of Public Health | CDPH-COVIDNet; UCLA Technology Center for Genomics & Bioinformatics |
| EPI_ISL_3144947 | Center for Virology | Center for Virology | Jeremy V. Camp; Monika Redlberger-Fritz; Stephan W. Aberle |
| EPI_ISL_2296515, EPI_ISL_2296516 | Centracare Laboratory Services | Minnesota Department of Health, Public Health Laboratory | Alexandra Lorentz; Jacob Garfin; Matt Plumb; and Xiong Wang |
| EPI_ISL_2502476, EPI_ISL_2502477, EPI_ISL_2502478, EPI_ISL_2502479, EPI_ISL_2502480, EPI_ISL_2502481, EPI_ISL_2502482, EPI_ISL_2502483, EPI_ISL_2502484, EPI_ISL_2502485, EPI_ISL_2502486, EPI_ISL_2502487, EPI_ISL_2502488, EPI_ISL_2502489, EPI_ISL_2502490, EPI_ISL_2502491, EPI_ISL_2502492, EPI_ISL_2502493, EPI_ISL_2502494, EPI_ISL_2502495, EPI_ISL_2502496, EPI_ISL_2502497, EPI_ISL_2502498, EPI_ISL_2502499, EPI_ISL_2502500, EPI_ISL_2502501, EPI_ISL_2502502, EPI_ISL_2502503, EPI_ISL_2502504, EPI_ISL_2502505, EPI_ISL_2502506, EPI_ISL_2502508, EPI_ISL_3462589, EPI_ISL_3462622, EPI_ISL_3462623, EPI_ISL_3462624, EPI_ISL_3462625, EPI_ISL_3462626 | | | |
| see above | Central Laboratory, Bureau of Public Health (BOG) and Academic Hospital Paramaribo | Erasmus Medical Center | Bas B Oude Munnink; Cherise Beek; Consuella Partowidjojo; Dion Gajadin; Ed PF IJzerman; Emmanuelle Munger; Gary Gummels; Ingrid SK Krishnadath; Lycke Woittiez; Marion PG Koopmans; Mireille Van de Veer; Phyllis Pinas; Princes Wongsowidjojo; Radjesh Ori; Ranisha Doerbalie; Rohma Banwari; Soeradj Harkisoen; Stephen Vreden; Tilotmadebie Ramlal; Verne Nanhoe |
| EPI_ISL_3255060, EPI_ISL_3255062, EPI_ISL_3255065, EPI_ISL_3255066, EPI_ISL_3255069, EPI_ISL_3255071, EPI_ISL_3255072, EPI_ISL_3255073, EPI_ISL_3255076, EPI_ISL_3255080, EPI_ISL_3255083, EPI_ISL_3255086, EPI_ISL_3255087, EPI_ISL_3255088, EPI_ISL_3255089, EPI_ISL_3255090, EPI_ISL_3255091, EPI_ISL_3255092, EPI_ISL_3255093, EPI_ISL_3255094, EPI_ISL_3255095, EPI_ISL_3255096, EPI_ISL_3255097, EPI_ISL_3255099, EPI_ISL_3255100, EPI_ISL_3255102, EPI_ISL_3255103, EPI_ISL_3255104, EPI_ISL_3255106, EPI_ISL_3255107, EPI_ISL_3255108, EPI_ISL_3255109, EPI_ISL_3255110, EPI_ISL_3255111, EPI_ISL_3255112, EPI_ISL_3255113, EPI_ISL_3255114, EPI_ISL_3255115, EPI_ISL_3255119, EPI_ISL_3255120, EPI_ISL_3255121, EPI_ISL_3255123, EPI_ISL_3255125, EPI_ISL_3255126, EPI_ISL_3255127, EPI_ISL_3255128, EPI_ISL_3255129, EPI_ISL_3255130, EPI_ISL_3255131, EPI_ISL_3255133, EPI_ISL_3255136, EPI_ISL_3255139, EPI_ISL_3255143, EPI_ISL_3255150, EPI_ISL_3255153, EPI_ISL_3255161, EPI_ISL_3255165, EPI_ISL_3255172, EPI_ISL_3255173, EPI_ISL_3255177, EPI_ISL_3255178, EPI_ISL_3255191, EPI_ISL_3255194, EPI_ISL_3255201, EPI_ISL_3255207, EPI_ISL_3255219, EPI_ISL_3255223, EPI_ISL_3255233 | | | |
| see above | Central Public Health Laboratory - LACEN -Bahia, Salvador, Brazil | Central Public Health Laboratory - LACEN -Bahia, Salvador, Brazil | Arabela Leal; Breno Dominguez; Felicidade Pereira; Jaqueline Gomes; Luciana Oliveira; Luiz Alcantara; Marcela Gómez; Marta Giovanetti; Patrícia Cajado; Stephane Tosta; Vagner Fonseca; Vanessa Nardy |
| EPI_ISL_1853299, EPI_ISL_2002663, EPI_ISL_2002664, EPI_ISL_2134734 | Centre Hospitalier Universitaire Clermont-Ferrand | CHU Clermont-Ferrand, service de virologie | Bisseux Maxime; Combes Patricia; Henquell Cécile; Mirand Audrey |
| EPI_ISL_2671553 | Centro de Investigacion Biomedica de Occidente (CIBO) | Unidad de Genomica Avanzada | ; Alejandra Garcia-Gasca; Alejandra Hernandez-Teran; Alejandro Sanchez-Flores; Alfredo Herrera-Estrella; Alicia Ocaña-Mondragon; Andreu Comas-Garcia; Angel Gustavo Salas-Lais; Antonio Loza Roman; Bernardo Martinez-Miguel; Blanca Taboada; Brenda Irasema Maldonado-Meza; Bruno Gomez-Gil; Carla Ivon Herrera-Najera; Carlos F. Arias; Celia Boukadida; Celida Duque Molina; Celida Martinez- Rodriguez; Clara Esperanza Santacruz-Tinoco; Concepcion Grajales-Muñiz; Consorcio Mexicano de Vigilancia Genomica (CoViGen-Mex). Authors (in alphabetical order): Julio Elias Alvarado-Yaah; Cristobal Chaidez-Quiroz; Daniel Fregoso-Rueda; Daniel Lira Morales; Eduardo Becerril-Vargas; Fernando Fontove-Herrera; Fidencio Mejia-Nepomuceno; Francisco Pulido; Gloria Elena Espinosa-Ayala; Gloria Maria Molina-Salinas; Gloria Vazquez; Hector Esteban Paz-Juarez; Hector Montoya-Fuentes; Helen Haydee Fernanda Ramirez-Plascencia; Irvin Gonzalez-Lopez; Jean Pierre Gonzalez; Jesus Hernandez; Joel Armando Vazquez-Perez.; Jorge Salas-Hernandez; Jose Antonio Enciso-Moreno; Jose Arturo Martinez-Orozco; Jose Esteban Muñoz-Medina; Jose de Jesus Nuñez-Contreras; Juan Bautista Chale-Dzul; Julissa Enciso-Ibarra; Luis Alberto Ochoa-Carrera; Margarita Matias-Florentino; Maria Guadalupe Santiago-Mauricio; Maria Guadalupe de Jesus Mireles-Rivera; Mario Mujica-Sanchez; Marissa Perez-Garcia; Nelly Selem-Mojica; Pavel Isa; Ricardo Ciria Merce; Ricardo Grande; Rosa Maria Gutierrez Rios; Santiago avila-Rios; Selene Zarate; Susana Lopez; Veronica Mata-Haro; Victor Eduardo Garcia-Arias; Victor Hugo Borja-Aburto |
| EPI_ISL_2671543, EPI_ISL_2671619, EPI_ISL_2671624, EPI_ISL_2671716, EPI_ISL_2671719 | Centro de Investigacion Biomedica del Noreste (CIBIN) | Unidad de Genomica Avanzada | ; Alejandra Garcia-Gasca; Alejandra Hernandez-Teran; Alejandro Sanchez-Flores; Alfredo Herrera-Estrella; Alicia Ocaña-Mondragon; Andreu Comas-Garcia; Angel Gustavo Salas-Lais; Antonio Loza Roman; Bernardo Martinez-Miguel; Blanca Taboada; Brenda Irasema Maldonado-Meza; Bruno Gomez-Gil; Carla Ivon Herrera-Najera; Carlos F. Arias; Celia Boukadida; Celida Duque Molina; Celida Martinez- Rodriguez; Clara Esperanza Santacruz-Tinoco; Concepcion Grajales-Muñiz; Consorcio Mexicano de Vigilancia Genomica (CoViGen-Mex). Authors (in alphabetical order): Julio Elias Alvarado-Yaah; Cristobal Chaidez-Quiroz; Daniel Fregoso-Rueda; Daniel Lira Morales; Eduardo Becerril-Vargas; Fernando Fontove-Herrera; Fidencio Mejia-Nepomuceno; Francisco Pulido; Gloria Elena Espinosa-Ayala; Gloria Maria Molina-Salinas; Gloria Vazquez; Hector Esteban Paz-Juarez; Hector Montoya-Fuentes; Helen Haydee Fernanda Ramirez-Plascencia; Irvin Gonzalez-Lopez; Jean Pierre Gonzalez; Jesus Hernandez; Joel Armando Vazquez-Perez.; Jorge Salas-Hernandez; Jose Antonio Enciso-Moreno; Jose Arturo Martinez-Orozco; Jose Esteban Muñoz-Medina; Jose de Jesus Nuñez-Contreras; Juan Bautista Chale-Dzul; Julissa Enciso-Ibarra; Luis Alberto Ochoa-Carrera; Margarita Matias-Florentino; Maria Guadalupe Santiago-Mauricio; Maria Guadalupe de Jesus Mireles-Rivera; Mario Mujica-Sanchez; Marissa Perez-Garcia; Nelly Selem-Mojica; Pavel Isa; Ricardo Ciria Merce; Ricardo Grande; Rosa Maria Gutierrez Rios; Santiago avila-Rios; Selene Zarate; Susana Lopez; Veronica Mata-Haro; Victor Eduardo Garcia-Arias; Victor Hugo Borja-Aburto |
| EPI_ISL_2490382, | Centro de Investigación Biomédica de | Centro de Investigación en | ; Alejandra Garcia-Gasca; Alejandra Hernández-Terán; Alejandro Sanchez-Flores; Alfredo Herrera-Estrella; Alicia Ocaña-Mondragón; Andreu Comas-García; Angel Gustavo Salas-Lais; Antonio Loza Román; Bernardo Martínez-Miguel; Blanca Taboada; Brenda Irasema Maldonado-Meza; Bruno Gomez-Gil; Carla |

| | | | |
|---|---|---|---|
| EPI_ISL_2490383, EPI_ISL_2490384 | Occidente (CIBO) | Enfermedades Infecciosas (CIENI), Instituto Nacional de Enfermedades Respiratorias (INER) | Ivón Herrera-Najera; Carlos F. Arias; Celia Boukadida; Clara Esperanza Santacruz-Tinoco; Concepción Grajales-Muñiz; Consorcio Mexicano de Vigilancia Genómica (CoViGen-Mex). Authors (in alphabetical order): Julio Elias Alvarado-Yaah; Cristóbal Cháidez-Quiróz; Célida Duque Molina; Célida Martínez-Rodríguez; Daniel Fregoso-Rueda; Daniel Lira Morales; Eduardo Becerril-Vargas; Fernando Fontove-Herrera; Fidencio Mejía-Nepomuceno; Francisco Pulido; Gloria Elena Espinosa-Ayala; Gloria María Molina-Salinas; Gloria Vazquez; Hector Esteban Paz-Juárez; Hector Montoya-Fuentes; Helen Haydee Fernanda Ramírez-Plascencia; Irvin González-López; Jean Pierre González; Joel Armando Vázquez-Pérez.; Jorge Salas-Hernández; José Antonio Enciso-Moreno; José Arturo Martínez-Orozco; José Esteban Muñoz-Medina; José de Jesús Nuñez-Contreras; Juan Bautista Chale-Dzul; Julissa Enciso-Ibarra; Luis Alberto Ochoa-Carrera; Margarita Matías-Florentino; Mario Mújica-Sánchez; Marissa Perez-Garcia; María Guadalupe Santiago-Mauricio; María Guadalupe de Jesús Mireles-Rivera; Nelly Sélem-Mojica; Pavel Isa; Ricardo Ciria Merce; Ricardo Grande; Rosa María Gutierrez Rios; Santiago Ávila-Ríos; Selene Zárate; Susana Lopez; Victor Eduardo Garcia-Arias; Víctor Hugo Borja-Aburto |
| EPI_ISL_2402171 | Centro de Investigación Biomédica de Occidente (CIBO) | Unidad de Genomica Avanzada | Alejandro Sanchez-Flores; Alfredo Herrera-Estrella; Alicia Ocana-Mondragon; Angel Gustavo Salas-Lais; Bernardo Martinez-Miguel; Blanca Taboada; Brenda Irasema Maldonado-Meza; Carla Ivon Herrera-Najera; Carlos F. Arias; Celia Boukadida; Clara Esperanza Santacruz-Tinoco; Concepcion Grajales-Muniz; Consorcio Mexicano de Vigilancia Genomica (CoViGen-Mex). Authors (in alphabetical order): Julio Elias Alvarado-Yaah; Fernando Fontove-Herrera; Francisco Pulido; Gloria Elena Espinoza-Ayala; Gloria Maria Molina-Salinas; Gloria Vazquez; Hector Esteban Paz-Juarez; Hector Montoya-Fuentes; Helen Haydee Fernanda Ramirez-Plascencia; Jorge Ivan Salinal-Nevarez; Jose Antonio Enciso-Moreno; Jose Esteban Munoz-Medina; Jose de Jesus Nunez-Contreras; Juan Bautista Chale-Dzul; Luis Alberto Ochoa-Carrera; Margarita Matias-Florentino; Maria Guadalupe Santiago-Mauricio; Maria Guadalupe de Jesus Mireles-Rivera; Nelly Selem-Mojica; Pavel Isa; Ricardo Grande; Santiago Avila-Rios; Victor Hugo Borja-Aburto |
| EPI_ISL_2490359, EPI_ISL_2490360, EPI_ISL_2490361, EPI_ISL_2490364, EPI_ISL_2490365, EPI_ISL_2490366 | Centro de Investigación Biomédica del Noreste (CIBIN) | Centro de Investigación en Enfermedades Infecciosas (CIENI), Instituto Nacional de Enfermedades Respiratorias (INER) | ; Alejandra Garcia-Gasca; Alejandra Hernández-Terán; Alejandro Sanchez-Flores; Alfredo Herrera-Estrella; Alicia Ocaña-Mondragón; Andreu Comas-García; Angel Gustavo Salas-Lais; Antonio Loza Román; Bernardo Martínez-Miguel; Blanca Taboada; Brenda Irasema Maldonado-Meza; Bruno Gomez-Gil; Carla Ivón Herrera-Najera; Carlos F. Arias; Celia Boukadida; Clara Esperanza Santacruz-Tinoco; Concepción Grajales-Muñiz; Consorcio Mexicano de Vigilancia Genómica (CoViGen-Mex). Authors (in alphabetical order): Julio Elias Alvarado-Yaah; Cristóbal Cháidez-Quiróz; Célida Duque Molina; Célida Martínez-Rodríguez; Daniel Fregoso-Rueda; Daniel Lira Morales; Eduardo Becerril-Vargas; Fernando Fontove-Herrera; Fidencio Mejía-Nepomuceno; Francisco Pulido; Gloria Elena Espinosa-Ayala; Gloria María Molina-Salinas; Gloria Vazquez; Hector Esteban Paz-Juárez; Hector Montoya-Fuentes; Helen Haydee Fernanda Ramírez-Plascencia; Irvin González-López; Jean Pierre González; Joel Armando Vázquez-Pérez.; Jorge Salas-Hernández; José Antonio Enciso-Moreno; José Arturo Martínez-Orozco; José Esteban Muñoz-Medina; José de Jesús Nuñez-Contreras; Juan Bautista Chale-Dzul; Julissa Enciso-Ibarra; Luis Alberto Ochoa-Carrera; Margarita Matías-Florentino; Mario Mújica-Sánchez; Marissa Perez-Garcia; María Guadalupe Santiago-Mauricio; María Guadalupe de Jesús Mireles-Rivera; Nelly Sélem-Mojica; Pavel Isa; Ricardo Ciria Merce; Ricardo Grande; Rosa María Gutierrez Rios; Santiago Ávila-Ríos; Selene Zárate; Susana Lopez; Victor Eduardo Garcia-Arias; Víctor Hugo Borja-Aburto |
| EPI_ISL_2401953, EPI_ISL_2401979, EPI_ISL_2401991 | Centro de Investigación Biomédica del Noreste (CIBIN) | Unidad de Genomica Avanzada | Alejandro Sanchez-Flores; Alfredo Herrera-Estrella; Alicia Ocana-Mondragon; Angel Gustavo Salas-Lais; Bernardo Martinez-Miguel; Blanca Taboada; Brenda Irasema Maldonado-Meza; Carla Ivon Herrera-Najera; Carlos F. Arias; Celia Boukadida; Clara Esperanza Santacruz-Tinoco; Concepcion Grajales-Muniz; Consorcio Mexicano de Vigilancia Genomica (CoViGen-Mex). Authors (in alphabetical order): Julio Elias Alvarado-Yaah; Fernando Fontove-Herrera; Francisco Pulido; Gloria Elena Espinoza-Ayala; Gloria Maria Molina-Salinas; Gloria Vazquez; Hector Esteban Paz-Juarez; Hector Montoya-Fuentes; Helen Haydee Fernanda Ramirez-Plascencia; Jorge Ivan Salinal-Nevarez; Jose Antonio Enciso-Moreno; Jose Esteban Munoz-Medina; Jose de Jesus Nunez-Contreras; Juan Bautista Chale-Dzul; Luis Alberto Ochoa-Carrera; Margarita Matias-Florentino; Maria Guadalupe Santiago-Mauricio; Maria Guadalupe de Jesus Mireles-Rivera; Nelly Selem-Mojica; Pavel Isa; Ricardo Grande; Santiago Avila-Rios; Victor Hugo Borja-Aburto |
| EPI_ISL_2004300, EPI_ISL_2004301 | Cintramedica | Instituto Nacional de Saude (INSA) | Borges et al |
| EPI_ISL_2258223, EPI_ISL_2258229, EPI_ISL_2258231, EPI_ISL_2258240, EPI_ISL_2258241, EPI_ISL_2258252, EPI_ISL_2273859, EPI_ISL_2617566, EPI_ISL_3074103 | | | |
| see above | City of Milwaukee Health Department Laboratory | City of Milwaukee Health Department Laboratory | Amy Bauer; Jennifer Lentz; Manjeet Khubbar; Samantha Scott; Sanjib Bhattacharyya |
| EPI_ISL_2467143, EPI_ISL_2467152, EPI_ISL_2467174, EPI_ISL_2467217 | Clinica INDISA | "Facultad de Ciencias de la Vida, UNAB" | "Claudio Meneses; Ariel Orellana"; Claudio Olmos; Daniel Leon; Dayan Sanhueza; Eduardo Castro; Gonzalo Campaña; Macarena Bastias; Paola Pidal; Ricardo Yusta; Sebastian Wolter; Susana Saez; Víctor Monreal; Waldo Diaz |
| EPI_ISL_2098734, EPI_ISL_2380856 | Clinical Molecular Microbiology Laboratory, UNC Hospitals | Jeremy Wang | Alexander Rubinsteyn; Brianna de la Houssaye; Colleen Rice; Corbin Jones; Jason Smedberg; Jeremy Wang; Jose Luis Torres Castillo; Melissa Miller; Robert Hagan; Shawn Hawken |
| EPI_ISL_1805786 | Clinical Reference Laboratory | Kansas Health and Environmental Lab | Ben Olsen; Jonathan Barnell; Mike Grose; and Phil Adam |
| EPI_ISL_2408426, EPI_ISL_2466571 | Clinical Virology | Clinical Bacteriology | Adrian Egli; Alfredo Mari; Fanny Wegner; Hans Hirsch; Helena MB Seth-Smith; Julia Bielicki; Karoline Leuzinger; Manuel Battegay; Tim Roloff |
| EPI_ISL_2309507, EPI_ISL_2309567, EPI_ISL_2309879, EPI_ISL_2309958, EPI_ISL_2309972, EPI_ISL_2310745, EPI_ISL_2549913, EPI_ISL_2549925, EPI_ISL_2549993, EPI_ISL_2550187, EPI_ISL_2550261, EPI_ISL_2550275, EPI_ISL_2550311, EPI_ISL_2987419, EPI_ISL_3160136, EPI_ISL_3160137, EPI_ISL_3583885, EPI_ISL_3842256, EPI_ISL_3842258 | | | |
| see above | Colorado Department of Public Health and Environment | Colorado Department of Public Health and Environment | Alexandria Rossheim; Diana Ir; Emily A. Travanty; Laura Bankers; Mandy Waters; Michael Martin; Molly C. Hetherington-Rauth; Sarah Elizabeth Totten; Shannon R. Matzinger |
| EPI_ISL_2285125 | Complejo Hospitalario Universitario de Pontevedra | Microbiology Department. Complexo Hospitalario Universitario de Vigo | Campello M; Perez S; Regueiro B; Trigo M |
| EPI_ISL_2496914 | Contra Costa County Public Health Lab | Chan-Zuckerberg Biohub | CZB Cliahub Consortium |
| EPI_ISL_3663778, EPI_ISL_3663779, EPI_ISL_3663781, EPI_ISL_3663784, EPI_ISL_3663785, EPI_ISL_3663786, EPI_ISL_3663789, EPI_ISL_3663791, EPI_ISL_3663792, EPI_ISL_3663793, EPI_ISL_3663795, EPI_ISL_3663796, EPI_ISL_3663797, EPI_ISL_3663798, EPI_ISL_3663799, EPI_ISL_3663800, EPI_ISL_3663801, EPI_ISL_3663802, EPI_ISL_3663804, EPI_ISL_3663805, EPI_ISL_3663806, EPI_ISL_3663808, EPI_ISL_3663809, EPI_ISL_3663810, EPI_ISL_3663811, EPI_ISL_3663812, EPI_ISL_3663813, EPI_ISL_3663816, EPI_ISL_3663822, EPI_ISL_3663823, EPI_ISL_3663824, EPI_ISL_3663826, EPI_ISL_3663828, EPI_ISL_3663829, EPI_ISL_3663830, EPI_ISL_3663832, EPI_ISL_3663833, EPI_ISL_3663835, EPI_ISL_3663836, EPI_ISL_3663837, EPI_ISL_3663839, EPI_ISL_3663840, EPI_ISL_3663841, EPI_ISL_3663845, EPI_ISL_3663846, EPI_ISL_3663848, EPI_ISL_3663850, EPI_ISL_3663851, EPI_ISL_3663852, EPI_ISL_3663853, EPI_ISL_3663854, EPI_ISL_3663859, EPI_ISL_3663861, EPI_ISL_3663862, EPI_ISL_3663863, EPI_ISL_3663865, EPI_ISL_3663866, EPI_ISL_3663867, EPI_ISL_3663870, EPI_ISL_3663871, EPI_ISL_3663872, EPI_ISL_3663874, EPI_ISL_3663878, EPI_ISL_3663879, EPI_ISL_3663880, EPI_ISL_3663881, EPI_ISL_3663882, EPI_ISL_3663883, EPI_ISL_3663886, EPI_ISL_3663888, EPI_ISL_3663889, EPI_ISL_3663891, EPI_ISL_3663893, EPI_ISL_3663894, EPI_ISL_3663900, EPI_ISL_3663906, EPI_ISL_3663907, EPI_ISL_3663910, EPI_ISL_3663911, EPI_ISL_3663912, EPI_ISL_3663913, EPI_ISL_3663916, EPI_ISL_3663917, EPI_ISL_3663918, EPI_ISL_3663920, EPI_ISL_3663922, EPI_ISL_3663923, EPI_ISL_3663924, EPI_ISL_3663926, EPI_ISL_3663927, EPI_ISL_3663928, EPI_ISL_3663930, EPI_ISL_3663931, EPI_ISL_3663932, EPI_ISL_3663934, EPI_ISL_3663935, EPI_ISL_3663937, EPI_ISL_3663939, EPI_ISL_3663941, EPI_ISL_3663942, EPI_ISL_3663943, EPI_ISL_3663944, EPI_ISL_3663947, EPI_ISL_3663950, EPI_ISL_3663951, EPI_ISL_3663952, EPI_ISL_3663953, EPI_ISL_3663954, EPI_ISL_3663955, EPI_ISL_3663956, EPI_ISL_3663957, EPI_ISL_3663958, EPI_ISL_3663959, EPI_ISL_3663962, EPI_ISL_3663964, EPI_ISL_3663965, EPI_ISL_3663966, EPI_ISL_3663967, EPI_ISL_3663968, EPI_ISL_3663969, EPI_ISL_3663971, EPI_ISL_3663975, EPI_ISL_3663977, EPI_ISL_3663978, EPI_ISL_3663980, EPI_ISL_3663981, EPI_ISL_3663982, EPI_ISL_3663983, EPI_ISL_3663984, EPI_ISL_3663985, EPI_ISL_3663986, EPI_ISL_3663989, EPI_ISL_3663991, EPI_ISL_3663992, EPI_ISL_3663995, EPI_ISL_3663996, EPI_ISL_3663997, EPI_ISL_3663998, EPI_ISL_3664001, EPI_ISL_3664002, EPI_ISL_3664003, EPI_ISL_3664004, EPI_ISL_3664005, EPI_ISL_3664006, EPI_ISL_3664007, EPI_ISL_3664009, EPI_ISL_3664010, EPI_ISL_3664012, EPI_ISL_3664014, EPI_ISL_3664016, EPI_ISL_3664017, EPI_ISL_3664018, EPI_ISL_3664020, EPI_ISL_3664021, EPI_ISL_3664025, EPI_ISL_3664026, EPI_ISL_3664027, EPI_ISL_3664028, EPI_ISL_3664029, EPI_ISL_3664030, EPI_ISL_3664032, EPI_ISL_3664033, EPI_ISL_3664034, EPI_ISL_3664036, EPI_ISL_3664037, EPI_ISL_3664038, EPI_ISL_3664043, EPI_ISL_3664045, EPI_ISL_3664046, EPI_ISL_3664048, EPI_ISL_3664049, EPI_ISL_3664050, EPI_ISL_3664051, EPI_ISL_3664053, EPI_ISL_3664054, EPI_ISL_3664055, EPI_ISL_3664057, EPI_ISL_3664058, EPI_ISL_3664059, EPI_ISL_3664062, EPI_ISL_3664063, EPI_ISL_3664065, EPI_ISL_3664067, EPI_ISL_3664068, EPI_ISL_3664070, EPI_ISL_3664073, EPI_ISL_3664077, EPI_ISL_3664078, EPI_ISL_3664079, EPI_ISL_3664080, EPI_ISL_3664084, EPI_ISL_3664086, EPI_ISL_3664087, EPI_ISL_3664088, EPI_ISL_3664089, EPI_ISL_3664093, EPI_ISL_3664094, EPI_ISL_3664095, EPI_ISL_3664096, EPI_ISL_3664098, EPI_ISL_3664101, EPI_ISL_3664106, EPI_ISL_3664107, EPI_ISL_3664108, EPI_ISL_3664109, EPI_ISL_3664110, EPI_ISL_3664111, EPI_ISL_3664113, EPI_ISL_3664114, EPI_ISL_3664124, EPI_ISL_3664125, EPI_ISL_3664127, EPI_ISL_3664130, EPI_ISL_3664132, EPI_ISL_3664133, EPI_ISL_3664134, EPI_ISL_3664135, EPI_ISL_3664136, EPI_ISL_3664138, EPI_ISL_3664139, EPI_ISL_3664140, EPI_ISL_3664141, EPI_ISL_3664142, EPI_ISL_3664143, EPI_ISL_3664144, EPI_ISL_3664148, EPI_ISL_3664149, EPI_ISL_3664150, EPI_ISL_3664151, EPI_ISL_3664154, EPI_ISL_3664155, EPI_ISL_3664158, EPI_ISL_3664159, EPI_ISL_3664161, EPI_ISL_3664162, EPI_ISL_3664164, EPI_ISL_3664165, EPI_ISL_3664170, EPI_ISL_3664173, EPI_ISL_3664177, EPI_ISL_3664178, EPI_ISL_3664180, EPI_ISL_3664181, EPI_ISL_3664182, EPI_ISL_3664183, EPI_ISL_3664187, EPI_ISL_3664188, EPI_ISL_3664190, EPI_ISL_3664192, EPI_ISL_3664193, EPI_ISL_3664194, EPI_ISL_3664195, EPI_ISL_3664196, EPI_ISL_3664199, EPI_ISL_3664200, EPI_ISL_3664202, EPI_ISL_3664203, EPI_ISL_3664204, EPI_ISL_3664205, EPI_ISL_3664208, EPI_ISL_3664210, EPI_ISL_3664211, EPI_ISL_3664212, EPI_ISL_3664213, EPI_ISL_3664214, EPI_ISL_3664215, EPI_ISL_3664216, EPI_ISL_3664217, EPI_ISL_3664218, EPI_ISL_3664221, EPI_ISL_3664222, EPI_ISL_3664223, EPI_ISL_3664224, EPI_ISL_3664225, EPI_ISL_3664226, EPI_ISL_3664227, EPI_ISL_3664228, EPI_ISL_3664229, EPI_ISL_3664230, EPI_ISL_3664231, EPI_ISL_3664232, EPI_ISL_3664234, EPI_ISL_3664235, EPI_ISL_3664236, EPI_ISL_3664237, EPI_ISL_3664238, EPI_ISL_3664239, EPI_ISL_3664244, EPI_ISL_3664245, EPI_ISL_3664246, EPI_ISL_3664249, EPI_ISL_3664251, EPI_ISL_3664252, EPI_ISL_3664253, EPI_ISL_3664256, EPI_ISL_3664257, EPI_ISL_3664258, EPI_ISL_3664259, EPI_ISL_3664261, EPI_ISL_3664263, EPI_ISL_3664264, EPI_ISL_3664267, EPI_ISL_3664268, EPI_ISL_3664269, EPI_ISL_3664271, EPI_ISL_3664273 | | | |
| see above | DASA | DASA | Adriano Bonaldi; Angelica Hristov; Annelise Lopes; Bianca Cota; Cristina Oliveira; Jose Levi; Lidia Yamamoto; Paulo Pierry; Rodrigo Guarischi; Rodrigo Salazar |
| EPI_ISL_2229246, EPI_ISL_2451775 | DC Public Health Lab/ Dept. of Forensic Sciences | Centers for Disease Control and Prevention Division of Viral Diseases, Pathogen Discovery | Alison Laufer Halpin; Ben L. Rambo-Martin; Clinton R. Paden; Dakota Howard; Darlene Wagner; Dave Wentworth; Dhwani Batra; Jasmine Padilla; Justin Lee; Katie Dillon; Krista Queen; Kristen Knipe; Kristine Lacek; Mark Burroughs; Matthew Schmerer; Mili Sheth; Peter Cook; Sam Shepard; Sarah Nobles; Shoshona Le; Suxiang Tong; Vivien Dugan; Yvette Unoarumhi |
| EPI_ISL_2228976, EPI_ISL_2452011, EPI_ISL_2840444 | DE Public Health Laboratory | Centers for Disease Control and Prevention Division of Viral Diseases, Pathogen Discovery | Alison Laufer Halpin; Ben L. Rambo-Martin; Clinton R. Paden; Dakota Howard; Darlene Wagner; Dave Wentworth; Dhwani Batra; Jasmine Padilla; Justin Lee; Katie Dillon; Krista Queen; Kristen Knipe; Kristine Lacek; Mark Burroughs; Matthew Schmerer; Mili Sheth; Peter Cook; Sam Shepard; Sarah Nobles; Shoshona Le; Suxiang Tong; Vivien Dugan; Yvette Unoarumhi |
| EPI_ISL_2493631, EPI_ISL_2493632, EPI_ISL_2493634, EPI_ISL_2493636, EPI_ISL_2493638, EPI_ISL_2493639, EPI_ISL_2493641, EPI_ISL_2493642 | | | |
| see above | DEPARTAMENTO DE SAUDE MUNICIPAL SOCORRO SP | Instituto Butantan | Antonio Jorge Martins; Claudia Renata dos Santos Barros; David Schlesinger; Debora Botequio Moretti; Dimas Tadeu Covas; Elaine Cristina Marqueze; Elaine Vieira Santos; Evandra Strazza Rodrigues; Heidge Fukumasu; Jayme Augusto de Souza-Neto; José Salvatore Leister Patané; Luiz Alcantara; Luiz Lehmann Coutinho; Maria Carolina Elias; Maurício Lacerda Nogueira; Rafael dos Santos Bezerra; Raul Machado Neto; Rejane Maria Tommasini Grotto; Ricardo Haddad; Sandra Coccuzzo Sampaio Vessoni; Simone Kashima; Svetoslav Nanev Slavov; Vincent Louis Viala |
| EPI_ISL_2345240, EPI_ISL_2345241, EPI_ISL_2345243, EPI_ISL_2345244, EPI_ISL_2345249, EPI_ISL_2345252, EPI_ISL_2345254, EPI_ISL_2345255, EPI_ISL_2345257, EPI_ISL_2345258, EPI_ISL_2345259, EPI_ISL_2345260 | | | |
| see above | DEPARTAMENTO DE SAUDE MUNICIPAL SOCORRO SP | Instituto Butantan / ESALQ-Piracicaba | Antonio Jorge Martins; Claudia Renata dos Santos Barros; David Schlesinger; Debora Botequio Moretti; Dimas Tadeu Covas; Elaine Cristina Marqueze; Elaine Vieira Santos; Evandra Strazza Rodrigues; Heidge Fukumasu; Jayme Augusto de Souza-Neto; José Salvatore Leister Patané; Luiz Alcantara; Luiz Lehmann Coutinho; Maria Carolina Elias; Maurício Lacerda Nogueira; Rafael dos Santos Bezerra; Raul Machado Neto; Rejane Maria Tommasini Grotto; Ricardo Haddad; Sandra Coccuzzo Sampaio Vessoni; Simone Kashima; Svetoslav Nanev Slavov; Vincent Louis Viala |
| EPI_ISL_2209045, EPI_ISL_2209370, EPI_ISL_2209739, EPI_ISL_2345679, EPI_ISL_2345683, EPI_ISL_2345732, EPI_ISL_2345776, EPI_ISL_2345799, EPI_ISL_2345827, EPI_ISL_2345871 | | | |
| see above | DEPARTAMENTO DE SAUDE MUNICIPAL SOCORRO SP | Instituto Butantan / Mendelics | Antonio Jorge Martins; Claudia Renata dos Santos Barros; David Schlesinger; Debora Botequio Moretti; Dimas Tadeu Covas; Elaine Cristina Marqueze; Elaine Vieira Santos; Evandra Strazza Rodrigues; Heidge Fukumasu; Jayme Augusto de Souza-Neto; José Salvatore Leister Patané; Luiz Alcantara; Luiz Lehmann Coutinho; Maria Carolina Elias; Maurício Lacerda Nogueira; Rafael dos Santos Bezerra; Raul Machado Neto; Rejane Maria Tommasini Grotto; Ricardo Haddad; Sandra Coccuzzo Sampaio Vessoni; Simone Kashima; Svetoslav Nanev Slavov; Vincent Louis Viala |
| EPI_ISL_2445177, EPI_ISL_2445178, EPI_ISL_2445180, EPI_ISL_2445182, EPI_ISL_2445184, EPI_ISL_2445186 | DIRETORIA MUNICIPAL DE SAUDE | Instituto Butantan | Antonio Jorge Martins; Claudia Renata dos Santos Barros; David Schlesinger; Debora Botequio Moretti; Dimas Tadeu Covas; Elaine Cristina Marqueze; Elaine Vieira Santos; Evandra Strazza Rodrigues; Heidge Fukumasu; Jayme Augusto de Souza-Neto; José Salvatore Leister Patané; Luiz Alcantara; Luiz Lehmann Coutinho; Maria Carolina Elias; Maurício Lacerda Nogueira; Rafael dos Santos Bezerra; Raul Machado Neto; Rejane Maria Tommasini Grotto; Ricardo Haddad; Sandra Coccuzzo Sampaio Vessoni; Simone Kashima; Svetoslav Nanev Slavov; Vincent Louis Viala |
| EPI_ISL_2493374 | DIRETORIA MUNICIPAL DE SAUDE DE ENGENHEIRO COELHO | Instituto Butantan | Antonio Jorge Martins; Claudia Renata dos Santos Barros; David Schlesinger; Debora Botequio Moretti; Dimas Tadeu Covas; Elaine Cristina Marqueze; Elaine Vieira Santos; Evandra Strazza Rodrigues; Heidge Fukumasu; Jayme Augusto de Souza-Neto; José Salvatore Leister Patané; Luiz Alcantara; Luiz Lehmann Coutinho; Maria Carolina Elias; Maurício Lacerda Nogueira; Rafael dos Santos Bezerra; Raul Machado Neto; Rejane Maria Tommasini Grotto; Ricardo Haddad; Sandra Coccuzzo Sampaio Vessoni; Simone Kashima; Svetoslav Nanev Slavov; Vincent Louis Viala |
| EPI_ISL_2322910 | DOHMH PHL | New York City Public Health Laboratory | Jade Wang; et al. |
| EPI_ISL_2383713, EPI_ISL_2383852, EPI_ISL_2383864, EPI_ISL_2383877 | DPHL | Delaware Public Health Lab | Rebecca Savage |
| EPI_ISL_2429555, EPI_ISL_2430305, EPI_ISL_2430615, EPI_ISL_2431115, EPI_ISL_2431486, EPI_ISL_2431537 | Department of Bacteria, Parasites and Fungi, Statens Serum Institut, Copenhagen, Denmark | Statens Serum Institut Bioinformatics and Microbial Genomics | Danish Covid-19 Genome Consortium |
| EPI_ISL_2099850, EPI_ISL_2099851 | Department of Public Health Bucharest | National Institute of Infectious Diseases-Prof. Dr. Matei Bals Molecular Diagnostics Laboratory | Andreea Tudor; Corina Casangiu; Dan Otelea; Leontina Banica; Marius Surleac; Ovidiu Vlaicu; Simona Paraschiv |
| EPI_ISL_2608834 | Department of Virology and | Department of Virology, Faculty | Essi Korhonen; Hanna Jarva; Hanna Liimatainen; Hannimari Kallio-Kokko; Harri Kangas; Hussein Alburkat; Jenni Virtanen; Maija Lappalainen; Maija Suvanto; Olli Vapalahti; Pekka Ellonen; Phuoc Truong; Ravi Kant; Sari Hannula; Satu Kurkela; Teemu Smura |

| | | | |
|---|---|---|---|
| | Immunology, University of Helsinki and Helsinki University Hospital, Huslab Finland | of Medicine, University of Helsinki, Helsinki, Finland | |
| EPI_ISL_2403852, EPI_ISL_2403858, EPI_ISL_2403859 | Dianalabs SA | Genesupport | Tanguy ARAUD |
| EPI_ISL_3578151, EPI_ISL_3578168, EPI_ISL_3578189 | Dr. Risch Ostschweiz AG | Microbiology, Dr. Risch | Dominique Fabien Hilti; Faina Wehrli; Lorenz Risch; Martin Risch; Nadia Wohlwend; Sinem Kas; Thomas Bodmer |
| EPI_ISL_1962255, EPI_ISL_1962278, EPI_ISL_1962279, EPI_ISL_2093856, EPI_ISL_2093859, EPI_ISL_2093861, EPI_ISL_2093862, EPI_ISL_2093863, EPI_ISL_2093870, EPI_ISL_2093881, EPI_ISL_2093883, EPI_ISL_2093885, EPI_ISL_2217848, EPI_ISL_2219019, EPI_ISL_2219022, EPI_ISL_2219024, EPI_ISL_2219035, EPI_ISL_2219050, EPI_ISL_2219052, EPI_ISL_2219059, EPI_ISL_2219068, EPI_ISL_2219102, EPI_ISL_2303365, EPI_ISL_2303375, EPI_ISL_2303390, EPI_ISL_2303391, EPI_ISL_2405523, EPI_ISL_2405534, EPI_ISL_2405886, EPI_ISL_2405922, EPI_ISL_2406150, EPI_ISL_2406247, EPI_ISL_2406269, EPI_ISL_2406366, EPI_ISL_2406457, EPI_ISL_2475681, EPI_ISL_2475682, EPI_ISL_2475689, EPI_ISL_2475705, EPI_ISL_2475708, EPI_ISL_2673227, EPI_ISL_2673236 | | | |
| see above | Dutch COVID-19 response team | National Institute for Public Health and the Environment (RIVM) | Adam Meijer; AnneMarie van den Brandt; Annelies Kroneman; Bas van der Veer; Chantal Reusken; Dennis Schmitz; Dirk Eggink; Euníce Then; Florian Zwagemaker; Harry Vennema; James Groot; Jeroen Cremer; Karim Hajji; Kim Freriks; Linda van de Nes; Lisa Wijsman; Lynn Aarts; Melissa van Tuil; Ryanne Jaarsma; Sanne Bos; Sharon van den Brink; Sjoerd Kuiling; on behalf of the national COVID-19 response team |
| EPI_ISL_4060100, EPI_ISL_4060104, EPI_ISL_4060105 | Dynacare Manitoba | Manitoba Cadham Provincial Laboratory | David Alexander; Hilary Racher; Hongbin Li; Melissa Desaulnier; Tintu Abraham |
| EPI_ISL_2344869 | E S F DR EDSON GIANETTO JD CARLOS CASSETARI | Instituto Butantan / ESALQ-Piracicaba | Antonio Jorge Martins; Claudia Renata dos Santos Barros; David Schlesinger; Debora Botequio Moretti; Dimas Tadeu Covas; Elaine Cristina Marqueze; Elaine Vieira Santos; Evandra Strazza Rodrigues; Heidge Fukumasu; Jayme Augusto de Souza-Neto; José Salvatore Leister Patané; Luiz Alcantara; Luiz Lehmann Coutinho; Maria Carolina Elias; Maurício Lacerda Nogueira; Rafael dos Santos Bezerra; Raul Machado Neto; Rejane Maria Tommasini Grotto; Ricardo Haddad; Sandra Coccuzzo Sampaio Vessoni; Simone Kashima; Svetoslav Nanev Slavov; Vincent Louis Viala |
| EPI_ISL_2344877 | E S F WALDO DSTEFANO JD PRIMAVERA JOSE BONIFACIO | Instituto Butantan / ESALQ-Piracicaba | Antonio Jorge Martins; Claudia Renata dos Santos Barros; David Schlesinger; Debora Botequio Moretti; Dimas Tadeu Covas; Elaine Cristina Marqueze; Elaine Vieira Santos; Evandra Strazza Rodrigues; Heidge Fukumasu; Jayme Augusto de Souza-Neto; José Salvatore Leister Patané; Luiz Alcantara; Luiz Lehmann Coutinho; Maria Carolina Elias; Maurício Lacerda Nogueira; Rafael dos Santos Bezerra; Raul Machado Neto; Rejane Maria Tommasini Grotto; Ricardo Haddad; Sandra Coccuzzo Sampaio Vessoni; Simone Kashima; Svetoslav Nanev Slavov; Vincent Louis Viala |
| EPI_ISL_1941841 | EHNV | Laboratory of genomics and metagenomics | Claire Bertelli; Damien Jacot; Gilbert Greub; Sebastien Aeby; Trestan Pillonel |
| EPI_ISL_2493401, EPI_ISL_2493408 | EMERGENCIA RESPIRATORIA DE NOVA GRANADA | Instituto Butantan | Antonio Jorge Martins; Claudia Renata dos Santos Barros; David Schlesinger; Debora Botequio Moretti; Dimas Tadeu Covas; Elaine Cristina Marqueze; Elaine Vieira Santos; Evandra Strazza Rodrigues; Heidge Fukumasu; Jayme Augusto de Souza-Neto; José Salvatore Leister Patané; Luiz Alcantara; Luiz Lehmann Coutinho; Maria Carolina Elias; Maurício Lacerda Nogueira; Rafael dos Santos Bezerra; Raul Machado Neto; Rejane Maria Tommasini Grotto; Ricardo Haddad; Sandra Coccuzzo Sampaio Vessoni; Simone Kashima; Svetoslav Nanev Slavov; Vincent Louis Viala |
| EPI_ISL_2344834, EPI_ISL_2344835, EPI_ISL_2344845, EPI_ISL_2344861 | EMERGENCIA RESPIRATORIA DE NOVA GRANADA | Instituto Butantan / ESALQ-Piracicaba | Antonio Jorge Martins; Claudia Renata dos Santos Barros; David Schlesinger; Debora Botequio Moretti; Dimas Tadeu Covas; Elaine Cristina Marqueze; Elaine Vieira Santos; Evandra Strazza Rodrigues; Heidge Fukumasu; Jayme Augusto de Souza-Neto; José Salvatore Leister Patané; Luiz Alcantara; Luiz Lehmann Coutinho; Maria Carolina Elias; Maurício Lacerda Nogueira; Rafael dos Santos Bezerra; Raul Machado Neto; Rejane Maria Tommasini Grotto; Ricardo Haddad; Sandra Coccuzzo Sampaio Vessoni; Simone Kashima; Svetoslav Nanev Slavov; Vincent Louis Viala |
| EPI_ISL_2345128 | ESF COHAB IV | Instituto Butantan / ESALQ-Piracicaba | Antonio Jorge Martins; Claudia Renata dos Santos Barros; David Schlesinger; Debora Botequio Moretti; Dimas Tadeu Covas; Elaine Cristina Marqueze; Elaine Vieira Santos; Evandra Strazza Rodrigues; Heidge Fukumasu; Jayme Augusto de Souza-Neto; José Salvatore Leister Patané; Luiz Alcantara; Luiz Lehmann Coutinho; Maria Carolina Elias; Maurício Lacerda Nogueira; Rafael dos Santos Bezerra; Raul Machado Neto; Rejane Maria Tommasini Grotto; Ricardo Haddad; Sandra Coccuzzo Sampaio Vessoni; Simone Kashima; Svetoslav Nanev Slavov; Vincent Louis Viala |
| EPI_ISL_2445535, EPI_ISL_2473790 | ESF I E II | Instituto Butantan | Antonio Jorge Martins; Claudia Renata dos Santos Barros; David Schlesinger; Debora Botequio Moretti; Dimas Tadeu Covas; Elaine Cristina Marqueze; Elaine Vieira Santos; Evandra Strazza Rodrigues; Heidge Fukumasu; Jayme Augusto de Souza-Neto; José Salvatore Leister Patané; Luiz Alcantara; Luiz Lehmann Coutinho; Maria Carolina Elias; Maurício Lacerda Nogueira; Rafael dos Santos Bezerra; Raul Machado Neto; Rejane Maria Tommasini Grotto; Ricardo Haddad; Sandra Coccuzzo Sampaio Vessoni; Simone Kashima; Svetoslav Nanev Slavov; Vincent Louis Viala |
| EPI_ISL_2445515 | ESF NOVA TANABI II | Instituto Butantan | Antonio Jorge Martins; Claudia Renata dos Santos Barros; David Schlesinger; Debora Botequio Moretti; Dimas Tadeu Covas; Elaine Cristina Marqueze; Elaine Vieira Santos; Evandra Strazza Rodrigues; Heidge Fukumasu; Jayme Augusto de Souza-Neto; José Salvatore Leister Patané; Luiz Alcantara; Luiz Lehmann Coutinho; Maria Carolina Elias; Maurício Lacerda Nogueira; Rafael dos Santos Bezerra; Raul Machado Neto; Rejane Maria Tommasini Grotto; Ricardo Haddad; Sandra Coccuzzo Sampaio Vessoni; Simone Kashima; Svetoslav Nanev Slavov; Vincent Louis Viala |
| EPI_ISL_2445157, EPI_ISL_2473706 | ESF PARQUE COLINAS | Instituto Butantan | Antonio Jorge Martins; Claudia Renata dos Santos Barros; David Schlesinger; Debora Botequio Moretti; Dimas Tadeu Covas; Elaine Cristina Marqueze; Elaine Vieira Santos; Evandra Strazza Rodrigues; Heidge Fukumasu; Jayme Augusto de Souza-Neto; José Salvatore Leister Patané; Luiz Alcantara; Luiz Lehmann Coutinho; Maria Carolina Elias; Maurício Lacerda Nogueira; Rafael dos Santos Bezerra; Raul Machado Neto; Rejane Maria Tommasini Grotto; Ricardo Haddad; Sandra Coccuzzo Sampaio Vessoni; Simone Kashima; Svetoslav Nanev Slavov; Vincent Louis Viala |
| EPI_ISL_2445191, EPI_ISL_2445192, EPI_ISL_2473712 | ESF PQ UNIVERSITARIO DE ASSIS | Instituto Butantan | Antonio Jorge Martins; Claudia Renata dos Santos Barros; David Schlesinger; Debora Botequio Moretti; Dimas Tadeu Covas; Elaine Cristina Marqueze; Elaine Vieira Santos; Evandra Strazza Rodrigues; Heidge Fukumasu; Jayme Augusto de Souza-Neto; José Salvatore Leister Patané; Luiz Alcantara; Luiz Lehmann Coutinho; Maria Carolina Elias; Maurício Lacerda Nogueira; Rafael dos Santos Bezerra; Raul Machado Neto; Rejane Maria Tommasini Grotto; Ricardo Haddad; Sandra Coccuzzo Sampaio Vessoni; Simone Kashima; Svetoslav Nanev Slavov; Vincent Louis Viala |
| EPI_ISL_2344870 | ESF RURAL DR JOSE CICERO FONTES XAVIER | Instituto Butantan / ESALQ-Piracicaba | Antonio Jorge Martins; Claudia Renata dos Santos Barros; David Schlesinger; Debora Botequio Moretti; Dimas Tadeu Covas; Elaine Cristina Marqueze; Elaine Vieira Santos; Evandra Strazza Rodrigues; Heidge Fukumasu; Jayme Augusto de Souza-Neto; José Salvatore Leister Patané; Luiz Alcantara; Luiz Lehmann Coutinho; Maria Carolina Elias; Maurício Lacerda Nogueira; Rafael dos Santos Bezerra; Raul Machado Neto; Rejane Maria Tommasini Grotto; Ricardo Haddad; Sandra Coccuzzo Sampaio Vessoni; Simone Kashima; Svetoslav Nanev Slavov; Vincent Louis Viala |
| EPI_ISL_2445169, EPI_ISL_2445176 | ESF VILA GLORIA I | Instituto Butantan | Antonio Jorge Martins; Claudia Renata dos Santos Barros; David Schlesinger; Debora Botequio Moretti; Dimas Tadeu Covas; Elaine Cristina Marqueze; Elaine Vieira Santos; Evandra Strazza Rodrigues; Heidge Fukumasu; Jayme Augusto de Souza-Neto; José Salvatore Leister Patané; Luiz Alcantara; Luiz Lehmann Coutinho; Maria Carolina Elias; Maurício Lacerda Nogueira; Rafael dos Santos Bezerra; Raul Machado Neto; Rejane Maria Tommasini Grotto; Ricardo Haddad; Sandra Coccuzzo Sampaio Vessoni; Simone Kashima; Svetoslav Nanev Slavov; Vincent Louis Viala |
| EPI_ISL_2445171, EPI_ISL_2473816 | ESF VILA PROGRESSO | Instituto Butantan | Antonio Jorge Martins; Claudia Renata dos Santos Barros; David Schlesinger; Debora Botequio Moretti; Dimas Tadeu Covas; Elaine Cristina Marqueze; Elaine Vieira Santos; Evandra Strazza Rodrigues; Heidge Fukumasu; Jayme Augusto de Souza-Neto; José Salvatore Leister Patané; Luiz Alcantara; Luiz Lehmann Coutinho; Maria Carolina Elias; Maurício Lacerda Nogueira; Rafael dos Santos Bezerra; Raul Machado Neto; Rejane Maria Tommasini Grotto; Ricardo Haddad; Sandra Coccuzzo Sampaio Vessoni; Simone Kashima; Svetoslav Nanev Slavov; Vincent Louis Viala |
| EPI_ISL_2445158, EPI_ISL_2445160 | ESF VITORIA | Instituto Butantan | Antonio Jorge Martins; Claudia Renata dos Santos Barros; David Schlesinger; Debora Botequio Moretti; Dimas Tadeu Covas; Elaine Cristina Marqueze; Elaine Vieira Santos; Evandra Strazza Rodrigues; Heidge Fukumasu; Jayme Augusto de Souza-Neto; José Salvatore Leister Patané; Luiz Alcantara; Luiz Lehmann Coutinho; Maria Carolina Elias; Maurício Lacerda Nogueira; Rafael dos Santos Bezerra; Raul Machado Neto; Rejane Maria Tommasini Grotto; Ricardo Haddad; Sandra Coccuzzo Sampaio Vessoni; Simone Kashima; Svetoslav Nanev Slavov; Vincent Louis Viala |
| EPI_ISL_3215305, EPI_ISL_3215593, EPI_ISL_3215602, EPI_ISL_3215627, EPI_ISL_3215650, EPI_ISL_3215668, EPI_ISL_3215669, EPI_ISL_3215674, EPI_ISL_3215691, EPI_ISL_3215692, EPI_ISL_3215700, EPI_ISL_3215715, EPI_ISL_3215725, EPI_ISL_3215743, EPI_ISL_3215746 | | | |
| see above | EXCITE Lab | Andersen lab at Scripps Research | Abigail Schnapper; Angela Scioscia; Cheryl Anderson; Chip Schooley; David Pride; Greg Humphrey; Helena Tubb; Natasha Martin; Natasha Martin Cheryl Anderson; Sawyer Farmer; Sharon Reed; Smruthi Karthikeyan; Tommy Valles + SEARCH; Tommy Valles + SEARCH |
| EPI_ISL_2346154, EPI_ISL_2346155, EPI_ISL_3758745 | Emory Molecular Diagnostics Laboratory, Emory Healthcare | Piantadosi Lab, Emory Department of Pathology | Ahmed Babiker; Anne Piantadosi; Dara Khosravi |
| EPI_ISL_2316877, EPI_ISL_2316950, EPI_ISL_2316952, EPI_ISL_2316955, EPI_ISL_2317019, EPI_ISL_2317031, EPI_ISL_2471625, EPI_ISL_2471628, EPI_ISL_2471639, EPI_ISL_2471642, EPI_ISL_2471656 | | | |
| see above | Eurofins LifeCodexx GmbH | Robert Koch Institute | |
| EPI_ISL_2445114, EPI_ISL_2493708, EPI_ISL_2493709, EPI_ISL_2493710, EPI_ISL_2493712 | FACULDADE DE MEDICINA ABC | Instituto Butantan | Antonio Jorge Martins; Claudia Renata dos Santos Barros; David Schlesinger; Debora Botequio Moretti; Dimas Tadeu Covas; Elaine Cristina Marqueze; Elaine Vieira Santos; Evandra Strazza Rodrigues; Heidge Fukumasu; Jayme Augusto de Souza-Neto; José Salvatore Leister Patané; Luiz Alcantara; Luiz Lehmann Coutinho; Maria Carolina Elias; Maurício Lacerda Nogueira; Rafael dos Santos Bezerra; Raul Machado Neto; Rejane Maria Tommasini Grotto; Ricardo Haddad; Sandra Coccuzzo Sampaio Vessoni; Simone Kashima; Svetoslav Nanev Slavov; Vincent Louis Viala |
| EPI_ISL_3355485 | FUNDACIÓN HOSPITAL SAN PEDRO | Corporacion CorpoGen-Universidad de los Andes-Universidad Central | Christian Romero; Jorge Duitama; Juan Manuel Anzola; Laura González; Maryam Chaib De Mares; María Mercedes Zambrano; Nelly Díaz; Patricia Del Portillo; Silvia Restrepo |
| EPI_ISL_2008736, EPI_ISL_2277740, EPI_ISL_2277956, EPI_ISL_2277957, EPI_ISL_2277962, EPI_ISL_2277963, EPI_ISL_2277965, EPI_ISL_2277968, EPI_ISL_2277969, EPI_ISL_2277971, EPI_ISL_2277973, EPI_ISL_2277974, EPI_ISL_2277975, EPI_ISL_2277976, EPI_ISL_2442745, EPI_ISL_2442785, EPI_ISL_2442795, EPI_ISL_2442809, EPI_ISL_2442831, EPI_ISL_2442833, EPI_ISL_2442946, EPI_ISL_2442947, EPI_ISL_2442965, EPI_ISL_2567092, EPI_ISL_2567093, EPI_ISL_2567099, EPI_ISL_2567147, EPI_ISL_2567164, EPI_ISL_2567165, EPI_ISL_2567178, EPI_ISL_2836028, EPI_ISL_2836031, EPI_ISL_2836105, EPI_ISL_2836330, EPI_ISL_2836449, EPI_ISL_2836676 | | | |
| see above | Florida Bureau of Public Health Laboratories | Florida Bureau of Public Health Laboratories | Jason Blanton; Sarah Schmedes |
| EPI_ISL_2620708, EPI_ISL_2620954 | Fondazione IRCCS Ca' Granda Ospedale Maggiore Policlinico | Fondazione IRCCS Ca' Granda Ospedale Maggiore Policlinico | Ferruccio Ceriotti; Sara Uceda Renteria |
| EPI_ISL_1839261, EPI_ISL_1839596, EPI_ISL_1839893, EPI_ISL_1839980, EPI_ISL_1839997, EPI_ISL_1840063, EPI_ISL_1840076, EPI_ISL_1840092, EPI_ISL_1840093, EPI_ISL_1840116, EPI_ISL_1992053, EPI_ISL_1992082, EPI_ISL_1992088, EPI_ISL_1992152, EPI_ISL_1992174, EPI_ISL_1992201, EPI_ISL_1992202, EPI_ISL_1992203, EPI_ISL_1992206, EPI_ISL_1992212, EPI_ISL_1992287, EPI_ISL_1992317, EPI_ISL_1992345, EPI_ISL_1992348, EPI_ISL_1992448, EPI_ISL_1992449, EPI_ISL_1992457, EPI_ISL_1992542, EPI_ISL_1992587, EPI_ISL_1992643, EPI_ISL_1992800, EPI_ISL_1992805, EPI_ISL_1992813, EPI_ISL_1992855, EPI_ISL_1992892, EPI_ISL_1992921, EPI_ISL_1992981, EPI_ISL_1992984, EPI_ISL_1992985, EPI_ISL_1993040, EPI_ISL_1993067, EPI_ISL_1993076, EPI_ISL_1993170, EPI_ISL_1993173, EPI_ISL_1993176, EPI_ISL_1993177, EPI_ISL_1993183, EPI_ISL_1993190, EPI_ISL_1993195, EPI_ISL_1993217, EPI_ISL_1993249, EPI_ISL_1993252, EPI_ISL_1993255, EPI_ISL_1993270, EPI_ISL_1993357, EPI_ISL_2043693, EPI_ISL_2043701, EPI_ISL_2043712, EPI_ISL_2043777, EPI_ISL_2043865, EPI_ISL_2043880, EPI_ISL_2043953, EPI_ISL_2043969, EPI_ISL_2043976, EPI_ISL_2043977, EPI_ISL_2043988, EPI_ISL_2133941, EPI_ISL_2133988, EPI_ISL_2134052, EPI_ISL_2134084, EPI_ISL_2134148, EPI_ISL_2134151, EPI_ISL_2134177, EPI_ISL_2134189, EPI_ISL_2134191, EPI_ISL_2134192, EPI_ISL_2134194, EPI_ISL_2134225, EPI_ISL_2134245, EPI_ISL_2134312, EPI_ISL_2134321, EPI_ISL_2134332, EPI_ISL_2134337, EPI_ISL_2134345, EPI_ISL_2134433, EPI_ISL_2134434, EPI_ISL_2134442, EPI_ISL_2134464, EPI_ISL_2134465, EPI_ISL_2134466, EPI_ISL_2134471, EPI_ISL_2134499, EPI_ISL_2134502, EPI_ISL_2134514, EPI_ISL_2160765, EPI_ISL_2160778, EPI_ISL_2160829, EPI_ISL_2160837, EPI_ISL_2160845, EPI_ISL_2160903, EPI_ISL_2160904, EPI_ISL_2160909, EPI_ISL_2160935, EPI_ISL_2160988, EPI_ISL_2161001, EPI_ISL_2161024, EPI_ISL_2179864, EPI_ISL_2179894, EPI_ISL_2179900, EPI_ISL_2269221, EPI_ISL_2269251, EPI_ISL_2269256, EPI_ISL_2269277, EPI_ISL_2269314, EPI_ISL_2269360, EPI_ISL_2269368, EPI_ISL_2269371, EPI_ISL_2269436, EPI_ISL_2269502, EPI_ISL_2269503, EPI_ISL_2269509, EPI_ISL_2269525, EPI_ISL_2280263, EPI_ISL_2368135, EPI_ISL_2371518, EPI_ISL_2371573, EPI_ISL_2371575, EPI_ISL_2371576, EPI_ISL_2371578, EPI_ISL_2371579, EPI_ISL_2371580, EPI_ISL_2371584, EPI_ISL_2371585, EPI_ISL_2371587, EPI_ISL_2371639, EPI_ISL_2441831, EPI_ISL_2480073, EPI_ISL_2525557, EPI_ISL_2525633, EPI_ISL_2526057 | | | |
| see above | Fulgent Genetics | Centers for Disease Control and Prevention Division of Viral Diseases, Pathogen Discovery | Adrian Paskey; Becky Tsai; Benafsh Sapra; Benjamin Rambo-Martin; Christopher Gulvick; Clinton R. Paden; Dakota Howard; Darlene Wagner; Dhwani Batra; Doreen Ng; Duncan MacCannell; Harry Gao; James Xie; Jason Caravas; John Gao; Joseph Fierro; Kara Moser; Matthew Schmerer; Mickey Li; Peter W. Cook; Scott Sammons; Shatavia Morrison; Yan Meng; Yvette Unoarumhi |
| EPI_ISL_2550708 | GA Department of Public Health | GA Department of Public Health | Aliyah Fields; Cynthia Dixey; Jonathan Edwards; Stacy Reeves; Taylor Smith; Tonia Parrott |
| EPI_ISL_2229306 | GA Department of Public Health Laboratory | Centers for Disease Control and Prevention Division of Viral Diseases, Pathogen Discovery | Alison Laufer Halpin; Ben L. Rambo-Martin; Clinton R. Paden; Dakota Howard; Darlene Wagner; Dave Wentworth; Dhwani Batra; Jasmine Padilla; Justin Lee; Katie Dillon; Krista Queen; Kristen Knipe; Kristine Lacek; Mark Burroughs; Matthew Schmerer; Mili Sheth; Peter Cook; Sam Shepard; Sarah Nobles; Shoshona Le; Suxiang Tong; Vivien Dugan; Yvette Unoarumhi |
| EPI_ISL_2311749, EPI_ISL_2422457 | GA Department of Public Health Laboratory | Genomics and Discovery, Respiratory Viruses Branch, Division of Viral Diseases, Centers for Disease Control and Prevention | Adam Retchless; Anna Kelleher; Anna Uehara; Brian Lynch; Clinton R. Paden; Dhwani Batra; Haibin Wang; Han Jia Justin Ng; Jasmine Padilla; Jing Zhang; Justin Lee; Krista Queen; Mark Burroughs; Mili Sheth; Morgan Davis; Peter Cook; Rachel Marine; Sarah Nobles; Suxiang Tong; Tara Coalter; Yan Li; Ying Tao |
| EPI_ISL_2228873 | GH A.CHENEVIER-H.MONDOR | Department of Virology, Henri Mondor University Hospital, Assistance Publique Hôpitaux de Paris, Université Paris-Est Créteil, INSERM U955 | Alexandre Soulier; Christophe Rodriguez; Elisabeth Trawinski; Guillaume Gricourt; Jean-Michel Pawlotsky; Melissa N'Debi; Slim Fourati; Vanessa Demontant |

| | | | |
|---|---|---|---|
| EPI_ISL_2421160, EPI_ISL_3031056 | GLENS FALLS HOSPITAL LABORATORY | Wadsworth Center, New York State Department of Health | Alexis Russell; Catharine Prussing; Daryl M. Lamson; Erasmus Schneider; Erica Lasek-Nesselquist; John Kelly; Jonathan Plitnick; Kirsten St. George; Matthew Shudt; Melissa A Leisner; Navjot Singh |
| EPI_ISL_2345663, EPI_ISL_2345701, EPI_ISL_2345709, EPI_ISL_2345756 | GRIPARIO FALCAO | Instituto Butantan / Mendelics | Antonio Jorge Martins; Claudia Renata dos Santos Barros; David Schlesinger; Debora Botequio Moretti; Dimas Tadeu Covas; Elaine Cristina Marqueze; Elaine Vieira Santos; Evandra Strazza Rodrigues; Heidge Fukumasu; Jayme Augusto de Souza-Neto; José Salvatore Leister Patané; Luiz Alcantara; Luiz Lehmann Coutinho; Maria Carolina Elias; Maurício Lacerda Nogueira; Rafael dos Santos Bezerra; Raul Machado Neto; Rejane Maria Tommasini Grotto; Ricardo Haddad; Sandra Coccuzzo Sampaio Vessoni; Simone Kashima; Svetoslav Nanev Slavov; Vincent Louis Viala |
| EPI_ISL_2228704, EPI_ISL_2333688 | GROUPE HOSPITALIER SUD ILE DE FRANCE | Department of Virology, Henri Mondor University Hospital, Assistance Publique Hôpitaux de Paris, Université Paris-Est Créteil, INSERM U955 | Alexandre Soulier; Christophe Rodriguez; Elisabeth Trawinski; Guillaume Gricourt; Jean-Michel Pawlotsky; Melissa N'Debi; Slim Fourati; Vanessa Demontant |
| EPI_ISL_2421228 | Garnet Health Medical Center Laboratory | Wadsworth Center, New York State Department of Health | Alexis Russell; Catharine Prussing; Daryl M. Lamson; Erasmus Schneider; Erica Lasek-Nesselquist; John Kelly; Jonathan Plitnick; Kirsten St. George; Matthew Shudt; Melissa A Leisner; Navjot Singh |
| EPI_ISL_3491798, EPI_ISL_3491799, EPI_ISL_3491800, EPI_ISL_3491803, EPI_ISL_3491805, EPI_ISL_3491810, EPI_ISL_3491820, EPI_ISL_3491822, EPI_ISL_3491824, EPI_ISL_3491825, EPI_ISL_3491835 | | | |
| see above | Gencore - Universidad de los Andes | Gencore - Universidad de los Andes | Catherine Jaller; Cristian Barrera; David González; Luisa Sacristan; Marcela Guevara; Silvia Restrepo |
| EPI_ISL_2009545, EPI_ISL_2009556, EPI_ISL_2009558, EPI_ISL_2009559, EPI_ISL_2391013, EPI_ISL_2391030, EPI_ISL_2391049, EPI_ISL_2391068, EPI_ISL_2391086, EPI_ISL_2391087, EPI_ISL_2391093, EPI_ISL_2391099, EPI_ISL_2391100, EPI_ISL_2391124, EPI_ISL_2391125, EPI_ISL_2391133, EPI_ISL_2391153, EPI_ISL_2391157, EPI_ISL_2391173, EPI_ISL_2391180, EPI_ISL_2391185, EPI_ISL_2391187, EPI_ISL_2391190, EPI_ISL_2391191, EPI_ISL_2391196, EPI_ISL_2391208, EPI_ISL_2391214, EPI_ISL_2391217, EPI_ISL_2391238, EPI_ISL_2391241, EPI_ISL_2391248, EPI_ISL_2391250, EPI_ISL_2391252, EPI_ISL_2391253, EPI_ISL_2391254, EPI_ISL_2391255, EPI_ISL_2391259, EPI_ISL_2391260, EPI_ISL_2391261, EPI_ISL_2391263, EPI_ISL_2391266, EPI_ISL_2391268, EPI_ISL_2391270, EPI_ISL_2391293, EPI_ISL_2391296, EPI_ISL_2391297, EPI_ISL_2391299, EPI_ISL_2391303, EPI_ISL_2391306, EPI_ISL_2391310, EPI_ISL_2391319, EPI_ISL_2391322, EPI_ISL_2391323, EPI_ISL_2391326, EPI_ISL_2391327, EPI_ISL_2391336, EPI_ISL_2391342, EPI_ISL_2391345, EPI_ISL_2391347, EPI_ISL_2391348, EPI_ISL_2391352, EPI_ISL_2391356, EPI_ISL_2391393, EPI_ISL_2391407, EPI_ISL_2391408, EPI_ISL_2391415, EPI_ISL_2391416, EPI_ISL_2391419, EPI_ISL_2391420, EPI_ISL_2391437, EPI_ISL_2391439, EPI_ISL_2391442, EPI_ISL_2391446, EPI_ISL_2391448, EPI_ISL_2391449, EPI_ISL_2391470, EPI_ISL_2391471, EPI_ISL_2391475, EPI_ISL_2391476, EPI_ISL_2508551, EPI_ISL_2508613, EPI_ISL_2508616, EPI_ISL_2509122, EPI_ISL_2509130, EPI_ISL_2509131, EPI_ISL_2509132, EPI_ISL_2509133, EPI_ISL_2509134, EPI_ISL_2509135, EPI_ISL_2509136, EPI_ISL_2509138, EPI_ISL_2509139, EPI_ISL_2509141, EPI_ISL_2509145, EPI_ISL_2509155, EPI_ISL_2509156, EPI_ISL_2509166, EPI_ISL_2509167, EPI_ISL_2509282, EPI_ISL_2509288, EPI_ISL_2509300, EPI_ISL_2509302, EPI_ISL_2509308, EPI_ISL_2509324, EPI_ISL_2543449, EPI_ISL_2543452, EPI_ISL_2543454, EPI_ISL_2543457, EPI_ISL_2543458, EPI_ISL_2543461, EPI_ISL_2543464, EPI_ISL_2543465, EPI_ISL_2543469, EPI_ISL_2543470, EPI_ISL_2543471, EPI_ISL_2543482, EPI_ISL_2543483, EPI_ISL_2543484, EPI_ISL_2543489, EPI_ISL_2543492, EPI_ISL_2543494, EPI_ISL_2543496, EPI_ISL_2543497, EPI_ISL_2543499, EPI_ISL_2543500, EPI_ISL_2543509, EPI_ISL_2543510, EPI_ISL_2543515, EPI_ISL_2543542, EPI_ISL_2543543, EPI_ISL_2543548, EPI_ISL_2543549, EPI_ISL_2543551, EPI_ISL_2543617, EPI_ISL_2543620, EPI_ISL_2557264, EPI_ISL_2557268, EPI_ISL_2566533, EPI_ISL_2597267, EPI_ISL_2597268, EPI_ISL_2597271, EPI_ISL_2597272, EPI_ISL_2597273, EPI_ISL_2597274, EPI_ISL_2597278, EPI_ISL_2597279, EPI_ISL_2597283, EPI_ISL_2892726 | | | |
| see above | Genetica Molecular and Subdepartamento de Virologia ISP Chile | Instituto de Salud Publica de Chile | Andres Castillo; Barbara Parra; Constanza Campano; Gisselle Barra; Javier Tognarelli; Jorge Fernandez; Karen Orostica; Loredana Arata; Patricia Bustos; Rodrigo Fasce; Soledad Ulloa |
| EPI_ISL_2248888, EPI_ISL_2340184, EPI_ISL_2340221, EPI_ISL_2340225 | Germano de Sousa | Instituto Nacional de Saude (INSA) | Borges et al |
| EPI_ISL_2340344 | Germano de Sousa | Instituto Nacional de Saude (INSA) and BioSystems & Integrative Sciences Institute (BioISI) Genomics Unit, FCUL | Borges et al |
| EPI_ISL_2360225 | Gesundheitsdirektion Zürich | Institute of Medical Virology | Alexandra Trkola; Annette Audigé; Cyril Shah; Gabriela Ziltener; Guido Bloemberg; Jon Huder; Jürg Böni; Kevin Steiner; Maria Grünberg; Maryam Zaheri; Michael Huber; Riccarda Capaul; Stefan Schmutz; Verena Kufner |
| EPI_ISL_2535843, EPI_ISL_2535845, EPI_ISL_2535863, EPI_ISL_2535873, EPI_ISL_2535878, EPI_ISL_2535904, EPI_ISL_2535977, EPI_ISL_2536023, EPI_ISL_2536025, EPI_ISL_2536036, EPI_ISL_2537097, EPI_ISL_2537099, EPI_ISL_2537108, EPI_ISL_2537109, EPI_ISL_2601808, EPI_ISL_2601821, EPI_ISL_2601869, EPI_ISL_2601908, EPI_ISL_2601911, EPI_ISL_2602027, EPI_ISL_2602034, EPI_ISL_2602055, EPI_ISL_2602060, EPI_ISL_2602076, EPI_ISL_2602079, EPI_ISL_2602080, EPI_ISL_2602099, EPI_ISL_2602149, EPI_ISL_2602248, EPI_ISL_2602278, EPI_ISL_2602309, EPI_ISL_2602381, EPI_ISL_2613840, EPI_ISL_2613860, EPI_ISL_2613863, EPI_ISL_2613873, EPI_ISL_2613877, EPI_ISL_2613885, EPI_ISL_2613889, EPI_ISL_2613892, EPI_ISL_2613908, EPI_ISL_2614004, EPI_ISL_2614011, EPI_ISL_2614012, EPI_ISL_2614024, EPI_ISL_2614027, EPI_ISL_2614039, EPI_ISL_2614055, EPI_ISL_2854839, EPI_ISL_2854858, EPI_ISL_2854860, EPI_ISL_2854917 | | | |
| see above | Gravity Diagnostics, LLC | Gravity Diagnostics, LLC | Gravity Diagnostics |
| EPI_ISL_1745232, EPI_ISL_1960121, EPI_ISL_2157237, EPI_ISL_2157239, EPI_ISL_2157248, EPI_ISL_2376238, EPI_ISL_2376239, EPI_ISL_2376243, EPI_ISL_2376247, EPI_ISL_2376249, EPI_ISL_2500991 | | | |
| see above | Gundersen Clinical Microbiology Laboratory | Kabara Cancer Research Institute | Craig S. Richmond; Paraic A. Kenny |
| EPI_ISL_3102213 | HGCC HOSPITAL GERAL DR CESAR CALS | Analytical Competence Molecular Epidemiology Lab/ACME, Oswaldo Cruz Foundation, Ceara (FIOCRUZ CE) | Cleber Furtado Aksenen; Fabio Miyajima; Fernanda Braga Stehling; Francisco Eder de Moura Lopes; Jamille Maria Mendes Bezerra; Joaquim César do Nascimento Sousa Junior; Pedro Miguel Carneiro Jeronimo; Suzana Porto Almeida e Lucas Delerino; Thais Ferreira de Oliveira; Thais de Oliveira Costa; Ticiane Cavalcante de Souza; Veridiana Pessoa Miyajima |
| EPI_ISL_1941834 | HIB | Laboratory of genomics and metagenomics | Claire Bertelli; Damien Jacot; Gilbert Greub; Sebastien Aeby; Trestan Pillonel |
| EPI_ISL_2017370, EPI_ISL_2017372, EPI_ISL_2017384, EPI_ISL_2017385, EPI_ISL_2017386, EPI_ISL_2017387, EPI_ISL_2017389, EPI_ISL_2017390, EPI_ISL_2017432, EPI_ISL_2017434, EPI_ISL_2017435, EPI_ISL_2017437, EPI_ISL_2017438, EPI_ISL_2017439, EPI_ISL_2187985, EPI_ISL_2187987, EPI_ISL_2187988, EPI_ISL_2187990, EPI_ISL_2187991, EPI_ISL_2187992, EPI_ISL_2187994, EPI_ISL_2187995, EPI_ISL_2187997, EPI_ISL_2187999, EPI_ISL_2188001, EPI_ISL_2188002, EPI_ISL_2188004, EPI_ISL_2188005, EPI_ISL_2188012, EPI_ISL_2188013, EPI_ISL_2188014, EPI_ISL_2188015, EPI_ISL_2188016, EPI_ISL_2188017, EPI_ISL_2188018, EPI_ISL_2188019, EPI_ISL_2188020, EPI_ISL_2188021, EPI_ISL_2188023, EPI_ISL_2188024, EPI_ISL_2188025, EPI_ISL_2188027, EPI_ISL_2222900, EPI_ISL_2222902, EPI_ISL_2222905, EPI_ISL_2222911, EPI_ISL_2222916, EPI_ISL_2222917, EPI_ISL_2222918, EPI_ISL_2227564, EPI_ISL_2227565, EPI_ISL_2348604, EPI_ISL_2348605, EPI_ISL_2348606, EPI_ISL_2348607, EPI_ISL_2348608, EPI_ISL_2348609, EPI_ISL_2348610, EPI_ISL_2348611, EPI_ISL_2348612, EPI_ISL_2348614, EPI_ISL_2348615, EPI_ISL_2466448, EPI_ISL_2466449, EPI_ISL_2466450, EPI_ISL_2466451, EPI_ISL_2466452, EPI_ISL_2466454, EPI_ISL_2466455, EPI_ISL_2466456, EPI_ISL_2466457, EPI_ISL_2466458, EPI_ISL_2466459, EPI_ISL_2466460, EPI_ISL_2466461, EPI_ISL_2466462, EPI_ISL_2466464, EPI_ISL_2466465, EPI_ISL_2497437, EPI_ISL_2497438, EPI_ISL_2497439, EPI_ISL_2497441, EPI_ISL_2497442, EPI_ISL_2497465, EPI_ISL_2617584, EPI_ISL_2617585, EPI_ISL_2617586, EPI_ISL_3254316, EPI_ISL_3254321, EPI_ISL_3254465, EPI_ISL_4061646 | | | |
| see above | HLAGYN - Laboratorio de Imunologia de Transplantes de Goias | HLAGYN - Laboratorio de Imunologia de Transplantes de Goias | Alessandro Leonardo Alvares Magalhaes; Daniel Ferreira de Sousa; Danielle de Paiva Rezende; Erika Lopes Rocha Batista; Fernando Antonio Vinhal dos Santos; Frederico Rodrigues Vinhal; Kamila Oliveira Reis De Freitas.; Lucas Carlos Gomes Pereira; Paola Cristina Resende Silva; Sabrina Sara Moreira Duarte |
| EPI_ISL_2209313, EPI_ISL_2344914, EPI_ISL_2344948 | HOSP E MATERNIDADE MUNICIPAL N SRA MONTE SERRAT | Instituto Butantan / ESALQ-Piracicaba | Antonio Jorge Martins; Claudia Renata dos Santos Barros; David Schlesinger; Debora Botequio Moretti; Dimas Tadeu Covas; Elaine Cristina Marqueze; Elaine Vieira Santos; Evandra Strazza Rodrigues; Heidge Fukumasu; Jayme Augusto de Souza-Neto; José Salvatore Leister Patané; Luiz Alcantara; Luiz Lehmann Coutinho; Maria Carolina Elias; Maurício Lacerda Nogueira; Rafael dos Santos Bezerra; Raul Machado Neto; Rejane Maria Tommasini Grotto; Ricardo Haddad; Sandra Coccuzzo Sampaio Vessoni; Simone Kashima; Svetoslav Nanev Slavov; Vincent Louis Viala |
| EPI_ISL_2345764, EPI_ISL_2362254 | HOSP E MATERNIDADE MUNICIPAL N SRA MONTE SERRAT | Instituto Butantan / Mendelics | Antonio Jorge Martins; Claudia Renata dos Santos Barros; David Schlesinger; Debora Botequio Moretti; Dimas Tadeu Covas; Elaine Cristina Marqueze; Elaine Vieira Santos; Evandra Strazza Rodrigues; Heidge Fukumasu; Jayme Augusto de Souza-Neto; José Salvatore Leister Patané; Luiz Alcantara; Luiz Lehmann Coutinho; Maria Carolina Elias; Maurício Lacerda Nogueira; Rafael dos Santos Bezerra; Raul Machado Neto; Rejane Maria Tommasini Grotto; Ricardo Haddad; Sandra Coccuzzo Sampaio Vessoni; Simone Kashima; Svetoslav Nanev Slavov; Vincent Louis Viala |
| EPI_ISL_2473728, EPI_ISL_2473729, EPI_ISL_2473749, EPI_ISL_2473758, EPI_ISL_2473759 | HOSP GERAL DE SAO PAULO | Instituto Butantan | Antonio Jorge Martins; Claudia Renata dos Santos Barros; David Schlesinger; Debora Botequio Moretti; Dimas Tadeu Covas; Elaine Cristina Marqueze; Elaine Vieira Santos; Evandra Strazza Rodrigues; Heidge Fukumasu; Jayme Augusto de Souza-Neto; José Salvatore Leister Patané; Luiz Alcantara; Luiz Lehmann Coutinho; Maria Carolina Elias; Maurício Lacerda Nogueira; Rafael dos Santos Bezerra; Raul Machado Neto; Rejane Maria Tommasini Grotto; Ricardo Haddad; Sandra Coccuzzo Sampaio Vessoni; Simone Kashima; Svetoslav Nanev Slavov; Vincent Louis Viala |
| EPI_ISL_2345110 | HOSP MAT 8 DE MAIO | Instituto Butantan / ESALQ-Piracicaba | Antonio Jorge Martins; Claudia Renata dos Santos Barros; David Schlesinger; Debora Botequio Moretti; Dimas Tadeu Covas; Elaine Cristina Marqueze; Elaine Vieira Santos; Evandra Strazza Rodrigues; Heidge Fukumasu; Jayme Augusto de Souza-Neto; José Salvatore Leister Patané; Luiz Alcantara; Luiz Lehmann Coutinho; Maria Carolina Elias; Maurício Lacerda Nogueira; Rafael dos Santos Bezerra; Raul Machado Neto; Rejane Maria Tommasini Grotto; Ricardo Haddad; Sandra Coccuzzo Sampaio Vessoni; Simone Kashima; Svetoslav Nanev Slavov; Vincent Louis Viala |
| EPI_ISL_2209024, EPI_ISL_2345061, EPI_ISL_2345064, EPI_ISL_2345074, EPI_ISL_2345075 | HOSP MUN CIDADE TIRADENTES CARMEN PRUDENTE | Instituto Butantan / ESALQ-Piracicaba | Antonio Jorge Martins; Claudia Renata dos Santos Barros; David Schlesinger; Debora Botequio Moretti; Dimas Tadeu Covas; Elaine Cristina Marqueze; Elaine Vieira Santos; Evandra Strazza Rodrigues; Heidge Fukumasu; Jayme Augusto de Souza-Neto; José Salvatore Leister Patané; Luiz Alcantara; Luiz Lehmann Coutinho; Maria Carolina Elias; Maurício Lacerda Nogueira; Rafael dos Santos Bezerra; Raul Machado Neto; Rejane Maria Tommasini Grotto; Ricardo Haddad; Sandra Coccuzzo Sampaio Vessoni; Simone Kashima; Svetoslav Nanev Slavov; Vincent Louis Viala |
| EPI_ISL_2209383, EPI_ISL_2345043, EPI_ISL_2345067, EPI_ISL_2345068, EPI_ISL_2345071, EPI_ISL_2345072, EPI_ISL_2345073 | | | |
| see above | HOSP MUN DE MOGI DAS CRUZES PREF WALDEMAR COSTA FILHO | Instituto Butantan / ESALQ-Piracicaba | Antonio Jorge Martins; Claudia Renata dos Santos Barros; David Schlesinger; Debora Botequio Moretti; Dimas Tadeu Covas; Elaine Cristina Marqueze; Elaine Vieira Santos; Evandra Strazza Rodrigues; Heidge Fukumasu; Jayme Augusto de Souza-Neto; José Salvatore Leister Patané; Luiz Alcantara; Luiz Lehmann Coutinho; Maria Carolina Elias; Maurício Lacerda Nogueira; Rafael dos Santos Bezerra; Raul Machado Neto; Rejane Maria Tommasini Grotto; Ricardo Haddad; Sandra Coccuzzo Sampaio Vessoni; Simone Kashima; Svetoslav Nanev Slavov; Vincent Louis Viala |
| EPI_ISL_2345076, EPI_ISL_2345081, EPI_ISL_2345104, EPI_ISL_2345108 | HOSP MUN JOSANIAS CASTANHA BRAGA | Instituto Butantan / ESALQ-Piracicaba | Antonio Jorge Martins; Claudia Renata dos Santos Barros; David Schlesinger; Debora Botequio Moretti; Dimas Tadeu Covas; Elaine Cristina Marqueze; Elaine Vieira Santos; Evandra Strazza Rodrigues; Heidge Fukumasu; Jayme Augusto de Souza-Neto; José Salvatore Leister Patané; Luiz Alcantara; Luiz Lehmann Coutinho; Maria Carolina Elias; Maurício Lacerda Nogueira; Rafael dos Santos Bezerra; Raul Machado Neto; Rejane Maria Tommasini Grotto; Ricardo Haddad; Sandra Coccuzzo Sampaio Vessoni; Simone Kashima; Svetoslav Nanev Slavov; Vincent Louis Viala |
| EPI_ISL_3102518 | HOSPITAL MATERNIDADE DR LUIS DE GONZAGA DA FONSECA MOTA | Analytical Competence Molecular Epidemiology Lab/ACME, Oswaldo Cruz Foundation, Ceara (FIOCRUZ CE) | Cleber Furtado Aksenen; Fabio Miyajima; Fernanda Braga Stehling; Francisco Eder de Moura Lopes; Jamille Maria Mendes Bezerra; Joaquim César do Nascimento Sousa Junior; Pedro Miguel Carneiro Jeronimo; Suzana Porto Almeida e Lucas Delerino; Thais Ferreira de Oliveira; Thais de Oliveira Costa; Ticiane Cavalcante de Souza; Veridiana Pessoa Miyajima |
| EPI_ISL_2100346 | HOSPITAL CAN MISSES | HOSPITAL UNIVERSITARIO SON ESPASES | Antonio Oliver; Carla López-Causapé; Pablo Fraile-Ribot; SeqCovid |
| EPI_ISL_3102255 | HOSPITAL CURA DARS | Analytical Competence Molecular Epidemiology Lab/ACME, Oswaldo Cruz Foundation, Ceara (FIOCRUZ CE) | Cleber Furtado Aksenen; Fabio Miyajima; Fernanda Braga Stehling; Francisco Eder de Moura Lopes; Jamille Maria Mendes Bezerra; Joaquim César do Nascimento Sousa Junior; Pedro Miguel Carneiro Jeronimo; Suzana Porto Almeida e Lucas Delerino; Thais Ferreira de Oliveira; Thais de Oliveira Costa; Ticiane Cavalcante de Souza; Veridiana Pessoa Miyajima |
| EPI_ISL_2345044, EPI_ISL_2345045, EPI_ISL_2345066 | HOSPITAL DE CAMPANHA COVID 19 CAIEIRAS | Instituto Butantan / ESALQ-Piracicaba | Antonio Jorge Martins; Claudia Renata dos Santos Barros; David Schlesinger; Debora Botequio Moretti; Dimas Tadeu Covas; Elaine Cristina Marqueze; Elaine Vieira Santos; Evandra Strazza Rodrigues; Heidge Fukumasu; Jayme Augusto de Souza-Neto; José Salvatore Leister Patané; Luiz Alcantara; Luiz Lehmann Coutinho; Maria Carolina Elias; Maurício Lacerda Nogueira; Rafael dos Santos Bezerra; Raul Machado Neto; Rejane Maria Tommasini Grotto; Ricardo Haddad; Sandra Coccuzzo Sampaio Vessoni; Simone Kashima; Svetoslav Nanev Slavov; Vincent Louis Viala |
| EPI_ISL_2445571, EPI_ISL_2445572, EPI_ISL_2445577, EPI_ISL_2445579, EPI_ISL_2445583, EPI_ISL_2445588, EPI_ISL_2445591, EPI_ISL_2473800 | | | |
| see above | HOSPITAL DE CAMPANHA COVID 19 MUNICIPIO DE TAUBATE | Instituto Butantan | Antonio Jorge Martins; Claudia Renata dos Santos Barros; David Schlesinger; Debora Botequio Moretti; Dimas Tadeu Covas; Elaine Cristina Marqueze; Elaine Vieira Santos; Evandra Strazza Rodrigues; Heidge Fukumasu; Jayme Augusto de Souza-Neto; José Salvatore Leister Patané; Luiz Alcantara; Luiz Lehmann Coutinho; Maria Carolina Elias; Maurício Lacerda Nogueira; Rafael dos Santos Bezerra; Raul Machado Neto; Rejane Maria Tommasini Grotto; Ricardo Haddad; Sandra Coccuzzo Sampaio Vessoni; Simone Kashima; Svetoslav Nanev Slavov; Vincent Louis Viala |
| EPI_ISL_2344955, EPI_ISL_2344956, EPI_ISL_2344957, EPI_ISL_2344973, EPI_ISL_2344975, EPI_ISL_2344977, EPI_ISL_2344978, EPI_ISL_2344980, EPI_ISL_2344997, EPI_ISL_2345005, EPI_ISL_2345008, EPI_ISL_2345011, EPI_ISL_2345018, EPI_ISL_2345021 | | | |

| | | | |
|---|---|---|---|
| see above | HOSPITAL DE CAMPANHA COVID 19 MUNICIPIO DE TAUBATE | Instituto Butantan / ESALQ-Piracicaba | Antonio Jorge Martins; Claudia Renata dos Santos Barros; David Schlesinger; Debora Botequio Moretti; Dimas Tadeu Covas; Elaine Cristina Marqueze; Elaine Vieira Santos; Evandra Strazza Rodrigues; Heidge Fukumasu; Jayme Augusto de Souza-Neto; José Salvatore Leister Patané; Luiz Alcantara; Luiz Lehmann Coutinho; Maria Carolina Elias; Maurício Lacerda Nogueira; Rafael dos Santos Bezerra; Raul Machado Neto; Rejane Maria Tommasini Grotto; Ricardo Haddad; Sandra Coccuzzo Sampaio Vessoni; Simone Kashima; Svetoslav Nanev Slavov; Vincent Louis Viala |
| EPI_ISL_2209101, EPI_ISL_2209363, EPI_ISL_2209883, EPI_ISL_2210101, EPI_ISL_2210236, EPI_ISL_2345722, EPI_ISL_2345731, EPI_ISL_2345819, EPI_ISL_2345826, EPI_ISL_2345870 | | | |
| see above | HOSPITAL DE CAMPANHA COVID 19 MUNICIPIO DE TAUBATE | Instituto Butantan / Mendelics | Antonio Jorge Martins; Claudia Renata dos Santos Barros; David Schlesinger; Debora Botequio Moretti; Dimas Tadeu Covas; Elaine Cristina Marqueze; Elaine Vieira Santos; Evandra Strazza Rodrigues; Heidge Fukumasu; Jayme Augusto de Souza-Neto; José Salvatore Leister Patané; Luiz Alcantara; Luiz Lehmann Coutinho; Maria Carolina Elias; Maurício Lacerda Nogueira; Rafael dos Santos Bezerra; Raul Machado Neto; Rejane Maria Tommasini Grotto; Ricardo Haddad; Sandra Coccuzzo Sampaio Vessoni; Simone Kashima; Svetoslav Nanev Slavov; Vincent Louis Viala |
| EPI_ISL_2344848, EPI_ISL_2344849 | HOSPITAL DE MISERICORDIA ALTINOPOLIS | Instituto Butantan / ESALQ-Piracicaba | Antonio Jorge Martins; Claudia Renata dos Santos Barros; David Schlesinger; Debora Botequio Moretti; Dimas Tadeu Covas; Elaine Cristina Marqueze; Elaine Vieira Santos; Evandra Strazza Rodrigues; Heidge Fukumasu; Jayme Augusto de Souza-Neto; José Salvatore Leister Patané; Luiz Alcantara; Luiz Lehmann Coutinho; Maria Carolina Elias; Maurício Lacerda Nogueira; Rafael dos Santos Bezerra; Raul Machado Neto; Rejane Maria Tommasini Grotto; Ricardo Haddad; Sandra Coccuzzo Sampaio Vessoni; Simone Kashima; Svetoslav Nanev Slavov; Vincent Louis Viala |
| EPI_ISL_2493305, EPI_ISL_2493306, EPI_ISL_2493307, EPI_ISL_2493308, EPI_ISL_2493309, EPI_ISL_2493312, EPI_ISL_2493316, EPI_ISL_2493318, EPI_ISL_2493320, EPI_ISL_2493321, EPI_ISL_2493322, EPI_ISL_2493323, EPI_ISL_2493324, EPI_ISL_2493325, EPI_ISL_2493330, EPI_ISL_2493331, EPI_ISL_2493334 | | | |
| see above | HOSPITAL DO RIM E HIPERTENSAO | Instituto Butantan | Antonio Jorge Martins; Claudia Renata dos Santos Barros; David Schlesinger; Debora Botequio Moretti; Dimas Tadeu Covas; Elaine Cristina Marqueze; Elaine Vieira Santos; Evandra Strazza Rodrigues; Heidge Fukumasu; Jayme Augusto de Souza-Neto; José Salvatore Leister Patané; Luiz Alcantara; Luiz Lehmann Coutinho; Maria Carolina Elias; Maurício Lacerda Nogueira; Rafael dos Santos Bezerra; Raul Machado Neto; Rejane Maria Tommasini Grotto; Ricardo Haddad; Sandra Coccuzzo Sampaio Vessoni; Simone Kashima; Svetoslav Nanev Slavov; Vincent Louis Viala |
| EPI_ISL_2345131 | HOSPITAL DR OSIRIS FLORINDO COELHO FERRAZ DE VASCONCELOS | Instituto Butantan / ESALQ-Piracicaba | Antonio Jorge Martins; Claudia Renata dos Santos Barros; David Schlesinger; Debora Botequio Moretti; Dimas Tadeu Covas; Elaine Cristina Marqueze; Elaine Vieira Santos; Evandra Strazza Rodrigues; Heidge Fukumasu; Jayme Augusto de Souza-Neto; José Salvatore Leister Patané; Luiz Alcantara; Luiz Lehmann Coutinho; Maria Carolina Elias; Maurício Lacerda Nogueira; Rafael dos Santos Bezerra; Raul Machado Neto; Rejane Maria Tommasini Grotto; Ricardo Haddad; Sandra Coccuzzo Sampaio Vessoni; Simone Kashima; Svetoslav Nanev Slavov; Vincent Louis Viala |
| EPI_ISL_2493370 | HOSPITAL E MATERNIDADE MADRE VANNINI | Instituto Butantan | Antonio Jorge Martins; Claudia Renata dos Santos Barros; David Schlesinger; Debora Botequio Moretti; Dimas Tadeu Covas; Elaine Cristina Marqueze; Elaine Vieira Santos; Evandra Strazza Rodrigues; Heidge Fukumasu; Jayme Augusto de Souza-Neto; José Salvatore Leister Patané; Luiz Alcantara; Luiz Lehmann Coutinho; Maria Carolina Elias; Maurício Lacerda Nogueira; Rafael dos Santos Bezerra; Raul Machado Neto; Rejane Maria Tommasini Grotto; Ricardo Haddad; Sandra Coccuzzo Sampaio Vessoni; Simone Kashima; Svetoslav Nanev Slavov; Vincent Louis Viala |
| EPI_ISL_2344982 | HOSPITAL E MATERNIDADE NOSSA SENHORA DA AJUDA | Instituto Butantan / ESALQ-Piracicaba | Antonio Jorge Martins; Claudia Renata dos Santos Barros; David Schlesinger; Debora Botequio Moretti; Dimas Tadeu Covas; Elaine Cristina Marqueze; Elaine Vieira Santos; Evandra Strazza Rodrigues; Heidge Fukumasu; Jayme Augusto de Souza-Neto; José Salvatore Leister Patané; Luiz Alcantara; Luiz Lehmann Coutinho; Maria Carolina Elias; Maurício Lacerda Nogueira; Rafael dos Santos Bezerra; Raul Machado Neto; Rejane Maria Tommasini Grotto; Ricardo Haddad; Sandra Coccuzzo Sampaio Vessoni; Simone Kashima; Svetoslav Nanev Slavov; Vincent Louis Viala |
| EPI_ISL_2209292 | HOSPITAL E MATERNIDADE NOSSA SENHORA DA AJUDA | Instituto Butantan / Mendelics | Antonio Jorge Martins; Claudia Renata dos Santos Barros; David Schlesinger; Debora Botequio Moretti; Dimas Tadeu Covas; Elaine Cristina Marqueze; Elaine Vieira Santos; Evandra Strazza Rodrigues; Heidge Fukumasu; Jayme Augusto de Souza-Neto; José Salvatore Leister Patané; Luiz Alcantara; Luiz Lehmann Coutinho; Maria Carolina Elias; Maurício Lacerda Nogueira; Rafael dos Santos Bezerra; Raul Machado Neto; Rejane Maria Tommasini Grotto; Ricardo Haddad; Sandra Coccuzzo Sampaio Vessoni; Simone Kashima; Svetoslav Nanev Slavov; Vincent Louis Viala |
| EPI_ISL_2209269, EPI_ISL_2209315, EPI_ISL_2344899, EPI_ISL_2344911, EPI_ISL_2344944, EPI_ISL_2344945, EPI_ISL_2344946 | | | |
| see above | HOSPITAL E PRONTO SOCORRO PORTINARI | Instituto Butantan | Antonio Jorge Martins; Claudia Renata dos Santos Barros; David Schlesinger; Debora Botequio Moretti; Dimas Tadeu Covas; Elaine Cristina Marqueze; Elaine Vieira Santos; Evandra Strazza Rodrigues; Heidge Fukumasu; Jayme Augusto de Souza-Neto; José Salvatore Leister Patané; Luiz Alcantara; Luiz Lehmann Coutinho; Maria Carolina Elias; Maurício Lacerda Nogueira; Rafael dos Santos Bezerra; Raul Machado Neto; Rejane Maria Tommasini Grotto; Ricardo Haddad; Sandra Coccuzzo Sampaio Vessoni; Simone Kashima; Svetoslav Nanev Slavov; Vincent Louis Viala; Vincent Louis Viala. |
| EPI_ISL_1967299 | HOSPITAL E PRONTO SOCORRO PORTINARI | Instituto Butantan / Mendelics | Antonio Jorge Martins; Bianca Cechetto Carlos. Mendelics: Bibiana Santos; Claudia Renata dos Santos Barros; Cíntia Bittar; David Schlesinger. Hemocentro Ribeirão Preto: Simone Kashima; Debora Botequio Moretti; Elaine Cristina Marqueze; Elaine Vieira dos Santos; Elisangela Chicaroni Mattos; Erika Freitas; Evandra Strazza Rodrigues; Felipe Allan da Silva da Costa; Flavia Aburjaile; Fábio Sossai Possebon; Guilherme Campos; Guilherme Targino Valente; Heidge Fukumasu. USP-Botucatu: Rejane Maria Tommasini Grotto; Helena Lage Ferreira; Instituto Butantan: Dimas Tadeu Covas; Jardelina de Souza Todao Bernardino; Jayme A. Souza-Neto; Jessika Cristina Chagas Lesbon; Jorge A. Petroli Marchesi; José Salvatore Leister Patané; João Paulo Kitajima; João Pessoa Araújo Jr.; Leila Sabrina Ullmann; Loyze Paola Oliveira de Lima; Luiz Aurelio de Campos Crispin. Centro de Genômica Funcional da ESALQ: Luiz Lehmann Coutinho; Luiz Carlos Junior de Alcantara; Lívia Sacchetto; Maisa C. Pereira Parra; Maria Carolina Elias; Marta Giovanetti; Marília Moraes; Maurício Lacerda Nogueira. Prefeitura de Sao Paulo: Melissa Palmieri.; Patricia Akemi Assato; Paula Rahal; Paulo Inacio da Costa; Rafael dos Santos Bezerra; Raquel de Lello Rocha Campos Cassano. NGS Soluções Genômicas: Pilar Drummond Sampaio Corrêa Mariani. FZEA-USP Pirassununga: Mirele Daiana Poleti; Raul Machado Neto; Ricardo Augusto Brassaloti; Ricardo Haddad; Rodrigo Tocantins Calado. FAMERP-SJRP: Cecília Artico Banho; Sandra Coccuzzo Sampaio; Svetoslav Nanev Slavov; Vagner Fonseca; Vincent Louis Viala |
| EPI_ISL_2345029 | HOSPITAL ESCOLA DA UNIVERSIDADE DE TAUBATE | Instituto Butantan / ESALQ-Piracicaba | Antonio Jorge Martins; Claudia Renata dos Santos Barros; David Schlesinger; Debora Botequio Moretti; Dimas Tadeu Covas; Elaine Cristina Marqueze; Elaine Vieira Santos; Evandra Strazza Rodrigues; Heidge Fukumasu; Jayme Augusto de Souza-Neto; José Salvatore Leister Patané; Luiz Alcantara; Luiz Lehmann Coutinho; Maria Carolina Elias; Maurício Lacerda Nogueira; Rafael dos Santos Bezerra; Raul Machado Neto; Rejane Maria Tommasini Grotto; Ricardo Haddad; Sandra Coccuzzo Sampaio Vessoni; Simone Kashima; Svetoslav Nanev Slavov; Vincent Louis Viala |
| EPI_ISL_3102276, EPI_ISL_3102289, EPI_ISL_3102290, EPI_ISL_3102317 | HOSPITAL ESTADUAL LEONARDO DA VINCI | Analytical Competence Molecular Epidemiology Lab/ACME, Oswaldo Cruz Foundation, Ceara (FIOCRUZ CE) | Cleber Furtado Aksenen; Fabio Miyajima; Fernando Braga Stehling; Francisco Eder de Moura Lopes; Jamille Maria Mendes Bezerra; Joaquim César do Nascimento Sousa Junior; Pedro Miguel Carneiro Jeronimo; Suzana Porto Almeida e Lucas Delerino; Thais Ferreira de Oliveira; Thais de Oliveira Costa; Ticiane Cavalcante de Souza; Veridiana Pessoa Miyajima |
| EPI_ISL_3260869 | HOSPITAL GENERAL UNIVERSITARIO DE ALBACETE | Instituto de Salud Carlos III | A. Monzón; CARIDAD; F. Casas; I. Jiménez; I. SAINZ DE BARANDA CAMINO; M. Camarero; P. Zaballos; S. Cuesta; S. Iglesias-Caballero; S. Pozo; S. Varona; Sandonís; V. Vázquez-Morón |
| EPI_ISL_3260824, EPI_ISL_3260876 | HOSPITAL GENERAL UNIVERSITARIO DE GUADALAJARA | Instituto de Salud Carlos III | A. Monzón; ALEJANDRO; F. Casas; I. Jiménez; I.GONZALEZ PRAETORIUS; Iglesias-Caballero; M. Sandonís; P. Zaballos; S. Camarero; S. Cuesta; S. Iglesias-Caballero; S. Pozo; S. Varona; V. Camarero; V. Vázquez-Morón; Vázquez-Morón |
| EPI_ISL_3260840 | HOSPITAL GENERAL UNIVERSITARIO SANTA LUCIA | Instituto de Salud Carlos III | A. Monzón; ANA; F. Casas; I. Jiménez; I.BLAZQUEZ ABELLAN; M. Sandonís; P. Zaballos; S. Cuesta; S. Iglesias-Caballero; S. Pozo; S. Varona; V. Camarero; Vázquez-Morón |
| EPI_ISL_2345143 | HOSPITAL GERAL JESUS TEIXEIRA DA COSTA GUAIANASES SAO PAULO | Instituto Butantan / ESALQ-Piracicaba | Antonio Jorge Martins; Claudia Renata dos Santos Barros; David Schlesinger; Debora Botequio Moretti; Dimas Tadeu Covas; Elaine Cristina Marqueze; Elaine Vieira Santos; Evandra Strazza Rodrigues; Heidge Fukumasu; Jayme Augusto de Souza-Neto; José Salvatore Leister Patané; Luiz Alcantara; Luiz Lehmann Coutinho; Maria Carolina Elias; Maurício Lacerda Nogueira; Rafael dos Santos Bezerra; Raul Machado Neto; Rejane Maria Tommasini Grotto; Ricardo Haddad; Sandra Coccuzzo Sampaio Vessoni; Simone Kashima; Svetoslav Nanev Slavov; Vincent Louis Viala |
| EPI_ISL_3102287, EPI_ISL_3102320 | HOSPITAL MATERNIDADE SANTA RITA | Analytical Competence Molecular Epidemiology Lab/ACME, Oswaldo Cruz Foundation, Ceara (FIOCRUZ CE) | Cleber Furtado Aksenen; Fabio Miyajima; Fernando Braga Stehling; Francisco Eder de Moura Lopes; Jamille Maria Mendes Bezerra; Joaquim César do Nascimento Sousa Junior; Pedro Miguel Carneiro Jeronimo; Suzana Porto Almeida e Lucas Delerino; Thais Ferreira de Oliveira; Thais de Oliveira Costa; Ticiane Cavalcante de Souza; Veridiana Pessoa Miyajima |
| EPI_ISL_2493687, EPI_ISL_2493689 | HOSPITAL MUNICIPAL DE IBIUNA IBIUNA SP | Instituto Butantan | Antonio Jorge Martins; Claudia Renata dos Santos Barros; David Schlesinger; Debora Botequio Moretti; Dimas Tadeu Covas; Elaine Cristina Marqueze; Elaine Vieira Santos; Evandra Strazza Rodrigues; Heidge Fukumasu; Jayme Augusto de Souza-Neto; José Salvatore Leister Patané; Luiz Alcantara; Luiz Lehmann Coutinho; Maria Carolina Elias; Maurício Lacerda Nogueira; Rafael dos Santos Bezerra; Raul Machado Neto; Rejane Maria Tommasini Grotto; Ricardo Haddad; Sandra Coccuzzo Sampaio Vessoni; Simone Kashima; Svetoslav Nanev Slavov; Vincent Louis Viala |
| EPI_ISL_2208890 | HOSPITAL MUNICIPAL DE IBIUNA IBIUNA SP | Instituto Butantan / Mendelics | Antonio Jorge Martins; Claudia Renata dos Santos Barros; David Schlesinger; Debora Botequio Moretti; Dimas Tadeu Covas; Elaine Cristina Marqueze; Elaine Vieira Santos; Evandra Strazza Rodrigues; Heidge Fukumasu; Jayme Augusto de Souza-Neto; José Salvatore Leister Patané; Luiz Alcantara; Luiz Lehmann Coutinho; Maria Carolina Elias; Maurício Lacerda Nogueira; Rafael dos Santos Bezerra; Raul Machado Neto; Rejane Maria Tommasini Grotto; Ricardo Haddad; Sandra Coccuzzo Sampaio Vessoni; Simone Kashima; Svetoslav Nanev Slavov; Vincent Louis Viala |
| EPI_ISL_2493253, EPI_ISL_2493254, EPI_ISL_2493592, EPI_ISL_2493593, EPI_ISL_2493609, EPI_ISL_2493650, EPI_ISL_2493651 | | | |
| see above | HOSPITAL MUNICIPAL DE ILHABELA GOV MARIO COVAS JR | Instituto Butantan | Antonio Jorge Martins; Claudia Renata dos Santos Barros; David Schlesinger; Debora Botequio Moretti; Dimas Tadeu Covas; Elaine Cristina Marqueze; Elaine Vieira Santos; Evandra Strazza Rodrigues; Heidge Fukumasu; Jayme Augusto de Souza-Neto; José Salvatore Leister Patané; Luiz Alcantara; Luiz Lehmann Coutinho; Maria Carolina Elias; Maurício Lacerda Nogueira; Rafael dos Santos Bezerra; Raul Machado Neto; Rejane Maria Tommasini Grotto; Ricardo Haddad; Sandra Coccuzzo Sampaio Vessoni; Simone Kashima; Svetoslav Nanev Slavov; Vincent Louis Viala |
| EPI_ISL_2493628 | HOSPITAL MUNICIPAL DE ITAPIRA | Instituto Butantan | Antonio Jorge Martins; Claudia Renata dos Santos Barros; David Schlesinger; Debora Botequio Moretti; Dimas Tadeu Covas; Elaine Cristina Marqueze; Elaine Vieira Santos; Evandra Strazza Rodrigues; Heidge Fukumasu; Jayme Augusto de Souza-Neto; José Salvatore Leister Patané; Luiz Alcantara; Luiz Lehmann Coutinho; Maria Carolina Elias; Maurício Lacerda Nogueira; Rafael dos Santos Bezerra; Raul Machado Neto; Rejane Maria Tommasini Grotto; Ricardo Haddad; Sandra Coccuzzo Sampaio Vessoni; Simone Kashima; Svetoslav Nanev Slavov; Vincent Louis Viala |
| EPI_ISL_2209603, EPI_ISL_2345774 | HOSPITAL MUNICIPAL DE ITAPIRA | Instituto Butantan / Mendelics | Antonio Jorge Martins; Claudia Renata dos Santos Barros; David Schlesinger; Debora Botequio Moretti; Dimas Tadeu Covas; Elaine Cristina Marqueze; Elaine Vieira Santos; Evandra Strazza Rodrigues; Heidge Fukumasu; Jayme Augusto de Souza-Neto; José Salvatore Leister Patané; Luiz Alcantara; Luiz Lehmann Coutinho; Maria Carolina Elias; Maurício Lacerda Nogueira; Rafael dos Santos Bezerra; Raul Machado Neto; Rejane Maria Tommasini Grotto; Ricardo Haddad; Sandra Coccuzzo Sampaio Vessoni; Simone Kashima; Svetoslav Nanev Slavov; Vincent Louis Viala |
| EPI_ISL_2445394, EPI_ISL_2445397, EPI_ISL_2445400, EPI_ISL_2445403, EPI_ISL_2445408, EPI_ISL_2445410, EPI_ISL_2445414, EPI_ISL_2493519, EPI_ISL_2493520, EPI_ISL_2493522 | | | |
| see above | HOSPITAL MUNICIPAL DR MARIO GATTI CAMPINAS | Instituto Butantan | Antonio Jorge Martins; Claudia Renata dos Santos Barros; David Schlesinger; Debora Botequio Moretti; Dimas Tadeu Covas; Elaine Cristina Marqueze; Elaine Vieira Santos; Evandra Strazza Rodrigues; Heidge Fukumasu; Jayme Augusto de Souza-Neto; José Salvatore Leister Patané; Luiz Alcantara; Luiz Lehmann Coutinho; Maria Carolina Elias; Maurício Lacerda Nogueira; Rafael dos Santos Bezerra; Raul Machado Neto; Rejane Maria Tommasini Grotto; Ricardo Haddad; Sandra Coccuzzo Sampaio Vessoni; Simone Kashima; Svetoslav Nanev Slavov; Vincent Louis Viala |
| EPI_ISL_2445506, EPI_ISL_2445507, EPI_ISL_2445509, EPI_ISL_2493392, EPI_ISL_2493523, EPI_ISL_2493524 | HOSPITAL MUNICIPAL DR TABAJARA RAMOS | Instituto Butantan | Antonio Jorge Martins; Claudia Renata dos Santos Barros; David Schlesinger; Debora Botequio Moretti; Dimas Tadeu Covas; Elaine Cristina Marqueze; Elaine Vieira Santos; Evandra Strazza Rodrigues; Heidge Fukumasu; Jayme Augusto de Souza-Neto; José Salvatore Leister Patané; Luiz Alcantara; Luiz Lehmann Coutinho; Maria Carolina Elias; Maurício Lacerda Nogueira; Rafael dos Santos Bezerra; Raul Machado Neto; Rejane Maria Tommasini Grotto; Ricardo Haddad; Sandra Coccuzzo Sampaio Vessoni; Simone Kashima; Svetoslav Nanev Slavov; Vincent Louis Viala |
| EPI_ISL_2362259 | HOSPITAL MUNICIPAL DR WALDEMAR TEBALDI | Instituto Butantan / Mendelics | Antonio Jorge Martins; Claudia Renata dos Santos Barros; David Schlesinger; Debora Botequio Moretti; Dimas Tadeu Covas; Elaine Cristina Marqueze; Elaine Vieira Santos; Evandra Strazza Rodrigues; Heidge Fukumasu; Jayme Augusto de Souza-Neto; José Salvatore Leister Patané; Luiz Alcantara; Luiz Lehmann Coutinho; Maria Carolina Elias; Maurício Lacerda Nogueira; Rafael dos Santos Bezerra; Raul Machado Neto; Rejane Maria Tommasini Grotto; Ricardo Haddad; Sandra Coccuzzo Sampaio Vessoni; Simone Kashima; Svetoslav Nanev Slavov; Vincent Louis Viala |
| EPI_ISL_2493441 | HOSPITAL MUNICIPAL JORGE CURY DE SANTA MARIA DA SERRA | Instituto Butantan | Antonio Jorge Martins; Claudia Renata dos Santos Barros; David Schlesinger; Debora Botequio Moretti; Dimas Tadeu Covas; Elaine Cristina Marqueze; Elaine Vieira Santos; Evandra Strazza Rodrigues; Heidge Fukumasu; Jayme Augusto de Souza-Neto; José Salvatore Leister Patané; Luiz Alcantara; Luiz Lehmann Coutinho; Maria Carolina Elias; Maurício Lacerda Nogueira; Rafael dos Santos Bezerra; Raul Machado Neto; Rejane Maria Tommasini Grotto; Ricardo Haddad; Sandra Coccuzzo Sampaio Vessoni; Simone Kashima; Svetoslav Nanev Slavov; Vincent Louis Viala |
| EPI_ISL_2493688 | HOSPITAL MUNICIPAL LEONARDUS VAN MELLIS | Instituto Butantan | Antonio Jorge Martins; Claudia Renata dos Santos Barros; David Schlesinger; Debora Botequio Moretti; Dimas Tadeu Covas; Elaine Cristina Marqueze; Elaine Vieira Santos; Evandra Strazza Rodrigues; Heidge Fukumasu; Jayme Augusto de Souza-Neto; José Salvatore Leister Patané; Luiz Alcantara; Luiz Lehmann Coutinho; Maria Carolina Elias; Maurício Lacerda Nogueira; Rafael dos Santos Bezerra; Raul Machado Neto; Rejane Maria Tommasini Grotto; Ricardo Haddad; Sandra Coccuzzo Sampaio Vessoni; Simone Kashima; Svetoslav Nanev Slavov; Vincent Louis Viala |
| EPI_ISL_2801335, EPI_ISL_2801336, EPI_ISL_2801345, EPI_ISL_2801352, EPI_ISL_2801358, EPI_ISL_2801361, EPI_ISL_2801362, EPI_ISL_2801368, EPI_ISL_3102296, EPI_ISL_3102517 | | | |
| see above | HOSPITAL MUNICIPAL NOSSA SENHORA DOS MILAGRES | Analytical Competence Molecular Epidemiology Lab/ACME, Oswaldo Cruz Foundation, Ceara (FIOCRUZ CE) | Cleber Furtado Aksenen; Cleber Furtado Aksenen e Suzana Porto Almeida; Fabio Miyajima; Fernando Braga Stehling; Francisco Eder de Moura Lopes; Jamille Maria Mendes Bezerra; Joaquim César do Nascimento Sousa Junior; Pedro Miguel Carneiro Jeronimo; Suzana Porto Almeida e Lucas Delerino; Thais Ferreira de Oliveira; Thais de Oliveira Costa; Ticiane Cavalcante de Souza; Veridiana Pessoa Miyajima |
| EPI_ISL_2445230 | HOSPITAL MUNICIPAL REYNALDO GUERRA CAJATI | Instituto Butantan | Antonio Jorge Martins; Claudia Renata dos Santos Barros; David Schlesinger; Debora Botequio Moretti; Dimas Tadeu Covas; Elaine Cristina Marqueze; Elaine Vieira Santos; Evandra Strazza Rodrigues; Heidge Fukumasu; Jayme Augusto de Souza-Neto; José Salvatore Leister Patané; Luiz Alcantara; Luiz Lehmann Coutinho; Maria Carolina Elias; Maurício Lacerda Nogueira; Rafael dos Santos Bezerra; Raul Machado Neto; Rejane Maria Tommasini Grotto; Ricardo Haddad; Sandra Coccuzzo Sampaio Vessoni; Simone Kashima; Svetoslav Nanev Slavov; Vincent Louis Viala |
| EPI_ISL_2344960, EPI_ISL_2344969 | HOSPITAL MUNICIPAL REYNALDO GUERRA CAJATI | Instituto Butantan / ESALQ-Piracicaba | Antonio Jorge Martins; Claudia Renata dos Santos Barros; David Schlesinger; Debora Botequio Moretti; Dimas Tadeu Covas; Elaine Cristina Marqueze; Elaine Vieira Santos; Evandra Strazza Rodrigues; Heidge Fukumasu; Jayme Augusto de Souza-Neto; José Salvatore Leister Patané; Luiz Alcantara; Luiz Lehmann Coutinho; Maria Carolina Elias; Maurício Lacerda Nogueira; Rafael dos Santos Bezerra; Raul Machado Neto; Rejane Maria Tommasini Grotto; Ricardo Haddad; Sandra Coccuzzo Sampaio Vessoni; Simone Kashima; Svetoslav Nanev Slavov; Vincent Louis Viala |
| EPI_ISL_2493296 | HOSPITAL MUNICIPAL SANTO ANTONIO | Instituto Butantan | Antonio Jorge Martins; Claudia Renata dos Santos Barros; David Schlesinger; Debora Botequio Moretti; Dimas Tadeu Covas; Elaine Cristina Marqueze; Elaine Vieira Santos; Evandra Strazza Rodrigues; Heidge Fukumasu; Jayme Augusto de Souza-Neto; José Salvatore Leister Patané; Luiz Alcantara; Luiz Lehmann Coutinho; Maria Carolina Elias; Maurício Lacerda Nogueira; Rafael dos Santos Bezerra; Raul Machado Neto; Rejane Maria Tommasini Grotto; Ricardo Haddad; Sandra Coccuzzo Sampaio Vessoni; Simone Kashima; Svetoslav Nanev Slavov; Vincent Louis Viala |
| EPI_ISL_2345130 | HOSPITAL NOSSA SENHORA APARECIDA | Instituto Butantan / ESALQ-Piracicaba | Antonio Jorge Martins; Claudia Renata dos Santos Barros; David Schlesinger; Debora Botequio Moretti; Dimas Tadeu Covas; Elaine Cristina Marqueze; Elaine Vieira Santos; Evandra Strazza Rodrigues; Heidge Fukumasu; Jayme Augusto de Souza-Neto; José Salvatore Leister Patané; Luiz Alcantara; Luiz Lehmann Coutinho; Maria Carolina Elias; Maurício Lacerda Nogueira; Rafael dos Santos Bezerra; Raul Machado Neto; Rejane Maria Tommasini Grotto; Ricardo Haddad; Sandra Coccuzzo Sampaio Vessoni; Simone Kashima; Svetoslav Nanev Slavov; Vincent Louis Viala |
| EPI_ISL_2801367, EPI_ISL_3102319, EPI_ISL_3102513 | HOSPITAL OTOCLINICA | Analytical Competence Molecular Epidemiology Lab/ACME, Oswaldo Cruz Foundation, Ceara (FIOCRUZ CE) | Cleber Furtado Aksenen; Cleber Furtado Aksenen e Suzana Porto Almeida; Fabio Miyajima; Fernando Braga Stehling; Francisco Eder de Moura Lopes; Jamille Maria Mendes Bezerra; Joaquim César do Nascimento Sousa Junior; Pedro Miguel Carneiro Jeronimo; Suzana Porto Almeida e Lucas Delerino; Thais Ferreira de Oliveira; Thais de Oliveira Costa; Ticiane Cavalcante de Souza; Veridiana Pessoa Miyajima |
| EPI_ISL_2445550 | HOSPITAL REGIONAL DE ITAPETININGA | Instituto Butantan | Antonio Jorge Martins; Claudia Renata dos Santos Barros; David Schlesinger; Debora Botequio Moretti; Dimas Tadeu Covas; Elaine Cristina Marqueze; Elaine Vieira Santos; Evandra Strazza Rodrigues; Heidge Fukumasu; Jayme Augusto de Souza-Neto; José Salvatore Leister Patané; Luiz Alcantara; Luiz Lehmann Coutinho; Maria Carolina Elias; Maurício Lacerda Nogueira; Rafael dos Santos Bezerra; Raul Machado Neto; Rejane Maria Tommasini Grotto; Ricardo Haddad; Sandra Coccuzzo Sampaio Vessoni; Simone Kashima; Svetoslav Nanev Slavov; Vincent Louis Viala |
| EPI_ISL_2801334 | HOSPITAL REGIONAL DO CARIRI | Analytical Competence Molecular Epidemiology | Cleber Furtado Aksenen e Suzana Porto Almeida; Fabio Miyajima; Fernando Braga Stehling; Francisco Eder de Moura Lopes; Jamille Maria Mendes Bezerra; Joaquim César do Nascimento Sousa Junior; Pedro Miguel Carneiro Jeronimo; Thais Ferreira de Oliveira; Thais de Oliveira Costa; Ticiane Cavalcante de Souza; Veridiana Pessoa Miyajima |

| | | | |
|---|---|---|---|
| | | Lab/ACME, Oswaldo Cruz Foundation, Ceara (FIOCRUZ CE) | |
| EPI_ISL_3102267 | HOSPITAL REGIONAL NORTE | Analytical Competence Molecular Epidemiology Lab/ACME, Oswaldo Cruz Foundation, Ceara (FIOCRUZ CE) | Cleber Furtado Aksenen; Fabio Miyajima; Fernando Braga Stehling; Francisco Eder de Moura Lopes; Jamille Maria Mendes Bezerra; Joaquim César do Nascimento Sousa Junior; Pedro Miguel Carneiro Jeronimo; Suzana Porto Almeida e Lucas Delerino; Thais Ferreira de Oliveira; Thais de Oliveira Costa; Ticiane Cavalcante de Souza; Veridiana Pessoa Miyajima |
| EPI_ISL_2658278 | HOSPITAL SAN FRANCISCO DE ASIS | Inciensa, Instituto Costarricense de Investigación y Enseñanza en Nutrición y Salud | Adriana Godínez; Claudio Soto-Garita; Estela Cordero; Francisco Duarte; Hebleen Porras; Joselyn Prado & Juan Carlos Villalobos-Ugalde; José Luis Vargas; Mariela Gutiérrez; Melany Calderón |
| EPI_ISL_2502760, EPI_ISL_2502762 | HOSPITAL SAN JUAN DE DIOS | Inciensa, Instituto Costarricense de InvestigaciOn y Enseñanza en NutriciOn y Salud | Adriana Godínez; Claudio Soto-Garita; Estela Cordero; Francisco Duarte; Hebleen Porras; Jose Luis Vargas; Joselyn Prado & Marco Cháves-Otárola; Mariela Gutierrez; Melany CalderOn |
| EPI_ISL_2502719 | HOSPITAL SAN VICENTE DE PAUL | Inciensa, Instituto Costarricense de InvestigaciOn y Enseñanza en NutriciOn y Salud | Adriana Godínez; Claudio Soto-Garita; Estela Cordero; Francisco Duarte; Hebleen Porras; Jose Luis Vargas; Joselyn Prado & Silvia Saenz-Garcia; Mariela Gutierrez; Melany CalderOn |
| EPI_ISL_2445220 | HOSPITAL SANTO ANTONIO DE JUQUIA JUQUIA | Instituto Butantan | Antonio Jorge Martins; Claudia Renata dos Santos Barros; David Schlesinger; Debora Botequio Moretti; Dimas Tadeu Covas; Elaine Cristina Marqueze; Elaine Vieira Santos; Evandra Strazza Rodrigues; Heidge Fukumasu; Jayme Augusto de Souza-Neto; José Salvatore Leister Patané; Luiz Alcantara; Luiz Lehmann Coutinho; Maria Carolina Elias; Maurício Lacerda Nogueira; Rafael dos Santos Bezerra; Raul Machado Neto; Rejane Maria Tommasini Grotto; Ricardo Haddad; Sandra Coccuzzo Sampaio Vessoni; Simone Kashima; Svetoslav Nanev Slavov; Vincent Louis Viala |
| EPI_ISL_2445242 | HOSPITAL SAO FRANCISCO SC LTDA | Instituto Butantan | Antonio Jorge Martins; Claudia Renata dos Santos Barros; David Schlesinger; Debora Botequio Moretti; Dimas Tadeu Covas; Elaine Cristina Marqueze; Elaine Vieira Santos; Evandra Strazza Rodrigues; Heidge Fukumasu; Jayme Augusto de Souza-Neto; José Salvatore Leister Patané; Luiz Alcantara; Luiz Lehmann Coutinho; Maria Carolina Elias; Maurício Lacerda Nogueira; Rafael dos Santos Bezerra; Raul Machado Neto; Rejane Maria Tommasini Grotto; Ricardo Haddad; Sandra Coccuzzo Sampaio Vessoni; Simone Kashima; Svetoslav Nanev Slavov; Vincent Louis Viala |
| EPI_ISL_2801369 | HOSPITAL SAO MATEUS | Analytical Competence Molecular Epidemiology Lab/ACME, Oswaldo Cruz Foundation, Ceara (FIOCRUZ CE) | Cleber Furtado Aksenen e Suzana Porto Almeida; Fabio Miyajima; Fernando Braga Stehling; Francisco Eder de Moura Lopes; Jamille Maria Mendes Bezerra; Joaquim César do Nascimento Sousa Junior; Pedro Miguel Carneiro Jeronimo; Thais Ferreira de Oliveira; Thais de Oliveira Costa; Ticiane Cavalcante de Souza; Veridiana Pessoa Miyajima |
| EPI_ISL_3502489, EPI_ISL_3502727, EPI_ISL_3502728, EPI_ISL_3502729, EPI_ISL_3502741, EPI_ISL_3502744, EPI_ISL_3502746, EPI_ISL_3502750, EPI_ISL_3502754, EPI_ISL_3502760, EPI_ISL_3532472 | | | |
| see above | HOSPITAL UNIVERSITARIO 12 DE OCTUBRE | HOSPITAL UNIVERSITARIO 12 DE OCTUBRE | Carmen Martín-Higuera; Esther Viedma; Irene Múñoz-Gallego; M.ª Dolores Folgueira; Mar Aguilera; Noelia Moral; Rafael Delgado; Sagrario Zurita |
| EPI_ISL_2100300, EPI_ISL_2100328, EPI_ISL_2100340, EPI_ISL_2100342, EPI_ISL_2134884, EPI_ISL_2134934, EPI_ISL_2241439, EPI_ISL_2241449, EPI_ISL_2241457 | | | |
| see above | HOSPITAL UNIVERSITARIO SON ESPASES | HOSPITAL UNIVERSITARIO SON ESPASES | Antonio Oliver; Carla López-Causapé; Pablo Fraile-Ribot; SeqCovid |
| EPI_ISL_2241452 | HOSPITAL UNIVERSITARIO SON LLATZER | HOSPITAL UNIVERSITARIO SON ESPASES | Antonio Oliver; Carla López-Causapé; Pablo Fraile-Ribot; SeqCovid |
| EPI_ISL_2738508, EPI_ISL_2738515, EPI_ISL_2738517, EPI_ISL_3235352, EPI_ISL_3235354, EPI_ISL_3235356, EPI_ISL_3235358 | | | |
| see above | Hackensack Medical Center | New York Genome Center | Amy Baldwin; Andre Corvelo; Barry Kreiswirth; David Perlin; Dayna M. Oschwald; Jose Mediavilla; Kaelea Composto; Kar Chow; Liang Chen; Marcus Cunningham; Michael Zody; Samantha Fennessey; Tom Maniatis |
| EPI_ISL_2006656 | Health Services Laboratories | Wellcome Sanger Institute for the COVID-19 Genomics UK (COG-UK) Consortium | Cordelia Langford; David K. Jackson; Dominic Kwiatkowski; Ewan Harrison; Health Services Laboratories and Alex Alderton; Ian Johnston; Jeffrey Barrett; John Sillitoe on behalf of the Wellcome Sanger Institute COVID-19 Surveillance Team; Roberto Amato; Sonia Goncalves |
| EPI_ISL_2727870 | HealthPartners Central Lab | Minnesota Department of Health, Public Health Laboratory | Alexandra Lorentz; Jacob Garfin; Matt Plumb; and Xiong Wang |
| EPI_ISL_1990624, EPI_ISL_1990644, EPI_ISL_1990668, EPI_ISL_1990737, EPI_ISL_1990757, EPI_ISL_1990804, EPI_ISL_1990814, EPI_ISL_1990827, EPI_ISL_1991374, EPI_ISL_1991493, EPI_ISL_1991524, EPI_ISL_1991580, EPI_ISL_1991601, EPI_ISL_1991667, EPI_ISL_1991677, EPI_ISL_2009737, EPI_ISL_2009745, EPI_ISL_2009777, EPI_ISL_2009807, EPI_ISL_2009875, EPI_ISL_2009880, EPI_ISL_2009889, EPI_ISL_2009898, EPI_ISL_2009900, EPI_ISL_2009921, EPI_ISL_2009932, EPI_ISL_2009951, EPI_ISL_2010001, EPI_ISL_2010013, EPI_ISL_2010031, EPI_ISL_2010037, EPI_ISL_2010084, EPI_ISL_2010096, EPI_ISL_2010122, EPI_ISL_2010132, EPI_ISL_2010149, EPI_ISL_2010185, EPI_ISL_2010274, EPI_ISL_2010282, EPI_ISL_2010312, EPI_ISL_2010351, EPI_ISL_2010395, EPI_ISL_2010400, EPI_ISL_2010420, EPI_ISL_2010422, EPI_ISL_2010428, EPI_ISL_2010442, EPI_ISL_2010462, EPI_ISL_2010485, EPI_ISL_2010515, EPI_ISL_2010521, EPI_ISL_2010534, EPI_ISL_2010580, EPI_ISL_2010622, EPI_ISL_2010671, EPI_ISL_2010741, EPI_ISL_2010749, EPI_ISL_2010807, EPI_ISL_2035280, EPI_ISL_2097347, EPI_ISL_2097388, EPI_ISL_2097390, EPI_ISL_2097397, EPI_ISL_2097399, EPI_ISL_2097425, EPI_ISL_2097433, EPI_ISL_2097435, EPI_ISL_2097448, EPI_ISL_2097462, EPI_ISL_2097463, EPI_ISL_2097466, EPI_ISL_2097467, EPI_ISL_2097498, EPI_ISL_2097573, EPI_ISL_2097593, EPI_ISL_2097594, EPI_ISL_2097635, EPI_ISL_2097639, EPI_ISL_2097654, EPI_ISL_2097659, EPI_ISL_2097668, EPI_ISL_2097747, EPI_ISL_2097753, EPI_ISL_2097762, EPI_ISL_2097850, EPI_ISL_2097857, EPI_ISL_2097862, EPI_ISL_2097874, EPI_ISL_2097895, EPI_ISL_2097898, EPI_ISL_2097911, EPI_ISL_2097973, EPI_ISL_2098075, EPI_ISL_2098086, EPI_ISL_2098097, EPI_ISL_2098108, EPI_ISL_2098165, EPI_ISL_2098205, EPI_ISL_2098243, EPI_ISL_2098255, EPI_ISL_2132610, EPI_ISL_2144059, EPI_ISL_2144084, EPI_ISL_2144100, EPI_ISL_2144116, EPI_ISL_2144121, EPI_ISL_2144126, EPI_ISL_2144140, EPI_ISL_2144194, EPI_ISL_2144205, EPI_ISL_2144211, EPI_ISL_2144233, EPI_ISL_2144276, EPI_ISL_2144299, EPI_ISL_2144346, EPI_ISL_2144372, EPI_ISL_2144376, EPI_ISL_2144391, EPI_ISL_2144393, EPI_ISL_2144396, EPI_ISL_2144401, EPI_ISL_2144404, EPI_ISL_2144417, EPI_ISL_2144451, EPI_ISL_2144540, EPI_ISL_2144545, EPI_ISL_2144568, EPI_ISL_2144598, EPI_ISL_2144605, EPI_ISL_2144635, EPI_ISL_2144708, EPI_ISL_2144724, EPI_ISL_2144742, EPI_ISL_2144759, EPI_ISL_2144774, EPI_ISL_2144934, EPI_ISL_2144959, EPI_ISL_2144982, EPI_ISL_2144998, EPI_ISL_2145002, EPI_ISL_2145018, EPI_ISL_2145029, EPI_ISL_2145031, EPI_ISL_2145061, EPI_ISL_2145144, EPI_ISL_2145185, EPI_ISL_2145201, EPI_ISL_2145236, EPI_ISL_2145320, EPI_ISL_2145352, EPI_ISL_2145372, EPI_ISL_2145384, EPI_ISL_2145556, EPI_ISL_2145761, EPI_ISL_2159478, EPI_ISL_2159479, EPI_ISL_2159485, EPI_ISL_2159490, EPI_ISL_2159492, EPI_ISL_2159497, EPI_ISL_2159508, EPI_ISL_2159518, EPI_ISL_2159535, EPI_ISL_2159538, EPI_ISL_2159557, EPI_ISL_2159571, EPI_ISL_2159590, EPI_ISL_2159635, EPI_ISL_2159736, EPI_ISL_2159778, EPI_ISL_2159786, EPI_ISL_2159824, EPI_ISL_2159831, EPI_ISL_2159865, EPI_ISL_2159889, EPI_ISL_2159906, EPI_ISL_2159940, EPI_ISL_2159943, EPI_ISL_2159950, EPI_ISL_2159977, EPI_ISL_2159997, EPI_ISL_2160037, EPI_ISL_2160048, EPI_ISL_2160052, EPI_ISL_2160054, EPI_ISL_2160093, EPI_ISL_2160102, EPI_ISL_2160106, EPI_ISL_2160147, EPI_ISL_2160163, EPI_ISL_2160181, EPI_ISL_2160182, EPI_ISL_2160206, EPI_ISL_2160212, EPI_ISL_2160218, EPI_ISL_2160235, EPI_ISL_2160241, EPI_ISL_2160255, EPI_ISL_2160271, EPI_ISL_2160279, EPI_ISL_2160312, EPI_ISL_2160351, EPI_ISL_2160367, EPI_ISL_2160388, EPI_ISL_2160394, EPI_ISL_2160408, EPI_ISL_2160418, EPI_ISL_2160434, EPI_ISL_2160493, EPI_ISL_2160524, EPI_ISL_2160526, EPI_ISL_2160527, EPI_ISL_2160537, EPI_ISL_2160545, EPI_ISL_2160617, EPI_ISL_2160634, EPI_ISL_2160645, EPI_ISL_2160683, EPI_ISL_2160722, EPI_ISL_2187320, EPI_ISL_2187339, EPI_ISL_2187623, EPI_ISL_2187632, EPI_ISL_2187633, EPI_ISL_2187638, EPI_ISL_2247374, EPI_ISL_2247512, EPI_ISL_2247521, EPI_ISL_2247539, EPI_ISL_2247561, EPI_ISL_2247588, EPI_ISL_2247628, EPI_ISL_2247631, EPI_ISL_2247657, EPI_ISL_2247692, EPI_ISL_2247714, EPI_ISL_2247715, EPI_ISL_2247807, EPI_ISL_2247820, EPI_ISL_2247834, EPI_ISL_2247850, EPI_ISL_2247864, EPI_ISL_2247872, EPI_ISL_2247874, EPI_ISL_2247941, EPI_ISL_2248015, EPI_ISL_2248057, EPI_ISL_2248069, EPI_ISL_2248070, EPI_ISL_2248072, EPI_ISL_2248089, EPI_ISL_2248148, EPI_ISL_2248151, EPI_ISL_2248156, EPI_ISL_2248232, EPI_ISL_2248233, EPI_ISL_2248293, EPI_ISL_2248335, EPI_ISL_2248355, EPI_ISL_2248368, EPI_ISL_2248384, EPI_ISL_2248409, EPI_ISL_2248420, EPI_ISL_2248432, EPI_ISL_2248434, EPI_ISL_2268352, EPI_ISL_2269627, EPI_ISL_2269635, EPI_ISL_2269637, EPI_ISL_2269649, EPI_ISL_2269669, EPI_ISL_2269670, EPI_ISL_2269709, EPI_ISL_2269750, EPI_ISL_2269768, EPI_ISL_2269773, EPI_ISL_2269813, EPI_ISL_2269839, EPI_ISL_2269844, EPI_ISL_2269873, EPI_ISL_2269888, EPI_ISL_2269947, EPI_ISL_2269976, EPI_ISL_2269982, EPI_ISL_2269986, EPI_ISL_2269988, EPI_ISL_2270014, EPI_ISL_2270047, EPI_ISL_2270105, EPI_ISL_2270111, EPI_ISL_2270172, EPI_ISL_2270199, EPI_ISL_2270203, EPI_ISL_2270224, EPI_ISL_2270231, EPI_ISL_2270241, EPI_ISL_2270257, EPI_ISL_2270302, EPI_ISL_2270309, EPI_ISL_2270315, EPI_ISL_2270316, EPI_ISL_2270317, EPI_ISL_2270333, EPI_ISL_2270360, EPI_ISL_2270387, EPI_ISL_2270388, EPI_ISL_2270412, EPI_ISL_2270445, EPI_ISL_2270456, EPI_ISL_2270461, EPI_ISL_2270474, EPI_ISL_2270479, EPI_ISL_2270499, EPI_ISL_2270508, EPI_ISL_2270519, EPI_ISL_2270529, EPI_ISL_2270534, EPI_ISL_2270536, EPI_ISL_2270584, EPI_ISL_2270595, EPI_ISL_2270597, EPI_ISL_2270598, EPI_ISL_2270603, EPI_ISL_2270628, EPI_ISL_2270655, EPI_ISL_2270692, EPI_ISL_2270703, EPI_ISL_2270706, EPI_ISL_2270714, EPI_ISL_2270719, EPI_ISL_2270739, EPI_ISL_2270761, EPI_ISL_2270779, EPI_ISL_2270783, EPI_ISL_2270792, EPI_ISL_2270800, EPI_ISL_2270830, EPI_ISL_2270833, EPI_ISL_2280178, EPI_ISL_2280209, EPI_ISL_2280219, EPI_ISL_2319996, EPI_ISL_2320016, EPI_ISL_2320018, EPI_ISL_2320033, EPI_ISL_2320046, EPI_ISL_2320054, EPI_ISL_2320100, EPI_ISL_2320126, EPI_ISL_2320167, EPI_ISL_2320174, EPI_ISL_2320199, EPI_ISL_2320218, EPI_ISL_2320223, EPI_ISL_2320274, EPI_ISL_2320278, EPI_ISL_2320303, EPI_ISL_2320330, EPI_ISL_2320378, EPI_ISL_2320387, EPI_ISL_2320394, EPI_ISL_2320427, EPI_ISL_2320454, EPI_ISL_2320455, EPI_ISL_2320479, EPI_ISL_2320506, EPI_ISL_2320513, EPI_ISL_2320540, EPI_ISL_2320541, EPI_ISL_2320548, EPI_ISL_2320583, EPI_ISL_2320584, EPI_ISL_2320595, EPI_ISL_2320600, EPI_ISL_2320687, EPI_ISL_2320691, EPI_ISL_2320694, EPI_ISL_2320702, EPI_ISL_2320709, EPI_ISL_2320722, EPI_ISL_2320731, EPI_ISL_2320743, EPI_ISL_2320752, EPI_ISL_2320772, EPI_ISL_2320784, EPI_ISL_2320792, EPI_ISL_2320799, EPI_ISL_2320851, EPI_ISL_2320857, EPI_ISL_2320862, EPI_ISL_2320877, EPI_ISL_2320882, EPI_ISL_2320905, EPI_ISL_2320929, EPI_ISL_2320939, EPI_ISL_2320958, EPI_ISL_2320974, EPI_ISL_2320993, EPI_ISL_2321015, EPI_ISL_2321016, EPI_ISL_2321033, EPI_ISL_2321062, EPI_ISL_2321082, EPI_ISL_2321110, EPI_ISL_2321114, EPI_ISL_2321128, EPI_ISL_2367609, EPI_ISL_2367611, EPI_ISL_2367614, EPI_ISL_2367617, EPI_ISL_2367638, EPI_ISL_2440073, EPI_ISL_2440075, EPI_ISL_2440091, EPI_ISL_2440106, EPI_ISL_2440132, EPI_ISL_2440167, EPI_ISL_2440176, EPI_ISL_2440187, EPI_ISL_2440195, EPI_ISL_2440203, EPI_ISL_2440227, EPI_ISL_2440236, EPI_ISL_2440242, EPI_ISL_2440245, EPI_ISL_2440265, EPI_ISL_2440284, EPI_ISL_2440309, EPI_ISL_2440344, EPI_ISL_2440350, EPI_ISL_2440352, EPI_ISL_2440358, EPI_ISL_2440367, EPI_ISL_2440382, EPI_ISL_2440384, EPI_ISL_2440395, EPI_ISL_2440399, EPI_ISL_2440406, EPI_ISL_2440412, EPI_ISL_2440429, EPI_ISL_2440461, EPI_ISL_2440466, EPI_ISL_2440467, EPI_ISL_2440483, EPI_ISL_2440516, EPI_ISL_2440523, EPI_ISL_2440526, EPI_ISL_2451495, EPI_ISL_2451500, EPI_ISL_2598855, EPI_ISL_2598856, EPI_ISL_2598860, EPI_ISL_2598880, EPI_ISL_2598886, EPI_ISL_2598907, EPI_ISL_2598912, EPI_ISL_2598914, EPI_ISL_2598936, EPI_ISL_2598960, EPI_ISL_2598964, EPI_ISL_2598968, EPI_ISL_2598975, EPI_ISL_2598977, EPI_ISL_2598988, EPI_ISL_2598997, EPI_ISL_2599002, EPI_ISL_2599019, EPI_ISL_2599023, EPI_ISL_2599035, EPI_ISL_2599039, EPI_ISL_2599057, EPI_ISL_2599060, EPI_ISL_2599077, EPI_ISL_2599098, EPI_ISL_2599100, EPI_ISL_2599120, EPI_ISL_2599123, EPI_ISL_2599239, EPI_ISL_2599240, EPI_ISL_2599260, EPI_ISL_2599280, EPI_ISL_2599289, EPI_ISL_2599298, EPI_ISL_2599304, EPI_ISL_2599320, EPI_ISL_2599342, EPI_ISL_2599345, EPI_ISL_2599347, EPI_ISL_2599402, EPI_ISL_2599416, EPI_ISL_2599417, EPI_ISL_2599431, EPI_ISL_2599441, EPI_ISL_2599450, EPI_ISL_2599452, EPI_ISL_2599496, EPI_ISL_2599516, EPI_ISL_2599538, EPI_ISL_2599549, EPI_ISL_2599563, EPI_ISL_2599611, EPI_ISL_2599623, EPI_ISL_2649916, EPI_ISL_2649917, EPI_ISL_2649926, EPI_ISL_2649936 | | | |
| see above | Helix/Illumina | Centers for Disease Control and Prevention Division of Viral Diseases, Pathogen Discovery | Adrian Paskey; Alexandre Bolze; Ary Ascencio; Benjamin Rambo-Martin; Brad Sickler; Charlotte Rivera-Garcia; Christine Tran; Christopher Gulvick; Clinton R. Paden; Dakota Howard; Darlene Wagner; David Becker; Dhwani Batra; Duncan MacCannell; Efren Sandoval; Eileen de Feo; Elizabeth Cirulli; Eric Allen; Geraint Levan; James Lu; Jan Antico; Jason Caravas; Jason Nguyen; Jimmy Ramirez; Jingtao Liu; Kara Moser; Kelly Schiabor Barrett; Kim Gietzen; Magnus Isaksson; Marc Laurent; Matthew Schmerer; Matthew Tolentino; Nicole L. Washington; Peter W. Cook; Phil Febbo; Ryan Cho; Scott Sammons; Shannon Wickline; Shatavia Morrison; Sherry Wang; Simon White; Tyler Cassens; William Lee; Yvette Unoarumhi |
| EPI_ISL_2029808, EPI_ISL_2029930, EPI_ISL_2030070, EPI_ISL_2188427, EPI_ISL_2279645, EPI_ISL_2374380, EPI_ISL_2462941 | | | |
| see above | Hospital | National Reference Center for Viruses of Respiratory Infections, Institut Pasteur, Paris | Alain Delepine; Amaury Vaysse; Angela Brisebarre; Axelle Paquin; Camille Capel; Christophe Malabat; Corinne Maufrais; Emmanuelle Permal; Eric Farfour; Etienne Simon-Lorière; Frédéric Lemoine; Louise Lefrançois; Marion Barbet; Maud Vanpeene; Méline Bizard; Nabil Gastli; Pascale Martres; StéPhanie Marque Juillet; Sylvie Behillil; Sylvie Van der Werf; Vincent Enouf |
| EPI_ISL_2812290 | Hospital Cangas y Severo Ochoa | Laboratorio de Virología HUCA | Abreu F; Alvarez-Arguelles ME; Boga JA; Castelló C; Costales I; Coto E; Gómez de Oña J; Martín-Rodríguez G; Melón S; Perez-Martínez Z; Rojo S; Sandoval M |
| EPI_ISL_2812439 | Hospital Carmen y Severo Ochoa | Laboratorio de Virología HUCA | Abreu F; Alvarez-Arguelles ME; Boga JA; Castelló C; Costales I; Coto E; Gómez de Oña J; Martín-Rodríguez G; Melón S; Perez-Martínez Z; Rojo S; Sandoval M |
| EPI_ISL_2439984 | Hospital Francesc de Borja (Gandia) | SeqCOVID-SPAIN consortium/IBV(CSIC) | Inmaculada Vidal Catalá; Isabel García Nava and SeqCOVID-SPAIN consortium; Mª Rosa Navarro Cots; Nieves Orta Mira |
| EPI_ISL_1914327, EPI_ISL_2080923, EPI_ISL_2178828, EPI_ISL_2304183, EPI_ISL_2304185, EPI_ISL_2304186, EPI_ISL_3505909, EPI_ISL_3505911, EPI_ISL_3505912, EPI_ISL_3505913, EPI_ISL_3505915, EPI_ISL_3505917, EPI_ISL_3505921, EPI_ISL_3505923, EPI_ISL_3505927, EPI_ISL_3505932, EPI_ISL_3505935, EPI_ISL_3505940, EPI_ISL_3505943, EPI_ISL_3505948, EPI_ISL_3505949, EPI_ISL_3505957, EPI_ISL_3505961, EPI_ISL_3505963, EPI_ISL_3505967 | | | |
| see above | Hospital General Universitario Gregorio Marañón | Hospital General Universitario Gregorio Marañón | Cristina Rodriguez-Grande; Darío García de Viedma; Julia Suárez; Laura Pérez-Lago; Marta Herranz Martin; Patricia Muñoz; Pedro Sola Campoy; Pilar Catalán; Sergio Buenestado Serrano; Victor Manuel de la Cueva; Victor de la Cueva |
| EPI_ISL_2271588 | Hospital General Universitario de Alicante - Instituto de Investigación Sanitaria y Biomédica de Alicante | SeqCOVID-SPAIN consortium/IBV(CSIC) | Carmen Molina Pardines and SeqCOVID-SPAIN consortium; Maripaz Ventero Martín |
| EPI_ISL_2614581 | Hospital Geral do Grajau | Instituto Adolfo Lutz, Interdiciplinary Procedures Center, Strategic Laboratory | Caio Vinicius Dias Lopes; Claudia Regina Gonçalves; Claudio Tavares Sacchi; Erica Valessa Ramos Gomes; Karoline Rodrigues Campos; Leonardo Jose Tadeu de Araujo |
| EPI_ISL_3033046, EPI_ISL_3033047, EPI_ISL_3033048, EPI_ISL_3033050, EPI_ISL_3033051, EPI_ISL_3260091 | Hospital Jaime Ferre - SAMCO Rafaela | Grupo de Genómica y Bioinformática del Instituto de Investigación de la Cadena Láctea CONICET-INTA on behalf of 'Proyecto Argentino Interinstitucional de genomica de SARS-CoV-2' (PAIS Consortium) | AF; Aliprandi D; Amadio; C; Eberhardt; Irazoqui; Isaia; JF; JM; MF; Pandolfi; Quaranta; Soratti R; V |
| EPI_ISL_2365419, EPI_ISL_2662822 | Hospital San Agustín | Laboratorio de Virología HUCA | Abreu F; Alvarez-Arguelles ME; Boga JA; Castelló C; Costales I; Coto E; Gómez de Oña J; Martín-Rodríguez G; Melón S; Perez-Martínez Z; Rojo S; Sandoval M |
| EPI_ISL_3230021 | Hospital Señor del Milagro | Instituto de Patologia Vegetal (CIAP-INTA) on behalf of 'Proyecto Argentino Interinstitucional de genomica de SARS-CoV-2' (PAIS | Debat, HJ.; FD; Fernanda Aguero; Fernández; Lavaque Esteban; Marquez, N.; Pamela Cajal; Raskovsky Viviana; Veronica Lesser |

| | | | |
|---|---|---|---|
| | | Consortium) | |
| EPI_ISL_2284955, EPI_ISL_2284956, EPI_ISL_2284957, EPI_ISL_2284958, EPI_ISL_2284968, EPI_ISL_2284969, EPI_ISL_2284974, EPI_ISL_2284975, EPI_ISL_2391949, EPI_ISL_2391950 | | | |
| see above | Hospital Universitari Arnau de Vilanova | Hospital Universitari Vall d'Hebron - Vall d'Hebron Institut de Recerca | Alejandra González-Sánchez; Andrés Antón; Ariadna Rando; Carla Castillo; Cristina Andrés; Damir Garcia-Cehic; Josep Quer; Juliana Esperalba; Maria Carmen Martin; Maria Gema Codina; Maria Piñana; Tomàs Pumarola |
| EPI_ISL_2017673, EPI_ISL_2351935, EPI_ISL_2516404, EPI_ISL_2516527 | Hospital Universitari Bellvitge | Microbiology Department | Aida Gonzalez-Diaz; Carmen Ardanuy; Jordi Camara; Jordi Niubó; Laura Calatayud; M Angeles Domínguez; Miguel Fernandez-Huerta; Sara Marti |
| EPI_ISL_2652049, EPI_ISL_2652054 | Hospital Universitari Joan XXIII | Hospital Universitari Joan XXIII | Carla Martín; Clara Benavent; Cristina Gutiérrez; Ester Picó; Gemma Recio; Margarida Terrón; Natalia Bastón |
| EPI_ISL_1908893, EPI_ISL_2047850, EPI_ISL_2284977, EPI_ISL_2391939 | Hospital Universitari Joan XXIII de Tarragona | Hospital Universitari Vall d'Hebron - Vall d'Hebron Institut de Recerca | Alejandra González-Sánchez; Andrés Antón; Ariadna Rando; Carla Castillo; Cristina Andrés; Damir Garcia-Cehic; Josep Quer; Juliana Esperalba; Maria Carmen Martin; Maria Gema Codina; Maria Piñana; Tomàs Pumarola |
| EPI_ISL_2047711, EPI_ISL_2047712, EPI_ISL_2047721, EPI_ISL_2162247, EPI_ISL_2162253, EPI_ISL_2200155 | Hospital Universitari Vall d'Hebron - Vall d'Hebron Institut de Recerca | Hospital Universitari Vall d'Hebron - Vall d'Hebron Institut de Recerca | Alejandra González-Sánchez; Andrés Antón; Ariadna Rando; Carla Castillo; Cristina Andrés; Damir Garcia-Cehic; Josep Quer; Juliana Esperalba; Maria Carmen Martin; Maria Gema Codina; Maria Piñana; Tomàs Pumarola |
| EPI_ISL_2323396 | Hospital Universitari i Politècnic La Fe de València | SeqCOVID-SPAIN consortium/IBV(CSIC) | Ana Gil Brusola; Eva González Barberá; José Luis López Hontangas and SeqCOVID-SPAIN consortium; María Dolores Gómez Ruiz; Salvador Giner Almaraz |
| EPI_ISL_2465946 | Hospital Universitario Virgen de las Nieves de Granada-SAS | SeqCOVID-SPAIN consortium/IBV(CSIC) | Irene Pedrosa Corral; José M. Navarro-Marí and SeqCOVID-SPAIN consortium; Sara Sanbonmatsu Gámez |
| EPI_ISL_2238703, EPI_ISL_2238704 | Hospital de Cabueñes | Laboratorio de Virología HUCA | Abreu F; Alvarez-Arguelles ME; Boga JA; Castelló C; Costales I; Coto E; Gómez de Oña J; Martín-Rodríguez G; Melón S; Perez-Martínez Z; Rojo S; Sandoval M |
| EPI_ISL_2238720 | Hospital de San Agustin | Laboratorio de Virología HUCA | Abreu F; Alvarez-Arguelles ME; Boga JA; Castelló C; Costales I; Coto E; Gómez de Oña J; Martín-Rodríguez G; Melón S; Perez-Martínez Z; Rojo S; Sandoval M |
| EPI_ISL_2407652 | Hospital de San Agustín | Laboratorio de Virología HUCA | Abreu F; Alvarez-Arguelles ME; Boga JA; Castelló C; Costales I; Coto E; Gómez de Oña J; Martín-Rodríguez G; Melón S; Perez-Martínez Z; Rojo S; Sandoval M |
| EPI_ISL_3022946, EPI_ISL_3022992, EPI_ISL_3023015, EPI_ISL_3023037 | Hospital of the University of Pennsylvania Molecular Pathology Lab | Bushman Lab - University of Pennsylvania | Aoife M. Roche; Arupa Ganguly; Ayannah S. Fitzgerald; Brendan Kelly; Jevon Graham-Wooten; John K. Everett; Kyle Rodino; Layla A. Khatib; Mike Feldman; Ronald G. Collman and Frederic Bushan; Samantha A. Whiteside; Scott Sherrill-Mix; Shantan Reddy; Young Hwang |
| EPI_ISL_1961020 | Houston Health Dept. | Houston Health Dept. | Adolpho Lara; Pamela Brown; Ryker Penn; Yanlai Lai |
| EPI_ISL_2224488, EPI_ISL_2224693, EPI_ISL_2224718, EPI_ISL_2225125, EPI_ISL_2225228, EPI_ISL_2225308, EPI_ISL_2225349, EPI_ISL_2225382, EPI_ISL_2225387, EPI_ISL_2225419, EPI_ISL_2225425 | | | |
| see above | Houston Methodist Hospital | Houston Methodist Hospital | Ilya J. Finkelstein; James J. Davis; Jessica Cambric; Jimmy Gollihar; Kristina Reppond; Layne Pruitt; Madison N. Shyer; Marcus Nguyen; Matthew Ojeda Saavedra; Paul A. Christensen; Prasanti Yerramilli; Randall J. Olsen; Robert Olson; Ryan Gadd; S. Wesley Long; Sishir Subedi; and James M. Musser |
| EPI_ISL_2497657, EPI_ISL_2497660, EPI_ISL_2497663, EPI_ISL_2497664, EPI_ISL_2500025, EPI_ISL_2500061, EPI_ISL_2658738 | | | |
| see above | Humboldt County Public Health Laboratory | Chan-Zuckerberg Biohub | CZB Cliahub Consortium |
| EPI_ISL_2445617, EPI_ISL_2445620, EPI_ISL_2445630, EPI_ISL_2680474, EPI_ISL_2680483 | Humboldt County Public Health Laboratory | Humboldt County Public Health Laboratory | Jeremy Corrigan |
| EPI_ISL_1937747 | IA State Hygienic Laboratory | Centers for Disease Control and Prevention Division of Viral Diseases, Pathogen Discovery | Alison Laufer Halpin; Ben L. Rambo-Martin; Clinton R. Paden; Dakota Howard; Darlene Wagner; Dave Wentworth; Dhwani Batra; Jasmine Padilla; Justin Lee; Katie Dillon; Krista Queen; Kristen Knipe; Kristine Lacek; Mark Burroughs; Matthew Schmerer; Mili Sheth; Peter Cook; Sam Shepard; Sarah Nobles; Shoshona Le; Suxiang Tong; Vivien Dugan; Yvette Unoarumhi |
| EPI_ISL_2452079, EPI_ISL_2690066, EPI_ISL_2690070, EPI_ISL_2690072, EPI_ISL_2690082, EPI_ISL_2690083 | ID Bureau of Laboratories | Centers for Disease Control and Prevention Division of Viral Diseases, Pathogen Discovery | Alison Laufer Halpin; Ben L. Rambo-Martin; Clinton R. Paden; Dakota Howard; Darlene Wagner; Dave Wentworth; Dhwani Batra; Jasmine Padilla; Justin Lee; Katie Dillon; Krista Queen; Kristen Knipe; Kristine Lacek; Mark Burroughs; Matthew Schmerer; Mili Sheth; Peter Cook; Sam Shepard; Sarah Nobles; Shoshona Le; Suxiang Tong; Vivien Dugan; Yvette Unoarumhi |
| EPI_ISL_2444813, EPI_ISL_2444815, EPI_ISL_2444819, EPI_ISL_2444820, EPI_ISL_2444821, EPI_ISL_2444822, EPI_ISL_2444823, EPI_ISL_2444824, EPI_ISL_2444827, EPI_ISL_2444828, EPI_ISL_2444829, EPI_ISL_2444830 | | | |
| see above | IICS-UNA | IICS-UNA | Adriana Valenzuela; Alejandra Rojas; Chyntia Diaz; Eva Nara; Fatima Cardozo; Florencia del Puerto; Joel Ortiz; Jonas Fernandez; Laura Franco; Laura Mendoza; Leticia Rojas; Magaly Martinez; Maria Eugenia Galeano. |
| EPI_ISL_2229216, EPI_ISL_2229221, EPI_ISL_2229223, EPI_ISL_2229226, EPI_ISL_2229237, EPI_ISL_2229238, EPI_ISL_2229239, EPI_ISL_2229241, EPI_ISL_2248732, EPI_ISL_2451787, EPI_ISL_2451791, EPI_ISL_2451792, EPI_ISL_2451798, EPI_ISL_2451803, EPI_ISL_2451804, EPI_ISL_2451808, EPI_ISL_2451809, EPI_ISL_2451811, EPI_ISL_2463002 | | | |
| see above | IL Dept. of Public Health Springfield Laboratory | Centers for Disease Control and Prevention Division of Viral Diseases, Pathogen Discovery | Alison Laufer Halpin; Ben L. Rambo-Martin; Clinton R. Paden; Dakota Howard; Darlene Wagner; Dave Wentworth; Dhwani Batra; Jasmine Padilla; Justin Lee; Katie Dillon; Krista Queen; Kristen Knipe; Kristine Lacek; Mark Burroughs; Matthew Schmerer; Mili Sheth; Peter Cook; Sam Shepard; Sarah Nobles; Shoshona Le; Suxiang Tong; Vivien Dugan; Yvette Unoarumhi |
| EPI_ISL_2463004, EPI_ISL_2689991 | IN State Department of Health Laboratory Services | Centers for Disease Control and Prevention Division of Viral Diseases, Pathogen Discovery | Alison Laufer Halpin; Ben L. Rambo-Martin; Clinton R. Paden; Dakota Howard; Darlene Wagner; Dave Wentworth; Dhwani Batra; Jasmine Padilla; Justin Lee; Katie Dillon; Krista Queen; Kristen Knipe; Kristine Lacek; Mark Burroughs; Matthew Schmerer; Mili Sheth; Peter Cook; Sam Shepard; Sarah Nobles; Shoshona Le; Suxiang Tong; Vivien Dugan; Yvette Unoarumhi |
| EPI_ISL_1994879, EPI_ISL_1994906, EPI_ISL_1994908, EPI_ISL_1994909, EPI_ISL_1994911, EPI_ISL_1994915, EPI_ISL_1994920, EPI_ISL_2097176, EPI_ISL_2097192, EPI_ISL_2097206, EPI_ISL_2097213, EPI_ISL_2242547, EPI_ISL_2304496, EPI_ISL_2361369, EPI_ISL_2361371, EPI_ISL_2361378, EPI_ISL_2361384, EPI_ISL_2361389, EPI_ISL_2361413, EPI_ISL_2361434, EPI_ISL_2361449, EPI_ISL_2420155, EPI_ISL_2420158, EPI_ISL_2420163, EPI_ISL_2420170, EPI_ISL_2420208, EPI_ISL_2420213, EPI_ISL_2420216, EPI_ISL_2439149, EPI_ISL_2439150, EPI_ISL_2439166, EPI_ISL_2439178, EPI_ISL_2439181, EPI_ISL_2768054 | | | |
| see above | IN State Department of Health Laboratory Services | IN State Department of Health Laboratory Services | Brian Pope; Cassandra Campion; Jamie Yeadon; Kyle Brownlee; Lixia Liu; Mark Glazier; Melissa Hindenlang |
| EPI_ISL_2319321, EPI_ISL_2319349, EPI_ISL_2319369, EPI_ISL_2319370, EPI_ISL_2319372, EPI_ISL_2319373 | INDRE | Instituto Nacional de Medicina Genomica | Cedro-Tanda A; Garcia-Cardenas FJ; Gisela Barrera-Badillo; Gonzalez-Barrera D; Gonzalez-Woge MA; Herrera-Montalvo LA.; Hidalgo-Miranda A; Irma Lopez-Martinez; Jose Ernesto Ramirez González; Mendoza-Vargas A; Munguia-Garza P; Rangel-DeLeon D; Reyes-Grajeda JP |
| EPI_ISL_2628297, EPI_ISL_2628307 | INHRR | Laboratorio de Virología Molecular | Carmen L Loureiro; CoViVen Group; Domingo J Garzaro; Esmeralda Vizzi; Flor H Pujol; Héctor R Rangel; José Luis Zambrano; Lieska Rodríguez; Mariana Hidalgo; Pierina D´Angelo; Rossana C Jaspe; Víctor Alarcón; Yoneira Sulbaran; Zoila Moros |
| EPI_ISL_2493969, EPI_ISL_2494037 | INSIDE CRSN FREGUESIA/BRASILÂNDIA | Instituto Butantan | Antonio Jorge Martins; Claudia Renata dos Santos Barros; David Schlesinger; Debora Botequio Moretti; Dimas Tadeu Covas; Elaine Cristina Marqueze; Elaine Vieira Santos; Evandra Strazza Rodrigues; Heidge Fukumasu; Jayme Augusto de Souza-Neto; José Salvatore Leister Patané; Luiz Alcantara; Luiz Lehmann Coutinho; Maria Carolina Elias; Maurício Lacerda Nogueira; Rafael dos Santos Bezerra; Raul Machado Neto; Rejane Maria Tommasini Grotto; Ricardo Haddad; Sandra Coccuzzo Sampaio Vessoni; Simone Kashima; Svetoslav Nanev Slavov; Vincent Louis Viala |
| EPI_ISL_2493974 | INSIDE CRSN PERUS | Instituto Butantan | Antonio Jorge Martins; Claudia Renata dos Santos Barros; David Schlesinger; Debora Botequio Moretti; Dimas Tadeu Covas; Elaine Cristina Marqueze; Elaine Vieira Santos; Evandra Strazza Rodrigues; Heidge Fukumasu; Jayme Augusto de Souza-Neto; José Salvatore Leister Patané; Luiz Alcantara; Luiz Lehmann Coutinho; Maria Carolina Elias; Maurício Lacerda Nogueira; Rafael dos Santos Bezerra; Raul Machado Neto; Rejane Maria Tommasini Grotto; Ricardo Haddad; Sandra Coccuzzo Sampaio Vessoni; Simone Kashima; Svetoslav Nanev Slavov; Vincent Louis Viala |
| EPI_ISL_2493966 | INSIDE CRSN VILA MARIA/VILA GUILHERME | Instituto Butantan | Antonio Jorge Martins; Claudia Renata dos Santos Barros; David Schlesinger; Debora Botequio Moretti; Dimas Tadeu Covas; Elaine Cristina Marqueze; Elaine Vieira Santos; Evandra Strazza Rodrigues; Heidge Fukumasu; Jayme Augusto de Souza-Neto; José Salvatore Leister Patané; Luiz Alcantara; Luiz Lehmann Coutinho; Maria Carolina Elias; Maurício Lacerda Nogueira; Rafael dos Santos Bezerra; Raul Machado Neto; Rejane Maria Tommasini Grotto; Ricardo Haddad; Sandra Coccuzzo Sampaio Vessoni; Simone Kashima; Svetoslav Nanev Slavov; Vincent Louis Viala |
| EPI_ISL_2493991 | INSIDE CRSSU MOOCA/ARICANDUVA | Instituto Butantan | Antonio Jorge Martins; Claudia Renata dos Santos Barros; David Schlesinger; Debora Botequio Moretti; Dimas Tadeu Covas; Elaine Cristina Marqueze; Elaine Vieira Santos; Evandra Strazza Rodrigues; Heidge Fukumasu; Jayme Augusto de Souza-Neto; José Salvatore Leister Patané; Luiz Alcantara; Luiz Lehmann Coutinho; Maria Carolina Elias; Maurício Lacerda Nogueira; Rafael dos Santos Bezerra; Raul Machado Neto; Rejane Maria Tommasini Grotto; Ricardo Haddad; Sandra Coccuzzo Sampaio Vessoni; Simone Kashima; Svetoslav Nanev Slavov; Vincent Louis Viala |
| EPI_ISL_2494013, EPI_ISL_2494021 | INSIDE CRSSU SAPOPEMBA/VILA PRUDENTE | Instituto Butantan | Antonio Jorge Martins; Claudia Renata dos Santos Barros; David Schlesinger; Debora Botequio Moretti; Dimas Tadeu Covas; Elaine Cristina Marqueze; Elaine Vieira Santos; Evandra Strazza Rodrigues; Heidge Fukumasu; Jayme Augusto de Souza-Neto; José Salvatore Leister Patané; Luiz Alcantara; Luiz Lehmann Coutinho; Maria Carolina Elias; Maurício Lacerda Nogueira; Rafael dos Santos Bezerra; Raul Machado Neto; Rejane Maria Tommasini Grotto; Ricardo Haddad; Sandra Coccuzzo Sampaio Vessoni; Simone Kashima; Svetoslav Nanev Slavov; Vincent Louis Viala |
| EPI_ISL_2493247, EPI_ISL_2493713, EPI_ISL_2493714, EPI_ISL_2493715, EPI_ISL_2493716, EPI_ISL_2493718, EPI_ISL_2493719, EPI_ISL_2493720, EPI_ISL_2493721, EPI_ISL_2493724, EPI_ISL_2493725, EPI_ISL_2493726, EPI_ISL_2493727, EPI_ISL_2493728, EPI_ISL_2493729, EPI_ISL_2493732, EPI_ISL_2493733, EPI_ISL_2493735, EPI_ISL_2493736, EPI_ISL_2493737, EPI_ISL_2493738, EPI_ISL_2494318, EPI_ISL_2500972, EPI_ISL_2500973 | | | |
| see above | INSIDE DIAGNÓSTICOS | Instituto Butantan | Antonio Jorge Martins; Claudia Renata dos Santos Barros; David Schlesinger; Debora Botequio Moretti; Dimas Tadeu Covas; Elaine Cristina Marqueze; Elaine Vieira Santos; Evandra Strazza Rodrigues; Heidge Fukumasu; Jayme Augusto de Souza-Neto; José Salvatore Leister Patané; Luiz Alcantara; Luiz Lehmann Coutinho; Maria Carolina Elias; Maurício Lacerda Nogueira; Rafael dos Santos Bezerra; Raul Machado Neto; Rejane Maria Tommasini Grotto; Ricardo Haddad; Sandra Coccuzzo Sampaio Vessoni; Simone Kashima; Svetoslav Nanev Slavov; Vincent Louis Viala |
| EPI_ISL_2493704, EPI_ISL_2493705, EPI_ISL_2493707 | INSIDE DIAGNÓSTICOS NORTE BRASILÂNDIA/FREGUESIA | Instituto Butantan | Antonio Jorge Martins; Claudia Renata dos Santos Barros; David Schlesinger; Debora Botequio Moretti; Dimas Tadeu Covas; Elaine Cristina Marqueze; Elaine Vieira Santos; Evandra Strazza Rodrigues; Heidge Fukumasu; Jayme Augusto de Souza-Neto; José Salvatore Leister Patané; Luiz Alcantara; Luiz Lehmann Coutinho; Maria Carolina Elias; Maurício Lacerda Nogueira; Rafael dos Santos Bezerra; Raul Machado Neto; Rejane Maria Tommasini Grotto; Ricardo Haddad; Sandra Coccuzzo Sampaio Vessoni; Simone Kashima; Svetoslav Nanev Slavov; Vincent Louis Viala |
| EPI_ISL_2445050, EPI_ISL_2445081 | INSIDE DIAGNÓSTICOS NORTE BRASILÂNDIA/FREGUESIA DO Ó | Instituto Butantan | Antonio Jorge Martins; Claudia Renata dos Santos Barros; David Schlesinger; Debora Botequio Moretti; Dimas Tadeu Covas; Elaine Cristina Marqueze; Elaine Vieira Santos; Evandra Strazza Rodrigues; Heidge Fukumasu; Jayme Augusto de Souza-Neto; José Salvatore Leister Patané; Luiz Alcantara; Luiz Lehmann Coutinho; Maria Carolina Elias; Maurício Lacerda Nogueira; Rafael dos Santos Bezerra; Raul Machado Neto; Rejane Maria Tommasini Grotto; Ricardo Haddad; Sandra Coccuzzo Sampaio Vessoni; Simone Kashima; Svetoslav Nanev Slavov; Vincent Louis Viala |

| | | | |
|---|---|---|---|
| EPI_ISL_2445059, EPI_ISL_2445065, EPI_ISL_2445070, EPI_ISL_2493706 | INSIDE DIAGNÓSTICOS NORTE CACHOEIRINHA/CASA VERDE | Instituto Butantan | Antonio Jorge Martins; Claudia Renata dos Santos Barros; David Schlesinger; Debora Botequio Moretti; Dimas Tadeu Covas; Elaine Cristina Marqueze; Elaine Vieira Santos; Evandra Strazza Rodrigues; Heidge Fukumasu; Jayme Augusto de Souza-Neto; José Salvatore Leister Patané; Luiz Alcantara; Luiz Lehmann Coutinho; Maria Carolina Elias; Maurício Lacerda Nogueira; Rafael dos Santos Bezerra; Raul Machado Neto; Rejane Maria Tommasini Grotto; Ricardo Haddad; Sandra Coccuzzo Sampaio Vessoni; Simone Kashima; Svetoslav Nanev Slavov; Vincent Louis Viala |
| EPI_ISL_2445062, EPI_ISL_2445071 | INSIDE DIAGNÓSTICOS NORTE PERUS | Instituto Butantan | Antonio Jorge Martins; Claudia Renata dos Santos Barros; David Schlesinger; Debora Botequio Moretti; Dimas Tadeu Covas; Elaine Cristina Marqueze; Elaine Vieira Santos; Evandra Strazza Rodrigues; Heidge Fukumasu; Jayme Augusto de Souza-Neto; José Salvatore Leister Patané; Luiz Alcantara; Luiz Lehmann Coutinho; Maria Carolina Elias; Maurício Lacerda Nogueira; Rafael dos Santos Bezerra; Raul Machado Neto; Rejane Maria Tommasini Grotto; Ricardo Haddad; Sandra Coccuzzo Sampaio Vessoni; Simone Kashima; Svetoslav Nanev Slavov; Vincent Louis Viala |
| EPI_ISL_2445058 | INSIDE DIAGNÓSTICOS NORTE PIRITUBA | Instituto Butantan | Antonio Jorge Martins; Claudia Renata dos Santos Barros; David Schlesinger; Debora Botequio Moretti; Dimas Tadeu Covas; Elaine Cristina Marqueze; Elaine Vieira Santos; Evandra Strazza Rodrigues; Heidge Fukumasu; Jayme Augusto de Souza-Neto; José Salvatore Leister Patané; Luiz Alcantara; Luiz Lehmann Coutinho; Maria Carolina Elias; Maurício Lacerda Nogueira; Rafael dos Santos Bezerra; Raul Machado Neto; Rejane Maria Tommasini Grotto; Ricardo Haddad; Sandra Coccuzzo Sampaio Vessoni; Simone Kashima; Svetoslav Nanev Slavov; Vincent Louis Viala |
| EPI_ISL_2445047, EPI_ISL_2445060 | INSIDE DIAGNÓSTICOS NORTE VILA MARIA/VILA GUILHERME | Instituto Butantan | Antonio Jorge Martins; Claudia Renata dos Santos Barros; David Schlesinger; Debora Botequio Moretti; Dimas Tadeu Covas; Elaine Cristina Marqueze; Elaine Vieira Santos; Evandra Strazza Rodrigues; Heidge Fukumasu; Jayme Augusto de Souza-Neto; José Salvatore Leister Patané; Luiz Alcantara; Luiz Lehmann Coutinho; Maria Carolina Elias; Maurício Lacerda Nogueira; Rafael dos Santos Bezerra; Raul Machado Neto; Rejane Maria Tommasini Grotto; Ricardo Haddad; Sandra Coccuzzo Sampaio Vessoni; Simone Kashima; Svetoslav Nanev Slavov; Vincent Louis Viala |
| EPI_ISL_2445074 | INSIDE DIAGNÓSTICOS OESTE BUTANTÃ | Instituto Butantan | Antonio Jorge Martins; Claudia Renata dos Santos Barros; David Schlesinger; Debora Botequio Moretti; Dimas Tadeu Covas; Elaine Cristina Marqueze; Elaine Vieira Santos; Evandra Strazza Rodrigues; Heidge Fukumasu; Jayme Augusto de Souza-Neto; José Salvatore Leister Patané; Luiz Alcantara; Luiz Lehmann Coutinho; Maria Carolina Elias; Maurício Lacerda Nogueira; Rafael dos Santos Bezerra; Raul Machado Neto; Rejane Maria Tommasini Grotto; Ricardo Haddad; Sandra Coccuzzo Sampaio Vessoni; Simone Kashima; Svetoslav Nanev Slavov; Vincent Louis Viala |
| EPI_ISL_2445073 | INSIDE DIAGNÓSTICOS OESTE LAPA/PINHEIROS | Instituto Butantan | Antonio Jorge Martins; Claudia Renata dos Santos Barros; David Schlesinger; Debora Botequio Moretti; Dimas Tadeu Covas; Elaine Cristina Marqueze; Elaine Vieira Santos; Evandra Strazza Rodrigues; Heidge Fukumasu; Jayme Augusto de Souza-Neto; José Salvatore Leister Patané; Luiz Alcantara; Luiz Lehmann Coutinho; Maria Carolina Elias; Maurício Lacerda Nogueira; Rafael dos Santos Bezerra; Raul Machado Neto; Rejane Maria Tommasini Grotto; Ricardo Haddad; Sandra Coccuzzo Sampaio Vessoni; Simone Kashima; Svetoslav Nanev Slavov; Vincent Louis Viala |
| EPI_ISL_2445067, EPI_ISL_2445072, EPI_ISL_2445076, EPI_ISL_2445084 | INSIDE DIAGNÓSTICOS SUDESTE IPIRANGA | Instituto Butantan | Antonio Jorge Martins; Claudia Renata dos Santos Barros; David Schlesinger; Debora Botequio Moretti; Dimas Tadeu Covas; Elaine Cristina Marqueze; Elaine Vieira Santos; Evandra Strazza Rodrigues; Heidge Fukumasu; Jayme Augusto de Souza-Neto; José Salvatore Leister Patané; Luiz Alcantara; Luiz Lehmann Coutinho; Maria Carolina Elias; Maurício Lacerda Nogueira; Rafael dos Santos Bezerra; Raul Machado Neto; Rejane Maria Tommasini Grotto; Ricardo Haddad; Sandra Coccuzzo Sampaio Vessoni; Simone Kashima; Svetoslav Nanev Slavov; Vincent Louis Viala |
| EPI_ISL_2445056 | INSIDE DIAGNÓSTICOS SUDESTE MOOCA/ARICANDUVA | Instituto Butantan | Antonio Jorge Martins; Claudia Renata dos Santos Barros; David Schlesinger; Debora Botequio Moretti; Dimas Tadeu Covas; Elaine Cristina Marqueze; Elaine Vieira Santos; Evandra Strazza Rodrigues; Heidge Fukumasu; Jayme Augusto de Souza-Neto; José Salvatore Leister Patané; Luiz Alcantara; Luiz Lehmann Coutinho; Maria Carolina Elias; Maurício Lacerda Nogueira; Rafael dos Santos Bezerra; Raul Machado Neto; Rejane Maria Tommasini Grotto; Ricardo Haddad; Sandra Coccuzzo Sampaio Vessoni; Simone Kashima; Svetoslav Nanev Slavov; Vincent Louis Viala |
| EPI_ISL_2445052, EPI_ISL_2473815 | INSIDE DIAGNÓSTICOS SUDESTE VILA MARIANA/JABAQUARA | Instituto Butantan | Antonio Jorge Martins; Claudia Renata dos Santos Barros; David Schlesinger; Debora Botequio Moretti; Dimas Tadeu Covas; Elaine Cristina Marqueze; Elaine Vieira Santos; Evandra Strazza Rodrigues; Heidge Fukumasu; Jayme Augusto de Souza-Neto; José Salvatore Leister Patané; Luiz Alcantara; Luiz Lehmann Coutinho; Maria Carolina Elias; Maurício Lacerda Nogueira; Rafael dos Santos Bezerra; Raul Machado Neto; Rejane Maria Tommasini Grotto; Ricardo Haddad; Sandra Coccuzzo Sampaio Vessoni; Simone Kashima; Svetoslav Nanev Slavov; Vincent Louis Viala |
| EPI_ISL_2445054, EPI_ISL_2445057, EPI_ISL_2445080 | INSIDE DIAGNÓSTICOS SUL CAPELA DO SOCORRO | Instituto Butantan | Antonio Jorge Martins; Claudia Renata dos Santos Barros; David Schlesinger; Debora Botequio Moretti; Dimas Tadeu Covas; Elaine Cristina Marqueze; Elaine Vieira Santos; Evandra Strazza Rodrigues; Heidge Fukumasu; Jayme Augusto de Souza-Neto; José Salvatore Leister Patané; Luiz Alcantara; Luiz Lehmann Coutinho; Maria Carolina Elias; Maurício Lacerda Nogueira; Rafael dos Santos Bezerra; Raul Machado Neto; Rejane Maria Tommasini Grotto; Ricardo Haddad; Sandra Coccuzzo Sampaio Vessoni; Simone Kashima; Svetoslav Nanev Slavov; Vincent Louis Viala |
| EPI_ISL_2445051, EPI_ISL_2445075 | INSIDE DIAGNÓSTICOS SUL PARELHEIROS | Instituto Butantan | Antonio Jorge Martins; Claudia Renata dos Santos Barros; David Schlesinger; Debora Botequio Moretti; Dimas Tadeu Covas; Elaine Cristina Marqueze; Elaine Vieira Santos; Evandra Strazza Rodrigues; Heidge Fukumasu; Jayme Augusto de Souza-Neto; José Salvatore Leister Patané; Luiz Alcantara; Luiz Lehmann Coutinho; Maria Carolina Elias; Maurício Lacerda Nogueira; Rafael dos Santos Bezerra; Raul Machado Neto; Rejane Maria Tommasini Grotto; Ricardo Haddad; Sandra Coccuzzo Sampaio Vessoni; Simone Kashima; Svetoslav Nanev Slavov; Vincent Louis Viala |
| EPI_ISL_2445046 | INSIDE LABORATÓRIOS NORTE PIRITUBA | Instituto Butantan | Antonio Jorge Martins; Claudia Renata dos Santos Barros; David Schlesinger; Debora Botequio Moretti; Dimas Tadeu Covas; Elaine Cristina Marqueze; Elaine Vieira Santos; Evandra Strazza Rodrigues; Heidge Fukumasu; Jayme Augusto de Souza-Neto; José Salvatore Leister Patané; Luiz Alcantara; Luiz Lehmann Coutinho; Maria Carolina Elias; Maurício Lacerda Nogueira; Rafael dos Santos Bezerra; Raul Machado Neto; Rejane Maria Tommasini Grotto; Ricardo Haddad; Sandra Coccuzzo Sampaio Vessoni; Simone Kashima; Svetoslav Nanev Slavov; Vincent Louis Viala |
| EPI_ISL_2458114, EPI_ISL_2458115, EPI_ISL_2488728, EPI_ISL_2757630 | INSPI-CRN DE INFLUENZA Y OTROS VIRUS RESPIRATORIOS | NIC-INSPI | Alfredo Bruno; Domenica de Mora.; Jimmy Garcés; Johanna Laines; Lizbeth Patiño; Manuel Gonzalez; Maritza Olmedo; Michelle Páez |
| EPI_ISL_2444184 | IPIGUA | Instituto Butantan / FAMERP | Antonio Jorge Martins; Claudia Renata dos Santos Barros; David Schlesinger; Debora Botequio Moretti; Dimas Tadeu Covas; Elaine Cristina Marqueze; Elaine Vieira Santos; Evandra Strazza Rodrigues; Heidge Fukumasu; Jayme Augusto de Souza-Neto; José Salvatore Leister Patané; Luiz Alcantara; Luiz Lehmann Coutinho; Maria Carolina Elias; Maurício Lacerda Nogueira; Rafael dos Santos Bezerra; Raul Machado Neto; Rejane Maria Tommasini Grotto; Ricardo Haddad; Sandra Coccuzzo Sampaio Vessoni; Simone Kashima; Svetoslav Nanev Slavov; Vincent Louis Viala |
| EPI_ISL_2612491, EPI_ISL_2693359, EPI_ISL_2693365, EPI_ISL_2693367, EPI_ISL_2712986 | ISDH | Quantigen Biosciences | Paul J. Childress |
| EPI_ISL_2628306 | IVIC | Laboratorio de Virología Molecular | Carmen L Loureiro; CoViVen Group; Domingo J Garzaro; Esmeralda Vizzi; Flor H Pujol; Héctor R Rangel; José Luis Zambrano; Lieska Rodríguez; Mariana Hidalgo; Pierina D´Angelo; Rossana C Jaspe; Víctor Alarcón; Yoneira Sulbaran; Zoila Moros |
| EPI_ISL_2427311, EPI_ISL_2500373, EPI_ISL_2500407, EPI_ISL_2500420, EPI_ISL_2598272, EPI_ISL_2598284, EPI_ISL_2789975, EPI_ISL_2789977 | | | |
| see above | IVY3 Central Lab, Vanderbilt University Medical Center | Lauring Lab, University of Michigan, Department of Microbiology and Immunology | Gilbert; Valesano |
| EPI_ISL_2786785, EPI_ISL_2786786 | Ibaraki Prefectural Institute of Public Health | Ibaraki Prefectural Institute of Public Health | Keiko Goto; Kentaro Itokawa; Makoto Kuroda; Masanori Hashino; Rina Tanaka; Tsuyoshi Sekizuka |
| EPI_ISL_2598130, EPI_ISL_2834650, EPI_ISL_2834656 | Idaho Bureau of Laboratories | Center for Global Health, University of New Mexico Health Sciences Center | Christopher Ball; Darrell Dinwiddie; Daryl Domman; Kurt Schwalm; Matthew Burns; Robert Voermans; Valerie Morley |
| EPI_ISL_2295874, EPI_ISL_2295877, EPI_ISL_2295878, EPI_ISL_2295879, EPI_ISL_2295882, EPI_ISL_2295917, EPI_ISL_2801135, EPI_ISL_2801136, EPI_ISL_2801137, EPI_ISL_2801138, EPI_ISL_2801139, EPI_ISL_2801142, EPI_ISL_2960186, EPI_ISL_3670779 | | | |
| see above | Illinois Department of Public Health | Gagnon Lab, Southern Illinois University | Keith Gagnon |
| EPI_ISL_1965031, EPI_ISL_1965071, EPI_ISL_1965077, EPI_ISL_1965081, EPI_ISL_1965085, EPI_ISL_1965090, EPI_ISL_1965100, EPI_ISL_1965116, EPI_ISL_2140025, EPI_ISL_2140026, EPI_ISL_2140027, EPI_ISL_2140028, EPI_ISL_2140030, EPI_ISL_2140031, EPI_ISL_2300283, EPI_ISL_2300286, EPI_ISL_2300328, EPI_ISL_2300353, EPI_ISL_2300387, EPI_ISL_2300396, EPI_ISL_2300414, EPI_ISL_2300519, EPI_ISL_2300525, EPI_ISL_2300531, EPI_ISL_2300536, EPI_ISL_2300582, EPI_ISL_2300631, EPI_ISL_2467401, EPI_ISL_2467406, EPI_ISL_2467414, EPI_ISL_2467450, EPI_ISL_2467452, EPI_ISL_2467453, EPI_ISL_2467455, EPI_ISL_2467459, EPI_ISL_2467461, EPI_ISL_2467467, EPI_ISL_2467472, EPI_ISL_2467474, EPI_ISL_2467483, EPI_ISL_2467486, EPI_ISL_2467488, EPI_ISL_2467502, EPI_ISL_2467508, EPI_ISL_2800858, EPI_ISL_2800860, EPI_ISL_2800866 | | | |
| see above | Illinois Department of Public Health | Illinois Department of Public Health - Chicago Lab | Ira Heimler; Joel Price; Vineet K. Dhiman |
| EPI_ISL_1825800, EPI_ISL_1825807, EPI_ISL_1825837, EPI_ISL_1825842, EPI_ISL_1825844, EPI_ISL_1825852, EPI_ISL_1825865, EPI_ISL_1938505, EPI_ISL_1938507, EPI_ISL_1938517, EPI_ISL_1938532, EPI_ISL_1938551, EPI_ISL_1938557, EPI_ISL_1938567, EPI_ISL_1938571, EPI_ISL_1938575, EPI_ISL_1938577, EPI_ISL_1938578, EPI_ISL_1938581, EPI_ISL_1938582, EPI_ISL_1938584, EPI_ISL_1938585, EPI_ISL_1938591, EPI_ISL_1938594, EPI_ISL_1938600, EPI_ISL_1938605, EPI_ISL_1938606, EPI_ISL_1938607, EPI_ISL_1938611, EPI_ISL_1938613, EPI_ISL_1938621, EPI_ISL_1938631, EPI_ISL_1938634, EPI_ISL_1938635, EPI_ISL_1938636, EPI_ISL_1938639, EPI_ISL_1938640, EPI_ISL_1938641, EPI_ISL_1938650, EPI_ISL_1938654, EPI_ISL_1938656, EPI_ISL_1938662, EPI_ISL_1938664, EPI_ISL_1938665, EPI_ISL_2000780, EPI_ISL_2000787, EPI_ISL_2000795, EPI_ISL_2000796, EPI_ISL_2000799, EPI_ISL_2000800, EPI_ISL_2000803, EPI_ISL_2000827, EPI_ISL_2000828, EPI_ISL_2000830, EPI_ISL_2000831, EPI_ISL_2000841, EPI_ISL_2000847, EPI_ISL_2000851, EPI_ISL_2086492, EPI_ISL_2086496, EPI_ISL_2086499, EPI_ISL_2086500, EPI_ISL_2086507, EPI_ISL_2086517, EPI_ISL_2086543, EPI_ISL_2086560, EPI_ISL_2086561, EPI_ISL_2086562, EPI_ISL_2135039, EPI_ISL_2135040, EPI_ISL_2135041, EPI_ISL_2135044, EPI_ISL_2135047, EPI_ISL_2135048, EPI_ISL_2135051, EPI_ISL_2135052, EPI_ISL_2135058, EPI_ISL_2135064, EPI_ISL_2135069, EPI_ISL_2135089, EPI_ISL_2135103, EPI_ISL_2135106, EPI_ISL_2135110, EPI_ISL_2135113, EPI_ISL_2135114, EPI_ISL_2135119, EPI_ISL_2135122, EPI_ISL_2135123, EPI_ISL_2135131, EPI_ISL_2203492, EPI_ISL_2203498, EPI_ISL_2203505, EPI_ISL_2203506, EPI_ISL_2203519, EPI_ISL_2203521, EPI_ISL_2203524, EPI_ISL_2203525, EPI_ISL_2203531, EPI_ISL_2203533, EPI_ISL_2203537, EPI_ISL_2203542, EPI_ISL_2203553, EPI_ISL_2203555, EPI_ISL_2203558, EPI_ISL_2203559, EPI_ISL_2203562, EPI_ISL_2203564, EPI_ISL_2203565, EPI_ISL_2203567, EPI_ISL_2203573, EPI_ISL_2203577, EPI_ISL_2203580, EPI_ISL_2203594, EPI_ISL_2203596, EPI_ISL_2203619, EPI_ISL_2203626, EPI_ISL_2203627, EPI_ISL_2203630, EPI_ISL_2203632, EPI_ISL_2203635, EPI_ISL_2203639, EPI_ISL_2203653, EPI_ISL_2203663, EPI_ISL_2203671, EPI_ISL_2203689, EPI_ISL_2203692, EPI_ISL_2203696, EPI_ISL_2203700, EPI_ISL_2203726, EPI_ISL_2203736, EPI_ISL_2203744, EPI_ISL_2203755, EPI_ISL_2203763, EPI_ISL_2339343, EPI_ISL_2339346, EPI_ISL_2339352, EPI_ISL_2339354, EPI_ISL_2339355, EPI_ISL_2339357, EPI_ISL_2339362, EPI_ISL_2339367, EPI_ISL_2339374, EPI_ISL_2339378, EPI_ISL_2339379, EPI_ISL_2339387, EPI_ISL_2339389, EPI_ISL_2339392, EPI_ISL_2339393, EPI_ISL_2339394, EPI_ISL_2339396, EPI_ISL_2339397, EPI_ISL_2339399, EPI_ISL_2339400, EPI_ISL_2339401, EPI_ISL_2339405, EPI_ISL_2339410, EPI_ISL_2339412, EPI_ISL_2339415, EPI_ISL_2339416, EPI_ISL_2339419, EPI_ISL_2339426, EPI_ISL_2339432, EPI_ISL_2339433, EPI_ISL_2339434, EPI_ISL_2339435, EPI_ISL_2403013, EPI_ISL_2403015, EPI_ISL_2403016, EPI_ISL_2403017, EPI_ISL_2403021, EPI_ISL_2403026, EPI_ISL_2403027, EPI_ISL_2403031, EPI_ISL_2403034, EPI_ISL_2403036, EPI_ISL_2403037, EPI_ISL_2403043, EPI_ISL_2403044, EPI_ISL_2403058, EPI_ISL_2403072, EPI_ISL_2403073, EPI_ISL_2403088, EPI_ISL_2403090, EPI_ISL_2403091, EPI_ISL_2403092, EPI_ISL_2403097, EPI_ISL_2403098, EPI_ISL_2403101, EPI_ISL_2403103, EPI_ISL_2403104, EPI_ISL_2403106, EPI_ISL_2403107, EPI_ISL_2403127, EPI_ISL_2403131, EPI_ISL_2403132, EPI_ISL_2403138, EPI_ISL_2403155, EPI_ISL_2598209 | | | |
| see above | Illinois Department of Public Health - Springfield Lab | Illinois Department of Public Health - Springfield Lab | Bryan Sim; Gordon McCall |
| EPI_ISL_2987712 | Imágenes Diagnósticas de Mazatlán | Microbial Genomics Laboratory | ; Alejandra García-Gasca; Alejandra Hernández-Terán; Alejandro Sánchez-Flores; Alfredo Herrera-Estrella; Alicia Ocaña-Mondragón; Andreu Comas-García; Angel Gustavo Salas-Lais; Antonio Loza Román; Bernardo Martínez-Miguel; Blanca Taboada; Brenda Irasema Maldonado-Meza; Bruno Gómez-Gil; Carla Ivón Herrera-Najera; Carlos F. Arias; Celia Boukadida; Clara Esperanza Santacruz-Tinoco; Concepción Grajales-Muñiz; Consorcio Mexicano de Vigilancia Genómica (CoViGen-Mex). Authors (in alphabetical order): Julio Elias Alvarado-Yaah; Cristóbal Cháidez-Quiróz; Célida Duque Molina; Célida Martínez-Rodríguez; Daniel Fregoso-Rueda; Daniel Lira Morales; Eduardo Becerril-Vargas; Fernando Fontove-Herrera; Fidencio Mejía-Nepomuceno; Francisco Pulido; Gloria Elena Espinosa-Ayala; Gloria María Molina-Salinas; Gloria Vazquez; Hector Esteban Paz-Juárez; Hector Montoya-Fuentes; Helen Haydee Fernanda Ramírez-Plascencia; Irvin González-López; Jean Pierre González; Jesús Hernández; Joel Armando Vázquez-Pérez.; Jorge Salas-Hernández; José Antonio Enciso-Moreno; José Arturo Martínez-Orozco; José Esteban Muñoz-Medina; José de Jesús Nuñez-Contreras; Juan Bautista Chale-Dzul; Julissa Enciso-Ibarra; Luis Alberto Ochoa-Carrera; Margarita Matías-Florentino; Mario Mújica-Sánchez; Marissa Perez-Garcia; María Guadalupe Santiago-Mauricio; María Guadalupe de Jesús Mireles-Rivera; Nelly Sélem-Mojica; Pavel Isa; Ricardo Ciria Merce; Ricardo Grande; Rosa María Gutiérrez Rios; Santiago Ávila-Ríos; Selene Zárate; Susana Lopez; Verónica Mata-Haro; Víctor Eduardo García-Arias; Víctor Hugo Borja-Aburto |
| EPI_ISL_2230897 | InDRE | Instituto Nacional de Medicina Genomica | Cedro-Tanda A; Garcia-Cardenas FJ; Gisela Barrera-Badillo; Gonzalez-Barrera D; Gonzalez-Woge MA; Herrera-Montalvo LA.; Hidalgo-Miranda A; Irma Lopez-Martinez; Jose Ernesto Ramirez González; Mendoza-Vargas A; Munguia-Garza P; Rangel-DeLeon D; Reyes-Grajeda JP |
| EPI_ISL_2011564, EPI_ISL_2011567, EPI_ISL_2011623, EPI_ISL_2011633, EPI_ISL_2136745, EPI_ISL_2490282, EPI_ISL_2490285, EPI_ISL_2536388, EPI_ISL_2536390, EPI_ISL_2536392, EPI_ISL_2536418, EPI_ISL_2536424, EPI_ISL_2536428, EPI_ISL_2536437, EPI_ISL_2536438, EPI_ISL_2536459, EPI_ISL_2536466, EPI_ISL_2536469, EPI_ISL_2692422, EPI_ISL_2692433, EPI_ISL_2714927, EPI_ISL_2714937, EPI_ISL_2714941, EPI_ISL_2714943 | | | |
| see above | Incyte Diagnostics Spokane | Seattle Flu Study | Amanda Adler; Barry R. Lutz; Benjamin Pelle; Brian Hiatt; Caitlin R. Wolf; Chris D. Frazar; Deborah A. Nickerson; Elisabeth Brandstetter; Erica Ryke; Geoff Melly; Helen Y. Chu; Janet A. Englund; Jay Shendure; Jover Lee; Kairsten Fay; Kirsten Lacombe; Lea M. Starita; Mark J. Rieder; Matthew Richardson; Matthew Thompson; Melissa Truong; Michael Boeckh; Michael Famulare; Misja Ilcisin; Peter D. Han; Philip Dykema; Romesh Gautom; Scott Lindquist; Thomas R. Sibley; Trevor Bedford |
| EPI_ISL_2304418, EPI_ISL_2304419, EPI_ISL_2304448, EPI_ISL_2304449, EPI_ISL_2304451, EPI_ISL_2304452, EPI_ISL_2304454 | | | |
| see above | Indiana Animal Disease Diagnostic Laboratory | Carpi Laboratory - Purdue University | Abebe A Fola; G Kenitra Hendrix; Giovanna Carpi; Ilinca I Ciubotariu; Jack Dorman; Jobin J Kattoor; Lev Gorenstein; Nicole M Perry; Rebecca P Wilkes |
| EPI_ISL_2444235 | Indiana State Department of Health | Quantigen Biosciences | Paul J. Chidress |
| EPI_ISL_2660339, EPI_ISL_2660351, EPI_ISL_2660355, EPI_ISL_2660363 | Infectious Disease Program, Broad Institute of Harvard and MIT | Infectious Disease Program, Broad Institute of Harvard and MIT | A.E.; Adams, G.; Anahtar, M.; B.L.; B.W.; Bauer, M.; Birren; Branda, J.; Carter, A.; Cerrato, F.; Chaluvadi, S.; Chapman; Cusick, C.; D.J.; DeRuff, K.; Flowers, K.; Gallagher, G.; Gladden-Young, A.; Gnirke, A.; Harris, J.; J.E.; K.J.; LaRocque, R.; Lagerborg, K.; Lemieux; Lin; Loreth, C.; MacInnis; Neumann, A.; Normandin, E.; P.C.; Park; Pierce, V.; Reilly, S.; Rosenberg, E.; Rudy, M.; Ryan, E.; S.B.; Sabeti; Shaw, B.; Siddle; Slater, D.; Smole, S.; Tomkins-Tinch, C.; Turbett, S. |
| EPI_ISL_2087169, EPI_ISL_2087180, EPI_ISL_2087204, EPI_ISL_2087239, EPI_ISL_2087245, EPI_ISL_2087284, EPI_ISL_2087351, EPI_ISL_2087374, EPI_ISL_2087463, EPI_ISL_2087567, EPI_ISL_2087621, EPI_ISL_2087640, EPI_ISL_2087772, EPI_ISL_2087783, EPI_ISL_2087803, EPI_ISL_2087827, EPI_ISL_2087900, EPI_ISL_2087935, EPI_ISL_2088221, EPI_ISL_2088241, EPI_ISL_2088250, EPI_ISL_2088376, EPI_ISL_2088412, EPI_ISL_2088418, EPI_ISL_2088456, EPI_ISL_2088514, EPI_ISL_2088564, EPI_ISL_2088619, EPI_ISL_2088651, EPI_ISL_2088742, EPI_ISL_2088832, EPI_ISL_2088858, EPI_ISL_2088951, EPI_ISL_2089330, EPI_ISL_2089416, EPI_ISL_2132374, EPI_ISL_2186443, EPI_ISL_2186561, EPI_ISL_2186568, EPI_ISL_2186620, EPI_ISL_2186815, EPI_ISL_2186862, EPI_ISL_2187001, EPI_ISL_2187222, EPI_ISL_2187223, EPI_ISL_2282269, EPI_ISL_2282561, EPI_ISL_2282601, EPI_ISL_2282639, EPI_ISL_2282640, EPI_ISL_2282655, EPI_ISL_2282714, EPI_ISL_2282787, EPI_ISL_2282899, EPI_ISL_2323487, EPI_ISL_2323648, EPI_ISL_2323673, EPI_ISL_2323677, EPI_ISL_2323678, EPI_ISL_2323698, EPI_ISL_2323701, EPI_ISL_2323705, EPI_ISL_2323758, EPI_ISL_2323812, EPI_ISL_2323904, EPI_ISL_2323958, EPI_ISL_2324039, EPI_ISL_2324062, EPI_ISL_2324078, EPI_ISL_2324094, EPI_ISL_2324110, EPI_ISL_2375552, EPI_ISL_2375555, EPI_ISL_2375684, EPI_ISL_2652549, EPI_ISL_2652797, EPI_ISL_2652818, EPI_ISL_2652884, EPI_ISL_2652896, EPI_ISL_2652938, EPI_ISL_2652968 | | | |

| | | | |
|---|---|---|---|
| see above | Infinity Biologix | Centers for Disease Control and Prevention Division of Viral Diseases, Pathogen Discovery | Adrian Paskey; Benjamin Rambo-Martin; Chirayu Goswami; Christian Bixby; Christopher Gulvick; Clinton R. Paden; Dakota Howard; Darlene Wagner; Dhwani Batra; Duncan MacCannell; Jason Caravas; Jonathan Schultz; Kara Moser; Matthew Schmerer; Peter W. Cook; Robin Grimwood; Russ Hager; Scott Sammons; Shatavia Morrison; Yihe Wang; Yvette Unoarumhi |
| EPI_ISL_2828662, EPI_ISL_2828663, EPI_ISL_2828664, EPI_ISL_2828665, EPI_ISL_2828666, EPI_ISL_2828667 | Instituo Adolfo Lutz - Reginal de Ribeirao Preto | Instituto Adolfo Lutz, Interdiciplinary Procedures Center, Strategic Laboratory | Caio Vinicius Dias Lopes; Claudia Regina Gonçalves; Claudio Tavares Sacchi; Erica Valessa Ramos Gomes; Karoline Rodrigues Campos |
| EPI_ISL_2691151, EPI_ISL_2691152, EPI_ISL_2691153, EPI_ISL_2691154, EPI_ISL_2691157, EPI_ISL_2691158, EPI_ISL_2691160, EPI_ISL_2691161, EPI_ISL_2691162, EPI_ISL_2691163, EPI_ISL_2691164, EPI_ISL_2691165, EPI_ISL_2691166, EPI_ISL_2691167, EPI_ISL_2691168, EPI_ISL_2691169, EPI_ISL_2828654, EPI_ISL_2828655, EPI_ISL_2828656, EPI_ISL_2828657, EPI_ISL_2828658, EPI_ISL_2828660 | | | |
| see above | Instituo Adolfo Lutz Central | Instituto Adolfo Lutz, Interdiciplinary Procedures Center, Strategic Laboratory | Caio Vinicius Dias Lopes; Claudia Regina Gonçalves; Claudio Tavares Sacchi; Erica Valessa Ramos Gomes; Karoline Rodrigues Campos; Leonardo Jose Tadeu de Araujo |
| EPI_ISL_1739244, EPI_ISL_2361461 | Institute of Microbiology, Universidad San Francisco de Quito | Institute of Microbiology, Universidad San Francisco de Quito | Belén Prado-Vivar; Bernardo Gutiérrez; Ernesto Pazos; Fernada Zurita; Gabriel Trueba; Gabriela Zambrano; Juan José Guadalupe; Michelle Grunauer; Monica Becerra-Wong; Patricio Rojas-Silva; Paúl Cárdenas; Sully Márquez; Verónica Barragán |
| EPI_ISL_2691171, EPI_ISL_2691172, EPI_ISL_2691173, EPI_ISL_2691176, EPI_ISL_2691177, EPI_ISL_2691178, EPI_ISL_2691180, EPI_ISL_2691181, EPI_ISL_2691182, EPI_ISL_2691183, EPI_ISL_2691184, EPI_ISL_2691185, EPI_ISL_2691186, EPI_ISL_2691187, EPI_ISL_2691188, EPI_ISL_2691189, EPI_ISL_2691190, EPI_ISL_2691191, EPI_ISL_2691192, EPI_ISL_2691193, EPI_ISL_2691195, EPI_ISL_2691196, EPI_ISL_2691197, EPI_ISL_2691198, EPI_ISL_2691199, EPI_ISL_2691200, EPI_ISL_2691201, EPI_ISL_2691202, EPI_ISL_2691203, EPI_ISL_2828647, EPI_ISL_2828648, EPI_ISL_2828649, EPI_ISL_2828651, EPI_ISL_2828652, EPI_ISL_2828653 | | | |
| see above | Instituto Adolfo Lutz - Reginal de Sao José do Rio Preto | Instituto Adolfo Lutz, Interdiciplinary Procedures Center, Strategic Laboratory | Caio Vinicius Dias Lopes; Claudia Regina Gonçalves; Claudio Tavares Sacchi; Erica Valessa Ramos Gomes; Karoline Rodrigues Campos; Leonardo Jose Tadeu de Araujo |
| EPI_ISL_2691110, EPI_ISL_2691116, EPI_ISL_2691118, EPI_ISL_2919240, EPI_ISL_2919241, EPI_ISL_2919243, EPI_ISL_2919244, EPI_ISL_2919248, EPI_ISL_2919250, EPI_ISL_2919252, EPI_ISL_2919254 | | | |
| see above | Instituto Adolfo Lutz - Regional de Bauru | Instituto Adolfo Lutz, Interdiciplinary Procedures Center, Strategic Laboratory | Caio Vinicius Dias Lopes; Claudia Regina Gonçalves; Claudio Tavares Sacchi; Erica Valessa Ramos Gomes; Karoline Rodrigues Campos; Leonardo Jose Tadeu de Araujo |
| EPI_ISL_2828686, EPI_ISL_2828691, EPI_ISL_2828692, EPI_ISL_2828693, EPI_ISL_2828694, EPI_ISL_2828695, EPI_ISL_3316200, EPI_ISL_3316207, EPI_ISL_3316216 | | | |
| see above | Instituto Adolfo Lutz - Regional de Campinas | Instituto Adolfo Lutz, Interdiciplinary Procedures Center, Strategic Laboratory | Caio Vinicius Dias Lopes; Claudia Regina Gonçalves; Claudio Tavares Sacchi; Erica Valessa Ramos Gomes; Karoline Rodrigues Campos; Leonardo Tadeu de Araujo; Marlon Benedito Nascomento Santos |
| EPI_ISL_2614627, EPI_ISL_2614628, EPI_ISL_2691089, EPI_ISL_2691090, EPI_ISL_2691091, EPI_ISL_2691092, EPI_ISL_2691093, EPI_ISL_2691094, EPI_ISL_2691095 | | | |
| see above | Instituto Adolfo Lutz - Regional de Marilia | Instituto Adolfo Lutz, Interdiciplinary Procedures Center, Strategic Laboratory | Caio Vinicius Dias Lopes; Claudia Regina Gonçalves; Claudio Tavares Sacchi; Erica Valessa Ramos Gomes; Karoline Rodrigues Campos; Leonardo Jose Tadeu de Araujo |
| EPI_ISL_2614618, EPI_ISL_2614619, EPI_ISL_2614620, EPI_ISL_2614621, EPI_ISL_2614622, EPI_ISL_2614623, EPI_ISL_2614624, EPI_ISL_2614625, EPI_ISL_3316154, EPI_ISL_3316156, EPI_ISL_3316174, EPI_ISL_3316228, EPI_ISL_3316234, EPI_ISL_3316235 | | | |
| see above | Instituto Adolfo Lutz - Regional de Presidente Prudente | Instituto Adolfo Lutz, Interdiciplinary Procedures Center, Strategic Laboratory | Caio Vinicius Dias Lopes; Claudia Regina Gonçalves; Claudio Tavares Sacchi; Erica Valessa Ramos Gomes; Karoline Rodrigues Campos; Leonardo Jose Tadeu de Araujo; Leonardo Tadeu de Araujo; Marlon Benedito Nascomento Santos |
| EPI_ISL_2691123, EPI_ISL_2691124, EPI_ISL_2691125 | Instituto Adolfo Lutz - Regional de Ribeirao Preto | Instituto Adolfo Lutz, Interdiciplinary Procedures Center, Strategic Laboratory | Caio Vinicius Dias Lopes; Claudia Regina Gonçalves; Claudio Tavares Sacchi; Erica Valessa Ramos Gomes; Karoline Rodrigues Campos; Leonardo Jose Tadeu de Araujo |
| EPI_ISL_2828697, EPI_ISL_2828698, EPI_ISL_2828700, EPI_ISL_2828701, EPI_ISL_2828703 | Instituto Adolfo Lutz - Regional de Santos | Instituto Adolfo Lutz, Interdiciplinary Procedures Center, Strategic Laboratory | Caio Vinicius Dias Lopes; Claudia Regina Gonçalves; Claudio Tavares Sacchi; Erica Valessa Ramos Gomes; Karoline Rodrigues Campos |
| EPI_ISL_2919264, EPI_ISL_2958834, EPI_ISL_2958835, EPI_ISL_2958836, EPI_ISL_2958837, EPI_ISL_2958838, EPI_ISL_2958839 | | | |
| see above | Instituto Adolfo Lutz - Regional de Sao Jose do Rio Preto | Instituto Adolfo Lutz, Interdiciplinary Procedures Center, Strategic Laboratory | Caio Vinicius Dias Lopes; Claudia Regina Gonçalves; Claudio Tavares Sacchi; Erica Valessa Ramos Gomes; Karoline Rodrigues Campos |
| EPI_ISL_2958844, EPI_ISL_2958845, EPI_ISL_2958846 | Instituto Adolfo Lutz - Regional de Sorocaba | Instituto Adolfo Lutz, Interdiciplinary Procedures Center, Strategic Laboratory | Caio Vinicius Dias Lopes; Claudia Regina Gonçalves; Claudio Tavares Sacchi; Erica Valessa Ramos Gomes; Karoline Rodrigues Campos |
| EPI_ISL_2691126, EPI_ISL_2691127, EPI_ISL_2691128, EPI_ISL_2691129, EPI_ISL_2691130, EPI_ISL_2691131, EPI_ISL_2691132, EPI_ISL_2691134, EPI_ISL_2691135, EPI_ISL_2691136, EPI_ISL_2691139, EPI_ISL_2691140, EPI_ISL_2691141, EPI_ISL_2691142, EPI_ISL_2691143, EPI_ISL_2756427, EPI_ISL_2756428, EPI_ISL_2756429, EPI_ISL_2756430, EPI_ISL_2756431, EPI_ISL_2958848, EPI_ISL_2958849, EPI_ISL_2958850, EPI_ISL_2958851, EPI_ISL_2958852, EPI_ISL_2958853 | | | |
| see above | Instituto Adolfo Lutz Central | Instituto Adolfo Lutz, Interdiciplinary Procedures Center, Strategic Laboratory | Caio Vinicius Dias Lopes; Claudia Regina Gonçalves; Claudio Tavares Sacchi; Erica Valessa Ramos Gomes; Karoline Rodrigues Campos; Leonardo Jose Tadeu de Araujo |
| EPI_ISL_2241317, EPI_ISL_2241318, EPI_ISL_2241319, EPI_ISL_2241322, EPI_ISL_2241323, EPI_ISL_2241324, EPI_ISL_2241325, EPI_ISL_2241326, EPI_ISL_4104677, EPI_ISL_4104681, EPI_ISL_4104691, EPI_ISL_4104695, EPI_ISL_4104704, EPI_ISL_4104705, EPI_ISL_4104709, EPI_ISL_4104715, EPI_ISL_4104716, EPI_ISL_4104719, EPI_ISL_4104725, EPI_ISL_4104727, EPI_ISL_4104734 | | | |
| see above | Instituto Butantan | Instituto Butantan | Antonio Jorge Martins; Claudia Renata dos Santos Barros; David Schlesinger; Debora Botequio Moretti; Dimas Tadeu Covas; Elaine Cristina Marqueze; Elaine Vieira Santos; Evandra Strazza Rodrigues; Heidge Fukumasu; Jayme Augusto de Souza-Neto; José Salvatore Leister Patané; Luiz Alcantara; Luiz Lehmann Coutinho; Maria Carolina Elias; Maurício Lacerda Nogueira; Rafael dos Santos Bezerra; Raul Machado Neto; Rejane Maria Tommasini Grotto; Ricardo Haddad; Sandra Coccuzzo Sampaio Vessoni; Simone Kashima; Svetoslav Nanev Slavov; Vincent Louis Viala |
| EPI_ISL_2348761, EPI_ISL_2348790 | Instituto Nacional De Investigación En Salud Pública-Crn De Influenza Y Otros Virus Respiratorios | NIC-Instituto Nacional de Investigación en Salud Pública | Alfredo Bruno; Domenica de Mora.; Jimmy Garcés; Johanna Laines; Lizbeth Patiño; Manuel Gonzalez; Maritza Olmedo; Michelle Páez |
| EPI_ISL_2346467, EPI_ISL_2490588 | Instituto Nacional de Enfermedades Respiratorias (INER) | Centro de Investigación en Enfermedades Infecciosas (CIENI), Instituto Nacional de Enfermedades Respiratorias (INER) | ; Alejandra Garcia-Gasca; Alejandra Hernández-Terán; Alejandro Sanchez-Flores; Alfredo Herrera-Estrella; Alicia Ocaña-Mondragón; Andreu Comas-García; Angel Gustavo Salas-Lais; Antonio Loza Román; Bernardo Martínez-Miguel; Blanca Taboada; Brenda Irasema Maldonado-Meza; Bruno Gomez-Gil; Carla Ivón Herrera-Najera; Carlos F. Arias; Celia Boukadida; Clara Esperanza Santacruz-Tinoco; Concepción Grajales-Muñiz; Consorcio Mexicano de Vigilancia Genómica (CoViGen-Mex). Authors (in alphabetical order): Julio Elias Alvarado-Yaah; Cristóbal Cháidez-Quiróz; Célida Duque Molina; Célida Martínez-Rodríguez; Daniel Fregoso-Rueda; Daniel Lira Morales; Eduardo Becerril-Vargas; Fernando Fontove-Herrera; Fidencio Mejía-Nepomuceno; Francisco Pulido; Gloria Elena Espinosa-Ayala; Gloria María Molina-Salinas; Gloria Vazquez; Hector Esteban Paz-Juárez; Hector Montoya-Fuentes; Helen Haydee Fernanda Ramírez-Plascencia; Irvin González-López; Jean Pierre González; Joel Armando Vázquez-Pérez; Joel Armando Vázquez-Pérez.; Jorge Salas-Hernández; José Antonio Enciso-Moreno; José Arturo Martínez-Orozco; José Esteban Muñoz-Medina; José de Jesús Nuñez-Contreras; Juan Bautista Chale-Dzul; Julissa Enciso-Ibarra; Luis Alberto Ochoa-Carrera; Margarita Matías-Florentino; Mario Mújica-Sánchez; Marissa Perez-Garcia; María Guadalupe Santiago-Mauricio; María Guadalupe de Jesús Mireles-Rivera; Nelly Sélem-Mojica; Pavel Isa; Ricardo Ciria Merce; Ricardo Grande; Rosa María Gutierrez Rios; Santiago Ávila-Ríos; Selene Zárate; Susana Lopez; Victor Eduardo Garcia-Arias; Víctor Hugo Borja-Aburto |
| EPI_ISL_2646131 | Instituto Nacional de Medicina Genomica | Centro de Investigación en Enfermedades Infecciosas (CIENI), Instituto Nacional de Enfermedades Respiratorias (INER) | Avila-Rios S; Boukadida C; Cedro-Tanda A; Herrera-Montalvo LA; Hidalgo-Miranda A; Matias-Florentino M; Mendoza-Vargas A; Perez-Garcia M; Reyes-Grajeda JP; Reyes-Teran G |
| EPI_ISL_2295417, EPI_ISL_2295418, EPI_ISL_2295419, EPI_ISL_2295420, EPI_ISL_2295423, EPI_ISL_2295425, EPI_ISL_2295427, EPI_ISL_2295428, EPI_ISL_2295432, EPI_ISL_2295434, EPI_ISL_2295435, EPI_ISL_2295436, EPI_ISL_2295437, EPI_ISL_2295438, EPI_ISL_2295439, EPI_ISL_2295440, EPI_ISL_2295442, EPI_ISL_2295444, EPI_ISL_2295447, EPI_ISL_2295449, EPI_ISL_2295450, EPI_ISL_2295451, EPI_ISL_2378643, EPI_ISL_2378646, EPI_ISL_2378649, EPI_ISL_2378650, EPI_ISL_2378653, EPI_ISL_2378656, EPI_ISL_2378659, EPI_ISL_2378661, EPI_ISL_2378664, EPI_ISL_2378665, EPI_ISL_2378666, EPI_ISL_2378667, EPI_ISL_2378668, EPI_ISL_2378669, EPI_ISL_2378672, EPI_ISL_2378673, EPI_ISL_2378674, EPI_ISL_2378676, EPI_ISL_2378678, EPI_ISL_2378679, EPI_ISL_2378680, EPI_ISL_2378681, EPI_ISL_2378684, EPI_ISL_2378686, EPI_ISL_2378688, EPI_ISL_2378689, EPI_ISL_2378690, EPI_ISL_2378691, EPI_ISL_2378692, EPI_ISL_2378693, EPI_ISL_2378694, EPI_ISL_3118820, EPI_ISL_3118828, EPI_ISL_3118833, EPI_ISL_3118834, EPI_ISL_3118835, EPI_ISL_3118840, EPI_ISL_3118842, EPI_ISL_3118846 | | | |
| see above | Instituto de Biotecnologia - UNESP-Botucatu-SP | Instituto de Biotecnologia - UNESP-Botucatu-SP | Cecília Artico Banho; Cíntia Bittar; Fábio Sossai Possebon; Guilherme Campos; Helena Lage Ferreira; Jorge A. Petroli Marchesi; João Pessoa Araújo Jr.; Leila Sabrina Ullmann; Lívia Sacchetto; Maisa C. Pereira Parra; Marília Moraes; Maurício L. Nogueira; Paula Rahal; Paulo Inacio da Costa |
| EPI_ISL_3104701, EPI_ISL_3104702, EPI_ISL_3104706, EPI_ISL_3104707, EPI_ISL_3104708, EPI_ISL_3236435 | Instituto de Medicina Tropical & Salud Global Universidad Iberoamericana | Grubaugh Lab - Yale School of Public Health | Alejandaro Vallejo Degaudenzi; Anderson Brito; Andrea Firpo; Annie Watkins; Chaney Kalinich; Chantal Vogels; David Rivera Aponte; Elisa Contreras; Ernesto Vazquez; Esperanza Mendoza; Fabiola Cruz-Lopez; Fabiola Fontanet; Isabel Ott; Jessica Rothman; Joseph Fauver; Karolane Gonzalez; Kendall Billig; Mallery Breban; Mary Petrone; Natalie Machargo; Nathan Grubaugh; Patricia Serrano; Robert Paulino-Ramirez; Tania Mitwalli; Tara Alpert; Tobias Koch; Vianca Aponte; Victor Virgilio Calderon |
| EPI_ISL_2536526, EPI_ISL_2536536, EPI_ISL_2715007 | Interpath Laboratory | Seattle Flu Study | Amanda Adler; Barry R. Lutz; Benjamin Pelle; Brian Hiatt; Caitlin R. Wolf; Chris D. Frazar; Deborah A. Nickerson; Elisabeth Brandstetter; Erica Ryke; Geoff Melly; Helen Y. Chu; Janet A. Englund; Jay Shendure; Jover Lee; Kairsten Fay; Kirsten Lacombe; Lea M. Starita; Mark J. Rieder; Matthew Richardson; Matthew Thompson; Melissa Truong; Michael Boeckh; Michael Famulare; Misja Ilcisin; Peter D. Han; Philip Dykema; Romesh Gautom; Scott Lindquist; Thomas R. Sibley; Trevor Bedford |
| EPI_ISL_2691146 | Intituto Adolfo Lutz - Regional de Sorocaba | Instituto Adolfo Lutz, Interdiciplinary Procedures Center, Strategic Laboratory | Caio Vinicius Dias Lopes; Claudia Regina Gonçalves; Claudio Tavares Sacchi; Erica Valessa Ramos Gomes; Karoline Rodrigues Campos; Leonardo Jose Tadeu de Araujo |
| EPI_ISL_2636150 | Israel Central Virology laboratory | Israel National Consortium for SARS-CoV-2 sequencing | Dana Bar-Ilan; Efrat Dahan Bucris; Efrat Glick-Saar; Ella Mendelson; Gideon Rechavi; Michal Mandelboim; Miranda Geva; Neta Zuckerman; Omri Nayshool; Oran Erster; Orna Mor |
| EPI_ISL_2194355, EPI_ISL_2194401, EPI_ISL_2194436, | Istituto Zooprofilattico Sperimentale del Mezzogiorno | TIGEM | Antonio Grimaldi Patrizia Annunziata Francesco Panariello Biancamaria Pierri Claudia Tiberio Teresa Giuliano Valentina Bouche Chiara Colantuono Maria Concetta Cuomo Denise Di Concilio Lucio Di Filippo Anna Manfredi Marcello Salvi Antonio Limone Luigi Atripaldi Pellegrino Cerino Andrea Ballabio Davide Cacchiarelli |

| | | | |
|---|---|---|---|
| EPI_ISL_2282039, EPI_ISL_2282090, EPI_ISL_2282125 | | | |
| EPI_ISL_2615918, EPI_ISL_2615919 | J.W. Ruby Memorial Hospital | WVU and Marshall University Combined Genomics Core Facilities | James Denvir; Peter Perrotta; Peter Stoilov; Ryan Percifield; Wesley Kimble |
| EPI_ISL_2444186 | JOSE BONIFACIO | Instituto Butantan / FAMERP | Antonio Jorge Martins; Claudia Renata dos Santos Barros; David Schlesinger; Debora Botequio Moretti; Dimas Tadeu Covas; Elaine Cristina Marqueze; Elaine Vieira Santos; Evandra Strazza Rodrigues; Heidge Fukumasu; Jayme Augusto de Souza-Neto; José Salvatore Leister Patané; Luiz Alcantara; Luiz Lehmann Coutinho; Maria Carolina Elias; Maurício Lacerda Nogueira; Rafael dos Santos Bezerra; Raul Machado Neto; Rejane Maria Tommasini Grotto; Ricardo Haddad; Sandra Coccuzzo Sampaio Vessoni; Simone Kashima; Svetoslav Nanev Slavov; Vincent Louis Viala |
| EPI_ISL_2322377 | Jessa | Jessa | Berden et al. on behalf of the Jessa_cmdLab |
| EPI_ISL_2248906, EPI_ISL_2248907, EPI_ISL_2248910 | Joaquim Chaves Saude | Instituto Nacional de Saude (INSA) | Borges et al |
| EPI_ISL_2476376, EPI_ISL_2476380, EPI_ISL_2476384, EPI_ISL_2476386, EPI_ISL_2692054, EPI_ISL_2692057, EPI_ISL_2692060, EPI_ISL_2692063, EPI_ISL_2692064, EPI_ISL_2692066, EPI_ISL_2692068, EPI_ISL_2692070, EPI_ISL_2692072, EPI_ISL_2692074, EPI_ISL_2692076 | | | |
| see above | John H Stroger Jr Hospital of Cook County | RIPHL at Rush University Medical Center | Ah-Young Sung; Felix Araujo Perez; Kevin Kunstman; Laura Furtado; Marieta Hyde; Max Kolton; Sharon Welbel; Stefan Green |
| EPI_ISL_2331499, EPI_ISL_2331546 | Johns Hopkins Hospital Department of Pathology | Johns Hopkins Hospital Department of Pathology | Adannaya Amadi; C. Paul Morris; Chun Huai Luo; Heba H. Mostafa; Matthew Schwartz; Nicholas Gallagher |
| EPI_ISL_2384201, EPI_ISL_2645292, EPI_ISL_2645297 | KS Health and Environmental Laboratories | Centers for Disease Control and Prevention Division of Viral Diseases, Pathogen Discovery | Alison Laufer Halpin; Ben L. Rambo-Martin; Clinton R. Paden; Dakota Howard; Darlene Wagner; Dave Wentworth; Dhwani Batra; Jasmine Padilla; Justin Lee; Katie Dillon; Krista Queen; Kristen Knipe; Kristine Lacek; Mark Burroughs; Matthew Schmerer; Mili Sheth; Peter Cook; Sam Shepard; Sarah Nobles; Shoshona Le; Suxiang Tong; Vivien Dugan; Yvette Unoarumhi |
| EPI_ISL_2425155, EPI_ISL_2726430, EPI_ISL_2726467, EPI_ISL_2726473, EPI_ISL_2832723, EPI_ISL_2833071, EPI_ISL_2833419, EPI_ISL_2833426 | | | |
| see above | KU Leuven, Rega Institute, Clinical and Epidemiological Virology | KU Leuven, Rega Institute, Clinical and Epidemiological Virology | Bert Vanmechelen; Joan Marti-Carerras; Piet Maes; Tony Wawina-Bokalanga |
| EPI_ISL_2383979 | KY State Public Health Lab | Centers for Disease Control and Prevention Division of Viral Diseases, Pathogen Discovery | Alison Laufer Halpin; Ben L. Rambo-Martin; Clinton R. Paden; Dakota Howard; Darlene Wagner; Dave Wentworth; Dhwani Batra; Jasmine Padilla; Justin Lee; Katie Dillon; Krista Queen; Kristen Knipe; Kristine Lacek; Mark Burroughs; Matthew Schmerer; Mili Sheth; Peter Cook; Sam Shepard; Sarah Nobles; Shoshona Le; Suxiang Tong; Vivien Dugan; Yvette Unoarumhi |
| EPI_ISL_3561093, EPI_ISL_3561231, EPI_ISL_3561270, EPI_ISL_3561295, EPI_ISL_3561318, EPI_ISL_3561509, EPI_ISL_3561554 | | | |
| see above | Kaiser Permanente NW Reginal Lab | OHSU MM Lab | Yun Wu |
| EPI_ISL_3505123, EPI_ISL_3505124, EPI_ISL_3505125, EPI_ISL_3505126, EPI_ISL_3505127, EPI_ISL_3505128, EPI_ISL_3505129, EPI_ISL_3505130 | | | |
| see above | Kaiser Permanente NW Reginal Lab | OHSU-MM Lab | Yun Wu |
| EPI_ISL_2934066, EPI_ISL_2934068 | Kaiser Regional | Santa Clara County Public Health Laboratory | Santa Clara County Public Health Department |
| EPI_ISL_1805462, EPI_ISL_1964227, EPI_ISL_2037393, EPI_ISL_2092832, EPI_ISL_2092834, EPI_ISL_2101374, EPI_ISL_2156041, EPI_ISL_2230472, EPI_ISL_2271719, EPI_ISL_2289071, EPI_ISL_2289072, EPI_ISL_2289073, EPI_ISL_2289074 | | | |
| see above | Kansas Health and Environmental Lab | Kansas Health and Environmental Lab | Ben Olsen; Jonathan Barnell; Katherine Wiggins; Mike Grose; and Phil Adam |
| EPI_ISL_1964231 | Kansas Health and Environmental Lab | Wichita State University - Molecular Diagnostics Lab | Ben Olsen; Jonathan Barnell; Mike Grose; and Phil Adam |
| EPI_ISL_2898414 | Karolinska University Hospital Solna | Karolinska University Hospital | Annelie Bjerkner; Isak Sylvin; Jan Albert; Karolina Ininbergs; Lina Guerra Blomqvist; Lynda Eneh; Martin Ekman; Martina Wahlund; Robert Dyrdak; Sandra Broddesson; Tanja Normark; Tobias Allander; Valtteri Wirta; Zhibing Yun |
| EPI_ISL_3737713, EPI_ISL_3737714, EPI_ISL_3737715, EPI_ISL_3742793, EPI_ISL_3758007, EPI_ISL_4080577, EPI_ISL_4080578, EPI_ISL_4080579, EPI_ISL_4080580, EPI_ISL_4080726 | | | |
| see above | Kentucky State Public Health Lab | Kentucky State Public Health Lab | Anjayya Tummala; Chitralipi Kattunga; Joshua Tobias; Karim George; Matthew Johnson; Rachel Zinner; Rhonda Lucas; Vaneet Arora; William Grooms |
| EPI_ISL_3354895 | LA Office of Public Health Laboratories | Centers for Disease Control and Prevention Division of Viral Diseases, Pathogen Discovery | Alex Burgin; Ben Rambo-Martin; Clinton Paden; Dakota Howard; Dave Wentworth; Dhwani Batra; Jasmine Padilla; Justin Lee; Krista Queen; Kristen Knipe; Kristine Lacek; Mark Burroughs; Matthew Schmerer; Meghan Bentz; Mili Sheth; Peter Cook; Sam Shepard; Sarah Nobles; Suxiang Tong; Vivien Dugan; Yvette Unoarumhi |
| EPI_ISL_2482673 | LABORATOIRE CREAVALLEE | CNR Virus des Infections Respiratoires - France SUD | Antonin Bal; Bruno Lina; Gregory Destras; Gwendolyne Burfin; Hadrien Regue; Laurence Josset; Martine Valette; Quentin Semanas |
| EPI_ISL_2445495, EPI_ISL_2445497, EPI_ISL_2473783, EPI_ISL_2493646, EPI_ISL_2493647, EPI_ISL_2493648 | LABORATORIO DE FRANCA | Instituto Butantan | Antonio Jorge Martins; Claudia Renata dos Santos Barros; David Schlesinger; Debora Botequio Moretti; Dimas Tadeu Covas; Elaine Cristina Marqueze; Elaine Vieira Santos; Evandra Strazza Rodrigues; Heidge Fukumasu; Jayme Augusto de Souza-Neto; José Salvatore Leister Patané; Luiz Alcantara; Luiz Lehmann Coutinho; Maria Carolina Elias; Maurício Lacerda Nogueira; Rafael dos Santos Bezerra; Raul Machado Neto; Rejane Maria Tommasini Grotto; Ricardo Haddad; Sandra Coccuzzo Sampaio Vessoni; Simone Kashima; Svetoslav Nanev Slavov; Vincent Louis Viala |
| EPI_ISL_2445561, EPI_ISL_2445562 | LABORATORIO DR PAULO EMILIO DALESSANDRO PINDAMONHANGABA | Instituto Butantan | Antonio Jorge Martins; Claudia Renata dos Santos Barros; David Schlesinger; Debora Botequio Moretti; Dimas Tadeu Covas; Elaine Cristina Marqueze; Elaine Vieira Santos; Evandra Strazza Rodrigues; Heidge Fukumasu; Jayme Augusto de Souza-Neto; José Salvatore Leister Patané; Luiz Alcantara; Luiz Lehmann Coutinho; Maria Carolina Elias; Maurício Lacerda Nogueira; Rafael dos Santos Bezerra; Raul Machado Neto; Rejane Maria Tommasini Grotto; Ricardo Haddad; Sandra Coccuzzo Sampaio Vessoni; Simone Kashima; Svetoslav Nanev Slavov; Vincent Louis Viala |
| EPI_ISL_2651229 | LABORATORIO HEMATOLOGICO | Universidad Nacional de Colombia - Laboratorio Genómico One Health | Andres F. Cardona-Rios; Carlos Franco-Muñoz; Carolina Muñoz-Arango; Celeny Ortiz; Daniel O. Maldonado-Perez; Diego A. Álvarez-Díaz; Hector Alejandro Ruiz-Moreno; Idabely Betancur Ortiz; Jorge E. Osorio; Juan P. Hernandez-Ortiz; Karl A Ciuoderis; Katherine Laiton-Donato; Laura Silvana Perez; Lina M. Hurtado; Marcela Mercado-Reyes; Maria Angélica Maya; Maria Stella López; Rita Almanza Payares; Sandra Ines Cano; Simón Villegas Velásquez |
| EPI_ISL_2345141, EPI_ISL_2345142, EPI_ISL_2345149, EPI_ISL_2345150, EPI_ISL_2345152 | LABORATORIO LOCAL DE ITAPECERICA DA SERRA | Instituto Butantan / ESALQ-Piracicaba | Antonio Jorge Martins; Claudia Renata dos Santos Barros; David Schlesinger; Debora Botequio Moretti; Dimas Tadeu Covas; Elaine Cristina Marqueze; Elaine Vieira Santos; Evandra Strazza Rodrigues; Heidge Fukumasu; Jayme Augusto de Souza-Neto; José Salvatore Leister Patané; Luiz Alcantara; Luiz Lehmann Coutinho; Maria Carolina Elias; Maurício Lacerda Nogueira; Rafael dos Santos Bezerra; Raul Machado Neto; Rejane Maria Tommasini Grotto; Ricardo Haddad; Sandra Coccuzzo Sampaio Vessoni; Simone Kashima; Svetoslav Nanev Slavov; Vincent Louis Viala |
| EPI_ISL_2493684, EPI_ISL_2493703 | LABORATORIO MUNICIPAL DE ANALISES CLINICAS DE ITANHAEM | Instituto Butantan | Antonio Jorge Martins; Claudia Renata dos Santos Barros; David Schlesinger; Debora Botequio Moretti; Dimas Tadeu Covas; Elaine Cristina Marqueze; Elaine Vieira Santos; Evandra Strazza Rodrigues; Heidge Fukumasu; Jayme Augusto de Souza-Neto; José Salvatore Leister Patané; Luiz Alcantara; Luiz Lehmann Coutinho; Maria Carolina Elias; Maurício Lacerda Nogueira; Rafael dos Santos Bezerra; Raul Machado Neto; Rejane Maria Tommasini Grotto; Ricardo Haddad; Sandra Coccuzzo Sampaio Vessoni; Simone Kashima; Svetoslav Nanev Slavov; Vincent Louis Viala |
| EPI_ISL_2493368, EPI_ISL_2493379, EPI_ISL_2493380, EPI_ISL_2493381, EPI_ISL_2493397, EPI_ISL_2493398, EPI_ISL_2493403, EPI_ISL_2493440, EPI_ISL_2494151 | | | |
| see above | LABORATORIO MUNICIPAL DE ANALISES CLINICAS DE RIO CLARO | Instituto Butantan | Antonio Jorge Martins; Claudia Renata dos Santos Barros; David Schlesinger; Debora Botequio Moretti; Dimas Tadeu Covas; Elaine Cristina Marqueze; Elaine Vieira Santos; Evandra Strazza Rodrigues; Heidge Fukumasu; Jayme Augusto de Souza-Neto; José Salvatore Leister Patané; Luiz Alcantara; Luiz Lehmann Coutinho; Maria Carolina Elias; Maurício Lacerda Nogueira; Rafael dos Santos Bezerra; Raul Machado Neto; Rejane Maria Tommasini Grotto; Ricardo Haddad; Sandra Coccuzzo Sampaio Vessoni; Simone Kashima; Svetoslav Nanev Slavov; Vincent Louis Viala |
| EPI_ISL_2493449, EPI_ISL_2493451, EPI_ISL_2493453, EPI_ISL_2493454, EPI_ISL_2493649, EPI_ISL_2493653, EPI_ISL_2494154, EPI_ISL_2494157 | | | |
| see above | LABORATORIO MUNICIPAL DE PIRACICABA | Instituto Butantan | Antonio Jorge Martins; Claudia Renata dos Santos Barros; David Schlesinger; Debora Botequio Moretti; Dimas Tadeu Covas; Elaine Cristina Marqueze; Elaine Vieira Santos; Evandra Strazza Rodrigues; Heidge Fukumasu; Jayme Augusto de Souza-Neto; José Salvatore Leister Patané; Luiz Alcantara; Luiz Lehmann Coutinho; Maria Carolina Elias; Maurício Lacerda Nogueira; Rafael dos Santos Bezerra; Raul Machado Neto; Rejane Maria Tommasini Grotto; Ricardo Haddad; Sandra Coccuzzo Sampaio Vessoni; Simone Kashima; Svetoslav Nanev Slavov; Vincent Louis Viala |
| EPI_ISL_2344838, EPI_ISL_2344864, EPI_ISL_2344917, EPI_ISL_2344971, EPI_ISL_2344972, EPI_ISL_2344979, EPI_ISL_2344983, EPI_ISL_2344988, EPI_ISL_2345034, EPI_ISL_2345118, EPI_ISL_2345169, EPI_ISL_2345170, EPI_ISL_2345186, EPI_ISL_2345248, EPI_ISL_2345250, EPI_ISL_2345251, EPI_ISL_2345271, EPI_ISL_2345288, EPI_ISL_2345289, EPI_ISL_2345290, EPI_ISL_2345292, EPI_ISL_2345293, EPI_ISL_2345297, EPI_ISL_2345298 | | | |
| see above | LABORATORIO MUNICIPAL DE PIRACICABA | Instituto Butantan / ESALQ-Piracicaba | Antonio Jorge Martins; Claudia Renata dos Santos Barros; David Schlesinger; Debora Botequio Moretti; Dimas Tadeu Covas; Elaine Cristina Marqueze; Elaine Vieira Santos; Evandra Strazza Rodrigues; Heidge Fukumasu; Jayme Augusto de Souza-Neto; José Salvatore Leister Patané; Luiz Alcantara; Luiz Lehmann Coutinho; Maria Carolina Elias; Maurício Lacerda Nogueira; Rafael dos Santos Bezerra; Raul Machado Neto; Rejane Maria Tommasini Grotto; Ricardo Haddad; Sandra Coccuzzo Sampaio Vessoni; Simone Kashima; Svetoslav Nanev Slavov; Vincent Louis Viala |
| EPI_ISL_2210259, EPI_ISL_2345741 | LABORATORIO MUNICIPAL DE SUZANO | Instituto Butantan / Mendelics | Antonio Jorge Martins; Claudia Renata dos Santos Barros; David Schlesinger; Debora Botequio Moretti; Dimas Tadeu Covas; Elaine Cristina Marqueze; Elaine Vieira Santos; Evandra Strazza Rodrigues; Heidge Fukumasu; Jayme Augusto de Souza-Neto; José Salvatore Leister Patané; Luiz Alcantara; Luiz Lehmann Coutinho; Maria Carolina Elias; Maurício Lacerda Nogueira; Rafael dos Santos Bezerra; Raul Machado Neto; Rejane Maria Tommasini Grotto; Ricardo Haddad; Sandra Coccuzzo Sampaio Vessoni; Simone Kashima; Svetoslav Nanev Slavov; Vincent Louis Viala |
| EPI_ISL_2488814, EPI_ISL_2488815, EPI_ISL_2488816, EPI_ISL_2488817, EPI_ISL_2488818, EPI_ISL_2488819 | LACEN - Laboratório Central de Saúde Pública do Para | Evandro Chagas Institute | A.M.; Barbagelata; E.C.; E.M.A.; Ferreira; J.A.; Junior; K.C.; L.C.; L.S.; M.C.; P.S.; Pinheiro; Santos; Silva; Sousa; Sousa Junior; W.D.C.; da Silva |
| EPI_ISL_3316308, EPI_ISL_3316309, EPI_ISL_3316310, EPI_ISL_3316311, EPI_ISL_3316312, EPI_ISL_3316313, EPI_ISL_3316314, EPI_ISL_3316315, EPI_ISL_3316316, EPI_ISL_3671906 | | | |
| see above | LACEN do Distrito Federal | Instituto Adolfo Lutz, Interdiciplinary Procedures Center, Strategic Laboratory | Caio Vinicius Dias Lopes; Claudia Regina Gonçalves; Claudio Tavares Sacchi; Karoline Rodrigues Campos; Leonardo Tadeu de Araujo; Marlon Benedito Nascomento Santos |
| EPI_ISL_2691206, EPI_ISL_2691207, EPI_ISL_2691208, EPI_ISL_2691209, EPI_ISL_2691210, EPI_ISL_2691211, EPI_ISL_2691212, EPI_ISL_2691213, EPI_ISL_2691216, EPI_ISL_2691217, EPI_ISL_2691218 | | | |
| see above | LACEN do Estado de Goias | Instituto Adolfo Lutz, Interdiciplinary Procedures Center, Strategic Laboratory | Caio Vinicius Dias Lopes; Claudia Regina Gonçalves; Claudio Tavares Sacchi; Erica Valessa Ramos Gomes; Karoline Rodrigues Campos; Leonardo Jose Tadeu de Araujo |
| EPI_ISL_3316160, EPI_ISL_3316166, EPI_ISL_3316176, EPI_ISL_3316177, EPI_ISL_3316180, EPI_ISL_3316182, EPI_ISL_3316185, EPI_ISL_3316188, EPI_ISL_3316190, EPI_ISL_3316203, EPI_ISL_3316211, EPI_ISL_3316212, EPI_ISL_3316217, EPI_ISL_3316219, EPI_ISL_3316223, EPI_ISL_3316226, EPI_ISL_3316229, EPI_ISL_3316230, EPI_ISL_3316231, EPI_ISL_3316233, EPI_ISL_3316241 | | | |

| | | | |
|---|---|---|---|
| see above | LACEN do Estado de Mato Grosso | Instituto Adolfo Lutz, Interdiciplinary Procedures Center, Strategic Laboratory | Caio Vinicius Dias Lopes; Claudia Regina Gonçalves; Claudio Tavares Sacchi; Karoline Rodrigues Campos; Leonardo Tadeu de Araujo; Marlon Benedito Nascomento Santos |
| EPI_ISL_2919205, EPI_ISL_2919207, EPI_ISL_2919209, EPI_ISL_2919210, EPI_ISL_2919211, EPI_ISL_2919212, EPI_ISL_2919213, EPI_ISL_2919214, EPI_ISL_2919215, EPI_ISL_2919218, EPI_ISL_2919226 | | | |
| see above | LACEN do Estado de Tocantins | Instituto Adolfo Lutz, Interdiciplinary Procedures Center, Strategic Laboratory | Caio Vinicius Dias Lopes; Claudia Regina Gonçalves; Claudio Tavares Sacchi; Erica Valessa Ramos Gomes; Karoline Rodrigues Campos |
| EPI_ISL_3545806, EPI_ISL_3545808, EPI_ISL_3545809, EPI_ISL_3545810, EPI_ISL_3545811, EPI_ISL_3545812, EPI_ISL_3545813, EPI_ISL_3545814, EPI_ISL_3691363, EPI_ISL_3691364, EPI_ISL_3691365, EPI_ISL_3691367, EPI_ISL_3691368, EPI_ISL_3691369, EPI_ISL_3691371, EPI_ISL_3691372, EPI_ISL_3691373, EPI_ISL_3691374, EPI_ISL_3691375, EPI_ISL_3691376, EPI_ISL_3691377, EPI_ISL_3691378, EPI_ISL_3691379, EPI_ISL_3691380, EPI_ISL_3691381 | | | |
| see above | LACEN do Estado do Mato Grosso do Sul | Instituto Adolfo Lutz, Interdiciplinary Procedures Center, Strategic Laboratory | Caio Vinicius Dias Lopes; Claudia Regina Gonçalves; Claudio Tavares Sacchi; Karoline Rodrigues Campos; Leonardo Tadeu de Araujo; Marlon Benedito Nascomento Santos |
| EPI_ISL_3691685, EPI_ISL_3691686, EPI_ISL_3691687 | LACEN do Estado do Piaui | Instituto Adolfo Lutz, Interdiciplinary Procedures Center, Strategic Laboratory | Caio Vinicius Dias Lopes; Claudia Regina Gonçalves; Claudio Tavares Sacchi; Karoline Rodrigues Campos; Leonardo Tadeu de Araujo; Marlon Benedito Nascomento Santos |
| EPI_ISL_2958858, EPI_ISL_2958859, EPI_ISL_2958860, EPI_ISL_2958861, EPI_ISL_2958862, EPI_ISL_2958863, EPI_ISL_2958864, EPI_ISL_2958866, EPI_ISL_2958867, EPI_ISL_2958868, EPI_ISL_2958869, EPI_ISL_2958870, EPI_ISL_2958871, EPI_ISL_2958873, EPI_ISL_2958874, EPI_ISL_2958875, EPI_ISL_2958876, EPI_ISL_2958877, EPI_ISL_2958878, EPI_ISL_2958879 | | | |
| see above | LACEN do Mato Grosso do Sul | Instituto Adolfo Lutz, Interdiciplinary Procedures Center, Strategic Laboratory | Caio Vinicius Dias Lopes; Claudia Regina Gonçalves; Claudio Tavares Sacchi; Erica Valessa Ramos Gomes; Karoline Rodrigues Campos |
| EPI_ISL_3835228 | LACEN/PB | WallauLab on behalf of Fiocruz COVID-19 Genomic Surveillance Network | Alexandre Freitas da Silva; Cassia Docena; Constância Flávia Junqueira Ayres; Filipe Zimmer Dezordi; Gabriel Luz Wallau; Gustavo Barbosa de Lima; Lais Ceschini Machado; Lilian Caroliny Amorim Silva; Marcelo Henrique dos Santos Paiva; Matheus Filgueira Bezerra; Sinval Pinto Brandão Filho |
| EPI_ISL_2821274, EPI_ISL_2821275, EPI_ISL_3134550, EPI_ISL_3134551, EPI_ISL_3134552, EPI_ISL_3134554, EPI_ISL_3134555, EPI_ISL_3134556, EPI_ISL_3134557, EPI_ISL_3134558, EPI_ISL_3134559, EPI_ISL_3134560, EPI_ISL_3134561, EPI_ISL_3134562, EPI_ISL_3134563, EPI_ISL_3134564, EPI_ISL_3134565, EPI_ISL_3134566, EPI_ISL_3134567, EPI_ISL_3134568, EPI_ISL_3134569, EPI_ISL_3134570, EPI_ISL_3134571, EPI_ISL_3134572, EPI_ISL_3134573, EPI_ISL_3134574, EPI_ISL_3134575, EPI_ISL_3134576, EPI_ISL_3134577, EPI_ISL_3134578, EPI_ISL_3134580, EPI_ISL_3134581, EPI_ISL_3134582, EPI_ISL_3134583, EPI_ISL_3134584, EPI_ISL_3134585, EPI_ISL_3134586, EPI_ISL_3134587, EPI_ISL_3134588, EPI_ISL_3134590, EPI_ISL_3134591, EPI_ISL_3134592, EPI_ISL_3134593, EPI_ISL_3134594, EPI_ISL_3134596, EPI_ISL_3134597, EPI_ISL_3134598, EPI_ISL_3134599, EPI_ISL_3134600, EPI_ISL_3134601, EPI_ISL_3134602, EPI_ISL_3134603, EPI_ISL_3134604, EPI_ISL_3134605, EPI_ISL_3134606, EPI_ISL_3134607, EPI_ISL_3134608, EPI_ISL_3134610, EPI_ISL_3134611, EPI_ISL_3134612, EPI_ISL_3134613, EPI_ISL_3134614, EPI_ISL_3134615, EPI_ISL_3134616, EPI_ISL_3134617, EPI_ISL_3134618, EPI_ISL_3134619, EPI_ISL_3134620, EPI_ISL_3134621, EPI_ISL_3134622, EPI_ISL_3134623, EPI_ISL_3134624, EPI_ISL_3134625, EPI_ISL_3134626, EPI_ISL_3134627, EPI_ISL_3134628, EPI_ISL_3134629, EPI_ISL_3134630, EPI_ISL_3134632, EPI_ISL_3134633, EPI_ISL_3134634, EPI_ISL_3134635, EPI_ISL_3134637, EPI_ISL_3134638, EPI_ISL_3134639, EPI_ISL_3134640, EPI_ISL_3134641, EPI_ISL_3134643, EPI_ISL_3134645, EPI_ISL_3134646, EPI_ISL_3134647, EPI_ISL_3134648, EPI_ISL_3134649, EPI_ISL_3134650, EPI_ISL_3134652, EPI_ISL_3134653, EPI_ISL_3134654, EPI_ISL_3134655, EPI_ISL_3134656, EPI_ISL_3134657, EPI_ISL_3134658, EPI_ISL_3134659, EPI_ISL_3134660, EPI_ISL_3134661, EPI_ISL_3134662, EPI_ISL_3134663, EPI_ISL_3134836, EPI_ISL_3134837, EPI_ISL_3134838, EPI_ISL_3134840, EPI_ISL_3134841, EPI_ISL_3134842, EPI_ISL_3134845, EPI_ISL_3134846, EPI_ISL_3134848, EPI_ISL_3134849, EPI_ISL_3134850, EPI_ISL_3134851, EPI_ISL_3134852, EPI_ISL_3134856, EPI_ISL_3134857, EPI_ISL_3134859, EPI_ISL_3152237, EPI_ISL_3447589, EPI_ISL_3447590, EPI_ISL_3447591, EPI_ISL_3703158, EPI_ISL_3703169, EPI_ISL_3703170, EPI_ISL_3703171, EPI_ISL_3703172, EPI_ISL_3703173, EPI_ISL_3703174, EPI_ISL_3703175, EPI_ISL_3703176, EPI_ISL_3703177, EPI_ISL_3703178, EPI_ISL_3703179, EPI_ISL_3703180, EPI_ISL_3703181, EPI_ISL_3703182, EPI_ISL_3703183, EPI_ISL_3703184, EPI_ISL_3703185, EPI_ISL_3703188, EPI_ISL_3703197, EPI_ISL_3703207, EPI_ISL_3703216, EPI_ISL_3703226, EPI_ISL_3703236, EPI_ISL_3703246, EPI_ISL_3703255, EPI_ISL_3703265, EPI_ISL_3703275, EPI_ISL_3703285, EPI_ISL_3703295, EPI_ISL_3703306, EPI_ISL_3703316, EPI_ISL_3703334, EPI_ISL_3703344, EPI_ISL_3703353, EPI_ISL_3703362, EPI_ISL_3703370, EPI_ISL_3703374, EPI_ISL_3703385, EPI_ISL_3703396, EPI_ISL_3703399, EPI_ISL_3703403, EPI_ISL_3703407, EPI_ISL_3703417, EPI_ISL_3703421, EPI_ISL_3703424, EPI_ISL_3703428, EPI_ISL_3703432, EPI_ISL_3703436, EPI_ISL_3703451, EPI_ISL_3703461, EPI_ISL_3703471, EPI_ISL_3703475, EPI_ISL_3703479, EPI_ISL_3703490, EPI_ISL_3703498, EPI_ISL_3703508, EPI_ISL_3703511, EPI_ISL_3703515, EPI_ISL_3703519, EPI_ISL_3703523, EPI_ISL_3703527, EPI_ISL_3703540, EPI_ISL_3703544, EPI_ISL_3703550, EPI_ISL_3703558, EPI_ISL_3703569, EPI_ISL_3703572, EPI_ISL_3703575, EPI_ISL_3703580, EPI_ISL_3703584, EPI_ISL_3703588, EPI_ISL_3703600, EPI_ISL_3703603, EPI_ISL_3703607, EPI_ISL_3703612, EPI_ISL_3703621, EPI_ISL_3703635, EPI_ISL_3703638, EPI_ISL_3703642, EPI_ISL_3703646, EPI_ISL_3703650, EPI_ISL_3703654, EPI_ISL_3703657, EPI_ISL_3703661, EPI_ISL_3703665 | | | |
| see above | LACEN/PE | WallauLab on behalf of Fiocruz COVID-19 Genomic Surveillance Network | Alexandre Freitas da Silva; Antonio Mauro Rezende; Cassia Docena; Constância Flávia Junqueira Ayres; Cássia Docena; Duschinka Ribeiro Duarte Guedes; Elisama Helvecio; Filipe Zimmer Dezordi; Gabriel Luz Wallau; Gustavo Barbosa de Lima; Lais Ceschini Machado; Larissa Krokovsky; Laís Ceschini Machado; Lilian Caroliny Amorim Silva; Marcelo Henrique dos Santos Paiva; Matheus Filgueira Bezerra; Sinval Pinto Brandão Filho |
| EPI_ISL_3010762, EPI_ISL_3010765, EPI_ISL_3010770, EPI_ISL_3010771, EPI_ISL_3010773, EPI_ISL_3010774, EPI_ISL_3010778, EPI_ISL_3010779, EPI_ISL_3010780, EPI_ISL_3010785, EPI_ISL_3010787, EPI_ISL_3010788, EPI_ISL_3010789, EPI_ISL_3010790, EPI_ISL_3010792, EPI_ISL_3010795, EPI_ISL_3010796, EPI_ISL_3010797, EPI_ISL_3010812, EPI_ISL_3010813, EPI_ISL_3010815, EPI_ISL_3010821, EPI_ISL_3010822, EPI_ISL_3010829, EPI_ISL_3010834, EPI_ISL_3010838, EPI_ISL_3010844, EPI_ISL_3010850, EPI_ISL_3010855, EPI_ISL_3010857, EPI_ISL_3010858, EPI_ISL_3010872, EPI_ISL_3010873, EPI_ISL_3010878, EPI_ISL_3010880, EPI_ISL_3010882, EPI_ISL_3010883, EPI_ISL_3010884, EPI_ISL_3010887, EPI_ISL_3010908, EPI_ISL_3010916, EPI_ISL_3010919, EPI_ISL_3010927, EPI_ISL_3010929, EPI_ISL_3010930, EPI_ISL_3261103 | | | |
| see above | LATE - Laboratório de Técnicas Especiais - Hospital Israelita Albert Einstein | LATE - Laboratório de Técnicas Especiais - Hospital Israelita Albert Einstein | Alexandre Hideaki Takara; Ana Paula Moreira Salles; Anelisie da Silva Santos; Deyvid Amgarten; Erick Gustavo Dorlass; Fernanda de Mello Malta; João Renato Rebello Pinho; Marcio Anunciacao Menezes; Pedro Henrique Sebe Rodrigues; Raquel Riyuzo |
| EPI_ISL_2483064 | LBM CBM Le Chesnay | Eurofins Biomnis | Véronique Jacomo; Xavier Naudot |
| EPI_ISL_2373664 | LBM UNIBIO VALENTIN | CNR Virus des Infections Respiratoires - France SUD | Antonin Bal; Bruno Lina; Gregory Destras; Gwendolyne Burfin; Hadrien Regue; Laurence Josset; Martine Valette; Quentin Semanas |
| EPI_ISL_2296086, EPI_ISL_2476439 | LESP Baja California | Instituto de Diagnostico y Referencia Epidemiologicos (INDRE) | Abril Rodriguez-Maldonado; Ariadna Medina-Benitez; Claudia Wong-Arambula; Ernesto Ramirez-Gonzalez.; Gisela Barrera-Badillo; Irma Lopez-Martinez; Joaquin Quiroz-Mercado; Lucia Hernandez-Rivas; Natividad Cruz-Ortiz; Sergio Rangel-Guerrero; Tatiana Nunez-Garcia; Vanessa Rivero-Arredondo |
| EPI_ISL_2296085 | LESP Chiapas | Instituto de Diagnostico y Referencia Epidemiologicos (INDRE) | Abril Rodriguez-Maldonado; Ariadna Medina-Benitez; Claudia Wong-Arambula; Ernesto Ramirez-Gonzalez.; Gisela Barrera-Badillo; Irma Lopez-Martinez; Joaquin Quiroz-Mercado; Lucia Hernandez-Rivas; Natividad Cruz-Ortiz; Sergio Rangel-Guerrero; Tatiana Nunez-Garcia; Vanessa Rivero-Arredondo |
| EPI_ISL_2105694, EPI_ISL_2105716, EPI_ISL_2105717, EPI_ISL_2105718, EPI_ISL_2296060, EPI_ISL_2296074, EPI_ISL_2296075, EPI_ISL_2296078, EPI_ISL_2476409, EPI_ISL_2476410, EPI_ISL_2476411, EPI_ISL_2476412, EPI_ISL_2476427, EPI_ISL_2476428, EPI_ISL_2476429, EPI_ISL_2545695, EPI_ISL_2545696, EPI_ISL_2545697 | | | |
| see above | LESP Ciudad de Mexico | Instituto de Diagnostico y Referencia Epidemiologicos (INDRE) | Abril Rodriguez-Maldonado; Ariadna Medina-Benitez; Claudia Wong-Arambula; Ernesto Ramirez-Gonzalez.; Gisela Barrera-Badillo; Irma Lopez-Martinez; Joaquin Quiroz-Mercado; Lucia Hernandez-Rivas; Natividad Cruz-Ortiz; Sergio Rangel-Guerrero; Tatiana Nunez-Garcia; Vanessa Rivero-Arredondo |
| EPI_ISL_2476438 | LESP Colima | Instituto de Diagnostico y Referencia Epidemiologicos (INDRE) | Abril Rodriguez-Maldonado; Ariadna Medina-Benitez; Claudia Wong-Arambula; Ernesto Ramirez-Gonzalez.; Gisela Barrera-Badillo; Irma Lopez-Martinez; Joaquin Quiroz-Mercado; Lucia Hernandez-Rivas; Natividad Cruz-Ortiz; Sergio Rangel-Guerrero; Tatiana Nunez-Garcia; Vanessa Rivero-Arredondo |
| EPI_ISL_2296073, EPI_ISL_2545698 | LESP Estado de Mexico | Instituto de Diagnostico y Referencia Epidemiologicos (INDRE) | Abril Rodriguez-Maldonado; Ariadna Medina-Benitez; Claudia Wong-Arambula; Ernesto Ramirez-Gonzalez.; Gisela Barrera-Badillo; Irma Lopez-Martinez; Joaquin Quiroz-Mercado; Lucia Hernandez-Rivas; Natividad Cruz-Ortiz; Sergio Rangel-Guerrero; Tatiana Nunez-Garcia; Vanessa Rivero-Arredondo |
| EPI_ISL_2105686, EPI_ISL_2105687, EPI_ISL_2296093, EPI_ISL_2476415, EPI_ISL_2476418, EPI_ISL_2476419, EPI_ISL_2476420, EPI_ISL_2476421, EPI_ISL_2476422, EPI_ISL_2476424, EPI_ISL_2476426, EPI_ISL_2674656 | | | |
| see above | LESP Guanajuato | Instituto de Diagnostico y Referencia Epidemiologicos (INDRE) | Abril Rodriguez-Maldonado; Ariadna Medina-Benitez; Claudia Wong-Arambula; Ernesto Ramirez-Gonzalez.; Gisela Barrera-Badillo; Irma Lopez-Martinez; Joaquin Quiroz-Mercado; Lucia Hernandez-Rivas; Natividad Cruz-Ortiz; Sergio Rangel-Guerrero; Tatiana Nunez-Garcia; Vanessa Rivero-Arredondo |
| EPI_ISL_2105715, EPI_ISL_2296070 | LESP Jalisco | Instituto de Diagnostico y Referencia Epidemiologicos (INDRE) | Abril Rodriguez-Maldonado; Ariadna Medina-Benitez; Claudia Wong-Arambula; Ernesto Ramirez-Gonzalez.; Gisela Barrera-Badillo; Irma Lopez-Martinez; Joaquin Quiroz-Mercado; Lucia Hernandez-Rivas; Natividad Cruz-Ortiz; Sergio Rangel-Guerrero; Tatiana Nunez-Garcia; Vanessa Rivero-Arredondo |
| EPI_ISL_2296094, EPI_ISL_2476440 | LESP Morelos | Instituto de Diagnostico y Referencia Epidemiologicos (INDRE) | Abril Rodriguez-Maldonado; Ariadna Medina-Benitez; Claudia Wong-Arambula; Ernesto Ramirez-Gonzalez.; Gisela Barrera-Badillo; Irma Lopez-Martinez; Joaquin Quiroz-Mercado; Lucia Hernandez-Rivas; Natividad Cruz-Ortiz; Sergio Rangel-Guerrero; Tatiana Nunez-Garcia; Vanessa Rivero-Arredondo |
| EPI_ISL_2296068 | LESP Nuevo Leon | Instituto de Diagnostico y Referencia Epidemiologicos (INDRE) | Abril Rodriguez-Maldonado; Ariadna Medina-Benitez; Claudia Wong-Arambula; Ernesto Ramirez-Gonzalez.; Gisela Barrera-Badillo; Irma Lopez-Martinez; Joaquin Quiroz-Mercado; Lucia Hernandez-Rivas; Natividad Cruz-Ortiz; Sergio Rangel-Guerrero; Tatiana Nunez-Garcia; Vanessa Rivero-Arredondo |
| EPI_ISL_2476430 | LESP Oaxaca | Instituto de Diagnostico y Referencia Epidemiologicos (INDRE) | Abril Rodriguez-Maldonado; Ariadna Medina-Benitez; Claudia Wong-Arambula; Ernesto Ramirez-Gonzalez.; Gisela Barrera-Badillo; Irma Lopez-Martinez; Joaquin Quiroz-Mercado; Lucia Hernandez-Rivas; Natividad Cruz-Ortiz; Sergio Rangel-Guerrero; Tatiana Nunez-Garcia; Vanessa Rivero-Arredondo |
| EPI_ISL_2545682 | LESP Queretaro | Instituto de Diagnostico y Referencia Epidemiologicos (INDRE) | Abril Rodriguez-Maldonado; Ariadna Medina-Benitez; Claudia Wong-Arambula; Ernesto Ramirez-Gonzalez.; Gisela Barrera-Badillo; Irma Lopez-Martinez; Joaquin Quiroz-Mercado; Lucia Hernandez-Rivas; Natividad Cruz-Ortiz; Sergio Rangel-Guerrero; Tatiana Nunez-Garcia; Vanessa Rivero-Arredondo |
| EPI_ISL_2296087 | LESP Quintana Roo | Instituto de Diagnostico y Referencia Epidemiologicos (INDRE) | Abril Rodriguez-Maldonado; Ariadna Medina-Benitez; Claudia Wong-Arambula; Ernesto Ramirez-Gonzalez.; Gisela Barrera-Badillo; Irma Lopez-Martinez; Joaquin Quiroz-Mercado; Lucia Hernandez-Rivas; Natividad Cruz-Ortiz; Sergio Rangel-Guerrero; Tatiana Nunez-Garcia; Vanessa Rivero-Arredondo |
| EPI_ISL_2476437 | LESP Sinaloa | Instituto de Diagnostico y Referencia Epidemiologicos (INDRE) | Abril Rodriguez-Maldonado; Ariadna Medina-Benitez; Claudia Wong-Arambula; Ernesto Ramirez-Gonzalez.; Gisela Barrera-Badillo; Irma Lopez-Martinez; Joaquin Quiroz-Mercado; Lucia Hernandez-Rivas; Natividad Cruz-Ortiz; Sergio Rangel-Guerrero; Tatiana Nunez-Garcia; Vanessa Rivero-Arredondo |
| EPI_ISL_2296065, EPI_ISL_2476416, EPI_ISL_2476425, EPI_ISL_2476431, EPI_ISL_2476432, EPI_ISL_2476433, EPI_ISL_2476434, EPI_ISL_2476435, EPI_ISL_2476436, EPI_ISL_2545680, EPI_ISL_2545681, EPI_ISL_2545690, EPI_ISL_2545691 | | | |
| see above | LESP Tamaulipas | Instituto de Diagnostico y Referencia Epidemiologicos (INDRE) | Abril Rodriguez-Maldonado; Ariadna Medina-Benitez; Claudia Wong-Arambula; Ernesto Ramirez-Gonzalez.; Gisela Barrera-Badillo; Irma Lopez-Martinez; Joaquin Quiroz-Mercado; Lucia Hernandez-Rivas; Natividad Cruz-Ortiz; Sergio Rangel-Guerrero; Tatiana Nunez-Garcia; Vanessa Rivero-Arredondo |
| EPI_ISL_2476414, EPI_ISL_2545684, EPI_ISL_2545685, EPI_ISL_2545686, EPI_ISL_2545687, EPI_ISL_2545688 | LESP Yucatan | Instituto de Diagnostico y Referencia Epidemiologicos (INDRE) | Abril Rodriguez-Maldonado; Ariadna Medina-Benitez; Claudia Wong-Arambula; Ernesto Ramirez-Gonzalez.; Gisela Barrera-Badillo; Irma Lopez-Martinez; Joaquin Quiroz-Mercado; Lucia Hernandez-Rivas; Natividad Cruz-Ortiz; Sergio Rangel-Guerrero; Tatiana Nunez-Garcia; Vanessa Rivero-Arredondo |

| | | | |
|---|---|---|---|
| EPI_ISL_2476417 | LESP Zacatecas | Instituto de Diagnostico y Referencia Epidemiologicos (INDRE) | Abril Rodriguez-Maldonado; Ariadna Medina-Benitez; Claudia Wong-Arambula; Ernesto Ramirez-Gonzalez.; Gisela Barrera-Badillo; Irma Lopez-Martinez; Joaquin Quiroz-Mercado; Lucia Hernandez-Rivas; Natividad Cruz-Ortiz; Sergio Rangel-Guerrero; Tatiana Nunez-Garcia; Vanessa Rivero-Arredondo |
| EPI_ISL_2029739 | Labo Analyses Med | National Reference Center for Viruses of Respiratory Infections, Institut Pasteur, Paris | Angela Brisebarre; Arditi Cocco; Camille Capel; Christophe Malabat; Corinne Maufrais; Etienne Simon-Lorière; Frédéric Lemoine; Louise Lefrançois; Marion Barbet; Maud Vanpeene; Méline Bizard; Pierre Lechat; Sylvie Behillil; Sylvie Van der Werf; Vincent Enouf |
| EPI_ISL_2678400 | Labor Becker & Kollegen (Standort München) | Robert Koch Institute | |
| EPI_ISL_2384910, EPI_ISL_2384917 | Labor Dr. Fenner und Kollegen | Heinrich Pette Institute, Leibniz Institute for Experimental Virology | Adam Grundhoff; Alexis Robitaille; Johannes Knobloch; Martin Aepfelbacher; Nicole Fischer; Thomas Günther |
| EPI_ISL_2129701, EPI_ISL_2389341 | Labor Dr. Heidrich & Kollegen MVZ GmbH Hamburg | Robert Koch Institute | |
| EPI_ISL_2129083 | Labor Monchengladbach MVZ Dr. Stein + Kollegen GbR | Robert Koch Institute | |
| EPI_ISL_2125029 | Labor Prof. Dr. G. Enders MVZ GbR | Robert Koch Institute | |
| EPI_ISL_2316147 | Labor für medizinische Mikrobiologie Bakteriologie Mykologie Virologie & Infektionsserologie - Labor Mölbis | Robert Koch Institute | |
| EPI_ISL_2122501 | Laborarztpraxis Osnabruck | Robert Koch Institute | |
| EPI_ISL_2227905, EPI_ISL_2362537, EPI_ISL_2362553, EPI_ISL_2426980 | Laboratoire Carage | Institut Pasteur de la Guyane | A. Enfissi; A. Salmier; Anne Lavergne; Antoine Enfissi; Arielle Salmier; Dominique Rousset |
| EPI_ISL_2492500, EPI_ISL_2492502, EPI_ISL_2492503, EPI_ISL_2492506, EPI_ISL_2492507, EPI_ISL_2492509, EPI_ISL_2492510, EPI_ISL_2492512, EPI_ISL_2492513, EPI_ISL_2492515, EPI_ISL_2492517, EPI_ISL_2492518, EPI_ISL_2492519, EPI_ISL_2492520, EPI_ISL_2492521, EPI_ISL_2492522, EPI_ISL_2492523, EPI_ISL_2492524, EPI_ISL_2492525, EPI_ISL_2492526, EPI_ISL_2492527, EPI_ISL_2492531, EPI_ISL_2492533, EPI_ISL_2492534, EPI_ISL_2492536, EPI_ISL_2492538, EPI_ISL_2492539, EPI_ISL_2492540, EPI_ISL_2492541, EPI_ISL_2492543, EPI_ISL_2492544, EPI_ISL_2492546 | | | |
| see above | Laboratoire National de Santé Publique – LNSP (HAITI - LNSP) | Laboratory of Respiratory Viruses and Measles, Oswaldo Cruz Institute, FIOCRUZ | Alice Sampaio Rocha; Ana Carolina Mendonca; Anna Carolina Paixao; Elisa Cavalcante Pereira; Fernando Motta; Ito Journel; Jaques Boncy; Luciana Appolinario; Marilda Siqueira on behalf of the Fiocruz COVID-19 Genomic Surveillance Network; Paola Resende; Patrick Delly; Renata Serrano Lopes; Taina Venas |
| EPI_ISL_3458621 | Laboratoire de santé publique du Québec | Laboratoire de santé publique du Québec | Guillaume Bourque; Ioannis Ragoussis; Jesse Shapiro; Mark Lathrop and Michel Roger on behalf of the CoVSeQ research group; Sandrine Moreira |
| EPI_ISL_2401051, EPI_ISL_2401055, EPI_ISL_2401076, EPI_ISL_2401143, EPI_ISL_2401159, EPI_ISL_2401180, EPI_ISL_2401181, EPI_ISL_2401183 | | | |
| see above | Laboratoire national de sante, Microbiology, Virology | Laboratoire national de sante, Microbiology, Microbial Genomics Platform | Anke Wienecke-Baldacchino; Catherine Ragimbeau; Fatu Djabi; Jessica Tapp; Lise Pignon; Raoul Salmon; Tamir Abdelrahman; Trung Nguyen Nguyen |
| EPI_ISL_2400867 | Laboratoires Reunis | Laboratoire national de sante, Microbiology, Microbial Genomics Platform | Anke Wienecke-Baldacchino; Bernard Weber; Catherine Ragimbeau; Fatu Djabi; Jessica Tapp; Lise Pignon; Raoul Salmon; Tamir Abdelrahman |
| EPI_ISL_2400675, EPI_ISL_2400677, EPI_ISL_2400715, EPI_ISL_2400731, EPI_ISL_2400775, EPI_ISL_3148148, EPI_ISL_3148149 | | | |
| see above | Laboratoires d'analyses medicales - Ketterthill | Laboratoire national de sante, Microbiology, Microbial Genomics Platform | Anke Wienecke-Baldacchino; Caroline Scheiber; Catherine Ragimbeau; Elodie Solarino; Fatu Djabi; Jessica Tapp; Lise Pignon; Raoul Salmon; Serge Vedy; Tamir Abdelrahman; Virginie Jover |
| EPI_ISL_2497876, EPI_ISL_2497877, EPI_ISL_2626433 | Laboratori de Referencia de Catalunya | Laboratori de Referencia de Catalunya | Bellosillo B.; Canal M.; Hernandez JJ.; Padilla E.; Ramirez A.; Vilas A. |
| EPI_ISL_2983179, EPI_ISL_3190350 | Laboratorio Central de Saude Publica do Estado de Goias (LACEN/GO) | Laboratory of Respiratory Viruses and Measles, Oswaldo Cruz Institute, FIOCRUZ | Agatha Cristinne Prudencio; Alice Sampaio Rocha; Ana Carolina Mendonca; Ana Flavia Mendonça; Anna Carolina Paixao; Carmen Helena Ramos; Cassiane Casanova; Elisa Cavalcante Pereira; Fernando Motta; Flavia Pereira Amorim da Silva; Ighor Leonardo Arantes Gomes; Luciana Appolinario; Luiz Augusto Pereira; Marilda Siqueira on behalf of the Fiocruz COVID-19 Genomic Surveillance Network; Paola Resende; Rafael Souza Guedes; Renata Serrano Lopes; Taina Venas; Vinicius Lemes da Silva |
| EPI_ISL_2196304, EPI_ISL_2196305, EPI_ISL_2443561, EPI_ISL_2443562, EPI_ISL_2645421, EPI_ISL_2645422, EPI_ISL_2645424, EPI_ISL_2645425, EPI_ISL_2645426, EPI_ISL_2645428, EPI_ISL_2645429, EPI_ISL_2645430, EPI_ISL_2645431, EPI_ISL_2645432, EPI_ISL_2645433, EPI_ISL_2645437, EPI_ISL_2645438, EPI_ISL_2645439, EPI_ISL_4080767 | | | |
| see above | Laboratorio Central de Saude Publica do Estado do Para (LACEN/PA) | Laboratory of Respiratory Viruses and Measles, Oswaldo Cruz Institute, FIOCRUZ | Agatha Soares; Alice Sampaio Rocha; Ana Carolina Mendonca; Anna Carolina Paixao; Elisa Cavalcante Pereira; Fernando Motta; Ighor Arantes; Luciana Appolinario; Marilda Siqueira on behalf of the Fiocruz COVID-19 Genomic Surveillance Network; Paola Resende; Renata Serrano Lopes; Taina Venas; Valnete Andrade |
| EPI_ISL_1923562, EPI_ISL_2047600, EPI_ISL_2156063, EPI_ISL_2156081, EPI_ISL_2340131 | Laboratorio Aziendale di Microbiologia e Virologia, Azienda Sanitaria dell'Alto Adige | Laboratorio Aziendale di Microbiologia e Virologia, Azienda Sanitaria dell'Alto Adige | Anne Picard; Bartolomeo Mobilio Rodriguez; Chiara Cantaloni; Claudia Volpato; Elisa Masi; Elisabetta Giacobazzi; Elisabetta Pagani; Irene Bianconi; Stefanie Wieser |
| EPI_ISL_2101403, EPI_ISL_2101404, EPI_ISL_2101405, EPI_ISL_2101406, EPI_ISL_2101409, EPI_ISL_2101411, EPI_ISL_2101412, EPI_ISL_2101413, EPI_ISL_2101414, EPI_ISL_2101415, EPI_ISL_2101416, EPI_ISL_2101417, EPI_ISL_2101418, EPI_ISL_2101419, EPI_ISL_2101420, EPI_ISL_2101421, EPI_ISL_2101422, EPI_ISL_2101423, EPI_ISL_2101425, EPI_ISL_2101426, EPI_ISL_2101434, EPI_ISL_2101435, EPI_ISL_2101436, EPI_ISL_2101437, EPI_ISL_2101438, EPI_ISL_2101439, EPI_ISL_2101440, EPI_ISL_2101441, EPI_ISL_2101442, EPI_ISL_2101445, EPI_ISL_2101446, EPI_ISL_2101447, EPI_ISL_2101448, EPI_ISL_2101449, EPI_ISL_2101451, EPI_ISL_2101452, EPI_ISL_2101453, EPI_ISL_2101464, EPI_ISL_2101466, EPI_ISL_2101467, EPI_ISL_2101478, EPI_ISL_2101479, EPI_ISL_2101480, EPI_ISL_2101481, EPI_ISL_2101482, EPI_ISL_2101489, EPI_ISL_2101490, EPI_ISL_2101491, EPI_ISL_2101492, EPI_ISL_2101493, EPI_ISL_2101500, EPI_ISL_2101501, EPI_ISL_2101502, EPI_ISL_2101503, EPI_ISL_2101504, EPI_ISL_2101507, EPI_ISL_2101514, EPI_ISL_2101515, EPI_ISL_2101516, EPI_ISL_2101517, EPI_ISL_2101518, EPI_ISL_2101519, EPI_ISL_2101528, EPI_ISL_2101529, EPI_ISL_2101530, EPI_ISL_2101531, EPI_ISL_2101532, EPI_ISL_2101533, EPI_ISL_2101534, EPI_ISL_2101535, EPI_ISL_2101536, EPI_ISL_2101537, EPI_ISL_2101538, EPI_ISL_2101539, EPI_ISL_2101540, EPI_ISL_2101544, EPI_ISL_2101545, EPI_ISL_2101548, EPI_ISL_2101563, EPI_ISL_2101564, EPI_ISL_2101565, EPI_ISL_2101573, EPI_ISL_2101574, EPI_ISL_2101575, EPI_ISL_2101576, EPI_ISL_2101577, EPI_ISL_2101578, EPI_ISL_2101579, EPI_ISL_2101580, EPI_ISL_2101581, EPI_ISL_2101582, EPI_ISL_2101584, EPI_ISL_2101585, EPI_ISL_2101586, EPI_ISL_2101590, EPI_ISL_2101592, EPI_ISL_2101594, EPI_ISL_2101596, EPI_ISL_2101597, EPI_ISL_2101598, EPI_ISL_2101599, EPI_ISL_2101600, EPI_ISL_2101601, EPI_ISL_2101603, EPI_ISL_2101604, EPI_ISL_2101605, EPI_ISL_2101606, EPI_ISL_2101608, EPI_ISL_2101609, EPI_ISL_2101610, EPI_ISL_2101611, EPI_ISL_2101613, EPI_ISL_2101615, EPI_ISL_2101616, EPI_ISL_2101617, EPI_ISL_2101618, EPI_ISL_2101619, EPI_ISL_2101620, EPI_ISL_2101622, EPI_ISL_2101623, EPI_ISL_2101624, EPI_ISL_2101625, EPI_ISL_2101626, EPI_ISL_2101627, EPI_ISL_2101628, EPI_ISL_2101629, EPI_ISL_2101630, EPI_ISL_2101631, EPI_ISL_2101632, EPI_ISL_2101633, EPI_ISL_2101634, EPI_ISL_2101635, EPI_ISL_2101636, EPI_ISL_2101637, EPI_ISL_2101672, EPI_ISL_2101674, EPI_ISL_2101675, EPI_ISL_2101690, EPI_ISL_2101691, EPI_ISL_2101697, EPI_ISL_2101698, EPI_ISL_2101699, EPI_ISL_2101700, EPI_ISL_2101701, EPI_ISL_2101702, EPI_ISL_2101703, EPI_ISL_2101704, EPI_ISL_2101705, EPI_ISL_2101706, EPI_ISL_2101707, EPI_ISL_2101708, EPI_ISL_2101709, EPI_ISL_2101710, EPI_ISL_2101711, EPI_ISL_2101712, EPI_ISL_2101713, EPI_ISL_2101716, EPI_ISL_2101717, EPI_ISL_2101718, EPI_ISL_2101719, EPI_ISL_2101725, EPI_ISL_2101730, EPI_ISL_2101732, EPI_ISL_2101733, EPI_ISL_2101736, EPI_ISL_2101737, EPI_ISL_2101738, EPI_ISL_2101745, EPI_ISL_2385432, EPI_ISL_2385433, EPI_ISL_2385434, EPI_ISL_2385435, EPI_ISL_2385436, EPI_ISL_2385437, EPI_ISL_2385438, EPI_ISL_2385440, EPI_ISL_2385441, EPI_ISL_2385442, EPI_ISL_2385443, EPI_ISL_2385444, EPI_ISL_2385445, EPI_ISL_2385446, EPI_ISL_2385447, EPI_ISL_2385448, EPI_ISL_2385449, EPI_ISL_2385450, EPI_ISL_2385451, EPI_ISL_2385452, EPI_ISL_2385453, EPI_ISL_2385454, EPI_ISL_2385455, EPI_ISL_2385456, EPI_ISL_2385457, EPI_ISL_2385458, EPI_ISL_2385459, EPI_ISL_2385460, EPI_ISL_2385461, EPI_ISL_2385462, EPI_ISL_2385463, EPI_ISL_2385464, EPI_ISL_2385465, EPI_ISL_2385466, EPI_ISL_2385467, EPI_ISL_2385468, EPI_ISL_2385469, EPI_ISL_2385470, EPI_ISL_2385471, EPI_ISL_2385472, EPI_ISL_2385474, EPI_ISL_2385475, EPI_ISL_2385476, EPI_ISL_2385477, EPI_ISL_2385478, EPI_ISL_2385480, EPI_ISL_2385482, EPI_ISL_2385483, EPI_ISL_2385484, EPI_ISL_2385485, EPI_ISL_2385486, EPI_ISL_2385487, EPI_ISL_2385488, EPI_ISL_2385489, EPI_ISL_2385490, EPI_ISL_2385491, EPI_ISL_2385492, EPI_ISL_2385493, EPI_ISL_2385496, EPI_ISL_2385498, EPI_ISL_2385499, EPI_ISL_2385500, EPI_ISL_2385501, EPI_ISL_2385504, EPI_ISL_2385505, EPI_ISL_2385506, EPI_ISL_2385507, EPI_ISL_2385508, EPI_ISL_2385510, EPI_ISL_2385511, EPI_ISL_2385512, EPI_ISL_2385513, EPI_ISL_2385514, EPI_ISL_2385515, EPI_ISL_2385516, EPI_ISL_2385517, EPI_ISL_2385518, EPI_ISL_2385519, EPI_ISL_2385521, EPI_ISL_2385522, EPI_ISL_2385523, EPI_ISL_2385527, EPI_ISL_2385528, EPI_ISL_2385530, EPI_ISL_2385531, EPI_ISL_2385532, EPI_ISL_2385533, EPI_ISL_2385534, EPI_ISL_2385535, EPI_ISL_2385536, EPI_ISL_2385537, EPI_ISL_2385538, EPI_ISL_2385539, EPI_ISL_2385541, EPI_ISL_2385542, EPI_ISL_2385543, EPI_ISL_2385544, EPI_ISL_2385545, EPI_ISL_2385546, EPI_ISL_2385547, EPI_ISL_2385548, EPI_ISL_2385549, EPI_ISL_2385550, EPI_ISL_2385551, EPI_ISL_2385552, EPI_ISL_2385553, EPI_ISL_2385554, EPI_ISL_2385558, EPI_ISL_2385559, EPI_ISL_2385561, EPI_ISL_2385562, EPI_ISL_2385563, EPI_ISL_2385567, EPI_ISL_2385568, EPI_ISL_2385570, EPI_ISL_2385571, EPI_ISL_2385572, EPI_ISL_2385573, EPI_ISL_2385574, EPI_ISL_2385575, EPI_ISL_2385576, EPI_ISL_2385577, EPI_ISL_2385578, EPI_ISL_2385579, EPI_ISL_2385580, EPI_ISL_2385581, EPI_ISL_2385583, EPI_ISL_2385584, EPI_ISL_2385586, EPI_ISL_2385587, EPI_ISL_2385589, EPI_ISL_2385590, EPI_ISL_2385592, EPI_ISL_2385593, EPI_ISL_2385594, EPI_ISL_2385595, EPI_ISL_2385596, EPI_ISL_2385597, EPI_ISL_2385598, EPI_ISL_2385599, EPI_ISL_2385600, EPI_ISL_2385601, EPI_ISL_2385602, EPI_ISL_2385603, EPI_ISL_2385604, EPI_ISL_2385605, EPI_ISL_2385606, EPI_ISL_2385607, EPI_ISL_2385608, EPI_ISL_2385609, EPI_ISL_2385610, EPI_ISL_2385611, EPI_ISL_2385612, EPI_ISL_2385613, EPI_ISL_2385616, EPI_ISL_2385620, EPI_ISL_2385624, EPI_ISL_2385625, EPI_ISL_2385627, EPI_ISL_2385629, EPI_ISL_2385630, EPI_ISL_2385631, EPI_ISL_2385632, EPI_ISL_2385633, EPI_ISL_2385634, EPI_ISL_2385636, EPI_ISL_2385637, EPI_ISL_2385639, EPI_ISL_2385641, EPI_ISL_2385642, EPI_ISL_2385644, EPI_ISL_2385647, EPI_ISL_2385648, EPI_ISL_2385649, EPI_ISL_2385651, EPI_ISL_2385652, EPI_ISL_2385654, EPI_ISL_2385655, EPI_ISL_2385656, EPI_ISL_2385657, EPI_ISL_2385658, EPI_ISL_2385659, EPI_ISL_2385660, EPI_ISL_2385661, EPI_ISL_2385662, EPI_ISL_2385663, EPI_ISL_2385664, EPI_ISL_2385665, EPI_ISL_2385666, EPI_ISL_2385667, EPI_ISL_2385668, EPI_ISL_2534862, EPI_ISL_2534863, EPI_ISL_2534865, EPI_ISL_2534866, EPI_ISL_2534868, EPI_ISL_2534869, EPI_ISL_2534870, EPI_ISL_2534871, EPI_ISL_2534872, EPI_ISL_2534873, EPI_ISL_2534874, EPI_ISL_2534875, EPI_ISL_2534876, EPI_ISL_2534877, EPI_ISL_2534878, EPI_ISL_2534879, EPI_ISL_2534880, EPI_ISL_2534883, EPI_ISL_2534884, EPI_ISL_2534885, EPI_ISL_2534886, EPI_ISL_2534887, EPI_ISL_2534888, EPI_ISL_2534889, EPI_ISL_2534890, EPI_ISL_2534891, EPI_ISL_2534892, EPI_ISL_2534893, EPI_ISL_2534894, EPI_ISL_2534896, EPI_ISL_2534897, EPI_ISL_2534898, EPI_ISL_2534899, EPI_ISL_2534900, EPI_ISL_2534901, EPI_ISL_2534902, EPI_ISL_2534903, EPI_ISL_2534904, EPI_ISL_2534905, EPI_ISL_2534906, EPI_ISL_2534907, EPI_ISL_2534908, EPI_ISL_2534909, EPI_ISL_2534910, EPI_ISL_2534911, EPI_ISL_2534912, EPI_ISL_2534913, EPI_ISL_2534914, EPI_ISL_2534915, EPI_ISL_2534916, EPI_ISL_2534917, EPI_ISL_2534918, EPI_ISL_2534919, EPI_ISL_2534920, EPI_ISL_2534921, EPI_ISL_2534922, EPI_ISL_2534925, EPI_ISL_2534927, EPI_ISL_2534928, EPI_ISL_2534929, EPI_ISL_2534930, EPI_ISL_2534931, EPI_ISL_2534933, EPI_ISL_2534934, EPI_ISL_2534935, EPI_ISL_2534936, EPI_ISL_2534938, EPI_ISL_2534939, EPI_ISL_2534941, EPI_ISL_2534942, EPI_ISL_2534944, EPI_ISL_2534946, EPI_ISL_2534947, EPI_ISL_2534948, EPI_ISL_2534949, EPI_ISL_2534950, EPI_ISL_2534951, EPI_ISL_2534952, EPI_ISL_2534955, EPI_ISL_2534956, EPI_ISL_2534957, EPI_ISL_2534959, EPI_ISL_2534960, EPI_ISL_2534961, EPI_ISL_2534962, EPI_ISL_2534963, EPI_ISL_2534964, EPI_ISL_2534965, EPI_ISL_2534966, EPI_ISL_2534967, EPI_ISL_2534968, EPI_ISL_2534971, EPI_ISL_2534973, EPI_ISL_2534974, EPI_ISL_2534975, EPI_ISL_2534976, EPI_ISL_2534977, EPI_ISL_2534978, EPI_ISL_2534979, EPI_ISL_2534980, EPI_ISL_2534981, EPI_ISL_2534982, EPI_ISL_2534983, EPI_ISL_2534984, EPI_ISL_2534985, EPI_ISL_2534986, EPI_ISL_2534987, EPI_ISL_2534988, EPI_ISL_2534989, EPI_ISL_2534990, EPI_ISL_2534991, EPI_ISL_2534992, EPI_ISL_2534993, EPI_ISL_2534994, EPI_ISL_2534996, EPI_ISL_2534997, EPI_ISL_2534999, EPI_ISL_2535000, EPI_ISL_2535001, EPI_ISL_2535002, EPI_ISL_2535003, EPI_ISL_2535004, EPI_ISL_2535005, EPI_ISL_2535006, EPI_ISL_2535007, EPI_ISL_2535009, EPI_ISL_2535010, EPI_ISL_2535011, EPI_ISL_2535012, EPI_ISL_2535014, EPI_ISL_2535015, EPI_ISL_2535017, EPI_ISL_2535021, EPI_ISL_2535022, EPI_ISL_2535023, EPI_ISL_2535024, EPI_ISL_2535025, EPI_ISL_2535027, EPI_ISL_2535028, EPI_ISL_2535029, EPI_ISL_2535030, EPI_ISL_2535031, EPI_ISL_2535032, EPI_ISL_2535033, EPI_ISL_2535034, EPI_ISL_2535035, EPI_ISL_2535036, EPI_ISL_2535039, EPI_ISL_2535040, EPI_ISL_2535041, EPI_ISL_2535042, EPI_ISL_2535043, EPI_ISL_2535044, EPI_ISL_2535045, EPI_ISL_2535047, EPI_ISL_2535048, EPI_ISL_2535049, EPI_ISL_2535050, EPI_ISL_2535051, EPI_ISL_2535052, EPI_ISL_2535053, EPI_ISL_2535054, EPI_ISL_2535055, EPI_ISL_2535056, EPI_ISL_2535058, EPI_ISL_2535059, EPI_ISL_2535060, EPI_ISL_2535061, EPI_ISL_2535062, EPI_ISL_2535063, EPI_ISL_2535064, EPI_ISL_2535065, EPI_ISL_2535066, EPI_ISL_2535067, EPI_ISL_2535068, EPI_ISL_2535069, EPI_ISL_2535070, EPI_ISL_2535072, EPI_ISL_2535073, EPI_ISL_2535074, EPI_ISL_2535075, EPI_ISL_2535076, EPI_ISL_2535077, EPI_ISL_2535080, EPI_ISL_2535081, EPI_ISL_2535082, EPI_ISL_2535083, EPI_ISL_2535084, EPI_ISL_2535085, EPI_ISL_2535089, EPI_ISL_2535090, EPI_ISL_2535091, EPI_ISL_2535092, EPI_ISL_2535093, EPI_ISL_2535094, EPI_ISL_2535095, EPI_ISL_2535096, EPI_ISL_2535097, EPI_ISL_2535098, EPI_ISL_2535099, EPI_ISL_2535100, EPI_ISL_2535102, EPI_ISL_2535106, EPI_ISL_2691353, EPI_ISL_2691354, EPI_ISL_2691355, EPI_ISL_2691356 | | | |
| see above | Laboratorio Central Noel Nutels | Bioinformatics Laboratory / LNCC | Alessandra P Lamarca; Alexandra L Gerber; Amilcar Tanuri; Ana Paula de C Guimaraes; Ana Tereza R Vasconcelos; Andrea Cony Cavalcanti; Caio Luiz Pereira Ribeiro; Cassia Alves; Cintia Policarpo; Claudia Maria Braga de Mello; Cristiane Gomes da Silva; Diana Mariani; Douglas Terra Machado; Flavio Dias da Silva; Gleidson da Silva de Oliveira; Leandro Magalhaes de Souza; Liliane Cavalcante; Luiz G P de Almeida; Marcio Henrique de Oliveira Garcia; Mario Sergio Ribeiro; Ronaldo da Silva F Jr; Silvia Carvalho |
| EPI_ISL_2671535, EPI_ISL_2671637, EPI_ISL_2671645 | Laboratorio Central de Epidemiologia (LCE) | Unidad de Genomica Avanzada | ; Alejandra Garcia-Gasca; Alejandra Hernandez-Teran; Alejandro Sanchez-Flores; Alfredo Herrera-Estrella; Alicia Ocaña-Mondragon; Andreu Comas-Garcia; Angel Gustavo Salas-Lais; Antonio Loza Roman; Bernardo Martinez-Miguel; Blanca Taboada; Brenda Irasema Maldonado-Meza; Bruno Gomez-Gil; Carla Ivon Herrera-Najera; Carlos F. Arias; Celia Boukadida; Celida Duque Molina; Celida Martinez- Rodriguez; Clara Esperanza Santacruz-Tinoco; Concepcion Grajales-Muñiz; Consorcio Mexicano de Vigilancia Genomica (CoViGen-Mex). Authors (in alphabetical order): Julio Elias Alvarado-Yaah; Cristobal Chaidez-Quiroz; Daniel Fregoso-Rueda; Daniel Lira Morales; Eduardo Becerril-Vargas; Fernando Fontove-Herrera; Fidencio Mejia-Nepomuceno; Francisco Pulido; Gloria Elena Espinosa-Ayala; Gloria Maria Molina-Salinas; Gloria Vazquez; Hector Esteban Paz-Juarez; Hector Montoya-Fuentes; Helen Haydee Fernanda Ramirez-Plascencia; Irvin Gonzalez-Lopez; Jean Pierre Gonzalez; Jesus Hernandez; Joel Armando Vazquez-Perez.; Jorge Salas-Hernandez; Jose Antonio Enciso-Moreno; Jose Arturo Martinez-Orozco; Jose Esteban Muñoz-Medina; Jose de Jesus Nuñez-Contreras; Juan Bautista Chale-Dzul; Julissa Enciso-Ibarra; Luis Alberto Ochoa-Carrera; Margarita Matias-Florentino; Maria Guadalupe Santiago-Mauricio; Maria Guadalupe de Jesus Mireles-Rivera; Mario Mujica-Sanchez; Marissa Perez-Garcia; Nelly Selem-Mojica; Pavel Isa; Ricardo Ciria Merce; Ricardo Grande; Rosa Maria Gutierrez Rios; Santiago avila-Rios; Selene Zarate; Susana Lopez; Veronica Mata-Haro; Victor Eduardo Garcia-Arias; Victor Hugo Borja-Aburto |
| EPI_ISL_2490388, EPI_ISL_2490391 | Laboratorio Central de Epidemiología (LCE) | Centro de Investigación en Enfermedades Infecciosas (CIENI), Instituto Nacional de Enfermedades Respiratorias (INER) | ; Alejandra Garcia-Gasca; Alejandra Hernández-Terán; Alejandro Sanchez-Flores; Alfredo Herrera-Estrella; Alicia Ocaña-Mondragón; Andreu Comas-García; Angel Gustavo Salas-Lais; Antonio Loza Román; Bernardo Martínez-Miguel; Blanca Taboada; Brenda Irasema Maldonado-Meza; Bruno Gomez-Gil; Carla Ivón Herrera-Najera; Carlos F. Arias; Celia Boukadida; Clara Esperanza Santacruz-Tinoco; Concepción Grajales-Muñiz; Consorcio Mexicano de Vigilancia Genómica (CoViGen-Mex). Authors (in alphabetical order): Julio Elias Alvarado-Yaah; Cristóbal Cháidez-Quiróz; Célida Duque Molina; Célida Martínez-Rodríguez; Daniel Fregoso-Rueda; Daniel Lira Morales; Eduardo Becerril-Vargas; Fernando Fontove-Herrera; Fidencio Mejía-Nepomuceno; Francisco Pulido; Gloria Elena Espinosa-Ayala; Gloria María Molina-Salinas; Gloria Vazquez; Hector Esteban Paz-Juárez; Hector Montoya-Fuentes; Helen Haydee Fernanda Ramírez-Plascencia; Irvin González-López; Jean Pierre González; Joel Armando Vázquez-Pérez.; Jorge Salas-Hernández; José Antonio Enciso-Moreno; José Arturo Martínez-Orozco; José Esteban Muñoz-Medina; José de Jesús Nuñez-Contreras; Juan Bautista Chale-Dzul; Julissa Enciso-Ibarra; Luis Alberto Ochoa-Carrera; Margarita Matías-Florentino; Mario Mújica-Sánchez; Marissa Perez-Garcia; María Guadalupe Santiago-Mauricio; María Guadalupe de Jesús Mireles-Rivera; Nelly Sélem-Mojica; Pavel Isa; Ricardo Ciria Merce; Ricardo Grande; Rosa María Gutierrez Rios; Santiago Ávila-Ríos; Selene Zárate; Susana Lopez; Victor Eduardo Garcia-Arias; Víctor Hugo Borja-Aburto |
| EPI_ISL_2391526, | Laboratorio Central de Epidemiología | Instituto de Biotecnología de la | ; Alejandra Garcia-Gasca; Alejandra García-Gasca; Alejandra Hernández-Terán; Alejandro Sanchez-Flores; Alejandro Sánchez-Flores; Alfredo Herrera-Estrella; Alicia Ocaña-Mondragón; Andreu Comas-García; Angel Gustavo Salas-Lais; Antonio Loza Román; Bernardo Martínez-Miguel; Blanca Taboada; Brenda |

| | | | |
|---|---|---|---|
| EPI_ISL_2391549, EPI_ISL_2391609, EPI_ISL_2391610, EPI_ISL_2681081 | (LCE) | UNAM | Irasema Maldonado-Meza; Bruno Gomez-Gil; Bruno Gómez-Gil; Carla Ivón Herrera-Najera; Carlos F. Arias; Celia Boukadida; Clara Esperanza Santacruz-Tinoco; Concepción Grajales-Muñiz; Consorcio Mexicano de Vigilancia Genómica (CoViGen-Mex). Authors (in alphabetical order): Julio Elias Alvarado-Yaah; Cristóbal Cháidez-Quiróz; Célida Duque Molina; Célida Martínez- Rodríguez; Daniel Fregoso-Rueda; Daniel Lira Morales; Eduardo Becerril-Vargas; Fernando Fontove-Herrera; Fidencio Mejía-Nepomuceno; Francisco Pulido; Gloria Elena Espinosa-Ayala; Gloria María Molina-Salinas; Gloria Vazquez; Hector Esteban Paz-Juárez; Hector Montoya-Fuentes; Helen Haydee Fernanda Ramírez-Plascencia; Irvin González-López; Jean Pierre González; Jesús Hernández; Joel Armando Vázquez-Pérez.; Jorge Salas-Hernández; José Antonio Enciso-Moreno; José Arturo Martínez-Orozco; José Esteban Muñoz-Medina; José de Jesús Nuñez-Contreras; Juan Bautista Chale-Dzul; Julissa Enciso-Ibarra; Luis Alberto Ochoa-Carrera; Margarita Matías-Florentino; Mario Mújica-Sánchez; Marissa Perez-Garcia; María Guadalupe Santiago-Mauricio; María Guadalupe de Jesús Mireles-Rivera; Nelly Sélem-Mojica; Pavel Isa; Ricardo Ciria Merce; Ricardo Grande; Rosa María Gutierrez Rios; Rosa María Gutiérrez Rios; Santiago Ávila-Ríos; Selene Zárate; Susana Lopez; Verónica Mata-Haro; Victor Eduardo Garcia-Arias; Victor Eduardo García-Arias; Víctor Hugo Borja-Aburto |
| EPI_ISL_2402216 | Laboratorio Central de Epidemiología (LCE) | Unidad de Genomica Avanzada | Alejandro Sanchez-Flores; Alfredo Herrera-Estrella; Alicia Ocana-Mondragon; Angel Gustavo Salas-Lais; Bernardo Martinez-Miguel; Blanca Taboada; Brenda Irasema Maldonado-Meza; Carla Ivon Herrera-Najera; Carlos F. Arias; Celia Boukadida; Clara Esperanza Santacruz-Tinoco; Concepcion Grajales-Muniz; Consorcio Mexicano de Vigilancia Genomica (CoViGen-Mex). Authors (in alphabetical order): Julio Elias Alvarado-Yaah; Fernando Fontove-Herrera; Francisco Pulido; Gloria Elena Espinoza-Ayala; Gloria Maria Molina-Salinas; Gloria Vazquez; Hector Esteban Paz-Juarez; Hector Montoya-Fuentes; Helen Haydee Fernanda Ramirez-Plascencia; Jorge Ivan Salinal-Nevarez; Jose Antonio Enciso-Moreno; Jose Esteban Munoz-Medina; Jose de Jesus Nunez-Contreras; Juan Bautista Chale-Dzul; Luis Alberto Ochoa-Carrera; Margarita Matias-Florentino; Maria Guadalupe Santiago-Mauricio; Maria Guadalupe de Jesus Mireles-Rivera; Nelly Selem-Mojica; Pavel Isa; Ricardo Grande; Santiago Avila-Rios; Victor Hugo Borja-Aburto |
| EPI_ISL_3149735, EPI_ISL_3149736, EPI_ISL_3388827 | Laboratorio Central de Salud Publica | Laboratorio de Biología Molecular, Instituto de Medicina Regional on behalf of 'Proyecto Argentino Interinstitucional de genomica de SARS-CoV-2' (PAIS Consortium) | Andrea Ayala; Bettina Brusés; Erica Struss; Esteban Paredes; Griselda Oria; Horacio Lucero.; Javier Mussin; Laura Formichelli; Melina Lorenzini Campos; Raúl Maximiliano Acevedo; Verónica Gómez; Victoria Femenías |
| EPI_ISL_2196253, EPI_ISL_2196260, EPI_ISL_2196261, EPI_ISL_2274072, EPI_ISL_2274073, EPI_ISL_2274074, EPI_ISL_2274075, EPI_ISL_2466262, EPI_ISL_2466263, EPI_ISL_2466264, EPI_ISL_2557337, EPI_ISL_2557338, EPI_ISL_2557339 | | | |
| see above | Laboratorio Central de Saude Publica do Estado da Bahia (LACEN/BA) | Laboratory of Respiratory Viruses and Measles, Oswaldo Cruz Institute, FIOCRUZ | Alice Sampaio Rocha; Ana Carolina Mendonca; Anderson Brandao Leite; Anna Carolina Paixao; Elisa Cavalcante Pereira; Felicidade Pereira; Fernando Motta; Luciana Appolinario; Marilda Siqueira on behalf of the Fiocruz COVID-19 Genomic Surveillance Network; Paola Resende; Renata Serrano Lopes; Taina Venas |
| EPI_ISL_2536323, EPI_ISL_2536338, EPI_ISL_2536344, EPI_ISL_3061882, EPI_ISL_3061889, EPI_ISL_3061890 | Laboratorio Central de Saude Publica do Estado da Paraiba (LACEN-PB) | Laboratory of Respiratory Viruses and Measles, Oswaldo Cruz Institute, FIOCRUZ | Alice Sampaio Rocha; Ana Carolina Mendonca; Anna Carolina Paixao; Dalane Loudal Florentino Teixeira; Elisa Cavalcante Pereira; Fernando Motta; Joao Felipe Bezerra; Luciana Appolinario; Marilda Siqueira on behalf of the Fiocruz COVID-19 Genomic Surveillance Network; Paola Resende; Renata Serrano Lopes; Taina Venas |
| EPI_ISL_3190189 | Laboratorio Central de Saude Publica do Estado da Paraiba (LACEN/PB) | Laboratory of Respiratory Viruses and Measles, Oswaldo Cruz Institute, FIOCRUZ | Alice Sampaio Rocha; Ana Carolina Mendonca; Anna Carolina Paixao; Dalane Loudal Florentino Teixeira; Elisa Cavalcante Pereira; Fernando Motta; Joao Felipe Bezerra; Luciana Appolinario; Marilda Siqueira on behalf of the Fiocruz COVID-19 Genomic Surveillance Network; Paola Resende; Renata Serrano Lopes; Taina Venas |
| EPI_ISL_2274069, EPI_ISL_2274071, EPI_ISL_2466231, EPI_ISL_2466232, EPI_ISL_2466233, EPI_ISL_2466235, EPI_ISL_2466236, EPI_ISL_2466237, EPI_ISL_2466238, EPI_ISL_2466239, EPI_ISL_2466240, EPI_ISL_2466241, EPI_ISL_2466242, EPI_ISL_2466243, EPI_ISL_2466244, EPI_ISL_2466245, EPI_ISL_2466246, EPI_ISL_2466247, EPI_ISL_2466248, EPI_ISL_2466249, EPI_ISL_2466250, EPI_ISL_2466251, EPI_ISL_2466252, EPI_ISL_2466253, EPI_ISL_2466254, EPI_ISL_2466255, EPI_ISL_2466256, EPI_ISL_2466257, EPI_ISL_2466258, EPI_ISL_2466259, EPI_ISL_2557310, EPI_ISL_2557311, EPI_ISL_2557312, EPI_ISL_2557313, EPI_ISL_2557314, EPI_ISL_2557315, EPI_ISL_2557316, EPI_ISL_2557317, EPI_ISL_2557318, EPI_ISL_2557319, EPI_ISL_2557320, EPI_ISL_2557321, EPI_ISL_2557322, EPI_ISL_2557323, EPI_ISL_2557324, EPI_ISL_2557325, EPI_ISL_2557326, EPI_ISL_2557327, EPI_ISL_2557328, EPI_ISL_2557329, EPI_ISL_2557330, EPI_ISL_2557331, EPI_ISL_2557332, EPI_ISL_2557333, EPI_ISL_2557334, EPI_ISL_2557335, EPI_ISL_2557336, EPI_ISL_2645384, EPI_ISL_2645388, EPI_ISL_2645389, EPI_ISL_2645390, EPI_ISL_2645391, EPI_ISL_2645392, EPI_ISL_2645394, EPI_ISL_2645395, EPI_ISL_2645396, EPI_ISL_2645402, EPI_ISL_2645406, EPI_ISL_2645407 | | | |
| see above | Laboratorio Central de Saude Publica do Estado de Alagoas (LACEN/AL) | Laboratory of Respiratory Viruses and Measles, Oswaldo Cruz Institute, FIOCRUZ | Alice Sampaio Rocha; Ana Carolina Mendonca; Anderson Brandao Leite; Anna Carolina Paixao; Elisa Cavalcante Pereira; Fernando Motta; Luciana Appolinario; Marilda Siqueira on behalf of the Fiocruz COVID-19 Genomic Surveillance Network; Paola Resende; Renata Serrano Lopes; Taina Venas |
| EPI_ISL_3235302, EPI_ISL_3832396, EPI_ISL_3832439, EPI_ISL_3832440, EPI_ISL_3832441, EPI_ISL_3832442, EPI_ISL_3832443, EPI_ISL_4061434, EPI_ISL_4061435, EPI_ISL_4061436 | | | |
| see above | Laboratorio Central de Saude Publica do Estado de Santa Catarina (LACEN/SC) | Laboratory of Respiratory Viruses and Measles, Oswaldo Cruz Institute, FIOCRUZ | Alice Sampaio Rocha; Ana Carolina Mendonca; Anna Carolina Paixao; Darcita Buerger Rovaris; Elisa Cavalcante Pereira; Fernando Motta; Luciana Appolinario; Marilda Siqueira on behalf of the Fiocruz COVID-19 Genomic Surveillance Network; Paola Resende; Renata Serrano Lopes; Sandra Bianchini Fernandes; Taina Venas |
| EPI_ISL_2863618, EPI_ISL_2863619, EPI_ISL_2863625, EPI_ISL_2863628, EPI_ISL_2863635 | Laboratorio Central de Saude Publica do Estado de Sergipe (LACEN/SE) | Laboratory of Respiratory Viruses and Measles, Oswaldo Cruz Institute, FIOCRUZ | Alice Sampaio Rocha; Ana Carolina Mendonca; Anna Carolina Paixao; Cliomar Alves dos Santos; Elisa Cavalcante Pereira; Fernando Motta; Luciana Appolinario; Marilda Siqueira on behalf of the Fiocruz COVID-19 Genomic Surveillance Network; Paola Resende; Renata Serrano Lopes; Tainá Moreira Martins Venas |
| EPI_ISL_2196262, EPI_ISL_3045432, EPI_ISL_3045433, EPI_ISL_3045448, EPI_ISL_3045462, EPI_ISL_3190238, EPI_ISL_3190239, EPI_ISL_3190241, EPI_ISL_3190242, EPI_ISL_3190243 | | | |
| see above | Laboratorio Central de Saude Publica do Estado do Espirito Santo (LACEN/ES) | Laboratory of Respiratory Viruses and Measles, Oswaldo Cruz Institute, FIOCRUZ | Alice Sampaio Rocha; Ana Carolina Mendonca; Anna Carolina Paixao; Eilisa Cavalcante Pereira; Elisa Cavalcante Pereira; Fernando Motta; Luciana Appolinario; Marilda Siqueira on behalf of the Fiocruz COVID-19 Genomic Surveillance Network; Paola Resende; Renata Serrano Lopes; Rodrigo Ribeiro Rodrigues; Taina Venas |
| EPI_ISL_2983189, EPI_ISL_2983190, EPI_ISL_2983191, EPI_ISL_2983193, EPI_ISL_2983195, EPI_ISL_2983241, EPI_ISL_2983243, EPI_ISL_2983250, EPI_ISL_2983251, EPI_ISL_2983253, EPI_ISL_2983254, EPI_ISL_2983268, EPI_ISL_2983269, EPI_ISL_2983270, EPI_ISL_2983272, EPI_ISL_2983273, EPI_ISL_2983274, EPI_ISL_2983275, EPI_ISL_2983276, EPI_ISL_2983279, EPI_ISL_2983282, EPI_ISL_2983283, EPI_ISL_2983284, EPI_ISL_2983285, EPI_ISL_2983286, EPI_ISL_2983287, EPI_ISL_2983288, EPI_ISL_2983289, EPI_ISL_2983291, EPI_ISL_2983292, EPI_ISL_2983293, EPI_ISL_2983294, EPI_ISL_2983295, EPI_ISL_2983296, EPI_ISL_2983297, EPI_ISL_2983298, EPI_ISL_2983300, EPI_ISL_2983301, EPI_ISL_2983303, EPI_ISL_2983304, EPI_ISL_2983325, EPI_ISL_2983326, EPI_ISL_2983327, EPI_ISL_2983328, EPI_ISL_2983333, EPI_ISL_2983334, EPI_ISL_2983336, EPI_ISL_3190255, EPI_ISL_3190257, EPI_ISL_3190258 | | | |
| see above | Laboratorio Central de Saude Publica do Estado do Maranhao (LACEN-MA) | Laboratory of Respiratory Viruses and Measles, Oswaldo Cruz Institute, FIOCRUZ | Agatha Cristinne Prudencio; Agatha Soares; Alice Sampaio Rocha; Ana Carolina Mendonca; Anna Carolina Paixao; Elisa Cavalcante Pereira; Fernando Motta; Ighor Arantes; Ighor Leonardo Arantes Gomes; Lidio Gonçalves Lima Neto; Luciana Appolinario; Marilda Siqueira on behalf of the Fiocruz COVID-19 Genomic Surveillance Network; Paola Resende; Renata Serrano Lopes; Taina Moreira Venas; Tainá Venas |
| EPI_ISL_2536354 | Laboratorio Central de Saude Publica do Estado do Para (LACEN/PA) | Laboratory of Respiratory Viruses and Measles, Oswaldo Cruz Institute, FIOCRUZ | Alice Sampaio Rocha; Ana Carolina Mendonca; Anna Carolina Paixao; Elisa Cavalcante Pereira; Fernando Motta; Luciana Appolinario; Marilda Siqueira on behalf of the Fiocruz COVID-19 Genomic Surveillance Network; Paola Resende; Renata Serrano Lopes; Taina Venas; Valnete Andrade |
| EPI_ISL_2645458, EPI_ISL_2645461, EPI_ISL_2645463, EPI_ISL_2645465, EPI_ISL_2645466, EPI_ISL_2645470, EPI_ISL_2645471, EPI_ISL_2645472, EPI_ISL_2645474, EPI_ISL_2645482, EPI_ISL_2645484, EPI_ISL_2645485, EPI_ISL_2645487, EPI_ISL_2645491, EPI_ISL_2645492, EPI_ISL_2645494, EPI_ISL_2645495, EPI_ISL_2645497, EPI_ISL_2645498, EPI_ISL_2645499, EPI_ISL_2645500, EPI_ISL_2645501, EPI_ISL_2645503, EPI_ISL_2645504, EPI_ISL_2983447, EPI_ISL_2983448, EPI_ISL_2983450, EPI_ISL_4061299, EPI_ISL_4061300, EPI_ISL_4061301, EPI_ISL_4061302, EPI_ISL_4061305, EPI_ISL_4061306, EPI_ISL_4061308, EPI_ISL_4061309, EPI_ISL_4061312, EPI_ISL_4061313, EPI_ISL_4061314, EPI_ISL_4061315, EPI_ISL_4061317, EPI_ISL_4061318, EPI_ISL_4061320, EPI_ISL_4061321, EPI_ISL_4061322, EPI_ISL_4061323, EPI_ISL_4061324, EPI_ISL_4061325, EPI_ISL_4061326 | | | |
| see above | Laboratorio Central de Saude Publica do Estado do Parana (LACEN/PR) | Laboratory of Respiratory Viruses and Measles, Oswaldo Cruz Institute, FIOCRUZ | Agatha Soares; Alice Sampaio Rocha; Ana Carolina Mendonca; Anna Carolina Paixao; Elisa Cavalcante Pereira; Fernando Motta; Ighor Leonardo Arantes; Irina Riediger; Luciana Appolinario; Marilda Siqueira on behalf of the Fiocruz COVID-19 Genomic Surveillance Network; Paola Resende; Renata Serrano Lopes; Taina Venas |
| EPI_ISL_2603432, EPI_ISL_2603433, EPI_ISL_2603436, EPI_ISL_2603442, EPI_ISL_2603443, EPI_ISL_2603444, EPI_ISL_2603445, EPI_ISL_2603446, EPI_ISL_2603447, EPI_ISL_2603448, EPI_ISL_2603450, EPI_ISL_2603536, EPI_ISL_2603537, EPI_ISL_2603538, EPI_ISL_2603539, EPI_ISL_2603540, EPI_ISL_2603541, EPI_ISL_2603542, EPI_ISL_2603547, EPI_ISL_2603548, EPI_ISL_2603550, EPI_ISL_2603551, EPI_ISL_2603552, EPI_ISL_2603553, EPI_ISL_2603554, EPI_ISL_2603555, EPI_ISL_2603556, EPI_ISL_2603559, EPI_ISL_2603560, EPI_ISL_2603562, EPI_ISL_2603563, EPI_ISL_2603564, EPI_ISL_2603565, EPI_ISL_2603566, EPI_ISL_2603567, EPI_ISL_2603568, EPI_ISL_2603569, EPI_ISL_2603571, EPI_ISL_2603572, EPI_ISL_2603573, EPI_ISL_2603574, EPI_ISL_2603575, EPI_ISL_2603576, EPI_ISL_2603577, EPI_ISL_2603578, EPI_ISL_2603579, EPI_ISL_2603581, EPI_ISL_2603583, EPI_ISL_2603586, EPI_ISL_2603587, EPI_ISL_2603588, EPI_ISL_2603589, EPI_ISL_2603590, EPI_ISL_2982757, EPI_ISL_2982758, EPI_ISL_2982759, EPI_ISL_2982760, EPI_ISL_2982761, EPI_ISL_2982770, EPI_ISL_2982771, EPI_ISL_2982772, EPI_ISL_2982773, EPI_ISL_2982774, EPI_ISL_2982778, EPI_ISL_2982782, EPI_ISL_2982783, EPI_ISL_2982785, EPI_ISL_2982787, EPI_ISL_2982789, EPI_ISL_2982790, EPI_ISL_2982791, EPI_ISL_2982793 | | | |
| see above | Laboratorio Central de Saude Publica do Estado do Rio Grande do Sul (LACEN-RS) | Laboratory of Respiratory Viruses and Measles, Oswaldo Cruz Institute, FIOCRUZ | Alice Sampaio Rocha; Ana Carolina Mendonca; Anderson Brandao Leite; Anna Carolina Paixao; Elisa Cavalcante Pereira; Fernando Motta; Luciana Appolinario; Marilda Siqueira on behalf of the Fiocruz COVID-19 Genomic Surveillance Network; Paola Resende; Renata Serrano Lopes; Taina Venas |
| EPI_ISL_3048803, EPI_ISL_3048805, EPI_ISL_3048807, EPI_ISL_3048808 | Laboratorio Central de Saude Publica do Estado do Rio Grande do Sul (LACEN-RS) | Laboratório de Biologia Molecular da Universidade Federal de Ciências da Saúde de Porto Alegre | Adriana Seixas; Ana B. G. Veiga; Ana Paula Muterle Varela; Fabiana Quoos Mayer; Fernando Hayashi Sant'Anna; Janira Prichula; Letícia Garay Martins; Richard Steiner Salvato; Tatiana Schäffer Gregianini |
| EPI_ISL_2983213 | Laboratorio Central de Saude Publica do Estado do Rio de Janeiro (LACEN-RJ) | Laboratory of Respiratory Viruses and Measles, Oswaldo Cruz Institute, FIOCRUZ | Agatha Cristinne Prudencio; Alice Sampaio Rocha; Ana Carolina Mendonca; Andrea Cony Cavalcanti; Anna Carolina Paixao; Elisa Cavalcante Pereira; Fernando Motta; Ighor Leonardo Arantes Gomes; Luciana Appolinario; Marilda Siqueira on behalf of the Fiocruz COVID-19 Genomic Surveillance Network; Paola Resende; Renata Serrano Lopes; Taina Venas |
| EPI_ISL_3033037, EPI_ISL_3033040, EPI_ISL_3033041, EPI_ISL_3033044, EPI_ISL_3033052, EPI_ISL_3033053, EPI_ISL_3033054, EPI_ISL_3033055, EPI_ISL_3033057, EPI_ISL_3033060, EPI_ISL_3033062, EPI_ISL_3033063 | | | |
| see above | Laboratorio Central de la Ciudad de Santa Fe | Grupo de Genómica y Bioinformática del Instituto de Investigación de la Cadena Láctea CONICET-INTA on behalf of 'Proyecto Argentino Interinstitucional de genomica de SARS-CoV-2' (PAIS Consortium) | AF; Amadio; C; Eberhardt; G; Irazoqui; JM; MF; Mugna; Ojeda; Pastor; Rompato; V |
| EPI_ISL_2271688, EPI_ISL_2271690, EPI_ISL_2271708 | Laboratorio Central, Ministerio de Salud Cordoba | Instituto de Patologia Vegetal (CIAP-INTA) on behalf of 'Proyecto Argentino Interinstitucional de genomica de SARS-CoV-2' (PAIS Consortium) | Barbas, G.; Castro, G.; Debat, HJ.; FD; Fernandez; M; M.B.; Marquez, N.; Pisano; Re, V. |
| EPI_ISL_3333567 | Laboratorio Clínico Gamma | MELISA Institute Genomics and Proteomics SpA | Cristian Vargas; Elard Koch; Francisco Alvarez; Guillermo Nourdin; Mauricio Hernandez |
| EPI_ISL_3045409, EPI_ISL_3045415, EPI_ISL_3045418, EPI_ISL_3045419, | Laboratorio Nacional de Salud Pública Dr. Defilló - LNSPDD | Laboratory of Respiratory Viruses and Measles, Oswaldo Cruz Institute, FIOCRUZ | Alice Sampaio Rocha; Ana Carolina Mendonca; Anna Carolina Paixao; Elisa Cavalcante Pereira; Fernando Motta; Grey Benoit Vasquez; Isaac Miguel Sanchez; Ivonne Imbert; Lucia de la Cruz; Luciana Appolinario; Marilda Siqueira on behalf of the Fiocruz COVID-19 Genomic Surveillance Network; Nurys de Castro; Paola Resende; Renata Serrano Lopes; Ronald Skewes; Taina Venas |

| | | | |
|---|---|---|---|
| EPI_ISL_3045420 | | | |
| EPI_ISL_3275219 | Laboratorio Nacional de Salud, Ministerio de Salud Publica y Asistencia Social | Genomics and Proteomics Departament, Gorgas Memorial Institute For Health Studies | Alexander A Martinez; Ambar Moreno; Claudia Estrada; Claudia Gonzalez V; César Roberto Conde Pereira; Jessica Gondola; Leyda Abrego; Marlene Castillo; Oris Chavarria |
| EPI_ISL_3307458 | Laboratorio SYNLAB Colombia | Corporación para Investigaciones Biológicas-CIB | Jeanneth Mosquera Rendon; Jenny Santiago Cuesta; Marcela Mercado Reyes; Uriel A. Hurtado Paez |
| EPI_ISL_2777494, EPI_ISL_4030325, EPI_ISL_4030327, EPI_ISL_4030335, EPI_ISL_4030337, EPI_ISL_4030339, EPI_ISL_4030346, EPI_ISL_4030349, EPI_ISL_4030355, EPI_ISL_4030358, EPI_ISL_4030361, EPI_ISL_4030364 | | | |
| see above | Laboratorio de Ecologia de Doencas Transmissiveis na Amazonia, Instituto Leonidas e Maria Deane - Fiocruz Amazonia | Laboratorio de Ecologia de Doencas Transmissiveis na Amazonia, Instituto Leonidas e Maria Deane - Fiocruz Amazonia | Alex Martins; André Corado; Debora Duarte; Fabiola Mendonça da Silva Chui; Felipe Naveca; Fernanda Nascimento; Fernando Fonseca de Almeida Val; George Silva; Gisely Cardoso de Melo; Karina Pessoa; Luciana Gonçalves; Marcus Vinicius Guimarães de Lacerda; Maria Júlia Brandão; Maria Paula Gomes Mourão; Mariana Xavier Simão; Matilde Mejía; Michele Jesus; Valdinete Nascimento; Vanderson de Souza Sampaio; Victor Souza; Ágatha Costa |
| EPI_ISL_2728553 | Laboratorio de Infectologia y Virologia Molecular | Laboratory of Molecular Virology, School of Medicine, Pontificia Universidad Catolica de Chile | Alejandro Bhrun; Ana Maria Contreras; Andres E. Munoz-Marcos; Carlos Palma; Catalina Pardo-Roa; Constanza Maldonado; Constanza Martinez-Valdevenito; Eileen Serrano; Erick Salinas; Estefany Poblete; Francisco Melo; Jennifer Angulo; Jorge Levican; Leonardo I. Almonacid; M. Belen Leyton; Magdalena Vera; Marcela Ferres; Maria Jose Avendano; Rafael A. Medina; Tamara Garcia-Salum |
| EPI_ISL_2854140 | Laboratorio de Medicina Molecular, Universidad de Magallanes | Centro Asistencial Docente y de Investigacion, Universidad de Magallanes | Constanza Ceroni; Diego Alvarez; Jacqueline Aldridge; Jorge González; Marcelo Navarrete; Roberto Uribe-Paredes |
| EPI_ISL_2008913, EPI_ISL_2008915, EPI_ISL_2008923, EPI_ISL_2008927, EPI_ISL_2008931, EPI_ISL_3048959, EPI_ISL_3048960, EPI_ISL_3048961, EPI_ISL_3048962, EPI_ISL_3048964, EPI_ISL_3048966, EPI_ISL_3048968, EPI_ISL_3048969, EPI_ISL_3048970, EPI_ISL_3048971, EPI_ISL_3048972, EPI_ISL_3048973, EPI_ISL_3048974, EPI_ISL_3048976, EPI_ISL_3048977, EPI_ISL_3048978, EPI_ISL_3761613, EPI_ISL_3761616, EPI_ISL_3761636, EPI_ISL_3761645, EPI_ISL_3761646, EPI_ISL_3761749, EPI_ISL_3761752, EPI_ISL_3761763 | | | |
| see above | Laboratorio de Pesquisa em Virologia, FAMERP, SJRP | Laboratorio de Pesquisa em Virologia, FAMERP, SJRP | Beatriz de Carvalho Marques; Cecília Artico Banho; Cíntia Bittar; Fábio Sossai Possebon; Guilherme Campos; Helena Lage Ferreira; Jorge A. Petroli Marchesi; João Pessoa Araújo Jr.; Leila Sabrina Ullmann; Lívia Sacchetto; Maisa C. Pereira Parra; Marília Moraes; Maurício L. Nogueira.; Paula Rahal; Paulo Inacio da Costa |
| EPI_ISL_3401433, EPI_ISL_3401453, EPI_ISL_3401455, EPI_ISL_3401458, EPI_ISL_3401459, EPI_ISL_3401463, EPI_ISL_3401464, EPI_ISL_3672413, EPI_ISL_3672414, EPI_ISL_3672415, EPI_ISL_3672418, EPI_ISL_3826392, EPI_ISL_3826393, EPI_ISL_3826394, EPI_ISL_3826395, EPI_ISL_3826396, EPI_ISL_3826397, EPI_ISL_3826398, EPI_ISL_3826399, EPI_ISL_3826400, EPI_ISL_3826401, EPI_ISL_3826403, EPI_ISL_3826404, EPI_ISL_3826407, EPI_ISL_3826409, EPI_ISL_3826410, EPI_ISL_3826411, EPI_ISL_3826413, EPI_ISL_4052702, EPI_ISL_4052714, EPI_ISL_4052715, EPI_ISL_4052717, EPI_ISL_4052719, EPI_ISL_4052720, EPI_ISL_4052721, EPI_ISL_4052722, EPI_ISL_4052724, EPI_ISL_4052725, EPI_ISL_4052726, EPI_ISL_4052728, EPI_ISL_4052729 | | | |
| see above | Laboratorio de Referencia Nacional de Virus Respiratorios. Centro Nacional de Salud Publica. Instituto Nacional de Salud Peru. | Laboratorio de Referencia Nacional de Virus Respiratorios. Centro Nacional de Salud Publica. Instituto Nacional de Salud Peru. | Carlos Padilla Rojas; Henri Bailon Calderon; Iris Silva Molina; Joseph Huayra Niquen; Lely Solari Zerpa; Luis Barcena Flores; Marco Galarza Perez; Nancy Rojas Serrano; Nieves Sevilla Castañeda; Omar Caceres Rey; Orson Mestanza Millones; Princesa Medrano Alhuay; Priscila Lope Pari; Sandra Morales Ruiz; Sara Gordillo Vilchez; Steve Acedo Lazo; Veronica Hurtado Vela; Victor Jimenez Vasquez; Wendy Lizarraga Olivares |
| EPI_ISL_3023587, EPI_ISL_3023588, EPI_ISL_3023599, EPI_ISL_3375938, EPI_ISL_3376007, EPI_ISL_3376013, EPI_ISL_3376020, EPI_ISL_3376021, EPI_ISL_3376023, EPI_ISL_3376031, EPI_ISL_3376382, EPI_ISL_3376383 | | | |
| see above | Laboratorio de Referencial Nacional de Virus Respiratorios | Laboratorio de Referencial Nacional de Virus Respiratorios | Carlos Padilla Rojas; Henri Bailon Calderon; Iris Silva Molina; Joseph Huayra Niquen; Lely Solari Zerpa; Luis Barcena Flores; Marco Galarza Perez; Nancy Rojas Serrano; Omar Caceres Rey; Orson Mestanza Millones; Priscila Lope Pari; Sandra Morales Ruiz; Steve Acedo Lazo; Veronica Hurtado Vela |
| EPI_ISL_1926315, EPI_ISL_1926316, EPI_ISL_1926589, EPI_ISL_1926612, EPI_ISL_1926667, EPI_ISL_1926675, EPI_ISL_1926678, EPI_ISL_1926685, EPI_ISL_1926690, EPI_ISL_1926829, EPI_ISL_1926835, EPI_ISL_1926839, EPI_ISL_1926989, EPI_ISL_1927003, EPI_ISL_1927009, EPI_ISL_1927018, EPI_ISL_1927055, EPI_ISL_1927072, EPI_ISL_1927085, EPI_ISL_1927142, EPI_ISL_1927259, EPI_ISL_1927307, EPI_ISL_1928731, EPI_ISL_1928733, EPI_ISL_1928741, EPI_ISL_1928742, EPI_ISL_1928787, EPI_ISL_1928803, EPI_ISL_1928809, EPI_ISL_1928811, EPI_ISL_1928814, EPI_ISL_1928818, EPI_ISL_1928828, EPI_ISL_1928879, EPI_ISL_1928897, EPI_ISL_1928907, EPI_ISL_1928908, EPI_ISL_1928909, EPI_ISL_1928928, EPI_ISL_1928929, EPI_ISL_1928980, EPI_ISL_1928986, EPI_ISL_1928987, EPI_ISL_1929039, EPI_ISL_1929111, EPI_ISL_1929137, EPI_ISL_1929161, EPI_ISL_1929198, EPI_ISL_1929201, EPI_ISL_1929323, EPI_ISL_1929370, EPI_ISL_1929553, EPI_ISL_1929984, EPI_ISL_1930106, EPI_ISL_1930112, EPI_ISL_1930276, EPI_ISL_1930412, EPI_ISL_1930439, EPI_ISL_1930531, EPI_ISL_1930579, EPI_ISL_2044024, EPI_ISL_2044065, EPI_ISL_2044267, EPI_ISL_2044278, EPI_ISL_2044295, EPI_ISL_2044456, EPI_ISL_2044471, EPI_ISL_2044472, EPI_ISL_2044487, EPI_ISL_2044489, EPI_ISL_2044496, EPI_ISL_2044499, EPI_ISL_2044501, EPI_ISL_2044505, EPI_ISL_2044555, EPI_ISL_2044602, EPI_ISL_2044671, EPI_ISL_2044683, EPI_ISL_2044693, EPI_ISL_2044697, EPI_ISL_2044698, EPI_ISL_2044699, EPI_ISL_2044720, EPI_ISL_2044853, EPI_ISL_2045023, EPI_ISL_2045127, EPI_ISL_2045128, EPI_ISL_2045132, EPI_ISL_2045145, EPI_ISL_2045155, EPI_ISL_2045159, EPI_ISL_2045183, EPI_ISL_2045202, EPI_ISL_2045204, EPI_ISL_2045215, EPI_ISL_2045231, EPI_ISL_2045239, EPI_ISL_2045247, EPI_ISL_2045250, EPI_ISL_2045257, EPI_ISL_2045275, EPI_ISL_2045311, EPI_ISL_2045318, EPI_ISL_2045323, EPI_ISL_2045324, EPI_ISL_2045334, EPI_ISL_2045404, EPI_ISL_2045410, EPI_ISL_2045412, EPI_ISL_2045416, EPI_ISL_2045458, EPI_ISL_2045470, EPI_ISL_2045588, EPI_ISL_2045763, EPI_ISL_2045771, EPI_ISL_2045779, EPI_ISL_2045784, EPI_ISL_2045858, EPI_ISL_2045872, EPI_ISL_2046300, EPI_ISL_2046385, EPI_ISL_2046456, EPI_ISL_2046488, EPI_ISL_2046524, EPI_ISL_2046569, EPI_ISL_2046623, EPI_ISL_2046739, EPI_ISL_2046796, EPI_ISL_2182100, EPI_ISL_2182182, EPI_ISL_2182211, EPI_ISL_2182217, EPI_ISL_2182231, EPI_ISL_2182239, EPI_ISL_2182313, EPI_ISL_2182319, EPI_ISL_2182333, EPI_ISL_2182341, EPI_ISL_2182349, EPI_ISL_2182399, EPI_ISL_2182437, EPI_ISL_2182452, EPI_ISL_2182463, EPI_ISL_2182535, EPI_ISL_2182628, EPI_ISL_2182658, EPI_ISL_2182666, EPI_ISL_2182725, EPI_ISL_2182728, EPI_ISL_2182755, EPI_ISL_2182761, EPI_ISL_2182805, EPI_ISL_2182853, EPI_ISL_2182883, EPI_ISL_2182976, EPI_ISL_2182990, EPI_ISL_2183097, EPI_ISL_2183177, EPI_ISL_2183380, EPI_ISL_2183411, EPI_ISL_2183480, EPI_ISL_2183486, EPI_ISL_2183491, EPI_ISL_2183509, EPI_ISL_2183512, EPI_ISL_2183537, EPI_ISL_2183540, EPI_ISL_2183561, EPI_ISL_2183603, EPI_ISL_2183617, EPI_ISL_2183623, EPI_ISL_2183626, EPI_ISL_2183634, EPI_ISL_2183639, EPI_ISL_2183645, EPI_ISL_2183656, EPI_ISL_2183664, EPI_ISL_2183757, EPI_ISL_2183778, EPI_ISL_2183781, EPI_ISL_2183798, EPI_ISL_2183816, EPI_ISL_2183821, EPI_ISL_2183826, EPI_ISL_2183840, EPI_ISL_2183847, EPI_ISL_2183855, EPI_ISL_2183860, EPI_ISL_2183862, EPI_ISL_2183868, EPI_ISL_2183870, EPI_ISL_2183883, EPI_ISL_2183884, EPI_ISL_2183898, EPI_ISL_2183902, EPI_ISL_2183912, EPI_ISL_2183918, EPI_ISL_2183987, EPI_ISL_2183988, EPI_ISL_2184005, EPI_ISL_2184013, EPI_ISL_2184016, EPI_ISL_2184032, EPI_ISL_2184033, EPI_ISL_2184056, EPI_ISL_2184174, EPI_ISL_2184190, EPI_ISL_2184195, EPI_ISL_2184257, EPI_ISL_2184325, EPI_ISL_2184348, EPI_ISL_2184440, EPI_ISL_2184550, EPI_ISL_2184551, EPI_ISL_2184559, EPI_ISL_2184563, EPI_ISL_2184588, EPI_ISL_2184596, EPI_ISL_2184612, EPI_ISL_2184620, EPI_ISL_2184639, EPI_ISL_2184712, EPI_ISL_2184725, EPI_ISL_2184726, EPI_ISL_2184730, EPI_ISL_2184731, EPI_ISL_2184732, EPI_ISL_2184736, EPI_ISL_2184753, EPI_ISL_2184766, EPI_ISL_2184770, EPI_ISL_2184775, EPI_ISL_2184788, EPI_ISL_2184811, EPI_ISL_2184820, EPI_ISL_2184833, EPI_ISL_2184915, EPI_ISL_2185069, EPI_ISL_2185076, EPI_ISL_2185084, EPI_ISL_2185086, EPI_ISL_2185097, EPI_ISL_2185134, EPI_ISL_2185161, EPI_ISL_2185470, EPI_ISL_2185479, EPI_ISL_2185483, EPI_ISL_2304837, EPI_ISL_2304918, EPI_ISL_2304923, EPI_ISL_2304925, EPI_ISL_2304926, EPI_ISL_2304959, EPI_ISL_2304961, EPI_ISL_2304963, EPI_ISL_2305051, EPI_ISL_2305057, EPI_ISL_2305060, EPI_ISL_2305063, EPI_ISL_2305074, EPI_ISL_2305095, EPI_ISL_2305191, EPI_ISL_2305257, EPI_ISL_2305293, EPI_ISL_2305336, EPI_ISL_2305339, EPI_ISL_2305682, EPI_ISL_2305684, EPI_ISL_2305729, EPI_ISL_2305733, EPI_ISL_2305735, EPI_ISL_2305738, EPI_ISL_2305741, EPI_ISL_2305745, EPI_ISL_2305766, EPI_ISL_2305790, EPI_ISL_2305791, EPI_ISL_2305805, EPI_ISL_2305810, EPI_ISL_2305814, EPI_ISL_2305818, EPI_ISL_2305834, EPI_ISL_2305842, EPI_ISL_2305860, EPI_ISL_2305904, EPI_ISL_2305905, EPI_ISL_2305906, EPI_ISL_2305974, EPI_ISL_2306026, EPI_ISL_2306028, EPI_ISL_2306030, EPI_ISL_2306032, EPI_ISL_2306051, EPI_ISL_2306054, EPI_ISL_2306057, EPI_ISL_2306059, EPI_ISL_2306071, EPI_ISL_2306082, EPI_ISL_2306120, EPI_ISL_2306131, EPI_ISL_2306186, EPI_ISL_2306193, EPI_ISL_2306218, EPI_ISL_2306276, EPI_ISL_2306286, EPI_ISL_2306329, EPI_ISL_2306335, EPI_ISL_2306336, EPI_ISL_2306414, EPI_ISL_2306417, EPI_ISL_2306422, EPI_ISL_2306426, EPI_ISL_2306429, EPI_ISL_2306432, EPI_ISL_2306433, EPI_ISL_2306435, EPI_ISL_2306439, EPI_ISL_2306442, EPI_ISL_2306455, EPI_ISL_2306461, EPI_ISL_2306468, EPI_ISL_2306471, EPI_ISL_2306475, EPI_ISL_2306478, EPI_ISL_2306481, EPI_ISL_2306489, EPI_ISL_2306504, EPI_ISL_2306540, EPI_ISL_2306550, EPI_ISL_2306552, EPI_ISL_2306555, EPI_ISL_2306557, EPI_ISL_2306564, EPI_ISL_2306565, EPI_ISL_2306566, EPI_ISL_2306574, EPI_ISL_2306599, EPI_ISL_2306603, EPI_ISL_2306616, EPI_ISL_2306621, EPI_ISL_2306623, EPI_ISL_2306634, EPI_ISL_2306640, EPI_ISL_2306671, EPI_ISL_2306681, EPI_ISL_2306718, EPI_ISL_2306727, EPI_ISL_2306729, EPI_ISL_2306743, EPI_ISL_2306748, EPI_ISL_2306759, EPI_ISL_2306761, EPI_ISL_2306762, EPI_ISL_2306777, EPI_ISL_2306811, EPI_ISL_2306849, EPI_ISL_2306885, EPI_ISL_2306917, EPI_ISL_2306919, EPI_ISL_2306930, EPI_ISL_2306942, EPI_ISL_2306976, EPI_ISL_2306979, EPI_ISL_2306988, EPI_ISL_2306993, EPI_ISL_2306997, EPI_ISL_2307053, EPI_ISL_2324462, EPI_ISL_2324468, EPI_ISL_2324469, EPI_ISL_2324474, EPI_ISL_2324481, EPI_ISL_2324487, EPI_ISL_2324493, EPI_ISL_2324495, EPI_ISL_2324519, EPI_ISL_2324530, EPI_ISL_2324536, EPI_ISL_2324538, EPI_ISL_2324544, EPI_ISL_2324565, EPI_ISL_2324570, EPI_ISL_2324577, EPI_ISL_2324580, EPI_ISL_2324588, EPI_ISL_2397857, EPI_ISL_2397866, EPI_ISL_2397903, EPI_ISL_2397944, EPI_ISL_2397955, EPI_ISL_2397956, EPI_ISL_2397957, EPI_ISL_2397998, EPI_ISL_2398020, EPI_ISL_2398042, EPI_ISL_2398055, EPI_ISL_2398076, EPI_ISL_2398079, EPI_ISL_2398083, EPI_ISL_2398097, EPI_ISL_2398111, EPI_ISL_2398114, EPI_ISL_2398115, EPI_ISL_2398153, EPI_ISL_2398158, EPI_ISL_2398166, EPI_ISL_2398168, EPI_ISL_2398177, EPI_ISL_2398192, EPI_ISL_2398200, EPI_ISL_2398208, EPI_ISL_2398220, EPI_ISL_2398222, EPI_ISL_2398225, EPI_ISL_2398235, EPI_ISL_2398244, EPI_ISL_2398245, EPI_ISL_2398247, EPI_ISL_2398248, EPI_ISL_2398252, EPI_ISL_2398255, EPI_ISL_2398256, EPI_ISL_2398258, EPI_ISL_2398274, EPI_ISL_2398276, EPI_ISL_2398281, EPI_ISL_2398318, EPI_ISL_2398319, EPI_ISL_2398324, EPI_ISL_2398325, EPI_ISL_2398334, EPI_ISL_2398338, EPI_ISL_2398343, EPI_ISL_2398351, EPI_ISL_2398352, EPI_ISL_2398355, EPI_ISL_2398364, EPI_ISL_2398372, EPI_ISL_2398373, EPI_ISL_2398376, EPI_ISL_2398386, EPI_ISL_2398401, EPI_ISL_2398451, EPI_ISL_2398458, EPI_ISL_2398459, EPI_ISL_2398476, EPI_ISL_2398479, EPI_ISL_2398481, EPI_ISL_2398482, EPI_ISL_2398483, EPI_ISL_2398491, EPI_ISL_2398502, EPI_ISL_2398503, EPI_ISL_2398508, EPI_ISL_2398513, EPI_ISL_2398520, EPI_ISL_2398526, EPI_ISL_2398530, EPI_ISL_2398531, EPI_ISL_2398567, EPI_ISL_2398632, EPI_ISL_2398765, EPI_ISL_2398769, EPI_ISL_2398801, EPI_ISL_2398803, EPI_ISL_2398804, EPI_ISL_2398842, EPI_ISL_2398845, EPI_ISL_2398884, EPI_ISL_2398902, EPI_ISL_2398917, EPI_ISL_2398922, EPI_ISL_2398928, EPI_ISL_2398935, EPI_ISL_2398936, EPI_ISL_2398941, EPI_ISL_2398948, EPI_ISL_2398957, EPI_ISL_2398969, EPI_ISL_2398979, EPI_ISL_2398981, EPI_ISL_2399009, EPI_ISL_2399041, EPI_ISL_2399050, EPI_ISL_2399052, EPI_ISL_2399065, EPI_ISL_2399149, EPI_ISL_2399155, EPI_ISL_2399166, EPI_ISL_2399193, EPI_ISL_2399204, EPI_ISL_2399233, EPI_ISL_2399234, EPI_ISL_2399254, EPI_ISL_2399255, EPI_ISL_2399265, EPI_ISL_2399267, EPI_ISL_2399268, EPI_ISL_2399272, EPI_ISL_2399290, EPI_ISL_2399302, EPI_ISL_2399336, EPI_ISL_2399338, EPI_ISL_2399351, EPI_ISL_2399355, EPI_ISL_2399367, EPI_ISL_2480199, EPI_ISL_2480638, EPI_ISL_2480639, EPI_ISL_2480646, EPI_ISL_2480702, EPI_ISL_2480790, EPI_ISL_2480791, EPI_ISL_2480987, EPI_ISL_2481019, EPI_ISL_2481039, EPI_ISL_2481044, EPI_ISL_2481058, EPI_ISL_2481084, EPI_ISL_2481110, EPI_ISL_2481130, EPI_ISL_2481143, EPI_ISL_2481144, EPI_ISL_2481514, EPI_ISL_2481521, EPI_ISL_2481597, EPI_ISL_2481875, EPI_ISL_2611267, EPI_ISL_2611268, EPI_ISL_2611271, EPI_ISL_2611272, EPI_ISL_2611280 | | | |
| see above | Laboratory Corporation of America | Centers for Disease Control and Prevention Division of Viral Diseases, Pathogen Discovery | Adrian Paskey; Amanda Douglas; Amanda Suchanek; Andrea Throop; Ayla Burns; Benjamin Rambo-Martin; Bobbi Croy; Brian Krueger; Brian Norvell; Christopher Gulvick; Christos Petropoulos; Clinton R. Paden; Craig Lukasik; Dakota Howard; Darlene Wagner; Debbie Boles; Dhwani Batra; Duncan MacCannell; Eyad Almasri; Goran Stevovic; Howard Engler; Hrushikesh Deshmukh; Jake Humphrey; Jana Schroth; Jason Caravas; Joe Voshell; John Pruitt; Jonathan Meltzer; Jonathan Williams; Kara Moser; Kimberly Wagner; Lax Iyer; Lisa Pfefferle; Lyndon Tilson; Manoj Jain; Marcia Eisenberg; Mary Ann Cristobal; Mary Williamson; Matthew Robinson; Matthew Schmerer; Michael Levandoski; Mike Sapeta; Mindy Nye; Minoo Agarwal; Mohan Kolli; Nuthawin Charoensri; Oren Cohen; Peter W. Cook; Prashant Gupta; Qian Zeng; Rama Ghatti; Scott Parker; Scott Ryan; Scott Sammons; Shatavia Morrison; Stanley Letovsky; Steven Ragan; Suresh Babu Selvaraju; Susan Countryman; Susan Hicks; Suzanne Dale; Thomas Urban; Tim Kuphal; Tricia Zwiefelhofer; Vincent Drouillon; Yvette Unoarumhi |
| EPI_ISL_2835679 | Laboratory Medicine, University of Washington | Laboratory Medicine, University of Washington | Castor; J.L. |
| EPI_ISL_2157545, EPI_ISL_2196251, EPI_ISL_2443601, EPI_ISL_2443605, EPI_ISL_2443606, EPI_ISL_2443607, EPI_ISL_2443610, EPI_ISL_2443611, EPI_ISL_2443613, EPI_ISL_2443634, EPI_ISL_2443659, EPI_ISL_2443660, EPI_ISL_2443661, EPI_ISL_2443662, EPI_ISL_2557403, EPI_ISL_3045381, EPI_ISL_3045471, EPI_ISL_3434727, EPI_ISL_3434728, EPI_ISL_3539937 | | | |
| see above | Laboratory of Respiratory Viruses and Measles, Oswaldo Cruz Institute, FIOCRUZ | Laboratory of Respiratory Viruses and Measles, Oswaldo Cruz Institute, FIOCRUZ | Agatha Cristinne Prudencio; Agatha Cristinne Prudencio Soares; Alice Sampaio Rocha; Ana Carolina Mendonca; Andrea Cony Cavalcanti; Anna Carolina Paixao; Elisa Cavalcante Pereira; Fernando Motta; Ighor Leonardo Arantes Gomes; Luciana Appolinario; Marilda Siqueira on behalf of the Fiocruz COVID-19 Genomic Surveillance Network; Paola Resende; Renata Serrano Lopes; Taina Venas |
| EPI_ISL_2696345, EPI_ISL_2696352, EPI_ISL_2696373, EPI_ISL_2696378, EPI_ISL_2696394, EPI_ISL_2696396, EPI_ISL_2696397, EPI_ISL_2696417, EPI_ISL_2696466, EPI_ISL_2696555, EPI_ISL_2696567, EPI_ISL_2696612, EPI_ISL_2696651, EPI_ISL_2696654, EPI_ISL_2696754, EPI_ISL_2696783, EPI_ISL_2696815, EPI_ISL_2696877, EPI_ISL_3025502, EPI_ISL_3025522, EPI_ISL_3025524, EPI_ISL_3025584 | | | |
| see above | Laboratory, Gravity Diagnostics | Laboratory, Gravity Diagnostics | Diagnostics, G. |
| EPI_ISL_2157542, EPI_ISL_2157543, EPI_ISL_2157544 | Laboratório Central de Saude Publica do Estado da Bahia (LACEN/BA) | Laboratory of Respiratory Viruses and Measles, Oswaldo Cruz Institute, FIOCRUZ | Alice Sampaio Rocha; Ana Carolina Mendonca; Anna Carolina Paixao; Elisa Cavalcante Pereira; Felicidade Pereira; Fernando Motta; Luciana Appolinario; Marilda Siqueira on behalf of the Fiocruz COVID-19 Genomic Surveillance Network; Paola Resende; Renata Serrano Lopes; Taina Venas |
| EPI_ISL_2777242, EPI_ISL_2777243, EPI_ISL_2777244, EPI_ISL_2777245, EPI_ISL_2777246, EPI_ISL_2777280, EPI_ISL_2777281, EPI_ISL_2777282, EPI_ISL_2777283, EPI_ISL_2777284, EPI_ISL_2777285, EPI_ISL_2777292, EPI_ISL_2777295, EPI_ISL_2777297, EPI_ISL_2777299, EPI_ISL_2777313, EPI_ISL_2777318, EPI_ISL_2777334, EPI_ISL_2777345, EPI_ISL_2777346, EPI_ISL_2777347, EPI_ISL_2777348, EPI_ISL_2777511, EPI_ISL_2777513, EPI_ISL_2777515, EPI_ISL_2777517, EPI_ISL_2777524, EPI_ISL_2777527, EPI_ISL_2777530, EPI_ISL_2777532, EPI_ISL_2777545, EPI_ISL_2777546, EPI_ISL_2777553, EPI_ISL_2777556, EPI_ISL_2777559, EPI_ISL_2777561, EPI_ISL_2777562, EPI_ISL_2777570, EPI_ISL_2777573, EPI_ISL_2777575, EPI_ISL_2777584, EPI_ISL_2777585, EPI_ISL_2777586, EPI_ISL_2777591, EPI_ISL_2777593, EPI_ISL_2777600, EPI_ISL_2777601, EPI_ISL_2777603, EPI_ISL_2777604, EPI_ISL_2777605, EPI_ISL_2777606, EPI_ISL_2777612, EPI_ISL_2777622, EPI_ISL_2777624, EPI_ISL_2777627, EPI_ISL_2777628, EPI_ISL_2777630, EPI_ISL_2777632, EPI_ISL_2777634, EPI_ISL_2777635, EPI_ISL_2777636, EPI_ISL_2777637, EPI_ISL_2777639, EPI_ISL_2777641, EPI_ISL_2777642, EPI_ISL_2777643, EPI_ISL_2777645, EPI_ISL_2777654, EPI_ISL_2777659, EPI_ISL_2777675, EPI_ISL_2777683, EPI_ISL_2777686, EPI_ISL_2777687, EPI_ISL_2777693, EPI_ISL_2777702, EPI_ISL_2777703, EPI_ISL_2777711, EPI_ISL_2777757, EPI_ISL_2777764, EPI_ISL_2777765, EPI_ISL_2777833, EPI_ISL_2777834, EPI_ISL_2777837, EPI_ISL_2777839, EPI_ISL_2777844, EPI_ISL_2777845, EPI_ISL_2777849, EPI_ISL_2777850, EPI_ISL_2777875, EPI_ISL_2777876, EPI_ISL_2777877, EPI_ISL_2777878, EPI_ISL_2777879, EPI_ISL_2777880, EPI_ISL_2777881, EPI_ISL_2777882, EPI_ISL_2777883, EPI_ISL_2777884, EPI_ISL_2777885, EPI_ISL_2777886, EPI_ISL_2777888, EPI_ISL_2777892, EPI_ISL_2777900, EPI_ISL_2777903, EPI_ISL_2777911, EPI_ISL_2777913, EPI_ISL_2777915, EPI_ISL_2777916, EPI_ISL_2777917, EPI_ISL_2777919, EPI_ISL_2777922, EPI_ISL_2777923, EPI_ISL_2777932, EPI_ISL_2777934, EPI_ISL_2777935, EPI_ISL_2777936, EPI_ISL_2777938, EPI_ISL_2777939, EPI_ISL_2777940, EPI_ISL_2777950, EPI_ISL_2777952, EPI_ISL_2777954, EPI_ISL_2777955, EPI_ISL_2777962, EPI_ISL_2777963, EPI_ISL_2777964, EPI_ISL_2777965, EPI_ISL_2777980, EPI_ISL_2777989, EPI_ISL_2777990, EPI_ISL_2777994, EPI_ISL_2778006, EPI_ISL_2778007 | | | |
| see above | Laboratório Central de Saúde Pública do Amazonas - LACEN-AM | Laboratorio de Ecologia de Doencas Transmissiveis na Amazonia, Instituto Leonidas e Maria Deane - Fiocruz Amazonia | André Corado; Debora Duarte; Felipe Naveca; Fernanda Nascimento; George Silva; Karina Pessoa; Luciana Gonçalves; Maria Júlia Brandão; Matilde Mejía; Michele Jesus; Valdinete Nascimento; Victor Souza; Ágatha Costa |
| EPI_ISL_2466190, EPI_ISL_2466191, EPI_ISL_2466192, EPI_ISL_2466193, EPI_ISL_2466194, EPI_ISL_2466195, EPI_ISL_2466196, EPI_ISL_2466197, EPI_ISL_2466198, EPI_ISL_2466199 | | | |
| see above | Laboratório de Biologia Molecular de Doenças Infecciosas e do Câncer (LADIC - UFRN) | Laboratory of Respiratory Viruses and Measles, Oswaldo Cruz Institute, FIOCRUZ | Alice Sampaio Rocha; Ana Carolina Mendonca; Anna Carolina Paixao; Elisa Cavalcante Pereira; Fernando Motta; Josélio Araújo; Luciana Appolinario; Marilda Siqueira on behalf of the Fiocruz COVID-19 Genomic Surveillance Network; Paola Resende; Renata Serrano Lopes; Taina Venas |
| EPI_ISL_2731508, EPI_ISL_2731509, EPI_ISL_2731513, EPI_ISL_2731514, EPI_ISL_2731516, EPI_ISL_2731517, EPI_ISL_2731519, EPI_ISL_2731520, EPI_ISL_2731521, EPI_ISL_2731524, EPI_ISL_2731526, EPI_ISL_2731527, EPI_ISL_2731528, EPI_ISL_2731529, EPI_ISL_2731530, EPI_ISL_2731531, EPI_ISL_2731532, EPI_ISL_2731533, EPI_ISL_2731536, EPI_ISL_2731538, EPI_ISL_2731540, EPI_ISL_2731542, EPI_ISL_2731544, EPI_ISL_2731545, EPI_ISL_2731546, EPI_ISL_2731548, EPI_ISL_2731549, EPI_ISL_2731551, EPI_ISL_2731553, EPI_ISL_2731554, EPI_ISL_2731555, EPI_ISL_2731556, EPI_ISL_2731557, EPI_ISL_2731558, EPI_ISL_2731559, EPI_ISL_2731567, EPI_ISL_2731568, EPI_ISL_2731569, EPI_ISL_2731575, EPI_ISL_2731578, EPI_ISL_2731580, EPI_ISL_2731581, EPI_ISL_2731582, EPI_ISL_2731584, EPI_ISL_2731585, EPI_ISL_2731586, EPI_ISL_2731588, EPI_ISL_2731589, EPI_ISL_2731590, EPI_ISL_2731591, EPI_ISL_2731593, EPI_ISL_2731595, EPI_ISL_2731598, EPI_ISL_2731599, EPI_ISL_2731600, EPI_ISL_2731601, EPI_ISL_2731602, EPI_ISL_2731603, EPI_ISL_2731604, EPI_ISL_2731610, EPI_ISL_2731611, EPI_ISL_2731612, EPI_ISL_2731613, EPI_ISL_2731614, EPI_ISL_2731615, EPI_ISL_2731616, EPI_ISL_2731617, EPI_ISL_2731618, EPI_ISL_2731620, EPI_ISL_2731621, EPI_ISL_2731622, EPI_ISL_2731632, EPI_ISL_2731634, EPI_ISL_2731640, EPI_ISL_2731642, EPI_ISL_2731643, EPI_ISL_2731644, EPI_ISL_2731645, EPI_ISL_2731646, EPI_ISL_2731649, EPI_ISL_2731650, EPI_ISL_2731651, EPI_ISL_2731654, EPI_ISL_2731655, EPI_ISL_2731656, EPI_ISL_2731660, EPI_ISL_2731663, EPI_ISL_2731664, EPI_ISL_2731665, EPI_ISL_2731667, EPI_ISL_2731668, EPI_ISL_2731669, EPI_ISL_2731671, EPI_ISL_2731672, EPI_ISL_2731673, EPI_ISL_2731675, EPI_ISL_2731676, EPI_ISL_2731677, EPI_ISL_2790137, EPI_ISL_2790139, EPI_ISL_2790140, EPI_ISL_2790144, EPI_ISL_2790145, EPI_ISL_2790146, EPI_ISL_2790147, EPI_ISL_2790148, EPI_ISL_2790149, EPI_ISL_2790150, EPI_ISL_2790151, EPI_ISL_2790152, EPI_ISL_2790153, EPI_ISL_2790154, EPI_ISL_2790155, EPI_ISL_2790156, EPI_ISL_2790157, EPI_ISL_2790158, EPI_ISL_2790159, EPI_ISL_2790165, EPI_ISL_2790166, EPI_ISL_2790168, EPI_ISL_2790169, EPI_ISL_2790171, EPI_ISL_2790172, EPI_ISL_2790173, EPI_ISL_2790174, EPI_ISL_2835107, EPI_ISL_2835108, EPI_ISL_2835110, EPI_ISL_2835111, EPI_ISL_2835112, EPI_ISL_2835113, EPI_ISL_2835116, EPI_ISL_2835118, EPI_ISL_2835119, EPI_ISL_2835120, EPI_ISL_2835124, EPI_ISL_2835125, EPI_ISL_2835126, EPI_ISL_2835129, EPI_ISL_2835130, EPI_ISL_2835132, EPI_ISL_2835135, EPI_ISL_2835141, EPI_ISL_2835145, EPI_ISL_2835146, EPI_ISL_2835147, EPI_ISL_2835151, EPI_ISL_2835152, EPI_ISL_2835153, EPI_ISL_2835154, EPI_ISL_2835155, EPI_ISL_2835156, EPI_ISL_2835157, EPI_ISL_2835159, EPI_ISL_2835160, EPI_ISL_2835161, EPI_ISL_2835162, EPI_ISL_2835163, EPI_ISL_2835164, EPI_ISL_2835165, EPI_ISL_2835167, EPI_ISL_2835171, EPI_ISL_2835172, EPI_ISL_2835174, EPI_ISL_2835175, EPI_ISL_2835176, EPI_ISL_2835177, EPI_ISL_2835178, EPI_ISL_2835179, EPI_ISL_2835180, EPI_ISL_2835181, EPI_ISL_2835182, EPI_ISL_2835183, EPI_ISL_2835184, EPI_ISL_2835188, EPI_ISL_2835189, EPI_ISL_2835191, EPI_ISL_2835192, EPI_ISL_2835193, EPI_ISL_2835194, EPI_ISL_2835196, EPI_ISL_2835198, EPI_ISL_2835202, EPI_ISL_2835203, EPI_ISL_2835207, EPI_ISL_2835208, EPI_ISL_2835214, EPI_ISL_2983471 | | | |
| see above | Laboratório de Biotecnologia Aplicada (LBA) - Laboratório de Biologia Molecular - Hospital das Clínicas, Faculdade de | Laboratory of Respiratory Viruses and Measles, Oswaldo Cruz Institute, FIOCRUZ | Alice Sampaio Rocha; Ana Carolina Mendonca; Anna Carolina Paixao; Elisa Cavalcante Pereira; Felipe Allan da Silva Costa; Fernando Motta; Jayme Augusto de Souza Neto; Leonardo Nazario de Moraes; Luciana Appolinario; Marilda Siqueira on behalf of the Fiocruz COVID-19 Genomic Surveillance Network; Paola Resende; Patricia Akemi Assato; Rejane Maria Tommasini; Renata Serrano Lopes; Taina Venas |

| | | | |
|---|---|---|---|
| | Medicina de Botucatu, Departamento de Bioprocessos e Biotecnologia - Faculdade de Ciências Agronômicas, UNESP – Botucatu/SP | | |
| EPI_ISL_2375845, EPI_ISL_2382535, EPI_ISL_2382536, EPI_ISL_3873650, EPI_ISL_3873656, EPI_ISL_3873658, EPI_ISL_3873660, EPI_ISL_4051953, EPI_ISL_4051955 | | | |
| see above | Laboratório de Microbiologia Molecular - Universidade FEEVALE | Molecular Microbiology Laboratory | Alana Witt Hansen; Fernando Rosado Spilki; Flávio Silveira; Fágner Henrique Heldt; Juliana Schons Gularte; Juliane Deise Fleck; Mariana Soares da Silva; Matheus Nunes Weber; Meriane Demoliner; Micheli Filippi; Micheli Filippi.; Paula Rodrigues de Almeida; Vyctoria Malayhka de Abreu Góes Pereira. |
| EPI_ISL_2629833, EPI_ISL_2629834, EPI_ISL_2629835, EPI_ISL_2629838 | Laboratório de Virologia Molecular - Universidade Federal do Rio de Janeiro | Laboratório de Virologia Molecular - Universidade Federal do Rio de Janeiro | ; Alice Laschuk Herlinger; Amilcar Tanuri; André Felipe Andrade dos Santos; Carolina Moreira Voloch; Cássia Cristina Alves Gonçalves; Diana Mariani; Débora Souza Faffe; Filipe Romero Rebello Moreira; Francine Bittencourt Schiffler; Isabela de Carvalho Leitão; Marcelo Calado de Paula Tôrres; Matheus Augusto Calvano Cosentino; Mirela D'arc; Orlando da Costa Ferreira Junior; Rafael Mello Galliez; Raíssa Mirella dos Santos Cunha da Costa; Renato Santana de Aguiar; Terezinha Marta Pereira Pinto Castineiras; Thamiris dos Santos Miranda; Átila Duque Rossi |
| EPI_ISL_2112830 | Labormedizin Darmstadt | Robert Koch Institute | |
| EPI_ISL_2443529, EPI_ISL_2443675, EPI_ISL_2443676, EPI_ISL_2443678, EPI_ISL_2443679, EPI_ISL_2443680, EPI_ISL_2443681, EPI_ISL_2443682, EPI_ISL_2443683, EPI_ISL_2443708, EPI_ISL_2443710, EPI_ISL_2466260, EPI_ISL_2536227, EPI_ISL_2536229, EPI_ISL_2536230, EPI_ISL_2536231, EPI_ISL_2536232, EPI_ISL_2536233, EPI_ISL_2536234, EPI_ISL_2536235, EPI_ISL_2536236, EPI_ISL_2536237, EPI_ISL_2536238, EPI_ISL_2536239, EPI_ISL_2536240, EPI_ISL_2536241, EPI_ISL_2536242, EPI_ISL_2536243, EPI_ISL_2536244, EPI_ISL_2536245, EPI_ISL_2536246, EPI_ISL_2536247, EPI_ISL_2536248, EPI_ISL_2536249, EPI_ISL_2536250, EPI_ISL_2536251, EPI_ISL_2536252, EPI_ISL_2536253, EPI_ISL_2536254, EPI_ISL_2536255, EPI_ISL_2536256, EPI_ISL_2536257, EPI_ISL_2536258, EPI_ISL_2536259, EPI_ISL_2536260, EPI_ISL_2536261, EPI_ISL_2536263, EPI_ISL_2557414, EPI_ISL_2557416, EPI_ISL_2557417, EPI_ISL_2557418, EPI_ISL_2557419, EPI_ISL_2557420, EPI_ISL_2557421, EPI_ISL_3061899 | | | |
| see above | Labortorio Central de Saude Publica do Estado de Santa Catarina (LACEN/SC) | Laboratory of Respiratory Viruses and Measles, Oswaldo Cruz Institute, FIOCRUZ | Alice Sampaio Rocha; Ana Carolina Mendonca; Anna Carolina Paixao; Darcita Buerger Rovaris; Elisa Cavalcante Pereira; Fernando Motta; Luciana Appolinario; Marilda Siqueira on behalf of the Fiocruz COVID-19 Genomic Surveillance Network; Paola Resende; Renata Serrano Lopes; Sandra Bianchini Fernandes; Taina Venas |
| EPI_ISL_2196333, EPI_ISL_2196334, EPI_ISL_2274098, EPI_ISL_2443639, EPI_ISL_2443640, EPI_ISL_2443641, EPI_ISL_2443642, EPI_ISL_2443643, EPI_ISL_2443644, EPI_ISL_2443645, EPI_ISL_2443646, EPI_ISL_2443647, EPI_ISL_2443648, EPI_ISL_2443649, EPI_ISL_2443650, EPI_ISL_2443651, EPI_ISL_2443652, EPI_ISL_2443653, EPI_ISL_2443654, EPI_ISL_2443655, EPI_ISL_2443656, EPI_ISL_2443657, EPI_ISL_2443658, EPI_ISL_2466230, EPI_ISL_2536275, EPI_ISL_2536276, EPI_ISL_2536277, EPI_ISL_2536278, EPI_ISL_2536279, EPI_ISL_2536281, EPI_ISL_2536282, EPI_ISL_2536283, EPI_ISL_2536284, EPI_ISL_2536285, EPI_ISL_2536286, EPI_ISL_2536287, EPI_ISL_2536288, EPI_ISL_2536289, EPI_ISL_2536290, EPI_ISL_2536291, EPI_ISL_2536292, EPI_ISL_2536293, EPI_ISL_2536294, EPI_ISL_2536295, EPI_ISL_2536296, EPI_ISL_2536297, EPI_ISL_2536298, EPI_ISL_2536299, EPI_ISL_2536300, EPI_ISL_2536301, EPI_ISL_2536302, EPI_ISL_2557376, EPI_ISL_2557377, EPI_ISL_2557378, EPI_ISL_2557379, EPI_ISL_2557380, EPI_ISL_2557381, EPI_ISL_2557382, EPI_ISL_2557383, EPI_ISL_2557384, EPI_ISL_2557404, EPI_ISL_2557406, EPI_ISL_3061894, EPI_ISL_3061895, EPI_ISL_3061897 | | | |
| see above | Labortorio Central de Saude Publica do Estado do Rio de Janeiro (LACEN/RJ) | Laboratory of Respiratory Viruses and Measles, Oswaldo Cruz Institute, FIOCRUZ | Alice Sampaio Rocha; Ana Carolina Mendonca; Andrea Cony Cavalcanti; Anna Carolina Paixao; Elisa Cavalcante Pereira; Fernando Motta; Luciana Appolinario; Marilda Siqueira on behalf of the Fiocruz COVID-19 Genomic Surveillance Network; Paola Resende; Renata Serrano Lopes; Taina Venas |
| EPI_ISL_2645510 | Labortorio Central de Saude Publica do Estado do Tocantins (LACEN/TO) | Laboratory of Respiratory Viruses and Measles, Oswaldo Cruz Institute, FIOCRUZ | Alice Sampaio Rocha; Ana Carolina Mendonca; Anna Carolina Paixao; Elisa Cavalcante Pereira; Fernando Motta; Jucimaria Dantas Galvao; Luciana Appolinario; Marilda Siqueira on behalf of the Fiocruz COVID-19 Genomic Surveillance Network; Paola Resende; Renata Serrano Lopes; Taina Venas |
| EPI_ISL_2443528, EPI_ISL_2443543, EPI_ISL_2443544, EPI_ISL_2443547, EPI_ISL_2443548, EPI_ISL_2443549, EPI_ISL_2443550, EPI_ISL_2443554, EPI_ISL_2443573, EPI_ISL_2443574, EPI_ISL_2443575, EPI_ISL_2443576, EPI_ISL_2557359, EPI_ISL_2557361, EPI_ISL_2557363, EPI_ISL_2557364, EPI_ISL_2557365, EPI_ISL_2557366, EPI_ISL_2557367, EPI_ISL_2557368, EPI_ISL_2557369, EPI_ISL_2557370, EPI_ISL_2557371, EPI_ISL_2557372, EPI_ISL_2557373, EPI_ISL_2557375, EPI_ISL_2603430, EPI_ISL_2603439, EPI_ISL_2603454, EPI_ISL_2603455, EPI_ISL_2603460, EPI_ISL_2603461, EPI_ISL_2603464, EPI_ISL_2603466, EPI_ISL_2603470, EPI_ISL_2645440, EPI_ISL_2645441, EPI_ISL_2645442, EPI_ISL_2645443, EPI_ISL_2863682, EPI_ISL_2863684, EPI_ISL_2863685, EPI_ISL_2863687, EPI_ISL_2863725, EPI_ISL_2863795, EPI_ISL_2863855, EPI_ISL_2863856, EPI_ISL_2863860, EPI_ISL_2863861, EPI_ISL_2863862, EPI_ISL_2863863, EPI_ISL_2863864, EPI_ISL_2863865, EPI_ISL_2863866, EPI_ISL_2863867, EPI_ISL_2863868, EPI_ISL_2863869, EPI_ISL_2863870, EPI_ISL_2863871, EPI_ISL_2863872, EPI_ISL_2863873, EPI_ISL_2863874, EPI_ISL_2863875, EPI_ISL_2863876, EPI_ISL_2863877, EPI_ISL_2863878, EPI_ISL_2863880, EPI_ISL_2863881, EPI_ISL_2863882, EPI_ISL_2863883, EPI_ISL_2863884, EPI_ISL_2863885, EPI_ISL_2863886, EPI_ISL_2863887, EPI_ISL_2863888, EPI_ISL_2863889, EPI_ISL_2863895, EPI_ISL_2863896 | | | |
| see above | Lboratorio Central de Saude Publica do Estado do Parana (LACEN/PR) | Laboratory of Respiratory Viruses and Measles, Oswaldo Cruz Institute, FIOCRUZ | Agatha Cristinne Prudencio; Agatha Cristinne Prudencio Soares; Alice Sampaio Rocha; Ana Carolina Mendonca; Anna Carolina Paixao; Elisa Cavalcante Pereira; Fernando Motta; Ighor Leonardo Arantes Gomes; Irina Riediger; Luciana Appolinario; Marilda Siqueira on behalf of the Fiocruz COVID-19 Genomic Surveillance Network; Paola Resende; Renata Serrano Lopes; Taina Venas |
| EPI_ISL_2692136, EPI_ISL_2692138, EPI_ISL_2692139, EPI_ISL_2692140, EPI_ISL_2692141 | Lifescan | RIPHL at Rush University Medical Center | Chicago Department of Public Health; Felix Araujo Perez; Kevin Kunstman; Laura Furtado; Marieta Hyde; Max Kolton; Stefan Green |
| EPI_ISL_2117614, EPI_ISL_2117967, EPI_ISL_2118029 | Lighthouse Lab in Alderley Park | Wellcome Sanger Institute for the COVID-19 Genomics UK (COG-UK) Consortium | Cordelia Langford; David K. Jackson; Dominic Kwiatkowski; Ewan Harrison; Ian Johnston; Jacquelyn Wynn; Jeffrey Barrett; John Sillitoe on behalf of the Wellcome Sanger Institute COVID-19 Surveillance Team; Mairead Hyland; Roberto Amato; Sonia Goncalves; The Lighthouse Lab in Alderley Park and Alex Alderton |
| EPI_ISL_1740541, EPI_ISL_1740689 | Lighthouse Lab in Cambridge | Wellcome Sanger Institute for the COVID-19 Genomics UK (COG-UK) Consortium | Cordelia Langford; David K. Jackson; Dominic Kwiatkowski; Ewan Harrison; Ian Johnston; Jeffrey Barrett; John Sillitoe on behalf of the Wellcome Sanger Institute COVID-19 Surveillance Team; Rob Howes; Roberto Amato; Sonia Goncalves; The Lighthouse Lab in Cambridge and Alex Alderton |
| EPI_ISL_1921451, EPI_ISL_1921677, EPI_ISL_2004797, EPI_ISL_2021062, EPI_ISL_2021666, EPI_ISL_2022037, EPI_ISL_2117733, EPI_ISL_2117840, EPI_ISL_2118938, EPI_ISL_2119812, EPI_ISL_2172155, EPI_ISL_2172194, EPI_ISL_2316530 | | | |
| see above | Lighthouse Lab in Milton Keynes | Wellcome Sanger Institute for the COVID-19 Genomics UK (COG-UK) Consortium | Cordelia Langford; David K. Jackson; Dominic Kwiatkowski; Ewan Harrison; Ian Johnston; Jeffrey Barrett; John Sillitoe on behalf of the Wellcome Sanger Institute COVID-19 Surveillance Team; Roberto Amato; Sonia Goncalves; The Lighthouse Lab in Milton Keynes and Alex Alderton |
| EPI_ISL_2266523 | Limbach - MVZ Humangenetik Ulm | Robert Koch Institute | |
| EPI_ISL_2373062, EPI_ISL_2373069, EPI_ISL_2373076, EPI_ISL_2373094, EPI_ISL_2373100, EPI_ISL_2373103, EPI_ISL_2373104, EPI_ISL_2373107, EPI_ISL_2842585, EPI_ISL_2842589 | | | |
| see above | Lurie Children's Hospital of Chicago | Northwestern University - Ozer Lab | Egon A. Ozer; Judd F. Hultquist; Lacy M. Simons; Larry K. Kociolek; Michael G. Ison; Ramon Lorenzo-Redondo; Taylor J. Dean; William J. Muller; Xiaotian; Zheng |
| EPI_ISL_2249573, EPI_ISL_2249576 | M Health Fairview | Minnesota Department of Health, Public Health Laboratory | Alexandra Lorentz; Jacob Garfin; Matt Plumb; and Xiong Wang |
| EPI_ISL_2451813 | MA State Public Health Laboratory | Centers for Disease Control and Prevention Division of Viral Diseases, Pathogen Discovery | Alison Laufer Halpin; Ben L. Rambo-Martin; Clinton R. Paden; Dakota Howard; Darlene Wagner; Dave Wentworth; Dhwani Batra; Jasmine Padilla; Justin Lee; Katie Dillon; Krista Queen; Kristen Knipe; Kristine Lacek; Mark Burroughs; Matthew Schmerer; Mili Sheth; Peter Cook; Sam Shepard; Sarah Nobles; Shoshona Le; Suxiang Tong; Vivien Dugan; Yvette Unoarumhi |
| EPI_ISL_2801332, EPI_ISL_2801333 | MATERNIDADE ESCOLA ASSIS CHATEAUBRIAND | Analytical Competence Molecular Epidemiology Lab/ACME, Oswaldo Cruz Foundation, Ceara (FIOCRUZ CE) | Cleber Furtado Aksenen e Suzana Porto Almeida; Fabio Miyajima; Fernando Braga Stehling; Francisco Eder de Moura Lopes; Jamille Maria Mendes Bezerra; Joaquim César do Nascimento Sousa Junior; Pedro Miguel Carneiro Jeronimo; Thais Ferreira de Oliveira; Thais de Oliveira Costa; Ticiane Cavalcante de Souza; Veridiana Pessoa Miyajima |
| EPI_ISL_1811581, EPI_ISL_2227429 | MD PHL | MD PHL | Maryland Department of Health Laboratories Administration |
| EPI_ISL_2383931 | ME Health and Environmental Testing Laboratory | Centers for Disease Control and Prevention Division of Viral Diseases, Pathogen Discovery | Alison Laufer Halpin; Ben L. Rambo-Martin; Clinton R. Paden; Dakota Howard; Darlene Wagner; Dave Wentworth; Dhwani Batra; Jasmine Padilla; Justin Lee; Katie Dillon; Krista Queen; Kristen Knipe; Kristine Lacek; Mark Burroughs; Matthew Schmerer; Mili Sheth; Peter Cook; Sam Shepard; Sarah Nobles; Shoshona Le; Suxiang Tong; Vivien Dugan; Yvette Unoarumhi |
| EPI_ISL_2453794, EPI_ISL_2453796, EPI_ISL_2453806, EPI_ISL_2453816 | MEPHI, Aix Marseille University | MEPHI, Aix Marseille University | Anthony LEVASSEUR |
| EPI_ISL_2451618 | MI - Michigan Department of Health and Human Services - Bureau of Laboratories | Centers for Disease Control and Prevention Division of Viral Diseases, Pathogen Discovery | Alison Laufer Halpin; Ben L. Rambo-Martin; Clinton R. Paden; Dakota Howard; Darlene Wagner; Dave Wentworth; Dhwani Batra; Jasmine Padilla; Justin Lee; Katie Dillon; Krista Queen; Kristen Knipe; Kristine Lacek; Mark Burroughs; Matthew Schmerer; Mili Sheth; Peter Cook; Sam Shepard; Sarah Nobles; Shoshona Le; Suxiang Tong; Vivien Dugan; Yvette Unoarumhi |
| EPI_ISL_2229261, EPI_ISL_3354883 | MO State Public Health Laboratory | Centers for Disease Control and Prevention Division of Viral Diseases, Pathogen Discovery | Alex Burgin; Alison Laufer Halpin; Ben L. Rambo-Martin; Ben Rambo-Martin; Clinton Paden; Clinton R. Paden; Dakota Howard; Darlene Wagner; Dave Wentworth; Dhwani Batra; Jasmine Padilla; Justin Lee; Katie Dillon; Krista Queen; Kristen Knipe; Kristine Lacek; Mark Burroughs; Matthew Schmerer; Meghan Bentz; Mili Sheth; Peter Cook; Sam Shepard; Sarah Nobles; Shoshona Le; Suxiang Tong; Vivien Dugan; Yvette Unoarumhi |
| EPI_ISL_2444203, EPI_ISL_2444210 | MONTE APRAZIVEL | Instituto Butantan / FAMERP | Antonio Jorge Martins; Claudia Renata dos Santos Barros; David Schlesinger; Debora Botequio Moretti; Dimas Tadeu Covas; Elaine Cristina Marqueze; Elaine Vieira Santos; Evandra Strazza Rodrigues; Heidge Fukumasu; Jayme Augusto de Souza-Neto; José Salvatore Leister Patané; Luiz Alcantara; Luiz Lehmann Coutinho; Maria Carolina Elias; Maurício Lacerda Nogueira; Rafael dos Santos Bezerra; Raul Machado Neto; Rejane Maria Tommasini Grotto; Ricardo Haddad; Sandra Coccuzzo Sampaio Vessoni; Simone Kashima; Svetoslav Nanev Slavov; Vincent Louis Viala |
| EPI_ISL_2274608 | MONTEFIORE MEDICAL CENTER LABORATORIES | Wadsworth Center, New York State Department of Health | Alexis Russell; Catharine Prussing; Daryl M. Lamson; Erasmus Schneider; Erica Lasek-Nesselquist; John Kelly; Jonathan Plitnick; Kirsten St. George; Matthew Shudt; Melissa A Leisner; Navjot Singh |
| EPI_ISL_2451640 | MT Public Health Laboratory | Centers for Disease Control and Prevention Division of Viral Diseases, Pathogen Discovery | Alison Laufer Halpin; Ben L. Rambo-Martin; Clinton R. Paden; Dakota Howard; Darlene Wagner; Dave Wentworth; Dhwani Batra; Jasmine Padilla; Justin Lee; Katie Dillon; Krista Queen; Kristen Knipe; Kristine Lacek; Mark Burroughs; Matthew Schmerer; Mili Sheth; Peter Cook; Sam Shepard; Sarah Nobles; Shoshona Le; Suxiang Tong; Vivien Dugan; Yvette Unoarumhi |
| EPI_ISL_2111276 | MVZ Labor Dr. Fenner und Kollegen (Standort Hamburg) | Robert Koch Institute | |
| EPI_ISL_1849414, EPI_ISL_1849483 | MVZ Labor Krone GbR | Robert Koch Institute | |
| EPI_ISL_2314745 | MVZ für Laboratoriumsmedizin und Mikrobiologie Koblenz-Mittelrhein (Labor Koblenz) | Robert Koch Institute | |
| EPI_ISL_2101010, EPI_ISL_2101014, EPI_ISL_2501190 | Maine HETL | Health and Environmental Testing Laboratory | Barter, M.; Dewey, H.; Grieser, H.; H. and Tewhey, R.; Lynch, R.; Martha, J.; Matluk, N.; Meak, S.; Munger |
| EPI_ISL_1826331, EPI_ISL_1826332, EPI_ISL_2245512, EPI_ISL_2245527, EPI_ISL_2245568, EPI_ISL_2245596, EPI_ISL_2802401, EPI_ISL_2802402, EPI_ISL_2802436, EPI_ISL_2802525, EPI_ISL_2802526, EPI_ISL_2802527, EPI_ISL_2802528, EPI_ISL_2802529, EPI_ISL_2802530, EPI_ISL_2802531, EPI_ISL_2802532, EPI_ISL_2802691, EPI_ISL_3298815, EPI_ISL_3298842, EPI_ISL_3298847, EPI_ISL_3298856, | | | |

| | | | |
|---|---|---|---|
| EPI_ISL_3298857, EPI_ISL_3298874, EPI_ISL_3298878, EPI_ISL_3298879, EPI_ISL_3298890, EPI_ISL_4083775, EPI_ISL_4083810 | | | |
| see above | Maine Health and Environmental Testing Laboratory | Tewhey Lab, The Jackson Laboratory | Barter, M.; Dewey, H.; H. and Tewhey, R.; Iosue, F.; Lynch, R.; Matluk, N.; Munger |
| EPI_ISL_2285048, EPI_ISL_2307182, EPI_ISL_2307204, EPI_ISL_2307272, EPI_ISL_2307289, EPI_ISL_2441051, EPI_ISL_2450966, EPI_ISL_2450988, EPI_ISL_2451021, EPI_ISL_2451027, EPI_ISL_2451029, EPI_ISL_2451088, EPI_ISL_2451212, EPI_ISL_2488897, EPI_ISL_2488952, EPI_ISL_2488979, EPI_ISL_2489013, EPI_ISL_2489040, EPI_ISL_2489042, EPI_ISL_2525127, EPI_ISL_2525324, EPI_ISL_2525355, EPI_ISL_2646386, EPI_ISL_2646418, EPI_ISL_2646444, EPI_ISL_2646447, EPI_ISL_2646459, EPI_ISL_2646463, EPI_ISL_2646464, EPI_ISL_2646472, EPI_ISL_2646474, EPI_ISL_2646505, EPI_ISL_2646507, EPI_ISL_2646516, EPI_ISL_2646523, EPI_ISL_2646573, EPI_ISL_2646578, EPI_ISL_2646619, EPI_ISL_2646685, EPI_ISL_2646692, EPI_ISL_2646724, EPI_ISL_2646732, EPI_ISL_2646801, EPI_ISL_2646872, EPI_ISL_2646888, EPI_ISL_2646959, EPI_ISL_2647100, EPI_ISL_2647303, EPI_ISL_2647314, EPI_ISL_2647321, EPI_ISL_2647323, EPI_ISL_2647325, EPI_ISL_2647376, EPI_ISL_2647434, EPI_ISL_2647439, EPI_ISL_2647451, EPI_ISL_2647917, EPI_ISL_2647965, EPI_ISL_2786548 | | | |
| see above | Mako Medical | Centers for Disease Control and Prevention Division of Viral Diseases, Pathogen Discovery | Adrian Paskey; Benjamin Rambo-Martin; Christopher Gulvick; Clinton R. Paden; Dakota Howard; Darlene Wagner; Dhwani Batra; Duncan MacCannell; Jason Caravas; Kara Moser; Lauren Moon; Matthew Schmerer; Matthew Tugwell; Peter W. Cook; Scott Sammons; Shatavia Morrison; Yvette Unoarumhi |
| EPI_ISL_2501831, EPI_ISL_2598490, EPI_ISL_3011863 | Maryland Genomics, Institute for Genome Sciences, University of Maryland School of Medicine | Maryland Genomics, Institute for Genome Sciences, University of Maryland School of Medicine | Claire M; Fraser; Hazen; Holly; Humphrys; Ivette; Jacques; Jonathan; Kranthi; Lim; Lisa D; Luke J; Mike; Ott; Ravel; Roussey; Sadzewicz; Sandra; Santana-Cruz; Tallon; Tracy; Vavikolanu |
| EPI_ISL_2550760 | Maryland Public Health Laboratory (MD PHL) | Maryland Public Health Laboratory (MD PHL) | Maryland Department of Health Laboratories Administration |
| EPI_ISL_2016473, EPI_ISL_2016481, EPI_ISL_2016483, EPI_ISL_2362372 | Massachusetts General Hospital | Infectious Disease Program, Broad Institute of Harvard and MIT | A.E.; Adams, G.; Anahtar, M.; B.L.; B.W.; Bauer, M.; Birren; Branda, J.; Carter, A.; Cerrato, F.; Chaluvadi, S.; Chapman; Cusick, C.; D.J.; DeRuff, K.; E. and Sabeti; Flowers, K.; Gallagher, G.; Gladden-Young, A.; Gnirke, A.; Harris, J.; J.E.; K.J.; LaRocque, R.; Lagerborg, K.; Lemieux; Lin; Loreth, C.; MacInnis; Neumann, A.; Normandin, E.; P.C.; Park; Pierce, V.; Reilly, S.; Rosenberg; Rudy, M.; Ryan, E.; S.B.; Shaw, B.; Siddle; Slater, D.; Smole, S.; Tomkins-Tinch, C.; Turbett, S. |
| EPI_ISL_2008341, EPI_ISL_2008344, EPI_ISL_2229517, EPI_ISL_2229808, EPI_ISL_2400245, EPI_ISL_2483777, EPI_ISL_2483781, EPI_ISL_2483790, EPI_ISL_2483799, EPI_ISL_2483801, EPI_ISL_2483803, EPI_ISL_2484937 | | | |
| see above | Massachusetts State Public Health Laboratory | Massachusetts State Public Health Laboratory | Andrew Lang; Glen Gallagher; Sandra Smole; Timelia Fink |
| EPI_ISL_2136412 | Mayo Clinic & Mayo Clinic Laboratories | Minnesota Department of Health, Public Health Laboratory | Alexandra Lorentz; Jacob Garfin; Matt Plumb; and Xiong Wang |
| EPI_ISL_2389368 | Med. Labor Prof. Schenk Dr. Ansorge & Kollegen | Robert Koch Institute | |
| EPI_ISL_1999698, EPI_ISL_1999734, EPI_ISL_1999766, EPI_ISL_1999797, EPI_ISL_2433981, EPI_ISL_2434027, EPI_ISL_2434039 | | | |
| see above | Microbiologia CATLAB | Can Ruti SARS-CoV-2 Sequencing Hub (HUGTiP/IrsiCaixa/IGTP) | Alba Sánchez; Anna Not; Antoni E Bordoy; Bonaventura Clotet; Cristina Casañ; Cristina Esteban; Francesc Catala-Moll; Gemma Clara; Ignacio Blanco; Marc Noguera-Julian; Maria Casadellà; Mariona Parera; Mercedes Guerrero; Montserrat Giménez; Pere-Joan Cardona; Pilar Armengol; Roger Paredes; Verónica Saludes; and Elisa Martró on behalf of the Can Ruti SARS-CoV-2 Sequencing Hub. |
| EPI_ISL_1892991, EPI_ISL_2000122, EPI_ISL_2136876, EPI_ISL_2136877, EPI_ISL_2136900, EPI_ISL_2136912, EPI_ISL_2136924, EPI_ISL_2136947 | | | |
| see above | Microbiologia CATLAB | Can Ruti SARS-CoV-2 Sequencing Hub (HUGTiP/IrsiCaixa/IGTP) | Alba Sánchez; Anna Not; Antoni E Bordoy; Bonaventura Clotet; Cristina Casañ; Cristina Esteban; Francesc Catala-Moll; Gemma Clara; Ignacio Blanco; Marc Noguera-Julian; Maria Casadellà; Mariona Parera; Mercedes Guerrero; Montserrat Giménez; Pere-Joan Cardona; Pilar Armengol; Roger Paredes; Verónica Saludes; and Elisa Martró on behalf of the Can Ruti SARS-CoV-2 Sequencing Hub. |
| EPI_ISL_3138668 | Microbiology Department - University Hospital Brussel | Microbiology Department - University Hospital Brussel | Thomas Demuyser |
| EPI_ISL_2140625, EPI_ISL_2372690, EPI_ISL_2448032 | Microbiology Department, Laboratori Clínic Metropolitana Nord. Hospital Universitari Germans Trias i Pujol | Can Ruti SARS-CoV-2 Sequencing Hub (HUGTiP/IrsiCaixa/IGTP) | Alba Snchez; Alba Sánchez; Anna Not; Antoni E Bordoy; Bonaventura Clotet; Cristina Casa; Cristina Casañ; Cristina Esteban; Francesc Catala-Moll; Gemma Clara; Ignacio Blanco; Marc Noguera-Julian; Maria Casadell; Maria Casadellà; Mariona Parera; Mercedes Guerrero; Montserrat Gimnez; Montserrat Giménez; Pere-Joan Cardona; Pilar Armengol; Roger Pared; Roger Paredes; Vernica Saludes; Verónica Saludes; and Elisa Martr on behalf of the Can Ruti SARS-CoV-2 Sequencing Hub.; and Elisa Martró on behalf of the Can Ruti SARS-CoV-2 Sequencing Hub. |
| EPI_ISL_1831601, EPI_ISL_1831662, EPI_ISL_1892968, EPI_ISL_1893046, EPI_ISL_2380035, EPI_ISL_2380037, EPI_ISL_2380046 | | | |
| see above | Microbiology Department, Laboratori Clínic Metropolitana Nord. Hospital Universitari Germans Trias i Pujol. | Can Ruti SARS-CoV-2 Sequencing Hub (HUGTiP/IrsiCaixa/IGTP) | Alba Sánchez; Anna Not; Antoni E Bordoy; Bonaventura Clotet; Cristina Casañ; Cristina Esteban; Francesc Catala-Moll; Gemma Clara; Ignacio Blanco; Marc Noguera-Julian; Maria Casadellà; Mariona Parera; Mercedes Guerrero; Montserrat Giménez; Pere-Joan Cardona; Pilar Armengol; Roger Pared; Roger Paredes; Verónica Saludes; and Elisa Martró on behalf of the Can Ruti SARS-CoV-2 Sequencing Hub. |
| EPI_ISL_2036092, EPI_ISL_2325183, EPI_ISL_2502699 | Microbiology Department, University Hospital Donostia | Microbiology Department, University Hospital Donostia | Cilla G.; Gomez M; Grandioso D; Marimon JM; Martin-Peñaranda T; Montes M; Piñeiro L; Sorarrain A |
| EPI_ISL_2086155, EPI_ISL_2086156, EPI_ISL_2086157, EPI_ISL_2086158, EPI_ISL_2086159, EPI_ISL_2086163, EPI_ISL_2086164, EPI_ISL_2086165, EPI_ISL_2086171, EPI_ISL_2086172, EPI_ISL_2086173, EPI_ISL_2967793, EPI_ISL_2967838 | | | |
| see above | Microbiology Department. Complexo Hospitalario Universitario de Vigo | Microbiology Department. Complexo Hospitalario Universitario de Vigo | Alfaya N; Alonso I; Alvarez M; Cabrera JJ; Carballo R; Cores O; Cortizo S; Davina C; Martinez L; Mediero G; Perez S; Potel C; Regueiro B; Rey S; Vasallo FJ; del-Campo V |
| EPI_ISL_2312976 | Microbiology Division, SC DHEC | Microbiology Division, SC DHEC | Flores, H.; Freeman, J. |
| EPI_ISL_2029100, EPI_ISL_2029101, EPI_ISL_2259193 | Microvida | Microvida | Jaco Verweij; Joep Stohr; Joep Stöhr; Suzan D. Pas |
| EPI_ISL_2406491 | Middlemore Hospital | Institute of Environmental Science and Research (ESR) | Anja Werno; Antje van der Linden; Arlo Upton; Chris Mansell; David Hammer; Dragana Drinkovic; Erasmus Smit; Gary McAuliffe; Hana Sofia Andersson; Hermes Perez; James Ussher; Jill Sherwood; Jing Wang; Joep de Ligt; Josh Freeman; Julia Howard; Juliet Elvy; Lauren Jelly; Mary DeAlmeida; Matt Blakiston; Matt Storey; Matthew Rogers; Max Bloomfield; Michael Addidle; Michelle Balm; Muhammad Faisal; Nikki Freed; Olin Silander; Olivia Stroeven; Rachel Boyle; Sally Roberts; SallyAnn Harbison; Sarah Jefferies; Sharmini Muttaiyah; Susan Morpeth; Susan Taylor; Timothy Blackmore; Vani Sathyendran; Veronica Playle; Virginia Hope; Xiaoyun Ren |
| EPI_ISL_2092803, EPI_ISL_2136384, EPI_ISL_2136395, EPI_ISL_2136397, EPI_ISL_2136426, EPI_ISL_2178660, EPI_ISL_2244913, EPI_ISL_2249519, EPI_ISL_2249537, EPI_ISL_2249545, EPI_ISL_2249547, EPI_ISL_2249548, EPI_ISL_2325100, EPI_ISL_2332698, EPI_ISL_2403691, EPI_ISL_2425394, EPI_ISL_2658542 | | | |
| see above | Minnesota Department of Health, Public Health Laboratory | Minnesota Department of Health, Public Health Laboratory | Alexandra Lorentz; Jacob Garfin; Matt Plumb; and Xiong Wang |
| EPI_ISL_2035571, EPI_ISL_2035591, EPI_ISL_2158679, EPI_ISL_2158682, EPI_ISL_2158684, EPI_ISL_2158690, EPI_ISL_2304242, EPI_ISL_2304250, EPI_ISL_2304260, EPI_ISL_2501926 | | | |
| see above | Missouri State Public Health Laboratory | Missouri State Public Health Laboratory | Ashley New; Joshua Barry; Matthew Sinn |
| EPI_ISL_2105943, EPI_ISL_2105944, EPI_ISL_2105945, EPI_ISL_2105949, EPI_ISL_2105956, EPI_ISL_2107889 | Molecular Diagnostics Pathology Department Mater Dei Hospital Malta | Molecular Diagnostics Pathology Department Mater Dei Hospital Malta | C Cilia; G Zahra; L Grech; R Borg |
| EPI_ISL_2646995, EPI_ISL_2941746, EPI_ISL_2941753 | Montana Public Health Laboratory | Montana Public Health Laboratory | Carrie Biskupiak; Deborah Gibson; Joy Ritter; Michael Dills; Michelle Mozer |
| EPI_ISL_3355029 | ND Dept. of Health Laboratory Services-Microbiology | Centers for Disease Control and Prevention Division of Viral Diseases, Pathogen Discovery | Alex Burgin; Ben Rambo-Martin; Clinton Paden; Dakota Howard; Dave Wentworth; Dhwani Batra; Jasmine Padilla; Justin Lee; Krista Queen; Kristen Knipe; Kristine Lacek; Mark Burroughs; Matthew Schmerer; Meghan Bentz; Mili Sheth; Peter Cook; Sam Shepard; Sarah Nobles; Suxiang Tong; Vivien Dugan; Yvette Unoarumhi |
| EPI_ISL_2384008, EPI_ISL_3354969 | NH Dept. of Health and Human Services Public Health Labs | Centers for Disease Control and Prevention Division of Viral Diseases, Pathogen Discovery | Alex Burgin; Alison Laufer Halpin; Ben L. Rambo-Martin; Ben Rambo-Martin; Clinton Paden; Clinton R. Paden; Dakota Howard; Darlene Wagner; Dave Wentworth; Dhwani Batra; Jasmine Padilla; Justin Lee; Katie Dillon; Krista Queen; Kristen Knipe; Kristine Lacek; Mark Burroughs; Matthew Schmerer; Meghan Bentz; Mili Sheth; Peter Cook; Sam Shepard; Sarah Nobles; Shoshona Le; Suxiang Tong; Vivien Dugan; Yvette Unoarumhi |
| EPI_ISL_1969075, EPI_ISL_2185595, EPI_ISL_2281856, EPI_ISL_2557099 | NJDOH, Public Health and Environmental Laboratories | NJ_PHEL | Allison Roder; Byeong Jeong; Chelsea San Filippo; Dana Woell; Jacquelyn Deverell; Lindsey Bodnar; Matthew Scarnati; Ryan Pachucki; Shiv K. Verma |
| EPI_ISL_2097278, EPI_ISL_2097289 | NORTHWELL HEALTH LABORATORIES | Wadsworth Center, New York State Department of Health | Alexis Russell; Catharine Prussing; Daryl M. Lamson; Erasmus Schneider; Erica Lasek-Nesselquist; John Kelly; Jonathan Plitnick; Kirsten St. George; Matthew Shudt; Melissa A Leisner; Navjot Singh |
| EPI_ISL_2345165, EPI_ISL_2345174, EPI_ISL_2345176, EPI_ISL_2345177 | NUCLEO DE ESPECIALIDADES EM SAUDE | Instituto Butantan / ESALQ-Piracicaba | Antonio Jorge Martins; Claudia Renata dos Santos Barros; David Schlesinger; Debora Botequio Moretti; Dimas Tadeu Covas; Elaine Cristina Marqueze; Elaine Vieira Santos; Evandra Strazza Rodrigues; Heidge Fukumasu; Jayme Augusto de Souza-Neto; José Salvatore Leister Patané; Luiz Alcantara; Luiz Lehmann Coutinho; Maria Carolina Elias; Maurício Lacerda Nogueira; Rafael dos Santos Bezerra; Raul Machado Neto; Rejane Maria Tommasini Grotto; Ricardo Haddad; Sandra Coccuzzo Sampaio Vessoni; Simone Kashima; Svetoslav Nanev Slavov; Vincent Louis Viala |
| EPI_ISL_2345800 | NUCLEO DE SAUDE MARY DOTA DE BAURU | Instituto Butantan / Mendelics | Antonio Jorge Martins; Claudia Renata dos Santos Barros; David Schlesinger; Debora Botequio Moretti; Dimas Tadeu Covas; Elaine Cristina Marqueze; Elaine Vieira Santos; Evandra Strazza Rodrigues; Heidge Fukumasu; Jayme Augusto de Souza-Neto; José Salvatore Leister Patané; Luiz Alcantara; Luiz Lehmann Coutinho; Maria Carolina Elias; Maurício Lacerda Nogueira; Rafael dos Santos Bezerra; Raul Machado Neto; Rejane Maria Tommasini Grotto; Ricardo Haddad; Sandra Coccuzzo Sampaio Vessoni; Simone Kashima; Svetoslav Nanev Slavov; Vincent Louis Viala |
| EPI_ISL_2445138, EPI_ISL_2473751 | NUCLEO DE SAUDE VILA FALCAO DE BAURU | Instituto Butantan | Antonio Jorge Martins; Claudia Renata dos Santos Barros; David Schlesinger; Debora Botequio Moretti; Dimas Tadeu Covas; Elaine Cristina Marqueze; Elaine Vieira Santos; Evandra Strazza Rodrigues; Heidge Fukumasu; Jayme Augusto de Souza-Neto; José Salvatore Leister Patané; Luiz Alcantara; Luiz Lehmann Coutinho; Maria Carolina Elias; Maurício Lacerda Nogueira; Rafael dos Santos Bezerra; Raul Machado Neto; Rejane Maria Tommasini Grotto; Ricardo Haddad; Sandra Coccuzzo Sampaio Vessoni; Simone Kashima; Svetoslav Nanev Slavov; Vincent Louis Viala |
| EPI_ISL_2345215, EPI_ISL_2345216, | NUCLEO DE SAUDE VILA FALCAO DE BAURU | Instituto Butantan / ESALQ-Piracicaba | Antonio Jorge Martins; Claudia Renata dos Santos Barros; David Schlesinger; Debora Botequio Moretti; Dimas Tadeu Covas; Elaine Cristina Marqueze; Elaine Vieira Santos; Evandra Strazza Rodrigues; Heidge Fukumasu; Jayme Augusto de Souza-Neto; José Salvatore Leister Patané; Luiz Alcantara; Luiz Lehmann Coutinho; Maria Carolina Elias; Maurício Lacerda Nogueira; Rafael dos Santos Bezerra; Raul Machado Neto; Rejane Maria Tommasini Grotto; Ricardo Haddad; Sandra Coccuzzo Sampaio Vessoni; Simone Kashima; Svetoslav Nanev Slavov; Vincent Louis Viala |

| | | | |
|---|---|---|---|
| EPI_ISL_2345217, EPI_ISL_2345218, EPI_ISL_2345242, EPI_ISL_2345263 | | | |
| EPI_ISL_2345084, EPI_ISL_2345091, EPI_ISL_2345132, EPI_ISL_2345145, EPI_ISL_2345146, EPI_ISL_2345147, EPI_ISL_2345148, EPI_ISL_2345164 | | | |
| see above | NUCLEO SAUDE III N HABITACIONAL PRESIDENTE GEISEL BAURU | Instituto Butantan / ESALQ-Piracicaba | Antonio Jorge Martins; Claudia Renata dos Santos Barros; David Schlesinger; Debora Botequio Moretti; Dimas Tadeu Covas; Elaine Cristina Marqueze; Elaine Vieira Santos; Evandra Strazza Rodrigues; Heidge Fukumasu; Jayme Augusto de Souza-Neto; José Salvatore Leister Patané; Luiz Alcantara; Luiz Lehmann Coutinho; Maria Carolina Elias; Maurício Lacerda Nogueira; Rafael dos Santos Bezerra; Raul Machado Neto; Rejane Maria Tommasini Grotto; Ricardo Haddad; Sandra Coccuzzo Sampaio Vessoni; Simone Kashima; Svetoslav Nanev Slavov; Vincent Louis Viala |
| EPI_ISL_2451588 | NV-Southern Nevada Public Health Laboratory | Centers for Disease Control and Prevention Division of Viral Diseases, Pathogen Discovery | Alison Laufer Halpin; Ben L. Rambo-Martin; Clinton R. Paden; Dakota Howard; Darlene Wagner; Dave Wentworth; Dhwani Batra; Jasmine Padilla; Justin Lee; Katie Dillon; Krista Queen; Kristen Knipe; Kristine Lacek; Mark Burroughs; Matthew Schmerer; Mili Sheth; Peter Cook; Sam Shepard; Sarah Nobles; Shoshona Le; Suxiang Tong; Vivien Dugan; Yvette Unoarumhi |
| EPI_ISL_2098603, EPI_ISL_2098618, EPI_ISL_2274698 | NYC Pandemic Response Lab | Wadsworth Center, New York State Department of Health | Alexis Russell; Catharine Prussing; Daryl M. Lamson; Erasmus Schneider; Erica Lasek-Nesselquist; John Kelly; Jonathan Plitnick; Kirsten St. George; Matthew Shudt; Melissa A Leisner; Navjot Singh |
| EPI_ISL_2018244 | NYSDOH Wadsworth Center, Virology Lab | Centers for Disease Control and Prevention Division of Viral Diseases, Pathogen Discovery | Alison Laufer Halpin; Ben L. Rambo-Martin; Clinton R. Paden; Dakota Howard; Darlene Wagner; Dave Wentworth; Dhwani Batra; Jasmine Padilla; Justin Lee; Katie Dillon; Krista Queen; Kristen Knipe; Kristine Lacek; Mark Burroughs; Matthew Schmerer; Mili Sheth; Peter Cook; Sam Shepard; Sarah Nobles; Shoshona Le; Suxiang Tong; Vivien Dugan; Yvette Unoarumhi |
| EPI_ISL_2161718, EPI_ISL_2161747, EPI_ISL_2161775, EPI_ISL_2427404, EPI_ISL_2427406, EPI_ISL_2427408, EPI_ISL_2427410, EPI_ISL_2427432, EPI_ISL_2427464, EPI_ISL_2427468 | | | |
| see above | NYU Langone Health | Departments of Pathology and Medicine, New York University School of Medicine | Adriana Heguy; Christian Marier; Dacia Dimartino; Emily Guzman; Gael Westby; Guiqing Wang; Paolo Cotzia; Paul Zappile; Peter Meyn; Sitharam Ramaswami; Yutong Zhang |
| EPI_ISL_2023408, EPI_ISL_2023409, EPI_ISL_2233081, EPI_ISL_2400270, EPI_ISL_2400274, EPI_ISL_2689668 | Nebraska Public Health Laboratory | NPHL COVID-19 Response Team | NPHL COVID-19 Response Team |
| EPI_ISL_2277638 | Nevada State Public Health Laboratory | Nevada State Public Health Laboratory | Andrew Gorzalski; Mark Pandori |
| EPI_ISL_1970970, EPI_ISL_2322768, EPI_ISL_2322770, EPI_ISL_2322789, EPI_ISL_2322792, EPI_ISL_2322848, EPI_ISL_2381909, EPI_ISL_2381943, EPI_ISL_2652062 | | | |
| see above | New Mexico Department of Health Scientific Laboratory | New Mexico Department of Health Scientific Laboratory | Anastacia Griego-Fisher; D'eldra Malone; Ellie Johnson; Jennifer Benoit; Keila Gutierrez; Linda Salazar; Mark Willmon; Ratheesh Rajan |
| EPI_ISL_1920010 | Northwell Health Laboratories | Wadsworth Center, New York State Department of Health | Alexis Russell; Catharine Prussing; Daryl M. Lamson; Erasmus Schneider; Erica Lasek-Nesselquist; John Kelly; Jonathan Plitnick; Kirsten St. George; Matthew Shudt; Melissa A Leisner; Navjot Singh |
| EPI_ISL_2490159, EPI_ISL_2490251, EPI_ISL_2692356, EPI_ISL_2692368 | Northwest Laboratory | Seattle Flu Study | Amanda Adler; Barry R. Lutz; Benjamin Pelle; Brian Hiatt; Caitlin R. Wolf; Chris D. Frazar; Deborah A. Nickerson; Elisabeth Brandstetter; Erica Ryke; Geoff Melly; Helen Y. Chu; Janet A. Englund; Jay Shendure; Jover Lee; Kairsten Fay; Kirsten Lacombe; Lea M. Starita; Mark J. Rieder; Matthew Richardson; Matthew Thompson; Melissa Truong; Michael Boeckh; Michael Famulare; Misja Ilcisin; Peter D. Han; Philip Dykema; Romesh Gautom; Scott Lindquist; Thomas R. Sibley; Trevor Bedford |
| EPI_ISL_2009333, EPI_ISL_2009339, EPI_ISL_2009347, EPI_ISL_2009351, EPI_ISL_2009352, EPI_ISL_2372956, EPI_ISL_2372972, EPI_ISL_2372974, EPI_ISL_2372978, EPI_ISL_2372984, EPI_ISL_2372992, EPI_ISL_2372993, EPI_ISL_2373002, EPI_ISL_2373008, EPI_ISL_2373009, EPI_ISL_2373019, EPI_ISL_2373023, EPI_ISL_2373031, EPI_ISL_2373032, EPI_ISL_2373040, EPI_ISL_2373042 | | | |
| see above | Northwestern Memorial Hospital | Northwestern University - Ozer Lab | Chad J. Achenbach; Chao Qi; Egon A. Ozer; Judd F. Hultquist; Lacy M. Simons; Lawrence J. Jennings; Michael G. Ison; Ramon Lorenzo-Redondo; Taylor J. Dean |
| EPI_ISL_2692034, EPI_ISL_2692045, EPI_ISL_2692046, EPI_ISL_3388716, EPI_ISL_3388717, EPI_ISL_3388718, EPI_ISL_3388722, EPI_ISL_3388802 | | | |
| see above | Northwestern Memorial Hospital | RIPHL at Rush University Medical Center | Amber Kimble; Chao Qi; Edith Perez; Felix Araujo Perez; Kevin Kunstman; Laura Furtado; Marieta Hyde; Max Kolton; Stefan Green |
| EPI_ISL_2227690, EPI_ISL_2227691, EPI_ISL_2227692, EPI_ISL_2227693, EPI_ISL_2378934, EPI_ISL_2378977, EPI_ISL_2378981, EPI_ISL_2462076, EPI_ISL_2462090, EPI_ISL_2462091, EPI_ISL_2462105, EPI_ISL_2462117, EPI_ISL_2462120, EPI_ISL_2462135, EPI_ISL_2462136, EPI_ISL_2462137, EPI_ISL_2462138, EPI_ISL_2462140, EPI_ISL_2462147, EPI_ISL_2462153, EPI_ISL_2462173, EPI_ISL_2462179, EPI_ISL_2462182, EPI_ISL_2462184, EPI_ISL_2462185, EPI_ISL_2462187, EPI_ISL_2462198, EPI_ISL_2462202, EPI_ISL_2462221, EPI_ISL_2462223, EPI_ISL_2462230, EPI_ISL_2462231, EPI_ISL_2462232, EPI_ISL_2462241, EPI_ISL_2462253, EPI_ISL_2462263, EPI_ISL_2462270, EPI_ISL_2462272, EPI_ISL_2462280, EPI_ISL_2462281, EPI_ISL_2462285, EPI_ISL_2462294, EPI_ISL_2462297, EPI_ISL_2462302, EPI_ISL_2462304, EPI_ISL_2462305, EPI_ISL_2462308, EPI_ISL_2663619, EPI_ISL_2663623, EPI_ISL_2663642, EPI_ISL_2663643, EPI_ISL_2663648, EPI_ISL_2663651, EPI_ISL_2663652, EPI_ISL_2663669, EPI_ISL_2663671, EPI_ISL_2663672, EPI_ISL_2663679, EPI_ISL_2663687, EPI_ISL_2663703, EPI_ISL_2663706, EPI_ISL_2663707, EPI_ISL_2663716, EPI_ISL_2663722, EPI_ISL_2663723, EPI_ISL_2663736, EPI_ISL_2663738, EPI_ISL_2663740, EPI_ISL_2663741, EPI_ISL_2663752, EPI_ISL_2663755, EPI_ISL_2663764, EPI_ISL_2663765, EPI_ISL_2838825, EPI_ISL_2838826, EPI_ISL_2838827, EPI_ISL_2838828, EPI_ISL_2838829, EPI_ISL_2838830, EPI_ISL_2838831, EPI_ISL_2838832, EPI_ISL_2838833, EPI_ISL_2838834, EPI_ISL_2838835, EPI_ISL_2838836, EPI_ISL_2838837, EPI_ISL_2838838, EPI_ISL_2838839, EPI_ISL_2838840, EPI_ISL_2838841, EPI_ISL_2838842, EPI_ISL_2838843, EPI_ISL_2838844, EPI_ISL_2838845, EPI_ISL_2838846, EPI_ISL_2838847, EPI_ISL_2838848, EPI_ISL_2838849, EPI_ISL_2838850, EPI_ISL_2838851, EPI_ISL_2838852, EPI_ISL_2838853, EPI_ISL_2838854, EPI_ISL_2838855 | | | |
| see above | OHSU Lab Services Molecular Microbiology Lab | Oregon SARS-CoV-2 Genome Sequencing Center | '[UsersoroakBox SyncPHI_SARS-COV-2_Sample_Sheet_infoGISAID_Submission_Infometa_master_2021.xlsx]release20_final'!D:AC; 26; Alec J. Hirsch; Andrew C. Adey; Benjamin N. Bimber; Brendan L. O'Connell; Brian J. O'Roak; Cierra LaBlanc; Daniel N. Streblow; Donna Hansel; FALSE); Guang Fan; Kayla Carter; Ruth V. Nichols; Sally Grindstaff; Sonia Acharya; VLOOKUP(C378; VLOOKUP(C379; VLOOKUP(C389; VLOOKUP(C390; VLOOKUP(C391; VLOOKUP(C395; VLOOKUP(C396; VLOOKUP(C397; VLOOKUP(C398; VLOOKUP(C410; VLOOKUP(C412; VLOOKUP(C413; VLOOKUP(C416; VLOOKUP(C418; VLOOKUP(C419; VLOOKUP(C421; VLOOKUP(C422; VLOOKUP(C423; VLOOKUP(C424; VLOOKUP(C427; VLOOKUP(C428; VLOOKUP(C429; VLOOKUP(C430; VLOOKUP(C433; VLOOKUP(C434; VLOOKUP(C438; VLOOKUP(C440; VLOOKUP(C441; VLOOKUP(C454; VLOOKUP(C459; VLOOKUP(C463; William B. Messer; Xuan Qin |
| EPI_ISL_2467884 | OK Public Health Laboratory, Oklahoma State DOH | Genomics and Discovery, Respiratory Viruses Branch, Division of Viral Diseases, Centers for Disease Control and Prevention | Adam Retchless; Anna Kelleher; Anna Uehara; Brian Lynch; Clinton R. Paden; Dhwani Batra; Haibin Wang; Han Jia Justin Ng; Jasmine Padilla; Jing Zhang; Justin Lee; Krista Queen; Mark Burroughs; Mili Sheth; Morgan Davis; Peter Cook; Rachel Marine; Sarah Nobles; Suxiang Tong; Tara Coalter; Yan Li; Ying Tao |
| EPI_ISL_2384211, EPI_ISL_2384217, EPI_ISL_2384220, EPI_ISL_2399429, EPI_ISL_2451980, EPI_ISL_2451989, EPI_ISL_2451994, EPI_ISL_2689978, EPI_ISL_2689988, EPI_ISL_3354886, EPI_ISL_3354890, EPI_ISL_3354911, EPI_ISL_3354936 | | | |
| see above | OR State PHL-Virology/Immunology Section | Centers for Disease Control and Prevention Division of Viral Diseases, Pathogen Discovery | Alex Burgin; Alison Laufer Halpin; Ben L. Rambo-Martin; Ben Rambo-Martin; Clinton Paden; Clinton R. Paden; Dakota Howard; Darlene Wagner; Dave Wentworth; Dhwani Batra; Jasmine Padilla; Justin Lee; Katie Dillon; Krista Queen; Kristen Knipe; Kristine Lacek; Mark Burroughs; Matthew Schmerer; Meghan Bentz; Mili Sheth; Peter Cook; Sam Shepard; Sarah Nobles; Shoshona Le; Suxiang Tong; Vivien Dugan; Yvette Unoarumhi |
| EPI_ISL_2753965, EPI_ISL_2755100, EPI_ISL_2768100, EPI_ISL_2858702, EPI_ISL_2858703 | Ohio Department of Health Laboratory | Ohio Department of Health Laboratory | Allison Black; Brent Lee; Caitlin McDonnell; Eric Brandt; Erica Leasure; Glen McGillivary; Heather Blankenship; Holmes; Jade Mowery; Jennifer; Kelsey Florek; Keoni Omura; Kirtana Ramadugu; Quanta Brown; Stephanie Mccracken; Tyler Payne; and Tammy Bannerman |
| EPI_ISL_2566359, EPI_ISL_2566360, EPI_ISL_2566361 | Operation Infectious Diseases, Naval Health Research Center | Operation Infectious Diseases, Naval Health Research Center | A.E.; Balansay-Ames; C.A.; Fierro, M.; Iniguez-Stevens, E.; Kriner, P.; M.S.; Moser, K.; Myers; Pan; Phippard; R.S.; R.W.; Underwood |
| EPI_ISL_2500059, EPI_ISL_2500104 | Orange County Public Health Lab | Chan-Zuckerberg Biohub | CZB Cliahub Consortium |
| EPI_ISL_1897467, EPI_ISL_1967717, EPI_ISL_1967718, EPI_ISL_2023470, EPI_ISL_2023471, EPI_ISL_2023472, EPI_ISL_2023477, EPI_ISL_2081513, EPI_ISL_2081522, EPI_ISL_2086677, EPI_ISL_2086696, EPI_ISL_2138891, EPI_ISL_2138892, EPI_ISL_2139551, EPI_ISL_2139560, EPI_ISL_2139565, EPI_ISL_2203066, EPI_ISL_2203067, EPI_ISL_2203070, EPI_ISL_2203071, EPI_ISL_2203095, EPI_ISL_2250168, EPI_ISL_2339321, EPI_ISL_2339322, EPI_ISL_2339323, EPI_ISL_2339324, EPI_ISL_2339325, EPI_ISL_2339326, EPI_ISL_2339327, EPI_ISL_2339335, EPI_ISL_2346112, EPI_ISL_2346113, EPI_ISL_2346120, EPI_ISL_2348927, EPI_ISL_2348931, EPI_ISL_2348933, EPI_ISL_2348949, EPI_ISL_2348953, EPI_ISL_2404739, EPI_ISL_2404748, EPI_ISL_2424247, EPI_ISL_2424248, EPI_ISL_2424255, EPI_ISL_2473661, EPI_ISL_2473662, EPI_ISL_2473663, EPI_ISL_2473671, EPI_ISL_2473672, EPI_ISL_2557921, EPI_ISL_2727477 | | | |
| see above | Oregon State Public Health Laboratory | Oregon State Public Health Laboratory | Eugene Yeboah; John Fontana and Shane Sevey; Laura Tsaknaridis; Rafia Razzaque; Vanda Makris |
| EPI_ISL_2295791 | Oregon State University TRACE Project | OSU Center for Genome Research and Biocomputing | Oregon State University TRACE Project |
| EPI_ISL_2293349, EPI_ISL_2293357, EPI_ISL_2293358, EPI_ISL_2293359, EPI_ISL_2293360, EPI_ISL_2293361, EPI_ISL_2293362, EPI_ISL_2293363, EPI_ISL_2293364, EPI_ISL_2293365, EPI_ISL_2293367, EPI_ISL_2293368, EPI_ISL_2293369, EPI_ISL_2293370, EPI_ISL_2293371, EPI_ISL_2293373, EPI_ISL_2293374, EPI_ISL_2293375, EPI_ISL_2293376, EPI_ISL_2293377, EPI_ISL_2293378, EPI_ISL_2293379, EPI_ISL_2293380, EPI_ISL_2293381, EPI_ISL_2293382 | | | |
| see above | Outre mer | National Reference Center for Viruses of Respiratory Infections, Institut Pasteur, Paris | Angela Brisebarre; Camille Capel; Christophe Malabat; Corinne Maufrais; Dominique Rousset; Etienne Simon-Lorière; Frédéric Lemoine; Louise Lefrançois; Marion Barbet; Maud Vanpeene; Méline Bizard; Pierre Lechat; Sylvie Behillil; Sylvie Van der Werf; Vincent Enouf |
| EPI_ISL_2445407, EPI_ISL_2445439 | PA CAPELA | Instituto Butantan | Antonio Jorge Martins; Claudia Renata dos Santos Barros; David Schlesinger; Debora Botequio Moretti; Dimas Tadeu Covas; Elaine Cristina Marqueze; Elaine Vieira Santos; Evandra Strazza Rodrigues; Heidge Fukumasu; Jayme Augusto de Souza-Neto; José Salvatore Leister Patané; Luiz Alcantara; Luiz Lehmann Coutinho; Maria Carolina Elias; Maurício Lacerda Nogueira; Rafael dos Santos Bezerra; Raul Machado Neto; Rejane Maria Tommasini Grotto; Ricardo Haddad; Sandra Coccuzzo Sampaio Vessoni; Simone Kashima; Svetoslav Nanev Slavov; Vincent Louis Viala |
| EPI_ISL_2493489, EPI_ISL_2493490, EPI_ISL_2493501, EPI_ISL_2493504 | PA MATAO | Instituto Butantan | Antonio Jorge Martins; Claudia Renata dos Santos Barros; David Schlesinger; Debora Botequio Moretti; Dimas Tadeu Covas; Elaine Cristina Marqueze; Elaine Vieira Santos; Evandra Strazza Rodrigues; Heidge Fukumasu; Jayme Augusto de Souza-Neto; José Salvatore Leister Patané; Luiz Alcantara; Luiz Lehmann Coutinho; Maria Carolina Elias; Maurício Lacerda Nogueira; Rafael dos Santos Bezerra; Raul Machado Neto; Rejane Maria Tommasini Grotto; Ricardo Haddad; Sandra Coccuzzo Sampaio Vessoni; Simone Kashima; Svetoslav Nanev Slavov; Vincent Louis Viala |
| EPI_ISL_2493356, EPI_ISL_2493357, EPI_ISL_2493358, EPI_ISL_2493359 | PAS JOAO ANTONIO DO NASCIMENTO | Instituto Butantan | Antonio Jorge Martins; Claudia Renata dos Santos Barros; David Schlesinger; Debora Botequio Moretti; Dimas Tadeu Covas; Elaine Cristina Marqueze; Elaine Vieira Santos; Evandra Strazza Rodrigues; Heidge Fukumasu; Jayme Augusto de Souza-Neto; José Salvatore Leister Patané; Luiz Alcantara; Luiz Lehmann Coutinho; Maria Carolina Elias; Maurício Lacerda Nogueira; Rafael dos Santos Bezerra; Raul Machado Neto; Rejane Maria Tommasini Grotto; Ricardo Haddad; Sandra Coccuzzo Sampaio Vessoni; Simone Kashima; Svetoslav Nanev Slavov; Vincent Louis Viala |
| EPI_ISL_2345675 | PAS JOAO ANTONIO DO NASCIMENTO | Instituto Butantan / Mendelics | Antonio Jorge Martins; Claudia Renata dos Santos Barros; David Schlesinger; Debora Botequio Moretti; Dimas Tadeu Covas; Elaine Cristina Marqueze; Elaine Vieira Santos; Evandra Strazza Rodrigues; Heidge Fukumasu; Jayme Augusto de Souza-Neto; José Salvatore Leister Patané; Luiz Alcantara; Luiz Lehmann Coutinho; Maria Carolina Elias; Maurício Lacerda Nogueira; Rafael dos Santos Bezerra; Raul Machado Neto; Rejane Maria Tommasini Grotto; Ricardo Haddad; Sandra Coccuzzo Sampaio Vessoni; Simone Kashima; Svetoslav Nanev Slavov; Vincent Louis Viala |
| EPI_ISL_2500981 | PAS JULIANIA DE HERCULANDIA | Instituto Butantan | Antonio Jorge Martins; Claudia Renata dos Santos Barros; David Schlesinger; Debora Botequio Moretti; Dimas Tadeu Covas; Elaine Cristina Marqueze; Elaine Vieira Santos; Evandra Strazza Rodrigues; Heidge Fukumasu; Jayme Augusto de Souza-Neto; José Salvatore Leister Patané; Luiz Alcantara; Luiz Lehmann |

| | | | |
|---|---|---|---|
| | | | Coutinho; Maria Carolina Elias; Maurício Lacerda Nogueira; Rafael dos Santos Bezerra; Raul Machado Neto; Rejane Maria Tommasini Grotto; Ricardo Haddad; Sandra Coccuzzo Sampaio Vessoni; Simone Kashima; Svetoslav Nanev Slavov; Vincent Louis Viala |
| EPI_ISL_2208911 | PENITENCIARIA DE PRESIDENTE PRUDENTE | Instituto Butantan / Mendelics | Antonio Jorge Martins; Claudia Renata dos Santos Barros; David Schlesinger; Debora Botequio Moretti; Dimas Tadeu Covas; Elaine Cristina Marqueze; Elaine Vieira Santos; Evandra Strazza Rodrigues; Heidge Fukumasu; Jayme Augusto de Souza-Neto; José Salvatore Leister Patané; Luiz Alcantara; Luiz Lehmann Coutinho; Maria Carolina Elias; Maurício Lacerda Nogueira; Rafael dos Santos Bezerra; Raul Machado Neto; Rejane Maria Tommasini Grotto; Ricardo Haddad; Sandra Coccuzzo Sampaio Vessoni; Simone Kashima; Svetoslav Nanev Slavov; Vincent Louis Viala |
| EPI_ISL_2445596, EPI_ISL_2493667, EPI_ISL_2493673 | POLICLINICA COVID 19 ITAPETININGA | Instituto Butantan | Antonio Jorge Martins; Claudia Renata dos Santos Barros; David Schlesinger; Debora Botequio Moretti; Dimas Tadeu Covas; Elaine Cristina Marqueze; Elaine Vieira Santos; Evandra Strazza Rodrigues; Heidge Fukumasu; Jayme Augusto de Souza-Neto; José Salvatore Leister Patané; Luiz Alcantara; Luiz Lehmann Coutinho; Maria Carolina Elias; Maurício Lacerda Nogueira; Rafael dos Santos Bezerra; Raul Machado Neto; Rejane Maria Tommasini Grotto; Ricardo Haddad; Sandra Coccuzzo Sampaio Vessoni; Simone Kashima; Svetoslav Nanev Slavov; Vincent Louis Viala |
| EPI_ISL_2344927, EPI_ISL_2344941 | POLICLINICA COVID 19 ITAPETININGA | Instituto Butantan / ESALQ-Piracicaba | Antonio Jorge Martins; Claudia Renata dos Santos Barros; David Schlesinger; Debora Botequio Moretti; Dimas Tadeu Covas; Elaine Cristina Marqueze; Elaine Vieira Santos; Evandra Strazza Rodrigues; Heidge Fukumasu; Jayme Augusto de Souza-Neto; José Salvatore Leister Patané; Luiz Alcantara; Luiz Lehmann Coutinho; Maria Carolina Elias; Maurício Lacerda Nogueira; Rafael dos Santos Bezerra; Raul Machado Neto; Rejane Maria Tommasini Grotto; Ricardo Haddad; Sandra Coccuzzo Sampaio Vessoni; Simone Kashima; Svetoslav Nanev Slavov; Vincent Louis Viala |
| EPI_ISL_2345809, EPI_ISL_2345814, EPI_ISL_2345859 | POLICLINICA COVID 19 ITAPETININGA | Instituto Butantan / Mendelics | Antonio Jorge Martins; Claudia Renata dos Santos Barros; David Schlesinger; Debora Botequio Moretti; Dimas Tadeu Covas; Elaine Cristina Marqueze; Elaine Vieira Santos; Evandra Strazza Rodrigues; Heidge Fukumasu; Jayme Augusto de Souza-Neto; José Salvatore Leister Patané; Luiz Alcantara; Luiz Lehmann Coutinho; Maria Carolina Elias; Maurício Lacerda Nogueira; Rafael dos Santos Bezerra; Raul Machado Neto; Rejane Maria Tommasini Grotto; Ricardo Haddad; Sandra Coccuzzo Sampaio Vessoni; Simone Kashima; Svetoslav Nanev Slavov; Vincent Louis Viala |
| EPI_ISL_2493198, EPI_ISL_2493246, EPI_ISL_2493468, EPI_ISL_2493471, EPI_ISL_2493478, EPI_ISL_2493481, EPI_ISL_2493493, EPI_ISL_2493499, EPI_ISL_2493502, EPI_ISL_2493503 | | | |
| see above | POLICLINICA HORTOLANDIA | Instituto Butantan | Antonio Jorge Martins; Claudia Renata dos Santos Barros; David Schlesinger; Debora Botequio Moretti; Dimas Tadeu Covas; Elaine Cristina Marqueze; Elaine Vieira Santos; Evandra Strazza Rodrigues; Heidge Fukumasu; Jayme Augusto de Souza-Neto; José Salvatore Leister Patané; Luiz Alcantara; Luiz Lehmann Coutinho; Maria Carolina Elias; Maurício Lacerda Nogueira; Rafael dos Santos Bezerra; Raul Machado Neto; Rejane Maria Tommasini Grotto; Ricardo Haddad; Sandra Coccuzzo Sampaio Vessoni; Simone Kashima; Svetoslav Nanev Slavov; Vincent Louis Viala |
| EPI_ISL_2345265 | POLICLINICA HORTOLANDIA | Instituto Butantan / ESALQ-Piracicaba | Antonio Jorge Martins; Claudia Renata dos Santos Barros; David Schlesinger; Debora Botequio Moretti; Dimas Tadeu Covas; Elaine Cristina Marqueze; Elaine Vieira Santos; Evandra Strazza Rodrigues; Heidge Fukumasu; Jayme Augusto de Souza-Neto; José Salvatore Leister Patané; Luiz Alcantara; Luiz Lehmann Coutinho; Maria Carolina Elias; Maurício Lacerda Nogueira; Rafael dos Santos Bezerra; Raul Machado Neto; Rejane Maria Tommasini Grotto; Ricardo Haddad; Sandra Coccuzzo Sampaio Vessoni; Simone Kashima; Svetoslav Nanev Slavov; Vincent Louis Viala |
| EPI_ISL_2445574, EPI_ISL_2445576 | POSTO DE SAUDE BENVIRA E S F 4 | Instituto Butantan | Antonio Jorge Martins; Claudia Renata dos Santos Barros; David Schlesinger; Debora Botequio Moretti; Dimas Tadeu Covas; Elaine Cristina Marqueze; Elaine Vieira Santos; Evandra Strazza Rodrigues; Heidge Fukumasu; Jayme Augusto de Souza-Neto; José Salvatore Leister Patané; Luiz Alcantara; Luiz Lehmann Coutinho; Maria Carolina Elias; Maurício Lacerda Nogueira; Rafael dos Santos Bezerra; Raul Machado Neto; Rejane Maria Tommasini Grotto; Ricardo Haddad; Sandra Coccuzzo Sampaio Vessoni; Simone Kashima; Svetoslav Nanev Slavov; Vincent Louis Viala |
| EPI_ISL_3102280 | POSTO DE SAUDE MARIA DE LOURDES MAGALHAES MAIA | Analytical Competence Molecular Epidemiology Lab/ACME, Oswaldo Cruz Foundation, Ceara (FIOCRUZ CE) | Cleber Furtado Aksenen; Fabio Miyajima; Fernando Braga Stehling; Francisco Eder de Moura Lopes; Jamille Maria Mendes Bezerra; Joaquim César do Nascimento Sousa Junior; Pedro Miguel Carneiro Jeronimo; Suzana Porto Almeida e Lucas Delerino; Thais Ferreira de Oliveira; Thais de Oliveira Costa; Ticiane Cavalcante de Souza; Veridiana Pessoa Miyajima |
| EPI_ISL_2493393 | POSTO MEDICO CIDADE NOVA | Instituto Butantan | Antonio Jorge Martins; Claudia Renata dos Santos Barros; David Schlesinger; Debora Botequio Moretti; Dimas Tadeu Covas; Elaine Cristina Marqueze; Elaine Vieira Santos; Evandra Strazza Rodrigues; Heidge Fukumasu; Jayme Augusto de Souza-Neto; José Salvatore Leister Patané; Luiz Alcantara; Luiz Lehmann Coutinho; Maria Carolina Elias; Maurício Lacerda Nogueira; Rafael dos Santos Bezerra; Raul Machado Neto; Rejane Maria Tommasini Grotto; Ricardo Haddad; Sandra Coccuzzo Sampaio Vessoni; Simone Kashima; Svetoslav Nanev Slavov; Vincent Louis Viala |
| EPI_ISL_2493589, EPI_ISL_2493590, EPI_ISL_2493629 | POSTO MEDICO NATAL DIEGUES DE ESTIVA GERBI | Instituto Butantan | Antonio Jorge Martins; Claudia Renata dos Santos Barros; David Schlesinger; Debora Botequio Moretti; Dimas Tadeu Covas; Elaine Cristina Marqueze; Elaine Vieira Santos; Evandra Strazza Rodrigues; Heidge Fukumasu; Jayme Augusto de Souza-Neto; José Salvatore Leister Patané; Luiz Alcantara; Luiz Lehmann Coutinho; Maria Carolina Elias; Maurício Lacerda Nogueira; Rafael dos Santos Bezerra; Raul Machado Neto; Rejane Maria Tommasini Grotto; Ricardo Haddad; Sandra Coccuzzo Sampaio Vessoni; Simone Kashima; Svetoslav Nanev Slavov; Vincent Louis Viala |
| EPI_ISL_3354889 | PR Public Health Lab | Centers for Disease Control and Prevention Division of Viral Diseases, Pathogen Discovery | Alex Burgin; Ben Rambo-Martin; Clinton Paden; Dakota Howard; Dave Wentworth; Dhwani Batra; Jasmine Padilla; Justin Lee; Krista Queen; Kristen Knipe; Kristine Lacek; Mark Burroughs; Matthew Schmerer; Meghan Bentz; Mili Sheth; Peter Cook; Sam Shepard; Sarah Nobles; Suxiang Tong; Vivien Dugan; Yvette Unoarumhi |
| EPI_ISL_2493277, EPI_ISL_2493355, EPI_ISL_2493694, EPI_ISL_2493695 | PROGRAMA SAUDE DA FAMILIA I IEPE | Instituto Butantan | Antonio Jorge Martins; Claudia Renata dos Santos Barros; David Schlesinger; Debora Botequio Moretti; Dimas Tadeu Covas; Elaine Cristina Marqueze; Elaine Vieira Santos; Evandra Strazza Rodrigues; Heidge Fukumasu; Jayme Augusto de Souza-Neto; José Salvatore Leister Patané; Luiz Alcantara; Luiz Lehmann Coutinho; Maria Carolina Elias; Maurício Lacerda Nogueira; Rafael dos Santos Bezerra; Raul Machado Neto; Rejane Maria Tommasini Grotto; Ricardo Haddad; Sandra Coccuzzo Sampaio Vessoni; Simone Kashima; Svetoslav Nanev Slavov; Vincent Louis Viala |
| EPI_ISL_2445241, EPI_ISL_2445244, EPI_ISL_2445252, EPI_ISL_2445369 | PRONTO ATENDIMENTO DE VARGEM GRANDE PAULISTA | Instituto Butantan | Antonio Jorge Martins; Claudia Renata dos Santos Barros; David Schlesinger; Debora Botequio Moretti; Dimas Tadeu Covas; Elaine Cristina Marqueze; Elaine Vieira Santos; Evandra Strazza Rodrigues; Heidge Fukumasu; Jayme Augusto de Souza-Neto; José Salvatore Leister Patané; Luiz Alcantara; Luiz Lehmann Coutinho; Maria Carolina Elias; Maurício Lacerda Nogueira; Rafael dos Santos Bezerra; Raul Machado Neto; Rejane Maria Tommasini Grotto; Ricardo Haddad; Sandra Coccuzzo Sampaio Vessoni; Simone Kashima; Svetoslav Nanev Slavov; Vincent Louis Viala |
| EPI_ISL_2345714 | PRONTO ATENDIMENTO MUNICIPAL | Instituto Butantan / Mendelics | Antonio Jorge Martins; Claudia Renata dos Santos Barros; David Schlesinger; Debora Botequio Moretti; Dimas Tadeu Covas; Elaine Cristina Marqueze; Elaine Vieira Santos; Evandra Strazza Rodrigues; Heidge Fukumasu; Jayme Augusto de Souza-Neto; José Salvatore Leister Patané; Luiz Alcantara; Luiz Lehmann Coutinho; Maria Carolina Elias; Maurício Lacerda Nogueira; Rafael dos Santos Bezerra; Raul Machado Neto; Rejane Maria Tommasini Grotto; Ricardo Haddad; Sandra Coccuzzo Sampaio Vessoni; Simone Kashima; Svetoslav Nanev Slavov; Vincent Louis Viala |
| EPI_ISL_2493470 | PRONTO ATENDIMENTO SAO JOSE | Instituto Butantan | Antonio Jorge Martins; Claudia Renata dos Santos Barros; David Schlesinger; Debora Botequio Moretti; Dimas Tadeu Covas; Elaine Cristina Marqueze; Elaine Vieira Santos; Evandra Strazza Rodrigues; Heidge Fukumasu; Jayme Augusto de Souza-Neto; José Salvatore Leister Patané; Luiz Alcantara; Luiz Lehmann Coutinho; Maria Carolina Elias; Maurício Lacerda Nogueira; Rafael dos Santos Bezerra; Raul Machado Neto; Rejane Maria Tommasini Grotto; Ricardo Haddad; Sandra Coccuzzo Sampaio Vessoni; Simone Kashima; Svetoslav Nanev Slavov; Vincent Louis Viala |
| EPI_ISL_2493552, EPI_ISL_2493556, EPI_ISL_2493564 | PRONTO SOCORRO DE AGENOR DE CAMPOS MONGAGUA | Instituto Butantan | Antonio Jorge Martins; Claudia Renata dos Santos Barros; David Schlesinger; Debora Botequio Moretti; Dimas Tadeu Covas; Elaine Cristina Marqueze; Elaine Vieira Santos; Evandra Strazza Rodrigues; Heidge Fukumasu; Jayme Augusto de Souza-Neto; José Salvatore Leister Patané; Luiz Alcantara; Luiz Lehmann Coutinho; Maria Carolina Elias; Maurício Lacerda Nogueira; Rafael dos Santos Bezerra; Raul Machado Neto; Rejane Maria Tommasini Grotto; Ricardo Haddad; Sandra Coccuzzo Sampaio Vessoni; Simone Kashima; Svetoslav Nanev Slavov; Vincent Louis Viala |
| EPI_ISL_2493594, EPI_ISL_2493595, EPI_ISL_2493613, EPI_ISL_2493618, EPI_ISL_2493624, EPI_ISL_2493626 | PRONTO SOCORRO MUNICIPAL DE CRAVINHOS | Instituto Butantan | Antonio Jorge Martins; Claudia Renata dos Santos Barros; David Schlesinger; Debora Botequio Moretti; Dimas Tadeu Covas; Elaine Cristina Marqueze; Elaine Vieira Santos; Evandra Strazza Rodrigues; Heidge Fukumasu; Jayme Augusto de Souza-Neto; José Salvatore Leister Patané; Luiz Alcantara; Luiz Lehmann Coutinho; Maria Carolina Elias; Maurício Lacerda Nogueira; Rafael dos Santos Bezerra; Raul Machado Neto; Rejane Maria Tommasini Grotto; Ricardo Haddad; Sandra Coccuzzo Sampaio Vessoni; Simone Kashima; Svetoslav Nanev Slavov; Vincent Louis Viala |
| EPI_ISL_2345199 | PRONTO SOCORRO MUNICIPAL DE SEVERINIA | Instituto Butantan / ESALQ-Piracicaba | Antonio Jorge Martins; Claudia Renata dos Santos Barros; David Schlesinger; Debora Botequio Moretti; Dimas Tadeu Covas; Elaine Cristina Marqueze; Elaine Vieira Santos; Evandra Strazza Rodrigues; Heidge Fukumasu; Jayme Augusto de Souza-Neto; José Salvatore Leister Patané; Luiz Alcantara; Luiz Lehmann Coutinho; Maria Carolina Elias; Maurício Lacerda Nogueira; Rafael dos Santos Bezerra; Raul Machado Neto; Rejane Maria Tommasini Grotto; Ricardo Haddad; Sandra Coccuzzo Sampaio Vessoni; Simone Kashima; Svetoslav Nanev Slavov; Vincent Louis Viala |
| EPI_ISL_2493527 | PRONTO SOCORRO MUNICIPAL TAMBAU | Instituto Butantan | Antonio Jorge Martins; Claudia Renata dos Santos Barros; David Schlesinger; Debora Botequio Moretti; Dimas Tadeu Covas; Elaine Cristina Marqueze; Elaine Vieira Santos; Evandra Strazza Rodrigues; Heidge Fukumasu; Jayme Augusto de Souza-Neto; José Salvatore Leister Patané; Luiz Alcantara; Luiz Lehmann Coutinho; Maria Carolina Elias; Maurício Lacerda Nogueira; Rafael dos Santos Bezerra; Raul Machado Neto; Rejane Maria Tommasini Grotto; Ricardo Haddad; Sandra Coccuzzo Sampaio Vessoni; Simone Kashima; Svetoslav Nanev Slavov; Vincent Louis Viala |
| EPI_ISL_2345729 | PRONTO SOCORRO MUNICIPAL TAMBAU | Instituto Butantan / Mendelics | Antonio Jorge Martins; Claudia Renata dos Santos Barros; David Schlesinger; Debora Botequio Moretti; Dimas Tadeu Covas; Elaine Cristina Marqueze; Elaine Vieira Santos; Evandra Strazza Rodrigues; Heidge Fukumasu; Jayme Augusto de Souza-Neto; José Salvatore Leister Patané; Luiz Alcantara; Luiz Lehmann Coutinho; Maria Carolina Elias; Maurício Lacerda Nogueira; Rafael dos Santos Bezerra; Raul Machado Neto; Rejane Maria Tommasini Grotto; Ricardo Haddad; Sandra Coccuzzo Sampaio Vessoni; Simone Kashima; Svetoslav Nanev Slavov; Vincent Louis Viala |
| EPI_ISL_2209716 | PSF DO PARQUE DOS PINHEIROS | Instituto Butantan / Mendelics | Antonio Jorge Martins; Claudia Renata dos Santos Barros; David Schlesinger; Debora Botequio Moretti; Dimas Tadeu Covas; Elaine Cristina Marqueze; Elaine Vieira Santos; Evandra Strazza Rodrigues; Heidge Fukumasu; Jayme Augusto de Souza-Neto; José Salvatore Leister Patané; Luiz Alcantara; Luiz Lehmann Coutinho; Maria Carolina Elias; Maurício Lacerda Nogueira; Rafael dos Santos Bezerra; Raul Machado Neto; Rejane Maria Tommasini Grotto; Ricardo Haddad; Sandra Coccuzzo Sampaio Vessoni; Simone Kashima; Svetoslav Nanev Slavov; Vincent Louis Viala |
| EPI_ISL_2344910 | PSF MARIA JOSE SALTO DE PIRAPORA | Instituto Butantan / ESALQ-Piracicaba | Antonio Jorge Martins; Claudia Renata dos Santos Barros; David Schlesinger; Debora Botequio Moretti; Dimas Tadeu Covas; Elaine Cristina Marqueze; Elaine Vieira Santos; Evandra Strazza Rodrigues; Heidge Fukumasu; Jayme Augusto de Souza-Neto; José Salvatore Leister Patané; Luiz Alcantara; Luiz Lehmann Coutinho; Maria Carolina Elias; Maurício Lacerda Nogueira; Rafael dos Santos Bezerra; Raul Machado Neto; Rejane Maria Tommasini Grotto; Ricardo Haddad; Sandra Coccuzzo Sampaio Vessoni; Simone Kashima; Svetoslav Nanev Slavov; Vincent Louis Viala |
| EPI_ISL_2345672 | PSF MARIA JOSE SALTO DE PIRAPORA | Instituto Butantan / Mendelics | Antonio Jorge Martins; Claudia Renata dos Santos Barros; David Schlesinger; Debora Botequio Moretti; Dimas Tadeu Covas; Elaine Cristina Marqueze; Elaine Vieira Santos; Evandra Strazza Rodrigues; Heidge Fukumasu; Jayme Augusto de Souza-Neto; José Salvatore Leister Patané; Luiz Alcantara; Luiz Lehmann Coutinho; Maria Carolina Elias; Maurício Lacerda Nogueira; Rafael dos Santos Bezerra; Raul Machado Neto; Rejane Maria Tommasini Grotto; Ricardo Haddad; Sandra Coccuzzo Sampaio Vessoni; Simone Kashima; Svetoslav Nanev Slavov; Vincent Louis Viala |
| EPI_ISL_2344918 | PSF PALMITALZINHO APIAI | Instituto Butantan / ESALQ-Piracicaba | Antonio Jorge Martins; Claudia Renata dos Santos Barros; David Schlesinger; Debora Botequio Moretti; Dimas Tadeu Covas; Elaine Cristina Marqueze; Elaine Vieira Santos; Evandra Strazza Rodrigues; Heidge Fukumasu; Jayme Augusto de Souza-Neto; José Salvatore Leister Patané; Luiz Alcantara; Luiz Lehmann Coutinho; Maria Carolina Elias; Maurício Lacerda Nogueira; Rafael dos Santos Bezerra; Raul Machado Neto; Rejane Maria Tommasini Grotto; Ricardo Haddad; Sandra Coccuzzo Sampaio Vessoni; Simone Kashima; Svetoslav Nanev Slavov; Vincent Louis Viala |
| EPI_ISL_2493640, EPI_ISL_2493644, EPI_ISL_2493645 | PSF VIRGINIO BASSO | Instituto Butantan | Antonio Jorge Martins; Claudia Renata dos Santos Barros; David Schlesinger; Debora Botequio Moretti; Dimas Tadeu Covas; Elaine Cristina Marqueze; Elaine Vieira Santos; Evandra Strazza Rodrigues; Heidge Fukumasu; Jayme Augusto de Souza-Neto; José Salvatore Leister Patané; Luiz Alcantara; Luiz Lehmann Coutinho; Maria Carolina Elias; Maurício Lacerda Nogueira; Rafael dos Santos Bezerra; Raul Machado Neto; Rejane Maria Tommasini Grotto; Ricardo Haddad; Sandra Coccuzzo Sampaio Vessoni; Simone Kashima; Svetoslav Nanev Slavov; Vincent Louis Viala |
| EPI_ISL_1717818, EPI_ISL_1717859, EPI_ISL_1717950, EPI_ISL_1828239, EPI_ISL_1828491, EPI_ISL_1828520, EPI_ISL_1828569, EPI_ISL_1828614, EPI_ISL_1828615, EPI_ISL_1828683, EPI_ISL_2036997, EPI_ISL_2037071, EPI_ISL_2037072, EPI_ISL_2037091, EPI_ISL_2037109, EPI_ISL_2037153, EPI_ISL_2037165, EPI_ISL_2037181, EPI_ISL_2037216, EPI_ISL_2037300, EPI_ISL_2037338, EPI_ISL_2037375, EPI_ISL_2106316, EPI_ISL_2106360, EPI_ISL_2106365, EPI_ISL_2106383, EPI_ISL_2106488, EPI_ISL_2106568, EPI_ISL_2235006, EPI_ISL_2235042, EPI_ISL_2235090, EPI_ISL_2235158, EPI_ISL_2235168, EPI_ISL_2235191, EPI_ISL_2235194, EPI_ISL_2235205, EPI_ISL_2235215 | | | |
| see above | Pandemic Response Lab - NYC | Pandemic Response Lab, R&D | Cybill del Castillo; Dylan Law; Haiping Hao; Henry Lee; Jon Laurent; Katharine Nelson; Melissa Hopkins; Michael Hammerling; Pradeep Bugga; Shinyoung Clair Kang; Sol Rey; William Ward |
| EPI_ISL_3194067 | Pathogen Genomics Center, National Institute of Infectious Diseases | Pathogen Genomics Center, National Institute of Infectious Diseases | Kentaro Itokawa; Makoto Kuroda; Masanori Hashino; Rina Tanaka; Tsuyoshi Sekizuka |
| EPI_ISL_2993757 | Pathology, UAB | Pathology, UAB | E.J.; Hendrickson; Hernandez, J.; Leal; Lefkowitz; Moates, D.; R.C.; S.M. |
| EPI_ISL_2451606 | Philadelphia Department of Public Health | Centers for Disease Control and Prevention Division of Viral Diseases, Pathogen Discovery | Alison Laufer Halpin; Ben L. Rambo-Martin; Clinton R. Paden; Dakota Howard; Darlene Wagner; Dave Wentworth; Dhwani Batra; Jasmine Padilla; Justin Lee; Katie Dillon; Krista Queen; Kristen Knipe; Kristine Lacek; Mark Burroughs; Matthew Schmerer; Mili Sheth; Peter Cook; Sam Shepard; Sarah Nobles; Shoshona Le; Suxiang Tong; Vivien Dugan; Yvette Unoarumhi |
| EPI_ISL_2663311 | Plataforma de Vigilancia Molecular (PVM) - FIOCRUZ/BA | Plataforma de Vigilancia Molecular (PVM) - FIOCRUZ/BA | Bruno Bezerril Andrade; Camila I. de Oliveira on behalf of the Fiocruz COVID-19 Genomic Surveillance Network.; Clarissa Araújo Gurgel; Leonardo Paiva Farias; Marina Cucco; Ricardo Khouri; Tiago Graf |
| EPI_ISL_2131313, EPI_ISL_2272771, EPI_ISL_2362593 | Platform BIS UZA/UAntwerpen | Labo Klinische Biologie, UZA | Basil Britto Xavier; Christine Lammens; Herman Goossens; Jasmine Coppens; Marie Le Mercier; Veerle Matheeussen |
| EPI_ISL_2391780, EPI_ISL_2405204 | Pomona Valley Hospital Medical Center | Los Angeles County Public Health Laboratories | P. Hemarajata et al. |
| EPI_ISL_2759149 | Population Medicine and Diagnostic Sciences, Cornell University | Population Medicine and Diagnostic Sciences, Cornell University | Anderson; B.D.; Caserta; Cronk; D.G.; Diel; L.C.; Laverack, M.; Mitchell; P.K.; Plocharczyk, E.; R.R.; Venugopalan, R. |
| EPI_ISL_4052654 | Precision Genomics | MUSC Molecular Pathology Laboratory | Ana Olivo; Ana Vallari; Barbara Harris; Dariusz Pytel; Frederick S. Nolte; Gavin Cloherty; Jaclyn Dunne; Jeremy Stuart; Jim Madory; Julie W. Hirschhorn; Kristen Maurer; Mary Rodgers; Scott Curry; Todd Meyer; W. Bailey Glen Jr |
| EPI_ISL_2345907, EPI_ISL_2345911, EPI_ISL_2345915, EPI_ISL_2345916, EPI_ISL_2345917, EPI_ISL_2345920, EPI_ISL_2345923, EPI_ISL_2345926, EPI_ISL_2345927, EPI_ISL_2345928, EPI_ISL_2345932, EPI_ISL_2345933, EPI_ISL_2345934, EPI_ISL_2345935, EPI_ISL_2345937, EPI_ISL_2345938, EPI_ISL_2346094, EPI_ISL_2346095, EPI_ISL_2346096, EPI_ISL_2346097 | | | |
| see above | Prefeirtura de SP | Instituto Butantan | Antonio Jorge Martins; Claudia Renata dos Santos Barros; David Schlesinger; Debora Botequio Moretti; Dimas Tadeu Covas; Elaine Cristina Marqueze; Elaine Vieira Santos; Evandra Strazza Rodrigues; Heidge Fukumasu; Jayme Augusto de Souza-Neto; José Salvatore Leister Patané; Luiz Alcantara; Luiz Lehmann Coutinho; Maria Carolina Elias; Maurício Lacerda Nogueira; Rafael dos Santos Bezerra; Raul Machado Neto; Rejane Maria Tommasini Grotto; Ricardo Haddad; Sandra Coccuzzo Sampaio Vessoni; Simone Kashima; Svetoslav Nanev Slavov; Vincent Louis Viala |

| | | | |
|---|---|---|---|
| EPI_ISL_2232606 | Premier Medical Lab | Minnesota Department of Health, Public Health Laboratory | Alexandra Lorentz; Jacob Garfin; Matt Plumb; and Xiong Wang |
| EPI_ISL_2551453 | Providence Regional Medical Center Everett | Providence St. Joseph Health Molecular Genomics Laboratory | Alexa K Dowdell; Brian D Piening; Carlo B Bifulco; Fred L Robinson; Mary Campbell; Rogan Rattray |
| EPI_ISL_2294568, EPI_ISL_2697716, EPI_ISL_2697779, EPI_ISL_2697800, EPI_ISL_2697856, EPI_ISL_2697864, EPI_ISL_2698216, EPI_ISL_2698232, EPI_ISL_2698234, EPI_ISL_2698247, EPI_ISL_2698317, EPI_ISL_2698363, EPI_ISL_2698387, EPI_ISL_2698423, EPI_ISL_2698429, EPI_ISL_2698546, EPI_ISL_2698728, EPI_ISL_2699183, EPI_ISL_2699253, EPI_ISL_2699399, EPI_ISL_2699437, EPI_ISL_2709021, EPI_ISL_2709031, EPI_ISL_2709048 | | | |
| see above | Public Health Laboratory, Minnesota Department of Health | University of Minnesota Genomics Center | Corbin Dirkx; Daryl M. Gohl; Jaquelyn Kuriger-Laber; John Garbe; and Sean Wang |
| EPI_ISL_2196082, EPI_ISL_2196150 | QLabs | WVU and Marshall University Combined Genomics Core Facilities | James Denvir; Peter Perrotta; Peter Stoilov; Ryan Percifield; Wesley Kimble |
| EPI_ISL_1942187, EPI_ISL_1989981, EPI_ISL_1989982, EPI_ISL_1989986, EPI_ISL_1989988, EPI_ISL_1989990, EPI_ISL_2034720, EPI_ISL_2034724, EPI_ISL_2034758, EPI_ISL_2034799, EPI_ISL_2034815, EPI_ISL_2034829, EPI_ISL_2035303, EPI_ISL_2035307, EPI_ISL_2035309, EPI_ISL_2035311, EPI_ISL_2035312, EPI_ISL_2035315, EPI_ISL_2035316, EPI_ISL_2035317, EPI_ISL_2035318, EPI_ISL_2035320, EPI_ISL_2035321, EPI_ISL_2035322, EPI_ISL_2035323, EPI_ISL_2035329, EPI_ISL_2035330, EPI_ISL_2035331, EPI_ISL_2035335, EPI_ISL_2035336, EPI_ISL_2035337, EPI_ISL_2035338, EPI_ISL_2035340, EPI_ISL_2035343, EPI_ISL_2035346, EPI_ISL_2035348, EPI_ISL_2035349, EPI_ISL_2035350, EPI_ISL_2035351, EPI_ISL_2035352, EPI_ISL_2035356, EPI_ISL_2035357, EPI_ISL_2035360, EPI_ISL_2035363, EPI_ISL_2035364, EPI_ISL_2132576, EPI_ISL_2132581, EPI_ISL_2132583, EPI_ISL_2132586, EPI_ISL_2132619, EPI_ISL_2132620, EPI_ISL_2132621, EPI_ISL_2132623, EPI_ISL_2132624, EPI_ISL_2132625, EPI_ISL_2132626, EPI_ISL_2132627, EPI_ISL_2132631, EPI_ISL_2140733, EPI_ISL_2140734, EPI_ISL_2140735, EPI_ISL_2140737, EPI_ISL_2140738, EPI_ISL_2140739, EPI_ISL_2140742, EPI_ISL_2140743, EPI_ISL_2140744, EPI_ISL_2140745, EPI_ISL_2140746, EPI_ISL_2140747, EPI_ISL_2140749, EPI_ISL_2140750, EPI_ISL_2140751, EPI_ISL_2140752, EPI_ISL_2140753, EPI_ISL_2140757, EPI_ISL_2140760, EPI_ISL_2140766, EPI_ISL_2140767, EPI_ISL_2140769, EPI_ISL_2140774, EPI_ISL_2140775, EPI_ISL_2140777, EPI_ISL_2140778, EPI_ISL_2140779, EPI_ISL_2140780, EPI_ISL_2140782, EPI_ISL_2140785, EPI_ISL_2140786, EPI_ISL_2140787, EPI_ISL_2140788, EPI_ISL_2140789, EPI_ISL_2140790, EPI_ISL_2140791, EPI_ISL_2140792, EPI_ISL_2140797, EPI_ISL_2187231, EPI_ISL_2187233, EPI_ISL_2187235, EPI_ISL_2187236, EPI_ISL_2187237, EPI_ISL_2187238, EPI_ISL_2187240, EPI_ISL_2187243, EPI_ISL_2187244, EPI_ISL_2187245, EPI_ISL_2187246, EPI_ISL_2187247, EPI_ISL_2187249, EPI_ISL_2187250, EPI_ISL_2187251, EPI_ISL_2187252, EPI_ISL_2187253, EPI_ISL_2187254, EPI_ISL_2187255, EPI_ISL_2187257, EPI_ISL_2187258, EPI_ISL_2187259, EPI_ISL_2187260, EPI_ISL_2187261, EPI_ISL_2187262, EPI_ISL_2187263, EPI_ISL_2187265, EPI_ISL_2187267, EPI_ISL_2187268, EPI_ISL_2187269, EPI_ISL_2187270, EPI_ISL_2187273, EPI_ISL_2187275, EPI_ISL_2187278, EPI_ISL_2187279, EPI_ISL_2187280, EPI_ISL_2187281, EPI_ISL_2187282, EPI_ISL_2187284, EPI_ISL_2187285, EPI_ISL_2187288, EPI_ISL_2187289, EPI_ISL_2187290, EPI_ISL_2187291, EPI_ISL_2187293, EPI_ISL_2187296, EPI_ISL_2187299, EPI_ISL_2187300, EPI_ISL_2187301, EPI_ISL_2187303, EPI_ISL_2187305, EPI_ISL_2187306, EPI_ISL_2187309, EPI_ISL_2187312, EPI_ISL_2187313, EPI_ISL_2187314, EPI_ISL_2241627, EPI_ISL_2241629, EPI_ISL_2241630, EPI_ISL_2241633, EPI_ISL_2241635, EPI_ISL_2241636, EPI_ISL_2241637, EPI_ISL_2241638, EPI_ISL_2241640, EPI_ISL_2268144, EPI_ISL_2268146, EPI_ISL_2268148, EPI_ISL_2268152, EPI_ISL_2268159, EPI_ISL_2268161, EPI_ISL_2268164, EPI_ISL_2268166, EPI_ISL_2268169, EPI_ISL_2268171, EPI_ISL_2268174, EPI_ISL_2268178, EPI_ISL_2268181, EPI_ISL_2268184, EPI_ISL_2268186, EPI_ISL_2268189, EPI_ISL_2268271, EPI_ISL_2268275, EPI_ISL_2268278, EPI_ISL_2268282, EPI_ISL_2268287, EPI_ISL_2268297, EPI_ISL_2268299, EPI_ISL_2268302, EPI_ISL_2268304, EPI_ISL_2268307, EPI_ISL_2268311, EPI_ISL_2268314, EPI_ISL_2268321, EPI_ISL_2268323, EPI_ISL_2268325, EPI_ISL_2268328, EPI_ISL_2268331, EPI_ISL_2268335, EPI_ISL_2268366, EPI_ISL_2268367, EPI_ISL_2268368, EPI_ISL_2268369, EPI_ISL_2268370, EPI_ISL_2268374, EPI_ISL_2268375, EPI_ISL_2268379, EPI_ISL_2268380, EPI_ISL_2268381, EPI_ISL_2268382, EPI_ISL_2268383, EPI_ISL_2268384, EPI_ISL_2268387, EPI_ISL_2268388, EPI_ISL_2268390, EPI_ISL_2268391, EPI_ISL_2268392, EPI_ISL_2268393, EPI_ISL_2280082, EPI_ISL_2280084, EPI_ISL_2280085, EPI_ISL_2280086, EPI_ISL_2280088, EPI_ISL_2280090, EPI_ISL_2280091, EPI_ISL_2280092, EPI_ISL_2280093, EPI_ISL_2280094, EPI_ISL_2280095, EPI_ISL_2280096, EPI_ISL_2280097, EPI_ISL_2280099, EPI_ISL_2280100, EPI_ISL_2280101, EPI_ISL_2280102, EPI_ISL_2280103, EPI_ISL_2280104, EPI_ISL_2280106, EPI_ISL_2280108, EPI_ISL_2280110, EPI_ISL_2280111, EPI_ISL_2280112, EPI_ISL_2280113, EPI_ISL_2280117, EPI_ISL_2280118, EPI_ISL_2280120, EPI_ISL_2280121, EPI_ISL_2280123, EPI_ISL_2280124, EPI_ISL_2280125, EPI_ISL_2280127, EPI_ISL_2280128, EPI_ISL_2280129, EPI_ISL_2280130, EPI_ISL_2280131, EPI_ISL_2280132, EPI_ISL_2280133, EPI_ISL_2280134, EPI_ISL_2280135, EPI_ISL_2280136, EPI_ISL_2280137, EPI_ISL_2280139, EPI_ISL_2280143, EPI_ISL_2280144, EPI_ISL_2280145, EPI_ISL_2280146, EPI_ISL_2280152, EPI_ISL_2280153, EPI_ISL_2280155, EPI_ISL_2280157, EPI_ISL_2280158, EPI_ISL_2280159, EPI_ISL_2280160, EPI_ISL_2280161, EPI_ISL_2280163, EPI_ISL_2280167, EPI_ISL_2280168, EPI_ISL_2280169, EPI_ISL_2280171, EPI_ISL_2280172, EPI_ISL_2280173, EPI_ISL_2280174, EPI_ISL_2307143, EPI_ISL_2307146, EPI_ISL_2307149, EPI_ISL_2307150, EPI_ISL_2307152, EPI_ISL_2307153, EPI_ISL_2307154, EPI_ISL_2307155, EPI_ISL_2307156, EPI_ISL_2307157, EPI_ISL_2307158, EPI_ISL_2307160, EPI_ISL_2307161, EPI_ISL_2307162, EPI_ISL_2307163, EPI_ISL_2307164, EPI_ISL_2371691, EPI_ISL_2371692, EPI_ISL_2371693, EPI_ISL_2371694, EPI_ISL_2371697, EPI_ISL_2371698, EPI_ISL_2371700, EPI_ISL_2371702, EPI_ISL_2371703, EPI_ISL_2371706, EPI_ISL_2371707, EPI_ISL_2371708, EPI_ISL_2371709, EPI_ISL_2371710, EPI_ISL_2371711, EPI_ISL_2371714, EPI_ISL_2371715, EPI_ISL_2371717, EPI_ISL_2371719, EPI_ISL_2371720, EPI_ISL_2371722, EPI_ISL_2371723, EPI_ISL_2371724, EPI_ISL_2371725, EPI_ISL_2371726, EPI_ISL_2371727, EPI_ISL_2371729, EPI_ISL_2371730, EPI_ISL_2371732, EPI_ISL_2371734, EPI_ISL_2371735, EPI_ISL_2372410, EPI_ISL_2372411, EPI_ISL_2372412, EPI_ISL_2372413, EPI_ISL_2372414, EPI_ISL_2372415, EPI_ISL_2372416, EPI_ISL_2372417, EPI_ISL_2372418, EPI_ISL_2372419, EPI_ISL_2372420, EPI_ISL_2372421, EPI_ISL_2372422, EPI_ISL_2372423, EPI_ISL_2372900, EPI_ISL_2372902, EPI_ISL_2372903, EPI_ISL_2372904, EPI_ISL_2372905, EPI_ISL_2372906, EPI_ISL_2372908, EPI_ISL_2372909, EPI_ISL_2372911, EPI_ISL_2372912, EPI_ISL_2372914, EPI_ISL_2372915, EPI_ISL_2372916, EPI_ISL_2372919, EPI_ISL_2372920, EPI_ISL_2372921, EPI_ISL_2372922, EPI_ISL_2372923, EPI_ISL_2372924, EPI_ISL_2372927, EPI_ISL_2372928, EPI_ISL_2372929, EPI_ISL_2372930, EPI_ISL_2372932, EPI_ISL_2372933, EPI_ISL_2372934, EPI_ISL_2372936, EPI_ISL_2372937, EPI_ISL_2372938, EPI_ISL_2372939, EPI_ISL_2372940, EPI_ISL_2372942, EPI_ISL_2372943, EPI_ISL_2372945, EPI_ISL_2372946, EPI_ISL_2372948, EPI_ISL_2399463, EPI_ISL_2399464, EPI_ISL_2399467, EPI_ISL_2399468, EPI_ISL_2399469, EPI_ISL_2399471, EPI_ISL_2399472, EPI_ISL_2399473, EPI_ISL_2399474, EPI_ISL_2399475, EPI_ISL_2399476, EPI_ISL_2399477, EPI_ISL_2399480, EPI_ISL_2399481, EPI_ISL_2399482, EPI_ISL_2399483, EPI_ISL_2399485, EPI_ISL_2399487, EPI_ISL_2399488, EPI_ISL_2399490, EPI_ISL_2399491, EPI_ISL_2399493, EPI_ISL_2399495, EPI_ISL_2399496, EPI_ISL_2399499, EPI_ISL_2399500, EPI_ISL_2399503, EPI_ISL_2399504, EPI_ISL_2399505, EPI_ISL_2399507, EPI_ISL_2399509, EPI_ISL_2399510, EPI_ISL_2399511, EPI_ISL_2439774, EPI_ISL_2451404, EPI_ISL_2451405, EPI_ISL_2451408, EPI_ISL_2451410, EPI_ISL_2451411, EPI_ISL_2451413, EPI_ISL_2451415, EPI_ISL_2451417, EPI_ISL_2451418, EPI_ISL_2451419, EPI_ISL_2451420, EPI_ISL_2451422, EPI_ISL_2451424, EPI_ISL_2451425, EPI_ISL_2451427, EPI_ISL_2451429, EPI_ISL_2451430, EPI_ISL_2451431, EPI_ISL_2451432, EPI_ISL_2451434, EPI_ISL_2451435, EPI_ISL_2451436, EPI_ISL_2451437, EPI_ISL_2451438, EPI_ISL_2451439, EPI_ISL_2451440, EPI_ISL_2451443, EPI_ISL_2451445, EPI_ISL_2451446, EPI_ISL_2451447, EPI_ISL_2451448, EPI_ISL_2451449, EPI_ISL_2451451, EPI_ISL_2451452, EPI_ISL_2451453, EPI_ISL_2451454, EPI_ISL_2451455, EPI_ISL_2451456, EPI_ISL_2451457, EPI_ISL_2451458, EPI_ISL_2451459, EPI_ISL_2451460, EPI_ISL_2451461, EPI_ISL_2451462, EPI_ISL_2451468, EPI_ISL_2451469, EPI_ISL_2451471, EPI_ISL_2451472, EPI_ISL_2451474, EPI_ISL_2451475, EPI_ISL_2451477, EPI_ISL_2451479, EPI_ISL_2451480, EPI_ISL_2451481, EPI_ISL_2451484, EPI_ISL_2451485, EPI_ISL_2451486, EPI_ISL_2451488, EPI_ISL_2451489, EPI_ISL_2451507, EPI_ISL_2451508, EPI_ISL_2451510, EPI_ISL_2451512, EPI_ISL_2451513, EPI_ISL_2451514, EPI_ISL_2451515, EPI_ISL_2451516, EPI_ISL_2451519, EPI_ISL_2451521, EPI_ISL_2451522, EPI_ISL_2451523, EPI_ISL_2451526, EPI_ISL_2451528, EPI_ISL_2451531, EPI_ISL_2451532, EPI_ISL_2451539, EPI_ISL_2451545, EPI_ISL_2451547, EPI_ISL_2451548, EPI_ISL_2451553, EPI_ISL_2451554, EPI_ISL_2451556, EPI_ISL_2451560, EPI_ISL_2451561, EPI_ISL_2451562 | | | |
| see above | Quest Diagnostics Incorporated | Centers for Disease Control and Prevention Division of Viral Diseases, Pathogen Discovery | A. Gerasimova; A. Perez; Adrian Paskey; B. Anderson; Benjamin Rambo-Martin; Christopher Gulvick; Clinton R. Paden; Dakota Howard; Darlene Wagner; Dhwani Batra; Duncan MacCannell; F. Lacbawan; I. A. Shlyakhter; Jason Caravas; K.E. Livingston; Kara Moser; L.E. Bernstein; M. Hua; Matthew Schmerer; P. Tanpaiboon; Peter W. Cook; R. M. Kagan; R. Owen; R. V. Rolando; S. H. Rosenthal; Scott Sammons; Shatavia Morrison; Y. Liu; Yvette Unoarumhi |
| EPI_ISL_2105113, EPI_ISL_2105129, EPI_ISL_2105133, EPI_ISL_2105144, EPI_ISL_2384137, EPI_ISL_2384147, EPI_ISL_2384148, EPI_ISL_2384149, EPI_ISL_2452233, EPI_ISL_2452241, EPI_ISL_2452242, EPI_ISL_2452256, EPI_ISL_2452268 | | | |
| see above | RI State Health Laboratories | Centers for Disease Control and Prevention Division of Viral Diseases, Pathogen Discovery | Alison Laufer Halpin; Ben L. Rambo-Martin; Clinton R. Paden; Dakota Howard; Darlene Wagner; Dave Wentworth; Dhwani Batra; Jasmine Padilla; Justin Lee; Katie Dillon; Krista Queen; Kristen Knipe; Kristine Lacek; Mark Burroughs; Matthew Schmerer; Mili Sheth; Peter Cook; Sam Shepard; Sarah Nobles; Shoshona Le; Suxiang Tong; Vivien Dugan; Yvette Unoarumhi |
| EPI_ISL_2004447 | Randox Laboratories | Wellcome Sanger Institute for the COVID-19 Genomics UK (COG-UK) Consortium | Cordelia Langford; David K. Jackson; Dominic Kwiatkowski; Ewan Harrison; Ian Johnston; Jeffrey Barrett; John Sillitoe on behalf of the Wellcome Sanger Institute COVID-19 Surveillance Team; Randox Laboratories and Alex Alderton; Roberto Amato; Sonia Goncalves |
| EPI_ISL_2319845, EPI_ISL_2319865, EPI_ISL_2319885, EPI_ISL_2319895, EPI_ISL_2319910, EPI_ISL_2319911, EPI_ISL_2319915, EPI_ISL_2319919, EPI_ISL_2319935, EPI_ISL_2319943, EPI_ISL_2319946, EPI_ISL_2319948, EPI_ISL_2319956, EPI_ISL_2319957, EPI_ISL_2319959, EPI_ISL_2319960, EPI_ISL_2319963, EPI_ISL_2319964, EPI_ISL_2319965, EPI_ISL_2319967, EPI_ISL_2319969, EPI_ISL_2319975, EPI_ISL_2319976, EPI_ISL_2319977, EPI_ISL_2474032, EPI_ISL_2474062, EPI_ISL_2474066, EPI_ISL_2474069, EPI_ISL_2474070, EPI_ISL_2474072, EPI_ISL_2474074, EPI_ISL_2474076, EPI_ISL_2474077, EPI_ISL_2474078, EPI_ISL_2474083, EPI_ISL_2474096, EPI_ISL_2474106, EPI_ISL_2474126, EPI_ISL_2474136, EPI_ISL_2474151, EPI_ISL_2474172, EPI_ISL_2474173, EPI_ISL_2474176, EPI_ISL_2474177, EPI_ISL_2662183, EPI_ISL_2662190, EPI_ISL_2662191, EPI_ISL_2662195, EPI_ISL_2662197, EPI_ISL_2662202, EPI_ISL_2662204, EPI_ISL_2662205, EPI_ISL_2662221, EPI_ISL_2662226, EPI_ISL_2662234, EPI_ISL_2662241, EPI_ISL_2662253, EPI_ISL_2662255, EPI_ISL_2662262, EPI_ISL_2662265, EPI_ISL_2662266, EPI_ISL_2662267, EPI_ISL_2662271, EPI_ISL_2662273, EPI_ISL_2662280, EPI_ISL_2662282, EPI_ISL_2662283, EPI_ISL_2662284 | | | |
| see above | Reditus Laboratories | Reditus Laboratories | Alexa Eichelberger; Cassy Philips; Joshua J. Geltz; M.S.; Ph.D.; Rex Dyer; Robert M. Sgambelluri |
| EPI_ISL_3776073, EPI_ISL_3777139, EPI_ISL_3777148 | Respiratory Virus Unit, Microbiology Services Colindale, Public Health England | COVID-19 Genomics UK (COG-UK) Consortium | PHE Covid Sequencing Team |
| EPI_ISL_2106026, EPI_ISL_2106030, EPI_ISL_2106049, EPI_ISL_2106100, EPI_ISL_2106103, EPI_ISL_2253127, EPI_ISL_2253128, EPI_ISL_2253129, EPI_ISL_2253130, EPI_ISL_2253131, EPI_ISL_2253132, EPI_ISL_2253133, EPI_ISL_2253135, EPI_ISL_2253136, EPI_ISL_2253139, EPI_ISL_2253141, EPI_ISL_2253142, EPI_ISL_2253144, EPI_ISL_2253145, EPI_ISL_2253147, EPI_ISL_2253151, EPI_ISL_2253154, EPI_ISL_2253156, EPI_ISL_2253158, EPI_ISL_2253159, EPI_ISL_2253161, EPI_ISL_2253162, EPI_ISL_2254274, EPI_ISL_2254320, EPI_ISL_2254331, EPI_ISL_2254351, EPI_ISL_2254358, EPI_ISL_2254370, EPI_ISL_2254375, EPI_ISL_2254377, EPI_ISL_2254457, EPI_ISL_2254461, EPI_ISL_2254467, EPI_ISL_2254484 | | | |
| see above | Respiratory Viruses Branch, Centers for Disease Control and Prevention | Respiratory Viruses Branch, Centers for Disease Control and Prevention | Anderson, B.; Baker; Bakhash; Batra, D.; Bernstein, L.; Breit, N.; C.R.; Campbell, P.; Caravas, J.; Case, R.; Clark, C.; Cook; Ellis, S.; Gerasimova, A.; Ghorpade, V.; Greninger, A.; Gulvick, C.; Hardison, M.; Houdeshell, H.; Howard, D.; Hua, M.; Huang; I.A.; Jerome; K.R.; Kagan; Kvalvaag, O.; Lacbawan, F.; Lin, M.; Liu, Y.; Livingston; Livingston, K.; M.-L.; M.S.; MacCannell, D.; Mathias, P.; Morrison, S.; Moser, K.; N.R.; Nall, D.; Nguyen; Owen, R.; P.W.; Paden; Paskey, A.; Perez, A.; Perez, R.; R.J.; R.M.; R.V.; Rambo-Martin, B.; Rolando; Rosenthal; Roychoudhury; S.H.; S.M.; Sammons, S.; Sanders, E.; Schmerer, M.; Shlyakhter; Shrestha, L.; T.V.; Tanpaiboon, P.; Unoarumhi, Y.; Vest, A.; Wagner, D.; Westlund, S.; Xie, H. |
| EPI_ISL_2016490, EPI_ISL_2016491, EPI_ISL_2016504, EPI_ISL_2016515, EPI_ISL_2016516, EPI_ISL_2016523, EPI_ISL_2016554, EPI_ISL_2016556, EPI_ISL_2016581, EPI_ISL_2016597, EPI_ISL_2133808, EPI_ISL_2133814, EPI_ISL_2133879, EPI_ISL_2133886, EPI_ISL_2133888, EPI_ISL_2133897, EPI_ISL_2133911, EPI_ISL_2133926, EPI_ISL_2232415, EPI_ISL_2232434, EPI_ISL_2232435, EPI_ISL_2232439, EPI_ISL_2232443, EPI_ISL_2232444, EPI_ISL_2232445, EPI_ISL_2232451, EPI_ISL_2232530, EPI_ISL_2232532, EPI_ISL_2232537, EPI_ISL_2232538, EPI_ISL_2362382, EPI_ISL_2362390, EPI_ISL_2362397, EPI_ISL_2362403, EPI_ISL_2362406, EPI_ISL_2362416, EPI_ISL_2362417, EPI_ISL_2362440 | | | |
| see above | Rhode Island Department of Health | Infectious Disease Program, Broad Institute of Harvard and MIT | Adams, G.; Azevedo, K.; B.L.; B.W.; Bauer, M.; Birren; Carter, A.; Chaluvadi, S.; D.J.; DeRuff, K.; Gladden-Young, A.; Huard, R.; J.E.; K.J.; King, E.; Lagerborg, K.; Lemieux; Loreth, C.; Miller, A.; Normandin, E.; P.C.; Park; Pearlman, L.; Reilly, S.; Rudy, M.; Sabeti; Siddle; Tomkins-Tinch, C.; and MacInnis |
| EPI_ISL_2604798, EPI_ISL_2604813, EPI_ISL_2604861 | Rhode Island State Health Laboratories | Rhode Island State Health Laboratories | Ewa King; Kristin Carpenter-Azevedo; Richard C. Huard |
| EPI_ISL_2425504, EPI_ISL_2425519, EPI_ISL_2425522, EPI_ISL_2426847, EPI_ISL_2426848, EPI_ISL_2426849, EPI_ISL_2725578 | | | |
| see above | Rhode Island State Health Laboratory | Rhode Island State Health Laboratory | Ewa King; Kristin Carpenter-Azevedo; Richard C. Huard |
| EPI_ISL_2691916, EPI_ISL_2691926, EPI_ISL_2691936, EPI_ISL_2691937, EPI_ISL_2691939, EPI_ISL_2691940, EPI_ISL_2691944, EPI_ISL_2691948, EPI_ISL_2691952 | | | |
| see above | Rush University Medical Center | RIPHL at Rush University Medical Center | Diane Springer; Felix Araujo Perez; Joyce Houlihan; Kevin Kunstman; Laura Furtado; Marieta Hyde; Mary Hayden; Max Kolton; Stefan Green |
| EPI_ISL_2445155, EPI_ISL_2445156, EPI_ISL_2445159, EPI_ISL_2445161, EPI_ISL_2445162, EPI_ISL_2445163, EPI_ISL_2445166, EPI_ISL_2445170, EPI_ISL_2445174, EPI_ISL_2445409, EPI_ISL_2445411, EPI_ISL_2445413, EPI_ISL_2473708, EPI_ISL_2473709 | | | |
| see above | SAE CTA SERV ATEND ESPEC TESTAGEM E ACONSELHAMENTO | Instituto Butantan | Antonio Jorge Martins; Claudia Renata dos Santos Barros; David Schlesinger; Debora Botequio Moretti; Dimas Tadeu Covas; Elaine Cristina Marqueze; Elaine Vieira Santos; Evandra Strazza Rodrigues; Heidge Fukumasu; Jayme Augusto de Souza-Neto; José Salvatore Leister Patané; Luiz Alcantara; Luiz Lehmann Coutinho; Maria Carolina Elias; Maurício Lacerda Nogueira; Rafael dos Santos Bezerra; Raul Machado Neto; Rejane Maria Tommasini Grotto; Ricardo Haddad; Sandra Coccuzzo Sampaio Vessoni; Simone Kashima; Svetoslav Nanev Slavov; Vincent Louis Viala |
| EPI_ISL_2344802, EPI_ISL_2344803, EPI_ISL_2344804, EPI_ISL_2344805, EPI_ISL_2344810, EPI_ISL_2344811, EPI_ISL_2344812, EPI_ISL_2344813, EPI_ISL_2344815, EPI_ISL_2344816, EPI_ISL_2344817, EPI_ISL_2344819 | | | |
| see above | SAE CTA SERV ATEND ESPEC TESTAGEM E ACONSELHAMENTO | Instituto Butantan / ESALQ-Piracicaba | Antonio Jorge Martins; Claudia Renata dos Santos Barros; David Schlesinger; Debora Botequio Moretti; Dimas Tadeu Covas; Elaine Cristina Marqueze; Elaine Vieira Santos; Evandra Strazza Rodrigues; Heidge Fukumasu; Jayme Augusto de Souza-Neto; José Salvatore Leister Patané; Luiz Alcantara; Luiz Lehmann Coutinho; Maria Carolina Elias; Maurício Lacerda Nogueira; Rafael dos Santos Bezerra; Raul Machado Neto; Rejane Maria Tommasini Grotto; Ricardo Haddad; Sandra Coccuzzo Sampaio Vessoni; Simone Kashima; Svetoslav Nanev Slavov; Vincent Louis Viala |
| EPI_ISL_2445139, EPI_ISL_2445140, EPI_ISL_2445141, EPI_ISL_2473755 | SANTA CASA DE DOIS CORREGOS | Instituto Butantan | Antonio Jorge Martins; Claudia Renata dos Santos Barros; David Schlesinger; Debora Botequio Moretti; Dimas Tadeu Covas; Elaine Cristina Marqueze; Elaine Vieira Santos; Evandra Strazza Rodrigues; Heidge Fukumasu; Jayme Augusto de Souza-Neto; José Salvatore Leister Patané; Luiz Alcantara; Luiz Lehmann Coutinho; Maria Carolina Elias; Maurício Lacerda Nogueira; Rafael dos Santos Bezerra; Raul Machado Neto; Rejane Maria Tommasini Grotto; Ricardo Haddad; Sandra Coccuzzo Sampaio Vessoni; Simone Kashima; Svetoslav Nanev Slavov; Vincent Louis Viala |
| EPI_ISL_2345037 | SANTA CASA DE GUARAREMA | Instituto Butantan | Antonio Jorge Martins; Claudia Renata dos Santos Barros; David Schlesinger; Debora Botequio Moretti; Dimas Tadeu Covas; Elaine Cristina Marqueze; Elaine Vieira Santos; Evandra Strazza Rodrigues; Heidge Fukumasu; Jayme Augusto de Souza-Neto; José Salvatore Leister Patané; Luiz Alcantara; Luiz Lehmann Coutinho; Maria Carolina Elias; Maurício Lacerda Nogueira; Rafael dos Santos Bezerra; Raul Machado Neto; Rejane Maria Tommasini Grotto; Ricardo Haddad; Sandra Coccuzzo Sampaio Vessoni; Simone Kashima; Svetoslav Nanev Slavov; Vincent Louis Viala |
| EPI_ISL_2345115, EPI_ISL_2345121, EPI_ISL_2345124 | SANTA CASA DE GUARAREMA | Instituto Butantan / ESALQ-Piracicaba | Antonio Jorge Martins; Claudia Renata dos Santos Barros; David Schlesinger; Debora Botequio Moretti; Dimas Tadeu Covas; Elaine Cristina Marqueze; Elaine Vieira Santos; Evandra Strazza Rodrigues; Heidge Fukumasu; Jayme Augusto de Souza-Neto; José Salvatore Leister Patané; Luiz Alcantara; Luiz Lehmann Coutinho; Maria Carolina Elias; Maurício Lacerda Nogueira; Rafael dos Santos Bezerra; Raul Machado Neto; Rejane Maria Tommasini Grotto; Ricardo Haddad; Sandra Coccuzzo Sampaio Vessoni; Simone Kashima; Svetoslav Nanev Slavov; Vincent Louis Viala |
| EPI_ISL_2170997 | SANTA CASA DE GUARAREMA | Instituto Butantan / Mendelics | Antonio Jorge Martins; Bianca Cechetto Carlos. Mendelics: Bibiana Santos; Claudia Renata dos Santos Barros; Cíntia Bittar; David Schlesinger. Hemocentro Ribeirão Preto: Simone Kashima; Debora Botequio Moretti; Elaine Cristina Marqueze; Elaine Vieira dos Santos; Elisangela Chicaroni Mattos; Erika Freitas; Evandra Strazza Rodrigues; Felipe Allan da Silva da Costa; Flavia Aburjaile; Fábio Sossai Possebon; Guilherme Campos; Guilherme Targino Valente; Heidge Fukumasu. USP-Botucatu: Rejane Maria Tommasini Grotto; Helena Lage Ferreira; Instituto Butantan: Dimas Tadeu Covas; Jardelina de Souza Todao Bernardino; Jayme A. Souza-Neto; Jessika Cristina Chagas Lesbon; Jorge A. Petroli Marchesi; José Salvatore Leister Patané; João Paulo Kitajima; João Pessoa Araújo Jr.; Leila Sabrina Ullmann; Loyze Paola Oliveira de Lima; Luiz Aurelio de Campos Crispin. Centro de Genômica Funcional da ESALQ: Luiz Lehmann Coutinho; Luiz Carlos Junior de Alcantara; Lívia Sacchetto; Maisa C. Pereira Parra; Maria Carolina Elias; Marta Giovanetti; Marília Moraes; Maurício Lacerda Nogueira. Prefeitura de Sao Paulo: Melissa Palmieri.; Patricia Akemi Assato; Paula Rahal; Paulo Inacio da Costa; Rafael dos Santos Bezerra; Raquel de Lello Rocha Campos Cassano. NGS Soluções Genômicas: Pilar Drummond Sampaio Corrêa Mariani. FZEA-USP Pirassununga: Mirele Daiana Poleti; Raul Machado Neto; Ricardo Augusto Brassaloti; Ricardo Haddad; Rodrigo Tocantins Calado. FAMERP-SJRP: Cecília Artico Banho; Sandra Coccuzzo Sampaio; Svetoslav Nanev Slavov; Vagner Fonseca; Vincent Louis Viala |
| EPI_ISL_2344865, EPI_ISL_2344876, EPI_ISL_2344878, EPI_ISL_2344880, | SANTA CASA DE JOSE BONIFACIO | Instituto Butantan / ESALQ-Piracicaba | Antonio Jorge Martins; Claudia Renata dos Santos Barros; David Schlesinger; Debora Botequio Moretti; Dimas Tadeu Covas; Elaine Cristina Marqueze; Elaine Vieira Santos; Evandra Strazza Rodrigues; Heidge Fukumasu; Jayme Augusto de Souza-Neto; José Salvatore Leister Patané; Luiz Alcantara; Luiz Lehmann Coutinho; Maria Carolina Elias; Maurício Lacerda Nogueira; Rafael dos Santos Bezerra; Raul Machado Neto; Rejane Maria Tommasini Grotto; Ricardo Haddad; Sandra Coccuzzo Sampaio Vessoni; Simone Kashima; Svetoslav Nanev Slavov; Vincent Louis Viala |

| | | | |
|---|---|---|---|
| EPI_ISL_2344883 | | | |
| EPI_ISL_2493681 | SANTA CASA DE MISERICORDIA DE TIETE | Instituto Butantan | Antonio Jorge Martins; Claudia Renata dos Santos Barros; David Schlesinger; Debora Botequio Moretti; Dimas Tadeu Covas; Elaine Cristina Marqueze; Elaine Vieira Santos; Evandra Strazza Rodrigues; Heidge Fukumasu; Jayme Augusto de Souza-Neto; José Salvatore Leister Patané; Luiz Alcantara; Luiz Lehmann Coutinho; Maria Carolina Elias; Maurício Lacerda Nogueira; Rafael dos Santos Bezerra; Raul Machado Neto; Rejane Maria Tommasini Grotto; Ricardo Haddad; Sandra Coccuzzo Sampaio Vessoni; Simone Kashima; Svetoslav Nanev Slavov; Vincent Louis Viala |
| EPI_ISL_2209872, EPI_ISL_2210133 | SANTA CASA DE MISERICORDIA DE UBATUBA | Instituto Butantan / Mendelics | Antonio Jorge Martins; Claudia Renata dos Santos Barros; David Schlesinger; Debora Botequio Moretti; Dimas Tadeu Covas; Elaine Cristina Marqueze; Elaine Vieira Santos; Evandra Strazza Rodrigues; Heidge Fukumasu; Jayme Augusto de Souza-Neto; José Salvatore Leister Patané; Luiz Alcantara; Luiz Lehmann Coutinho; Maria Carolina Elias; Maurício Lacerda Nogueira; Rafael dos Santos Bezerra; Raul Machado Neto; Rejane Maria Tommasini Grotto; Ricardo Haddad; Sandra Coccuzzo Sampaio Vessoni; Simone Kashima; Svetoslav Nanev Slavov; Vincent Louis Viala |
| EPI_ISL_2209108, EPI_ISL_2345004, EPI_ISL_2345007, EPI_ISL_2345009, EPI_ISL_2345010, EPI_ISL_2345012 | SANTA CASA DE MISERICORDIA SAO JOSE | Instituto Butantan / ESALQ-Piracicaba | Antonio Jorge Martins; Claudia Renata dos Santos Barros; David Schlesinger; Debora Botequio Moretti; Dimas Tadeu Covas; Elaine Cristina Marqueze; Elaine Vieira Santos; Evandra Strazza Rodrigues; Heidge Fukumasu; Jayme Augusto de Souza-Neto; José Salvatore Leister Patané; Luiz Alcantara; Luiz Lehmann Coutinho; Maria Carolina Elias; Maurício Lacerda Nogueira; Rafael dos Santos Bezerra; Raul Machado Neto; Rejane Maria Tommasini Grotto; Ricardo Haddad; Sandra Coccuzzo Sampaio Vessoni; Simone Kashima; Svetoslav Nanev Slavov; Vincent Louis Viala |
| EPI_ISL_2209888, EPI_ISL_2210250, EPI_ISL_2345682, EPI_ISL_2345727, EPI_ISL_2345739, EPI_ISL_2345783, EPI_ISL_2362269 | | | |
| see above | SANTA CASA DE MISERICORDIA SAO JOSE | Instituto Butantan / Mendelics | Antonio Jorge Martins; Claudia Renata dos Santos Barros; David Schlesinger; Debora Botequio Moretti; Dimas Tadeu Covas; Elaine Cristina Marqueze; Elaine Vieira Santos; Evandra Strazza Rodrigues; Heidge Fukumasu; Jayme Augusto de Souza-Neto; José Salvatore Leister Patané; Luiz Alcantara; Luiz Lehmann Coutinho; Maria Carolina Elias; Maurício Lacerda Nogueira; Rafael dos Santos Bezerra; Raul Machado Neto; Rejane Maria Tommasini Grotto; Ricardo Haddad; Sandra Coccuzzo Sampaio Vessoni; Simone Kashima; Svetoslav Nanev Slavov; Vincent Louis Viala |
| EPI_ISL_2344891, EPI_ISL_2344892, EPI_ISL_2344894 | SANTA CASA DE PORTO FELIZ | Instituto Butantan / ESALQ-Piracicaba | Antonio Jorge Martins; Claudia Renata dos Santos Barros; David Schlesinger; Debora Botequio Moretti; Dimas Tadeu Covas; Elaine Cristina Marqueze; Elaine Vieira Santos; Evandra Strazza Rodrigues; Heidge Fukumasu; Jayme Augusto de Souza-Neto; José Salvatore Leister Patané; Luiz Alcantara; Luiz Lehmann Coutinho; Maria Carolina Elias; Maurício Lacerda Nogueira; Rafael dos Santos Bezerra; Raul Machado Neto; Rejane Maria Tommasini Grotto; Ricardo Haddad; Sandra Coccuzzo Sampaio Vessoni; Simone Kashima; Svetoslav Nanev Slavov; Vincent Louis Viala |
| EPI_ISL_2444154, EPI_ISL_2444157, EPI_ISL_2444159, EPI_ISL_2444160, EPI_ISL_2444161, EPI_ISL_2444163, EPI_ISL_2444164, EPI_ISL_2444166, EPI_ISL_2444167, EPI_ISL_2444168, EPI_ISL_2444169, EPI_ISL_2444171, EPI_ISL_2444174, EPI_ISL_2444176, EPI_ISL_2444178, EPI_ISL_2444179, EPI_ISL_2444180, EPI_ISL_2444182, EPI_ISL_2444185, EPI_ISL_2444187, EPI_ISL_2444188, EPI_ISL_2444190, EPI_ISL_2444193, EPI_ISL_2444196, EPI_ISL_2444197, EPI_ISL_2444198, EPI_ISL_2444200, EPI_ISL_2444201, EPI_ISL_2444204, EPI_ISL_2444205, EPI_ISL_2444206, EPI_ISL_2444207, EPI_ISL_2444209, EPI_ISL_2444211, EPI_ISL_2444213, EPI_ISL_2444214, EPI_ISL_2444215, EPI_ISL_2444216, EPI_ISL_2444218, EPI_ISL_2444219, EPI_ISL_2444220, EPI_ISL_2444221, EPI_ISL_2444222, EPI_ISL_2455396, EPI_ISL_2455397 | | | |
| see above | SAO JOSE DO RIO PRETO | Instituto Butantan / FAMERP | Antonio Jorge Martins; Claudia Renata dos Santos Barros; David Schlesinger; Debora Botequio Moretti; Dimas Tadeu Covas; Elaine Cristina Marqueze; Elaine Vieira Santos; Evandra Strazza Rodrigues; Heidge Fukumasu; Jayme Augusto de Souza-Neto; José Salvatore Leister Patané; Luiz Alcantara; Luiz Lehmann Coutinho; Maria Carolina Elias; Maurício Lacerda Nogueira; Rafael dos Santos Bezerra; Raul Machado Neto; Rejane Maria Tommasini Grotto; Ricardo Haddad; Sandra Coccuzzo Sampaio Vessoni; Simone Kashima; Svetoslav Nanev Slavov; Vincent Louis Viala |
| EPI_ISL_2098626 | SARATOGA HOSPITAL LABORATORY | Wadsworth Center, New York State Department of Health | Alexis Russell; Catharine Prussing; Daryl M. Lamson; Erasmus Schneider; Erica Lasek-Nesselquist; John Kelly; Jonathan Plitnick; Kirsten St. George; Matthew Shudt; Melissa A Leisner; Navjot Singh |
| EPI_ISL_2791991, EPI_ISL_2986088 | SARS-CoV-2 Sequencing Castilla y Leon-Spain Consortium | SARS-CoV-2 Sequencing Castilla y Leon-Spain Consortium | Antonio Orduña-Domingo; Carlos Fuster Foz; Carmen Aldea-Mansilla; Carmen Gimeno Crespo; David Abad; Gabriel March Rossello; Gregoria Megías Lobón; Jose María Eiros Bouza; Laura Sánchez de Prada; M. Isabel Fernandez-Natal; Marta Dominguez-Gil; Marta Hernandez; María Antonia García Castro; Mª Fe Brezmes-Valdivieso; Noelia Arenal Andrés; Silvia Rojo; Sonsoles Garcinuño Pérez |
| EPI_ISL_2131666, EPI_ISL_2336130, EPI_ISL_2336133, EPI_ISL_2337801, EPI_ISL_2447611, EPI_ISL_2447612 | SARS-CoV-2 testing team, National Institute of Infectious Diseases | Pathogen Genomics Center, National Institute of Infectious Diseases | Hazuka Y Furihata; Kentaro Itokawa; Makoto Kuroda; Masanori Hashino; Masumichi Saito; Naomi Nojiri; Nozomu Hanaoka; Rina Tanaka; Sana Uchikoba; Tsuguto Fujimoto; Tsuyoshi Sekizuka |
| EPI_ISL_2018303 | SC Dept of Health and Env. Control-Bureau of Laboratories | Centers for Disease Control and Prevention Division of Viral Diseases, Pathogen Discovery | Alison Laufer Halpin; Ben L. Rambo-Martin; Clinton R. Paden; Dakota Howard; Darlene Wagner; Dave Wentworth; Dhwani Batra; Jasmine Padilla; Justin Lee; Katie Dillon; Krista Queen; Kristen Knipe; Kristine Lacek; Mark Burroughs; Matthew Schmerer; Mili Sheth; Peter Cook; Sam Shepard; Sarah Nobles; Shoshona Le; Suxiang Tong; Vivien Dugan; Yvette Unoarumhi |
| EPI_ISL_3355017 | SD Public Health Laboratory | Centers for Disease Control and Prevention Division of Viral Diseases, Pathogen Discovery | Alex Burgin; Ben Rambo-Martin; Clinton Paden; Dakota Howard; Dave Wentworth; Dhwani Batra; Jasmine Padilla; Justin Lee; Krista Queen; Kristen Knipe; Kristine Lacek; Mark Burroughs; Matthew Schmerer; Meghan Bentz; Mili Sheth; Peter Cook; Sam Shepard; Sarah Nobles; Suxiang Tong; Vivien Dugan; Yvette Unoarumhi |
| EPI_ISL_2345050, EPI_ISL_2345051 | SECAO CENTRO DE DIAGNOSTICO SECEDI | Instituto Butantan / ESALQ-Piracicaba | Antonio Jorge Martins; Claudia Renata dos Santos Barros; David Schlesinger; Debora Botequio Moretti; Dimas Tadeu Covas; Elaine Cristina Marqueze; Elaine Vieira Santos; Evandra Strazza Rodrigues; Heidge Fukumasu; Jayme Augusto de Souza-Neto; José Salvatore Leister Patané; Luiz Alcantara; Luiz Lehmann Coutinho; Maria Carolina Elias; Maurício Lacerda Nogueira; Rafael dos Santos Bezerra; Raul Machado Neto; Rejane Maria Tommasini Grotto; Ricardo Haddad; Sandra Coccuzzo Sampaio Vessoni; Simone Kashima; Svetoslav Nanev Slavov; Vincent Louis Viala |
| EPI_ISL_2345667, EPI_ISL_2345854 | SECAO CENTRO DE DIAGNOSTICO SECEDI | Instituto Butantan / Mendelics | Antonio Jorge Martins; Claudia Renata dos Santos Barros; David Schlesinger; Debora Botequio Moretti; Dimas Tadeu Covas; Elaine Cristina Marqueze; Elaine Vieira Santos; Evandra Strazza Rodrigues; Heidge Fukumasu; Jayme Augusto de Souza-Neto; José Salvatore Leister Patané; Luiz Alcantara; Luiz Lehmann Coutinho; Maria Carolina Elias; Maurício Lacerda Nogueira; Rafael dos Santos Bezerra; Raul Machado Neto; Rejane Maria Tommasini Grotto; Ricardo Haddad; Sandra Coccuzzo Sampaio Vessoni; Simone Kashima; Svetoslav Nanev Slavov; Vincent Louis Viala |
| EPI_ISL_2493621, EPI_ISL_2493627 | SECRETARIA DE SAUDE | Instituto Butantan | Antonio Jorge Martins; Claudia Renata dos Santos Barros; David Schlesinger; Debora Botequio Moretti; Dimas Tadeu Covas; Elaine Cristina Marqueze; Elaine Vieira Santos; Evandra Strazza Rodrigues; Heidge Fukumasu; Jayme Augusto de Souza-Neto; José Salvatore Leister Patané; Luiz Alcantara; Luiz Lehmann Coutinho; Maria Carolina Elias; Maurício Lacerda Nogueira; Rafael dos Santos Bezerra; Raul Machado Neto; Rejane Maria Tommasini Grotto; Ricardo Haddad; Sandra Coccuzzo Sampaio Vessoni; Simone Kashima; Svetoslav Nanev Slavov; Vincent Louis Viala |
| EPI_ISL_2445464, EPI_ISL_2445468, EPI_ISL_2445470, EPI_ISL_2445474, EPI_ISL_2445475, EPI_ISL_2445477 | SECRETARIA DE SAUDE DE LOUVEIRA | Instituto Butantan | Antonio Jorge Martins; Claudia Renata dos Santos Barros; David Schlesinger; Debora Botequio Moretti; Dimas Tadeu Covas; Elaine Cristina Marqueze; Elaine Vieira Santos; Evandra Strazza Rodrigues; Heidge Fukumasu; Jayme Augusto de Souza-Neto; José Salvatore Leister Patané; Luiz Alcantara; Luiz Lehmann Coutinho; Maria Carolina Elias; Maurício Lacerda Nogueira; Rafael dos Santos Bezerra; Raul Machado Neto; Rejane Maria Tommasini Grotto; Ricardo Haddad; Sandra Coccuzzo Sampaio Vessoni; Simone Kashima; Svetoslav Nanev Slavov; Vincent Louis Viala |
| EPI_ISL_2494148 | SECRETARIA DE SAUDE DE RAFARD | Instituto Butantan | Antonio Jorge Martins; Claudia Renata dos Santos Barros; David Schlesinger; Debora Botequio Moretti; Dimas Tadeu Covas; Elaine Cristina Marqueze; Elaine Vieira Santos; Evandra Strazza Rodrigues; Heidge Fukumasu; Jayme Augusto de Souza-Neto; José Salvatore Leister Patané; Luiz Alcantara; Luiz Lehmann Coutinho; Maria Carolina Elias; Maurício Lacerda Nogueira; Rafael dos Santos Bezerra; Raul Machado Neto; Rejane Maria Tommasini Grotto; Ricardo Haddad; Sandra Coccuzzo Sampaio Vessoni; Simone Kashima; Svetoslav Nanev Slavov; Vincent Louis Viala |
| EPI_ISL_2493389, EPI_ISL_2493390, EPI_ISL_2493447, EPI_ISL_2494156 | SECRETARIA DE SAUDE DE SANTA GERTRUDES | Instituto Butantan | Antonio Jorge Martins; Claudia Renata dos Santos Barros; David Schlesinger; Debora Botequio Moretti; Dimas Tadeu Covas; Elaine Cristina Marqueze; Elaine Vieira Santos; Evandra Strazza Rodrigues; Heidge Fukumasu; Jayme Augusto de Souza-Neto; José Salvatore Leister Patané; Luiz Alcantara; Luiz Lehmann Coutinho; Maria Carolina Elias; Maurício Lacerda Nogueira; Rafael dos Santos Bezerra; Raul Machado Neto; Rejane Maria Tommasini Grotto; Ricardo Haddad; Sandra Coccuzzo Sampaio Vessoni; Simone Kashima; Svetoslav Nanev Slavov; Vincent Louis Viala |
| EPI_ISL_2494155 | SECRETARIA DE SAUDE DE SAO PEDRO | Instituto Butantan | Antonio Jorge Martins; Claudia Renata dos Santos Barros; David Schlesinger; Debora Botequio Moretti; Dimas Tadeu Covas; Elaine Cristina Marqueze; Elaine Vieira Santos; Evandra Strazza Rodrigues; Heidge Fukumasu; Jayme Augusto de Souza-Neto; José Salvatore Leister Patané; Luiz Alcantara; Luiz Lehmann Coutinho; Maria Carolina Elias; Maurício Lacerda Nogueira; Rafael dos Santos Bezerra; Raul Machado Neto; Rejane Maria Tommasini Grotto; Ricardo Haddad; Sandra Coccuzzo Sampaio Vessoni; Simone Kashima; Svetoslav Nanev Slavov; Vincent Louis Viala |
| EPI_ISL_2209329, EPI_ISL_2209516, EPI_ISL_2209905, EPI_ISL_2210305, EPI_ISL_2345703, EPI_ISL_2345802 | SECRETARIA DE SAUDE PUBLICA DE PRAIA GRANDE | Instituto Butantan / Mendelics | Antonio Jorge Martins; Claudia Renata dos Santos Barros; David Schlesinger; Debora Botequio Moretti; Dimas Tadeu Covas; Elaine Cristina Marqueze; Elaine Vieira Santos; Evandra Strazza Rodrigues; Heidge Fukumasu; Jayme Augusto de Souza-Neto; José Salvatore Leister Patané; Luiz Alcantara; Luiz Lehmann Coutinho; Maria Carolina Elias; Maurício Lacerda Nogueira; Rafael dos Santos Bezerra; Raul Machado Neto; Rejane Maria Tommasini Grotto; Ricardo Haddad; Sandra Coccuzzo Sampaio Vessoni; Simone Kashima; Svetoslav Nanev Slavov; Vincent Louis Viala |
| EPI_ISL_2493598, EPI_ISL_2493601, EPI_ISL_2493603, EPI_ISL_2493612 | SECRETARIA MUNICIPAL DA SAUDE DE GUARIBA | Instituto Butantan | Antonio Jorge Martins; Claudia Renata dos Santos Barros; David Schlesinger; Debora Botequio Moretti; Dimas Tadeu Covas; Elaine Cristina Marqueze; Elaine Vieira Santos; Evandra Strazza Rodrigues; Heidge Fukumasu; Jayme Augusto de Souza-Neto; José Salvatore Leister Patané; Luiz Alcantara; Luiz Lehmann Coutinho; Maria Carolina Elias; Maurício Lacerda Nogueira; Rafael dos Santos Bezerra; Raul Machado Neto; Rejane Maria Tommasini Grotto; Ricardo Haddad; Sandra Coccuzzo Sampaio Vessoni; Simone Kashima; Svetoslav Nanev Slavov; Vincent Louis Viala |
| EPI_ISL_2344896 | SECRETARIA MUNICIPAL DA SAUDE DE ITAPEVA SP | Instituto Butantan / ESALQ-Piracicaba | Antonio Jorge Martins; Claudia Renata dos Santos Barros; David Schlesinger; Debora Botequio Moretti; Dimas Tadeu Covas; Elaine Cristina Marqueze; Elaine Vieira Santos; Evandra Strazza Rodrigues; Heidge Fukumasu; Jayme Augusto de Souza-Neto; José Salvatore Leister Patané; Luiz Alcantara; Luiz Lehmann Coutinho; Maria Carolina Elias; Maurício Lacerda Nogueira; Rafael dos Santos Bezerra; Raul Machado Neto; Rejane Maria Tommasini Grotto; Ricardo Haddad; Sandra Coccuzzo Sampaio Vessoni; Simone Kashima; Svetoslav Nanev Slavov; Vincent Louis Viala |
| EPI_ISL_2445599 | SECRETARIA MUNICIPAL DA SAUDE DE ITARARE | Instituto Butantan | Antonio Jorge Martins; Claudia Renata dos Santos Barros; David Schlesinger; Debora Botequio Moretti; Dimas Tadeu Covas; Elaine Cristina Marqueze; Elaine Vieira Santos; Evandra Strazza Rodrigues; Heidge Fukumasu; Jayme Augusto de Souza-Neto; José Salvatore Leister Patané; Luiz Alcantara; Luiz Lehmann Coutinho; Maria Carolina Elias; Maurício Lacerda Nogueira; Rafael dos Santos Bezerra; Raul Machado Neto; Rejane Maria Tommasini Grotto; Ricardo Haddad; Sandra Coccuzzo Sampaio Vessoni; Simone Kashima; Svetoslav Nanev Slavov; Vincent Louis Viala |
| EPI_ISL_2344903, EPI_ISL_2344912 | SECRETARIA MUNICIPAL DA SAUDE DE ITARARE | Instituto Butantan / ESALQ-Piracicaba | Antonio Jorge Martins; Claudia Renata dos Santos Barros; David Schlesinger; Debora Botequio Moretti; Dimas Tadeu Covas; Elaine Cristina Marqueze; Elaine Vieira Santos; Evandra Strazza Rodrigues; Heidge Fukumasu; Jayme Augusto de Souza-Neto; José Salvatore Leister Patané; Luiz Alcantara; Luiz Lehmann Coutinho; Maria Carolina Elias; Maurício Lacerda Nogueira; Rafael dos Santos Bezerra; Raul Machado Neto; Rejane Maria Tommasini Grotto; Ricardo Haddad; Sandra Coccuzzo Sampaio Vessoni; Simone Kashima; Svetoslav Nanev Slavov; Vincent Louis Viala |
| EPI_ISL_2345855 | SECRETARIA MUNICIPAL DA SAUDE DE ITARARE | Instituto Butantan / Mendelics | Antonio Jorge Martins; Claudia Renata dos Santos Barros; David Schlesinger; Debora Botequio Moretti; Dimas Tadeu Covas; Elaine Cristina Marqueze; Elaine Vieira Santos; Evandra Strazza Rodrigues; Heidge Fukumasu; Jayme Augusto de Souza-Neto; José Salvatore Leister Patané; Luiz Alcantara; Luiz Lehmann Coutinho; Maria Carolina Elias; Maurício Lacerda Nogueira; Rafael dos Santos Bezerra; Raul Machado Neto; Rejane Maria Tommasini Grotto; Ricardo Haddad; Sandra Coccuzzo Sampaio Vessoni; Simone Kashima; Svetoslav Nanev Slavov; Vincent Louis Viala |
| EPI_ISL_2345875 | SECRETARIA MUNICIPAL DA SAUDE DE JOANOPOLIS | Instituto Butantan / Mendelics | Antonio Jorge Martins; Claudia Renata dos Santos Barros; David Schlesinger; Debora Botequio Moretti; Dimas Tadeu Covas; Elaine Cristina Marqueze; Elaine Vieira Santos; Evandra Strazza Rodrigues; Heidge Fukumasu; Jayme Augusto de Souza-Neto; José Salvatore Leister Patané; Luiz Alcantara; Luiz Lehmann Coutinho; Maria Carolina Elias; Maurício Lacerda Nogueira; Rafael dos Santos Bezerra; Raul Machado Neto; Rejane Maria Tommasini Grotto; Ricardo Haddad; Sandra Coccuzzo Sampaio Vessoni; Simone Kashima; Svetoslav Nanev Slavov; Vincent Louis Viala |
| EPI_ISL_2493513, EPI_ISL_2493577, EPI_ISL_2493579, EPI_ISL_2493581, EPI_ISL_2493630, EPI_ISL_2493633, EPI_ISL_2493635, EPI_ISL_2493637 | | | |
| see above | SECRETARIA MUNICIPAL DE SAUDE | Instituto Butantan | Antonio Jorge Martins; Claudia Renata dos Santos Barros; David Schlesinger; Debora Botequio Moretti; Dimas Tadeu Covas; Elaine Cristina Marqueze; Elaine Vieira Santos; Evandra Strazza Rodrigues; Heidge Fukumasu; Jayme Augusto de Souza-Neto; José Salvatore Leister Patané; Luiz Alcantara; Luiz Lehmann Coutinho; Maria Carolina Elias; Maurício Lacerda Nogueira; Rafael dos Santos Bezerra; Raul Machado Neto; Rejane Maria Tommasini Grotto; Ricardo Haddad; Sandra Coccuzzo Sampaio Vessoni; Simone Kashima; Svetoslav Nanev Slavov; Vincent Louis Viala |
| EPI_ISL_2493367, EPI_ISL_2493396, EPI_ISL_2493427, EPI_ISL_2493428, EPI_ISL_2493452, EPI_ISL_2494158 | SECRETARIA MUNICIPAL DE SAUDE DE CORDEIROPOLIS | Instituto Butantan | Antonio Jorge Martins; Claudia Renata dos Santos Barros; David Schlesinger; Debora Botequio Moretti; Dimas Tadeu Covas; Elaine Cristina Marqueze; Elaine Vieira Santos; Evandra Strazza Rodrigues; Heidge Fukumasu; Jayme Augusto de Souza-Neto; José Salvatore Leister Patané; Luiz Alcantara; Luiz Lehmann Coutinho; Maria Carolina Elias; Maurício Lacerda Nogueira; Rafael dos Santos Bezerra; Raul Machado Neto; Rejane Maria Tommasini Grotto; Ricardo Haddad; Sandra Coccuzzo Sampaio Vessoni; Simone Kashima; Svetoslav Nanev Slavov; Vincent Louis Viala |
| EPI_ISL_2344858 | SECRETARIA MUNICIPAL DE SAUDE DE CORDEIROPOLIS | Instituto Butantan / ESALQ-Piracicaba | Antonio Jorge Martins; Claudia Renata dos Santos Barros; David Schlesinger; Debora Botequio Moretti; Dimas Tadeu Covas; Elaine Cristina Marqueze; Elaine Vieira Santos; Evandra Strazza Rodrigues; Heidge Fukumasu; Jayme Augusto de Souza-Neto; José Salvatore Leister Patané; Luiz Alcantara; Luiz Lehmann Coutinho; Maria Carolina Elias; Maurício Lacerda Nogueira; Rafael dos Santos Bezerra; Raul Machado Neto; Rejane Maria Tommasini Grotto; Ricardo Haddad; Sandra Coccuzzo Sampaio Vessoni; Simone Kashima; Svetoslav Nanev Slavov; Vincent Louis Viala |
| EPI_ISL_2345116, EPI_ISL_2345117, EPI_ISL_2345125, EPI_ISL_2345237, EPI_ISL_2345238 | SECRETARIA MUNICIPAL DE SAUDE DE JARINU | Instituto Butantan / ESALQ-Piracicaba | Antonio Jorge Martins; Claudia Renata dos Santos Barros; David Schlesinger; Debora Botequio Moretti; Dimas Tadeu Covas; Elaine Cristina Marqueze; Elaine Vieira Santos; Evandra Strazza Rodrigues; Heidge Fukumasu; Jayme Augusto de Souza-Neto; José Salvatore Leister Patané; Luiz Alcantara; Luiz Lehmann Coutinho; Maria Carolina Elias; Maurício Lacerda Nogueira; Rafael dos Santos Bezerra; Raul Machado Neto; Rejane Maria Tommasini Grotto; Ricardo Haddad; Sandra Coccuzzo Sampaio Vessoni; Simone Kashima; Svetoslav Nanev Slavov; Vincent Louis Viala |
| EPI_ISL_2801338, EPI_ISL_2801339, EPI_ISL_2801340, EPI_ISL_2801351, EPI_ISL_2801353, EPI_ISL_2801354, EPI_ISL_2801356 | | | |

| | | | |
|---|---|---|---|
| see above | SECRETARIA MUNICIPAL DE SAUDE DE TIANGUA | Analytical Competence Molecular Epidemiology Lab/ACME, Oswaldo Cruz Foundation, Ceara (FIOCRUZ CE) | Cleber Furtado Aksenen e Suzana Porto Almeida; Fabio Miyajima; Fernando Braga Stehling; Francisco Eder de Moura Lopes; Jamille Maria Mendes Bezerra; Joaquim César do Nascimento Sousa Junior; Pedro Miguel Carneiro Jeronimo; Thais Ferreira de Oliveira; Thais de Oliveira Costa; Ticiane Cavalcante de Souza; Veridiana Pessoa Miyajima |
| EPI_ISL_2445386 | SECRETARIA MUNICIPAL DE SAUDE DE VALPARAISO SP | Instituto Butantan | Antonio Jorge Martins; Claudia Renata dos Santos Barros; David Schlesinger; Debora Botequio Moretti; Dimas Tadeu Covas; Elaine Cristina Marqueze; Elaine Vieira Santos; Evandra Strazza Rodrigues; Heidge Fukumasu; Jayme Augusto de Souza-Neto; José Salvatore Leister Patané; Luiz Alcantara; Luiz Lehmann Coutinho; Maria Carolina Elias; Maurício Lacerda Nogueira; Rafael dos Santos Bezerra; Raul Machado Neto; Rejane Maria Tommasini Grotto; Ricardo Haddad; Sandra Coccuzzo Sampaio Vessoni; Simone Kashima; Svetoslav Nanev Slavov; Vincent Louis Viala |
| EPI_ISL_3102520 | SECRETARIA MUNICIPAL DE SAUDE DE VARZEA ALEGRE | Analytical Competence Molecular Epidemiology Lab/ACME, Oswaldo Cruz Foundation, Ceara (FIOCRUZ CE) | Cleber Furtado Aksenen; Fabio Miyajima; Fernando Braga Stehling; Francisco Eder de Moura Lopes; Jamille Maria Mendes Bezerra; Joaquim César do Nascimento Sousa Junior; Pedro Miguel Carneiro Jeronimo; Suzana Porto Almeida e Lucas Delerino; Thais Ferreira de Oliveira; Thais de Oliveira Costa; Ticiane Cavalcante de Souza; Veridiana Pessoa Miyajima |
| EPI_ISL_2493679 | SECRETARIA MUNICIPAL DE SAUDE PORTO FERREIRA | Instituto Butantan | Antonio Jorge Martins; Claudia Renata dos Santos Barros; David Schlesinger; Debora Botequio Moretti; Dimas Tadeu Covas; Elaine Cristina Marqueze; Elaine Vieira Santos; Evandra Strazza Rodrigues; Heidge Fukumasu; Jayme Augusto de Souza-Neto; José Salvatore Leister Patané; Luiz Alcantara; Luiz Lehmann Coutinho; Maria Carolina Elias; Maurício Lacerda Nogueira; Rafael dos Santos Bezerra; Raul Machado Neto; Rejane Maria Tommasini Grotto; Ricardo Haddad; Sandra Coccuzzo Sampaio Vessoni; Simone Kashima; Svetoslav Nanev Slavov; Vincent Louis Viala |
| EPI_ISL_2493654, EPI_ISL_2493657, EPI_ISL_2493660, EPI_ISL_2493668, EPI_ISL_2493693, EPI_ISL_2493698, EPI_ISL_2500979, EPI_ISL_2500983 | | | |
| see above | SECRETARIA MUNICIPAL DE SAUDE SOROCABA | Instituto Butantan | Antonio Jorge Martins; Claudia Renata dos Santos Barros; David Schlesinger; Debora Botequio Moretti; Dimas Tadeu Covas; Elaine Cristina Marqueze; Elaine Vieira Santos; Evandra Strazza Rodrigues; Heidge Fukumasu; Jayme Augusto de Souza-Neto; José Salvatore Leister Patané; Luiz Alcantara; Luiz Lehmann Coutinho; Maria Carolina Elias; Maurício Lacerda Nogueira; Rafael dos Santos Bezerra; Raul Machado Neto; Rejane Maria Tommasini Grotto; Ricardo Haddad; Sandra Coccuzzo Sampaio Vessoni; Simone Kashima; Svetoslav Nanev Slavov; Vincent Louis Viala |
| EPI_ISL_2344897, EPI_ISL_2344919, EPI_ISL_2344921, EPI_ISL_2344922, EPI_ISL_2344923, EPI_ISL_2344928, EPI_ISL_2344929, EPI_ISL_2344930, EPI_ISL_2344931, EPI_ISL_2344933, EPI_ISL_2344934, EPI_ISL_2344935, EPI_ISL_2344937, EPI_ISL_2344938, EPI_ISL_2344939 | | | |
| see above | SECRETARIA MUNICIPAL DE SAUDE SOROCABA | Instituto Butantan / ESALQ-Piracicaba | Antonio Jorge Martins; Claudia Renata dos Santos Barros; David Schlesinger; Debora Botequio Moretti; Dimas Tadeu Covas; Elaine Cristina Marqueze; Elaine Vieira Santos; Evandra Strazza Rodrigues; Heidge Fukumasu; Jayme Augusto de Souza-Neto; José Salvatore Leister Patané; Luiz Alcantara; Luiz Lehmann Coutinho; Maria Carolina Elias; Maurício Lacerda Nogueira; Rafael dos Santos Bezerra; Raul Machado Neto; Rejane Maria Tommasini Grotto; Ricardo Haddad; Sandra Coccuzzo Sampaio Vessoni; Simone Kashima; Svetoslav Nanev Slavov; Vincent Louis Viala |
| EPI_ISL_2209664, EPI_ISL_2345655, EPI_ISL_2345664, EPI_ISL_2345700, EPI_ISL_2345805, EPI_ISL_2345848, EPI_ISL_2345852 | | | |
| see above | SECRETARIA MUNICIPAL DE SAUDE SOROCABA | Instituto Butantan / Mendelics | Antonio Jorge Martins; Claudia Renata dos Santos Barros; David Schlesinger; Debora Botequio Moretti; Dimas Tadeu Covas; Elaine Cristina Marqueze; Elaine Vieira Santos; Evandra Strazza Rodrigues; Heidge Fukumasu; Jayme Augusto de Souza-Neto; José Salvatore Leister Patané; Luiz Alcantara; Luiz Lehmann Coutinho; Maria Carolina Elias; Maurício Lacerda Nogueira; Rafael dos Santos Bezerra; Raul Machado Neto; Rejane Maria Tommasini Grotto; Ricardo Haddad; Sandra Coccuzzo Sampaio Vessoni; Simone Kashima; Svetoslav Nanev Slavov; Vincent Louis Viala |
| EPI_ISL_2493562, EPI_ISL_2493565, EPI_ISL_2493567, EPI_ISL_2493568, EPI_ISL_2493569 | SERV DE VIG SANITARIA EPIDEMIO E CTRL DE ZOONOZES GUARUJA | Instituto Butantan | Antonio Jorge Martins; Claudia Renata dos Santos Barros; David Schlesinger; Debora Botequio Moretti; Dimas Tadeu Covas; Elaine Cristina Marqueze; Elaine Vieira Santos; Evandra Strazza Rodrigues; Heidge Fukumasu; Jayme Augusto de Souza-Neto; José Salvatore Leister Patané; Luiz Alcantara; Luiz Lehmann Coutinho; Maria Carolina Elias; Maurício Lacerda Nogueira; Rafael dos Santos Bezerra; Raul Machado Neto; Rejane Maria Tommasini Grotto; Ricardo Haddad; Sandra Coccuzzo Sampaio Vessoni; Simone Kashima; Svetoslav Nanev Slavov; Vincent Louis Viala |
| EPI_ISL_2209341, EPI_ISL_2345810 | SERV DE VIG SANITARIA EPIDEMIO E CTRL DE ZOONOZES GUARUJA | Instituto Butantan / Mendelics | Antonio Jorge Martins; Claudia Renata dos Santos Barros; David Schlesinger; Debora Botequio Moretti; Dimas Tadeu Covas; Elaine Cristina Marqueze; Elaine Vieira Santos; Evandra Strazza Rodrigues; Heidge Fukumasu; Jayme Augusto de Souza-Neto; José Salvatore Leister Patané; Luiz Alcantara; Luiz Lehmann Coutinho; Maria Carolina Elias; Maurício Lacerda Nogueira; Rafael dos Santos Bezerra; Raul Machado Neto; Rejane Maria Tommasini Grotto; Ricardo Haddad; Sandra Coccuzzo Sampaio Vessoni; Simone Kashima; Svetoslav Nanev Slavov; Vincent Louis Viala |
| EPI_ISL_2344940 | SMS IPERO | Instituto Butantan / ESALQ-Piracicaba | Antonio Jorge Martins; Claudia Renata dos Santos Barros; David Schlesinger; Debora Botequio Moretti; Dimas Tadeu Covas; Elaine Cristina Marqueze; Elaine Vieira Santos; Evandra Strazza Rodrigues; Heidge Fukumasu; Jayme Augusto de Souza-Neto; José Salvatore Leister Patané; Luiz Alcantara; Luiz Lehmann Coutinho; Maria Carolina Elias; Maurício Lacerda Nogueira; Rafael dos Santos Bezerra; Raul Machado Neto; Rejane Maria Tommasini Grotto; Ricardo Haddad; Sandra Coccuzzo Sampaio Vessoni; Simone Kashima; Svetoslav Nanev Slavov; Vincent Louis Viala |
| EPI_ISL_2209419 | SMS IPERO | Instituto Butantan / Mendelics | Antonio Jorge Martins; Claudia Renata dos Santos Barros; David Schlesinger; Debora Botequio Moretti; Dimas Tadeu Covas; Elaine Cristina Marqueze; Elaine Vieira Santos; Evandra Strazza Rodrigues; Heidge Fukumasu; Jayme Augusto de Souza-Neto; José Salvatore Leister Patané; Luiz Alcantara; Luiz Lehmann Coutinho; Maria Carolina Elias; Maurício Lacerda Nogueira; Rafael dos Santos Bezerra; Raul Machado Neto; Rejane Maria Tommasini Grotto; Ricardo Haddad; Sandra Coccuzzo Sampaio Vessoni; Simone Kashima; Svetoslav Nanev Slavov; Vincent Louis Viala |
| EPI_ISL_2344888, EPI_ISL_2344913 | SMS SECRETARIA MUNICIPAL DE SAUDE DE BOITUVA | Instituto Butantan / ESALQ-Piracicaba | Antonio Jorge Martins; Claudia Renata dos Santos Barros; David Schlesinger; Debora Botequio Moretti; Dimas Tadeu Covas; Elaine Cristina Marqueze; Elaine Vieira Santos; Evandra Strazza Rodrigues; Heidge Fukumasu; Jayme Augusto de Souza-Neto; José Salvatore Leister Patané; Luiz Alcantara; Luiz Lehmann Coutinho; Maria Carolina Elias; Maurício Lacerda Nogueira; Rafael dos Santos Bezerra; Raul Machado Neto; Rejane Maria Tommasini Grotto; Ricardo Haddad; Sandra Coccuzzo Sampaio Vessoni; Simone Kashima; Svetoslav Nanev Slavov; Vincent Louis Viala |
| EPI_ISL_2210223, EPI_ISL_2345713, EPI_ISL_2345718 | SMS SECRETARIA MUNICIPAL DE SAUDE DE BOITUVA | Instituto Butantan / Mendelics | Antonio Jorge Martins; Claudia Renata dos Santos Barros; David Schlesinger; Debora Botequio Moretti; Dimas Tadeu Covas; Elaine Cristina Marqueze; Elaine Vieira Santos; Evandra Strazza Rodrigues; Heidge Fukumasu; Jayme Augusto de Souza-Neto; José Salvatore Leister Patané; Luiz Alcantara; Luiz Lehmann Coutinho; Maria Carolina Elias; Maurício Lacerda Nogueira; Rafael dos Santos Bezerra; Raul Machado Neto; Rejane Maria Tommasini Grotto; Ricardo Haddad; Sandra Coccuzzo Sampaio Vessoni; Simone Kashima; Svetoslav Nanev Slavov; Vincent Louis Viala |
| EPI_ISL_2097297, EPI_ISL_2274580, EPI_ISL_2421052, EPI_ISL_2422317 | STONY BROOK UNIVERSITY HOSPITAL | Wadsworth Center, New York State Department of Health | Alexis Russell; Catharine Prussing; Daryl M. Lamson; Erasmus Schneider; Erica Lasek-Nesselquist; John Kelly; Jonathan Plitnick; Kirsten St. George; Matthew Shudt; Melissa A Leisner; Navjot Singh |
| EPI_ISL_2098138 | SUNY UPSTATE MEDICAL UNIVERSITY | Wadsworth Center, New York State Department of Health | Alexis Russell; Catharine Prussing; Daryl M. Lamson; Erasmus Schneider; Erica Lasek-Nesselquist; John Kelly; Jonathan Plitnick; Kirsten St. George; Matthew Shudt; Melissa A Leisner; Navjot Singh |
| EPI_ISL_2004328 | SYNLAB | Instituto Nacional de Saude (INSA) | Borges et al |
| EPI_ISL_1848074 | SYNLAB Jena Oncoscreen | Robert Koch Institute | |
| EPI_ISL_2105731, EPI_ISL_2105794, EPI_ISL_2105847, EPI_ISL_2105848, EPI_ISL_2105857, EPI_ISL_2230900, EPI_ISL_2230903, EPI_ISL_2319198, EPI_ISL_2319216, EPI_ISL_2319233, EPI_ISL_2319236, EPI_ISL_2319286, EPI_ISL_2349882, EPI_ISL_2349884, EPI_ISL_2350133, EPI_ISL_2444402, EPI_ISL_2444506, EPI_ISL_2444518, EPI_ISL_2444545 | | | |
| see above | Salud Digna | Instituto Nacional de Medicina Genomica | Abraham Campos-Romero; Cedro-Tanda A; Escobar-Arrazola; Gonzalez-Barrera D; Herrera-Montalvo LA.; Hidalgo-Miranda A; Luna-Ruiz Marco; M; Mendoza-Vargas A; Moreno-Camacho José Luis; Munguia-Garza P; Ramirez-Vega O; Rangel-DeLeon D; Reyes-Grajeda JP; Rodriguez-Gallegos Jorge |
| EPI_ISL_2231178, EPI_ISL_2329537, EPI_ISL_2329595, EPI_ISL_2329601, EPI_ISL_2329608, EPI_ISL_2422993, EPI_ISL_2423003, EPI_ISL_2423011, EPI_ISL_2423013, EPI_ISL_2423017, EPI_ISL_2628598, EPI_ISL_2628614, EPI_ISL_2628627, EPI_ISL_2628630, EPI_ISL_2628677, EPI_ISL_2628786, EPI_ISL_2628844, EPI_ISL_2628904 | | | |
| see above | San Diego County Public Health Laboratory | Andersen lab at Scripps Research | Brett Austin; Jovan Shephard; SEARCH Alliance San Diego with Ashleigh Murphy |
| EPI_ISL_2840867, EPI_ISL_2840895 | San Diego County Public Health Laboratory | San Diego County Public Health Laboratory | Ashleigh Murphy Schafer; Brett Austin |
| EPI_ISL_1970975, EPI_ISL_1970978, EPI_ISL_2287924 | San Juan Regional Medical Center | New Mexico Department of Health Scientific Laboratory | Anastacia Griego-Fisher; D'eldra Malone; Ellie Johnson; Jennifer Benoit; Linda Salazar; Mark Willmon; Ratheesh Rajan |
| EPI_ISL_2631213, EPI_ISL_2659047, EPI_ISL_2659075, EPI_ISL_2659111, EPI_ISL_2659114 | Santa Clara Valley Medical Center | Santa Clara County Public Health Laboratory | Santa Clara County Public Health Department |
| EPI_ISL_1969513, EPI_ISL_1969518, EPI_ISL_1969532, EPI_ISL_3373746, EPI_ISL_3543807 | Scripps Medical Laboratory | Andersen lab at Scripps Research | Ellen Stefanski; Ian Mchardy; SEARCH Alliance San Diego with Michael Quigley |
| EPI_ISL_2179729, EPI_ISL_2179731, EPI_ISL_2179732, EPI_ISL_2179752, EPI_ISL_2179753, EPI_ISL_2179754, EPI_ISL_2179758, EPI_ISL_2179794, EPI_ISL_2179796, EPI_ISL_2179797, EPI_ISL_2179802 | | | |
| see above | Servicio Microbiología Hospital La Paz | Servicio Microbiología Hospital La Paz | Elie Dahdouh; Fernando Lázaro; Jesús Mingorance Cruz; Rubén Cáceres |
| EPI_ISL_2136127, EPI_ISL_2136130, EPI_ISL_2136132, EPI_ISL_2140045, EPI_ISL_2140060, EPI_ISL_2140078, EPI_ISL_2140084, EPI_ISL_2140086, EPI_ISL_2140093, EPI_ISL_2140106, EPI_ISL_2140135, EPI_ISL_2140143, EPI_ISL_2140144, EPI_ISL_2158741, EPI_ISL_2158831, EPI_ISL_2158843, EPI_ISL_4082187, EPI_ISL_4082200, EPI_ISL_4082202, EPI_ISL_4082206, EPI_ISL_4082207, EPI_ISL_4082215, EPI_ISL_4082216, EPI_ISL_4082219, EPI_ISL_4082221, EPI_ISL_4082246, EPI_ISL_4082286, EPI_ISL_4082295, EPI_ISL_4082298, EPI_ISL_4082310, EPI_ISL_4082330, EPI_ISL_4082338, EPI_ISL_4082344, EPI_ISL_4082354, EPI_ISL_4082356, EPI_ISL_4082358, EPI_ISL_4082372, EPI_ISL_4082405, EPI_ISL_4082426, EPI_ISL_4082459, EPI_ISL_4082489, EPI_ISL_4082493, EPI_ISL_4082525, EPI_ISL_4082526, EPI_ISL_4082527, EPI_ISL_4082539, EPI_ISL_4082624, EPI_ISL_4082648, EPI_ISL_4082653, EPI_ISL_4082661, EPI_ISL_4082721, EPI_ISL_4082766, EPI_ISL_4083068, EPI_ISL_4083069 | | | |
| see above | Servicio Virosis Respiratorias-Departamento Virología-INEI | Instituto Nacional Enfermedades Infecciosas C.G.Malbran | Avaro M.; Baumeister E.; Benedetti E.; Campos J.; Cisterna D.; Dattero ME; De Belder D.; Haim MS.; Lorenzo F.; Molina V.; Perandones C.; Poklepovich T.; Pontoriero A.; Russo M.; Sanchez Loria J.; Tuduri E. |
| EPI_ISL_2104452, EPI_ISL_2426641, EPI_ISL_2426651, EPI_ISL_2558027 | Servicio de Microbiologia Hospital Ramon y Cajal | Servicio de Microbiologia Hospital Ramon y Cajal | Galan JC; Martinez L. Abreu M; Ponce M; y Gonzalez-Alba JM |
| EPI_ISL_1922315, EPI_ISL_1922316, EPI_ISL_1922318, EPI_ISL_1922321, EPI_ISL_1922394, EPI_ISL_1922397, EPI_ISL_1922479, EPI_ISL_2133001, EPI_ISL_2133002, EPI_ISL_2510576, EPI_ISL_2510579, EPI_ISL_2510590, EPI_ISL_2510591, EPI_ISL_2510593 | | | |
| see above | Servicio de Microbiología Clínica (Complejo Hospitalario de Navarra, Pamplona) | Centro de Secuenciación NASERTIC | Ana Miqueleiz; Ana Navascués; Carmen Ezpeleta Baquedano |
| EPI_ISL_2187013 | Servicio de Microbiología, Hospital General Universitario de Ciudad Real | Servicio de Microbiología, Hospital General Universitario de Ciudad Real | Jose Martinez Alarcon; Soledad Illescas |
| EPI_ISL_2545376, EPI_ISL_2545401, EPI_ISL_2545405 | Servicio de Microbiología, Laboratori Clínic Metropolitana Nord. Hospital Universitari Germans Trias i Pujol. Institut d'Investigació en Ciències de la Salut Germans Trias i Pujol (IGTP) | SeqCOVID-SPAIN consortium/IBV(CSIC) | Adrián Antuori; Anabel Fernández; Anna Not; Antoni E. Bordoy; Cristina Casañ and SeqCOVID-SPAIN consortium; Elisa Martró; Nona Romaní; Verónica Saludes |
| EPI_ISL_2271613, EPI_ISL_2362312, | Servicio de Microbiología. Consorcio Hospital General Universitario de | SeqCOVID-SPAIN consortium/IBV(CSIC) | Begoña Fuster Escrivá; Carme Salvador García; Concepción Gimeno Cardona and SeqCOVID-SPAIN consortium; María Dolores Ocete; Rafael Medina González |

| | | | |
|---|---|---|---|
| EPI_ISL_2658196, EPI_ISL_2658198 | Valencia | | |
| EPI_ISL_2016629 | Servicio de Microbiología. Hospital Clínico Universitario de Valencia | SeqCOVID-SPAIN consortium/IBV(CSIC) | David Navarro Ortega; Eliseo Albert Vicent; Ignacio Torres and SeqCOVID-SPAIN consortium |
| EPI_ISL_2627507 | Servicio de Microbiología. Hospital Universitario Doctor Peset | SeqCOVID-SPAIN consortium/IBV(CSIC) | José Miguel Nogueira Coito and SeqCOVID-SPAIN consortium; Juan Alberola Enguídanos; Juan José Camarena Miñana; Rosa González Pellicer |
| EPI_ISL_2162847 | Servizo de Microbioloxía. Complexo Hospitalario de Santiago de Compostela | Servizo de Microbioloxía. Complexo Hospitalario de Santiago de Compostela | Amparo Coira; Antonio Aguilera; Carlos García_Riestra; Daniel Navarro; Gema Barbeito; Iria Roca; José Llovo; Laura Millán; Manuela Hernández; María Luisa Pérez_del_Molino; Mercedes Treviño; Rocío trastoy; Teresa Lopez_Valiño; Xosé Costa |
| EPI_ISL_2231176, EPI_ISL_2231177, EPI_ISL_2231179, EPI_ISL_2231180, EPI_ISL_2231181, EPI_ISL_2231182, EPI_ISL_2231184, EPI_ISL_2231185, EPI_ISL_2329450, EPI_ISL_2329452, EPI_ISL_2329453, EPI_ISL_2329459, EPI_ISL_2329462, EPI_ISL_2329463, EPI_ISL_2422816, EPI_ISL_2422833 | | | |
| see above | Sharp HealthCare Laboratory | Andersen lab at Scripps Research | Art Mendoza; Cathy Woerle; Jacquelyn Berumen; Liam McGinnis; Omid Bakhtar; SEARCH Alliance San Diego with Aaron Harding |
| EPI_ISL_2296014, EPI_ISL_2296019, EPI_ISL_2296024, EPI_ISL_2296034, EPI_ISL_2296044, EPI_ISL_2801143, EPI_ISL_2801144, EPI_ISL_2801145, EPI_ISL_2801146, EPI_ISL_2801147, EPI_ISL_2801148, EPI_ISL_2801149, EPI_ISL_2801150, EPI_ISL_2801151 | | | |
| see above | Simple Laboratories | Gagnon Lab, Southern Illinois University | Keith Gagnon |
| EPI_ISL_2230739, EPI_ISL_2484886 | Southern Nevada Public Health Laboratory | Southern Nevada Public Health Laboratory | Michael Picker |
| EPI_ISL_2102287, EPI_ISL_2510856, EPI_ISL_2510888, EPI_ISL_2510973, EPI_ISL_2510976 | Spital Limmattal | Institute of Medical Virology | Alexandra Trkola; Annette AudigŽ; Catharine Aquino; Cyril Shah; Daniel Ehrsam; Gabriela Ziltener; Guido Bloemberg; Hubert Rehrauer; Isabel StŸrmer; Joel Wirz; Jon Huder; JŸrg Bšni; Kevin Steiner; Maria GrŸnberg; Maryam Zaheri; Michael Huber; Riccarda Capaul; Stefan Schmutz; Verena Kufner; Weihong Qi |
| EPI_ISL_2136954, EPI_ISL_2136986, EPI_ISL_2136995, EPI_ISL_2449879 | Spital MŠnnedorf AG | Institute of Medical Virology | Alexandra Trkola; Annette AudigŽ; Catharine Aquino; Cyril Shah; Daniel Ehrsam; Gabriela Ziltener; Guido Bloemberg; Hubert Rehrauer; Isabel StŸrmer; Joel Wirz; Jon Huder; JŸrg Bšni; Kevin Steiner; Maria GrŸnberg; Maryam Zaheri; Michael Huber; Riccarda Capaul; Stefan Schmutz; Verena Kufner; Weihong Qi |
| EPI_ISL_3418888, EPI_ISL_3418895, EPI_ISL_3418896, EPI_ISL_3418957, EPI_ISL_3418973 | StarMed Healthcare | UNC Charlotte Environmental Monitoring Laboratory | Adam France; Cynthia Gibas; Erin Stiers; Jannatul Ferdous; Jessica Schlueter; Kevin Lambirth; Sam Kunkleman |
| EPI_ISL_2007997, EPI_ISL_2135446, EPI_ISL_2135447, EPI_ISL_2135474, EPI_ISL_2724535, EPI_ISL_2724590, EPI_ISL_2724624, EPI_ISL_2724635, EPI_ISL_2724646, EPI_ISL_2724669, EPI_ISL_2724720, EPI_ISL_2724730, EPI_ISL_2724739, EPI_ISL_2724807 | | | |
| see above | State Laboratories Division, Hawaii State Department of Health | State Laboratories Division, Hawaii State Department of Health | Ayana Garnet; Drew Kuwazaki; Edward Desmond; Pamela O'Brien; Razvan Sultana |
| EPI_ISL_2378181, EPI_ISL_2378182, EPI_ISL_2378546, EPI_ISL_2378592, EPI_ISL_2472922, EPI_ISL_2472954, EPI_ISL_2473147, EPI_ISL_2473153, EPI_ISL_2473183, EPI_ISL_2473255, EPI_ISL_2473270, EPI_ISL_2473299, EPI_ISL_2473310, EPI_ISL_2473366, EPI_ISL_2473368, EPI_ISL_2473382, EPI_ISL_2473390, EPI_ISL_2473409, EPI_ISL_2473423, EPI_ISL_2473478, EPI_ISL_2473489, EPI_ISL_2473495, EPI_ISL_2473523, EPI_ISL_2473526, EPI_ISL_2473537, EPI_ISL_2473557, EPI_ISL_2473561, EPI_ISL_2473579, EPI_ISL_2473638, EPI_ISL_2800583 | | | |
| see above | State Testing Facility | Altius Institute for Biomedical Research | Alex Isner; Alex Nguyen; Amanda Gale; Audra Johnson; Clem Green; Daniel Bates; Eric Thorland; Jacob Rodriguez; Jean Robinston; Jemma Nelson; Jessica Kunder; John Stamatoyannopoulos; Joshua Richards; Julia Wald; Kneshay Harper; Lauren Mitchell; Mark Frerker; Matt Hartman; Michael Buckley; Muhammad Halimun; Rebecca Bruders; Sadie Patraw; Sofia Olsson; Tobias Ragoczy |
| EPI_ISL_2391791 | Sun Clinical Laboratory | Los Angeles County Public Health Laboratories | P. Hemarajata et al. |
| EPI_ISL_2692135 | Swedish Covenant Hospital | RIPHL at Rush University Medical Center | Chicago Department of Public Health; Felix Araujo Perez; Kevin Kunstman; Laura Furtado; Marieta Hyde; Max Kolton; Stefan Green |
| EPI_ISL_2033275, EPI_ISL_2033311, EPI_ISL_2257985, EPI_ISL_2413880, EPI_ISL_2413987, EPI_ISL_2413998, EPI_ISL_2414004, EPI_ISL_2414029, EPI_ISL_2414091, EPI_ISL_2414220, EPI_ISL_2414270, EPI_ISL_2581053, EPI_ISL_2581084, EPI_ISL_2581124, EPI_ISL_2581130, EPI_ISL_2581444, EPI_ISL_2980127, EPI_ISL_3157893 | | | |
| see above | Swedish national genomic surveillance program of SARS-CoV-2 | The Public Health Agency of Sweden | Alma Brolund; Maria Lind Karlberg; Maximilian Riess; Swedish national genomic surveillance program of SARS-CoV-2 |
| EPI_ISL_2421182 | TEMPUS LABS INC | Wadsworth Center, New York State Department of Health | Alexis Russell; Catharine Prussing; Daryl M. Lamson; Erasmus Schneider; Erica Lasek-Nesselquist; John Kelly; Jonathan Plitnick; Kirsten St. George; Matthew Shudt; Melissa A Leisner; Navjot Singh |
| EPI_ISL_2281451, EPI_ISL_2281501, EPI_ISL_2423702, EPI_ISL_2424098, EPI_ISL_2491282, EPI_ISL_3088567 | TGen North | TGen North | "Jolene Bowers; Ashlyn Pfeiffer; Brett Van Tassel; Chris French; Darrin Lemmer; Dave Engelthaler; Dave Engelthaler"; Hayley Yaglom; Heather Centner; Jolene Bowers; The Arizona COVID Genomics Union (ACGU); The Arizona COVID Genomics Union (ACGU)" |
| EPI_ISL_2450366 | TN Division of Laboratory Services | Genomics and Discovery, Respiratory Viruses Branch, Division of Viral Diseases, Centers for Disease Control and Prevention | Adam Retchless; Anna Kelleher; Anna Uehara; Brian Lynch; Clinton R. Paden; Dhwani Batra; Haibin Wang; Han Jia Justin Ng; Jasmine Padilla; Jing Zhang; Justin Lee; Krista Queen; Mark Burroughs; Mili Sheth; Morgan Davis; Peter Cook; Rachel Marine; Sarah Nobles; Suxiang Tong; Tara Coalter; Yan Li; Ying Tao |
| EPI_ISL_2451912 | TX DSHS, Lab Services Section MC 1947 | Centers for Disease Control and Prevention Division of Viral Diseases, Pathogen Discovery | Alison Laufer Halpin; Ben L. Rambo-Martin; Clinton R. Paden; Dakota Howard; Darlene Wagner; Dave Wentworth; Dhwani Batra; Jasmine Padilla; Justin Lee; Katie Dillon; Krista Queen; Kristen Knipe; Kristine Lacek; Mark Burroughs; Matthew Schmerer; Mili Sheth; Peter Cook; Sam Shepard; Sarah Nobles; Shoshona Le; Suxiang Tong; Vivien Dugan; Yvette Unoarumhi |
| EPI_ISL_2649802, EPI_ISL_2649809, EPI_ISL_2649815, EPI_ISL_2790522, EPI_ISL_2790532, EPI_ISL_2790544, EPI_ISL_2966835 | | | |
| see above | Tampa General Hospital Esoteric Lab | Tampa General Hospital Esoteric Research & Development Lab | Amorce Lima; Deanna Becker; Dominic Uy; Elaine Vendrone; Jon Faughn; Jorge Mecias-Frias; Suzane Silbert; Vicki Healer |
| EPI_ISL_2501617 | Texas Children's Hospital | Texas Children's Microbiome Center | Ila Singh; James Dunn; James Versalovic; Jennifer K. Spinler; Ruth Ann Luna |
| EPI_ISL_1826530, EPI_ISL_1970663, EPI_ISL_1970674, EPI_ISL_2133353, EPI_ISL_2230024, EPI_ISL_2230026, EPI_ISL_2346333, EPI_ISL_2346334, EPI_ISL_2346335, EPI_ISL_2346338 | | | |
| see above | The Jackson Laboratory | The Jackson Laboratory | Adams M; Bergeron D; Kelly K; Li L; Long J; Omerza G; Renzette N |
| EPI_ISL_3930751 | The National University Hospital of Iceland | deCODE genetics | Agnar Helgason; Alma Moller; Arna B Agustsdottir; Arnaldur Gylfason; Asgeir Sigurdsson; Aslaug Jonasdottir; Berglind Eiriksdottir; Bjarni Thorbjornsson; Brynjar O Jensson; Daniel F Gudbjartsson; Droplaug N Magnusdottir; Elisabet E Gardarsdottir; Emil A Thorarensen; Gardar Sveinbjornsson; Gisli Masson; Gudmundur Georgsson; Gudmundur L Norddahl; Gudrun Sigmundsdottir; Hakon Jonsson; Hannes Eggertsson; Hilma Holm; Ingileif Jonsdottir; Jona Saemundsdottir; Kamilla S Josefsdottir; Kari Stefansson; Karl G Kristinsson; Kjartan R Gudmundsson; Kristin E Sveinsdottir; Kristjan E Hjorleifsson; Louise le Roux; Maney Sveinsdottir; Olafia S Gretarsdottir; Olafur T Magnusson; Pall Melsted; Patrick Sulem; Run Fridriksdottir; Solvi Rognvaldsson; Thora R Gunnarsdottir; Thordur Kristjansson; Thorolfur Gudnason; Unnur Thorsteinsdottir |
| EPI_ISL_2834212 | The Ohio State University Applied Microbiology Services Laboratory | The Ohio State University Applied Microbiology Services Laboratory | Seth A. Faith PhD |
| EPI_ISL_2454361 | TriCore Reference Laboratories | Center for Global Health, University of New Mexico Health Sciences Center | Cecilia Thompson; Darrell Dinwiddie; Daryl Domman; Karissa Culbreath; Kendra Pesko; Kurt Schwalm; Valerie Morley |
| EPI_ISL_2598094, EPI_ISL_2598100 | TriCore Reference Laboratories | Center for Global Health, University of New Mexico Health Sciences Center | Cecilia Thompson; Darrell Dinwiddie; Daryl Domman; Karissa Culbreath; Kendra Pesko; Kurt Schwalm; Valerie Morley |
| EPI_ISL_2208930 | UBS ALAMBARI | Instituto Butantan / Mendelics | Antonio Jorge Martins; Claudia Renata dos Santos Barros; David Schlesinger; Debora Botequio Moretti; Dimas Tadeu Covas; Elaine Cristina Marqueze; Elaine Vieira Santos; Evandra Strazza Rodrigues; Heidge Fukumasu; Jayme Augusto de Souza-Neto; José Salvatore Leister Patané; Luiz Alcantara; Luiz Lehmann Coutinho; Maria Carolina Elias; Maurício Lacerda Nogueira; Rafael dos Santos Bezerra; Raul Machado Neto; Rejane Maria Tommasini Grotto; Ricardo Haddad; Sandra Coccuzzo Sampaio Vessoni; Simone Kashima; Svetoslav Nanev Slavov; Vincent Louis Viala |
| EPI_ISL_2345154, EPI_ISL_2345155, EPI_ISL_2345157 | UBS BONFIM | Instituto Butantan / ESALQ-Piracicaba | Antonio Jorge Martins; Claudia Renata dos Santos Barros; David Schlesinger; Debora Botequio Moretti; Dimas Tadeu Covas; Elaine Cristina Marqueze; Elaine Vieira Santos; Evandra Strazza Rodrigues; Heidge Fukumasu; Jayme Augusto de Souza-Neto; José Salvatore Leister Patané; Luiz Alcantara; Luiz Lehmann Coutinho; Maria Carolina Elias; Maurício Lacerda Nogueira; Rafael dos Santos Bezerra; Raul Machado Neto; Rejane Maria Tommasini Grotto; Ricardo Haddad; Sandra Coccuzzo Sampaio Vessoni; Simone Kashima; Svetoslav Nanev Slavov; Vincent Louis Viala |
| EPI_ISL_2445173 | UBS CAIC OURINHOS | Instituto Butantan | Antonio Jorge Martins; Claudia Renata dos Santos Barros; David Schlesinger; Debora Botequio Moretti; Dimas Tadeu Covas; Elaine Cristina Marqueze; Elaine Vieira Santos; Evandra Strazza Rodrigues; Heidge Fukumasu; Jayme Augusto de Souza-Neto; José Salvatore Leister Patané; Luiz Alcantara; Luiz Lehmann Coutinho; Maria Carolina Elias; Maurício Lacerda Nogueira; Rafael dos Santos Bezerra; Raul Machado Neto; Rejane Maria Tommasini Grotto; Ricardo Haddad; Sandra Coccuzzo Sampaio Vessoni; Simone Kashima; Svetoslav Nanev Slavov; Vincent Louis Viala |
| EPI_ISL_2445207, EPI_ISL_2473718 | UBS CELESTINO CARNIELO | Instituto Butantan | Antonio Jorge Martins; Claudia Renata dos Santos Barros; David Schlesinger; Debora Botequio Moretti; Dimas Tadeu Covas; Elaine Cristina Marqueze; Elaine Vieira Santos; Evandra Strazza Rodrigues; Heidge Fukumasu; Jayme Augusto de Souza-Neto; José Salvatore Leister Patané; Luiz Alcantara; Luiz Lehmann Coutinho; Maria Carolina Elias; Maurício Lacerda Nogueira; Rafael dos Santos Bezerra; Raul Machado Neto; Rejane Maria Tommasini Grotto; Ricardo Haddad; Sandra Coccuzzo Sampaio Vessoni; Simone Kashima; Svetoslav Nanev Slavov; Vincent Louis Viala |
| EPI_ISL_2445208, EPI_ISL_2473719, EPI_ISL_2493655, EPI_ISL_2493656, | UBS DE PARAISO | Instituto Butantan | Antonio Jorge Martins; Claudia Renata dos Santos Barros; David Schlesinger; Debora Botequio Moretti; Dimas Tadeu Covas; Elaine Cristina Marqueze; Elaine Vieira Santos; Evandra Strazza Rodrigues; Heidge Fukumasu; Jayme Augusto de Souza-Neto; José Salvatore Leister Patané; Luiz Alcantara; Luiz Lehmann Coutinho; Maria Carolina Elias; Maurício Lacerda Nogueira; Rafael dos Santos Bezerra; Raul Machado Neto; Rejane Maria Tommasini Grotto; Ricardo Haddad; Sandra Coccuzzo Sampaio Vessoni; Simone Kashima; Svetoslav Nanev Slavov; Vincent Louis Viala |

| | | | |
|---|---|---|---|
| EPI_ISL_2493658 | | | |
| EPI_ISL_2445528, EPI_ISL_2493418 | UBS DE PEDRANOPOLIS | Instituto Butantan | Antonio Jorge Martins; Claudia Renata dos Santos Barros; David Schlesinger; Debora Botequio Moretti; Dimas Tadeu Covas; Elaine Cristina Marqueze; Elaine Vieira Santos; Evandra Strazza Rodrigues; Heidge Fukumasu; Jayme Augusto de Souza-Neto; José Salvatore Leister Patané; Luiz Alcantara; Luiz Lehmann Coutinho; Maria Carolina Elias; Maurício Lacerda Nogueira; Rafael dos Santos Bezerra; Raul Machado Neto; Rejane Maria Tommasini Grotto; Ricardo Haddad; Sandra Coccuzzo Sampaio Vessoni; Simone Kashima; Svetoslav Nanev Slavov; Vincent Louis Viala |
| EPI_ISL_2344847 | UBS DE PEDRANOPOLIS | Instituto Butantan / ESALQ-Piracicaba | Antonio Jorge Martins; Claudia Renata dos Santos Barros; David Schlesinger; Debora Botequio Moretti; Dimas Tadeu Covas; Elaine Cristina Marqueze; Elaine Vieira Santos; Evandra Strazza Rodrigues; Heidge Fukumasu; Jayme Augusto de Souza-Neto; José Salvatore Leister Patané; Luiz Alcantara; Luiz Lehmann Coutinho; Maria Carolina Elias; Maurício Lacerda Nogueira; Rafael dos Santos Bezerra; Raul Machado Neto; Rejane Maria Tommasini Grotto; Ricardo Haddad; Sandra Coccuzzo Sampaio Vessoni; Simone Kashima; Svetoslav Nanev Slavov; Vincent Louis Viala |
| EPI_ISL_2344850 | UBS DE SAO JOAO DAS DUAS PONTES | Instituto Butantan / ESALQ-Piracicaba | Antonio Jorge Martins; Claudia Renata dos Santos Barros; David Schlesinger; Debora Botequio Moretti; Dimas Tadeu Covas; Elaine Cristina Marqueze; Elaine Vieira Santos; Evandra Strazza Rodrigues; Heidge Fukumasu; Jayme Augusto de Souza-Neto; José Salvatore Leister Patané; Luiz Alcantara; Luiz Lehmann Coutinho; Maria Carolina Elias; Maurício Lacerda Nogueira; Rafael dos Santos Bezerra; Raul Machado Neto; Rejane Maria Tommasini Grotto; Ricardo Haddad; Sandra Coccuzzo Sampaio Vessoni; Simone Kashima; Svetoslav Nanev Slavov; Vincent Louis Viala |
| EPI_ISL_2445195 | UBS DE SUD MENNUCCI | Instituto Butantan | Antonio Jorge Martins; Claudia Renata dos Santos Barros; David Schlesinger; Debora Botequio Moretti; Dimas Tadeu Covas; Elaine Cristina Marqueze; Elaine Vieira Santos; Evandra Strazza Rodrigues; Heidge Fukumasu; Jayme Augusto de Souza-Neto; José Salvatore Leister Patané; Luiz Alcantara; Luiz Lehmann Coutinho; Maria Carolina Elias; Maurício Lacerda Nogueira; Rafael dos Santos Bezerra; Raul Machado Neto; Rejane Maria Tommasini Grotto; Ricardo Haddad; Sandra Coccuzzo Sampaio Vessoni; Simone Kashima; Svetoslav Nanev Slavov; Vincent Louis Viala |
| EPI_ISL_2445206, EPI_ISL_2493402, EPI_ISL_2493404, EPI_ISL_2493409, EPI_ISL_2493423, EPI_ISL_2493425, EPI_ISL_2493426, EPI_ISL_2494337 | | | |
| see above | UBS DE TABAPUA | Instituto Butantan | Antonio Jorge Martins; Claudia Renata dos Santos Barros; David Schlesinger; Debora Botequio Moretti; Dimas Tadeu Covas; Elaine Cristina Marqueze; Elaine Vieira Santos; Evandra Strazza Rodrigues; Heidge Fukumasu; Jayme Augusto de Souza-Neto; José Salvatore Leister Patané; Luiz Alcantara; Luiz Lehmann Coutinho; Maria Carolina Elias; Maurício Lacerda Nogueira; Rafael dos Santos Bezerra; Raul Machado Neto; Rejane Maria Tommasini Grotto; Ricardo Haddad; Sandra Coccuzzo Sampaio Vessoni; Simone Kashima; Svetoslav Nanev Slavov; Vincent Louis Viala |
| EPI_ISL_2445536, EPI_ISL_2493659, EPI_ISL_2493665, EPI_ISL_2493666 | UBS DE UBARANA | Instituto Butantan | Antonio Jorge Martins; Claudia Renata dos Santos Barros; David Schlesinger; Debora Botequio Moretti; Dimas Tadeu Covas; Elaine Cristina Marqueze; Elaine Vieira Santos; Evandra Strazza Rodrigues; Heidge Fukumasu; Jayme Augusto de Souza-Neto; José Salvatore Leister Patané; Luiz Alcantara; Luiz Lehmann Coutinho; Maria Carolina Elias; Maurício Lacerda Nogueira; Rafael dos Santos Bezerra; Raul Machado Neto; Rejane Maria Tommasini Grotto; Ricardo Haddad; Sandra Coccuzzo Sampaio Vessoni; Simone Kashima; Svetoslav Nanev Slavov; Vincent Louis Viala |
| EPI_ISL_2345190, EPI_ISL_2345191 | UBS DR ABELARDO PINHEIRO GUIMARAES SCRPARDO | Instituto Butantan / ESALQ-Piracicaba | Antonio Jorge Martins; Claudia Renata dos Santos Barros; David Schlesinger; Debora Botequio Moretti; Dimas Tadeu Covas; Elaine Cristina Marqueze; Elaine Vieira Santos; Evandra Strazza Rodrigues; Heidge Fukumasu; Jayme Augusto de Souza-Neto; José Salvatore Leister Patané; Luiz Alcantara; Luiz Lehmann Coutinho; Maria Carolina Elias; Maurício Lacerda Nogueira; Rafael dos Santos Bezerra; Raul Machado Neto; Rejane Maria Tommasini Grotto; Ricardo Haddad; Sandra Coccuzzo Sampaio Vessoni; Simone Kashima; Svetoslav Nanev Slavov; Vincent Louis Viala |
| EPI_ISL_2345156, EPI_ISL_2345158, EPI_ISL_2345159, EPI_ISL_2345162, EPI_ISL_2345163, EPI_ISL_2345166, EPI_ISL_2345167, EPI_ISL_2345168, EPI_ISL_2345173, EPI_ISL_2345175, EPI_ISL_2345179 | | | |
| see above | UBS DR ALFREDO DANTAS DE SOUZA UMUARAMA | Instituto Butantan / ESALQ-Piracicaba | Antonio Jorge Martins; Claudia Renata dos Santos Barros; David Schlesinger; Debora Botequio Moretti; Dimas Tadeu Covas; Elaine Cristina Marqueze; Elaine Vieira Santos; Evandra Strazza Rodrigues; Heidge Fukumasu; Jayme Augusto de Souza-Neto; José Salvatore Leister Patané; Luiz Alcantara; Luiz Lehmann Coutinho; Maria Carolina Elias; Maurício Lacerda Nogueira; Rafael dos Santos Bezerra; Raul Machado Neto; Rejane Maria Tommasini Grotto; Ricardo Haddad; Sandra Coccuzzo Sampaio Vessoni; Simone Kashima; Svetoslav Nanev Slavov; Vincent Louis Viala |
| EPI_ISL_1967205, EPI_ISL_1967206, EPI_ISL_1967207, EPI_ISL_1967208, EPI_ISL_1967209, EPI_ISL_2345646, EPI_ISL_2345745 | | | |
| see above | UBS DR ALFREDO DANTAS DE SOUZA UMUARAMA | Instituto Butantan / Mendelics | Antonio Jorge Martins; Bianca Cechetto Carlos. Mendelics: Bibiana Santos; Claudia Renata dos Santos Barros; Cíntia Bittar; David Schlesinger; David Schlesinger. Hemocentro Ribeirão Preto: Simone Kashima; Debora Botequio Moretti; Dimas Tadeu Covas; Elaine Cristina Marqueze; Elaine Vieira Santos; Elaine Vieira dos Santos; Elisangela Chicaroni Mattos; Erika Freitas; Evandra Strazza Rodrigues; Felipe Allan da Silva da Costa; Flavia Aburjaile; Fábio Sossai Possebon; Guilherme Campos; Guilherme Targino Valente; Heidge Fukumasu; Heidge Fukumasu. USP-Botucatu: Rejane Maria Tommasini Grotto; Helena Lage Ferreira; Instituto Butantan: Dimas Tadeu Covas; Jardelina de Souza Todao Bernardino; Jayme A. Souza-Neto; Jayme Augusto de Souza-Neto; Jessika Cristina Chagas Lesbon; Jorge A. Petroli Marchesi; José Salvatore Leister Patané; João Paulo Kitajima; João Pessoa Araújo Jr.; Leila Sabrina Ullmann; Loyze Paola Oliveira de Lima; Luiz Alcantara; Luiz Aurelio de Campos Crispin. Centro de Genômica Funcional da ESALQ: Luiz Lehmann Coutinho; Luiz Carlos Junior de Alcantara; Luiz Lehmann Coutinho; Lívia Sacchetto; Maisa C. Pereira Parra; Maria Carolina Elias; Marta Giovanetti; Marília Moraes; Maurício Lacerda Nogueira; Maurício Lacerda Nogueira. Prefeitura de Sao Paulo: Melissa Palmieri.; Patricia Akemi Assato; Paula Rahal; Paulo Inacio da Costa; Rafael dos Santos Bezerra; Raquel de Lello Rocha Campos Cassano. NGS Soluções Genômicas: Pilar Drummond Sampaio Corrêa Mariani. FZEA-USP Pirassununga: Mirele Daiana Poleti; Raul Machado Neto; Rejane Maria Tommasini Grotto; Ricardo Augusto Brassaloti; Ricardo Haddad; Rodrigo Tocantins Calado. FAMERP-SJRP: Cecília Artico Banho; Sandra Coccuzzo Sampaio; Sandra Coccuzzo Sampaio Vessoni; Simone Kashima; Svetoslav Nanev Slavov; Vagner Fonseca; Vincent Louis Viala |
| EPI_ISL_2345803 | UBS DR ERNESTO GAION | Instituto Butantan / Mendelics | Antonio Jorge Martins; Claudia Renata dos Santos Barros; David Schlesinger; Debora Botequio Moretti; Dimas Tadeu Covas; Elaine Cristina Marqueze; Elaine Vieira Santos; Evandra Strazza Rodrigues; Heidge Fukumasu; Jayme Augusto de Souza-Neto; José Salvatore Leister Patané; Luiz Alcantara; Luiz Lehmann Coutinho; Maria Carolina Elias; Maurício Lacerda Nogueira; Rafael dos Santos Bezerra; Raul Machado Neto; Rejane Maria Tommasini Grotto; Ricardo Haddad; Sandra Coccuzzo Sampaio Vessoni; Simone Kashima; Svetoslav Nanev Slavov; Vincent Louis Viala |
| EPI_ISL_2345757 | UBS DR WALDOMIRO FERREIRA NEVES SCRPARDO | Instituto Butantan / Mendelics | Antonio Jorge Martins; Claudia Renata dos Santos Barros; David Schlesinger; Debora Botequio Moretti; Dimas Tadeu Covas; Elaine Cristina Marqueze; Elaine Vieira Santos; Evandra Strazza Rodrigues; Heidge Fukumasu; Jayme Augusto de Souza-Neto; José Salvatore Leister Patané; Luiz Alcantara; Luiz Lehmann Coutinho; Maria Carolina Elias; Maurício Lacerda Nogueira; Rafael dos Santos Bezerra; Raul Machado Neto; Rejane Maria Tommasini Grotto; Ricardo Haddad; Sandra Coccuzzo Sampaio Vessoni; Simone Kashima; Svetoslav Nanev Slavov; Vincent Louis Viala |
| EPI_ISL_2345822, EPI_ISL_2345858 | UBS II DE ALVARES MACHADO | Instituto Butantan / Mendelics | Antonio Jorge Martins; Claudia Renata dos Santos Barros; David Schlesinger; Debora Botequio Moretti; Dimas Tadeu Covas; Elaine Cristina Marqueze; Elaine Vieira Santos; Evandra Strazza Rodrigues; Heidge Fukumasu; Jayme Augusto de Souza-Neto; José Salvatore Leister Patané; Luiz Alcantara; Luiz Lehmann Coutinho; Maria Carolina Elias; Maurício Lacerda Nogueira; Rafael dos Santos Bezerra; Raul Machado Neto; Rejane Maria Tommasini Grotto; Ricardo Haddad; Sandra Coccuzzo Sampaio Vessoni; Simone Kashima; Svetoslav Nanev Slavov; Vincent Louis Viala |
| EPI_ISL_2493278, EPI_ISL_2493326, EPI_ISL_2493327, EPI_ISL_2493328, EPI_ISL_2493341, EPI_ISL_2493343, EPI_ISL_2493346, EPI_ISL_2493348, EPI_ISL_2493349, EPI_ISL_2493350, EPI_ISL_2493351, EPI_ISL_2493352, EPI_ISL_2493353, EPI_ISL_2500984 | | | |
| see above | UBS II DE PIRAPOZINHO C SERVICO DE EACS E ESF | Instituto Butantan | Antonio Jorge Martins; Claudia Renata dos Santos Barros; David Schlesinger; Debora Botequio Moretti; Dimas Tadeu Covas; Elaine Cristina Marqueze; Elaine Vieira Santos; Evandra Strazza Rodrigues; Heidge Fukumasu; Jayme Augusto de Souza-Neto; José Salvatore Leister Patané; Luiz Alcantara; Luiz Lehmann Coutinho; Maria Carolina Elias; Maurício Lacerda Nogueira; Rafael dos Santos Bezerra; Raul Machado Neto; Rejane Maria Tommasini Grotto; Ricardo Haddad; Sandra Coccuzzo Sampaio Vessoni; Simone Kashima; Svetoslav Nanev Slavov; Vincent Louis Viala |
| EPI_ISL_2445482, EPI_ISL_2445484 | UBS II DE REGENTE FEIJO | Instituto Butantan | Antonio Jorge Martins; Claudia Renata dos Santos Barros; David Schlesinger; Debora Botequio Moretti; Dimas Tadeu Covas; Elaine Cristina Marqueze; Elaine Vieira Santos; Evandra Strazza Rodrigues; Heidge Fukumasu; Jayme Augusto de Souza-Neto; José Salvatore Leister Patané; Luiz Alcantara; Luiz Lehmann Coutinho; Maria Carolina Elias; Maurício Lacerda Nogueira; Rafael dos Santos Bezerra; Raul Machado Neto; Rejane Maria Tommasini Grotto; Ricardo Haddad; Sandra Coccuzzo Sampaio Vessoni; Simone Kashima; Svetoslav Nanev Slavov; Vincent Louis Viala |
| EPI_ISL_2209618 | UBS II DE REGENTE FEIJO | Instituto Butantan / Mendelics | Antonio Jorge Martins; Claudia Renata dos Santos Barros; David Schlesinger; Debora Botequio Moretti; Dimas Tadeu Covas; Elaine Cristina Marqueze; Elaine Vieira Santos; Evandra Strazza Rodrigues; Heidge Fukumasu; Jayme Augusto de Souza-Neto; José Salvatore Leister Patané; Luiz Alcantara; Luiz Lehmann Coutinho; Maria Carolina Elias; Maurício Lacerda Nogueira; Rafael dos Santos Bezerra; Raul Machado Neto; Rejane Maria Tommasini Grotto; Ricardo Haddad; Sandra Coccuzzo Sampaio Vessoni; Simone Kashima; Svetoslav Nanev Slavov; Vincent Louis Viala |
| EPI_ISL_2445202, EPI_ISL_2445516, EPI_ISL_2445518, EPI_ISL_2445520, EPI_ISL_2445534 | UBS II DE TANABI MILTON MARTINS PERCHES | Instituto Butantan | Antonio Jorge Martins; Claudia Renata dos Santos Barros; David Schlesinger; Debora Botequio Moretti; Dimas Tadeu Covas; Elaine Cristina Marqueze; Elaine Vieira Santos; Evandra Strazza Rodrigues; Heidge Fukumasu; Jayme Augusto de Souza-Neto; José Salvatore Leister Patané; Luiz Alcantara; Luiz Lehmann Coutinho; Maria Carolina Elias; Maurício Lacerda Nogueira; Rafael dos Santos Bezerra; Raul Machado Neto; Rejane Maria Tommasini Grotto; Ricardo Haddad; Sandra Coccuzzo Sampaio Vessoni; Simone Kashima; Svetoslav Nanev Slavov; Vincent Louis Viala |
| EPI_ISL_2445552, EPI_ISL_2445555, EPI_ISL_2473796 | UBS II DR JOSE CARLOS MACUCO JANINI | Instituto Butantan | Antonio Jorge Martins; Claudia Renata dos Santos Barros; David Schlesinger; Debora Botequio Moretti; Dimas Tadeu Covas; Elaine Cristina Marqueze; Elaine Vieira Santos; Evandra Strazza Rodrigues; Heidge Fukumasu; Jayme Augusto de Souza-Neto; José Salvatore Leister Patané; Luiz Alcantara; Luiz Lehmann Coutinho; Maria Carolina Elias; Maurício Lacerda Nogueira; Rafael dos Santos Bezerra; Raul Machado Neto; Rejane Maria Tommasini Grotto; Ricardo Haddad; Sandra Coccuzzo Sampaio Vessoni; Simone Kashima; Svetoslav Nanev Slavov; Vincent Louis Viala |
| EPI_ISL_2209600, EPI_ISL_2210148 | UBS II DR JOSE CARLOS MACUCO JANINI | Instituto Butantan / Mendelics | Antonio Jorge Martins; Claudia Renata dos Santos Barros; David Schlesinger; Debora Botequio Moretti; Dimas Tadeu Covas; Elaine Cristina Marqueze; Elaine Vieira Santos; Evandra Strazza Rodrigues; Heidge Fukumasu; Jayme Augusto de Souza-Neto; José Salvatore Leister Patané; Luiz Alcantara; Luiz Lehmann Coutinho; Maria Carolina Elias; Maurício Lacerda Nogueira; Rafael dos Santos Bezerra; Raul Machado Neto; Rejane Maria Tommasini Grotto; Ricardo Haddad; Sandra Coccuzzo Sampaio Vessoni; Simone Kashima; Svetoslav Nanev Slavov; Vincent Louis Viala |
| EPI_ISL_2493531, EPI_ISL_2493545, EPI_ISL_2493550 | UBS III DE PARIQUERA ACU PARIQUERA ACU | Instituto Butantan | Antonio Jorge Martins; Claudia Renata dos Santos Barros; David Schlesinger; Debora Botequio Moretti; Dimas Tadeu Covas; Elaine Cristina Marqueze; Elaine Vieira Santos; Evandra Strazza Rodrigues; Heidge Fukumasu; Jayme Augusto de Souza-Neto; José Salvatore Leister Patané; Luiz Alcantara; Luiz Lehmann Coutinho; Maria Carolina Elias; Maurício Lacerda Nogueira; Rafael dos Santos Bezerra; Raul Machado Neto; Rejane Maria Tommasini Grotto; Ricardo Haddad; Sandra Coccuzzo Sampaio Vessoni; Simone Kashima; Svetoslav Nanev Slavov; Vincent Louis Viala |
| EPI_ISL_2209494 | UBS III DE PARIQUERA ACU PARIQUERA ACU | Instituto Butantan / Mendelics | Antonio Jorge Martins; Claudia Renata dos Santos Barros; David Schlesinger; Debora Botequio Moretti; Dimas Tadeu Covas; Elaine Cristina Marqueze; Elaine Vieira Santos; Evandra Strazza Rodrigues; Heidge Fukumasu; Jayme Augusto de Souza-Neto; José Salvatore Leister Patané; Luiz Alcantara; Luiz Lehmann Coutinho; Maria Carolina Elias; Maurício Lacerda Nogueira; Rafael dos Santos Bezerra; Raul Machado Neto; Rejane Maria Tommasini Grotto; Ricardo Haddad; Sandra Coccuzzo Sampaio Vessoni; Simone Kashima; Svetoslav Nanev Slavov; Vincent Louis Viala |
| EPI_ISL_2493686, EPI_ISL_2493699 | UBS III DE RANCHARIA | Instituto Butantan | Antonio Jorge Martins; Claudia Renata dos Santos Barros; David Schlesinger; Debora Botequio Moretti; Dimas Tadeu Covas; Elaine Cristina Marqueze; Elaine Vieira Santos; Evandra Strazza Rodrigues; Heidge Fukumasu; Jayme Augusto de Souza-Neto; José Salvatore Leister Patané; Luiz Alcantara; Luiz Lehmann Coutinho; Maria Carolina Elias; Maurício Lacerda Nogueira; Rafael dos Santos Bezerra; Raul Machado Neto; Rejane Maria Tommasini Grotto; Ricardo Haddad; Sandra Coccuzzo Sampaio Vessoni; Simone Kashima; Svetoslav Nanev Slavov; Vincent Louis Viala |
| EPI_ISL_2345666, EPI_ISL_2345710 | UBS III DE RANCHARIA | Instituto Butantan / Mendelics | Antonio Jorge Martins; Claudia Renata dos Santos Barros; David Schlesinger; Debora Botequio Moretti; Dimas Tadeu Covas; Elaine Cristina Marqueze; Elaine Vieira Santos; Evandra Strazza Rodrigues; Heidge Fukumasu; Jayme Augusto de Souza-Neto; José Salvatore Leister Patané; Luiz Alcantara; Luiz Lehmann Coutinho; Maria Carolina Elias; Maurício Lacerda Nogueira; Rafael dos Santos Bezerra; Raul Machado Neto; Rejane Maria Tommasini Grotto; Ricardo Haddad; Sandra Coccuzzo Sampaio Vessoni; Simone Kashima; Svetoslav Nanev Slavov; Vincent Louis Viala |
| EPI_ISL_2445543 | UBS III DR BENITO MALZONE | Instituto Butantan | Antonio Jorge Martins; Claudia Renata dos Santos Barros; David Schlesinger; Debora Botequio Moretti; Dimas Tadeu Covas; Elaine Cristina Marqueze; Elaine Vieira Santos; Evandra Strazza Rodrigues; Heidge Fukumasu; Jayme Augusto de Souza-Neto; José Salvatore Leister Patané; Luiz Alcantara; Luiz Lehmann Coutinho; Maria Carolina Elias; Maurício Lacerda Nogueira; Rafael dos Santos Bezerra; Raul Machado Neto; Rejane Maria Tommasini Grotto; Ricardo Haddad; Sandra Coccuzzo Sampaio Vessoni; Simone Kashima; Svetoslav Nanev Slavov; Vincent Louis Viala |
| EPI_ISL_2345193, EPI_ISL_2345196 | UBS III MURUTINGA DO SUL | Instituto Butantan / ESALQ-Piracicaba | Antonio Jorge Martins; Claudia Renata dos Santos Barros; David Schlesinger; Debora Botequio Moretti; Dimas Tadeu Covas; Elaine Cristina Marqueze; Elaine Vieira Santos; Evandra Strazza Rodrigues; Heidge Fukumasu; Jayme Augusto de Souza-Neto; José Salvatore Leister Patané; Luiz Alcantara; Luiz Lehmann Coutinho; Maria Carolina Elias; Maurício Lacerda Nogueira; Rafael dos Santos Bezerra; Raul Machado Neto; Rejane Maria Tommasini Grotto; Ricardo Haddad; Sandra Coccuzzo Sampaio Vessoni; Simone Kashima; Svetoslav Nanev Slavov; Vincent Louis Viala |
| EPI_ISL_2345223, EPI_ISL_2345225, EPI_ISL_2345228, EPI_ISL_2345229, EPI_ISL_2345230, EPI_ISL_2345231, EPI_ISL_2345232, EPI_ISL_2345233, EPI_ISL_2345234, EPI_ISL_2345235 | | | |
| see above | UBS IV GUARARAPES | Instituto Butantan / ESALQ-Piracicaba | Antonio Jorge Martins; Claudia Renata dos Santos Barros; David Schlesinger; Debora Botequio Moretti; Dimas Tadeu Covas; Elaine Cristina Marqueze; Elaine Vieira Santos; Evandra Strazza Rodrigues; Heidge Fukumasu; Jayme Augusto de Souza-Neto; José Salvatore Leister Patané; Luiz Alcantara; Luiz Lehmann Coutinho; Maria Carolina Elias; Maurício Lacerda Nogueira; Rafael dos Santos Bezerra; Raul Machado Neto; Rejane Maria Tommasini Grotto; Ricardo Haddad; Sandra Coccuzzo Sampaio Vessoni; Simone Kashima; Svetoslav Nanev Slavov; Vincent Louis Viala |
| EPI_ISL_1967285, EPI_ISL_1967286 | UBS JARDIM CABUCU | Instituto Butantan / Mendelics | Antonio Jorge Martins; Bianca Cechetto Carlos. Mendelics: Bibiana Santos; Claudia Renata dos Santos Barros; Cíntia Bittar; David Schlesinger. Hemocentro Ribeirão Preto: Simone Kashima; Debora Botequio Moretti; Elaine Cristina Marqueze; Elaine Vieira dos Santos; Elisangela Chicaroni Mattos; Erika Freitas; Evandra Strazza Rodrigues; Felipe Allan da Silva da Costa; Flavia Aburjaile; Fábio Sossai Possebon; Guilherme Campos; Guilherme Targino Valente; Heidge Fukumasu. USP-Botucatu: Rejane Maria Tommasini Grotto; Helena Lage Ferreira; Instituto Butantan: Dimas Tadeu Covas; Jardelina de Souza Todao Bernardino; Jayme A. Souza-Neto; Jessika Cristina Chagas Lesbon; Jorge A. Petroli Marchesi; José Salvatore Leister Patané; João Paulo Kitajima; João Pessoa Araújo Jr.; Leila Sabrina Ullmann; Loyze Paola Oliveira de Lima; Luiz Aurelio de Campos Crispin. Centro de Genômica Funcional da ESALQ: Luiz Lehmann Coutinho; Luiz Carlos Junior de Alcantara; Lívia Sacchetto; Maisa C. Pereira Parra; Maria Carolina Elias; Marta Giovanetti; Marília Moraes; Maurício Lacerda Nogueira. Prefeitura de Sao Paulo: Melissa Palmieri.; Patricia Akemi Assato; Paula Rahal; Paulo Inacio da Costa; Rafael dos Santos Bezerra; Raquel de Lello Rocha Campos Cassano. NGS Soluções Genômicas: Pilar Drummond Sampaio Corrêa Mariani. FZEA-USP Pirassununga: Mirele Daiana Poleti; Raul Machado Neto; Ricardo Augusto Brassaloti; Ricardo Haddad; Rodrigo Tocantins Calado. FAMERP-SJRP: Cecília Artico Banho; Sandra Coccuzzo Sampaio; Svetoslav Nanev Slavov; Vagner Fonseca; Vincent Louis Viala |
| EPI_ISL_2171000 | UBS JARDIM MUNHOZ | Instituto Butantan / Mendelics | Antonio Jorge Martins; Bianca Cechetto Carlos. Mendelics: Bibiana Santos; Claudia Renata dos Santos Barros; Cíntia Bittar; David Schlesinger. Hemocentro Ribeirão Preto: Simone Kashima; Debora Botequio Moretti; Elaine Cristina Marqueze; Elaine Vieira dos Santos; Elisangela Chicaroni Mattos; Erika Freitas; Evandra Strazza Rodrigues; Felipe Allan da Silva da Costa; Flavia Aburjaile; Fábio Sossai Possebon; Guilherme Campos; Guilherme Targino Valente; Heidge Fukumasu. USP-Botucatu: Rejane Maria Tommasini Grotto; Helena Lage Ferreira; Instituto Butantan: Dimas Tadeu Covas; Jardelina de Souza Todao Bernardino; Jayme A. Souza-Neto; Jessika Cristina Chagas Lesbon; Jorge A. Petroli Marchesi; José Salvatore Leister Patané; João Paulo Kitajima; João Pessoa Araújo Jr.; Leila Sabrina Ullmann; Loyze Paola Oliveira de Lima; Luiz Aurelio de Campos Crispin. Centro de Genômica Funcional da ESALQ: Luiz Lehmann Coutinho; Luiz Carlos Junior de Alcantara; Lívia Sacchetto; Maisa C. Pereira Parra; Maria Carolina Elias; Marta Giovanetti; Marília Moraes; Maurício Lacerda Nogueira. Prefeitura de Sao Paulo: Melissa Palmieri.; Patricia Akemi Assato; Paula Rahal; Paulo Inacio da Costa; Rafael dos Santos Bezerra; Raquel de Lello Rocha Campos Cassano. NGS Soluções Genômicas: Pilar Drummond Sampaio Corrêa Mariani. FZEA-USP Pirassununga: Mirele Daiana Poleti; Raul Machado Neto; Ricardo Augusto Brassaloti; Ricardo Haddad; Rodrigo Tocantins Calado. FAMERP-SJRP: Cecília Artico Banho; Sandra Coccuzzo Sampaio; Svetoslav Nanev Slavov; Vagner Fonseca; Vincent Louis Viala |
| EPI_ISL_2344902 | UBS JARDIM VITORINO IPERO | Instituto Butantan / ESALQ-Piracicaba | Antonio Jorge Martins; Claudia Renata dos Santos Barros; David Schlesinger; Debora Botequio Moretti; Dimas Tadeu Covas; Elaine Cristina Marqueze; Elaine Vieira Santos; Evandra Strazza Rodrigues; Heidge Fukumasu; Jayme Augusto de Souza-Neto; José Salvatore Leister Patané; Luiz Alcantara; Luiz Lehmann Coutinho; Maria Carolina Elias; Maurício Lacerda Nogueira; Rafael dos Santos Bezerra; Raul Machado Neto; Rejane Maria Tommasini Grotto; Ricardo Haddad; Sandra Coccuzzo Sampaio Vessoni; Simone Kashima; Svetoslav Nanev Slavov; Vincent Louis Viala |
| EPI_ISL_2344825 | UBS JD SALTENSE | Instituto Butantan / ESALQ-Piracicaba | Antonio Jorge Martins; Claudia Renata dos Santos Barros; David Schlesinger; Debora Botequio Moretti; Dimas Tadeu Covas; Elaine Cristina Marqueze; Elaine Vieira Santos; Evandra Strazza Rodrigues; Heidge Fukumasu; Jayme Augusto de Souza-Neto; José Salvatore Leister Patané; Luiz Alcantara; Luiz Lehmann Coutinho; Maria Carolina Elias; Maurício Lacerda Nogueira; Rafael dos Santos Bezerra; Raul Machado Neto; Rejane Maria Tommasini Grotto; Ricardo Haddad; Sandra Coccuzzo Sampaio Vessoni; Simone Kashima; Svetoslav Nanev Slavov; Vincent Louis Viala |
| EPI_ISL_2493663 | UBS LUIS FACHIN IPIGUA | Instituto Butantan | Antonio Jorge Martins; Claudia Renata dos Santos Barros; David Schlesinger; Debora Botequio Moretti; Dimas Tadeu Covas; Elaine Cristina Marqueze; Elaine Vieira Santos; Evandra Strazza Rodrigues; Heidge Fukumasu; Jayme Augusto de Souza-Neto; José Salvatore Leister Patané; Luiz Alcantara; Luiz Lehmann Coutinho; Maria Carolina Elias; Maurício Lacerda Nogueira; Rafael dos Santos Bezerra; Raul Machado Neto; Rejane Maria Tommasini Grotto; Ricardo Haddad; Sandra Coccuzzo Sampaio Vessoni; Simone Kashima; Svetoslav Nanev Slavov; Vincent Louis Viala |
| EPI_ISL_2493584 | UBS MARIA IZABEL | Instituto Butantan | Antonio Jorge Martins; Claudia Renata dos Santos Barros; David Schlesinger; Debora Botequio Moretti; Dimas Tadeu Covas; Elaine Cristina Marqueze; Elaine Vieira Santos; Evandra Strazza Rodrigues; Heidge Fukumasu; Jayme Augusto de Souza-Neto; José Salvatore Leister Patané; Luiz Alcantara; Luiz Lehmann Coutinho; Maria Carolina Elias; Maurício Lacerda Nogueira; Rafael dos Santos Bezerra; Raul Machado Neto; Rejane Maria Tommasini Grotto; Ricardo Haddad; Sandra Coccuzzo Sampaio Vessoni; Simone Kashima; Svetoslav Nanev Slavov; Vincent Louis Viala |
| EPI_ISL_2445198, EPI_ISL_2445199, EPI_ISL_2445200, EPI_ISL_2445201, EPI_ISL_2445522, EPI_ISL_2445524, EPI_ISL_2473714, EPI_ISL_2473715 | | | |
| see above | UBS PASTOR MARCELINO DEUNGARO RIO GRANDE | Instituto Butantan | Antonio Jorge Martins; Claudia Renata dos Santos Barros; David Schlesinger; Debora Botequio Moretti; Dimas Tadeu Covas; Elaine Cristina Marqueze; Elaine Vieira Santos; Evandra Strazza Rodrigues; Heidge Fukumasu; Jayme Augusto de Souza-Neto; José Salvatore Leister Patané; Luiz Alcantara; Luiz Lehmann Coutinho; Maria Carolina Elias; Maurício Lacerda Nogueira; Rafael dos Santos Bezerra; Raul Machado Neto; Rejane Maria Tommasini Grotto; Ricardo Haddad; Sandra Coccuzzo Sampaio Vessoni; Simone Kashima; Svetoslav Nanev Slavov; Vincent Louis Viala |
| EPI_ISL_2493293 | UBS PAULO HUMBERTO DE FIGUEIREDO | Instituto Butantan | Antonio Jorge Martins; Claudia Renata dos Santos Barros; David Schlesinger; Debora Botequio Moretti; Dimas Tadeu Covas; Elaine Cristina Marqueze; Elaine Vieira Santos; Evandra Strazza Rodrigues; Heidge Fukumasu; Jayme Augusto de Souza-Neto; José Salvatore Leister Patané; Luiz Alcantara; Luiz Lehmann |

| | | | |
|---|---|---|---|
| | | | Coutinho; Maria Carolina Elias; Maurício Lacerda Nogueira; Rafael dos Santos Bezerra; Raul Machado Neto; Rejane Maria Tommasini Grotto; Ricardo Haddad; Sandra Coccuzzo Sampaio Vessoni; Simone Kashima; Svetoslav Nanev Slavov; Vincent Louis Viala |
| EPI_ISL_2445254 | UBS SAO LOURENCO DA SERRA | Instituto Butantan | Antonio Jorge Martins; Claudia Renata dos Santos Barros; David Schlesinger; Debora Botequio Moretti; Dimas Tadeu Covas; Elaine Cristina Marqueze; Elaine Vieira Santos; Evandra Strazza Rodrigues; Heidge Fukumasu; Jayme Augusto de Souza-Neto; José Salvatore Leister Patané; Luiz Alcantara; Luiz Lehmann Coutinho; Maria Carolina Elias; Maurício Lacerda Nogueira; Rafael dos Santos Bezerra; Raul Machado Neto; Rejane Maria Tommasini Grotto; Ricardo Haddad; Sandra Coccuzzo Sampaio Vessoni; Simone Kashima; Svetoslav Nanev Slavov; Vincent Louis Viala |
| EPI_ISL_2345054 | UBS SAO LOURENCO DA SERRA | Instituto Butantan / ESALQ-Piracicaba | Antonio Jorge Martins; Claudia Renata dos Santos Barros; David Schlesinger; Debora Botequio Moretti; Dimas Tadeu Covas; Elaine Cristina Marqueze; Elaine Vieira Santos; Evandra Strazza Rodrigues; Heidge Fukumasu; Jayme Augusto de Souza-Neto; José Salvatore Leister Patané; Luiz Alcantara; Luiz Lehmann Coutinho; Maria Carolina Elias; Maurício Lacerda Nogueira; Rafael dos Santos Bezerra; Raul Machado Neto; Rejane Maria Tommasini Grotto; Ricardo Haddad; Sandra Coccuzzo Sampaio Vessoni; Simone Kashima; Svetoslav Nanev Slavov; Vincent Louis Viala |
| EPI_ISL_2445188, EPI_ISL_2473710 | UBS VILA RIBEIRO | Instituto Butantan | Antonio Jorge Martins; Claudia Renata dos Santos Barros; David Schlesinger; Debora Botequio Moretti; Dimas Tadeu Covas; Elaine Cristina Marqueze; Elaine Vieira Santos; Evandra Strazza Rodrigues; Heidge Fukumasu; Jayme Augusto de Souza-Neto; José Salvatore Leister Patané; Luiz Alcantara; Luiz Lehmann Coutinho; Maria Carolina Elias; Maurício Lacerda Nogueira; Rafael dos Santos Bezerra; Raul Machado Neto; Rejane Maria Tommasini Grotto; Ricardo Haddad; Sandra Coccuzzo Sampaio Vessoni; Simone Kashima; Svetoslav Nanev Slavov; Vincent Louis Viala |
| EPI_ISL_2345134 | UBS VILA RIBEIRO | Instituto Butantan / ESALQ-Piracicaba | Antonio Jorge Martins; Claudia Renata dos Santos Barros; David Schlesinger; Debora Botequio Moretti; Dimas Tadeu Covas; Elaine Cristina Marqueze; Elaine Vieira Santos; Evandra Strazza Rodrigues; Heidge Fukumasu; Jayme Augusto de Souza-Neto; José Salvatore Leister Patané; Luiz Alcantara; Luiz Lehmann Coutinho; Maria Carolina Elias; Maurício Lacerda Nogueira; Rafael dos Santos Bezerra; Raul Machado Neto; Rejane Maria Tommasini Grotto; Ricardo Haddad; Sandra Coccuzzo Sampaio Vessoni; Simone Kashima; Svetoslav Nanev Slavov; Vincent Louis Viala |
| EPI_ISL_3419583 | UC Davis Genome Center and Healthy Davis Together | UC Davis Genome Center | UC Davis |
| EPI_ISL_3321297, EPI_ISL_3321303, EPI_ISL_3535720 | UC-Christus Clinical Hospital Laboratory | Laboratory of Molecular Virology, School of Medicine, Pontificia Universidad Catolica de Chile | Ana Maira Guzman; Andres E. Munoz-Marcos; Catalina Pardo-Roa; Eileen Serrano; Erick Salinas; Estefany Poblete; Francisco Melo; Jorge Levican; Leonardo I. Almonacid; Maria Jose Avendano; Maria Patricia Vega; Rafael A. Medina; Ricardo Enrique de la Barra; Tamara Garcia-Salum |
| EPI_ISL_3104695, EPI_ISL_3104696, EPI_ISL_3104697, EPI_ISL_3104698, EPI_ISL_3104699, EPI_ISL_3104700 | UCC (Universidad Central del Caribe) | Grubaugh Lab - Yale School of Public Health | Anderson Brito; Andrea Firpo; Annie Watkins; Chaney Kalinich; Chantal Vogels; David Rivera Aponte; Ernesto Vazquez; Fabiola Cruz-Lopez; Fabiola Fontanet; Isabel Ott; Jessica Rothman; Joseph Fauver; Karolane Gonzalez; Kendall Billig; Mallery Breban; Mary Petrone; Natalie Machargo; Nathan Grubaugh; Patricia Serrano; Tania Mitwalli; Tara Alpert; Tobias Koch; Vianca Aponte |
| EPI_ISL_2444195 | UCHOA | Instituto Butantan / FAMERP | Antonio Jorge Martins; Claudia Renata dos Santos Barros; David Schlesinger; Debora Botequio Moretti; Dimas Tadeu Covas; Elaine Cristina Marqueze; Elaine Vieira Santos; Evandra Strazza Rodrigues; Heidge Fukumasu; Jayme Augusto de Souza-Neto; José Salvatore Leister Patané; Luiz Alcantara; Luiz Lehmann Coutinho; Maria Carolina Elias; Maurício Lacerda Nogueira; Rafael dos Santos Bezerra; Raul Machado Neto; Rejane Maria Tommasini Grotto; Ricardo Haddad; Sandra Coccuzzo Sampaio Vessoni; Simone Kashima; Svetoslav Nanev Slavov; Vincent Louis Viala |
| EPI_ISL_2405167, EPI_ISL_2617950, EPI_ISL_2620751 | UCLA Clinical Micro Lab | Los Angeles County Public Health Laboratories | P. Hemarajata et al. |
| EPI_ISL_2547290, EPI_ISL_2547291, EPI_ISL_2547292, EPI_ISL_2547299, EPI_ISL_2547312, EPI_ISL_2547314, EPI_ISL_2547315, EPI_ISL_2547316 | | | |
| see above | UHTL, University Hospitals | UHTL, University Hospitals | Alouani, D.; Sadri, N.; Song, X. |
| EPI_ISL_2448598 | ULSS 2 Marca Trevigiana | Istituto Zooprofilattico Sperimentale delle Venezie | Adelaide Milani; Alessia Schivo; Alice Fusaro; Ambra Pastori; Annalisa Salviato; Antonia Ricci; Calogero Terregino; Edoardo Giussani; Elisa Palumbo; Erika Giorgia Quaranta; Isabella Monne; Luca Tassoni |
| EPI_ISL_2448471 | ULSS 5 Polesana | Istituto Zooprofilattico Sperimentale delle Venezie | Adelaide Milani; Alessia Schivo; Alice Fusaro; Ambra Pastori; Annalisa Salviato; Antonia Ricci; Calogero Terregino; Edoardo Giussani; Elisa Palumbo; Erika Giorgia Quaranta; Isabella Monne; Luca Tassoni |
| EPI_ISL_3409608, EPI_ISL_3410080 | UMass Memorial Medical Center | Center for Microbiome Research | Doyle V. Ward; Karl J. Simon; Richard T. Ellison III |
| EPI_ISL_2493585, EPI_ISL_2493588 | UNIDADE BASICA DE SAUDE DE TARUMA | Instituto Butantan | Antonio Jorge Martins; Claudia Renata dos Santos Barros; David Schlesinger; Debora Botequio Moretti; Dimas Tadeu Covas; Elaine Cristina Marqueze; Elaine Vieira Santos; Evandra Strazza Rodrigues; Heidge Fukumasu; Jayme Augusto de Souza-Neto; José Salvatore Leister Patané; Luiz Alcantara; Luiz Lehmann Coutinho; Maria Carolina Elias; Maurício Lacerda Nogueira; Rafael dos Santos Bezerra; Raul Machado Neto; Rejane Maria Tommasini Grotto; Ricardo Haddad; Sandra Coccuzzo Sampaio Vessoni; Simone Kashima; Svetoslav Nanev Slavov; Vincent Louis Viala |
| EPI_ISL_2345203, EPI_ISL_2345204 | UNIDADE BASICA DE SAUDE DE TARUMA | Instituto Butantan / ESALQ-Piracicaba | Antonio Jorge Martins; Claudia Renata dos Santos Barros; David Schlesinger; Debora Botequio Moretti; Dimas Tadeu Covas; Elaine Cristina Marqueze; Elaine Vieira Santos; Evandra Strazza Rodrigues; Heidge Fukumasu; Jayme Augusto de Souza-Neto; José Salvatore Leister Patané; Luiz Alcantara; Luiz Lehmann Coutinho; Maria Carolina Elias; Maurício Lacerda Nogueira; Rafael dos Santos Bezerra; Raul Machado Neto; Rejane Maria Tommasini Grotto; Ricardo Haddad; Sandra Coccuzzo Sampaio Vessoni; Simone Kashima; Svetoslav Nanev Slavov; Vincent Louis Viala |
| EPI_ISL_2494152 | UNIDADE BASICA DE SAUDE DR MATHEUS GABRIEL BONASSA | Instituto Butantan | Antonio Jorge Martins; Claudia Renata dos Santos Barros; David Schlesinger; Debora Botequio Moretti; Dimas Tadeu Covas; Elaine Cristina Marqueze; Elaine Vieira Santos; Evandra Strazza Rodrigues; Heidge Fukumasu; Jayme Augusto de Souza-Neto; José Salvatore Leister Patané; Luiz Alcantara; Luiz Lehmann Coutinho; Maria Carolina Elias; Maurício Lacerda Nogueira; Rafael dos Santos Bezerra; Raul Machado Neto; Rejane Maria Tommasini Grotto; Ricardo Haddad; Sandra Coccuzzo Sampaio Vessoni; Simone Kashima; Svetoslav Nanev Slavov; Vincent Louis Viala |
| EPI_ISL_2493196, EPI_ISL_2493197, EPI_ISL_2493311, EPI_ISL_2493314, EPI_ISL_2493319, EPI_ISL_2493457, EPI_ISL_2493458 | | | |
| see above | UNIDADE BASICA DE SAUDE HORTOLANDIA PACS | Instituto Butantan | Antonio Jorge Martins; Claudia Renata dos Santos Barros; David Schlesinger; Debora Botequio Moretti; Dimas Tadeu Covas; Elaine Cristina Marqueze; Elaine Vieira Santos; Evandra Strazza Rodrigues; Heidge Fukumasu; Jayme Augusto de Souza-Neto; José Salvatore Leister Patané; Luiz Alcantara; Luiz Lehmann Coutinho; Maria Carolina Elias; Maurício Lacerda Nogueira; Rafael dos Santos Bezerra; Raul Machado Neto; Rejane Maria Tommasini Grotto; Ricardo Haddad; Sandra Coccuzzo Sampaio Vessoni; Simone Kashima; Svetoslav Nanev Slavov; Vincent Louis Viala |
| EPI_ISL_2209732, EPI_ISL_2210126, EPI_ISL_2345770 | UNIDADE BASICA DE SAUDE II ESTRELA DO NORTE | Instituto Butantan / Mendelics | Antonio Jorge Martins; Claudia Renata dos Santos Barros; David Schlesinger; Debora Botequio Moretti; Dimas Tadeu Covas; Elaine Cristina Marqueze; Elaine Vieira Santos; Evandra Strazza Rodrigues; Heidge Fukumasu; Jayme Augusto de Souza-Neto; José Salvatore Leister Patané; Luiz Alcantara; Luiz Lehmann Coutinho; Maria Carolina Elias; Maurício Lacerda Nogueira; Rafael dos Santos Bezerra; Raul Machado Neto; Rejane Maria Tommasini Grotto; Ricardo Haddad; Sandra Coccuzzo Sampaio Vessoni; Simone Kashima; Svetoslav Nanev Slavov; Vincent Louis Viala |
| EPI_ISL_2210141, EPI_ISL_2345673, EPI_ISL_2345719 | UNIDADE BASICA DE SAUDE JARDIM DO LAGO | Instituto Butantan / Mendelics | Antonio Jorge Martins; Claudia Renata dos Santos Barros; David Schlesinger; Debora Botequio Moretti; Dimas Tadeu Covas; Elaine Cristina Marqueze; Elaine Vieira Santos; Evandra Strazza Rodrigues; Heidge Fukumasu; Jayme Augusto de Souza-Neto; José Salvatore Leister Patané; Luiz Alcantara; Luiz Lehmann Coutinho; Maria Carolina Elias; Maurício Lacerda Nogueira; Rafael dos Santos Bezerra; Raul Machado Neto; Rejane Maria Tommasini Grotto; Ricardo Haddad; Sandra Coccuzzo Sampaio Vessoni; Simone Kashima; Svetoslav Nanev Slavov; Vincent Louis Viala |
| EPI_ISL_2445463, EPI_ISL_2493294, EPI_ISL_2493295 | UNIDADE BASICA SAUDE NATAL ABADIO LACERDA | Instituto Butantan | Antonio Jorge Martins; Claudia Renata dos Santos Barros; David Schlesinger; Debora Botequio Moretti; Dimas Tadeu Covas; Elaine Cristina Marqueze; Elaine Vieira Santos; Evandra Strazza Rodrigues; Heidge Fukumasu; Jayme Augusto de Souza-Neto; José Salvatore Leister Patané; Luiz Alcantara; Luiz Lehmann Coutinho; Maria Carolina Elias; Maurício Lacerda Nogueira; Rafael dos Santos Bezerra; Raul Machado Neto; Rejane Maria Tommasini Grotto; Ricardo Haddad; Sandra Coccuzzo Sampaio Vessoni; Simone Kashima; Svetoslav Nanev Slavov; Vincent Louis Viala |
| EPI_ISL_2345789 | UNIDADE BASICA SAUDE NATAL ABADIO LACERDA | Instituto Butantan / Mendelics | Antonio Jorge Martins; Claudia Renata dos Santos Barros; David Schlesinger; Debora Botequio Moretti; Dimas Tadeu Covas; Elaine Cristina Marqueze; Elaine Vieira Santos; Evandra Strazza Rodrigues; Heidge Fukumasu; Jayme Augusto de Souza-Neto; José Salvatore Leister Patané; Luiz Alcantara; Luiz Lehmann Coutinho; Maria Carolina Elias; Maurício Lacerda Nogueira; Rafael dos Santos Bezerra; Raul Machado Neto; Rejane Maria Tommasini Grotto; Ricardo Haddad; Sandra Coccuzzo Sampaio Vessoni; Simone Kashima; Svetoslav Nanev Slavov; Vincent Louis Viala |
| EPI_ISL_2493473, EPI_ISL_2493480, EPI_ISL_2493484, EPI_ISL_2493492 | UNIDADE DE PRONTO ATENDIMENTO | Instituto Butantan | Antonio Jorge Martins; Claudia Renata dos Santos Barros; David Schlesinger; Debora Botequio Moretti; Dimas Tadeu Covas; Elaine Cristina Marqueze; Elaine Vieira Santos; Evandra Strazza Rodrigues; Heidge Fukumasu; Jayme Augusto de Souza-Neto; José Salvatore Leister Patané; Luiz Alcantara; Luiz Lehmann Coutinho; Maria Carolina Elias; Maurício Lacerda Nogueira; Rafael dos Santos Bezerra; Raul Machado Neto; Rejane Maria Tommasini Grotto; Ricardo Haddad; Sandra Coccuzzo Sampaio Vessoni; Simone Kashima; Svetoslav Nanev Slavov; Vincent Louis Viala |
| EPI_ISL_2344885, EPI_ISL_2344886 | UNIDADE DE PRONTO ATENDIMENTO DE JALES | Instituto Butantan / ESALQ-Piracicaba | Antonio Jorge Martins; Claudia Renata dos Santos Barros; David Schlesinger; Debora Botequio Moretti; Dimas Tadeu Covas; Elaine Cristina Marqueze; Elaine Vieira Santos; Evandra Strazza Rodrigues; Heidge Fukumasu; Jayme Augusto de Souza-Neto; José Salvatore Leister Patané; Luiz Alcantara; Luiz Lehmann Coutinho; Maria Carolina Elias; Maurício Lacerda Nogueira; Rafael dos Santos Bezerra; Raul Machado Neto; Rejane Maria Tommasini Grotto; Ricardo Haddad; Sandra Coccuzzo Sampaio Vessoni; Simone Kashima; Svetoslav Nanev Slavov; Vincent Louis Viala |
| EPI_ISL_2345085, EPI_ISL_2345089, EPI_ISL_2345092, EPI_ISL_2345098, EPI_ISL_2345102, EPI_ISL_2345103, EPI_ISL_2345106 | | | |
| see above | UNIDADE DE PRONTO ATENDIMENTO DE VARZEA PAULISTA UPA II | Instituto Butantan / ESALQ-Piracicaba | Antonio Jorge Martins; Claudia Renata dos Santos Barros; David Schlesinger; Debora Botequio Moretti; Dimas Tadeu Covas; Elaine Cristina Marqueze; Elaine Vieira Santos; Evandra Strazza Rodrigues; Heidge Fukumasu; Jayme Augusto de Souza-Neto; José Salvatore Leister Patané; Luiz Alcantara; Luiz Lehmann Coutinho; Maria Carolina Elias; Maurício Lacerda Nogueira; Rafael dos Santos Bezerra; Raul Machado Neto; Rejane Maria Tommasini Grotto; Ricardo Haddad; Sandra Coccuzzo Sampaio Vessoni; Simone Kashima; Svetoslav Nanev Slavov; Vincent Louis Viala |
| EPI_ISL_2345080, EPI_ISL_2345083, EPI_ISL_2345100, EPI_ISL_2345105, EPI_ISL_2345119 | UNIDADE DE PRONTO ATENDIMENTO UPA DRA ANA OLIVIA BENTIVOGLIO | Instituto Butantan / ESALQ-Piracicaba | Antonio Jorge Martins; Claudia Renata dos Santos Barros; David Schlesinger; Debora Botequio Moretti; Dimas Tadeu Covas; Elaine Cristina Marqueze; Elaine Vieira Santos; Evandra Strazza Rodrigues; Heidge Fukumasu; Jayme Augusto de Souza-Neto; José Salvatore Leister Patané; Luiz Alcantara; Luiz Lehmann Coutinho; Maria Carolina Elias; Maurício Lacerda Nogueira; Rafael dos Santos Bezerra; Raul Machado Neto; Rejane Maria Tommasini Grotto; Ricardo Haddad; Sandra Coccuzzo Sampaio Vessoni; Simone Kashima; Svetoslav Nanev Slavov; Vincent Louis Viala |
| EPI_ISL_2345127, EPI_ISL_2345135 | UNIDADE DE PRONTO ATENDIMENTO UPA I SANTA ISABEL | Instituto Butantan / ESALQ-Piracicaba | Antonio Jorge Martins; Claudia Renata dos Santos Barros; David Schlesinger; Debora Botequio Moretti; Dimas Tadeu Covas; Elaine Cristina Marqueze; Elaine Vieira Santos; Evandra Strazza Rodrigues; Heidge Fukumasu; Jayme Augusto de Souza-Neto; José Salvatore Leister Patané; Luiz Alcantara; Luiz Lehmann Coutinho; Maria Carolina Elias; Maurício Lacerda Nogueira; Rafael dos Santos Bezerra; Raul Machado Neto; Rejane Maria Tommasini Grotto; Ricardo Haddad; Sandra Coccuzzo Sampaio Vessoni; Simone Kashima; Svetoslav Nanev Slavov; Vincent Louis Viala |
| EPI_ISL_2494305 | UNIDADE DE SAUDE DA FAMILIA NOVO HORIZONTE | Instituto Butantan | Antonio Jorge Martins; Claudia Renata dos Santos Barros; David Schlesinger; Debora Botequio Moretti; Dimas Tadeu Covas; Elaine Cristina Marqueze; Elaine Vieira Santos; Evandra Strazza Rodrigues; Heidge Fukumasu; Jayme Augusto de Souza-Neto; José Salvatore Leister Patané; Luiz Alcantara; Luiz Lehmann Coutinho; Maria Carolina Elias; Maurício Lacerda Nogueira; Rafael dos Santos Bezerra; Raul Machado Neto; Rejane Maria Tommasini Grotto; Ricardo Haddad; Sandra Coccuzzo Sampaio Vessoni; Simone Kashima; Svetoslav Nanev Slavov; Vincent Louis Viala |
| EPI_ISL_2345212, EPI_ISL_2345227 | UNIDADE DE SAUDE DA FAMILIA NOVO HORIZONTE | Instituto Butantan / ESALQ-Piracicaba | Antonio Jorge Martins; Claudia Renata dos Santos Barros; David Schlesinger; Debora Botequio Moretti; Dimas Tadeu Covas; Elaine Cristina Marqueze; Elaine Vieira Santos; Evandra Strazza Rodrigues; Heidge Fukumasu; Jayme Augusto de Souza-Neto; José Salvatore Leister Patané; Luiz Alcantara; Luiz Lehmann Coutinho; Maria Carolina Elias; Maurício Lacerda Nogueira; Rafael dos Santos Bezerra; Raul Machado Neto; Rejane Maria Tommasini Grotto; Ricardo Haddad; Sandra Coccuzzo Sampaio Vessoni; Simone Kashima; Svetoslav Nanev Slavov; Vincent Louis Viala |
| EPI_ISL_2493526 | UNIDADE DE SAUDE DE ITOBI ALCIBIADES PIRES | Instituto Butantan | Antonio Jorge Martins; Claudia Renata dos Santos Barros; David Schlesinger; Debora Botequio Moretti; Dimas Tadeu Covas; Elaine Cristina Marqueze; Elaine Vieira Santos; Evandra Strazza Rodrigues; Heidge Fukumasu; Jayme Augusto de Souza-Neto; José Salvatore Leister Patané; Luiz Alcantara; Luiz Lehmann Coutinho; Maria Carolina Elias; Maurício Lacerda Nogueira; Rafael dos Santos Bezerra; Raul Machado Neto; Rejane Maria Tommasini Grotto; Ricardo Haddad; Sandra Coccuzzo Sampaio Vessoni; Simone Kashima; Svetoslav Nanev Slavov; Vincent Louis Viala |
| EPI_ISL_2493297, EPI_ISL_2493298 | UNIDADE DE SAUDE DR PHEBO DE OLIVEIRA ROGE FERREIRA | Instituto Butantan | Antonio Jorge Martins; Claudia Renata dos Santos Barros; David Schlesinger; Debora Botequio Moretti; Dimas Tadeu Covas; Elaine Cristina Marqueze; Elaine Vieira Santos; Evandra Strazza Rodrigues; Heidge Fukumasu; Jayme Augusto de Souza-Neto; José Salvatore Leister Patané; Luiz Alcantara; Luiz Lehmann Coutinho; Maria Carolina Elias; Maurício Lacerda Nogueira; Rafael dos Santos Bezerra; Raul Machado Neto; Rejane Maria Tommasini Grotto; Ricardo Haddad; Sandra Coccuzzo Sampaio Vessoni; Simone Kashima; Svetoslav Nanev Slavov; Vincent Louis Viala |
| EPI_ISL_2445480 | UNIDADE DE VIGILANCIA EM SAUDE | Instituto Butantan | Antonio Jorge Martins; Claudia Renata dos Santos Barros; David Schlesinger; Debora Botequio Moretti; Dimas Tadeu Covas; Elaine Cristina Marqueze; Elaine Vieira Santos; Evandra Strazza Rodrigues; Heidge Fukumasu; Jayme Augusto de Souza-Neto; José Salvatore Leister Patané; Luiz Alcantara; Luiz Lehmann Coutinho; Maria Carolina Elias; Maurício Lacerda Nogueira; Rafael dos Santos Bezerra; Raul Machado Neto; Rejane Maria Tommasini Grotto; Ricardo Haddad; Sandra Coccuzzo Sampaio Vessoni; Simone Kashima; Svetoslav Nanev Slavov; Vincent Louis Viala |
| EPI_ISL_2345269, EPI_ISL_2345270, EPI_ISL_2345272, EPI_ISL_2345275, EPI_ISL_2345277, EPI_ISL_2345279, EPI_ISL_2345281, EPI_ISL_2345285, EPI_ISL_2345286, EPI_ISL_2345287 | | | |
| see above | UNIDADE DE VIGILANCIA EM SAUDE | Instituto Butantan / ESALQ-Piracicaba | Antonio Jorge Martins; Claudia Renata dos Santos Barros; David Schlesinger; Debora Botequio Moretti; Dimas Tadeu Covas; Elaine Cristina Marqueze; Elaine Vieira Santos; Evandra Strazza Rodrigues; Heidge Fukumasu; Jayme Augusto de Souza-Neto; José Salvatore Leister Patané; Luiz Alcantara; Luiz Lehmann Coutinho; Maria Carolina Elias; Maurício Lacerda Nogueira; Rafael dos Santos Bezerra; Raul Machado Neto; Rejane Maria Tommasini Grotto; Ricardo Haddad; Sandra Coccuzzo Sampaio Vessoni; Simone Kashima; Svetoslav Nanev Slavov; Vincent Louis Viala |
| EPI_ISL_2445236, EPI_ISL_2445237 | UNIDADE MISTA DE IGUAPE | Instituto Butantan | Antonio Jorge Martins; Claudia Renata dos Santos Barros; David Schlesinger; Debora Botequio Moretti; Dimas Tadeu Covas; Elaine Cristina Marqueze; Elaine Vieira Santos; Evandra Strazza Rodrigues; Heidge Fukumasu; Jayme Augusto de Souza-Neto; José Salvatore Leister Patané; Luiz Alcantara; Luiz Lehmann Coutinho; Maria Carolina Elias; Maurício Lacerda Nogueira; Rafael dos Santos Bezerra; Raul Machado Neto; Rejane Maria Tommasini Grotto; Ricardo Haddad; Sandra Coccuzzo Sampaio Vessoni; Simone Kashima; Svetoslav Nanev Slavov; Vincent Louis Viala |
| EPI_ISL_2493602, EPI_ISL_2493604, EPI_ISL_2493605, EPI_ISL_2493607, EPI_ISL_2493611, EPI_ISL_2493614, EPI_ISL_2493616, EPI_ISL_2493617, EPI_ISL_2493620, EPI_ISL_2493623 | | | |
| see above | UNIDADE MISTA DE LUIZ ANTONIO | Instituto Butantan | Antonio Jorge Martins; Claudia Renata dos Santos Barros; David Schlesinger; Debora Botequio Moretti; Dimas Tadeu Covas; Elaine Cristina Marqueze; Elaine Vieira Santos; Evandra Strazza Rodrigues; Heidge Fukumasu; Jayme Augusto de Souza-Neto; José Salvatore Leister Patané; Luiz Alcantara; Luiz Lehmann Coutinho; Maria Carolina Elias; Maurício Lacerda Nogueira; Rafael dos Santos Bezerra; Raul Machado Neto; Rejane Maria Tommasini Grotto; Ricardo Haddad; Sandra Coccuzzo Sampaio Vessoni; Simone Kashima; Svetoslav Nanev Slavov; Vincent Louis Viala |
| EPI_ISL_3102542 | UNIDADE MISTA DE SAUDE DE MISSAO VELHA | Analytical Competence Molecular Epidemiology Lab/ACME, Oswaldo Cruz Foundation, Ceara (FIOCRUZ CE) | Cleber Furtado Aksenen; Fabio Miyajima; Fernando Braga Stehling; Francisco Eder de Moura Lopes; Jamille Maria Mendes Bezerra; Joaquim César do Nascimento Sousa Junior; Pedro Miguel Carneiro Jeronimo; Suzana Porto Almeida e Lucas Delerino; Thais Ferreira de Oliveira; Thais de Oliveira Costa; Ticiane Cavalcante de Souza; Veridiana Pessoa Miyajima |
| EPI_ISL_2445196 | UNIDADE MISTA DE SAUDE DE ORIENTE | Instituto Butantan | Antonio Jorge Martins; Claudia Renata dos Santos Barros; David Schlesinger; Debora Botequio Moretti; Dimas Tadeu Covas; Elaine Cristina Marqueze; Elaine Vieira Santos; Evandra Strazza Rodrigues; Heidge Fukumasu; Jayme Augusto de Souza-Neto; José Salvatore Leister Patané; Luiz Alcantara; Luiz Lehmann |

| | | | |
|---|---|---|---|
| | | | Coutinho; Maria Carolina Elias; Maurício Lacerda Nogueira; Rafael dos Santos Bezerra; Raul Machado Neto; Rejane Maria Tommasini Grotto; Ricardo Haddad; Sandra Coccuzzo Sampaio Vessoni; Simone Kashima; Svetoslav Nanev Slavov; Vincent Louis Viala |
| EPI_ISL_2493387, EPI_ISL_2493442 | UNIDADE MISTA DE SAUDE MOMBUCA | Instituto Butantan | Antonio Jorge Martins; Claudia Renata dos Santos Barros; David Schlesinger; Debora Botequio Moretti; Dimas Tadeu Covas; Elaine Cristina Marqueze; Elaine Vieira Santos; Evandra Strazza Rodrigues; Heidge Fukumasu; Jayme Augusto de Souza-Neto; José Salvatore Leister Patané; Luiz Alcantara; Luiz Lehmann Coutinho; Maria Carolina Elias; Maurício Lacerda Nogueira; Rafael dos Santos Bezerra; Raul Machado Neto; Rejane Maria Tommasini Grotto; Ricardo Haddad; Sandra Coccuzzo Sampaio Vessoni; Simone Kashima; Svetoslav Nanev Slavov; Vincent Louis Viala |
| EPI_ISL_2445585 | UNIDADE MISTA DE SAUDE ROSEIRA DR PAULO GUIMARAES CASTRO | Instituto Butantan | Antonio Jorge Martins; Claudia Renata dos Santos Barros; David Schlesinger; Debora Botequio Moretti; Dimas Tadeu Covas; Elaine Cristina Marqueze; Elaine Vieira Santos; Evandra Strazza Rodrigues; Heidge Fukumasu; Jayme Augusto de Souza-Neto; José Salvatore Leister Patané; Luiz Alcantara; Luiz Lehmann Coutinho; Maria Carolina Elias; Maurício Lacerda Nogueira; Rafael dos Santos Bezerra; Raul Machado Neto; Rejane Maria Tommasini Grotto; Ricardo Haddad; Sandra Coccuzzo Sampaio Vessoni; Simone Kashima; Svetoslav Nanev Slavov; Vincent Louis Viala |
| EPI_ISL_2493372 | UNIDADE MISTA OSVALDO P CAMARGO ELIAS FAUSTO | Instituto Butantan | Antonio Jorge Martins; Claudia Renata dos Santos Barros; David Schlesinger; Debora Botequio Moretti; Dimas Tadeu Covas; Elaine Cristina Marqueze; Elaine Vieira Santos; Evandra Strazza Rodrigues; Heidge Fukumasu; Jayme Augusto de Souza-Neto; José Salvatore Leister Patané; Luiz Alcantara; Luiz Lehmann Coutinho; Maria Carolina Elias; Maurício Lacerda Nogueira; Rafael dos Santos Bezerra; Raul Machado Neto; Rejane Maria Tommasini Grotto; Ricardo Haddad; Sandra Coccuzzo Sampaio Vessoni; Simone Kashima; Svetoslav Nanev Slavov; Vincent Louis Viala |
| EPI_ISL_2801360 | UNIDADE PRONTO ATENDIMENTO CONJUNTO CEARA | Analytical Competence Molecular Epidemiology Lab/ACME, Oswaldo Cruz Foundation, Ceara (FIOCRUZ CE) | Cleber Furtado Aksenen e Suzana Porto Almeida; Fabio Miyajima; Fernando Braga Stehling; Francisco Eder de Moura Lopes; Jamille Maria Mendes Bezerra; Joaquim César do Nascimento Sousa Junior; Pedro Miguel Carneiro Jeronimo; Thais Ferreira de Oliveira; Thais de Oliveira Costa; Ticiane Cavalcante de Souza; Veridiana Pessoa Miyajima |
| EPI_ISL_2801357 | UNIDADE PRONTO ATENDIMENTO ITAPERI | Analytical Competence Molecular Epidemiology Lab/ACME, Oswaldo Cruz Foundation, Ceara (FIOCRUZ CE) | Cleber Furtado Aksenen e Suzana Porto Almeida; Fabio Miyajima; Fernando Braga Stehling; Francisco Eder de Moura Lopes; Jamille Maria Mendes Bezerra; Joaquim César do Nascimento Sousa Junior; Pedro Miguel Carneiro Jeronimo; Thais Ferreira de Oliveira; Thais de Oliveira Costa; Ticiane Cavalcante de Souza; Veridiana Pessoa Miyajima |
| EPI_ISL_2344916 | UNIDADE PSF PINHEIROS APIAI | Instituto Butantan / ESALQ-Piracicaba | Antonio Jorge Martins; Claudia Renata dos Santos Barros; David Schlesinger; Debora Botequio Moretti; Dimas Tadeu Covas; Elaine Cristina Marqueze; Elaine Vieira Santos; Evandra Strazza Rodrigues; Heidge Fukumasu; Jayme Augusto de Souza-Neto; José Salvatore Leister Patané; Luiz Alcantara; Luiz Lehmann Coutinho; Maria Carolina Elias; Maurício Lacerda Nogueira; Rafael dos Santos Bezerra; Raul Machado Neto; Rejane Maria Tommasini Grotto; Ricardo Haddad; Sandra Coccuzzo Sampaio Vessoni; Simone Kashima; Svetoslav Nanev Slavov; Vincent Louis Viala |
| EPI_ISL_2445154, EPI_ISL_2445175, EPI_ISL_2445181, EPI_ISL_2445185, EPI_ISL_2473707, EPI_ISL_2473724, EPI_ISL_2473812 | | | |
| see above | UNIDADE REFERENCIAL SUDOESTE | Instituto Butantan | Antonio Jorge Martins; Claudia Renata dos Santos Barros; David Schlesinger; Debora Botequio Moretti; Dimas Tadeu Covas; Elaine Cristina Marqueze; Elaine Vieira Santos; Evandra Strazza Rodrigues; Heidge Fukumasu; Jayme Augusto de Souza-Neto; José Salvatore Leister Patané; Luiz Alcantara; Luiz Lehmann Coutinho; Maria Carolina Elias; Maurício Lacerda Nogueira; Rafael dos Santos Bezerra; Raul Machado Neto; Rejane Maria Tommasini Grotto; Ricardo Haddad; Sandra Coccuzzo Sampaio Vessoni; Simone Kashima; Svetoslav Nanev Slavov; Vincent Louis Viala |
| EPI_ISL_2345079, EPI_ISL_2345109, EPI_ISL_2345136, EPI_ISL_2345139, EPI_ISL_2345160 | UNIDADE REFERENCIAL SUDOESTE | Instituto Butantan / ESALQ-Piracicaba | Antonio Jorge Martins; Claudia Renata dos Santos Barros; David Schlesinger; Debora Botequio Moretti; Dimas Tadeu Covas; Elaine Cristina Marqueze; Elaine Vieira Santos; Evandra Strazza Rodrigues; Heidge Fukumasu; Jayme Augusto de Souza-Neto; José Salvatore Leister Patané; Luiz Alcantara; Luiz Lehmann Coutinho; Maria Carolina Elias; Maurício Lacerda Nogueira; Rafael dos Santos Bezerra; Raul Machado Neto; Rejane Maria Tommasini Grotto; Ricardo Haddad; Sandra Coccuzzo Sampaio Vessoni; Simone Kashima; Svetoslav Nanev Slavov; Vincent Louis Viala |
| EPI_ISL_2493249, EPI_ISL_2493250, EPI_ISL_2493461, EPI_ISL_2493474, EPI_ISL_2493477, EPI_ISL_2493483, EPI_ISL_2493507, EPI_ISL_2493511, EPI_ISL_2493515, EPI_ISL_2493517, EPI_ISL_2493518, EPI_ISL_2494292 | | | |
| see above | UNIDADE RESPIRATORIA NOVA HORTOLANDIA | Instituto Butantan | Antonio Jorge Martins; Claudia Renata dos Santos Barros; David Schlesinger; Debora Botequio Moretti; Dimas Tadeu Covas; Elaine Cristina Marqueze; Elaine Vieira Santos; Evandra Strazza Rodrigues; Heidge Fukumasu; Jayme Augusto de Souza-Neto; José Salvatore Leister Patané; Luiz Alcantara; Luiz Lehmann Coutinho; Maria Carolina Elias; Maurício Lacerda Nogueira; Rafael dos Santos Bezerra; Raul Machado Neto; Rejane Maria Tommasini Grotto; Ricardo Haddad; Sandra Coccuzzo Sampaio Vessoni; Simone Kashima; Svetoslav Nanev Slavov; Vincent Louis Viala |
| EPI_ISL_2208922, EPI_ISL_2345042, EPI_ISL_2345047, EPI_ISL_2345049, EPI_ISL_2345052, EPI_ISL_2345053 | UNIDADE RESPIRATORIA NOVA HORTOLANDIA | Instituto Butantan / ESALQ-Piracicaba | Antonio Jorge Martins; Claudia Renata dos Santos Barros; David Schlesinger; Debora Botequio Moretti; Dimas Tadeu Covas; Elaine Cristina Marqueze; Elaine Vieira Santos; Evandra Strazza Rodrigues; Heidge Fukumasu; Jayme Augusto de Souza-Neto; José Salvatore Leister Patané; Luiz Alcantara; Luiz Lehmann Coutinho; Maria Carolina Elias; Maurício Lacerda Nogueira; Rafael dos Santos Bezerra; Raul Machado Neto; Rejane Maria Tommasini Grotto; Ricardo Haddad; Sandra Coccuzzo Sampaio Vessoni; Simone Kashima; Svetoslav Nanev Slavov; Vincent Louis Viala |
| EPI_ISL_2208916 | UNIDADE SENTINELA COVID19 | Instituto Butantan / Mendelics | Antonio Jorge Martins; Claudia Renata dos Santos Barros; David Schlesinger; Debora Botequio Moretti; Dimas Tadeu Covas; Elaine Cristina Marqueze; Elaine Vieira Santos; Evandra Strazza Rodrigues; Heidge Fukumasu; Jayme Augusto de Souza-Neto; José Salvatore Leister Patané; Luiz Alcantara; Luiz Lehmann Coutinho; Maria Carolina Elias; Maurício Lacerda Nogueira; Rafael dos Santos Bezerra; Raul Machado Neto; Rejane Maria Tommasini Grotto; Ricardo Haddad; Sandra Coccuzzo Sampaio Vessoni; Simone Kashima; Svetoslav Nanev Slavov; Vincent Louis Viala |
| EPI_ISL_2445329, EPI_ISL_2493272, EPI_ISL_2493273, EPI_ISL_2493566 | UPA DE BEBEDOURO | Instituto Butantan | Antonio Jorge Martins; Claudia Renata dos Santos Barros; David Schlesinger; Debora Botequio Moretti; Dimas Tadeu Covas; Elaine Cristina Marqueze; Elaine Vieira Santos; Evandra Strazza Rodrigues; Heidge Fukumasu; Jayme Augusto de Souza-Neto; José Salvatore Leister Patané; Luiz Alcantara; Luiz Lehmann Coutinho; Maria Carolina Elias; Maurício Lacerda Nogueira; Rafael dos Santos Bezerra; Raul Machado Neto; Rejane Maria Tommasini Grotto; Ricardo Haddad; Sandra Coccuzzo Sampaio Vessoni; Simone Kashima; Svetoslav Nanev Slavov; Vincent Louis Viala |
| EPI_ISL_2208944 | UPA DE BEBEDOURO | Instituto Butantan / Mendelics | Antonio Jorge Martins; Claudia Renata dos Santos Barros; David Schlesinger; Debora Botequio Moretti; Dimas Tadeu Covas; Elaine Cristina Marqueze; Elaine Vieira Santos; Evandra Strazza Rodrigues; Heidge Fukumasu; Jayme Augusto de Souza-Neto; José Salvatore Leister Patané; Luiz Alcantara; Luiz Lehmann Coutinho; Maria Carolina Elias; Maurício Lacerda Nogueira; Rafael dos Santos Bezerra; Raul Machado Neto; Rejane Maria Tommasini Grotto; Ricardo Haddad; Sandra Coccuzzo Sampaio Vessoni; Simone Kashima; Svetoslav Nanev Slavov; Vincent Louis Viala |
| EPI_ISL_2445245, EPI_ISL_2445249, EPI_ISL_2445250, EPI_ISL_2445256, EPI_ISL_2445260, EPI_ISL_2445261, EPI_ISL_2445264 | | | |
| see above | UPA DR AKIRA TADA | Instituto Butantan | Antonio Jorge Martins; Claudia Renata dos Santos Barros; David Schlesinger; Debora Botequio Moretti; Dimas Tadeu Covas; Elaine Cristina Marqueze; Elaine Vieira Santos; Evandra Strazza Rodrigues; Heidge Fukumasu; Jayme Augusto de Souza-Neto; José Salvatore Leister Patané; Luiz Alcantara; Luiz Lehmann Coutinho; Maria Carolina Elias; Maurício Lacerda Nogueira; Rafael dos Santos Bezerra; Raul Machado Neto; Rejane Maria Tommasini Grotto; Ricardo Haddad; Sandra Coccuzzo Sampaio Vessoni; Simone Kashima; Svetoslav Nanev Slavov; Vincent Louis Viala |
| EPI_ISL_2345056, EPI_ISL_2345057, EPI_ISL_2345058 | UPA DR AKIRA TADA | Instituto Butantan / ESALQ-Piracicaba | Antonio Jorge Martins; Claudia Renata dos Santos Barros; David Schlesinger; Debora Botequio Moretti; Dimas Tadeu Covas; Elaine Cristina Marqueze; Elaine Vieira Santos; Evandra Strazza Rodrigues; Heidge Fukumasu; Jayme Augusto de Souza-Neto; José Salvatore Leister Patané; Luiz Alcantara; Luiz Lehmann Coutinho; Maria Carolina Elias; Maurício Lacerda Nogueira; Rafael dos Santos Bezerra; Raul Machado Neto; Rejane Maria Tommasini Grotto; Ricardo Haddad; Sandra Coccuzzo Sampaio Vessoni; Simone Kashima; Svetoslav Nanev Slavov; Vincent Louis Viala |
| EPI_ISL_2171026 | UPA DR AKIRA TADA | Instituto Butantan / Mendelics | Antonio Jorge Martins; Bianca Cechetto Carlos. Mendelics: Bibiana Santos; Claudia Renata dos Santos Barros; Cíntia Bittar; David Schlesinger. Hemocentro Ribeirão Preto: Simone Kashima; Debora Botequio Moretti; Elaine Cristina Marqueze; Elaine Vieira dos Santos; Elisangela Chicaroni Mattos; Erika Freitas; Evandra Strazza Rodrigues; Felipe Allan da Silva da Costa; Flavia Aburjaile; Fábio Sossai Possebon; Guilherme Campos; Guilherme Targino Valente; Heidge Fukumasu. USP-Botucatu: Rejane Maria Tommasini Grotto; Helena Lage Ferreira; Instituto Butantan: Dimas Tadeu Covas; Jardelina de Souza Todao Bernardino; Jayme A. Souza-Neto; Jessika Cristina Chagas Lesbon; Jorge A. Petroli Marchesi; José Salvatore Leister Patané; João Paulo Kitajima; João Pessoa Araújo Jr.; Leila Sabrina Ullmann; Loyze Paola Oliveira de Lima; Luiz Aurelio de Campos Crispin. Centro de Genômica Funcional da ESALQ: Luiz Lehmann Coutinho; Luiz Carlos Junior de Alcantara; Lívia Sacchetto; Maisa C. Pereira Parra; Maria Carolina Elias; Marta Giovanetti; Marília Moraes; Maurício Lacerda Nogueira. Prefeitura de Sao Paulo: Melissa Palmieri.; Patricia Akemi Assato; Paula Rahal; Paulo Inacio da Costa; Rafael dos Santos Bezerra; Raquel de Lello Rocha Campos Cassano. NGS Soluções Genômicas: Pilar Drummond Sampaio Corrêa Mariani. FZEA-USP Pirassununga: Mirele Daiana Poleti; Raul Machado Neto; Ricardo Augusto Brassaloti; Ricardo Haddad; Rodrigo Tocantins Calado. FAMERP-SJRP: Cecília Artico Banho; Sandra Coccuzzo Sampaio; Svetoslav Nanev Slavov; Vagner Fonseca; Vincent Louis Viala |
| EPI_ISL_2345014 | UPA DR FRANCO DA ROCHA | Instituto Butantan | Antonio Jorge Martins; Claudia Renata dos Santos Barros; David Schlesinger; Debora Botequio Moretti; Dimas Tadeu Covas; Elaine Cristina Marqueze; Elaine Vieira Santos; Evandra Strazza Rodrigues; Heidge Fukumasu; Jayme Augusto de Souza-Neto; José Salvatore Leister Patané; Luiz Alcantara; Luiz Lehmann Coutinho; Maria Carolina Elias; Maurício Lacerda Nogueira; Rafael dos Santos Bezerra; Raul Machado Neto; Rejane Maria Tommasini Grotto; Ricardo Haddad; Sandra Coccuzzo Sampaio Vessoni; Simone Kashima; Svetoslav Nanev Slavov; Vincent Louis Viala |
| EPI_ISL_2493510, EPI_ISL_2493514, EPI_ISL_2494303 | UPA II JUNDIAI | Instituto Butantan | Antonio Jorge Martins; Claudia Renata dos Santos Barros; David Schlesinger; Debora Botequio Moretti; Dimas Tadeu Covas; Elaine Cristina Marqueze; Elaine Vieira Santos; Evandra Strazza Rodrigues; Heidge Fukumasu; Jayme Augusto de Souza-Neto; José Salvatore Leister Patané; Luiz Alcantara; Luiz Lehmann Coutinho; Maria Carolina Elias; Maurício Lacerda Nogueira; Rafael dos Santos Bezerra; Raul Machado Neto; Rejane Maria Tommasini Grotto; Ricardo Haddad; Sandra Coccuzzo Sampaio Vessoni; Simone Kashima; Svetoslav Nanev Slavov; Vincent Louis Viala |
| EPI_ISL_2209746 | UPA MARY DOTA | Instituto Butantan / Mendelics | Antonio Jorge Martins; Claudia Renata dos Santos Barros; David Schlesinger; Debora Botequio Moretti; Dimas Tadeu Covas; Elaine Cristina Marqueze; Elaine Vieira Santos; Evandra Strazza Rodrigues; Heidge Fukumasu; Jayme Augusto de Souza-Neto; José Salvatore Leister Patané; Luiz Alcantara; Luiz Lehmann Coutinho; Maria Carolina Elias; Maurício Lacerda Nogueira; Rafael dos Santos Bezerra; Raul Machado Neto; Rejane Maria Tommasini Grotto; Ricardo Haddad; Sandra Coccuzzo Sampaio Vessoni; Simone Kashima; Svetoslav Nanev Slavov; Vincent Louis Viala |
| EPI_ISL_2210275 | UPA UNIDADE DE PRONTO ATENDIMENTO 24 HORAS BOM JESUS | Instituto Butantan / Mendelics | Antonio Jorge Martins; Claudia Renata dos Santos Barros; David Schlesinger; Debora Botequio Moretti; Dimas Tadeu Covas; Elaine Cristina Marqueze; Elaine Vieira Santos; Evandra Strazza Rodrigues; Heidge Fukumasu; Jayme Augusto de Souza-Neto; José Salvatore Leister Patané; Luiz Alcantara; Luiz Lehmann Coutinho; Maria Carolina Elias; Maurício Lacerda Nogueira; Rafael dos Santos Bezerra; Raul Machado Neto; Rejane Maria Tommasini Grotto; Ricardo Haddad; Sandra Coccuzzo Sampaio Vessoni; Simone Kashima; Svetoslav Nanev Slavov; Vincent Louis Viala |
| EPI_ISL_2344999, EPI_ISL_2345000, EPI_ISL_2345001, EPI_ISL_2445275, EPI_ISL_2445300, EPI_ISL_2445302 | UPA VILA SANTA CATARINA | Instituto Butantan | Antonio Jorge Martins; Claudia Renata dos Santos Barros; David Schlesinger; Debora Botequio Moretti; Dimas Tadeu Covas; Elaine Cristina Marqueze; Elaine Vieira Santos; Evandra Strazza Rodrigues; Heidge Fukumasu; Jayme Augusto de Souza-Neto; José Salvatore Leister Patané; Luiz Alcantara; Luiz Lehmann Coutinho; Maria Carolina Elias; Maurício Lacerda Nogueira; Rafael dos Santos Bezerra; Raul Machado Neto; Rejane Maria Tommasini Grotto; Ricardo Haddad; Sandra Coccuzzo Sampaio Vessoni; Simone Kashima; Svetoslav Nanev Slavov; Vincent Louis Viala |
| EPI_ISL_1967298 | UPA VILA SANTA CATARINA | Instituto Butantan / Mendelics | Antonio Jorge Martins; Bianca Cechetto Carlos. Mendelics: Bibiana Santos; Claudia Renata dos Santos Barros; Cíntia Bittar; David Schlesinger. Hemocentro Ribeirão Preto: Simone Kashima; Debora Botequio Moretti; Elaine Cristina Marqueze; Elaine Vieira dos Santos; Elisangela Chicaroni Mattos; Erika Freitas; Evandra Strazza Rodrigues; Felipe Allan da Silva da Costa; Flavia Aburjaile; Fábio Sossai Possebon; Guilherme Campos; Guilherme Targino Valente; Heidge Fukumasu. USP-Botucatu: Rejane Maria Tommasini Grotto; Helena Lage Ferreira; Instituto Butantan: Dimas Tadeu Covas; Jardelina de Souza Todao Bernardino; Jayme A. Souza-Neto; Jessika Cristina Chagas Lesbon; Jorge A. Petroli Marchesi; José Salvatore Leister Patané; João Paulo Kitajima; João Pessoa Araújo Jr.; Leila Sabrina Ullmann; Loyze Paola Oliveira de Lima; Luiz Aurelio de Campos Crispin. Centro de Genômica Funcional da ESALQ: Luiz Lehmann Coutinho; Luiz Carlos Junior de Alcantara; Lívia Sacchetto; Maisa C. Pereira Parra; Maria Carolina Elias; Marta Giovanetti; Marília Moraes; Maurício Lacerda Nogueira. Prefeitura de Sao Paulo: Melissa Palmieri.; Patricia Akemi Assato; Paula Rahal; Paulo Inacio da Costa; Rafael dos Santos Bezerra; Raquel de Lello Rocha Campos Cassano. NGS Soluções Genômicas: Pilar Drummond Sampaio Corrêa Mariani. FZEA-USP Pirassununga: Mirele Daiana Poleti; Raul Machado Neto; Ricardo Augusto Brassaloti; Ricardo Haddad; Rodrigo Tocantins Calado. FAMERP-SJRP: Cecília Artico Banho; Sandra Coccuzzo Sampaio; Svetoslav Nanev Slavov; Vagner Fonseca; Vincent Louis Viala |
| EPI_ISL_2426773, EPI_ISL_2426774, EPI_ISL_2426783, EPI_ISL_2450425, EPI_ISL_2661479, EPI_ISL_2661486 | US Air Force School of Aerospace Medicine | US Air Force School of Aerospace Medicine | Amanda Javorina; Anthony Fries; Carol Garrett; Clarise Starr; Cole Anderson; Elizabeth Macias; Fritz Castillo; Jennifer Meyer; Sarah Purves; William Gruner |
| EPI_ISL_2493546 | USAFA FORTE | Instituto Butantan | Antonio Jorge Martins; Claudia Renata dos Santos Barros; David Schlesinger; Debora Botequio Moretti; Dimas Tadeu Covas; Elaine Cristina Marqueze; Elaine Vieira Santos; Evandra Strazza Rodrigues; Heidge Fukumasu; Jayme Augusto de Souza-Neto; José Salvatore Leister Patané; Luiz Alcantara; Luiz Lehmann Coutinho; Maria Carolina Elias; Maurício Lacerda Nogueira; Rafael dos Santos Bezerra; Raul Machado Neto; Rejane Maria Tommasini Grotto; Ricardo Haddad; Sandra Coccuzzo Sampaio Vessoni; Simone Kashima; Svetoslav Nanev Slavov; Vincent Louis Viala |
| EPI_ISL_2493536, EPI_ISL_2494304 | USAFA GUILHERMINA | Instituto Butantan | Antonio Jorge Martins; Claudia Renata dos Santos Barros; David Schlesinger; Debora Botequio Moretti; Dimas Tadeu Covas; Elaine Cristina Marqueze; Elaine Vieira Santos; Evandra Strazza Rodrigues; Heidge Fukumasu; Jayme Augusto de Souza-Neto; José Salvatore Leister Patané; Luiz Alcantara; Luiz Lehmann Coutinho; Maria Carolina Elias; Maurício Lacerda Nogueira; Rafael dos Santos Bezerra; Raul Machado Neto; Rejane Maria Tommasini Grotto; Ricardo Haddad; Sandra Coccuzzo Sampaio Vessoni; Simone Kashima; Svetoslav Nanev Slavov; Vincent Louis Viala |
| EPI_ISL_2493555 | USAFA MELVI | Instituto Butantan | Antonio Jorge Martins; Claudia Renata dos Santos Barros; David Schlesinger; Debora Botequio Moretti; Dimas Tadeu Covas; Elaine Cristina Marqueze; Elaine Vieira Santos; Evandra Strazza Rodrigues; Heidge Fukumasu; Jayme Augusto de Souza-Neto; José Salvatore Leister Patané; Luiz Alcantara; Luiz Lehmann Coutinho; Maria Carolina Elias; Maurício Lacerda Nogueira; Rafael dos Santos Bezerra; Raul Machado Neto; Rejane Maria Tommasini Grotto; Ricardo Haddad; Sandra Coccuzzo Sampaio Vessoni; Simone Kashima; Svetoslav Nanev Slavov; Vincent Louis Viala |
| EPI_ISL_2493557 | USAFA OCIAN | Instituto Butantan | Antonio Jorge Martins; Claudia Renata dos Santos Barros; David Schlesinger; Debora Botequio Moretti; Dimas Tadeu Covas; Elaine Cristina Marqueze; Elaine Vieira Santos; Evandra Strazza Rodrigues; Heidge Fukumasu; Jayme Augusto de Souza-Neto; José Salvatore Leister Patané; Luiz Alcantara; Luiz Lehmann Coutinho; Maria Carolina Elias; Maurício Lacerda Nogueira; Rafael dos Santos Bezerra; Raul Machado Neto; Rejane Maria Tommasini Grotto; Ricardo Haddad; Sandra Coccuzzo Sampaio Vessoni; Simone Kashima; Svetoslav Nanev Slavov; Vincent Louis Viala |
| EPI_ISL_2493559, EPI_ISL_2493561 | USAFA PRINCESA | Instituto Butantan | Antonio Jorge Martins; Claudia Renata dos Santos Barros; David Schlesinger; Debora Botequio Moretti; Dimas Tadeu Covas; Elaine Cristina Marqueze; Elaine Vieira Santos; Evandra Strazza Rodrigues; Heidge Fukumasu; Jayme Augusto de Souza-Neto; José Salvatore Leister Patané; Luiz Alcantara; Luiz Lehmann Coutinho; Maria Carolina Elias; Maurício Lacerda Nogueira; Rafael dos Santos Bezerra; Raul Machado Neto; Rejane Maria Tommasini Grotto; Ricardo Haddad; Sandra Coccuzzo Sampaio Vessoni; Simone Kashima; Svetoslav Nanev Slavov; Vincent Louis Viala |
| EPI_ISL_2493560, EPI_ISL_2493563 | USAFA SAO JORGE | Instituto Butantan | Antonio Jorge Martins; Claudia Renata dos Santos Barros; David Schlesinger; Debora Botequio Moretti; Dimas Tadeu Covas; Elaine Cristina Marqueze; Elaine Vieira Santos; Evandra Strazza Rodrigues; Heidge Fukumasu; Jayme Augusto de Souza-Neto; José Salvatore Leister Patané; Luiz Alcantara; Luiz Lehmann Coutinho; Maria Carolina Elias; Maurício Lacerda Nogueira; Rafael dos Santos Bezerra; Raul Machado Neto; Rejane Maria Tommasini Grotto; Ricardo Haddad; Sandra Coccuzzo Sampaio Vessoni; Simone Kashima; Svetoslav Nanev Slavov; Vincent Louis Viala |
| EPI_ISL_2493551 | USAFA TUDE BASTOS | Instituto Butantan | Antonio Jorge Martins; Claudia Renata dos Santos Barros; David Schlesinger; Debora Botequio Moretti; Dimas Tadeu Covas; Elaine Cristina Marqueze; Elaine Vieira Santos; Evandra Strazza Rodrigues; Heidge Fukumasu; Jayme Augusto de Souza-Neto; José Salvatore Leister Patané; Luiz Alcantara; Luiz Lehmann Coutinho; Maria Carolina Elias; Maurício Lacerda Nogueira; Rafael dos Santos Bezerra; Raul Machado Neto; Rejane Maria Tommasini Grotto; Ricardo Haddad; Sandra Coccuzzo Sampaio Vessoni; Simone Kashima; Svetoslav Nanev Slavov; Vincent Louis Viala |

| | | | |
|---|---|---|---|
| EPI_ISL_2391784 | USC Clinical Lab | Los Angeles County Public Health Laboratories | P. Hemarajata et al. |
| EPI_ISL_2345746 | USF ARCADAS | Instituto Butantan / Mendelics | Antonio Jorge Martins; Claudia Renata dos Santos Barros; David Schlesinger; Debora Botequio Moretti; Dimas Tadeu Covas; Elaine Cristina Marqueze; Elaine Vieira Santos; Evandra Strazza Rodrigues; Heidge Fukumasu; Jayme Augusto de Souza-Neto; José Salvatore Leister Patané; Luiz Alcantara; Luiz Lehmann Coutinho; Maria Carolina Elias; Maurício Lacerda Nogueira; Rafael dos Santos Bezerra; Raul Machado Neto; Rejane Maria Tommasini Grotto; Ricardo Haddad; Sandra Coccuzzo Sampaio Vessoni; Simone Kashima; Svetoslav Nanev Slavov; Vincent Louis Viala |
| EPI_ISL_2445456 | USF II DR JOSE RODOLFO ZIMMERMANN DIAS | Instituto Butantan | Antonio Jorge Martins; Claudia Renata dos Santos Barros; David Schlesinger; Debora Botequio Moretti; Dimas Tadeu Covas; Elaine Cristina Marqueze; Elaine Vieira Santos; Evandra Strazza Rodrigues; Heidge Fukumasu; Jayme Augusto de Souza-Neto; José Salvatore Leister Patané; Luiz Alcantara; Luiz Lehmann Coutinho; Maria Carolina Elias; Maurício Lacerda Nogueira; Rafael dos Santos Bezerra; Raul Machado Neto; Rejane Maria Tommasini Grotto; Ricardo Haddad; Sandra Coccuzzo Sampaio Vessoni; Simone Kashima; Svetoslav Nanev Slavov; Vincent Louis Viala |
| EPI_ISL_2473711 | USF ITAJUBY | Instituto Butantan | Antonio Jorge Martins; Claudia Renata dos Santos Barros; David Schlesinger; Debora Botequio Moretti; Dimas Tadeu Covas; Elaine Cristina Marqueze; Elaine Vieira Santos; Evandra Strazza Rodrigues; Heidge Fukumasu; Jayme Augusto de Souza-Neto; José Salvatore Leister Patané; Luiz Alcantara; Luiz Lehmann Coutinho; Maria Carolina Elias; Maurício Lacerda Nogueira; Rafael dos Santos Bezerra; Raul Machado Neto; Rejane Maria Tommasini Grotto; Ricardo Haddad; Sandra Coccuzzo Sampaio Vessoni; Simone Kashima; Svetoslav Nanev Slavov; Vincent Louis Viala |
| EPI_ISL_2445203, EPI_ISL_2445204, EPI_ISL_2445205, EPI_ISL_2445526, EPI_ISL_2473716, EPI_ISL_2473717, EPI_ISL_2473817 | | | |
| see above | USF PAULISTA FERNANDOPOLIS ANTONIO PIVATO | Instituto Butantan | Antonio Jorge Martins; Claudia Renata dos Santos Barros; David Schlesinger; Debora Botequio Moretti; Dimas Tadeu Covas; Elaine Cristina Marqueze; Elaine Vieira Santos; Evandra Strazza Rodrigues; Heidge Fukumasu; Jayme Augusto de Souza-Neto; José Salvatore Leister Patané; Luiz Alcantara; Luiz Lehmann Coutinho; Maria Carolina Elias; Maurício Lacerda Nogueira; Rafael dos Santos Bezerra; Raul Machado Neto; Rejane Maria Tommasini Grotto; Ricardo Haddad; Sandra Coccuzzo Sampaio Vessoni; Simone Kashima; Svetoslav Nanev Slavov; Vincent Louis Viala |
| EPI_ISL_2445392, EPI_ISL_2493463, EPI_ISL_2493469, EPI_ISL_2493475, EPI_ISL_2493479 | USF SALERNO | Instituto Butantan | Antonio Jorge Martins; Claudia Renata dos Santos Barros; David Schlesinger; Debora Botequio Moretti; Dimas Tadeu Covas; Elaine Cristina Marqueze; Elaine Vieira Santos; Evandra Strazza Rodrigues; Heidge Fukumasu; Jayme Augusto de Souza-Neto; José Salvatore Leister Patané; Luiz Alcantara; Luiz Lehmann Coutinho; Maria Carolina Elias; Maurício Lacerda Nogueira; Rafael dos Santos Bezerra; Raul Machado Neto; Rejane Maria Tommasini Grotto; Ricardo Haddad; Sandra Coccuzzo Sampaio Vessoni; Simone Kashima; Svetoslav Nanev Slavov; Vincent Louis Viala |
| EPI_ISL_2383988 | UT-Unified State Labs: Public Health Utah DOH | Centers for Disease Control and Prevention Division of Viral Diseases, Pathogen Discovery | Alison Laufer Halpin; Ben L. Rambo-Martin; Clinton R. Paden; Dakota Howard; Darlene Wagner; Dave Wentworth; Dhwani Batra; Jasmine Padilla; Justin Lee; Katie Dillon; Krista Queen; Kristen Knipe; Kristine Lacek; Mark Burroughs; Matthew Schmerer; Mili Sheth; Peter Cook; Sam Shepard; Sarah Nobles; Shoshona Le; Suxiang Tong; Vivien Dugan; Yvette Unoarumhi |
| EPI_ISL_2002420, EPI_ISL_2002816, EPI_ISL_2002881, EPI_ISL_2031481, EPI_ISL_2031482, EPI_ISL_2031483, EPI_ISL_2031487, EPI_ISL_2031494, EPI_ISL_2037449, EPI_ISL_2037584, EPI_ISL_2037587, EPI_ISL_2037652, EPI_ISL_2037654, EPI_ISL_2037656, EPI_ISL_2037664, EPI_ISL_2037666, EPI_ISL_2037667, EPI_ISL_2037682, EPI_ISL_2037684, EPI_ISL_2082514, EPI_ISL_2082569, EPI_ISL_2082570, EPI_ISL_2082572, EPI_ISL_2082573, EPI_ISL_2082641, EPI_ISL_2082671, EPI_ISL_2082675, EPI_ISL_2082677, EPI_ISL_2082681, EPI_ISL_2082699, EPI_ISL_2086397, EPI_ISL_2086422, EPI_ISL_2104837, EPI_ISL_2104840, EPI_ISL_2104842, EPI_ISL_2104885, EPI_ISL_2104889, EPI_ISL_2104956, EPI_ISL_2104977, EPI_ISL_2104979, EPI_ISL_2105034, EPI_ISL_2105055, EPI_ISL_2161842, EPI_ISL_2194050, EPI_ISL_2194051, EPI_ISL_2194174, EPI_ISL_2233293, EPI_ISL_2272617, EPI_ISL_2272656, EPI_ISL_2273779, EPI_ISL_2273807, EPI_ISL_2299454, EPI_ISL_2332880, EPI_ISL_2332923, EPI_ISL_2332940, EPI_ISL_2332988, EPI_ISL_2332989, EPI_ISL_2333008, EPI_ISL_2333034, EPI_ISL_2333061, EPI_ISL_2340712, EPI_ISL_2340743, EPI_ISL_2340768, EPI_ISL_2340804, EPI_ISL_2426281, EPI_ISL_2426347, EPI_ISL_2426424, EPI_ISL_2426430, EPI_ISL_2426433, EPI_ISL_2426436, EPI_ISL_2426507, EPI_ISL_2426515, EPI_ISL_2426518, EPI_ISL_2432639, EPI_ISL_2432675, EPI_ISL_2432766, EPI_ISL_2432776, EPI_ISL_2432783, EPI_ISL_2432785, EPI_ISL_2432844, EPI_ISL_2432849, EPI_ISL_2432859, EPI_ISL_2432877, EPI_ISL_2432902, EPI_ISL_2432911, EPI_ISL_2502847, EPI_ISL_2502913, EPI_ISL_2502924, EPI_ISL_2502944, EPI_ISL_2503028, EPI_ISL_2503066, EPI_ISL_2503133, EPI_ISL_2544938, EPI_ISL_2544942, EPI_ISL_2544958, EPI_ISL_2544968, EPI_ISL_2618311, EPI_ISL_2618363, EPI_ISL_2618412, EPI_ISL_2618442, EPI_ISL_2618585, EPI_ISL_2618637, EPI_ISL_2618660, EPI_ISL_2618665, EPI_ISL_2618669, EPI_ISL_2618702, EPI_ISL_2618728 | | | |
| see above | UW Virology Lab | UW Virology Lab | Alexander Greninger; Hong Xie; Keith R Jerome; Lasata Shrestha; Meei-Li Huang; Michelle Lin; Nathan Breit; Noah R. Baker; Patrick Mathias; Pavitra Roychoudhury; Ricardo Perez; Robert J. Livingston; Sean Ellis; Shah Mohamed Bakhash; Tien V. Nguyen |
| EPI_ISL_2671499, EPI_ISL_2671670, EPI_ISL_2671672, EPI_ISL_2671678 | Unidad de Investigacion Medica de Yucatan (UIMY) | Unidad de Genomica Avanzada | ; Alejandra Garcia-Gasca; Alejandra Hernandez-Teran; Alejandro Sanchez-Flores; Alfredo Herrera-Estrella; Alicia Ocaña-Mondragon; Andreu Comas-Garcia; Angel Gustavo Salas-Lais; Antonio Loza Roman; Bernardo Martinez-Miguel; Blanca Taboada; Brenda Irasema Maldonado-Meza; Bruno Gomez-Gil; Carla Ivon Herrera-Najera; Carlos F. Arias; Celia Boukadida; Celida Duque Molina; Celida Martinez- Rodriguez; Clara Esperanza Santacruz-Tinoco; Concepcion Grajales-Muñiz; Consorcio Mexicano de Vigilancia Genomica (CoViGen-Mex). Authors (in alphabetical order): Julio Elias Alvarado-Yaah; Cristobal Chaidez-Quiroz; Daniel Fregoso-Rueda; Daniel Lira Morales; Eduardo Becerril-Vargas; Fernando Fontove-Herrera; Fidencio Mejia-Nepomuceno; Francisco Pulido; Gloria Elena Espinosa-Ayala; Gloria Maria Molina-Salinas; Gloria Vazquez; Hector Esteban Paz-Juarez; Hector Montoya-Fuentes; Helen Haydee Fernanda Ramirez-Plascencia; Irvin Gonzalez-Lopez; Jean Pierre Gonzalez; Jesus Hernandez; Joel Armando Vazquez-Perez.; Jorge Salas-Hernandez; Jose Antonio Enciso-Moreno; Jose Arturo Martinez-Orozco; Jose Esteban Muñoz-Medina; Jose de Jesus Nuñez-Contreras; Juan Bautista Chale-Dzul; Julissa Enciso-Ibarra; Luis Alberto Ochoa-Carrera; Margarita Matias-Florentino; Maria Guadalupe Santiago-Mauricio; Maria Guadalupe de Jesus Mireles-Rivera; Mario Mujica-Sanchez; Marissa Perez-Garcia; Nelly Selem-Mojica; Pavel Isa; Ricardo Ciria Merce; Ricardo Grande; Rosa Maria Gutierrez Rios; Santiago avila-Rios; Selene Zarate; Susana Lopez; Veronica Mata-Haro; Victor Eduardo Garcia-Arias; Victor Hugo Borja-Aburto |
| EPI_ISL_2490385, EPI_ISL_2490386 | Unidad de Investigación Biomédica de Zacatecas (UIBZ) | Centro de Investigación en Enfermedades Infecciosas (CIENI), Instituto Nacional de Enfermedades Respiratorias (INER) | ; Alejandra Garcia-Gasca; Alejandra Hernández-Terán; Alejandro Sanchez-Flores; Alfredo Herrera-Estrella; Alicia Ocaña-Mondragón; Andreu Comas-García; Angel Gustavo Salas-Lais; Antonio Loza Román; Bernardo Martínez-Miguel; Blanca Taboada; Brenda Irasema Maldonado-Meza; Bruno Gomez-Gil; Carla Ivón Herrera-Najera; Carlos F. Arias; Celia Boukadida; Clara Esperanza Santacruz-Tinoco; Concepción Grajales-Muñiz; Consorcio Mexicano de Vigilancia Genómica (CoViGen-Mex). Authors (in alphabetical order): Julio Elias Alvarado-Yaah; Cristóbal Cháidez-Quiróz; Célida Duque Molina; Célida Martínez-Rodríguez; Daniel Fregoso-Rueda; Daniel Lira Morales; Eduardo Becerril-Vargas; Fernando Fontove-Herrera; Fidencio Mejía-Nepomuceno; Francisco Pulido; Gloria Elena Espinosa-Ayala; Gloria María Molina-Salinas; Gloria Vazquez; Hector Esteban Paz-Juárez; Hector Montoya-Fuentes; Helen Haydee Fernanda Ramírez-Plascencia; Irvin González-López; Jean Pierre González; Joel Armando Vázquez-Pérez.; Jorge Salas-Hernández; José Antonio Enciso-Moreno; José Arturo Martínez-Orozco; José Esteban Muñoz-Medina; José de Jesús Nuñez-Contreras; Juan Bautista Chale-Dzul; Julissa Enciso-Ibarra; Luis Alberto Ochoa-Carrera; Margarita Matías-Florentino; Mario Mújica-Sánchez; Marissa Perez-Garcia; María Guadalupe Santiago-Mauricio; María Guadalupe de Jesús Mireles-Rivera; Nelly Sélem-Mojica; Pavel Isa; Ricardo Ciria Merce; Ricardo Grande; Rosa María Gutierrez Rios; Santiago Ávila-Ríos; Selene Zárate; Susana Lopez; Victor Eduardo Garcia-Arias; Víctor Hugo Borja-Aburto |
| EPI_ISL_2490376, EPI_ISL_2490381 | Unidad de Investigación Médica de Yucatán (UIMY) | Centro de Investigación en Enfermedades Infecciosas (CIENI), Instituto Nacional de Enfermedades Respiratorias (INER) | ; Alejandra Garcia-Gasca; Alejandra Hernández-Terán; Alejandro Sanchez-Flores; Alfredo Herrera-Estrella; Alicia Ocaña-Mondragón; Andreu Comas-García; Angel Gustavo Salas-Lais; Antonio Loza Román; Bernardo Martínez-Miguel; Blanca Taboada; Brenda Irasema Maldonado-Meza; Bruno Gomez-Gil; Carla Ivón Herrera-Najera; Carlos F. Arias; Celia Boukadida; Clara Esperanza Santacruz-Tinoco; Concepción Grajales-Muñiz; Consorcio Mexicano de Vigilancia Genómica (CoViGen-Mex). Authors (in alphabetical order): Julio Elias Alvarado-Yaah; Cristóbal Cháidez-Quiróz; Célida Duque Molina; Célida Martínez-Rodríguez; Daniel Fregoso-Rueda; Daniel Lira Morales; Eduardo Becerril-Vargas; Fernando Fontove-Herrera; Fidencio Mejía-Nepomuceno; Francisco Pulido; Gloria Elena Espinosa-Ayala; Gloria María Molina-Salinas; Gloria Vazquez; Hector Esteban Paz-Juárez; Hector Montoya-Fuentes; Helen Haydee Fernanda Ramírez-Plascencia; Irvin González-López; Jean Pierre González; Joel Armando Vázquez-Pérez.; Jorge Salas-Hernández; José Antonio Enciso-Moreno; José Arturo Martínez-Orozco; José Esteban Muñoz-Medina; José de Jesús Nuñez-Contreras; Juan Bautista Chale-Dzul; Julissa Enciso-Ibarra; Luis Alberto Ochoa-Carrera; Margarita Matías-Florentino; Mario Mújica-Sánchez; Marissa Perez-Garcia; María Guadalupe Santiago-Mauricio; María Guadalupe de Jesús Mireles-Rivera; Nelly Sélem-Mojica; Pavel Isa; Ricardo Ciria Merce; Ricardo Grande; Rosa María Gutierrez Rios; Santiago Ávila-Ríos; Selene Zárate; Susana Lopez; Victor Eduardo Garcia-Arias; Víctor Hugo Borja-Aburto |
| EPI_ISL_2535111, EPI_ISL_2535112, EPI_ISL_2535113, EPI_ISL_2535114, EPI_ISL_2535115, EPI_ISL_2535116, EPI_ISL_2535117, EPI_ISL_2535118, EPI_ISL_2535119, EPI_ISL_2535120, EPI_ISL_2535121, EPI_ISL_2535124, EPI_ISL_2535125, EPI_ISL_2535126, EPI_ISL_2535127, EPI_ISL_2535128, EPI_ISL_2535131, EPI_ISL_2535132, EPI_ISL_2535133, EPI_ISL_2535134, EPI_ISL_2535135, EPI_ISL_2535136, EPI_ISL_2535137, EPI_ISL_2535138, EPI_ISL_2535140, EPI_ISL_2535141, EPI_ISL_2535142, EPI_ISL_2535143, EPI_ISL_2535144, EPI_ISL_2535145, EPI_ISL_2535146, EPI_ISL_2535147, EPI_ISL_2535148, EPI_ISL_2535151, EPI_ISL_2535152, EPI_ISL_2535154, EPI_ISL_2535155, EPI_ISL_2535156, EPI_ISL_2535157, EPI_ISL_2535158, EPI_ISL_2535159, EPI_ISL_2535160, EPI_ISL_2535161, EPI_ISL_2535162, EPI_ISL_2535163, EPI_ISL_2535164, EPI_ISL_2535165, EPI_ISL_2535166, EPI_ISL_2535167, EPI_ISL_2535168, EPI_ISL_2535169, EPI_ISL_2535170, EPI_ISL_2535171, EPI_ISL_2535172, EPI_ISL_2535173, EPI_ISL_2535174, EPI_ISL_2535176, EPI_ISL_2535178, EPI_ISL_2535179, EPI_ISL_2535180, EPI_ISL_2535181, EPI_ISL_2535182, EPI_ISL_2535184, EPI_ISL_2535185, EPI_ISL_2535187, EPI_ISL_2535188, EPI_ISL_2535190, EPI_ISL_2535191, EPI_ISL_2535192, EPI_ISL_2535193, EPI_ISL_2535194, EPI_ISL_2535195, EPI_ISL_2535196, EPI_ISL_2535197, EPI_ISL_2535198, EPI_ISL_2535199, EPI_ISL_2535200, EPI_ISL_2535201, EPI_ISL_2535202, EPI_ISL_2535203, EPI_ISL_2535204, EPI_ISL_2535205, EPI_ISL_2535206, EPI_ISL_2535207, EPI_ISL_2535208, EPI_ISL_2535209, EPI_ISL_2535210, EPI_ISL_2535211, EPI_ISL_2535212, EPI_ISL_2535213, EPI_ISL_2535216, EPI_ISL_2535217, EPI_ISL_2535218, EPI_ISL_2535221, EPI_ISL_2535222, EPI_ISL_2535224, EPI_ISL_2535225, EPI_ISL_2535226, EPI_ISL_2535227, EPI_ISL_2535228, EPI_ISL_2535229, EPI_ISL_2535230, EPI_ISL_2535231, EPI_ISL_2535232, EPI_ISL_2535233, EPI_ISL_2535234, EPI_ISL_2535235, EPI_ISL_2691623, EPI_ISL_2691624, EPI_ISL_2691626, EPI_ISL_2691641, EPI_ISL_2691645, EPI_ISL_2691646 | | | |
| see above | Unidade de apoio ao diagnostico da COVID - UNADIG | Bioinformatics Laboratory / LNCC | Alessandra P Lamarca; Alexandra L Gerber; Amilcar Tanuri; Ana Paula de C Guimaraes; Ana Tereza R Vasconcelos; Andrea Cony Cavalcanti; Caio Luiz Pereira Ribeiro; Cassia Alves; Cintia Policarpo; Claudia Maria Braga de Mello; Cristiane Gomes da Silva; Diana Mariani; Douglas Terra Machado; Flavio Dias da Silva; Gleidson da Silva de Oliveira; Leandro Magalhaes de Souza; Liliane Cavalcante; Luiz G P de Almeida; Marcio Henrique de Oliveira Garcia; Mario Sergio Ribeiro; Ronaldo da Silva F Jr; Silvia Carvalho |
| EPI_ISL_2101508, EPI_ISL_2101509, EPI_ISL_2101511, EPI_ISL_2101513, EPI_ISL_2101671, EPI_ISL_2101714, EPI_ISL_2385669, EPI_ISL_2385670, EPI_ISL_2385671, EPI_ISL_2385674, EPI_ISL_2385675, EPI_ISL_2385676, EPI_ISL_2385677, EPI_ISL_2385678, EPI_ISL_2385679, EPI_ISL_2385680, EPI_ISL_2385681, EPI_ISL_2385682, EPI_ISL_2385683, EPI_ISL_2385684, EPI_ISL_2385685, EPI_ISL_2385686, EPI_ISL_2385689, EPI_ISL_2385690, EPI_ISL_2385691, EPI_ISL_2385692, EPI_ISL_2385693, EPI_ISL_2385694, EPI_ISL_2385695, EPI_ISL_2385697, EPI_ISL_2385698, EPI_ISL_2385699, EPI_ISL_2385700, EPI_ISL_2385701, EPI_ISL_2385702, EPI_ISL_2385704, EPI_ISL_2385705, EPI_ISL_2385707, EPI_ISL_2385708, EPI_ISL_2385709, EPI_ISL_2385711, EPI_ISL_2385712, EPI_ISL_2385713, EPI_ISL_2385714, EPI_ISL_2385717, EPI_ISL_2385718, EPI_ISL_2385719, EPI_ISL_2385720, EPI_ISL_2385723, EPI_ISL_2385724, EPI_ISL_2385725, EPI_ISL_2385726, EPI_ISL_2385727, EPI_ISL_2385728, EPI_ISL_2385729, EPI_ISL_2385730, EPI_ISL_2385731, EPI_ISL_2385733, EPI_ISL_2385734, EPI_ISL_2385735, EPI_ISL_2385736, EPI_ISL_2385737, EPI_ISL_2385739, EPI_ISL_2385740, EPI_ISL_2385741, EPI_ISL_2385742, EPI_ISL_2385743, EPI_ISL_2385745, EPI_ISL_2385748, EPI_ISL_2385749, EPI_ISL_2385750, EPI_ISL_2385751, EPI_ISL_2385752, EPI_ISL_2385753, EPI_ISL_2385754, EPI_ISL_2385755, EPI_ISL_2385757, EPI_ISL_2385758, EPI_ISL_2385759, EPI_ISL_2385760, EPI_ISL_2385761, EPI_ISL_2385764, EPI_ISL_2385765, EPI_ISL_2385766, EPI_ISL_2385768, EPI_ISL_2385769, EPI_ISL_2385770, EPI_ISL_2385771, EPI_ISL_2385772, EPI_ISL_2385773, EPI_ISL_2385774, EPI_ISL_2385776, EPI_ISL_2385777, EPI_ISL_2385778, EPI_ISL_2385779, EPI_ISL_2385780, EPI_ISL_2385781, EPI_ISL_2385782, EPI_ISL_2385784, EPI_ISL_2385785, EPI_ISL_2385786 | | | |
| see above | Unidade de apoio ao diagnóstico da COVID – UNADIG | Bioinformatics Laboratory / LNCC | Alessandra P Lamarca; Alexandra L Gerber; Amilcar Tanuri; Ana Paula de C Guimaraes; Ana Tereza R Vasconcelos; Andrea Cony Cavalcanti; Caio Luiz Pereira Ribeiro; Cassia Alves; Cintia Policarpo; Claudia Maria Braga de Mello; Cristiane Gomes da Silva; Diana Mariani; Douglas Terra Machado; Flavio Dias da Silva; Gleidson da Silva de Oliveira; Leandro Magalhaes de Souza; Liliane Cavalcante; Luiz G P de Almeida; Marcio Henrique de Oliveira Garcia; Mario Sergio Ribeiro; Ronaldo da Silva F Jr; Silvia Carvalho |
| EPI_ISL_2204905 | United Hospital Center | WVU and Marshall University Combined Genomics Core Facilities | James Denvir; Peter Perrotta; Peter Stoilov; Ryan Percifield; Wesley Kimble |
| EPI_ISL_3216907, EPI_ISL_3216908, EPI_ISL_3477562, EPI_ISL_3477566 | Universidad del Valle, LDAB-Laboratorio de Diagnostico de Agentes Biologicos | Universidad del Valle, TAO-Lab, VIREM, NEAS Network | Andres Castillo; Beatriz Parra & COVID-19 Team Univalle; Diana López-Alvarez; Nelson Rivera Franco |
| EPI_ISL_2091058, EPI_ISL_2091059, EPI_ISL_2091061, EPI_ISL_2091062, EPI_ISL_2091063, EPI_ISL_2091064, EPI_ISL_2091078, EPI_ISL_2091079, EPI_ISL_2091080, EPI_ISL_2091108, EPI_ISL_2091109, EPI_ISL_2091111, EPI_ISL_2091112, EPI_ISL_2091113, EPI_ISL_2091114, EPI_ISL_2193388, EPI_ISL_2193389, EPI_ISL_2193390, EPI_ISL_2193391, EPI_ISL_2193393, EPI_ISL_2193401, EPI_ISL_2193402, EPI_ISL_2193403, EPI_ISL_2193404, EPI_ISL_2193405, EPI_ISL_2193406, EPI_ISL_2193407, EPI_ISL_2193411, EPI_ISL_2193414, EPI_ISL_2193417, EPI_ISL_2193435, EPI_ISL_2193437, EPI_ISL_2193438, EPI_ISL_2193439 | | | |
| see above | University Hospitals Translational Laboratory (UHTL), University Hospitals | University Hospitals Translational Laboratory (UHTL), University Hospitals | Alouani, D.; Sadri, N.; Song, X. |
| EPI_ISL_1963270, EPI_ISL_1963272 | University Hospitals of Geneva, Laboratory of Virology | HUG, Laboratory of Virology and the Health2030 Genome Center | Ana Rita Goncalves; Deborah Penet; Emmanouil Dermitzakis; Henri Pegeot; Ioannis Xenarios; Keith Harshman; Laurent Kaiser; Lorenzo Cerutti; Melyssa Elies; Samuel Cordey |
| EPI_ISL_2931009 | University of Arkansas for Medical Sciences | Center for Global Health, University of New Mexico Health Sciences Center | Catherine Kirkpatrick; Darrell Dinwiddie; Daryl Domman; David Ussery; Jennifer Laudadio; Joshua L. Kennedy; Kurt Schwalm; Susan Harley; Uday Chalwadi; Valerie Morley |
| EPI_ISL_2408064 | University of Bari Biomedical Sciences and Human Oncology | University of Bari Biomedical Sciences and Human Oncology | Accogli M.; Chironna M.; Loconsole D.; Sallustio A. |
| EPI_ISL_2476350, EPI_ISL_2476359, EPI_ISL_2476365, EPI_ISL_2476370, EPI_ISL_2691960, EPI_ISL_2691964, EPI_ISL_2691967, EPI_ISL_2691988, EPI_ISL_2691991, EPI_ISL_2692011, EPI_ISL_2931108, EPI_ISL_3388709 | | | |
| see above | University of Chicago Medicine | RIPHL at Rush University Medical Center | Edith Perez; Felix Araujo Perez; Kevin Kunstman; Laura Furtado; Lauren Megger; Marieta Hyde; Max Kolton; Stefan Green |
| EPI_ISL_2476390, EPI_ISL_2476391, EPI_ISL_2476393, EPI_ISL_2476400, EPI_ISL_2692086, EPI_ISL_2692096, EPI_ISL_2692100, EPI_ISL_2692101, EPI_ISL_2692103, EPI_ISL_2692104, EPI_ISL_2692107, EPI_ISL_2692108, EPI_ISL_2692113, EPI_ISL_2692114, EPI_ISL_2692122, EPI_ISL_2692125, EPI_ISL_2692129, EPI_ISL_2692131, EPI_ISL_2692132 | | | |
| see above | University of Illinois at Chicago | RIPHL at Rush University Medical Center | Felix Araujo Perez; Julie Bentea; Kevin Kunstman; Laura Furtado; Marieta Hyde; Max Kolton; Omar Perez; Stefan Green |
| EPI_ISL_2001220, EPI_ISL_2001240, EPI_ISL_2001258, EPI_ISL_2100396, EPI_ISL_2136244, EPI_ISL_2376042, EPI_ISL_2400358, EPI_ISL_2400362, EPI_ISL_2400366, EPI_ISL_2400372 | | | |
| see above | University of Michigan Clinical Microbiology Laboratory | Lauring Lab, University of Michigan, Department of Microbiology and Immunology | Gilbert; Valesano |
| EPI_ISL_2404840, EPI_ISL_2404846 | University of Oregon COVID-19 MAP Laboratory | University of Oregon Genomics and Cell Charactericzation Core Facility (GC3F) | Ariana White; Douglas Turnbull; Jason Carriere; Jason Sydes; Jeff Bishop; Peter Batzel |
| EPI_ISL_1850441 | UniversitÃ¤tsklinikum MÃ¼nster Institut fÃ¼r Virologie | Robert Koch Institute | |
| EPI_ISL_2086795, EPI_ISL_2086872, EPI_ISL_2136993, EPI_ISL_2449934, EPI_ISL_2510857, EPI_ISL_2510860, EPI_ISL_2510880, EPI_ISL_2510900, EPI_ISL_2510924, EPI_ISL_2510943, EPI_ISL_2510979, EPI_ISL_2510999, EPI_ISL_2511002, EPI_ISL_2511008 | | | |
| see above | UniversitŠtsSpital ZŸrich | Institute of Medical Virology | Alexandra Trkola; Annette AudigŽ; Catharine Aquino; Cyril Shah; Daniel Ehrsam; Gabriela Ziltener; Guido Bloemberg; Hubert Rehrauer; Isabel StŸrmer; Joel Wirz; Jon Huder; JŸrg Bšni; Kevin Steiner; Maria GrŸnberg; Maryam Zaheri; Michael Huber; Riccarda Capaul; Stefan Schmutz; Verena Kufner; Weihong Qi |
| EPI_ISL_2152435, | Usansolo-Galdakao University Hospital | Cruces University Hospital | Ana Belén de la Hoz; Ana Gual-de-Torrella; Izaskun Alejo-Cancho; Mikel Gallego |

| | | | |
|---|---|---|---|
| EPI_ISL_2408062, EPI_ISL_2408063, EPI_ISL_2483459 | | | |
| EPI_ISL_2284852, EPI_ISL_2292200, EPI_ISL_2292801, EPI_ISL_2456321, EPI_ISL_2456350, EPI_ISL_2456508, EPI_ISL_2456649, EPI_ISL_2456657, EPI_ISL_2458490, EPI_ISL_2679471, EPI_ISL_2679601, EPI_ISL_2679632, EPI_ISL_2855894, EPI_ISL_2856068, EPI_ISL_2856073, EPI_ISL_2856080, EPI_ISL_2856095, EPI_ISL_2856452, EPI_ISL_2856473, EPI_ISL_2856496, EPI_ISL_2857455, EPI_ISL_2857920, EPI_ISL_2857948, EPI_ISL_2857955, EPI_ISL_2857979, EPI_ISL_2857990, EPI_ISL_2858009, EPI_ISL_2858015, EPI_ISL_2858028, EPI_ISL_3269714 | | | |
| see above | Utah Public Health Laboratory | Utah Public Health Laboratory | Erin L. Young; Kelly F. Oakeson; Olinto Linares-Perdomo; Tara Gallagher |
| EPI_ISL_2210083, EPI_ISL_2344951, EPI_ISL_2344958, EPI_ISL_2344959, EPI_ISL_2344962, EPI_ISL_2344964, EPI_ISL_2344968, EPI_ISL_2344970, EPI_ISL_2345039, EPI_ISL_2345040 | | | |
| see above | VIGILANCIA EM SAUDE DE FCO MORATO | Instituto Butantan | Antonio Jorge Martins; Claudia Renata dos Santos Barros; David Schlesinger; Debora Botequio Moretti; Dimas Tadeu Covas; Elaine Cristina Marqueze; Elaine Vieira Santos; Evandra Strazza Rodrigues; Heidge Fukumasu; Jayme Augusto de Souza-Neto; José Salvatore Leister Patané; Luiz Alcantara; Luiz Lehmann Coutinho; Maria Carolina Elias; Maurício Lacerda Nogueira; Rafael dos Santos Bezerra; Raul Machado Neto; Rejane Maria Tommasini Grotto; Ricardo Haddad; Sandra Coccuzzo Sampaio Vessoni; Simone Kashima; Svetoslav Nanev Slavov; Vincent Louis Viala; Vincent Louis Viala. |
| EPI_ISL_2170994 | VIGILANCIA EM SAUDE DE FCO MORATO | Instituto Butantan / Mendelics | Antonio Jorge Martins; Bianca Cechetto Carlos. Mendelics: Bibiana Santos; Claudia Renata dos Santos Barros; Cíntia Bittar; David Schlesinger. Hemocentro Ribeirão Preto: Simone Kashima; Debora Botequio Moretti; Elaine Cristina Marqueze; Elaine Vieira dos Santos; Elisangela Chicaroni Mattos; Erika Freitas; Evandra Strazza Rodrigues; Felipe Allan da Silva da Costa; Flavia Aburjaile; Fábio Sossai Possebon; Guilherme Campos; Guilherme Targino Valente; Heidge Fukumasu. USP-Botucatu: Rejane Maria Tommasini Grotto; Helena Lage Ferreira; Instituto Butantan: Dimas Tadeu Covas; Jardelina de Souza Todao Bernardino; Jayme A. Souza-Neto; Jessika Cristina Chagas Lesbon; Jorge A. Petroli Marchesi; José Salvatore Leister Patané; João Paulo Kitajima; João Pessoa Araújo Jr.; Leila Sabrina Ullmann; Loyze Paola Oliveira de Lima; Luiz Aurelio de Campos Crispin. Centro de Genômica Funcional da ESALQ: Luiz Lehmann Coutinho; Luiz Carlos Junior de Alcantara; Lívia Sacchetto; Maisa C. Pereira Parra; Maria Carolina Elias; Marta Giovanetti; Marília Moraes; Maurício Lacerda Nogueira. Prefeitura de Sao Paulo: Melissa Palmieri.; Patricia Akemi Assato; Paula Rahal; Paulo Inacio da Costa; Rafael dos Santos Bezerra; Raquel de Lello Rocha Campos Cassano. NGS Soluções Genômicas: Pilar Drummond Sampaio Corrêa Mariani. FZEA-USP Pirassununga: Mirele Daiana Poleti; Raul Machado Neto; Ricardo Augusto Brassaloti; Ricardo Haddad; Rodrigo Tocantins Calado. FAMERP-SJRP: Cecília Artico Banho; Sandra Coccuzzo Sampaio; Svetoslav Nanev Slavov; Vagner Fonseca; Vincent Louis Viala |
| EPI_ISL_2209304, EPI_ISL_2209309, EPI_ISL_2345651, EPI_ISL_2362250 | VIGILANCIA EM SAUDE SAE VIGILANCIA EPIDEMIOLOGICA | Instituto Butantan / Mendelics | Antonio Jorge Martins; Claudia Renata dos Santos Barros; David Schlesinger; Debora Botequio Moretti; Dimas Tadeu Covas; Elaine Cristina Marqueze; Elaine Vieira Santos; Evandra Strazza Rodrigues; Heidge Fukumasu; Jayme Augusto de Souza-Neto; José Salvatore Leister Patané; Luiz Alcantara; Luiz Lehmann Coutinho; Maria Carolina Elias; Maurício Lacerda Nogueira; Rafael dos Santos Bezerra; Raul Machado Neto; Rejane Maria Tommasini Grotto; Ricardo Haddad; Sandra Coccuzzo Sampaio Vessoni; Simone Kashima; Svetoslav Nanev Slavov; Vincent Louis Viala |
| EPI_ISL_2493366, EPI_ISL_2493395, EPI_ISL_2493399, EPI_ISL_2493450, EPI_ISL_2494335 | VIGILANCIA EPIDEMIOLOGICA DE LEME | Instituto Butantan | Antonio Jorge Martins; Claudia Renata dos Santos Barros; David Schlesinger; Debora Botequio Moretti; Dimas Tadeu Covas; Elaine Cristina Marqueze; Elaine Vieira Santos; Evandra Strazza Rodrigues; Heidge Fukumasu; Jayme Augusto de Souza-Neto; José Salvatore Leister Patané; Luiz Alcantara; Luiz Lehmann Coutinho; Maria Carolina Elias; Maurício Lacerda Nogueira; Rafael dos Santos Bezerra; Raul Machado Neto; Rejane Maria Tommasini Grotto; Ricardo Haddad; Sandra Coccuzzo Sampaio Vessoni; Simone Kashima; Svetoslav Nanev Slavov; Vincent Louis Viala |
| EPI_ISL_2493365, EPI_ISL_2493391, EPI_ISL_2493432, EPI_ISL_2493434, EPI_ISL_2493448, EPI_ISL_2500982 | VIGILANCIA EPIDEMIOLOGICA E CONTROLE DE VETORES PIRASSUNUN | Instituto Butantan | Antonio Jorge Martins; Claudia Renata dos Santos Barros; David Schlesinger; Debora Botequio Moretti; Dimas Tadeu Covas; Elaine Cristina Marqueze; Elaine Vieira Santos; Evandra Strazza Rodrigues; Heidge Fukumasu; Jayme Augusto de Souza-Neto; José Salvatore Leister Patané; Luiz Alcantara; Luiz Lehmann Coutinho; Maria Carolina Elias; Maurício Lacerda Nogueira; Rafael dos Santos Bezerra; Raul Machado Neto; Rejane Maria Tommasini Grotto; Ricardo Haddad; Sandra Coccuzzo Sampaio Vessoni; Simone Kashima; Svetoslav Nanev Slavov; Vincent Louis Viala |
| EPI_ISL_2344830 | VIGILANCIA EPIDEMIOLOGICA E CONTROLE DE VETORES PIRASSUNUN | Instituto Butantan / ESALQ-Piracicaba | Antonio Jorge Martins; Claudia Renata dos Santos Barros; David Schlesinger; Debora Botequio Moretti; Dimas Tadeu Covas; Elaine Cristina Marqueze; Elaine Vieira Santos; Evandra Strazza Rodrigues; Heidge Fukumasu; Jayme Augusto de Souza-Neto; José Salvatore Leister Patané; Luiz Alcantara; Luiz Lehmann Coutinho; Maria Carolina Elias; Maurício Lacerda Nogueira; Rafael dos Santos Bezerra; Raul Machado Neto; Rejane Maria Tommasini Grotto; Ricardo Haddad; Sandra Coccuzzo Sampaio Vessoni; Simone Kashima; Svetoslav Nanev Slavov; Vincent Louis Viala |
| EPI_ISL_2493599, EPI_ISL_2493610 | VIGILANCIA EPIDEMIOLOGICA JARDINOPOLIS SP | Instituto Butantan | Antonio Jorge Martins; Claudia Renata dos Santos Barros; David Schlesinger; Debora Botequio Moretti; Dimas Tadeu Covas; Elaine Cristina Marqueze; Elaine Vieira Santos; Evandra Strazza Rodrigues; Heidge Fukumasu; Jayme Augusto de Souza-Neto; José Salvatore Leister Patané; Luiz Alcantara; Luiz Lehmann Coutinho; Maria Carolina Elias; Maurício Lacerda Nogueira; Rafael dos Santos Bezerra; Raul Machado Neto; Rejane Maria Tommasini Grotto; Ricardo Haddad; Sandra Coccuzzo Sampaio Vessoni; Simone Kashima; Svetoslav Nanev Slavov; Vincent Louis Viala |
| EPI_ISL_2209094, EPI_ISL_2345645, EPI_ISL_2345744, EPI_ISL_2345782, EPI_ISL_2345787, EPI_ISL_2345869 | VIGILANCIA EPIDEMIOLOGICA JARDINOPOLIS SP | Instituto Butantan / Mendelics | Antonio Jorge Martins; Claudia Renata dos Santos Barros; David Schlesinger; Debora Botequio Moretti; Dimas Tadeu Covas; Elaine Cristina Marqueze; Elaine Vieira Santos; Evandra Strazza Rodrigues; Heidge Fukumasu; Jayme Augusto de Souza-Neto; José Salvatore Leister Patané; Luiz Alcantara; Luiz Lehmann Coutinho; Maria Carolina Elias; Maurício Lacerda Nogueira; Rafael dos Santos Bezerra; Raul Machado Neto; Rejane Maria Tommasini Grotto; Ricardo Haddad; Sandra Coccuzzo Sampaio Vessoni; Simone Kashima; Svetoslav Nanev Slavov; Vincent Louis Viala |
| EPI_ISL_2445607 | VIGILANCIA EPIDIMIOLOGICA DE ARACARIGUAMA | Instituto Butantan | Antonio Jorge Martins; Claudia Renata dos Santos Barros; David Schlesinger; Debora Botequio Moretti; Dimas Tadeu Covas; Elaine Cristina Marqueze; Elaine Vieira Santos; Evandra Strazza Rodrigues; Heidge Fukumasu; Jayme Augusto de Souza-Neto; José Salvatore Leister Patané; Luiz Alcantara; Luiz Lehmann Coutinho; Maria Carolina Elias; Maurício Lacerda Nogueira; Rafael dos Santos Bezerra; Raul Machado Neto; Rejane Maria Tommasini Grotto; Ricardo Haddad; Sandra Coccuzzo Sampaio Vessoni; Simone Kashima; Svetoslav Nanev Slavov; Vincent Louis Viala |
| EPI_ISL_2345668 | VIGILANCIA EPIDIMIOLOGICA DE ARACARIGUAMA | Instituto Butantan / Mendelics | Antonio Jorge Martins; Claudia Renata dos Santos Barros; David Schlesinger; Debora Botequio Moretti; Dimas Tadeu Covas; Elaine Cristina Marqueze; Elaine Vieira Santos; Evandra Strazza Rodrigues; Heidge Fukumasu; Jayme Augusto de Souza-Neto; José Salvatore Leister Patané; Luiz Alcantara; Luiz Lehmann Coutinho; Maria Carolina Elias; Maurício Lacerda Nogueira; Rafael dos Santos Bezerra; Raul Machado Neto; Rejane Maria Tommasini Grotto; Ricardo Haddad; Sandra Coccuzzo Sampaio Vessoni; Simone Kashima; Svetoslav Nanev Slavov; Vincent Louis Viala |
| EPI_ISL_2445282, EPI_ISL_2445285, EPI_ISL_2445286, EPI_ISL_2445290, EPI_ISL_2445291, EPI_ISL_2445293, EPI_ISL_2445294, EPI_ISL_2445296, EPI_ISL_2445303, EPI_ISL_2473822 | | | |
| see above | VIGILANCIA SANITARIA E VIG EPIDEMIOLOGICA DE ITAPEVI | Instituto Butantan | Antonio Jorge Martins; Claudia Renata dos Santos Barros; David Schlesinger; Debora Botequio Moretti; Dimas Tadeu Covas; Elaine Cristina Marqueze; Elaine Vieira Santos; Evandra Strazza Rodrigues; Heidge Fukumasu; Jayme Augusto de Souza-Neto; José Salvatore Leister Patané; Luiz Alcantara; Luiz Lehmann Coutinho; Maria Carolina Elias; Maurício Lacerda Nogueira; Rafael dos Santos Bezerra; Raul Machado Neto; Rejane Maria Tommasini Grotto; Ricardo Haddad; Sandra Coccuzzo Sampaio Vessoni; Simone Kashima; Svetoslav Nanev Slavov; Vincent Louis Viala |
| EPI_ISL_2109687 | VIROLOGY LABORATORY-CHU NICE | VIROLOGY LABORATORY-CHU NICE | Aicha El Yakine; Geraldine Gonfrier; Jean Machowiak; Sebastien Vitale; Valerie Giordanengo; Virginie Flahou |
| EPI_ISL_2444192 | VOTUPORANGA | Instituto Butantan / FAMERP | Antonio Jorge Martins; Claudia Renata dos Santos Barros; David Schlesinger; Debora Botequio Moretti; Dimas Tadeu Covas; Elaine Cristina Marqueze; Elaine Vieira Santos; Evandra Strazza Rodrigues; Heidge Fukumasu; Jayme Augusto de Souza-Neto; José Salvatore Leister Patané; Luiz Alcantara; Luiz Lehmann Coutinho; Maria Carolina Elias; Maurício Lacerda Nogueira; Rafael dos Santos Bezerra; Raul Machado Neto; Rejane Maria Tommasini Grotto; Ricardo Haddad; Sandra Coccuzzo Sampaio Vessoni; Simone Kashima; Svetoslav Nanev Slavov; Vincent Louis Viala |
| EPI_ISL_1914207 | Viollier AG | Department of Biosystems Science and Engineering, ETH Zurich | Chaoran Chen; Christian Beisel; Christiane Beckmann; Christoph Noppen; David Dreifuss; Elodie Burcklen; Ina Nissen; Ivan Topolsky; Katharina Jahn; Lara Fuhrmann; Maurice Redondo; Mirjam Feldkamp; Natascha Santacroce; Niko Beerenwinkel; Noemie Santamaria de Souza; Olivier Kobel; Philipp Jablonski; Rebecca Denes; Sarah Nadeau; Sophie Seidel; Tanja Stadler |
| EPI_ISL_2151992, EPI_ISL_2212753, EPI_ISL_2307457, EPI_ISL_2307461, EPI_ISL_2307612, EPI_ISL_2307706, EPI_ISL_2307750, EPI_ISL_2307771, EPI_ISL_2307844, EPI_ISL_2307864, EPI_ISL_2308026, EPI_ISL_2308074 | | | |
| see above | Viollier AG | Department of Biosystems Science and Engineering, ETH Zürich | Andrea Patrignani; Andreia Cabral de Gouvea; Catharine Aquino; Chaoran Chen; Christian Beisel; Christiane Beckmann; Christoph Noppen; Daniel Ehrsam; David Dreifuss; Doris Popovic; Elodie Burcklen; Griffin White; Ina Nissen; Isabel Stürmer; Ivan Topolsky; Jay Tracy; Katharina Jahn; Lara Fuhrmann; Laura Neff; Lennart Opitz; Maria Domenica Moccia; Maurice Redondo; Mirjam Feldkamp; Natascha Santacroce; Niko Beerenwinkel; Noemie Santamaria de Souza; Olivier Kobel; Philipp Jablonski; Ralph Schlapbach; Rebecca Denes; Sarah Nadeau; Simon Grüter; Sophie Seidel; Tanja Stadler; Timothy Sykes |
| EPI_ISL_2226439 | Viollier AG | Viollier AG | Andrea Patrizia Salzmann; Christiane Beckmann; Christoph Noppen; Henriette Kurth; Maurice Redondo; Olivier Kobel |
| EPI_ISL_2003686, EPI_ISL_2101937, EPI_ISL_2101940, EPI_ISL_2344478, EPI_ISL_2344479, EPI_ISL_2344485, EPI_ISL_2344490, EPI_ISL_2344505, EPI_ISL_2603207, EPI_ISL_2603208, EPI_ISL_2603217 | | | |
| see above | Virginia Division of Consolidated Laboratory Services | Virginia Division of Consolidated Laboratory Services | Virginia DCLS |
| EPI_ISL_2105163, EPI_ISL_2451955 | WA State Department of Health | Centers for Disease Control and Prevention Division of Viral Diseases, Pathogen Discovery | Alison Laufer Halpin; Ben L. Rambo-Martin; Clinton R. Paden; Dakota Howard; Darlene Wagner; Dave Wentworth; Dhwani Batra; Jasmine Padilla; Justin Lee; Katie Dillon; Krista Queen; Kristen Knipe; Kristine Lacek; Mark Burroughs; Matthew Schmerer; Mili Sheth; Peter Cook; Sam Shepard; Sarah Nobles; Shoshona Le; Suxiang Tong; Vivien Dugan; Yvette Unoarumhi |
| EPI_ISL_2097314, EPI_ISL_2274650, EPI_ISL_2627011 | WESTCHESTER MEDICAL CENTER | Wadsworth Center, New York State Department of Health | Alexis Russell; Catharine Prussing; Daryl M. Lamson; Erasmus Schneider; Erica Lasek-Nesselquist; John Kelly; Jonathan Plitnick; Kirsten St. George; Matthew Shudt; Melissa A Leisner; Navjot Singh |
| EPI_ISL_2385106 | WHO National Influenza Centre Russian Federation | WHO National Influenza Centre Russian Federation | Andrey Komissarov; Artem Fadeev; Daria Danilenko; Dmitry Lioznov; Elena Nabieva; Georgii Bazykin; Kirill Varchenko; Ksenia Safina; Kseniya Komissarova; Maria Pisareva; Maria Timofeeva; Mikhail Bakaev; Nikita Yolshin; Oula Masour; Tamila Musaeva; Veronika Eder |
| EPI_ISL_2248729, EPI_ISL_2452015 | WI State Laboratory of Hygiene | Centers for Disease Control and Prevention Division of Viral Diseases, Pathogen Discovery | Alison Laufer Halpin; Ben L. Rambo-Martin; Clinton R. Paden; Dakota Howard; Darlene Wagner; Dave Wentworth; Dhwani Batra; Jasmine Padilla; Justin Lee; Katie Dillon; Krista Queen; Kristen Knipe; Kristine Lacek; Mark Burroughs; Matthew Schmerer; Mili Sheth; Peter Cook; Sam Shepard; Sarah Nobles; Shoshona Le; Suxiang Tong; Vivien Dugan; Yvette Unoarumhi |
| EPI_ISL_2533758 | WLA VAMC | Genomics and Discovery, Respiratory Viruses Branch, Division of Viral Diseases, Centers for Disease Control and Prevention | Adam Retchless; Anna Kelleher; Anna Uehara; Brian Lynch; Clinton R. Paden; Dhwani Batra; Haibin Wang; Han Jia Justin Ng; Jasmine Padilla; Jing Zhang; Justin Lee; Krista Queen; Mark Burroughs; Mili Sheth; Morgan Davis; Peter Cook; Rachel Marine; Sarah Nobles; Suxiang Tong; Tara Coalter; Yan Li; Ying Tao |
| EPI_ISL_2452097 | WY Public Health Laboratory | Centers for Disease Control and Prevention Division of Viral Diseases, Pathogen Discovery | Alison Laufer Halpin; Ben L. Rambo-Martin; Clinton R. Paden; Dakota Howard; Darlene Wagner; Dave Wentworth; Dhwani Batra; Jasmine Padilla; Justin Lee; Katie Dillon; Krista Queen; Kristen Knipe; Kristine Lacek; Mark Burroughs; Matthew Schmerer; Mili Sheth; Peter Cook; Sam Shepard; Sarah Nobles; Shoshona Le; Suxiang Tong; Vivien Dugan; Yvette Unoarumhi |
| EPI_ISL_1897403, EPI_ISL_1897404, EPI_ISL_1897405, EPI_ISL_1897407, EPI_ISL_1897408, EPI_ISL_1897409, EPI_ISL_1897416, EPI_ISL_1897429, EPI_ISL_2096103, EPI_ISL_2096104, EPI_ISL_2096105, EPI_ISL_2096109, EPI_ISL_2096110, EPI_ISL_2096111, EPI_ISL_2096112, EPI_ISL_2096113, EPI_ISL_2336157, EPI_ISL_2336164, EPI_ISL_2336170, EPI_ISL_2336171, EPI_ISL_2336173, EPI_ISL_2336196, EPI_ISL_2336207, EPI_ISL_2336212, EPI_ISL_2336242, EPI_ISL_2336244, EPI_ISL_2336245, EPI_ISL_2378831, EPI_ISL_2378838, EPI_ISL_2378842, EPI_ISL_2378843, EPI_ISL_2378844, EPI_ISL_2378845, EPI_ISL_2378846, EPI_ISL_2378847, EPI_ISL_2378848, EPI_ISL_2378849, EPI_ISL_2378881, EPI_ISL_2378883, EPI_ISL_2378884, EPI_ISL_2378885, EPI_ISL_2378886, EPI_ISL_2378891, EPI_ISL_2443465, EPI_ISL_2557050, EPI_ISL_2651041 | | | |
| see above | Washington State Department of Health Public Health Laboratories | Washington State Department of Health Public Health Laboratories | Avi Singh; Darren Lucas; Denny Russell; Drew MacKellar; Geoff Melly; Hannah Gray; Joenice Gonzalez; JohnAric Peterson; Philip Dykema; Rebecca Cao; Vanessa De Los Santos |
| EPI_ISL_2402825 | Weill Cornell Medicine | New York Genome Center | Andre Corvelo; Arryn Craney; Chris Mason; Dayna M. Oschwald; Hanna Rennert; Lars F Westblade; Margaret Elizabeth Ross; Melissa Cushing; Michael Zody; Olivier Elemento; Priya Velu; Samantha Fennessey; Tom Maniatis |
| EPI_ISL_2376267, EPI_ISL_2399759, EPI_ISL_2657692, EPI_ISL_2657693, EPI_ISL_2657695 | Wexner Medical Center | The Ohio State University College of Medicine | Koenig, S.; Seminetta, J. |
| EPI_ISL_1760181, EPI_ISL_1805458, EPI_ISL_1805780, EPI_ISL_1807191, EPI_ISL_1807199, EPI_ISL_1819295, EPI_ISL_1819296, EPI_ISL_1854743, EPI_ISL_1854746, EPI_ISL_1854748, EPI_ISL_1854751, EPI_ISL_1857044, EPI_ISL_1857061, EPI_ISL_1857070, EPI_ISL_1857087, EPI_ISL_1940143, EPI_ISL_1961015, EPI_ISL_2023321, EPI_ISL_2023341, EPI_ISL_2023343, EPI_ISL_2086433, EPI_ISL_2086435, | | | |

| | | | |
|---|---|---|---|
| EPI_ISL_2092839, EPI_ISL_2156025, EPI_ISL_2156026, EPI_ISL_2156034, EPI_ISL_2227391, EPI_ISL_2248756, EPI_ISL_2276085, EPI_ISL_2284856, EPI_ISL_2284866 | | | |
| see above | Wichita State University - Molecular Diagnostics Lab | Kansas Health and Environmental Lab | Ben Olsen; Jonathan Barnell; Katherine Wiggins; Mike Grose; and Phil Adam |
| EPI_ISL_2279001, EPI_ISL_2279054, EPI_ISL_2279087, EPI_ISL_2279132, EPI_ISL_2279137, EPI_ISL_2279143, EPI_ISL_2279180, EPI_ISL_2279286, EPI_ISL_2279292, EPI_ISL_2279316, EPI_ISL_2279327, EPI_ISL_2279337, EPI_ISL_2279367, EPI_ISL_2279394, EPI_ISL_2279430, EPI_ISL_2279435, EPI_ISL_2279446, EPI_ISL_2279454, EPI_ISL_2279460, EPI_ISL_2279464, EPI_ISL_2279466, EPI_ISL_2279470, EPI_ISL_2279474, EPI_ISL_2279482, EPI_ISL_2279497, EPI_ISL_2558085, EPI_ISL_2558092, EPI_ISL_2558094, EPI_ISL_2558097, EPI_ISL_2558150, EPI_ISL_2558168, EPI_ISL_2558171, EPI_ISL_2558174, EPI_ISL_2558222, EPI_ISL_2558243, EPI_ISL_2558268, EPI_ISL_2558327, EPI_ISL_2558332, EPI_ISL_2558340, EPI_ISL_2558359, EPI_ISL_2558361, EPI_ISL_2558375, EPI_ISL_2558379, EPI_ISL_2558384, EPI_ISL_2558436, EPI_ISL_2558447, EPI_ISL_2558481, EPI_ISL_2558484, EPI_ISL_2558562, EPI_ISL_2558569, EPI_ISL_2558605, EPI_ISL_2558627 | | | |
| see above | Wisconsin State Laboratory of Hygiene Communicable Disease Division | Wisconsin State Laboratory of Hygiene Communicable Disease Division | Abigail C. Shockey; Alicia J. Mooney; Erika M. Hanson; Kelsey R. Florek; Richard Griesser; Sara Wagner; Tonya Danz |
| EPI_ISL_2023097, EPI_ISL_2023184, EPI_ISL_2023211, EPI_ISL_2296383, EPI_ISL_2296387, EPI_ISL_2296389, EPI_ISL_2426129, EPI_ISL_2501685, EPI_ISL_2501703, EPI_ISL_2501704, EPI_ISL_2501705 | | | |
| see above | Yale Clinical Virology Lab | Grubaugh Lab - Yale School of Public Health | Anderson Brito; Annie Watkins; Chaney Kalinich; Chantal Vogels; Isabel Ott; Jessica Rothman; Joseph Fauver; Mallery Breban; Marie L. Landry; Mary Petrone; Nathan Grubaugh; Tara Alpert |
| EPI_ISL_2360232 | Zentrallabor Zürich | Institute of Medical Virology | Alexandra Trkola; Annette Audigé; Cyril Shah; Gabriela Ziltener; Guido Bloemberg; Jon Huder; Jürg Böni; Kevin Steiner; Maria Grünberg; Maryam Zaheri; Michael Huber; Riccarda Capaul; Stefan Schmutz; Verena Kufner |
| EPI_ISL_2445858 | unknown | Instituto Nacional de Saude (INSA) | Borges et al |
| EPI_ISL_2488813 | Evandro Chagas Institute | Evandro Chagas Institute | A.M.; Barbagelata; E.C.; E.M.A.; Ferreira; J.A.; Junior; K.C.; L.C.; L.S.; M.C.; P.S.; Pinheiro; Santos; Silva; Sousa; Sousa Junior; W.D.C.; da Silva |

We gratefully acknowledge the following Authors from the Originating laboratories responsible for obtaining the specimens, as well as the Submitting laboratories where the genome data were generated and shared via GISAID, on which this research is based.

All Submitters of data may be contacted directly via www.gisaid.org

Authors are sorted alphabetically.

| Accession ID | Originating Laboratory | Submitting Laboratory | Authors |
|---|---|---|---|
| EPI_ISL_3354027 | "AK State Public Health Lab, State Health Department" | Centers for Disease Control and Prevention Division of Viral Diseases, Pathogen Discovery | Alex Burgin; Ben Rambo-Martin; Clinton Paden; Dakota Howard; Dave Wentworth; Dhwani Batra; Jasmine Padilla; Justin Lee; Krista Queen; Kristen Knipe; Kristine Lacek; Mark Burroughs; Matthew Schmerer; Meghan Bentz; Mili Sheth; Peter Cook; Sam Shepard; Sarah Nobles; Suxiang Tong; Vivien Dugan; Yvette Unoarumhi |
| EPI_ISL_2645302, EPI_ISL_2645305 | "CO Dept. of Public Health and Environment, Lab Services Division" | Centers for Disease Control and Prevention Division of Viral Diseases, Pathogen Discovery | Alison Laufer Halpin; Ben L. Rambo-Martin; Clinton R. Paden; Dakota Howard; Darlene Wagner; Dave Wentworth; Dhwani Batra; Jasmine Padilla; Justin Lee; Katie Dillon; Krista Queen; Kristen Knipe; Kristine Lacek; Mark Burroughs; Matthew Schmerer; Mili Sheth; Peter Cook; Sam Shepard; Sarah Nobles; Shoshona Le; Suxiang Tong; Vivien Dugan; Yvette Unoarumhi |
| EPI_ISL_1711723 | "NM Dept. Health, Scientific Laboratory Division " | Centers for Disease Control and Prevention Division of Viral Diseases, Pathogen Discovery | Alison Laufer Halpin; Ben L. Rambo-Martin; Clinton R. Paden; Dakota Howard; Darlene Wagner; Dave Wentworth; Dhwani Batra; Jasmine Padilla; Justin Lee; Katie Dillon; Krista Queen; Kristen Knipe; Kristine Lacek; Mark Burroughs; Matthew Schmerer; Mili Sheth; Peter Cook; Sam Shepard; Sarah Nobles; Shoshona Le; Suxiang Tong; Vivien Dugan; Yvette Unoarumhi |
| EPI_ISL_2720456 | A.S.L. VERCELLI | Fondazione del Piemonte per l'Oncologia IRCCS | Antonino Sottile; Giorgio Giardina; Paola Marino; Silvia Brossa |
| EPI_ISL_1499576, EPI_ISL_1499577 | ABC Algarve | Instituto Nacional de Saude (INSA) | Borges et al |
| EPI_ISL_3347192, EPI_ISL_3347193 | ADILAB | Laboratorio Departamental de Salud Publica de Antioquia | Andres F. Cardona-Rios; Gloria Isabel Escobar; Idabely Betancur Ortiz; Juan P. Hernandez-Ortiz; Maria Stella López |
| EPI_ISL_3354677 | AL Dept. of Public Health Bureau of Clinical Laboratories | Centers for Disease Control and Prevention Division of Viral Diseases, Pathogen Discovery | Alex Burgin; Ben Rambo-Martin; Clinton Paden; Dakota Howard; Dave Wentworth; Dhwani Batra; Jasmine Padilla; Justin Lee; Krista Queen; Kristen Knipe; Kristine Lacek; Mark Burroughs; Matthew Schmerer; Meghan Bentz; Mili Sheth; Peter Cook; Sam Shepard; Sarah Nobles; Suxiang Tong; Vivien Dugan; Yvette Unoarumhi |
| EPI_ISL_1756002, EPI_ISL_1756003, EPI_ISL_1756015 | ALBANY MEDICAL CENTER | Wadsworth Center, New York State Department of Health | Alexis Russell; Catharine Prussing; Daryl M. Lamson; Erasmus Schneider; Erica Lasek-Nesselquist; John Kelly; Jonathan Plitnick; Kirsten St. George; Matthew Shudt; Melissa A Leisner; Navjot Singh |
| EPI_ISL_1336264 | ALGEMEEN MEDISCH LABO | UAntwerp, Laboratory of Medical Microbiology | Basil Britto Xavier; Christine Lammens; Herman Goossens; Jasmine Coppens; Marie Le Mercier; Veerle Matheeussen |
| EPI_ISL_1795113, EPI_ISL_2345312 | AMBULATORIO DE ATENDIMENTO DE DST DE GUARIBA | Instituto Butantan / ESALQ-Piracicaba | Antonio Jorge Martins; Bianca Cechetto Carlos. Mendelics: Bibiana Santos; Claudia Renata dos Santos Barros; David Schlesinger; David Schlesinger. Hemocentro Ribeirão Preto: Simone Kashima; Debora Botequio Moretti; Debora Botequio Moretti. Centro de Genômica Funcional da ESALQ: Luiz Lehmann Coutinho; Dimas Tadeu Covas; Elaine Cristina Marqueze; Elaine Vieira Santos; Elaine Vieira dos Santos; Elisangela Chicaroni Mattos; Erika Freitas; Evandra Strazza Rodrigues; Felipe Allan da Silva da Costa; Flavia Aburjaile; Guilherme Targino Valente; Heidge Fukumasu; Heidge Fukumasu. USP-Botucatu: Rejane Maria Tommasini Grotto; Instituto Butantan: Alexander Roberto Precioso; Jayme A. Souza-Neto; Jayme Augusto de Souza-Neto; Jessika Cristina Chagas Lesbon; José Salvatore Leister Patané; João Paulo Kitajima; Luiz Alcantara; Luiz Carlos Junior de Alcantara; Luiz Lehmann Coutinho; Maria Carolina Elias; Marta Giovanetti; Maurício Lacerda Nogueira; Patricia Akemi Assato; Rafael dos Santos Bezerra; Raquel de Lello Rocha Campos Cassano. NGS Soluções Genômicas: Pilar Drummond Sampaio Corrêa Mariani. FZEA-USP Pirassununga: Mirele Daiana Poleti; Raul Machado Neto; Rejane Maria Tommasini Grotto; Ricardo Augusto Brassaloti; Ricardo Haddad; Rodrigo Tocantins Calado.; Sandra Coccuzzo Sampaio; Sandra Coccuzzo Sampaio Vessoni; Simone Kashima; Svetoslav Nanev Slavov; Vagner Fonseca; Vincent Louis Viala |
| EPI_ISL_1795200, EPI_ISL_1795207, EPI_ISL_1795292, EPI_ISL_2345423, EPI_ISL_2345430, EPI_ISL_2345532 | AMBULATORIO DE ESPECIALIDADE V E MOGI MIRIM | Instituto Butantan / ESALQ-Piracicaba | Antonio Jorge Martins; Bianca Cechetto Carlos. Mendelics: Bibiana Santos; Claudia Renata dos Santos Barros; David Schlesinger; David Schlesinger. Hemocentro Ribeirão Preto: Simone Kashima; Debora Botequio Moretti; Debora Botequio Moretti. Centro de Genômica Funcional da ESALQ: Luiz Lehmann Coutinho; Dimas Tadeu Covas; Elaine Cristina Marqueze; Elaine Vieira Santos; Elaine Vieira dos Santos; Elisangela Chicaroni Mattos; Erika Freitas; Evandra Strazza Rodrigues; Felipe Allan da Silva da Costa; Flavia Aburjaile; Guilherme Targino Valente; Heidge Fukumasu; Heidge Fukumasu. USP-Botucatu: Rejane Maria Tommasini Grotto; Instituto Butantan: Alexander Roberto Precioso; Jayme A. Souza-Neto; Jayme Augusto de Souza-Neto; Jessika Cristina Chagas Lesbon; José Salvatore Leister Patané; João Paulo Kitajima; Luiz Alcantara; Luiz Carlos Junior de Alcantara; Luiz Lehmann Coutinho; Maria Carolina Elias; Marta Giovanetti; Maurício Lacerda Nogueira; Patricia Akemi Assato; Rafael dos Santos Bezerra; Raquel de Lello Rocha Campos Cassano. NGS Soluções Genômicas: Pilar Drummond Sampaio Corrêa Mariani. FZEA-USP Pirassununga: Mirele Daiana Poleti; Raul Machado Neto; Rejane Maria Tommasini Grotto; Ricardo Augusto Brassaloti; Ricardo Haddad; Rodrigo Tocantins Calado.; Sandra Coccuzzo Sampaio; Sandra Coccuzzo Sampaio Vessoni; Simone Kashima; Svetoslav Nanev Slavov; Vagner Fonseca; Vincent Louis Viala |
| EPI_ISL_1967297 | AMBULATORIO DE ESPECIALIDADE V E MOGI MIRIM | Instituto Butantan / Mendelics | Antonio Jorge Martins; Bianca Cechetto Carlos. Mendelics: Bibiana Santos; Claudia Renata dos Santos Barros; Cíntia Bittar; David Schlesinger. Hemocentro Ribeirão Preto: Simone Kashima; Debora Botequio Moretti; Elaine Cristina Marqueze; Elaine Vieira dos Santos; Elisangela Chicaroni Mattos; Erika Freitas; Evandra Strazza Rodrigues; Felipe Allan da Silva da Costa; Flavia Aburjaile; Fábio Sossai Possebon; Guilherme Campos; Guilherme Targino Valente; Heidge Fukumasu. USP-Botucatu: Rejane Maria Tommasini Grotto; Helena Lage Ferreira; Instituto Butantan: Dimas Tadeu Covas; Jardelina de Souza Todao Bernardino; Jayme A. Souza-Neto; Jessika Cristina Chagas Lesbon; Jorge A. Petroli Marchesi; José Salvatore Leister Patané; João Paulo Kitajima; João Pessoa Araújo Jr.; Leila Sabrina Ullmann; Loyze Paola Oliveira de Lima; Luiz Aurelio de Campos Crispin. Centro de Genômica Funcional da ESALQ: Luiz Lehmann Coutinho; Luiz Carlos Junior de Alcantara; Lívia Sacchetto; Maisa C. Pereira Parra; Maria Carolina Elias; Marta Giovanetti; Marília Moraes; Maurício Lacerda Nogueira. Prefeitura de Sao Paulo: Melissa Palmieri.; Patricia Akemi Assato; Paula Rahal; Paulo Inacio da Costa; Rafael dos Santos Bezerra; Raquel de Lello Rocha Campos Cassano. NGS Soluções Genômicas: Pilar Drummond Sampaio Corrêa Mariani. FZEA-USP Pirassununga: Mirele Daiana Poleti; Raul Machado Neto; Ricardo Augusto Brassaloti; Ricardo Haddad; Rodrigo Tocantins Calado. FAMERP-SJRP: Cecília Artico Banho; Sandra Coccuzzo Sampaio; Svetoslav Nanev Slavov; Vagner Fonseca; Vincent Louis Viala |
| EPI_ISL_1967028 | AMBULATORIO DE MOLESTIAS INFECCIOSAS DE SALTO | Instituto Butantan / ESALQ-USP (Piracicaba) | Antonio Jorge Martins; Bianca Cechetto Carlos. Mendelics: Bibiana Santos; Claudia Renata dos Santos Barros; Cíntia Bittar; David Schlesinger. Hemocentro Ribeirão Preto: Simone Kashima; Debora Botequio Moretti; Elaine Cristina Marqueze; Elaine Vieira dos Santos; Elisangela Chicaroni Mattos; Erika Freitas; Evandra Strazza Rodrigues; Felipe Allan da Silva da Costa; Flavia Aburjaile; Fábio Sossai Possebon; Guilherme Campos; Guilherme Targino Valente; Heidge Fukumasu. USP-Botucatu: Rejane Maria Tommasini Grotto; Helena Lage Ferreira; Instituto Butantan: Dimas Tadeu Covas; Jardelina de Souza Todao Bernardino; Jayme A. Souza-Neto; Jessika Cristina Chagas Lesbon; Jorge A. Petroli Marchesi; José Salvatore Leister Patané; João Paulo Kitajima; João Pessoa Araújo Jr.; Leila Sabrina Ullmann; Loyze Paola Oliveira de Lima; Luiz Aurelio de Campos Crispin. Centro de Genômica Funcional da ESALQ: Luiz Lehmann Coutinho; Luiz Carlos Junior de Alcantara; Lívia Sacchetto; Maisa C. Pereira Parra; Maria Carolina Elias; Marta Giovanetti; Marília Moraes; Maurício Lacerda Nogueira. Prefeitura de Sao Paulo: Melissa Palmieri.; Patricia Akemi Assato; Paula Rahal; Paulo Inacio da Costa; Rafael dos Santos Bezerra; Raquel de Lello Rocha Campos Cassano. NGS Soluções Genômicas: Pilar Drummond Sampaio Corrêa Mariani. FZEA-USP Pirassununga: Mirele Daiana Poleti; Raul Machado Neto; Ricardo Augusto Brassaloti; Ricardo Haddad; Rodrigo Tocantins Calado. FAMERP-SJRP: Cecília Artico Banho; Sandra Coccuzzo Sampaio; Svetoslav Nanev Slavov; Vagner Fonseca; Vincent Louis Viala |
| EPI_ISL_1966095 | AMBULATORIO MEDICO DE ESPECIALIDADES DE PERUIBE | Instituto Butantan / Mendelics | Antonio Jorge Martins; Bianca Cechetto Carlos. Mendelics: Bibiana Santos; Claudia Renata dos Santos Barros; Cíntia Bittar; David Schlesinger. Hemocentro Ribeirão Preto: Simone Kashima; Debora Botequio Moretti; Elaine Cristina Marqueze; Elaine Vieira dos Santos; Elisangela Chicaroni Mattos; Erika Freitas; Evandra Strazza Rodrigues; Felipe Allan da Silva da Costa; Flavia Aburjaile; Fábio Sossai Possebon; Guilherme Campos; Guilherme Targino Valente; Heidge Fukumasu. USP-Botucatu: Rejane Maria Tommasini Grotto; Helena Lage Ferreira; Instituto Butantan: Dimas Tadeu Covas; Jardelina de Souza Todao Bernardino; Jayme A. Souza-Neto; Jessika Cristina Chagas Lesbon; Jorge A. Petroli Marchesi; José Salvatore Leister Patané; João Paulo Kitajima; João Pessoa Araújo Jr.; Leila Sabrina Ullmann; Loyze Paola Oliveira de Lima; Luiz Aurelio de Campos Crispin. Centro de Genômica Funcional da ESALQ: Luiz Lehmann Coutinho; Luiz Carlos Junior de Alcantara; Lívia Sacchetto; Maisa C. Pereira Parra; Maria Carolina Elias; Marta Giovanetti; Marília Moraes; Maurício Lacerda Nogueira. Prefeitura de Sao Paulo: Melissa Palmieri.; Patricia Akemi Assato; Paula Rahal; Paulo Inacio da Costa; Rafael dos Santos Bezerra; Raquel de Lello Rocha Campos Cassano. NGS Soluções Genômicas: Pilar Drummond Sampaio Corrêa Mariani. FZEA-USP Pirassununga: Mirele Daiana Poleti; Raul Machado Neto; Ricardo Augusto Brassaloti; Ricardo Haddad; Rodrigo Tocantins Calado. FAMERP-SJRP: Cecília Artico Banho; Sandra Coccuzzo Sampaio; Svetoslav Nanev Slavov; Vagner Fonseca; Vincent Louis Viala |
| EPI_ISL_1795367, EPI_ISL_2345940 | AMBULATORIO MEDICO MUNICIPAL DE AGUDOS | Instituto Butantan / ESALQ-Piracicaba | Antonio Jorge Martins; Bianca Cechetto Carlos. Mendelics: Bibiana Santos; Claudia Renata dos Santos Barros; David Schlesinger; David Schlesinger. Hemocentro Ribeirão Preto: Simone Kashima; Debora Botequio Moretti; Debora Botequio Moretti. Centro de Genômica Funcional da ESALQ: Luiz Lehmann Coutinho; Dimas Tadeu Covas; Elaine Cristina Marqueze; Elaine Vieira Santos; Elaine Vieira dos Santos; Elisangela Chicaroni Mattos; Erika Freitas; Evandra Strazza Rodrigues; Felipe Allan da Silva da Costa; Flavia Aburjaile; Guilherme Targino Valente; Heidge Fukumasu; Heidge Fukumasu. USP-Botucatu: Rejane Maria Tommasini Grotto; Instituto Butantan: Alexander Roberto Precioso; Jayme A. Souza-Neto; Jayme Augusto de Souza-Neto; Jessika Cristina Chagas Lesbon; José Salvatore Leister Patané; João Paulo Kitajima; Luiz Alcantara; Luiz Carlos Junior de Alcantara; Luiz Lehmann Coutinho; Maria Carolina Elias; Marta Giovanetti; Maurício Lacerda Nogueira; Patricia Akemi Assato; Rafael dos Santos Bezerra; Raquel de Lello Rocha Campos Cassano. NGS Soluções Genômicas: Pilar Drummond Sampaio Corrêa Mariani. FZEA-USP Pirassununga: Mirele Daiana Poleti; Raul Machado Neto; Rejane Maria Tommasini Grotto; Ricardo Augusto Brassaloti; Ricardo Haddad; Rodrigo Tocantins Calado.; Sandra Coccuzzo Sampaio; Sandra Coccuzzo Sampaio Vessoni; Simone Kashima; Svetoslav Nanev Slavov; Vagner Fonseca; Vincent Louis Viala |
| EPI_ISL_2018198, EPI_ISL_2018203 | AR Dept. of Health-PHL, Molecular Diagnostics | Centers for Disease Control and Prevention Division of Viral Diseases, Pathogen Discovery | Alison Laufer Halpin; Ben L. Rambo-Martin; Clinton R. Paden; Dakota Howard; Darlene Wagner; Dave Wentworth; Dhwani Batra; Jasmine Padilla; Justin Lee; Katie Dillon; Krista Queen; Kristen Knipe; Kristine Lacek; Mark Burroughs; Matthew Schmerer; Mili Sheth; Peter Cook; Sam Shepard; Sarah Nobles; Shoshona Le; Suxiang Tong; Vivien Dugan; Yvette Unoarumhi |
| EPI_ISL_2272980, EPI_ISL_2272981 | AREA DE SALUD CARRILLO | Inciensa, Instituto Costarricense de Investigación y Enseñanza en Nutrición y Salud | Adriana Godínez; Claudio Soto-Garita; Estela Cordero; Francisco Duarte; Hebleen Porras; Joselyn Prado & Adriana Bemúdez Espinoza; José Luis Vargas; Mariela Gutiérrez; Melany Calderón |
| EPI_ISL_1827536 | AREA DE SALUD COTO BRUS | Inciensa, Instituto Costarricense de Investigación y Enseñanza en Nutrición y Salud | Adriana Godínez; Claudio Soto-Garita; Estela Cordero; Francisco Duarte; Hebleen Porras; Joselyn Prado & Fabricio Aguilar; José Luis Vargas; Mariela Gutiérrez; Melany Calderón |
| EPI_ISL_1827523 | AREA DE SALUD CURRIDABAT 2 | Inciensa, Instituto Costarricense de Investigación y Enseñanza en Nutrición y Salud | Adriana Godínez; Claudio Soto-Garita; Estela Cordero; Francisco Duarte; Hebleen Porras; José Luis Vargas; Mariela Gutiérrez & Joselyn Prado; Melany Calderón |
| EPI_ISL_2272999 | AREA DE SALUD DESAMPARADOS 1 - CLINICA DR. MARCIAL FALLAS | Inciensa, Instituto Costarricense de Investigación y Enseñanza en Nutrición | Adriana Godínez; Claudio Soto-Garita; Estela Cordero; Francisco Duarte; Hebleen Porras; Joselyn Prado & Andony Cordero-Jiménez; José Luis Vargas; Mariela Gutiérrez; Melany Calderón |

| | | | |
|---|---|---|---|
| | | y Salud | |
| EPI_ISL_2103398 | AREA DE SALUD GOICOECHEA 2 - CLINICA DR. JIMENEZ NUÑEZ | Inciensa, Instituto Costarricense de Investigación y Enseñanza en Nutrición y Salud | Adriana Godínez; Caterina Guzmán; Claudio Soto-Garita; Estela Cordero; Francisco Duarte; Hebleen Porras; Joselyn Prado; José Luis Vargas; Mariela Gutiérrez; Melany Calderón; Nazareth Ruiz & Mariel López |
| EPI_ISL_2272960 | AREA DE SALUD LA CRUZ | Inciensa, Instituto Costarricense de Investigación y Enseñanza en Nutrición y Salud | Adriana Godínez; Claudio Soto-Garita; Estela Cordero; Francisco Duarte; Hebleen Porras; Joselyn Prado & Arnoldo Zamora-Villegas; José Luis Vargas; Mariela Gutiérrez; Melany Calderón |
| EPI_ISL_1827525 | AREA DE SALUD LOS CHILES | Inciensa, Instituto Costarricense de Investigación y Enseñanza en Nutrición y Salud | Adriana Godínez; Claudio Soto-Garita; Estela Cordero; Francisco Duarte; Hebleen Porras; Joselyn Prado & Teresita Somogyi; José Luis Vargas; Mariela Gutiérrez; Melany Calderón |
| EPI_ISL_1117051 | ARS Algarve - Laboratorio Laura Ayres | Instituto Nacional de Saude (INSA) | Borges et al |
| EPI_ISL_1811234 | AS Santa Barbara | Inciensa, Instituto Costarricense de Investigación y Enseñanza en Nutrición y Salud | Pérez-Corrales C & Aguilar-Monge R |
| EPI_ISL_1418241 | AX BIO OCEAN | CNR Virus des Infections Respiratoires - France SUD | Antonin Bal; Bruno Lina; Gregory Destras; Gwendolyne Burfin; Hadrien Regue; Laurence Josset; Martine Valette; Quentin Semanas |
| EPI_ISL_1176304, EPI_ISL_1824637 | AZ Klina | AZ Klina | Carl Vael - Lynsey Berckmans; Dr. C. Vael |
| EPI_ISL_1493499, EPI_ISL_1493500, EPI_ISL_1493501, EPI_ISL_1493553, EPI_ISL_1550407, EPI_ISL_1550762, EPI_ISL_1550831, EPI_ISL_1550859, EPI_ISL_1551112, EPI_ISL_1551279, EPI_ISL_1551325, EPI_ISL_1551366, EPI_ISL_1551415, EPI_ISL_1551610, EPI_ISL_1559815, EPI_ISL_1559994, EPI_ISL_1560203, EPI_ISL_1560507, EPI_ISL_1560534, EPI_ISL_1560587, EPI_ISL_1560646, EPI_ISL_1560762, EPI_ISL_1560783, EPI_ISL_1560846, EPI_ISL_1560857, EPI_ISL_1560865, EPI_ISL_1561024, EPI_ISL_1561065, EPI_ISL_1561280, EPI_ISL_1561621, EPI_ISL_1561714, EPI_ISL_1562168, EPI_ISL_1562577, EPI_ISL_1562578, EPI_ISL_1562633, EPI_ISL_1562647, EPI_ISL_1562648, EPI_ISL_1562745, EPI_ISL_1562882, EPI_ISL_1563030, EPI_ISL_1563037, EPI_ISL_1563197, EPI_ISL_1563451, EPI_ISL_1563484, EPI_ISL_1563485, EPI_ISL_1563567, EPI_ISL_1648968, EPI_ISL_1649227, EPI_ISL_1649229, EPI_ISL_1649311, EPI_ISL_1649343, EPI_ISL_1649457, EPI_ISL_1649748, EPI_ISL_1649929, EPI_ISL_1649947, EPI_ISL_1649978, EPI_ISL_1649984, EPI_ISL_1650057, EPI_ISL_1650105, EPI_ISL_1650172, EPI_ISL_1650219, EPI_ISL_1650262, EPI_ISL_1650314, EPI_ISL_1650376, EPI_ISL_1650389, EPI_ISL_1650398, EPI_ISL_1650399, EPI_ISL_1650441, EPI_ISL_1650469, EPI_ISL_1650489, EPI_ISL_1650624, EPI_ISL_1650644, EPI_ISL_1650668, EPI_ISL_1650750, EPI_ISL_1651013, EPI_ISL_1651035, EPI_ISL_1651095, EPI_ISL_1651186, EPI_ISL_1667450, EPI_ISL_1667530, EPI_ISL_1667626, EPI_ISL_1667658, EPI_ISL_1667693, EPI_ISL_1667694, EPI_ISL_1667718, EPI_ISL_1667793, EPI_ISL_1667799, EPI_ISL_1667800, EPI_ISL_1667935, EPI_ISL_1667953, EPI_ISL_1668005, EPI_ISL_1668007, EPI_ISL_1668010, EPI_ISL_1668012, EPI_ISL_1668078, EPI_ISL_1668099, EPI_ISL_1668107, EPI_ISL_1668208, EPI_ISL_1668233, EPI_ISL_1668276, EPI_ISL_1668327, EPI_ISL_1668426, EPI_ISL_1668442, EPI_ISL_1685525, EPI_ISL_1685548, EPI_ISL_1685650, EPI_ISL_1685679, EPI_ISL_1685686, EPI_ISL_1685735, EPI_ISL_1685870, EPI_ISL_1685896, EPI_ISL_1685935, EPI_ISL_1685956, EPI_ISL_1685971, EPI_ISL_1685973, EPI_ISL_1686002, EPI_ISL_1686005, EPI_ISL_1686186, EPI_ISL_1686249, EPI_ISL_1686252, EPI_ISL_1686319, EPI_ISL_1686339, EPI_ISL_1686355, EPI_ISL_1686356, EPI_ISL_1686391, EPI_ISL_1686412, EPI_ISL_1686484, EPI_ISL_1686487, EPI_ISL_1686489, EPI_ISL_1686554, EPI_ISL_1686581, EPI_ISL_1686609, EPI_ISL_1686626, EPI_ISL_1686670, EPI_ISL_1686678, EPI_ISL_1686683, EPI_ISL_1686692, EPI_ISL_1686858, EPI_ISL_1686866, EPI_ISL_1686870, EPI_ISL_1686946, EPI_ISL_1686996, EPI_ISL_1687001, EPI_ISL_1687015, EPI_ISL_1687097, EPI_ISL_1687245, EPI_ISL_1687312, EPI_ISL_1687428, EPI_ISL_1687429, EPI_ISL_1687449, EPI_ISL_1687541, EPI_ISL_1687693, EPI_ISL_1687700, EPI_ISL_1687884, EPI_ISL_1688102, EPI_ISL_1688174, EPI_ISL_1688305, EPI_ISL_1688362, EPI_ISL_1688422, EPI_ISL_1688611, EPI_ISL_1688647, EPI_ISL_1688668, EPI_ISL_1688712, EPI_ISL_1688775, EPI_ISL_1688803, EPI_ISL_1688843, EPI_ISL_1688846, EPI_ISL_1688852, EPI_ISL_1688970, EPI_ISL_1689070, EPI_ISL_1689089, EPI_ISL_1689214, EPI_ISL_1689314, EPI_ISL_1689333, EPI_ISL_1689343, EPI_ISL_1689544, EPI_ISL_1689616, EPI_ISL_1689783, EPI_ISL_1689795, EPI_ISL_1689800, EPI_ISL_1689835, EPI_ISL_1689902, EPI_ISL_1689933, EPI_ISL_1690148, EPI_ISL_1690158, EPI_ISL_1695136, EPI_ISL_1695204, EPI_ISL_1695299, EPI_ISL_1695330, EPI_ISL_1695521, EPI_ISL_1701897, EPI_ISL_1701962, EPI_ISL_1701964, EPI_ISL_1702109, EPI_ISL_1702112, EPI_ISL_1702120, EPI_ISL_1702722, EPI_ISL_1702841, EPI_ISL_1702935, EPI_ISL_1702936, EPI_ISL_1702937, EPI_ISL_1702943, EPI_ISL_1702959, EPI_ISL_1711975, EPI_ISL_1735706, EPI_ISL_1735708, EPI_ISL_1735737, EPI_ISL_1735739, EPI_ISL_1735779, EPI_ISL_1735796, EPI_ISL_1735828, EPI_ISL_1735838, EPI_ISL_1735874, EPI_ISL_1735883, EPI_ISL_1735889, EPI_ISL_1735953, EPI_ISL_1735979, EPI_ISL_1735987, EPI_ISL_1736123, EPI_ISL_1736134, EPI_ISL_1736224, EPI_ISL_1736249, EPI_ISL_1736312, EPI_ISL_1736359, EPI_ISL_1736432, EPI_ISL_1736463, EPI_ISL_1736522, EPI_ISL_1736543, EPI_ISL_1736594, EPI_ISL_1736597, EPI_ISL_1736604, EPI_ISL_1736638, EPI_ISL_1736657, EPI_ISL_1736712, EPI_ISL_1736755, EPI_ISL_1736757, EPI_ISL_1736761, EPI_ISL_1736762, EPI_ISL_1736801, EPI_ISL_1736807, EPI_ISL_1736812, EPI_ISL_1736861, EPI_ISL_1736876, EPI_ISL_1736879, EPI_ISL_1736973, EPI_ISL_1737044, EPI_ISL_1737060, EPI_ISL_1737113, EPI_ISL_1737163, EPI_ISL_1737358, EPI_ISL_1737385, EPI_ISL_1737414, EPI_ISL_1737459, EPI_ISL_1737476, EPI_ISL_1737492, EPI_ISL_1737531, EPI_ISL_1737532, EPI_ISL_1737575, EPI_ISL_1737587, EPI_ISL_1737599, EPI_ISL_1737616, EPI_ISL_1737640, EPI_ISL_1737662, EPI_ISL_1737673, EPI_ISL_1753545, EPI_ISL_1753558, EPI_ISL_1834452, EPI_ISL_1834477, EPI_ISL_1834479, EPI_ISL_1834524, EPI_ISL_1834597, EPI_ISL_1834789, EPI_ISL_1834822, EPI_ISL_1834853, EPI_ISL_1834865, EPI_ISL_1834874, EPI_ISL_1834944, EPI_ISL_1834993, EPI_ISL_1835043, EPI_ISL_1835044, EPI_ISL_1835045, EPI_ISL_1835046, EPI_ISL_1835092, EPI_ISL_1835153, EPI_ISL_1835185, EPI_ISL_1835258, EPI_ISL_1835267, EPI_ISL_1835310, EPI_ISL_1835388, EPI_ISL_1835395, EPI_ISL_1835396, EPI_ISL_1835413, EPI_ISL_1835419, EPI_ISL_1835420, EPI_ISL_1835421, EPI_ISL_1835423, EPI_ISL_1835424, EPI_ISL_1835425, EPI_ISL_1835427, EPI_ISL_1835445, EPI_ISL_1835462, EPI_ISL_1835466, EPI_ISL_1835567, EPI_ISL_1835602, EPI_ISL_1835614, EPI_ISL_1835826, EPI_ISL_1835857, EPI_ISL_1835915, EPI_ISL_1835982, EPI_ISL_1835992, EPI_ISL_1836083, EPI_ISL_1836086, EPI_ISL_1836115, EPI_ISL_1836213, EPI_ISL_1836270, EPI_ISL_1836279, EPI_ISL_1836317, EPI_ISL_1836323, EPI_ISL_1836326, EPI_ISL_1836464, EPI_ISL_1836649, EPI_ISL_1836670, EPI_ISL_1836731, EPI_ISL_1836735, EPI_ISL_1836839, EPI_ISL_1836841, EPI_ISL_1836879, EPI_ISL_1836897, EPI_ISL_1836955, EPI_ISL_1837138, EPI_ISL_1837172, EPI_ISL_1837218, EPI_ISL_1837308, EPI_ISL_1837343, EPI_ISL_1837427, EPI_ISL_1837492, EPI_ISL_1837525, EPI_ISL_1837526, EPI_ISL_1837537, EPI_ISL_1837677, EPI_ISL_1837729, EPI_ISL_1837747, EPI_ISL_1837794, EPI_ISL_1837806, EPI_ISL_1837819, EPI_ISL_1837820, EPI_ISL_1837822, EPI_ISL_1837948, EPI_ISL_1838108, EPI_ISL_1838177, EPI_ISL_1838181, EPI_ISL_1838300, EPI_ISL_1838394, EPI_ISL_1900196, EPI_ISL_1900212, EPI_ISL_1900275, EPI_ISL_1900345, EPI_ISL_1900379, EPI_ISL_1900384, EPI_ISL_1900402, EPI_ISL_1900415, EPI_ISL_1900417, EPI_ISL_1922902, EPI_ISL_1922930, EPI_ISL_1922950, EPI_ISL_1922958, EPI_ISL_1922976, EPI_ISL_1922977, EPI_ISL_1923120, EPI_ISL_1923146, EPI_ISL_1925151, EPI_ISL_1925241, EPI_ISL_1925284, EPI_ISL_1925476, EPI_ISL_1925492, EPI_ISL_1994977, EPI_ISL_1994999, EPI_ISL_1995021, EPI_ISL_1995094, EPI_ISL_1995155, EPI_ISL_1995255, EPI_ISL_1995285, EPI_ISL_1995293, EPI_ISL_1995305, EPI_ISL_1995325, EPI_ISL_1995348, EPI_ISL_1995362, EPI_ISL_1995418, EPI_ISL_1995434, EPI_ISL_1995438, EPI_ISL_1995476, EPI_ISL_1995521, EPI_ISL_1995533, EPI_ISL_1995561, EPI_ISL_1995565, EPI_ISL_1995588, EPI_ISL_1995612, EPI_ISL_1995615, EPI_ISL_1995635, EPI_ISL_1995664, EPI_ISL_1995683, EPI_ISL_1995699, EPI_ISL_1995811, EPI_ISL_1995838, EPI_ISL_1995846, EPI_ISL_1995879, EPI_ISL_1995922, EPI_ISL_1995926, EPI_ISL_1995939, EPI_ISL_1995986, EPI_ISL_1996000, EPI_ISL_1996001, EPI_ISL_1996015, EPI_ISL_1996026, EPI_ISL_1996030, EPI_ISL_1996071, EPI_ISL_1996121, EPI_ISL_1996152, EPI_ISL_1996203, EPI_ISL_1996222, EPI_ISL_1996269, EPI_ISL_1996340, EPI_ISL_1996351, EPI_ISL_1996361, EPI_ISL_1996373, EPI_ISL_1996378, EPI_ISL_1996379, EPI_ISL_1996455, EPI_ISL_1996469, EPI_ISL_1996480, EPI_ISL_1996496, EPI_ISL_1996524, EPI_ISL_1996624, EPI_ISL_1996685, EPI_ISL_1996690, EPI_ISL_1996701, EPI_ISL_1996714, EPI_ISL_1996772, EPI_ISL_1996773, EPI_ISL_1996785, EPI_ISL_1996834, EPI_ISL_1996835, EPI_ISL_1996836, EPI_ISL_1996858, EPI_ISL_1996872, EPI_ISL_1996874, EPI_ISL_1996881, EPI_ISL_1996887, EPI_ISL_1996942, EPI_ISL_1997051, EPI_ISL_1997205, EPI_ISL_1997207, EPI_ISL_1997252, EPI_ISL_1997345, EPI_ISL_1997360, EPI_ISL_1997461, EPI_ISL_1997492, EPI_ISL_1997602, EPI_ISL_1997607, EPI_ISL_1997640, EPI_ISL_1997642, EPI_ISL_1997738, EPI_ISL_1997769, EPI_ISL_1997791, EPI_ISL_1997838, EPI_ISL_1997887, EPI_ISL_1997889, EPI_ISL_1997913, EPI_ISL_1998098, EPI_ISL_1998179, EPI_ISL_1998188, EPI_ISL_1998194, EPI_ISL_1998195, EPI_ISL_1998241, EPI_ISL_1998348, EPI_ISL_1998350, EPI_ISL_1998374, EPI_ISL_1998518, EPI_ISL_1998555, EPI_ISL_1998563, EPI_ISL_1998564, EPI_ISL_1998573, EPI_ISL_1998612, EPI_ISL_1998636, EPI_ISL_1998679, EPI_ISL_1998735, EPI_ISL_1998787, EPI_ISL_1998795, EPI_ISL_1998802, EPI_ISL_1998808, EPI_ISL_1998810, EPI_ISL_1998855, EPI_ISL_1998868, EPI_ISL_1998955, EPI_ISL_1999015, EPI_ISL_1999034, EPI_ISL_1999085, EPI_ISL_1999086, EPI_ISL_1999161, EPI_ISL_1999177, EPI_ISL_1999178, EPI_ISL_1999239, EPI_ISL_1999267, EPI_ISL_1999322, EPI_ISL_1999334, EPI_ISL_1999488, EPI_ISL_1999582, EPI_ISL_1999607, EPI_ISL_1999668, EPI_ISL_1999879, EPI_ISL_1999949, EPI_ISL_2000154, EPI_ISL_2000155, EPI_ISL_2000167, EPI_ISL_2000195, EPI_ISL_2000208, EPI_ISL_2000336, EPI_ISL_2000345, EPI_ISL_2000349, EPI_ISL_2000370, EPI_ISL_2000373, EPI_ISL_2000413, EPI_ISL_2000425, EPI_ISL_2000427, EPI_ISL_2000458, EPI_ISL_2000489, EPI_ISL_2000539, EPI_ISL_2035114, EPI_ISL_2035138, EPI_ISL_2035156, EPI_ISL_2035204, EPI_ISL_2035219, EPI_ISL_2035227, EPI_ISL_2038984, EPI_ISL_2039001, EPI_ISL_2039174, EPI_ISL_2039191, EPI_ISL_2039192, EPI_ISL_2039194, EPI_ISL_2039297, EPI_ISL_2039301, EPI_ISL_2039314, EPI_ISL_2039404, EPI_ISL_2039551, EPI_ISL_2039579, EPI_ISL_2039716, EPI_ISL_2040012, EPI_ISL_2040080, EPI_ISL_2040085, EPI_ISL_2040104, EPI_ISL_2040108, EPI_ISL_2040423, EPI_ISL_2040513, EPI_ISL_2040629, EPI_ISL_2040657, EPI_ISL_2040689, EPI_ISL_2040725, EPI_ISL_2040851, EPI_ISL_2040885, EPI_ISL_2040899, EPI_ISL_2040900, EPI_ISL_2041010, EPI_ISL_2041012, EPI_ISL_2041018, EPI_ISL_2041046, EPI_ISL_2041123, EPI_ISL_2041155, EPI_ISL_2041179, EPI_ISL_2041215, EPI_ISL_2041216, EPI_ISL_2041280, EPI_ISL_2041362, EPI_ISL_2041377, EPI_ISL_2041564, EPI_ISL_2041601, EPI_ISL_2041633, EPI_ISL_2041721, EPI_ISL_2041723, EPI_ISL_2041730, EPI_ISL_2041739, EPI_ISL_2041980, EPI_ISL_2041984, EPI_ISL_2042007, EPI_ISL_2042009, EPI_ISL_2042128, EPI_ISL_2042129, EPI_ISL_2042150, EPI_ISL_2042170, EPI_ISL_2042171, EPI_ISL_2042180, EPI_ISL_2042293, EPI_ISL_2042390, EPI_ISL_2042393, EPI_ISL_2042423, EPI_ISL_2042466, EPI_ISL_2042470, EPI_ISL_2042482, EPI_ISL_2149527, EPI_ISL_2149536, EPI_ISL_2149538, EPI_ISL_2149539, EPI_ISL_2149971, EPI_ISL_2150312, EPI_ISL_2186024, EPI_ISL_2186039, EPI_ISL_2186091, EPI_ISL_2186157, EPI_ISL_2201173, EPI_ISL_2201195, EPI_ISL_2201369, EPI_ISL_2201449, EPI_ISL_2201547, EPI_ISL_2201774, EPI_ISL_2201864, EPI_ISL_2201886, EPI_ISL_2201920, EPI_ISL_2201925, EPI_ISL_2201942, EPI_ISL_2202002, EPI_ISL_2202004, EPI_ISL_2202051, EPI_ISL_2202058, EPI_ISL_2202110, EPI_ISL_2202115, EPI_ISL_2202284, EPI_ISL_2202565, EPI_ISL_2202609, EPI_ISL_2202618, EPI_ISL_2202699, EPI_ISL_2202760, EPI_ISL_2202821, EPI_ISL_2202961, EPI_ISL_2241697, EPI_ISL_2241753, EPI_ISL_2241758, EPI_ISL_2241761, EPI_ISL_2241780, EPI_ISL_2241821, EPI_ISL_2242042, EPI_ISL_2242079, EPI_ISL_2242099, EPI_ISL_2242148, EPI_ISL_2242150, EPI_ISL_2242182, EPI_ISL_2242265, EPI_ISL_2242272, EPI_ISL_2242344, EPI_ISL_2242362, EPI_ISL_2242422, EPI_ISL_2242903, EPI_ISL_2242974, EPI_ISL_2242976, EPI_ISL_2242991, EPI_ISL_2421304, EPI_ISL_2421336, EPI_ISL_2421341, EPI_ISL_2421365, EPI_ISL_2421418, EPI_ISL_2421490, EPI_ISL_2421516, EPI_ISL_2421551, EPI_ISL_2421556, EPI_ISL_2421586, EPI_ISL_2421598, EPI_ISL_2421614, EPI_ISL_2421641, EPI_ISL_2421733, EPI_ISL_2421740, EPI_ISL_2421767, EPI_ISL_2442303, EPI_ISL_2442316, EPI_ISL_2442319, EPI_ISL_2442321, EPI_ISL_2482363, EPI_ISL_2482383, EPI_ISL_2482422, EPI_ISL_2600021, EPI_ISL_2600207, EPI_ISL_2649903, EPI_ISL_2649914 | | | |
| see above | Aegis Sciences Corporation | Centers for Disease Control and Prevention Division of Viral Diseases, Pathogen Discovery | Adrian Paskey; Alec Vest; Benjamin Rambo-Martin; Christopher Gulvick; Clinton R. Paden; Cyndi Clark; Dakota Howard; Darlene Wagner; Dhwani Batra; Dillon Nall; Duncan MacCannell; Ethan Sanders; Holly Houdeshell; Jason Caravas; Kara Moser; Matthew Hardison; Matthew Schmerer; Ola Kvalvaag; Patrick Campbell; Peter W. Cook; Rob Case; Scott Sammons; Shatavia Morrison; Shaun Westlund; Vikramsinha Ghorpade; Yvette Unoarumhi |
| EPI_ISL_1201884, EPI_ISL_1219029, EPI_ISL_1219030, EPI_ISL_1219033, EPI_ISL_1219036 | Aeroporto Internacional de Guarulhos | Instituto Adolfo Lutz, Interdiciplinary Procedures Center, Strategic Laboratory | Caio Vinicius Dias Lopes; Claudia Regina Gonçalves; Claudio Tavares Sacchi; Erica Valessa Ramos Gomes; Karoline Rodrigues Campos |
| EPI_ISL_1739043 | Affidea | Instituto Nacional de Saude (INSA) | Borges et al |
| EPI_ISL_1854251 | Affidea | Instituto Nacional de Saude (INSA) and Institute of Biomedicine (iBiMed), Universidade de Aveiro | Borges et al |
| EPI_ISL_1061282, EPI_ISL_1182734, EPI_ISL_1182768, EPI_ISL_1182769, EPI_ISL_1483589, EPI_ISL_1652201, EPI_ISL_1653895, EPI_ISL_1675103, EPI_ISL_1789630, EPI_ISL_1893796 | | | |
| see above | Alaska State Virology Laboratory | Alaska State Virology Laboratory | Elva House; Jack Chen; Jacob Zidek; Lisa Smith; Ph.D.; Stephanie DeRonde |
| EPI_ISL_1587072 | Altius Institute | Seattle Flu Study | Alex Nguyen; Amanda Adler; Andrew Meuser; Barry R. Lutz; Benjamin Pelle; Caitlin R. Wolf; Chris D. Frazar; Clem Green; Daniel Bates; Deborah A. Nickerson; Elisabeth Brandstetter; Erica Ryke; Hannah Petersen; Helen Y. Chu; Jacob Rodriguez; Janet A. Englund; Jay Shendure; Jessica Halow; John Stamatoyannopoulos; Joshua Richards; Jover Lee; Julia Wald; Kairsten Fay; Kirsten Lacombe; Kneshay Harper; Lea M. Starita; Mark J. Rieder; Matt Hartman; Matthew Richardson; Matthew Thompson; Melissa Truong; Michael Boeckh; Michael Famulare; Misja Ilcisin; Muhammad Halimun; Olivia Waltner; Peter D. Han; Rebecca Bruders; Ryan Alexander; Sadie Patraw; Sofia Olsson; Stephanie DeBaun; Thomas R. Sibley; Tobias Ragoczy; Trevor Bedford; Truong Nguyen |
| EPI_ISL_1625972 | Ama J Angela | Instituto Adolfo Lutz, Interdiciplinary Procedures Center, Strategic Laboratory | Caio Vinicius Dias Lopes; Claudia Regina Gonçalves; Claudio Tavares Sacchi; Erica Valessa Ramos Gomes; Karoline Rodrigues Campos; Katia Correa de Oliveira Santos; Leonardo Jose Tadeu de Araujo |
| EPI_ISL_1966328, EPI_ISL_1966331, EPI_ISL_1966340, EPI_ISL_1966497, EPI_ISL_1966499, EPI_ISL_1966500, EPI_ISL_1966506 | | | |
| see above | Ambulatório IB | Instituto Butantan / Mendelics | Antonio Jorge Martins; Bianca Cechetto Carlos. Mendelics: Bibiana Santos; Claudia Renata dos Santos Barros; Cíntia Bittar; David Schlesinger. Hemocentro Ribeirão Preto: Simone Kashima; Debora Botequio Moretti; Elaine Cristina Marqueze; Elaine Vieira dos Santos; Elisangela Chicaroni Mattos; Erika Freitas; Evandra Strazza Rodrigues; Felipe Allan da Silva da Costa; Flavia Aburjaile; Fábio Sossai Possebon; Guilherme Campos; Guilherme Targino Valente; Heidge Fukumasu. USP-Botucatu: Rejane Maria Tommasini Grotto; Helena Lage Ferreira; Instituto Butantan: Dimas Tadeu Covas; Jardelina de Souza Todao Bernardino; Jayme A. Souza-Neto; Jessika Cristina Chagas Lesbon; Jorge A. Petroli Marchesi; José Salvatore Leister Patané; João Paulo Kitajima; João Pessoa Araújo Jr.; Leila Sabrina Ullmann; Loyze Paola Oliveira de Lima; Luiz Aurelio de Campos Crispin. Centro de Genômica Funcional da ESALQ: Luiz Lehmann Coutinho; Luiz Carlos Junior de Alcantara; Lívia Sacchetto; Maisa C. Pereira Parra; Maria Carolina Elias; Marta Giovanetti; Marília Moraes; Maurício Lacerda Nogueira. Prefeitura de Sao Paulo: Melissa Palmieri.; Patricia Akemi Assato; Paula Rahal; Paulo Inacio da Costa; Rafael dos Santos Bezerra; Raquel de Lello Rocha Campos Cassano. NGS Soluções Genômicas: Pilar Drummond Sampaio Corrêa Mariani. FZEA-USP Pirassununga: Mirele Daiana Poleti; Raul Machado Neto; Ricardo Augusto Brassaloti; Ricardo Haddad; Rodrigo Tocantins Calado. FAMERP-SJRP: Cecília Artico Banho; Sandra Coccuzzo Sampaio; Svetoslav Nanev Slavov; Vagner Fonseca; Vincent Louis Viala |
| EPI_ISL_1574966 | American Indian Health Service of Chicago | Illinois Department of Public Health - Chicago Lab | Ira Heimler; Vineet K. Dhiman |
| EPI_ISL_1121976 | Area of Virology, Serology and Virology Division (SAViD), New South Wales Health Pathology Randwick | Virology Research Laboratory; Area of Virology, Serology and Virology Division (SAViD), New South | Au, J.; Bull, R.; Deveson, I.; Foster, C.; Rawlinson, W.; Ruiz Silva, M.; Van Hal, S. |

| | | | |
|---|---|---|---|
| | | Wales Health Pathology Randwick | |
| EPI_ISL_2263452, EPI_ISL_2263453, EPI_ISL_2499835 | Arizona State Public Health Laboratory | Arizona State Public Health Laboratory | Jessica Escobar; Katherine Fullerton; Linda Getsinger; Nobuko Fukushima; Stacy White; Trung Huynh; Victor Waddell |
| EPI_ISL_1743682, EPI_ISL_1743683, EPI_ISL_1743684, EPI_ISL_1743685, EPI_ISL_1743686, EPI_ISL_1910773, EPI_ISL_1910806, EPI_ISL_1910821, EPI_ISL_2007179, EPI_ISL_2153662, EPI_ISL_2716683 | | | |
| see above | Arizona State University | Arizona State University | Efrem S. Lim; Joshua LaBaer; Joy M. Blain; LaRinda A. Holland; Matthew F. Smith; Nicholas J. Mellor; Peter T. Skidmore; Rabia Maqsood; Valerie Harris; Vel Murugan |
| EPI_ISL_2105735 | Arión Genética | Instituto Nacional de Medicina Genomica | Cedro-Tanda A; Escobar-Arrazola M; Gonzalez-Barrera D; Herrera-Montalvo LA.; Hidalgo-Miranda A; Mendoza-Vargas A; Munguia-Garza P; Ramirez-Vega O; Rangel-DeLeon D; Reyes-Grajeda JP; Roldan-Castillo Magaly; Uribe-Figueroa Laura; Verea Jazmin |
| EPI_ISL_1365747 | Associação Fundo de Incentivo a Pesquisa | Associação Fundo de Incentivo à Pesquisa (AFIP) | Debora Ribeiro Ramadan; Erika Rodrigues de Oliveira; Juliana Nogueira Martins Rodrigues; Priscila Farias Tempaku; Sergio Tufik; Soraya Sgambatti de Andrade |
| EPI_ISL_1499105 | Associação Fundo de Incentivo à Pesquisa (AFIP) | Associação Fundo de Incentivo à Pesquisa (AFIP) | Debora R. Ramadan; Erika Rodrigues de Oliveira; Juliana Nogueira Martins Rodrigues; Priscila Farias Tempaku; Sergio Tufik.; Soraya Sgambatti de Andrade |
| EPI_ISL_1498380 | Associação Fundo de Incentivo à Pesquisa (AFIP). | Associação Fundo de Incentivo à Pesquisa (AFIP) | Debora R. Ramadan; Erika Rodrigues de Oliveira; Juliana Nogueira Martins Rodrigues; Priscila Farias Tempaku; Sergio Tufik.; Soraya Sgambatti de Andrade |
| EPI_ISL_2423529 | Atlanta VA Medical Center | Genomics and Discovery, Respiratory Viruses Branch, Division of Viral Diseases, Centers for Disease Control and Prevention | Adam Retchless; Anna Kelleher; Anna Uehara; Brian Lynch; Clinton R. Paden; Dhwani Batra; Haibin Wang; Han Jia Justin Ng; Jasmine Padilla; Jing Zhang; Justin Lee; Krista Queen; Mark Burroughs; Mili Sheth; Morgan Davis; Peter Cook; Rachel Marine; Sarah Nobles; Suxiang Tong; Tara Coalter; Yan Li; Ying Tao |
| EPI_ISL_1854228, EPI_ISL_1854244, EPI_ISL_1854245, EPI_ISL_1854246, EPI_ISL_1854247, EPI_ISL_1854248 | Avelab | Instituto Nacional de Saude (INSA) and BioSystems & Integrative Sciences Institute (BioISI) Genomics Unit, FCUL | Borges et al |
| EPI_ISL_3341942 | Azienda Ospedaliera Regionale San Carlo Potenza | Laboratorio di patologia clinica e microbiologia A.O.R - San Carlo Potenza | Angela Menchise; Anna Curci; Antonio Picerno; Chiara Autilio; Sara Pascale; Teresa Lopizzo |
| EPI_ISL_1745318, EPI_ISL_1745320, EPI_ISL_1840710, EPI_ISL_1840728, EPI_ISL_1941565, EPI_ISL_1941574, EPI_ISL_2007319 | | | |
| see above | Azienda Sanitaria dell'Alto Adige Laboratorio Aziendale di Microbiologia e Virologia | Istituto di Genomica Applicata | Davide Scaglione; Eleonora Paparelli; Elisa Masi; Elisabetta Giacobazzi; Elisabetta Pagani; Gabriele Magris; Irena Jurman; Irene Bianconi; Michele Morgante; Stefanie Wieser; Vera Vendramin |
| EPI_ISL_2525481, EPI_ISL_2525549, EPI_ISL_2527711, EPI_ISL_2529847, EPI_ISL_2653581, EPI_ISL_2718844 | BCCDC Public Health Laboratory | BCCDC Public Health Laboratory | Ana Pacagnella; Corrinne Ng; Dan Fornika; John Tyson; Kim Macdonald; Kimia Kamelian; Linda Hoang; Loretta Janz; Mel Krajden; Prystajecky Natalie; Robert Azana; Shannon Russell |
| EPI_ISL_1756356 | BIO-REFERENCE LABORATORIES | Wadsworth Center, New York State Department of Health | Alexis Russell; Catharine Prussing; Daryl M. Lamson; Erasmus Schneider; Erica Lasek-Nesselquist; John Kelly; Jonathan Plitnick; Kirsten St. George; Matthew Shudt; Melissa A Leisner; Navjot Singh |
| EPI_ISL_1359892 | BIOLAC - SIEGE | CNR Virus des Infections Respiratoires - France SUD | Antonin Bal; Bruno Lina; Gregory Destras; Gwendolyne Burfin; Hadrien Regue; Laurence Josset; Martine Valette; Quentin Semanas |
| EPI_ISL_1200661, EPI_ISL_1313158, EPI_ISL_1418249, EPI_ISL_1418250 | BIOMNIS LYON | CNR Virus des Infections Respiratoires - France SUD | Antonin Bal; Bruno Lina; Gregory Destras; Gwendolyne Burfin; Hadrien Regue; Laurence Josset; Martine Valette; Quentin Semanas |
| EPI_ISL_1538414 | Basurto University Hospital | Biocruces | Ana de la Hoz; Estibaliz Ugalde Zarraga; José Luís Díaz de Tuesta del Arco; Mikel Gallego Rodrigo; Mikel Urrutikoetxea-Gutierrez; Mª Carmen Nieto Toboso |
| EPI_ISL_1739331 | Basurto University Hospital | Biocruces Bizkaia | Ana de la Hoz; Estibaliz Ugalde Zarraga; Jose Luis Díaz de Tuesta del Arco; Mikel Gallego Rodrigo; Mikel Urrutikoetxea-Gutiérrez; Mº Carmen Nieto Toboso |
| EPI_ISL_1706360 | Basurto University Hospital Clinical Microbiology Lab | Biocruces Bizkaia | Ana de la Hoz; Estibaliz Ugalde Zarraga; José Luis Díaz de Tuesta; Mikel Gallego Rodrigo; Mikel Urrutikoetxea-Gutierrez; Mª Carmen Nieto Toboso |
| EPI_ISL_1706277 | Basurto University Hospital Clinical Microbiology Laboratory | Biocruces Bizkaia | Ana de la Hoz; Estibaliz Ugalde Zarraga; José Luis Díaz de Tuesta del Arco.; Mikel Gallego Rodrigo; Mikel Urrutikoetxea-Gutierrez; Mª Carmen Nieto Toboso |
| EPI_ISL_1538413 | Basurto University Hospital Microbiology Lab | Biocruces | Ana de la Hoz; Estibalez Ugalde Zarrraga; Jose Luis Díaz de Tuesta del Arco; Mikel Gallego Rodrigo; Mikel Urrutikoetxea-Gutierrez; Mª Carmen Nieto Toboso |
| EPI_ISL_1736905, EPI_ISL_1739330 | Basurto University Hospital Microbiology Lab | Biocruces Bizkaia | Ana de la Hoz; Estibaliz Ugalde Zarraga; Jose Luis Díaz de Tuesta del Arco; Jose Luís Díaz de Tuesta del Arco; Mikel Gallego Rodrigo; Mikel Urrutikoetxea-Gutierrez; Mº Carmen Nieto Toboso |
| EPI_ISL_1615556, EPI_ISL_1615557, EPI_ISL_2037414, EPI_ISL_2037443 | Basurto University Hospital: Clinical Microbiology Laboratory | Biocruces Bizkaia | Ana de la Hoz; Estibaliz Ugalde Zarraga; José Luis Díaz de Tuesta del Arco; José Luis Díaz de Tuesta del Arco.; Mikel Gallego Rodrigo; Mikel Urrutikoetxea-Gutierrez; Mikel Urrutikoetxea-Gutiérrez; Mª Carmen Nieto Toboso |
| EPI_ISL_1280655 | Bayerisches Landesamt für Gesundheit und Lebensmittelsicherheit (LGL) | Robert Koch Institute | |
| EPI_ISL_1627453, EPI_ISL_1627454 | Baylor Scott & White-Temple | Baylor Scott & White-Temple | Ari Rao; Caitlin Maloney; Kimberly Walker; Linden Morales; Marcus Volz; Shelby Hendrickson |
| EPI_ISL_1747611 | Bioanalytika AG | Clinical Bacteriology | Adrian Egli; Adrian Härri; Alfredo Mari; Hans Hirsch; Helena MB Seth-Smith; Julia Bielicki; Karoline Leuzinger; Madlen Stange; Manuel Battegay; Tim Roloff |
| EPI_ISL_1756620, EPI_ISL_1757461 | Biogroup Bio Lam-LCD Saint-Denis | Department of Virology, Henri Mondor University Hospital, Assistance Publique Hôpitaux de Paris, Université Paris-Est Créteil, INSERM U955 | Alexandre Soulier; Christophe Rodriguez; Elisabeth Trawinski; Guillaume Gricourt; Jean-Michel Pawlotsky; Melissa N'Debi; Slim Fourati; Vanessa Demontant |
| EPI_ISL_1406151, EPI_ISL_1406179, EPI_ISL_1406183, EPI_ISL_4053500, EPI_ISL_4053502, EPI_ISL_4053505, EPI_ISL_4053520, EPI_ISL_4053522 | | | |
| see above | Biolab Diagnostic Laboratories | Biolab Diagnostic Laboratories | Ahmad Tibi; Amid Abdelnour; Badia Saddedin; Eiad Atwa; Issa Abu-Dayyeh; Lama Hussein; Shaima Ali; Shayma Ali |

| | | | |
|---|---|---|---|
| EPI_ISL_1749270 | Biology Lab, HIA PERCY | IRBA, 2MI | CHAPUS C.; DEPEILLE A.; FOISSAUD V.; GORGE O.; GRANDPERRET V.; JARJAVAL F.; NOLENT F.; SARILAR V.; VERGUET N. |
| EPI_ISL_2698100, EPI_ISL_2698101 | Biology, UFLA, Universidade Federal de Lavras | Biology, UFLA | Barcante, J.; Cherem, J.; Fernandes, G.; Luciano, P.; Melo, D.; Pylro, V. |
| EPI_ISL_2106184 | Biology, UFLA, Universidade Federal de Lavras | Biology, UFLA, Universidade Federal de Lavras | Barcante, J.; Cherem, J.; Fernandes, G.; Luciano, P.; Melo, D.; Pylro, V. |
| EPI_ISL_1917064 | BioneXt Lab | Laboratoire national de sante, Microbiology, Microbial Genomics Platform | Anke Wienecke-Baldacchino; Catherine Ragimbeau; Fatu Djabi; Jessica Tapp; Lise Pignon; Raoul Salmon; Tamir Abdelrahman; Thibault Ferrandon |
| EPI_ISL_3494231, EPI_ISL_3494238, EPI_ISL_3494241 | Boston University CTL | Boston University/National Emerging Infectious Disease Laboratories | Catherine Klapperich; Jacquelyn Turcinovic; John H. Connor; Lena Landeverde; Lynn Doucette-Stamm |
| EPI_ISL_1413799, EPI_ISL_1413803, EPI_ISL_1413805, EPI_ISL_1413813, EPI_ISL_1413823, EPI_ISL_1413829, EPI_ISL_1413837, EPI_ISL_1413840, EPI_ISL_1413842, EPI_ISL_1413845, EPI_ISL_1413851, EPI_ISL_1413856, EPI_ISL_1413859, EPI_ISL_1413861, EPI_ISL_1413864, EPI_ISL_1516491, EPI_ISL_1516492, EPI_ISL_1516498, EPI_ISL_1516700, EPI_ISL_1516701, EPI_ISL_1516720, EPI_ISL_1578342, EPI_ISL_1578346, EPI_ISL_1578347, EPI_ISL_1578348, EPI_ISL_1757215, EPI_ISL_1757235, EPI_ISL_1757243, EPI_ISL_1919016, EPI_ISL_2016413, EPI_ISL_2016415 | | | |
| see above | Broad Institute Clinical Research Sequencing Platform | Infectious Disease Program, Broad Institute of Harvard and MIT | Adams, G.; B.L.; B.W.; Bauer, M.; Birren; Blumenstiel, B.; Brown, C.; Carter, A.; Chaluvadi, S.; D.J.; DeFelice, M.; DeRuff, K.; Dodge, S.; Gabriel, S.; Gallagher, G.; Gladden-Young, A.; Granger, B.; J.E.; K.J.; Lagerborg, K.; Larkin, K.; Lee, M.; Lemieux; Lennon, N.; Loreth, C.; Madoff, L.; McGovern, S.; Meldrim, J.; Normandin, E.; P.C.; Park; Pearlman, L.; Reilly, S.; Rudy, M.; Sabeti; Siddle; Smole, S.; Tomkins-Tinch, C.; Vicente, G.; and MacInnis |
| EPI_ISL_1501135, EPI_ISL_1501265 | Broussais | HEGP - Laboratoire de Virologie | David Veyer |
| EPI_ISL_1701128, EPI_ISL_2103132, EPI_ISL_2103137 | CA DPH Viral and Rickettsial Disease Laboratory | Chan-Zuckerberg Biohub | CZB Cliahub Consortium |
| EPI_ISL_1937829 | CA-Los Angeles County Public Health Laboratory | Centers for Disease Control and Prevention Division of Viral Diseases, Pathogen Discovery | Alison Laufer Halpin; Ben L. Rambo-Martin; Clinton R. Paden; Dakota Howard; Darlene Wagner; Dave Wentworth; Dhwani Batra; Jasmine Padilla; Justin Lee; Katie Dillon; Krista Queen; Kristen Knipe; Kristine Lacek; Mark Burroughs; Matthew Schmerer; Mili Sheth; Peter Cook; Sam Shepard; Sarah Nobles; Shoshona Le; Suxiang Tong; Vivien Dugan; Yvette Unoarumhi |
| EPI_ISL_1761303 | CAP EL REMEI | Banc de Sang i Teixits | Carlos Hobeich; Francisco Vidal; Irene Corrales; Lorena Ramírez; Maria Glòria Soria; Natália Comes; Nina Borràs; Noemí Gonzalez; Silvia Sauleda |
| EPI_ISL_1761184, EPI_ISL_1761216 | CAP SANTA EUGÈNIA DE BERGA | Banc de Sang i Teixits | Carlos Hobeich; Francisco Vidal; Irene Corrales; Lorena Ramírez; Maria Glòria Soria; Natália Comes; Nina Borràs; Noemí Gonzalez; Silvia Sauleda |
| EPI_ISL_2825836, EPI_ISL_2825838, EPI_ISL_2825846, EPI_ISL_2825854, EPI_ISL_2825874, EPI_ISL_2858320, EPI_ISL_2858570, EPI_ISL_2882791, EPI_ISL_2882859, EPI_ISL_2882880, EPI_ISL_2882953, EPI_ISL_2883042, EPI_ISL_2890356, EPI_ISL_2890358, EPI_ISL_2890362, EPI_ISL_2891028, EPI_ISL_2891189 | | | |
| see above | CDPH VBL | California Department of Public Health | CDPH-COVIDNet; UCLA Technology Center for Genomics & Bioinformatics |
| EPI_ISL_1798770, EPI_ISL_2018315 | CDPH, Viral and Rickettsial Disease Laboratory | Centers for Disease Control and Prevention Division of Viral Diseases, Pathogen Discovery | Alison Laufer Halpin; Ben L. Rambo-Martin; Clinton R. Paden; Dakota Howard; Darlene Wagner; Dave Wentworth; Dhwani Batra; Jasmine Padilla; Justin Lee; Katie Dillon; Krista Queen; Kristen Knipe; Kristine Lacek; Mark Burroughs; Matthew Schmerer; Mili Sheth; Peter Cook; Sam Shepard; Sarah Nobles; Shoshona Le; Suxiang Tong; Vivien Dugan; Yvette Unoarumhi |
| EPI_ISL_3102254 | CENTRAL DE TRANSPLANTES DO CEARA | Analytical Competence Molecular Epidemiology Lab/ACME, Oswaldo Cruz Foundation, Ceara (FIOCRUZ CE) | Cleber Furtado Aksenen; Fabio Miyajima; Fernando Braga Stehling; Francisco Eder de Moura Lopes; Jamille Maria Mendes Bezerra; Joaquim César do Nascimento Sousa Junior; Pedro Miguel Carneiro Jeronimo; Suzana Porto Almeida e Lucas Delerino; Thais Ferreira de Oliveira; Thais de Oliveira Costa; Ticiane Cavalcante de Souza; Veridiana Pessoa Miyajima |
| EPI_ISL_1966895, EPI_ISL_1966896, EPI_ISL_1966897, EPI_ISL_1966898, EPI_ISL_1966899, EPI_ISL_1966900, EPI_ISL_1966902, EPI_ISL_1966903, EPI_ISL_1966904 | | | |
| see above | CENTRO DE APOIO EPIDEMIOLOGICO | Instituto Butantan / ESALQ-USP (Piracicaba) | Antonio Jorge Martins; Bianca Cechetto Carlos. Mendelics: Bibiana Santos; Claudia Renata dos Santos Barros; Cíntia Bittar; David Schlesinger. Hemocentro Ribeirão Preto: Simone Kashima; Debora Botequio Moretti; Elaine Cristina Marqueze; Elaine Vieira dos Santos; Elisangela Chicaroni Mattos; Erika Freitas; Evandra Strazza Rodrigues; Felipe Allan da Silva da Costa; Flavia Aburjaile; Fábio Sossai Possebon; Guilherme Campos; Guilherme Targino Valente; Heidge Fukumasu. USP-Botucatu: Rejane Maria Tommasini Grotto; Helena Lage Ferreira; Instituto Butantan: Dimas Tadeu Covas; Jardelina de Souza Todao Bernardino; Jayme A. Souza-Neto; Jessika Cristina Chagas Lesbon; Jorge A. Petroli Marchesi; José Salvatore Leister Patané; João Paulo Kitajima; João Pessoa Araújo Jr.; Leila Sabrina Ullmann; Loyze Paola Oliveira de Lima; Luiz Aurelio de Campos Crispin. Centro de Genômica Funcional da ESALQ: Luiz Lehmann Coutinho; Luiz Carlos Junior de Alcantara; Lívia Sacchetto; Maisa C. Pereira Parra; Maria Carolina Elias; Marta Giovanetti; Marília Moraes; Maurício Lacerda Nogueira. Prefeitura de Sao Paulo: Melissa Palmieri.; Patricia Akemi Assato; Paula Rahal; Paulo Inacio da Costa; Rafael dos Santos Bezerra; Raquel de Lello Rocha Campos Cassano. NGS Soluções Genômicas: Pilar Drummond Sampaio Corrêa Mariani. FZEA-USP Pirassununga: Mirele Daiana Poleti; Raul Machado Neto; Ricardo Augusto Brassaloti; Ricardo Haddad; Rodrigo Tocantins Calado. FAMERP-SJRP: Cecília Artico Banho; Sandra Coccuzzo Sampaio; Svetoslav Nanev Slavov; Vagner Fonseca; Vincent Louis Viala |
| EPI_ISL_3912133 | CENTRO DE ATENDIMENTO COVID 19 TEREZINHA NUNES DA ROCHA | Analytical Competence Molecular Epidemiology Lab/ACME, Oswaldo Cruz Foundation, Ceara (FIOCRUZ CE) | Cleber Furtado Aksenen; Fabio Miyajima; Fernando Braga Stehling; Francisco Eder de Moura Lopes; Jamille Maria Mendes Bezerra; Joaquim Cesar do Nascimento Sousa Junior; Pedro Miguel Carneiro Jeronimo; Suzana Porto Almeida & Lucas Delerino on behalf of COVID-19 FIOCRUZ Genomic Network; Thais Ferreira de Oliveira; Thais de Oliveira Costa; Ticiane Cavalcante de Souza; Veridiana Pessoa Miyajima |
| EPI_ISL_3102322, EPI_ISL_3102538 | CENTRO DE ATENDIMENTO PARA ENFRENTAMENTO AO COVID 19 | Analytical Competence Molecular Epidemiology Lab/ACME, Oswaldo Cruz Foundation, Ceara (FIOCRUZ CE) | Cleber Furtado Aksenen; Fabio Miyajima; Fernando Braga Stehling; Francisco Eder de Moura Lopes; Jamille Maria Mendes Bezerra; Joaquim César do Nascimento Sousa Junior; Pedro Miguel Carneiro Jeronimo; Suzana Porto Almeida e Lucas Delerino; Thais Ferreira de Oliveira; Thais de Oliveira Costa; Ticiane Cavalcante de Souza; Veridiana Pessoa Miyajima |
| EPI_ISL_2344786, EPI_ISL_2344792 | CENTRO DE ESPECIALIDADES DE AGUAI AGUAI | Instituto Butantan / ESALQ-Piracicaba | Antonio Jorge Martins; Claudia Renata dos Santos Barros; David Schlesinger; Debora Botequio Moretti; Dimas Tadeu Covas; Elaine Cristina Marqueze; Elaine Vieira Santos; Evandra Strazza Rodrigues; Heidge Fukumasu; Jayme Augusto de Souza-Neto; José Salvatore Leister Patané; Luiz Alcantara; Luiz Lehmann Coutinho; Maria Carolina Elias; Maurício Lacerda Nogueira; Rafael dos Santos Bezerra; Raul Machado Neto; Rejane Maria Tommasini Grotto; Ricardo Haddad; Sandra Coccuzzo Sampaio Vessoni; Simone Kashima; Svetoslav Nanev Slavov; Vincent Louis Viala |
| EPI_ISL_1795325, EPI_ISL_1795333, EPI_ISL_2345586, EPI_ISL_2345594 | CENTRO DE ESPECIALIDADES DE PRIMAVERA | Instituto Butantan / ESALQ-Piracicaba | Antonio Jorge Martins; Bianca Cechetto Carlos. Mendelics: Bibiana Santos; Claudia Renata dos Santos Barros; David Schlesinger; David Schlesinger. Hemocentro Ribeirão Preto: Simone Kashima; Debora Botequio Moretti; Debora Botequio Moretti. Centro de Genômica Funcional da ESALQ: Luiz Lehmann Coutinho; Dimas Tadeu Covas; Elaine Cristina Marqueze; Elaine Vieira Santos; Elaine Vieira dos Santos; Elisangela Chicaroni Mattos; Erika Freitas; Evandra Strazza Rodrigues; Felipe Allan da Silva da Costa; Flavia Aburjaile; Guilherme Targino Valente; Heidge Fukumasu; Heidge Fukumasu. USP-Botucatu: Rejane Maria Tommasini Grotto; Instituto Butantan: Alexander Roberto Precioso; Jayme A. Souza-Neto; Jayme Augusto de Souza-Neto; Jessika Cristina Chagas Lesbon; José Salvatore Leister Patané; João Paulo Kitajima; Luiz Alcantara; Luiz Carlos Junior de Alcantara; Luiz Lehmann Coutinho; Maria Carolina Elias; Marta Giovanetti; Maurício Lacerda Nogueira; Patricia Akemi Assato; Rafael dos Santos Bezerra; Raquel de Lello Rocha Campos Cassano. NGS Soluções Genômicas: Pilar Drummond Sampaio Corrêa Mariani. FZEA-USP Pirassununga: Mirele Daiana Poleti; Raul Machado Neto; Rejane Maria Tommasini Grotto; Ricardo Augusto Brassaloti; Ricardo Haddad; Rodrigo Tocantins Calado.; Sandra Coccuzzo Sampaio; Sandra Coccuzzo Sampaio Vessoni; Simone Kashima; Svetoslav Nanev Slavov; Vagner Fonseca; Vincent Louis Viala |
| EPI_ISL_3912145 | CENTRO DE ESPECIALIDADES MEDICAS DR NESTOR DE PAULA PESSOA | Analytical Competence Molecular Epidemiology Lab/ACME, Oswaldo Cruz Foundation, Ceara (FIOCRUZ CE) | Cleber Furtado Aksenen; Fabio Miyajima; Fernando Braga Stehling; Francisco Eder de Moura Lopes; Jamille Maria Mendes Bezerra; Joaquim Cesar do Nascimento Sousa Junior; Pedro Miguel Carneiro Jeronimo; Suzana Porto Almeida & Lucas Delerino on behalf of COVID-19 FIOCRUZ Genomic Network; Thais Ferreira de Oliveira; Thais de Oliveira Costa; Ticiane Cavalcante de Souza; Veridiana Pessoa Miyajima |
| EPI_ISL_1795109, EPI_ISL_2344683 | CENTRO DE ESPECIALIDDS MEDICAS IRMA LEOPOLDINA PIRASSUNUNGA | Instituto Butantan / ESALQ-Piracicaba | Antonio Jorge Martins; Bianca Cechetto Carlos. Mendelics: Bibiana Santos; Claudia Renata dos Santos Barros; David Schlesinger; David Schlesinger. Hemocentro Ribeirão Preto: Simone Kashima; Debora Botequio Moretti; Debora Botequio Moretti. Centro de Genômica Funcional da ESALQ: Luiz Lehmann Coutinho; Dimas Tadeu Covas; Elaine Cristina Marqueze; Elaine Vieira Santos; Elaine Vieira dos Santos; Elisangela Chicaroni Mattos; Erika Freitas; Evandra Strazza Rodrigues; Felipe Allan da Silva da Costa; Flavia Aburjaile; Guilherme Targino Valente; Heidge Fukumasu; Heidge Fukumasu. USP-Botucatu: Rejane Maria Tommasini Grotto; Instituto Butantan: Alexander Roberto Precioso; Jayme A. Souza-Neto; Jayme Augusto de Souza-Neto; Jessika Cristina Chagas Lesbon; José Salvatore Leister Patané; João Paulo Kitajima; Luiz Alcantara; Luiz Carlos Junior de Alcantara; Luiz Lehmann Coutinho; Maria Carolina Elias; Marta Giovanetti; Maurício Lacerda Nogueira; Patricia Akemi Assato; Rafael dos Santos Bezerra; Raquel de Lello Rocha Campos Cassano. NGS Soluções Genômicas: Pilar Drummond Sampaio Corrêa Mariani. FZEA-USP Pirassununga: Mirele Daiana Poleti; Raul Machado Neto; Rejane Maria Tommasini Grotto; Ricardo Augusto Brassaloti; Ricardo Haddad; Rodrigo Tocantins Calado.; Sandra Coccuzzo Sampaio; Sandra Coccuzzo Sampaio Vessoni; Simone Kashima; Svetoslav Nanev Slavov; Vagner Fonseca; Vincent Louis Viala |
| EPI_ISL_2344665, EPI_ISL_2344666, EPI_ISL_2344678, EPI_ISL_2344679, EPI_ISL_2344680 | CENTRO DE ESPECIALIDDS MEDICAS IRMA LEOPOLDINA PIRASSUNUNGA | Instituto Butantan / FZEA-USP-Pirassununga | Antonio Jorge Martins; Claudia Renata dos Santos Barros; David Schlesinger; Debora Botequio Moretti; Dimas Tadeu Covas; Elaine Cristina Marqueze; Elaine Vieira Santos; Evandra Strazza Rodrigues; Heidge Fukumasu; Jayme Augusto de Souza-Neto; José Salvatore Leister Patané; Luiz Alcantara; Luiz Lehmann Coutinho; Maria Carolina Elias; Maurício Lacerda Nogueira; Rafael dos Santos Bezerra; Raul Machado Neto; Rejane Maria Tommasini Grotto; Ricardo Haddad; Sandra Coccuzzo Sampaio Vessoni; Simone Kashima; Svetoslav Nanev Slavov; Vincent Louis Viala |
| EPI_ISL_1795228, EPI_ISL_1795229, EPI_ISL_2345459, EPI_ISL_2345460 | CENTRO DE REFERENCIA DO IDOSO DR HUMBERTO MENDES DE CARVALHO | Instituto Butantan / ESALQ-Piracicaba | Antonio Jorge Martins; Bianca Cechetto Carlos. Mendelics: Bibiana Santos; Claudia Renata dos Santos Barros; David Schlesinger; David Schlesinger. Hemocentro Ribeirão Preto: Simone Kashima; Debora Botequio Moretti; Debora Botequio Moretti. Centro de Genômica Funcional da ESALQ: Luiz Lehmann Coutinho; Dimas Tadeu Covas; Elaine Cristina Marqueze; Elaine Vieira Santos; Elaine Vieira dos Santos; Elisangela Chicaroni Mattos; Erika Freitas; Evandra Strazza Rodrigues; Felipe Allan da Silva da Costa; Flavia Aburjaile; Guilherme Targino Valente; Heidge Fukumasu; Heidge Fukumasu. USP-Botucatu: Rejane Maria Tommasini Grotto; Instituto Butantan: Alexander Roberto Precioso; Jayme A. Souza-Neto; Jayme Augusto de Souza-Neto; Jessika Cristina Chagas Lesbon; José Salvatore Leister Patané; João Paulo Kitajima; Luiz Alcantara; Luiz Carlos Junior de Alcantara; Luiz Lehmann Coutinho; Maria Carolina Elias; Marta Giovanetti; Maurício Lacerda Nogueira; Patricia Akemi Assato; Rafael dos Santos Bezerra; Raquel de Lello Rocha Campos Cassano. NGS Soluções Genômicas: Pilar Drummond Sampaio Corrêa Mariani. FZEA-USP Pirassununga: Mirele Daiana Poleti; Raul Machado Neto; Rejane Maria Tommasini Grotto; Ricardo Augusto Brassaloti; Ricardo Haddad; Rodrigo Tocantins Calado.; Sandra Coccuzzo Sampaio; Sandra Coccuzzo Sampaio Vessoni; Simone Kashima; Svetoslav Nanev Slavov; Vagner Fonseca; Vincent Louis Viala |
| EPI_ISL_1966522 | CENTRO DE SAUDE DE ADAMANTINA | Instituto Butantan / FZEA-USP (Pirassununga) | Antonio Jorge Martins; Bianca Cechetto Carlos. Mendelics: Bibiana Santos; Claudia Renata dos Santos Barros; Cíntia Bittar; David Schlesinger. Hemocentro Ribeirão Preto: Simone Kashima; Debora Botequio Moretti; Elaine Cristina Marqueze; Elaine Vieira dos Santos; Elisangela Chicaroni Mattos; Erika Freitas; Evandra Strazza Rodrigues; Felipe Allan da Silva da Costa; Flavia Aburjaile; Fábio Sossai Possebon; Guilherme Campos; Guilherme Targino Valente; Heidge Fukumasu. USP-Botucatu: Rejane Maria Tommasini Grotto; Helena Lage Ferreira; Instituto Butantan: Dimas Tadeu Covas; Jardelina de Souza Todao Bernardino; Jayme A. Souza-Neto; Jessika Cristina Chagas Lesbon; Jorge A. Petroli Marchesi; José Salvatore Leister Patané; João Paulo Kitajima; João Pessoa Araújo Jr.; Leila Sabrina Ullmann; Loyze Paola Oliveira de Lima; Luiz Aurelio de Campos Crispin. Centro de Genômica Funcional da ESALQ: Luiz Lehmann Coutinho; Luiz Carlos Junior de Alcantara; Lívia Sacchetto; Maisa C. Pereira Parra; Maria Carolina Elias; Marta Giovanetti; Marília Moraes; Maurício Lacerda Nogueira. Prefeitura de Sao Paulo: Melissa Palmieri.; Patricia Akemi Assato; Paula Rahal; Paulo Inacio da Costa; Rafael dos Santos Bezerra; Raquel de Lello Rocha Campos Cassano. NGS Soluções Genômicas: Pilar Drummond Sampaio Corrêa Mariani. FZEA-USP Pirassununga: Mirele Daiana Poleti; Raul Machado Neto; Ricardo Augusto Brassaloti; Ricardo Haddad; Rodrigo Tocantins Calado. FAMERP-SJRP: Cecília Artico Banho; Sandra Coccuzzo Sampaio; Svetoslav Nanev Slavov; Vagner Fonseca; Vincent Louis Viala |

| | | | |
|---|---|---|---|
| EPI_ISL_2344774 | CENTRO DE SAUDE DE CHAVANTES | Instituto Butantan / ESALQ-Piracicaba | Antonio Jorge Martins; Claudia Renata dos Santos Barros; David Schlesinger; Debora Botequio Moretti; Dimas Tadeu Covas; Elaine Cristina Marqueze; Elaine Vieira Santos; Evandra Strazza Rodrigues; Heidge Fukumasu; Jayme Augusto de Souza-Neto; José Salvatore Leister Patané; Luiz Alcantara; Luiz Lehmann Coutinho; Maria Carolina Elias; Maurício Lacerda Nogueira; Rafael dos Santos Bezerra; Raul Machado Neto; Rejane Maria Tommasini Grotto; Ricardo Haddad; Sandra Coccuzzo Sampaio Vessoni; Simone Kashima; Svetoslav Nanev Slavov; Vincent Louis Viala |
| EPI_ISL_1795167, EPI_ISL_1795168, EPI_ISL_1795169, EPI_ISL_2345378, EPI_ISL_2345379, EPI_ISL_2345380 | CENTRO DE SAUDE DE FLORINEA | Instituto Butantan / ESALQ-Piracicaba | Antonio Jorge Martins; Bianca Cechetto Carlos. Mendelics: Bibiana Santos; Claudia Renata dos Santos Barros; David Schlesinger; David Schlesinger. Hemocentro Ribeirão Preto: Simone Kashima; Debora Botequio Moretti; Debora Botequio Moretti. Centro de Genômica Funcional da ESALQ: Luiz Lehmann Coutinho; Dimas Tadeu Covas; Elaine Cristina Marqueze; Elaine Vieira Santos; Elaine Vieira dos Santos; Elisangela Chicaroni Mattos; Erika Freitas; Evandra Strazza Rodrigues; Felipe Allan da Silva da Costa; Flavia Aburjaile; Guilherme Targino Valente; Heidge Fukumasu; Heidge Fukumasu. USP-Botucatu: Rejane Maria Tommasini Grotto; Instituto Butantan: Alexander Roberto Precioso; Jayme A. Souza-Neto; Jayme Augusto de Souza-Neto; Jessika Cristina Chagas Lesbon; José Salvatore Leister Patané; João Paulo Kitajima; Luiz Alcantara; Luiz Carlos Junior de Alcantara; Luiz Lehmann Coutinho; Maria Carolina Elias; Marta Giovanetti; Maurício Lacerda Nogueira; Patricia Akemi Assato; Rafael dos Santos Bezerra; Raquel de Lello Rocha Campos Cassano. NGS Soluções Genômicas: Pilar Drummond Sampaio Corrêa Mariani. FZEA-USP Pirassununga: Mirele Daiana Poleti; Raul Machado Neto; Rejane Maria Tommasini Grotto; Ricardo Augusto Brassaloti; Ricardo Haddad; Rodrigo Tocantins Calado.; Sandra Coccuzzo Sampaio; Sandra Coccuzzo Sampaio Vessoni; Simone Kashima; Svetoslav Nanev Slavov; Vagner Fonseca; Vincent Louis Viala |
| EPI_ISL_2209396 | CENTRO DE SAUDE DE IBIRAREMA | Instituto Butantan / ESALQ-Piracicaba | Antonio Jorge Martins; Claudia Renata dos Santos Barros; David Schlesinger; Debora Botequio Moretti; Dimas Tadeu Covas; Elaine Cristina Marqueze; Elaine Vieira Santos; Evandra Strazza Rodrigues; Heidge Fukumasu; Jayme Augusto de Souza-Neto; José Salvatore Leister Patané; Luiz Alcantara; Luiz Lehmann Coutinho; Maria Carolina Elias; Maurício Lacerda Nogueira; Rafael dos Santos Bezerra; Raul Machado Neto; Rejane Maria Tommasini Grotto; Ricardo Haddad; Sandra Coccuzzo Sampaio Vessoni; Simone Kashima; Svetoslav Nanev Slavov; Vincent Louis Viala |
| EPI_ISL_2344787 | CENTRO DE SAUDE DE LUTECIA | Instituto Butantan / ESALQ-Piracicaba | Antonio Jorge Martins; Claudia Renata dos Santos Barros; David Schlesinger; Debora Botequio Moretti; Dimas Tadeu Covas; Elaine Cristina Marqueze; Elaine Vieira Santos; Evandra Strazza Rodrigues; Heidge Fukumasu; Jayme Augusto de Souza-Neto; José Salvatore Leister Patané; Luiz Alcantara; Luiz Lehmann Coutinho; Maria Carolina Elias; Maurício Lacerda Nogueira; Rafael dos Santos Bezerra; Raul Machado Neto; Rejane Maria Tommasini Grotto; Ricardo Haddad; Sandra Coccuzzo Sampaio Vessoni; Simone Kashima; Svetoslav Nanev Slavov; Vincent Louis Viala |
| EPI_ISL_1795170, EPI_ISL_1795172, EPI_ISL_2345381, EPI_ISL_2345387 | CENTRO DE SAUDE DE MARACAI | Instituto Butantan / ESALQ-Piracicaba | Antonio Jorge Martins; Bianca Cechetto Carlos. Mendelics: Bibiana Santos; Claudia Renata dos Santos Barros; David Schlesinger; David Schlesinger. Hemocentro Ribeirão Preto: Simone Kashima; Debora Botequio Moretti; Debora Botequio Moretti. Centro de Genômica Funcional da ESALQ: Luiz Lehmann Coutinho; Dimas Tadeu Covas; Elaine Cristina Marqueze; Elaine Vieira Santos; Elaine Vieira dos Santos; Elisangela Chicaroni Mattos; Erika Freitas; Evandra Strazza Rodrigues; Felipe Allan da Silva da Costa; Flavia Aburjaile; Guilherme Targino Valente; Heidge Fukumasu; Heidge Fukumasu. USP-Botucatu: Rejane Maria Tommasini Grotto; Instituto Butantan: Alexander Roberto Precioso; Jayme A. Souza-Neto; Jayme Augusto de Souza-Neto; Jessika Cristina Chagas Lesbon; José Salvatore Leister Patané; João Paulo Kitajima; Luiz Alcantara; Luiz Carlos Junior de Alcantara; Luiz Lehmann Coutinho; Maria Carolina Elias; Marta Giovanetti; Maurício Lacerda Nogueira; Patricia Akemi Assato; Rafael dos Santos Bezerra; Raquel de Lello Rocha Campos Cassano. NGS Soluções Genômicas: Pilar Drummond Sampaio Corrêa Mariani. FZEA-USP Pirassununga: Mirele Daiana Poleti; Raul Machado Neto; Rejane Maria Tommasini Grotto; Ricardo Augusto Brassaloti; Ricardo Haddad; Rodrigo Tocantins Calado.; Sandra Coccuzzo Sampaio; Sandra Coccuzzo Sampaio Vessoni; Simone Kashima; Svetoslav Nanev Slavov; Vagner Fonseca; Vincent Louis Viala |
| EPI_ISL_1966957, EPI_ISL_1966959, EPI_ISL_1966960 | CENTRO DE SAUDE DE MARACAI | Instituto Butantan / ESALQ-USP (Piracicaba) | Antonio Jorge Martins; Bianca Cechetto Carlos. Mendelics: Bibiana Santos; Claudia Renata dos Santos Barros; Cíntia Bittar; David Schlesinger. Hemocentro Ribeirão Preto: Simone Kashima; Debora Botequio Moretti; Elaine Cristina Marqueze; Elaine Vieira dos Santos; Elisangela Chicaroni Mattos; Erika Freitas; Evandra Strazza Rodrigues; Felipe Allan da Silva da Costa; Flavia Aburjaile; Fábio Sossai Possebon; Guilherme Campos; Guilherme Targino Valente; Heidge Fukumasu. USP-Botucatu: Rejane Maria Tommasini Grotto; Helena Lage Ferreira; Instituto Butantan: Dimas Tadeu Covas; Jardelina de Souza Todao Bernardino; Jayme A. Souza-Neto; Jessika Cristina Chagas Lesbon; Jorge A. Petroli Marchesi; José Salvatore Leister Patané; João Paulo Kitajima; João Pessoa Araújo Jr.; Leila Sabrina Ullmann; Loyze Paola Oliveira de Lima; Luiz Aurelio de Campos Crispin. Centro de Genômica Funcional da ESALQ: Luiz Lehmann Coutinho; Luiz Carlos Junior de Alcantara; Lívia Sacchetto; Maisa C. Pereira Parra; Maria Carolina Elias; Marta Giovanetti; Marília Moraes; Maurício Lacerda Nogueira. Prefeitura de Sao Paulo: Melissa Palmieri.; Patricia Akemi Assato; Paula Rahal; Paulo Inacio da Costa; Rafael dos Santos Bezerra; Raquel de Lello Rocha Campos Cassano. NGS Soluções Genômicas: Pilar Drummond Sampaio Corrêa Mariani. FZEA-USP Pirassununga: Mirele Daiana Poleti; Raul Machado Neto; Ricardo Augusto Brassaloti; Ricardo Haddad; Rodrigo Tocantins Calado. FAMERP-SJRP: Cecília Artico Banho; Sandra Coccuzzo Sampaio; Svetoslav Nanev Slavov; Vagner Fonseca; Vincent Louis Viala |
| EPI_ISL_2344785 | CENTRO DE SAUDE DE OCAUCU | Instituto Butantan / ESALQ-Piracicaba | Antonio Jorge Martins; Claudia Renata dos Santos Barros; David Schlesinger; Debora Botequio Moretti; Dimas Tadeu Covas; Elaine Cristina Marqueze; Elaine Vieira Santos; Evandra Strazza Rodrigues; Heidge Fukumasu; Jayme Augusto de Souza-Neto; José Salvatore Leister Patané; Luiz Alcantara; Luiz Lehmann Coutinho; Maria Carolina Elias; Maurício Lacerda Nogueira; Rafael dos Santos Bezerra; Raul Machado Neto; Rejane Maria Tommasini Grotto; Ricardo Haddad; Sandra Coccuzzo Sampaio Vessoni; Simone Kashima; Svetoslav Nanev Slavov; Vincent Louis Viala |
| EPI_ISL_2344791 | CENTRO DE SAUDE DE SPEDRO DO TURVO | Instituto Butantan / ESALQ-Piracicaba | Antonio Jorge Martins; Claudia Renata dos Santos Barros; David Schlesinger; Debora Botequio Moretti; Dimas Tadeu Covas; Elaine Cristina Marqueze; Elaine Vieira Santos; Evandra Strazza Rodrigues; Heidge Fukumasu; Jayme Augusto de Souza-Neto; José Salvatore Leister Patané; Luiz Alcantara; Luiz Lehmann Coutinho; Maria Carolina Elias; Maurício Lacerda Nogueira; Rafael dos Santos Bezerra; Raul Machado Neto; Rejane Maria Tommasini Grotto; Ricardo Haddad; Sandra Coccuzzo Sampaio Vessoni; Simone Kashima; Svetoslav Nanev Slavov; Vincent Louis Viala |
| EPI_ISL_2344744 | CENTRO DE SAUDE DO INDAIA DO AGUAPEI FLORIDA PAULISTA | Instituto Butantan / ESALQ-Piracicaba | Antonio Jorge Martins; Claudia Renata dos Santos Barros; David Schlesinger; Debora Botequio Moretti; Dimas Tadeu Covas; Elaine Cristina Marqueze; Elaine Vieira Santos; Evandra Strazza Rodrigues; Heidge Fukumasu; Jayme Augusto de Souza-Neto; José Salvatore Leister Patané; Luiz Alcantara; Luiz Lehmann Coutinho; Maria Carolina Elias; Maurício Lacerda Nogueira; Rafael dos Santos Bezerra; Raul Machado Neto; Rejane Maria Tommasini Grotto; Ricardo Haddad; Sandra Coccuzzo Sampaio Vessoni; Simone Kashima; Svetoslav Nanev Slavov; Vincent Louis Viala |
| EPI_ISL_3912146, EPI_ISL_3912147 | CENTRO DE SAUDE DR EDIJANIR GARCIA DA SILVA | Analytical Competence Molecular Epidemiology Lab/ACME, Oswaldo Cruz Foundation, Ceara (FIOCRUZ CE) | Cleber Furtado Aksenen; Fabio Miyajima; Fernando Braga Stehling; Francisco Eder de Moura Lopes; Jamille Maria Mendes Bezerra; Joaquim Cesar do Nascimento Sousa Junior; Pedro Miguel Carneiro Jeronimo; Suzana Porto Almeida & Lucas Delerino on behalf of COVID-19 FIOCRUZ Genomic Network; Thais Ferreira de Oliveira; Thais de Oliveira Costa; Ticiane Cavalcante de Souza; Veridiana Pessoa Miyajima |
| EPI_ISL_3912148 | CENTRO DE SAUDE DR GUARANY MONTALVERNE | Analytical Competence Molecular Epidemiology Lab/ACME, Oswaldo Cruz Foundation, Ceara (FIOCRUZ CE) | Cleber Furtado Aksenen; Fabio Miyajima; Fernando Braga Stehling; Francisco Eder de Moura Lopes; Jamille Maria Mendes Bezerra; Joaquim Cesar do Nascimento Sousa Junior; Pedro Miguel Carneiro Jeronimo; Suzana Porto Almeida & Lucas Delerino on behalf of COVID-19 FIOCRUZ Genomic Network; Thais Ferreira de Oliveira; Thais de Oliveira Costa; Ticiane Cavalcante de Souza; Veridiana Pessoa Miyajima |
| EPI_ISL_2209265, EPI_ISL_2344710 | CENTRO DE SAUDE DR MARIO DIAS DE AGUIAR CAPIVARI | Instituto Butantan / FZEA-USP-Pirassununga | Antonio Jorge Martins; Claudia Renata dos Santos Barros; David Schlesinger; Debora Botequio Moretti; Dimas Tadeu Covas; Elaine Cristina Marqueze; Elaine Vieira Santos; Evandra Strazza Rodrigues; Heidge Fukumasu; Jayme Augusto de Souza-Neto; José Salvatore Leister Patané; Luiz Alcantara; Luiz Lehmann Coutinho; Maria Carolina Elias; Maurício Lacerda Nogueira; Rafael dos Santos Bezerra; Raul Machado Neto; Rejane Maria Tommasini Grotto; Ricardo Haddad; Sandra Coccuzzo Sampaio Vessoni; Simone Kashima; Svetoslav Nanev Slavov; Vincent Louis Viala |
| EPI_ISL_3912149 | CENTRO DE SAUDE DR MIRANDA TAVARES | Analytical Competence Molecular Epidemiology Lab/ACME, Oswaldo Cruz Foundation, Ceara (FIOCRUZ CE) | Cleber Furtado Aksenen; Fabio Miyajima; Fernando Braga Stehling; Francisco Eder de Moura Lopes; Jamille Maria Mendes Bezerra; Joaquim Cesar do Nascimento Sousa Junior; Pedro Miguel Carneiro Jeronimo; Suzana Porto Almeida & Lucas Delerino on behalf of COVID-19 FIOCRUZ Genomic Network; Thais Ferreira de Oliveira; Thais de Oliveira Costa; Ticiane Cavalcante de Souza; Veridiana Pessoa Miyajima |
| EPI_ISL_2344797 | CENTRO DE SAUDE DR NELSON DA CUNHA BASTOS | Instituto Butantan / ESALQ-Piracicaba | Antonio Jorge Martins; Claudia Renata dos Santos Barros; David Schlesinger; Debora Botequio Moretti; Dimas Tadeu Covas; Elaine Cristina Marqueze; Elaine Vieira Santos; Evandra Strazza Rodrigues; Heidge Fukumasu; Jayme Augusto de Souza-Neto; José Salvatore Leister Patané; Luiz Alcantara; Luiz Lehmann Coutinho; Maria Carolina Elias; Maurício Lacerda Nogueira; Rafael dos Santos Bezerra; Raul Machado Neto; Rejane Maria Tommasini Grotto; Ricardo Haddad; Sandra Coccuzzo Sampaio Vessoni; Simone Kashima; Svetoslav Nanev Slavov; Vincent Louis Viala |
| EPI_ISL_2208950 | CENTRO DE SAUDE DR RENATO DE CARVALHO RIBEIRO ANGATUBA | Instituto Butantan | Antonio Jorge Martins; Claudia Renata dos Santos Barros; David Schlesinger; Debora Botequio Moretti; Dimas Tadeu Covas; Elaine Cristina Marqueze; Elaine Vieira Santos; Evandra Strazza Rodrigues; Heidge Fukumasu; Jayme Augusto de Souza-Neto; José Salvatore Leister Patané; Luiz Alcantara; Luiz Lehmann Coutinho; Maria Carolina Elias; Maurício Lacerda Nogueira; Rafael dos Santos Bezerra; Raul Machado Neto; Rejane Maria Tommasini Grotto; Ricardo Haddad; Sandra Coccuzzo Sampaio Vessoni; Simone Kashima; Svetoslav Nanev Slavov; Vincent Louis Viala. |
| EPI_ISL_1966824, EPI_ISL_1966825, EPI_ISL_1966826, EPI_ISL_1966827, EPI_ISL_1966828, EPI_ISL_1966829 | CENTRO DE SAUDE DR RENATO DE CARVALHO RIBEIRO ANGATUBA | Instituto Butantan / ESALQ-USP (Piracicaba) | Antonio Jorge Martins; Bianca Cechetto Carlos. Mendelics: Bibiana Santos; Claudia Renata dos Santos Barros; Cíntia Bittar; David Schlesinger. Hemocentro Ribeirão Preto: Simone Kashima; Debora Botequio Moretti; Elaine Cristina Marqueze; Elaine Vieira dos Santos; Elisangela Chicaroni Mattos; Erika Freitas; Evandra Strazza Rodrigues; Felipe Allan da Silva da Costa; Flavia Aburjaile; Fábio Sossai Possebon; Guilherme Campos; Guilherme Targino Valente; Heidge Fukumasu. USP-Botucatu: Rejane Maria Tommasini Grotto; Helena Lage Ferreira; Instituto Butantan: Dimas Tadeu Covas; Jardelina de Souza Todao Bernardino; Jayme A. Souza-Neto; Jessika Cristina Chagas Lesbon; Jorge A. Petroli Marchesi; José Salvatore Leister Patané; João Paulo Kitajima; João Pessoa Araújo Jr.; Leila Sabrina Ullmann; Loyze Paola Oliveira de Lima; Luiz Aurelio de Campos Crispin. Centro de Genômica Funcional da ESALQ: Luiz Lehmann Coutinho; Luiz Carlos Junior de Alcantara; Lívia Sacchetto; Maisa C. Pereira Parra; Maria Carolina Elias; Marta Giovanetti; Marília Moraes; Maurício Lacerda Nogueira. Prefeitura de Sao Paulo: Melissa Palmieri.; Patricia Akemi Assato; Paula Rahal; Paulo Inacio da Costa; Rafael dos Santos Bezerra; Raquel de Lello Rocha Campos Cassano. NGS Soluções Genômicas: Pilar Drummond Sampaio Corrêa Mariani. FZEA-USP Pirassununga: Mirele Daiana Poleti; Raul Machado Neto; Ricardo Augusto Brassaloti; Ricardo Haddad; Rodrigo Tocantins Calado. FAMERP-SJRP: Cecília Artico Banho; Sandra Coccuzzo Sampaio; Svetoslav Nanev Slavov; Vagner Fonseca; Vincent Louis Viala |
| EPI_ISL_2344587 | CENTRO DE SAUDE DR RENATO DE CARVALHO RIBEIRO ANGATUBA | Instituto Butantan / UNESP-Botucatu | Antonio Jorge Martins; Claudia Renata dos Santos Barros; David Schlesinger; Debora Botequio Moretti; Dimas Tadeu Covas; Elaine Cristina Marqueze; Elaine Vieira Santos; Evandra Strazza Rodrigues; Heidge Fukumasu; Jayme Augusto de Souza-Neto; José Salvatore Leister Patané; Luiz Alcantara; Luiz Lehmann Coutinho; Maria Carolina Elias; Maurício Lacerda Nogueira; Rafael dos Santos Bezerra; Raul Machado Neto; Rejane Maria Tommasini Grotto; Ricardo Haddad; Sandra Coccuzzo Sampaio Vessoni; Simone Kashima; Svetoslav Nanev Slavov; Vincent Louis Viala |
| EPI_ISL_2344779 | CENTRO DE SAUDE ESF IV ZONA RURAL DOMINGOS DE S SJRIOPARDO | Instituto Butantan / ESALQ-Piracicaba | Antonio Jorge Martins; Claudia Renata dos Santos Barros; David Schlesinger; Debora Botequio Moretti; Dimas Tadeu Covas; Elaine Cristina Marqueze; Elaine Vieira Santos; Evandra Strazza Rodrigues; Heidge Fukumasu; Jayme Augusto de Souza-Neto; José Salvatore Leister Patané; Luiz Alcantara; Luiz Lehmann Coutinho; Maria Carolina Elias; Maurício Lacerda Nogueira; Rafael dos Santos Bezerra; Raul Machado Neto; Rejane Maria Tommasini Grotto; Ricardo Haddad; Sandra Coccuzzo Sampaio Vessoni; Simone Kashima; Svetoslav Nanev Slavov; Vincent Louis Viala |
| EPI_ISL_1967059 | CENTRO DE SAUDE ESF IV ZONA RURAL DOMINGOS DE S SJRIOPARDO | Instituto Butantan / ESALQ-USP (Piracicaba) | Antonio Jorge Martins; Bianca Cechetto Carlos. Mendelics: Bibiana Santos; Claudia Renata dos Santos Barros; Cíntia Bittar; David Schlesinger. Hemocentro Ribeirão Preto: Simone Kashima; Debora Botequio Moretti; Elaine Cristina Marqueze; Elaine Vieira dos Santos; Elisangela Chicaroni Mattos; Erika Freitas; Evandra Strazza Rodrigues; Felipe Allan da Silva da Costa; Flavia Aburjaile; Fábio Sossai Possebon; Guilherme Campos; Guilherme Targino Valente; Heidge Fukumasu. USP-Botucatu: Rejane Maria Tommasini Grotto; Helena Lage Ferreira; Instituto Butantan: Dimas Tadeu Covas; Jardelina de Souza Todao Bernardino; Jayme A. Souza-Neto; Jessika Cristina Chagas Lesbon; Jorge A. Petroli Marchesi; José Salvatore Leister Patané; João Paulo Kitajima; João Pessoa Araújo Jr.; Leila Sabrina Ullmann; Loyze Paola Oliveira de Lima; Luiz Aurelio de Campos Crispin. Centro de Genômica Funcional da ESALQ: Luiz Lehmann Coutinho; Luiz Carlos Junior de Alcantara; Lívia Sacchetto; Maisa C. Pereira Parra; Maria Carolina Elias; Marta Giovanetti; Marília Moraes; Maurício Lacerda Nogueira. Prefeitura de Sao Paulo: Melissa Palmieri.; Patricia Akemi Assato; Paula Rahal; Paulo Inacio da Costa; Rafael dos Santos Bezerra; Raquel de Lello Rocha Campos Cassano. NGS Soluções Genômicas: Pilar Drummond Sampaio Corrêa Mariani. FZEA-USP Pirassununga: Mirele Daiana Poleti; Raul Machado Neto; Ricardo Augusto Brassaloti; Ricardo Haddad; Rodrigo Tocantins Calado. FAMERP-SJRP: Cecília Artico Banho; Sandra Coccuzzo Sampaio; Svetoslav Nanev Slavov; Vagner Fonseca; Vincent Louis Viala |
| EPI_ISL_1966536, EPI_ISL_1966538, EPI_ISL_1966539, EPI_ISL_1966540 | CENTRO DE SAUDE I TACITO LEITE DE CARVALHO E SILVA | Instituto Butantan / FZEA-USP (Pirassununga) | Antonio Jorge Martins; Bianca Cechetto Carlos. Mendelics: Bibiana Santos; Claudia Renata dos Santos Barros; Cíntia Bittar; David Schlesinger. Hemocentro Ribeirão Preto: Simone Kashima; Debora Botequio Moretti; Elaine Cristina Marqueze; Elaine Vieira dos Santos; Elisangela Chicaroni Mattos; Erika Freitas; Evandra Strazza Rodrigues; Felipe Allan da Silva da Costa; Flavia Aburjaile; Fábio Sossai Possebon; Guilherme Campos; Guilherme Targino Valente; Heidge Fukumasu. USP-Botucatu: Rejane Maria Tommasini Grotto; Helena Lage Ferreira; Instituto Butantan: Dimas Tadeu Covas; Jardelina de Souza Todao Bernardino; Jayme A. Souza-Neto; Jessika Cristina Chagas Lesbon; Jorge A. Petroli Marchesi; José Salvatore Leister Patané; João Paulo Kitajima; João Pessoa Araújo Jr.; Leila Sabrina Ullmann; Loyze Paola Oliveira de Lima; Luiz Aurelio de Campos Crispin. Centro de Genômica Funcional da ESALQ: Luiz Lehmann Coutinho; Luiz Carlos Junior de Alcantara; Lívia Sacchetto; Maisa C. Pereira Parra; Maria Carolina Elias; Marta Giovanetti; Marília Moraes; Maurício Lacerda Nogueira. Prefeitura de Sao Paulo: Melissa Palmieri.; Patricia Akemi Assato; Paula Rahal; Paulo Inacio da Costa; Rafael dos Santos Bezerra; Raquel de Lello Rocha Campos Cassano. NGS Soluções Genômicas: Pilar Drummond Sampaio Corrêa Mariani. FZEA-USP Pirassununga: Mirele Daiana Poleti; Raul Machado Neto; Ricardo Augusto Brassaloti; Ricardo Haddad; Rodrigo Tocantins Calado. FAMERP-SJRP: Cecília Artico Banho; Sandra Coccuzzo Sampaio; Svetoslav Nanev Slavov; Vagner Fonseca; Vincent Louis Viala |
| EPI_ISL_1795212, EPI_ISL_2345440 | CENTRO DE SAUDE II DR ALCIDES FACUNDO ARROYO | Instituto Butantan / ESALQ-Piracicaba | Antonio Jorge Martins; Bianca Cechetto Carlos. Mendelics: Bibiana Santos; Claudia Renata dos Santos Barros; David Schlesinger; David Schlesinger. Hemocentro Ribeirão Preto: Simone Kashima; Debora Botequio Moretti; Debora Botequio Moretti. Centro de Genômica Funcional da ESALQ: Luiz Lehmann Coutinho; Dimas Tadeu Covas; Elaine Cristina Marqueze; Elaine Vieira Santos; Elaine Vieira dos Santos; Elisangela Chicaroni Mattos; Erika Freitas; Evandra Strazza Rodrigues; Felipe Allan da Silva da Costa; Flavia Aburjaile; Guilherme Targino Valente; Heidge Fukumasu; Heidge Fukumasu. USP-Botucatu: Rejane Maria Tommasini Grotto; Instituto Butantan: Alexander Roberto Precioso; Jayme A. Souza-Neto; Jayme Augusto de Souza-Neto; Jessika Cristina Chagas Lesbon; José Salvatore Leister Patané; João Paulo Kitajima; Luiz Alcantara; Luiz Carlos Junior de Alcantara; Luiz Lehmann Coutinho; Maria Carolina Elias; Marta Giovanetti; Maurício Lacerda Nogueira; Patricia Akemi Assato; Rafael dos Santos Bezerra; Raquel de Lello Rocha Campos Cassano. NGS Soluções Genômicas: Pilar Drummond Sampaio Corrêa Mariani. FZEA-USP Pirassununga: Mirele Daiana Poleti; Raul Machado Neto; Rejane Maria Tommasini Grotto; Ricardo Augusto Brassaloti; Ricardo Haddad; Rodrigo Tocantins Calado.; Sandra Coccuzzo Sampaio; Sandra Coccuzzo Sampaio Vessoni; Simone Kashima; Svetoslav Nanev Slavov; Vagner Fonseca; Vincent Louis Viala |
| EPI_ISL_1795327, EPI_ISL_1795331, EPI_ISL_2345588, EPI_ISL_2345592 | CENTRO DE SAUDE II JUNQUEIROPOLIS | Instituto Butantan / ESALQ-Piracicaba | Antonio Jorge Martins; Bianca Cechetto Carlos. Mendelics: Bibiana Santos; Claudia Renata dos Santos Barros; David Schlesinger; David Schlesinger. Hemocentro Ribeirão Preto: Simone Kashima; Debora Botequio Moretti; Debora Botequio Moretti. Centro de Genômica Funcional da ESALQ: Luiz Lehmann Coutinho; Dimas Tadeu Covas; Elaine Cristina Marqueze; Elaine Vieira Santos; Elaine Vieira dos Santos; Elisangela Chicaroni Mattos; Erika Freitas; Evandra Strazza Rodrigues; Felipe Allan da Silva da Costa; Flavia Aburjaile; Guilherme Targino Valente; Heidge Fukumasu; Heidge Fukumasu. USP-Botucatu: Rejane Maria Tommasini Grotto; Instituto Butantan: Alexander Roberto Precioso; Jayme A. Souza-Neto; Jayme Augusto de Souza-Neto; Jessika Cristina Chagas Lesbon; José Salvatore Leister Patané; João Paulo Kitajima; Luiz Alcantara; Luiz Carlos Junior de Alcantara; Luiz Lehmann Coutinho; Maria Carolina Elias; Marta Giovanetti; Maurício Lacerda Nogueira; Patricia Akemi Assato; Rafael dos Santos Bezerra; Raquel de Lello Rocha Campos Cassano. NGS Soluções Genômicas: Pilar Drummond Sampaio Corrêa Mariani. FZEA-USP Pirassununga: Mirele Daiana Poleti; Raul Machado Neto; Rejane Maria Tommasini Grotto; Ricardo Augusto Brassaloti; Ricardo Haddad; Rodrigo Tocantins Calado.; Sandra Coccuzzo Sampaio; Sandra Coccuzzo Sampaio Vessoni; Simone Kashima; Svetoslav Nanev Slavov; Vagner Fonseca; Vincent Louis Viala |
| EPI_ISL_1966955 | CENTRO DE SAUDE II MAIRINQUE MAIRINQUE | Instituto Butantan / ESALQ-USP (Piracicaba) | Antonio Jorge Martins; Bianca Cechetto Carlos. Mendelics: Bibiana Santos; Claudia Renata dos Santos Barros; Cíntia Bittar; David Schlesinger. Hemocentro Ribeirão Preto: Simone Kashima; Debora Botequio Moretti; Elaine Cristina Marqueze; Elaine Vieira dos Santos; Elisangela Chicaroni Mattos; Erika Freitas; Evandra Strazza Rodrigues; Felipe Allan da Silva da Costa; Flavia Aburjaile; Fábio Sossai Possebon; Guilherme Campos; Guilherme Targino Valente; Heidge Fukumasu. USP-Botucatu: Rejane Maria Tommasini Grotto; Helena Lage Ferreira; Instituto Butantan: Dimas Tadeu Covas; Jardelina de Souza Todao Bernardino; Jayme A. Souza-Neto; Jessika Cristina Chagas Lesbon; Jorge A. Petroli Marchesi; José Salvatore Leister Patané; João Paulo Kitajima; João Pessoa Araújo Jr.; Leila Sabrina Ullmann; Loyze Paola Oliveira de Lima; Luiz Aurelio de Campos Crispin. Centro de Genômica Funcional da ESALQ: Luiz Lehmann Coutinho; Luiz Carlos Junior de Alcantara; Lívia Sacchetto; Maisa C. Pereira Parra; Maria Carolina Elias; Marta Giovanetti; Marília Moraes; Maurício Lacerda Nogueira. Prefeitura de Sao Paulo: Melissa Palmieri.; Patricia Akemi Assato; Paula Rahal; Paulo Inacio da Costa; Rafael dos Santos Bezerra; Raquel de Lello Rocha Campos Cassano. NGS Soluções Genômicas: Pilar Drummond Sampaio Corrêa Mariani. FZEA-USP Pirassununga: Mirele Daiana Poleti; Raul Machado Neto; Ricardo Augusto Brassaloti; Ricardo Haddad; Rodrigo Tocantins Calado. FAMERP-SJRP: Cecília Artico Banho; Sandra Coccuzzo Sampaio; Svetoslav Nanev Slavov; Vagner Fonseca; Vincent Louis Viala |
| EPI_ISL_1967062, EPI_ISL_1967063, EPI_ISL_1967064 | CENTRO DE SAUDE II SAO MIGUEL ARCANJO | Instituto Butantan / ESALQ-USP (Piracicaba) | Antonio Jorge Martins; Bianca Cechetto Carlos. Mendelics: Bibiana Santos; Claudia Renata dos Santos Barros; Cíntia Bittar; David Schlesinger. Hemocentro Ribeirão Preto: Simone Kashima; Debora Botequio Moretti; Elaine Cristina Marqueze; Elaine Vieira dos Santos; Elisangela Chicaroni Mattos; Erika Freitas; Evandra Strazza Rodrigues; Felipe Allan da Silva da Costa; Flavia Aburjaile; Fábio Sossai Possebon; Guilherme Campos; Guilherme Targino Valente; Heidge Fukumasu. USP-Botucatu: Rejane Maria Tommasini Grotto; Helena Lage Ferreira; Instituto Butantan: Dimas Tadeu Covas; Jardelina de Souza Todao Bernardino; Jayme A. Souza-Neto; Jessika Cristina Chagas Lesbon; Jorge A. Petroli Marchesi; José Salvatore Leister Patané; João Paulo Kitajima; João Pessoa Araújo Jr.; Leila Sabrina Ullmann; Loyze Paola Oliveira de Lima; Luiz Aurelio de Campos Crispin. Centro de Genômica Funcional da ESALQ: Luiz Lehmann Coutinho; Luiz Carlos Junior de Alcantara; Lívia Sacchetto; Maisa C. Pereira Parra; Maria Carolina Elias; Marta Giovanetti; Marília Moraes; Maurício Lacerda Nogueira. Prefeitura de Sao Paulo: Melissa Palmieri.; Patricia Akemi Assato; Paula Rahal; Paulo Inacio da Costa; Rafael dos Santos Bezerra; Raquel de Lello Rocha Campos Cassano. NGS Soluções Genômicas: Pilar Drummond Sampaio Corrêa Mariani. FZEA-USP Pirassununga: Mirele Daiana Poleti; Raul Machado Neto; Ricardo Augusto Brassaloti; Ricardo Haddad; Rodrigo Tocantins Calado. FAMERP-SJRP: Cecília Artico Banho; Sandra Coccuzzo Sampaio; Svetoslav Nanev Slavov; Vagner Fonseca; Vincent Louis Viala |

| | | | |
|---|---|---|---|
| EPI_ISL_1795081, EPI_ISL_1795297, EPI_ISL_1795299, EPI_ISL_2345539, EPI_ISL_2345542, EPI_ISL_2345546 | CENTRO DE SAUDE III AFFONSO LUZZI SANTA CRUZ DAS PALMEIRAS | Instituto Butantan / ESALQ-Piracicaba | Antonio Jorge Martins; Bianca Cechetto Carlos. Mendelics: Bibiana Santos; Claudia Renata dos Santos Barros; David Schlesinger; David Schlesinger. Hemocentro Ribeirão Preto: Simone Kashima; Debora Botequio Moretti; Debora Botequio Moretti. Centro de Genômica Funcional da ESALQ: Luiz Lehmann Coutinho; Dimas Tadeu Covas; Elaine Cristina Marqueze; Elaine Vieira Santos; Elaine Vieira dos Santos; Elisangela Chicaroni Mattos; Erika Freitas; Evandra Strazza Rodrigues; Felipe Allan da Silva da Costa; Flavia Aburjaile; Guilherme Targino Valente; Heidge Fukumasu; Heidge Fukumasu. USP-Botucatu: Rejane Maria Tommasini Grotto; Instituto Butantan: Alexander Roberto Precioso; Jayme A. Souza-Neto; Jayme Augusto de Souza-Neto; Jessika Cristina Chagas Lesbon; José Salvatore Leister Patané; João Paulo Kitajima; Luiz Alcantara; Luiz Lehmann Coutinho; Maria Carolina Elias; Marta Giovanetti; Maurício Lacerda Nogueira; Patricia Akemi Assato; Rafael dos Santos Bezerra; Raquel de Lello Rocha Campos Cassano. NGS Soluções Genômicas: Pilar Drummond Sampaio Corrêa Mariani. FZEA-USP Pirassununga: Mirele Daiana Poleti; Raul Machado Neto; Rejane Maria Tommasini Grotto; Ricardo Augusto Brassaloti; Ricardo Haddad; Rodrigo Tocantins Calado.; Sandra Coccuzzo Sampaio; Sandra Coccuzzo Sampaio Vessoni; Simone Kashima; Svetoslav Nanev Slavov; Vagner Fonseca; Vincent Louis Viala |
| EPI_ISL_2344567 | CENTRO DE SAUDE III AFFONSO LUZZI SANTA CRUZ DAS PALMEIRAS | Instituto Butantan / FZEA-USP-Pirassununga | Antonio Jorge Martins; Claudia Renata dos Santos Barros; David Schlesinger; Debora Botequio Moretti; Dimas Tadeu Covas; Elaine Cristina Marqueze; Elaine Vieira Santos; Evandra Strazza Rodrigues; Heidge Fukumasu; Jayme Augusto de Souza-Neto; José Salvatore Leister Patané; Luiz Alcantara; Luiz Lehmann Coutinho; Maria Carolina Elias; Maurício Lacerda Nogueira; Rafael dos Santos Bezerra; Raul Machado Neto; Rejane Maria Tommasini Grotto; Ricardo Haddad; Sandra Coccuzzo Sampaio Vessoni; Simone Kashima; Svetoslav Nanev Slavov; Vincent Louis Viala |
| EPI_ISL_1795114, EPI_ISL_1795115, EPI_ISL_2345313, EPI_ISL_2345314 | CENTRO DE SAUDE III BORBOREMA | Instituto Butantan / ESALQ-Piracicaba | Antonio Jorge Martins; Bianca Cechetto Carlos. Mendelics: Bibiana Santos; Claudia Renata dos Santos Barros; David Schlesinger; David Schlesinger. Hemocentro Ribeirão Preto: Simone Kashima; Debora Botequio Moretti; Debora Botequio Moretti. Centro de Genômica Funcional da ESALQ: Luiz Lehmann Coutinho; Dimas Tadeu Covas; Elaine Cristina Marqueze; Elaine Vieira Santos; Elaine Vieira dos Santos; Elisangela Chicaroni Mattos; Erika Freitas; Evandra Strazza Rodrigues; Felipe Allan da Silva da Costa; Flavia Aburjaile; Guilherme Targino Valente; Heidge Fukumasu; Heidge Fukumasu. USP-Botucatu: Rejane Maria Tommasini Grotto; Instituto Butantan: Alexander Roberto Precioso; Jayme A. Souza-Neto; Jayme Augusto de Souza-Neto; Jessika Cristina Chagas Lesbon; José Salvatore Leister Patané; João Paulo Kitajima; Luiz Alcantara; Luiz Carlos Junior de Alcantara; Luiz Lehmann Coutinho; Maria Carolina Elias; Marta Giovanetti; Maurício Lacerda Nogueira; Patricia Akemi Assato; Rafael dos Santos Bezerra; Raquel de Lello Rocha Campos Cassano. NGS Soluções Genômicas: Pilar Drummond Sampaio Corrêa Mariani. FZEA-USP Pirassununga: Mirele Daiana Poleti; Raul Machado Neto; Rejane Maria Tommasini Grotto; Ricardo Augusto Brassaloti; Ricardo Haddad; Rodrigo Tocantins Calado.; Sandra Coccuzzo Sampaio; Sandra Coccuzzo Sampaio Vessoni; Simone Kashima; Svetoslav Nanev Slavov; Vagner Fonseca; Vincent Louis Viala |
| EPI_ISL_1795125, EPI_ISL_1795127, EPI_ISL_2345327, EPI_ISL_2345329 | CENTRO DE SAUDE III CANDIDO RODRIGUES | Instituto Butantan / ESALQ-Piracicaba | Antonio Jorge Martins; Bianca Cechetto Carlos. Mendelics: Bibiana Santos; Claudia Renata dos Santos Barros; David Schlesinger; David Schlesinger. Hemocentro Ribeirão Preto: Simone Kashima; Debora Botequio Moretti; Debora Botequio Moretti. Centro de Genômica Funcional da ESALQ: Luiz Lehmann Coutinho; Dimas Tadeu Covas; Elaine Cristina Marqueze; Elaine Vieira Santos; Elaine Vieira dos Santos; Elisangela Chicaroni Mattos; Erika Freitas; Evandra Strazza Rodrigues; Felipe Allan da Silva da Costa; Flavia Aburjaile; Guilherme Targino Valente; Heidge Fukumasu; Heidge Fukumasu. USP-Botucatu: Rejane Maria Tommasini Grotto; Instituto Butantan: Alexander Roberto Precioso; Jayme A. Souza-Neto; Jayme Augusto de Souza-Neto; Jessika Cristina Chagas Lesbon; José Salvatore Leister Patané; João Paulo Kitajima; Luiz Alcantara; Luiz Carlos Junior de Alcantara; Luiz Lehmann Coutinho; Maria Carolina Elias; Marta Giovanetti; Maurício Lacerda Nogueira; Patricia Akemi Assato; Rafael dos Santos Bezerra; Raquel de Lello Rocha Campos Cassano. NGS Soluções Genômicas: Pilar Drummond Sampaio Corrêa Mariani. FZEA-USP Pirassununga: Mirele Daiana Poleti; Raul Machado Neto; Rejane Maria Tommasini Grotto; Ricardo Augusto Brassaloti; Ricardo Haddad; Rodrigo Tocantins Calado.; Sandra Coccuzzo Sampaio; Sandra Coccuzzo Sampaio Vessoni; Simone Kashima; Svetoslav Nanev Slavov; Vagner Fonseca; Vincent Louis Viala |
| EPI_ISL_2344689 | CENTRO DE SAUDE III DE AGUAS DE SAO PEDRO | Instituto Butantan / FZEA-USP-Pirassununga | Antonio Jorge Martins; Claudia Renata dos Santos Barros; David Schlesinger; Debora Botequio Moretti; Dimas Tadeu Covas; Elaine Cristina Marqueze; Elaine Vieira Santos; Evandra Strazza Rodrigues; Heidge Fukumasu; Jayme Augusto de Souza-Neto; José Salvatore Leister Patané; Luiz Alcantara; Luiz Lehmann Coutinho; Maria Carolina Elias; Maurício Lacerda Nogueira; Rafael dos Santos Bezerra; Raul Machado Neto; Rejane Maria Tommasini Grotto; Ricardo Haddad; Sandra Coccuzzo Sampaio Vessoni; Simone Kashima; Svetoslav Nanev Slavov; Vincent Louis Viala |
| EPI_ISL_1795126, EPI_ISL_2345328 | CENTRO DE SAUDE III SALES OLIVEIRA | Instituto Butantan / ESALQ-Piracicaba | Antonio Jorge Martins; Bianca Cechetto Carlos. Mendelics: Bibiana Santos; Claudia Renata dos Santos Barros; David Schlesinger; David Schlesinger. Hemocentro Ribeirão Preto: Simone Kashima; Debora Botequio Moretti; Debora Botequio Moretti. Centro de Genômica Funcional da ESALQ: Luiz Lehmann Coutinho; Dimas Tadeu Covas; Elaine Cristina Marqueze; Elaine Vieira Santos; Elaine Vieira dos Santos; Elisangela Chicaroni Mattos; Erika Freitas; Evandra Strazza Rodrigues; Felipe Allan da Silva da Costa; Flavia Aburjaile; Guilherme Targino Valente; Heidge Fukumasu; Heidge Fukumasu. USP-Botucatu: Rejane Maria Tommasini Grotto; Instituto Butantan: Alexander Roberto Precioso; Jayme A. Souza-Neto; Jayme Augusto de Souza-Neto; Jessika Cristina Chagas Lesbon; José Salvatore Leister Patané; João Paulo Kitajima; Luiz Alcantara; Luiz Carlos Junior de Alcantara; Luiz Lehmann Coutinho; Maria Carolina Elias; Marta Giovanetti; Maurício Lacerda Nogueira; Patricia Akemi Assato; Rafael dos Santos Bezerra; Raquel de Lello Rocha Campos Cassano. NGS Soluções Genômicas: Pilar Drummond Sampaio Corrêa Mariani. FZEA-USP Pirassununga: Mirele Daiana Poleti; Raul Machado Neto; Rejane Maria Tommasini Grotto; Ricardo Augusto Brassaloti; Ricardo Haddad; Rodrigo Tocantins Calado.; Sandra Coccuzzo Sampaio; Sandra Coccuzzo Sampaio Vessoni; Simone Kashima; Svetoslav Nanev Slavov; Vagner Fonseca; Vincent Louis Viala |
| EPI_ISL_1966345 | CENTRO DE SAUDE III TABATINGA | Instituto Butantan / Mendelics | Antonio Jorge Martins; Bianca Cechetto Carlos. Mendelics: Bibiana Santos; Claudia Renata dos Santos Barros; Cíntia Bittar; David Schlesinger. Hemocentro Ribeirão Preto: Simone Kashima; Debora Botequio Moretti; Elaine Cristina Marqueze; Elaine Vieira dos Santos; Elisangela Chicaroni Mattos; Erika Freitas; Evandra Strazza Rodrigues; Felipe Allan da Silva da Costa; Flavia Aburjaile; Fábio Sossai Possebon; Guilherme Campos; Guilherme Targino Valente; Heidge Fukumasu. USP-Botucatu: Rejane Maria Tommasini Grotto; Helena Lage Ferreira; Instituto Butantan: Dimas Tadeu Covas; Jardelina de Souza Todao Bernardino; Jayme A. Souza-Neto; Jessika Cristina Chagas Lesbon; Jorge A. Petroli Marchesi; José Salvatore Leister Patané; João Paulo Kitajima; João Pessoa Araújo Jr.; Leila Sabrina Ullmann; Loyze Paola Oliveira de Lima; Luiz Aurelio de Campos Crispin. Centro de Genômica Funcional da ESALQ: Luiz Lehmann Coutinho; Luiz Carlos Junior de Alcantara; Lívia Sacchetto; Maisa C. Pereira Parra; Maria Carolina Elias; Marta Giovanetti; Marília Moraes; Maurício Lacerda Nogueira. Prefeitura de Sao Paulo: Melissa Palmieri.; Patricia Akemi Assato; Paula Rahal; Paulo Inacio da Costa; Rafael dos Santos Bezerra; Raquel de Lello Rocha Campos Cassano. NGS Soluções Genômicas: Pilar Drummond Sampaio Corrêa Mariani. FZEA-USP Pirassununga: Mirele Daiana Poleti; Raul Machado Neto; Ricardo Augusto Brassaloti; Ricardo Haddad; Rodrigo Tocantins Calado. FAMERP-SJRP: Cecília Artico Banho; Sandra Coccuzzo Sampaio; Svetoslav Nanev Slavov; Vagner Fonseca; Vincent Louis Viala |
| EPI_ISL_2344735 | CENTRO DE SAUDE PE CESAR CORNEO | Instituto Butantan / ESALQ-Piracicaba | Antonio Jorge Martins; Claudia Renata dos Santos Barros; David Schlesinger; Debora Botequio Moretti; Dimas Tadeu Covas; Elaine Cristina Marqueze; Elaine Vieira Santos; Evandra Strazza Rodrigues; Heidge Fukumasu; Jayme Augusto de Souza-Neto; José Salvatore Leister Patané; Luiz Alcantara; Luiz Lehmann Coutinho; Maria Carolina Elias; Maurício Lacerda Nogueira; Rafael dos Santos Bezerra; Raul Machado Neto; Rejane Maria Tommasini Grotto; Ricardo Haddad; Sandra Coccuzzo Sampaio Vessoni; Simone Kashima; Svetoslav Nanev Slavov; Vincent Louis Viala |
| EPI_ISL_1967096, EPI_ISL_1967097 | CENTRO DE SAUDE SAO ROQUE DR JOSE CARVALHO BRITO | Instituto Butantan / ESALQ-USP (Piracicaba) | Antonio Jorge Martins; Bianca Cechetto Carlos. Mendelics: Bibiana Santos; Claudia Renata dos Santos Barros; Cíntia Bittar; David Schlesinger. Hemocentro Ribeirão Preto: Simone Kashima; Debora Botequio Moretti; Elaine Cristina Marqueze; Elaine Vieira dos Santos; Elisangela Chicaroni Mattos; Erika Freitas; Evandra Strazza Rodrigues; Felipe Allan da Silva da Costa; Flavia Aburjaile; Fábio Sossai Possebon; Guilherme Campos; Guilherme Targino Valente; Heidge Fukumasu. USP-Botucatu: Rejane Maria Tommasini Grotto; Helena Lage Ferreira; Instituto Butantan: Dimas Tadeu Covas; Jardelina de Souza Todao Bernardino; Jayme A. Souza-Neto; Jessika Cristina Chagas Lesbon; Jorge A. Petroli Marchesi; José Salvatore Leister Patané; João Paulo Kitajima; João Pessoa Araújo Jr.; Leila Sabrina Ullmann; Loyze Paola Oliveira de Lima; Luiz Aurelio de Campos Crispin. Centro de Genômica Funcional da ESALQ: Luiz Lehmann Coutinho; Luiz Carlos Junior de Alcantara; Lívia Sacchetto; Maisa C. Pereira Parra; Maria Carolina Elias; Marta Giovanetti; Marília Moraes; Maurício Lacerda Nogueira. Prefeitura de Sao Paulo: Melissa Palmieri.; Patricia Akemi Assato; Paula Rahal; Paulo Inacio da Costa; Rafael dos Santos Bezerra; Raquel de Lello Rocha Campos Cassano. NGS Soluções Genômicas: Pilar Drummond Sampaio Corrêa Mariani. FZEA-USP Pirassununga: Mirele Daiana Poleti; Raul Machado Neto; Ricardo Augusto Brassaloti; Ricardo Haddad; Rodrigo Tocantins Calado. FAMERP-SJRP: Cecília Artico Banho; Sandra Coccuzzo Sampaio; Svetoslav Nanev Slavov; Vagner Fonseca; Vincent Louis Viala |
| EPI_ISL_2344749, EPI_ISL_2344752, EPI_ISL_2344757, EPI_ISL_2344759 | CENTRO DE TRIAGEM COVID 19 | Instituto Butantan / ESALQ-Piracicaba | Antonio Jorge Martins; Claudia Renata dos Santos Barros; David Schlesinger; Debora Botequio Moretti; Dimas Tadeu Covas; Elaine Cristina Marqueze; Elaine Vieira Santos; Evandra Strazza Rodrigues; Heidge Fukumasu; Jayme Augusto de Souza-Neto; José Salvatore Leister Patané; Luiz Alcantara; Luiz Lehmann Coutinho; Maria Carolina Elias; Maurício Lacerda Nogueira; Rafael dos Santos Bezerra; Raul Machado Neto; Rejane Maria Tommasini Grotto; Ricardo Haddad; Sandra Coccuzzo Sampaio Vessoni; Simone Kashima; Svetoslav Nanev Slavov; Vincent Louis Viala |
| EPI_ISL_2344690 | CENTRO MEDICO DR NELSON SALOME DE CONCHAL | Instituto Butantan / FZEA-USP-Pirassununga | Antonio Jorge Martins; Claudia Renata dos Santos Barros; David Schlesinger; Debora Botequio Moretti; Dimas Tadeu Covas; Elaine Cristina Marqueze; Elaine Vieira Santos; Evandra Strazza Rodrigues; Heidge Fukumasu; Jayme Augusto de Souza-Neto; José Salvatore Leister Patané; Luiz Alcantara; Luiz Lehmann Coutinho; Maria Carolina Elias; Maurício Lacerda Nogueira; Rafael dos Santos Bezerra; Raul Machado Neto; Rejane Maria Tommasini Grotto; Ricardo Haddad; Sandra Coccuzzo Sampaio Vessoni; Simone Kashima; Svetoslav Nanev Slavov; Vincent Louis Viala |
| EPI_ISL_1795274, EPI_ISL_1795275, EPI_ISL_2345510, EPI_ISL_2345513 | CENTRO MEDICO PMESP | Instituto Butantan / ESALQ-Piracicaba | Antonio Jorge Martins; Bianca Cechetto Carlos. Mendelics: Bibiana Santos; Claudia Renata dos Santos Barros; David Schlesinger; David Schlesinger. Hemocentro Ribeirão Preto: Simone Kashima; Debora Botequio Moretti; Debora Botequio Moretti. Centro de Genômica Funcional da ESALQ: Luiz Lehmann Coutinho; Dimas Tadeu Covas; Elaine Cristina Marqueze; Elaine Vieira Santos; Elaine Vieira dos Santos; Elisangela Chicaroni Mattos; Erika Freitas; Evandra Strazza Rodrigues; Felipe Allan da Silva da Costa; Flavia Aburjaile; Guilherme Targino Valente; Heidge Fukumasu; Heidge Fukumasu. USP-Botucatu: Rejane Maria Tommasini Grotto; Instituto Butantan: Alexander Roberto Precioso; Jayme A. Souza-Neto; Jayme Augusto de Souza-Neto; Jessika Cristina Chagas Lesbon; José Salvatore Leister Patané; João Paulo Kitajima; Luiz Alcantara; Luiz Carlos Junior de Alcantara; Luiz Lehmann Coutinho; Maria Carolina Elias; Marta Giovanetti; Maurício Lacerda Nogueira; Patricia Akemi Assato; Rafael dos Santos Bezerra; Raquel de Lello Rocha Campos Cassano. NGS Soluções Genômicas: Pilar Drummond Sampaio Corrêa Mariani. FZEA-USP Pirassununga: Mirele Daiana Poleti; Raul Machado Neto; Rejane Maria Tommasini Grotto; Ricardo Augusto Brassaloti; Ricardo Haddad; Rodrigo Tocantins Calado.; Sandra Coccuzzo Sampaio; Sandra Coccuzzo Sampaio Vessoni; Simone Kashima; Svetoslav Nanev Slavov; Vagner Fonseca; Vincent Louis Viala |
| EPI_ISL_2209074 | CENTRO MUNICIPAL DE EPIDEMIOLOGIA E IMUNIZACOES LIMEIRA | Instituto Butantan | Antonio Jorge Martins; Claudia Renata dos Santos Barros; David Schlesinger; Debora Botequio Moretti; Dimas Tadeu Covas; Elaine Cristina Marqueze; Elaine Vieira Santos; Evandra Strazza Rodrigues; Heidge Fukumasu; Jayme Augusto de Souza-Neto; José Salvatore Leister Patané; Luiz Alcantara; Luiz Lehmann Coutinho; Maria Carolina Elias; Maurício Lacerda Nogueira; Rafael dos Santos Bezerra; Raul Machado Neto; Rejane Maria Tommasini Grotto; Ricardo Haddad; Sandra Coccuzzo Sampaio Vessoni; Simone Kashima; Svetoslav Nanev Slavov; Vincent Louis Viala. |
| EPI_ISL_2209982 | CENTRO MUNICIPAL DE EPIDEMIOLOGIA E IMUNIZACOES LIMEIRA | Instituto Butantan / FZEA-USP-Pirassununga | Antonio Jorge Martins; Claudia Renata dos Santos Barros; David Schlesinger; Debora Botequio Moretti; Dimas Tadeu Covas; Elaine Cristina Marqueze; Elaine Vieira Santos; Evandra Strazza Rodrigues; Heidge Fukumasu; Jayme Augusto de Souza-Neto; José Salvatore Leister Patané; Luiz Alcantara; Luiz Lehmann Coutinho; Maria Carolina Elias; Maurício Lacerda Nogueira; Rafael dos Santos Bezerra; Raul Machado Neto; Rejane Maria Tommasini Grotto; Ricardo Haddad; Sandra Coccuzzo Sampaio Vessoni; Simone Kashima; Svetoslav Nanev Slavov; Vincent Louis Viala |
| EPI_ISL_1265833 | CERBALLIANCE PACA | CNR Virus des Infections Respiratoires - France SUD | Antonin Bal; Bruno Lina; Gregory Destras; Gwendolyne Burfin; Hadrien Regue; Laurence Josset; Martine Valette; Quentin Semanas |
| EPI_ISL_1526351 | CH ANTIBES - JUAN LES PINS | CNR Virus des Infections Respiratoires - France SUD | Antonin Bal; Bruno Lina; Gregory Destras; Gwendolyne Burfin; Hadrien Regue; Laurence Josset; Martine Valette; Quentin Semanas |
| EPI_ISL_2562029, EPI_ISL_2562030 | CH Franck Joly | Cerba lab | Aude Lessenne; Bénédicte Roquebert; Emmanuel Lecorche; Kader Merah; Laura Verdurme; Patrice Herisson; Sabine Trombert-Paolantoni; Stéphanie Haïm-Boukobza; Thierry Collin |
| EPI_ISL_1399630 | CH Porto - H Sto Antonio | Instituto Nacional de Saude (INSA) and BioSystems & Integrative Sciences Institute (BioISI) Genomics Unit, FCUL | Borges et al |
| EPI_ISL_2226343, EPI_ISL_2226354, EPI_ISL_2226358 | CH SAINTONGE | CHU Poitiers | Agnes BEBY-DEFAUX; Birama N'DIAYE; Caroline MICHAUD; Magali GARCIA; Manon PRAT; Maxime PICHON; Nicolas LEVEQUE; Valentin BON-BARET |
| EPI_ISL_1593466, EPI_ISL_1628339, EPI_ISL_1628340, EPI_ISL_1628342 | CH de l'Ouest Guyanais | Institut Pasteur de la Guyane | A. Salmier; Anne Lavergne; Dominique Rousset |
| EPI_ISL_1085358, EPI_ISL_1085912, EPI_ISL_1672684, EPI_ISL_1672741, EPI_ISL_1706628, EPI_ISL_1706945, EPI_ISL_1706948, EPI_ISL_1757696 | | | |
| see above | CH.INTERCOMMUNAL DE CRETEIL | Department of Virology, Henri Mondor University Hospital, Assistance Publique Hôpitaux de Paris, Université Paris-Est Créteil, INSERM U955 | Alexandre Soulier; Christophe Rodriguez; Elisabeth Trawinski; Guillaume Gricourt; Jean-Michel Pawlotsky; Melissa N'Debi; Slim Fourati; Vanessa Demontant |
| EPI_ISL_1593463, EPI_ISL_1593464, EPI_ISL_1593980, EPI_ISL_1627097, EPI_ISL_1627102, EPI_ISL_1627104, EPI_ISL_1627105, EPI_ISL_1628334, EPI_ISL_1628335, EPI_ISL_1628343, EPI_ISL_2227910 | | | |
| see above | CHC Andrée Rosemon | Institut Pasteur de la Guyane | A. Salmier; Anne Lavergne; Dominique Rousset |
| EPI_ISL_1738809, EPI_ISL_1739049 | CHLC | Instituto Nacional de Saude (INSA) | Borges et al |
| EPI_ISL_1171967 | CHLN | Instituto Nacional de Saude (INSA) | Borges et al |
| EPI_ISL_2194032 | CHU Nantes Virology | CHU Nantes Virology | Audrey Rodallec; Berthe-Marie Imbert-Marcille; Celine Bressollette-Bodin; Thomas Drumel |
| EPI_ISL_1399631, EPI_ISL_1399632 | CHU Sao Joao, Porto | Instituto Nacional de Saude (INSA) and Instituto Gulbenkian de | Borges et al |

| | | | |
|---|---|---|---|
| | | Ciencia (IGC) | |
| EPI_ISL_1854234, EPI_ISL_1854235, EPI_ISL_1854236 | CHULN - H Santa Maria | Instituto Nacional de Saude (INSA) | Borges et al |
| EPI_ISL_1963930, EPI_ISL_1963933, EPI_ISL_1963936 | CLILAB | Microbiology Department | Aida Gonzalez-Diaz; Carmen Ardanuy; Jordi Camara; Jordi Niubó; Laura Calatayud; M Angeles Domínguez; Miguel Fernandez-Huerta; Sara Marti |
| EPI_ISL_3912150 | CLINICA ESCOLA DE SAUDE | Analytical Competence Molecular Epidemiology Lab/ACME, Oswaldo Cruz Foundation, Ceara (FIOCRUZ CE) | Cleber Furtado Aksenen; Fabio Miyajima; Fernando Braga Stehling; Francisco Eder de Moura Lopes; Jamille Maria Mendes Bezerra; Joaquim Cesar do Nascimento Sousa Junior; Pedro Miguel Carneiro Jeronimo; Suzana Porto Almeida & Lucas Delerino on behalf of COVID-19 FIOCRUZ Genomic Network; Thais Ferreira de Oliveira; Thais de Oliveira Costa; Ticiane Cavalcante de Souza; Veridiana Pessoa Miyajima |
| EPI_ISL_1593470, EPI_ISL_1593471, EPI_ISL_1627100, EPI_ISL_1627101, EPI_ISL_1628336, EPI_ISL_1628337, EPI_ISL_1628338, EPI_ISL_2227911, EPI_ISL_2227912, EPI_ISL_2227913, EPI_ISL_2227914, EPI_ISL_2227915, EPI_ISL_2227917, EPI_ISL_2227918 | | | |
| see above | CNR Institut Pasteur de la Guyane | Institut Pasteur de la Guyane | A. Salmier; Anne Lavergne; Dominique Rousset |
| EPI_ISL_1418235, EPI_ISL_1695421 | CNR Virus des Infections Respiratoires - France SUD | CNR Virus des Infections Respiratoires - France SUD | Antonin Bal; Bruno Lina; Gregory Destras; Gwendolyne Burfin; Hadrien Regue; Laurence Josset; Martine Valette; Quentin Semanas |
| EPI_ISL_2383912 | CO Dept. of Public Health and Environment, Lab Services Division | Centers for Disease Control and Prevention Division of Viral Diseases, Pathogen Discovery | Alison Laufer Halpin; Ben L. Rambo-Martin; Clinton R. Paden; Dakota Howard; Darlene Wagner; Dave Wentworth; Dhwani Batra; Jasmine Padilla; Justin Lee; Katie Dillon; Krista Queen; Kristen Knipe; Kristine Lacek; Mark Burroughs; Matthew Schmerer; Mili Sheth; Peter Cook; Sam Shepard; Sarah Nobles; Shoshona Le; Suxiang Tong; Vivien Dugan; Yvette Unoarumhi |
| EPI_ISL_1445109 | COMPLEXO HOSPITALAR OURO VERDE DE CAMPINAS | Instituto Butantan / Mendelics | Antonio Jorge Martins; Bibiana Santos; Claudia Renata dos Santos Barros; David Schlesinger; Debora Botequio Moretti; Dimas Tadeu Covas; Elaine Cristina Marqueze; Elaine Vieira dos Santos; Erika Freitas; Evandra Strazza Rodrigues; Flavia Aburjaile; José Salvatore Leister Patané; João Paulo Kitajima; Luiz Carlos Junior de Alcantara; Maria Carolina Elias; Marta Giovanetti; Rafael dos Santos Bezerra; Raul Machado Neto; Ricardo Haddad; Rodrigo Tocantins Calado.; Sandra Coccuzzo Sampaio; Simone Kashima; Svetoslav Nanev Slavov; Vagner Fonseca; Vincent Louis Viala |
| EPI_ISL_2209080 | COORDENADORIA MUNICIPAL DE SAUDE DE IRACEMAPOLIS | Instituto Butantan | Antonio Jorge Martins; Claudia Renata dos Santos Barros; David Schlesinger; Debora Botequio Moretti; Dimas Tadeu Covas; Elaine Cristina Marqueze; Elaine Vieira Santos; Evandra Strazza Rodrigues; Heidge Fukumasu; Jayme Augusto de Souza-Neto; José Salvatore Leister Patané; Luiz Alcantara; Luiz Lehmann Coutinho; Maria Carolina Elias; Maurício Lacerda Nogueira; Rafael dos Santos Bezerra; Raul Machado Neto; Rejane Maria Tommasini Grotto; Ricardo Haddad; Sandra Coccuzzo Sampaio Vessoni; Simone Kashima; Svetoslav Nanev Slavov; Vincent Louis Viala. |
| EPI_ISL_1795107, EPI_ISL_2344660 | COORDENADORIA MUNICIPAL DE SAUDE DE IRACEMAPOLIS | Instituto Butantan / ESALQ-Piracicaba | Antonio Jorge Martins; Bianca Cechetto Carlos. Mendelics: Bibiana Santos; Claudia Renata dos Santos Barros; David Schlesinger; David Schlesinger. Hemocentro Ribeirão Preto: Simone Kashima; Debora Botequio Moretti; Debora Botequio Moretti. Centro de Genômica Funcional da ESALQ: Luiz Lehmann Coutinho; Dimas Tadeu Covas; Elaine Cristina Marqueze; Elaine Vieira Santos; Elaine Vieira dos Santos; Elisangela Chicaroni Mattos; Erika Freitas; Evandra Strazza Rodrigues; Felipe Allan da Silva da Costa; Flavia Aburjaile; Guilherme Targino Valente; Heidge Fukumasu; Heidge Fukumasu. USP-Botucatu: Rejane Maria Tommasini Grotto; Instituto Butantan: Alexander Roberto Precioso; Jayme A. Souza-Neto; Jayme Augusto de Souza-Neto; Jessika Cristina Chagas Lesbon; José Salvatore Leister Patané; João Paulo Kitajima; Luiz Alcantara; Luiz Carlos Junior de Alcantara; Luiz Lehmann Coutinho; Maria Carolina Elias; Marta Giovanetti; Maurício Lacerda Nogueira; Patricia Akemi Assato; Rafael dos Santos Bezerra; Raquel de Lello Rocha Campos Cassano. NGS Soluções Genômicas: Pilar Drummond Sampaio Corrêa Mariani. FZEA-USP Pirassununga: Mirele Daiana Poleti; Raul Machado Neto; Rejane Maria Tommasini Grotto; Ricardo Augusto Brassaloti; Ricardo Haddad; Rodrigo Tocantins Calado.; Sandra Coccuzzo Sampaio; Sandra Coccuzzo Sampaio Vessoni; Simone Kashima; Svetoslav Nanev Slavov; Vagner Fonseca; Vincent Louis Viala |
| EPI_ISL_2209275, EPI_ISL_2344685, EPI_ISL_2344702 | COORDENADORIA MUNICIPAL DE SAUDE DE IRACEMAPOLIS | Instituto Butantan / FZEA-USP-Pirassununga | Antonio Jorge Martins; Claudia Renata dos Santos Barros; David Schlesinger; Debora Botequio Moretti; Dimas Tadeu Covas; Elaine Cristina Marqueze; Elaine Vieira Santos; Evandra Strazza Rodrigues; Heidge Fukumasu; Jayme Augusto de Souza-Neto; José Salvatore Leister Patané; Luiz Alcantara; Luiz Lehmann Coutinho; Maria Carolina Elias; Maurício Lacerda Nogueira; Rafael dos Santos Bezerra; Raul Machado Neto; Rejane Maria Tommasini Grotto; Ricardo Haddad; Sandra Coccuzzo Sampaio Vessoni; Simone Kashima; Svetoslav Nanev Slavov; Vincent Louis Viala |
| EPI_ISL_1795234, EPI_ISL_1795235, EPI_ISL_1795236, EPI_ISL_1795237, EPI_ISL_1795238, EPI_ISL_1795241, EPI_ISL_1795389, EPI_ISL_2345466, EPI_ISL_2345467, EPI_ISL_2345468, EPI_ISL_2345469, EPI_ISL_2345470, EPI_ISL_2345473, EPI_ISL_2345475 | | | |
| see above | CS DE BALSAMO | Instituto Butantan / ESALQ-Piracicaba | Antonio Jorge Martins; Bianca Cechetto Carlos. Mendelics: Bibiana Santos; Claudia Renata dos Santos Barros; David Schlesinger; David Schlesinger. Hemocentro Ribeirão Preto: Simone Kashima; Debora Botequio Moretti; Debora Botequio Moretti. Centro de Genômica Funcional da ESALQ: Luiz Lehmann Coutinho; Dimas Tadeu Covas; Elaine Cristina Marqueze; Elaine Vieira Santos; Elaine Vieira dos Santos; Elisangela Chicaroni Mattos; Erika Freitas; Evandra Strazza Rodrigues; Felipe Allan da Silva da Costa; Flavia Aburjaile; Guilherme Targino Valente; Heidge Fukumasu; Heidge Fukumasu. USP-Botucatu: Rejane Maria Tommasini Grotto; Instituto Butantan: Alexander Roberto Precioso; Jayme A. Souza-Neto; Jayme Augusto de Souza-Neto; Jessika Cristina Chagas Lesbon; José Salvatore Leister Patané; João Paulo Kitajima; Luiz Alcantara; Luiz Carlos Junior de Alcantara; Luiz Lehmann Coutinho; Maria Carolina Elias; Marta Giovanetti; Maurício Lacerda Nogueira; Patricia Akemi Assato; Rafael dos Santos Bezerra; Raquel de Lello Rocha Campos Cassano. NGS Soluções Genômicas: Pilar Drummond Sampaio Corrêa Mariani. FZEA-USP Pirassununga: Mirele Daiana Poleti; Raul Machado Neto; Rejane Maria Tommasini Grotto; Ricardo Augusto Brassaloti; Ricardo Haddad; Rodrigo Tocantins Calado.; Sandra Coccuzzo Sampaio; Sandra Coccuzzo Sampaio Vessoni; Simone Kashima; Svetoslav Nanev Slavov; Vagner Fonseca; Vincent Louis Viala |
| EPI_ISL_1966834 | CS DE BALSAMO | Instituto Butantan / ESALQ-USP (Piracicaba) | Antonio Jorge Martins; Bianca Cechetto Carlos. Mendelics: Bibiana Santos; Claudia Renata dos Santos Barros; Cíntia Bittar; David Schlesinger. Hemocentro Ribeirão Preto: Simone Kashima; Debora Botequio Moretti; Elaine Cristina Marqueze; Elaine Vieira dos Santos; Elisangela Chicaroni Mattos; Erika Freitas; Evandra Strazza Rodrigues; Felipe Allan da Silva da Costa; Flavia Aburjaile; Fábio Sossai Possebon; Guilherme Campos; Guilherme Targino Valente; Heidge Fukumasu. USP-Botucatu: Rejane Maria Tommasini Grotto; Helena Lage Ferreira; Instituto Butantan: Dimas Tadeu Covas; Jardelina de Souza Todao Bernardino; Jayme A. Souza-Neto; Jessika Cristina Chagas Lesbon; Jorge A. Petroli Marchesi; José Salvatore Leister Patané; João Paulo Kitajima; João Pessoa Araújo Jr.; Leila Sabrina Ullmann; Loyze Paola Oliveira de Lima; Luiz Aurelio de Campos Crispin. Centro de Genômica Funcional da ESALQ: Luiz Lehmann Coutinho; Luiz Carlos Junior de Alcantara; Lívia Sacchetto; Maisa C. Pereira Parra; Maria Carolina Elias; Marta Giovanetti; Marília Moraes; Maurício Lacerda Nogueira. Prefeitura de Sao Paulo: Melissa Palmieri.; Patricia Akemi Assato; Paula Rahal; Paulo Inacio da Costa; Rafael dos Santos Bezerra; Raquel de Lello Rocha Campos Cassano. NGS Soluções Genômicas: Pilar Drummond Sampaio Corrêa Mariani. FZEA-USP Pirassununga: Mirele Daiana Poleti; Raul Machado Neto; Ricardo Augusto Brassaloti; Ricardo Haddad; Rodrigo Tocantins Calado. FAMERP-SJRP: Cecília Artico Banho; Sandra Coccuzzo Sampaio; Svetoslav Nanev Slavov; Vagner Fonseca; Vincent Louis Viala |
| EPI_ISL_1966524 | CS DE BALSAMO | Instituto Butantan / FZEA-USP (Pirassununga) | Antonio Jorge Martins; Bianca Cechetto Carlos. Mendelics: Bibiana Santos; Claudia Renata dos Santos Barros; Cíntia Bittar; David Schlesinger. Hemocentro Ribeirão Preto: Simone Kashima; Debora Botequio Moretti; Elaine Cristina Marqueze; Elaine Vieira dos Santos; Elisangela Chicaroni Mattos; Erika Freitas; Evandra Strazza Rodrigues; Felipe Allan da Silva da Costa; Flavia Aburjaile; Fábio Sossai Possebon; Guilherme Campos; Guilherme Targino Valente; Heidge Fukumasu. USP-Botucatu: Rejane Maria Tommasini Grotto; Helena Lage Ferreira; Instituto Butantan: Dimas Tadeu Covas; Jardelina de Souza Todao Bernardino; Jayme A. Souza-Neto; Jessika Cristina Chagas Lesbon; Jorge A. Petroli Marchesi; José Salvatore Leister Patané; João Paulo Kitajima; João Pessoa Araújo Jr.; Leila Sabrina Ullmann; Loyze Paola Oliveira de Lima; Luiz Aurelio de Campos Crispin. Centro de Genômica Funcional da ESALQ: Luiz Lehmann Coutinho; Luiz Carlos Junior de Alcantara; Lívia Sacchetto; Maisa C. Pereira Parra; Maria Carolina Elias; Marta Giovanetti; Marília Moraes; Maurício Lacerda Nogueira. Prefeitura de Sao Paulo: Melissa Palmieri.; Patricia Akemi Assato; Paula Rahal; Paulo Inacio da Costa; Rafael dos Santos Bezerra; Raquel de Lello Rocha Campos Cassano. NGS Soluções Genômicas: Pilar Drummond Sampaio Corrêa Mariani. FZEA-USP Pirassununga: Mirele Daiana Poleti; Raul Machado Neto; Ricardo Augusto Brassaloti; Ricardo Haddad; Rodrigo Tocantins Calado. FAMERP-SJRP: Cecília Artico Banho; Sandra Coccuzzo Sampaio; Svetoslav Nanev Slavov; Vagner Fonseca; Vincent Louis Viala |
| EPI_ISL_1966868, EPI_ISL_1966869 | CS DE CATIGUA | Instituto Butantan / ESALQ-USP (Piracicaba) | Antonio Jorge Martins; Bianca Cechetto Carlos. Mendelics: Bibiana Santos; Claudia Renata dos Santos Barros; Cíntia Bittar; David Schlesinger. Hemocentro Ribeirão Preto: Simone Kashima; Debora Botequio Moretti; Elaine Cristina Marqueze; Elaine Vieira dos Santos; Elisangela Chicaroni Mattos; Erika Freitas; Evandra Strazza Rodrigues; Felipe Allan da Silva da Costa; Flavia Aburjaile; Fábio Sossai Possebon; Guilherme Campos; Guilherme Targino Valente; Heidge Fukumasu. USP-Botucatu: Rejane Maria Tommasini Grotto; Helena Lage Ferreira; Instituto Butantan: Dimas Tadeu Covas; Jardelina de Souza Todao Bernardino; Jayme A. Souza-Neto; Jessika Cristina Chagas Lesbon; Jorge A. Petroli Marchesi; José Salvatore Leister Patané; João Paulo Kitajima; João Pessoa Araújo Jr.; Leila Sabrina Ullmann; Loyze Paola Oliveira de Lima; Luiz Aurelio de Campos Crispin. Centro de Genômica Funcional da ESALQ: Luiz Lehmann Coutinho; Luiz Carlos Junior de Alcantara; Lívia Sacchetto; Maisa C. Pereira Parra; Maria Carolina Elias; Marta Giovanetti; Marília Moraes; Maurício Lacerda Nogueira. Prefeitura de Sao Paulo: Melissa Palmieri.; Patricia Akemi Assato; Paula Rahal; Paulo Inacio da Costa; Rafael dos Santos Bezerra; Raquel de Lello Rocha Campos Cassano. NGS Soluções Genômicas: Pilar Drummond Sampaio Corrêa Mariani. FZEA-USP Pirassununga: Mirele Daiana Poleti; Raul Machado Neto; Ricardo Augusto Brassaloti; Ricardo Haddad; Rodrigo Tocantins Calado. FAMERP-SJRP: Cecília Artico Banho; Sandra Coccuzzo Sampaio; Svetoslav Nanev Slavov; Vagner Fonseca; Vincent Louis Viala |
| EPI_ISL_1795259, EPI_ISL_1795260, EPI_ISL_1795262, EPI_ISL_1795263, EPI_ISL_2345493, EPI_ISL_2345494, EPI_ISL_2345496, EPI_ISL_2345497 | | | |
| see above | CS DE ICEM | Instituto Butantan / ESALQ-Piracicaba | Antonio Jorge Martins; Bianca Cechetto Carlos. Mendelics: Bibiana Santos; Claudia Renata dos Santos Barros; David Schlesinger; David Schlesinger. Hemocentro Ribeirão Preto: Simone Kashima; Debora Botequio Moretti; Debora Botequio Moretti. Centro de Genômica Funcional da ESALQ: Luiz Lehmann Coutinho; Dimas Tadeu Covas; Elaine Cristina Marqueze; Elaine Vieira Santos; Elaine Vieira dos Santos; Elisangela Chicaroni Mattos; Erika Freitas; Evandra Strazza Rodrigues; Felipe Allan da Silva da Costa; Flavia Aburjaile; Guilherme Targino Valente; Heidge Fukumasu; Heidge Fukumasu. USP-Botucatu: Rejane Maria Tommasini Grotto; Instituto Butantan: Alexander Roberto Precioso; Jayme A. Souza-Neto; Jayme Augusto de Souza-Neto; Jessika Cristina Chagas Lesbon; José Salvatore Leister Patané; João Paulo Kitajima; Luiz Alcantara; Luiz Carlos Junior de Alcantara; Luiz Lehmann Coutinho; Maria Carolina Elias; Marta Giovanetti; Maurício Lacerda Nogueira; Patricia Akemi Assato; Rafael dos Santos Bezerra; Raquel de Lello Rocha Campos Cassano. NGS Soluções Genômicas: Pilar Drummond Sampaio Corrêa Mariani. FZEA-USP Pirassununga: Mirele Daiana Poleti; Raul Machado Neto; Rejane Maria Tommasini Grotto; Ricardo Augusto Brassaloti; Ricardo Haddad; Rodrigo Tocantins Calado.; Sandra Coccuzzo Sampaio; Sandra Coccuzzo Sampaio Vessoni; Simone Kashima; Svetoslav Nanev Slavov; Vagner Fonseca; Vincent Louis Viala |
| EPI_ISL_1966785 | CS DE MIRASSOLANDIA | Instituto Butantan / Mendelics | Antonio Jorge Martins; Bianca Cechetto Carlos. Mendelics: Bibiana Santos; Claudia Renata dos Santos Barros; Cíntia Bittar; David Schlesinger. Hemocentro Ribeirão Preto: Simone Kashima; Debora Botequio Moretti; Elaine Cristina Marqueze; Elaine Vieira dos Santos; Elisangela Chicaroni Mattos; Erika Freitas; Evandra Strazza Rodrigues; Felipe Allan da Silva da Costa; Flavia Aburjaile; Fábio Sossai Possebon; Guilherme Campos; Guilherme Targino Valente; Heidge Fukumasu. USP-Botucatu: Rejane Maria Tommasini Grotto; Helena Lage Ferreira; Instituto Butantan: Dimas Tadeu Covas; Jardelina de Souza Todao Bernardino; Jayme A. Souza-Neto; Jessika Cristina Chagas Lesbon; Jorge A. Petroli Marchesi; José Salvatore Leister Patané; João Paulo Kitajima; João Pessoa Araújo Jr.; Leila Sabrina Ullmann; Loyze Paola Oliveira de Lima; Luiz Aurelio de Campos Crispin. Centro de Genômica Funcional da ESALQ: Luiz Lehmann Coutinho; Luiz Carlos Junior de Alcantara; Lívia Sacchetto; Maisa C. Pereira Parra; Maria Carolina Elias; Marta Giovanetti; Marília Moraes; Maurício Lacerda Nogueira. Prefeitura de Sao Paulo: Melissa Palmieri.; Patricia Akemi Assato; Paula Rahal; Paulo Inacio da Costa; Rafael dos Santos Bezerra; Raquel de Lello Rocha Campos Cassano. NGS Soluções Genômicas: Pilar Drummond Sampaio Corrêa Mariani. FZEA-USP Pirassununga: Mirele Daiana Poleti; Raul Machado Neto; Ricardo Augusto Brassaloti; Ricardo Haddad; Rodrigo Tocantins Calado. FAMERP-SJRP: Cecília Artico Banho; Sandra Coccuzzo Sampaio; Svetoslav Nanev Slavov; Vagner Fonseca; Vincent Louis Viala |
| EPI_ISL_1966961 | CS DE MONTE APRAZIVEL | Instituto Butantan / ESALQ-USP (Piracicaba) | Antonio Jorge Martins; Bianca Cechetto Carlos. Mendelics: Bibiana Santos; Claudia Renata dos Santos Barros; Cíntia Bittar; David Schlesinger. Hemocentro Ribeirão Preto: Simone Kashima; Debora Botequio Moretti; Elaine Cristina Marqueze; Elaine Vieira dos Santos; Elisangela Chicaroni Mattos; Erika Freitas; Evandra Strazza Rodrigues; Felipe Allan da Silva da Costa; Flavia Aburjaile; Fábio Sossai Possebon; Guilherme Campos; Guilherme Targino Valente; Heidge Fukumasu. USP-Botucatu: Rejane Maria Tommasini Grotto; Helena Lage Ferreira; Instituto Butantan: Dimas Tadeu Covas; Jardelina de Souza Todao Bernardino; Jayme A. Souza-Neto; Jessika Cristina Chagas Lesbon; Jorge A. Petroli Marchesi; José Salvatore Leister Patané; João Paulo Kitajima; João Pessoa Araújo Jr.; Leila Sabrina Ullmann; Loyze Paola Oliveira de Lima; Luiz Aurelio de Campos Crispin. Centro de Genômica Funcional da ESALQ: Luiz Lehmann Coutinho; Luiz Carlos Junior de Alcantara; Lívia Sacchetto; Maisa C. Pereira Parra; Maria Carolina Elias; Marta Giovanetti; Marília Moraes; Maurício Lacerda Nogueira. Prefeitura de Sao Paulo: Melissa Palmieri.; Patricia Akemi Assato; Paula Rahal; Paulo Inacio da Costa; Rafael dos Santos Bezerra; Raquel de Lello Rocha Campos Cassano. NGS Soluções Genômicas: Pilar Drummond Sampaio Corrêa Mariani. FZEA-USP Pirassununga: Mirele Daiana Poleti; Raul Machado Neto; Ricardo Augusto Brassaloti; Ricardo Haddad; Rodrigo Tocantins Calado. FAMERP-SJRP: Cecília Artico Banho; Sandra Coccuzzo Sampaio; Svetoslav Nanev Slavov; Vagner Fonseca; Vincent Louis Viala |
| EPI_ISL_1966984, EPI_ISL_1966985 | CS DE NEVES PAULISTA | Instituto Butantan / ESALQ-USP (Piracicaba) | Antonio Jorge Martins; Bianca Cechetto Carlos. Mendelics: Bibiana Santos; Claudia Renata dos Santos Barros; Cíntia Bittar; David Schlesinger. Hemocentro Ribeirão Preto: Simone Kashima; Debora Botequio Moretti; Elaine Cristina Marqueze; Elaine Vieira dos Santos; Elisangela Chicaroni Mattos; Erika Freitas; Evandra Strazza Rodrigues; Felipe Allan da Silva da Costa; Flavia Aburjaile; Fábio Sossai Possebon; Guilherme Campos; Guilherme Targino Valente; Heidge Fukumasu. USP-Botucatu: Rejane Maria Tommasini Grotto; Helena Lage Ferreira; Instituto Butantan: Dimas Tadeu Covas; Jardelina de Souza Todao Bernardino; Jayme A. Souza-Neto; Jessika Cristina Chagas Lesbon; Jorge A. Petroli Marchesi; José Salvatore Leister Patané; João Paulo Kitajima; João Pessoa Araújo Jr.; Leila Sabrina Ullmann; Loyze Paola Oliveira de Lima; Luiz Aurelio de Campos Crispin. Centro de Genômica Funcional da ESALQ: Luiz Lehmann Coutinho; Luiz Carlos Junior de Alcantara; Lívia Sacchetto; Maisa C. Pereira Parra; Maria Carolina Elias; Marta Giovanetti; Marília Moraes; Maurício Lacerda Nogueira. Prefeitura de Sao Paulo: Melissa Palmieri.; Patricia Akemi Assato; Paula Rahal; Paulo Inacio da Costa; Rafael dos Santos Bezerra; Raquel de Lello Rocha Campos Cassano. NGS Soluções Genômicas: Pilar Drummond Sampaio Corrêa Mariani. FZEA-USP Pirassununga: Mirele Daiana Poleti; Raul Machado Neto; Ricardo Augusto Brassaloti; Ricardo Haddad; Rodrigo Tocantins Calado. FAMERP-SJRP: Cecília Artico Banho; Sandra Coccuzzo Sampaio; Svetoslav Nanev Slavov; Vagner Fonseca; Vincent Louis Viala |
| EPI_ISL_1795288, EPI_ISL_2345526 | CS DE NIPOA | Instituto Butantan / ESALQ-Piracicaba | Antonio Jorge Martins; Bianca Cechetto Carlos. Mendelics: Bibiana Santos; Claudia Renata dos Santos Barros; David Schlesinger; David Schlesinger. Hemocentro Ribeirão Preto: Simone Kashima; Debora Botequio Moretti; Debora Botequio Moretti. Centro de Genômica Funcional da ESALQ: Luiz Lehmann Coutinho; Dimas Tadeu Covas; Elaine Cristina Marqueze; Elaine Vieira Santos; Elaine Vieira dos Santos; Elisangela Chicaroni Mattos; Erika Freitas; Evandra Strazza Rodrigues; Felipe Allan da Silva da Costa; Flavia Aburjaile; Guilherme Targino Valente; Heidge Fukumasu; Heidge Fukumasu. USP-Botucatu: Rejane Maria Tommasini Grotto; Instituto Butantan: Alexander Roberto Precioso; Jayme A. Souza-Neto; Jayme Augusto de Souza-Neto; Jessika Cristina Chagas Lesbon; José Salvatore Leister Patané; João Paulo Kitajima; Luiz Alcantara; Luiz Carlos Junior de Alcantara; Luiz Lehmann Coutinho; Maria Carolina Elias; Marta Giovanetti; Maurício Lacerda Nogueira; Patricia Akemi Assato; Rafael dos Santos Bezerra; Raquel de Lello Rocha Campos Cassano. NGS Soluções Genômicas: Pilar Drummond Sampaio Corrêa Mariani. FZEA-USP Pirassununga: Mirele Daiana Poleti; Raul Machado Neto; Rejane Maria Tommasini Grotto; Ricardo Augusto Brassaloti; Ricardo Haddad; Rodrigo Tocantins Calado.; Sandra Coccuzzo Sampaio; Sandra Coccuzzo Sampaio Vessoni; Simone Kashima; Svetoslav Nanev Slavov; Vagner Fonseca; Vincent Louis Viala |
| EPI_ISL_1966788, EPI_ISL_1966789 | CS DE NIPOA | Instituto Butantan / Mendelics | Antonio Jorge Martins; Bianca Cechetto Carlos. Mendelics: Bibiana Santos; Claudia Renata dos Santos Barros; Cíntia Bittar; David Schlesinger. Hemocentro Ribeirão Preto: Simone Kashima; Debora Botequio Moretti; Elaine Cristina Marqueze; Elaine Vieira dos Santos; Elisangela Chicaroni Mattos; Erika Freitas; Evandra Strazza Rodrigues; Felipe Allan da Silva da Costa; Flavia Aburjaile; Fábio Sossai Possebon; Guilherme Campos; Guilherme Targino Valente; Heidge Fukumasu. USP-Botucatu: Rejane Maria Tommasini Grotto; Helena Lage Ferreira; Instituto Butantan: Dimas Tadeu Covas; Jardelina de Souza Todao Bernardino; Jayme A. Souza-Neto; Jessika Cristina Chagas Lesbon; Jorge A. Petroli Marchesi; José Salvatore Leister Patané; João Paulo Kitajima; João Pessoa Araújo Jr.; Leila Sabrina Ullmann; Loyze Paola Oliveira de Lima; Luiz Aurelio de Campos Crispin. Centro de Genômica Funcional da ESALQ: Luiz Lehmann Coutinho; Luiz Carlos Junior de Alcantara; Lívia Sacchetto; Maisa C. Pereira Parra; Maria Carolina Elias; Marta Giovanetti; Marília Moraes; Maurício Lacerda Nogueira. Prefeitura de Sao Paulo: Melissa Palmieri.; Patricia Akemi Assato; Paula Rahal; Paulo Inacio da Costa; Rafael dos Santos Bezerra; Raquel de Lello Rocha Campos Cassano. NGS Soluções Genômicas: Pilar Drummond Sampaio Corrêa Mariani. FZEA-USP Pirassununga: Mirele Daiana Poleti; Raul Machado Neto; Ricardo Augusto Brassaloti; Ricardo Haddad; Rodrigo Tocantins Calado. FAMERP-SJRP: Cecília Artico Banho; Sandra Coccuzzo Sampaio; Svetoslav Nanev Slavov; Vagner Fonseca; Vincent Louis Viala |
| EPI_ISL_1966988 | CS DE ORINDIUVA | Instituto Butantan / ESALQ-USP (Piracicaba) | Antonio Jorge Martins; Bianca Cechetto Carlos. Mendelics: Bibiana Santos; Claudia Renata dos Santos Barros; Cíntia Bittar; David Schlesinger. Hemocentro Ribeirão Preto: Simone Kashima; Debora Botequio Moretti; Elaine Cristina Marqueze; Elaine Vieira dos Santos; Elisangela Chicaroni Mattos; Erika Freitas; Evandra Strazza Rodrigues; Felipe Allan da Silva da Costa; Flavia Aburjaile; Fábio Sossai Possebon; Guilherme Campos; Guilherme Targino Valente; Heidge Fukumasu. USP-Botucatu: Rejane Maria Tommasini Grotto; Helena Lage Ferreira; Instituto Butantan: Dimas Tadeu Covas; Jardelina de Souza Todao Bernardino; Jayme A. Souza-Neto; Jessika Cristina Chagas Lesbon; Jorge A. Petroli Marchesi; José Salvatore Leister Patané; João Paulo Kitajima; João Pessoa Araújo Jr.; Leila Sabrina Ullmann; Loyze Paola Oliveira de Lima; Luiz Aurelio de Campos Crispin. Centro de Genômica Funcional da ESALQ: Luiz Lehmann Coutinho; Luiz Carlos Junior de Alcantara; Lívia Sacchetto; Maisa C. Pereira Parra; Maria Carolina Elias; Marta Giovanetti; Marília Moraes; Maurício Lacerda Nogueira. Prefeitura de Sao Paulo: Melissa Palmieri.; Patricia Akemi Assato; Paula Rahal; Paulo Inacio da Costa; Rafael dos Santos Bezerra; Raquel de Lello Rocha Campos Cassano. NGS Soluções Genômicas: Pilar Drummond Sampaio Corrêa Mariani. FZEA-USP Pirassununga: Mirele Daiana Poleti; Raul Machado Neto; Ricardo Augusto Brassaloti; Ricardo Haddad; Rodrigo Tocantins Calado. FAMERP-SJRP: Cecília Artico Banho; Sandra Coccuzzo Sampaio; Svetoslav Nanev Slavov; Vagner Fonseca; Vincent Louis Viala |
| EPI_ISL_1795171, EPI_ISL_1795173, EPI_ISL_1795390, EPI_ISL_2345383, | CS DE OSCAR BRESSANE PSF | Instituto Butantan / ESALQ-Piracicaba | Antonio Jorge Martins; Bianca Cechetto Carlos. Mendelics: Bibiana Santos; Claudia Renata dos Santos Barros; David Schlesinger; David Schlesinger. Hemocentro Ribeirão Preto: Simone Kashima; Debora Botequio Moretti; Debora Botequio Moretti. Centro de Genômica Funcional da ESALQ: Luiz Lehmann Coutinho; Dimas Tadeu Covas; Elaine Cristina Marqueze; Elaine Vieira Santos; Elaine Vieira dos Santos; Elisangela Chicaroni Mattos; Erika Freitas; Evandra Strazza Rodrigues; Felipe Allan da Silva da Costa; Flavia Aburjaile; Guilherme Targino Valente; Heidge Fukumasu; Heidge Fukumasu. USP-Botucatu: Rejane Maria Tommasini Grotto; Instituto Butantan: Alexander Roberto Precioso; Jayme A. Souza-Neto; Jayme Augusto de Souza-Neto; Jessika Cristina Chagas Lesbon; José Salvatore Leister Patané; João Paulo Kitajima; Luiz Alcantara; Luiz Carlos Junior de Alcantara; Luiz Lehmann Coutinho; Maria Carolina Elias; Marta Giovanetti; Maurício Lacerda Nogueira; Patricia Akemi Assato; Rafael dos Santos Bezerra; Raquel de Lello Rocha Campos Cassano. NGS Soluções Genômicas: Pilar Drummond Sampaio Corrêa Mariani. FZEA-USP Pirassununga: Mirele Daiana Poleti; Raul Machado Neto; Rejane Maria Tommasini Grotto; Ricardo Augusto Brassaloti; Ricardo Haddad; Rodrigo Tocantins Calado.; Sandra Coccuzzo Sampaio; Sandra Coccuzzo |

| | | | |
|---|---|---|---|
| EPI_ISL_2345388, EPI_ISL_2345389 | | | Sampaio Vessoni; Simone Kashima; Svetoslav Nanev Slavov; Vagner Fonseca; Vincent Louis Viala |
| EPI_ISL_1795276, EPI_ISL_1795277, EPI_ISL_1795278, EPI_ISL_1795279, EPI_ISL_1795280, EPI_ISL_1795281, EPI_ISL_1795282, EPI_ISL_1795283, EPI_ISL_1795284, EPI_ISL_1795285, EPI_ISL_1795286, EPI_ISL_2345514, EPI_ISL_2345515, EPI_ISL_2345516, EPI_ISL_2345517, EPI_ISL_2345518, EPI_ISL_2345519, EPI_ISL_2345520, EPI_ISL_2345521, EPI_ISL_2345522, EPI_ISL_2345523, EPI_ISL_2345524 | | | |
| see above | CS DE PALESTINA | Instituto Butantan / ESALQ-Piracicaba | Antonio Jorge Martins; Bianca Cechetto Carlos. Mendelics: Bibiana Santos; Claudia Renata dos Santos Barros; David Schlesinger; David Schlesinger. Hemocentro Ribeirão Preto: Simone Kashima; Debora Botequio Moretti; Debora Botequio Moretti. Centro de Genômica Funcional da ESALQ: Luiz Lehmann Coutinho; Dimas Tadeu Covas; Elaine Cristina Marqueze; Elaine Vieira Santos; Elaine Vieira dos Santos; Elisangela Chicaroni Mattos; Erika Freitas; Evandra Strazza Rodrigues; Felipe Allan da Silva da Costa; Flavia Aburjaile; Guilherme Targino Valente; Heidge Fukumasu; Heidge Fukumasu. USP-Botucatu: Rejane Maria Tommasini Grotto; Instituto Butantan: Alexander Roberto Precioso; Jayme A. Souza-Neto; Jayme Augusto de Souza-Neto; Jessika Cristina Chagas Lesbon; José Salvatore Leister Patané; João Paulo Kitajima; Luiz Alcantara; Luiz Carlos Junior de Alcantara; Luiz Lehmann Coutinho; Maria Carolina Elias; Marta Giovanetti; Maurício Lacerda Nogueira; Patricia Akemi Assato; Rafael dos Santos Bezerra; Raquel de Lello Rocha Campos Cassano. NGS Soluções Genômicas: Pilar Drummond Sampaio Corrêa Mariani. FZEA-USP Pirassununga: Mirele Daiana Poleti; Raul Machado Neto; Rejane Maria Tommasini Grotto; Ricardo Augusto Brassaloti; Ricardo Haddad; Rodrigo Tocantins Calado.; Sandra Coccuzzo Sampaio; Sandra Coccuzzo Sampaio Vessoni; Simone Kashima; Svetoslav Nanev Slavov; Vagner Fonseca; Vincent Louis Viala |
| EPI_ISL_1795264, EPI_ISL_1795271, EPI_ISL_2345498, EPI_ISL_2345506 | CS DE PARANAPUA | Instituto Butantan / ESALQ-Piracicaba | Antonio Jorge Martins; Bianca Cechetto Carlos. Mendelics: Bibiana Santos; Claudia Renata dos Santos Barros; David Schlesinger; David Schlesinger. Hemocentro Ribeirão Preto: Simone Kashima; Debora Botequio Moretti; Debora Botequio Moretti. Centro de Genômica Funcional da ESALQ: Luiz Lehmann Coutinho; Dimas Tadeu Covas; Elaine Cristina Marqueze; Elaine Vieira Santos; Elaine Vieira dos Santos; Elisangela Chicaroni Mattos; Erika Freitas; Evandra Strazza Rodrigues; Felipe Allan da Silva da Costa; Flavia Aburjaile; Guilherme Targino Valente; Heidge Fukumasu; Heidge Fukumasu. USP-Botucatu: Rejane Maria Tommasini Grotto; Instituto Butantan: Alexander Roberto Precioso; Jayme A. Souza-Neto; Jayme Augusto de Souza-Neto; Jessika Cristina Chagas Lesbon; José Salvatore Leister Patané; João Paulo Kitajima; Luiz Alcantara; Luiz Carlos Junior de Alcantara; Luiz Lehmann Coutinho; Maria Carolina Elias; Marta Giovanetti; Maurício Lacerda Nogueira; Patricia Akemi Assato; Rafael dos Santos Bezerra; Raquel de Lello Rocha Campos Cassano. NGS Soluções Genômicas: Pilar Drummond Sampaio Corrêa Mariani. FZEA-USP Pirassununga: Mirele Daiana Poleti; Raul Machado Neto; Rejane Maria Tommasini Grotto; Ricardo Augusto Brassaloti; Ricardo Haddad; Rodrigo Tocantins Calado.; Sandra Coccuzzo Sampaio; Sandra Coccuzzo Sampaio Vessoni; Simone Kashima; Svetoslav Nanev Slavov; Vagner Fonseca; Vincent Louis Viala |
| EPI_ISL_1795258, EPI_ISL_2345492 | CS DE PLANALTO | Instituto Butantan / ESALQ-Piracicaba | Antonio Jorge Martins; Bianca Cechetto Carlos. Mendelics: Bibiana Santos; Claudia Renata dos Santos Barros; David Schlesinger; David Schlesinger. Hemocentro Ribeirão Preto: Simone Kashima; Debora Botequio Moretti; Debora Botequio Moretti. Centro de Genômica Funcional da ESALQ: Luiz Lehmann Coutinho; Dimas Tadeu Covas; Elaine Cristina Marqueze; Elaine Vieira Santos; Elaine Vieira dos Santos; Elisangela Chicaroni Mattos; Erika Freitas; Evandra Strazza Rodrigues; Felipe Allan da Silva da Costa; Flavia Aburjaile; Guilherme Targino Valente; Heidge Fukumasu; Heidge Fukumasu. USP-Botucatu: Rejane Maria Tommasini Grotto; Instituto Butantan: Alexander Roberto Precioso; Jayme A. Souza-Neto; Jayme Augusto de Souza-Neto; Jessika Cristina Chagas Lesbon; José Salvatore Leister Patané; João Paulo Kitajima; Luiz Alcantara; Luiz Carlos Junior de Alcantara; Luiz Lehmann Coutinho; Maria Carolina Elias; Marta Giovanetti; Maurício Lacerda Nogueira; Patricia Akemi Assato; Rafael dos Santos Bezerra; Raquel de Lello Rocha Campos Cassano. NGS Soluções Genômicas: Pilar Drummond Sampaio Corrêa Mariani. FZEA-USP Pirassununga: Mirele Daiana Poleti; Raul Machado Neto; Rejane Maria Tommasini Grotto; Ricardo Augusto Brassaloti; Ricardo Haddad; Rodrigo Tocantins Calado.; Sandra Coccuzzo Sampaio; Sandra Coccuzzo Sampaio Vessoni; Simone Kashima; Svetoslav Nanev Slavov; Vagner Fonseca; Vincent Louis Viala |
| EPI_ISL_2344769, EPI_ISL_2344772 | CS DE RINOPOLIS | Instituto Butantan / ESALQ-Piracicaba | Antonio Jorge Martins; Claudia Renata dos Santos Barros; David Schlesinger; Debora Botequio Moretti; Dimas Tadeu Covas; Elaine Cristina Marqueze; Elaine Vieira Santos; Evandra Strazza Rodrigues; Heidge Fukumasu; Jayme Augusto de Souza-Neto; José Salvatore Leister Patané; Luiz Alcantara; Luiz Lehmann Coutinho; Maria Carolina Elias; Maurício Lacerda Nogueira; Rafael dos Santos Bezerra; Raul Machado Neto; Rejane Maria Tommasini Grotto; Ricardo Haddad; Sandra Coccuzzo Sampaio Vessoni; Simone Kashima; Svetoslav Nanev Slavov; Vincent Louis Viala |
| EPI_ISL_1966257 | CS DE RUBINEIA | Instituto Butantan / Mendelics | Antonio Jorge Martins; Bianca Cechetto Carlos. Mendelics: Bibiana Santos; Claudia Renata dos Santos Barros; Cíntia Bittar; David Schlesinger. Hemocentro Ribeirão Preto: Simone Kashima; Debora Botequio Moretti; Elaine Cristina Marqueze; Elaine Vieira dos Santos; Elisangela Chicaroni Mattos; Erika Freitas; Evandra Strazza Rodrigues; Felipe Allan da Silva da Costa; Flavia Aburjaile; Fábio Sossai Possebon; Guilherme Campos; Guilherme Targino Valente; Heidge Fukumasu. USP-Botucatu: Rejane Maria Tommasini Grotto; Helena Lage Ferreira; Instituto Butantan: Dimas Tadeu Covas; Jardelina de Souza Todao Bernardino; Jayme A. Souza-Neto; Jessika Cristina Chagas Lesbon; Jorge A. Petroli Marchesi; José Salvatore Leister Patané; João Paulo Kitajima; João Pessoa Araújo Jr.; Leila Sabrina Ullmann; Loyze Paola Oliveira de Lima; Luiz Aurelio de Campos Crispin. Centro de Genômica Funcional da ESALQ: Luiz Lehmann Coutinho; Luiz Carlos Junior de Alcantara; Lívia Sacchetto; Maisa C. Pereira Parra; Maria Carolina Elias; Marta Giovanetti; Marília Moraes; Maurício Lacerda Nogueira. Prefeitura de Sao Paulo: Melissa Palmieri.; Patricia Akemi Assato; Paula Rahal; Paulo Inacio da Costa; Rafael dos Santos Bezerra; Raquel de Lello Rocha Campos Cassano. NGS Soluções Genômicas: Pilar Drummond Sampaio Corrêa Mariani. FZEA-USP Pirassununga: Mirele Daiana Poleti; Raul Machado Neto; Ricardo Augusto Brassaloti; Ricardo Haddad; Rodrigo Tocantins Calado. FAMERP-SJRP: Cecília Artico Banho; Sandra Coccuzzo Sampaio; Svetoslav Nanev Slavov; Vagner Fonseca; Vincent Louis Viala |
| EPI_ISL_1967050 | CS DE SANTA RITA DOESTE | Instituto Butantan / ESALQ-USP (Piracicaba) | Antonio Jorge Martins; Bianca Cechetto Carlos. Mendelics: Bibiana Santos; Claudia Renata dos Santos Barros; Cíntia Bittar; David Schlesinger. Hemocentro Ribeirão Preto: Simone Kashima; Debora Botequio Moretti; Elaine Cristina Marqueze; Elaine Vieira dos Santos; Elisangela Chicaroni Mattos; Erika Freitas; Evandra Strazza Rodrigues; Felipe Allan da Silva da Costa; Flavia Aburjaile; Fábio Sossai Possebon; Guilherme Campos; Guilherme Targino Valente; Heidge Fukumasu. USP-Botucatu: Rejane Maria Tommasini Grotto; Helena Lage Ferreira; Instituto Butantan: Dimas Tadeu Covas; Jardelina de Souza Todao Bernardino; Jayme A. Souza-Neto; Jessika Cristina Chagas Lesbon; Jorge A. Petroli Marchesi; José Salvatore Leister Patané; João Paulo Kitajima; João Pessoa Araújo Jr.; Leila Sabrina Ullmann; Loyze Paola Oliveira de Lima; Luiz Aurelio de Campos Crispin. Centro de Genômica Funcional da ESALQ: Luiz Lehmann Coutinho; Luiz Carlos Junior de Alcantara; Lívia Sacchetto; Maisa C. Pereira Parra; Maria Carolina Elias; Marta Giovanetti; Marília Moraes; Maurício Lacerda Nogueira. Prefeitura de Sao Paulo: Melissa Palmieri.; Patricia Akemi Assato; Paula Rahal; Paulo Inacio da Costa; Rafael dos Santos Bezerra; Raquel de Lello Rocha Campos Cassano. NGS Soluções Genômicas: Pilar Drummond Sampaio Corrêa Mariani. FZEA-USP Pirassununga: Mirele Daiana Poleti; Raul Machado Neto; Ricardo Augusto Brassaloti; Ricardo Haddad; Rodrigo Tocantins Calado. FAMERP-SJRP: Cecília Artico Banho; Sandra Coccuzzo Sampaio; Svetoslav Nanev Slavov; Vagner Fonseca; Vincent Louis Viala |
| EPI_ISL_1966547 | CS DE SANTA RITA DOESTE | Instituto Butantan / FZEA-USP (Pirassununga) | Antonio Jorge Martins; Bianca Cechetto Carlos. Mendelics: Bibiana Santos; Claudia Renata dos Santos Barros; Cíntia Bittar; David Schlesinger. Hemocentro Ribeirão Preto: Simone Kashima; Debora Botequio Moretti; Elaine Cristina Marqueze; Elaine Vieira dos Santos; Elisangela Chicaroni Mattos; Erika Freitas; Evandra Strazza Rodrigues; Felipe Allan da Silva da Costa; Flavia Aburjaile; Fábio Sossai Possebon; Guilherme Campos; Guilherme Targino Valente; Heidge Fukumasu. USP-Botucatu: Rejane Maria Tommasini Grotto; Helena Lage Ferreira; Instituto Butantan: Dimas Tadeu Covas; Jardelina de Souza Todao Bernardino; Jayme A. Souza-Neto; Jessika Cristina Chagas Lesbon; Jorge A. Petroli Marchesi; José Salvatore Leister Patané; João Paulo Kitajima; João Pessoa Araújo Jr.; Leila Sabrina Ullmann; Loyze Paola Oliveira de Lima; Luiz Aurelio de Campos Crispin. Centro de Genômica Funcional da ESALQ: Luiz Lehmann Coutinho; Luiz Carlos Junior de Alcantara; Lívia Sacchetto; Maisa C. Pereira Parra; Maria Carolina Elias; Marta Giovanetti; Marília Moraes; Maurício Lacerda Nogueira. Prefeitura de Sao Paulo: Melissa Palmieri.; Patricia Akemi Assato; Paula Rahal; Paulo Inacio da Costa; Rafael dos Santos Bezerra; Raquel de Lello Rocha Campos Cassano. NGS Soluções Genômicas: Pilar Drummond Sampaio Corrêa Mariani. FZEA-USP Pirassununga: Mirele Daiana Poleti; Raul Machado Neto; Ricardo Augusto Brassaloti; Ricardo Haddad; Rodrigo Tocantins Calado. FAMERP-SJRP: Cecília Artico Banho; Sandra Coccuzzo Sampaio; Svetoslav Nanev Slavov; Vagner Fonseca; Vincent Louis Viala |
| EPI_ISL_1795240, EPI_ISL_2345472 | CS DE SEBASTIANOPOLIS DO SUL | Instituto Butantan / ESALQ-Piracicaba | Antonio Jorge Martins; Bianca Cechetto Carlos. Mendelics: Bibiana Santos; Claudia Renata dos Santos Barros; David Schlesinger; David Schlesinger. Hemocentro Ribeirão Preto: Simone Kashima; Debora Botequio Moretti; Debora Botequio Moretti. Centro de Genômica Funcional da ESALQ: Luiz Lehmann Coutinho; Dimas Tadeu Covas; Elaine Cristina Marqueze; Elaine Vieira Santos; Elaine Vieira dos Santos; Elisangela Chicaroni Mattos; Erika Freitas; Evandra Strazza Rodrigues; Felipe Allan da Silva da Costa; Flavia Aburjaile; Guilherme Targino Valente; Heidge Fukumasu; Heidge Fukumasu. USP-Botucatu: Rejane Maria Tommasini Grotto; Instituto Butantan: Alexander Roberto Precioso; Jayme A. Souza-Neto; Jayme Augusto de Souza-Neto; Jessika Cristina Chagas Lesbon; José Salvatore Leister Patané; João Paulo Kitajima; Luiz Alcantara; Luiz Carlos Junior de Alcantara; Luiz Lehmann Coutinho; Maria Carolina Elias; Marta Giovanetti; Maurício Lacerda Nogueira; Patricia Akemi Assato; Rafael dos Santos Bezerra; Raquel de Lello Rocha Campos Cassano. NGS Soluções Genômicas: Pilar Drummond Sampaio Corrêa Mariani. FZEA-USP Pirassununga: Mirele Daiana Poleti; Raul Machado Neto; Rejane Maria Tommasini Grotto; Ricardo Augusto Brassaloti; Ricardo Haddad; Rodrigo Tocantins Calado.; Sandra Coccuzzo Sampaio; Sandra Coccuzzo Sampaio Vessoni; Simone Kashima; Svetoslav Nanev Slavov; Vagner Fonseca; Vincent Louis Viala |
| EPI_ISL_1795232, EPI_ISL_1795233, EPI_ISL_1795239, EPI_ISL_1795242, EPI_ISL_1795243, EPI_ISL_1795244, EPI_ISL_2345464, EPI_ISL_2345465, EPI_ISL_2345471, EPI_ISL_2345474, EPI_ISL_2345476, EPI_ISL_2345477 | | | |
| see above | CS DE URUPES | Instituto Butantan / ESALQ-Piracicaba | Antonio Jorge Martins; Bianca Cechetto Carlos. Mendelics: Bibiana Santos; Claudia Renata dos Santos Barros; David Schlesinger; David Schlesinger. Hemocentro Ribeirão Preto: Simone Kashima; Debora Botequio Moretti; Debora Botequio Moretti. Centro de Genômica Funcional da ESALQ: Luiz Lehmann Coutinho; Dimas Tadeu Covas; Elaine Cristina Marqueze; Elaine Vieira Santos; Elaine Vieira dos Santos; Elisangela Chicaroni Mattos; Erika Freitas; Evandra Strazza Rodrigues; Felipe Allan da Silva da Costa; Flavia Aburjaile; Guilherme Targino Valente; Heidge Fukumasu; Heidge Fukumasu. USP-Botucatu: Rejane Maria Tommasini Grotto; Instituto Butantan: Alexander Roberto Precioso; Jayme A. Souza-Neto; Jayme Augusto de Souza-Neto; Jessika Cristina Chagas Lesbon; José Salvatore Leister Patané; João Paulo Kitajima; Luiz Alcantara; Luiz Carlos Junior de Alcantara; Luiz Lehmann Coutinho; Maria Carolina Elias; Marta Giovanetti; Maurício Lacerda Nogueira; Patricia Akemi Assato; Rafael dos Santos Bezerra; Raquel de Lello Rocha Campos Cassano. NGS Soluções Genômicas: Pilar Drummond Sampaio Corrêa Mariani. FZEA-USP Pirassununga: Mirele Daiana Poleti; Raul Machado Neto; Rejane Maria Tommasini Grotto; Ricardo Augusto Brassaloti; Ricardo Haddad; Rodrigo Tocantins Calado.; Sandra Coccuzzo Sampaio; Sandra Coccuzzo Sampaio Vessoni; Simone Kashima; Svetoslav Nanev Slavov; Vagner Fonseca; Vincent Louis Viala |
| EPI_ISL_1966992, EPI_ISL_1966993 | CS I DE OURINHOS | Instituto Butantan / ESALQ-USP (Piracicaba) | Antonio Jorge Martins; Bianca Cechetto Carlos. Mendelics: Bibiana Santos; Claudia Renata dos Santos Barros; Cíntia Bittar; David Schlesinger. Hemocentro Ribeirão Preto: Simone Kashima; Debora Botequio Moretti; Elaine Cristina Marqueze; Elaine Vieira dos Santos; Elisangela Chicaroni Mattos; Erika Freitas; Evandra Strazza Rodrigues; Felipe Allan da Silva da Costa; Flavia Aburjaile; Fábio Sossai Possebon; Guilherme Campos; Guilherme Targino Valente; Heidge Fukumasu. USP-Botucatu: Rejane Maria Tommasini Grotto; Helena Lage Ferreira; Instituto Butantan: Dimas Tadeu Covas; Jardelina de Souza Todao Bernardino; Jayme A. Souza-Neto; Jessika Cristina Chagas Lesbon; Jorge A. Petroli Marchesi; José Salvatore Leister Patané; João Paulo Kitajima; João Pessoa Araújo Jr.; Leila Sabrina Ullmann; Loyze Paola Oliveira de Lima; Luiz Aurelio de Campos Crispin. Centro de Genômica Funcional da ESALQ: Luiz Lehmann Coutinho; Luiz Carlos Junior de Alcantara; Lívia Sacchetto; Maisa C. Pereira Parra; Maria Carolina Elias; Marta Giovanetti; Marília Moraes; Maurício Lacerda Nogueira. Prefeitura de Sao Paulo: Melissa Palmieri.; Patricia Akemi Assato; Paula Rahal; Paulo Inacio da Costa; Rafael dos Santos Bezerra; Raquel de Lello Rocha Campos Cassano. NGS Soluções Genômicas: Pilar Drummond Sampaio Corrêa Mariani. FZEA-USP Pirassununga: Mirele Daiana Poleti; Raul Machado Neto; Ricardo Augusto Brassaloti; Ricardo Haddad; Rodrigo Tocantins Calado. FAMERP-SJRP: Cecília Artico Banho; Sandra Coccuzzo Sampaio; Svetoslav Nanev Slavov; Vagner Fonseca; Vincent Louis Viala |
| EPI_ISL_2344778 | CS IACRI | Instituto Butantan / ESALQ-Piracicaba | Antonio Jorge Martins; Claudia Renata dos Santos Barros; David Schlesinger; Debora Botequio Moretti; Dimas Tadeu Covas; Elaine Cristina Marqueze; Elaine Vieira Santos; Evandra Strazza Rodrigues; Heidge Fukumasu; Jayme Augusto de Souza-Neto; José Salvatore Leister Patané; Luiz Alcantara; Luiz Lehmann Coutinho; Maria Carolina Elias; Maurício Lacerda Nogueira; Rafael dos Santos Bezerra; Raul Machado Neto; Rejane Maria Tommasini Grotto; Ricardo Haddad; Sandra Coccuzzo Sampaio Vessoni; Simone Kashima; Svetoslav Nanev Slavov; Vincent Louis Viala |
| EPI_ISL_1966909 | CS II DR JAHYR DE PAULA RIBEIRO GUARA | Instituto Butantan / ESALQ-USP (Piracicaba) | Antonio Jorge Martins; Bianca Cechetto Carlos. Mendelics: Bibiana Santos; Claudia Renata dos Santos Barros; Cíntia Bittar; David Schlesinger. Hemocentro Ribeirão Preto: Simone Kashima; Debora Botequio Moretti; Elaine Cristina Marqueze; Elaine Vieira dos Santos; Elisangela Chicaroni Mattos; Erika Freitas; Evandra Strazza Rodrigues; Felipe Allan da Silva da Costa; Flavia Aburjaile; Fábio Sossai Possebon; Guilherme Campos; Guilherme Targino Valente; Heidge Fukumasu. USP-Botucatu: Rejane Maria Tommasini Grotto; Helena Lage Ferreira; Instituto Butantan: Dimas Tadeu Covas; Jardelina de Souza Todao Bernardino; Jayme A. Souza-Neto; Jessika Cristina Chagas Lesbon; Jorge A. Petroli Marchesi; José Salvatore Leister Patané; João Paulo Kitajima; João Pessoa Araújo Jr.; Leila Sabrina Ullmann; Loyze Paola Oliveira de Lima; Luiz Aurelio de Campos Crispin. Centro de Genômica Funcional da ESALQ: Luiz Lehmann Coutinho; Luiz Carlos Junior de Alcantara; Lívia Sacchetto; Maisa C. Pereira Parra; Maria Carolina Elias; Marta Giovanetti; Marília Moraes; Maurício Lacerda Nogueira. Prefeitura de Sao Paulo: Melissa Palmieri.; Patricia Akemi Assato; Paula Rahal; Paulo Inacio da Costa; Rafael dos Santos Bezerra; Raquel de Lello Rocha Campos Cassano. NGS Soluções Genômicas: Pilar Drummond Sampaio Corrêa Mariani. FZEA-USP Pirassununga: Mirele Daiana Poleti; Raul Machado Neto; Ricardo Augusto Brassaloti; Ricardo Haddad; Rodrigo Tocantins Calado. FAMERP-SJRP: Cecília Artico Banho; Sandra Coccuzzo Sampaio; Svetoslav Nanev Slavov; Vagner Fonseca; Vincent Louis Viala |
| EPI_ISL_1966808 | CS II DR JOSE CARQUEIJO SCRPARDO | Instituto Butantan / Mendelics | Antonio Jorge Martins; Bianca Cechetto Carlos. Mendelics: Bibiana Santos; Claudia Renata dos Santos Barros; Cíntia Bittar; David Schlesinger. Hemocentro Ribeirão Preto: Simone Kashima; Debora Botequio Moretti; Elaine Cristina Marqueze; Elaine Vieira dos Santos; Elisangela Chicaroni Mattos; Erika Freitas; Evandra Strazza Rodrigues; Felipe Allan da Silva da Costa; Flavia Aburjaile; Fábio Sossai Possebon; Guilherme Campos; Guilherme Targino Valente; Heidge Fukumasu. USP-Botucatu: Rejane Maria Tommasini Grotto; Helena Lage Ferreira; Instituto Butantan: Dimas Tadeu Covas; Jardelina de Souza Todao Bernardino; Jayme A. Souza-Neto; Jessika Cristina Chagas Lesbon; Jorge A. Petroli Marchesi; José Salvatore Leister Patané; João Paulo Kitajima; João Pessoa Araújo Jr.; Leila Sabrina Ullmann; Loyze Paola Oliveira de Lima; Luiz Aurelio de Campos Crispin. Centro de Genômica Funcional da ESALQ: Luiz Lehmann Coutinho; Luiz Carlos Junior de Alcantara; Lívia Sacchetto; Maisa C. Pereira Parra; Maria Carolina Elias; Marta Giovanetti; Marília Moraes; Maurício Lacerda Nogueira. Prefeitura de Sao Paulo: Melissa Palmieri.; Patricia Akemi Assato; Paula Rahal; Paulo Inacio da Costa; Rafael dos Santos Bezerra; Raquel de Lello Rocha Campos Cassano. NGS Soluções Genômicas: Pilar Drummond Sampaio Corrêa Mariani. FZEA-USP Pirassununga: Mirele Daiana Poleti; Raul Machado Neto; Ricardo Augusto Brassaloti; Ricardo Haddad; Rodrigo Tocantins Calado. FAMERP-SJRP: Cecília Artico Banho; Sandra Coccuzzo Sampaio; Svetoslav Nanev Slavov; Vagner Fonseca; Vincent Louis Viala |
| EPI_ISL_1966989 | CS II DR MIGUEL VITALIANO ORLANDIA | Instituto Butantan / ESALQ-USP (Piracicaba) | Antonio Jorge Martins; Bianca Cechetto Carlos. Mendelics: Bibiana Santos; Claudia Renata dos Santos Barros; Cíntia Bittar; David Schlesinger. Hemocentro Ribeirão Preto: Simone Kashima; Debora Botequio Moretti; Elaine Cristina Marqueze; Elaine Vieira dos Santos; Elisangela Chicaroni Mattos; Erika Freitas; Evandra Strazza Rodrigues; Felipe Allan da Silva da Costa; Flavia Aburjaile; Fábio Sossai Possebon; Guilherme Campos; Guilherme Targino Valente; Heidge Fukumasu. USP-Botucatu: Rejane Maria Tommasini Grotto; Helena Lage Ferreira; Instituto Butantan: Dimas Tadeu Covas; Jardelina de Souza Todao Bernardino; Jayme A. Souza-Neto; Jessika Cristina Chagas Lesbon; Jorge A. Petroli Marchesi; José Salvatore Leister Patané; João Paulo Kitajima; João Pessoa Araújo Jr.; Leila Sabrina Ullmann; Loyze Paola Oliveira de Lima; Luiz Aurelio de Campos Crispin. Centro de Genômica Funcional da ESALQ: Luiz Lehmann Coutinho; Luiz Carlos Junior de Alcantara; Lívia Sacchetto; Maisa C. Pereira Parra; Maria Carolina Elias; Marta Giovanetti; Marília Moraes; Maurício Lacerda Nogueira. Prefeitura de Sao Paulo: Melissa Palmieri.; Patricia Akemi Assato; Paula Rahal; Paulo Inacio da Costa; Rafael dos Santos Bezerra; Raquel de Lello Rocha Campos Cassano. NGS Soluções Genômicas: Pilar Drummond Sampaio Corrêa Mariani. FZEA-USP Pirassununga: Mirele Daiana Poleti; Raul Machado Neto; Ricardo Augusto Brassaloti; Ricardo Haddad; Rodrigo Tocantins Calado. FAMERP-SJRP: Cecília Artico Banho; Sandra Coccuzzo Sampaio; Svetoslav Nanev Slavov; Vagner Fonseca; Vincent Louis Viala |
| EPI_ISL_1625975 | CS II Dr Antonio Vicoso Moreira De Rezende Sumare | Instituto Adolfo Lutz, Interdiciplinary Procedures Center, Strategic Laboratory | Caio Vinicius Dias Lopes; Claudia Regina Gonçalves; Claudio Tavares Sacchi; Erica Valessa Ramos Gomes; Karoline Rodrigues Campos; Katia Correa de Oliveira Santos; Leonardo Jose Tadeu de Araujo |
| EPI_ISL_1715135, EPI_ISL_1715136 | CS II Dr Jose Ferreira Telles | Instituto Adolfo Lutz, Interdiciplinary Procedures Center, Strategic Laboratory | Caio Vinicius Dias Lopes; Claudia Regina Gonçalves; Claudio Tavares Sacchi; Erica Valessa Ramos Gomes; Karoline Rodrigues Campos; Katia Correa de Oliveira Santos; Leonardo Jose Tadeu de Araujo |
| EPI_ISL_1493580, EPI_ISL_1493582 | CS II Dr Miguel Vitaliano Orlandia | Instituto Adolfo Lutz, Interdiciplinary Procedures Center, Strategic Laboratory | Caio Vinicius Dias Lopes; Claudia Regina Gonçalves; Claudio Tavares Sacchi; Erica Valessa Ramos Gomes; Karoline Rodrigues Campos |
| EPI_ISL_1966965, EPI_ISL_1966966, EPI_ISL_1966967, EPI_ISL_1966968, EPI_ISL_1966970, EPI_ISL_1966971, EPI_ISL_1966972, EPI_ISL_1966973, EPI_ISL_1966974, EPI_ISL_1966975, EPI_ISL_1966977, EPI_ISL_1966978, EPI_ISL_1966979, EPI_ISL_1966980, EPI_ISL_1966981, EPI_ISL_1966982, EPI_ISL_1966983 | | | |
| see above | CS II EGIDIO BRUNHARA MORRO AGUDO | Instituto Butantan / ESALQ-USP (Piracicaba) | Antonio Jorge Martins; Bianca Cechetto Carlos. Mendelics: Bibiana Santos; Claudia Renata dos Santos Barros; Cíntia Bittar; David Schlesinger. Hemocentro Ribeirão Preto: Simone Kashima; Debora Botequio Moretti; Elaine Cristina Marqueze; Elaine Vieira dos Santos; Elisangela Chicaroni Mattos; Erika Freitas; Evandra Strazza Rodrigues; Felipe Allan da Silva da Costa; Flavia Aburjaile; Fábio Sossai Possebon; Guilherme Campos; Guilherme Targino Valente; Heidge Fukumasu. USP-Botucatu: Rejane Maria Tommasini Grotto; Helena Lage Ferreira; Instituto Butantan: Dimas Tadeu Covas; Jardelina de Souza Todao Bernardino; Jayme A. Souza-Neto; Jessika Cristina Chagas Lesbon; Jorge A. Petroli Marchesi; José Salvatore Leister Patané; João Paulo Kitajima; João Pessoa Araújo Jr.; Leila Sabrina Ullmann; Loyze Paola Oliveira de Lima; Luiz Aurelio de Campos Crispin. Centro de Genômica Funcional da ESALQ: Luiz Lehmann Coutinho; Luiz Carlos Junior de Alcantara; Lívia Sacchetto; Maisa C. Pereira Parra; Maria Carolina Elias; Marta Giovanetti; Marília Moraes; Maurício Lacerda Nogueira. Prefeitura de Sao Paulo: Melissa Palmieri.; Patricia Akemi Assato; Paula Rahal; Paulo Inacio da Costa; Rafael dos Santos Bezerra; Raquel de Lello Rocha Campos Cassano. NGS Soluções Genômicas: Pilar Drummond Sampaio Corrêa Mariani. FZEA-USP Pirassununga: Mirele Daiana Poleti; Raul Machado Neto; Ricardo Augusto Brassaloti; Ricardo Haddad; Rodrigo Tocantins Calado. FAMERP-SJRP: Cecília Artico Banho; Sandra Coccuzzo Sampaio; Svetoslav Nanev Slavov; Vagner Fonseca; Vincent Louis Viala |
| EPI_ISL_1966480 | CS III SANTOPOLIS DO AGUAPEI | Instituto Butantan / Mendelics | Antonio Jorge Martins; Bianca Cechetto Carlos. Mendelics: Bibiana Santos; Claudia Renata dos Santos Barros; Cíntia Bittar; David Schlesinger. Hemocentro Ribeirão Preto: Simone Kashima; Debora Botequio Moretti; Elaine Cristina Marqueze; Elaine Vieira dos Santos; Elisangela Chicaroni Mattos; Erika Freitas; Evandra Strazza Rodrigues; Felipe Allan da Silva da Costa; Flavia Aburjaile; Fábio Sossai Possebon; Guilherme Campos; Guilherme Targino Valente; Heidge Fukumasu. USP-Botucatu: Rejane Maria Tommasini Grotto; Helena Lage Ferreira; Instituto Butantan: Dimas Tadeu Covas; Jardelina de Souza Todao Bernardino; Jayme A. Souza-Neto; Jessika Cristina Chagas Lesbon; Jorge A. Petroli Marchesi; José Salvatore Leister Patané; João Paulo Kitajima; João Pessoa Araújo Jr.; Leila Sabrina Ullmann; Loyze Paola Oliveira de Lima; Luiz Aurelio de Campos Crispin. Centro de Genômica Funcional da ESALQ: Luiz Lehmann Coutinho; Luiz Carlos Junior de Alcantara; Lívia Sacchetto; Maisa C. Pereira Parra; Maria Carolina Elias; Marta Giovanetti; Marília Moraes; Maurício Lacerda Nogueira. Prefeitura de Sao Paulo: Melissa Palmieri.; Patricia Akemi Assato; Paula Rahal; Paulo Inacio da Costa; Rafael dos Santos Bezerra; Raquel de Lello Rocha Campos Cassano. NGS Soluções Genômicas: Pilar Drummond Sampaio Corrêa Mariani. FZEA-USP Pirassununga: Mirele Daiana Poleti; Raul Machado Neto; Ricardo Augusto Brassaloti; Ricardo Haddad; Rodrigo Tocantins Calado. FAMERP-SJRP: Cecília Artico Banho; Sandra Coccuzzo Sampaio; Svetoslav Nanev Slavov; Vagner Fonseca; Vincent Louis Viala |
| EPI_ISL_1795161, EPI_ISL_2345372 | CS III VILA ODILON | Instituto Butantan / ESALQ-Piracicaba | Antonio Jorge Martins; Bianca Cechetto Carlos. Mendelics: Bibiana Santos; Claudia Renata dos Santos Barros; David Schlesinger; David Schlesinger. Hemocentro Ribeirão Preto: Simone Kashima; Debora Botequio Moretti; Debora Botequio Moretti. Centro de Genômica Funcional da ESALQ: Luiz Lehmann Coutinho; Dimas Tadeu Covas; Elaine Cristina Marqueze; Elaine Vieira Santos; Elaine Vieira dos Santos; Elisangela Chicaroni Mattos; Erika Freitas; Evandra Strazza Rodrigues; Felipe Allan da Silva da Costa; Flavia Aburjaile; Guilherme Targino Valente; Heidge Fukumasu; Heidge Fukumasu. USP-Botucatu: Rejane Maria Tommasini Grotto; Instituto Butantan: Alexander Roberto Precioso; Jayme A. Souza-Neto; Jayme Augusto de Souza-Neto; Jessika Cristina Chagas Lesbon; José Salvatore Leister Patané; João Paulo Kitajima; Luiz Alcantara; Luiz Carlos Junior de Alcantara; Luiz Lehmann Coutinho; Maria Carolina Elias; Marta Giovanetti; Maurício Lacerda Nogueira; Patricia Akemi Assato; Rafael dos Santos Bezerra; Raquel de Lello Rocha Campos Cassano. NGS Soluções Genômicas: Pilar Drummond Sampaio Corrêa Mariani. FZEA-USP Pirassununga: Mirele Daiana Poleti; Raul Machado Neto; Rejane Maria Tommasini Grotto; Ricardo Augusto Brassaloti; Ricardo Haddad; Rodrigo Tocantins Calado.; Sandra Coccuzzo Sampaio; Sandra Coccuzzo Sampaio Vessoni; Simone Kashima; Svetoslav Nanev Slavov; Vagner Fonseca; Vincent Louis Viala |
| EPI_ISL_1628366, EPI_ISL_1715142 | CS III de Patrocinio Paulista | Instituto Adolfo Lutz, Interdiciplinary Procedures Center, | Caio Vinicius Dias Lopes; Claudia Regina Gonçalves; Claudio Tavares Sacchi; Erica Valessa Ramos Gomes; Karoline Rodrigues Campos; Katia Correa de Oliveira Santos; Leonardo Jose Tadeu de Araujo |

| | | | |
|---|---|---|---|
| | | Strategic Laboratory | |
| EPI_ISL_1821209, EPI_ISL_2003112 | CS de Paulo de Faria | Instituto Adolfo Lutz, Interdiciplinary Procedures Center, Strategic Laboratory | Caio Vinicius Dias Lopes; Claudia Regina Gonçalves; Claudio Tavares Sacchi; Erica Valessa Ramos Gomes; Karoline Rodrigues Campos; Leonardo Jose Tadeu de Araujo |
| EPI_ISL_3912154 | CSF COHAB II | Analytical Competence Molecular Epidemiology Lab/ACME, Oswaldo Cruz Foundation, Ceara (FIOCRUZ CE) | Cleber Furtado Aksenen; Fabio Miyajima; Fernando Braga Stehling; Francisco Eder de Moura Lopes; Jamille Maria Mendes Bezerra; Joaquim Cesar do Nascimento Sousa Junior; Pedro Miguel Carneiro Jeronimo; Suzana Porto Almeida & Lucas Delerino on behalf of COVID-19 FIOCRUZ Genomic Network; Thais Ferreira de Oliveira; Thais de Oliveira Costa; Ticiane Cavalcante de Souza; Veridiana Pessoa Miyajima |
| EPI_ISL_3912156 | CSF SINHA SABOIA | Analytical Competence Molecular Epidemiology Lab/ACME, Oswaldo Cruz Foundation, Ceara (FIOCRUZ CE) | Cleber Furtado Aksenen; Fabio Miyajima; Fernando Braga Stehling; Francisco Eder de Moura Lopes; Jamille Maria Mendes Bezerra; Joaquim Cesar do Nascimento Sousa Junior; Pedro Miguel Carneiro Jeronimo; Suzana Porto Almeida & Lucas Delerino on behalf of COVID-19 FIOCRUZ Genomic Network; Thais Ferreira de Oliveira; Thais de Oliveira Costa; Ticiane Cavalcante de Souza; Veridiana Pessoa Miyajima |
| EPI_ISL_3912157 | CSF TAPERUABA | Analytical Competence Molecular Epidemiology Lab/ACME, Oswaldo Cruz Foundation, Ceara (FIOCRUZ CE) | Cleber Furtado Aksenen; Fabio Miyajima; Fernando Braga Stehling; Francisco Eder de Moura Lopes; Jamille Maria Mendes Bezerra; Joaquim Cesar do Nascimento Sousa Junior; Pedro Miguel Carneiro Jeronimo; Suzana Porto Almeida & Lucas Delerino on behalf of COVID-19 FIOCRUZ Genomic Network; Thais Ferreira de Oliveira; Thais de Oliveira Costa; Ticiane Cavalcante de Souza; Veridiana Pessoa Miyajima |
| EPI_ISL_1967004, EPI_ISL_1967006, EPI_ISL_1967007, EPI_ISL_1967008, EPI_ISL_1967010, EPI_ISL_1967011, EPI_ISL_1967012, EPI_ISL_1967013 | | | |
| see above | CSII DR WASHINGTON LUIS M RODRIGUES DA SILVA PITANGUEIRAS | Instituto Butantan / ESALQ-USP (Piracicaba) | Antonio Jorge Martins; Bianca Cechetto Carlos. Mendelics: Bibiana Santos; Claudia Renata dos Santos Barros; Cíntia Bittar; David Schlesinger. Hemocentro Ribeirão Preto: Simone Kashima; Debora Botequio Moretti; Elaine Cristina Marqueze; Elaine Vieira dos Santos; Elisangela Chicaroni Mattos; Erika Freitas; Evandra Strazza Rodrigues; Felipe Allan da Silva da Costa; Flavia Aburjaile; Fábio Sossai Possebon; Guilherme Campos; Guilherme Targino Valente; Heidge Fukumasu. USP-Botucatu: Rejane Maria Tommasini Grotto; Helena Lage Ferreira; Instituto Butantan: Dimas Tadeu Covas; Jardelina de Souza Todao Bernardino; Jayme A. Souza-Neto; Jessika Cristina Chagas Lesbon; Jorge A. Petroli Marchesi; José Salvatore Leister Patané; João Paulo Kitajima; João Pessoa Araújo Jr.; Leila Sabrina Ullmann; Loyze Paola Oliveira de Lima; Luiz Aurelio de Campos Crispin. Centro de Genômica Funcional da ESALQ: Luiz Lehmann Coutinho; Luiz Carlos Junior de Alcantara; Lívia Sacchetto; Maisa C. Pereira Parra; Maria Carolina Elias; Marta Giovanetti; Marília Moraes; Maurício Lacerda Nogueira. Prefeitura de Sao Paulo: Melissa Palmieri.; Patricia Akemi Assato; Paula Rahal; Paulo Inacio da Costa; Rafael dos Santos Bezerra; Raquel de Lello Rocha Campos Cassano. NGS Soluções Genômicas: Pilar Drummond Sampaio Corrêa Mariani. FZEA-USP Pirassununga: Mirele Daiana Poleti; Raul Machado Neto; Ricardo Augusto Brassaloti; Ricardo Haddad; Rodrigo Tocantins Calado. FAMERP-SJRP: Cecília Artico Banho; Sandra Coccuzzo Sampaio; Svetoslav Nanev Slavov; Vagner Fonseca; Vincent Louis Viala |
| EPI_ISL_2105160 | CT-Dr. Katherine A. Kelley State Public Health Lab | Centers for Disease Control and Prevention Division of Viral Diseases, Pathogen Discovery | Alison Laufer Halpin; Ben L. Rambo-Martin; Clinton R. Paden; Dakota Howard; Darlene Wagner; Dave Wentworth; Dhwani Batra; Jasmine Padilla; Justin Lee; Katie Dillon; Krista Queen; Kristen Knipe; Kristine Lacek; Mark Burroughs; Matthew Schmerer; Mili Sheth; Peter Cook; Sam Shepard; Sarah Nobles; Shoshona Le; Suxiang Tong; Vivien Dugan; Yvette Unoarumhi |
| EPI_ISL_2234349, EPI_ISL_2537462 | California Department of Public Health | California Department of Public Health | CDPH IDLB COVIDNet et al |
| EPI_ISL_2837494, EPI_ISL_2837516, EPI_ISL_2837724, EPI_ISL_2837733, EPI_ISL_2837789, EPI_ISL_2837807, EPI_ISL_2837808, EPI_ISL_2837809, EPI_ISL_2837817, EPI_ISL_2837820, EPI_ISL_2837908 | | | |
| see above | California Department of Public Health Valencia Branch Laboratory (CDPH VBL) | California Department of Public Health | CDPH-COVIDNet |
| EPI_ISL_1731575, EPI_ISL_1752638 | Casa de Caridade Sao Vicente de Paulo Cajuru | Instituto Adolfo Lutz, Interdiciplinary Procedures Center, Strategic Laboratory | Caio Vinicius Dias Lopes; Claudia Regina Gonçalves; Claudio Tavares Sacchi; Erica Valessa Ramos Gomes; Karoline Rodrigues Campos; Katia Correa de Oliveira Santos; Leonardo Jose Tadeu de Araujo |
| EPI_ISL_1067728, EPI_ISL_1067736 | Center for Biotechnology and Cell Therapy, São Rafael Hospital, Salvador, Brazil | Central Public Health Laboratory - LACEN - Bahia, Salvador, Brazil | Arabela Leal; Breno Dominguez; Felicidade Pereira; Jaqueline Gomes; Luciana Oliveira; Luiz Alcantara; Marcela Gómez; Marta Giovanetti; Patrícia Cajado; Stephane Tosta; Vagner Fonseca; Vanessa Nardy |
| EPI_ISL_1760267, EPI_ISL_1760280, EPI_ISL_1760281, EPI_ISL_1760284 | Centers for Disease Control and Prevention, Dengue Branch | Centers for Disease Control and Prevention, Dengue Branch | Betzabel Flores; Gabriela Paz-Bailey; Gilberto A. Santiago; Glenda Gonzalez; Jorge L. Munoz-Jordan; Keyla Charriez |
| EPI_ISL_2108582 | Centogene; Dr. Bauer Laboratoriums GmbH | Robert Koch Institute | |
| EPI_ISL_984619, EPI_ISL_984620, EPI_ISL_984621 | Central Laboratory of Public Health of Rio Grande do Sul (Lacen-RS) | State Center for Health Surveillance of the Health Department of the State of Rio Grande do Sul (CEVS/SES-RS) | Aline Campos; Cynthia Molina; Lara Crescente; Leticia Garay; Ludmila Fiorenzano Baethgen; Richard Salvato; Tatiana Gregianini |
| EPI_ISL_2502442, EPI_ISL_2502443, EPI_ISL_2502444, EPI_ISL_2502445, EPI_ISL_2502446, EPI_ISL_2502447, EPI_ISL_2502448, EPI_ISL_2502449, EPI_ISL_2502450, EPI_ISL_2502451, EPI_ISL_2502452, EPI_ISL_2502453, EPI_ISL_2502454, EPI_ISL_2502455, EPI_ISL_2502456, EPI_ISL_2502457, EPI_ISL_2502458, EPI_ISL_2502459, EPI_ISL_2502460, EPI_ISL_2502461, EPI_ISL_2502462, EPI_ISL_2502463, EPI_ISL_2502464, EPI_ISL_2502465, EPI_ISL_2502466, EPI_ISL_2502467, EPI_ISL_2502468, EPI_ISL_2502469, EPI_ISL_2502470, EPI_ISL_2502471, EPI_ISL_2502472, EPI_ISL_2502473, EPI_ISL_2502474, EPI_ISL_2502475, EPI_ISL_2549287 | | | |
| see above | Central Laboratory, Bureau of Public Health (BOG) and Academic Hospital Paramaribo | Erasmus Medical Center | Bas B Oude Munnink; Cherise Beek; Consuella Partowidjojo; Dion Gajadin; Ed PF IJzerman; Emmanuelle Munger; Gary Gummels; Ingrid SK Krishnadath; Lycke Woittiez; Marion PG Koopmans; Mireille Van de Veer; Phyllis Pinas; Princes Wongsowidjojo; Radjesh Ori; Ranisha Doerbalie; Rohma Banwari; Soeradj Harkisoen; Stephen Vreden; Tilotmadebie Ramlal; Verne Nanhoe |
| EPI_ISL_1067729, EPI_ISL_1067730, EPI_ISL_1067731, EPI_ISL_1067733, EPI_ISL_1067734, EPI_ISL_1067735, EPI_ISL_1067737, EPI_ISL_1067738, EPI_ISL_1583650, EPI_ISL_1583656, EPI_ISL_1583665, EPI_ISL_1583673, EPI_ISL_1583674, EPI_ISL_1583676, EPI_ISL_1583677, EPI_ISL_1583679, EPI_ISL_1583680, EPI_ISL_1583681, EPI_ISL_1583682, EPI_ISL_1583683, EPI_ISL_1583694, EPI_ISL_1583697, EPI_ISL_1583700, EPI_ISL_1583719, EPI_ISL_1583722, EPI_ISL_1583725, EPI_ISL_1583730, EPI_ISL_1583733, EPI_ISL_3255057, EPI_ISL_3255058, EPI_ISL_3255061, EPI_ISL_3255064, EPI_ISL_3255077, EPI_ISL_3255078, EPI_ISL_3255079, EPI_ISL_3255134, EPI_ISL_3255187, EPI_ISL_3255193, EPI_ISL_3255195, EPI_ISL_3266079, EPI_ISL_3266081, EPI_ISL_3266087, EPI_ISL_3266089, EPI_ISL_3266090, EPI_ISL_3266091, EPI_ISL_3266092, EPI_ISL_3266094, EPI_ISL_3266099, EPI_ISL_3266103, EPI_ISL_3266106, EPI_ISL_3266107, EPI_ISL_3266108, EPI_ISL_3266109, EPI_ISL_3266110, EPI_ISL_3266111, EPI_ISL_3266112, EPI_ISL_3266113, EPI_ISL_3266114, EPI_ISL_3266117, EPI_ISL_3266118, EPI_ISL_3944576 | | | |
| see above | Central Public Health Laboratory - LACEN - Bahia, Salvador, Brazil | Central Public Health Laboratory - LACEN - Bahia, Salvador, Brazil | Arabela Leal; Breno Dominguez; Felicidade Pereira; Gabriela Menezes; Jaqueline Gomes; Lenisa Dandara; Luciana Oliveira; Luiz Alcantara; Marcela Gómez; Marta Giovanetti; Patrícia Cajado; Stephane Tosta; Vagner Fonseca; Vanessa Nardy |
| EPI_ISL_1547454 | Centre De Prelevement COVID RIOM | CHU Clermont-Ferrand, service de virologie | Bisseux Maxime; Combes Patricia; Henquell Cécile; Mirand Audrey |
| EPI_ISL_1483773, EPI_ISL_1623086 | Centre Hospitalier Universitaire Clermont-Ferrand | CHU Clermont-Ferrand, service de virologie | Bisseux Maxime; Combes Patricia; Henquell Cécile; Mirand Audrey |
| EPI_ISL_1628377 | Centro De Saude II Ibitinga | Instituto Adolfo Lutz, Interdiciplinary Procedures Center, Strategic Laboratory | Caio Vinicius Dias Lopes; Claudia Regina Gonçalves; Claudio Tavares Sacchi; Erica Valessa Ramos Gomes; Karoline Rodrigues Campos; Katia Correa de Oliveira Santos; Leonardo Jose Tadeu de Araujo |
| EPI_ISL_1628368 | Centro Medico Social Comunitario Januario Teodoro de Souza | Instituto Adolfo Lutz, Interdiciplinary Procedures Center, Strategic Laboratory | Caio Vinicius Dias Lopes; Claudia Regina Gonçalves; Claudio Tavares Sacchi; Erica Valessa Ramos Gomes; Karoline Rodrigues Campos; Katia Correa de Oliveira Santos; Leonardo Jose Tadeu de Araujo |
| EPI_ISL_2274030, EPI_ISL_2274031, EPI_ISL_2274032, EPI_ISL_2274034, EPI_ISL_2274035, EPI_ISL_2274037, EPI_ISL_2274038, EPI_ISL_2274039 | | | |
| see above | Centro Nacional de Enfermidades Tropicales (CENETROP) | Laboratory of Respiratory Viruses and Measles, Oswaldo Cruz Institute, FIOCRUZ | Alice Sampaio Rocha; Ana Carolina Mendonca; Anna Carolina Paixao; Cinthia Avila; Elisa Cavalcante Pereira; Fernando Motta; Luciana Appolinario; Marilda Siqueira on behalf of the Fiocruz COVID-19 Genomic Surveillance Network; Paola Resende; Renata Serrano Lopes; Roxana Loayza; Taina Venas |
| EPI_ISL_2612325, EPI_ISL_2612358, EPI_ISL_2612363, EPI_ISL_2612365, EPI_ISL_2612366, EPI_ISL_2612372, EPI_ISL_2612373, EPI_ISL_2612377, EPI_ISL_2612378, EPI_ISL_2612379, EPI_ISL_2612380, EPI_ISL_2612391, EPI_ISL_2612392, EPI_ISL_2612398, EPI_ISL_2612399, EPI_ISL_2612400, EPI_ISL_2612402, EPI_ISL_2612407, EPI_ISL_2612408 | | | |
| see above | Centro de Infectologia Charles Mérieux/ Laboratório Rodolphe Mérieux, FUNDHACRE | Bioinformatics Laboratory / LNCC | Alessandra P Lamarca; Alexandra L Gerber; Ana Paula de C Guimarães; Ana Tereza R Vasconcelos; Andreas Stocker; Cirley Maria de Oliveira Lobato; Douglas Terra Machado; Luiz Fellype Alves de Souza; Luiz G P de Almeida; Ronaldo da Silva F Jr |
| EPI_ISL_2031715, EPI_ISL_2031719, EPI_ISL_2031720, EPI_ISL_2031721, EPI_ISL_2031722, EPI_ISL_2031725, EPI_ISL_2031727, EPI_ISL_2031728, EPI_ISL_2031737, EPI_ISL_2031739, EPI_ISL_2031740, EPI_ISL_2031742, EPI_ISL_2031744 | | | |

| | | | |
|---|---|---|---|
| see above | Centro de Innovación en Vigilancia Epidemiológica (CiVE), Institut Pasteur Montevideo, Uruguay | Centro de Innovación en Vigilancia Epidemiológica (CiVE), Institut Pasteur Montevideo, Uruguay | Alicia Costábile; Alvaro Fajardo; Andrés Lizosaín; Belén González; Bernardina Rivera; Cecilia Alonso; Cecilia Salazar; Gonzalo Moratorio; Gregorio Iraola; Henry Albornoz; Ignacio Ferrés; Inés Bellini; Juan Zanetti; Julio Medina; Lucia Bilbao; Luciana Griffero; Lucía Spangenberg; Ma Noel Bentancor; Ma Pía Techera; Mailen Arleo; Martina Alonso; María José Benítez; Matías Maidana; Mauricio Méndez; Melissa Duquía; Mercedes Paz; Natalia Rego; Natalia Reyes; Odhille Chappos; Paula Perbolianachis; Pilar Moreno; Rodney Colina; Rodrigo Arce; Tamara Fernández; Tania Possi |
| EPI_ISL_2091312, EPI_ISL_2091317 | Centro de Investigación Biomédica de Occidente (CIBO) | Centro de Investigación en Enfermedades Infecciosas (CIENI), Instituto Nacional de Enfermedades Respiratorias (INER) | Alejandro Sanchez-Flores; Alfredo Herrera-Estrella; Alicia Ocaña-Mondragón; Angel Gustavo Salas-Lais; Bernardo Martínez-Miguel; Blanca Taboada; Brenda Irasema Maldonado-Meza; Carla Ivón Herrera-Najera; Carlos F. Arias; Celia Boukadida; Clara Esperanza Santacruz-Tinoco; Concepción Grajales-Muñiz; Consorcio Mexicano de Vigilancia Genómica (CoViGen-Mex). Authors (in alphabetical order): Julio Elias Alvarado-Yaah; Célida Duque Molina; Fernando Fontove-Herrera; Francisco Pulido; Gloria Elena Espinosa-Ayala; Gloria María Molina-Salinas; Gloria Vazquez; Hector Esteban Paz-Juárez; Hector Montoya-Fuentes; Helen Haydee Fernanda Ramírez-Plascencia; José Antonio Enciso-Moreno; José Esteban Muñoz-Medina; José de Jesús Nuñez-Contreras; Juan Bautista Chale-Dzul; Luis Alberto Ochoa-Carrera; Margarita Matías-Florentino; María Guadalupe Santiago-Mauricio; María Guadalupe de Jesús Mireles-Rivera; Nelly Sélem-Mojica; Pavel Isa; Ricardo Grande; Santiago Ávila-Ríos; Victor Eduardo Garcia-Arias; Víctor Hugo Borja-Aburto |
| EPI_ISL_2402153 | Centro de Investigación Biomédica de Occidente (CIBO) | Unidad de Genomica Avanzada | Alejandro Sanchez-Flores; Alfredo Herrera-Estrella; Alicia Ocana-Mondragon; Angel Gustavo Salas-Lais; Bernardo Martinez-Miguel; Blanca Taboada; Brenda Irasema Maldonado-Meza; Carla Ivon Herrera-Najera; Carlos F. Arias; Celia Boukadida; Clara Esperanza Santacruz-Tinoco; Concepcion Grajales-Muniz; Consorcio Mexicano de Vigilancia Genomica (CoViGen-Mex). Authors (in alphabetical order): Julio Elias Alvarado-Yaah; Fernando Fontove-Herrera; Francisco Pulido; Gloria Elena Espinoza-Ayala; Gloria Maria Molina-Salinas; Gloria Vazquez; Hector Esteban Paz-Juarez; Hector Montoya-Fuentes; Helen Haydee Fernanda Ramirez-Plascencia; Jorge Ivan Salinal-Nevarez; Jose Antonio Enciso-Moreno; Jose Esteban Munoz-Medina; Jose de Jesus Nunez-Contreras; Juan Bautista Chale-Dzul; Luis Alberto Ochoa-Carrera; Margarita Matias-Florentino; Maria Guadalupe Santiago-Mauricio; Maria Guadalupe de Jesus Mireles-Rivera; Nelly Selem-Mojica; Pavel Isa; Ricardo Grande; Santiago Avila-Rios; Victor Hugo Borja-Aburto |
| EPI_ISL_1279276 | Centro de Investigación Biomédica del Noreste (CIBIN) | Instituto Nacional de Enfermedades Respiratorias (INER): Centro de Investigación en Enfermedades Infecciosas (CIENI) | Alejandro Sanchez-Flores; Alfredo Herrera-Estrella; Alicia Ocaña-Mondragón; Angel Gustavo Salas-Lais; Bernardo Martínez-Miguel; Blanca Taboada; Brenda Irasema Maldonado-Meza; Carla Ivón Herrera-Najera; Carlos F. Arias; Celia Boukadida; Clara Esperanza Santacruz-Tinoco; Concepción Grajales-Muñiz; Consorcio Mexicano de Vigilancia Genómica (CoViGen-Mex). Authors (in alphabetical order): Julio Elias Alvarado-Yaah; Fernando Fontove-Herrera; Francisco Pulido; Gloria Elena Espinoza-Ayala; Gloria María Molina-Salinas; Gloria Vazquez; Hector Esteban Paz-Juárez; Hector Montoya-Fuentes; Helen Haydee Fernanda Ramírez-Plascencia; Jorge Ivan Salinal-Nevarez; José Antonio Enciso-Moreno; José Esteban Muñoz-Medina; José de Jesús Nuñez-Contreras; Juan Bautista Chale-Dzul; Luis Alberto Ochoa-Carrera; Margarita Matías-Florentino; María Guadalupe Santiago-Mauricio; María Guadalupe de Jesús Mireles-Rivera; Nelly Sélem-Mojica; Pavel Isa; Ricardo Grande; Santiago Ávila-Ríos; Víctor Hugo Borja-Aburto |
| EPI_ISL_2491713, EPI_ISL_2491714, EPI_ISL_2491715, EPI_ISL_2491716, EPI_ISL_2491717, EPI_ISL_2491718 | Centro de Pesquisa Gonçalo Moniz (CPqGM - FIOCRUZ/BA) | Laboratory of Respiratory Viruses and Measles, Oswaldo Cruz Institute, FIOCRUZ | Alice Sampaio Rocha; Ana Carolina Mendonca; Anna Carolina Paixao; Camila I. de Oliveira; Elisa Cavalcante Pereira; Fernando Motta; Luciana Appolinario; Marilda Siqueira on behalf of the Fiocruz COVID-19 Genomic Surveillance Network; Paola Resende; Renata Serrano Lopes; Ricardo Khouri; Taina Venas |
| EPI_ISL_1171648, EPI_ISL_1171649, EPI_ISL_1171650 | Centro de Saude Dr. Jose Paione em Mococa | Instituto Adolfo Lutz, Interdiciplinary Procedures Center, Strategic Laboratory | Caio Vinicius Dias Lopes; Claudia Regina Gonçalves; Claudio Tavares Sacchi; Erica Valessa Ramos Gomes; Karoline Rodrigues Campos |
| EPI_ISL_1520129 | Centro de Saude II Dr Jose Paione Mococa | Instituto Adolfo Lutz, Interdiciplinary Procedures Center, Strategic Laboratory | Caio Vinicius Dias Lopes; Claudia Regina Gonçalves; Claudio Tavares Sacchi; Erica Valessa Ramos Gomes; Karoline Rodrigues Campos |
| EPI_ISL_1196296 | Centro de Saude II Dr. Jose Paione Mococa | Instituto Adolfo Lutz, Interdiciplinary Procedures Center, Strategic Laboratory | Caio Vinicius Dias Lopes; Claudia Regina Gonçalves; Claudio Tavares Sacchi; Erica Valessa Ramos Gomes; Karoline Rodrigues Campos |
| EPI_ISL_1715134 | Centro de Saude II Ibitinga | Instituto Adolfo Lutz, Interdiciplinary Procedures Center, Strategic Laboratory | Caio Vinicius Dias Lopes; Claudia Regina Gonçalves; Claudio Tavares Sacchi; Erica Valessa Ramos Gomes; Karoline Rodrigues Campos; Katia Correa de Oliveira Santos; Leonardo Jose Tadeu de Araujo |
| EPI_ISL_1468427 | Centro de Saude II Matao | Instituto Adolfo Lutz, Interdiciplinary Procedures Center, Strategic Laboratory | Caio Vinicius Dias Lopes; Claudia Regina Gonçalves; Claudio Tavares Sacchi; Erica Valessa Ramos Gomes; Karoline Rodrigues Campos |
| EPI_ISL_1358285 | Centro de Treinamento e Referencia DST AIDS | Instituto Adolfo Lutz, Interdiciplinary Procedures Center, Strategic Laboratory | Caio Vinicius Dias Lopes; Claudia Regina Gonçalves; Claudio Tavares Sacchi; Erica Valessa Ramos Gomes; Karoline Rodrigues Campos |
| EPI_ISL_1005684 | Cerballiance Monthléry | Cerba | Benazra M; Collin T; Delaplace E; Gresset N; Haim-Boukobza S; Lecorche E; Lesenne A; Olivi M; Ricard B; Roquebert B; Trombert-Paolantoni S |
| EPI_ISL_1005685 | Cerballiance Montlhéry | Cerba | Benazra M; Collin T; Delaplace E; Gresset N; Haim-Boukobza S; Lecorche E; Lesenne A; Olivi M; Ricard B; Roquebert B; Trombert-Paolantoni S |
| EPI_ISL_1738812, EPI_ISL_1738813, EPI_ISL_1854239 | Cintramedica | Instituto Nacional de Saude (INSA) | Borges et al |
| EPI_ISL_2234845 | City of Milwaukee Health Department Laboratory | City of Milwaukee Health Department Laboratory | Amy Bauer; Jennifer Lentz; Manjeet Khubbar; Samantha Scott; Sanjib Bhattacharyya |
| EPI_ISL_1164183 | Clinical Microbiology, Nationwide Children's Hospital | James Molecular Laboratory | Amy Leber; Dan Jones; Huanyu Wang; Pam Snyder; Zachary Rudy |
| EPI_ISL_1664592, EPI_ISL_2098779 | Clinical Molecular Microbiology Laboratory, UNC Hospitals | Jeremy Wang | Alexander Rubinsteyn; Brianna de la Houssaye; Colleen Rice; Corbin Jones; Jason Smedberg; Jeremy Wang; Jose Luis Torres Castillo; Melissa Miller; Robert Hagan; Shawn Hawken |
| EPI_ISL_1617331, EPI_ISL_1617332, EPI_ISL_1665881, EPI_ISL_2195081 | Colorado Department of Public Health and Environment | Colorado Department of Public Health and Environment | Alexandria Rossheim; Diana Ir; Emily A. Travanty; Laura Bankers; Molly C. Hetherington-Rauth; Sarah Elizabeth Totten; Shannon Ely; Shannon R. Matzinger |
| EPI_ISL_1582265, EPI_ISL_1582267, EPI_ISL_1755861, EPI_ISL_1855286 | Columbia University Irving Medical Center | Wadsworth Center, New York State Department of Health | Alexis Russell; Catharine Prussing; Daryl M. Lamson; Erasmus Schneider; Erica Lasek-Nesselquist; John Kelly; Jonathan Plitnick; Kirsten St. George; Matthew Shudt; Melissa A Leisner; Navjot Singh |
| EPI_ISL_1595844 | Compass Laboratory Services, LLC | Compass Laboratory Services | PhD; William Budd |
| EPI_ISL_872191, EPI_ISL_872192, EPI_ISL_1381068 | Conjunto Hospitalar do Mandaqui de Sao Paulo | Instituto Adolfo Lutz, Interdiciplinary Procedures Center, Strategic Laboratory | Ana Lucia de Carvalho Avelino; Caio Vinicius Dias Lopes; Claudia Regina Gonçalves; Claudio Tavares Sacchi; Erica Valessa Ramos Gomes; Fabiana Cristina Pereira dos Santos; Karoline Rodrigues Campos; Katia Correa de Oliveira Santos |
| EPI_ISL_2034730 | Connecticut DPH | Yale Center for Genomic Analysis | Anderson Brito; Annie Watkins; Anthony Muyombwe; Brooke Sullivan; Chaney Kalinich; Chantal Vogels; Curt Scharfe; Irina Tikhonova; Isabel Ott; Jafar Razeq; Jessica Rothman; Joseph Fauver; Kaya Bilguvar; Mallery Breban; Mary Petrone; Nathan Grubaugh; Randy Downing; Shrikant Mane; Stephen M. Bart; Tara Alpert |
| EPI_ISL_2497667 | Contra Costa County Public Health Lab | Chan-Zuckerberg Biohub | CZB Cliahub Consortium |
| EPI_ISL_2102890 | County of San Luis Obispo Public Health Laboratory | Chan-Zuckerberg Biohub | CZB Cliahub Consortium |
| EPI_ISL_2658458, EPI_ISL_2658466 | Croatian Institute of Public Health | Croatian Institute of Public Health | Irena Tabain; Ivana Ferenčak |
| EPI_ISL_1575080 | Cruces University Hospital | Biocruces Bizkaia | Ana Belen de la Hoz Rastrollo; Mikel Gallego Rodrigo |
| EPI_ISL_3243124, EPI_ISL_3452406 | Curative Labs | New Mexico Department of Health Scientific Laboratory | Anastacia Griego-Fisher; D'eldra Malone; Ellie Johnson; Jennifer Benoit; Justin Griego; Linda Salazar; Linda Salzar; Michael Sam; Monica Manginell; Ratheesh Rajan; T. Mark Willmon |
| EPI_ISL_3664118, EPI_ISL_3664123, | DASA | DASA | Adriano Bonaldi; Angelica Hristov; Annelise Lopes; Bianca Cota; Cristina Oliveira; Jose Levi; Lidia Yamamoto; Paulo Pierry; Rodrigo Guarischi; Rodrigo Salazar |

| | | | |
|---|---|---|---|
| EPI_ISL_3664137 | | | |
| EPI_ISL_833167, EPI_ISL_833169, EPI_ISL_833170, EPI_ISL_833171, EPI_ISL_833172, EPI_ISL_833173, EPI_ISL_833174 | | | |
| see above | DB Diagnosticos do Brasil | Instituto Adolfo Lutz, Interdiciplinary Procedures Center, Strategic Laboratory | Claudia Regina Gonçalves; Claudio Tavares Sacchi; Erica Valessa Ramos Gomes; Karoline Rodrigues Campos |
| EPI_ISL_3482011 | DC Public Health Lab/ Dept. of Forensic Sciences | DC Public Health Lab/ Dept. of Forensic Sciences | Brittany Hamilton; Connie Maza; David Payne; Elizabeth Zelaya; Janis Doss; Jocelyn Hauser; Monica Mann; Sarah Scott; Scott Nguyen |
| EPI_ISL_2209152 | DEPARTAMENTO DE SAUDE COLETIVA | Instituto Butantan | Antonio Jorge Martins; Claudia Renata dos Santos Barros; David Schlesinger; Debora Botequio Moretti; Dimas Tadeu Covas; Elaine Cristina Marqueze; Elaine Vieira Santos; Evandra Strazza Rodrigues; Heidge Fukumasu; Jayme Augusto de Souza-Neto; José Salvatore Leister Patané; Luiz Alcantara; Luiz Lehmann Coutinho; Maria Carolina Elias; Maurício Lacerda Nogueira; Rafael dos Santos Bezerra; Raul Machado Neto; Rejane Maria Tommasini Grotto; Ricardo Haddad; Sandra Coccuzzo Sampaio Vessoni; Simone Kashima; Svetoslav Nanev Slavov; Vincent Louis Viala. |
| EPI_ISL_1967197 | DEPARTAMENTO DE SAUDE COLETIVA | Instituto Butantan / ESALQ-USP (Piracicaba) | Antonio Jorge Martins; Bianca Cechetto Carlos. Mendelics: Bibiana Santos; Claudia Renata dos Santos Barros; Cíntia Bittar; David Schlesinger. Hemocentro Ribeirão Preto: Simone Kashima; Debora Botequio Moretti; Elaine Cristina Marqueze; Elaine Vieira dos Santos; Elisangela Chicaroni Mattos; Erika Freitas; Evandra Strazza Rodrigues; Felipe Allan da Silva da Costa; Flavia Aburjaile; Fábio Sossai Possebon; Guilherme Campos; Guilherme Targino Valente; Heidge Fukumasu. USP-Botucatu: Rejane Maria Tommasini Grotto; Helena Lage Ferreira; Instituto Butantan: Dimas Tadeu Covas; Jardelina de Souza Todao Bernardino; Jayme A. Souza-Neto; Jessika Cristina Chagas Lesbon; Jorge A. Petroli Marchesi; José Salvatore Leister Patané; João Paulo Kitajima; João Pessoa Araújo Jr.; Leila Sabrina Ullmann; Loyze Paola Oliveira de Lima; Luiz Aurelio de Campos Crispin. Centro de Genômica Funcional da ESALQ: Luiz Lehmann Coutinho; Luiz Carlos Junior de Alcantara; Lívia Sacchetto; Maisa C. Pereira Parra; Maria Carolina Elias; Marta Giovanetti; Marília Moraes; Maurício Lacerda Nogueira. Prefeitura de Sao Paulo: Melissa Palmieri.; Patricia Akemi Assato; Paula Rahal; Paulo Inacio da Costa; Rafael dos Santos Bezerra; Raquel de Lello Rocha Campos Cassano. NGS Soluções Genômicas: Pilar Drummond Sampaio Corrêa Mariani. FZEA-USP Pirassununga: Mirele Daiana Poleti; Raul Machado Neto; Ricardo Augusto Brassaloti; Ricardo Haddad; Rodrigo Tocantins Calado. FAMERP-SJRP: Cecília Artico Banho; Sandra Coccuzzo Sampaio; Svetoslav Nanev Slavov; Vagner Fonseca; Vincent Louis Viala |
| EPI_ISL_2344545 | DEPARTAMENTO DE SAUDE COLETIVA | Instituto Butantan / UNESP-Botucatu | Antonio Jorge Martins; Claudia Renata dos Santos Barros; David Schlesinger; Debora Botequio Moretti; Dimas Tadeu Covas; Elaine Cristina Marqueze; Elaine Vieira Santos; Evandra Strazza Rodrigues; Heidge Fukumasu; Jayme Augusto de Souza-Neto; José Salvatore Leister Patané; Luiz Alcantara; Luiz Lehmann Coutinho; Maria Carolina Elias; Maurício Lacerda Nogueira; Rafael dos Santos Bezerra; Raul Machado Neto; Rejane Maria Tommasini Grotto; Ricardo Haddad; Sandra Coccuzzo Sampaio Vessoni; Simone Kashima; Svetoslav Nanev Slavov; Vincent Louis Viala |
| EPI_ISL_1795182, EPI_ISL_2345400 | DEPARTAMENTO DE SAUDE MUNICIPAL SOCORRO SP | Instituto Butantan / ESALQ-Piracicaba | Antonio Jorge Martins; Bianca Cechetto Carlos. Mendelics: Bibiana Santos; Claudia Renata dos Santos Barros; David Schlesinger; David Schlesinger. Hemocentro Ribeirão Preto: Simone Kashima; Debora Botequio Moretti; Debora Botequio Moretti. Centro de Genômica Funcional da ESALQ: Luiz Lehmann Coutinho; Dimas Tadeu Covas; Elaine Cristina Marqueze; Elaine Vieira Santos; Elaine Vieira dos Santos; Elisangela Chicaroni Mattos; Erika Freitas; Evandra Strazza Rodrigues; Felipe Allan da Silva da Costa; Flavia Aburjaile; Guilherme Targino Valente; Heidge Fukumasu; Heidge Fukumasu. USP-Botucatu: Rejane Maria Tommasini Grotto; Instituto Butantan: Alexander Roberto Precioso; Jayme A. Souza-Neto; Jayme Augusto de Souza-Neto; Jessika Cristina Chagas Lesbon; José Salvatore Leister Patané; João Paulo Kitajima; Luiz Alcantara; Luiz Carlos Junior de Alcantara; Luiz Lehmann Coutinho; Maria Carolina Elias; Marta Giovanetti; Maurício Lacerda Nogueira; Patricia Akemi Assato; Rafael dos Santos Bezerra; Raquel de Lello Rocha Campos Cassano. NGS Soluções Genômicas: Pilar Drummond Sampaio Corrêa Mariani. FZEA-USP Pirassununga: Mirele Daiana Poleti; Raul Machado Neto; Rejane Maria Tommasini Grotto; Ricardo Augusto Brassaloti; Ricardo Haddad; Rodrigo Tocantins Calado.; Sandra Coccuzzo Sampaio; Sandra Coccuzzo Sampaio Vessoni; Simone Kashima; Svetoslav Nanev Slavov; Vagner Fonseca; Vincent Louis Viala |
| EPI_ISL_2362237 | DIRETORIA MUNICIPAL DE SAUDE DE ENGENHEIRO COELHO | Instituto Butantan / FZEA-USP-Pirassununga | Antonio Jorge Martins; Claudia Renata dos Santos Barros; David Schlesinger; Debora Botequio Moretti; Dimas Tadeu Covas; Elaine Cristina Marqueze; Elaine Vieira Santos; Evandra Strazza Rodrigues; Heidge Fukumasu; Jayme Augusto de Souza-Neto; José Salvatore Leister Patané; Luiz Alcantara; Luiz Lehmann Coutinho; Maria Carolina Elias; Maurício Lacerda Nogueira; Rafael dos Santos Bezerra; Raul Machado Neto; Rejane Maria Tommasini Grotto; Ricardo Haddad; Sandra Coccuzzo Sampaio Vessoni; Simone Kashima; Svetoslav Nanev Slavov; Vincent Louis Viala |
| EPI_ISL_2820233 | DNAGYN | LGBio - Laboratório de Genética e Biodiversidade | Alex Honda Bernardes; Amanda Alves de Melo; Aparecido Divino da Cruz; Cintia Pelegrineti Targueta de Azevedo Brito; Daniela de Melo e Silva; Elisangela de Paula Silveira Lacerda; Juliana Santana de Curcio; Luiz Augusto Pereira; Marc Alexandre Duarte Gigonzac; Mariana Pires de Campos Telles; Ramilla dos Santos Braga; Renata de Oliveira Dias; Rhewter Nunes; Thais Cidália Vieira Gigonzac; Thais Guimarães Castro; Thays Millena Alves Pedroso |
| EPI_ISL_1583675 | DNA Laboratory | Central Public Health Laboratory - LACEN - Bahia, Salvador, Brazil | Arabela Leal; Breno Dominguez; Felicidade Pereira; Jaqueline Gomes; Luciana Oliveira; Luiz Alcantara; Marcela Gómez; Marta Giovanetti; Patrícia Cajado; Stephane Tosta; Vagner Fonseca; Vanessa Nardy |
| EPI_ISL_1447144 | DOHMH Corona | New York City Public Health Laboratory | Jade Wang; et al. |
| EPI_ISL_2262606 | DOHMH PHL | New York City Public Health Laboratory | Jade Wang; et al. |
| EPI_ISL_1908950 | DPHL | Delaware Public Health Lab | Rebecca Savage |
| EPI_ISL_1012923 | Department of Infectious Diseases, Istituto Superiore di Sanità, Rome, Italy; ASST Sette Laghi, Varese, Italy | Istituto Superiore di Sanità (ISS) | Alessandra Lo Presti; Andreina Baj; Angela Di Martino; Angelo Genoni; Fabrizio Maggi; Federica Novazzi; Manuela Marra; Marco Crescenzi; Maria Carollo; Paola Stefanelli; Stefano Fiore |
| EPI_ISL_1864209, EPI_ISL_1868237 | Department of Virus and Microbiological Special Diagnostics, Statens Serum Institut, Copenhagen, Denmark | Aalborg University | Danish Covid-19 Genome Consortium |
| EPI_ISL_1086034, EPI_ISL_1086037, EPI_ISL_1086038, EPI_ISL_1086039, EPI_ISL_1086040, EPI_ISL_1086041, EPI_ISL_1086042, EPI_ISL_1086043, EPI_ISL_1096121, EPI_ISL_1096135 | | | |
| see above | Diagnosticos da America - DASA | Instituto Adolfo Lutz, Interdiciplinary Procedures Center, Strategic Laboratory | Caio Vinicius Dias Lopes; Claudia Regina Gonçalves; Claudio Tavares Sacchi; Erica Valessa Ramos Gomes; Karoline Rodrigues Campos |
| EPI_ISL_1821211, EPI_ISL_1821212, EPI_ISL_1821213 | Diagnóstico da America S/A | Instituto Adolfo Lutz, Interdiciplinary Procedures Center, Strategic Laboratory | Caio Vinicius Dias Lopes; Claudia Regina Gonçalves; Claudio Tavares Sacchi; Erica Valessa Ramos Gomes; Karoline Rodrigues Campos; Leonardo Jose Tadeu de Araujo |
| EPI_ISL_1622497 | Division of Emerging Infectious Diseases, Bureau of Infectious Diseases Diagnosis Control, Korea Disease Control and Prevention Agency | Division of Emerging Infectious Diseases, Bureau of Infectious Diseases Diagnosis Control, Korea Disease Control and Prevention Agency | Ae Kyung Park; Chae Young Lee; Eun-Jin Kim; Heui Man Kim; Il-Hwan Kim; Jeong-Ah Kim; Jeong-Min Kim; Jin Sun No |
| EPI_ISL_2145333, EPI_ISL_2145336, EPI_ISL_2145338, EPI_ISL_2145424, EPI_ISL_2145438, EPI_ISL_2145475, EPI_ISL_2145478, EPI_ISL_2145479, EPI_ISL_2145481, EPI_ISL_2145588, EPI_ISL_2145589, EPI_ISL_2145630, EPI_ISL_2145632, EPI_ISL_2449986 | | | |
| see above | Dutch COVID-19 response team | Erasmus Medical Center | Anne van der Linden; Annemiek van der Eijk; Bas Oude Munnink; Corine GeurtsvanKessel; David Nieuwenhuijse; Emmanuelle Munger; Irina Chestakova; Marion Koopmans; Marjan Boter; Reina Sikkema; Richard Molenkamp; on behalf of the Dutch national COVID-19 respo |
| EPI_ISL_2030599 | Dutch COVID-19 response team | Medical Microbiology, Maastricht University Medical Centre | Brian van der Veer*; Carmen Reumkens; Christian Hoebe; Erik Beuken; Jozef Dingemans*; Lieke van Alphen; Paul Savelkoul |
| EPI_ISL_943045, EPI_ISL_943046, EPI_ISL_1014338, EPI_ISL_1035203, EPI_ISL_1165565, EPI_ISL_1289219, EPI_ISL_1289220, EPI_ISL_1289221, EPI_ISL_1289222, EPI_ISL_1289224, EPI_ISL_1370577, EPI_ISL_1370615, EPI_ISL_1370834, EPI_ISL_1370893, EPI_ISL_1370903, EPI_ISL_1371081, EPI_ISL_1371282, EPI_ISL_1371513, EPI_ISL_1371702, EPI_ISL_1371705, EPI_ISL_1456631, EPI_ISL_1456632, EPI_ISL_1456635, EPI_ISL_1456640, EPI_ISL_1456641, EPI_ISL_1456642, EPI_ISL_1456643, EPI_ISL_1456644, EPI_ISL_1456645, EPI_ISL_1456653, EPI_ISL_1456654, EPI_ISL_1456655, EPI_ISL_1456659, EPI_ISL_1456660, EPI_ISL_1456661, EPI_ISL_1521295, EPI_ISL_1521296, EPI_ISL_1521297, EPI_ISL_1521298, EPI_ISL_1521299, EPI_ISL_1521300, EPI_ISL_1521301, EPI_ISL_1521302, EPI_ISL_1521303, EPI_ISL_1521304, EPI_ISL_1521306, EPI_ISL_1521321, EPI_ISL_1521322, EPI_ISL_1521323, EPI_ISL_1521324, EPI_ISL_1521325, EPI_ISL_1521326, EPI_ISL_1521327, EPI_ISL_1521328, EPI_ISL_1521331, EPI_ISL_1521332, EPI_ISL_1521337, EPI_ISL_1596022, EPI_ISL_1596027, EPI_ISL_1596162, EPI_ISL_1596280, EPI_ISL_1596609, EPI_ISL_1596777, EPI_ISL_1596936, EPI_ISL_1597044, EPI_ISL_1597233, EPI_ISL_1597244, EPI_ISL_1597245, EPI_ISL_1597256, EPI_ISL_1597268, EPI_ISL_1597291, EPI_ISL_1597302, EPI_ISL_1597313, EPI_ISL_1597412, EPI_ISL_1597424, EPI_ISL_1597436, EPI_ISL_1597446, EPI_ISL_1597522, EPI_ISL_1597525, EPI_ISL_1597590, EPI_ISL_1703183, EPI_ISL_1703184, EPI_ISL_1703193, EPI_ISL_1703204, EPI_ISL_1703214, EPI_ISL_1703215, EPI_ISL_1703225, EPI_ISL_1703226, EPI_ISL_1703251, EPI_ISL_1703252, EPI_ISL_1703253, EPI_ISL_1703260, EPI_ISL_1703269, EPI_ISL_1703279, EPI_ISL_1703289, EPI_ISL_1703313, EPI_ISL_1703324, EPI_ISL_1703330, EPI_ISL_1703336, EPI_ISL_1703620, EPI_ISL_1704983, EPI_ISL_1705088, EPI_ISL_1705097, EPI_ISL_1705115, EPI_ISL_1705388, EPI_ISL_1705398, EPI_ISL_1705409, EPI_ISL_1705413, EPI_ISL_1705414, EPI_ISL_1705443, EPI_ISL_1705477, EPI_ISL_1705502, EPI_ISL_1705513, EPI_ISL_1705525, EPI_ISL_1705536, EPI_ISL_1705556, EPI_ISL_1705566, EPI_ISL_1705853, EPI_ISL_1705864, EPI_ISL_1792429, EPI_ISL_1792430, EPI_ISL_1792431, EPI_ISL_1792433, EPI_ISL_1792435, EPI_ISL_1792436, EPI_ISL_1792438, EPI_ISL_1792447, EPI_ISL_1792448, EPI_ISL_1792449, EPI_ISL_1792450, EPI_ISL_1792451, EPI_ISL_1792452, EPI_ISL_1792453, EPI_ISL_1961155, EPI_ISL_1962254, EPI_ISL_1962256, EPI_ISL_1962272, EPI_ISL_1962273, EPI_ISL_1962274, EPI_ISL_1962275, EPI_ISL_2093857, EPI_ISL_2475704 | | | |
| see above | Dutch COVID-19 response team | National Institute for Public Health and the Environment (RIVM) | Adam Meijer; AnneMarie van den Brandt; Annelies Kroneman; Bas van der Veer; Chantal Reusken; Dennis Schmitz; Dirk Eggink; Euníce Then; Florian Zwagemaker; Harry Vennema; James Groot; Jeroen Cremer; Jolienke Hardeman; Karim Hajji; Kim Freriks; Linda van de Nes; Lisa Wijsman; Lynn Aarts; Melissa van Tuil; Robert Kohl; Ryanne Jaarsma; Sanne Bos; Sharon van den Brink; Sjoerd Kuiling; on behalf of the national COVID-19 response team |
| EPI_ISL_4060096, EPI_ISL_4060097, EPI_ISL_4060098 | Dynacare Manitoba | Manitoba Cadham Provincial Laboratory | David Alexander; Hilary Racher; Hongbin Li; Melissa Desaulnier; Tintu Abraham |
| EPI_ISL_1795245, EPI_ISL_1795246, EPI_ISL_1795247, EPI_ISL_2345478, EPI_ISL_2345480, EPI_ISL_2345481 | EMERGENCIA RESPIRATORIA DE NOVA GRANADA | Instituto Butantan / ESALQ-Piracicaba | Antonio Jorge Martins; Bianca Cechetto Carlos. Mendelics: Bibiana Santos; Claudia Renata dos Santos Barros; David Schlesinger; David Schlesinger. Hemocentro Ribeirão Preto: Simone Kashima; Debora Botequio Moretti; Debora Botequio Moretti. Centro de Genômica Funcional da ESALQ: Luiz Lehmann Coutinho; Dimas Tadeu Covas; Elaine Cristina Marqueze; Elaine Vieira Santos; Elaine Vieira dos Santos; Elisangela Chicaroni Mattos; Erika Freitas; Evandra Strazza Rodrigues; Felipe Allan da Silva da Costa; Flavia Aburjaile; Guilherme Targino Valente; Heidge Fukumasu; Heidge Fukumasu. USP-Botucatu: Rejane Maria Tommasini Grotto; Instituto Butantan: Alexander Roberto Precioso; Jayme A. Souza-Neto; Jayme Augusto de Souza-Neto; Jessika Cristina Chagas Lesbon; José Salvatore Leister Patané; João Paulo Kitajima; Luiz Alcantara; Luiz Carlos Junior de Alcantara; Luiz Lehmann Coutinho; Maria Carolina Elias; Marta Giovanetti; Maurício Lacerda Nogueira; Patricia Akemi Assato; Rafael dos Santos Bezerra; Raquel de Lello Rocha Campos Cassano. NGS Soluções Genômicas: Pilar Drummond Sampaio Corrêa Mariani. FZEA-USP Pirassununga: Mirele Daiana Poleti; Raul Machado Neto; Rejane Maria Tommasini Grotto; Ricardo Augusto Brassaloti; Ricardo Haddad; Rodrigo Tocantins Calado.; Sandra Coccuzzo Sampaio; Sandra Coccuzzo Sampaio Vessoni; Simone Kashima; Svetoslav Nanev Slavov; Vagner Fonseca; Vincent Louis Viala |
| EPI_ISL_1966533 | EMERGENCIA RESPIRATORIA DE NOVA GRANADA | Instituto Butantan / FZEA-USP (Pirassununga) | Antonio Jorge Martins; Bianca Cechetto Carlos. Mendelics: Bibiana Santos; Claudia Renata dos Santos Barros; Cíntia Bittar; David Schlesinger. Hemocentro Ribeirão Preto: Simone Kashima; Debora Botequio Moretti; Elaine Cristina Marqueze; Elaine Vieira dos Santos; Elisangela Chicaroni Mattos; Erika Freitas; Evandra Strazza Rodrigues; Felipe Allan da Silva da Costa; Flavia Aburjaile; Fábio Sossai Possebon; Guilherme Campos; Guilherme Targino Valente; Heidge Fukumasu. USP-Botucatu: Rejane Maria Tommasini Grotto; Helena Lage Ferreira; Instituto Butantan: Dimas Tadeu Covas; Jardelina de Souza Todao Bernardino; Jayme A. Souza-Neto; Jessika Cristina Chagas Lesbon; Jorge A. Petroli Marchesi; José Salvatore Leister Patané; João Paulo Kitajima; João Pessoa Araújo Jr.; Leila Sabrina Ullmann; Loyze Paola Oliveira de Lima; Luiz Aurelio de Campos Crispin. Centro de Genômica Funcional da ESALQ: Luiz Lehmann Coutinho; Luiz Carlos Junior de Alcantara; Lívia Sacchetto; Maisa C. Pereira Parra; Maria Carolina Elias; Marta Giovanetti; Marília Moraes; Maurício Lacerda Nogueira. Prefeitura de Sao Paulo: Melissa Palmieri.; Patricia Akemi Assato; Paula Rahal; Paulo Inacio da Costa; Rafael dos Santos Bezerra; Raquel de Lello Rocha Campos Cassano. NGS Soluções Genômicas: Pilar Drummond Sampaio Corrêa Mariani. FZEA-USP Pirassununga: Mirele Daiana Poleti; Raul Machado Neto; Ricardo Augusto Brassaloti; Ricardo Haddad; Rodrigo Tocantins Calado. FAMERP-SJRP: Cecília Artico Banho; Sandra Coccuzzo Sampaio; Svetoslav Nanev Slavov; Vagner Fonseca; Vincent Louis Viala |
| EPI_ISL_1795098, | ESALQ | Instituto Butantan / | Antonio Jorge Martins; Bianca Cechetto Carlos. Mendelics: Bibiana Santos; Claudia Renata dos Santos Barros; David Schlesinger; David Schlesinger. Hemocentro Ribeirão Preto: Simone Kashima; Debora Botequio Moretti; Debora Botequio Moretti. Centro de Genômica Funcional da ESALQ: Luiz Lehmann Coutinho; Dimas Tadeu Covas; |

| | | | |
|---|---|---|---|
| EPI_ISL_1795099, EPI_ISL_2344544, EPI_ISL_2344547 | | ESALQ-Piracicaba | Elaine Cristina Marqueze; Elaine Vieira Santos; Elaine Vieira dos Santos; Elisangela Chicaroni Mattos; Erika Freitas; Evandra Strazza Rodrigues; Felipe Allan da Silva da Costa; Flavia Aburjaile; Guilherme Targino Valente; Heidge Fukumasu; Heidge Fukumasu. USP-Botucatu: Rejane Maria Tommasini Grotto; Instituto Butantan: Alexander Roberto Precioso; Jayme A. Souza-Neto; Jayme Augusto de Souza-Neto; Jessika Cristina Chagas Lesbon; José Salvatore Leister Patané; João Paulo Kitajima; Luiz Alcantara; Luiz Carlos Junior de Alcantara; Luiz Lehmann Coutinho; Maria Carolina Elias; Marta Giovanetti; Maurício Lacerda Nogueira; Patricia Akemi Assato; Rafael dos Santos Bezerra; Raquel de Lello Rocha Campos Cassano. NGS Soluções Genômicas: Pilar Drummond Sampaio Corrêa Mariani. FZEA-USP Pirassununga: Mirele Daiana Poleti; Raul Machado Neto; Rejane Maria Tommasini Grotto; Ricardo Augusto Brassaloti; Ricardo Haddad; Rodrigo Tocantins Calado.; Sandra Coccuzzo Sampaio; Sandra Coccuzzo Sampaio Vessoni; Simone Kashima; Svetoslav Nanev Slavov; Vagner Fonseca; Vincent Louis Viala |
| EPI_ISL_1966251 | ESF DR LUIS ERNESTO SANDI MORI JALES | Instituto Butantan / Mendelics | Antonio Jorge Martins; Bianca Cechetto Carlos. Mendelics: Bibiana Santos; Claudia Renata dos Santos Barros; Cíntia Bittar; David Schlesinger. Hemocentro Ribeirão Preto: Simone Kashima; Debora Botequio Moretti; Elaine Cristina Marqueze; Elaine Vieira dos Santos; Elisangela Chicaroni Mattos; Erika Freitas; Evandra Strazza Rodrigues; Felipe Allan da Silva da Costa; Flavia Aburjaile; Fábio Sossai Possebon; Guilherme Campos; Guilherme Targino Valente; Heidge Fukumasu. USP-Botucatu: Rejane Maria Tommasini Grotto; Helena Lage Ferreira; Instituto Butantan: Dimas Tadeu Covas; Jardelina de Souza Todao Bernardino; Jayme A. Souza-Neto; Jessika Cristina Chagas Lesbon; Jorge A. Petroli Marchesi; José Salvatore Leister Patané; João Paulo Kitajima; João Pessoa Araújo Jr.; Leila Sabrina Ullmann; Loyze Paola Oliveira de Lima; Luiz Aurelio de Campos Crispin. Centro de Genômica Funcional da ESALQ: Luiz Lehmann Coutinho; Luiz Carlos Junior de Alcantara; Lívia Sacchetto; Maisa C. Pereira Parra; Maria Carolina Elias; Marta Giovanetti; Marília Moraes; Maurício Lacerda Nogueira. Prefeitura de Sao Paulo: Melissa Palmieri.; Patricia Akemi Assato; Paula Rahal; Paulo Inacio da Costa; Rafael dos Santos Bezerra; Raquel de Lello Rocha Campos Cassano. NGS Soluções Genômicas: Pilar Drummond Sampaio Corrêa Mariani. FZEA-USP Pirassununga: Mirele Daiana Poleti; Raul Machado Neto; Ricardo Augusto Brassaloti; Ricardo Haddad; Rodrigo Tocantins Calado. FAMERP-SJRP: Cecília Artico Banho; Sandra Coccuzzo Sampaio; Svetoslav Nanev Slavov; Vagner Fonseca; Vincent Louis Viala |
| EPI_ISL_1795376, EPI_ISL_1795377, EPI_ISL_2345952, EPI_ISL_2345953 | ESF MINEIROS DO TIETE | Instituto Butantan / ESALQ-Piracicaba | Antonio Jorge Martins; Bianca Cechetto Carlos. Mendelics: Bibiana Santos; Claudia Renata dos Santos Barros; David Schlesinger; David Schlesinger. Hemocentro Ribeirão Preto: Simone Kashima; Debora Botequio Moretti; Debora Botequio Moretti. Centro de Genômica Funcional da ESALQ: Luiz Lehmann Coutinho; Dimas Tadeu Covas; Elaine Cristina Marqueze; Elaine Vieira Santos; Elaine Vieira dos Santos; Elisangela Chicaroni Mattos; Erika Freitas; Evandra Strazza Rodrigues; Felipe Allan da Silva da Costa; Flavia Aburjaile; Guilherme Targino Valente; Heidge Fukumasu; Heidge Fukumasu. USP-Botucatu: Rejane Maria Tommasini Grotto; Instituto Butantan: Alexander Roberto Precioso; Jayme A. Souza-Neto; Jayme Augusto de Souza-Neto; Jessika Cristina Chagas Lesbon; José Salvatore Leister Patané; João Paulo Kitajima; Luiz Alcantara; Luiz Carlos Junior de Alcantara; Luiz Lehmann Coutinho; Maria Carolina Elias; Marta Giovanetti; Maurício Lacerda Nogueira; Patricia Akemi Assato; Rafael dos Santos Bezerra; Raquel de Lello Rocha Campos Cassano. NGS Soluções Genômicas: Pilar Drummond Sampaio Corrêa Mariani. FZEA-USP Pirassununga: Mirele Daiana Poleti; Raul Machado Neto; Rejane Maria Tommasini Grotto; Ricardo Augusto Brassaloti; Ricardo Haddad; Rodrigo Tocantins Calado.; Sandra Coccuzzo Sampaio; Sandra Coccuzzo Sampaio Vessoni; Simone Kashima; Svetoslav Nanev Slavov; Vagner Fonseca; Vincent Louis Viala |
| EPI_ISL_1795248, EPI_ISL_1795249, EPI_ISL_1795250, EPI_ISL_2345482, EPI_ISL_2345483, EPI_ISL_2345484 | ESF NOVA TANABI II | Instituto Butantan / ESALQ-Piracicaba | Antonio Jorge Martins; Bianca Cechetto Carlos. Mendelics: Bibiana Santos; Claudia Renata dos Santos Barros; David Schlesinger; David Schlesinger. Hemocentro Ribeirão Preto: Simone Kashima; Debora Botequio Moretti; Debora Botequio Moretti. Centro de Genômica Funcional da ESALQ: Luiz Lehmann Coutinho; Dimas Tadeu Covas; Elaine Cristina Marqueze; Elaine Vieira Santos; Elaine Vieira dos Santos; Elisangela Chicaroni Mattos; Erika Freitas; Evandra Strazza Rodrigues; Felipe Allan da Silva da Costa; Flavia Aburjaile; Guilherme Targino Valente; Heidge Fukumasu; Heidge Fukumasu. USP-Botucatu: Rejane Maria Tommasini Grotto; Instituto Butantan: Alexander Roberto Precioso; Jayme A. Souza-Neto; Jayme Augusto de Souza-Neto; Jessika Cristina Chagas Lesbon; José Salvatore Leister Patané; João Paulo Kitajima; Luiz Alcantara; Luiz Carlos Junior de Alcantara; Luiz Lehmann Coutinho; Maria Carolina Elias; Marta Giovanetti; Maurício Lacerda Nogueira; Patricia Akemi Assato; Rafael dos Santos Bezerra; Raquel de Lello Rocha Campos Cassano. NGS Soluções Genômicas: Pilar Drummond Sampaio Corrêa Mariani. FZEA-USP Pirassununga: Mirele Daiana Poleti; Raul Machado Neto; Rejane Maria Tommasini Grotto; Ricardo Augusto Brassaloti; Ricardo Haddad; Rodrigo Tocantins Calado.; Sandra Coccuzzo Sampaio; Sandra Coccuzzo Sampaio Vessoni; Simone Kashima; Svetoslav Nanev Slavov; Vagner Fonseca; Vincent Louis Viala |
| EPI_ISL_1966853 | ESF ROSA ROSSI MAIA | Instituto Butantan / ESALQ-USP (Piracicaba) | Antonio Jorge Martins; Bianca Cechetto Carlos. Mendelics: Bibiana Santos; Claudia Renata dos Santos Barros; Cíntia Bittar; David Schlesinger. Hemocentro Ribeirão Preto: Simone Kashima; Debora Botequio Moretti; Elaine Cristina Marqueze; Elaine Vieira dos Santos; Elisangela Chicaroni Mattos; Erika Freitas; Evandra Strazza Rodrigues; Felipe Allan da Silva da Costa; Flavia Aburjaile; Fábio Sossai Possebon; Guilherme Campos; Guilherme Targino Valente; Heidge Fukumasu. USP-Botucatu: Rejane Maria Tommasini Grotto; Helena Lage Ferreira; Instituto Butantan: Dimas Tadeu Covas; Jardelina de Souza Todao Bernardino; Jayme A. Souza-Neto; Jessika Cristina Chagas Lesbon; Jorge A. Petroli Marchesi; José Salvatore Leister Patané; João Paulo Kitajima; João Pessoa Araújo Jr.; Leila Sabrina Ullmann; Loyze Paola Oliveira de Lima; Luiz Aurelio de Campos Crispin. Centro de Genômica Funcional da ESALQ: Luiz Lehmann Coutinho; Luiz Carlos Junior de Alcantara; Lívia Sacchetto; Maisa C. Pereira Parra; Maria Carolina Elias; Marta Giovanetti; Marília Moraes; Maurício Lacerda Nogueira. Prefeitura de Sao Paulo: Melissa Palmieri.; Patricia Akemi Assato; Paula Rahal; Paulo Inacio da Costa; Rafael dos Santos Bezerra; Raquel de Lello Rocha Campos Cassano. NGS Soluções Genômicas: Pilar Drummond Sampaio Corrêa Mariani. FZEA-USP Pirassununga: Mirele Daiana Poleti; Raul Machado Neto; Ricardo Augusto Brassaloti; Ricardo Haddad; Rodrigo Tocantins Calado. FAMERP-SJRP: Cecília Artico Banho; Sandra Coccuzzo Sampaio; Svetoslav Nanev Slavov; Vagner Fonseca; Vincent Louis Viala |
| EPI_ISL_3215463, EPI_ISL_3215466, EPI_ISL_3215596, EPI_ISL_3215666, EPI_ISL_3215740 | EXCITE Lab | Andersen lab at Scripps Research | Abigail Schnapper; Angela Scioscia; Chip Schooley; David Pride; Helena Tubb; Natasha Martin Cheryl Anderson; Sawyer Farmer; Sharon Reed; Tommy Valles + SEARCH |
| EPI_ISL_2477521 | Edmonton Provincial Lab | Public Health Agency of Canada (PHAC) National Microbiology Laboratory | Buss; Croxen M; Deo A; Dieu P; E; Ferrato C; Gill K; Khan F; Koleva P; Li V; Lloyd C; Lynch T; Ma R; Murphy S; Pabbaraju K; Shokoples S; Thayer J; Tipples G; Whitehouse M; Wong A; Yu C; Zelyas N |
| EPI_ISL_2107410, EPI_ISL_2107414, EPI_ISL_2107415 | Emory Molecular Diagnostics Laboratory, Emory Healthcare | Piantadosi Lab, Emory Department of Pathology | Ahmed Babiker; Anne Piantadosi |
| EPI_ISL_2112527 | Epidemiology of Microbial Diseases, Yale School of Public Health | Epidemiology of Microbial Diseases, Yale School of Public Health | A.E.; A.F.; Alpert, T.; Breban; Brito; C.B.; C.C.; Fauver; Grubaugh; I.M.; Iwasaki, A.; J.E.; J.R.; Kalinich; Landry; Lucas, C.; M.E.; M.I.; M.L.; N.D.; Ott; Petrone; Rothman; Vogels; Watkins |
| EPI_ISL_1571326, EPI_ISL_1845999, EPI_ISL_2110509, EPI_ISL_2110530, EPI_ISL_2110534, EPI_ISL_2110542, EPI_ISL_2110571, EPI_ISL_2110576, EPI_ISL_2123732 | | | |
| see above | Eurofins LifeCodexx GmbH | Robert Koch Institute | |
| EPI_ISL_3354635 | FL Bur. of Public Health Laboratories-Jacksonville | Centers for Disease Control and Prevention Division of Viral Diseases, Pathogen Discovery | Alex Burgin; Ben Rambo-Martin; Clinton Paden; Dakota Howard; Dave Wentworth; Dhwani Batra; Jasmine Padilla; Justin Lee; Krista Queen; Kristen Knipe; Kristine Lacek; Mark Burroughs; Matthew Schmerer; Meghan Bentz; Mili Sheth; Peter Cook; Sam Shepard; Sarah Nobles; Suxiang Tong; Vivien Dugan; Yvette Unoarumhi |
| EPI_ISL_2018168, EPI_ISL_3354607 | FL Bureau of Public Health Laboratories-Miami | Centers for Disease Control and Prevention Division of Viral Diseases, Pathogen Discovery | Alex Burgin; Alison Laufer Halpin; Ben L. Rambo-Martin; Ben Rambo-Martin; Clinton Paden; Clinton R. Paden; Dakota Howard; Darlene Wagner; Dave Wentworth; Dhwani Batra; Jasmine Padilla; Justin Lee; Katie Dillon; Krista Queen; Kristen Knipe; Kristine Lacek; Mark Burroughs; Matthew Schmerer; Meghan Bentz; Mili Sheth; Peter Cook; Sam Shepard; Sarah Nobles; Shoshona Le; Suxiang Tong; Vivien Dugan; Yvette Unoarumhi |
| EPI_ISL_854594 | Faroese National Reference Laboratory for Fish and Animal Diseases | Faroese National Reference Laboratory for Fish and Animal Diseases | Arnfinnur Kallsberg Junior; Debes Hammershaimb Christiansen; Maria Marjunardóttir Dahl; Petra Elisabeth Petersen |
| EPI_ISL_2156293 | First Aide Diagnostic Center | Philippine Genome Center | Alethea R. de Guzman; Anna Ong-Lim; Arianne A. Zamora; Asia Louisa U. Chong; Benedict A. Maralit; Candice Francheska B. Tambaoan; Carlo M. Lapid; Celia Carlos; Devon Ray Pacial; Edsel Maurice Salvaña; El King D. Morado; Elcid Aaron R. Pangilinan; Eva Maria Cutiongco-de la Paz; Francis A. Tablizo; Irish Coleen A. Asin; Jaime C. Montoya; Jan Michael C. Yap; Jo-Hannah S. Llames; John Q. Wong; Joshua Gregor A. Dizon; Juan Antonio R. Magalang; Karol Sophia Agape R. Padilla; Kenneth M. Kim; Kris P. Punayan; Marc Edsel C. Ayes; Marc Jerrone R. Castro; Maria Rosario Singh-Vergeire and Cynthia P. Saloma; Maria Sofia L. Yangzon; Marissa Alejandria; Razel Nikka M. Hao; Rianna Patricia S. Cruz; Sheila Mae M. Araiza |
| EPI_ISL_1652237 | Fleury | Instituto Butantan / Mendelics | Alexander Roberto Precioso; Antonio Jorge Martins; Bibiana Santos; Claudia Renata dos Santos Barros; David Schlesinger; Debora Botequio Moretti; Dimas Tadeu Covas; Elaine Cristina Marqueze; Elaine Vieira dos Santos; Erika Freitas; Evandra Strazza Rodrigues; Flavia Aburjaile; José Salvatore Leister Patané; João Paulo Kitajima; Luiz Carlos Junior de Alcantara; Maria Carolina Elias; Marta Giovanetti; Rafael dos Santos Bezerra; Raul Machado Neto; Ricardo Haddad; Rodrigo Tocantins Calado.; Sandra Coccuzzo Sampaio; Simone Kashima; Svetoslav Nanev Slavov; Vagner Fonseca; Vincent Louis Viala |
| EPI_ISL_1239974, EPI_ISL_1531237, EPI_ISL_1651496, EPI_ISL_1651531, EPI_ISL_2008652, EPI_ISL_2008659, EPI_ISL_2008709, EPI_ISL_2008742, EPI_ISL_2008744, EPI_ISL_2008745, EPI_ISL_2008827, EPI_ISL_2229962, EPI_ISL_2229989, EPI_ISL_2277955, EPI_ISL_2442426, EPI_ISL_2836047, EPI_ISL_2836377 | | | |
| see above | Florida Bureau of Public Health Laboratories | Florida Bureau of Public Health Laboratories | Jason Blanton; Sarah Schmedes |
| EPI_ISL_1554964, EPI_ISL_1554965, EPI_ISL_1555018, EPI_ISL_1555035, EPI_ISL_1555060, EPI_ISL_1555064, EPI_ISL_1555082, EPI_ISL_1555099, EPI_ISL_1555137, EPI_ISL_1555146, EPI_ISL_1555187, EPI_ISL_1555264, EPI_ISL_1555282, EPI_ISL_1555330, EPI_ISL_1555406, EPI_ISL_1555504, EPI_ISL_1555538, EPI_ISL_1555591, EPI_ISL_1555674, EPI_ISL_1555688, EPI_ISL_1555693, EPI_ISL_1555728, EPI_ISL_1555759, EPI_ISL_1555775, EPI_ISL_1555901, EPI_ISL_1555910, EPI_ISL_1555974, EPI_ISL_1556010, EPI_ISL_1556163, EPI_ISL_1556170, EPI_ISL_1556202, EPI_ISL_1556218, EPI_ISL_1556224, EPI_ISL_1556227, EPI_ISL_1556267, EPI_ISL_1556324, EPI_ISL_1556395, EPI_ISL_1556396, EPI_ISL_1556452, EPI_ISL_1556486, EPI_ISL_1556646, EPI_ISL_1556649, EPI_ISL_1556682, EPI_ISL_1556720, EPI_ISL_1556729, EPI_ISL_1556737, EPI_ISL_1556787, EPI_ISL_1556805, EPI_ISL_1556907, EPI_ISL_1556918, EPI_ISL_1556919, EPI_ISL_1556924, EPI_ISL_1556947, EPI_ISL_1556998, EPI_ISL_1556999, EPI_ISL_1557007, EPI_ISL_1557095, EPI_ISL_1557096, EPI_ISL_1612943, EPI_ISL_1612964, EPI_ISL_1613001, EPI_ISL_1613006, EPI_ISL_1613068, EPI_ISL_1613339, EPI_ISL_1613410, EPI_ISL_1613534, EPI_ISL_1613543, EPI_ISL_1613836, EPI_ISL_1613957, EPI_ISL_1614028, EPI_ISL_1614045, EPI_ISL_1614083, EPI_ISL_1614101, EPI_ISL_1614119, EPI_ISL_1614154, EPI_ISL_1614179, EPI_ISL_1614213, EPI_ISL_1614415, EPI_ISL_1614436, EPI_ISL_1614500, EPI_ISL_1614512, EPI_ISL_1614589, EPI_ISL_1614598, EPI_ISL_1614667, EPI_ISL_1614680, EPI_ISL_1614698, EPI_ISL_1614734, EPI_ISL_1614747, EPI_ISL_1614905, EPI_ISL_1665906, EPI_ISL_1665945, EPI_ISL_1666026, EPI_ISL_1666085, EPI_ISL_1666100, EPI_ISL_1666101, EPI_ISL_1666131, EPI_ISL_1666224, EPI_ISL_1666274, EPI_ISL_1666322, EPI_ISL_1666323, EPI_ISL_1666325, EPI_ISL_1666346, EPI_ISL_1666376, EPI_ISL_1666383, EPI_ISL_1666389, EPI_ISL_1666391, EPI_ISL_1666396, EPI_ISL_1666432, EPI_ISL_1666492, EPI_ISL_1666515, EPI_ISL_1666657, EPI_ISL_1666808, EPI_ISL_1667066, EPI_ISL_1667068, EPI_ISL_1667118, EPI_ISL_1667119, EPI_ISL_1667186, EPI_ISL_1667199, EPI_ISL_1667221, EPI_ISL_1667226, EPI_ISL_1667245, EPI_ISL_1667284, EPI_ISL_1667360, EPI_ISL_1733818, EPI_ISL_1733866, EPI_ISL_1733903, EPI_ISL_1733971, EPI_ISL_1734014, EPI_ISL_1734124, EPI_ISL_1734136, EPI_ISL_1734154, EPI_ISL_1734267, EPI_ISL_1734447, EPI_ISL_1734481, EPI_ISL_1734512, EPI_ISL_1734526, EPI_ISL_1734530, EPI_ISL_1734540, EPI_ISL_1802677, EPI_ISL_1802694, EPI_ISL_1839089, EPI_ISL_1839119, EPI_ISL_1839128, EPI_ISL_1839145, EPI_ISL_1839146, EPI_ISL_1839284, EPI_ISL_1839305, EPI_ISL_1839313, EPI_ISL_1839328, EPI_ISL_1839333, EPI_ISL_1839379, EPI_ISL_1839384, EPI_ISL_1839386, EPI_ISL_1839392, EPI_ISL_1839393, EPI_ISL_1839414, EPI_ISL_1839513, EPI_ISL_1839525, EPI_ISL_1839530, EPI_ISL_1839538, EPI_ISL_1839656, EPI_ISL_1839662, EPI_ISL_1839693, EPI_ISL_1839736, EPI_ISL_1839743, EPI_ISL_1839870, EPI_ISL_1839910, EPI_ISL_1839937, EPI_ISL_1839938, EPI_ISL_1840157, EPI_ISL_1992484, EPI_ISL_1992524, EPI_ISL_1992528, EPI_ISL_1992529, EPI_ISL_2037730, EPI_ISL_2037765, EPI_ISL_2037773, EPI_ISL_2037774, EPI_ISL_2037776, EPI_ISL_2037784, EPI_ISL_2037786, EPI_ISL_2037823, EPI_ISL_2037875, EPI_ISL_2037951, EPI_ISL_2037956, EPI_ISL_2037963, EPI_ISL_2037966, EPI_ISL_2037999, EPI_ISL_2038010, EPI_ISL_2038036, EPI_ISL_2038042, EPI_ISL_2038059, EPI_ISL_2038062, EPI_ISL_2038083, EPI_ISL_2038087, EPI_ISL_2038103, EPI_ISL_2038129, EPI_ISL_2038141, EPI_ISL_2038145, EPI_ISL_2038155, EPI_ISL_2038161, EPI_ISL_2038174, EPI_ISL_2038214, EPI_ISL_2038232, EPI_ISL_2038235, EPI_ISL_2038255, EPI_ISL_2038310, EPI_ISL_2038398, EPI_ISL_2038399, EPI_ISL_2038400, EPI_ISL_2038434, EPI_ISL_2038440, EPI_ISL_2038456, EPI_ISL_2038473, EPI_ISL_2038491, EPI_ISL_2038549, EPI_ISL_2038576, EPI_ISL_2038645, EPI_ISL_2038670, EPI_ISL_2038683, EPI_ISL_2038712, EPI_ISL_2038734, EPI_ISL_2038761 | | | |
| see above | Fulgent Genetics | Centers for Disease Control and Prevention Division of Viral Diseases, Pathogen Discovery | Adrian Paskey; Becky Tsai; Benafsh Sapra; Benjamin Rambo-Martin; Christopher Gulvick; Clinton R. Paden; Dakota Howard; Darlene Wagner; Dhwani Batra; Doreen Ng; Duncan MacCannell; Harry Gao; James Xie; Jason Caravas; John Gao; Joseph Fierro; Kara Moser; Matthew Schmerer; Mickey Li; Peter W. Cook; Scott Sammons; Shatavia Morrison; Yan Meng; Yvette Unoarumhi |
| EPI_ISL_1937097, EPI_ISL_1937161, EPI_ISL_1937397 | Fulgent Genetics | Fulgent Genetics | Becky Tsai; Benafsh Sapra; Doreen Ng; Harry Gao; James Xie; John Gao; Joseph Fierro; Mickey Li; Yan Meng |
| EPI_ISL_1239137, EPI_ISL_1239138 | Fundação Ezequiel Dias | Coordenação Geral de Laboratórios de Saúde Pública (CGLAB) | ; Vagner Fonseca et al |
| EPI_ISL_2249397, EPI_ISL_2293021, EPI_ISL_2293022 | Fundação Ezequiel Dias | Coordenação Geral de Laboratórios de Saúde Pública (CGLAB/DAEVS/SVS/MS) | Vagner Fonseca; et al. |
| EPI_ISL_1182543, EPI_ISL_1182544, EPI_ISL_1182545 | Fundação Ezequiel Dias (FUNED) | Coordenação Geral de Laboratórios de Saúde Pública (CGLAB/DAEVS/SVS/MS) | Vagner Fonseca; et al. |
| EPI_ISL_3553531, EPI_ISL_3553532, EPI_ISL_3553533, EPI_ISL_3553534, EPI_ISL_3553535, EPI_ISL_3553536, EPI_ISL_3553537, EPI_ISL_3553538, EPI_ISL_3553539, EPI_ISL_3553540, EPI_ISL_3553541, EPI_ISL_3553542, EPI_ISL_3553543, EPI_ISL_3553544, EPI_ISL_3553545, EPI_ISL_3553546, EPI_ISL_3553547, EPI_ISL_3553548, EPI_ISL_3553549, EPI_ISL_3553550, EPI_ISL_3553551, EPI_ISL_3553552, | | | |

| | | | |
|---|---|---|---|
| EPI_ISL_3553553, EPI_ISL_3553554, EPI_ISL_3553555, EPI_ISL_3553556, EPI_ISL_3553557, EPI_ISL_3553558, EPI_ISL_3553559, EPI_ISL_3553560, EPI_ISL_3553561, EPI_ISL_3553562, EPI_ISL_3553563, EPI_ISL_3553564, EPI_ISL_3553565, EPI_ISL_3553566, EPI_ISL_3553567, EPI_ISL_3553568, EPI_ISL_3553569, EPI_ISL_3553570, EPI_ISL_3553571, EPI_ISL_3553572, EPI_ISL_3553573, EPI_ISL_3553574, EPI_ISL_3553575, EPI_ISL_3553576, EPI_ISL_3553577, EPI_ISL_3553578, EPI_ISL_3553579, EPI_ISL_3553580, EPI_ISL_3553581, EPI_ISL_3553582, EPI_ISL_3553583, EPI_ISL_3553688 | | | |
| see above | Fundação Ezequiel Dias (FUNED) | Fundação Ezequiel Dias | Andre Leal; Cynthia Vazquez; Elaine Cristina; Felipe Iani; Flavia Aburjaile; Gislene Garcia de Castro Lichs; Glauco Carvalho; Hegger Fritsch; Joilson Xavier; Luiz Alcantara.; Luiz Henrique Ferraz Demarchi; Luiz Takao Watanabe; Marina Castilhos Souza Umaki Zardin; Marta Giovanetti; Natalia Guimaraes; Raquel da Silva Ferreira; Talita Adelino; Vagner Fonseca; de Oliveira |
| EPI_ISL_2551523, EPI_ISL_2551524, EPI_ISL_2551530 | Fundação Ezequiel Dias - FUNED | Laboratório de Virologia Clínica e Molecular | Alex Fiorini; Ana Paula Salles Fernandes; Bruna Larotonda Telezynski; Danielle Bruna Leal Oliveira; Edison Luiz Durigon; Erick Gustavo Dorlass; Flavio Fonseca e Santuza Teixeira; Guilherme Pereira Scagion; Helena Perez Coelho; Hugo Sato; Karine Lima Lourenço; Luciano Matsumiya Thomazelli; Renata Peixoto; Rubens Daniel Miserani Magalhães; Tatiana Ometto |
| EPI_ISL_1708318, EPI_ISL_2499748 | Fundação Hospitalar de Hematologia e Hemoterapia do Amazonas | Laboratório de Estudos de Vírus Emergentes | Adriana S. S. Duarte; Alessandro S. Farias; Arilson Bernardo S. P. Gomes; Audrey B. Zangirolami; Bruno D. Benites; Camila A.M. Silva; Camila L. Simeoni; Camila Lopes Simeoni; Carolina Costa-Lima; Cecilia C. Camilo; Chieh-Hsi Wu; Christopher Dye; Clarice W. Arns; Daniel A. Toledo-Teixeira; Daniel Augusto de Toledo-Teixeira; Darlan S. Candido; Erika R. Manuli; Esmenia C. Rocha; Ester C. Sabino; Ester Cerdeira Sabino; Fabia; Fabiana Granja; Fernando R. Spilki; Flavia C. Sales; Gabriela Fabiano de Souza; Giulia M. Ferreira; Grazielle C. Maktura; Henrique Marques-Souza; Ingra M. Claro; Jaqueline G. de Jesus; José Luiz Proença Módena; Julia Forato; Karina Bispo dos Santos; Karina Bispo-dos-Santos; Lais D. Coimbra; Lais Durço Coimbra; Leandro M. Souza; Lucas A.M. Franco; Lucas I. Buscaratti; Luciana S. Mofatto; Luciana Souto Mofatto; Magnun N.N. Santos; Marcelo A.S. Mori; Marcelo Addas-Carvalho; Maria L. Moretti; Maria Luiza Moretti; Mariana C. Pinho; Mariana S. Ramundo; Mariene R. Amorim; Mariene Ribeiro Amorim; Michael S. Diamond; Myuki A.E. Crispim; Natalia S. Brunetti; Nelson Gaburo; Nuno R. Faria; Nuno Rodrigues Faria; Oliver G. Pybus; Pamela S. Andrade; Pierina L. Parise; Pierina Lorencini Parise; Priscilla P. Barbosa; Priscilla Paschoal Barbosa; Rafael E. Marques; Renata Sesti-Costa; Rodrigo N. Angerami; Rodrigo Nogueira Angerami; Stefanie Primon Muraro; Thais M. Coletti; Vitor A. Costa; William M. Souza; William Marciel de Souza |
| EPI_ISL_1579172 | Furst Medical Laboratory | Norwegian Institute of Public Health, Department of Virology | Atiya R Ali; Debech Nadia; Engebretsen Serina Beate; Garcia Llorente Ignacio; Hilde Elshaug; Hilde Vollan; Jon Bråte; Kamilla Heddeland Instefjord; Karoline Bragstad; Kathrine Stene-Johansen; Marie Paulsen Madsen; Olav Hungnes; Pedersen Benedikte Nevjen; Rasmus Riis Kopperud |
| EPI_ISL_1678367, EPI_ISL_1937673 | GA Department of Public Health Laboratory | Centers for Disease Control and Prevention Division of Viral Diseases, Pathogen Discovery | Alison Laufer Halpin; Ben L. Rambo-Martin; Clinton R. Paden; Dakota Howard; Darlene Wagner; Dave Wentworth; Dhwani Batra; Jasmine Padilla; Justin Lee; Katie Dillon; Krista Queen; Kristen Knipe; Kristine Lacek; Mark Burroughs; Matthew Schmerer; Mili Sheth; Peter Cook; Sam Shepard; Sarah Nobles; Shoshona Le; Suxiang Tong; Vivien Dugan; Yvette Unoarumhi |
| EPI_ISL_1754367, EPI_ISL_1754385, EPI_ISL_1754388, EPI_ISL_1756229, EPI_ISL_1756495, EPI_ISL_1756508 | GH A.CHENEVIER-H.MONDOR | Department of Virology, Henri Mondor University Hospital, Assistance Publique Hôpitaux de Paris, Université Paris-Est Créteil, INSERM U955 | Alexandre Soulier; Christophe Rodriguez; Elisabeth Trawinski; Guillaume Gricourt; Jean-Michel Pawlotsky; Melissa N'Debi; Slim Fourati; Vanessa Demontant |
| EPI_ISL_1755025 | GROUPE HOSPITALIER SUD ILE DE FRANCE | Department of Virology, Henri Mondor University Hospital, Assistance Publique Hôpitaux de Paris, Université Paris-Est Créteil, INSERM U955 | Alexandre Soulier; Christophe Rodriguez; Elisabeth Trawinski; Guillaume Gricourt; Jean-Michel Pawlotsky; Melissa N'Debi; Slim Fourati; Vanessa Demontant |
| EPI_ISL_1966724 | GRUPO TECNICO DE VIGILANCIA SANITARIA E EPIDEMIOLOGICA | Instituto Butantan | Antonio Jorge Martins; Bianca Cechetto Carlos. Mendelics: Bibiana Santos; Claudia Renata dos Santos Barros; Cíntia Bittar; David Schlesinger. Hemocentro Ribeirão Preto: Simone Kashima; Debora Botequio Moretti; Elaine Cristina Marqueze; Elaine Vieira dos Santos; Elisangela Chicaroni Mattos; Erika Freitas; Evandra Strazza Rodrigues; Felipe Allan da Silva da Costa; Flavia Aburjaile; Fábio Sossai Possebon; Guilherme Campos; Guilherme Targino Valente; Heidge Fukumasu. USP-Botucatu: Rejane Maria Tommasini Grotto; Helena Lage Ferreira; Instituto Butantan: Dimas Tadeu Covas; Jardelina de Souza Todao Bernardino; Jayme A. Souza-Neto; Jessika Cristina Chagas Lesbon; Jorge A. Petroli Marchesi; José Salvatore Leister Patané; João Paulo Kitajima; João Pessoa Araújo Jr.; Leila Sabrina Ullmann; Loyze Paola Oliveira de Lima; Luiz Aurelio de Campos Crispin. Centro de Genômica Funcional da ESALQ: Luiz Lehmann Coutinho; Luiz Carlos Junior de Alcantara; Lívia Sacchetto; Maisa C. Pereira Parra; Maria Carolina Elias; Marta Giovanetti; Marília Moraes; Maurício Lacerda Nogueira. Prefeitura de Sao Paulo: Melissa Palmieri.; Patricia Akemi Assato; Paula Rahal; Paulo Inacio da Costa; Rafael dos Santos Bezerra; Raquel de Lello Rocha Campos Cassano. NGS Soluções Genômicas: Pilar Drummond Sampaio Corrêa Mariani. FZEA-USP Pirassununga: Mirele Daiana Poleti; Raul Machado Neto; Ricardo Augusto Brassaloti; Ricardo Haddad; Rodrigo Tocantins Calado. FAMERP-SJRP: Cecília Artico Banho; Sandra Coccuzzo Sampaio; Svetoslav Nanev Slavov; Vagner Fonseca; Vincent Louis Viala |
| EPI_ISL_1966835, EPI_ISL_1966836, EPI_ISL_1966838, EPI_ISL_1966839 | GRUPO TECNICO DE VIGILANCIA SANITARIA E EPIDEMIOLOGICA | Instituto Butantan / ESALQ-USP (Piracicaba) | Antonio Jorge Martins; Bianca Cechetto Carlos. Mendelics: Bibiana Santos; Claudia Renata dos Santos Barros; Cíntia Bittar; David Schlesinger. Hemocentro Ribeirão Preto: Simone Kashima; Debora Botequio Moretti; Elaine Cristina Marqueze; Elaine Vieira dos Santos; Elisangela Chicaroni Mattos; Erika Freitas; Evandra Strazza Rodrigues; Felipe Allan da Silva da Costa; Flavia Aburjaile; Fábio Sossai Possebon; Guilherme Campos; Guilherme Targino Valente; Heidge Fukumasu. USP-Botucatu: Rejane Maria Tommasini Grotto; Helena Lage Ferreira; Instituto Butantan: Dimas Tadeu Covas; Jardelina de Souza Todao Bernardino; Jayme A. Souza-Neto; Jessika Cristina Chagas Lesbon; Jorge A. Petroli Marchesi; José Salvatore Leister Patané; João Paulo Kitajima; João Pessoa Araújo Jr.; Leila Sabrina Ullmann; Loyze Paola Oliveira de Lima; Luiz Aurelio de Campos Crispin. Centro de Genômica Funcional da ESALQ: Luiz Lehmann Coutinho; Luiz Carlos Junior de Alcantara; Lívia Sacchetto; Maisa C. Pereira Parra; Maria Carolina Elias; Marta Giovanetti; Marília Moraes; Maurício Lacerda Nogueira. Prefeitura de Sao Paulo: Melissa Palmieri.; Patricia Akemi Assato; Paula Rahal; Paulo Inacio da Costa; Rafael dos Santos Bezerra; Raquel de Lello Rocha Campos Cassano. NGS Soluções Genômicas: Pilar Drummond Sampaio Corrêa Mariani. FZEA-USP Pirassununga: Mirele Daiana Poleti; Raul Machado Neto; Ricardo Augusto Brassaloti; Ricardo Haddad; Rodrigo Tocantins Calado. FAMERP-SJRP: Cecília Artico Banho; Sandra Coccuzzo Sampaio; Svetoslav Nanev Slavov; Vagner Fonseca; Vincent Louis Viala |
| EPI_ISL_2345835, EPI_ISL_2362271 | GRUPO TECNICO DE VIGILANCIA SANITARIA E EPIDEMIOLOGICA | Instituto Butantan / Mendelics | Antonio Jorge Martins; Claudia Renata dos Santos Barros; David Schlesinger; Debora Botequio Moretti; Dimas Tadeu Covas; Elaine Cristina Marqueze; Elaine Vieira Santos; Evandra Strazza Rodrigues; Heidge Fukumasu; Jayme Augusto de Souza-Neto; José Salvatore Leister Patané; Luiz Alcantara; Luiz Lehmann Coutinho; Maria Carolina Elias; Maurício Lacerda Nogueira; Rafael dos Santos Bezerra; Raul Machado Neto; Rejane Maria Tommasini Grotto; Ricardo Haddad; Sandra Coccuzzo Sampaio Vessoni; Simone Kashima; Svetoslav Nanev Slavov; Vincent Louis Viala |
| EPI_ISL_1626613, EPI_ISL_1626615, EPI_ISL_1626616, EPI_ISL_2157195, EPI_ISL_2157196, EPI_ISL_2157197, EPI_ISL_2157198, EPI_ISL_2157201 | | | |
| see above | Gencore - Universidad de los Andes | Gencore - Universidad de los Andes | Ana Maria Palacio; Cristian Barrera; David Gonzalez; Erica Salguero; Gabriela Ariza; Luisa Sacristan; Marcela Guevara; Silvia Restrepo |
| EPI_ISL_1167929, EPI_ISL_1321468, EPI_ISL_1470430, EPI_ISL_1470440, EPI_ISL_1470451, EPI_ISL_1470457, EPI_ISL_1470478, EPI_ISL_1470501, EPI_ISL_1470517, EPI_ISL_1470531, EPI_ISL_1470548, EPI_ISL_1470549, EPI_ISL_1470550, EPI_ISL_1470551, EPI_ISL_1470554, EPI_ISL_1534575, EPI_ISL_1534590, EPI_ISL_1534603, EPI_ISL_1541015, EPI_ISL_1541055, EPI_ISL_1633360, EPI_ISL_1633365, EPI_ISL_1633367, EPI_ISL_1633368, EPI_ISL_1633379, EPI_ISL_1633388, EPI_ISL_1633392, EPI_ISL_1633430, EPI_ISL_1633433, EPI_ISL_1633523, EPI_ISL_1633524, EPI_ISL_1633525, EPI_ISL_1633529, EPI_ISL_1633544, EPI_ISL_1712326, EPI_ISL_1712329, EPI_ISL_1712347, EPI_ISL_1712351, EPI_ISL_1712353, EPI_ISL_1712354, EPI_ISL_1712359, EPI_ISL_1712360, EPI_ISL_1712362, EPI_ISL_1712364, EPI_ISL_1712367, EPI_ISL_1712368, EPI_ISL_1712370, EPI_ISL_1712372, EPI_ISL_2009215, EPI_ISL_2009227, EPI_ISL_2009236, EPI_ISL_2009237, EPI_ISL_2009239, EPI_ISL_2009245, EPI_ISL_2009255, EPI_ISL_2009266, EPI_ISL_2009287, EPI_ISL_2009603, EPI_ISL_2009604, EPI_ISL_2009605, EPI_ISL_2009608, EPI_ISL_2009611, EPI_ISL_2009617, EPI_ISL_2009620, EPI_ISL_2009624, EPI_ISL_2597248, EPI_ISL_2597252, EPI_ISL_2597260, EPI_ISL_2597299, EPI_ISL_2597301, EPI_ISL_3546385 | | | |
| see above | Genetica Molecular and Subdepartamento de Virologia ISP Chile | Instituto de Salud Publica de Chile | Andres Castillo; Barbara Parra; Constanza Campano; Gisselle Barra; Jaime Lagos; Javier Tognarelli; Jorge Fernandez; Karen Orostica; Loredana Arata; Patricia Bustos; Rodrigo Fasce; Soledad Ulloa |
| EPI_ISL_1511643 | Genomic and molecular Biology Group, A.C.Camargo Cancer Center | Laboratory of Bioinformatics and Computational Biology, A.C.Camargo Cancer Center | Alexandre Defelicibus; Dirce Carraro; Giovana Torrezan; Israel Tojal |
| EPI_ISL_1399624, EPI_ISL_1399625, EPI_ISL_1399626, EPI_ISL_1739105, EPI_ISL_1739106, EPI_ISL_1739107, EPI_ISL_1739135, EPI_ISL_1739136 | | | |
| see above | Germano de Sousa | Instituto Nacional de Saude (INSA) | Borges et al |
| EPI_ISL_1854229 | Germano de Sousa | Instituto Nacional de Saude (INSA) and Instituto Gulbenkian de Ciencia (IGC) | Borges et al |
| EPI_ISL_1219136 | Gonçalo Moniz Institute, FIOCRUZ, Bahia | Laboratory of Respiratory Viruses and Measles, Oswaldo Cruz Institute, FIOCRUZ | Alice Sampaio Rocha; Ana Carolina Mendonca; Anna Carolina Paixao; Fernando Motta; Luciana Appolinario; Marilda Siqueira on behalf of the Fiocruz COVID-19 Genomic Surveillance Network; Paola Resende; Renata Serrano Lopes; Ricardo Khouri; Tiago Graf |
| EPI_ISL_2550797 | Grady Memorial Hospital, Clinical Microbiology Laboratory | Piantadosi Lab, Emory Department of Pathology | Anne Piantadosi; Dara Khosravi; Yun F (Wayne) Wang |
| EPI_ISL_2339821, EPI_ISL_2339822, EPI_ISL_2339835, EPI_ISL_2339841, EPI_ISL_2603133, EPI_ISL_2603138, EPI_ISL_2603147, EPI_ISL_2603153, EPI_ISL_2603156 | | | |
| see above | Grupo de Investigación en Enfermedades Tropicales del Ejército (GINETEJ), Laboratorio de Referencia e Investigación, Dirección de Sanidad Ejército, Bogotá, Colombia | Centro de Investigaciones en Microbiología y Biotecnología-UR (CIMBIUR), Facultad de Ciencias Naturales, Universidad del Rosario, Bogotá, Colombia | Camilo A. Correa-Cárdenas; Carolina Oliveros; Claudia Méndez; Elizabeth K. Márquez; Frank de los Santos Ortiz; Juan David Ramírez; Julie Pérez; Lorena Albarracín; Luz H. Patiño; Maria Clara Duque; Marina Muñoz; María Teresa Alvarado; Nathalia Ballesteros; Sergio Castañeda; Sergio Gutiérrez-Riveros; Yanira Romero; Zulma Cucunubá |
| EPI_ISL_1632710 | Gundersen Clinical Microbiology Laboratory | Kabara Cancer Research Institute | Craig S. Richmond; Paraic A. Kenny |
| EPI_ISL_1854240 | H Evora | Instituto Nacional de Saude (INSA) | Borges et al |
| EPI_ISL_1854241 | H Garcia de Orta | Instituto Nacional de Saude (INSA) | Borges et al |
| EPI_ISL_3102237, EPI_ISL_3102238, EPI_ISL_3102458, | H J M A HOSPITAL JOSE MARTINIANO DE ALENCAR | Analytical Competence Molecular Epidemiology Lab/ACME, Oswaldo | Cleber Furtado Aksenen; Fabio Miyajima; Fernando Braga Stehling; Francisco Eder de Moura Lopes; Jamille Maria Mendes Bezerra; Joaquim Cesar do Nascimento Sousa Junior; Joaquim César do Nascimento Sousa Junior; Pedro Miguel Carneiro Jeronimo; Suzana Porto Almeida & Lucas Delerino on behalf of COVID-19 FIOCRUZ Genomic Network; Suzana Porto Almeida e Lucas Delerino; Thais Ferreira de Oliveira; Thais de Oliveira Costa; Ticiane Cavalcante de Souza; Veridiana Pessoa Miyajima |

| | | | |
|---|---|---|---|
| EPI_ISL_3912159, EPI_ISL_3912160 | | Cruz Foundation, Ceara (FIOCRUZ CE) | |
| EPI_ISL_1854249, EPI_ISL_1854250 | H Vila Franca Xira | Instituto Nacional de Saude (INSA) and Institute of Biomedicine (iBiMed), Universidade de Aveiro | Borges et al |
| EPI_ISL_3102216, EPI_ISL_3102218, EPI_ISL_3102219, EPI_ISL_3102220 | HEMOCE CENTRO DE HEMATOLOGIA E HEMOTERAPIA | Analytical Competence Molecular Epidemiology Lab/ACME, Oswaldo Cruz Foundation, Ceara (FIOCRUZ CE) | Cleber Furtado Aksenen; Fabio Miyajima; Fernando Braga Stehling; Francisco Eder de Moura Lopes; Jamille Maria Mendes Bezerra; Joaquim César do Nascimento Sousa Junior; Pedro Miguel Carneiro Jeronimo; Suzana Porto Almeida e Lucas Delerino; Thais Ferreira de Oliveira; Thais de Oliveira Costa; Ticiane Cavalcante de Souza; Veridiana Pessoa Miyajima |
| EPI_ISL_2801308, EPI_ISL_2801309, EPI_ISL_2801322, EPI_ISL_2801365, EPI_ISL_3102332, EPI_ISL_3102338, EPI_ISL_3102362, EPI_ISL_3102371, EPI_ISL_3102402 | | | |
| see above | HEMOCE CENTRO DE HEMATOLOGIA E HEMOTERAPIA DO CEARA | Analytical Competence Molecular Epidemiology Lab/ACME, Oswaldo Cruz Foundation, Ceara (FIOCRUZ CE) | Cleber Furtado Aksenen; Cleber Furtado Aksenen e Suzana Porto Almeida; Fabio Miyajima; Fernando Braga Stehling; Francisco Eder de Moura Lopes; Jamille Maria Mendes Bezerra; Joaquim César do Nascimento Sousa Junior; Pedro Miguel Carneiro Jeronimo; Suzana Porto Almeida e Lucas Delerino; Thais Ferreira de Oliveira; Thais de Oliveira Costa; Ticiane Cavalcante de Souza; Veridiana Pessoa Miyajima |
| EPI_ISL_2801307, EPI_ISL_3102380, EPI_ISL_3102388, EPI_ISL_3102404, EPI_ISL_3912163, EPI_ISL_3912164 | HGCC HOSPITAL GERAL DR CESAR CALS | Analytical Competence Molecular Epidemiology Lab/ACME, Oswaldo Cruz Foundation, Ceara (FIOCRUZ CE) | Cleber Furtado Aksenen; Cleber Furtado Aksenen e Suzana Porto Almeida; Fabio Miyajima; Fernando Braga Stehling; Francisco Eder de Moura Lopes; Jamille Maria Mendes Bezerra; Joaquim Cesar do Nascimento Sousa Junior; Joaquim César do Nascimento Sousa Junior; Pedro Miguel Carneiro Jeronimo; Suzana Porto Almeida & Lucas Delerino on behalf of COVID-19 FIOCRUZ Genomic Network; Suzana Porto Almeida e Lucas Delerino; Thais Ferreira de Oliveira; Thais de Oliveira Costa; Ticiane Cavalcante de Souza; Veridiana Pessoa Miyajima |
| EPI_ISL_3102344, EPI_ISL_3912166 | HGF HOSPITAL GERAL DE FORTALEZA | Analytical Competence Molecular Epidemiology Lab/ACME, Oswaldo Cruz Foundation, Ceara (FIOCRUZ CE) | Cleber Furtado Aksenen; Fabio Miyajima; Fernando Braga Stehling; Francisco Eder de Moura Lopes; Jamille Maria Mendes Bezerra; Joaquim Cesar do Nascimento Sousa Junior; Joaquim César do Nascimento Sousa Junior; Pedro Miguel Carneiro Jeronimo; Suzana Porto Almeida & Lucas Delerino on behalf of COVID-19 FIOCRUZ Genomic Network; Suzana Porto Almeida e Lucas Delerino; Thais Ferreira de Oliveira; Thais de Oliveira Costa; Ticiane Cavalcante de Souza; Veridiana Pessoa Miyajima |
| EPI_ISL_3102350, EPI_ISL_3102358, EPI_ISL_3102406 | HIAS HOSPITAL INFANTIL ALBERT SABIN | Analytical Competence Molecular Epidemiology Lab/ACME, Oswaldo Cruz Foundation, Ceara (FIOCRUZ CE) | Cleber Furtado Aksenen; Fabio Miyajima; Fernando Braga Stehling; Francisco Eder de Moura Lopes; Jamille Maria Mendes Bezerra; Joaquim César do Nascimento Sousa Junior; Pedro Miguel Carneiro Jeronimo; Suzana Porto Almeida e Lucas Delerino; Thais Ferreira de Oliveira; Thais de Oliveira Costa; Ticiane Cavalcante de Souza; Veridiana Pessoa Miyajima |
| EPI_ISL_2017282, EPI_ISL_2017283, EPI_ISL_2017284, EPI_ISL_2017285, EPI_ISL_2017286, EPI_ISL_2017287, EPI_ISL_2017288, EPI_ISL_2017290, EPI_ISL_2017292, EPI_ISL_2017293, EPI_ISL_2017294, EPI_ISL_2017295, EPI_ISL_2017296, EPI_ISL_2017298, EPI_ISL_2017299, EPI_ISL_2017300, EPI_ISL_2017301, EPI_ISL_2017303, EPI_ISL_2017304, EPI_ISL_2017305, EPI_ISL_2017324, EPI_ISL_2017325, EPI_ISL_2017326, EPI_ISL_2017327, EPI_ISL_2017328, EPI_ISL_2017329, EPI_ISL_2017330, EPI_ISL_2017331, EPI_ISL_2017332, EPI_ISL_2017333, EPI_ISL_2017334, EPI_ISL_2017335, EPI_ISL_2017336, EPI_ISL_2017338, EPI_ISL_2017339, EPI_ISL_2017340, EPI_ISL_2017341, EPI_ISL_2017342, EPI_ISL_2017343, EPI_ISL_2017344, EPI_ISL_2017345, EPI_ISL_2017346, EPI_ISL_2017347, EPI_ISL_2017348, EPI_ISL_2017349, EPI_ISL_2017350, EPI_ISL_2017351, EPI_ISL_2017352, EPI_ISL_2017353, EPI_ISL_2017354, EPI_ISL_2017355, EPI_ISL_2017356, EPI_ISL_2017357, EPI_ISL_2017358, EPI_ISL_2017359, EPI_ISL_2017360, EPI_ISL_2017361, EPI_ISL_2017362, EPI_ISL_2017365, EPI_ISL_2017368, EPI_ISL_2017371, EPI_ISL_2017373, EPI_ISL_2017374, EPI_ISL_2017375, EPI_ISL_2017376, EPI_ISL_2017377, EPI_ISL_2017378, EPI_ISL_2017379, EPI_ISL_2017380, EPI_ISL_2017382, EPI_ISL_2017393, EPI_ISL_2017395, EPI_ISL_2017396, EPI_ISL_2017399, EPI_ISL_2017404, EPI_ISL_2017407, EPI_ISL_2017408, EPI_ISL_2017413, EPI_ISL_2017414, EPI_ISL_2017417, EPI_ISL_2017418, EPI_ISL_2017419, EPI_ISL_2017421, EPI_ISL_2017424, EPI_ISL_2017425, EPI_ISL_2017426, EPI_ISL_2017427, EPI_ISL_2017428, EPI_ISL_2017429, EPI_ISL_2017446, EPI_ISL_2017448, EPI_ISL_2017451, EPI_ISL_2017452, EPI_ISL_2017453, EPI_ISL_2017454, EPI_ISL_2017456, EPI_ISL_2017459, EPI_ISL_2017462, EPI_ISL_2017463, EPI_ISL_2017464, EPI_ISL_2017467, EPI_ISL_2017468, EPI_ISL_2017469, EPI_ISL_2017470, EPI_ISL_2017471, EPI_ISL_2017472, EPI_ISL_2017473, EPI_ISL_2017474, EPI_ISL_2017475, EPI_ISL_2017477, EPI_ISL_2017479, EPI_ISL_2017483, EPI_ISL_2017484, EPI_ISL_2102518, EPI_ISL_2102519, EPI_ISL_2102520, EPI_ISL_2102521, EPI_ISL_2187689, EPI_ISL_2187713, EPI_ISL_2187722, EPI_ISL_2187723, EPI_ISL_2187733, EPI_ISL_2187734, EPI_ISL_2187741, EPI_ISL_2187748, EPI_ISL_2187749, EPI_ISL_2187754, EPI_ISL_2187757, EPI_ISL_2187762, EPI_ISL_2187763, EPI_ISL_2187764, EPI_ISL_2187768, EPI_ISL_2187769, EPI_ISL_2187770, EPI_ISL_2187771, EPI_ISL_2187773, EPI_ISL_2187775, EPI_ISL_2187779, EPI_ISL_2187780, EPI_ISL_2187782, EPI_ISL_2187784, EPI_ISL_2187785, EPI_ISL_2187787, EPI_ISL_2187792, EPI_ISL_2187794, EPI_ISL_2187796, EPI_ISL_2187797, EPI_ISL_2187798, EPI_ISL_2187803, EPI_ISL_2187804, EPI_ISL_2187806, EPI_ISL_2187807, EPI_ISL_2187808, EPI_ISL_2187809, EPI_ISL_2187811, EPI_ISL_2187812, EPI_ISL_2187813, EPI_ISL_2187815, EPI_ISL_2187816, EPI_ISL_2187817, EPI_ISL_2187820, EPI_ISL_2187821, EPI_ISL_2187822, EPI_ISL_2187824, EPI_ISL_2187825, EPI_ISL_2187826, EPI_ISL_2187829, EPI_ISL_2187830, EPI_ISL_2187832, EPI_ISL_2187833, EPI_ISL_2187834, EPI_ISL_2187835, EPI_ISL_2187836, EPI_ISL_2187837, EPI_ISL_2187838, EPI_ISL_2187839, EPI_ISL_2187840, EPI_ISL_2187842, EPI_ISL_2187845, EPI_ISL_2187846, EPI_ISL_2187847, EPI_ISL_2187848, EPI_ISL_2187849, EPI_ISL_2187851, EPI_ISL_2187852, EPI_ISL_2187853, EPI_ISL_2187855, EPI_ISL_2187856, EPI_ISL_2187857, EPI_ISL_2187858, EPI_ISL_2187859, EPI_ISL_2187861, EPI_ISL_2187862, EPI_ISL_2187863, EPI_ISL_2187864, EPI_ISL_2187865, EPI_ISL_2187866, EPI_ISL_2187867, EPI_ISL_2187870, EPI_ISL_2187873, EPI_ISL_2187874, EPI_ISL_2187876, EPI_ISL_2187877, EPI_ISL_2187878, EPI_ISL_2187880, EPI_ISL_2187884, EPI_ISL_2187885, EPI_ISL_2187886, EPI_ISL_2187887, EPI_ISL_2187888, EPI_ISL_2187891, EPI_ISL_2187892, EPI_ISL_2187893, EPI_ISL_2187894, EPI_ISL_2187895, EPI_ISL_2187898, EPI_ISL_2187899, EPI_ISL_2187900, EPI_ISL_2187901, EPI_ISL_2187902, EPI_ISL_2187905, EPI_ISL_2187906, EPI_ISL_2187907, EPI_ISL_2187912, EPI_ISL_2187913, EPI_ISL_2187914, EPI_ISL_2187915, EPI_ISL_2187919, EPI_ISL_2187920, EPI_ISL_2187921, EPI_ISL_2187925, EPI_ISL_2187926, EPI_ISL_2187927, EPI_ISL_2187928, EPI_ISL_2187929, EPI_ISL_2187931, EPI_ISL_2187932, EPI_ISL_2187933, EPI_ISL_2187934, EPI_ISL_2187935, EPI_ISL_2187936, EPI_ISL_2187937, EPI_ISL_2187938, EPI_ISL_2187939, EPI_ISL_2187940, EPI_ISL_2187941, EPI_ISL_2187942, EPI_ISL_2187944, EPI_ISL_2187945, EPI_ISL_2187946, EPI_ISL_2187947, EPI_ISL_2187948, EPI_ISL_2187950, EPI_ISL_2187954, EPI_ISL_2187955, EPI_ISL_2187956, EPI_ISL_2187957, EPI_ISL_2187958, EPI_ISL_2187960, EPI_ISL_2187961, EPI_ISL_2187962, EPI_ISL_2187963, EPI_ISL_2187964, EPI_ISL_2187966, EPI_ISL_2187967, EPI_ISL_2187968, EPI_ISL_2187971, EPI_ISL_2187973, EPI_ISL_2187975, EPI_ISL_2187976, EPI_ISL_2187977, EPI_ISL_2187978, EPI_ISL_2187979, EPI_ISL_2187980, EPI_ISL_2187981, EPI_ISL_2187982, EPI_ISL_2187983, EPI_ISL_2187984, EPI_ISL_2222885, EPI_ISL_2222894, EPI_ISL_2227562, EPI_ISL_2227563, EPI_ISL_2348599, EPI_ISL_2348600, EPI_ISL_2348601, EPI_ISL_2348602, EPI_ISL_2348603, EPI_ISL_2466444, EPI_ISL_2466445, EPI_ISL_2466467, EPI_ISL_2497436 | | | |
| see above | HLAGYN - Laboratorio de Imunologia de Transplantes de Goias | HLAGYN - Laboratorio de Imunologia de Transplantes de Goias | Alessandro Leonardo Alvares Magalhaes; Daniel Ferreira de Sousa; Danielle de Paiva Rezende; Erika Lopes Rocha Batista; Fernando Antonio Vinhal dos Santos; Frederico Rodrigues Vinhal; Lucas Carlos Gomes Pereira; Paola Cristina Resende Silva; Raphael Bessa Parmigiane; Sabrina Sara Moreira Duarte |
| EPI_ISL_2801311, EPI_ISL_2801323, EPI_ISL_3102226, EPI_ISL_3102228, EPI_ISL_3102229, EPI_ISL_3102230, EPI_ISL_3102231, EPI_ISL_3102232, EPI_ISL_3102233, EPI_ISL_3102234, EPI_ISL_3102235, EPI_ISL_3102236, EPI_ISL_3102271, EPI_ISL_3102415, EPI_ISL_3102416, EPI_ISL_3102418, EPI_ISL_3102425, EPI_ISL_3102433, EPI_ISL_3102443, EPI_ISL_3102445, EPI_ISL_3102447, EPI_ISL_3102452, EPI_ISL_3102467, EPI_ISL_3102468, EPI_ISL_3102470, EPI_ISL_3102473, EPI_ISL_3912174, EPI_ISL_3912175, EPI_ISL_3912176, EPI_ISL_3912177, EPI_ISL_3912178, EPI_ISL_3912179, EPI_ISL_3912180, EPI_ISL_3912181, EPI_ISL_3912182, EPI_ISL_3912183, EPI_ISL_3912185, EPI_ISL_3912186, EPI_ISL_3912187 | | | |
| see above | HM HOSPITAL DE MESSEJANA DR CARLOS ALBERTO STUDART GOMES | Analytical Competence Molecular Epidemiology Lab/ACME, Oswaldo Cruz Foundation, Ceara (FIOCRUZ CE) | Cleber Furtado Aksenen; Cleber Furtado Aksenen e Suzana Porto Almeida; Fabio Miyajima; Fernando Braga Stehling; Francisco Eder de Moura Lopes; Jamille Maria Mendes Bezerra; Joaquim Cesar do Nascimento Sousa Junior; Joaquim César do Nascimento Sousa Junior; Pedro Miguel Carneiro Jeronimo; Suzana Porto Almeida & Lucas Delerino on behalf of COVID-19 FIOCRUZ Genomic Network; Suzana Porto Almeida e Lucas Delerino; Thais Ferreira de Oliveira; Thais de Oliveira Costa; Ticiane Cavalcante de Souza; Veridiana Pessoa Miyajima |
| EPI_ISL_1811236 | HNN | Inciensa, Instituto Costarricense de Investigación y Enseñanza en Nutrición y Salud | Pérez-Corrales C & Catalán-Marín F |
| EPI_ISL_2142666 | HOPITAL SAINT ANDRE | CNR Virus des Infections Respiratoires - France SUD | Antonin Bal; Bruno Lina; Gregory Destras; Gwendolyne Burfin; Hadrien Regue; Laurence Josset; Martine Valette; Quentin Semanas |
| EPI_ISL_2345972 | HOSP DO SERV PUB ESTFCO MORATO DE OLIVEIRA SAO PAULO | Instituto Butantan | Antonio Jorge Martins; Claudia Renata dos Santos Barros; David Schlesinger; Debora Botequio Moretti; Dimas Tadeu Covas; Elaine Cristina Marqueze; Elaine Vieira Santos; Evandra Strazza Rodrigues; Heidge Fukumasu; Jayme Augusto de Souza-Neto; José Salvatore Leister Patané; Luiz Alcantara; Luiz Lehmann Coutinho; Maria Carolina Elias; Maurício Lacerda Nogueira; Rafael dos Santos Bezerra; Raul Machado Neto; Rejane Maria Tommasini Grotto; Ricardo Haddad; Sandra Coccuzzo Sampaio Vessoni; Simone Kashima; Svetoslav Nanev Slavov; Vincent Louis Viala |
| EPI_ISL_1967093, EPI_ISL_1967094, EPI_ISL_1967095 | HOSP DO SERV PUB ESTFCO MORATO DE OLIVEIRA SAO PAULO | Instituto Butantan / ESALQ-USP (Piracicaba) | Antonio Jorge Martins; Bianca Cechetto Carlos. Mendelics: Bibiana Santos; Claudia Renata dos Santos Barros; Cíntia Bittar; David Schlesinger. Hemocentro Ribeirão Preto: Simone Kashima; Debora Botequio Moretti; Elaine Cristina Marqueze; Elaine Vieira dos Santos; Elisangela Chicaroni Mattos; Erika Freitas; Evandra Strazza Rodrigues; Felipe Allan da Silva da Costa; Flavia Aburjaile; Fábio Sossai Possebon; Guilherme Campos; Guilherme Targino Valente; Heidge Fukumasu. USP-Botucatu: Rejane Maria Tommasini Grotto; Helena Lage Ferreira; Instituto Butantan: Dimas Tadeu Covas; Jardelina de Souza Todao Bernardino; Jayme A. Souza-Neto; Jessika Cristina Chagas Lesbon; Jorge A. Petroli Marchesi; José Salvatore Leister Patané; João Paulo Kitajima; João Pessoa Araújo Jr.; Leila Sabrina Ullmann; Loyze Paola Oliveira de Lima; Luiz Aurelio de Campos Crispin. Centro de Genômica Funcional da ESALQ: Luiz Lehmann Coutinho; Luiz Carlos Junior de Alcantara; Lívia Sacchetto; Maisa C. Pereira Parra; Maria Carolina Elias; Marta Giovanetti; Marília Moraes; Maurício Lacerda Nogueira. Prefeitura de Sao Paulo: Melissa Palmieri.; Patricia Akemi Assato; Paula Rahal; Paulo Inacio da Costa; Rafael dos Santos Bezerra; Raquel de Lello Rocha Campos Cassano. NGS Soluções Genômicas: Pilar Drummond Sampaio Corrêa Mariani. FZEA-USP Pirassununga: Mirele Daiana Poleti; Raul Machado Neto; Ricardo Augusto Brassaloti; Ricardo Haddad; Rodrigo Tocantins Calado. FAMERP-SJRP: Cecília Artico Banho; Sandra Coccuzzo Sampaio; Svetoslav Nanev Slavov; Vagner Fonseca; Vincent Louis Viala |
| EPI_ISL_2209204 | HOSP E MATERNIDADE MUNICIPAL N SRA MONTE SERRAT | Instituto Butantan | Antonio Jorge Martins; Claudia Renata dos Santos Barros; David Schlesinger; Debora Botequio Moretti; Dimas Tadeu Covas; Elaine Cristina Marqueze; Elaine Vieira Santos; Evandra Strazza Rodrigues; Heidge Fukumasu; Jayme Augusto de Souza-Neto; José Salvatore Leister Patané; Luiz Alcantara; Luiz Lehmann Coutinho; Maria Carolina Elias; Maurício Lacerda Nogueira; Rafael dos Santos Bezerra; Raul Machado Neto; Rejane Maria Tommasini Grotto; Ricardo Haddad; Sandra Coccuzzo Sampaio Vessoni; Simone Kashima; Svetoslav Nanev Slavov; Vincent Louis Viala. |
| EPI_ISL_2344947 | HOSP E MATERNIDADE MUNICIPAL N SRA MONTE SERRAT | Instituto Butantan / ESALQ-Piracicaba | Antonio Jorge Martins; Claudia Renata dos Santos Barros; David Schlesinger; Debora Botequio Moretti; Dimas Tadeu Covas; Elaine Cristina Marqueze; Elaine Vieira Santos; Evandra Strazza Rodrigues; Heidge Fukumasu; Jayme Augusto de Souza-Neto; José Salvatore Leister Patané; Luiz Alcantara; Luiz Lehmann Coutinho; Maria Carolina Elias; Maurício Lacerda Nogueira; Rafael dos Santos Bezerra; Raul Machado Neto; Rejane Maria Tommasini Grotto; Ricardo Haddad; Sandra Coccuzzo Sampaio Vessoni; Simone Kashima; Svetoslav Nanev Slavov; Vincent Louis Viala |
| EPI_ISL_1967029, EPI_ISL_1967030, EPI_ISL_1967031 | HOSP E MATERNIDADE MUNICIPAL N SRA MONTE SERRAT | Instituto Butantan / ESALQ-USP (Piracicaba) | Antonio Jorge Martins; Bianca Cechetto Carlos. Mendelics: Bibiana Santos; Claudia Renata dos Santos Barros; Cíntia Bittar; David Schlesinger. Hemocentro Ribeirão Preto: Simone Kashima; Debora Botequio Moretti; Elaine Cristina Marqueze; Elaine Vieira dos Santos; Elisangela Chicaroni Mattos; Erika Freitas; Evandra Strazza Rodrigues; Felipe Allan da Silva da Costa; Flavia Aburjaile; Fábio Sossai Possebon; Guilherme Campos; Guilherme Targino Valente; Heidge Fukumasu. USP-Botucatu: Rejane Maria Tommasini Grotto; Helena Lage Ferreira; Instituto Butantan: Dimas Tadeu Covas; Jardelina de Souza Todao Bernardino; Jayme A. Souza-Neto; Jessika Cristina Chagas Lesbon; Jorge A. Petroli Marchesi; José Salvatore Leister Patané; João Paulo Kitajima; João Pessoa Araújo Jr.; Leila Sabrina Ullmann; Loyze Paola Oliveira de Lima; Luiz Aurelio de Campos Crispin. Centro de Genômica Funcional da ESALQ: Luiz Lehmann Coutinho; Luiz Carlos Junior de Alcantara; Lívia Sacchetto; Maisa C. Pereira Parra; Maria Carolina Elias; Marta Giovanetti; Marília Moraes; Maurício Lacerda Nogueira. Prefeitura de Sao Paulo: Melissa Palmieri.; Patricia Akemi Assato; Paula Rahal; Paulo Inacio da Costa; Rafael dos Santos Bezerra; Raquel de Lello Rocha Campos Cassano. NGS Soluções Genômicas: Pilar Drummond Sampaio Corrêa Mariani. FZEA-USP Pirassununga: Mirele Daiana Poleti; Raul Machado Neto; Ricardo Augusto Brassaloti; Ricardo Haddad; Rodrigo Tocantins Calado. FAMERP-SJRP: Cecília Artico Banho; Sandra Coccuzzo Sampaio; Svetoslav Nanev Slavov; Vagner Fonseca; Vincent Louis Viala |
| EPI_ISL_2170937 | HOSP E MATERNIDADE MUNICIPAL N SRA MONTE SERRAT | Instituto Butantan / Mendelics | Antonio Jorge Martins; Bianca Cechetto Carlos. Mendelics: Bibiana Santos; Claudia Renata dos Santos Barros; Cíntia Bittar; David Schlesinger. Hemocentro Ribeirão Preto: Simone Kashima; Debora Botequio Moretti; Elaine Cristina Marqueze; Elaine Vieira dos Santos; Elisangela Chicaroni Mattos; Erika Freitas; Evandra Strazza Rodrigues; Felipe Allan da Silva da Costa; Flavia Aburjaile; Fábio Sossai Possebon; Guilherme Campos; Guilherme Targino Valente; Heidge Fukumasu. USP-Botucatu: Rejane Maria Tommasini Grotto; Helena Lage Ferreira; Instituto Butantan: Dimas Tadeu Covas; Jardelina de Souza Todao Bernardino; Jayme A. Souza-Neto; Jessika Cristina Chagas Lesbon; Jorge A. Petroli Marchesi; José Salvatore Leister Patané; João Paulo Kitajima; João Pessoa Araújo Jr.; Leila Sabrina Ullmann; Loyze Paola Oliveira de Lima; Luiz Aurelio de Campos Crispin. Centro de Genômica Funcional da ESALQ: Luiz Lehmann Coutinho; Luiz Carlos Junior de Alcantara; Lívia Sacchetto; Maisa C. Pereira Parra; Maria Carolina Elias; Marta Giovanetti; Marília Moraes; Maurício Lacerda Nogueira. Prefeitura de Sao Paulo: Melissa Palmieri.; Patricia Akemi Assato; Paula Rahal; Paulo Inacio da Costa; Rafael dos Santos Bezerra; Raquel de Lello Rocha Campos Cassano. NGS Soluções Genômicas: Pilar Drummond Sampaio Corrêa Mariani. FZEA-USP Pirassununga: Mirele Daiana Poleti; Raul Machado Neto; Ricardo Augusto Brassaloti; Ricardo Haddad; Rodrigo Tocantins Calado. FAMERP-SJRP: Cecília Artico Banho; Sandra Coccuzzo Sampaio; Svetoslav Nanev Slavov; Vagner Fonseca; Vincent Louis Viala |
| EPI_ISL_2344546 | HOSP E MATERNIDADE MUNICIPAL N SRA MONTE SERRAT | Instituto Butantan / UNESP-Botucatu | Antonio Jorge Martins; Claudia Renata dos Santos Barros; David Schlesinger; Debora Botequio Moretti; Dimas Tadeu Covas; Elaine Cristina Marqueze; Elaine Vieira Santos; Evandra Strazza Rodrigues; Heidge Fukumasu; Jayme Augusto de Souza-Neto; José Salvatore Leister Patané; Luiz Alcantara; Luiz Lehmann Coutinho; Maria Carolina Elias; Maurício Lacerda Nogueira; Rafael dos Santos Bezerra; Raul Machado Neto; Rejane Maria Tommasini Grotto; Ricardo Haddad; Sandra Coccuzzo Sampaio Vessoni; Simone Kashima; Svetoslav Nanev Slavov; Vincent Louis Viala |
| EPI_ISL_3912189 | HOSP MATERN JULIA JORGE | Analytical Competence Molecular Epidemiology Lab/ACME, Oswaldo Cruz Foundation, Ceara (FIOCRUZ CE) | Cleber Furtado Aksenen; Fabio Miyajima; Fernando Braga Stehling; Francisco Eder de Moura Lopes; Jamille Maria Mendes Bezerra; Joaquim Cesar do Nascimento Sousa Junior; Pedro Miguel Carneiro Jeronimo; Suzana Porto Almeida & Lucas Delerino on behalf of COVID-19 FIOCRUZ Genomic Network; Thais Ferreira de Oliveira; Thais de Oliveira Costa; Ticiane Cavalcante de Souza; Veridiana Pessoa Miyajima |

| | | | |
|---|---|---|---|
| EPI_ISL_3102259 | HOSP MATERN SANTA IZABEL ARACOIABA | Analytical Competence Molecular Epidemiology Lab/ACME, Oswaldo Cruz Foundation, Ceara (FIOCRUZ CE) | Cleber Furtado Aksenen; Fabio Miyajima; Fernando Braga Stehling; Francisco Eder de Moura Lopes; Jamille Maria Mendes Bezerra; Joaquim César do Nascimento Sousa Junior; Pedro Miguel Carneiro Jeronimo; Suzana Porto Almeida e Lucas Delerino; Thais Ferreira de Oliveira; Thais de Oliveira Costa; Ticiane Cavalcante de Souza; Veridiana Pessoa Miyajima |
| EPI_ISL_3912190 | HOSP MATERNIDADE MARIA TEREZA DE JESUS MATEUS | Analytical Competence Molecular Epidemiology Lab/ACME, Oswaldo Cruz Foundation, Ceara (FIOCRUZ CE) | Cleber Furtado Aksenen; Fabio Miyajima; Fernando Braga Stehling; Francisco Eder de Moura Lopes; Jamille Maria Mendes Bezerra; Joaquim Cesar do Nascimento Sousa Junior; Pedro Miguel Carneiro Jeronimo; Suzana Porto Almeida & Lucas Delerino on behalf of COVID-19 FIOCRUZ Genomic Network; Thais Ferreira de Oliveira; Thais de Oliveira Costa; Ticiane Cavalcante de Souza; Veridiana Pessoa Miyajima |
| EPI_ISL_3912192 | HOSP MUN ABELARDO GADELHA DA ROCHA | Analytical Competence Molecular Epidemiology Lab/ACME, Oswaldo Cruz Foundation, Ceara (FIOCRUZ CE) | Cleber Furtado Aksenen; Fabio Miyajima; Fernando Braga Stehling; Francisco Eder de Moura Lopes; Jamille Maria Mendes Bezerra; Joaquim Cesar do Nascimento Sousa Junior; Pedro Miguel Carneiro Jeronimo; Suzana Porto Almeida & Lucas Delerino on behalf of COVID-19 FIOCRUZ Genomic Network; Thais Ferreira de Oliveira; Thais de Oliveira Costa; Ticiane Cavalcante de Souza; Veridiana Pessoa Miyajima |
| EPI_ISL_2345997, EPI_ISL_2346003, EPI_ISL_2346004 | HOSP MUN CIDADE TIRADENTES CARMEN PRUDENTE | Instituto Butantan | Antonio Jorge Martins; Claudia Renata dos Santos Barros; David Schlesinger; Debora Botequio Moretti; Dimas Tadeu Covas; Elaine Cristina Marqueze; Elaine Vieira Santos; Evandra Strazza Rodrigues; Heidge Fukumasu; Jayme Augusto de Souza-Neto; José Salvatore Leister Patané; Luiz Alcantara; Luiz Lehmann Coutinho; Maria Carolina Elias; Maurício Lacerda Nogueira; Rafael dos Santos Bezerra; Raul Machado Neto; Rejane Maria Tommasini Grotto; Ricardo Haddad; Sandra Coccuzzo Sampaio Vessoni; Simone Kashima; Svetoslav Nanev Slavov; Vincent Louis Viala |
| EPI_ISL_1967070, EPI_ISL_1967071, EPI_ISL_1967072 | HOSP MUN CIDADE TIRADENTES CARMEN PRUDENTE | Instituto Butantan / ESALQ-USP (Piracicaba) | Antonio Jorge Martins; Bianca Cechetto Carlos. Mendelics: Bibiana Santos; Claudia Renata dos Santos Barros; Cíntia Bittar; David Schlesinger. Hemocentro Ribeirão Preto: Simone Kashima; Debora Botequio Moretti; Elaine Cristina Marqueze; Elaine Vieira dos Santos; Elisangela Chicaroni Mattos; Erika Freitas; Evandra Strazza Rodrigues; Felipe Allan da Silva da Costa; Flavia Aburjaile; Fábio Sossai Possebon; Guilherme Campos; Guilherme Targino Valente; Heidge Fukumasu. USP-Botucatu: Rejane Maria Tommasini Grotto; Helena Lage Ferreira; Instituto Butantan: Dimas Tadeu Covas; Jardelina de Souza Todao Bernardino; Jayme A. Souza-Neto; Jessika Cristina Chagas Lesbon; Jorge A. Petroli Marchesi; José Salvatore Leister Patané; João Paulo Kitajima; João Pessoa Araújo Jr.; Leila Sabrina Ullmann; Loyze Paola Oliveira de Lima; Luiz Aurelio de Campos Crispin. Centro de Genômica Funcional da ESALQ: Luiz Lehmann Coutinho; Luiz Carlos Junior de Alcantara; Lívia Sacchetto; Maisa C. Pereira Parra; Maria Carolina Elias; Marta Giovanetti; Marília Moraes; Maurício Lacerda Nogueira. Prefeitura de Sao Paulo: Melissa Palmieri.; Patricia Akemi Assato; Paula Rahal; Paulo Inacio da Costa; Rafael dos Santos Bezerra; Raquel de Lello Rocha Campos Cassano. NGS Soluções Genômicas: Pilar Drummond Sampaio Corrêa Mariani. FZEA-USP Pirassununga: Mirele Daiana Poleti; Raul Machado Neto; Ricardo Augusto Brassaloti; Ricardo Haddad; Rodrigo Tocantins Calado. FAMERP-SJRP: Cecília Artico Banho; Sandra Coccuzzo Sampaio; Svetoslav Nanev Slavov; Vagner Fonseca; Vincent Louis Viala |
| EPI_ISL_1966737 | HOSP MUN JOSANIAS CASTANHA BRAGA | Instituto Butantan | Antonio Jorge Martins; Bianca Cechetto Carlos. Mendelics: Bibiana Santos; Claudia Renata dos Santos Barros; Cíntia Bittar; David Schlesinger. Hemocentro Ribeirão Preto: Simone Kashima; Debora Botequio Moretti; Elaine Cristina Marqueze; Elaine Vieira dos Santos; Elisangela Chicaroni Mattos; Erika Freitas; Evandra Strazza Rodrigues; Felipe Allan da Silva da Costa; Flavia Aburjaile; Fábio Sossai Possebon; Guilherme Campos; Guilherme Targino Valente; Heidge Fukumasu. USP-Botucatu: Rejane Maria Tommasini Grotto; Helena Lage Ferreira; Instituto Butantan: Dimas Tadeu Covas; Jardelina de Souza Todao Bernardino; Jayme A. Souza-Neto; Jessika Cristina Chagas Lesbon; Jorge A. Petroli Marchesi; José Salvatore Leister Patané; João Paulo Kitajima; João Pessoa Araújo Jr.; Leila Sabrina Ullmann; Loyze Paola Oliveira de Lima; Luiz Aurelio de Campos Crispin. Centro de Genômica Funcional da ESALQ: Luiz Lehmann Coutinho; Luiz Carlos Junior de Alcantara; Lívia Sacchetto; Maisa C. Pereira Parra; Maria Carolina Elias; Marta Giovanetti; Marília Moraes; Maurício Lacerda Nogueira. Prefeitura de Sao Paulo: Melissa Palmieri.; Patricia Akemi Assato; Paula Rahal; Paulo Inacio da Costa; Rafael dos Santos Bezerra; Raquel de Lello Rocha Campos Cassano. NGS Soluções Genômicas: Pilar Drummond Sampaio Corrêa Mariani. FZEA-USP Pirassununga: Mirele Daiana Poleti; Raul Machado Neto; Ricardo Augusto Brassaloti; Ricardo Haddad; Rodrigo Tocantins Calado. FAMERP-SJRP: Cecília Artico Banho; Sandra Coccuzzo Sampaio; Svetoslav Nanev Slavov; Vagner Fonseca; Vincent Louis Viala |
| EPI_ISL_3912193 | HOSP MUNIC ANTONIO NERY FILHO | Analytical Competence Molecular Epidemiology Lab/ACME, Oswaldo Cruz Foundation, Ceara (FIOCRUZ CE) | Cleber Furtado Aksenen; Fabio Miyajima; Fernando Braga Stehling; Francisco Eder de Moura Lopes; Jamille Maria Mendes Bezerra; Joaquim Cesar do Nascimento Sousa Junior; Pedro Miguel Carneiro Jeronimo; Suzana Porto Almeida & Lucas Delerino on behalf of COVID-19 FIOCRUZ Genomic Network; Thais Ferreira de Oliveira; Thais de Oliveira Costa; Ticiane Cavalcante de Souza; Veridiana Pessoa Miyajima |
| EPI_ISL_1967067 | HOSP PRONTO SOCORRO ITAQUERA | Instituto Butantan / ESALQ-USP (Piracicaba) | Antonio Jorge Martins; Bianca Cechetto Carlos. Mendelics: Bibiana Santos; Claudia Renata dos Santos Barros; Cíntia Bittar; David Schlesinger. Hemocentro Ribeirão Preto: Simone Kashima; Debora Botequio Moretti; Elaine Cristina Marqueze; Elaine Vieira dos Santos; Elisangela Chicaroni Mattos; Erika Freitas; Evandra Strazza Rodrigues; Felipe Allan da Silva da Costa; Flavia Aburjaile; Fábio Sossai Possebon; Guilherme Campos; Guilherme Targino Valente; Heidge Fukumasu. USP-Botucatu: Rejane Maria Tommasini Grotto; Helena Lage Ferreira; Instituto Butantan: Dimas Tadeu Covas; Jardelina de Souza Todao Bernardino; Jayme A. Souza-Neto; Jessika Cristina Chagas Lesbon; Jorge A. Petroli Marchesi; José Salvatore Leister Patané; João Paulo Kitajima; João Pessoa Araújo Jr.; Leila Sabrina Ullmann; Loyze Paola Oliveira de Lima; Luiz Aurelio de Campos Crispin. Centro de Genômica Funcional da ESALQ: Luiz Lehmann Coutinho; Luiz Carlos Junior de Alcantara; Lívia Sacchetto; Maisa C. Pereira Parra; Maria Carolina Elias; Marta Giovanetti; Marília Moraes; Maurício Lacerda Nogueira. Prefeitura de Sao Paulo: Melissa Palmieri.; Patricia Akemi Assato; Paula Rahal; Paulo Inacio da Costa; Rafael dos Santos Bezerra; Raquel de Lello Rocha Campos Cassano. NGS Soluções Genômicas: Pilar Drummond Sampaio Corrêa Mariani. FZEA-USP Pirassununga: Mirele Daiana Poleti; Raul Machado Neto; Ricardo Augusto Brassaloti; Ricardo Haddad; Rodrigo Tocantins Calado. FAMERP-SJRP: Cecília Artico Banho; Sandra Coccuzzo Sampaio; Svetoslav Nanev Slavov; Vagner Fonseca; Vincent Louis Viala |
| EPI_ISL_3102217 | HOSPITAL ANTONIO PRUDENTE | Analytical Competence Molecular Epidemiology Lab/ACME, Oswaldo Cruz Foundation, Ceara (FIOCRUZ CE) | Cleber Furtado Aksenen; Fabio Miyajima; Fernando Braga Stehling; Francisco Eder de Moura Lopes; Jamille Maria Mendes Bezerra; Joaquim César do Nascimento Sousa Junior; Pedro Miguel Carneiro Jeronimo; Suzana Porto Almeida e Lucas Delerino; Thais Ferreira de Oliveira; Thais de Oliveira Costa; Ticiane Cavalcante de Souza; Veridiana Pessoa Miyajima |
| EPI_ISL_2801321 | HOSPITAL ANTONIO ROSENO DE MATOS | Analytical Competence Molecular Epidemiology Lab/ACME, Oswaldo Cruz Foundation, Ceara (FIOCRUZ CE) | Cleber Furtado Aksenen e Suzana Porto Almeida; Fabio Miyajima; Fernando Braga Stehling; Francisco Eder de Moura Lopes; Jamille Maria Mendes Bezerra; Joaquim César do Nascimento Sousa Junior; Pedro Miguel Carneiro Jeronimo; Thais Ferreira de Oliveira; Thais de Oliveira Costa; Ticiane Cavalcante de Souza; Veridiana Pessoa Miyajima |
| EPI_ISL_2020126 | HOSPITAL CAN MISSES | HOSPITAL UNIVERSITARIO SON ESPASES | Antonio Oliver; Carla López-Causapé; Pablo Fraile-Ribot; SeqCovid |
| EPI_ISL_3102308, EPI_ISL_3102316 | HOSPITAL CURA DARS | Analytical Competence Molecular Epidemiology Lab/ACME, Oswaldo Cruz Foundation, Ceara (FIOCRUZ CE) | Cleber Furtado Aksenen; Fabio Miyajima; Fernando Braga Stehling; Francisco Eder de Moura Lopes; Jamille Maria Mendes Bezerra; Joaquim César do Nascimento Sousa Junior; Pedro Miguel Carneiro Jeronimo; Suzana Porto Almeida e Lucas Delerino; Thais Ferreira de Oliveira; Thais de Oliveira Costa; Ticiane Cavalcante de Souza; Veridiana Pessoa Miyajima |
| EPI_ISL_2346002 | HOSPITAL DE CAMPANHA COVID 19 CAIEIRAS | Instituto Butantan | Antonio Jorge Martins; Claudia Renata dos Santos Barros; David Schlesinger; Debora Botequio Moretti; Dimas Tadeu Covas; Elaine Cristina Marqueze; Elaine Vieira Santos; Evandra Strazza Rodrigues; Heidge Fukumasu; Jayme Augusto de Souza-Neto; José Salvatore Leister Patané; Luiz Alcantara; Luiz Lehmann Coutinho; Maria Carolina Elias; Maurício Lacerda Nogueira; Rafael dos Santos Bezerra; Raul Machado Neto; Rejane Maria Tommasini Grotto; Ricardo Haddad; Sandra Coccuzzo Sampaio Vessoni; Simone Kashima; Svetoslav Nanev Slavov; Vincent Louis Viala |
| EPI_ISL_2210253, EPI_ISL_2362266 | HOSPITAL DE CAMPANHA COVID 19 CAIEIRAS | Instituto Butantan / Mendelics | Antonio Jorge Martins; Claudia Renata dos Santos Barros; David Schlesinger; Debora Botequio Moretti; Dimas Tadeu Covas; Elaine Cristina Marqueze; Elaine Vieira Santos; Evandra Strazza Rodrigues; Heidge Fukumasu; Jayme Augusto de Souza-Neto; José Salvatore Leister Patané; Luiz Alcantara; Luiz Lehmann Coutinho; Maria Carolina Elias; Maurício Lacerda Nogueira; Rafael dos Santos Bezerra; Raul Machado Neto; Rejane Maria Tommasini Grotto; Ricardo Haddad; Sandra Coccuzzo Sampaio Vessoni; Simone Kashima; Svetoslav Nanev Slavov; Vincent Louis Viala |
| EPI_ISL_1795345, EPI_ISL_2345612 | HOSPITAL DE CAMPANHA COVID 19 MUNICIPIO DE TAUBATE | Instituto Butantan / ESALQ-Piracicaba | Antonio Jorge Martins; Bianca Cechetto Carlos. Mendelics: Bibiana Santos; Claudia Renata dos Santos Barros; David Schlesinger; David Schlesinger. Hemocentro Ribeirão Preto: Simone Kashima; Debora Botequio Moretti; Debora Botequio Moretti. Centro de Genômica Funcional da ESALQ: Luiz Lehmann Coutinho; Dimas Tadeu Covas; Elaine Cristina Marqueze; Elaine Vieira Santos; Elaine Vieira dos Santos; Elisangela Chicaroni Mattos; Erika Freitas; Evandra Strazza Rodrigues; Felipe Allan da Silva da Costa; Flavia Aburjaile; Guilherme Targino Valente; Heidge Fukumasu; Heidge Fukumasu. USP-Botucatu: Rejane Maria Tommasini Grotto; Instituto Butantan: Alexander Roberto Precioso; Jayme A. Souza-Neto; Jayme Augusto de Souza-Neto; Jessika Cristina Chagas Lesbon; José Salvatore Leister Patané; João Paulo Kitajima; Luiz Alcantara; Luiz Carlos Junior de Alcantara; Luiz Lehmann Coutinho; Maria Carolina Elias; Marta Giovanetti; Maurício Lacerda Nogueira; Patricia Akemi Assato; Rafael dos Santos Bezerra; Raquel de Lello Rocha Campos Cassano. NGS Soluções Genômicas: Pilar Drummond Sampaio Corrêa Mariani. FZEA-USP Pirassununga: Mirele Daiana Poleti; Raul Machado Neto; Rejane Maria Tommasini Grotto; Ricardo Augusto Brassaloti; Ricardo Haddad; Rodrigo Tocantins Calado.; Sandra Coccuzzo Sampaio; Sandra Coccuzzo Sampaio Vessoni; Simone Kashima; Svetoslav Nanev Slavov; Vagner Fonseca; Vincent Louis Viala |
| EPI_ISL_1967165, EPI_ISL_1967166, EPI_ISL_1967167, EPI_ISL_1967168, EPI_ISL_1967169, EPI_ISL_1967170, EPI_ISL_1967171, EPI_ISL_1967172, EPI_ISL_1967173, EPI_ISL_1967174, EPI_ISL_1967175, EPI_ISL_1967176, EPI_ISL_1967178, EPI_ISL_1967179, EPI_ISL_1967180, EPI_ISL_1967181, EPI_ISL_1967182, EPI_ISL_1967183, EPI_ISL_1967185, EPI_ISL_1967186, EPI_ISL_1967187, EPI_ISL_1967188, EPI_ISL_1967189, EPI_ISL_1967190, EPI_ISL_1967191 | | | |
| see above | HOSPITAL DE CAMPANHA COVID 19 MUNICIPIO DE TAUBATE | Instituto Butantan / ESALQ-USP (Piracicaba) | Antonio Jorge Martins; Bianca Cechetto Carlos. Mendelics: Bibiana Santos; Claudia Renata dos Santos Barros; Cíntia Bittar; David Schlesinger. Hemocentro Ribeirão Preto: Simone Kashima; Debora Botequio Moretti; Elaine Cristina Marqueze; Elaine Vieira dos Santos; Elisangela Chicaroni Mattos; Erika Freitas; Evandra Strazza Rodrigues; Felipe Allan da Silva da Costa; Flavia Aburjaile; Fábio Sossai Possebon; Guilherme Campos; Guilherme Targino Valente; Heidge Fukumasu. USP-Botucatu: Rejane Maria Tommasini Grotto; Helena Lage Ferreira; Instituto Butantan: Dimas Tadeu Covas; Jardelina de Souza Todao Bernardino; Jayme A. Souza-Neto; Jessika Cristina Chagas Lesbon; Jorge A. Petroli Marchesi; José Salvatore Leister Patané; João Paulo Kitajima; João Pessoa Araújo Jr.; Leila Sabrina Ullmann; Loyze Paola Oliveira de Lima; Luiz Aurelio de Campos Crispin. Centro de Genômica Funcional da ESALQ: Luiz Lehmann Coutinho; Luiz Carlos Junior de Alcantara; Lívia Sacchetto; Maisa C. Pereira Parra; Maria Carolina Elias; Marta Giovanetti; Marília Moraes; Maurício Lacerda Nogueira. Prefeitura de Sao Paulo: Melissa Palmieri.; Patricia Akemi Assato; Paula Rahal; Paulo Inacio da Costa; Rafael dos Santos Bezerra; Raquel de Lello Rocha Campos Cassano. NGS Soluções Genômicas: Pilar Drummond Sampaio Corrêa Mariani. FZEA-USP Pirassununga: Mirele Daiana Poleti; Raul Machado Neto; Ricardo Augusto Brassaloti; Ricardo Haddad; Rodrigo Tocantins Calado. FAMERP-SJRP: Cecília Artico Banho; Sandra Coccuzzo Sampaio; Svetoslav Nanev Slavov; Vagner Fonseca; Vincent Louis Viala |
| EPI_ISL_3102272 | HOSPITAL DE PEQUENO PORTE SAO FRANCISCO SALITRE | Analytical Competence Molecular Epidemiology Lab/ACME, Oswaldo Cruz Foundation, Ceara (FIOCRUZ CE) | Cleber Furtado Aksenen; Fabio Miyajima; Fernando Braga Stehling; Francisco Eder de Moura Lopes; Jamille Maria Mendes Bezerra; Joaquim César do Nascimento Sousa Junior; Pedro Miguel Carneiro Jeronimo; Suzana Porto Almeida e Lucas Delerino; Thais Ferreira de Oliveira; Thais de Oliveira Costa; Ticiane Cavalcante de Souza; Veridiana Pessoa Miyajima |
| EPI_ISL_1795101, EPI_ISL_1795106, EPI_ISL_2344550, EPI_ISL_2344556 | HOSPITAL DOS FORNCEDORES | Instituto Butantan / ESALQ-Piracicaba | Antonio Jorge Martins; Bianca Cechetto Carlos. Mendelics: Bibiana Santos; Claudia Renata dos Santos Barros; David Schlesinger; David Schlesinger. Hemocentro Ribeirão Preto: Simone Kashima; Debora Botequio Moretti; Debora Botequio Moretti. Centro de Genômica Funcional da ESALQ: Luiz Lehmann Coutinho; Dimas Tadeu Covas; Elaine Cristina Marqueze; Elaine Vieira Santos; Elaine Vieira dos Santos; Elisangela Chicaroni Mattos; Erika Freitas; Evandra Strazza Rodrigues; Felipe Allan da Silva da Costa; Flavia Aburjaile; Guilherme Targino Valente; Heidge Fukumasu; Heidge Fukumasu. USP-Botucatu: Rejane Maria Tommasini Grotto; Instituto Butantan: Alexander Roberto Precioso; Jayme A. Souza-Neto; Jayme Augusto de Souza-Neto; Jessika Cristina Chagas Lesbon; José Salvatore Leister Patané; João Paulo Kitajima; Luiz Alcantara; Luiz Carlos Junior de Alcantara; Luiz Lehmann Coutinho; Maria Carolina Elias; Marta Giovanetti; Maurício Lacerda Nogueira; Patricia Akemi Assato; Rafael dos Santos Bezerra; Raquel de Lello Rocha Campos Cassano. NGS Soluções Genômicas: Pilar Drummond Sampaio Corrêa Mariani. FZEA-USP Pirassununga: Mirele Daiana Poleti; Raul Machado Neto; Rejane Maria Tommasini Grotto; Ricardo Augusto Brassaloti; Ricardo Haddad; Rodrigo Tocantins Calado.; Sandra Coccuzzo Sampaio; Sandra Coccuzzo Sampaio Vessoni; Simone Kashima; Svetoslav Nanev Slavov; Vagner Fonseca; Vincent Louis Viala |
| EPI_ISL_3102321 | HOSPITAL DR ALBERTO FEITOSA LIMA | Analytical Competence Molecular Epidemiology Lab/ACME, Oswaldo Cruz Foundation, Ceara (FIOCRUZ CE) | Cleber Furtado Aksenen; Fabio Miyajima; Fernando Braga Stehling; Francisco Eder de Moura Lopes; Jamille Maria Mendes Bezerra; Joaquim César do Nascimento Sousa Junior; Pedro Miguel Carneiro Jeronimo; Suzana Porto Almeida e Lucas Delerino; Thais Ferreira de Oliveira; Thais de Oliveira Costa; Ticiane Cavalcante de Souza; Veridiana Pessoa Miyajima |
| EPI_ISL_1712404, EPI_ISL_2272959 | HOSPITAL DR. ENRIQUE BALTODANO BRICEÑO | Inciensa, Instituto Costarricense de Investigación y Enseñanza en Nutrición y Salud | Adriana Godínez; Claudio Soto-Garita; Estela Cordero; Francisco Duarte; Hebleen Porras; Joselyn Prado & Adriana Bermúdez-Espinoza; Joselyn Prado & Arnoldo Zamora-Villegas; José Luis Vargas; Mariela Gutiérrez; Melany Calderón |
| EPI_ISL_2272986 | HOSPITAL DR. MAX PERALTA JIMENEZ | Inciensa, Instituto Costarricense de Investigación y Enseñanza en Nutrición y Salud | Adriana Godínez; Claudio Soto-Garita; Estela Cordero; Francisco Duarte; Hebleen Porras; Joselyn Prado & Mariana Piedra Calderón; José Luis Vargas; Mariela Gutiérrez; Melany Calderón |
| EPI_ISL_3102479 | HOSPITAL E | Analytical Competence | Cleber Furtado Aksenen; Fabio Miyajima; Fernando Braga Stehling; Francisco Eder de Moura Lopes; Jamille Maria Mendes Bezerra; Joaquim César do Nascimento Sousa Junior; Pedro Miguel Carneiro Jeronimo; Suzana Porto Almeida e Lucas Delerino; Thais Ferreira de Oliveira; Thais de Oliveira Costa; Ticiane Cavalcante de Souza; |

| | | | |
|---|---|---|---|
| | MATERNIDADE ADOLFO BEZERRA DE MENEZES | Molecular Epidemiology Lab/ACME, Oswaldo Cruz Foundation, Ceara (FIOCRUZ CE) | Veridiana Pessoa Miyajima |
| EPI_ISL_2801310, EPI_ISL_3102221, EPI_ISL_3102222, EPI_ISL_3102223, EPI_ISL_3102224, EPI_ISL_3102333, EPI_ISL_3102343, EPI_ISL_3102373, EPI_ISL_3102377, EPI_ISL_3102379, EPI_ISL_3102390, EPI_ISL_3912196, EPI_ISL_3912197 | | | |
| see above | HOSPITAL E MATERNIDADE DRA ZILDA ARNS NEUMANN | Analytical Competence Molecular Epidemiology Lab/ACME, Oswaldo Cruz Foundation, Ceara (FIOCRUZ CE) | Cleber Furtado Aksenen; Cleber Furtado Aksenen e Suzana Porto Almeida; Fabio Miyajima; Fernando Braga Stehling; Francisco Eder de Moura Lopes; Jamille Maria Mendes Bezerra; Joaquim Cesar do Nascimento Sousa Junior; Joaquim César do Nascimento Sousa Junior; Pedro Miguel Carneiro Jeronimo; Suzana Porto Almeida & Lucas Delerino on behalf of COVID-19 FIOCRUZ Genomic Network; Suzana Porto Almeida e Lucas Delerino; Thais Ferreira de Oliveira; Thais de Oliveira Costa; Ticiane Cavalcante de Souza; Veridiana Pessoa Miyajima |
| EPI_ISL_3102478 | HOSPITAL E MATERNIDADE GASTROCLINICA | Analytical Competence Molecular Epidemiology Lab/ACME, Oswaldo Cruz Foundation, Ceara (FIOCRUZ CE) | Cleber Furtado Aksenen; Fabio Miyajima; Fernando Braga Stehling; Francisco Eder de Moura Lopes; Jamille Maria Mendes Bezerra; Joaquim César do Nascimento Sousa Junior; Pedro Miguel Carneiro Jeronimo; Suzana Porto Almeida e Lucas Delerino; Thais Ferreira de Oliveira; Thais de Oliveira Costa; Ticiane Cavalcante de Souza; Veridiana Pessoa Miyajima |
| EPI_ISL_3912198 | HOSPITAL E MATERNIDADE JOSE GRANJA RIBEIRO | Analytical Competence Molecular Epidemiology Lab/ACME, Oswaldo Cruz Foundation, Ceara (FIOCRUZ CE) | Cleber Furtado Aksenen; Fabio Miyajima; Fernando Braga Stehling; Francisco Eder de Moura Lopes; Jamille Maria Mendes Bezerra; Joaquim Cesar do Nascimento Sousa Junior; Pedro Miguel Carneiro Jeronimo; Suzana Porto Almeida & Lucas Delerino on behalf of COVID-19 FIOCRUZ Genomic Network; Thais Ferreira de Oliveira; Thais de Oliveira Costa; Ticiane Cavalcante de Souza; Veridiana Pessoa Miyajima |
| EPI_ISL_1795344, EPI_ISL_2345610 | HOSPITAL E MATERNIDADE NOSSA SENHORA DA AJUDA | Instituto Butantan / ESALQ-Piracicaba | Antonio Jorge Martins; Bianca Cechetto Carlos. Mendelics: Bibiana Santos; Claudia Renata dos Santos Barros; David Schlesinger; David Schlesinger. Hemocentro Ribeirão Preto: Simone Kashima; Debora Botequio Moretti; Debora Botequio Moretti. Centro de Genômica Funcional da ESALQ: Luiz Lehmann Coutinho; Dimas Tadeu Covas; Elaine Cristina Marqueze; Elaine Vieira Santos; Elaine Vieira dos Santos; Elisangela Chicaroni Mattos; Erika Freitas; Evandra Strazza Rodrigues; Felipe Allan da Silva da Costa; Flavia Aburjaile; Guilherme Targino Valente; Heidge Fukumasu; Heidge Fukumasu. USP-Botucatu: Rejane Maria Tommasini Grotto; Instituto Butantan: Alexander Roberto Precioso; Jayme A. Souza-Neto; Jayme Augusto de Souza-Neto; Jessika Cristina Chagas Lesbon; José Salvatore Leister Patané; João Paulo Kitajima; Luiz Alcantara; Luiz Carlos Junior de Alcantara; Luiz Lehmann Coutinho; Maria Carolina Elias; Marta Giovanetti; Maurício Lacerda Nogueira; Patricia Akemi Assato; Rafael dos Santos Bezerra; Raquel de Lello Rocha Campos Cassano. NGS Soluções Genômicas: Pilar Drummond Sampaio Corrêa Mariani. FZEA-USP Pirassununga: Mirele Daiana Poleti; Raul Machado Neto; Rejane Maria Tommasini Grotto; Ricardo Augusto Brassaloti; Ricardo Haddad; Rodrigo Tocantins Calado.; Sandra Coccuzzo Sampaio; Sandra Coccuzzo Sampaio Vessoni; Simone Kashima; Svetoslav Nanev Slavov; Vagner Fonseca; Vincent Louis Viala |
| EPI_ISL_3102214, EPI_ISL_3102355, EPI_ISL_3102367, EPI_ISL_3102385, EPI_ISL_3102389, EPI_ISL_3102391, EPI_ISL_3102407, EPI_ISL_3102493, EPI_ISL_3102529, EPI_ISL_3102531, EPI_ISL_3102532, EPI_ISL_3102535, EPI_ISL_3912208, EPI_ISL_3912210 | | | |
| see above | HOSPITAL ESTADUAL LEONARDO DA VINCI | Analytical Competence Molecular Epidemiology Lab/ACME, Oswaldo Cruz Foundation, Ceara (FIOCRUZ CE) | Cleber Furtado Aksenen; Fabio Miyajima; Fernando Braga Stehling; Francisco Eder de Moura Lopes; Jamille Maria Mendes Bezerra; Joaquim Cesar do Nascimento Sousa Junior; Joaquim César do Nascimento Sousa Junior; Pedro Miguel Carneiro Jeronimo; Suzana Porto Almeida & Lucas Delerino on behalf of COVID-19 FIOCRUZ Genomic Network; Suzana Porto Almeida e Lucas Delerino; Thais Ferreira de Oliveira; Thais de Oliveira Costa; Ticiane Cavalcante de Souza; Veridiana Pessoa Miyajima |
| EPI_ISL_2150636 | HOSPITAL GENERAL UNIVERSITARIO DE GUADALAJARA | Instituto de Salud Carlos III | A. Monzón; ALEJANDRO; F. Casas; I. Jiménez; I.GONZALEZ PRAETORIUS; M. Sandonís; P. Zaballos; S. Cuesta; S. Iglesias-Caballero; S. Pozo; S. Varona; V. Camarero; Vázquez-Morón |
| EPI_ISL_2150639 | HOSPITAL GENERAL VIRGEN DE LA LUZ | Instituto de Salud Carlos III | A. Monzón; F. Casas; I. Jiménez; I.RODRIGUEZ ESCUDERO; Iglesias-Caballero; M. Sandonís; MARIA JOSE; P. Zaballos; S. Camarero; S. Cuesta; S. Pozo; S. Varona; V. Vázquez-Morón |
| EPI_ISL_1966738 | HOSPITAL GERAL DE VILA NOVA CACHOEIRINHA SAO PAULO | Instituto Butantan | Antonio Jorge Martins; Bianca Cechetto Carlos. Mendelics: Bibiana Santos; Claudia Renata dos Santos Barros; Cíntia Bittar; David Schlesinger. Hemocentro Ribeirão Preto: Simone Kashima; Debora Botequio Moretti; Elaine Cristina Marqueze; Elaine Vieira dos Santos; Elisangela Chicaroni Mattos; Erika Freitas; Evandra Strazza Rodrigues; Felipe Allan da Silva da Costa; Flavia Aburjaile; Fábio Sossai Possebon; Guilherme Campos; Guilherme Targino Valente; Heidge Fukumasu. USP-Botucatu: Rejane Maria Tommasini Grotto; Helena Lage Ferreira; Instituto Butantan: Dimas Tadeu Covas; Jardelina de Souza Todao Bernardino; Jayme A. Souza-Neto; Jessika Cristina Chagas Lesbon; Jorge A. Petroli Marchesi; José Salvatore Leister Patané; João Paulo Kitajima; João Pessoa Araújo Jr.; Leila Sabrina Ullmann; Loyze Paola Oliveira de Lima; Luiz Aurelio de Campos Crispin. Centro de Genômica Funcional da ESALQ: Luiz Lehmann Coutinho; Luiz Carlos Junior de Alcantara; Lívia Sacchetto; Maisa C. Pereira Parra; Maria Carolina Elias; Marta Giovanetti; Marília Moraes; Maurício Lacerda Nogueira. Prefeitura de Sao Paulo: Melissa Palmieri.; Patricia Akemi Assato; Paula Rahal; Paulo Inacio da Costa; Rafael dos Santos Bezerra; Raquel de Lello Rocha Campos Cassano. NGS Soluções Genômicas: Pilar Drummond Sampaio Corrêa Mariani. FZEA-USP Pirassununga: Mirele Daiana Poleti; Raul Machado Neto; Ricardo Augusto Brassaloti; Ricardo Haddad; Rodrigo Tocantins Calado. FAMERP-SJRP: Cecília Artico Banho; Sandra Coccuzzo Sampaio; Svetoslav Nanev Slavov; Vagner Fonseca; Vincent Louis Viala |
| EPI_ISL_1967078, EPI_ISL_1967080, EPI_ISL_1967081, EPI_ISL_1967084, EPI_ISL_1967086, EPI_ISL_1967088 | HOSPITAL GERAL JESUS TEIXEIRA DA COSTA GUAIANASES SAO PAULO | Instituto Butantan / ESALQ-USP (Piracicaba) | Antonio Jorge Martins; Bianca Cechetto Carlos. Mendelics: Bibiana Santos; Claudia Renata dos Santos Barros; Cíntia Bittar; David Schlesinger. Hemocentro Ribeirão Preto: Simone Kashima; Debora Botequio Moretti; Elaine Cristina Marqueze; Elaine Vieira dos Santos; Elisangela Chicaroni Mattos; Erika Freitas; Evandra Strazza Rodrigues; Felipe Allan da Silva da Costa; Flavia Aburjaile; Fábio Sossai Possebon; Guilherme Campos; Guilherme Targino Valente; Heidge Fukumasu. USP-Botucatu: Rejane Maria Tommasini Grotto; Helena Lage Ferreira; Instituto Butantan: Dimas Tadeu Covas; Jardelina de Souza Todao Bernardino; Jayme A. Souza-Neto; Jessika Cristina Chagas Lesbon; Jorge A. Petroli Marchesi; José Salvatore Leister Patané; João Paulo Kitajima; João Pessoa Araújo Jr.; Leila Sabrina Ullmann; Loyze Paola Oliveira de Lima; Luiz Aurelio de Campos Crispin. Centro de Genômica Funcional da ESALQ: Luiz Lehmann Coutinho; Luiz Carlos Junior de Alcantara; Lívia Sacchetto; Maisa C. Pereira Parra; Maria Carolina Elias; Marta Giovanetti; Marília Moraes; Maurício Lacerda Nogueira. Prefeitura de Sao Paulo: Melissa Palmieri.; Patricia Akemi Assato; Paula Rahal; Paulo Inacio da Costa; Rafael dos Santos Bezerra; Raquel de Lello Rocha Campos Cassano. NGS Soluções Genômicas: Pilar Drummond Sampaio Corrêa Mariani. FZEA-USP Pirassununga: Mirele Daiana Poleti; Raul Machado Neto; Ricardo Augusto Brassaloti; Ricardo Haddad; Rodrigo Tocantins Calado. FAMERP-SJRP: Cecília Artico Banho; Sandra Coccuzzo Sampaio; Svetoslav Nanev Slavov; Vagner Fonseca; Vincent Louis Viala |
| EPI_ISL_2801306 | HOSPITAL INFANTIL ALBERT SABIN | Analytical Competence Molecular Epidemiology Lab/ACME, Oswaldo Cruz Foundation, Ceara (FIOCRUZ CE) | Cleber Furtado Aksenen e Suzana Porto Almeida; Fabio Miyajima; Fernando Braga Stehling; Francisco Eder de Moura Lopes; Jamille Maria Mendes Bezerra; Joaquim César do Nascimento Sousa Junior; Pedro Miguel Carneiro Jeronimo; Thais Ferreira de Oliveira; Thais de Oliveira Costa; Ticiane Cavalcante de Souza; Veridiana Pessoa Miyajima |
| EPI_ISL_2020122 | HOSPITAL MANACOR | HOSPITAL UNIVERSITARIO SON ESPASES | Antonio Oliver; Carla López-Causapé; Pablo Fraile-Ribot; SeqCovid |
| EPI_ISL_2346037 | HOSPITAL MUN DR GUIDO GUIDA | Instituto Butantan | Antonio Jorge Martins; Claudia Renata dos Santos Barros; David Schlesinger; Debora Botequio Moretti; Dimas Tadeu Covas; Elaine Cristina Marqueze; Elaine Vieira Santos; Evandra Strazza Rodrigues; Heidge Fukumasu; Jayme Augusto de Souza-Neto; José Salvatore Leister Patané; Luiz Alcantara; Luiz Lehmann Coutinho; Maria Carolina Elias; Maurício Lacerda Nogueira; Rafael dos Santos Bezerra; Raul Machado Neto; Rejane Maria Tommasini Grotto; Ricardo Haddad; Sandra Coccuzzo Sampaio Vessoni; Simone Kashima; Svetoslav Nanev Slavov; Vincent Louis Viala |
| EPI_ISL_1966913, EPI_ISL_1966914 | HOSPITAL MUNICIPAL DE IBIUNA IBIUNA SP | Instituto Butantan / ESALQ-USP (Piracicaba) | Antonio Jorge Martins; Bianca Cechetto Carlos. Mendelics: Bibiana Santos; Claudia Renata dos Santos Barros; Cíntia Bittar; David Schlesinger. Hemocentro Ribeirão Preto: Simone Kashima; Debora Botequio Moretti; Elaine Cristina Marqueze; Elaine Vieira dos Santos; Elisangela Chicaroni Mattos; Erika Freitas; Evandra Strazza Rodrigues; Felipe Allan da Silva da Costa; Flavia Aburjaile; Fábio Sossai Possebon; Guilherme Campos; Guilherme Targino Valente; Heidge Fukumasu. USP-Botucatu: Rejane Maria Tommasini Grotto; Helena Lage Ferreira; Instituto Butantan: Dimas Tadeu Covas; Jardelina de Souza Todao Bernardino; Jayme A. Souza-Neto; Jessika Cristina Chagas Lesbon; Jorge A. Petroli Marchesi; José Salvatore Leister Patané; João Paulo Kitajima; João Pessoa Araújo Jr.; Leila Sabrina Ullmann; Loyze Paola Oliveira de Lima; Luiz Aurelio de Campos Crispin. Centro de Genômica Funcional da ESALQ: Luiz Lehmann Coutinho; Luiz Carlos Junior de Alcantara; Lívia Sacchetto; Maisa C. Pereira Parra; Maria Carolina Elias; Marta Giovanetti; Marília Moraes; Maurício Lacerda Nogueira. Prefeitura de Sao Paulo: Melissa Palmieri.; Patricia Akemi Assato; Paula Rahal; Paulo Inacio da Costa; Rafael dos Santos Bezerra; Raquel de Lello Rocha Campos Cassano. NGS Soluções Genômicas: Pilar Drummond Sampaio Corrêa Mariani. FZEA-USP Pirassununga: Mirele Daiana Poleti; Raul Machado Neto; Ricardo Augusto Brassaloti; Ricardo Haddad; Rodrigo Tocantins Calado. FAMERP-SJRP: Cecília Artico Banho; Sandra Coccuzzo Sampaio; Svetoslav Nanev Slavov; Vagner Fonseca; Vincent Louis Viala |
| EPI_ISL_1966919, EPI_ISL_1966920, EPI_ISL_1966921 | HOSPITAL MUNICIPAL DE ITABERA | Instituto Butantan / ESALQ-USP (Piracicaba) | Antonio Jorge Martins; Bianca Cechetto Carlos. Mendelics: Bibiana Santos; Claudia Renata dos Santos Barros; Cíntia Bittar; David Schlesinger. Hemocentro Ribeirão Preto: Simone Kashima; Debora Botequio Moretti; Elaine Cristina Marqueze; Elaine Vieira dos Santos; Elisangela Chicaroni Mattos; Erika Freitas; Evandra Strazza Rodrigues; Felipe Allan da Silva da Costa; Flavia Aburjaile; Fábio Sossai Possebon; Guilherme Campos; Guilherme Targino Valente; Heidge Fukumasu. USP-Botucatu: Rejane Maria Tommasini Grotto; Helena Lage Ferreira; Instituto Butantan: Dimas Tadeu Covas; Jardelina de Souza Todao Bernardino; Jayme A. Souza-Neto; Jessika Cristina Chagas Lesbon; Jorge A. Petroli Marchesi; José Salvatore Leister Patané; João Paulo Kitajima; João Pessoa Araújo Jr.; Leila Sabrina Ullmann; Loyze Paola Oliveira de Lima; Luiz Aurelio de Campos Crispin. Centro de Genômica Funcional da ESALQ: Luiz Lehmann Coutinho; Luiz Carlos Junior de Alcantara; Lívia Sacchetto; Maisa C. Pereira Parra; Maria Carolina Elias; Marta Giovanetti; Marília Moraes; Maurício Lacerda Nogueira. Prefeitura de Sao Paulo: Melissa Palmieri.; Patricia Akemi Assato; Paula Rahal; Paulo Inacio da Costa; Rafael dos Santos Bezerra; Raquel de Lello Rocha Campos Cassano. NGS Soluções Genômicas: Pilar Drummond Sampaio Corrêa Mariani. FZEA-USP Pirassununga: Mirele Daiana Poleti; Raul Machado Neto; Ricardo Augusto Brassaloti; Ricardo Haddad; Rodrigo Tocantins Calado. FAMERP-SJRP: Cecília Artico Banho; Sandra Coccuzzo Sampaio; Svetoslav Nanev Slavov; Vagner Fonseca; Vincent Louis Viala |
| EPI_ISL_2344767, EPI_ISL_2344788 | HOSPITAL MUNICIPAL DE ITAPIRA | Instituto Butantan / ESALQ-Piracicaba | Antonio Jorge Martins; Claudia Renata dos Santos Barros; David Schlesinger; Debora Botequio Moretti; Dimas Tadeu Covas; Elaine Cristina Marqueze; Elaine Vieira Santos; Evandra Strazza Rodrigues; Heidge Fukumasu; Jayme Augusto de Souza-Neto; José Salvatore Leister Patané; Luiz Alcantara; Luiz Lehmann Coutinho; Maria Carolina Elias; Maurício Lacerda Nogueira; Rafael dos Santos Bezerra; Raul Machado Neto; Rejane Maria Tommasini Grotto; Ricardo Haddad; Sandra Coccuzzo Sampaio Vessoni; Simone Kashima; Svetoslav Nanev Slavov; Vincent Louis Viala |
| EPI_ISL_1966942, EPI_ISL_1966945, EPI_ISL_1966946, EPI_ISL_1966947, EPI_ISL_1966949 | HOSPITAL MUNICIPAL DE ITAPIRA | Instituto Butantan / ESALQ-USP (Piracicaba) | Antonio Jorge Martins; Bianca Cechetto Carlos. Mendelics: Bibiana Santos; Claudia Renata dos Santos Barros; Cíntia Bittar; David Schlesinger. Hemocentro Ribeirão Preto: Simone Kashima; Debora Botequio Moretti; Elaine Cristina Marqueze; Elaine Vieira dos Santos; Elisangela Chicaroni Mattos; Erika Freitas; Evandra Strazza Rodrigues; Felipe Allan da Silva da Costa; Flavia Aburjaile; Fábio Sossai Possebon; Guilherme Campos; Guilherme Targino Valente; Heidge Fukumasu. USP-Botucatu: Rejane Maria Tommasini Grotto; Helena Lage Ferreira; Instituto Butantan: Dimas Tadeu Covas; Jardelina de Souza Todao Bernardino; Jayme A. Souza-Neto; Jessika Cristina Chagas Lesbon; Jorge A. Petroli Marchesi; José Salvatore Leister Patané; João Paulo Kitajima; João Pessoa Araújo Jr.; Leila Sabrina Ullmann; Loyze Paola Oliveira de Lima; Luiz Aurelio de Campos Crispin. Centro de Genômica Funcional da ESALQ: Luiz Lehmann Coutinho; Luiz Carlos Junior de Alcantara; Lívia Sacchetto; Maisa C. Pereira Parra; Maria Carolina Elias; Marta Giovanetti; Marília Moraes; Maurício Lacerda Nogueira. Prefeitura de Sao Paulo: Melissa Palmieri.; Patricia Akemi Assato; Paula Rahal; Paulo Inacio da Costa; Rafael dos Santos Bezerra; Raquel de Lello Rocha Campos Cassano. NGS Soluções Genômicas: Pilar Drummond Sampaio Corrêa Mariani. FZEA-USP Pirassununga: Mirele Daiana Poleti; Raul Machado Neto; Ricardo Augusto Brassaloti; Ricardo Haddad; Rodrigo Tocantins Calado. FAMERP-SJRP: Cecília Artico Banho; Sandra Coccuzzo Sampaio; Svetoslav Nanev Slavov; Vagner Fonseca; Vincent Louis Viala |
| EPI_ISL_2344571, EPI_ISL_2344573, EPI_ISL_2344574, EPI_ISL_2344578, EPI_ISL_2344579 | HOSPITAL MUNICIPAL DE ITAPIRA | Instituto Butantan / FZEA-USP-Pirassununga | Antonio Jorge Martins; Claudia Renata dos Santos Barros; David Schlesinger; Debora Botequio Moretti; Dimas Tadeu Covas; Elaine Cristina Marqueze; Elaine Vieira Santos; Evandra Strazza Rodrigues; Heidge Fukumasu; Jayme Augusto de Souza-Neto; José Salvatore Leister Patané; Luiz Alcantara; Luiz Lehmann Coutinho; Maria Carolina Elias; Maurício Lacerda Nogueira; Rafael dos Santos Bezerra; Raul Machado Neto; Rejane Maria Tommasini Grotto; Ricardo Haddad; Sandra Coccuzzo Sampaio Vessoni; Simone Kashima; Svetoslav Nanev Slavov; Vincent Louis Viala |
| EPI_ISL_1795211, EPI_ISL_2345439 | HOSPITAL MUNICIPAL DR MARIO GATTI CAMPINAS | Instituto Butantan / ESALQ-Piracicaba | Antonio Jorge Martins; Bianca Cechetto Carlos. Mendelics: Bibiana Santos; Claudia Renata dos Santos Barros; David Schlesinger; David Schlesinger. Hemocentro Ribeirão Preto: Simone Kashima; Debora Botequio Moretti; Debora Botequio Moretti. Centro de Genômica Funcional da ESALQ: Luiz Lehmann Coutinho; Dimas Tadeu Covas; Elaine Cristina Marqueze; Elaine Vieira Santos; Elaine Vieira dos Santos; Elisangela Chicaroni Mattos; Erika Freitas; Evandra Strazza Rodrigues; Felipe Allan da Silva da Costa; Flavia Aburjaile; Guilherme Targino Valente; Heidge Fukumasu; Heidge Fukumasu. USP-Botucatu: Rejane Maria Tommasini Grotto; Instituto Butantan: Alexander Roberto Precioso; Jayme A. Souza-Neto; Jayme Augusto de Souza-Neto; Jessika Cristina Chagas Lesbon; José Salvatore Leister Patané; João Paulo Kitajima; Luiz Alcantara; Luiz Carlos Junior de Alcantara; Luiz Lehmann Coutinho; Maria Carolina Elias; Marta Giovanetti; Maurício Lacerda Nogueira; Patricia Akemi Assato; Rafael dos Santos Bezerra; Raquel de Lello Rocha Campos Cassano. NGS Soluções Genômicas: Pilar Drummond Sampaio Corrêa Mariani. FZEA-USP Pirassununga: Mirele Daiana Poleti; Raul Machado Neto; Rejane Maria Tommasini Grotto; Ricardo Augusto Brassaloti; Ricardo Haddad; Rodrigo Tocantins Calado.; Sandra Coccuzzo Sampaio; Sandra Coccuzzo Sampaio Vessoni; Simone Kashima; Svetoslav Nanev Slavov; Vagner Fonseca; Vincent Louis Viala |
| EPI_ISL_2344800 | HOSPITAL MUNICIPAL DR TABAJARA RAMOS | Instituto Butantan / ESALQ-Piracicaba | Antonio Jorge Martins; Claudia Renata dos Santos Barros; David Schlesinger; Debora Botequio Moretti; Dimas Tadeu Covas; Elaine Cristina Marqueze; Elaine Vieira Santos; Evandra Strazza Rodrigues; Heidge Fukumasu; Jayme Augusto de Souza-Neto; José Salvatore Leister Patané; Luiz Alcantara; Luiz Lehmann Coutinho; Maria Carolina Elias; Maurício Lacerda Nogueira; Rafael dos Santos Bezerra; Raul Machado Neto; Rejane Maria Tommasini Grotto; Ricardo Haddad; Sandra Coccuzzo Sampaio Vessoni; Simone Kashima; Svetoslav Nanev Slavov; Vincent Louis Viala |
| EPI_ISL_1966645 | HOSPITAL MUNICIPAL DR WALDEMAR TEBALDI | Instituto Butantan / Mendelics | Antonio Jorge Martins; Bianca Cechetto Carlos. Mendelics: Bibiana Santos; Claudia Renata dos Santos Barros; Cíntia Bittar; David Schlesinger. Hemocentro Ribeirão Preto: Simone Kashima; Debora Botequio Moretti; Elaine Cristina Marqueze; Elaine Vieira dos Santos; Elisangela Chicaroni Mattos; Erika Freitas; Evandra Strazza Rodrigues; Felipe Allan da Silva da Costa; Flavia Aburjaile; Fábio Sossai Possebon; Guilherme Campos; Guilherme Targino Valente; Heidge Fukumasu. USP-Botucatu: Rejane Maria Tommasini Grotto; Helena Lage Ferreira; Instituto Butantan: Dimas Tadeu Covas; Jardelina de Souza Todao Bernardino; Jayme A. Souza-Neto; Jessika Cristina Chagas Lesbon; Jorge A. Petroli Marchesi; José Salvatore Leister Patané; João Paulo Kitajima; João Pessoa Araújo Jr.; Leila Sabrina Ullmann; Loyze Paola Oliveira de Lima; Luiz Aurelio de Campos Crispin. Centro de Genômica Funcional da ESALQ: Luiz Lehmann Coutinho; Luiz Carlos Junior de Alcantara; Lívia Sacchetto; Maisa C. Pereira Parra; Maria Carolina Elias; Marta Giovanetti; Marília Moraes; Maurício Lacerda Nogueira. Prefeitura de Sao Paulo: Melissa Palmieri.; Patricia Akemi Assato; Paula Rahal; Paulo Inacio da Costa; Rafael dos Santos Bezerra; Raquel de Lello Rocha Campos Cassano. NGS Soluções Genômicas: Pilar Drummond Sampaio Corrêa Mariani. FZEA-USP Pirassununga: Mirele Daiana Poleti; Raul Machado Neto; Ricardo Augusto Brassaloti; Ricardo Haddad; Rodrigo Tocantins Calado. FAMERP-SJRP: Cecília Artico Banho; Sandra Coccuzzo Sampaio; Svetoslav Nanev Slavov; Vagner Fonseca; Vincent Louis Viala |
| EPI_ISL_3102490 | HOSPITAL MUNICIPAL GOVERNADOR ADAUTO BEZERRA | Analytical Competence Molecular Epidemiology Lab/ACME, Oswaldo Cruz Foundation, Ceara (FIOCRUZ CE) | Cleber Furtado Aksenen; Fabio Miyajima; Fernando Braga Stehling; Francisco Eder de Moura Lopes; Jamille Maria Mendes Bezerra; Joaquim César do Nascimento Sousa Junior; Pedro Miguel Carneiro Jeronimo; Suzana Porto Almeida e Lucas Delerino; Thais Ferreira de Oliveira; Thais de Oliveira Costa; Ticiane Cavalcante de Souza; Veridiana Pessoa Miyajima |
| EPI_ISL_3912212, EPI_ISL_3912213 | HOSPITAL MUNICIPAL JOAO LEOPOLDO | Analytical Competence Molecular Epidemiology | Cleber Furtado Aksenen; Fabio Miyajima; Fernando Braga Stehling; Francisco Eder de Moura Lopes; Jamille Maria Mendes Bezerra; Joaquim Cesar do Nascimento Sousa Junior; Pedro Miguel Carneiro Jeronimo; Suzana Porto Almeida & Lucas Delerino on behalf of COVID-19 FIOCRUZ Genomic Network; Thais Ferreira de Oliveira; Thais de Oliveira Costa; Ticiane Cavalcante de Souza; Veridiana Pessoa Miyajima |

| | | | |
|---|---|---|---|
| | PINHEIRO LANDIM | Lab/ACME, Oswaldo Cruz Foundation, Ceara (FIOCRUZ CE) | |
| EPI_ISL_1967025, EPI_ISL_1967026 | HOSPITAL MUNICIPAL MARIA ROSA CARDOSO | Instituto Butantan / ESALQ-USP (Piracicaba) | Antonio Jorge Martins; Bianca Cechetto Carlos. Mendelics: Bibiana Santos; Claudia Renata dos Santos Barros; Cíntia Bittar; David Schlesinger. Hemocentro Ribeirão Preto: Simone Kashima; Debora Botequio Moretti; Elaine Cristina Marqueze; Elaine Vieira dos Santos; Elisangela Chicaroni Mattos; Erika Freitas; Evandra Strazza Rodrigues; Felipe Allan da Silva da Costa; Flavia Aburjaile; Fábio Sossai Possebon; Guilherme Campos; Guilherme Targino Valente; Heidge Fukumasu. USP-Botucatu: Rejane Maria Tommasini Grotto; Helena Lage Ferreira; Instituto Butantan: Dimas Tadeu Covas; Jardelina de Souza Todao Bernardino; Jayme A. Souza-Neto; Jessika Cristina Chagas Lesbon; Jorge A. Petroli Marchesi; José Salvatore Leister Patané; João Paulo Kitajima; João Pessoa Araújo Jr.; Leila Sabrina Ullmann; Loyze Paola Oliveira de Lima; Luiz Aurelio de Campos Crispin. Centro de Genômica Funcional da ESALQ: Luiz Lehmann Coutinho; Luiz Carlos Junior de Alcantara; Lívia Sacchetto; Maisa C. Pereira Parra; Maria Carolina Elias; Marta Giovanetti; Marília Moraes; Maurício Lacerda Nogueira. Prefeitura de Sao Paulo: Melissa Palmieri.; Patricia Akemi Assato; Paula Rahal; Paulo Inacio da Costa; Rafael dos Santos Bezerra; Raquel de Lello Rocha Campos Cassano. NGS Soluções Genômicas: Pilar Drummond Sampaio Corrêa Mariani. FZEA-USP Pirassununga: Mirele Daiana Poleti; Raul Machado Neto; Ricardo Augusto Brassaloti; Ricardo Haddad; Rodrigo Tocantins Calado. FAMERP-SJRP: Cecília Artico Banho; Sandra Coccuzzo Sampaio; Svetoslav Nanev Slavov; Vagner Fonseca; Vincent Louis Viala |
| EPI_ISL_1795337, EPI_ISL_1795338, EPI_ISL_1795341, EPI_ISL_2345603, EPI_ISL_2345604, EPI_ISL_2345607 | HOSPITAL MUNICIPAL REYNALDO GUERRA CAJATI | Instituto Butantan / ESALQ-Piracicaba | Antonio Jorge Martins; Bianca Cechetto Carlos. Mendelics: Bibiana Santos; Claudia Renata dos Santos Barros; David Schlesinger; David Schlesinger. Hemocentro Ribeirão Preto: Simone Kashima; Debora Botequio Moretti; Debora Botequio Moretti. Centro de Genômica Funcional da ESALQ: Luiz Lehmann Coutinho; Dimas Tadeu Covas; Elaine Cristina Marqueze; Elaine Vieira Santos; Elaine Vieira dos Santos; Elisangela Chicaroni Mattos; Erika Freitas; Evandra Strazza Rodrigues; Felipe Allan da Silva da Costa; Flavia Aburjaile; Guilherme Targino Valente; Heidge Fukumasu; Heidge Fukumasu. USP-Botucatu: Rejane Maria Tommasini Grotto; Instituto Butantan: Alexander Roberto Precioso; Jayme A. Souza-Neto; Jayme Augusto de Souza-Neto; Jessika Cristina Chagas Lesbon; José Salvatore Leister Patané; João Paulo Kitajima; Luiz Alcantara; Luiz Carlos Junior de Alcantara; Luiz Lehmann Coutinho; Maria Carolina Elias; Marta Giovanetti; Maurício Lacerda Nogueira; Patricia Akemi Assato; Rafael dos Santos Bezerra; Raquel de Lello Rocha Campos Cassano. NGS Soluções Genômicas: Pilar Drummond Sampaio Corrêa Mariani. FZEA-USP Pirassununga: Mirele Daiana Poleti; Raul Machado Neto; Rejane Maria Tommasini Grotto; Ricardo Augusto Brassaloti; Ricardo Haddad; Rodrigo Tocantins Calado.; Sandra Coccuzzo Sampaio; Sandra Coccuzzo Sampaio Vessoni; Simone Kashima; Svetoslav Nanev Slavov; Vagner Fonseca; Vincent Louis Viala |
| EPI_ISL_1966846 | HOSPITAL MUNICIPAL REYNALDO GUERRA CAJATI | Instituto Butantan / ESALQ-USP (Piracicaba) | Antonio Jorge Martins; Bianca Cechetto Carlos. Mendelics: Bibiana Santos; Claudia Renata dos Santos Barros; Cíntia Bittar; David Schlesinger. Hemocentro Ribeirão Preto: Simone Kashima; Debora Botequio Moretti; Elaine Cristina Marqueze; Elaine Vieira dos Santos; Elisangela Chicaroni Mattos; Erika Freitas; Evandra Strazza Rodrigues; Felipe Allan da Silva da Costa; Flavia Aburjaile; Fábio Sossai Possebon; Guilherme Campos; Guilherme Targino Valente; Heidge Fukumasu. USP-Botucatu: Rejane Maria Tommasini Grotto; Helena Lage Ferreira; Instituto Butantan: Dimas Tadeu Covas; Jardelina de Souza Todao Bernardino; Jayme A. Souza-Neto; Jessika Cristina Chagas Lesbon; Jorge A. Petroli Marchesi; José Salvatore Leister Patané; João Paulo Kitajima; João Pessoa Araújo Jr.; Leila Sabrina Ullmann; Loyze Paola Oliveira de Lima; Luiz Aurelio de Campos Crispin. Centro de Genômica Funcional da ESALQ: Luiz Lehmann Coutinho; Luiz Carlos Junior de Alcantara; Lívia Sacchetto; Maisa C. Pereira Parra; Maria Carolina Elias; Marta Giovanetti; Marília Moraes; Maurício Lacerda Nogueira. Prefeitura de Sao Paulo: Melissa Palmieri.; Patricia Akemi Assato; Paula Rahal; Paulo Inacio da Costa; Rafael dos Santos Bezerra; Raquel de Lello Rocha Campos Cassano. NGS Soluções Genômicas: Pilar Drummond Sampaio Corrêa Mariani. FZEA-USP Pirassununga: Mirele Daiana Poleti; Raul Machado Neto; Ricardo Augusto Brassaloti; Ricardo Haddad; Rodrigo Tocantins Calado. FAMERP-SJRP: Cecília Artico Banho; Sandra Coccuzzo Sampaio; Svetoslav Nanev Slavov; Vagner Fonseca; Vincent Louis Viala |
| EPI_ISL_3102323, EPI_ISL_3912221 | HOSPITAL OTOCLINICA | Analytical Competence Molecular Epidemiology Lab/ACME, Oswaldo Cruz Foundation, Ceara (FIOCRUZ CE) | Cleber Furtado Aksenen; Fabio Miyajima; Fernando Braga Stehling; Francisco Eder de Moura Lopes; Jamille Maria Mendes Bezerra; Joaquim Cesar do Nascimento Sousa Junior; Joaquim César do Nascimento Sousa Junior; Pedro Miguel Carneiro Jeronimo; Suzana Porto Almeida & Lucas Delerino on behalf of COVID-19 FIOCRUZ Genomic Network; Suzana Porto Almeida e Lucas Delerino; Thais Ferreira de Oliveira; Thais de Oliveira Costa; Ticiane Cavalcante de Souza; Veridiana Pessoa Miyajima |
| EPI_ISL_3912222 | HOSPITAL REGIONAL DE IGUATU | Analytical Competence Molecular Epidemiology Lab/ACME, Oswaldo Cruz Foundation, Ceara (FIOCRUZ CE) | Cleber Furtado Aksenen; Fabio Miyajima; Fernando Braga Stehling; Francisco Eder de Moura Lopes; Jamille Maria Mendes Bezerra; Joaquim Cesar do Nascimento Sousa Junior; Pedro Miguel Carneiro Jeronimo; Suzana Porto Almeida & Lucas Delerino on behalf of COVID-19 FIOCRUZ Genomic Network; Thais Ferreira de Oliveira; Thais de Oliveira Costa; Ticiane Cavalcante de Souza; Veridiana Pessoa Miyajima |
| EPI_ISL_1795221, EPI_ISL_2345451 | HOSPITAL REGIONAL DE ITAPETININGA | Instituto Butantan / ESALQ-Piracicaba | Antonio Jorge Martins; Bianca Cechetto Carlos. Mendelics: Bibiana Santos; Claudia Renata dos Santos Barros; David Schlesinger; David Schlesinger. Hemocentro Ribeirão Preto: Simone Kashima; Debora Botequio Moretti; Debora Botequio Moretti. Centro de Genômica Funcional da ESALQ: Luiz Lehmann Coutinho; Dimas Tadeu Covas; Elaine Cristina Marqueze; Elaine Vieira Santos; Elaine Vieira dos Santos; Elisangela Chicaroni Mattos; Erika Freitas; Evandra Strazza Rodrigues; Felipe Allan da Silva da Costa; Flavia Aburjaile; Guilherme Targino Valente; Heidge Fukumasu; Heidge Fukumasu. USP-Botucatu: Rejane Maria Tommasini Grotto; Instituto Butantan: Alexander Roberto Precioso; Jayme A. Souza-Neto; Jayme Augusto de Souza-Neto; Jessika Cristina Chagas Lesbon; José Salvatore Leister Patané; João Paulo Kitajima; Luiz Alcantara; Luiz Carlos Junior de Alcantara; Luiz Lehmann Coutinho; Maria Carolina Elias; Marta Giovanetti; Maurício Lacerda Nogueira; Patricia Akemi Assato; Rafael dos Santos Bezerra; Raquel de Lello Rocha Campos Cassano. NGS Soluções Genômicas: Pilar Drummond Sampaio Corrêa Mariani. FZEA-USP Pirassununga: Mirele Daiana Poleti; Raul Machado Neto; Rejane Maria Tommasini Grotto; Ricardo Augusto Brassaloti; Ricardo Haddad; Rodrigo Tocantins Calado.; Sandra Coccuzzo Sampaio; Sandra Coccuzzo Sampaio Vessoni; Simone Kashima; Svetoslav Nanev Slavov; Vagner Fonseca; Vincent Louis Viala |
| EPI_ISL_1966923, EPI_ISL_1966924, EPI_ISL_1966925 | HOSPITAL REGIONAL DE ITAPETININGA | Instituto Butantan / ESALQ-USP (Piracicaba) | Antonio Jorge Martins; Bianca Cechetto Carlos. Mendelics: Bibiana Santos; Claudia Renata dos Santos Barros; Cíntia Bittar; David Schlesinger. Hemocentro Ribeirão Preto: Simone Kashima; Debora Botequio Moretti; Elaine Cristina Marqueze; Elaine Vieira dos Santos; Elisangela Chicaroni Mattos; Erika Freitas; Evandra Strazza Rodrigues; Felipe Allan da Silva da Costa; Flavia Aburjaile; Fábio Sossai Possebon; Guilherme Campos; Guilherme Targino Valente; Heidge Fukumasu. USP-Botucatu: Rejane Maria Tommasini Grotto; Helena Lage Ferreira; Instituto Butantan: Dimas Tadeu Covas; Jardelina de Souza Todao Bernardino; Jayme A. Souza-Neto; Jessika Cristina Chagas Lesbon; Jorge A. Petroli Marchesi; José Salvatore Leister Patané; João Paulo Kitajima; João Pessoa Araújo Jr.; Leila Sabrina Ullmann; Loyze Paola Oliveira de Lima; Luiz Aurelio de Campos Crispin. Centro de Genômica Funcional da ESALQ: Luiz Lehmann Coutinho; Luiz Carlos Junior de Alcantara; Lívia Sacchetto; Maisa C. Pereira Parra; Maria Carolina Elias; Marta Giovanetti; Marília Moraes; Maurício Lacerda Nogueira. Prefeitura de Sao Paulo: Melissa Palmieri.; Patricia Akemi Assato; Paula Rahal; Paulo Inacio da Costa; Rafael dos Santos Bezerra; Raquel de Lello Rocha Campos Cassano. NGS Soluções Genômicas: Pilar Drummond Sampaio Corrêa Mariani. FZEA-USP Pirassununga: Mirele Daiana Poleti; Raul Machado Neto; Ricardo Augusto Brassaloti; Ricardo Haddad; Rodrigo Tocantins Calado. FAMERP-SJRP: Cecília Artico Banho; Sandra Coccuzzo Sampaio; Svetoslav Nanev Slavov; Vagner Fonseca; Vincent Louis Viala |
| EPI_ISL_2801320 | HOSPITAL REGIONAL DO CARIRI | Analytical Competence Molecular Epidemiology Lab/ACME, Oswaldo Cruz Foundation, Ceara (FIOCRUZ CE) | Cleber Furtado Aksenen e Suzana Porto Almeida; Fabio Miyajima; Fernando Braga Stehling; Francisco Eder de Moura Lopes; Jamille Maria Mendes Bezerra; Joaquim César do Nascimento Sousa Junior; Pedro Miguel Carneiro Jeronimo; Thais Ferreira de Oliveira; Thais de Oliveira Costa; Ticiane Cavalcante de Souza; Veridiana Pessoa Miyajima |
| EPI_ISL_3102326, EPI_ISL_3912224 | HOSPITAL REGIONAL DO SERTAO CENTRAL | Analytical Competence Molecular Epidemiology Lab/ACME, Oswaldo Cruz Foundation, Ceara (FIOCRUZ CE) | Cleber Furtado Aksenen; Fabio Miyajima; Fernando Braga Stehling; Francisco Eder de Moura Lopes; Jamille Maria Mendes Bezerra; Joaquim Cesar do Nascimento Sousa Junior; Joaquim César do Nascimento Sousa Junior; Pedro Miguel Carneiro Jeronimo; Suzana Porto Almeida & Lucas Delerino on behalf of COVID-19 FIOCRUZ Genomic Network; Suzana Porto Almeida e Lucas Delerino; Thais Ferreira de Oliveira; Thais de Oliveira Costa; Ticiane Cavalcante de Souza; Veridiana Pessoa Miyajima |
| EPI_ISL_2272969 | HOSPITAL SAN VICENTE DE PAUL | Inciensa, Instituto Costarricense de Investigación y Enseñanza en Nutrición y Salud | Adriana Godínez; Claudio Soto-Garita; Estela Cordero; Francisco Duarte; Hebleen Porras; Joselyn Prado & Silvia Sáenz-García; José Luis Vargas; Mariela Gutiérrez; Melany Calderón |
| EPI_ISL_1795372, EPI_ISL_2345946 | HOSPITAL SANTA THEREZINHA BROTAS | Instituto Butantan / ESALQ-Piracicaba | Antonio Jorge Martins; Bianca Cechetto Carlos. Mendelics: Bibiana Santos; Claudia Renata dos Santos Barros; David Schlesinger; David Schlesinger. Hemocentro Ribeirão Preto: Simone Kashima; Debora Botequio Moretti; Debora Botequio Moretti. Centro de Genômica Funcional da ESALQ: Luiz Lehmann Coutinho; Dimas Tadeu Covas; Elaine Cristina Marqueze; Elaine Vieira Santos; Elaine Vieira dos Santos; Elisangela Chicaroni Mattos; Erika Freitas; Evandra Strazza Rodrigues; Felipe Allan da Silva da Costa; Flavia Aburjaile; Guilherme Targino Valente; Heidge Fukumasu; Heidge Fukumasu. USP-Botucatu: Rejane Maria Tommasini Grotto; Instituto Butantan: Alexander Roberto Precioso; Jayme A. Souza-Neto; Jayme Augusto de Souza-Neto; Jessika Cristina Chagas Lesbon; José Salvatore Leister Patané; João Paulo Kitajima; Luiz Alcantara; Luiz Carlos Junior de Alcantara; Luiz Lehmann Coutinho; Maria Carolina Elias; Marta Giovanetti; Maurício Lacerda Nogueira; Patricia Akemi Assato; Rafael dos Santos Bezerra; Raquel de Lello Rocha Campos Cassano. NGS Soluções Genômicas: Pilar Drummond Sampaio Corrêa Mariani. FZEA-USP Pirassununga: Mirele Daiana Poleti; Raul Machado Neto; Rejane Maria Tommasini Grotto; Ricardo Augusto Brassaloti; Ricardo Haddad; Rodrigo Tocantins Calado.; Sandra Coccuzzo Sampaio; Sandra Coccuzzo Sampaio Vessoni; Simone Kashima; Svetoslav Nanev Slavov; Vagner Fonseca; Vincent Louis Viala |
| EPI_ISL_1966954 | HOSPITAL SANTO ANTONIO DE JUQUIA JUQUIA | Instituto Butantan / ESALQ-USP (Piracicaba) | Antonio Jorge Martins; Bianca Cechetto Carlos. Mendelics: Bibiana Santos; Claudia Renata dos Santos Barros; Cíntia Bittar; David Schlesinger. Hemocentro Ribeirão Preto: Simone Kashima; Debora Botequio Moretti; Elaine Cristina Marqueze; Elaine Vieira dos Santos; Elisangela Chicaroni Mattos; Erika Freitas; Evandra Strazza Rodrigues; Felipe Allan da Silva da Costa; Flavia Aburjaile; Fábio Sossai Possebon; Guilherme Campos; Guilherme Targino Valente; Heidge Fukumasu. USP-Botucatu: Rejane Maria Tommasini Grotto; Helena Lage Ferreira; Instituto Butantan: Dimas Tadeu Covas; Jardelina de Souza Todao Bernardino; Jayme A. Souza-Neto; Jessika Cristina Chagas Lesbon; Jorge A. Petroli Marchesi; José Salvatore Leister Patané; João Paulo Kitajima; João Pessoa Araújo Jr.; Leila Sabrina Ullmann; Loyze Paola Oliveira de Lima; Luiz Aurelio de Campos Crispin. Centro de Genômica Funcional da ESALQ: Luiz Lehmann Coutinho; Luiz Carlos Junior de Alcantara; Lívia Sacchetto; Maisa C. Pereira Parra; Maria Carolina Elias; Marta Giovanetti; Marília Moraes; Maurício Lacerda Nogueira. Prefeitura de Sao Paulo: Melissa Palmieri.; Patricia Akemi Assato; Paula Rahal; Paulo Inacio da Costa; Rafael dos Santos Bezerra; Raquel de Lello Rocha Campos Cassano. NGS Soluções Genômicas: Pilar Drummond Sampaio Corrêa Mariani. FZEA-USP Pirassununga: Mirele Daiana Poleti; Raul Machado Neto; Ricardo Augusto Brassaloti; Ricardo Haddad; Rodrigo Tocantins Calado. FAMERP-SJRP: Cecília Artico Banho; Sandra Coccuzzo Sampaio; Svetoslav Nanev Slavov; Vagner Fonseca; Vincent Louis Viala |
| EPI_ISL_3102239, EPI_ISL_3102240, EPI_ISL_3102241, EPI_ISL_3102242, EPI_ISL_3102244, EPI_ISL_3102245, EPI_ISL_3102246, EPI_ISL_3102247, EPI_ISL_3102249, EPI_ISL_3102250, EPI_ISL_3102251, EPI_ISL_3102410, EPI_ISL_3102411, EPI_ISL_3102414, EPI_ISL_3102421, EPI_ISL_3102423, EPI_ISL_3102437, EPI_ISL_3102438, EPI_ISL_3102439, EPI_ISL_3102441, EPI_ISL_3102446, EPI_ISL_3102456, EPI_ISL_3102459 | | | |
| see above | HOSPITAL SAO JOSE DE DOENCAS INFECCIOSAS | Analytical Competence Molecular Epidemiology Lab/ACME, Oswaldo Cruz Foundation, Ceara (FIOCRUZ CE) | Cleber Furtado Aksenen; Fabio Miyajima; Fernando Braga Stehling; Francisco Eder de Moura Lopes; Jamille Maria Mendes Bezerra; Joaquim César do Nascimento Sousa Junior; Pedro Miguel Carneiro Jeronimo; Suzana Porto Almeida e Lucas Delerino; Thais Ferreira de Oliveira; Thais de Oliveira Costa; Ticiane Cavalcante de Souza; Veridiana Pessoa Miyajima |
| EPI_ISL_2209319 | HOSPITAL SAO JOSE ITIRAPINA | Instituto Butantan | Antonio Jorge Martins; Claudia Renata dos Santos Barros; David Schlesinger; Debora Botequio Moretti; Dimas Tadeu Covas; Elaine Cristina Marqueze; Elaine Vieira Santos; Evandra Strazza Rodrigues; Heidge Fukumasu; Jayme Augusto de Souza-Neto; José Salvatore Leister Patané; Luiz Alcantara; Luiz Lehmann Coutinho; Maria Carolina Elias; Maurício Lacerda Nogueira; Rafael dos Santos Bezerra; Raul Machado Neto; Rejane Maria Tommasini Grotto; Ricardo Haddad; Sandra Coccuzzo Sampaio Vessoni; Simone Kashima; Svetoslav Nanev Slavov; Vincent Louis Viala. |
| EPI_ISL_2209975, EPI_ISL_2210091 | HOSPITAL SAO JOSE ITIRAPINA | Instituto Butantan / FZEA-USP-Pirassununga | Antonio Jorge Martins; Claudia Renata dos Santos Barros; David Schlesinger; Debora Botequio Moretti; Dimas Tadeu Covas; Elaine Cristina Marqueze; Elaine Vieira Santos; Evandra Strazza Rodrigues; Heidge Fukumasu; Jayme Augusto de Souza-Neto; José Salvatore Leister Patané; Luiz Alcantara; Luiz Lehmann Coutinho; Maria Carolina Elias; Maurício Lacerda Nogueira; Rafael dos Santos Bezerra; Raul Machado Neto; Rejane Maria Tommasini Grotto; Ricardo Haddad; Sandra Coccuzzo Sampaio Vessoni; Simone Kashima; Svetoslav Nanev Slavov; Vincent Louis Viala |
| EPI_ISL_3102489 | HOSPITAL SAO LUCAS | Analytical Competence Molecular Epidemiology Lab/ACME, Oswaldo Cruz Foundation, Ceara (FIOCRUZ CE) | Cleber Furtado Aksenen; Fabio Miyajima; Fernando Braga Stehling; Francisco Eder de Moura Lopes; Jamille Maria Mendes Bezerra; Joaquim César do Nascimento Sousa Junior; Pedro Miguel Carneiro Jeronimo; Suzana Porto Almeida e Lucas Delerino; Thais Ferreira de Oliveira; Thais de Oliveira Costa; Ticiane Cavalcante de Souza; Veridiana Pessoa Miyajima |
| EPI_ISL_3102486, EPI_ISL_3102487 | HOSPITAL SAO MATEUS | Analytical Competence Molecular Epidemiology Lab/ACME, Oswaldo Cruz Foundation, Ceara (FIOCRUZ CE) | Cleber Furtado Aksenen; Fabio Miyajima; Fernando Braga Stehling; Francisco Eder de Moura Lopes; Jamille Maria Mendes Bezerra; Joaquim César do Nascimento Sousa Junior; Pedro Miguel Carneiro Jeronimo; Suzana Porto Almeida e Lucas Delerino; Thais Ferreira de Oliveira; Thais de Oliveira Costa; Ticiane Cavalcante de Souza; Veridiana Pessoa Miyajima |
| EPI_ISL_1967065, EPI_ISL_1967066 | HOSPITAL SAO MIGUEL ARCANJO | Instituto Butantan / ESALQ-USP (Piracicaba) | Antonio Jorge Martins; Bianca Cechetto Carlos. Mendelics: Bibiana Santos; Claudia Renata dos Santos Barros; Cíntia Bittar; David Schlesinger. Hemocentro Ribeirão Preto: Simone Kashima; Debora Botequio Moretti; Elaine Cristina Marqueze; Elaine Vieira dos Santos; Elisangela Chicaroni Mattos; Erika Freitas; Evandra Strazza Rodrigues; Felipe Allan da Silva da Costa; Flavia Aburjaile; Fábio Sossai Possebon; Guilherme Campos; Guilherme Targino Valente; Heidge Fukumasu. USP-Botucatu: Rejane Maria Tommasini Grotto; Helena Lage Ferreira; Instituto Butantan: Dimas Tadeu Covas; Jardelina de Souza Todao Bernardino; Jayme A. Souza-Neto; Jessika Cristina Chagas Lesbon; Jorge A. Petroli Marchesi; José Salvatore Leister Patané; João Paulo Kitajima; João Pessoa Araújo Jr.; Leila Sabrina Ullmann; Loyze Paola Oliveira de Lima; Luiz Aurelio de Campos Crispin. Centro de Genômica Funcional da ESALQ: Luiz Lehmann Coutinho; Luiz Carlos Junior de Alcantara; Lívia Sacchetto; Maisa C. Pereira Parra; Maria Carolina Elias; Marta Giovanetti; Marília Moraes; Maurício Lacerda Nogueira. Prefeitura de Sao Paulo: Melissa Palmieri.; Patricia Akemi Assato; Paula Rahal; Paulo Inacio da Costa; Rafael dos Santos Bezerra; Raquel de Lello Rocha Campos Cassano. NGS Soluções Genômicas: Pilar Drummond Sampaio Corrêa Mariani. FZEA-USP Pirassununga: Mirele Daiana Poleti; Raul Machado Neto; Ricardo Augusto Brassaloti; Ricardo Haddad; Rodrigo Tocantins Calado. FAMERP-SJRP: Cecília Artico Banho; Sandra Coccuzzo Sampaio; Svetoslav Nanev Slavov; Vagner Fonseca; Vincent Louis Viala |
| EPI_ISL_2801312, EPI_ISL_2801324 | HOSPITAL SÃO JOSE DE DOENÇAS INFECCIOSAS | Analytical Competence Molecular Epidemiology Lab/ACME, Oswaldo Cruz Foundation, Ceara (FIOCRUZ CE) | Cleber Furtado Aksenen e Suzana Porto Almeida; Fabio Miyajima; Fernando Braga Stehling; Francisco Eder de Moura Lopes; Jamille Maria Mendes Bezerra; Joaquim César do Nascimento Sousa Junior; Pedro Miguel Carneiro Jeronimo; Thais Ferreira de Oliveira; Thais de Oliveira Costa; Ticiane Cavalcante de Souza; Veridiana Pessoa Miyajima |
| EPI_ISL_2801317 | HOSPITAL SÃO MATEUS | Analytical Competence Molecular Epidemiology Lab/ACME, Oswaldo Cruz Foundation, Ceara (FIOCRUZ CE) | Cleber Furtado Aksenen e Suzana Porto Almeida; Fabio Miyajima; Fernando Braga Stehling; Francisco Eder de Moura Lopes; Jamille Maria Mendes Bezerra; Joaquim César do Nascimento Sousa Junior; Pedro Miguel Carneiro Jeronimo; Thais Ferreira de Oliveira; Thais de Oliveira Costa; Ticiane Cavalcante de Souza; Veridiana Pessoa Miyajima |
| EPI_ISL_2150638 | HOSPITAL UNIV. | Instituto de Salud | A. Monzón; ANA MARIA; F. Casas; I. Jiménez; I.MILAGRO BEAMONTE; M. Sandonís; P. Zaballos; S. Cuesta; S. Iglesias-Caballero; S. Pozo; S. Varona; V. Camarero; Vázquez-Morón |

| | | | |
|---|---|---|---|
| | MIGUEL SERVET | Carlos III | |
| EPI_ISL_1908182 | HOSPITAL UNIVERS MARQUES DE VALDECILLA | Instituto de Salud Carlos III | A. Monzón; CARLOS ANTONIO; F. Casas; I. Jiménez; I.SALAS VENERO; M. Sandonís; P. Zaballos; S. Cuesta; S. Iglesias-Caballero; S. Pozo; S. Varona; V. Camarero; Vázquez-Morón |
| EPI_ISL_3502764, EPI_ISL_3532482 | HOSPITAL UNIVERSITARIO 12 DE OCTUBRE | HOSPITAL UNIVERSITARIO 12 DE OCTUBRE | Carmen Martín-Higuera; Esther Viedma; Irene Múñoz-Gallego; M.ª Dolores Folgueira; Mar Aguilera; Noelia Moral; Rafael Delgado; Sagrario Zurita |
| EPI_ISL_2020104, EPI_ISL_2020154, EPI_ISL_2020179, EPI_ISL_2100319 | HOSPITAL UNIVERSITARIO SON ESPASES | HOSPITAL UNIVERSITARIO SON ESPASES | Antonio Oliver; Carla López-Causapé; Pablo Fraile-Ribot; SeqCovid |
| EPI_ISL_1510754, EPI_ISL_2150635, EPI_ISL_2150641 | HOSPITAL UNIVERSITARIO VIRGEN DE LA ARRIXACA | Instituto de Salud Carlos III | A. Monzón; F. Casas; I. Jiménez; I. MORENO PARRADO; I.MORENO PARRADO; Iglesias-Caballero; LAURA; M. Camarero; M. Sandonís; P. Zaballos; S. Camarero; S. Cuesta; S. Iglesias-Caballero; S. Pozo; S. Varona; Sandonís; V. Vázquez-Morón |
| EPI_ISL_2801325 | HOSPITAL UNIVERSITÁRIO WALTER CANTIDIO | Analytical Competence Molecular Epidemiology Lab/ACME, Oswaldo Cruz Foundation, Ceara (FIOCRUZ CE) | Cleber Furtado Aksenen e Suzana Porto Almeida; Fabio Miyajima; Fernando Braga Stehling; Francisco Eder de Moura Lopes; Jamille Maria Mendes Bezerra; Joaquim César do Nascimento Sousa Junior; Pedro Miguel Carneiro Jeronimo; Thais Ferreira de Oliveira; Thais de Oliveira Costa; Ticiane Cavalcante de Souza; Veridiana Pessoa Miyajima |
| EPI_ISL_3102262 | HPP LUIZ ROBERTO PESSOA AIRES | Analytical Competence Molecular Epidemiology Lab/ACME, Oswaldo Cruz Foundation, Ceara (FIOCRUZ CE) | Cleber Furtado Aksenen; Fabio Miyajima; Fernando Braga Stehling; Francisco Eder de Moura Lopes; Jamille Maria Mendes Bezerra; Joaquim César do Nascimento Sousa Junior; Pedro Miguel Carneiro Jeronimo; Suzana Porto Almeida e Lucas Delerino; Thais Ferreira de Oliveira; Thais de Oliveira Costa; Ticiane Cavalcante de Souza; Veridiana Pessoa Miyajima |
| EPI_ISL_1971337, EPI_ISL_2081100, EPI_ISL_2081106, EPI_ISL_2081109, EPI_ISL_2081119, EPI_ISL_2483366, EPI_ISL_2483379, EPI_ISL_2483424, EPI_ISL_2483426, EPI_ISL_2483427, EPI_ISL_2483428, EPI_ISL_2483430, EPI_ISL_2483432, EPI_ISL_2483438, EPI_ISL_2483443, EPI_ISL_2738475 | | | |
| see above | Hackensack Medical Center | New York Genome Center | Andre Corvelo; Barry Kreiswirth; David Perlin; Dayna M. Oschwald; Jose Mediavilla; Kaelea Composto; Kar Chow; Liang Chen; Marcus Cunningham; Michael Zody; Samantha Fennessey; Tom Maniatis |
| EPI_ISL_1194849, EPI_ISL_1194852, EPI_ISL_1337165, EPI_ISL_1337333, EPI_ISL_1337346, EPI_ISL_1337348, EPI_ISL_1480020, EPI_ISL_1480124, EPI_ISL_1480193, EPI_ISL_1480204, EPI_ISL_1480211, EPI_ISL_1480288, EPI_ISL_1480420, EPI_ISL_1480437, EPI_ISL_1553319, EPI_ISL_1553399, EPI_ISL_1553453, EPI_ISL_1553537, EPI_ISL_1553576, EPI_ISL_1553622, EPI_ISL_1553640, EPI_ISL_1553671, EPI_ISL_1553907, EPI_ISL_1553916, EPI_ISL_1553947, EPI_ISL_1554010, EPI_ISL_1554059, EPI_ISL_1554095, EPI_ISL_1554100, EPI_ISL_1554122, EPI_ISL_1554156, EPI_ISL_1554167, EPI_ISL_1554231, EPI_ISL_1554323, EPI_ISL_1554341, EPI_ISL_1554396, EPI_ISL_1554596, EPI_ISL_1554602, EPI_ISL_1554637, EPI_ISL_1554734, EPI_ISL_1554751, EPI_ISL_1554758, EPI_ISL_1554779, EPI_ISL_1554811, EPI_ISL_1554823, EPI_ISL_1575765, EPI_ISL_1575901, EPI_ISL_1575923, EPI_ISL_1575934, EPI_ISL_1576009, EPI_ISL_1576082, EPI_ISL_1576104, EPI_ISL_1576139, EPI_ISL_1576253, EPI_ISL_1576393, EPI_ISL_1576405, EPI_ISL_1576414, EPI_ISL_1576530, EPI_ISL_1576599, EPI_ISL_1576618, EPI_ISL_1576647, EPI_ISL_1576731, EPI_ISL_1580837, EPI_ISL_1580975, EPI_ISL_1580989, EPI_ISL_1580995, EPI_ISL_1581008, EPI_ISL_1581042, EPI_ISL_1581070, EPI_ISL_1581109, EPI_ISL_1581249, EPI_ISL_1581305, EPI_ISL_1581378, EPI_ISL_1581490, EPI_ISL_1581546, EPI_ISL_1581567, EPI_ISL_1581626, EPI_ISL_1581731, EPI_ISL_1581760, EPI_ISL_1581778, EPI_ISL_1581803, EPI_ISL_1592426, EPI_ISL_1592434, EPI_ISL_1592486, EPI_ISL_1592539, EPI_ISL_1592666, EPI_ISL_1592803, EPI_ISL_1592825, EPI_ISL_1592894, EPI_ISL_1593008, EPI_ISL_1593020, EPI_ISL_1593159, EPI_ISL_1593163, EPI_ISL_1593172, EPI_ISL_1615255, EPI_ISL_1615324, EPI_ISL_1615327, EPI_ISL_1615338, EPI_ISL_1615350, EPI_ISL_1615371, EPI_ISL_1678891, EPI_ISL_1678910, EPI_ISL_1678978, EPI_ISL_1679026, EPI_ISL_1679027, EPI_ISL_1679074, EPI_ISL_1679117, EPI_ISL_1679121, EPI_ISL_1679207, EPI_ISL_1679340, EPI_ISL_1679452, EPI_ISL_1679551, EPI_ISL_1679579, EPI_ISL_1679595, EPI_ISL_1679599, EPI_ISL_1679862, EPI_ISL_1679942, EPI_ISL_1679959, EPI_ISL_1679973, EPI_ISL_1680144, EPI_ISL_1680182, EPI_ISL_1680227, EPI_ISL_1680266, EPI_ISL_1680271, EPI_ISL_1680325, EPI_ISL_1690332, EPI_ISL_1690441, EPI_ISL_1690458, EPI_ISL_1690564, EPI_ISL_1690718, EPI_ISL_1690748, EPI_ISL_1690877, EPI_ISL_1690982, EPI_ISL_1691032, EPI_ISL_1691069, EPI_ISL_1691081, EPI_ISL_1691095, EPI_ISL_1691114, EPI_ISL_1691123, EPI_ISL_1691348, EPI_ISL_1691353, EPI_ISL_1694681, EPI_ISL_1701841, EPI_ISL_1701864, EPI_ISL_1734583, EPI_ISL_1734594, EPI_ISL_1734697, EPI_ISL_1734717, EPI_ISL_1734718, EPI_ISL_1734724, EPI_ISL_1734793, EPI_ISL_1734946, EPI_ISL_1734976, EPI_ISL_1734985, EPI_ISL_1734993, EPI_ISL_1735009, EPI_ISL_1735113, EPI_ISL_1735114, EPI_ISL_1735219, EPI_ISL_1735276, EPI_ISL_1735289, EPI_ISL_1735431, EPI_ISL_1735449, EPI_ISL_1735480, EPI_ISL_1735492, EPI_ISL_1735628, EPI_ISL_1735647, EPI_ISL_1735676, EPI_ISL_1753502, EPI_ISL_1753511, EPI_ISL_1796142, EPI_ISL_1796210, EPI_ISL_1796288, EPI_ISL_1796386, EPI_ISL_1796438, EPI_ISL_1796446, EPI_ISL_1796488, EPI_ISL_1796491, EPI_ISL_1796498, EPI_ISL_1796516, EPI_ISL_1796524, EPI_ISL_1796547, EPI_ISL_1796608, EPI_ISL_1796655, EPI_ISL_1796677, EPI_ISL_1796724, EPI_ISL_1796737, EPI_ISL_1796769, EPI_ISL_1796776, EPI_ISL_1796789, EPI_ISL_1796801, EPI_ISL_1796817, EPI_ISL_1796839, EPI_ISL_1796856, EPI_ISL_1796866, EPI_ISL_1796942, EPI_ISL_1796971, EPI_ISL_1796997, EPI_ISL_1797023, EPI_ISL_1797033, EPI_ISL_1797131, EPI_ISL_1797138, EPI_ISL_1797191, EPI_ISL_1797209, EPI_ISL_1797219, EPI_ISL_1797237, EPI_ISL_1797262, EPI_ISL_1797381, EPI_ISL_1797398, EPI_ISL_1797409, EPI_ISL_1803536, EPI_ISL_1803537, EPI_ISL_1803553, EPI_ISL_1803568, EPI_ISL_1803580, EPI_ISL_1803603, EPI_ISL_1803624, EPI_ISL_1803625, EPI_ISL_1803642, EPI_ISL_1803656, EPI_ISL_1803673, EPI_ISL_1803720, EPI_ISL_1803777, EPI_ISL_1803800, EPI_ISL_1803802, EPI_ISL_1803816, EPI_ISL_1803855, EPI_ISL_1803888, EPI_ISL_1803946, EPI_ISL_1803999, EPI_ISL_1804036, EPI_ISL_1804110, EPI_ISL_1804112, EPI_ISL_1804117, EPI_ISL_1804120, EPI_ISL_1804144, EPI_ISL_1804146, EPI_ISL_1804160, EPI_ISL_1804216, EPI_ISL_1804339, EPI_ISL_1804434, EPI_ISL_1804524, EPI_ISL_1804544, EPI_ISL_1804573, EPI_ISL_1804581, EPI_ISL_1804583, EPI_ISL_1804584, EPI_ISL_1804594, EPI_ISL_1804607, EPI_ISL_1804613, EPI_ISL_1804615, EPI_ISL_1804678, EPI_ISL_1804721, EPI_ISL_1804727, EPI_ISL_1817999, EPI_ISL_1818024, EPI_ISL_1818042, EPI_ISL_1906887, EPI_ISL_1906925, EPI_ISL_1907002, EPI_ISL_1907008, EPI_ISL_1907016, EPI_ISL_1907090, EPI_ISL_1907118, EPI_ISL_1907124, EPI_ISL_1907164, EPI_ISL_1907165, EPI_ISL_1907181, EPI_ISL_1907197, EPI_ISL_1907235, EPI_ISL_1907265, EPI_ISL_1907293, EPI_ISL_1907351, EPI_ISL_1907373, EPI_ISL_1907406, EPI_ISL_1907412, EPI_ISL_1907420, EPI_ISL_1907450, EPI_ISL_1907508, EPI_ISL_1907514, EPI_ISL_1907517, EPI_ISL_1907538, EPI_ISL_1907561, EPI_ISL_1907579, EPI_ISL_1907627, EPI_ISL_1907660, EPI_ISL_1907729, EPI_ISL_1907743, EPI_ISL_1907806, EPI_ISL_1907808, EPI_ISL_1907826, EPI_ISL_1907867, EPI_ISL_1907889, EPI_ISL_1907985, EPI_ISL_1908017, EPI_ISL_1908053, EPI_ISL_1908071, EPI_ISL_1908076, EPI_ISL_1908116, EPI_ISL_1908137, EPI_ISL_1923204, EPI_ISL_1923216, EPI_ISL_1923224, EPI_ISL_1923261, EPI_ISL_1923376, EPI_ISL_1923383, EPI_ISL_1923474, EPI_ISL_1923487, EPI_ISL_1923513, EPI_ISL_1923524, EPI_ISL_1923525, EPI_ISL_1923542, EPI_ISL_1923577, EPI_ISL_1923605, EPI_ISL_1923608, EPI_ISL_1923629, EPI_ISL_1923648, EPI_ISL_1923652, EPI_ISL_1923683, EPI_ISL_1923747, EPI_ISL_1923749, EPI_ISL_1923758, EPI_ISL_1923771, EPI_ISL_1923775, EPI_ISL_1923804, EPI_ISL_1923816, EPI_ISL_1923844, EPI_ISL_1923875, EPI_ISL_1923894, EPI_ISL_1923917, EPI_ISL_1923929, EPI_ISL_1923989, EPI_ISL_1924003, EPI_ISL_1924035, EPI_ISL_1924042, EPI_ISL_1924048, EPI_ISL_1924079, EPI_ISL_1924114, EPI_ISL_1924148, EPI_ISL_1924156, EPI_ISL_1924167, EPI_ISL_1924255, EPI_ISL_1924258, EPI_ISL_1935596, EPI_ISL_1942151, EPI_ISL_1942160, EPI_ISL_1990577, EPI_ISL_1990589, EPI_ISL_1990612, EPI_ISL_1990887, EPI_ISL_1990933, EPI_ISL_1990973, EPI_ISL_1991018, EPI_ISL_1991027, EPI_ISL_1991232, EPI_ISL_1991243, EPI_ISL_1991422, EPI_ISL_1991526, EPI_ISL_1991540, EPI_ISL_1991564, EPI_ISL_1991702, EPI_ISL_1991705, EPI_ISL_1991711, EPI_ISL_1991716, EPI_ISL_1991721, EPI_ISL_1991805 | | | |
| see above | Helix/Illumina | Centers for Disease Control and Prevention Division of Viral Diseases, Pathogen Discovery | Adrian Paskey; Alexandre Bolze; Ary Ascencio; Ben L. Rambo-Martin; Benjamin Rambo-Martin; Brad Sickler; Charlotte Rivera-Garcia; Christine Tran; Christopher Gulvick; Clinton R. Paden; Dakota Howard; Darlene Wagner; David Becker; Dhwani Batra; Duncan MacCannell; Efren Sandoval; Eileen de Feo; Elizabeth Cirulli; Eric Allen; Geraint Levan; James Lu; Jan Antico; Jason Caravas; Jason Nguyen; Jimmy Ramirez; Jingtao Liu; Kara Moser; Kelly Schiabor Barrett; Kim Gietzen; Magnus Isaksson; Marc Laurent; Matthew Schmerer; Matthew Tolentino; Nicole L. Washington; Peter W. Cook; Phil Febbo; Ryan Cho; Scott Sammons; Shannon Wickline; Shatavia Morrison; Sherry Wang; Simon White; Summer Galloway; Suxiang Tong; Tyler Cassens; William Lee; Yvette Unoarumhi |
| EPI_ISL_3149997, EPI_ISL_3149998, EPI_ISL_3149999, EPI_ISL_3150000, EPI_ISL_3150001, EPI_ISL_3150002, EPI_ISL_3150003, EPI_ISL_3150004, EPI_ISL_3150005, EPI_ISL_3150006, EPI_ISL_3150007, EPI_ISL_3150008, EPI_ISL_3150009, EPI_ISL_3150010, EPI_ISL_3150011, EPI_ISL_3150012, EPI_ISL_3150013, EPI_ISL_3150014, EPI_ISL_3150015, EPI_ISL_3150016, EPI_ISL_3150017 | | | |
| see above | Hermes Pardini | Universidade Federal de Ciencias da Saude de Porto Alegre | Andrea Roberto de Souza; Andy Goren; Carlos Gustavo Wambier; Claudia Elizabeth Thompson; Daniel do Nascimento Fonseca; Emilyn Oliveira Guerreiro; Flávio Adsuara Cadegiani; Liane Nanci Rotta; Patrícia Aline Gröhs Ferrareze; Ricardo Ariel Zimerman; Zhihua Ren |
| EPI_ISL_1262988 | Hopital | National Reference Center for Viruses of Respiratory Infections, Institut Pasteur, Paris | Angela Brisebarre; Camille Capel; Etienne Simon-Lorière; Marion Barbet; Maud Vanpeene; Méline Bizard; Sylvie Behillil; Sylvie van der Werf; Vincent Enouf; Woercel Isabelle |
| EPI_ISL_1811228 | Horquetas Rio Frio | Inciensa, Instituto Costarricense de Investigación y Enseñanza en Nutrición y Salud | Pérez-Corrales C & Montiel-Ulloa G |
| EPI_ISL_1533707 | Hosp Mun Planalto Waldomiro de Paula | Instituto Adolfo Lutz, Interdiciplinary Procedures Center, Strategic Laboratory | Caio Vinicius Dias Lopes; Claudia Regina Gonçalves; Claudio Tavares Sacchi; Erica Valessa Ramos Gomes; Karoline Rodrigues Campos; Leonardo Jose Tadeu de Araujo |
| EPI_ISL_1381213, EPI_ISL_1381216, EPI_ISL_1381217, EPI_ISL_1516921, EPI_ISL_1554911, EPI_ISL_1583049, EPI_ISL_1621510, EPI_ISL_1621822, EPI_ISL_1675020, EPI_ISL_1700617, EPI_ISL_1700647, EPI_ISL_1700648, EPI_ISL_1700650, EPI_ISL_1821591, EPI_ISL_2029664, EPI_ISL_2030088, EPI_ISL_2279592, EPI_ISL_2279593, EPI_ISL_2790909 | | | |
| see above | Hospital | National Reference Center for Viruses of Respiratory Infections, Institut Pasteur, Paris | Adrien Pain; Amaury Vaysse; Angela Brisebarre; Camille Capel; Christophe Malabat; Corinne Maufrais; CéLine Ramanantsoa; Damien Mornico; Eleha Guillotel; Emmanuelle Permal; Etienne Simon-Lorière; Frédéric Lemoine; Gael Millot; Isabelle Woercel; JéRome Guinard; Laurence Louvet; Louise Lefrançois; Louvet Laurence; Marianne Burgard; Marion Barbet; Maud Vanpeene; Méline Bizard; Nabil Gastli; Pascale Martres; Pierre Lechat; Rousset Dominique; Serazin ValéRie; StéPhanie Marque Juillet; Sylvie Behillil; Sylvie Van der Werf; Sylvie van der Werf; Vincent Enouf |
| EPI_ISL_1443196, EPI_ISL_1443197, EPI_ISL_1443198 | Hospital Aliança | Hospital São Rafael - IDOR | Aquiles Assunção Camelier; Aurea Angelica Paste; Bruno Solano de Freitas Souza; Carolina Kymie Vasques Nonaka; Elves A.P. Maciel; Isadora Cristina de Siqueira; Karoline Almeida Félix de Sousa; Margarida Celia L. C. Neves; Tiago Gräf; Victor Costa Araujo; Yasmin Santos Freitas Macêdo |
| EPI_ISL_1219021 | Hospital Aviccena | Instituto Adolfo Lutz, Interdiciplinary Procedures Center, Strategic Laboratory | Caio Vinicius Dias Lopes; Claudia Regina Gonçalves; Claudio Tavares Sacchi; Erica Valessa Ramos Gomes; Karoline Rodrigues Campos |
| EPI_ISL_906080, EPI_ISL_906081 | Hospital Beneficiencia Portuguesa | Instituto Adolfo Lutz, Interdiciplinary Procedures Center, Strategic Laboratory | Claudia Regina Gonçalves; Claudio Tavares Sacchi; Erica Valessa Ramos Gomes; Karoline Rodrigues Campos |
| EPI_ISL_940626, EPI_ISL_940627 | Hospital Central Sao Caetano do Sul | Instituto Adolfo Lutz, Interdiciplinary Procedures Center, Strategic Laboratory | Claudia Regina Gonçalves; Claudio Tavares Sacchi; Erica Valessa Ramos Gomes; Karoline Rodrigues Campos |
| EPI_ISL_1121307 | Hospital E Antonio Policarpo de Oliveira | Instituto Adolfo Lutz, Interdiciplinary Procedures Center, Strategic Laboratory | Caio Vinicius Dias Lopes; Claudia Regina Gonçalves; Claudio Tavares Sacchi; Erica Valessa Ramos Gomes; Karoline Rodrigues Campos |
| EPI_ISL_1303542 | Hospital Estadual de Campanha Barradas | Instituto Adolfo Lutz, Interdiciplinary Procedures Center, Strategic Laboratory | Caio Vinicius Dias Lopes; Claudia Regina Gonçalves; Claudio Tavares Sacchi; Erica Valessa Ramos Gomes; Karoline Rodrigues Campos |
| EPI_ISL_1498917 | Hospital Estadual de Mirandopolis | Instituto Adolfo Lutz, Interdiciplinary Procedures Center, Strategic Laboratory | Caio Vinicius Dias Lopes; Claudia Regina Gonçalves; Claudio Tavares Sacchi; Erica Valessa Ramos Gomes; Karoline Rodrigues Campos |

| | | | |
|---|---|---|---|
| EPI_ISL_1303537 | Hospital Estadual de Vila Alpina | Instituto Adolfo Lutz, Interdiciplinary Procedures Center, Strategic Laboratory | Caio Vinicius Dias Lopes; Claudia Regina Gonçalves; Claudio Tavares Sacchi; Erica Valessa Ramos Gomes; Karoline Rodrigues Campos |
| EPI_ISL_1533709 | Hospital Estadual de Vila Alpina Org Social Seconci Sao Paulo | Instituto Adolfo Lutz, Interdiciplinary Procedures Center, Strategic Laboratory | Caio Vinicius Dias Lopes; Claudia Regina Gonçalves; Claudio Tavares Sacchi; Erica Valessa Ramos Gomes; Karoline Rodrigues Campos; Leonardo Jose Tadeu de Araujo |
| EPI_ISL_2016620, EPI_ISL_2016621, EPI_ISL_2016622, EPI_ISL_2016623 | Hospital Francesc de Borja (Gandia) | SeqCOVID-SPAIN consortium/IBV(CSIC) | Inmaculada Vidal Catalá; Isabel García Nava and SeqCOVID-SPAIN consortium; Mª Rosa Navarro Cots; Nieves Orta Mira |
| EPI_ISL_985404, EPI_ISL_1307715, EPI_ISL_1381872, EPI_ISL_1401467, EPI_ISL_1476981, EPI_ISL_1477061, EPI_ISL_1524839, EPI_ISL_1524840, EPI_ISL_1524841, EPI_ISL_1524842, EPI_ISL_1647783, EPI_ISL_1647841, EPI_ISL_1647852, EPI_ISL_1647854, EPI_ISL_1647855, EPI_ISL_1647858, EPI_ISL_1647859, EPI_ISL_1647945, EPI_ISL_1647946, EPI_ISL_1914363, EPI_ISL_1914364, EPI_ISL_1914366, EPI_ISL_1914367, EPI_ISL_1914368, EPI_ISL_3505904 | | | |
| see above | Hospital General Universitario Gregorio Marañón | Hospital General Universitario Gregorio Marañón | Cristina Rodriguez-Grande; Darío García de Viedma; Darío García de Viedma.; Julia Suárez; Laura Pérez-Lago; Marta Herranz Martin; Patricia Muñoz; Pedro Sola Campoy; Pilar Catalán; Sergio Buenestado Serrano; Victor Manuel de la Cueva |
| EPI_ISL_2271475 | Hospital General Universitario de Alicante - Instituto de Investigación Sanitaria y Biomédica de Alicante | SeqCOVID-SPAIN consortium/IBV(CSIC) | Carmen Molina Pardines and SeqCOVID-SPAIN consortium; Maripaz Ventero Martín |
| EPI_ISL_2614600 | Hospital Geral de Guaianazes | Instituto Adolfo Lutz, Interdiciplinary Procedures Center, Strategic Laboratory | Caio Vinicius Dias Lopes; Claudia Regina Gonçalves; Claudio Tavares Sacchi; Erica Valessa Ramos Gomes; Karoline Rodrigues Campos; Leonardo Jose Tadeu de Araujo |
| EPI_ISL_1533725 | Hospital Geral de Guarulhos | Instituto Adolfo Lutz, Interdiciplinary Procedures Center, Strategic Laboratory | Caio Vinicius Dias Lopes; Claudia Regina Gonçalves; Claudio Tavares Sacchi; Erica Valessa Ramos Gomes; Karoline Rodrigues Campos; Leonardo Jose Tadeu de Araujo |
| EPI_ISL_1533723 | Hospital Geral de Itaquaquecetuba | Instituto Adolfo Lutz, Interdiciplinary Procedures Center, Strategic Laboratory | Caio Vinicius Dias Lopes; Claudia Regina Gonçalves; Claudio Tavares Sacchi; Erica Valessa Ramos Gomes; Karoline Rodrigues Campos; Leonardo Jose Tadeu de Araujo |
| EPI_ISL_2614580 | Hospital Geral de Pedreira | Instituto Adolfo Lutz, Interdiciplinary Procedures Center, Strategic Laboratory | Caio Vinicius Dias Lopes; Claudia Regina Gonçalves; Claudio Tavares Sacchi; Erica Valessa Ramos Gomes; Karoline Rodrigues Campos; Leonardo Jose Tadeu de Araujo |
| EPI_ISL_940630, EPI_ISL_943967, EPI_ISL_943968, EPI_ISL_943969, EPI_ISL_943970, EPI_ISL_943971 | Hospital Geral de Sao Paulo | Instituto Adolfo Lutz, Interdiciplinary Procedures Center, Strategic Laboratory | Claudia Regina Gonçalves; Claudio Tavares Sacchi; Erica Valessa Ramos Gomes; Karoline Rodrigues Campos |
| EPI_ISL_906075 | Hospital Geral de Vila Penteado Dr Jose Pangella Sao Paulo | Instituto Adolfo Lutz, Interdiciplinary Procedures Center, Strategic Laboratory | Claudia Regina Gonçalves; Claudio Tavares Sacchi; Erica Valessa Ramos Gomes; Karoline Rodrigues Campos |
| EPI_ISL_1381069 | Hospital Heliopolis | Instituto Adolfo Lutz, Interdiciplinary Procedures Center, Strategic Laboratory | Caio Vinicius Dias Lopes; Claudia Regina Gonçalves; Claudio Tavares Sacchi; Erica Valessa Ramos Gomes; Karoline Rodrigues Campos |
| EPI_ISL_2551531 | Hospital Infantil Menino Jesus | Laboratório de Virologia Clínica e Molecular | Alex Fiorini; Ana Paula Salles Fernandes; Bruna Larotonda Telezynski; Danielle Bruna Leal Oliveira; Edison Luiz Durigon; Erick Gustavo Dorlass; Flavio Fonseca e Santuza Teixeira; Guilherme Pereira Scagion; Helena Perez Coelho; Hugo Sato; Karine Lima Lourenço; Luciano Matsumiya Thomazelli; Renata Peixoto; Rubens Daniel Miserani Magalhães; Tatiana Ometto |
| EPI_ISL_2551532 | Hospital Infantnil Darcy Vargas | Laboratório de Virologia Clínica e Molecular | Alex Fiorini; Ana Paula Salles Fernandes; Bruna Larotonda Telezynski; Danielle Bruna Leal Oliveira; Edison Luiz Durigon; Erick Gustavo Dorlass; Flavio Fonseca e Santuza Teixeira; Guilherme Pereira Scagion; Helena Perez Coelho; Hugo Sato; Karine Lima Lourenço; Luciano Matsumiya Thomazelli; Renata Peixoto; Rubens Daniel Miserani Magalhães; Tatiana Ometto |
| EPI_ISL_2363535, EPI_ISL_2363555, EPI_ISL_2363558, EPI_ISL_2363563, EPI_ISL_2363564, EPI_ISL_2363565 | Hospital Jaime Ferre - SAMCO Rafaela | Grupo de Genómica y Bioinformática del Instituto de Investigación de la Cadena Láctea CONICET-INTA on behalf of 'Proyecto Argentino Interinstitucional de genomica de SARS-CoV-2' (PAIS Consortium) | AF; Amadio; C; Eberhardt; Irazoqui; Isaia; JF; JM; MF; Pandolfi; Quaranta; V |
| EPI_ISL_3031345, EPI_ISL_3031346 | Hospital Metropolitano Dr. Célio de Castro | Instituto René Rachou / Fiocruz Minas | Anna Salim; Cristina Fonseca; Gabriel Fernandes; Mariana Melo; Núbia Fernandes; Pedro Alves; Rosiane Pereira; Rubens do Monte Neto; Sandra Gava; Thaís Santos; Thaís Silva; Wilma Patrícia Bernardes |
| EPI_ISL_2614577 | Hospital Municipal Antonio Giglio | Instituto Adolfo Lutz, Interdiciplinary Procedures Center, Strategic Laboratory | Caio Vinicius Dias Lopes; Claudia Regina Gonçalves; Claudio Tavares Sacchi; Erica Valessa Ramos Gomes; Karoline Rodrigues Campos; Leonardo Jose Tadeu de Araujo |
| EPI_ISL_1381070 | Hospital Municipal Cidade Tiradentes Carmem Prudente | Instituto Adolfo Lutz, Interdiciplinary Procedures Center, Strategic Laboratory | Caio Vinicius Dias Lopes; Claudia Regina Gonçalves; Claudio Tavares Sacchi; Erica Valessa Ramos Gomes; Karoline Rodrigues Campos |
| EPI_ISL_1303541 | Hospital Municipal Cidade Tiradentes Carmen Prudente | Instituto Adolfo Lutz, Interdiciplinary Procedures Center, Strategic Laboratory | Caio Vinicius Dias Lopes; Claudia Regina Gonçalves; Claudio Tavares Sacchi; Erica Valessa Ramos Gomes; Karoline Rodrigues Campos |
| EPI_ISL_1533695 | Hospital Municipal Dr Mario Gatti | Instituto Adolfo Lutz, Interdiciplinary Procedures Center, Strategic Laboratory | Caio Vinicius Dias Lopes; Claudia Regina Gonçalves; Claudio Tavares Sacchi; Erica Valessa Ramos Gomes; Karoline Rodrigues Campos; Leonardo Jose Tadeu de Araujo |
| EPI_ISL_1731577, EPI_ISL_2003113 | Hospital Municipal Dr Mario Gatti Campinas | Instituto Adolfo Lutz, Interdiciplinary Procedures Center, Strategic Laboratory | Caio Vinicius Dias Lopes; Claudia Regina Gonçalves; Claudio Tavares Sacchi; Erica Valessa Ramos Gomes; Karoline Rodrigues Campos; Katia Correa de Oliveira Santos; Leonardo Jose Tadeu de Araujo |
| EPI_ISL_2614590 | Hospital Municipal Dr. Guido Guida | Instituto Adolfo Lutz, Interdiciplinary Procedures Center, Strategic Laboratory | Caio Vinicius Dias Lopes; Claudia Regina Gonçalves; Claudio Tavares Sacchi; Erica Valessa Ramos Gomes; Karoline Rodrigues Campos; Leonardo Jose Tadeu de Araujo |
| EPI_ISL_1620638 | Hospital Municipal Dr. Ignacio Proença de Gouvea | Instituto Butantan / Mendelics | Alexander Roberto Precioso; Antonio Jorge Martins; Bibiana Santos; Claudia Renata dos Santos Barros; David Schlesinger; Debora Botequio Moretti; Dimas Tadeu Covas; Elaine Cristina Marqueze; Elaine Vieira dos Santos; Erika Freitas; Evandra Strazza Rodrigues; Flavia Aburjaile; José Salvatore Leister Patané; João Paulo Kitajima; Luiz Carlos Junior de Alcantara; Maria Carolina Elias; Marta Giovanetti; Rafael dos Santos Bezerra; Raul Machado Neto; Ricardo Haddad; Rodrigo Tocantins Calado.; Sandra Coccuzzo Sampaio; Simone Kashima; Svetoslav Nanev Slavov; Vagner Fonseca; Vincent Louis Viala |
| EPI_ISL_2614572 | Hospital Municipal Ermelino Matarazzo | Instituto Adolfo Lutz, Interdiciplinary | Caio Vinicius Dias Lopes; Claudia Regina Gonçalves; Claudio Tavares Sacchi; Erica Valessa Ramos Gomes; Karoline Rodrigues Campos; Leonardo Jose Tadeu de Araujo |

| | | | |
|---|---|---|---|
| | | Procedures Center, Strategic Laboratory | |
| EPI_ISL_2614597 | Hospital Municipal de Mogi das Cruzes | Instituto Adolfo Lutz, Interdiciplinary Procedures Center, Strategic Laboratory | Caio Vinicius Dias Lopes; Claudia Regina Gonçalves; Claudio Tavares Sacchi; Erica Valessa Ramos Gomes; Karoline Rodrigues Campos; Leonardo Jose Tadeu de Araujo |
| EPI_ISL_1533696 | Hospital Municipal de Pedreira | Instituto Adolfo Lutz, Interdiciplinary Procedures Center, Strategic Laboratory | Caio Vinicius Dias Lopes; Claudia Regina Gonçalves; Claudio Tavares Sacchi; Erica Valessa Ramos Gomes; Karoline Rodrigues Campos; Leonardo Jose Tadeu de Araujo |
| EPI_ISL_1358287 | Hospital Nipo Brasileiro | Instituto Adolfo Lutz, Interdiciplinary Procedures Center, Strategic Laboratory | Caio Vinicius Dias Lopes; Claudia Regina Gonçalves; Claudio Tavares Sacchi; Erica Valessa Ramos Gomes; Karoline Rodrigues Campos |
| EPI_ISL_2614594 | Hospital Regional de Cotia | Instituto Adolfo Lutz, Interdiciplinary Procedures Center, Strategic Laboratory | Caio Vinicius Dias Lopes; Claudia Regina Gonçalves; Claudio Tavares Sacchi; Erica Valessa Ramos Gomes; Karoline Rodrigues Campos; Leonardo Jose Tadeu de Araujo |
| EPI_ISL_940619, EPI_ISL_940620, EPI_ISL_940621, EPI_ISL_940622, EPI_ISL_940623, EPI_ISL_940624, EPI_ISL_940625 | | | |
| see above | Hospital Sao Joaquim - Beneficiencia Portuguesa | Instituto Adolfo Lutz, Interdiciplinary Procedures Center, Strategic Laboratory | Claudia Regina Gonçalves; Claudio Tavares Sacchi; Erica Valessa Ramos Gomes; Karoline Rodrigues Campos |
| EPI_ISL_906076, EPI_ISL_906077 | Hospital Sao Luiz Sao Caetano | Instituto Adolfo Lutz, Interdiciplinary Procedures Center, Strategic Laboratory | Claudia Regina Gonçalves; Claudio Tavares Sacchi; Erica Valessa Ramos Gomes; Karoline Rodrigues Campos |
| EPI_ISL_1628372 | Hospital Sao Marcos da Sama Morro Agudo | Instituto Adolfo Lutz, Interdiciplinary Procedures Center, Strategic Laboratory | Caio Vinicius Dias Lopes; Claudia Regina Gonçalves; Claudio Tavares Sacchi; Erica Valessa Ramos Gomes; Karoline Rodrigues Campos; Katia Correa de Oliveira Santos; Leonardo Jose Tadeu de Araujo |
| EPI_ISL_3860415 | Hospital Señor del Milagro | Instituto de Patologia Vegetal (CIAP-INTA) on behalf of 'Proyecto Argentino Interinstitucional de genomica de SARS-CoV-2' (PAIS Consortium) | Debat; FD; Fernanda Agüero; Fernández; HJ; Lavaque Esteban; Marquez, N.; Pamela Cajal; Raskovsky Viviana; Veronica Lesser |
| EPI_ISL_1608161, EPI_ISL_1608162, EPI_ISL_1608163, EPI_ISL_1608164, EPI_ISL_1608165, EPI_ISL_1608166, EPI_ISL_1608167, EPI_ISL_1608168, EPI_ISL_1608169, EPI_ISL_1608170, EPI_ISL_1608171 | | | |
| see above | Hospital São Rafael - IDOR | Hospital São Rafael - IDOR | Ana Verena Almeida Mendes; Bruno Solano de Freitas Souza; Camila Araújo de Lorenzo Barcia; Carolina Kymie Vasques Nonaka; Clarissa Araújo Gurgel Rocha; Ian Marinho Santos; Iasmin Nogueira Bastos; Isadora Cristina de Siqueira; Janderson Lopes de Oliveira; Karoline Almeida Felix de Sousa; Maria Clara Brito de Santana; Rogério da Hora Passos; Thamires Gomes Lopes Weber; Tiago Gräf; Vanessa Ferreira Costa |
| EPI_ISL_1963916 | Hospital Universitari Bellvitge | Microbiology Department | Aida Gonzalez-Diaz; Carmen Ardanuy; Jordi Camara; Jordi Niubó; Laura Calatayud; M Angeles Domínguez; Miguel Fernandez-Huerta; Sara Marti |
| EPI_ISL_1250798 | Hospital Universitari Vall d'Hebron - Vall d'Hebron Institut de Recerca | Hospital Universitari Vall d'Hebron - Vall d'Hebron Institut de Recerca | Andrés Antón; Ariadna Rando; Carla Castillo; Cristina Andrés; Damir Garcia-Cehic; Josep F Abril; Josep Quer; Juliana Esperalba; Maria Carmen Martin; Maria Gema Codina; Maria Piñana; Tomàs Pumarola |
| EPI_ISL_1538412 | Hospital Universitario Basurto Clinical Microbiology Service | Biocruces Bizkaia | Ana Belen de la Hoz; Estibaliza Ugalde Zarraga; Jose Luis Díaz de Tuesta del Arco; Mikel Gallego Rodrigo; Mikel Joseba Urrutikoetxea-Gutierrez; Mª Carmen Nieto Toboso |
| EPI_ISL_1533702, EPI_ISL_1533712 | Hospital Universitario da USP Sao Paulo | Instituto Adolfo Lutz, Interdiciplinary Procedures Center, Strategic Laboratory | Caio Vinicius Dias Lopes; Claudia Regina Gonçalves; Claudio Tavares Sacchi; Erica Valessa Ramos Gomes; Karoline Rodrigues Campos; Leonardo Jose Tadeu de Araujo |
| EPI_ISL_1821210 | Hospital Universitario de Marilia | Instituto Adolfo Lutz, Interdiciplinary Procedures Center, Strategic Laboratory | Caio Vinicius Dias Lopes; Claudia Regina Gonçalves; Claudio Tavares Sacchi; Erica Valessa Ramos Gomes; Karoline Rodrigues Campos; Leonardo Jose Tadeu de Araujo |
| EPI_ISL_2614569 | Hospital Universitário da USP | Instituto Adolfo Lutz, Interdiciplinary Procedures Center, Strategic Laboratory | Caio Vinicius Dias Lopes; Claudia Regina Gonçalves; Claudio Tavares Sacchi; Erica Valessa Ramos Gomes; Karoline Rodrigues Campos; Leonardo Jose Tadeu de Araujo |
| EPI_ISL_3031312, EPI_ISL_3031316, EPI_ISL_3031317, EPI_ISL_3031318 | Hospital da Baleia | Instituto René Rachou / Fiocruz Minas | Alana Oliveira; Anna Salim; Camila Corsini; Daniel Miranda; Gabriel Fernandes; Mozar de Castro; Nathalie Almeida; Pedro Alves; Priscilla Filgueiras; Rafaella Fortini; Raphael Silva; Raquel Vilela; Rubens do Monte Neto; Sarah Gomes; Thaís Silva; Wander Jeremias |
| EPI_ISL_1533699, EPI_ISL_2614575 | Hospital das Clinicas Luzia de Pinho Melo | Instituto Adolfo Lutz, Interdiciplinary Procedures Center, Strategic Laboratory | Caio Vinicius Dias Lopes; Claudia Regina Gonçalves; Claudio Tavares Sacchi; Erica Valessa Ramos Gomes; Karoline Rodrigues Campos; Leonardo Jose Tadeu de Araujo |
| EPI_ISL_2614598 | Hospital de Campanha Covid-19 Mairipora | Instituto Adolfo Lutz, Interdiciplinary Procedures Center, Strategic Laboratory | Caio Vinicius Dias Lopes; Claudia Regina Gonçalves; Claudio Tavares Sacchi; Erica Valessa Ramos Gomes; Karoline Rodrigues Campos; Leonardo Jose Tadeu de Araujo |
| EPI_ISL_875689 | Hospital do Servidor Publico | Instituto Adolfo Lutz, Interdiciplinary Procedures Center, Strategic Laboratory | Claudia Regina Gonçalves; Claudio Tavares Sacchi; Erica Valessa Ramos Gomes; Karoline Rodrigues Campos |
| EPI_ISL_1121308 | Hospital e Pronto Socorro Portinari | Instituto Adolfo Lutz, Interdiciplinary Procedures Center, Strategic Laboratory | Caio Vinicius Dias Lopes; Claudia Regina Gonçalves; Claudio Tavares Sacchi; Erica Valessa Ramos Gomes; Karoline Rodrigues Campos |
| EPI_ISL_2003926, EPI_ISL_2003956, EPI_ISL_2003966, EPI_ISL_2597572, EPI_ISL_2597577, EPI_ISL_2653036, EPI_ISL_3022828, EPI_ISL_3022865, EPI_ISL_3023231 | | | |
| see above | Hospital of the University of Pennsylvania Molecular Pathology Lab | Bushman Lab - University of Pennsylvania | Abigail Glascock; Aoife M. Roche; Arupa Ganguly; Ayannah S. Fitzgerald; Brendan Kelly; Jevon Graham-Wooten; John Everett; John K. Everett; Kyle Rodino; Layla A. Khatib; Mike Feldman; Pascha Hokama; Ronald G. Collman and Frederic Bushan; Ronald G. Collman and Frederic Bushman; Samantha A. Whiteside; Scott Sherrill-Mix; Shantan Reddy; Young Hwang |
| EPI_ISL_1479113, EPI_ISL_1672871 | Houston Health Dept. | Houston Health Dept. | Adolpho Lara; Pamela Brown; Ryker Penn; Yanlai Lai |
| EPI_ISL_1078954, EPI_ISL_1079206, EPI_ISL_1079246, EPI_ISL_1079917, EPI_ISL_1237447, EPI_ISL_1238196, EPI_ISL_1305221, EPI_ISL_1305222, EPI_ISL_1305226, EPI_ISL_2190495, EPI_ISL_2191170, EPI_ISL_2191231, EPI_ISL_2200291, EPI_ISL_2201494, EPI_ISL_2203377, EPI_ISL_2204974, EPI_ISL_2205669, EPI_ISL_2211977, EPI_ISL_2212003, EPI_ISL_2212042, EPI_ISL_2213726, EPI_ISL_2216599, EPI_ISL_2216645, EPI_ISL_2216933, EPI_ISL_2216950, EPI_ISL_2217100, EPI_ISL_2221466, EPI_ISL_2221525, EPI_ISL_2222823, EPI_ISL_2222893, EPI_ISL_2223003, EPI_ISL_2223017, EPI_ISL_2224440 | | | |
| see above | Houston Methodist Hospital | Houston Methodist Hospital | Ilya J. Finkelstein; James J. Davis; Jessica Cambric; Jimmy Gollihar; Kristina Reppond; Layne Pruitt; Madison N. Shyer; Marcus Nguyen; Matthew Ojeda Saavedra; Paul A. Christensen; Prasanti Yerramilli; Randall J. Olsen; Robert Olson; Ryan Gadd; S. Wesley Long; Sishir Subedi; and James M. Musser |
| EPI_ISL_1756322 | Hôpital Ambroise-Paré | Department of Virology, Henri Mondor University Hospital, | Alexandre Soulier; Christophe Rodriguez; Elisabeth Trawinski; Guillaume Gricourt; Jean-Michel Pawlotsky; Melissa N'Debi; Slim Fourati; Vanessa Demontant |

| | | | |
|---|---|---|---|
| | | Assistance Publique Hôpitaux de Paris, Université Paris-Est Créteil, INSERM U955 | |
| EPI_ISL_1754996, EPI_ISL_1755133, EPI_ISL_1755809, EPI_ISL_1757613 | Hôpital Avicenne | Department of Virology, Henri Mondor University Hospital, Assistance Publique Hôpitaux de Paris, Université Paris-Est Créteil, INSERM U955 | Alexandre Soulier; Christophe Rodriguez; Elisabeth Trawinski; Guillaume Gricourt; Jean-Michel Pawlotsky; Melissa N'Debi; Slim Fourati; Vanessa Demontant |
| EPI_ISL_1757581 | Hôpital Cochin | Department of Virology, Henri Mondor University Hospital, Assistance Publique Hôpitaux de Paris, Université Paris-Est Créteil, INSERM U955 | Alexandre Soulier; Christophe Rodriguez; Elisabeth Trawinski; Guillaume Gricourt; Jean-Michel Pawlotsky; Melissa N'Debi; Slim Fourati; Vanessa Demontant |
| EPI_ISL_1404536, EPI_ISL_1404538 | Hôpital Henri Mondor | Department of Virology, Henri Mondor University Hospital, Assistance Publique Hôpitaux de Paris, Université Paris-Est Créteil, INSERM U955 | Alexandre Soulier; Christophe Rodriguez; Elisabeth Trawinski; Guillaume Gricourt; Jean-Michel Pawlotsky; Melissa N'Debi; Slim Fourati; Vanessa Demontant |
| EPI_ISL_2614544, EPI_ISL_2614545, EPI_ISL_2614546, EPI_ISL_2614547, EPI_ISL_2614549, EPI_ISL_2614550, EPI_ISL_2614551, EPI_ISL_2614552, EPI_ISL_2614553, EPI_ISL_2614555, EPI_ISL_2614557, EPI_ISL_2614558, EPI_ISL_2614559, EPI_ISL_2614560, EPI_ISL_2614561, EPI_ISL_2614562, EPI_ISL_2614563 | | | |
| see above | IAL Presidente Prudente | Instituto Adolfo Lutz, Interdiciplinary Procedures Center, Strategic Laboratory | Caio Vinicius Dias Lopes; Claudia Regina Gonçalves; Claudio Tavares Sacchi; Erica Valessa Ramos Gomes; Karoline Rodrigues Campos; Leonardo Jose Tadeu de Araujo |
| EPI_ISL_1358288, EPI_ISL_1358289, EPI_ISL_1358290 | IAL Regional de Aracatuba | Instituto Adolfo Lutz, Interdiciplinary Procedures Center, Strategic Laboratory | Caio Vinicius Dias Lopes; Claudia Regina Gonçalves; Claudio Tavares Sacchi; Erica Valessa Ramos Gomes; Karoline Rodrigues Campos |
| EPI_ISL_1086044, EPI_ISL_1096131, EPI_ISL_1096132, EPI_ISL_1096134, EPI_ISL_1121310, EPI_ISL_1121313, EPI_ISL_1121314 | | | |
| see above | IAL Regional de Bauru | Instituto Adolfo Lutz, Interdiciplinary Procedures Center, Strategic Laboratory | Caio Vinicius Dias Lopes; Claudia Regina Gonçalves; Claudio Tavares Sacchi; Erica Valessa Ramos Gomes; Karoline Rodrigues Campos |
| EPI_ISL_1171619, EPI_ISL_1171653, EPI_ISL_1171656, EPI_ISL_1171657, EPI_ISL_1171658, EPI_ISL_1171666, EPI_ISL_1171672, EPI_ISL_1171674 | | | |
| see above | IAL Regional de Presidente Prudente | Instituto Adolfo Lutz, Interdiciplinary Procedures Center, Strategic Laboratory | Caio Vinicius Dias Lopes; Claudia Regina Gonçalves; Claudio Tavares Sacchi; Erica Valessa Ramos Gomes; Karoline Rodrigues Campos |
| EPI_ISL_1139070 | IAL Regional de Ribeirao Preto | Instituto Adolfo Lutz, Interdiciplinary Procedures Center, Strategic Laboratory | Caio Vinicius Dias Lopes; Claudia Regina Gonçalves; Claudio Tavares Sacchi; Erica Valessa Ramos Gomes; Karoline Rodrigues Campos |
| EPI_ISL_1381048, EPI_ISL_1381061, EPI_ISL_1381062, EPI_ISL_1381065 | IAL Regional de Santo Andre | Instituto Adolfo Lutz, Interdiciplinary Procedures Center, Strategic Laboratory | Caio Vinicius Dias Lopes; Claudia Regina Gonçalves; Claudio Tavares Sacchi; Erica Valessa Ramos Gomes; Karoline Rodrigues Campos |
| EPI_ISL_1219025, EPI_ISL_1293059, EPI_ISL_1293063, EPI_ISL_1293067, EPI_ISL_1293077, EPI_ISL_1293080 | IAL Regional de Sorocaba | Instituto Adolfo Lutz, Interdiciplinary Procedures Center, Strategic Laboratory | Caio Vinicius Dias Lopes; Claudia Regina Gonçalves; Claudio Tavares Sacchi; Erica Valessa Ramos Gomes; Karoline Rodrigues Campos |
| EPI_ISL_1303520, EPI_ISL_1303522, EPI_ISL_1303525, EPI_ISL_1303529 | IAL Regional de São Jose do Rio Preto | Instituto Adolfo Lutz, Interdiciplinary Procedures Center, Strategic Laboratory | Caio Vinicius Dias Lopes; Claudia Regina Gonçalves; Claudio Tavares Sacchi; Erica Valessa Ramos Gomes; Karoline Rodrigues Campos |
| EPI_ISL_1960074 | ICMT-APARTADO | Universidad Nacional de Colombia - Laboratorio Genómico One Health | Andres F. Cardona-Rios; Carlos Franco-Muñoz; Carolina Muñoz-Arango; Celeny Ortiz; Daniel O. Maldonado-Perez; Diego A. Álvarez-Díaz; Hector Alejandro Ruiz-Moreno; Idabely Betancur Ortiz; Jorge E. Osorio; Juan P. Hernandez-Ortiz; Karl A Ciuoderis; Katherine Laiton-Donato; Laura Silvana Perez; Lina M. Hurtado; Marcela Mercado-Reyes; Maria Angélica Maya; Maria Stella López; Rita Almanza Payares; Sandra Ines Cano; Simón Villegas Velásquez |
| EPI_ISL_2234873, EPI_ISL_2234874, EPI_ISL_2234875, EPI_ISL_2234886, EPI_ISL_2234900, EPI_ISL_2234903, EPI_ISL_2234905, EPI_ISL_2444776, EPI_ISL_2444778, EPI_ISL_2444779, EPI_ISL_2444781, EPI_ISL_2444782, EPI_ISL_2444787, EPI_ISL_2444789, EPI_ISL_2444790, EPI_ISL_2444791, EPI_ISL_2444798, EPI_ISL_2444801, EPI_ISL_2444805, EPI_ISL_2444808, EPI_ISL_2444809, EPI_ISL_2444811, EPI_ISL_2444816, EPI_ISL_2444818, EPI_ISL_2444831, EPI_ISL_2444834, EPI_ISL_2444837, EPI_ISL_4071892, EPI_ISL_4071893 | | | |
| see above | IICS-UNA | IICS-UNA | Adriana Valenzuela; Alejandra Rojas; Chyntia Diaz; Eva Nara; Fatima Cardozo; Florencia del Puerto; Joel Ortiz; Jonas Fernandez; Laura Franco; Laura Mendoza; Leticia Rojas; Magaly Martinez; Maria Eugenia Galeano. |
| EPI_ISL_3912230 | IJF INSTITUTO DR JOSE FROTA CENTRAL | Analytical Competence Molecular Epidemiology Lab/ACME, Oswaldo Cruz Foundation, Ceara (FIOCRUZ CE) | Cleber Furtado Aksenen; Fabio Miyajima; Fernando Braga Stehling; Francisco Eder de Moura Lopes; Jamille Maria Mendes Bezerra; Joaquim Cesar do Nascimento Sousa Junior; Pedro Miguel Carneiro Jeronimo; Suzana Porto Almeida & Lucas Delerino on behalf of COVID-19 FIOCRUZ Genomic Network; Thais Ferreira de Oliveira; Thais de Oliveira Costa; Ticiane Cavalcante de Souza; Veridiana Pessoa Miyajima |
| EPI_ISL_1446979, EPI_ISL_1823782, EPI_ISL_1823788 | IL Department of Public Health Chicago Laboratory | Centers for Disease Control and Prevention Division of Viral Diseases, Pathogen Discovery | Alison Laufer Halpin; Ben L. Rambo-Martin; Clinton R. Paden; Dakota Howard; Darlene Wagner; Dave Wentworth; Dhwani Batra; Jasmine Padilla; Justin Lee; Katie Dillon; Krista Queen; Kristen Knipe; Kristine Lacek; Mark Burroughs; Matthew Schmerer; Mili Sheth; Peter Cook; Sam Shepard; Sarah Nobles; Shoshona Le; Suxiang Tong; Vivien Dugan; Yvette Unoarumhi |
| EPI_ISL_1509934 | IL Department of Public Health Chicago Laboratory | Genomics and Discovery, Respiratory Viruses Branch, Division of Viral Diseases, Centers for Disease Control and Prevention | Adam Retchless; Anna Kelleher; Anna Montmayeur; Anna Uehara; Brian Lynch; Clinton R. Paden; Haibin Wang; Han Jia Justin Ng; Jing Zhang; Justin Lee; Krista Queen; Mark Burroughs; Peter Cook; Rachel Marine; Suxiang Tong; Yan Li; Ying Tao |
| EPI_ISL_1798496, EPI_ISL_1798498, EPI_ISL_1798508, EPI_ISL_1798561, EPI_ISL_1798583, EPI_ISL_1937768, EPI_ISL_1937770, EPI_ISL_1937771, EPI_ISL_1937777, EPI_ISL_1937780, EPI_ISL_1937781, EPI_ISL_1937783, EPI_ISL_1937784, EPI_ISL_1937785, EPI_ISL_1989977, EPI_ISL_3354527, EPI_ISL_3354528, EPI_ISL_3354530, EPI_ISL_3354531, EPI_ISL_3354532, EPI_ISL_3354536, EPI_ISL_3354545 | | | |
| see above | IL Dept. of Public Health Springfield Laboratory | Centers for Disease Control and Prevention Division of Viral Diseases, Pathogen Discovery | Alex Burgin; Alison Laufer Halpin; Ben L. Rambo-Martin; Ben Rambo-Martin; Clinton Paden; Clinton R. Paden; Dakota Howard; Darlene Wagner; Dave Wentworth; Dhwani Batra; Jasmine Padilla; Justin Lee; Katie Dillon; Krista Queen; Kristen Knipe; Kristine Lacek; Mark Burroughs; Matthew Schmerer; Meghan Bentz; Mili Sheth; Peter Cook; Sam Shepard; Sarah Nobles; Shoshona Le; Suxiang Tong; Vivien Dugan; Yvette Unoarumhi |
| EPI_ISL_1408896, EPI_ISL_1509935 | IL Dept. of Public Health Springfield Laboratory | Genomics and Discovery, Respiratory Viruses Branch, Division of Viral Diseases, Centers for Disease Control and Prevention | Adam Retchless; Anna Kelleher; Anna Montmayeur; Anna Uehara; Brian Lynch; Clinton R. Paden; Haibin Wang; Han Jia Justin Ng; Jing Zhang; Justin Lee; Krista Queen; Mark Burroughs; Peter Cook; Rachel Marine; Suxiang Tong; Yan Li; Ying Tao |
| EPI_ISL_1639313, | IMD - Medizinisches | Robert Koch Institute | |

| | | | |
|---|---|---|---|
| EPI_ISL_1639325, EPI_ISL_1722204, EPI_ISL_1722214, EPI_ISL_1722223 | Labor Rostock | | |
| EPI_ISL_1399627 | IMM | Instituto Nacional de Saude (INSA) | Borges et al |
| EPI_ISL_1213173, EPI_ISL_1213175, EPI_ISL_1213177, EPI_ISL_1213178, EPI_ISL_1213180, EPI_ISL_1213182, EPI_ISL_1213183, EPI_ISL_1213185, EPI_ISL_1213187, EPI_ISL_1213192, EPI_ISL_1213194, EPI_ISL_1213196, EPI_ISL_1213197, EPI_ISL_1213199 | | | |
| see above | IMT-UFRN/RN | Bioinformatics Laboratory / LNCC | Alessandra P Lamarca; Alexandra L Gerber; Ana Paula Melo Mariano; Ana Paula de C Guimarães; Ana Tereza R Vasconcelos; Angela Maria Guimarães Santos; Bianca Mendes Maciel; Danielle Angst Secco; Eduardo Sérgio Soares Sousa; Eloiza Helena Campana; Francisco Paulo Freire Neto; George Rego Albuquerque; Kátia Castanho Scortecci; Lucymara Fassarella Agnez Lima; Luiz G P de Almeida; Luís Cristóvão Porto; Otavio J. Brustolini; Paulo Ricardo Nascimento; Ronaldo da Silva Francisco Jr; Sandra Rocha Gadelha; Selma Maria Bezerra Jeronimo; Vinícius Pietta Perez |
| EPI_ISL_1824049 | IN State Department of Health Laboratory Services | Centers for Disease Control and Prevention Division of Viral Diseases, Pathogen Discovery | Alison Laufer Halpin; Ben L. Rambo-Martin; Clinton R. Paden; Dakota Howard; Darlene Wagner; Dave Wentworth; Dhwani Batra; Jasmine Padilla; Justin Lee; Katie Dillon; Krista Queen; Kristen Knipe; Kristine Lacek; Mark Burroughs; Matthew Schmerer; Mili Sheth; Peter Cook; Sam Shepard; Sarah Nobles; Shoshona Le; Suxiang Tong; Vivien Dugan; Yvette Unoarumhi |
| EPI_ISL_1648085, EPI_ISL_1818776, EPI_ISL_1841017, EPI_ISL_1841026, EPI_ISL_1841032 | IN State Department of Health Laboratory Services | IN State Department of Health Laboratory Services | Ankita Kashikar; Brian Pope; Cassandra Campion; Jamie Yeadon; Kyle Brownlee; Lixia Liu; Mark Glazier; Melissa Hindenlang |
| EPI_ISL_2628036, EPI_ISL_2628126, EPI_ISL_2628194, EPI_ISL_2628208 | INHRR | Laboratorio de Virología Molecular | Carmen L Loureiro; CoViVen Group; Domingo J Garzaro; Esmeralda Vizzi; Flor H Pujol; Héctor R Rangel; José Luis Zambrano; Lieska Rodríguez; Mariana Hidalgo; Pierina D´Angelo; Rossana C Jaspe; Víctor Alarcón; Yoneira Sulbaran; Zoila Moros |
| EPI_ISL_1055003, EPI_ISL_1547381 | INSA | Instituto Nacional de Saude (INSA) | Borges et al |
| EPI_ISL_1589853 | INSACOG-WB | National Institute of Biomedical Genomics – INSACOG | Ajay Chakraborti; Arindam Maitra; Bhaswati Bandyopadhyay; Nidhan Kumar Biswas; Saumitra Das; Sreedhar Chinnaswamy; Tamal Ghosh |
| EPI_ISL_1854237 | IPVC | Instituto Nacional de Saude (INSA) | Borges et al |
| EPI_ISL_2612488 | ISDH | Quantigen Biosciences | Paul J. Childress |
| EPI_ISL_2628271, EPI_ISL_2628298, EPI_ISL_2628301, EPI_ISL_2628302, EPI_ISL_2628303, EPI_ISL_2628304 | IVIC | Laboratorio de Virología Molecular | Carmen L Loureiro; CoViVen Group; Domingo J Garzaro; Esmeralda Vizzi; Flor H Pujol; Héctor R Rangel; José Luis Zambrano; Lieska Rodríguez; Mariana Hidalgo; Pierina D´Angelo; Rossana C Jaspe; Víctor Alarcón; Yoneira Sulbaran; Zoila Moros |
| EPI_ISL_2335108, EPI_ISL_2335136 | IVY3 Central Lab, Vanderbilt University Medical Center | Lauring Lab, University of Michigan, Department of Microbiology and Immunology | Gilbert |
| EPI_ISL_1663079, EPI_ISL_1663095, EPI_ISL_1663107, EPI_ISL_1663111, EPI_ISL_1663114, EPI_ISL_1663121, EPI_ISL_1663124, EPI_ISL_1663138, EPI_ISL_1663139, EPI_ISL_1742340, EPI_ISL_1742365, EPI_ISL_1742374, EPI_ISL_1742380, EPI_ISL_1742386, EPI_ISL_1742388, EPI_ISL_1742389, EPI_ISL_1742396, EPI_ISL_1742420, EPI_ISL_1809453, EPI_ISL_1809465, EPI_ISL_1809503, EPI_ISL_1809515, EPI_ISL_1809572, EPI_ISL_1809580, EPI_ISL_1809623, EPI_ISL_1809635, EPI_ISL_1809677, EPI_ISL_1809701, EPI_ISL_1809732, EPI_ISL_1964997, EPI_ISL_1964998, EPI_ISL_1965078, EPI_ISL_2800847 | | | |
| see above | Illinois Department of Public Health | Illinois Department of Public Health - Chicago Lab | Ira Heimler; Joel Price; Vineet K. Dhiman |
| EPI_ISL_1478879, EPI_ISL_1478906, EPI_ISL_1478920, EPI_ISL_1500158, EPI_ISL_1500175, EPI_ISL_1500177, EPI_ISL_1500184, EPI_ISL_1500221, EPI_ISL_1500223, EPI_ISL_1500224, EPI_ISL_1500225, EPI_ISL_1500233, EPI_ISL_1500243, EPI_ISL_1553175, EPI_ISL_1553176, EPI_ISL_1553179, EPI_ISL_1553184, EPI_ISL_1553205, EPI_ISL_1553207, EPI_ISL_1553209, EPI_ISL_1553227, EPI_ISL_1553236, EPI_ISL_1553238, EPI_ISL_1553239, EPI_ISL_1595524, EPI_ISL_1595531, EPI_ISL_1595533, EPI_ISL_1595535, EPI_ISL_1595554, EPI_ISL_1595560, EPI_ISL_1595569, EPI_ISL_1595578, EPI_ISL_1595583, EPI_ISL_1595585, EPI_ISL_1595592, EPI_ISL_1624091, EPI_ISL_1624095, EPI_ISL_1624117, EPI_ISL_1624119, EPI_ISL_1624127, EPI_ISL_1624129, EPI_ISL_1624142, EPI_ISL_1624158, EPI_ISL_1708240, EPI_ISL_1708241, EPI_ISL_1708243, EPI_ISL_1708244, EPI_ISL_1708248, EPI_ISL_1708254, EPI_ISL_1708257, EPI_ISL_1708258, EPI_ISL_1708259, EPI_ISL_1708262, EPI_ISL_1708274, EPI_ISL_1708280, EPI_ISL_1708281, EPI_ISL_1708282, EPI_ISL_1708285, EPI_ISL_1708304, EPI_ISL_1708309, EPI_ISL_1732553, EPI_ISL_1732558, EPI_ISL_1732562, EPI_ISL_1732575, EPI_ISL_1732578, EPI_ISL_1732601, EPI_ISL_1732603, EPI_ISL_1732606, EPI_ISL_1732608, EPI_ISL_1732612, EPI_ISL_1732615, EPI_ISL_1732617, EPI_ISL_1732618, EPI_ISL_1732627, EPI_ISL_1732628, EPI_ISL_1789122, EPI_ISL_1789141, EPI_ISL_1789152, EPI_ISL_1789174, EPI_ISL_1789175, EPI_ISL_1789176, EPI_ISL_1789177, EPI_ISL_1789200, EPI_ISL_1825694, EPI_ISL_1825695, EPI_ISL_1825696, EPI_ISL_1825701, EPI_ISL_1825704, EPI_ISL_1825705, EPI_ISL_1825707, EPI_ISL_1825714, EPI_ISL_1825716, EPI_ISL_1825720, EPI_ISL_1825723, EPI_ISL_1825726, EPI_ISL_1825731, EPI_ISL_1825742, EPI_ISL_1825743, EPI_ISL_1825744, EPI_ISL_1825748, EPI_ISL_1825749, EPI_ISL_1825750, EPI_ISL_1825758, EPI_ISL_1825761, EPI_ISL_1825762, EPI_ISL_1825763, EPI_ISL_1825764, EPI_ISL_1825766, EPI_ISL_1825771, EPI_ISL_1825776, EPI_ISL_1825779, EPI_ISL_1825781, EPI_ISL_1825783, EPI_ISL_1825786, EPI_ISL_1825792, EPI_ISL_1825795, EPI_ISL_1825796, EPI_ISL_1825797, EPI_ISL_1825804, EPI_ISL_1825806, EPI_ISL_1825810, EPI_ISL_1825811, EPI_ISL_1825812, EPI_ISL_1825821, EPI_ISL_1825847, EPI_ISL_1825858, EPI_ISL_1825863, EPI_ISL_1825864, EPI_ISL_1825866, EPI_ISL_1825869, EPI_ISL_1825873, EPI_ISL_1825874, EPI_ISL_1938496, EPI_ISL_1938501, EPI_ISL_1938521, EPI_ISL_1938540, EPI_ISL_2000783, EPI_ISL_2000811, EPI_ISL_2086537, EPI_ISL_2086538, EPI_ISL_2086547, EPI_ISL_2086566, EPI_ISL_2203511, EPI_ISL_2203512, EPI_ISL_2203516, EPI_ISL_2203518, EPI_ISL_2203534 | | | |
| see above | Illinois Department of Public Health - Springfield Lab | Illinois Department of Public Health - Springfield Lab | Bryan Sim; Gordon McCall |
| EPI_ISL_1628603, EPI_ISL_1810847 | InDRE | Instituto Nacional de Medicina Genomica | Alcaraz N; Canseco Mendez JC; Cedro-Tanda A; Garcia-Cardenas FJ; Gisela Barrera-Badillo; Gonzalez-Barrera D; Gonzalez-Woge MA; Herrera-Montalvo LA.; Hidalgo-Miranda A; Irma Lopez-Martinez; Jose Ernesto Ramirez González; Mendoza-Vargas A; Miranda-Ortiz H; Munguia-Garza P; Ramirez-Vega O; Rangel-DeLeon D; Reyes-Grajeda JP; Rosas-Escobar P |
| EPI_ISL_1517432, EPI_ISL_1517433 | Inciensa, Instituto Costarricense de Investigación y Enseñanza en Nutrición y Salud | Inciensa, Instituto Costarricense de Investigación y Enseñanza en Nutrición y Salud | Barboza-Arguedas E & Centeno-Miranda M; Cristian Peréz-Corrales; Cristian Peréz-Corrales & Barboza-Arguedas E |
| EPI_ISL_2011473, EPI_ISL_2011497, EPI_ISL_2011499, EPI_ISL_2011527, EPI_ISL_2136470, EPI_ISL_2136502, EPI_ISL_2136512 | | | |
| see above | Incyte Diagnostics Spokane | Seattle Flu Study | Amanda Adler; Barry R. Lutz; Benjamin Pelle; Brian Hiatt; Caitlin R. Wolf; Chris D. Frazar; Deborah A. Nickerson; Elisabeth Brandstetter; Erica Ryke; Geoff Melly; Helen Y. Chu; Janet A. Englund; Jay Shendure; Jover Lee; Kairsten Fay; Kirsten Lacombe; Lea M. Starita; Mark J. Rieder; Matthew Richardson; Matthew Thompson; Melissa Truong; Michael Boeckh; Michael Famulare; Misja Ilcisin; Peter D. Han; Philip Dykema; Romesh Gautom; Scott Lindquist; Thomas R. Sibley; Trevor Bedford |
| EPI_ISL_1691733, EPI_ISL_1691981, EPI_ISL_1692039, EPI_ISL_1693117, EPI_ISL_1693319, EPI_ISL_1693672, EPI_ISL_1693959, EPI_ISL_1694045, EPI_ISL_1694128, EPI_ISL_1694246, EPI_ISL_1694530, EPI_ISL_1702242, EPI_ISL_1702287, EPI_ISL_1732753, EPI_ISL_1732776, EPI_ISL_1732822, EPI_ISL_1732836, EPI_ISL_1795455, EPI_ISL_1795478, EPI_ISL_1795595, EPI_ISL_1802742, EPI_ISL_1802760, EPI_ISL_1802932, EPI_ISL_1803059, EPI_ISL_1803119, EPI_ISL_1803442, EPI_ISL_1817991, EPI_ISL_2013564, EPI_ISL_2013828, EPI_ISL_2013856, EPI_ISL_2014055, EPI_ISL_2014084, EPI_ISL_2014277, EPI_ISL_2014479, EPI_ISL_2014563, EPI_ISL_2047287, EPI_ISL_2047488, EPI_ISL_2283321, EPI_ISL_3305237 | | | |
| see above | Infinity Biologix | Centers for Disease Control and Prevention Division of Viral Diseases, Pathogen Discovery | Adrian Paskey; Benjamin Rambo-Martin; Chirayu Goswami; Christian Bixby; Christopher Gulvick; Clinton R. Paden; Dakota Howard; Darlene Wagner; Dhwani Batra; Duncan MacCannell; Jason Caravas; Jonathan Schultz; Kara Moser; Matthew Schmerer; Peter W. Cook; Robin Grimwood; Russ Hager; Scott Sammons; Shatavia Morrison; Yihe Wang; Yvette Unoarumhi |
| EPI_ISL_2280284 | Institut Pasteur de Guyane | Institut Pasteur | Angela Brisebarre; Camille Capel; Christophe Malabat; Corinne Maufrais; Dominique Rousset; Emmanuelle Permal; Etienne Simon-Lorière; Frédéric Lemoine; Louise Lefrançois; Marion Barbet; Maud Vanpeene; Méline Bizard; Sylvie Behillil; Sylvie Van der Werf; Vincent Enouf |
| EPI_ISL_1656147, EPI_ISL_1738807 | Institute of Microbiology, Universidad San Francisco de Quito | Institute of Microbiology, Universidad San Francisco de Quito | Belén Prado-Vivar; Bernardo Gutiérrez; Freddy Saldarriaga-Mera; Gabriel Trueba; Juan José Guadalupe; Manuel Jaramillo; Mayra Perero Intriago; Michelle Grunauer; Miguel Sacoto Mazini; Monica Becerra-Wong; Patricio Rojas-Silva; Paúl Cárdenas; Sully Márquez; Verónica Barragán |
| EPI_ISL_2245179, EPI_ISL_2245184, EPI_ISL_2245185 | Instituto Adolfo Lutz | Coordenação Geral de Laboratórios de Saúde Pública (CGLAB/DAEVS/SVS/MS) | Vagner Fonseca; et al. |
| EPI_ISL_1715146, EPI_ISL_1715147, EPI_ISL_1731578, EPI_ISL_1731579, EPI_ISL_1731580, EPI_ISL_1752642, EPI_ISL_1752643, EPI_ISL_1821214, EPI_ISL_1821215, EPI_ISL_1821216, EPI_ISL_1821217, EPI_ISL_1821218, EPI_ISL_1821219, EPI_ISL_1821220, EPI_ISL_1821221, EPI_ISL_1821222, EPI_ISL_1821223, EPI_ISL_1821224 | | | |
| see above | Instituto Adolfo Lutz - Regional de Aracatuba | Instituto Adolfo Lutz, Interdiciplinary Procedures Center, Strategic Laboratory | Caio Vinicius Dias Lopes; Claudia Regina Gonçalves; Claudio Tavares Sacchi; Erica Valessa Ramos Gomes; Karoline Rodrigues Campos; Katia Correa de Oliveira Santos; Leonardo Jose Tadeu de Araujo |
| EPI_ISL_2691109, EPI_ISL_2919236, EPI_ISL_2919239 | Instituto Adolfo Lutz - Regional de Bauru | Instituto Adolfo Lutz, Interdiciplinary Procedures Center, Strategic Laboratory | Caio Vinicius Dias Lopes; Claudia Regina Gonçalves; Claudio Tavares Sacchi; Erica Valessa Ramos Gomes; Karoline Rodrigues Campos; Leonardo Jose Tadeu de Araujo |
| EPI_ISL_906068, EPI_ISL_906069, EPI_ISL_2003162, EPI_ISL_2003163, EPI_ISL_2003167, EPI_ISL_2003168, EPI_ISL_2614520, EPI_ISL_2614521, EPI_ISL_2614525, EPI_ISL_2614529, EPI_ISL_2614530, EPI_ISL_2614531, EPI_ISL_2614533, EPI_ISL_2614534, EPI_ISL_2614535, EPI_ISL_2614536, EPI_ISL_2614537, EPI_ISL_2614538, EPI_ISL_2828688, EPI_ISL_2828689 | | | |
| see above | Instituto Adolfo Lutz - Regional de Campinas | Instituto Adolfo Lutz, Interdiciplinary Procedures Center, | Caio Vinicius Dias Lopes; Claudia Regina Gonçalves; Claudio Tavares Sacchi; Erica Valessa Ramos Gomes; Karoline Rodrigues Campos; Leonardo Jose Tadeu de Araujo |

| | | | |
|---|---|---|---|
| | | Strategic Laboratory | |
| EPI_ISL_1821234, EPI_ISL_1821235, EPI_ISL_1821236, EPI_ISL_1821237, EPI_ISL_1821239, EPI_ISL_1821240, EPI_ISL_1821241, EPI_ISL_1821242, EPI_ISL_2003152, EPI_ISL_2614626, EPI_ISL_2691096, EPI_ISL_2691097, EPI_ISL_2691100, EPI_ISL_2691101, EPI_ISL_2691102, EPI_ISL_2691103, EPI_ISL_2691104, EPI_ISL_2691105, EPI_ISL_2691106 | | | |
| see above | Instituto Adolfo Lutz - Regional de Marilia | Instituto Adolfo Lutz, Interdiciplinary Procedures Center, Strategic Laboratory | Caio Vinicius Dias Lopes; Claudia Regina Gonçalves; Claudio Tavares Sacchi; Erica Valessa Ramos Gomes; Karoline Rodrigues Campos; Leonardo Jose Tadeu de Araujo |
| EPI_ISL_2614614, EPI_ISL_2614615, EPI_ISL_2614616 | Instituto Adolfo Lutz - Regional de Presidente Prudente | Instituto Adolfo Lutz, Interdiciplinary Procedures Center, Strategic Laboratory | Caio Vinicius Dias Lopes; Claudia Regina Gonçalves; Claudio Tavares Sacchi; Erica Valessa Ramos Gomes; Karoline Rodrigues Campos; Leonardo Jose Tadeu de Araujo |
| EPI_ISL_1715148, EPI_ISL_1731581, EPI_ISL_1731582, EPI_ISL_1752645, EPI_ISL_1821246, EPI_ISL_1821247, EPI_ISL_1821248, EPI_ISL_1821249, EPI_ISL_1821250, EPI_ISL_1821251, EPI_ISL_1821252, EPI_ISL_1821253, EPI_ISL_1821254, EPI_ISL_1821255, EPI_ISL_1821256, EPI_ISL_1821258, EPI_ISL_1821259, EPI_ISL_1821260, EPI_ISL_1821261, EPI_ISL_1821262, EPI_ISL_1821263, EPI_ISL_1821264, EPI_ISL_1821265 | | | |
| see above | Instituto Adolfo Lutz - Regional de Ribeirao Preto | Instituto Adolfo Lutz, Interdiciplinary Procedures Center, Strategic Laboratory | Caio Vinicius Dias Lopes; Claudia Regina Gonçalves; Claudio Tavares Sacchi; Erica Valessa Ramos Gomes; Karoline Rodrigues Campos; Katia Correa de Oliveira Santos; Leonardo Jose Tadeu de Araujo |
| EPI_ISL_1625978, EPI_ISL_1625986, EPI_ISL_1625987, EPI_ISL_1625988, EPI_ISL_1625989, EPI_ISL_1625992, EPI_ISL_1625993, EPI_ISL_1625994, EPI_ISL_1625995, EPI_ISL_1625997, EPI_ISL_1625998, EPI_ISL_1625999, EPI_ISL_1626001, EPI_ISL_1626002, EPI_ISL_1626003, EPI_ISL_1626004, EPI_ISL_1626006, EPI_ISL_1626606 | | | |
| see above | Instituto Adolfo Lutz - Regional de Rio Claro | Instituto Adolfo Lutz, Interdiciplinary Procedures Center, Strategic Laboratory | Caio Vinicius Dias Lopes; Claudia Regina Gonçalves; Claudio Tavares Sacchi; Erica Valessa Ramos Gomes; Karoline Rodrigues Campos; Katia Correa de Oliveira Santos; Leonardo Jose Tadeu de Araujo |
| EPI_ISL_2003135, EPI_ISL_2003137, EPI_ISL_2003138, EPI_ISL_2003139, EPI_ISL_2003140, EPI_ISL_2003142, EPI_ISL_2003143, EPI_ISL_2003145, EPI_ISL_2003146, EPI_ISL_2003147, EPI_ISL_2003148 | | | |
| see above | Instituto Adolfo Lutz - Regional de Santos | Instituto Adolfo Lutz, Interdiciplinary Procedures Center, Strategic Laboratory | Caio Vinicius Dias Lopes; Claudia Regina Gonçalves; Claudio Tavares Sacchi; Erica Valessa Ramos Gomes; Karoline Rodrigues Campos; Leonardo Jose Tadeu de Araujo |
| EPI_ISL_2958842 | Instituto Adolfo Lutz - Regional de Sorocaba | Instituto Adolfo Lutz, Interdiciplinary Procedures Center, Strategic Laboratory | Caio Vinicius Dias Lopes; Claudia Regina Gonçalves; Claudio Tavares Sacchi; Erica Valessa Ramos Gomes; Karoline Rodrigues Campos |
| EPI_ISL_1628347, EPI_ISL_1628350, EPI_ISL_1628351, EPI_ISL_1628354, EPI_ISL_1628355, EPI_ISL_1628357, EPI_ISL_1628358, EPI_ISL_1628360, EPI_ISL_1628361, EPI_ISL_1628362, EPI_ISL_1715150, EPI_ISL_1715151, EPI_ISL_1715152, EPI_ISL_1715153, EPI_ISL_1715154, EPI_ISL_1715155, EPI_ISL_1715156, EPI_ISL_1715157, EPI_ISL_1715158, EPI_ISL_1731583, EPI_ISL_1731584, EPI_ISL_1731586, EPI_ISL_1731588, EPI_ISL_1731589, EPI_ISL_1731590, EPI_ISL_1731592, EPI_ISL_1731597, EPI_ISL_1731599, EPI_ISL_1731600, EPI_ISL_1731601, EPI_ISL_1731603, EPI_ISL_1731604, EPI_ISL_1731605, EPI_ISL_1731607, EPI_ISL_1731608, EPI_ISL_1731609, EPI_ISL_1731610, EPI_ISL_1731611, EPI_ISL_1731612, EPI_ISL_1752647, EPI_ISL_1752651, EPI_ISL_1752652, EPI_ISL_1752654, EPI_ISL_1752655, EPI_ISL_1752658, EPI_ISL_1752660, EPI_ISL_1752662, EPI_ISL_1752665, EPI_ISL_1752666, EPI_ISL_1752667, EPI_ISL_1752668, EPI_ISL_1821267, EPI_ISL_1821268, EPI_ISL_1821269, EPI_ISL_1821271, EPI_ISL_1821272, EPI_ISL_2003114, EPI_ISL_2003115, EPI_ISL_2003116, EPI_ISL_2003117, EPI_ISL_2003118, EPI_ISL_2003119, EPI_ISL_2003121, EPI_ISL_2003125, EPI_ISL_2003126, EPI_ISL_2003127, EPI_ISL_2003128, EPI_ISL_2003129, EPI_ISL_2003130, EPI_ISL_2003132, EPI_ISL_2756432, EPI_ISL_2756433, EPI_ISL_2756434, EPI_ISL_2756435, EPI_ISL_2756436, EPI_ISL_2756440, EPI_ISL_2756441, EPI_ISL_2756443, EPI_ISL_2756445, EPI_ISL_2756446, EPI_ISL_2756447, EPI_ISL_2756449, EPI_ISL_2756454, EPI_ISL_2756455, EPI_ISL_2756456, EPI_ISL_2756457, EPI_ISL_2756459, EPI_ISL_2756460, EPI_ISL_2756461, EPI_ISL_2756462, EPI_ISL_2756463, EPI_ISL_2756464, EPI_ISL_2756465, EPI_ISL_2756466, EPI_ISL_2756467, EPI_ISL_2756468, EPI_ISL_2756469, EPI_ISL_2756470, EPI_ISL_2756471, EPI_ISL_2756472, EPI_ISL_2756473, EPI_ISL_2756474, EPI_ISL_2756475, EPI_ISL_2756476, EPI_ISL_2756479, EPI_ISL_2756480, EPI_ISL_2756482, EPI_ISL_2756485, EPI_ISL_2756486, EPI_ISL_2756488, EPI_ISL_2756489, EPI_ISL_2756490, EPI_ISL_2756491, EPI_ISL_2756492, EPI_ISL_2756493 | | | |
| see above | Instituto Adolfo Lutz Central | Instituto Adolfo Lutz, Interdiciplinary Procedures Center, Strategic Laboratory | Caio Vinicius Dias Lopes; Claudia Regina Gonçalves; Claudio Tavares Sacchi; Erica Valessa Ramos Gomes; Karoline Rodrigues Campos; Katia Correa de Oliveira Santos; Leonardo Jose Tadeu de Araujo |
| EPI_ISL_2614602, EPI_ISL_2614603, EPI_ISL_2614604, EPI_ISL_2614605, EPI_ISL_2614606, EPI_ISL_2614608, EPI_ISL_2614609, EPI_ISL_2614610, EPI_ISL_2841607, EPI_ISL_2841609, EPI_ISL_2841611, EPI_ISL_2841612, EPI_ISL_2958833 | | | |
| see above | Instituto Biologico | Instituto Adolfo Lutz, Interdiciplinary Procedures Center, Strategic Laboratory | Caio Vinicius Dias Lopes; Claudia Regina Gonçalves; Claudio Tavares Sacchi; Erica Valessa Ramos Gomes; Karoline Rodrigues Campos; Leonardo Jose Tadeu de Araujo |
| EPI_ISL_4104674, EPI_ISL_4104678, EPI_ISL_4104680, EPI_ISL_4104682, EPI_ISL_4104686, EPI_ISL_4104687, EPI_ISL_4104689, EPI_ISL_4104690, EPI_ISL_4104694, EPI_ISL_4104698, EPI_ISL_4104699, EPI_ISL_4104701, EPI_ISL_4104703, EPI_ISL_4104708, EPI_ISL_4104717, EPI_ISL_4104718, EPI_ISL_4104722, EPI_ISL_4104724, EPI_ISL_4104728, EPI_ISL_4104729 | | | |
| see above | Instituto Butantan | Instituto Butantan | Antonio Jorge Martins; Claudia Renata dos Santos Barros; David Schlesinger; Debora Botequio Moretti; Dimas Tadeu Covas; Elaine Cristina Marqueze; Elaine Vieira Santos; Evandra Strazza Rodrigues; Heidge Fukumasu; Jayme Augusto de Souza-Neto; José Salvatore Leister Patané; Luiz Alcantara; Luiz Lehmann Coutinho; Maria Carolina Elias; Maurício Lacerda Nogueira; Rafael dos Santos Bezerra; Raul Machado Neto; Rejane Maria Tommasini Grotto; Ricardo Haddad; Sandra Coccuzzo Sampaio Vessoni; Simone Kashima; Svetoslav Nanev Slavov; Vincent Louis Viala |
| EPI_ISL_2344420 | Instituto Butantan | Instituto de Medicina Tropical de Sao Paulo | Brazil-UK Centre for Arbovirus Discovery Diagnosis Genomics and Epidemiology (CADDE) Genomic Network - Instituto de Medicina Tropical |
| EPI_ISL_3418671, EPI_ISL_3418673, EPI_ISL_3418676, EPI_ISL_3418678, EPI_ISL_3418679, EPI_ISL_3418680, EPI_ISL_3418681, EPI_ISL_3418682, EPI_ISL_3418683 | | | |
| see above | Instituto Carlos Chagas - ICC, FIOCRUZ Parana | Instituto Carlos Chagas - ICC, FIOCRUZ Parana | A.A.; A.M.; A.R.; Aguiar; Albrecht, L.; Alves; Avila; Balsanelli, E.; Becker, G.; Blanes, L.; Dallagiovanna, B.; Debur; E.M.; F.K.; F.O.; Faoro, H.; Graef, T.; H.G.; I.N.; L.G.; L.R.; M.M.; M.O.; Marchini; Md.C.; Morello; Nardeli; Oliveira; P.C.; Passetti, F.; Pedrosa; Resende; Riediger; S.C.; Schemberger; Suzukawa; V.A.; Zanette, D.; de Baura; de Souza; dos Santos |
| EPI_ISL_2493026 | Instituto Nacional de Investigação em Saúde | CERI, Centre for Epidemic Response and Innvoation, Stellenbosch University and KRISP, KZN Research Innovation and Sequencing Platform, UKZN. | Afonso P; David K; Emmanuel SJ; Freitas RH; Giandhari J; Inglês L; Lutucuta S; Miranda J; Morais J; Mufinda M; Naidoo Y; Neto Z; Paulo A Carralero RR Paixão JP; Pereira A; Pillay S; Tegally H; Wilkinson E; de Oliveira T |
| EPI_ISL_1824485, EPI_ISL_2160668 | Instituto Nacional de Medicina Genomica | Centro de Investigación en Enfermedades Infecciosas (CIENI), Instituto Nacional de Enfermedades Respiratorias (INER) | Arriaga-Canon C; Avila-Rios S; Boukadida C; Cedro-Tanda A; Herrera-Montalvo LA; Hidalgo-Miranda A; Matias-Florentino M; Mendoza-Vargas A; Perez-Garcia M; Reyes-Grajeda JP; Reyes-Teran G |
| EPI_ISL_1585916 | Instituto Nacional de Medicina Genomica | Instituto Nacional de Medicina Genomica | Alcaraz N; Canseco Mendez JC; Cedro-Tanda A; Garcia-Cardenas FJ; Gonzalez-Barrera D; Gonzalez-Woge MA; Herrera-Montalvo LA; Hidalgo-Miranda A; Mendoza-Vargas A; Miranda-Ortiz H; Munguia-Garza P; Ramirez-Vega O; Rangel-DeLeon D; Reyes-Grajeda JP; Rosas-Escobar P |
| EPI_ISL_956287, EPI_ISL_956289, EPI_ISL_956297 | Instituto Nacional de Salud- Dirección de Redes de Laboratorios de Salud Pública | Instituto Nacional de Salud- Dirección de Investigación en Salud Pública | Carlos Franco-Muñoz; Diego A. Álvarez-Díaz; Diego Andrés Prada; Gerardo Santamaría; Hector Alejandro Ruiz-Moreno; Jhonnatan Reales-González; Julian Naizaque; Katherine Laiton-Donato; Magdalena Wiesner; Marcela Mercado-Reyes; Maria T. Herrera-Sepúlveda; Martha Lucia Ospina Martinez; Mauricio Pacheco-Montealegre; Sheryll Corchuelo |
| EPI_ISL_1854230, EPI_ISL_1854231 | Instituto Nacional de Saude (INSA) and Instituto Gulbenkian de Ciencia (IGC) | Instituto Nacional de Saude (INSA) and Instituto Gulbenkian de Ciencia (IGC) | Borges et al |
| EPI_ISL_2004134, EPI_ISL_2004192 | Instituto Nacional de Saude (INSA) and i3S - Instituto de Investigação e Inovação em Saúde | Instituto Nacional de Saude (INSA) and i3S - Instituto de Investigação e Inovação em Saúde | Borges et al |
| EPI_ISL_2758988, EPI_ISL_2758992, EPI_ISL_2758994, EPI_ISL_2758995, EPI_ISL_2758997, EPI_ISL_2759001, EPI_ISL_2759002, EPI_ISL_2759004, EPI_ISL_2759006, EPI_ISL_2759011, EPI_ISL_2759012, EPI_ISL_2759016, EPI_ISL_2759017, EPI_ISL_2759020, EPI_ISL_2759021, EPI_ISL_2759022, EPI_ISL_2759024, EPI_ISL_2759025, EPI_ISL_2759026, EPI_ISL_2759028, EPI_ISL_2759033, EPI_ISL_2759035, EPI_ISL_2759037, EPI_ISL_2759041, EPI_ISL_2759045, EPI_ISL_2759046, EPI_ISL_2759047, EPI_ISL_2759050, EPI_ISL_2759051, EPI_ISL_2759052, EPI_ISL_2759054, EPI_ISL_2759056, EPI_ISL_2759057, EPI_ISL_2759059, EPI_ISL_2759061, EPI_ISL_2759063, EPI_ISL_2759065, EPI_ISL_2759066, EPI_ISL_2759068 | | | |
| see above | Instituto de Biologia Molecular do Paraná (LAC) | Instituto Carlos Chagas - Fiocruz | Alessandra De Melo Aguiar; Andreia Akemi Suzukawa; Andréa Rodrigues Ávila; Bruno Dallagiovanna; Dalila Zanette; Eduardo Balsanelli; Emanuel Maltempi de Souza; Fabio Passetti; Fabricio Klerynton Marchini; Fábio de Oliveira Pedrosa; Guilherme Becker; Helisson Faoro; Hellen Geremias dos Santos; Irina Nastassja Riediger; Letusa Albrecht; Lucas Blanes; Luis Gustavo Morello; Lysangela Ronalte Alves; Maria do Carmo Debur; Mauro de Medeiros Oliveira; Michelle Orane Schemberger; Paola Cristina Resende; Sheila Cristina Nardeli; Tiago Gräf; Valter Antônio de Baura |
| EPI_ISL_1000671, EPI_ISL_1000673, EPI_ISL_1734843, EPI_ISL_1734844, EPI_ISL_1734852, EPI_ISL_1734858, EPI_ISL_1734866, EPI_ISL_1734872, EPI_ISL_1734874, EPI_ISL_1734883, EPI_ISL_1911958, EPI_ISL_1911960, EPI_ISL_1911961, EPI_ISL_1911962, EPI_ISL_1911964, EPI_ISL_2223443, EPI_ISL_2223446, EPI_ISL_2223452, EPI_ISL_2295388, EPI_ISL_2295389, EPI_ISL_2295396, EPI_ISL_2295399, EPI_ISL_2295403, EPI_ISL_2295404, EPI_ISL_2295405, EPI_ISL_2295406, EPI_ISL_2295408, EPI_ISL_2295409, EPI_ISL_2295410, EPI_ISL_2295413, EPI_ISL_2295414, EPI_ISL_2295416, EPI_ISL_2308609, EPI_ISL_3118795, EPI_ISL_3118796, EPI_ISL_3118797, EPI_ISL_3118804, EPI_ISL_3118809, EPI_ISL_3118811, EPI_ISL_3118813 | | | |
| see above | Instituto de Biotecnologia - UNESP-Botucatu-SP | Instituto de Biotecnologia - UNESP-Botucatu-SP | Camila Dantas Malossi; Cecília Artico Banho; Cíntia Bittar; Fábio Sossai Possebon; Guilherme Campos; Helena Lage Ferreira; Jorge A. Petroli Marchesi; João Pessoa Araújo Jr.; Leila Sabrina Ullmann; Lívia Sacchetto; Maisa C. Pereira Parra; Marília Moraes; Maurício L. Nogueira; Paula Rahal; Paulo Inacio da Costa |
| EPI_ISL_2776166, EPI_ISL_2776171, EPI_ISL_2776172, EPI_ISL_2776174, | Instituto de Medicina Tropical & Salud Global (IMTSAG) | Grubaugh Lab - Yale School of Public Health | Alejandaro Vallejo Degaudenzi; Anderson Brito; Annie Watkins; Chaney Kalinich; Chantal Vogels; Elisa Contreras; Esperanza Mendoza; Isabel Ott; Jessica Rothman; Joseph Fauver; Kendall Billig; Mallery Breban; Mary Petrone; Nathan Grubaugh; Robert Paulino-Ramirez; Tara Alpert; Tobias Koch; Victor Virgilio Calderon |

| | | | |
|---|---|---|---|
| EPI_ISL_2776178, EPI_ISL_2776179 | | | |
| EPI_ISL_2037442 | Instituto de Virologia "Dr. J. M. Vanella", Facultad de Ciencias Médicas, Universidad Nacional de Córdoba. | Centro de Investigaciones Agropecuarias (CIAP), Instituto Nacional de Tecnologia Agropecuaria (INTA) Cordoba, Argentina, on behalf of 'Proyecto Argentino Interinstitucional de Genomica de SARS-CoV-2 (PAIS Consortium) | Adrian Diaz; Brenda Konigheim; Franco Fernandez; Gabriela Barbas; Gonzalo Castro; Humberto Debat; Javier Aguilar; Lorena Spinsanti; Maria Belen Pisano; Maria Elisa Rivarola; Mauricio Beranek; Nathalie Marquez; Sandra Gallego.; Sebastian Blanco; Viviana Re |
| EPI_ISL_2691149, EPI_ISL_2691150 | Intituto Adolfo Lutz - Regional de Sorocaba | Instituto Adolfo Lutz, Interdiciplinary Procedures Center, Strategic Laboratory | Caio Vinicius Dias Lopes; Claudia Regina Gonçalves; Claudio Tavares Sacchi; Erica Valessa Ramos Gomes; Karoline Rodrigues Campos; Leonardo Jose Tadeu de Araujo |
| EPI_ISL_1061413 | Istanbul University-Cerrahpasa, Cerrahpasa School of Medicine, COVID-19 Laboratory | Istanbul University-Cerrahpasa, Cerrahpasa School of Medicine, COVID-19 Laboratory | Ebru Yucebag; Fusun Can; Haluk Eraksoy; Kenan Midilli; Mert Ahmet Kuskucu; Okan Kadir Nohut; Serap Şimşek Yavuz; Yesim Tuyji Tok; Zarifa Abullayeva |
| EPI_ISL_1854242, EPI_ISL_1854243 | Joaquim Chaves Saude | Instituto Nacional de Saude (INSA) | Borges et al |
| EPI_ISL_1739011, EPI_ISL_1739012, EPI_ISL_1739013, EPI_ISL_1739041 | Joaquim Chaves Saude | Instituto Nacional de Saude (INSA) and Institute of Biomedicine (iBiMed), Universidade de Aveiro | Borges et al |
| EPI_ISL_2246720, EPI_ISL_2246721, EPI_ISL_2246725, EPI_ISL_2246726, EPI_ISL_2246728, EPI_ISL_2246731 | John H Stroger Jr Hospital of Cook County | RIPHL at Rush University Medical Center | Ah-Young Sung; Felix Araujo Perez; Kevin Kunstman; Laura Furtado; Marieta Hyde; Max Kolton; Sharon Welbel; Stefan Green |
| EPI_ISL_1973456, EPI_ISL_1973458 | Johns Hopkins Hospital Department of Pathology | Johns Hopkins Hospital Department of Pathology | Adannaya Amadi; C. Paul Morris; Chun Huai Luo; Heba H. Mostafa; Matthew Schwartz; Nicholas Gallagher |
| EPI_ISL_1312267, EPI_ISL_2404238, EPI_ISL_2404654, EPI_ISL_2425143, EPI_ISL_2425168, EPI_ISL_2425189, EPI_ISL_2425221, EPI_ISL_2832285, EPI_ISL_2832900 | | | |
| see above | KU Leuven, Rega Institute, Clinical and Epidemiological Virology | KU Leuven, Rega Institute, Clinical and Epidemiological Virology | Bert Vanmechelen; Joan Marti-Carerras; Piet Maes; Tony Wawina-Bokalanga |
| EPI_ISL_1823805, EPI_ISL_1823809 | LA Office of Public Health Laboratories | Centers for Disease Control and Prevention Division of Viral Diseases, Pathogen Discovery | Alison Laufer Halpin; Ben L. Rambo-Martin; Clinton R. Paden; Dakota Howard; Darlene Wagner; Dave Wentworth; Dhwani Batra; Jasmine Padilla; Justin Lee; Katie Dillon; Krista Queen; Kristen Knipe; Kristine Lacek; Mark Burroughs; Matthew Schmerer; Mili Sheth; Peter Cook; Sam Shepard; Sarah Nobles; Shoshona Le; Suxiang Tong; Vivien Dugan; Yvette Unoarumhi |
| EPI_ISL_1788362, EPI_ISL_2142664 | LABORATOIRE DE BIOLOGIE MEDICALE | CNR Virus des Infections Respiratoires - France SUD | Antonin Bal; Bruno Lina; Gregory Destras; Gwendolyne Burfin; Hadrien Regue; Laurence Josset; Martine Valette; Quentin Semanas |
| EPI_ISL_3102494, EPI_ISL_3102526 | LABORATORIO CLEMENTINO FRAGA | Analytical Competence Molecular Epidemiology Lab/ACME, Oswaldo Cruz Foundation, Ceara (FIOCRUZ CE) | Cleber Furtado Aksenen; Fabio Miyajima; Fernando Braga Stehling; Francisco Eder de Moura Lopes; Jamille Maria Mendes Bezerra; Joaquim César do Nascimento Sousa Junior; Pedro Miguel Carneiro Jeronimo; Suzana Porto Almeida e Lucas Delerino; Thais Ferreira de Oliveira; Thais de Oliveira Costa; Ticiane Cavalcante de Souza; Veridiana Pessoa Miyajima |
| EPI_ISL_1795303, EPI_ISL_1795306, EPI_ISL_1795308, EPI_ISL_1795309, EPI_ISL_1795310, EPI_ISL_1795311, EPI_ISL_1795313, EPI_ISL_1795316, EPI_ISL_1795317, EPI_ISL_1795319, EPI_ISL_1795320, EPI_ISL_1795322, EPI_ISL_1795323, EPI_ISL_2344795, EPI_ISL_2344796, EPI_ISL_2344828, EPI_ISL_2344836, EPI_ISL_2344840, EPI_ISL_2344841, EPI_ISL_2344842, EPI_ISL_2344843, EPI_ISL_2345549, EPI_ISL_2345554, EPI_ISL_2345557, EPI_ISL_2345558, EPI_ISL_2345559, EPI_ISL_2345560, EPI_ISL_2345563, EPI_ISL_2345568, EPI_ISL_2345569, EPI_ISL_2345570, EPI_ISL_2345576, EPI_ISL_2345578, EPI_ISL_2345582, EPI_ISL_2345584 | | | |
| see above | LABORATORIO DE FRANCA | Instituto Butantan / ESALQ-Piracicaba | Antonio Jorge Martins; Bianca Cechetto Carlos. Mendelics: Bibiana Santos; Claudia Renata dos Santos Barros; David Schlesinger; David Schlesinger. Hemocentro Ribeirão Preto: Simone Kashima; Debora Botequio Moretti; Debora Botequio Moretti. Centro de Genômica Funcional da ESALQ: Luiz Lehmann Coutinho; Dimas Tadeu Covas; Elaine Cristina Marqueze; Elaine Vieira Santos; Elaine Vieira dos Santos; Elisangela Chicaroni Mattos; Erika Freitas; Evandra Strazza Rodrigues; Felipe Allan da Silva da Costa; Flavia Aburjaile; Guilherme Targino Valente; Heidge Fukumasu; Heidge Fukumasu. USP-Botucatu: Rejane Maria Tommasini Grotto; Instituto Butantan: Alexander Roberto Precioso; Jayme A. Souza-Neto; Jayme Augusto de Souza-Neto; Jessika Cristina Chagas Lesbon; José Salvatore Leister Patané; João Paulo Kitajima; Luiz Alcantara; Luiz Carlos Junior de Alcantara; Luiz Lehmann Coutinho; Maria Carolina Elias; Marta Giovanetti; Maurício Lacerda Nogueira; Patricia Akemi Assato; Rafael dos Santos Bezerra; Raquel de Lello Rocha Campos Cassano. NGS Soluções Genômicas: Pilar Drummond Sampaio Corrêa Mariani. FZEA-USP Pirassununga: Mirele Daiana Poleti; Raul Machado Neto; Rejane Maria Tommasini Grotto; Ricardo Augusto Brassaloti; Ricardo Haddad; Rodrigo Tocantins Calado.; Sandra Coccuzzo Sampaio; Sandra Coccuzzo Sampaio Vessoni; Simone Kashima; Svetoslav Nanev Slavov; Vagner Fonseca; Vincent Louis Viala |
| EPI_ISL_2344583 | LABORATORIO DE FRANCA | Instituto Butantan / FZEA-USP-Pirassununga | Antonio Jorge Martins; Claudia Renata dos Santos Barros; David Schlesinger; Debora Botequio Moretti; Dimas Tadeu Covas; Elaine Cristina Marqueze; Elaine Vieira Santos; Evandra Strazza Rodrigues; Heidge Fukumasu; Jayme Augusto de Souza-Neto; José Salvatore Leister Patané; Luiz Alcantara; Luiz Lehmann Coutinho; Maria Carolina Elias; Maurício Lacerda Nogueira; Rafael dos Santos Bezerra; Raul Machado Neto; Rejane Maria Tommasini Grotto; Ricardo Haddad; Sandra Coccuzzo Sampaio Vessoni; Simone Kashima; Svetoslav Nanev Slavov; Vincent Louis Viala |
| EPI_ISL_1795346, EPI_ISL_1795347, EPI_ISL_1795348, EPI_ISL_1795349, EPI_ISL_1795350, EPI_ISL_1795351, EPI_ISL_1795352, EPI_ISL_1795353, EPI_ISL_1795354, EPI_ISL_1795355, EPI_ISL_1795356, EPI_ISL_1795358, EPI_ISL_1795359, EPI_ISL_1795365, EPI_ISL_1795366, EPI_ISL_2345613, EPI_ISL_2345615, EPI_ISL_2345616, EPI_ISL_2345617, EPI_ISL_2345618, EPI_ISL_2345619, EPI_ISL_2345621, EPI_ISL_2345622, EPI_ISL_2345627, EPI_ISL_2345628, EPI_ISL_2345629, EPI_ISL_2345633, EPI_ISL_2345634, EPI_ISL_2345640, EPI_ISL_2345641 | | | |
| see above | LABORATORIO DR PAULO EMILIO DALESSANDRO PINDAMONHANGABA | Instituto Butantan / ESALQ-Piracicaba | Antonio Jorge Martins; Bianca Cechetto Carlos. Mendelics: Bibiana Santos; Claudia Renata dos Santos Barros; David Schlesinger; David Schlesinger. Hemocentro Ribeirão Preto: Simone Kashima; Debora Botequio Moretti; Debora Botequio Moretti. Centro de Genômica Funcional da ESALQ: Luiz Lehmann Coutinho; Dimas Tadeu Covas; Elaine Cristina Marqueze; Elaine Vieira Santos; Elaine Vieira dos Santos; Elisangela Chicaroni Mattos; Erika Freitas; Evandra Strazza Rodrigues; Felipe Allan da Silva da Costa; Flavia Aburjaile; Guilherme Targino Valente; Heidge Fukumasu; Heidge Fukumasu. USP-Botucatu: Rejane Maria Tommasini Grotto; Instituto Butantan: Alexander Roberto Precioso; Jayme A. Souza-Neto; Jayme Augusto de Souza-Neto; Jessika Cristina Chagas Lesbon; José Salvatore Leister Patané; João Paulo Kitajima; Luiz Alcantara; Luiz Carlos Junior de Alcantara; Luiz Lehmann Coutinho; Maria Carolina Elias; Marta Giovanetti; Maurício Lacerda Nogueira; Patricia Akemi Assato; Rafael dos Santos Bezerra; Raquel de Lello Rocha Campos Cassano. NGS Soluções Genômicas: Pilar Drummond Sampaio Corrêa Mariani. FZEA-USP Pirassununga: Mirele Daiana Poleti; Raul Machado Neto; Rejane Maria Tommasini Grotto; Ricardo Augusto Brassaloti; Ricardo Haddad; Rodrigo Tocantins Calado.; Sandra Coccuzzo Sampaio; Sandra Coccuzzo Sampaio Vessoni; Simone Kashima; Svetoslav Nanev Slavov; Vagner Fonseca; Vincent Louis Viala |
| EPI_ISL_2345653, EPI_ISL_2345658, EPI_ISL_2345698, EPI_ISL_2362241, EPI_ISL_2362242, EPI_ISL_2362253, EPI_ISL_2362263, EPI_ISL_2362274 | | | |
| see above | LABORATORIO DR PAULO EMILIO DALESSANDRO PINDAMONHANGABA | Instituto Butantan / Mendelics | Antonio Jorge Martins; Claudia Renata dos Santos Barros; David Schlesinger; Debora Botequio Moretti; Dimas Tadeu Covas; Elaine Cristina Marqueze; Elaine Vieira Santos; Evandra Strazza Rodrigues; Heidge Fukumasu; Jayme Augusto de Souza-Neto; José Salvatore Leister Patané; Luiz Alcantara; Luiz Lehmann Coutinho; Maria Carolina Elias; Maurício Lacerda Nogueira; Rafael dos Santos Bezerra; Raul Machado Neto; Rejane Maria Tommasini Grotto; Ricardo Haddad; Sandra Coccuzzo Sampaio Vessoni; Simone Kashima; Svetoslav Nanev Slavov; Vincent Louis Viala |
| EPI_ISL_2209338, EPI_ISL_2209345 | LABORATORIO LOCAL DE ITAPECERICA DA SERRA | Instituto Butantan | Antonio Jorge Martins; Claudia Renata dos Santos Barros; David Schlesinger; Debora Botequio Moretti; Dimas Tadeu Covas; Elaine Cristina Marqueze; Elaine Vieira Santos; Evandra Strazza Rodrigues; Heidge Fukumasu; Jayme Augusto de Souza-Neto; José Salvatore Leister Patané; Luiz Alcantara; Luiz Lehmann Coutinho; Maria Carolina Elias; Maurício Lacerda Nogueira; Rafael dos Santos Bezerra; Raul Machado Neto; Rejane Maria Tommasini Grotto; Ricardo Haddad; Sandra Coccuzzo Sampaio Vessoni; Simone Kashima; Svetoslav Nanev Slavov; Vincent Louis Viala. |
| EPI_ISL_2345891, EPI_ISL_2345895, EPI_ISL_2345896, EPI_ISL_2345898 | LABORATORIO LOCAL DE ITAPECERICA DA SERRA | Instituto Butantan / FZEA-USP-Pirassununga | Antonio Jorge Martins; Claudia Renata dos Santos Barros; David Schlesinger; Debora Botequio Moretti; Dimas Tadeu Covas; Elaine Cristina Marqueze; Elaine Vieira Santos; Evandra Strazza Rodrigues; Heidge Fukumasu; Jayme Augusto de Souza-Neto; José Salvatore Leister Patané; Luiz Alcantara; Luiz Lehmann Coutinho; Maria Carolina Elias; Maurício Lacerda Nogueira; Rafael dos Santos Bezerra; Raul Machado Neto; Rejane Maria Tommasini Grotto; Ricardo Haddad; Sandra Coccuzzo Sampaio Vessoni; Simone Kashima; Svetoslav Nanev Slavov; Vincent Louis Viala |
| EPI_ISL_3102475, EPI_ISL_3102477, EPI_ISL_3102543 | LABORATORIO MUN ANALISES CLINICAS | Analytical Competence Molecular Epidemiology Lab/ACME, Oswaldo Cruz Foundation, Ceara (FIOCRUZ CE) | Cleber Furtado Aksenen; Fabio Miyajima; Fernando Braga Stehling; Francisco Eder de Moura Lopes; Jamille Maria Mendes Bezerra; Joaquim César do Nascimento Sousa Junior; Pedro Miguel Carneiro Jeronimo; Suzana Porto Almeida e Lucas Delerino; Thais Ferreira de Oliveira; Thais de Oliveira Costa; Ticiane Cavalcante de Souza; Veridiana Pessoa Miyajima |
| EPI_ISL_1966611 | LABORATORIO MUNICIPAL DE ANALISES CLINICAS DE ITANHAEM | Instituto Butantan / Mendelics | Antonio Jorge Martins; Bianca Cechetto Carlos. Mendelics: Bibiana Santos; Claudia Renata dos Santos Barros; Cíntia Bittar; David Schlesinger. Hemocentro Ribeirão Preto: Simone Kashima; Debora Botequio Moretti; Elaine Cristina Marqueze; Elaine Vieira dos Santos; Elisangela Chicaroni Mattos; Erika Freitas; Evandra Strazza Rodrigues; Felipe Allan da Silva da Costa; Flavia Aburjaile; Fábio Sossai Possebon; Guilherme Campos; Guilherme Targino Valente; Heidge Fukumasu. USP-Botucatu: Rejane Maria Tommasini Grotto; Helena Lage Ferreira; Instituto Butantan: Dimas Tadeu Covas; Jardelina de Souza Todao Bernardino; Jayme A. Souza-Neto; Jessika Cristina Chagas Lesbon; Jorge A. Petroli Marchesi; José Salvatore Leister Patané; João Paulo Kitajima; João Pessoa Araújo Jr.; Leila Sabrina Ullmann; Loyze Paola Oliveira de Lima; Luiz Aurelio de Campos Crispin. Centro de Genômica Funcional da ESALQ: Luiz Lehmann Coutinho; Luiz Carlos Junior de Alcantara; Lívia Sacchetto; Maisa C. Pereira Parra; Maria Carolina Elias; Marta Giovanetti; Marília Moraes; Maurício Lacerda Nogueira. Prefeitura de Sao Paulo: Melissa Palmieri.; Patricia Akemi Assato; Paula Rahal; Paulo Inacio da Costa; Rafael dos Santos Bezerra; Raquel de Lello Rocha Campos Cassano. NGS Soluções Genômicas: Pilar Drummond Sampaio Corrêa Mariani. FZEA-USP Pirassununga: Mirele Daiana Poleti; Raul Machado Neto; Ricardo Augusto Brassaloti; Ricardo Haddad; Rodrigo Tocantins Calado. FAMERP-SJRP: Cecília Artico Banho; Sandra Coccuzzo Sampaio; Svetoslav Nanev Slavov; Vagner Fonseca; Vincent Louis Viala |
| EPI_ISL_2209360 | LABORATORIO MUNICIPAL DE | Instituto Butantan | Antonio Jorge Martins; Claudia Renata dos Santos Barros; David Schlesinger; Debora Botequio Moretti; Dimas Tadeu Covas; Elaine Cristina Marqueze; Elaine Vieira Santos; Evandra Strazza Rodrigues; Heidge Fukumasu; Jayme Augusto de Souza-Neto; José Salvatore Leister Patané; Luiz Alcantara; Luiz Lehmann Coutinho; Maria Carolina Elias; Maurício Lacerda Nogueira; Rafael dos Santos Bezerra; Raul Machado Neto; Rejane Maria Tommasini Grotto; Ricardo Haddad; Sandra Coccuzzo Sampaio Vessoni; Simone Kashima; Svetoslav Nanev Slavov; Vincent Louis Viala. |

| | | | |
|---|---|---|---|
| | ANALISES CLINICAS DE RIO CLARO | | |
| EPI_ISL_2209122, EPI_ISL_2209252, EPI_ISL_2209278, EPI_ISL_2210088, EPI_ISL_2344728, EPI_ISL_2344729 | LABORATORIO MUNICIPAL DE ANALISES CLINICAS DE RIO CLARO | Instituto Butantan / FZEA-USP-Pirassununga | Antonio Jorge Martins; Claudia Renata dos Santos Barros; David Schlesinger; Debora Botequio Moretti; Dimas Tadeu Covas; Elaine Cristina Marqueze; Elaine Vieira Santos; Evandra Strazza Rodrigues; Heidge Fukumasu; Jayme Augusto de Souza-Neto; José Salvatore Leister Patané; Luiz Alcantara; Luiz Lehmann Coutinho; Maria Carolina Elias; Maurício Lacerda Nogueira; Rafael dos Santos Bezerra; Raul Machado Neto; Rejane Maria Tommasini Grotto; Ricardo Haddad; Sandra Coccuzzo Sampaio Vessoni; Simone Kashima; Svetoslav Nanev Slavov; Vincent Louis Viala |
| EPI_ISL_1795100, EPI_ISL_2344549, EPI_ISL_2344734, EPI_ISL_2344801, EPI_ISL_2344826, EPI_ISL_2344884 | LABORATORIO MUNICIPAL DE PIRACICABA | Instituto Butantan / ESALQ-Piracicaba | Antonio Jorge Martins; Bianca Cechetto Carlos. Mendelics: Bibiana Santos; Claudia Renata dos Santos Barros; David Schlesinger; David Schlesinger. Hemocentro Ribeirão Preto: Simone Kashima; Debora Botequio Moretti; Debora Botequio Moretti. Centro de Genômica Funcional da ESALQ: Luiz Lehmann Coutinho; Dimas Tadeu Covas; Elaine Cristina Marqueze; Elaine Vieira Santos; Elaine Vieira dos Santos; Elisangela Chicaroni Mattos; Erika Freitas; Evandra Strazza Rodrigues; Felipe Allan da Silva da Costa; Flavia Aburjaile; Guilherme Targino Valente; Heidge Fukumasu; Heidge Fukumasu. USP-Botucatu: Rejane Maria Tommasini Grotto; Instituto Butantan: Alexander Roberto Precioso; Jayme A. Souza-Neto; Jayme Augusto de Souza-Neto; Jessika Cristina Chagas Lesbon; José Salvatore Leister Patané; João Paulo Kitajima; Luiz Alcantara; Luiz Carlos Junior de Alcantara; Luiz Lehmann Coutinho; Maria Carolina Elias; Marta Giovanetti; Maurício Lacerda Nogueira; Patricia Akemi Assato; Rafael dos Santos Bezerra; Raquel de Lello Rocha Campos Cassano. NGS Soluções Genômicas: Pilar Drummond Sampaio Corrêa Mariani. FZEA-USP Pirassununga: Mirele Daiana Poleti; Raul Machado Neto; Rejane Maria Tommasini Grotto; Ricardo Augusto Brassaloti; Ricardo Haddad; Rodrigo Tocantins Calado.; Sandra Coccuzzo Sampaio; Sandra Coccuzzo Sampaio Vessoni; Simone Kashima; Svetoslav Nanev Slavov; Vagner Fonseca; Vincent Louis Viala |
| EPI_ISL_1966996, EPI_ISL_1966998, EPI_ISL_1966999, EPI_ISL_1967001, EPI_ISL_1967002, EPI_ISL_1967003 | LABORATORIO MUNICIPAL DE PIRACICABA | Instituto Butantan / ESALQ-USP (Piracicaba) | Antonio Jorge Martins; Bianca Cechetto Carlos. Mendelics: Bibiana Santos; Claudia Renata dos Santos Barros; Cíntia Bittar; David Schlesinger. Hemocentro Ribeirão Preto: Simone Kashima; Debora Botequio Moretti; Elaine Cristina Marqueze; Elaine Vieira dos Santos; Elisangela Chicaroni Mattos; Erika Freitas; Evandra Strazza Rodrigues; Felipe Allan da Silva da Costa; Flavia Aburjaile; Fábio Sossai Possebon; Guilherme Campos; Guilherme Targino Valente; Heidge Fukumasu. USP-Botucatu: Rejane Maria Tommasini Grotto; Helena Lage Ferreira; Instituto Butantan: Dimas Tadeu Covas; Jardelina de Souza Todao Bernardino; Jayme A. Souza-Neto; Jessika Cristina Chagas Lesbon; Jorge A. Petroli Marchesi; José Salvatore Leister Patané; João Paulo Kitajima; João Pessoa Araújo Jr.; Leila Sabrina Ullmann; Loyze Paola Oliveira de Lima; Luiz Aurelio de Campos Crispin. Centro de Genômica Funcional da ESALQ: Luiz Lehmann Coutinho; Luiz Carlos Junior de Alcantara; Lívia Sacchetto; Maisa C. Pereira Parra; Maria Carolina Elias; Marta Giovanetti; Marília Moraes; Maurício Lacerda Nogueira. Prefeitura de Sao Paulo: Melissa Palmieri.; Patricia Akemi Assato; Paula Rahal; Paulo Inacio da Costa; Rafael dos Santos Bezerra; Raquel de Lello Rocha Campos Cassano. NGS Soluções Genômicas: Pilar Drummond Sampaio Corrêa Mariani. FZEA-USP Pirassununga: Mirele Daiana Poleti; Raul Machado Neto; Ricardo Augusto Brassaloti; Ricardo Haddad; Rodrigo Tocantins Calado. FAMERP-SJRP: Cecília Artico Banho; Sandra Coccuzzo Sampaio; Svetoslav Nanev Slavov; Vagner Fonseca; Vincent Louis Viala |
| EPI_ISL_1967149, EPI_ISL_1967150, EPI_ISL_1967154, EPI_ISL_1967155 | LABORATORIO MUNICIPAL DE SUZANO | Instituto Butantan / ESALQ-USP (Piracicaba) | Antonio Jorge Martins; Bianca Cechetto Carlos. Mendelics: Bibiana Santos; Claudia Renata dos Santos Barros; Cíntia Bittar; David Schlesinger. Hemocentro Ribeirão Preto: Simone Kashima; Debora Botequio Moretti; Elaine Cristina Marqueze; Elaine Vieira dos Santos; Elisangela Chicaroni Mattos; Erika Freitas; Evandra Strazza Rodrigues; Felipe Allan da Silva da Costa; Flavia Aburjaile; Fábio Sossai Possebon; Guilherme Campos; Guilherme Targino Valente; Heidge Fukumasu. USP-Botucatu: Rejane Maria Tommasini Grotto; Helena Lage Ferreira; Instituto Butantan: Dimas Tadeu Covas; Jardelina de Souza Todao Bernardino; Jayme A. Souza-Neto; Jessika Cristina Chagas Lesbon; Jorge A. Petroli Marchesi; José Salvatore Leister Patané; João Paulo Kitajima; João Pessoa Araújo Jr.; Leila Sabrina Ullmann; Loyze Paola Oliveira de Lima; Luiz Aurelio de Campos Crispin. Centro de Genômica Funcional da ESALQ: Luiz Lehmann Coutinho; Luiz Carlos Junior de Alcantara; Lívia Sacchetto; Maisa C. Pereira Parra; Maria Carolina Elias; Marta Giovanetti; Marília Moraes; Maurício Lacerda Nogueira. Prefeitura de Sao Paulo: Melissa Palmieri.; Patricia Akemi Assato; Paula Rahal; Paulo Inacio da Costa; Rafael dos Santos Bezerra; Raquel de Lello Rocha Campos Cassano. NGS Soluções Genômicas: Pilar Drummond Sampaio Corrêa Mariani. FZEA-USP Pirassununga: Mirele Daiana Poleti; Raul Machado Neto; Ricardo Augusto Brassaloti; Ricardo Haddad; Rodrigo Tocantins Calado. FAMERP-SJRP: Cecília Artico Banho; Sandra Coccuzzo Sampaio; Svetoslav Nanev Slavov; Vagner Fonseca; Vincent Louis Viala |
| EPI_ISL_2801314 | LABORATÓRIO SÃO CARLOS | Analytical Competence Molecular Epidemiology Lab/ACME, Oswaldo Cruz Foundation, Ceara (FIOCRUZ CE) | Cleber Furtado Aksenen e Suzana Porto Almeida; Fabio Miyajima; Fernando Braga Stehling; Francisco Eder de Moura Lopes; Jamille Maria Mendes Bezerra; Joaquim César do Nascimento Sousa Junior; Pedro Miguel Carneiro Jeronimo; Thais Ferreira de Oliveira; Thais de Oliveira Costa; Ticiane Cavalcante de Souza; Veridiana Pessoa Miyajima |
| EPI_ISL_2488779, EPI_ISL_2488787, EPI_ISL_2488788, EPI_ISL_2488789, EPI_ISL_2488790 | LACEN - Laboratório Central de Saúde Pública de Pernambuco | Evandro Chagas Institute | A.M.; Barbagelata; E.C.; E.M.A.; Ferreira; J.A.; Junior; K.C.; L.C.; L.S.; M.C.; P.S.; Pinheiro; Santos; Silva; Sousa; Sousa Junior; W.D.C.; da Silva |
| EPI_ISL_1261687 | LACEN - Laboratório Central de Saúde Pública de Roraima | Evandro Chagas Institute | A.M.; Barbagelata; E.C.; E.M.A.; Ferreira; J.A.; Junior; K.C.; L.C.; L.S.; M.C.; P.S.; Pinheiro; Santos; Silva; Sousa; Sousa Junior; W.D.C.; da Silva |
| EPI_ISL_1164984, EPI_ISL_1261686, EPI_ISL_1261688, EPI_ISL_1261689, EPI_ISL_1261692, EPI_ISL_2488776, EPI_ISL_2488792, EPI_ISL_2488793, EPI_ISL_2488794, EPI_ISL_2488804, EPI_ISL_2488805, EPI_ISL_2488806, EPI_ISL_2488807 | | | |
| see above | LACEN - Laboratório Central de Saúde Pública do Amapá | Evandro Chagas Institute | A.M.; Barbagelata; E.C.; E.M.A.; Ferreira; J.A.; Junior; K.C.; L.C.; L.S.; M.C.; P.S.; Pinheiro; Santos; Silva; Sousa; Sousa Junior; W.D.C.; da Silva |
| EPI_ISL_918499, EPI_ISL_918500, EPI_ISL_918501, EPI_ISL_918502, EPI_ISL_918503, EPI_ISL_918504, EPI_ISL_918505, EPI_ISL_918506, EPI_ISL_918507, EPI_ISL_918508, EPI_ISL_918509, EPI_ISL_918510, EPI_ISL_918511, EPI_ISL_1261683, EPI_ISL_1261685, EPI_ISL_1261690, EPI_ISL_1261694, EPI_ISL_2488774, EPI_ISL_2488775 | | | |
| see above | LACEN - Laboratório Central de Saúde Pública do Amazonas | Evandro Chagas Institute | A.M.; Barbagelata; E.C.; E.M.A.; Ferreira; J.A.; Junior; K.C.; L.C.; L.S.; M.C.; P.S.; Pinheiro; Santos; Silva; Sousa; Sousa Junior; W.D.C.; da Silva |
| EPI_ISL_1164970, EPI_ISL_1164971, EPI_ISL_1164973, EPI_ISL_1164980, EPI_ISL_1261684, EPI_ISL_1261693, EPI_ISL_2488799, EPI_ISL_2488800, EPI_ISL_2488801, EPI_ISL_2488802, EPI_ISL_2488803 | | | |
| see above | LACEN - Laboratório Central de Saúde Pública do Ceará | Evandro Chagas Institute | A.M.; Barbagelata; E.C.; E.M.A.; Ferreira; J.A.; Junior; K.C.; L.C.; L.S.; M.C.; P.S.; Pinheiro; Santos; Silva; Sousa; Sousa Junior; W.D.C.; da Silva |
| EPI_ISL_2488808, EPI_ISL_2488809, EPI_ISL_2488811 | LACEN - Laboratório Central de Saúde Pública do Maranhao | Evandro Chagas Institute | A.M.; Barbagelata; E.C.; E.M.A.; Ferreira; J.A.; Junior; K.C.; L.C.; L.S.; M.C.; P.S.; Pinheiro; Santos; Silva; Sousa; Sousa Junior; W.D.C.; da Silva |
| EPI_ISL_1164991 | LACEN - Laboratório Central de Saúde Pública do Paraiba | Evandro Chagas Institute | A.M.; Barbagelata; E.C.; E.M.A.; Ferreira; J.A.; Junior; K.C.; L.C.; L.S.; M.C.; P.S.; Pinheiro; Santos; Silva; Sousa; Sousa Junior; W.D.C.; da Silva |
| EPI_ISL_904120, EPI_ISL_904121, EPI_ISL_1164972, EPI_ISL_1164978 | LACEN - Laboratório Central de Saúde Pública do Pará | Evandro Chagas Institute | A.M.; Barbagelata; E.C.; E.M.A.; Ferreira; J.A.; Junior; K.C.; L.C.; L.S.; M.C.; P.S.; Pinheiro; Santos; Silva; Sousa; Sousa Junior; W.D.C.; da Silva |
| EPI_ISL_1166615 | LACEN - Laboratório Central de Saúde Pública do Rio Grande do Norte | Evandro Chagas Institute Virology | A.M.; Barbagelata; E.C.; E.M.A.; Ferreira; J.A.; Junior; K.C.; L.C.; L.S.; M.C.; P.S.; Pinheiro; Santos; Silva; Sousa; Sousa Junior; W.D.C.; da Silva |
| EPI_ISL_2488812 | LACEN - Laboratório Central de Saúde Pública do Rio Grande do Norte | Evandro Chagas Institute | A.M.; Barbagelata; E.C.; E.M.A.; Ferreira; J.A.; Junior; K.C.; L.C.; L.S.; M.C.; P.S.; Pinheiro; Santos; Silva; Sousa; Sousa Junior; W.D.C.; da Silva |
| EPI_ISL_2488772, EPI_ISL_2488780, EPI_ISL_2488781, EPI_ISL_2488782, EPI_ISL_2488783, EPI_ISL_2488784, EPI_ISL_2488785, EPI_ISL_2488786 | | | |
| see above | LACEN - Laboratório Central de Saúde Pública do Roraima | Evandro Chagas Institute | A.M.; Barbagelata; E.C.; E.M.A.; Ferreira; J.A.; Junior; K.C.; L.C.; L.S.; M.C.; P.S.; Pinheiro; Santos; Silva; Sousa; Sousa Junior; W.D.C.; da Silva |
| EPI_ISL_1293053, EPI_ISL_1293054, EPI_ISL_1293055, EPI_ISL_1303502, EPI_ISL_1303503 | LACEN de Rondonia | Instituto Adolfo Lutz, Interdiciplinary Procedures Center, Strategic Laboratory | Caio Vinicius Dias Lopes; Claudia Regina Gonçalves; Claudio Tavares Sacchi; Erica Valessa Ramos Gomes; Karoline Rodrigues Campos |
| EPI_ISL_985318, EPI_ISL_985319 | LACEN de Santa Catarina | Instituto Adolfo Lutz, Interdiciplinary Procedures Center, Strategic Laboratory | Claudia Regina Gonçalves; Claudio Tavares Sacchi; Erica Valessa Ramos Gomes; Karoline Rodrigues Campos |
| EPI_ISL_1303506 | LACEN do Distrito Federal | Instituto Adolfo Lutz, Interdiciplinary Procedures Center, Strategic Laboratory | Caio Vinicius Dias Lopes; Claudia Regina Gonçalves; Claudio Tavares Sacchi; Erica Valessa Ramos Gomes; Karoline Rodrigues Campos |
| EPI_ISL_943990, EPI_ISL_985303, EPI_ISL_985304, EPI_ISL_985305, EPI_ISL_985306, EPI_ISL_985307, EPI_ISL_985308, EPI_ISL_985309, EPI_ISL_985310, EPI_ISL_985311, EPI_ISL_985312, EPI_ISL_985313, EPI_ISL_985314, EPI_ISL_985315, EPI_ISL_985316, EPI_ISL_985317, EPI_ISL_1039691, EPI_ISL_1039692, EPI_ISL_1039693, EPI_ISL_1039694, EPI_ISL_1039695, EPI_ISL_1041509, EPI_ISL_1303512, EPI_ISL_1303513, EPI_ISL_1303514, EPI_ISL_1303517, EPI_ISL_1468413, EPI_ISL_1468414, EPI_ISL_1468415, EPI_ISL_1493574, EPI_ISL_1493575, EPI_ISL_1493576, EPI_ISL_1493577, EPI_ISL_1628363, EPI_ISL_1628364, EPI_ISL_1628365, EPI_ISL_2614541, EPI_ISL_2614542, EPI_ISL_2691214, EPI_ISL_2691215, EPI_ISL_2828706, EPI_ISL_2919228, EPI_ISL_2919229, EPI_ISL_2919230, EPI_ISL_2919231, EPI_ISL_2919232 | | | |
| see above | LACEN do Estado de Goias | Instituto Adolfo Lutz, Interdiciplinary Procedures Center, Strategic Laboratory | Caio Vinicius Dias Lopes; Claudia Regina Gonçalves; Claudio Tavares Sacchi; Erica Valessa Ramos Gomes; Karoline Rodrigues Campos; Katia Correa de Oliveira Santos; Leonardo Jose Tadeu de Araujo |
| EPI_ISL_2958884, EPI_ISL_2958885, EPI_ISL_2958886, EPI_ISL_2958888, EPI_ISL_2958889, EPI_ISL_2958890, EPI_ISL_3316168, EPI_ISL_3316173, EPI_ISL_3316186, EPI_ISL_3316187, EPI_ISL_3316191, EPI_ISL_3316194, EPI_ISL_3316195, EPI_ISL_3316209, EPI_ISL_3316210, EPI_ISL_3316214, EPI_ISL_3316215, EPI_ISL_3316218, EPI_ISL_3316220, EPI_ISL_3316224 | | | |
| see above | LACEN do Estado de Mato Grosso | Instituto Adolfo Lutz, Interdiciplinary Procedures Center, Strategic Laboratory | Caio Vinicius Dias Lopes; Claudia Regina Gonçalves; Claudio Tavares Sacchi; Erica Valessa Ramos Gomes; Karoline Rodrigues Campos; Leonardo Tadeu de Araujo; Marlon Benedito Nascimento Santos |
| EPI_ISL_1493578, | LACEN do Estado de | Instituto Adolfo Lutz, | Caio Vinicius Dias Lopes; Claudia Regina Gonçalves; Claudio Tavares Sacchi; Erica Valessa Ramos Gomes; Karoline Rodrigues Campos |

| | | | |
|---|---|---|---|
| EPI_ISL_1493579, EPI_ISL_1494924, EPI_ISL_1520108, EPI_ISL_1520109 | Rondonia | Interdiciplinary Procedures Center, Strategic Laboratory | |
| EPI_ISL_943987, EPI_ISL_1303509, EPI_ISL_2919216, EPI_ISL_2919217, EPI_ISL_2919219, EPI_ISL_2919220, EPI_ISL_2919222, EPI_ISL_2919223, EPI_ISL_2919224, EPI_ISL_2919227 | | | |
| see above | LACEN do Estado de Tocantins | Instituto Adolfo Lutz, Interdiciplinary Procedures Center, Strategic Laboratory | Caio Vinicius Dias Lopes; Claudia Regina Gonçalves; Claudio Tavares Sacchi; Erica Valessa Ramos Gomes; Karoline Rodrigues Campos |
| EPI_ISL_3671908, EPI_ISL_3671909, EPI_ISL_3671911, EPI_ISL_3671912, EPI_ISL_3671914, EPI_ISL_3671915, EPI_ISL_3671916, EPI_ISL_3671917, EPI_ISL_3671918, EPI_ISL_3691382, EPI_ISL_3691383, EPI_ISL_3691384, EPI_ISL_3691385, EPI_ISL_3691386, EPI_ISL_3691387, EPI_ISL_3691388, EPI_ISL_3691389, EPI_ISL_3691390, EPI_ISL_3946597 | | | |
| see above | LACEN do Estado do Mato Grosso do Sul | Instituto Adolfo Lutz, Interdiciplinary Procedures Center, Strategic Laboratory | Caio Vinicius Dias Lopes; Claudia Regina Gonçalves; Claudio Tavares Sacchi; Karoline Rodrigues Campos; Leonardo Tadeu de Araujo; Marlon Benedito Nascomento Santos |
| EPI_ISL_3691681, EPI_ISL_3691682, EPI_ISL_3691683, EPI_ISL_3691684 | LACEN do Estado do Piaui | Instituto Adolfo Lutz, Interdiciplinary Procedures Center, Strategic Laboratory | Caio Vinicius Dias Lopes; Claudia Regina Gonçalves; Claudio Tavares Sacchi; Karoline Rodrigues Campos; Leonardo Tadeu de Araujo; Marlon Benedito Nascomento Santos |
| EPI_ISL_1358304, EPI_ISL_2761936, EPI_ISL_2958855, EPI_ISL_2958856 | LACEN do Mato Grosso do Sul | Instituto Adolfo Lutz, Interdiciplinary Procedures Center, Strategic Laboratory | Caio Vinicius Dias Lopes; Claudia Regina Gonçalves; Claudio Tavares Sacchi; Erica Valessa Ramos Gomes; Karoline Rodrigues Campos; Leonardo Jose Tadeu de Araujo |
| EPI_ISL_1121316 | LACEN do Rio Grande do Sul | Instituto Adolfo Lutz, Interdiciplinary Procedures Center, Strategic Laboratory | Caio Vinicius Dias Lopes; Claudia Regina Gonçalves; Claudio Tavares Sacchi; Erica Valessa Ramos Gomes; Karoline Rodrigues Campos |
| EPI_ISL_906071, EPI_ISL_940614, EPI_ISL_940615, EPI_ISL_940616, EPI_ISL_940617, EPI_ISL_940618 | LACEN-PI DR. Costa Alvarenga | Instituto Adolfo Lutz, Interdiciplinary Procedures Center, Strategic Laboratory | Claudia Regina Gonçalves; Claudio Tavares Sacchi; Erica Valessa Ramos Gomes; Karoline Rodrigues Campos |
| EPI_ISL_2221885, EPI_ISL_2821263, EPI_ISL_2821265, EPI_ISL_2821268, EPI_ISL_2821269, EPI_ISL_2821270, EPI_ISL_2821271, EPI_ISL_2821273, EPI_ISL_3134631, EPI_ISL_3703554 | | | |
| see above | LACEN/PE | WallauLab on behalf of Fiocruz COVID-19 Genomic Surveillance Network | Alexandre Freitas da Silva; Antonio Mauro Rezende; Cassia Docena; Constância Flávia Junqueira Ayres; Cássia Docena; Duschinka Ribeiro Duarte Guedes; Elisama Helvecio; Filipe Zimmer Dezordi; Gabriel Luz Wallau; Gustavo Barbosa de Lima; Lais Ceschini Machado; Larissa Krokovsky; Laís Ceschini Machado; Lilian Caroliny Amorim Silva; Marcelo Henrique dos Santos Paiva; Matheus Filgueira Bezerra; Sinval Pinto Brandão Filho |
| EPI_ISL_1213168 | LAFEM/UESC | Bioinformatics Laboratory / LNCC | Alessandra P Lamarca; Alexandra L Gerber; Ana Paula Melo Mariano; Ana Paula de C Guimarães; Ana Tereza R Vasconcelos; Angela Maria Guimarães Santos; Bianca Mendes Maciel; Danielle Angst Secco; Eduardo Sérgio Soares Sousa; Eloiza Helena Campana; Francisco Paulo Freire Neto; George Rego Albuquerque; Kátia Castanho Scortecci; Lucymara Fassarella Agnez Lima; Luiz G P de Almeida; Luís Cristóvão Porto; Otavio J. Brustolini; Paulo Ricardo Nascimento; Ronaldo da Silva Francisco Jr; Sandra Rocha Gadelha; Selma Maria Bezerra Jeronimo; Vinícius Pietta Perez |
| EPI_ISL_3134973 | LATE - Laboratório de Técnicas Especiais - Hospital Israelita Albert Einstein | LATE - Laboratório de Técnicas Especiais - Hospital Israelita Albert Einstein | Alexandre Hideaki Takara; Ana Paula Moreira Salles; Anelisie da Silva Santos; Deyvid Amgarten; Erick Gustavo Dorlass; Fernanda de Mello Malta; João Renato Rebello Pinho; Marcio Anunciacao Menezes; Pedro Henrique Sebe Rodrigues; Raquel Riyuzo |
| EPI_ISL_1313008 | LBA | CNR Virus des Infections Respiratoires - France SUD | Antonin Bal; Bruno Lina; Gregory Destras; Gwendolyne Burfin; Hadrien Regue; Laurence Josset; Martine Valette; Quentin Semanas |
| EPI_ISL_1608139 | LBM ALPHABIO, Marseille | LBM ALPHABIO, Marseille | Vincent GARCIA |
| EPI_ISL_1213189, EPI_ISL_1213190, EPI_ISL_1213201, EPI_ISL_1213202, EPI_ISL_1213204, EPI_ISL_1213206, EPI_ISL_1213207 | | | |
| see above | LBM/UFPB | Bioinformatics Laboratory / LNCC | Alessandra P Lamarca; Alexandra L Gerber; Ana Paula Melo Mariano; Ana Paula de C Guimarães; Ana Tereza R Vasconcelos; Angela Maria Guimarães Santos; Bianca Mendes Maciel; Danielle Angst Secco; Eduardo Sérgio Soares Sousa; Eloiza Helena Campana; Francisco Paulo Freire Neto; George Rego Albuquerque; Kátia Castanho Scortecci; Lucymara Fassarella Agnez Lima; Luiz G P de Almeida; Luís Cristóvão Porto; Otavio J. Brustolini; Paulo Ricardo Nascimento; Ronaldo da Silva Francisco Jr; Sandra Rocha Gadelha; Selma Maria Bezerra Jeronimo; Vinícius Pietta Perez |
| EPI_ISL_1960077, EPI_ISL_1960078 | LDSP | Universidad Nacional de Colombia - Laboratorio Genómico One Health | Andres F. Cardona-Rios; Carlos Franco-Muñoz; Carolina Muñoz-Arango; Celeny Ortiz; Daniel O. Maldonado-Perez; Diego A. Álvarez-Díaz; Hector Alejandro Ruiz-Moreno; Idabely Betancur Ortiz; Jorge E. Osorio; Juan P. Hernandez-Ortiz; Karl A Ciuoderis; Katherine Laiton-Donato; Laura Silvana Perez; Lina M. Hurtado; Marcela Mercado-Reyes; Maria Angélica Maya; Maria Stella López; Rita Almanza Payares; Sandra Ines Cano; Simón Villegas Velásquez |
| EPI_ISL_1805639 | LDSP FRONTERIZO | Instituto Nacional de Salud- Dirección de Investigación en Salud Pública | Carlos Franco-Muñoz; Carmen Osorio; Diana Malo; Diego A. Álvarez-Díaz; Diego Andrés Prada; Gerardo Santamaría; Hector Alejandro Ruiz-Moreno; Jhonnatan Reales-González; Jorge Rivera; Juan Camilo Martinez; Julian Naizaque; Katherine Laiton-Donato; Lisseth Pardo; Magdalena Wiesner; Marcela Mercado-Reyes; Maria T. Herrera-Sepúlveda; Marta Lopez Blanco; Martha Lucia Ospina Martinez; Paola Rojas; Sergio Gomez; Sheryll Corchuelo; Ángela Alarcon Cruz |
| EPI_ISL_1469941, EPI_ISL_2296079, EPI_ISL_2296080 | LESP Ciudad de Mexico | Instituto de Diagnostico y Referencia Epidemiologicos (INDRE) | Abril Rodriguez-Maldonado; Ariadna Medina-Benitez; Claudia Wong-Arambula; Ernesto Ramirez-Gonzalez.; Gisela Barrera-Badillo; Irma Lopez-Martinez; Joaquin Quiroz-Mercado; Lucia Hernandez-Rivas; Natividad Cruz-Ortiz; Sergio Rangel-Guerrero; Tatiana Nunez-Garcia; Vanessa Rivero-Arredondo |
| EPI_ISL_2296082 | LESP Estado de Mexico | Instituto de Diagnostico y Referencia Epidemiologicos (INDRE) | Abril Rodriguez-Maldonado; Ariadna Medina-Benitez; Claudia Wong-Arambula; Ernesto Ramirez-Gonzalez.; Gisela Barrera-Badillo; Irma Lopez-Martinez; Joaquin Quiroz-Mercado; Lucia Hernandez-Rivas; Natividad Cruz-Ortiz; Sergio Rangel-Guerrero; Tatiana Nunez-Garcia; Vanessa Rivero-Arredondo |
| EPI_ISL_2105682, EPI_ISL_2296069 | LESP Guanajuato | Instituto de Diagnostico y Referencia Epidemiologicos (INDRE) | Abril Rodriguez-Maldonado; Ariadna Medina-Benitez; Claudia Wong-Arambula; Ernesto Ramirez-Gonzalez.; Gisela Barrera-Badillo; Irma Lopez-Martinez; Joaquin Quiroz-Mercado; Lucia Hernandez-Rivas; Natividad Cruz-Ortiz; Sergio Rangel-Guerrero; Tatiana Nunez-Garcia; Vanessa Rivero-Arredondo |
| EPI_ISL_1805500, EPI_ISL_1805503, EPI_ISL_1805504 | LESP Nuevo Leon | Instituto de Diagnostico y Referencia Epidemiologicos (INDRE) | Abril Rodriguez-Maldonado; Ariadna Medina-Benitez; Claudia Wong-Arambula; Ernesto Ramirez-Gonzalez.; Gisela Barrera-Badillo; Irma Lopez-Martinez; Joaquin Quiroz-Mercado; Lucia Hernandez-Rivas; Natividad Cruz-Ortiz; Sergio Rangel-Guerrero; Tatiana Nunez-Garcia; Vanessa Rivero-Arredondo |
| EPI_ISL_2105693 | LESP Quintana Roo | Instituto de Diagnostico y Referencia Epidemiologicos (INDRE) | Abril Rodriguez-Maldonado; Ariadna Medina-Benitez; Claudia Wong-Arambula; Ernesto Ramirez-Gonzalez.; Gisela Barrera-Badillo; Irma Lopez-Martinez; Joaquin Quiroz-Mercado; Lucia Hernandez-Rivas; Natividad Cruz-Ortiz; Sergio Rangel-Guerrero; Tatiana Nunez-Garcia; Vanessa Rivero-Arredondo |
| EPI_ISL_2296084 | LESP Tamaulipas | Instituto de Diagnostico y Referencia Epidemiologicos (INDRE) | Abril Rodriguez-Maldonado; Ariadna Medina-Benitez; Claudia Wong-Arambula; Ernesto Ramirez-Gonzalez.; Gisela Barrera-Badillo; Irma Lopez-Martinez; Joaquin Quiroz-Mercado; Lucia Hernandez-Rivas; Natividad Cruz-Ortiz; Sergio Rangel-Guerrero; Tatiana Nunez-Garcia; Vanessa Rivero-Arredondo |
| EPI_ISL_1608054 | LHUB-ULB | Labo Klinische Biologie, UZA | Basil Britto Xavier; Christine Lammens; Herman Goossens; Jasmine Coppens; Marie Le Mercier; Veerle Matheeussen |
| EPI_ISL_1250700 | LabPLUS | Institute of Environmental Science and Research (ESR) | Anja Werno; Antje van der Linden; Arlo Upton; Chris Mansell; David Hammer; Dragana Drinkovic; Erasmus Smit; Gary McAuliffe; Hana Sofia Andersson; Hermes Perez; James Ussher; Jill Sherwood; Jing Wang; Joep de Ligt; Josh Freeman; Julia Howard; Juliet Elvy; Lauren Jelly; Mary DeAlmeida; Matt Blakiston; Matt Storey; Matthew Rogers; Max Bloomfield; Michael Addidle; Michelle Balm; Muhammad Faisal; Nikki Freed; Olin Silander; Olivia Stroeven; Rachel Boyle; Sally Roberts; SallyAnn Harbison; Sarah Jefferies; Sharmini Muttaiyah; Susan Morpeth; Susan Taylor; Timothy Blackmore; Vani Sathyendran; Veronica Playle; Virginia Hope; Xiaoyun Ren |
| EPI_ISL_1738825, EPI_ISL_1739172, EPI_ISL_1739174 | Labeto - CAB - Leiria | Instituto Nacional de Saude (INSA) | Borges et al |
| EPI_ISL_1381222, EPI_ISL_1417191, EPI_ISL_1417204, EPI_ISL_1417205, EPI_ISL_1594450, EPI_ISL_1788821, EPI_ISL_1788822, EPI_ISL_1788823, EPI_ISL_1788824, EPI_ISL_1788825, EPI_ISL_1788826, EPI_ISL_1788827, EPI_ISL_1788829, EPI_ISL_1788830, EPI_ISL_1788831, EPI_ISL_1788832, EPI_ISL_1788833, EPI_ISL_1788834, EPI_ISL_1788835, EPI_ISL_1788836, EPI_ISL_1788837, EPI_ISL_1788838, EPI_ISL_1821633, EPI_ISL_1821634, EPI_ISL_1821635, EPI_ISL_1821636, EPI_ISL_1821637, EPI_ISL_1821638, EPI_ISL_1821639, EPI_ISL_1821640, EPI_ISL_1821641, EPI_ISL_1821642, EPI_ISL_1821643, EPI_ISL_1821644, EPI_ISL_1821645, EPI_ISL_1821646, EPI_ISL_1821647, EPI_ISL_1821648, EPI_ISL_1821649, EPI_ISL_1821650, EPI_ISL_1821651, EPI_ISL_1821652, EPI_ISL_1821653, EPI_ISL_1821654, EPI_ISL_1821657 | | | |
| see above | Labo Analyses Med | National Reference Center for Viruses of Respiratory Infections, | Amaury Vaysse; Angela Brisebarre; Camille Capel; Catherine Coignard; Christophe Malabat; Corinne Maufrais; Dominique Rousset; Emmanuelle Permal; Etienne Simon-Lorière; Frédéric Lemoine; Louise Lefrançois; Marion Barbet; Maud Vanpeene; Méline Bizard; OphéLie Said-Delattre; Pierre-Yves Leonard; Rousset Dominique; Sylvie Behillil; Sylvie Van der Werf; Sylvie van der Werf; Vincent Enouf |

| | | | |
|---|---|---|---|
| | | Institut Pasteur, Paris | |
| EPI_ISL_1593984 | Labo Carage | Institut Pasteur de la Guyane | Anne Lavergne; Dominique Rousset |
| EPI_ISL_1727096 | Labor Dr. Heidrich & Kollegen MVZ GmbH Hamburg | Robert Koch Institute | |
| EPI_ISL_1217110 | Labor Dr. Wisplinghoff - Köln | Robert Koch Institute | |
| EPI_ISL_1153503 | Labor Lübeck bzw. Laborärztliche Gemeinschaftspraxis Lübeck | Robert Koch Institute | |
| EPI_ISL_1573258, EPI_ISL_1573260 | Labor Prof. Dr. G. Enders MVZ GbR | Robert Koch Institute | |
| EPI_ISL_1627106, EPI_ISL_1627107, EPI_ISL_1628330, EPI_ISL_1628331, EPI_ISL_2227919, EPI_ISL_2227920, EPI_ISL_2227921, EPI_ISL_2227922, EPI_ISL_2227923, EPI_ISL_2227924, EPI_ISL_2227925, EPI_ISL_2227926 | | | |
| see above | Laboratoire Carage | Institut Pasteur de la Guyane | A. Salmier; Anne Lavergne; Dominique Rousset |
| EPI_ISL_1259270 | Laboratoire Hospitalier Universitaire de Bruxelles (LHUB-ULB) | UAntwerp, Laboratory of Medical Microbiology | Basil Britto Xavier; Christine Lammens; Herman Goossens; Jasmine Coppens; Marie Le Mercier; Veerle Matheeussen |
| EPI_ISL_2374083, EPI_ISL_2374084, EPI_ISL_2374085, EPI_ISL_2374086 | Laboratoire Virologie Saint Louis APHP | Laboratoire Virologie Saint Louis APHP | Constance Delauguerre; Jérome Le Goff; Linda Feghoul; Marie Laure Chaix; Marie Laure Néré; Maud Salmona; Severine Mercier Delarue; Sophia Achaibou |
| EPI_ISL_1384156 | Laboratoire national de sante, Microbiology, Virology | Laboratoire national de sante, Microbiology, Microbial Genomics Platform | Anke Wienecke-Baldacchino; Catherine Ragimbeau; Fatu Djabi; Jessica Tapp; Lise Pignon; Raoul Salmon; Tamir Abdelrahman; Trung Nguyen Nguyen |
| EPI_ISL_1384478 | Laboratoires d'analyses medicales - Ketterthill | Laboratoire national de sante, Microbiology, Microbial Genomics Platform | Anke Wienecke-Baldacchino; Caroline Scheiber; Catherine Ragimbeau; Fatu Djabi; Jessica Tapp; Lise Pignon; Raoul Salmon; Serge Vedy; Tamir Abdelrahman |
| EPI_ISL_2289143, EPI_ISL_2308280 | Laboratori de Referencia de Catalunya | Laboratori de Referencia de Catalunya | Bellosillo B.; Canal M.; Hernandez JJ.; Padilla E.; Ramirez A.; Vilas A. |
| EPI_ISL_2157480, EPI_ISL_2157481, EPI_ISL_2157491, EPI_ISL_2196258, EPI_ISL_2196270, EPI_ISL_2196271, EPI_ISL_2196272, EPI_ISL_2196273, EPI_ISL_2196274, EPI_ISL_2196275, EPI_ISL_2196276, EPI_ISL_2196277, EPI_ISL_2196278, EPI_ISL_2196279, EPI_ISL_2196280, EPI_ISL_2557354, EPI_ISL_2557356 | | | |
| see above | Laboratorio Central de Saude Publica do Estado de Minas Gerais (LACEN/MG) | Laboratory of Respiratory Viruses and Measles, Oswaldo Cruz Institute, FIOCRUZ | Alice Sampaio Rocha; Ana Carolina Mendonca; Andre Felipe Leal Bernardes; Anna Carolina Paixao; Elisa Cavalcante Pereira; Fernando Motta; Luciana Appolinario; Marilda Siqueira on behalf of the Fiocruz COVID-19 Genomic Surveillance Network; Paola Resende; Renata Serrano Lopes; Taina Venas |
| EPI_ISL_2196243, EPI_ISL_2196281, EPI_ISL_2196282, EPI_ISL_2196283, EPI_ISL_2196284, EPI_ISL_2196285, EPI_ISL_2196286, EPI_ISL_2196287, EPI_ISL_2196288, EPI_ISL_2196289, EPI_ISL_2196290, EPI_ISL_2196291, EPI_ISL_2196292, EPI_ISL_2196293, EPI_ISL_2196294, EPI_ISL_2196295, EPI_ISL_2196296, EPI_ISL_2196297, EPI_ISL_2196298, EPI_ISL_2196299, EPI_ISL_2196300, EPI_ISL_2196301, EPI_ISL_2196302, EPI_ISL_2196303, EPI_ISL_2443557, EPI_ISL_2443558, EPI_ISL_2443559, EPI_ISL_2443560, EPI_ISL_2443563, EPI_ISL_2443564, EPI_ISL_2645819, EPI_ISL_2645820, EPI_ISL_2645821, EPI_ISL_2645822, EPI_ISL_2645823, EPI_ISL_2645824, EPI_ISL_2645826, EPI_ISL_2645827, EPI_ISL_2645828, EPI_ISL_2645829, EPI_ISL_2645830, EPI_ISL_2645831, EPI_ISL_2645832, EPI_ISL_2645838, EPI_ISL_2645840, EPI_ISL_2645842, EPI_ISL_2645844, EPI_ISL_2645845, EPI_ISL_2645847, EPI_ISL_2645848, EPI_ISL_2645849, EPI_ISL_2645850, EPI_ISL_2645851, EPI_ISL_2645852, EPI_ISL_2645853, EPI_ISL_2645854, EPI_ISL_2645856, EPI_ISL_2645862, EPI_ISL_2645863, EPI_ISL_2645864, EPI_ISL_2645865, EPI_ISL_2645866, EPI_ISL_2645867, EPI_ISL_2645868, EPI_ISL_2645869, EPI_ISL_2645871, EPI_ISL_2645872, EPI_ISL_2645873, EPI_ISL_2645874, EPI_ISL_2645875, EPI_ISL_2645876, EPI_ISL_2645877, EPI_ISL_2645878, EPI_ISL_2645879, EPI_ISL_2645880, EPI_ISL_2645883, EPI_ISL_2645884, EPI_ISL_2645886, EPI_ISL_2645887, EPI_ISL_2645888, EPI_ISL_2645889, EPI_ISL_2645890, EPI_ISL_2645891, EPI_ISL_2645892, EPI_ISL_2645893, EPI_ISL_2645894, EPI_ISL_2645895, EPI_ISL_2645896, EPI_ISL_2645897, EPI_ISL_2645898, EPI_ISL_2645899, EPI_ISL_2645900, EPI_ISL_2645901, EPI_ISL_2645902, EPI_ISL_2645903, EPI_ISL_2645905, EPI_ISL_2645906, EPI_ISL_2645907, EPI_ISL_2645908, EPI_ISL_2645910, EPI_ISL_2645911, EPI_ISL_2645912, EPI_ISL_2645913, EPI_ISL_2645914, EPI_ISL_2645915, EPI_ISL_2645916, EPI_ISL_2645918, EPI_ISL_2645919, EPI_ISL_2645920, EPI_ISL_2645921, EPI_ISL_2645922, EPI_ISL_2645923, EPI_ISL_2645924, EPI_ISL_2645925, EPI_ISL_2645926, EPI_ISL_2645930, EPI_ISL_2645931, EPI_ISL_2645932, EPI_ISL_4080768 | | | |
| see above | Laboratorio Central de Saude Publica do Estado do Para (LACEN/PA) | Laboratory of Respiratory Viruses and Measles, Oswaldo Cruz Institute, FIOCRUZ | Agatha Soares; Alice Sampaio Rocha; Ana Carolina Mendonca; Anna Carolina Paixao; Elisa Cavalcante Pereira; Fernando Motta; Ighor Arantes; Luciana Appolinario; Marilda Siqueira on behalf of the Fiocruz COVID-19 Genomic Surveillance Network; Paola Resende; Renata Serrano Lopes; Taina Venas; Valnete Andrade |
| EPI_ISL_3982691 | Laboratorio Antonello Pelotas Rio Grande do Sul | Hemocentro de Ribeirao Preto FMRP USP | Antonio Jorge Martins; Claudia Renata dos Santos Barros; David Schlesinger; Debora Botequio Moretti; Dimas Tadeu Covas; Elaine Cristina Marqueze; Elaine Vieira Santos; Evandra Strazza Rodrigues; Heidge Fukumasu; Jayme Augusto de Souza-Neto; José Salvatore Leister Patané; Luiz Alcantara; Luiz Lehmann Coutinho; Maria Carolina Elias; Maurício Lacerda Nogueira; Rafael dos Santos Bezerra; Raul Machado Neto; Rejane Maria Tommasini Grotto; Ricardo Haddad; Rodrigo Proto de Siqueira; Sandra Coccuzzo Sampaio Vessoni; Simone Kashima; Svetoslav Nanev Slavov; VV Cantarelli; Vincent Louis Viala |
| EPI_ISL_3982692 | Laboratorio Antonello Pelotas, Rio Grande do Sul | Hemocentro de Ribeirao Preto FMRP USP | Antonio Jorge Martins; Claudia Renata dos Santos Barros; David Schlesinger; Debora Botequio Moretti; Dimas Tadeu Covas; Elaine Cristina Marqueze; Elaine Vieira Santos; Evandra Strazza Rodrigues; Heidge Fukumasu; Jayme Augusto de Souza-Neto; José Salvatore Leister Patané; Luiz Alcantara; Luiz Lehmann Coutinho; Maria Carolina Elias; Maurício Lacerda Nogueira; Rafael dos Santos Bezerra; Raul Machado Neto; Rejane Maria Tommasini Grotto; Ricardo Haddad; Rodrigo Proto de Siqueira; Sandra Coccuzzo Sampaio Vessoni; Simone Kashima; Svetoslav Nanev Slavov; VV Cantarelli; Vincent Louis Viala |
| EPI_ISL_3982703, EPI_ISL_3982710, EPI_ISL_3982716, EPI_ISL_3982718, EPI_ISL_3982719, EPI_ISL_3982722, EPI_ISL_3982728, EPI_ISL_3982732, EPI_ISL_3982734, EPI_ISL_3982735, EPI_ISL_3982736, EPI_ISL_3982737, EPI_ISL_3982738, EPI_ISL_3982739, EPI_ISL_3982743, EPI_ISL_3982746, EPI_ISL_3982747, EPI_ISL_3982750, EPI_ISL_3982753, EPI_ISL_3982755, EPI_ISL_3982757, EPI_ISL_3982758, EPI_ISL_3982760, EPI_ISL_3982765 | | | |
| see above | Laboratorio Antonello, Pelotas, Rio Grande do Sul | Hemocentro de Ribeirao Preto FMRP USP | Antonio Jorge Martins; Claudia Renata dos Santos Barros; David Schlesinger; Debora Botequio Moretti; Dimas Tadeu Covas; Elaine Cristina Marqueze; Elaine Vieira Santos; Evandra Strazza Rodrigues; Heidge Fukumasu; Jayme Augusto de Souza-Neto; José Salvatore Leister Patané; Luiz Alcantara; Luiz Lehmann Coutinho; Maria Carolina Elias; Maurício Lacerda Nogueira; Rafael dos Santos Bezerra; Raul Machado Neto; Rejane Maria Tommasini Grotto; Ricardo Haddad; Rodrigo Proto de Siqueira; Sandra Coccuzzo Sampaio Vessoni; Simone Kashima; Svetoslav Nanev Slavov; VV Cantarelli; Vincent Louis Viala |
| EPI_ISL_3982713, EPI_ISL_3982733 | Laboratorio Antonello, Pelotas, Rio Grande do Sul | Hemocentro de Ribeirao Preto/FMRP-USP | Antonio Jorge Martins; Claudia Renata dos Santos Barros; David Schlesinger; Debora Botequio Moretti; Dimas Tadeu Covas; Elaine Cristina Marqueze; Elaine Vieira Santos; Evandra Strazza Rodrigues; Heidge Fukumasu; Jayme Augusto de Souza-Neto; José Salvatore Leister Patané; Luiz Alcantara; Luiz Lehmann Coutinho; Maria Carolina Elias; Maurício Lacerda Nogueira; Rafael dos Santos Bezerra; Raul Machado Neto; Rejane Maria Tommasini Grotto; Ricardo Haddad; Rodrigo Proto de Siqueira; Sandra Coccuzzo Sampaio Vessoni; Simone Kashima; Svetoslav Nanev Slavov; VV Cantarelli; Vincent Louis Viala |
| EPI_ISL_1804917 | Laboratorio Aziendale di Microbiologia e Virologia, Azienda Sanitaria dell'Alto Adige | Laboratorio Aziendale di Microbiologia e Virologia, Azienda Sanitaria dell'Alto Adige | Anne Picard; Bartolomeo Mobilio Rodriguez; Chiara Cantaloni; Claudia Volpato; Elisa Masi; Elisabetta Giacobazzi; Elisabetta Pagani; Irene Bianconi; Stefanie Wieser |
| EPI_ISL_1664113, EPI_ISL_1664114, EPI_ISL_1664116, EPI_ISL_1664117, EPI_ISL_1664118, EPI_ISL_1664119, EPI_ISL_1664120, EPI_ISL_1664122, EPI_ISL_1664123, EPI_ISL_1664124, EPI_ISL_1664125, EPI_ISL_1664127, EPI_ISL_1664128, EPI_ISL_1664129, EPI_ISL_1664130, EPI_ISL_1664131, EPI_ISL_1664132, EPI_ISL_1664133, EPI_ISL_1664135, EPI_ISL_1664136, EPI_ISL_1664137, EPI_ISL_1664139, EPI_ISL_1664140, EPI_ISL_1664141, EPI_ISL_1664142, EPI_ISL_1664143, EPI_ISL_1664145, EPI_ISL_1664146, EPI_ISL_1664147, EPI_ISL_1664148, EPI_ISL_1664149, EPI_ISL_1664151, EPI_ISL_1664152, EPI_ISL_1664153, EPI_ISL_1664154, EPI_ISL_1664155, EPI_ISL_1664156, EPI_ISL_1664157, EPI_ISL_1664158, EPI_ISL_1664159, EPI_ISL_1664160, EPI_ISL_1664161, EPI_ISL_1664162, EPI_ISL_1664163, EPI_ISL_1664174, EPI_ISL_1664176, EPI_ISL_1664177, EPI_ISL_1664178, EPI_ISL_1664179, EPI_ISL_1664180, EPI_ISL_1664181, EPI_ISL_1664182, EPI_ISL_1664183, EPI_ISL_1664186, EPI_ISL_1664187, EPI_ISL_1664188, EPI_ISL_1664189, EPI_ISL_1664190, EPI_ISL_1664191, EPI_ISL_1664192, EPI_ISL_1664193, EPI_ISL_1664194, EPI_ISL_1664195, EPI_ISL_1664196, EPI_ISL_1664197, EPI_ISL_1664198, EPI_ISL_1664199, EPI_ISL_1664200, EPI_ISL_1664201, EPI_ISL_1664202, EPI_ISL_1858267, EPI_ISL_1858269, EPI_ISL_1858271, EPI_ISL_1858273, EPI_ISL_1858275, EPI_ISL_1858277, EPI_ISL_1858279, EPI_ISL_1858282, EPI_ISL_1858284, EPI_ISL_1858286, EPI_ISL_1858288, EPI_ISL_1858290, EPI_ISL_1858292, EPI_ISL_1858294, EPI_ISL_1858295, EPI_ISL_1858297, EPI_ISL_1858299, EPI_ISL_1858301, EPI_ISL_1858304, EPI_ISL_1858306, EPI_ISL_1858308, EPI_ISL_1858310, EPI_ISL_1858312, EPI_ISL_1858314, EPI_ISL_1858315, EPI_ISL_1858317, EPI_ISL_1858319, EPI_ISL_1858321, EPI_ISL_1858323, EPI_ISL_1858325, EPI_ISL_1858327, EPI_ISL_1858329, EPI_ISL_1858331, EPI_ISL_1858333, EPI_ISL_1858334, EPI_ISL_1858336, EPI_ISL_1858338, EPI_ISL_1858340, EPI_ISL_1858342, EPI_ISL_1858344, EPI_ISL_1858346, EPI_ISL_1858348, EPI_ISL_1858350, EPI_ISL_1858352, EPI_ISL_1858354, EPI_ISL_1858356, EPI_ISL_1858361, EPI_ISL_1858363, EPI_ISL_1858365, EPI_ISL_1858371, EPI_ISL_1858373, EPI_ISL_1858375, EPI_ISL_1858379, EPI_ISL_1858384, EPI_ISL_1858389, EPI_ISL_1858391, EPI_ISL_1858392, EPI_ISL_1858397, EPI_ISL_1858399, EPI_ISL_1858401, EPI_ISL_1858403, EPI_ISL_1858409, EPI_ISL_1858411, EPI_ISL_1858414, EPI_ISL_1858416, EPI_ISL_1858418, EPI_ISL_1858420, EPI_ISL_1858422, EPI_ISL_1858423, EPI_ISL_1858426, EPI_ISL_1858428, EPI_ISL_1858430, EPI_ISL_1858432, EPI_ISL_1858434, EPI_ISL_1858436, EPI_ISL_1858438, EPI_ISL_1858440, EPI_ISL_1858442, EPI_ISL_1858444, EPI_ISL_1858446, EPI_ISL_1858449, EPI_ISL_1858451, EPI_ISL_1858453, EPI_ISL_1858454, EPI_ISL_1858456, EPI_ISL_1858458, EPI_ISL_1858460, EPI_ISL_1858464, EPI_ISL_1858466, EPI_ISL_1858468, EPI_ISL_1858473, EPI_ISL_1858475, EPI_ISL_1858477, EPI_ISL_1858479, EPI_ISL_1858481, EPI_ISL_1858483, EPI_ISL_1858488, EPI_ISL_1858490, EPI_ISL_1858492, EPI_ISL_1858494, EPI_ISL_1858496, EPI_ISL_1858498, EPI_ISL_1858500, EPI_ISL_1858504, EPI_ISL_1858508, EPI_ISL_1858512, EPI_ISL_1858514, EPI_ISL_1858516, EPI_ISL_1858518, EPI_ISL_1858520, EPI_ISL_1858522, EPI_ISL_1858526, EPI_ISL_1858528, EPI_ISL_1858530, EPI_ISL_1858532, EPI_ISL_1858536, EPI_ISL_1858538, EPI_ISL_1858540, EPI_ISL_1858542, EPI_ISL_1858546, EPI_ISL_1858548, EPI_ISL_1858549, EPI_ISL_1858551, EPI_ISL_1858554, EPI_ISL_1858555, EPI_ISL_1858557, EPI_ISL_1858559, EPI_ISL_1858561, EPI_ISL_1858563, EPI_ISL_1858565, EPI_ISL_1858567, EPI_ISL_1858571, EPI_ISL_1858572, EPI_ISL_1858575, EPI_ISL_1858577, EPI_ISL_1858578, EPI_ISL_1858580, EPI_ISL_1858582, EPI_ISL_1858584, EPI_ISL_1858586, EPI_ISL_1858590, EPI_ISL_1858592, EPI_ISL_1858594, EPI_ISL_1858596, EPI_ISL_1858598, EPI_ISL_1858599, EPI_ISL_1858603, EPI_ISL_1858606, EPI_ISL_1858607, EPI_ISL_1858610, EPI_ISL_1858611, EPI_ISL_1858613, EPI_ISL_1858615, EPI_ISL_1858617, EPI_ISL_1858619, EPI_ISL_1858621, EPI_ISL_1858625, EPI_ISL_1858629, EPI_ISL_1858641, EPI_ISL_1858643, EPI_ISL_1858645, EPI_ISL_1858648, EPI_ISL_1858650, EPI_ISL_1858651, EPI_ISL_1858655, EPI_ISL_1858656, EPI_ISL_1858658, EPI_ISL_1858660, EPI_ISL_1858662, EPI_ISL_1858664, EPI_ISL_1858666, EPI_ISL_1858668, EPI_ISL_1858674, EPI_ISL_1858675, EPI_ISL_1858677, EPI_ISL_1858680, EPI_ISL_1858682, EPI_ISL_1858684, EPI_ISL_1858686, EPI_ISL_1858688, EPI_ISL_1858691, EPI_ISL_1858693, EPI_ISL_1858694, EPI_ISL_1858697, EPI_ISL_1858701, EPI_ISL_1858705, EPI_ISL_1858707, EPI_ISL_1858709, EPI_ISL_1858711, EPI_ISL_1858713, EPI_ISL_1858715, EPI_ISL_1858717, EPI_ISL_1858719, EPI_ISL_1858721, EPI_ISL_1858722, EPI_ISL_1858724, EPI_ISL_1858728, EPI_ISL_1858730, EPI_ISL_1858732, EPI_ISL_1858734, EPI_ISL_1858736, EPI_ISL_1858738, EPI_ISL_1858740, EPI_ISL_1858743, EPI_ISL_1858745, EPI_ISL_1858747, EPI_ISL_1858751, EPI_ISL_1858752, EPI_ISL_1858754, EPI_ISL_1858756, EPI_ISL_2101410, EPI_ISL_2101427, EPI_ISL_2101428, EPI_ISL_2101465, EPI_ISL_2101468, EPI_ISL_2101469, EPI_ISL_2101474, EPI_ISL_2101477, EPI_ISL_2101485, EPI_ISL_2101486, EPI_ISL_2101487, EPI_ISL_2101488, EPI_ISL_2101496, EPI_ISL_2101497, EPI_ISL_2101498, EPI_ISL_2101499, EPI_ISL_2101541, EPI_ISL_2101542, EPI_ISL_2101546, EPI_ISL_2101547, EPI_ISL_2101549, EPI_ISL_2101550, EPI_ISL_2101551, EPI_ISL_2101552, EPI_ISL_2101553, EPI_ISL_2101554, EPI_ISL_2101555, EPI_ISL_2101556, EPI_ISL_2101557, EPI_ISL_2101558, EPI_ISL_2101559, EPI_ISL_2101560, EPI_ISL_2101561, EPI_ISL_2101562, EPI_ISL_2101621, EPI_ISL_2101673, EPI_ISL_2101721, EPI_ISL_2101723, EPI_ISL_2837041 | | | |
| see above | Laboratorio Central Noel Nutels | Bioinformatics Laboratory / LNCC | Alessandra P Lamarca; Alexandra L Gerber; Amilcar Tanuri; Ana Paula de C Guimaraes; Ana Paula de C Guimarães; Ana Tereza R Vasconcelos; Andrea Cony Cavalcanti; Andréa Cony Cavalcanti; Caio Luiz Pereira Ribeiro; Cassia Alves; Cintia Policarpo; Claudia Maria Braga de Mello; Cristiane Gomes da Silva; Diana Mariani; Douglas Terra Machado; Flavio Dias da Silva; Flávio Dias da Silva; Gleidson da Silva de Oliveira; Leandro Magalhaes de Souza; Leandro Magalhães de Souza; Liliane Cavalcante; Luiz G P de Almeida; Marcio Henrique de Oliveira Garcia; Mario Sergio Ribeiro; Ronaldo da Silva F Jr; Silvia Carvalho; Thais Felix Cruz |
| EPI_ISL_1662023 | Laboratorio Central de Epidemiologia (LCE) | Unidad de Genomica Avanzada | Alejandro Sanchez-Flores; Alfredo Herrera-Estrella; Alicia Ocana-Mondragon; Angel Gustavo Salas-Lais; Bernardo Martinez-Miguel; Blanca Taboada; Brenda Irasema Maldonado-Meza; Carla Ivon Herrera-Najera; Carlos F. Arias; Celia Boukadida; Celida Duque Molina; Clara Esperanza Santacruz-Tinoco; Concepcion Grajales-Muniz; Consorcio Mexicano de Vigilancia Genomica (CoViGen-Mex). Authors (in alphabetical order): Julio Elias Alvarado-Yaah; Fernando Fontove-Herrera; Francisco Pulido; Gloria Elena Espinosa-Ayala; Gloria Maria Molina-Salinas; Gloria Vazquez; Hector Esteban Paz-Juarez; Hector Montoya-Fuentes; Helen Haydee Fernanda Ramirez-Plascencia; Jose Antonio Enciso-Moreno; Jose Esteban Munoz-Medina; Jose de Jesus Nunez-Contreras; Juan Bautista Chale-Dzul; Luis Alberto Ochoa-Carrera; Margarita Matias-Florentino; Maria Guadalupe Santiago-Mauricio; Maria Guadalupe de Jesus Mireles-Rivera; Nelly Selem-Mojica; Pavel Isa; Ricardo Grande; Santiago Ávila-Rios; Victor Eduardo Garcia-Arias; Victor Hugo Borja-Aburto |
| EPI_ISL_2091410 | Laboratorio Central de Epidemiología (LCE) | Centro de Investigación en Enfermedades Infecciosas (CIENI), Instituto Nacional de Enfermedades Respiratorias (INER) | Alejandro Sanchez-Flores; Alfredo Herrera-Estrella; Alicia Ocaña-Mondragón; Angel Gustavo Salas-Lais; Bernardo Martínez-Miguel; Blanca Taboada; Brenda Irasema Maldonado-Meza; Carla Ivón Herrera-Najera; Carlos F. Arias; Celia Boukadida; Clara Esperanza Santacruz-Tinoco; Concepción Grajales-Muñiz; Consorcio Mexicano de Vigilancia Genómica (CoViGen-Mex). Authors (in alphabetical order): Julio Elias Alvarado-Yaah; Célida Duque Molina; Fernando Fontove-Herrera; Francisco Pulido; Gloria Elena Espinosa-Ayala; Gloria María Molina-Salinas; Gloria Vazquez; Hector Esteban Paz-Juárez; Hector Montoya-Fuentes; Helen Haydee Fernanda Ramírez-Plascencia; José Antonio Enciso-Moreno; José Esteban Muñoz-Medina; José de Jesús Nuñez-Contreras; Juan Bautista Chale-Dzul; Luis Alberto Ochoa-Carrera; Margarita Matías-Florentino; María Guadalupe Santiago-Mauricio; María Guadalupe de Jesús Mireles-Rivera; Nelly Sélem-Mojica; Pavel Isa; Ricardo Grande; Santiago Ávila-Ríos; Victor Eduardo Garcia-Arias; Víctor Hugo Borja-Aburto |
| EPI_ISL_2402180 | Laboratorio Central de | Unidad de Genomica | Alejandro Sanchez-Flores; Alfredo Herrera-Estrella; Alicia Ocana-Mondragon; Angel Gustavo Salas-Lais; Bernardo Martinez-Miguel; Blanca Taboada; Brenda Irasema Maldonado-Meza; Carla Ivon Herrera-Najera; Carlos F. Arias; Celia Boukadida; Clara Esperanza Santacruz-Tinoco; Concepcion Grajales-Muniz; Consorcio Mexicano de |

| | | | |
|---|---|---|---|
| | Epidemiología (LCE) | Avanzada | Vigilancia Genomica (CoViGen-Mex). Authors (in alphabetical order): Julio Elias Alvarado-Yaah; Fernando Fontove-Herrera; Francisco Pulido; Gloria Elena Espinoza-Ayala; Gloria Maria Molina-Salinas; Gloria Vazquez; Hector Esteban Paz-Juarez; Hector Montoya-Fuentes; Helen Haydee Fernanda Ramirez-Plascencia; Jorge Ivan Salinal-Nevarez; Jose Antonio Enciso-Moreno; Jose Esteban Munoz-Medina; Jose de Jesus Nunez-Contreras; Juan Bautista Chale-Dzul; Luis Alberto Ochoa-Carrera; Margarita Matias-Florentino; Maria Guadalupe Santiago-Mauricio; Maria Guadalupe de Jesus Mireles-Rivera; Nelly Selem-Mojica; Pavel Isa; Ricardo Grande; Santiago Avila-Rios; Victor Hugo Borja-Aburto |
| EPI_ISL_3553584 | Laboratorio Central de Saude Publica de Mato Grosso (LACEN-MT) | Fundação Ezequiel Dias | Andre Leal; Cynthia Vazquez; Elaine Cristina; Felipe Iani; Flavia Aburjaile; Gislene Garcia de Castro Lichs; Glauco Carvalho; Hegger Fritsch; Joilson Xavier; Luiz Alcantara.; Luiz Henrique Ferraz Demarchi; Luiz Takao Watanabe; Marina Castilhos Souza Umaki Zardin; Marta Giovanetti; Natalia Guimaraes; Raquel da Silva Ferreira; Talita Adelino; Vagner Fonseca; de Oliveira |
| EPI_ISL_4030307, EPI_ISL_4030312, EPI_ISL_4030318, EPI_ISL_4030368, EPI_ISL_4030369, EPI_ISL_4030370, EPI_ISL_4030371, EPI_ISL_4030372, EPI_ISL_4030373, EPI_ISL_4030374, EPI_ISL_4030375, EPI_ISL_4030376, EPI_ISL_4030377, EPI_ISL_4030378, EPI_ISL_4030379, EPI_ISL_4030380, EPI_ISL_4030381, EPI_ISL_4030382 | | | |
| see above | Laboratorio Central de Saude Publica do Amazonas - LACEN-AM | Laboratorio de Ecologia de Doencas Transmissiveis na Amazonia, Instituto Leonidas e Maria Deane - Fiocruz Amazonia | André Corado; Debora Duarte; Felipe Naveca; Fernanda Nascimento; George Silva; Karina Pessoa; Luciana Gonçalves; Maria Júlia Brandão; Matilde Mejía; Michele Jesus; Valdinete Nascimento; Victor Souza; Ágatha Costa |
| EPI_ISL_2157412, EPI_ISL_2157414, EPI_ISL_2157415, EPI_ISL_2157422, EPI_ISL_2157451, EPI_ISL_2157461, EPI_ISL_2157489, EPI_ISL_2157490, EPI_ISL_2157507, EPI_ISL_2157508, EPI_ISL_2157509, EPI_ISL_2157512, EPI_ISL_2157514, EPI_ISL_2157519, EPI_ISL_2157520, EPI_ISL_2157521, EPI_ISL_2157522, EPI_ISL_2157523, EPI_ISL_2157541 | | | |
| see above | Laboratorio Central de Saude Publica do Esatado de Alagoas (LACEN/AL) | Laboratory of Respiratory Viruses and Measles, Oswaldo Cruz Institute, FIOCRUZ | Alice Sampaio Rocha; Ana Carolina Mendonca; Anderson Brandao Leite; Anna Carolina Paixao; Elisa Cavalcante Pereira; Fernando Motta; Luciana Appolinario; Marilda Siqueira on behalf of the Fiocruz COVID-19 Genomic Surveillance Network; Paola Resende; Renata Serrano Lopes; Taina Venas |
| EPI_ISL_2196263, EPI_ISL_2196264, EPI_ISL_2196265, EPI_ISL_2196266, EPI_ISL_2196267, EPI_ISL_2196268, EPI_ISL_2196269, EPI_ISL_2196355, EPI_ISL_2196358, EPI_ISL_2274076, EPI_ISL_2274077, EPI_ISL_2274078, EPI_ISL_2274079, EPI_ISL_2274080, EPI_ISL_2274082, EPI_ISL_2274083, EPI_ISL_2274084, EPI_ISL_2274086, EPI_ISL_2274087, EPI_ISL_2274088, EPI_ISL_2274090, EPI_ISL_2443556, EPI_ISL_2536355 | | | |
| see above | Laboratorio Central de Saude Publica do Estado Maranhao (LACEN-MA) | Laboratory of Respiratory Viruses and Measles, Oswaldo Cruz Institute, FIOCRUZ | Alice Sampaio Rocha; Ana Carolina Mendonca; Anna Carolina Paixao; Elisa Cavalcante Pereira; Fernando Motta; Lidio Gonçalves Lima Neto; Luciana Appolinario; Marilda Siqueira on behalf of the Fiocruz COVID-19 Genomic Surveillance Network; Paola Resende; Renata Serrano Lopes; Taina Venas |
| EPI_ISL_2157531, EPI_ISL_2491697, EPI_ISL_2491698, EPI_ISL_2491699, EPI_ISL_2491700, EPI_ISL_2491701, EPI_ISL_2491702, EPI_ISL_2491703, EPI_ISL_2491704, EPI_ISL_2491705, EPI_ISL_2491706, EPI_ISL_2491707, EPI_ISL_2491708, EPI_ISL_2491709, EPI_ISL_2491710, EPI_ISL_2491711, EPI_ISL_2491712 | | | |
| see above | Laboratorio Central de Saude Publica do Estado da Bahia (LACEN/BA) | Laboratory of Respiratory Viruses and Measles, Oswaldo Cruz Institute, FIOCRUZ | Alice Sampaio Rocha; Ana Carolina Mendonca; Anna Carolina Paixao; Elisa Cavalcante Pereira; Felicidade Pereira; Fernando Motta; Luciana Appolinario; Marilda Siqueira on behalf of the Fiocruz COVID-19 Genomic Surveillance Network; Paola Resende; Renata Serrano Lopes; Taina Venas |
| EPI_ISL_2157379, EPI_ISL_2157380, EPI_ISL_2157383, EPI_ISL_2157394, EPI_ISL_2157395, EPI_ISL_2157401, EPI_ISL_2157425, EPI_ISL_2274092, EPI_ISL_2274093, EPI_ISL_2536303, EPI_ISL_2536304, EPI_ISL_2536305, EPI_ISL_2536306, EPI_ISL_2536307, EPI_ISL_2536308, EPI_ISL_2536309, EPI_ISL_2536310, EPI_ISL_2536312, EPI_ISL_2536313, EPI_ISL_2536314, EPI_ISL_2536315, EPI_ISL_2536316, EPI_ISL_2536317, EPI_ISL_2536318, EPI_ISL_2536319, EPI_ISL_2536320, EPI_ISL_2536321, EPI_ISL_2536322, EPI_ISL_2536324, EPI_ISL_2536325, EPI_ISL_2536326, EPI_ISL_2536328, EPI_ISL_2536329, EPI_ISL_2536331, EPI_ISL_2536332, EPI_ISL_2536333, EPI_ISL_2536334, EPI_ISL_2536336, EPI_ISL_2536337, EPI_ISL_2536339, EPI_ISL_2536340, EPI_ISL_2536341, EPI_ISL_2536342, EPI_ISL_2536343, EPI_ISL_2536345, EPI_ISL_2536346, EPI_ISL_2536347, EPI_ISL_2536348, EPI_ISL_2536349, EPI_ISL_2536350, EPI_ISL_2536351, EPI_ISL_2536353, EPI_ISL_3061880, EPI_ISL_3061881, EPI_ISL_3061883, EPI_ISL_3061884, EPI_ISL_3061885, EPI_ISL_3061886, EPI_ISL_3061887, EPI_ISL_3061888 | | | |
| see above | Laboratorio Central de Saude Publica do Estado da Paraiba (LACEN-PB) | Laboratory of Respiratory Viruses and Measles, Oswaldo Cruz Institute, FIOCRUZ | Alice Sampaio Rocha; Ana Carolina Mendonca; Anna Carolina Paixao; Dalane Loudal Florentino Teixeira; Elisa Cavalcante Pereira; Fernando Motta; Irina Riediger; Joao Felipe Bezerra; Luciana Appolinario; Marilda Siqueira on behalf of the Fiocruz COVID-19 Genomic Surveillance Network; Paola Resende; Renata Serrano Lopes; Taina Venas |
| EPI_ISL_2274041, EPI_ISL_2274042, EPI_ISL_2274043, EPI_ISL_2274044, EPI_ISL_2274045, EPI_ISL_2274046, EPI_ISL_2274047, EPI_ISL_2274048, EPI_ISL_2274049, EPI_ISL_2274050, EPI_ISL_2274051, EPI_ISL_2274052, EPI_ISL_2274053, EPI_ISL_2274054, EPI_ISL_2274055, EPI_ISL_2274056, EPI_ISL_2274057, EPI_ISL_2274058, EPI_ISL_2274059, EPI_ISL_2274060, EPI_ISL_2274061, EPI_ISL_2274062, EPI_ISL_2274063, EPI_ISL_2274064, EPI_ISL_2274065, EPI_ISL_2274066, EPI_ISL_2274067, EPI_ISL_2274068, EPI_ISL_2274070, EPI_ISL_2645654, EPI_ISL_2645658, EPI_ISL_2645659, EPI_ISL_2645660, EPI_ISL_2645661, EPI_ISL_2645662, EPI_ISL_2645663, EPI_ISL_2645664, EPI_ISL_2645666, EPI_ISL_2645668, EPI_ISL_2645671, EPI_ISL_2645674, EPI_ISL_2645680, EPI_ISL_2645683, EPI_ISL_2645689, EPI_ISL_2645690, EPI_ISL_2645691, EPI_ISL_2645692, EPI_ISL_2645694, EPI_ISL_2645696, EPI_ISL_2645698, EPI_ISL_2645700, EPI_ISL_2645701, EPI_ISL_2645702, EPI_ISL_2645703, EPI_ISL_2645706, EPI_ISL_2645707, EPI_ISL_2645708, EPI_ISL_2645709, EPI_ISL_2645718, EPI_ISL_2645724, EPI_ISL_2645729, EPI_ISL_2645732, EPI_ISL_2645733, EPI_ISL_2645734, EPI_ISL_2645738, EPI_ISL_2645739, EPI_ISL_2645740, EPI_ISL_2645741 | | | |
| see above | Laboratorio Central de Saude Publica do Estado de Alagoas (LACEN/AL) | Laboratory of Respiratory Viruses and Measles, Oswaldo Cruz Institute, FIOCRUZ | Alice Sampaio Rocha; Ana Carolina Mendonca; Anderson Brandao Leite; Anna Carolina Paixao; Elisa Cavalcante Pereira; Fernando Motta; Luciana Appolinario; Marilda Siqueira on behalf of the Fiocruz COVID-19 Genomic Surveillance Network; Paola Resende; Renata Serrano Lopes; Taina Venas |
| EPI_ISL_2038965 | Laboratorio Central de Saude Publica do Estado de Minas Gerais (LACEN-MG) | Laboratory of Respiratory Viruses and Measles, Oswaldo Cruz Institute, FIOCRUZ | Alice Sampaio Rocha; Ana Carolina Mendonca; Andre Felipe Leal Bernardes; Anna Carolina Paixao; Elisa Cavalcante Pereira; Fernando Motta; Luciana Appolinario; Marilda Siqueira on behalf of the Fiocruz COVID-19 Genomic Surveillance Network; Paola Resende; Renata Serrano Lopes; Taina Venas |
| EPI_ISL_2660453, EPI_ISL_2660470, EPI_ISL_2660471, EPI_ISL_2660472, EPI_ISL_2660486, EPI_ISL_2660488, EPI_ISL_2660490, EPI_ISL_2660517, EPI_ISL_2660521, EPI_ISL_2660523, EPI_ISL_2660524, EPI_ISL_2660526, EPI_ISL_2660527, EPI_ISL_2660528, EPI_ISL_2660529, EPI_ISL_2660530, EPI_ISL_2660531, EPI_ISL_2660533, EPI_ISL_2660534, EPI_ISL_2660535, EPI_ISL_2660536, EPI_ISL_2660537, EPI_ISL_2660538, EPI_ISL_2660539, EPI_ISL_2660540, EPI_ISL_2660541, EPI_ISL_2660542 | | | |
| see above | Laboratorio Central de Saude Publica do Estado de Minas Gerais (LACEN/MG) | Laboratory of Respiratory Viruses and Measles, Oswaldo Cruz Institute, FIOCRUZ | Alice Sampaio Rocha; Ana Carolina Mendonca; Andre Felipe Leal Bernardes; Anna Carolina Paixao; Elisa Cavalcante Pereira; Fernando Motta; Luciana Appolinario; Marilda Siqueira on behalf of the Fiocruz COVID-19 Genomic Surveillance Network; Paola Resende; Renata Serrano Lopes; Taina Venas |
| EPI_ISL_1533992, EPI_ISL_1533993, EPI_ISL_1533994, EPI_ISL_1533996, EPI_ISL_1534002, EPI_ISL_1534003, EPI_ISL_1534007, EPI_ISL_1534008, EPI_ISL_1534009, EPI_ISL_1534010, EPI_ISL_2038926, EPI_ISL_2038927, EPI_ISL_2038928, EPI_ISL_2038929, EPI_ISL_2038930, EPI_ISL_2038931, EPI_ISL_2038932, EPI_ISL_2038933, EPI_ISL_2038934, EPI_ISL_2038935, EPI_ISL_2038936, EPI_ISL_2038937, EPI_ISL_2038938, EPI_ISL_2038939, EPI_ISL_2038940, EPI_ISL_2038941, EPI_ISL_2038942, EPI_ISL_2038943, EPI_ISL_2038945, EPI_ISL_2038946, EPI_ISL_2038947, EPI_ISL_2038948, EPI_ISL_2038949, EPI_ISL_2038950, EPI_ISL_2038951, EPI_ISL_2038952, EPI_ISL_2038953, EPI_ISL_2038954, EPI_ISL_2038966, EPI_ISL_2038967 | | | |
| see above | Laboratorio Central de Saude Publica do Estado de Santa Catarina (LACEN-SC) | Laboratory of Respiratory Viruses and Measles, Oswaldo Cruz Institute, FIOCRUZ | Alice Sampaio Rocha; Ana Carolina Mendonca; Anna Carolina Paixao; Darcita Buerger Rovaris; Elisa Cavalcante Pereira; Fernando Motta; Luciana Appolinario; Marilda Siqueira on behalf of the Fiocruz COVID-19 Genomic Surveillance Network; Paola Resende; Renata Serrano Lopes; Sandra Bianchini Fernandes; Taina Venas |
| EPI_ISL_3190166, EPI_ISL_3434917 | Laboratorio Central de Saude Publica do Estado de Santa Catarina (LACEN/SC) | Laboratory of Respiratory Viruses and Measles, Oswaldo Cruz Institute, FIOCRUZ | Alice Sampaio Rocha; Ana Carolina Mendonca; Anna Carolina Paixao; Darcita Buerger Rovaris; Elisa Cavalcante Pereira; Fernando Motta; Luciana Appolinario; Marilda Siqueira on behalf of the Fiocruz COVID-19 Genomic Surveillance Network; Paola Resende; Renata Serrano Lopes; Sandra Bianchini Fernandes; Taina Venas |
| EPI_ISL_1534004 | Laboratorio Central de Saude Publica do Estado de Sergipe (LACEN-SE) | Laboratory of Respiratory Viruses and Measles, Oswaldo Cruz Institute, FIOCRUZ | Alice Sampaio Rocha; Ana Carolina Mendonca; Anna Carolina Paixao; Cliomar Alves dos Santos; Fernando Motta; Luciana Appolinario; Marilda Siqueira on behalf of the Fiocruz COVID-19 Genomic Surveillance Network; Paola Resende; Renata Serrano Lopes |
| EPI_ISL_2157381, EPI_ISL_2157384, EPI_ISL_2157385, EPI_ISL_2157389, EPI_ISL_2157427, EPI_ISL_2157430, EPI_ISL_2157432, EPI_ISL_2157433, EPI_ISL_2157434, EPI_ISL_2157436, EPI_ISL_2157437, EPI_ISL_2157438, EPI_ISL_2157439, EPI_ISL_2157440, EPI_ISL_2157441, EPI_ISL_2157443, EPI_ISL_2157445, EPI_ISL_2157446, EPI_ISL_2157454, EPI_ISL_2157456, EPI_ISL_2157457, EPI_ISL_2157458, EPI_ISL_2157459, EPI_ISL_2157460, EPI_ISL_2157462, EPI_ISL_2157463, EPI_ISL_2157464, EPI_ISL_2157465, EPI_ISL_2157466, EPI_ISL_2157467, EPI_ISL_2157468, EPI_ISL_2157469, EPI_ISL_2157470, EPI_ISL_2157472, EPI_ISL_2157475, EPI_ISL_2157476, EPI_ISL_2157477, EPI_ISL_2157546, EPI_ISL_2196210, EPI_ISL_2196212, EPI_ISL_2196214, EPI_ISL_2196216, EPI_ISL_2196217, EPI_ISL_2196218, EPI_ISL_2196219, EPI_ISL_2196221, EPI_ISL_2196222, EPI_ISL_2196223, EPI_ISL_2196224, EPI_ISL_2196225, EPI_ISL_2196226, EPI_ISL_2196227, EPI_ISL_2196228, EPI_ISL_2196229, EPI_ISL_2196230, EPI_ISL_2196231, EPI_ISL_2196232, EPI_ISL_2196233, EPI_ISL_2196234, EPI_ISL_2196235, EPI_ISL_2196237, EPI_ISL_2196351, EPI_ISL_2196352, EPI_ISL_2660563, EPI_ISL_2660564, EPI_ISL_2660565, EPI_ISL_2660566, EPI_ISL_2660569, EPI_ISL_2660573, EPI_ISL_2660576, EPI_ISL_2660578, EPI_ISL_2660580, EPI_ISL_2660583, EPI_ISL_2660584, EPI_ISL_2660587, EPI_ISL_2660589, EPI_ISL_2660594, EPI_ISL_2660595, EPI_ISL_2660599, EPI_ISL_2660601, EPI_ISL_2660602, EPI_ISL_2660603, EPI_ISL_2660604, EPI_ISL_2660609, EPI_ISL_2660611, EPI_ISL_2660612, EPI_ISL_2660613, EPI_ISL_2660614, EPI_ISL_2660615, EPI_ISL_2660617, EPI_ISL_2660618, EPI_ISL_2660620, EPI_ISL_2660625, EPI_ISL_2660626, EPI_ISL_2660627, EPI_ISL_2660629, EPI_ISL_2660630, EPI_ISL_2660631, EPI_ISL_2660633, EPI_ISL_2660636, EPI_ISL_2660639, EPI_ISL_2660640, EPI_ISL_2660641, EPI_ISL_2660642, EPI_ISL_2660643, EPI_ISL_2660644, EPI_ISL_2660646, EPI_ISL_2660648, EPI_ISL_2660650, EPI_ISL_2660651, EPI_ISL_2660652, EPI_ISL_2660653, EPI_ISL_2660655, EPI_ISL_2660656, EPI_ISL_2660659, EPI_ISL_2660661, EPI_ISL_2660691, EPI_ISL_2660692, EPI_ISL_2660693, EPI_ISL_2660694, EPI_ISL_2660695, EPI_ISL_2660696, EPI_ISL_3828019 | | | |
| see above | Laboratorio Central de Saude Publica do Estado de Sergipe (LACEN/SE) | Laboratory of Respiratory Viruses and Measles, Oswaldo Cruz Institute, FIOCRUZ | Agatha Soares; Alice Sampaio Rocha; Ana Carolina Mendonca; Anna Carolina Paixao; Cliomar Alves dos Santos; Elisa Cavalcante Pereira; Fernando Motta; Ighor Arantes; Luciana Appolinario; Marilda Siqueira on behalf of the Fiocruz COVID-19 Genomic Surveillance Network; Paola Resende; Renata Serrano Lopes; Tainá Moreira Martins Venas |
| EPI_ISL_1219134, EPI_ISL_1219135, EPI_ISL_2038964 | Laboratorio Central de Saude Publica do Estado do Alagoas (LACEN-AL) | Laboratory of Respiratory Viruses and Measles, Oswaldo Cruz Institute, FIOCRUZ | Alice Sampaio Rocha; Ana Carolina Mendonca; Anderson Brandao Leite; Anna Carolina Paixao; Elisa Cavalcante Pereira; Fernando Motta; Luciana Appolinario; Marilda Siqueira on behalf of the Fiocruz COVID-19 Genomic Surveillance Network; Paola Resende; Renata Serrano Lopes; Taina Venas |
| EPI_ISL_2983106, EPI_ISL_2983114, EPI_ISL_2983125, EPI_ISL_2983131, EPI_ISL_2983132, EPI_ISL_2983134, EPI_ISL_2983135, EPI_ISL_2983139, EPI_ISL_2983140, EPI_ISL_2983142, EPI_ISL_2983143, EPI_ISL_2983144, EPI_ISL_2983145, EPI_ISL_2983146, EPI_ISL_2983147, EPI_ISL_2983149, EPI_ISL_2983150, EPI_ISL_2983151, EPI_ISL_2983152, EPI_ISL_2983153, EPI_ISL_2983155, EPI_ISL_2983156, EPI_ISL_2983157, EPI_ISL_2983158, EPI_ISL_2983159, EPI_ISL_2983160, EPI_ISL_2983163, EPI_ISL_2983164, EPI_ISL_2983165, EPI_ISL_2983167, EPI_ISL_2983169, EPI_ISL_2983170, EPI_ISL_2983171, EPI_ISL_2983172, EPI_ISL_2983173, EPI_ISL_2983174, EPI_ISL_2983175, EPI_ISL_2983177, EPI_ISL_2983178 | | | |
| see above | Laboratorio Central de Saude Publica do Estado do Amapa (LACEN/AP) | Laboratory of Respiratory Viruses and Measles, Oswaldo Cruz Institute, FIOCRUZ | Agatha Cristinne Prudencio; Alice Sampaio Rocha; Ana Carolina Mendonca; Andreia Santos Costa; Anna Carolina Paixao; Anne Caroline da Silva Soledade; Elisa Cavalcante Pereira; Fernando Motta; Ighor Leonardo Arantes Gomes; Lindomar dos Anjos Silva; Luciana Appolinario; Marcia Socorro Pereira Cavalcante; Marilda Siqueira on behalf of the Fiocruz COVID-19 Genomic Surveillance Network; Paola Resende; Renata Serrano Lopes; Taina Venas |
| EPI_ISL_2645515, EPI_ISL_2645517, EPI_ISL_2645518, EPI_ISL_2645521, EPI_ISL_2645583, EPI_ISL_2645584, EPI_ISL_2645589, EPI_ISL_2645621, EPI_ISL_2645623, EPI_ISL_2645625, EPI_ISL_2645630, EPI_ISL_2645634, EPI_ISL_3045429, EPI_ISL_3045430, EPI_ISL_3045431, EPI_ISL_3045435, EPI_ISL_3045436, EPI_ISL_3045437, EPI_ISL_3045438, EPI_ISL_3045439, EPI_ISL_3045442, EPI_ISL_3045444, EPI_ISL_3045445, EPI_ISL_3045447, EPI_ISL_3045452, EPI_ISL_3045454, EPI_ISL_3045455, EPI_ISL_3045456, EPI_ISL_3045459, EPI_ISL_3061878 | | | |
| see above | Laboratorio Central de Saude Publica do Estado do Espirito Santo (LACEN/ES) | Laboratory of Respiratory Viruses and Measles, Oswaldo Cruz Institute, FIOCRUZ | Alice Sampaio Rocha; Ana Carolina Mendonca; Anna Carolina Paixao; Eilisa Cavalcante Pereira; Elisa Cavalcante Pereira; Fernando Motta; Luciana Appolinario; Marilda Siqueira on behalf of the Fiocruz COVID-19 Genomic Surveillance Network; Paola Resende; Renata Serrano Lopes; Rodrigo Ribeiro Rodrigues; Taina Venas |
| EPI_ISL_2038968 | Laboratorio Central de Saude Publica do | Laboratory of Respiratory Viruses and | Alice Sampaio Rocha; Ana Carolina Mendonca; Anna Carolina Paixao; Fernando Motta; Luciana Appolinario; Marilda Siqueira on behalf of the Fiocruz COVID-19 Genomic Surveillance Network; Paola Resende; Renata Serrano Lopes; Rodrigo Ribeiro Rodrigues |

| | | | |
|---|---|---|---|
| | Estado do Espitito Santo (LACEN-ES) | Measles, Oswaldo Cruz Institute, FIOCRUZ | |
| EPI_ISL_1465188, EPI_ISL_1465189, EPI_ISL_1465191, EPI_ISL_1465192, EPI_ISL_1465194, EPI_ISL_1465195, EPI_ISL_1465196, EPI_ISL_1465198, EPI_ISL_1465199, EPI_ISL_1465201, EPI_ISL_1465202, EPI_ISL_1465203, EPI_ISL_1465205, EPI_ISL_1465206, EPI_ISL_1465208, EPI_ISL_1465209, EPI_ISL_1465210, EPI_ISL_1465212, EPI_ISL_1465213, EPI_ISL_1465215, EPI_ISL_1465216, EPI_ISL_1465217, EPI_ISL_1465219, EPI_ISL_1465221, EPI_ISL_1465222, EPI_ISL_2983196, EPI_ISL_2983198, EPI_ISL_2983199, EPI_ISL_2983200, EPI_ISL_2983201, EPI_ISL_2983202, EPI_ISL_2983204, EPI_ISL_2983205, EPI_ISL_2983206, EPI_ISL_2983209, EPI_ISL_2983210, EPI_ISL_2983211, EPI_ISL_2983212, EPI_ISL_2983258, EPI_ISL_2983259, EPI_ISL_2983260, EPI_ISL_2983261, EPI_ISL_2983262, EPI_ISL_2983264, EPI_ISL_2983265, EPI_ISL_2983266, EPI_ISL_2983267, EPI_ISL_2983280, EPI_ISL_2983281, EPI_ISL_2983332, EPI_ISL_2983473 | | | |
| see above | Laboratorio Central de Saude Publica do Estado do Maranhao (LACEN-MA) | Laboratory of Respiratory Viruses and Measles, Oswaldo Cruz Institute, FIOCRUZ | Agatha Cristinne Prudencio; Alice Sampaio Rocha; Ana Carolina Mendonca; Anna Carolina Paixao; Elisa Cavalcante Pereira; Fernando Motta; Ighor Leonardo Arantes Gomes; Lidio Gonçalves Lima Neto; Luciana Appolinario; Marilda Siqueira on behalf of the Fiocruz COVID-19 Genomic Surveillance Network; Paola Resende; Renata Serrano Lopes; Taina Moreira Venas; Tainá Venas |
| EPI_ISL_2274091, EPI_ISL_2863796 | Laboratorio Central de Saude Publica do Estado do Mato Grosso do Sul (LACEN/MS) | Laboratory of Respiratory Viruses and Measles, Oswaldo Cruz Institute, FIOCRUZ | Agatha Cristinne Prudencio Soares; Alex Pauvolid-Corrêa; Alice Sampaio Rocha; Ana Beatriz Machado Lima; Ana Carolina Mendonca; Anna Carolina Paixao; Elisa Cavalcante Pereira; Fernando Motta; Gislene Lichs; Ighor Leonardo Arantes Gomes; Luciana Appolinario; Marilda Siqueira on behalf of the Fiocruz COVID-19 Genomic Surveillance Network; Mia Ferreira de Araujo; Natália Valente da Silva; Paola Resende; Renata Serrano Lopes; Taina Venas; Thayssa Keren Santos da Silva Neves |
| EPI_ISL_2758940, EPI_ISL_2758942, EPI_ISL_2758945, EPI_ISL_2758947, EPI_ISL_2758948, EPI_ISL_2758949, EPI_ISL_2758950, EPI_ISL_2758951, EPI_ISL_2758952, EPI_ISL_2758958, EPI_ISL_2758964, EPI_ISL_2758998, EPI_ISL_2759007, EPI_ISL_2759010, EPI_ISL_2759013, EPI_ISL_2759023, EPI_ISL_2759027, EPI_ISL_2759029, EPI_ISL_2759032, EPI_ISL_2759036, EPI_ISL_2759038, EPI_ISL_2759039, EPI_ISL_2759042, EPI_ISL_2759044, EPI_ISL_2759053, EPI_ISL_2759058, EPI_ISL_2759064, EPI_ISL_2759069 | | | |
| see above | Laboratorio Central de Saude Publica do Estado do Parana (Instituto de Biologia Molecular do Paraná (LAC)EN-PR) | Instituto Carlos Chagas - Fiocruz | Alessandra De Melo Aguiar; Andreia Akemi Suzukawa; Andréa Rodrigues Ávila; Bruno Dallagiovanna; Dalila Zanette; Eduardo Balsanelli; Emanuel Maltempi de Souza; Fabio Passetti; Fabricio Klerynton Marchini; Fábio de Oliveira Pedrosa; Guilherme Becker; Helisson Faoro; Hellen Geremias dos Santos; Irina Nastassja Riediger; Letusa Albrecht; Lucas Blanes; Luis Gustavo Morello; Lysangela Ronalte Alves; Maria do Carmo Debur; Mauro de Medeiros Oliveira; Michelle Orane Schemberger; Paola Cristina Resende; Sheila Cristina Nardeli; Tiago Gräf; Valter Antônio de Baura |
| EPI_ISL_1219132, EPI_ISL_1219133, EPI_ISL_1533999, EPI_ISL_1534000, EPI_ISL_1534001, EPI_ISL_2038955 | Laboratorio Central de Saude Publica do Estado do Parana (LACEN-PR) | Laboratory of Respiratory Viruses and Measles, Oswaldo Cruz Institute, FIOCRUZ | Alice Sampaio Rocha; Ana Carolina Mendonca; Anna Carolina Paixao; Elisa Cavalcante Pereira; Fernando Motta; Irina Nastassja Riediger; Irina Riediger; Luciana Appolinario; Maria do Carmo Debur; Marilda Siqueira on behalf of the Fiocruz COVID-19 Genomic Surveillance Network; Paola Resende; Renata Serrano Lopes; Taina Venas |
| EPI_ISL_2157482, EPI_ISL_3827992 | Laboratorio Central de Saude Publica do Estado do Parana (LACEN/PR) | Laboratory of Respiratory Viruses and Measles, Oswaldo Cruz Institute, FIOCRUZ | Agatha Soares; Alice Sampaio Rocha; Ana Carolina Mendonca; Anna Carolina Paixao; Elisa Cavalcante Pereira; Fernando Motta; Ighor Leonardo Arantes; Irina Riediger; Luciana Appolinario; Marilda Siqueira on behalf of the Fiocruz COVID-19 Genomic Surveillance Network; Paola Resende; Renata Serrano Lopes; Taina Venas |
| EPI_ISL_1534013, EPI_ISL_2038957, EPI_ISL_2038958, EPI_ISL_2038959, EPI_ISL_2038960, EPI_ISL_2274110, EPI_ISL_2274111, EPI_ISL_2274112, EPI_ISL_2274113, EPI_ISL_2274114, EPI_ISL_2274116, EPI_ISL_2274117, EPI_ISL_2274118, EPI_ISL_2274119, EPI_ISL_2274120, EPI_ISL_2274121, EPI_ISL_2274122, EPI_ISL_2274123, EPI_ISL_2274124, EPI_ISL_2274125, EPI_ISL_2274126, EPI_ISL_2274127, EPI_ISL_2274128, EPI_ISL_2274129, EPI_ISL_2274130, EPI_ISL_2274131, EPI_ISL_2274138, EPI_ISL_2443672, EPI_ISL_2466126, EPI_ISL_2466127, EPI_ISL_2466128, EPI_ISL_2466129, EPI_ISL_2466130, EPI_ISL_2466131, EPI_ISL_2466132, EPI_ISL_2466133, EPI_ISL_2466134, EPI_ISL_2466135, EPI_ISL_2466136, EPI_ISL_2466138, EPI_ISL_2466269, EPI_ISL_2603435, EPI_ISL_2603580, EPI_ISL_2661746, EPI_ISL_2661747, EPI_ISL_2661748, EPI_ISL_2661749, EPI_ISL_2661752, EPI_ISL_2661753, EPI_ISL_2661754, EPI_ISL_2661755, EPI_ISL_2661756, EPI_ISL_2661757, EPI_ISL_2661759, EPI_ISL_2661762, EPI_ISL_2661763, EPI_ISL_2661767, EPI_ISL_2661768, EPI_ISL_2661769, EPI_ISL_2661770, EPI_ISL_2661771, EPI_ISL_2661772, EPI_ISL_2661774, EPI_ISL_2661775, EPI_ISL_2661776, EPI_ISL_2661777, EPI_ISL_2661778, EPI_ISL_2661780, EPI_ISL_2661781, EPI_ISL_2661824, EPI_ISL_2661842, EPI_ISL_2661843, EPI_ISL_2661851, EPI_ISL_2661857, EPI_ISL_2661859, EPI_ISL_2661868, EPI_ISL_2661870 | | | |
| see above | Laboratorio Central de Saude Publica do Estado do Rio Grande do Sul (LACEN-RS) | Laboratory of Respiratory Viruses and Measles, Oswaldo Cruz Institute, FIOCRUZ | Alice Sampaio Rocha; Ana Carolina Mendonca; Anderson Brandao Leite; Anna Carolina Paixao; Elisa Cavalcante Pereira; Fernando Motta; Luciana Appolinario; Marilda Siqueira on behalf of the Fiocruz COVID-19 Genomic Surveillance Network; Paola Resende; Renata Serrano Lopes; Richard Salvato; Taina Venas; Tatiana Schaffer Gregianini |
| EPI_ISL_3048764, EPI_ISL_3048771, EPI_ISL_3048775, EPI_ISL_3048776, EPI_ISL_3048777, EPI_ISL_3048780, EPI_ISL_3048782, EPI_ISL_3048784, EPI_ISL_3048786, EPI_ISL_3048789, EPI_ISL_3048792, EPI_ISL_3048793, EPI_ISL_3048794, EPI_ISL_3048796, EPI_ISL_3048798, EPI_ISL_3048800, EPI_ISL_3048801 | | | |
| see above | Laboratorio Central de Saude Publica do Estado do Rio Grande do Sul (LACEN-RS) | Laboratório de Biologia Molecular da Universidade Federal de Ciências da Saúde de Porto Alegre | Adriana Seixas; Ana B. G. Veiga; Ana Paula Muterle Varela; Fabiana Quoos Mayer; Fernando Hayashi Sant'Anna; Janira Prichula; Letícia Garay Martins; Richard Steiner Salvato; Tatiana Schäffer Gregianini |
| EPI_ISL_1534012, EPI_ISL_2038956, EPI_ISL_2196189, EPI_ISL_2196197, EPI_ISL_2196198, EPI_ISL_2196199, EPI_ISL_2196200, EPI_ISL_2196201, EPI_ISL_2196202, EPI_ISL_2196203, EPI_ISL_2196204, EPI_ISL_2196205, EPI_ISL_2196206, EPI_ISL_2196207, EPI_ISL_2196208, EPI_ISL_2196209, EPI_ISL_2196236 | | | |
| see above | Laboratorio Central de Saude Publica do Estado do Rio de Janeiro (LACEN-RJ) | Laboratory of Respiratory Viruses and Measles, Oswaldo Cruz Institute, FIOCRUZ | Alice Sampaio Rocha; Ana Carolina Mendonca; Andrea Cony Cavalcanti; Anna Carolina Paixao; Elisa Cavalcante Pereira; Fernando Motta; Luciana Appolinario; Marilda Siqueira on behalf of the Fiocruz COVID-19 Genomic Surveillance Network; Paola Resende; Renata Serrano Lopes; Taina Venas |
| EPI_ISL_2363545, EPI_ISL_2363547, EPI_ISL_2363559, EPI_ISL_2363560, EPI_ISL_2363568, EPI_ISL_2363569, EPI_ISL_2363570, EPI_ISL_2363573, EPI_ISL_3033032 | | | |
| see above | Laboratorio Central de la Ciudad de Santa Fe | Grupo de Genómica y Bioinformática del Instituto de Investigación de la Cadena Láctea CONICET-INTA on behalf of 'Proyecto Argentino Interinstitucional de genomica de SARS-CoV-2' (PAIS Consortium) | AF; Amadio; C; Eberhardt; G; Irazoqui; JM; MF; Mugna; Ojeda; Pastor; Rompato; V |
| EPI_ISL_3997110, EPI_ISL_3997228, EPI_ISL_3997230, EPI_ISL_3997240, EPI_ISL_3997739, EPI_ISL_3997833, EPI_ISL_3997877, EPI_ISL_3998018, EPI_ISL_3998051, EPI_ISL_3998169, EPI_ISL_3999620, EPI_ISL_4000067, EPI_ISL_4000229, EPI_ISL_4000270, EPI_ISL_4000384, EPI_ISL_4000695, EPI_ISL_4000718, EPI_ISL_4000719, EPI_ISL_4000801, EPI_ISL_4000802, EPI_ISL_4000803, EPI_ISL_4000804, EPI_ISL_4001402, EPI_ISL_4001403, EPI_ISL_4001404, EPI_ISL_4001405, EPI_ISL_4001562, EPI_ISL_4001708, EPI_ISL_4009398 | | | |
| see above | Laboratorio Central do Estado do Parana | Laboratorio Central do Estado do Parana | Guilherme Nardi Becker; Irina Nastassja Riediger; Maria do Carmo Debur Rossa; Mayra Presibella Giacomini; Nelson Quallio Marques |
| EPI_ISL_3924929 | Laboratorio Clinico Toledo | Sequencing and Genomics facility (SGF) | Camille Zenon; Carlos Sariol; Filipa Godoy; Israel Candelaria Velez; Jose Carlos Velez; Jose Rodriguez Orengo; Limari Huertas; Marcia Cruz; Riccardo Papa; Silvia Planas; Steven Van Belleghem; Yadira Ortiz |
| EPI_ISL_2139494, EPI_ISL_2139496, EPI_ISL_2139498, EPI_ISL_2139499, EPI_ISL_2139500, EPI_ISL_2139501, EPI_ISL_2139502, EPI_ISL_2139504, EPI_ISL_2139505, EPI_ISL_2139506, EPI_ISL_2139507, EPI_ISL_2139509, EPI_ISL_2139510, EPI_ISL_2139511, EPI_ISL_2139516, EPI_ISL_2139518, EPI_ISL_2139520, EPI_ISL_2139521, EPI_ISL_2139522, EPI_ISL_2139525, EPI_ISL_2139528, EPI_ISL_2139529, EPI_ISL_2139530, EPI_ISL_2139532, EPI_ISL_2139534, EPI_ISL_2139535, EPI_ISL_2139538, EPI_ISL_2139539, EPI_ISL_2139540, EPI_ISL_2139541, EPI_ISL_2139542, EPI_ISL_2139543, EPI_ISL_2139545, EPI_ISL_2139546, EPI_ISL_2139547, EPI_ISL_2139548, EPI_ISL_2139549 | | | |
| see above | Laboratorio Exame | Universidade Federal de Ciencias da Saude de Porto Alegre | Gabriel Dickin Caldana et al.; Vinícius Bonetti Franceschi |
| EPI_ISL_2614585, EPI_ISL_2614589 | Laboratorio Municipal De Suzano | Instituto Adolfo Lutz, Interdiciplinary Procedures Center, Strategic Laboratory | Caio Vinicius Dias Lopes; Claudia Regina Gonçalves; Claudio Tavares Sacchi; Erica Valessa Ramos Gomes; Karoline Rodrigues Campos; Leonardo Jose Tadeu de Araujo |
| EPI_ISL_2274004, EPI_ISL_2274005 | Laboratorio Nacional de Salud Pública Dr. Defilló - LNSPDD | Laboratory of Respiratory Viruses and Measles, Oswaldo Cruz Institute, FIOCRUZ | Alice Sampaio Rocha; Ana Carolina Mendonca; Anna Carolina Paixao; Elisa Cavalcante Pereira; Fernando Motta; Grey Benoit Vasquez; Isaac Miguel Sanchez; Ivonne Imbert; Lucia de la Cruz; Luciana Appolinario; Marilda Siqueira on behalf of the Fiocruz COVID-19 Genomic Surveillance Network; Nurys de Castro; Paola Resende; Renata Serrano Lopes; Ronald Skewes; Taina Venas |
| EPI_ISL_1716407, EPI_ISL_1716408, EPI_ISL_1716441, EPI_ISL_1716442, EPI_ISL_1716443, EPI_ISL_1716445, EPI_ISL_1716447, EPI_ISL_1716448, EPI_ISL_1716453, EPI_ISL_1716455 | | | |
| see above | Laboratorio Saude | LGBio (Laboratorio de Genetica & Biodiversidade) | Amanda Alves de Melo; Cintia Pelegrineti Targueta de Azevedo Brito; Daniela de Melo e Silva; Elisangela de Paula Silveira Lacerda; Francyelli Mello Andrade; Mariana Pires de Campos Telles; Ramilla dos Santos Braga; Renata de Oliveira Dias; Rhewter Nunes; Thais Guimarães Castro; Thays Millena Alves Pedroso |
| EPI_ISL_2427505, EPI_ISL_2427506, EPI_ISL_2427522, EPI_ISL_2427526, EPI_ISL_2427533, EPI_ISL_2427538, EPI_ISL_2427567, EPI_ISL_2427573, EPI_ISL_2427574, EPI_ISL_2427582, EPI_ISL_2427590, EPI_ISL_2427657, EPI_ISL_2427673, EPI_ISL_2427693, EPI_ISL_2427702, EPI_ISL_2427724, EPI_ISL_2427725, EPI_ISL_2427729, EPI_ISL_2427731, EPI_ISL_2427735, EPI_ISL_2427751, EPI_ISL_2427759, EPI_ISL_2427762, EPI_ISL_2427768, EPI_ISL_2427772, EPI_ISL_2427777 | | | |
| see above | Laboratorio de Biología Molecular Médica Uruguaya | Departments of Pathology and Medicine, New York University School of Medicine | Adriana Heguy; Cecilia Sorhouet; Christian Marier; Dacia Dimartino; Gonzalo Manrique; Maria Cristina Mogdasy; Maria Noel Zubillaga; Maria Victoria Elizondo; Paul Zappile |
| EPI_ISL_811149, EPI_ISL_833136, EPI_ISL_833137, EPI_ISL_833138, EPI_ISL_833139, EPI_ISL_833140, EPI_ISL_1034304, EPI_ISL_1034306, EPI_ISL_1068110, EPI_ISL_1068111, EPI_ISL_1068112, EPI_ISL_1068114, EPI_ISL_1068149, EPI_ISL_1068150, EPI_ISL_1068151, EPI_ISL_1068154, EPI_ISL_1068156, EPI_ISL_1068157, EPI_ISL_1068158, EPI_ISL_1068159, EPI_ISL_1068160, EPI_ISL_1068169, EPI_ISL_1068198, EPI_ISL_1068221, EPI_ISL_1068222, EPI_ISL_1068225, EPI_ISL_1068226, EPI_ISL_1068243, EPI_ISL_1068248, EPI_ISL_1068249, EPI_ISL_1068256, EPI_ISL_1068258, EPI_ISL_1068260, EPI_ISL_1068261, EPI_ISL_1068262, EPI_ISL_1068263, EPI_ISL_1068264, EPI_ISL_1068266, EPI_ISL_1068268, EPI_ISL_1068269, EPI_ISL_1068270, EPI_ISL_1068271, EPI_ISL_1068272, EPI_ISL_1068273, EPI_ISL_1068274, EPI_ISL_1068275, EPI_ISL_1068276, EPI_ISL_1068278, EPI_ISL_1068279, EPI_ISL_1068280, EPI_ISL_1068281, EPI_ISL_1068282, EPI_ISL_1068283, EPI_ISL_1068284, EPI_ISL_1068285, EPI_ISL_1068286, EPI_ISL_1068287, EPI_ISL_1068288, EPI_ISL_1068289, EPI_ISL_1068290, EPI_ISL_1068291, EPI_ISL_1068292, EPI_ISL_1261122, EPI_ISL_1261123, EPI_ISL_1533609, EPI_ISL_2777363, EPI_ISL_2777364, EPI_ISL_2777373, EPI_ISL_2777374, EPI_ISL_2777375, EPI_ISL_2777376, EPI_ISL_2777377, EPI_ISL_2777378, EPI_ISL_2777380, EPI_ISL_2777381, EPI_ISL_2777382, EPI_ISL_2777383, EPI_ISL_2777384, EPI_ISL_2777385, EPI_ISL_2777388, EPI_ISL_2777393, EPI_ISL_2777395, EPI_ISL_2777396, EPI_ISL_2777397, EPI_ISL_2777398, EPI_ISL_2777399, EPI_ISL_2777400, EPI_ISL_2777401, | | | |

| | | | |
|---|---|---|---|
| EPI_ISL_2777402, EPI_ISL_2777403, EPI_ISL_2777404, EPI_ISL_2777405, EPI_ISL_2777406, EPI_ISL_2777407, EPI_ISL_2777408, EPI_ISL_2777409, EPI_ISL_2777410, EPI_ISL_2777411, EPI_ISL_2777412, EPI_ISL_2777413, EPI_ISL_2777414, EPI_ISL_2777415, EPI_ISL_2777416, EPI_ISL_2777417, EPI_ISL_2777418, EPI_ISL_2777419, EPI_ISL_2777420, EPI_ISL_2777421, EPI_ISL_2777423, EPI_ISL_2777424, EPI_ISL_2777425, EPI_ISL_2777426, EPI_ISL_2777427, EPI_ISL_2777428, EPI_ISL_2777429, EPI_ISL_2777430, EPI_ISL_2777431, EPI_ISL_2777432, EPI_ISL_2777454, EPI_ISL_2777455, EPI_ISL_2777456, EPI_ISL_2777457, EPI_ISL_2777458, EPI_ISL_2777459, EPI_ISL_2777460, EPI_ISL_2777461, EPI_ISL_2777462, EPI_ISL_2777463, EPI_ISL_2777464, EPI_ISL_2777465, EPI_ISL_2777466, EPI_ISL_2777467, EPI_ISL_2777468, EPI_ISL_2777469, EPI_ISL_2777470, EPI_ISL_2777471, EPI_ISL_2777472, EPI_ISL_2777473, EPI_ISL_2777474, EPI_ISL_2777475, EPI_ISL_2777476, EPI_ISL_2777477, EPI_ISL_2777478, EPI_ISL_2777479, EPI_ISL_2777480, EPI_ISL_2777481, EPI_ISL_2777482, EPI_ISL_2777483, EPI_ISL_2777484, EPI_ISL_2777485, EPI_ISL_2777486, EPI_ISL_2777487, EPI_ISL_2777488, EPI_ISL_2777489, EPI_ISL_2777490, EPI_ISL_2777491, EPI_ISL_2777492, EPI_ISL_2777493, EPI_ISL_2777995, EPI_ISL_2777996, EPI_ISL_2777997, EPI_ISL_2777998, EPI_ISL_2777999, EPI_ISL_2778000, EPI_ISL_2778003, EPI_ISL_4030308, EPI_ISL_4030309, EPI_ISL_4030310, EPI_ISL_4030319, EPI_ISL_4030333, EPI_ISL_4030340 | | | |
| see above | Laboratorio de Ecologia de Doencas Transmissiveis na Amazonia, Instituto Leonidas e Maria Deane - Fiocruz Amazonia | Laboratorio de Ecologia de Doencas Transmissiveis na Amazonia, Instituto Leonidas e Maria Deane - Fiocruz Amazonia | Alex Martins; André Corado; Debora Duarte; Fabiola Mendonça da Silva Chui; Felipe Naveca; Felipe Naveca on behalf of the Fiocruz COVID-19 Genomic Surveillance Network; Fernanda Nascimento; Fernando Fonseca de Almeida Val; George Silva; Gisely Cardoso de Melo; Karina Pessoa; Luciana Gonçalves; Marcus Vinicius Guimarães de Lacerda; Maria Júlia Brandão; Maria Paula Gomes Mourão; Mariana Xavier Simão; Matilde Mejía; Michele Jesus; Valdinete Nascimento; Vanderson de Souza Sampaio; Victor Souza; Ágatha Costa |
| EPI_ISL_1629793 | Laboratorio de Genómica Microbiana, Universidad Peruana Cayetano Heredia | Laboratorio de Genómica Microbiana, Universidad Peruana Cayetano Heredia | Alejandra Dávila-Barclay; Diego Cuicapuza; Guillermo Salvatierra; Janet Huancachoque; Lenin Maturrano; Luis González; Pablo Tsukayama; Pedro E. Romero; Pool Marcos |
| EPI_ISL_3707386 | Laboratorio de Genómica Microbiana, Universidad Peruana Cayetano Heredia | cov0891 | Alejandra Dávila-Barclay; Diego Cuicapuza; Guillermo Salvatierra; Janet Huancachoque; Luis González; Pablo Tsukayama; Pedro E. Romero; Pool Marcos |
| EPI_ISL_3707389 | Laboratorio de Genómica Microbiana, Universidad Peruana Cayetano Heredia | cov0892 | Alejandra Dávila-Barclay; Diego Cuicapuza; Guillermo Salvatierra; Janet Huancachoque; Luis González; Pablo Tsukayama; Pedro E. Romero; Pool Marcos |
| EPI_ISL_3707422 | Laboratorio de Genómica Microbiana, Universidad Peruana Cayetano Heredia | cov0910 | Alejandra Dávila-Barclay; Diego Cuicapuza; Guillermo Salvatierra; Janet Huancachoque; Luis González; Pablo Tsukayama; Pedro E. Romero; Pool Marcos |
| EPI_ISL_3707425 | Laboratorio de Genómica Microbiana, Universidad Peruana Cayetano Heredia | cov0929 | Alejandra Dávila-Barclay; Diego Cuicapuza; Guillermo Salvatierra; Janet Huancachoque; Luis González; Pablo Tsukayama; Pedro E. Romero; Pool Marcos |
| EPI_ISL_2728552, EPI_ISL_2728600 | Laboratorio de Infectologia y Virologia Molecular | Laboratory of Molecular Virology, School of Medicine, Pontificia Universidad Catolica de Chile | Alejandro Bhrun; Ana Maria Contreras; Andres E. Munoz-Marcos; Carlos Palma; Catalina Pardo-Roa; Constanza Maldonado; Constanza Martinez-Valdevenito; Eileen Serrano; Erick Salinas; Estefany Poblete; Francisco Melo; Jennifer Angulo; Jorge Levican; Leonardo I. Almonacid; M. Belen Leyton; Magdalena Vera; Marcela Ferres; Maria Jose Avendano; Rafael A. Medina; Tamara Garcia-Salum |
| EPI_ISL_2098938 | Laboratorio de Microbiología, Hospital Marina Baixa, Villajoyosa | SeqCOVID-SPAIN consortium/IBV(CSIC) | Bárbara Gomez Alonso; Carmen Martínez Peinado and SeqCOVID-SPAIN consortium; Francisco José Arjona Zaragozí; Mónica Parra Grande |
| EPI_ISL_2008941, EPI_ISL_2008946, EPI_ISL_2008947, EPI_ISL_2008949, EPI_ISL_2008950, EPI_ISL_2008952, EPI_ISL_2008953, EPI_ISL_2008954, EPI_ISL_2008955, EPI_ISL_2008957, EPI_ISL_2008958, EPI_ISL_2008959, EPI_ISL_2008960, EPI_ISL_2008963, EPI_ISL_2008964, EPI_ISL_2008967, EPI_ISL_2544864, EPI_ISL_3048954, EPI_ISL_3048956, EPI_ISL_3048957, EPI_ISL_3048958, EPI_ISL_3761750, EPI_ISL_3761751 | | | |
| see above | Laboratorio de Pesquisa em Virologia, FAMERP, SJRP | Laboratorio de Pesquisa em Virologia, FAMERP, SJRP | Beatriz de Carvalho Marques; Cecília Artico Banho; Cíntia Bittar; Fábio Sossai Possebon; Guilherme Campos; Helena Lage Ferreira; Jorge A. Petroli Marchesi; João Pessoa Araújo Jr.; Leila Sabrina Ullmann; Lívia Sacchetto; Maisa C. Pereira Parra; Marília Moraes; Maurício L. Nogueira.; Paula Rahal; Paulo Inacio da Costa |
| EPI_ISL_943570 | Laboratorio de Referencia Nacional de Virus Respiratorio. Instituto Nacional de Salud Perú | Laboratorio de Referencia Nacional de Biotecnología y Biología Molecular. Instituto Nacional de Salud Perú | Carlos Padilla Rojas; Henri Bailon Calderon; Johanna Balbuena Torrez; Karolyn Vega Chozo; Luis Barcena; Marco Galarza Perez; Maribel Huaringa Nuñez; Nancy Rojas Serrano; Omar Caceres Rey; Priscila Lope Pari |
| EPI_ISL_1111142, EPI_ISL_1111143, EPI_ISL_1111151, EPI_ISL_1111152, EPI_ISL_1111281, EPI_ISL_1111461, EPI_ISL_1111469, EPI_ISL_1111472, EPI_ISL_1111474, EPI_ISL_1111483, EPI_ISL_1111484, EPI_ISL_1111486, EPI_ISL_1111490, EPI_ISL_1111496 | | | |
| see above | Laboratorio de Referencia Nacional de Virus Respiratorio. Instituto Nacional de Salud Perú | Laboratorio de Referencia Nacional de Enteropatógenos. Instituto Nacional de Salud del Perú | Fiorella Orellana Peralta; Iris Silva Molina; Junior Caro Castro; Ronnie Gavilan Chavez; Veronica Hurtado Vela; Willi Quino Sifuentes |
| EPI_ISL_3401587, EPI_ISL_3401588, EPI_ISL_3401597, EPI_ISL_3401612 | Laboratorio de Referencia Nacional de Virus Respiratorios. Centro Nacional de Salud Publica. Instituto Nacional de Salud Peru. | Laboratorio de Referencia Nacional de Virus Respiratorios. Centro Nacional de Salud Publica. Instituto Nacional de Salud Peru. | Carlos Padilla Rojas; Henri Bailon Calderon; Iris Silva Molina; Joseph Huayra Niquen; Lely Solari Zerpa; Luis Barcena Flores; Marco Galarza Perez; Nancy Rojas Serrano; Nieves Sevilla Castañeda; Omar Caceres Rey; Orson Mestanza Millones; Princesa Medrano Alhuay; Priscila Lope Pari; Sandra Morales Ruiz; Sara Gordillo Vilchez; Steve Acedo Lazo; Veronica Hurtado Vela; Victor Jimenez Vasquez; Wendy Lizarraga Olivares |
| EPI_ISL_3023389, EPI_ISL_3023585, EPI_ISL_3375878, EPI_ISL_3375879, EPI_ISL_3375880, EPI_ISL_3375888, EPI_ISL_3375890, EPI_ISL_3375898, EPI_ISL_3375925, EPI_ISL_3375928, EPI_ISL_3375932, EPI_ISL_3375940, EPI_ISL_3375941, EPI_ISL_3375944, EPI_ISL_3375947, EPI_ISL_3375954, EPI_ISL_3375959, EPI_ISL_3375969, EPI_ISL_3375970, EPI_ISL_3375977, EPI_ISL_3375980, EPI_ISL_3375983, EPI_ISL_3375984, EPI_ISL_3375985, EPI_ISL_3375986, EPI_ISL_3375988, EPI_ISL_3375994, EPI_ISL_3375996, EPI_ISL_3376011, EPI_ISL_3376019, EPI_ISL_3376024, EPI_ISL_3376026, EPI_ISL_3376027, EPI_ISL_3376028, EPI_ISL_3376029, EPI_ISL_3376030, EPI_ISL_3376036, EPI_ISL_3376038, EPI_ISL_3376040, EPI_ISL_3376184, EPI_ISL_3376378, EPI_ISL_3376379, EPI_ISL_3376384, EPI_ISL_3376386, EPI_ISL_3376387, EPI_ISL_3376393, EPI_ISL_3376394, EPI_ISL_3376400, EPI_ISL_3376412, EPI_ISL_3376413 | | | |
| see above | Laboratorio de Referencial Nacional de Virus Respiratorios | Laboratorio de Referencial Nacional de Virus Respiratorios | Carlos Padilla Rojas; Henri Bailon Calderon; Iris Silva Molina; Joseph Huayra Niquen; Lely Solari Zerpa; Luis Barcena Flores; Marco Galarza Perez; Nancy Rojas Serrano; Omar Caceres Rey; Orson Mestanza Millones; Priscila Lope Pari; Sandra Morales Ruiz; Steve Acedo Lazo; Veronica Hurtado Vela |
| EPI_ISL_2157202, EPI_ISL_2157206, EPI_ISL_2157208, EPI_ISL_2157209 | Laboratorio de Salud Pública | Gencore - Universidad de los Andes | Alejandro Gomez; David Gonzalez; Gabriela Delgado; Johana Hernandez; Luisa Sacristan; Marcela Guevara; Silvia Restrepo |
| EPI_ISL_1626618, EPI_ISL_1626619 | Laboratorio de Salud Pública Bogota | Gencore - Universidad de los Andes | Alejandro Gomez; Ana Maria Palacio; David Gonzalez; Gabriela Delgado; Johana Hernandez; Luisa Sacristan; Marcela Guevara; Silvia Restrepo |
| EPI_ISL_2245177, EPI_ISL_2245178, EPI_ISL_2245180 | Laboratorio de Saude Publica Dr. Giovanni Cysneiros | Coordenação Geral de Laboratórios de Saúde Pública (CGLAB/DAEVS/SVS/MS) | Vagner Fonseca; et al. |
| EPI_ISL_1786560, EPI_ISL_1786561, EPI_ISL_1786562, EPI_ISL_1786564, EPI_ISL_1786565, EPI_ISL_1786567, EPI_ISL_1786568 | | | |
| see above | Laboratorio de Virologia Clinica do HCFMRP-USP | Hocentro de Ribeirao Preto - Laboratório de Biologia Molecular | Aparecida Yulie Yamamoto; Diego Villa Clé; Dimas Tadeu Covas; Elaine Vieira Santos; Evandra Strazza Rodrigues; Glauco de Carvalho Pereira; Joilson Xavier; Josiane Serrano Borges; Luiz Carlos Junior Alcantara; Mariane Evaristo; Marta Giovanetti; Rafael dos Santos Bezerra; Rodrigo Tocantins Calado; Simone Kashima; Svetoslav Nanev Slavov; Talita Adelino; Vagner Fonseca |
| EPI_ISL_1406598, EPI_ISL_1406689, EPI_ISL_1673278, EPI_ISL_1673282 | Laboratorio de Virología HUCA | Laboratorio de Virología HUCA | Abreu F; Alvarez-Arguelles ME; Boga JA; Castelló C; Costales I; Coto E; Gómez de Oña J; Martín-Rodríguez G; Melón S; Perez-Martínez Z; Rojo S; Sandoval M |
| EPI_ISL_2007527, EPI_ISL_2007528, EPI_ISL_2007529, EPI_ISL_2007531, EPI_ISL_2007533, EPI_ISL_2007549 | Laboratorio de Virología del Hospital de Niños Dr. Ricardo Gutierrez | Área de Secuenciación del Laboratorio de Virología del Hospital de Niños Dr. Ricardo Gutierrez on behalf of 'Proyecto Argentino Interinstitucional de genomica de SARS-CoV-2' (PAIS Consortium) | A; Acevedo; Acuña; Alexay; Alvarez Lopez; Barreda Frank; C; D; E; G; Goya; Grandis; Jacques; LE; Labarta; Lusso; M; ME; MI; Medina; Mistchenko; N; Nabaes Jodar; Natale; O; S; Streitenberger; Thomas; Valinotto; Viegas, M.; Villegas |
| EPI_ISL_2534043 | Laboratorio di Patologia Clinica, Ospedale San Paolo in | U.O. Igiene, Ospedale Policlinico San Martino | Bruzzone Bianca; Caligiuri Patrizia; De Pace Vanessa; Domnich Alexander; Icardi Giancarlo; Lillo Flavia; Orsi Andrea; Ricucci Valentina |

| | | | |
|---|---|---|---|
| | Valloria, ASL 2 Liguria | | |
| EPI_ISL_1481062, EPI_ISL_1548203, EPI_ISL_1548311, EPI_ISL_1548748, EPI_ISL_1548779, EPI_ISL_1548989, EPI_ISL_1549466, EPI_ISL_1549546, EPI_ISL_1549554, EPI_ISL_1549568, EPI_ISL_1549701, EPI_ISL_1549748, EPI_ISL_1550146, EPI_ISL_1550147, EPI_ISL_1550151, EPI_ISL_1550152, EPI_ISL_1550153, EPI_ISL_1608860, EPI_ISL_1608903, EPI_ISL_1608940, EPI_ISL_1608987, EPI_ISL_1609020, EPI_ISL_1609051, EPI_ISL_1609575, EPI_ISL_1610022, EPI_ISL_1610024, EPI_ISL_1611144, EPI_ISL_1611146, EPI_ISL_1611147, EPI_ISL_1611149, EPI_ISL_1611159, EPI_ISL_1611170, EPI_ISL_1611192, EPI_ISL_1611247, EPI_ISL_1611337, EPI_ISL_1611497, EPI_ISL_1611502, EPI_ISL_1611503, EPI_ISL_1611515, EPI_ISL_1611682, EPI_ISL_1611708, EPI_ISL_1680522, EPI_ISL_1680810, EPI_ISL_1680962, EPI_ISL_1681026, EPI_ISL_1681217, EPI_ISL_1681522, EPI_ISL_1681537, EPI_ISL_1681585, EPI_ISL_1681853, EPI_ISL_1681914, EPI_ISL_1681921, EPI_ISL_1681924, EPI_ISL_1681925, EPI_ISL_1681959, EPI_ISL_1681962, EPI_ISL_1681974, EPI_ISL_1682001, EPI_ISL_1682103, EPI_ISL_1682104, EPI_ISL_1682144, EPI_ISL_1682150, EPI_ISL_1682154, EPI_ISL_1682169, EPI_ISL_1682237, EPI_ISL_1682268, EPI_ISL_1682275, EPI_ISL_1682277, EPI_ISL_1682405, EPI_ISL_1682418, EPI_ISL_1682438, EPI_ISL_1682484, EPI_ISL_1682512, EPI_ISL_1682646, EPI_ISL_1682658, EPI_ISL_1682675, EPI_ISL_1682684, EPI_ISL_1682714, EPI_ISL_1682720, EPI_ISL_1682724, EPI_ISL_1682730, EPI_ISL_1682732, EPI_ISL_1682778, EPI_ISL_1682987, EPI_ISL_1683151, EPI_ISL_1683216, EPI_ISL_1683257, EPI_ISL_1683269, EPI_ISL_1683306, EPI_ISL_1683314, EPI_ISL_1683357, EPI_ISL_1683359, EPI_ISL_1683411, EPI_ISL_1683447, EPI_ISL_1683474, EPI_ISL_1683480, EPI_ISL_1683521, EPI_ISL_1683536, EPI_ISL_1683904, EPI_ISL_1683990, EPI_ISL_1684230, EPI_ISL_1684427, EPI_ISL_1684856, EPI_ISL_1798359, EPI_ISL_1798959, EPI_ISL_1798962, EPI_ISL_1799614, EPI_ISL_1799622, EPI_ISL_1799678, EPI_ISL_1799705, EPI_ISL_1799706, EPI_ISL_1799707, EPI_ISL_1799714, EPI_ISL_1799719, EPI_ISL_1799724, EPI_ISL_1799730, EPI_ISL_1799731, EPI_ISL_1799732, EPI_ISL_1799831, EPI_ISL_1799836, EPI_ISL_1799925, EPI_ISL_1799926, EPI_ISL_1799950, EPI_ISL_1799963, EPI_ISL_1800003, EPI_ISL_1800020, EPI_ISL_1800053, EPI_ISL_1800056, EPI_ISL_1800057, EPI_ISL_1800109, EPI_ISL_1800116, EPI_ISL_1800119, EPI_ISL_1800129, EPI_ISL_1800135, EPI_ISL_1800136, EPI_ISL_1800137, EPI_ISL_1800249, EPI_ISL_1800273, EPI_ISL_1800274, EPI_ISL_1800291, EPI_ISL_1800372, EPI_ISL_1800374, EPI_ISL_1800383, EPI_ISL_1800390, EPI_ISL_1800394, EPI_ISL_1800432, EPI_ISL_1800550, EPI_ISL_1800560, EPI_ISL_1800568, EPI_ISL_1800579, EPI_ISL_1800585, EPI_ISL_1800588, EPI_ISL_1800597, EPI_ISL_1800598, EPI_ISL_1800599, EPI_ISL_1800600, EPI_ISL_1800602, EPI_ISL_1800811, EPI_ISL_1800973, EPI_ISL_1800974, EPI_ISL_1801014, EPI_ISL_1801142, EPI_ISL_1801281, EPI_ISL_1801294, EPI_ISL_1801440, EPI_ISL_1801639, EPI_ISL_1801642, EPI_ISL_1801648, EPI_ISL_1801676, EPI_ISL_1801682, EPI_ISL_1801684, EPI_ISL_1801686, EPI_ISL_1801687, EPI_ISL_1801700, EPI_ISL_1801701, EPI_ISL_1801810, EPI_ISL_1801811, EPI_ISL_1801815, EPI_ISL_1801832, EPI_ISL_1801842, EPI_ISL_1801871, EPI_ISL_1801898, EPI_ISL_1801952, EPI_ISL_1802005, EPI_ISL_1802051, EPI_ISL_1802066, EPI_ISL_1802068, EPI_ISL_1802069, EPI_ISL_1802073, EPI_ISL_1802080, EPI_ISL_1802095, EPI_ISL_1802147, EPI_ISL_1802153, EPI_ISL_1802154, EPI_ISL_1802157, EPI_ISL_1802159, EPI_ISL_1802199, EPI_ISL_1802201, EPI_ISL_1802210, EPI_ISL_1802216, EPI_ISL_1802223, EPI_ISL_1802285, EPI_ISL_1802334, EPI_ISL_1802377, EPI_ISL_1802380, EPI_ISL_1802382, EPI_ISL_1802384, EPI_ISL_1802385, EPI_ISL_1802401, EPI_ISL_1802463, EPI_ISL_1802469, EPI_ISL_1802640, EPI_ISL_1925718, EPI_ISL_1925724, EPI_ISL_1925735, EPI_ISL_1925739, EPI_ISL_1926172, EPI_ISL_1926188, EPI_ISL_1926238, EPI_ISL_1926281, EPI_ISL_1926294, EPI_ISL_1926298, EPI_ISL_1926336, EPI_ISL_1926411, EPI_ISL_1926649, EPI_ISL_2046443 | | | |
| see above | Laboratory Corporation of America | Centers for Disease Control and Prevention Division of Viral Diseases, Pathogen Discovery | Adrian Paskey; Amanda Douglas; Amanda Suchanek; Andrea Throop; Ayla Burns; Benjamin Rambo-Martin; Bobbi Croy; Brian Krueger; Brian Norvell; Christopher Gulvick; Christos Petropoulos; Clinton R. Paden; Craig Lukasik; Dakota Howard; Darlene Wagner; Debbie Boles; Dhwani Batra; Duncan MacCannell; Eyad Almasri; Goran Stevovic; Howard Engler; Hrushikesh Deshmukh; Jake Humphrey; Jana Schroth; Jason Caravas; Joe Voshell; John Pruitt; Jonathan Meltzer; Jonathan Williams; Kara Moser; Kimberly Wagner; Lax Iyer; Lyndon Tilson; Manoj Jain; Marcia Eisenberg; Mary Ann Cristobal; Mary Williamson; Matthew Schmerer; Michael Levandoski; Mike Sapeta; Mindy Nye; Minoo Agarwal; Mohan Kolli; Nuthawin Charoensri; Oren Cohen; Peter W. Cook; Prashant Gupta; Qian Zeng; Rama Ghatti; Scott Parker; Scott Ryan; Scott Sammons; Shatavia Morrison; Stanley Letovsky; Steven Ragan; Suresh Babu Selvaraju; Susan Countryman; Susan Hicks; Suzanne Dale; Thomas Urban; Tim Kuphal; Tricia Zwiefelhofer; Vincent Drouillon; Yvette Unoarumhi |
| EPI_ISL_2249499, EPI_ISL_2249500, EPI_ISL_2249515, EPI_ISL_2249516 | Laboratory Medicine | Department of Laboratory Medicine, Lin-Kou Chang Gung Memorial Hospital, Taoyuan, Taiwan | Cheng-Hsun Chiu; Cheng-Yang Chen?; |
| EPI_ISL_3982770 | Laboratory of Molecular Biology, Hemocentro de Ribeirão Preto | Laboratory of Molecular Biology, Hemocentro de Ribeirão Preto | Antonio Jorge Martins; Claudia Renata dos Santos Barros; David Schlesinger; Debora Botequio Moretti; Dimas Tadeu Covas; Elaine Cristina Marqueze; Elaine Vieira Santos; Evandra Strazza Rodrigues; Heidge Fukumasu; Jayme Augusto de Souza-Neto; José Salvatore Leister Patané; Luiz Alcantara; Luiz Lehmann Coutinho; Maria Carolina Elias; Maurício Lacerda Nogueira; Rafael dos Santos Bezerra; Raul Machado Neto; Rejane Maria Tommasini Grotto; Ricardo Haddad; Rodrigo Proto de Siqueira; Sandra Coccuzzo Sampaio Vessoni; Simone Kashima; Svetoslav Nanev Slavov; VV Cantarelli; Vincent Louis Viala |
| EPI_ISL_2614074, EPI_ISL_2614076, EPI_ISL_2614077, EPI_ISL_2614078, EPI_ISL_2614080, EPI_ISL_2614081, EPI_ISL_2614082, EPI_ISL_2614083, EPI_ISL_2614087, EPI_ISL_2614088, EPI_ISL_2614089, EPI_ISL_2614090, EPI_ISL_2614091 | | | |
| see above | Laboratory of Molecular Virology, Federal University of Rio de Janeito, UFRJ | Laboratory of Respiratory Viruses and Measles, Oswaldo Cruz Institute, FIOCRUZ | Alice Sampaio Rocha; Amilcar Tanuri; Ana Carolina Mendonca; Anna Carolina Paixao; Elisa Cavalcante Pereira; Fernando Motta; Luciana Appolinario; Marilda Siqueira on behalf of the Fiocruz COVID-19 Genomic Surveillance Network; Paola Resende; Renata Serrano Lopes; Taina Venas |
| EPI_ISL_1402431, EPI_ISL_1534011, EPI_ISL_1534014, EPI_ISL_1534015, EPI_ISL_2038961, EPI_ISL_2038962, EPI_ISL_2038963, EPI_ISL_2196185, EPI_ISL_2196186, EPI_ISL_2196187, EPI_ISL_2196190, EPI_ISL_2196191, EPI_ISL_2196192, EPI_ISL_2196193, EPI_ISL_2196194, EPI_ISL_2196195, EPI_ISL_2196196, EPI_ISL_2274097, EPI_ISL_2274100, EPI_ISL_2274101, EPI_ISL_2274102, EPI_ISL_2274103, EPI_ISL_2274104, EPI_ISL_2274105, EPI_ISL_2274106, EPI_ISL_2274107, EPI_ISL_2443581, EPI_ISL_2443582, EPI_ISL_2443583, EPI_ISL_2443584, EPI_ISL_2443585, EPI_ISL_2443586, EPI_ISL_2443587, EPI_ISL_2443588, EPI_ISL_2443589, EPI_ISL_2443590, EPI_ISL_2443591, EPI_ISL_2443592, EPI_ISL_2443593, EPI_ISL_2443594, EPI_ISL_2443596, EPI_ISL_2443597, EPI_ISL_2443598, EPI_ISL_2536271, EPI_ISL_2536272, EPI_ISL_2536273, EPI_ISL_2536274, EPI_ISL_2557386, EPI_ISL_2557389, EPI_ISL_2557390, EPI_ISL_2557392, EPI_ISL_2557393, EPI_ISL_2614191, EPI_ISL_2614196, EPI_ISL_2614197, EPI_ISL_2614210, EPI_ISL_2614212, EPI_ISL_2614214, EPI_ISL_2614218, EPI_ISL_2614220, EPI_ISL_2614221, EPI_ISL_2614222, EPI_ISL_2614223, EPI_ISL_2614224, EPI_ISL_2614225, EPI_ISL_2614228, EPI_ISL_2614229, EPI_ISL_2614231, EPI_ISL_2614232, EPI_ISL_2614233, EPI_ISL_2614234, EPI_ISL_2614237, EPI_ISL_2614239, EPI_ISL_2614240, EPI_ISL_2614241, EPI_ISL_2614242, EPI_ISL_2614243, EPI_ISL_2614244, EPI_ISL_2614245, EPI_ISL_2614246, EPI_ISL_2614247, EPI_ISL_2614248, EPI_ISL_2614249, EPI_ISL_2614250, EPI_ISL_2614251, EPI_ISL_2614252, EPI_ISL_2614254, EPI_ISL_2614255, EPI_ISL_2614256, EPI_ISL_2614257, EPI_ISL_2614258, EPI_ISL_2614259, EPI_ISL_2614260, EPI_ISL_2614261, EPI_ISL_2614262, EPI_ISL_2614263, EPI_ISL_2614264, EPI_ISL_2614265, EPI_ISL_2614266, EPI_ISL_2614267, EPI_ISL_2614268, EPI_ISL_2614269, EPI_ISL_2614270, EPI_ISL_2614272, EPI_ISL_2614273, EPI_ISL_2614274, EPI_ISL_2614276, EPI_ISL_2614277, EPI_ISL_2614279, EPI_ISL_2614280, EPI_ISL_2614281, EPI_ISL_2614282, EPI_ISL_2614283, EPI_ISL_2614284, EPI_ISL_2614286, EPI_ISL_2614288, EPI_ISL_2614289, EPI_ISL_2614290, EPI_ISL_2614291, EPI_ISL_2614292, EPI_ISL_2614294, EPI_ISL_2614295, EPI_ISL_2614297, EPI_ISL_2614298, EPI_ISL_2614300, EPI_ISL_2614301, EPI_ISL_2614302, EPI_ISL_2614303, EPI_ISL_2614304, EPI_ISL_2614305, EPI_ISL_2614306, EPI_ISL_2614307, EPI_ISL_2614308, EPI_ISL_2614309, EPI_ISL_2614312, EPI_ISL_2614313, EPI_ISL_2614314, EPI_ISL_2614316, EPI_ISL_2614317, EPI_ISL_2614318, EPI_ISL_2614320, EPI_ISL_2614321, EPI_ISL_2614322, EPI_ISL_2614323, EPI_ISL_2614324, EPI_ISL_2614325, EPI_ISL_2614326, EPI_ISL_2614327, EPI_ISL_2614328, EPI_ISL_2614329, EPI_ISL_2614330, EPI_ISL_2614332, EPI_ISL_2614334, EPI_ISL_2614335, EPI_ISL_2614336, EPI_ISL_2614337, EPI_ISL_2614338, EPI_ISL_2614339, EPI_ISL_2614340, EPI_ISL_2614341, EPI_ISL_2614342, EPI_ISL_2614343, EPI_ISL_2614344, EPI_ISL_2614345, EPI_ISL_2614346, EPI_ISL_2614347, EPI_ISL_2614348, EPI_ISL_2614349, EPI_ISL_2614350, EPI_ISL_2677175, EPI_ISL_2677176, EPI_ISL_2677178, EPI_ISL_2677179, EPI_ISL_2677180, EPI_ISL_2677181, EPI_ISL_2677182, EPI_ISL_2677183, EPI_ISL_2677184, EPI_ISL_2677185, EPI_ISL_2677186, EPI_ISL_2677187, EPI_ISL_2677188, EPI_ISL_2677189, EPI_ISL_2677191, EPI_ISL_2677192, EPI_ISL_2677193, EPI_ISL_2677194, EPI_ISL_2677195, EPI_ISL_2677196, EPI_ISL_2677197, EPI_ISL_2677198, EPI_ISL_2677199, EPI_ISL_2677200, EPI_ISL_2677201, EPI_ISL_2677202, EPI_ISL_2677203, EPI_ISL_2677204, EPI_ISL_2677205, EPI_ISL_2677206, EPI_ISL_2677207, EPI_ISL_2677208, EPI_ISL_2677209, EPI_ISL_2677210, EPI_ISL_2677211, EPI_ISL_2677212, EPI_ISL_2677213, EPI_ISL_2677214, EPI_ISL_2677215, EPI_ISL_2677216, EPI_ISL_3190275, EPI_ISL_3190276, EPI_ISL_3190279, EPI_ISL_3832398, EPI_ISL_3832399 | | | |
| see above | Laboratory of Respiratory Viruses and Measles, Oswaldo Cruz Institute, FIOCRUZ | Laboratory of Respiratory Viruses and Measles, Oswaldo Cruz Institute, FIOCRUZ | Agatha Cristinne Prudencio; Agatha Soares; Alex Pauvolid-Correa; Alice Sampaio Rocha; Ana Beatriz Machado Lima; Ana Carolina Mendonca; Anna Carolina Paixao; Elisa Cavalcante Pereira; Felipe Naveca; Fernando Motta; Ighor Arantes; Ighor Leonardo Arantes Gomes; Luciana Appolinario; Marilda Siqueira on behalf of the Fiocruz COVID-19 Genomic Surveillance Network; Mia Ferreira Araujo; Paola Resende; Patricia Brasil; Renata Serrano Lopes; Taina Venas |
| EPI_ISL_1289960 | Laboratory of Virology, Ribeirão Preto General Hospital, Ribeirão Preto Medical School, University of São Paulo | Laboratory of Oncology, Blood Center of Ribeirão Preto, Ribeirão Preto School of Medicine, University of São Paulo | A.L.P.; A.Y.; B.A.L.; BELLISSIMO-RODRIGUES, F.; CAMPOS; COLLI; FONSECA; L.M.; MR; SANTOS; YAMAMOTO |
| EPI_ISL_2157532, EPI_ISL_2157533, EPI_ISL_2157534, EPI_ISL_2157535 | Laboratório Central de Saude Publica do Estado da Bahia (LACEN/BA) | Laboratory of Respiratory Viruses and Measles, Oswaldo Cruz Institute, FIOCRUZ | Alice Sampaio Rocha; Ana Carolina Mendonca; Anna Carolina Paixao; Elisa Cavalcante Pereira; Felicidade Pereira; Fernando Motta; Luciana Appolinario; Marilda Siqueira on behalf of the Fiocruz COVID-19 Genomic Surveillance Network; Paola Resende; Renata Serrano Lopes; Taina Venas |
| EPI_ISL_2157377, EPI_ISL_2157390, EPI_ISL_2157391, EPI_ISL_2157392, EPI_ISL_2157393, EPI_ISL_2157398, EPI_ISL_2157400, EPI_ISL_2157405, EPI_ISL_2157406, EPI_ISL_2157409, EPI_ISL_2157410, EPI_ISL_2157411, EPI_ISL_2157418, EPI_ISL_2157448, EPI_ISL_2157450, EPI_ISL_2157484, EPI_ISL_2157493, EPI_ISL_2157494, EPI_ISL_2157495, EPI_ISL_2157498, EPI_ISL_2157499, EPI_ISL_2157500, EPI_ISL_2157501, EPI_ISL_2157503, EPI_ISL_2157510, EPI_ISL_2157511, EPI_ISL_2157513, EPI_ISL_2157515, EPI_ISL_2157547, EPI_ISL_2157548, EPI_ISL_2274133, EPI_ISL_2274134, EPI_ISL_2274135, EPI_ISL_2274136 | | | |
| see above | Laboratório Central de Saude Publica do Estado de Santa Catarina (LACEN/SC) | Laboratory of Respiratory Viruses and Measles, Oswaldo Cruz Institute, FIOCRUZ | Alice Sampaio Rocha; Ana Carolina Mendonca; Anna Carolina Paixao; Darcita Buerger Rovaris; Elisa Cavalcante Pereira; Fernando Motta; Luciana Appolinario; Marilda Siqueira on behalf of the Fiocruz COVID-19 Genomic Surveillance Network; Paola Resende; Renata Serrano Lopes; Sandra Bianchini Fernandes; Taina Venas |
| EPI_ISL_2157506, EPI_ISL_2157516, EPI_ISL_2157517 | Laboratório Central de Saude Publica do Estado do Paraná (LACEN/PR) | Laboratory of Respiratory Viruses and Measles, Oswaldo Cruz Institute, FIOCRUZ | Alice Sampaio Rocha; Ana Carolina Mendonca; Anna Carolina Paixao; Elisa Cavalcante Pereira; Fernando Motta; Irina Nastassja Riediger; Luciana Appolinario; Maria do Carmo Debur; Marilda Siqueira on behalf of the Fiocruz COVID-19 Genomic Surveillance Network; Paola Resende; Renata Serrano Lopes; Taina Venas |
| EPI_ISL_2157525, EPI_ISL_2157526, EPI_ISL_2157528, EPI_ISL_2157529, EPI_ISL_2157530, EPI_ISL_2157536, EPI_ISL_2157537, EPI_ISL_2157538, EPI_ISL_2157539, EPI_ISL_2157549 | | | |
| see above | Laboratório Central de Saude Publica do Estado do Rio de Janeiro (LACEN/RJ) | Laboratory of Respiratory Viruses and Measles, Oswaldo Cruz Institute, FIOCRUZ | Alice Sampaio Rocha; Ana Carolina Mendonca; Andrea Cony Cavalcanti; Anna Carolina Paixao; Elisa Cavalcante Pereira; Fernando Motta; Luciana Appolinario; Marilda Siqueira on behalf of the Fiocruz COVID-19 Genomic Surveillance Network; Paola Resende; Renata Serrano Lopes; Taina Venas |
| EPI_ISL_1239111 | Laboratório Central de Saúde Pública Noel Nutels | Coordenação Geral de Laboratórios de Saúde Pública (CGLAB) | ; Vagner Fonseca et al |
| EPI_ISL_2241496, EPI_ISL_2241559, EPI_ISL_2241576, EPI_ISL_2241586, EPI_ISL_2241588, EPI_ISL_2246285 | Laboratório Central de Saúde Pública da Paraiba | Coordenação Geral de Laboratórios de Saúde Pública (CGLAB/DAEVS/SVS/MS) | Vagner Fonseca; et al. |
| EPI_ISL_2308400, EPI_ISL_2308401, EPI_ISL_2308421, EPI_ISL_2308424, EPI_ISL_2308463 | Laboratório Central de Saúde Pública de Alagoas | Coordenação Geral de Laboratórios de Saúde Pública (CGLAB/DAEVS/SVS/MS) | Vagner Fonseca; et al. |
| EPI_ISL_2241532, EPI_ISL_2241547, EPI_ISL_2241605, EPI_ISL_2308416, EPI_ISL_2308438, EPI_ISL_2308446 | Laboratório Central de Saúde Pública de Pernambuco | Coordenação Geral de Laboratórios de Saúde Pública (CGLAB/DAEVS/SVS/MS) | Vagner Fonseca; et al. |
| EPI_ISL_2245107, EPI_ISL_2245108, EPI_ISL_2245109, EPI_ISL_2245110, EPI_ISL_2245111, EPI_ISL_2245112, EPI_ISL_2245113, EPI_ISL_2298762, EPI_ISL_2298764, EPI_ISL_2298765, EPI_ISL_2298769, EPI_ISL_2298788, EPI_ISL_2298847, EPI_ISL_2298849 | | | |
| see above | Laboratório Central de Saúde Pública de Roraima | Coordenação Geral de Laboratórios de Saúde Pública (CGLAB/DAEVS/SVS/MS) | Vagner Fonseca; et al. |
| EPI_ISL_2249392, EPI_ISL_2249393, EPI_ISL_2249394, EPI_ISL_2249395, EPI_ISL_2249398, EPI_ISL_2249401, EPI_ISL_2249402, EPI_ISL_2249403, EPI_ISL_2292998, EPI_ISL_2292999, EPI_ISL_2293000, EPI_ISL_2293001, EPI_ISL_2293002, EPI_ISL_2293003, EPI_ISL_2293004, EPI_ISL_2293005, EPI_ISL_2293006 | | | |
| see above | Laboratório Central de Saúde Pública de | Coordenação Geral de Laboratórios de Saúde | Vagner Fonseca; et al. |

| | | | |
|---|---|---|---|
| | Santa Catarina | Pública (CGLAB/DAEVS/SVS/MS) | |
| EPI_ISL_2241541, EPI_ISL_2241555, EPI_ISL_2241578, EPI_ISL_2241587, EPI_ISL_2241602, EPI_ISL_2241604, EPI_ISL_2246286, EPI_ISL_2308402, EPI_ISL_2308403, EPI_ISL_2308411, EPI_ISL_2308413, EPI_ISL_2308414, EPI_ISL_2308415, EPI_ISL_2308417, EPI_ISL_2308437, EPI_ISL_2308454, EPI_ISL_2308457, EPI_ISL_2308465, EPI_ISL_2308466, EPI_ISL_2308467, EPI_ISL_2308470, EPI_ISL_2308474, EPI_ISL_2308476, EPI_ISL_2308477 | | | |
| see above | Laboratório Central de Saúde Pública de Sergipe | Coordenação Geral de Laboratórios de Saúde Pública (CGLAB/DAEVS/SVS/MS) | Vagner Fonseca; et al. |
| EPI_ISL_2245062, EPI_ISL_2248781 | Laboratório Central de Saúde Pública do Acre | Coordenação Geral de Laboratórios de Saúde Pública (CGLAB/DAEVS/SVS/MS) | Vagner Fonseca; et al. |
| EPI_ISL_2298795, EPI_ISL_2298801, EPI_ISL_2298817, EPI_ISL_2298819, EPI_ISL_2298821, EPI_ISL_2298824, EPI_ISL_2298828, EPI_ISL_2298830, EPI_ISL_2298832 | | | |
| see above | Laboratório Central de Saúde Pública do Amapá | Coordenação Geral de Laboratórios de Saúde Pública (CGLAB/DAEVS/SVS/MS) | Vagner Fonseca; et al. |
| EPI_ISL_2298850, EPI_ISL_2298852, EPI_ISL_2298854, EPI_ISL_2298856, EPI_ISL_2298859, EPI_ISL_2298861, EPI_ISL_2298863, EPI_ISL_2298867, EPI_ISL_2298871 | | | |
| see above | Laboratório Central de Saúde Pública do Amazonas | Coordenação Geral de Laboratórios de Saúde Pública (CGLAB/DAEVS/SVS/MS) | Vagner Fonseca; et al. |
| EPI_ISL_2777235, EPI_ISL_2777236, EPI_ISL_2777237, EPI_ISL_2777238, EPI_ISL_2777239, EPI_ISL_2777240, EPI_ISL_2777241, EPI_ISL_2777247, EPI_ISL_2777248, EPI_ISL_2777249, EPI_ISL_2777250, EPI_ISL_2777251, EPI_ISL_2777252, EPI_ISL_2777253, EPI_ISL_2777254, EPI_ISL_2777255, EPI_ISL_2777256, EPI_ISL_2777257, EPI_ISL_2777258, EPI_ISL_2777259, EPI_ISL_2777260, EPI_ISL_2777261, EPI_ISL_2777262, EPI_ISL_2777263, EPI_ISL_2777264, EPI_ISL_2777265, EPI_ISL_2777266, EPI_ISL_2777267, EPI_ISL_2777268, EPI_ISL_2777269, EPI_ISL_2777270, EPI_ISL_2777271, EPI_ISL_2777272, EPI_ISL_2777273, EPI_ISL_2777274, EPI_ISL_2777275, EPI_ISL_2777276, EPI_ISL_2777277, EPI_ISL_2777278, EPI_ISL_2777279, EPI_ISL_2777290, EPI_ISL_2777301, EPI_ISL_2777302, EPI_ISL_2777309, EPI_ISL_2777311, EPI_ISL_2777320, EPI_ISL_2777321, EPI_ISL_2777324, EPI_ISL_2777327, EPI_ISL_2777328, EPI_ISL_2777330, EPI_ISL_2777331, EPI_ISL_2777333, EPI_ISL_2777340, EPI_ISL_2777356, EPI_ISL_2777357, EPI_ISL_2777358, EPI_ISL_2777495, EPI_ISL_2777496, EPI_ISL_2777498, EPI_ISL_2777499, EPI_ISL_2777501, EPI_ISL_2777502, EPI_ISL_2777503, EPI_ISL_2777504, EPI_ISL_2777505, EPI_ISL_2777506, EPI_ISL_2777507, EPI_ISL_2777508, EPI_ISL_2777509, EPI_ISL_2777510, EPI_ISL_2777516, EPI_ISL_2777518, EPI_ISL_2777519, EPI_ISL_2777520, EPI_ISL_2777521, EPI_ISL_2777522, EPI_ISL_2777525, EPI_ISL_2777526, EPI_ISL_2777528, EPI_ISL_2777533, EPI_ISL_2777534, EPI_ISL_2777535, EPI_ISL_2777536, EPI_ISL_2777537, EPI_ISL_2777538, EPI_ISL_2777539, EPI_ISL_2777540, EPI_ISL_2777541, EPI_ISL_2777542, EPI_ISL_2777543, EPI_ISL_2777544, EPI_ISL_2777549, EPI_ISL_2777551, EPI_ISL_2777552, EPI_ISL_2777554, EPI_ISL_2777557, EPI_ISL_2777558, EPI_ISL_2777560, EPI_ISL_2777563, EPI_ISL_2777565, EPI_ISL_2777566, EPI_ISL_2777567, EPI_ISL_2777569, EPI_ISL_2777571, EPI_ISL_2777572, EPI_ISL_2777574, EPI_ISL_2777576, EPI_ISL_2777581, EPI_ISL_2777588, EPI_ISL_2777589, EPI_ISL_2777599, EPI_ISL_2777609, EPI_ISL_2777615, EPI_ISL_2777618, EPI_ISL_2777620, EPI_ISL_2777621, EPI_ISL_2777638, EPI_ISL_2777646, EPI_ISL_2777655, EPI_ISL_2777657, EPI_ISL_2777662, EPI_ISL_2777668, EPI_ISL_2777670, EPI_ISL_2777676, EPI_ISL_2777691, EPI_ISL_2777697, EPI_ISL_2777698, EPI_ISL_2777699, EPI_ISL_2777700, EPI_ISL_2777701, EPI_ISL_2777704, EPI_ISL_2777705, EPI_ISL_2777706, EPI_ISL_2777707, EPI_ISL_2777708, EPI_ISL_2777709, EPI_ISL_2777710, EPI_ISL_2777713, EPI_ISL_2777714, EPI_ISL_2777715, EPI_ISL_2777716, EPI_ISL_2777717, EPI_ISL_2777718, EPI_ISL_2777719, EPI_ISL_2777720, EPI_ISL_2777721, EPI_ISL_2777722, EPI_ISL_2777723, EPI_ISL_2777724, EPI_ISL_2777725, EPI_ISL_2777726, EPI_ISL_2777727, EPI_ISL_2777728, EPI_ISL_2777729, EPI_ISL_2777730, EPI_ISL_2777731, EPI_ISL_2777732, EPI_ISL_2777733, EPI_ISL_2777734, EPI_ISL_2777735, EPI_ISL_2777736, EPI_ISL_2777737, EPI_ISL_2777738, EPI_ISL_2777739, EPI_ISL_2777740, EPI_ISL_2777741, EPI_ISL_2777742, EPI_ISL_2777743, EPI_ISL_2777744, EPI_ISL_2777745, EPI_ISL_2777746, EPI_ISL_2777747, EPI_ISL_2777748, EPI_ISL_2777749, EPI_ISL_2777750, EPI_ISL_2777751, EPI_ISL_2777752, EPI_ISL_2777754, EPI_ISL_2777755, EPI_ISL_2777758, EPI_ISL_2777759, EPI_ISL_2777762, EPI_ISL_2777763, EPI_ISL_2777766, EPI_ISL_2777767, EPI_ISL_2777768, EPI_ISL_2777769, EPI_ISL_2777771, EPI_ISL_2777772, EPI_ISL_2777775, EPI_ISL_2777776, EPI_ISL_2777779, EPI_ISL_2777780, EPI_ISL_2777781, EPI_ISL_2777782, EPI_ISL_2777783, EPI_ISL_2777784, EPI_ISL_2777785, EPI_ISL_2777786, EPI_ISL_2777787, EPI_ISL_2777788, EPI_ISL_2777789, EPI_ISL_2777790, EPI_ISL_2777791, EPI_ISL_2777792, EPI_ISL_2777793, EPI_ISL_2777794, EPI_ISL_2777795, EPI_ISL_2777796, EPI_ISL_2777797, EPI_ISL_2777798, EPI_ISL_2777799, EPI_ISL_2777800, EPI_ISL_2777801, EPI_ISL_2777802, EPI_ISL_2777803, EPI_ISL_2777804, EPI_ISL_2777805, EPI_ISL_2777807, EPI_ISL_2777808, EPI_ISL_2777811, EPI_ISL_2777812, EPI_ISL_2777815, EPI_ISL_2777816, EPI_ISL_2777817, EPI_ISL_2777818, EPI_ISL_2777819, EPI_ISL_2777820, EPI_ISL_2777821, EPI_ISL_2777823, EPI_ISL_2777827, EPI_ISL_2777828, EPI_ISL_2777829, EPI_ISL_2777830, EPI_ISL_2777838, EPI_ISL_2777840, EPI_ISL_2777841, EPI_ISL_2777842, EPI_ISL_2777843, EPI_ISL_2777847, EPI_ISL_2777848, EPI_ISL_2777851, EPI_ISL_2777855, EPI_ISL_2777857, EPI_ISL_2777858, EPI_ISL_2777859, EPI_ISL_2777861, EPI_ISL_2777862, EPI_ISL_2777864, EPI_ISL_2777867, EPI_ISL_2777868, EPI_ISL_2777870, EPI_ISL_2777871, EPI_ISL_2777872, EPI_ISL_2777873, EPI_ISL_2777874, EPI_ISL_2777894, EPI_ISL_2777899, EPI_ISL_2777986, EPI_ISL_2777987, EPI_ISL_2777988, EPI_ISL_2777992, EPI_ISL_2777993, EPI_ISL_2778002, EPI_ISL_2778004, EPI_ISL_2778005 | | | |
| see above | Laboratório Central de Saúde Pública do Amazonas - LACEN-AM | Laboratorio de Ecologia de Doencas Transmissiveis na Amazonia, Instituto Leonidas e Maria Deane - Fiocruz Amazonia | André Corado; Debora Duarte; Felipe Naveca; Fernanda Nascimento; George Silva; Karina Pessoa; Luciana Gonçalves; Maria Júlia Brandão; Matilde Mejía; Michele Jesus; Valdinete Nascimento; Victor Souza; Ágatha Costa |
| EPI_ISL_2245080, EPI_ISL_2245082, EPI_ISL_2248788, EPI_ISL_2298779, EPI_ISL_2298781 | Laboratório Central de Saúde Pública do Ceará | Coordenação Geral de Laboratórios de Saúde Pública (CGLAB/DAEVS/SVS/MS) | Vagner Fonseca; et al. |
| EPI_ISL_1239119 | Laboratório Central de Saúde Pública do Espírito Santo | Coordenação Geral de Laboratórios de Saúde Pública (CGLAB) | ; Vagner Fonseca et al |
| EPI_ISL_2293011 | Laboratório Central de Saúde Pública do Espírito Santo | Coordenação Geral de Laboratórios de Saúde Pública (CGLAB/DAEVS/SVS/MS) | Vagner Fonseca; et al. |
| EPI_ISL_1239012 | Laboratório Central de Saúde Pública do Estado de Pernambuco (LACEN-PE) | WallauLab, Aggeu Magalhaes Institute | Alexandre Freitas da Silva; Antonio Mauro Rezende; Constância Flávia Junqueira Ayres; Cássia Docena; Duschinka Ribeiro Duarte Guedes; Elisama Helvecio; Filipe Zimmer Dezordi; Gabriel Luz Wallau on behalf of the Fiocruz COVID-19 Genomic Surveillance Network; Larissa Krokovsky; Laís Ceschini Machado; Marcelo Henrique dos Santos Paiva; Matheus Filgueira Bezerra; Sinval Pinto Brandão Filho |
| EPI_ISL_2298752, EPI_ISL_2298754, EPI_ISL_2298755, EPI_ISL_2298770, EPI_ISL_2298834, EPI_ISL_2298839, EPI_ISL_2298840, EPI_ISL_2298842, EPI_ISL_2298844, EPI_ISL_2298845 | | | |
| see above | Laboratório Central de Saúde Pública do Maranhão | Coordenação Geral de Laboratórios de Saúde Pública (CGLAB/DAEVS/SVS/MS) | Vagner Fonseca; et al. |
| EPI_ISL_2249450, EPI_ISL_2249454, EPI_ISL_2249455 | Laboratório Central de Saúde Pública do Paraá | Coordenação Geral de Laboratórios de Saúde Pública (CGLAB/DAEVS/SVS/MS) | Vagner Fonseca; et al. |
| EPI_ISL_2245089, EPI_ISL_2248763, EPI_ISL_2248766, EPI_ISL_2248782, EPI_ISL_2248783, EPI_ISL_2298758, EPI_ISL_2298783, EPI_ISL_2298787, EPI_ISL_2298810, EPI_ISL_2298812, EPI_ISL_2298814 | | | |
| see above | Laboratório Central de Saúde Pública do Pará | Coordenação Geral de Laboratórios de Saúde Pública (CGLAB/DAEVS/SVS/MS) | Vagner Fonseca; et al. |
| EPI_ISL_2241500, EPI_ISL_2241554, EPI_ISL_2241575 | Laboratório Central de Saúde Pública do Piaui | Coordenação Geral de Laboratórios de Saúde Pública (CGLAB/DAEVS/SVS/MS) | Vagner Fonseca; et al. |
| EPI_ISL_2241503, EPI_ISL_2241507, EPI_ISL_2241545, EPI_ISL_2241549, EPI_ISL_2241550, EPI_ISL_2241608, EPI_ISL_2308399, EPI_ISL_2308404, EPI_ISL_2308407, EPI_ISL_2308464 | | | |
| see above | Laboratório Central de Saúde Pública do Rio Grande do Norte | Coordenação Geral de Laboratórios de Saúde Pública (CGLAB/DAEVS/SVS/MS) | Vagner Fonseca; et al. |
| EPI_ISL_2249358, EPI_ISL_2249366, EPI_ISL_2249368, EPI_ISL_2249369, EPI_ISL_2249370, EPI_ISL_2249371, EPI_ISL_2249373, EPI_ISL_2249378, EPI_ISL_2249385 | | | |
| see above | Laboratório Central de Saúde Pública do Rio Grande do Sul | Coordenação Geral de Laboratórios de Saúde Pública (CGLAB/DAEVS/SVS/MS) | Vagner Fonseca; et al. |
| EPI_ISL_2249429, EPI_ISL_2249440, EPI_ISL_2249444 | Laboratório Central de Saúde Pública do Rio de Janeiro | Coordenação Geral de Laboratórios de Saúde Pública (CGLAB/DAEVS/SVS/MS) | Vagner Fonseca; et al. |
| EPI_ISL_1213171 | Laboratório HLA/UERJ | Bioinformatics Laboratory / LNCC | Alessandra P Lamarca; Alexandra L Gerber; Ana Paula Melo Mariano; Ana Paula de C Guimarães; Ana Tereza R Vasconcelos; Angela Maria Guimarães Santos; Bianca Mendes Maciel; Danielle Angst Secco; Eduardo Sérgio Soares Sousa; Eloiza Helena Campana; Francisco Paulo Freire Neto; George Rego Albuquerque; Kátia Castanho Scortecci; Lucymara Fassarella Agnez Lima; Luiz G P de Almeida; Luís Cristóvão Porto; Otavio J. Brustolini; Paulo Ricardo Nascimento; Ronaldo da Silva Francisco Jr; Sandra Rocha Gadelha; Selma Maria Bezerra Jeronimo; Vinícius Pietta Perez |
| EPI_ISL_3031297, EPI_ISL_3031298, EPI_ISL_3031299, EPI_ISL_3031300, EPI_ISL_3031301, EPI_ISL_3031302, EPI_ISL_3031303, EPI_ISL_3031304, EPI_ISL_3031305, EPI_ISL_3031306, EPI_ISL_3031307, EPI_ISL_3031308, EPI_ISL_3031309, EPI_ISL_3061854 | | | |
| see above | Laboratório Municipal de Biologia Molecular | Instituto René Rachou / Fiocruz Minas | André Menezes; Anna Salim; Eneida Oliveira; Gabriel Fernandes; Pedro Alves; Rubens do Monte Neto; Thaís Silva |
| EPI_ISL_1261691 | Laboratório Paulo C. | Evandro Chagas | A.M.; Barbagelata; E.C.; E.M.A.; Ferreira; J.A.; Junior; K.C.; L.C.; L.S.; M.C.; P.S.; Pinheiro; Santos; Silva; Sousa; Sousa Junior; W.D.C.; da Silva |

| | | | |
|---|---|---|---|
| | Azevedo | Institute | |
| EPI_ISL_1494977, EPI_ISL_1494986, EPI_ISL_1494994, EPI_ISL_1494998, EPI_ISL_1495005, EPI_ISL_1495010, EPI_ISL_1495015, EPI_ISL_1495016, EPI_ISL_1495019, EPI_ISL_1495023, EPI_ISL_1495031 | | | |
| see above | Laboratório de Biologia Integrativa | Laboratório de Biologia Integrativa | Alessandro Clayton de Souza Ferreira; Aline Brito de Lima; Carolina Moreira Voloch; Daniel Costa Queiroz; Danielle Alves Gomes Zauli; Diego Menezes Bonfim; Filipe Romero Rebello Moreira; Frederico Scott Varella Malta; Joice do Prado Silva; Lucyene Miguita Luiz; Nuno Rodrigues Faria; Paula Luize Camargos Fonseca; Rafael Marques de Souza; Renan Pedra de Souza; Renato Santana Aguiar; Rennan Garcias Moreira; Victor Cavalcanti Pardini; Victor Emmanuel Viana Geddes |
| EPI_ISL_2466200, EPI_ISL_2466201, EPI_ISL_2466202, EPI_ISL_2466203, EPI_ISL_2466205, EPI_ISL_2466206, EPI_ISL_2466208, EPI_ISL_2466209, EPI_ISL_2466210, EPI_ISL_2466211, EPI_ISL_2466212, EPI_ISL_2466213, EPI_ISL_2466214, EPI_ISL_2466215, EPI_ISL_2466216, EPI_ISL_2466217, EPI_ISL_2466218, EPI_ISL_2466220, EPI_ISL_2466222, EPI_ISL_2466223, EPI_ISL_2466225, EPI_ISL_2536268 | | | |
| see above | Laboratório de Biologia Molecular de Doenças Infecciosas e do Câncer (LADIC - UFRN) | Laboratory of Respiratory Viruses and Measles, Oswaldo Cruz Institute, FIOCRUZ | Alice Sampaio Rocha; Ana Carolina Mendonca; Anna Carolina Paixao; Elisa Cavalcante Pereira; Fernando Motta; Josélio Araújo; Luciana Appolinario; Marilda Siqueira on behalf of the Fiocruz COVID-19 Genomic Surveillance Network; Paola Resende; Renata Serrano Lopes; Taina Venas |
| EPI_ISL_1578453 | Laboratório de Biologia Molecular do Hospital das Clínicas da Faculdade de Medicina de Botucatu/SP | Laboratórios Genômica Funcional (FCA/UNESP) e Biologia Molecular (FMB-HC/UNESP) - Rede de Vigilância Genômica (Vigenômica)/UNESP | Bianca Cechetto Carlos; Felipe Allan da Silva da Costa; Flavia Hebner Barbosa Trovão; Guilherme Targino Valente; Jayme A. Souza-Neto.; Patrícia Akemi Assato; Rejane Maria Tommasini Grotto |
| EPI_ISL_1579258, EPI_ISL_1580502 | Laboratório de Biologia Molecular do Hospital das Clínicas da Faculdade de Medicina de Botucatu/SP | Laboratórios de Genômica Funcional (FCA/UNESP) e Biologia Molecular (FMB-HC/UNESP) - Rede de Vigilância Genômica (Vigenômica)/UNESP | Bianca Cechetto Carlos; Felipe Allan da Silva da Costa; Flavia Hebner Barbosa Trovão; Guilherme Targino Valente; Jayme A. Souza-Neto.; Patrícia Akemi Assato; Rejane Maria Tommasini Grotto |
| EPI_ISL_2466265, EPI_ISL_3061877 | Laboratório de Ecologia de Doenças Transmissíveis na Amazônia (EDTA), Instituto Leônidas e Maria Deane, FIOCRUZ, Manaus, Amazonas, Brazil. | Laboratory of Respiratory Viruses and Measles, Oswaldo Cruz Institute, FIOCRUZ | Alex Pauvolid-Corrêa; Alice Sampaio Rocha; Ana Beatriz Machado Lima; Ana Carolina Mendonca; Anna Carolina Paixao; Elisa Cavalcante Pereira; Felipe Naveca; Fernando Motta; Luciana Appolinario; Marilda Siqueira on behalf of the Fiocruz COVID-19 Genomic Surveillance Network; Mia Ferreira de Araujo; Paola Resende; Renata Serrano Lopes; Taina Venas |
| EPI_ISL_1630158 | Laboratório de Microbiologia Molecular - Universidade FEEVALE | Laboratório de Microbiologia Molecular - Universidade FEEVALE | Alana Witt Hansen; Fernando Rosado Spilki; Flávio Silveira; Fágner Henrique Heldt; Juliana Schons Gularte; Juliane Deise Fleck; Mariana Soares da Silva; Matheus Nunes Weber; Meriane Demoliner; Micheli Filippi.; Paula Rodrigues de Almeida |
| EPI_ISL_2431436, EPI_ISL_2431439, EPI_ISL_2431440, EPI_ISL_2431441, EPI_ISL_2431843, EPI_ISL_2431844, EPI_ISL_2431846, EPI_ISL_2431848, EPI_ISL_2431852, EPI_ISL_2431853, EPI_ISL_3873622, EPI_ISL_3873630, EPI_ISL_3873637, EPI_ISL_3873641, EPI_ISL_3873670, EPI_ISL_4051950, EPI_ISL_4051951, EPI_ISL_4051954, EPI_ISL_4051957 | | | |
| see above | Laboratório de Microbiologia Molecular - Universidade FEEVALE | Molecular Microbiology Laboratory | Alana Witt Hansen; Fernando Rosado Spilki; Flávio Silveira; Fágner Henrique Heldt; Juliana Schons Gularte; Juliane Deise Fleck; Mariana Soares da Silva; Matheus Nunes Weber; Meriane Demoliner; Micheli Filippi; Micheli Filippi.; Paula Rodrigues de Almeida; Vyctoria Malayhka de Abreu Góes Pereira. |
| EPI_ISL_1754186 | Laboratório de Pesquisa em Virologia, FAMERP, SJRP | Laboratório de Pesquisa em Virologia, FAMERP, SJRP | Cecília Artico Banho; Cíntia Bittar; Fábio Sossai Possebon; Guilherme Campos; Helena Lage Ferreira; Jorge A. Petroli Marchesi; João Pessoa Araújo Jr.; Leila Sabrina Ullmann; Lívia Sacchetto; Maisa C. Pereira Parra; Marília Moraes; Maurício L. Nogueira; Paula Rahal; Paulo Inacio da Costa |
| EPI_ISL_1464627, EPI_ISL_1464628, EPI_ISL_1464631, EPI_ISL_1464632, EPI_ISL_1464635, EPI_ISL_1464636, EPI_ISL_1464638, EPI_ISL_1464639, EPI_ISL_1464640, EPI_ISL_1464641, EPI_ISL_1464642, EPI_ISL_1464644, EPI_ISL_1464646, EPI_ISL_1464647, EPI_ISL_1464650, EPI_ISL_1464652, EPI_ISL_1464653, EPI_ISL_1464654, EPI_ISL_1464656, EPI_ISL_1464657, EPI_ISL_1464658, EPI_ISL_1464660, EPI_ISL_1464661, EPI_ISL_1464662, EPI_ISL_1464663, EPI_ISL_1464664, EPI_ISL_1464665, EPI_ISL_1464666, EPI_ISL_1464670, EPI_ISL_1464671 | | | |
| see above | Laboratório de Virologia - UNIFESP | Laboratory of Respiratory Viruses and Measles, Oswaldo Cruz Institute, FIOCRUZ | Alice Sampaio Rocha; Ana Carolina Mendonca; Anna Carolina Paixao; Fernando Motta; Luciana Appolinario; Marilda Siqueira on behalf of the Fiocruz COVID-19 Genomic Surveillance Network; Nancy Belei; Paola Resende; Renata Serrano Lopes |
| EPI_ISL_2629707, EPI_ISL_2629711, EPI_ISL_2629724, EPI_ISL_2629738, EPI_ISL_2629740, EPI_ISL_2629741, EPI_ISL_2629742, EPI_ISL_2629745, EPI_ISL_2629750, EPI_ISL_2629752, EPI_ISL_2629756, EPI_ISL_2629761, EPI_ISL_2629765, EPI_ISL_2629767, EPI_ISL_2629769, EPI_ISL_2629770, EPI_ISL_2629773, EPI_ISL_2629777, EPI_ISL_2629778, EPI_ISL_2629786, EPI_ISL_2629791, EPI_ISL_2629792, EPI_ISL_2629796, EPI_ISL_2629800, EPI_ISL_2629802, EPI_ISL_2629803, EPI_ISL_2629807, EPI_ISL_2629814, EPI_ISL_2629818, EPI_ISL_2629824, EPI_ISL_2629827 | | | |
| see above | Laboratório de Virologia Molecular - Universidade Federal do Rio de Janeiro | Laboratório de Virologia Molecular - Universidade Federal do Rio de Janeiro | ; Alice Laschuk Herlinger; Amilcar Tanuri; André Felipe Andrade dos Santos; Carolina Moreira Voloch; Cássia Cristina Alves Gonçalves; Diana Mariani; Débora Souza Faffe; Filipe Romero Rebello Moreira; Francine Bittencourt Schiffler; Isabela de Carvalho Leitão; Marcelo Calado de Paula Tôrres; Matheus Augusto Calvano Cosentino; Mirela D'arc; Orlando da Costa Ferreira Junior; Rafael Mello Galliez; Raíssa Mirella dos Santos Cunha da Costa; Renato Santana de Aguiar; Terezinha Marta Pereira Pinto Castineiras; Thamiris dos Santos Miranda; Átila Duque Rossi |
| EPI_ISL_1843311, EPI_ISL_2108265 | Labormedizin Darmstadt | Robert Koch Institute | |
| EPI_ISL_2196247, EPI_ISL_2196249, EPI_ISL_2196250, EPI_ISL_2196254, EPI_ISL_2196255, EPI_ISL_2196256, EPI_ISL_2196335, EPI_ISL_2196336, EPI_ISL_2196337, EPI_ISL_2196338, EPI_ISL_2196339, EPI_ISL_2196340, EPI_ISL_2196341, EPI_ISL_2196342, EPI_ISL_2196343, EPI_ISL_2196344, EPI_ISL_2196345, EPI_ISL_2196346, EPI_ISL_2196347, EPI_ISL_2196348, EPI_ISL_2196349, EPI_ISL_2196350, EPI_ISL_2196353, EPI_ISL_2196354, EPI_ISL_2196359, EPI_ISL_2443551, EPI_ISL_2443552, EPI_ISL_2443677, EPI_ISL_2443684, EPI_ISL_2443685, EPI_ISL_2443686, EPI_ISL_2443687, EPI_ISL_2443688, EPI_ISL_2443690, EPI_ISL_2443691, EPI_ISL_2443692, EPI_ISL_2443693, EPI_ISL_2443694, EPI_ISL_2443695, EPI_ISL_2443696, EPI_ISL_2443697, EPI_ISL_2443698, EPI_ISL_2443699, EPI_ISL_2443700, EPI_ISL_2443701, EPI_ISL_2443702, EPI_ISL_2443703, EPI_ISL_2443704, EPI_ISL_2443705, EPI_ISL_2443706, EPI_ISL_2443707, EPI_ISL_2443709, EPI_ISL_2443711, EPI_ISL_2466261, EPI_ISL_2536262, EPI_ISL_2536264, EPI_ISL_2536265, EPI_ISL_2677078, EPI_ISL_2677081, EPI_ISL_2677082, EPI_ISL_2677086, EPI_ISL_2677089, EPI_ISL_2677091, EPI_ISL_2677093, EPI_ISL_2677094, EPI_ISL_2677095, EPI_ISL_2677108, EPI_ISL_2677116, EPI_ISL_2677121, EPI_ISL_2677123, EPI_ISL_2677127, EPI_ISL_2677128, EPI_ISL_2677129, EPI_ISL_2677130, EPI_ISL_2677131, EPI_ISL_2677132, EPI_ISL_2677133, EPI_ISL_2677134, EPI_ISL_2677135, EPI_ISL_2677136, EPI_ISL_2677139, EPI_ISL_2677141, EPI_ISL_2677142, EPI_ISL_2677143, EPI_ISL_2677144, EPI_ISL_2677146, EPI_ISL_2677147, EPI_ISL_2677148, EPI_ISL_2677149, EPI_ISL_2677150, EPI_ISL_2677153, EPI_ISL_2677155, EPI_ISL_2677156, EPI_ISL_2677157, EPI_ISL_2677159, EPI_ISL_2677160, EPI_ISL_2677161, EPI_ISL_2677162, EPI_ISL_2677163, EPI_ISL_2677167, EPI_ISL_2677168, EPI_ISL_2677169, EPI_ISL_2677170, EPI_ISL_2677171, EPI_ISL_2677173, EPI_ISL_2677174, EPI_ISL_2677219, EPI_ISL_2677222, EPI_ISL_2677223, EPI_ISL_2677225, EPI_ISL_2677226, EPI_ISL_2677227, EPI_ISL_2677254 | | | |
| see above | Labortorio Central de Saude Publica do Estado de Santa Catarina (LACEN/SC) | Laboratory of Respiratory Viruses and Measles, Oswaldo Cruz Institute, FIOCRUZ | Alex Pauvolid-Corrêa; Alice Sampaio Rocha; Ana Beatriz Machado Lima; Ana Carolina Mendonca; Anna Carolina Paixao; Darcita Buerger Rovaris; Elisa Cavalcante Pereira; Fernando Motta; Luciana Appolinario; Marilda Siqueira on behalf of the Fiocruz COVID-19 Genomic Surveillance Network; Mia Ferreira de Araujo; Paola Resende; Renata Serrano Lopes; Sandra Bianchini Fernandes; Taina Venas |
| EPI_ISL_2196245, EPI_ISL_2196313, EPI_ISL_2196314, EPI_ISL_2196315, EPI_ISL_2196316, EPI_ISL_2196317, EPI_ISL_2196318, EPI_ISL_2196319, EPI_ISL_2196320, EPI_ISL_2196321, EPI_ISL_2196322, EPI_ISL_2196323, EPI_ISL_2196324, EPI_ISL_2196325, EPI_ISL_2196326, EPI_ISL_2196327, EPI_ISL_2196328, EPI_ISL_2196329, EPI_ISL_2196330, EPI_ISL_2196331, EPI_ISL_2196332, EPI_ISL_2196356, EPI_ISL_2274099, EPI_ISL_2443615, EPI_ISL_2443616, EPI_ISL_2443617, EPI_ISL_2443618, EPI_ISL_2443619, EPI_ISL_2443620, EPI_ISL_2443621, EPI_ISL_2443622, EPI_ISL_2443623, EPI_ISL_2443624, EPI_ISL_2443625, EPI_ISL_2443626, EPI_ISL_2443627, EPI_ISL_2443628, EPI_ISL_2443629, EPI_ISL_2443630, EPI_ISL_2443631, EPI_ISL_2443632, EPI_ISL_2443637, EPI_ISL_2443638, EPI_ISL_2614352, EPI_ISL_2614353, EPI_ISL_2614354, EPI_ISL_2614355, EPI_ISL_2614356, EPI_ISL_2614358, EPI_ISL_2614359, EPI_ISL_2614360, EPI_ISL_2614362, EPI_ISL_2614363, EPI_ISL_2614365, EPI_ISL_2614366, EPI_ISL_2614367, EPI_ISL_2614368, EPI_ISL_2614369, EPI_ISL_2614370, EPI_ISL_2614371, EPI_ISL_2614372, EPI_ISL_2614373, EPI_ISL_2614374, EPI_ISL_2614375, EPI_ISL_2614376, EPI_ISL_2614377, EPI_ISL_2614378, EPI_ISL_2614379, EPI_ISL_2614380, EPI_ISL_3061896 | | | |
| see above | Labortorio Central de Saude Publica do Estado do Rio de Janeiro (LACEN/RJ) | Laboratory of Respiratory Viruses and Measles, Oswaldo Cruz Institute, FIOCRUZ | Alice Sampaio Rocha; Ana Carolina Mendonca; Andrea Cony Cavalcanti; Anna Carolina Paixao; Elisa Cavalcante Pereira; Fernando Motta; Luciana Appolinario; Marilda Siqueira on behalf of the Fiocruz COVID-19 Genomic Surveillance Network; Paola Resende; Renata Serrano Lopes; Taina Venas |
| EPI_ISL_1358300, EPI_ISL_1358301 | Lacen de Tocantins | Instituto Adolfo Lutz, Interdiciplinary Procedures Center, Strategic Laboratory | Caio Vinicius Dias Lopes; Claudia Regina Gonçalves; Claudio Tavares Sacchi; Erica Valessa Ramos Gomes; Karoline Rodrigues Campos |
| EPI_ISL_2157386, EPI_ISL_2157387, EPI_ISL_2157388, EPI_ISL_2157397, EPI_ISL_2157402, EPI_ISL_2157403, EPI_ISL_2157408, EPI_ISL_2157413, EPI_ISL_2157416, EPI_ISL_2157417, EPI_ISL_2157419, EPI_ISL_2157420, EPI_ISL_2157421, EPI_ISL_2157424, EPI_ISL_2157431, EPI_ISL_2157452, EPI_ISL_2157479, EPI_ISL_2157483, EPI_ISL_2157485, EPI_ISL_2157486, EPI_ISL_2157487, EPI_ISL_2157488, EPI_ISL_2157492, EPI_ISL_2157496, EPI_ISL_2157504, EPI_ISL_2157527, EPI_ISL_2196252, EPI_ISL_2196259, EPI_ISL_2196306, EPI_ISL_2196307, EPI_ISL_2196308, EPI_ISL_2196309, EPI_ISL_2196310, EPI_ISL_2196311, EPI_ISL_2196312, EPI_ISL_2274095, EPI_ISL_2274096, EPI_ISL_2443530, EPI_ISL_2443532, EPI_ISL_2443533, EPI_ISL_2443534, EPI_ISL_2443535, EPI_ISL_2443536, EPI_ISL_2443537, EPI_ISL_2443538, EPI_ISL_2443540, EPI_ISL_2443541, EPI_ISL_2443546, EPI_ISL_2443553, EPI_ISL_2443565, EPI_ISL_2443566, EPI_ISL_2443567, EPI_ISL_2443568, EPI_ISL_2443569, EPI_ISL_2443570, EPI_ISL_2443571, EPI_ISL_2443572, EPI_ISL_2443577, EPI_ISL_2603424, EPI_ISL_2603425, EPI_ISL_2603426, EPI_ISL_2603427, EPI_ISL_2603428, EPI_ISL_2603431, EPI_ISL_2603437, EPI_ISL_2603440, EPI_ISL_2603452, EPI_ISL_2603457, EPI_ISL_2603458, EPI_ISL_2603459, EPI_ISL_2603465, EPI_ISL_2603467, EPI_ISL_2603468, EPI_ISL_2603469, EPI_ISL_2603471, EPI_ISL_2603543, EPI_ISL_2603545, EPI_ISL_2603546, EPI_ISL_2613816, EPI_ISL_2613817, EPI_ISL_2613819, EPI_ISL_2613820, EPI_ISL_2613821, EPI_ISL_2613822, EPI_ISL_2613823, EPI_ISL_2613824, EPI_ISL_2645488, EPI_ISL_2645489, EPI_ISL_2645490, EPI_ISL_2863857, EPI_ISL_2863858, EPI_ISL_2863859, EPI_ISL_2863890 | | | |
| see above | Lboratorio Central de Saude Publica do Estado do Parana (LACEN/PR) | Laboratory of Respiratory Viruses and Measles, Oswaldo Cruz Institute, FIOCRUZ | Agatha Cristinne Prudencio Soares; Alice Sampaio Rocha; Ana Carolina Mendonca; Anna Carolina Paixao; Elisa Cavalcante Pereira; Fernando Motta; Ighor Leonardo Arantes Gomes; Irina Riediger; Luciana Appolinario; Marilda Siqueira on behalf of the Fiocruz COVID-19 Genomic Surveillance Network; Paola Resende; Renata Serrano Lopes; Taina Venas |
| EPI_ISL_1718434 | Lighthouse Lab in Alderley Park | Wellcome Sanger Institute for the COVID-19 Genomics UK (COG-UK) Consortium | Cordelia Langford; David K. Jackson; Dominic Kwiatkowski; Ewan Harrison; Ian Johnston; Jacquelyn Wynn; Jeffrey Barrett; John Sillitoe on behalf of the Wellcome Sanger Institute COVID-19 Surveillance Team; Mairead Hyland; Roberto Amato; Sonia Goncalves; The Lighthouse Lab in Alderley Park and Alex Alderton |
| EPI_ISL_1116310, EPI_ISL_1187189, EPI_ISL_1315652, EPI_ISL_1327496, EPI_ISL_1327504, EPI_ISL_1327564, EPI_ISL_1327636, EPI_ISL_1327671, EPI_ISL_1329280, EPI_ISL_1365182, EPI_ISL_1374108, EPI_ISL_1409655, EPI_ISL_1409701, EPI_ISL_1454248, EPI_ISL_1519280, EPI_ISL_1653315, EPI_ISL_1719527, EPI_ISL_1741405, EPI_ISL_1741412 | | | |
| see above | Lighthouse Lab in Cambridge | Wellcome Sanger Institute for the COVID-19 Genomics UK (COG-UK) Consortium | Cordelia Langford; David K. Jackson; Dominic Kwiatkowski; Ewan Harrison; Ian Johnston; Jeffrey Barrett; John Sillitoe on behalf of the Wellcome Sanger Institute COVID-19 Surveillance Team; Rob Howes; Roberto Amato; Sonia Goncalves; The Lighthouse Lab in Cambridge and Alex Alderton |
| EPI_ISL_1277629 | Lighthouse Lab in | Wellcome Sanger | Anna Dominiczak and Alex Alderton; Carol Clugston; Cordelia Langford; David Gray; David K. Jackson; Dominic Kwiatkowski; Ewan Harrison; Harper VanSteenhouse; Ian Johnston; Jeffrey Barrett; John Sillitoe on behalf of the Wellcome Sanger Institute COVID-19 Surveillance Team; Roberto Amato; Sonia Goncalves; Yumi Kasai |

| | | | |
|---|---|---|---|
| | Glasgow | Institute for the COVID-19 Genomics UK (COG-UK) Consortium | |
| EPI_ISL_1084042, EPI_ISL_1084155, EPI_ISL_1207532, EPI_ISL_1718902, EPI_ISL_1718906, EPI_ISL_1741477, EPI_ISL_1831282, EPI_ISL_2005677 | | | |
| see above | Lighthouse Lab in Milton Keynes | Wellcome Sanger Institute for the COVID-19 Genomics UK (COG-UK) Consortium | Cordelia Langford; David K. Jackson; Dominic Kwiatkowski; Ewan Harrison; Ian Johnston; Jeffrey Barrett; John Sillitoe on behalf of the Wellcome Sanger Institute COVID-19 Surveillance Team; Roberto Amato; Sonia Goncalves; The Lighthouse Lab in Milton Keynes and Alex Alderton |
| EPI_ISL_1350502 | Limbach - MVZ Humangenetik Ulm | Robert Koch Institute | |
| EPI_ISL_1502021, EPI_ISL_1502027, EPI_ISL_1502040, EPI_ISL_1704729, EPI_ISL_1704738, EPI_ISL_1704741, EPI_ISL_1704744, EPI_ISL_1704751, EPI_ISL_1704772, EPI_ISL_2009369, EPI_ISL_2009371, EPI_ISL_2009389, EPI_ISL_2009394, EPI_ISL_2373057 | | | |
| see above | Lurie Children's Hospital of Chicago | Northwestern University - Ozer Lab | Egon A. Ozer; Judd F. Hultquist; Lacy M. Simons; Larry K. Kociolek; Michael G. Ison; Ramon Lorenzo-Redondo; Taylor J. Dean; William J. Muller; Xiaotian; Zheng |
| EPI_ISL_873257, EPI_ISL_1531712, EPI_ISL_1857182, EPI_ISL_2036066 | M Health Fairview | Minnesota Department of Health, Public Health Laboratory | Alexandra Lorentz; Jacob Garfin; Matt Plumb; and Xiong Wang |
| EPI_ISL_2801315, EPI_ISL_2801316, EPI_ISL_2801326, EPI_ISL_2801327, EPI_ISL_2801328, EPI_ISL_2801329, EPI_ISL_2801330, EPI_ISL_2801331 | | | |
| see above | MATERNIDADE ESCOLA ASSIS CHATEAUBRIAND | Analytical Competence Molecular Epidemiology Lab/ACME, Oswaldo Cruz Foundation, Ceara (FIOCRUZ CE) | Cleber Furtado Aksenen e Suzana Porto Almeida; Fabio Miyajima; Fernando Braga Stehling; Francisco Eder de Moura Lopes; Jamille Maria Mendes Bezerra; Joaquim César do Nascimento Sousa Junior; Pedro Miguel Carneiro Jeronimo; Thais Ferreira de Oliveira; Thais de Oliveira Costa; Ticiane Cavalcante de Souza; Veridiana Pessoa Miyajima |
| EPI_ISL_3102263 | MATERNIDADE QUITERIA DE LIMA | Analytical Competence Molecular Epidemiology Lab/ACME, Oswaldo Cruz Foundation, Ceara (FIOCRUZ CE) | Cleber Furtado Aksenen; Fabio Miyajima; Fernando Braga Stehling; Francisco Eder de Moura Lopes; Jamille Maria Mendes Bezerra; Joaquim César do Nascimento Sousa Junior; Pedro Miguel Carneiro Jeronimo; Suzana Porto Almeida e Lucas Delerino; Thais Ferreira de Oliveira; Thais de Oliveira Costa; Ticiane Cavalcante de Souza; Veridiana Pessoa Miyajima |
| EPI_ISL_1672366 | MD PHL | MD PHL | Maryland Department of Health Laboratories Administration |
| EPI_ISL_1759395 | ME Health and Environmental Testing Laboratory | Pathogen Discovery, Respiratory Viruses Branch, Division of Viral Diseases, Centers for Disease Control and Prevention | Adam Retchless; Anna Kelleher; Anna Montmayeur; Anna Uehara; Brian Lynch; Clinton R. Paden; Haibin Wang; Han Jia Justin Ng; Jing Zhang; Justin Lee; Krista Queen; Mark Burroughs; Peter Cook; Rachel Marine; Suxiang Tong; Yan Li; Ying Tao |
| EPI_ISL_982899 | MEPHI Aix Marseille University (AMU) | MEPHI Aix Marseille University (AMU) | Anthony LEVASSEUR |
| EPI_ISL_1711833 | MI - Michigan Department of Health and Human Services - Bureau of Laboratories | Centers for Disease Control and Prevention Division of Viral Diseases, Pathogen Discovery | Alison Laufer Halpin; Ben L. Rambo-Martin; Clinton R. Paden; Dakota Howard; Darlene Wagner; Dave Wentworth; Dhwani Batra; Jasmine Padilla; Justin Lee; Katie Dillon; Krista Queen; Kristen Knipe; Kristine Lacek; Mark Burroughs; Matthew Schmerer; Mili Sheth; Peter Cook; Sam Shepard; Sarah Nobles; Shoshona Le; Suxiang Tong; Vivien Dugan; Yvette Unoarumhi |
| EPI_ISL_1371772, EPI_ISL_1371773 | MIRIALIS CLUSES BECHET | CNR Virus des Infections Respiratoires - France SUD | Antonin Bal; Bruno Lina; Gregory Destras; Gwendolyne Burfin; Hadrien Regue; Laurence Josset; Martine Valette; Quentin Semanas |
| EPI_ISL_1755830 | MONTEFIORE MEDICAL CENTER LABORATORIES | Wadsworth Center, New York State Department of Health | Alexis Russell; Catharine Prussing; Daryl M. Lamson; Erasmus Schneider; Erica Lasek-Nesselquist; John Kelly; Jonathan Plitnick; Kirsten St. George; Matthew Shudt; Melissa A Leisner; Navjot Singh |
| EPI_ISL_2689876, EPI_ISL_2689885 | MS Public Health Laboratory | Centers for Disease Control and Prevention Division of Viral Diseases, Pathogen Discovery | Alison Laufer Halpin; Ben L. Rambo-Martin; Clinton R. Paden; Dakota Howard; Darlene Wagner; Dave Wentworth; Dhwani Batra; Jasmine Padilla; Justin Lee; Katie Dillon; Krista Queen; Kristen Knipe; Kristine Lacek; Mark Burroughs; Matthew Schmerer; Mili Sheth; Peter Cook; Sam Shepard; Sarah Nobles; Shoshona Le; Suxiang Tong; Vivien Dugan; Yvette Unoarumhi |
| EPI_ISL_1709345, EPI_ISL_1709625, EPI_ISL_1709863, EPI_ISL_1709940, EPI_ISL_1709956 | MSHS Clinical Microbiology Laboratories | MSHS Pathogen Surveillance Program | Adolfo García-Sastre; Adriana van de Guchte; Ajay Obla; Alberto Paniz-Mondolfi; Ana S. Gonzalez-Reiche; Angela Amoako; Ashley Salimbangon; Betsaida Salom Melo; Bremy Alburquerque; Brianne Ciferri; Charles Gleason; Daniel Floda; Deena R. Altman; Denise Jurczyszak; Emilia Mia Sordillo; Gintaras Deikus; Giulio Kleiner; Gopi Patel; Hala Alshammary; Harm van Bakel; Irina Oussenko; Jayeeta Dutta; Juan Soto; Julia Matthews; Katherine Beach; Kathryn Twyman; Kayla Russo; Komal Srivastava; Levy Sominsky; Mahmoud Awawda; Marta Luksza; Matthew M. Hernandez; Melissa Gitman; Michael D. Nowak; Mitchell J. Sullivan; Nancy Francoeur; Robert Sebra; Sarah Schaefer; Shelcie Fabre; Shwetha Hara Sridhar; Viviana Simon; Ying-Chih Wang; Zenab Khan |
| EPI_ISL_2690685 | MSK Microbiology Lab | MSK Microbiology Lab | Esther Babady; Krupa Jani; Tracy McMillen |
| EPI_ISL_1482931 | MUSC Molecular Pathology Laboratory | MUSC Molecular Pathology Laboratory | Dariusz Pytel; Frederick S. Nolte; Jaclyn Dunne; Julie W. Hirschhorn; Kristen Maurer; W. Bailey Glen Jr |
| EPI_ISL_1643478 | MVZ Dr. Eberhard & Partner Dortmund | Robert Koch Institute | |
| EPI_ISL_1638367 | MVZ Labor Dortmund | Robert Koch Institute | |
| EPI_ISL_1640683, EPI_ISL_1640692, EPI_ISL_1640733 | MVZ Labor Krone GbR | Robert Koch Institute | |
| EPI_ISL_1847231 | MVZ fÃ¼r Laboratoriumsmedizin und Mikrobiologie Koblenz-Mittelrhein (Labor Koblenz) | Robert Koch Institute | |
| EPI_ISL_1821053 | Maine HETL | Health and Environmental Testing Laboratory | Barter, M.; Dewey, H.; H. and Tewhey, R.; Lynch, R.; Martha, J.; Matluk, N.; Munger |
| EPI_ISL_1372646, EPI_ISL_1826325, EPI_ISL_1826326, EPI_ISL_1826327, EPI_ISL_1826329, EPI_ISL_1973041, EPI_ISL_1973064, EPI_ISL_1973065, EPI_ISL_1973066, EPI_ISL_2802277, EPI_ISL_3544238, EPI_ISL_3544240 | | | |
| see above | Maine Health and Environmental Testing Laboratory | Tewhey Lab, The Jackson Laboratory | Barter, M.; Dewey, H.; H. and Tewhey, R.; Iosue, F.; Lynch, R.; Matluk, N.; Munger |
| EPI_ISL_1015571 | Maryland Public Health Laboratory | Maryland Public Health Laboratory | Maryland Department of Health Laboratories Administration |
| EPI_ISL_1404620, EPI_ISL_1406704, EPI_ISL_1527218, EPI_ISL_1528149, EPI_ISL_1528150, EPI_ISL_1624697 | Massachusetts State Public Health Laboratory | Massachusetts State Public Health Laboratory | Andrew Lang; Glen Gallagher; Sandra Smole; Timelia Fink |
| EPI_ISL_1761103 | Medicine and Surgery, University of Insubria | Department of Molecular and Translational Medicine, Section of Microbiology, University of Brescia, ASST Spedali Civili, Brescia | Alberto Zani; Anna Bertelli; Arnaldo Caruso; Carlo Bonfanti; Francesca Caccuri; Serena Messali |
| EPI_ISL_1850411 | Medizinisches Labor Wahl LÃ¼denscheid | Robert Koch Institute | |
| EPI_ISL_2099218 | Michael E. DeBakey | Pathogen Discovery, | Adam Retchless; Anna Kelleher; Anna Uehara; Brian Lynch; Clinton R. Paden; Dhwani Batra; Haibin Wang; Han Jia Justin Ng; Jasmine Padilla; Jing Zhang; Justin Lee; Krista Queen; Mark Burroughs; Mili Sheth; Morgan Davis; Peter Cook; Rachel Marine; Sarah Nobles; Suxiang Tong; Tara Coalter; Yan Li; Ying Tao |

| | | | |
|---|---|---|---|
| | VA Medical Center | Respiratory Viruses Branch, Division of Viral Diseases, Centers for Disease Control and Prevention | |
| EPI_ISL_2374680 | Michigan Department of Health and Human Services, Bureau of Laboratories | Michigan Department of Health and Human Services, Bureau of Laboratories | Blankenship HM; Riner D; Soehnlen MK |
| EPI_ISL_1999828, EPI_ISL_1999848 | Microbiologia CATLAB | Can Ruti SARS-CoV-2 Sequencing Hub (HUGTiP/IrsiCaixa/IGTP) | Alba Sánchez; Anna Not; Antoni E Bordoy; Bonaventura Clotet; Cristina Casañ; Cristina Esteban; Francesc Catala-Moll; Gemma Clara; Ignacio Blanco; Marc Noguera-Julian; Maria Casadellà; Mariona Parera; Mercedes Guerrero; Montserrat Giménez; Pere-Joan Cardona; Pilar Armengol; Roger Paredes; Verónica Saludes; and Elisa Martró on behalf of the Can Ruti SARS-CoV-2 Sequencing Hub. |
| EPI_ISL_1761495, EPI_ISL_1761507, EPI_ISL_1761519, EPI_ISL_1761541, EPI_ISL_1892866, EPI_ISL_1892891, EPI_ISL_1892915, EPI_ISL_1892940, EPI_ISL_1892986, EPI_ISL_1893034 | | | |
| see above | Microbiologia CATLAB | Can Ruti SARS-CoV-2 Sequencing Hub (HUGTiP/IrsiCaixa/IGTP) | Alba Sánchez; Anna Not; Antoni E Bordoy; Bonaventura Clotet; Cristina Casañ; Cristina Esteban; Francesc Catala-Moll; Gemma Clara; Ignacio Blanco; Marc Noguera-Julian; Maria Casadellà; Mariona Parera; Mercedes Guerrero; Montserrat Giménez; Pere-Joan Cardona; Pilar Armengol; Roger Paredes; Verónica Saludes; and Elisa Martró on behalf of the Can Ruti SARS-CoV-2 Sequencing Hub. |
| EPI_ISL_2140526, EPI_ISL_2140535, EPI_ISL_2140538, EPI_ISL_2140613, EPI_ISL_2140636 | Microbiology Department, Laboratori Clínic Metropolitana Nord. Hospital Universitari Germans Trias i Pujol | Can Ruti SARS-CoV-2 Sequencing Hub (HUGTiP/IrsiCaixa/IGTP) | Alba Sánchez; Anna Not; Antoni E Bordoy; Bonaventura Clotet; Cristina Casañ; Cristina Esteban; Francesc Catala-Moll; Gemma Clara; Ignacio Blanco; Marc Noguera-Julian; Maria Casadellà; Mariona Parera; Mercedes Guerrero; Montserrat Giménez; Pere-Joan Cardona; Pilar Armengol; Roger Paredes; Verónica Saludes; and Elisa Martró on behalf of the Can Ruti SARS-CoV-2 Sequencing Hub. |
| EPI_ISL_1208676, EPI_ISL_1503906, EPI_ISL_1547724, EPI_ISL_1547769, EPI_ISL_1547780, EPI_ISL_1583556, EPI_ISL_1583601, EPI_ISL_1583612, EPI_ISL_1636419, EPI_ISL_1636443, EPI_ISL_1636450, EPI_ISL_1636453, EPI_ISL_1636455, EPI_ISL_1636477, EPI_ISL_1636478, EPI_ISL_1661922, EPI_ISL_1661933, EPI_ISL_1661943, EPI_ISL_1661956, EPI_ISL_1661968, EPI_ISL_1719794, EPI_ISL_1719796, EPI_ISL_1719797, EPI_ISL_1719819, EPI_ISL_1719827, EPI_ISL_1719843, EPI_ISL_1761474, EPI_ISL_1761504, EPI_ISL_1761547, EPI_ISL_2035851, EPI_ISL_2035860 | | | |
| see above | Microbiology Department, Laboratori Clínic Metropolitana Nord. Hospital Universitari Germans Trias i Pujol. | Can Ruti SARS-CoV-2 Sequencing Hub (HUGTiP/IrsiCaixa/IGTP) | Alba Sánchez; Anna Not; Antoni E Bordoy; Bonaventura Clotet; Carol Galvez Maria Casadellà; Cristina Casañ; Cristina Esteban; Francesc Catala-Moll; Gemma Clara; Ignacio Blanco; Irina Pey; Jordi Barretina; Julia G Prado; Marc Noguera-Julian; Maria Casadellà; Mariona Parera; Mercedes Guerrero; Montserrat Giménez; Pere-Joan Cardona; Pilar Armengol; Roger Paredes; Verónica Saludes; and Elisa Martró on behalf of the Can Ruti SARS-CoV-2 Sequencing Hub. |
| EPI_ISL_1670709, EPI_ISL_1908795 | Microbiology Department, University Hospital Donostia | Microbiology Department, University Hospital Donostia | Cilla G.; Gomez M; Marimon JM; Montes M; Piñeiro L; Sorarrain A |
| EPI_ISL_1595725, EPI_ISL_1595727, EPI_ISL_2086122, EPI_ISL_2086123, EPI_ISL_2086124, EPI_ISL_2086126, EPI_ISL_2086127, EPI_ISL_2086136, EPI_ISL_2086138, EPI_ISL_2086139, EPI_ISL_2086140, EPI_ISL_2086141, EPI_ISL_2086144, EPI_ISL_2086145, EPI_ISL_2086146, EPI_ISL_2086147, EPI_ISL_2086148, EPI_ISL_2086149, EPI_ISL_2086151, EPI_ISL_2086152, EPI_ISL_2086154, EPI_ISL_2086162, EPI_ISL_2086167, EPI_ISL_2086168, EPI_ISL_2086169, EPI_ISL_2086170, EPI_ISL_2516968, EPI_ISL_2516969, EPI_ISL_2516970, EPI_ISL_2516972, EPI_ISL_2516973, EPI_ISL_2516976 | | | |
| see above | Microbiology Department. Complexo Hospitalario Universitario de Vigo | Microbiology Department. Complexo Hospitalario Universitario de Vigo | Alfaya N; Alonso I; Alvarez M; Cabrera JJ; Carballo R; Cores O; Cortizo S; Martinez L; Mediero G; Perez S; Potel C; Regueiro B; Rey S; Vasallo FJ; del-Campo V |
| EPI_ISL_2106148, EPI_ISL_2289147 | Microbiology Division, SC DHEC | Microbiology Division, SC DHEC | Flores, H.; Freeman, J. |
| EPI_ISL_1661416 | Microvida | Microvida | Jaco Verweij; Joep Stöhr; Suzan D. Pas |
| EPI_ISL_1063969, EPI_ISL_1225863, EPI_ISL_1358351, EPI_ISL_1358352, EPI_ISL_1760136, EPI_ISL_2139609, EPI_ISL_2139610, EPI_ISL_2139611, EPI_ISL_2157996, EPI_ISL_2936139, EPI_ISL_2936164, EPI_ISL_2936169, EPI_ISL_3063486 | | | |
| see above | Ministry of Health Turkey | Ministry of Health Turkey | Ayşe Başak Altaş; Fatma Bayrakdar; Gulay Korukluoglu; Gülay Korukluoğlu; Suleyman Yalcin; Süleyman Yalcin; Yasemin Cosgun; Yasemin Coşgun |
| EPI_ISL_1531600, EPI_ISL_1710932, EPI_ISL_1711038, EPI_ISL_1733517, EPI_ISL_1733533, EPI_ISL_1821030, EPI_ISL_2015211 | | | |
| see above | Minnesota Department of Health, Public Health Laboratory | Minnesota Department of Health, Public Health Laboratory | Alexandra Lorentz; Jacob Garfin; Matt Plumb; and Xiong Wang |
| EPI_ISL_2443060 | Mirassol | Instituto Butantan / FAMERP | Antonio Jorge Martins; Claudia Renata dos Santos Barros; David Schlesinger; Debora Botequio Moretti; Dimas Tadeu Covas; Elaine Cristina Marqueze; Elaine Vieira Santos; Evandra Strazza Rodrigues; Heidge Fukumasu; Jayme Augusto de Souza-Neto; José Salvatore Leister Patané; Luiz Alcantara; Luiz Lehmann Coutinho; Maria Carolina Elias; Maurício Lacerda Nogueira; Rafael dos Santos Bezerra; Raul Machado Neto; Rejane Maria Tommasini Grotto; Ricardo Haddad; Sandra Coccuzzo Sampaio Vessoni; Simone Kashima; Svetoslav Nanev Slavov; Vincent Louis Viala |
| EPI_ISL_925031, EPI_ISL_925032 | Mirialis | CNR Virus des Infections Respiratoires - France SUD | Antonin Bal; Bruno Lina; Gregory Destras; Gwendolyne Burfin; Hadrien Règue; Laurence Josset; Martine Valette; Quentin Semanas |
| EPI_ISL_2035540, EPI_ISL_2035550, EPI_ISL_2035553 | Missouri State Public Health Laboratory | Missouri State Public Health Laboratory | Ashley New; Joshua Barry; Matthew Sinn |
| EPI_ISL_2101992 | Molecular Diagnostics Pathology Department Mater Dei Hospital Malta | Molecular Diagnostics Pathology Department Mater Dei Hospital Malta | C Cilia; G Zahra; L Grech; R Borg |
| EPI_ISL_1403611 | Molecular Virology Unit, Microbiology and Virology Depertment, Fondazione IRCCS Policlinico San Matteo, Pavia | Molecular Virology Unit, Microbiology and Virology Depertment, Fondazione IRCCS Policlinico San Matteo, Pavia | Antonio Piralla; Fausto Baldanti; Federica Giardina; Gherard Batisti Biffignandi; Guglielmo Ferrari; Stefania Paolucci; Stefano Gaiarsa |
| EPI_ISL_1920004 | Montefiore Medical Center Laboratories | Wadsworth Center, New York State Department of Health | Alexis Russell; Catharine Prussing; Daryl M. Lamson; Erasmus Schneider; Erica Lasek-Nesselquist; John Kelly; Jonathan Plitnick; Kirsten St. George; Matthew Shudt; Melissa A Leisner; Navjot Singh |
| EPI_ISL_1591098 | NAMRU-6 | Pathogen Discovery, Respiratory Viruses Branch, Division of Viral Diseases, Centers for Disease Control and Prevention | Adam Retchless; Anna Kelleher; Anna Montmayeur; Anna Uehara; Brian Lynch; Clinton R. Paden; Haibin Wang; Han Jia Justin Ng; Jing Zhang; Justin Lee; Krista Queen; Mark Burroughs; Peter Cook; Rachel Marine; Suxiang Tong; Yan Li; Ying Tao |
| EPI_ISL_3354647 | NC State Laboratory of Public Health | Centers for Disease Control and Prevention Division of Viral Diseases, Pathogen Discovery | Alex Burgin; Ben Rambo-Martin; Clinton Paden; Dakota Howard; Dave Wentworth; Dhwani Batra; Jasmine Padilla; Justin Lee; Krista Queen; Kristen Knipe; Kristine Lacek; Mark Burroughs; Matthew Schmerer; Meghan Bentz; Mili Sheth; Peter Cook; Sam Shepard; Sarah Nobles; Suxiang Tong; Vivien Dugan; Yvette Unoarumhi |
| EPI_ISL_1678371 | NH Dept. of Health and Human Services Public Health Labs | Centers for Disease Control and Prevention Division of Viral Diseases, Pathogen Discovery | Alison Laufer Halpin; Ben L. Rambo-Martin; Clinton R. Paden; Dakota Howard; Darlene Wagner; Dave Wentworth; Dhwani Batra; Jasmine Padilla; Justin Lee; Katie Dillon; Krista Queen; Kristen Knipe; Kristine Lacek; Mark Burroughs; Matthew Schmerer; Mili Sheth; Peter Cook; Sam Shepard; Sarah Nobles; Shoshona Le; Suxiang Tong; Vivien Dugan; Yvette Unoarumhi |
| EPI_ISL_3102314 | NIDADE MISTA DE SAUDE DE MISSAO VELHA | Analytical Competence Molecular Epidemiology Lab/ACME, Oswaldo Cruz Foundation, Ceara (FIOCRUZ CE) | Cleber Furtado Aksenen; Fabio Miyajima; Fernando Braga Stehling; Francisco Eder de Moura Lopes; Jamille Maria Mendes Bezerra; Joaquim César do Nascimento Sousa Junior; Pedro Miguel Carneiro Jeronimo; Suzana Porto Almeida e Lucas Delerino; Thais Ferreira de Oliveira; Thais de Oliveira Costa; Ticiane Cavalcante de Souza; Veridiana Pessoa Miyajima |
| EPI_ISL_1465755, EPI_ISL_1465756, EPI_ISL_1582291, EPI_ISL_1671640, EPI_ISL_1671641 | NORTHWELL HEALTH LABORATORIES | Wadsworth Center, New York State Department of Health | Alexis Russell; Catharine Prussing; Daryl M. Lamson; Erasmus Schneider; Erica Lasek-Nesselquist; John Kelly; Jonathan Plitnick; Kirsten St. George; Matthew Shudt; Melissa A Leisner; Navjot Singh |

| | | | |
|---|---|---|---|
| EPI_ISL_1711773 | NV State Public Health Laboratory | Centers for Disease Control and Prevention Division of Viral Diseases, Pathogen Discovery | Alison Laufer Halpin; Ben L. Rambo-Martin; Clinton R. Paden; Dakota Howard; Darlene Wagner; Dave Wentworth; Dhwani Batra; Jasmine Padilla; Justin Lee; Katie Dillon; Krista Queen; Kristen Knipe; Kristine Lacek; Mark Burroughs; Matthew Schmerer; Mili Sheth; Peter Cook; Sam Shepard; Sarah Nobles; Shoshona Le; Suxiang Tong; Vivien Dugan; Yvette Unoarumhi |
| EPI_ISL_1654584, EPI_ISL_1654617, EPI_ISL_1789918 | NYU Langone Health | Departments of Pathology and Medicine, New York University School of Medicine | Adriana Heguy; Christian Marier; Dacia Dimartino; Emily Guzman; Gael Westby; Guiqing Wang; Paolo Cotzia; Paul Zappile; Peter Meyn; Sitharam Ramaswami; Yutong Zhang |
| EPI_ISL_875688 | National Influenza Center - Instituto Adolfo Lutz | Instituto Adolfo Lutz, Interdiciplinary Procedures Center, Strategic Laboratory | Ana Lucia de Carvalho Avelino; Claudia Regina Gonçalves; Claudio Tavares Sacchi; Clovis Roberto Abe Constantino; Erica Valessa Ramos Gomes; Karoline Rodrigues Campos; Katia Correa de Oliveira Santos |
| EPI_ISL_1184825, EPI_ISL_1969931 | National Institute of Infectious Diseases-Prof. Dr. Matei Bals Molecular Diagnostics Laboratory | National Institute of Infectious Diseases-Prof. Dr. Matei Bals Molecular Diagnostics Laboratory | Andreea Tudor; Corina Casangiu; Dan Otelea; Leontina Banica; Marius Surleac; Ovidiu Vlaicu; Petre Milu; Simona Paraschiv |
| EPI_ISL_1055106, EPI_ISL_1180388, EPI_ISL_1301964, EPI_ISL_1323769, EPI_ISL_1357775, EPI_ISL_1357776, EPI_ISL_1499762, EPI_ISL_1499806, EPI_ISL_1499807, EPI_ISL_1499863, EPI_ISL_1499895, EPI_ISL_1499924, EPI_ISL_1577415, EPI_ISL_1577467, EPI_ISL_1577559, EPI_ISL_1623005, EPI_ISL_1623006, EPI_ISL_1651781 | | | |
| see above | National Virus Reference Laboratory | National Virus Reference Laboratory | Calum Walsh; Charlene Bennet; Charlene Bennett; Cillian F De Gascun; Fiona Crispie; Gabriel Gonzalez; Jonathan Dean; Matthew McCabe; Michael Carr; Paul Cotter; Zoe Yandle |
| EPI_ISL_1626543, EPI_ISL_1738803, EPI_ISL_2030453, EPI_ISL_2229831 | Nebraska Public Health Laboratory | NPHL COVID-19 Response Team | NPHL COVID-19 Response Team |
| EPI_ISL_1620692, EPI_ISL_1939412 | North Dakota Department of Health, Public Health Laboratory | North Dakota Department of Health, Public Health Laboratory | Lisa Wingerter |
| EPI_ISL_1920014, EPI_ISL_1920038 | Northwell Health Laboratories | Wadsworth Center, New York State Department of Health | Alexis Russell; Catharine Prussing; Daryl M. Lamson; Erasmus Schneider; Erica Lasek-Nesselquist; John Kelly; Jonathan Plitnick; Kirsten St. George; Matthew Shudt; Melissa A Leisner; Navjot Singh |
| EPI_ISL_2136545 | Northwest Laboratory | Seattle Flu Study | Amanda Adler; Barry R. Lutz; Benjamin Pelle; Brian Hiatt; Caitlin R. Wolf; Chris D. Frazar; Deborah A. Nickerson; Elisabeth Brandstetter; Erica Ryke; Geoff Melly; Helen Y. Chu; Janet A. Englund; Jay Shendure; Jover Lee; Kairsten Fay; Kirsten Lacombe; Lea M. Starita; Mark J. Rieder; Matthew Richardson; Matthew Thompson; Melissa Truong; Michael Boeckh; Michael Famulare; Misja Ilcisin; Peter D. Han; Philip Dykema; Romesh Gautom; Scott Lindquist; Thomas R. Sibley; Trevor Bedford |
| EPI_ISL_1157408, EPI_ISL_1340837, EPI_ISL_1340838, EPI_ISL_1340849, EPI_ISL_1340856, EPI_ISL_1502062, EPI_ISL_1704688, EPI_ISL_1704698, EPI_ISL_1704705, EPI_ISL_1704712, EPI_ISL_1704714, EPI_ISL_2009315, EPI_ISL_2009316, EPI_ISL_2009318, EPI_ISL_2009330 | | | |
| see above | Northwestern Memorial Hospital | Northwestern University - Ozer Lab | Chad J. Achenbach; Chao Qi; Egon A. Ozer; Judd F. Hultquist; Lacy M. Simons; Lawrence J. Jennings; Michael G. Ison; Ramon Lorenzo-Redondo; Taylor J. Dean |
| EPI_ISL_2090677, EPI_ISL_2090678, EPI_ISL_2246699, EPI_ISL_2692030, EPI_ISL_3388721 | Northwestern Memorial Hospital | RIPHL at Rush University Medical Center | Amber Kimble; Chao Qi; Edith Perez; Felix Araujo Perez; Kevin Kunstman; Laura Furtado; Marieta Hyde; Max Kolton; Stefan Green |
| EPI_ISL_1897570 | Norwegian Institute of Public Health, Department of Virology | Norwegian Institute of Public Health, Department of Virology | Atiya R Ali; Debech Nadia; Engebretsen Serina Beate; Garcia Llorente Ignacio; Hilde Elshaug; Hilde Vollan; Jon Bråte; Kamilla Heddeland Instefjord; Karoline Bragstad; Kathrine Stene-Johansen; Marie Paulsen Madsen; Olav Hungnes; Pedersen Benedikte Nevjen; Rasmus Riis Kopperud |
| EPI_ISL_2443063 | Nova Aliança | Instituto Butantan / FAMERP | Antonio Jorge Martins; Claudia Renata dos Santos Barros; David Schlesinger; Debora Botequio Moretti; Dimas Tadeu Covas; Elaine Cristina Marqueze; Elaine Vieira Santos; Evandra Strazza Rodrigues; Heidge Fukumasu; Jayme Augusto de Souza-Neto; José Salvatore Leister Patané; Luiz Alcantara; Luiz Lehmann Coutinho; Maria Carolina Elias; Maurício Lacerda Nogueira; Rafael dos Santos Bezerra; Raul Machado Neto; Rejane Maria Tommasini Grotto; Ricardo Haddad; Sandra Coccuzzo Sampaio Vessoni; Simone Kashima; Svetoslav Nanev Slavov; Vincent Louis Viala |
| EPI_ISL_2262575 | OCME Office Of Chief Medical Examiner | New York City Public Health Laboratory | Jade Wang; et al. |
| EPI_ISL_1542030, EPI_ISL_1701786, EPI_ISL_1701787, EPI_ISL_2080748, EPI_ISL_2080749, EPI_ISL_2080750, EPI_ISL_2080751, EPI_ISL_2080752, EPI_ISL_2080753, EPI_ISL_2080754, EPI_ISL_2080755, EPI_ISL_2080756, EPI_ISL_2080757, EPI_ISL_2080758, EPI_ISL_2080759, EPI_ISL_2080760, EPI_ISL_2080761, EPI_ISL_2080762, EPI_ISL_2227675, EPI_ISL_2227676, EPI_ISL_2227677, EPI_ISL_2227678, EPI_ISL_2227679, EPI_ISL_2227680, EPI_ISL_2227681, EPI_ISL_2227682, EPI_ISL_2227683, EPI_ISL_2227684, EPI_ISL_2227685, EPI_ISL_2227686, EPI_ISL_2227687, EPI_ISL_2227688, EPI_ISL_2227689 | | | |
| see above | OHSU Lab Services Molecular Microbiology Lab | Oregon SARS-CoV-2 Genome Sequencing Center | Alec J. Hirsch; Andrew C. Adey; Benjamin N. Bimber; Brendan L. O'Connell; Brian J. O'Roak; Cierra LaBlanc; Daniel N. Streblow; Donna Hansel; Guang Fan; Kayla Carter; Ruth V. Nichols; Sally Grindstaff; Sonia Acharya; William B. Messer; Xuan Qin |
| EPI_ISL_1759374 | OK Public Health Laboratory, Oklahoma State DOH | Pathogen Discovery, Respiratory Viruses Branch, Division of Viral Diseases, Centers for Disease Control and Prevention | Adam Retchless; Anna Kelleher; Anna Montmayeur; Anna Uehara; Brian Lynch; Clinton R. Paden; Haibin Wang; Han Jia Justin Ng; Jing Zhang; Justin Lee; Krista Queen; Mark Burroughs; Peter Cook; Rachel Marine; Suxiang Tong; Yan Li; Ying Tao |
| EPI_ISL_1701833, EPI_ISL_1711792, EPI_ISL_3354346 | OR State PHL-Virology/Immunology Section | Centers for Disease Control and Prevention Division of Viral Diseases, Pathogen Discovery | Alex Burgin; Alison Laufer Halpin; Ben L. Rambo-Martin; Ben Rambo-Martin; Clinton Paden; Clinton R. Paden; Dakota Howard; Darlene Wagner; Dave Wentworth; Dhwani Batra; Jasmine Padilla; Justin Lee; Katie Dillon; Krista Queen; Kristen Knipe; Kristine Lacek; Mark Burroughs; Matthew Schmerer; Meghan Bentz; Mili Sheth; Peter Cook; Sam Shepard; Sarah Nobles; Shoshona Le; Suxiang Tong; Vivien Dugan; Yvette Unoarumhi |
| EPI_ISL_1169498 | OR State PHL-Virology/Immunology Section | Genomics and Discovery, Respiratory Viruses Branch, Division of Viral Diseases, Centers for Disease Control and Prevention | Anna Montmayeur; Anna Uehara; Ben L. Rambo-Martin; Clinton R. Paden; Dhwani Batra; Haibin Wang; Jasmine Padilla; Jing Zhang; Justin Lee; Krista Queen; Lori Rowe; Mark Burroughs; Mili Sheth; Peter W. Cook; Rachel Marine; Sarah Nobles; Suxiang Tong; Yan Li; Ying Tao |
| EPI_ISL_1686350 | Ohio Department of Health Laboratory | Ohio Department of Health Laboratory | Allison Black; Brent Lee; Caitlin McDonnell; Eric Brandt; Erica Leasure; Glen McGillivary; Heather Blankenship; Holmes; Jade Mowery; Jennifer; Kelsey Florek; Keoni Omura; Kirtana Ramadugu; Quanta Brown; Stephanie Mccracken; Tyler Payne; and Tammy Bannerman |
| EPI_ISL_1310787, EPI_ISL_1310788 | Ohio State University Wexner Medical Center | James Polaris Molecular Laboratory | Chang Y-S; Chappell D; Corcoran S; Garee J; Jones D; Koenig S; Pancholi P; Ru P; Snyder P; Tu H |
| EPI_ISL_1968980 | Omics Sciences Laboratory | Omics Sciences Laboratory | Darlyn Amaya; Derly Andrade Molina; Gabriel Morey León; Juan Carlos Fernández Cadena; Katheryn Sacheri Viteri; Rubén Armas González |
| EPI_ISL_1511642, EPI_ISL_1585676, EPI_ISL_1623696, EPI_ISL_1897447, EPI_ISL_1964166, EPI_ISL_1964167 | Oregon State Public Health Laboratory | Oregon State Public Health Laboratory | Eugene Yeboah; John Fontana and Shane Sevey; Laura Tsaknaridis; Rafia Razzaque; Vanda Makris |
| EPI_ISL_1730802 | Originating lab: Wales Specialist Virology Centre Sequencing lab: Pathogen Genomics Unit | Public Health Wales Microbiology Cardiff Wales Specialist Virology Centre | Alec Birchley; Alexander Adams; Amy Gaskin; Angela Marchbank; Bree Gatica-Wilcox; Catherine Moore; Jason Coombes; Joanne Watkins; Joel Southgate; Johnathan Evans; Laura Gifford; Lauren Gilbert; Lee Graham; Malorie Perry; Matthew Bull; Nicole Pacchiarini; Sally Corden; Sara Kumziene-Summerhayes; Sara Rey; Sarah Taylor; Simon Cottrell; Sophie Jones; Tom Connor |
| EPI_ISL_1931621 | Osaka Institute of Public Health, Morinomiya Center | Pathogen Genomics Center, National Institute of Infectious Diseases | Hazuka Y Furihata; Kentaro Itokawa; Makoto Kuroda; Masanori Hashino; Masumichi Saito; Naomi Nojiri; Nozomu Hanaoka; Rina Tanaka; Sana Uchikoba; Tsuguto Fujimoto; Tsuyoshi Sekizuka |
| EPI_ISL_2661913, EPI_ISL_2661915, | Oswaldo Cruz Institute, FIOCRUZ/CE | Analytical Competence Molecular Epidemiology | Alice Sampaio Rocha; Ana Carolina Mendonca; Anna Carolina Paixao; Elisa Cavalcante Pereira; Fabio Miyajima; Fernando Motta; Luciana Appolinario; Marilda Siqueira on behalf of the Fiocruz COVID-19 Genomic Surveillance Network; Paola Resende; Renata Serrano Lopes; Taina Venas |

| | | | |
|---|---|---|---|
| EPI_ISL_2661917, EPI_ISL_2661922, EPI_ISL_2661928, EPI_ISL_2661931 | | Lab/ACME, Oswaldo Cruz Foundation, Ceara (FIOCRUZ CE) | |
| EPI_ISL_1262913 | Outre Mer | National Reference Center for Viruses of Respiratory Infections, Institut Pasteur, Paris | Angela Brisebarre; Camille Capel; Etienne Simon-Lorière; Marion Barbet; Maud Vanpeene; Méline Bizard; Rousset Dominique; Sylvie Behillil; Sylvie van der Werf; Vincent Enouf |
| EPI_ISL_1001384, EPI_ISL_1001385, EPI_ISL_1096262, EPI_ISL_1856984, EPI_ISL_1856985, EPI_ISL_1856986, EPI_ISL_1856987, EPI_ISL_1856989, EPI_ISL_1856990, EPI_ISL_1856993, EPI_ISL_2293328, EPI_ISL_2293329, EPI_ISL_2293330, EPI_ISL_2293332, EPI_ISL_2293334, EPI_ISL_2293335, EPI_ISL_2293336, EPI_ISL_2293337, EPI_ISL_2293338, EPI_ISL_2293339, EPI_ISL_2293340, EPI_ISL_2293341, EPI_ISL_2293342, EPI_ISL_2293343, EPI_ISL_2293345, EPI_ISL_2293346, EPI_ISL_2293347, EPI_ISL_2293348, EPI_ISL_2293351, EPI_ISL_2293352, EPI_ISL_2293354, EPI_ISL_2293355, EPI_ISL_2293356 | | | |
| see above | Outre mer | National Reference Center for Viruses of Respiratory Infections, Institut Pasteur, Paris | Angela Brisebarre; Camille Capel; Christophe Malabat; Corinne Maufrais; Dominique Rousset; Etienne Simon-Lorière; Frédéric Lemoine; Gael Millot; Louise Lefrançois; Marion Barbet; Maud Vanpeene; Méline Bizard; Pierre Lechat; Rousset (Guy) Dominique; Rousset Dominique; Sylvie Behillil; Sylvie Van der Werf; Sylvie van der Werf; Vincent Enouf |
| EPI_ISL_1967005 | PA DE IBITIUVA DR OTAVIO BENETTI PITANGUEIRAS | Instituto Butantan / ESALQ-USP (Piracicaba) | Antonio Jorge Martins; Bianca Cechetto Carlos. Mendelics: Bibiana Santos; Claudia Renata dos Santos Barros; Cíntia Bittar; David Schlesinger. Hemocentro Ribeirão Preto: Simone Kashima; Debora Botequio Moretti; Elaine Cristina Marqueze; Elaine Vieira dos Santos; Elisangela Chicaroni Mattos; Erika Freitas; Evandra Strazza Rodrigues; Felipe Allan da Silva da Costa; Flavia Aburjaile; Fábio Sossai Possebon; Guilherme Campos; Guilherme Targino Valente; Heidge Fukumasu. USP-Botucatu: Rejane Maria Tommasini Grotto; Helena Lage Ferreira; Instituto Butantan: Dimas Tadeu Covas; Jardelina de Souza Todao Bernardino; Jayme A. Souza-Neto; Jessika Cristina Chagas Lesbon; Jorge A. Petroli Marchesi; José Salvatore Leister Patané; João Paulo Kitajima; João Pessoa Araújo Jr.; Leila Sabrina Ullmann; Loyze Paola Oliveira de Lima; Luiz Aurelio de Campos Crispin. Centro de Genômica Funcional da ESALQ: Luiz Lehmann Coutinho; Luiz Carlos Junior de Alcantara; Lívia Sacchetto; Maisa C. Pereira Parra; Maria Carolina Elias; Marta Giovanetti; Marília Moraes; Maurício Lacerda Nogueira. Prefeitura de Sao Paulo: Melissa Palmieri.; Patricia Akemi Assato; Paula Rahal; Paulo Inacio da Costa; Rafael dos Santos Bezerra; Raquel de Lello Rocha Campos Cassano. NGS Soluções Genômicas: Pilar Drummond Sampaio Corrêa Mariani. FZEA-USP Pirassununga: Mirele Daiana Poleti; Raul Machado Neto; Ricardo Augusto Brassaloti; Ricardo Haddad; Rodrigo Tocantins Calado. FAMERP-SJRP: Cecília Artico Banho; Sandra Coccuzzo Sampaio; Svetoslav Nanev Slavov; Vagner Fonseca; Vincent Louis Viala |
| EPI_ISL_2467902 | PA Department of Health, Bureau of Laboratories | Genomics and Discovery, Respiratory Viruses Branch, Division of Viral Diseases, Centers for Disease Control and Prevention | Adam Retchless; Anna Kelleher; Anna Uehara; Brian Lynch; Clinton R. Paden; Dhwani Batra; Haibin Wang; Han Jia Justin Ng; Jasmine Padilla; Jing Zhang; Justin Lee; Krista Queen; Mark Burroughs; Mili Sheth; Morgan Davis; Peter Cook; Rachel Marine; Sarah Nobles; Suxiang Tong; Tara Coalter; Yan Li; Ying Tao |
| EPI_ISL_2345965, EPI_ISL_2345966 | PA NOVO OSASCO | Instituto Butantan | Antonio Jorge Martins; Claudia Renata dos Santos Barros; David Schlesinger; Debora Botequio Moretti; Dimas Tadeu Covas; Elaine Cristina Marqueze; Elaine Vieira Santos; Evandra Strazza Rodrigues; Heidge Fukumasu; Jayme Augusto de Souza-Neto; José Salvatore Leister Patané; Luiz Alcantara; Luiz Lehmann Coutinho; Maria Carolina Elias; Maurício Lacerda Nogueira; Rafael dos Santos Bezerra; Raul Machado Neto; Rejane Maria Tommasini Grotto; Ricardo Haddad; Sandra Coccuzzo Sampaio Vessoni; Simone Kashima; Svetoslav Nanev Slavov; Vincent Louis Viala |
| EPI_ISL_1795336, EPI_ISL_2345602 | PAS JOAO ANTONIO DO NASCIMENTO | Instituto Butantan / ESALQ-Piracicaba | Antonio Jorge Martins; Bianca Cechetto Carlos. Mendelics: Bibiana Santos; Claudia Renata dos Santos Barros; David Schlesinger; David Schlesinger. Hemocentro Ribeirão Preto: Simone Kashima; Debora Botequio Moretti; Debora Botequio Moretti. Centro de Genômica Funcional da ESALQ: Luiz Lehmann Coutinho; Dimas Tadeu Covas; Elaine Cristina Marqueze; Elaine Vieira Santos; Elaine Vieira dos Santos; Elisangela Chicaroni Mattos; Erika Freitas; Evandra Strazza Rodrigues; Felipe Allan da Silva da Costa; Flavia Aburjaile; Guilherme Targino Valente; Heidge Fukumasu; Heidge Fukumasu. USP-Botucatu: Rejane Maria Tommasini Grotto; Instituto Butantan: Alexander Roberto Precioso; Jayme A. Souza-Neto; Jayme Augusto de Souza-Neto; Jessika Cristina Chagas Lesbon; José Salvatore Leister Patané; João Paulo Kitajima; Luiz Alcantara; Luiz Carlos Junior de Alcantara; Luiz Lehmann Coutinho; Maria Carolina Elias; Marta Giovanetti; Maurício Lacerda Nogueira; Patricia Akemi Assato; Rafael dos Santos Bezerra; Raquel de Lello Rocha Campos Cassano. NGS Soluções Genômicas: Pilar Drummond Sampaio Corrêa Mariani. FZEA-USP Pirassununga: Mirele Daiana Poleti; Raul Machado Neto; Rejane Maria Tommasini Grotto; Ricardo Augusto Brassaloti; Ricardo Haddad; Rodrigo Tocantins Calado.; Sandra Coccuzzo Sampaio; Sandra Coccuzzo Sampaio Vessoni; Simone Kashima; Svetoslav Nanev Slavov; Vagner Fonseca; Vincent Louis Viala |
| EPI_ISL_1960076 | PECET | Universidad Nacional de Colombia - Laboratorio Genómico One Health | Andres F. Cardona-Rios; Carlos Franco-Muñoz; Carolina Muñoz-Arango; Celeny Ortiz; Daniel O. Maldonado-Perez; Diego A. Álvarez-Díaz; Hector Alejandro Ruiz-Moreno; Idabely Betancur Ortiz; Jorge E. Osorio; Juan P. Hernandez-Ortiz; Karl A Ciuoderis; Katherine Laiton-Donato; Laura Silvana Perez; Lina M. Hurtado; Marcela Mercado-Reyes; Maria Angélica Maya; Maria Stella López; Rita Almanza Payares; Sandra Ines Cano; Simón Villegas Velásquez |
| EPI_ISL_1966916 | PENITENCIARIA ODON RAMOS MARANHAO IPERO | Instituto Butantan / ESALQ-USP (Piracicaba) | Antonio Jorge Martins; Bianca Cechetto Carlos. Mendelics: Bibiana Santos; Claudia Renata dos Santos Barros; Cíntia Bittar; David Schlesinger. Hemocentro Ribeirão Preto: Simone Kashima; Debora Botequio Moretti; Elaine Cristina Marqueze; Elaine Vieira dos Santos; Elisangela Chicaroni Mattos; Erika Freitas; Evandra Strazza Rodrigues; Felipe Allan da Silva da Costa; Flavia Aburjaile; Fábio Sossai Possebon; Guilherme Campos; Guilherme Targino Valente; Heidge Fukumasu. USP-Botucatu: Rejane Maria Tommasini Grotto; Helena Lage Ferreira; Instituto Butantan: Dimas Tadeu Covas; Jardelina de Souza Todao Bernardino; Jayme A. Souza-Neto; Jessika Cristina Chagas Lesbon; Jorge A. Petroli Marchesi; José Salvatore Leister Patané; João Paulo Kitajima; João Pessoa Araújo Jr.; Leila Sabrina Ullmann; Loyze Paola Oliveira de Lima; Luiz Aurelio de Campos Crispin. Centro de Genômica Funcional da ESALQ: Luiz Lehmann Coutinho; Luiz Carlos Junior de Alcantara; Lívia Sacchetto; Maisa C. Pereira Parra; Maria Carolina Elias; Marta Giovanetti; Marília Moraes; Maurício Lacerda Nogueira. Prefeitura de Sao Paulo: Melissa Palmieri.; Patricia Akemi Assato; Paula Rahal; Paulo Inacio da Costa; Rafael dos Santos Bezerra; Raquel de Lello Rocha Campos Cassano. NGS Soluções Genômicas: Pilar Drummond Sampaio Corrêa Mariani. FZEA-USP Pirassununga: Mirele Daiana Poleti; Raul Machado Neto; Ricardo Augusto Brassaloti; Ricardo Haddad; Rodrigo Tocantins Calado. FAMERP-SJRP: Cecília Artico Banho; Sandra Coccuzzo Sampaio; Svetoslav Nanev Slavov; Vagner Fonseca; Vincent Louis Viala |
| EPI_ISL_1966927, EPI_ISL_1966928, EPI_ISL_1966929, EPI_ISL_1966930, EPI_ISL_1966931, EPI_ISL_1966932, EPI_ISL_1966933, EPI_ISL_1966934 | | | |
| see above | POLICLINICA COVID 19 ITAPETININGA | Instituto Butantan / ESALQ-USP (Piracicaba) | Antonio Jorge Martins; Bianca Cechetto Carlos. Mendelics: Bibiana Santos; Claudia Renata dos Santos Barros; Cíntia Bittar; David Schlesinger. Hemocentro Ribeirão Preto: Simone Kashima; Debora Botequio Moretti; Elaine Cristina Marqueze; Elaine Vieira dos Santos; Elisangela Chicaroni Mattos; Erika Freitas; Evandra Strazza Rodrigues; Felipe Allan da Silva da Costa; Flavia Aburjaile; Fábio Sossai Possebon; Guilherme Campos; Guilherme Targino Valente; Heidge Fukumasu. USP-Botucatu: Rejane Maria Tommasini Grotto; Helena Lage Ferreira; Instituto Butantan: Dimas Tadeu Covas; Jardelina de Souza Todao Bernardino; Jayme A. Souza-Neto; Jessika Cristina Chagas Lesbon; Jorge A. Petroli Marchesi; José Salvatore Leister Patané; João Paulo Kitajima; João Pessoa Araújo Jr.; Leila Sabrina Ullmann; Loyze Paola Oliveira de Lima; Luiz Aurelio de Campos Crispin. Centro de Genômica Funcional da ESALQ: Luiz Lehmann Coutinho; Luiz Carlos Junior de Alcantara; Lívia Sacchetto; Maisa C. Pereira Parra; Maria Carolina Elias; Marta Giovanetti; Marília Moraes; Maurício Lacerda Nogueira. Prefeitura de Sao Paulo: Melissa Palmieri.; Patricia Akemi Assato; Paula Rahal; Paulo Inacio da Costa; Rafael dos Santos Bezerra; Raquel de Lello Rocha Campos Cassano. NGS Soluções Genômicas: Pilar Drummond Sampaio Corrêa Mariani. FZEA-USP Pirassununga: Mirele Daiana Poleti; Raul Machado Neto; Ricardo Augusto Brassaloti; Ricardo Haddad; Rodrigo Tocantins Calado. FAMERP-SJRP: Cecília Artico Banho; Sandra Coccuzzo Sampaio; Svetoslav Nanev Slavov; Vagner Fonseca; Vincent Louis Viala |
| EPI_ISL_1966465, EPI_ISL_1966467, EPI_ISL_2345643 | POLICLINICA COVID 19 ITAPETININGA | Instituto Butantan / Mendelics | Antonio Jorge Martins; Bianca Cechetto Carlos. Mendelics: Bibiana Santos; Claudia Renata dos Santos Barros; Cíntia Bittar; David Schlesinger; David Schlesinger. Hemocentro Ribeirão Preto: Simone Kashima; Debora Botequio Moretti; Dimas Tadeu Covas; Elaine Cristina Marqueze; Elaine Vieira Santos; Elaine Vieira dos Santos; Elisangela Chicaroni Mattos; Erika Freitas; Evandra Strazza Rodrigues; Felipe Allan da Silva da Costa; Flavia Aburjaile; Fábio Sossai Possebon; Guilherme Campos; Guilherme Targino Valente; Heidge Fukumasu; Heidge Fukumasu. USP-Botucatu: Rejane Maria Tommasini Grotto; Helena Lage Ferreira; Instituto Butantan: Dimas Tadeu Covas; Jardelina de Souza Todao Bernardino; Jayme A. Souza-Neto; Jayme Augusto de Souza-Neto; Jessika Cristina Chagas Lesbon; Jorge A. Petroli Marchesi; José Salvatore Leister Patané; João Paulo Kitajima; João Pessoa Araújo Jr.; Leila Sabrina Ullmann; Loyze Paola Oliveira de Lima; Luiz Alcantara; Luiz Aurelio de Campos Crispin. Centro de Genômica Funcional da ESALQ: Luiz Lehmann Coutinho; Luiz Carlos Junior de Alcantara; Luiz Lehmann Coutinho; Lívia Sacchetto; Maisa C. Pereira Parra; Maria Carolina Elias; Marta Giovanetti; Marília Moraes; Maurício Lacerda Nogueira; Maurício Lacerda Nogueira. Prefeitura de Sao Paulo: Melissa Palmieri.; Patricia Akemi Assato; Paula Rahal; Paulo Inacio da Costa; Rafael dos Santos Bezerra; Raquel de Lello Rocha Campos Cassano. NGS Soluções Genômicas: Pilar Drummond Sampaio Corrêa Mariani. FZEA-USP Pirassununga: Mirele Daiana Poleti; Raul Machado Neto; Rejane Maria Tommasini Grotto; Ricardo Augusto Brassaloti; Ricardo Haddad; Rodrigo Tocantins Calado. FAMERP-SJRP: Cecília Artico Banho; Sandra Coccuzzo Sampaio; Sandra Coccuzzo Sampaio Vessoni; Simone Kashima; Svetoslav Nanev Slavov; Vagner Fonseca; Vincent Louis Viala |
| EPI_ISL_1795082, EPI_ISL_1795083, EPI_ISL_1795141, EPI_ISL_1795142, EPI_ISL_1795143, EPI_ISL_1795144, EPI_ISL_1795145, EPI_ISL_1795146, EPI_ISL_1795148, EPI_ISL_1795152, EPI_ISL_1795153, EPI_ISL_1795154, EPI_ISL_1795155, EPI_ISL_1795156, EPI_ISL_1795157, EPI_ISL_1795158, EPI_ISL_1795159, EPI_ISL_1795177, EPI_ISL_1795180, EPI_ISL_1795181, EPI_ISL_1795183, EPI_ISL_1795184, EPI_ISL_1795185, EPI_ISL_1795186, EPI_ISL_1795188, EPI_ISL_1795190, EPI_ISL_1795387, EPI_ISL_1795388, EPI_ISL_2345344, EPI_ISL_2345345, EPI_ISL_2345346, EPI_ISL_2345350, EPI_ISL_2345351, EPI_ISL_2345352, EPI_ISL_2345353, EPI_ISL_2345356, EPI_ISL_2345357, EPI_ISL_2345361, EPI_ISL_2345362, EPI_ISL_2345363, EPI_ISL_2345364, EPI_ISL_2345365, EPI_ISL_2345366, EPI_ISL_2345367, EPI_ISL_2345368, EPI_ISL_2345369, EPI_ISL_2345394, EPI_ISL_2345397, EPI_ISL_2345398, EPI_ISL_2345399, EPI_ISL_2345401, EPI_ISL_2345402, EPI_ISL_2345403, EPI_ISL_2345404, EPI_ISL_2345409, EPI_ISL_2345411, EPI_ISL_2345412 | | | |
| see above | POLICLINICA HORTOLANDIA | Instituto Butantan / ESALQ-Piracicaba | Antonio Jorge Martins; Bianca Cechetto Carlos. Mendelics: Bibiana Santos; Claudia Renata dos Santos Barros; David Schlesinger; David Schlesinger. Hemocentro Ribeirão Preto: Simone Kashima; Debora Botequio Moretti; Debora Botequio Moretti. Centro de Genômica Funcional da ESALQ: Luiz Lehmann Coutinho; Dimas Tadeu Covas; Elaine Cristina Marqueze; Elaine Vieira Santos; Elaine Vieira dos Santos; Elisangela Chicaroni Mattos; Erika Freitas; Evandra Strazza Rodrigues; Felipe Allan da Silva da Costa; Flavia Aburjaile; Guilherme Targino Valente; Heidge Fukumasu; Heidge Fukumasu. USP-Botucatu: Rejane Maria Tommasini Grotto; Instituto Butantan: Alexander Roberto Precioso; Jayme A. Souza-Neto; Jayme Augusto de Souza-Neto; Jessika Cristina Chagas Lesbon; José Salvatore Leister Patané; João Paulo Kitajima; Luiz Alcantara; Luiz Carlos Junior de Alcantara; Luiz Lehmann Coutinho; Maria Carolina Elias; Marta Giovanetti; Maurício Lacerda Nogueira; Patricia Akemi Assato; Rafael dos Santos Bezerra; Raquel de Lello Rocha Campos Cassano. NGS Soluções Genômicas: Pilar Drummond Sampaio Corrêa Mariani. FZEA-USP Pirassununga: Mirele Daiana Poleti; Raul Machado Neto; Rejane Maria Tommasini Grotto; Ricardo Augusto Brassaloti; Ricardo Haddad; Rodrigo Tocantins Calado.; Sandra Coccuzzo Sampaio; Sandra Coccuzzo Sampaio Vessoni; Simone Kashima; Svetoslav Nanev Slavov; Vagner Fonseca; Vincent Louis Viala |
| EPI_ISL_1966623, EPI_ISL_2171016 | POLICLINICA HORTOLANDIA | Instituto Butantan / Mendelics | Antonio Jorge Martins; Bianca Cechetto Carlos. Mendelics: Bibiana Santos; Claudia Renata dos Santos Barros; Cíntia Bittar; David Schlesinger. Hemocentro Ribeirão Preto: Simone Kashima; Debora Botequio Moretti; Elaine Cristina Marqueze; Elaine Vieira dos Santos; Elisangela Chicaroni Mattos; Erika Freitas; Evandra Strazza Rodrigues; Felipe Allan da Silva da Costa; Flavia Aburjaile; Fábio Sossai Possebon; Guilherme Campos; Guilherme Targino Valente; Heidge Fukumasu. USP-Botucatu: Rejane Maria Tommasini Grotto; Helena Lage Ferreira; Instituto Butantan: Dimas Tadeu Covas; Jardelina de Souza Todao Bernardino; Jayme A. Souza-Neto; Jessika Cristina Chagas Lesbon; Jorge A. Petroli Marchesi; José Salvatore Leister Patané; João Paulo Kitajima; João Pessoa Araújo Jr.; Leila Sabrina Ullmann; Loyze Paola Oliveira de Lima; Luiz Aurelio de Campos Crispin. Centro de Genômica Funcional da ESALQ: Luiz Lehmann Coutinho; Luiz Carlos Junior de Alcantara; Lívia Sacchetto; Maisa C. Pereira Parra; Maria Carolina Elias; Marta Giovanetti; Marília Moraes; Maurício Lacerda Nogueira. Prefeitura de Sao Paulo: Melissa Palmieri.; Patricia Akemi Assato; Paula Rahal; Paulo Inacio da Costa; Rafael dos Santos Bezerra; Raquel de Lello Rocha Campos Cassano. NGS Soluções Genômicas: Pilar Drummond Sampaio Corrêa Mariani. FZEA-USP Pirassununga: Mirele Daiana Poleti; Raul Machado Neto; Ricardo Augusto Brassaloti; Ricardo Haddad; Rodrigo Tocantins Calado. FAMERP-SJRP: Cecília Artico Banho; Sandra Coccuzzo Sampaio; Svetoslav Nanev Slavov; Vagner Fonseca; Vincent Louis Viala |
| EPI_ISL_2345978, EPI_ISL_2345980 | POLICLINICA JARDIM PARAISO | Instituto Butantan | Antonio Jorge Martins; Claudia Renata dos Santos Barros; David Schlesinger; Debora Botequio Moretti; Dimas Tadeu Covas; Elaine Cristina Marqueze; Elaine Vieira Santos; Evandra Strazza Rodrigues; Heidge Fukumasu; Jayme Augusto de Souza-Neto; José Salvatore Leister Patané; Luiz Alcantara; Luiz Lehmann Coutinho; Maria Carolina Elias; Maurício Lacerda Nogueira; Rafael dos Santos Bezerra; Raul Machado Neto; Rejane Maria Tommasini Grotto; Ricardo Haddad; Sandra Coccuzzo Sampaio Vessoni; Simone Kashima; Svetoslav Nanev Slavov; Vincent Louis Viala |
| EPI_ISL_2142665 | POLYCLINIQUE BORDEAUX NORD AQUITAINE | CNR Virus des Infections Respiratoires - France SUD | Antonin Bal; Bruno Lina; Gregory Destras; Gwendolyne Burfin; Hadrien Regue; Laurence Josset; Martine Valette; Quentin Semanas |
| EPI_ISL_2080492 | POOLE MICROBIOLOGY LABORATORY | COVID-19 Genomics UK (COG-UK) Consortium | PHE Covid Sequencing Team |
| EPI_ISL_3102492 | POSTO DE SAUDE MARIA DE LOURDES MAGALHAES MAIA | Analytical Competence Molecular Epidemiology Lab/ACME, Oswaldo Cruz Foundation, Ceara (FIOCRUZ CE) | Cleber Furtado Aksenen; Fabio Miyajima; Fernando Braga Stehling; Francisco Eder de Moura Lopes; Jamille Maria Mendes Bezerra; Joaquim César do Nascimento Sousa Junior; Pedro Miguel Carneiro Jeronimo; Suzana Porto Almeida e Lucas Delerino; Thais Ferreira de Oliveira; Thais de Oliveira Costa; Ticiane Cavalcante de Souza; Veridiana Pessoa Miyajima |
| EPI_ISL_3102481 | POSTO SAUDE DE VICOSA | Analytical Competence Molecular Epidemiology Lab/ACME, Oswaldo Cruz Foundation, Ceara (FIOCRUZ CE) | Cleber Furtado Aksenen; Fabio Miyajima; Fernando Braga Stehling; Francisco Eder de Moura Lopes; Jamille Maria Mendes Bezerra; Joaquim César do Nascimento Sousa Junior; Pedro Miguel Carneiro Jeronimo; Suzana Porto Almeida e Lucas Delerino; Thais Ferreira de Oliveira; Thais de Oliveira Costa; Ticiane Cavalcante de Souza; Veridiana Pessoa Miyajima |
| EPI_ISL_1445224, EPI_ISL_1966218 | PPA FENELON GUEDES PEREIRA | Instituto Butantan / Mendelics | Antonio Jorge Martins; Bianca Cechetto Carlos. Mendelics: Bibiana Santos; Bibiana Santos; Claudia Renata dos Santos Barros; Cíntia Bittar; David Schlesinger; David Schlesinger. Hemocentro Ribeirão Preto: Simone Kashima; Debora Botequio Moretti; Dimas Tadeu Covas; Elaine Cristina Marqueze; Elaine Vieira dos Santos; Elisangela Chicaroni Mattos; Erika Freitas; Evandra Strazza Rodrigues; Felipe Allan da Silva da Costa; Flavia Aburjaile; Fábio Sossai Possebon; Guilherme Campos; Guilherme Targino Valente; Heidge Fukumasu. USP-Botucatu: Rejane Maria Tommasini Grotto; Helena Lage Ferreira; Instituto Butantan: Dimas Tadeu Covas; Jardelina de Souza Todao Bernardino; Jayme A. Souza-Neto; Jessika Cristina Chagas Lesbon; Jorge A. Petroli Marchesi; José Salvatore Leister Patané; João Paulo Kitajima; João Pessoa Araújo Jr.; Leila Sabrina Ullmann; Loyze Paola Oliveira de Lima; Luiz Aurelio de Campos Crispin. Centro de Genômica Funcional da ESALQ: Luiz Lehmann Coutinho; Luiz Carlos Junior de Alcantara; Lívia Sacchetto; Maisa C. Pereira Parra; Maria Carolina Elias; Marta Giovanetti; Marília Moraes; Maurício Lacerda Nogueira. Prefeitura de Sao Paulo: Melissa Palmieri.; Patricia Akemi Assato; Paula Rahal; Paulo Inacio da Costa; Rafael dos Santos Bezerra; Raquel de Lello Rocha Campos Cassano. NGS Soluções Genômicas: Pilar Drummond Sampaio Corrêa Mariani. FZEA-USP Pirassununga: Mirele Daiana Poleti; Raul Machado Neto; Ricardo Augusto Brassaloti; Ricardo Haddad; Rodrigo Tocantins Calado.; Rodrigo Tocantins Calado. FAMERP-SJRP: Cecília Artico Banho; Sandra Coccuzzo Sampaio; Simone Kashima; Svetoslav Nanev Slavov; Vagner Fonseca; Vincent Louis Viala |
| EPI_ISL_1966727 | PPA MARCIO VALDEVINO BATISTA | Instituto Butantan | Antonio Jorge Martins; Bianca Cechetto Carlos. Mendelics: Bibiana Santos; Claudia Renata dos Santos Barros; Cíntia Bittar; David Schlesinger. Hemocentro Ribeirão Preto: Simone Kashima; Debora Botequio Moretti; Elaine Cristina Marqueze; Elaine Vieira dos Santos; Elisangela Chicaroni Mattos; Erika Freitas; Evandra Strazza Rodrigues; Felipe Allan da Silva da Costa; Flavia Aburjaile; Fábio Sossai Possebon; Guilherme Campos; Guilherme Targino Valente; Heidge Fukumasu. USP-Botucatu: Rejane Maria Tommasini Grotto; Helena Lage Ferreira; Instituto Butantan: Dimas Tadeu Covas; Jardelina de Souza Todao Bernardino; Jayme A. Souza-Neto; Jessika Cristina Chagas Lesbon; Jorge A. Petroli Marchesi; José Salvatore Leister Patané; João Paulo Kitajima; João Pessoa Araújo Jr.; Leila Sabrina Ullmann; Loyze Paola Oliveira de Lima; Luiz Aurelio de Campos Crispin. Centro de Genômica Funcional da ESALQ: Luiz Lehmann Coutinho; Luiz Carlos Junior de Alcantara; Lívia Sacchetto; Maisa C. Pereira Parra; Maria Carolina Elias; Marta Giovanetti; Marília Moraes; Maurício Lacerda Nogueira. Prefeitura de Sao Paulo: Melissa Palmieri.; Patricia Akemi Assato; Paula Rahal; Paulo Inacio da Costa; Rafael dos Santos Bezerra; Raquel de Lello Rocha Campos Cassano. NGS Soluções Genômicas: Pilar Drummond Sampaio Corrêa Mariani. FZEA-USP Pirassununga: Mirele Daiana Poleti; Raul Machado Neto; Ricardo Augusto Brassaloti; Ricardo Haddad; Rodrigo Tocantins Calado. FAMERP-SJRP: Cecília Artico Banho; Sandra Coccuzzo Sampaio; Svetoslav Nanev Slavov; Vagner Fonseca; Vincent Louis Viala |
| EPI_ISL_1711685, EPI_ISL_3354801, EPI_ISL_3354802 | PR Public Health Lab | Centers for Disease Control and Prevention Division of Viral Diseases, Pathogen | Alex Burgin; Alison Laufer Halpin; Ben L. Rambo-Martin; Ben Rambo-Martin; Clinton Paden; Clinton R. Paden; Dakota Howard; Darlene Wagner; Dave Wentworth; Dhwani Batra; Jasmine Padilla; Justin Lee; Katie Dillon; Krista Queen; Kristen Knipe; Kristine Lacek; Mark Burroughs; Matthew Schmerer; Meghan Bentz; Mili Sheth; Peter Cook; Sam Shepard; Sarah Nobles; Shoshona Le; Suxiang Tong; Vivien Dugan; Yvette Unoarumhi |

| | | | |
|---|---|---|---|
| | | Discovery | |
| EPI_ISL_1966910, EPI_ISL_1966911 | PR S DA FAMILIA UNIDADE DE SAUDE ADALBERTO ROCHA GUAREI | Instituto Butantan / ESALQ-USP (Piracicaba) | Antonio Jorge Martins; Bianca Cechetto Carlos. Mendelics: Bibiana Santos; Claudia Renata dos Santos Barros; Cíntia Bittar; David Schlesinger. Hemocentro Ribeirão Preto: Simone Kashima; Debora Botequio Moretti; Elaine Cristina Marqueze; Elaine Vieira dos Santos; Elisangela Chicaroni Mattos; Erika Freitas; Evandra Strazza Rodrigues; Felipe Allan da Silva da Costa; Flavia Aburjaile; Fábio Sossai Possebon; Guilherme Campos; Guilherme Targino Valente; Heidge Fukumasu. USP-Botucatu: Rejane Maria Tommasini Grotto; Helena Lage Ferreira; Instituto Butantan: Dimas Tadeu Covas; Jardelina de Souza Todao Bernardino; Jayme A. Souza-Neto; Jessika Cristina Chagas Lesbon; Jorge A. Petroli Marchesi; José Salvatore Leister Patané; João Paulo Kitajima; João Pessoa Araújo Jr.; Leila Sabrina Ullmann; Loyze Paola Oliveira de Lima; Luiz Aurelio de Campos Crispin. Centro de Genômica Funcional da ESALQ: Luiz Lehmann Coutinho; Luiz Carlos Junior de Alcantara; Lívia Sacchetto; Maisa C. Pereira Parra; Maria Carolina Elias; Marta Giovanetti; Marília Moraes; Maurício Lacerda Nogueira. Prefeitura de Sao Paulo: Melissa Palmieri.; Patricia Akemi Assato; Paula Rahal; Paulo Inacio da Costa; Rafael dos Santos Bezerra; Raquel de Lello Rocha Campos Cassano. NGS Soluções Genômicas: Pilar Drummond Sampaio Corrêa Mariani. FZEA-USP Pirassununga: Mirele Daiana Poleti; Raul Machado Neto; Ricardo Augusto Brassaloti; Ricardo Haddad; Rodrigo Tocantins Calado. FAMERP-SJRP: Cecília Artico Banho; Sandra Coccuzzo Sampaio; Svetoslav Nanev Slavov; Vagner Fonseca; Vincent Louis Viala |
| EPI_ISL_2345991, EPI_ISL_2345992, EPI_ISL_2346013, EPI_ISL_2346014 | PRONTO ATENDIMENTO DE VARGEM GRANDE PAULISTA | Instituto Butantan | Antonio Jorge Martins; Claudia Renata dos Santos Barros; David Schlesinger; Debora Botequio Moretti; Dimas Tadeu Covas; Elaine Cristina Marqueze; Elaine Vieira Santos; Evandra Strazza Rodrigues; Heidge Fukumasu; Jayme Augusto de Souza-Neto; José Salvatore Leister Patané; Luiz Alcantara; Luiz Lehmann Coutinho; Maria Carolina Elias; Maurício Lacerda Nogueira; Rafael dos Santos Bezerra; Raul Machado Neto; Rejane Maria Tommasini Grotto; Ricardo Haddad; Sandra Coccuzzo Sampaio Vessoni; Simone Kashima; Svetoslav Nanev Slavov; Vincent Louis Viala |
| EPI_ISL_1967281 | PRONTO ATENDIMENTO MUNICIPAL | Instituto Butantan / Mendelics | Antonio Jorge Martins; Bianca Cechetto Carlos. Mendelics: Bibiana Santos; Claudia Renata dos Santos Barros; Cíntia Bittar; David Schlesinger. Hemocentro Ribeirão Preto: Simone Kashima; Debora Botequio Moretti; Elaine Cristina Marqueze; Elaine Vieira dos Santos; Elisangela Chicaroni Mattos; Erika Freitas; Evandra Strazza Rodrigues; Felipe Allan da Silva da Costa; Flavia Aburjaile; Fábio Sossai Possebon; Guilherme Campos; Guilherme Targino Valente; Heidge Fukumasu. USP-Botucatu: Rejane Maria Tommasini Grotto; Helena Lage Ferreira; Instituto Butantan: Dimas Tadeu Covas; Jardelina de Souza Todao Bernardino; Jayme A. Souza-Neto; Jessika Cristina Chagas Lesbon; Jorge A. Petroli Marchesi; José Salvatore Leister Patané; João Paulo Kitajima; João Pessoa Araújo Jr.; Leila Sabrina Ullmann; Loyze Paola Oliveira de Lima; Luiz Aurelio de Campos Crispin. Centro de Genômica Funcional da ESALQ: Luiz Lehmann Coutinho; Luiz Carlos Junior de Alcantara; Lívia Sacchetto; Maisa C. Pereira Parra; Maria Carolina Elias; Marta Giovanetti; Marília Moraes; Maurício Lacerda Nogueira. Prefeitura de Sao Paulo: Melissa Palmieri.; Patricia Akemi Assato; Paula Rahal; Paulo Inacio da Costa; Rafael dos Santos Bezerra; Raquel de Lello Rocha Campos Cassano. NGS Soluções Genômicas: Pilar Drummond Sampaio Corrêa Mariani. FZEA-USP Pirassununga: Mirele Daiana Poleti; Raul Machado Neto; Ricardo Augusto Brassaloti; Ricardo Haddad; Rodrigo Tocantins Calado. FAMERP-SJRP: Cecília Artico Banho; Sandra Coccuzzo Sampaio; Svetoslav Nanev Slavov; Vagner Fonseca; Vincent Louis Viala |
| EPI_ISL_2344789 | PRONTO ATENDIMENTO MUNICIPAL DE JACUPIRANGA | Instituto Butantan / ESALQ-Piracicaba | Antonio Jorge Martins; Claudia Renata dos Santos Barros; David Schlesinger; Debora Botequio Moretti; Dimas Tadeu Covas; Elaine Cristina Marqueze; Elaine Vieira Santos; Evandra Strazza Rodrigues; Heidge Fukumasu; Jayme Augusto de Souza-Neto; José Salvatore Leister Patané; Luiz Alcantara; Luiz Lehmann Coutinho; Maria Carolina Elias; Maurício Lacerda Nogueira; Rafael dos Santos Bezerra; Raul Machado Neto; Rejane Maria Tommasini Grotto; Ricardo Haddad; Sandra Coccuzzo Sampaio Vessoni; Simone Kashima; Svetoslav Nanev Slavov; Vincent Louis Viala |
| EPI_ISL_1966952, EPI_ISL_1966953 | PRONTO ATENDIMENTO MUNICIPAL DE JACUPIRANGA | Instituto Butantan / ESALQ-USP (Piracicaba) | Antonio Jorge Martins; Bianca Cechetto Carlos. Mendelics: Bibiana Santos; Claudia Renata dos Santos Barros; Cíntia Bittar; David Schlesinger. Hemocentro Ribeirão Preto: Simone Kashima; Debora Botequio Moretti; Elaine Cristina Marqueze; Elaine Vieira dos Santos; Elisangela Chicaroni Mattos; Erika Freitas; Evandra Strazza Rodrigues; Felipe Allan da Silva da Costa; Flavia Aburjaile; Fábio Sossai Possebon; Guilherme Campos; Guilherme Targino Valente; Heidge Fukumasu. USP-Botucatu: Rejane Maria Tommasini Grotto; Helena Lage Ferreira; Instituto Butantan: Dimas Tadeu Covas; Jardelina de Souza Todao Bernardino; Jayme A. Souza-Neto; Jessika Cristina Chagas Lesbon; Jorge A. Petroli Marchesi; José Salvatore Leister Patané; João Paulo Kitajima; João Pessoa Araújo Jr.; Leila Sabrina Ullmann; Loyze Paola Oliveira de Lima; Luiz Aurelio de Campos Crispin. Centro de Genômica Funcional da ESALQ: Luiz Lehmann Coutinho; Luiz Carlos Junior de Alcantara; Lívia Sacchetto; Maisa C. Pereira Parra; Maria Carolina Elias; Marta Giovanetti; Marília Moraes; Maurício Lacerda Nogueira. Prefeitura de Sao Paulo: Melissa Palmieri.; Patricia Akemi Assato; Paula Rahal; Paulo Inacio da Costa; Rafael dos Santos Bezerra; Raquel de Lello Rocha Campos Cassano. NGS Soluções Genômicas: Pilar Drummond Sampaio Corrêa Mariani. FZEA-USP Pirassununga: Mirele Daiana Poleti; Raul Machado Neto; Ricardo Augusto Brassaloti; Ricardo Haddad; Rodrigo Tocantins Calado. FAMERP-SJRP: Cecília Artico Banho; Sandra Coccuzzo Sampaio; Svetoslav Nanev Slavov; Vagner Fonseca; Vincent Louis Viala |
| EPI_ISL_1966845 | PRONTO ATENDIMENTO MUNICIPAL ITALO SANTUCCI | Instituto Butantan / ESALQ-USP (Piracicaba) | Antonio Jorge Martins; Bianca Cechetto Carlos. Mendelics: Bibiana Santos; Claudia Renata dos Santos Barros; Cíntia Bittar; David Schlesinger. Hemocentro Ribeirão Preto: Simone Kashima; Debora Botequio Moretti; Elaine Cristina Marqueze; Elaine Vieira dos Santos; Elisangela Chicaroni Mattos; Erika Freitas; Evandra Strazza Rodrigues; Felipe Allan da Silva da Costa; Flavia Aburjaile; Fábio Sossai Possebon; Guilherme Campos; Guilherme Targino Valente; Heidge Fukumasu. USP-Botucatu: Rejane Maria Tommasini Grotto; Helena Lage Ferreira; Instituto Butantan: Dimas Tadeu Covas; Jardelina de Souza Todao Bernardino; Jayme A. Souza-Neto; Jessika Cristina Chagas Lesbon; Jorge A. Petroli Marchesi; José Salvatore Leister Patané; João Paulo Kitajima; João Pessoa Araújo Jr.; Leila Sabrina Ullmann; Loyze Paola Oliveira de Lima; Luiz Aurelio de Campos Crispin. Centro de Genômica Funcional da ESALQ: Luiz Lehmann Coutinho; Luiz Carlos Junior de Alcantara; Lívia Sacchetto; Maisa C. Pereira Parra; Maria Carolina Elias; Marta Giovanetti; Marília Moraes; Maurício Lacerda Nogueira. Prefeitura de Sao Paulo: Melissa Palmieri.; Patricia Akemi Assato; Paula Rahal; Paulo Inacio da Costa; Rafael dos Santos Bezerra; Raquel de Lello Rocha Campos Cassano. NGS Soluções Genômicas: Pilar Drummond Sampaio Corrêa Mariani. FZEA-USP Pirassununga: Mirele Daiana Poleti; Raul Machado Neto; Ricardo Augusto Brassaloti; Ricardo Haddad; Rodrigo Tocantins Calado. FAMERP-SJRP: Cecília Artico Banho; Sandra Coccuzzo Sampaio; Svetoslav Nanev Slavov; Vagner Fonseca; Vincent Louis Viala |
| EPI_ISL_1795219, EPI_ISL_2345449 | PRONTO ATENDIMENTO UNIDADE SAUDE ADALBERTO ROCHA GUAREI | Instituto Butantan / ESALQ-Piracicaba | Antonio Jorge Martins; Bianca Cechetto Carlos. Mendelics: Bibiana Santos; Claudia Renata dos Santos Barros; David Schlesinger; David Schlesinger. Hemocentro Ribeirão Preto: Simone Kashima; Debora Botequio Moretti; Debora Botequio Moretti. Centro de Genômica Funcional da ESALQ: Luiz Lehmann Coutinho; Dimas Tadeu Covas; Elaine Cristina Marqueze; Elaine Vieira Santos; Elaine Vieira dos Santos; Elisangela Chicaroni Mattos; Erika Freitas; Evandra Strazza Rodrigues; Felipe Allan da Silva da Costa; Flavia Aburjaile; Guilherme Targino Valente; Heidge Fukumasu; Heidge Fukumasu. USP-Botucatu: Rejane Maria Tommasini Grotto; Instituto Butantan: Alexander Roberto Precioso; Jayme A. Souza-Neto; Jayme Augusto de Souza-Neto; Jessika Cristina Chagas Lesbon; José Salvatore Leister Patané; João Paulo Kitajima; Luiz Alcantara; Luiz Carlos Junior de Alcantara; Luiz Lehmann Coutinho; Maria Carolina Elias; Marta Giovanetti; Maurício Lacerda Nogueira; Patricia Akemi Assato; Rafael dos Santos Bezerra; Raquel de Lello Rocha Campos Cassano. NGS Soluções Genômicas: Pilar Drummond Sampaio Corrêa Mariani. FZEA-USP Pirassununga: Mirele Daiana Poleti; Raul Machado Neto; Rejane Maria Tommasini Grotto; Ricardo Augusto Brassaloti; Ricardo Haddad; Rodrigo Tocantins Calado.; Sandra Coccuzzo Sampaio; Sandra Coccuzzo Sampaio Vessoni; Simone Kashima; Svetoslav Nanev Slavov; Vagner Fonseca; Vincent Louis Viala |
| EPI_ISL_1445104, EPI_ISL_1966134, EPI_ISL_1966135 | PRONTO ATENDIMENTO VILA PADRE ANCHIETA | Instituto Butantan / Mendelics | Antonio Jorge Martins; Bianca Cechetto Carlos. Mendelics: Bibiana Santos; Bibiana Santos; Claudia Renata dos Santos Barros; Cíntia Bittar; David Schlesinger; David Schlesinger. Hemocentro Ribeirão Preto: Simone Kashima; Debora Botequio Moretti; Dimas Tadeu Covas; Elaine Cristina Marqueze; Elaine Vieira dos Santos; Elisangela Chicaroni Mattos; Erika Freitas; Evandra Strazza Rodrigues; Felipe Allan da Silva da Costa; Flavia Aburjaile; Fábio Sossai Possebon; Guilherme Campos; Guilherme Targino Valente; Heidge Fukumasu. USP-Botucatu: Rejane Maria Tommasini Grotto; Helena Lage Ferreira; Instituto Butantan: Dimas Tadeu Covas; Jardelina de Souza Todao Bernardino; Jayme A. Souza-Neto; Jessika Cristina Chagas Lesbon; Jorge A. Petroli Marchesi; José Salvatore Leister Patané; João Paulo Kitajima; João Pessoa Araújo Jr.; Leila Sabrina Ullmann; Loyze Paola Oliveira de Lima; Luiz Aurelio de Campos Crispin. Centro de Genômica Funcional da ESALQ: Luiz Lehmann Coutinho; Luiz Carlos Junior de Alcantara; Lívia Sacchetto; Maisa C. Pereira Parra; Maria Carolina Elias; Marta Giovanetti; Marília Moraes; Maurício Lacerda Nogueira. Prefeitura de Sao Paulo: Melissa Palmieri.; Patricia Akemi Assato; Paula Rahal; Paulo Inacio da Costa; Rafael dos Santos Bezerra; Raquel de Lello Rocha Campos Cassano. NGS Soluções Genômicas: Pilar Drummond Sampaio Corrêa Mariani. FZEA-USP Pirassununga: Mirele Daiana Poleti; Raul Machado Neto; Ricardo Augusto Brassaloti; Ricardo Haddad; Rodrigo Tocantins Calado.; Rodrigo Tocantins Calado. FAMERP-SJRP: Cecília Artico Banho; Sandra Coccuzzo Sampaio; Simone Kashima; Svetoslav Nanev Slavov; Vagner Fonseca; Vincent Louis Viala |
| EPI_ISL_1966876 | PRONTO SOCORRO CAUCAIA DO ALTO | Instituto Butantan / ESALQ-USP (Piracicaba) | Antonio Jorge Martins; Bianca Cechetto Carlos. Mendelics: Bibiana Santos; Claudia Renata dos Santos Barros; Cíntia Bittar; David Schlesinger. Hemocentro Ribeirão Preto: Simone Kashima; Debora Botequio Moretti; Elaine Cristina Marqueze; Elaine Vieira dos Santos; Elisangela Chicaroni Mattos; Erika Freitas; Evandra Strazza Rodrigues; Felipe Allan da Silva da Costa; Flavia Aburjaile; Fábio Sossai Possebon; Guilherme Campos; Guilherme Targino Valente; Heidge Fukumasu. USP-Botucatu: Rejane Maria Tommasini Grotto; Helena Lage Ferreira; Instituto Butantan: Dimas Tadeu Covas; Jardelina de Souza Todao Bernardino; Jayme A. Souza-Neto; Jessika Cristina Chagas Lesbon; Jorge A. Petroli Marchesi; José Salvatore Leister Patané; João Paulo Kitajima; João Pessoa Araújo Jr.; Leila Sabrina Ullmann; Loyze Paola Oliveira de Lima; Luiz Aurelio de Campos Crispin. Centro de Genômica Funcional da ESALQ: Luiz Lehmann Coutinho; Luiz Carlos Junior de Alcantara; Lívia Sacchetto; Maisa C. Pereira Parra; Maria Carolina Elias; Marta Giovanetti; Marília Moraes; Maurício Lacerda Nogueira. Prefeitura de Sao Paulo: Melissa Palmieri.; Patricia Akemi Assato; Paula Rahal; Paulo Inacio da Costa; Rafael dos Santos Bezerra; Raquel de Lello Rocha Campos Cassano. NGS Soluções Genômicas: Pilar Drummond Sampaio Corrêa Mariani. FZEA-USP Pirassununga: Mirele Daiana Poleti; Raul Machado Neto; Ricardo Augusto Brassaloti; Ricardo Haddad; Rodrigo Tocantins Calado. FAMERP-SJRP: Cecília Artico Banho; Sandra Coccuzzo Sampaio; Svetoslav Nanev Slavov; Vagner Fonseca; Vincent Louis Viala |
| EPI_ISL_1967239 | PRONTO SOCORRO CENTRAL GUIOMAR FERREIRA ROEBBELEN | Instituto Butantan / Mendelics | Antonio Jorge Martins; Bianca Cechetto Carlos. Mendelics: Bibiana Santos; Claudia Renata dos Santos Barros; Cíntia Bittar; David Schlesinger. Hemocentro Ribeirão Preto: Simone Kashima; Debora Botequio Moretti; Elaine Cristina Marqueze; Elaine Vieira dos Santos; Elisangela Chicaroni Mattos; Erika Freitas; Evandra Strazza Rodrigues; Felipe Allan da Silva da Costa; Flavia Aburjaile; Fábio Sossai Possebon; Guilherme Campos; Guilherme Targino Valente; Heidge Fukumasu. USP-Botucatu: Rejane Maria Tommasini Grotto; Helena Lage Ferreira; Instituto Butantan: Dimas Tadeu Covas; Jardelina de Souza Todao Bernardino; Jayme A. Souza-Neto; Jessika Cristina Chagas Lesbon; Jorge A. Petroli Marchesi; José Salvatore Leister Patané; João Paulo Kitajima; João Pessoa Araújo Jr.; Leila Sabrina Ullmann; Loyze Paola Oliveira de Lima; Luiz Aurelio de Campos Crispin. Centro de Genômica Funcional da ESALQ: Luiz Lehmann Coutinho; Luiz Carlos Junior de Alcantara; Lívia Sacchetto; Maisa C. Pereira Parra; Maria Carolina Elias; Marta Giovanetti; Marília Moraes; Maurício Lacerda Nogueira. Prefeitura de Sao Paulo: Melissa Palmieri.; Patricia Akemi Assato; Paula Rahal; Paulo Inacio da Costa; Rafael dos Santos Bezerra; Raquel de Lello Rocha Campos Cassano. NGS Soluções Genômicas: Pilar Drummond Sampaio Corrêa Mariani. FZEA-USP Pirassununga: Mirele Daiana Poleti; Raul Machado Neto; Ricardo Augusto Brassaloti; Ricardo Haddad; Rodrigo Tocantins Calado. FAMERP-SJRP: Cecília Artico Banho; Sandra Coccuzzo Sampaio; Svetoslav Nanev Slavov; Vagner Fonseca; Vincent Louis Viala |
| EPI_ISL_1966315 | PRONTO SOCORRO DE AGENOR DE CAMPOS MONGAGUA | Instituto Butantan / Mendelics | Antonio Jorge Martins; Bianca Cechetto Carlos. Mendelics: Bibiana Santos; Claudia Renata dos Santos Barros; Cíntia Bittar; David Schlesinger. Hemocentro Ribeirão Preto: Simone Kashima; Debora Botequio Moretti; Elaine Cristina Marqueze; Elaine Vieira dos Santos; Elisangela Chicaroni Mattos; Erika Freitas; Evandra Strazza Rodrigues; Felipe Allan da Silva da Costa; Flavia Aburjaile; Fábio Sossai Possebon; Guilherme Campos; Guilherme Targino Valente; Heidge Fukumasu. USP-Botucatu: Rejane Maria Tommasini Grotto; Helena Lage Ferreira; Instituto Butantan: Dimas Tadeu Covas; Jardelina de Souza Todao Bernardino; Jayme A. Souza-Neto; Jessika Cristina Chagas Lesbon; Jorge A. Petroli Marchesi; José Salvatore Leister Patané; João Paulo Kitajima; João Pessoa Araújo Jr.; Leila Sabrina Ullmann; Loyze Paola Oliveira de Lima; Luiz Aurelio de Campos Crispin. Centro de Genômica Funcional da ESALQ: Luiz Lehmann Coutinho; Luiz Carlos Junior de Alcantara; Lívia Sacchetto; Maisa C. Pereira Parra; Maria Carolina Elias; Marta Giovanetti; Marília Moraes; Maurício Lacerda Nogueira. Prefeitura de Sao Paulo: Melissa Palmieri.; Patricia Akemi Assato; Paula Rahal; Paulo Inacio da Costa; Rafael dos Santos Bezerra; Raquel de Lello Rocha Campos Cassano. NGS Soluções Genômicas: Pilar Drummond Sampaio Corrêa Mariani. FZEA-USP Pirassununga: Mirele Daiana Poleti; Raul Machado Neto; Ricardo Augusto Brassaloti; Ricardo Haddad; Rodrigo Tocantins Calado. FAMERP-SJRP: Cecília Artico Banho; Sandra Coccuzzo Sampaio; Svetoslav Nanev Slavov; Vagner Fonseca; Vincent Louis Viala |
| EPI_ISL_2345904 | PRONTO SOCORRO JARDIM JACIRA | Instituto Butantan / FZEA-USP-Pirassununga | Antonio Jorge Martins; Claudia Renata dos Santos Barros; David Schlesinger; Debora Botequio Moretti; Dimas Tadeu Covas; Elaine Cristina Marqueze; Elaine Vieira Santos; Evandra Strazza Rodrigues; Heidge Fukumasu; Jayme Augusto de Souza-Neto; José Salvatore Leister Patané; Luiz Alcantara; Luiz Lehmann Coutinho; Maria Carolina Elias; Maurício Lacerda Nogueira; Rafael dos Santos Bezerra; Raul Machado Neto; Rejane Maria Tommasini Grotto; Ricardo Haddad; Sandra Coccuzzo Sampaio Vessoni; Simone Kashima; Svetoslav Nanev Slavov; Vincent Louis Viala |
| EPI_ISL_1795214, EPI_ISL_2345442 | PRONTO SOCORRO MUNICIPAL DE SEVERINIA | Instituto Butantan / ESALQ-Piracicaba | Antonio Jorge Martins; Bianca Cechetto Carlos. Mendelics: Bibiana Santos; Claudia Renata dos Santos Barros; David Schlesinger; David Schlesinger. Hemocentro Ribeirão Preto: Simone Kashima; Debora Botequio Moretti; Debora Botequio Moretti. Centro de Genômica Funcional da ESALQ: Luiz Lehmann Coutinho; Dimas Tadeu Covas; Elaine Cristina Marqueze; Elaine Vieira Santos; Elaine Vieira dos Santos; Elisangela Chicaroni Mattos; Erika Freitas; Evandra Strazza Rodrigues; Felipe Allan da Silva da Costa; Flavia Aburjaile; Guilherme Targino Valente; Heidge Fukumasu; Heidge Fukumasu. USP-Botucatu: Rejane Maria Tommasini Grotto; Instituto Butantan: Alexander Roberto Precioso; Jayme A. Souza-Neto; Jayme Augusto de Souza-Neto; Jessika Cristina Chagas Lesbon; José Salvatore Leister Patané; João Paulo Kitajima; Luiz Alcantara; Luiz Carlos Junior de Alcantara; Luiz Lehmann Coutinho; Maria Carolina Elias; Marta Giovanetti; Maurício Lacerda Nogueira; Patricia Akemi Assato; Rafael dos Santos Bezerra; Raquel de Lello Rocha Campos Cassano. NGS Soluções Genômicas: Pilar Drummond Sampaio Corrêa Mariani. FZEA-USP Pirassununga: Mirele Daiana Poleti; Raul Machado Neto; Rejane Maria Tommasini Grotto; Ricardo Augusto Brassaloti; Ricardo Haddad; Rodrigo Tocantins Calado.; Sandra Coccuzzo Sampaio; Sandra Coccuzzo Sampaio Vessoni; Simone Kashima; Svetoslav Nanev Slavov; Vagner Fonseca; Vincent Louis Viala |
| EPI_ISL_1966508 | PRONTO SOCORRO MUNICIPAL DE SEVERINIA | Instituto Butantan / Mendelics | Antonio Jorge Martins; Bianca Cechetto Carlos. Mendelics: Bibiana Santos; Claudia Renata dos Santos Barros; Cíntia Bittar; David Schlesinger. Hemocentro Ribeirão Preto: Simone Kashima; Debora Botequio Moretti; Elaine Cristina Marqueze; Elaine Vieira dos Santos; Elisangela Chicaroni Mattos; Erika Freitas; Evandra Strazza Rodrigues; Felipe Allan da Silva da Costa; Flavia Aburjaile; Fábio Sossai Possebon; Guilherme Campos; Guilherme Targino Valente; Heidge Fukumasu. USP-Botucatu: Rejane Maria Tommasini Grotto; Helena Lage Ferreira; Instituto Butantan: Dimas Tadeu Covas; Jardelina de Souza Todao Bernardino; Jayme A. Souza-Neto; Jessika Cristina Chagas Lesbon; Jorge A. Petroli Marchesi; José Salvatore Leister Patané; João Paulo Kitajima; João Pessoa Araújo Jr.; Leila Sabrina Ullmann; Loyze Paola Oliveira de Lima; Luiz Aurelio de Campos Crispin. Centro de Genômica Funcional da ESALQ: Luiz Lehmann Coutinho; Luiz Carlos Junior de Alcantara; Lívia Sacchetto; Maisa C. Pereira Parra; Maria Carolina Elias; Marta Giovanetti; Marília Moraes; Maurício Lacerda Nogueira. Prefeitura de Sao Paulo: Melissa Palmieri.; Patricia Akemi Assato; Paula Rahal; Paulo Inacio da Costa; Rafael dos Santos Bezerra; Raquel de Lello Rocha Campos Cassano. NGS Soluções Genômicas: Pilar Drummond Sampaio Corrêa Mariani. FZEA-USP Pirassununga: Mirele Daiana Poleti; Raul Machado Neto; Ricardo Augusto Brassaloti; Ricardo Haddad; Rodrigo Tocantins Calado. FAMERP-SJRP: Cecília Artico Banho; Sandra Coccuzzo Sampaio; Svetoslav Nanev Slavov; Vagner Fonseca; Vincent Louis Viala |
| EPI_ISL_1795208, EPI_ISL_1795209, EPI_ISL_1795300, EPI_ISL_1795301, EPI_ISL_1795302, EPI_ISL_1795304, EPI_ISL_2345435, EPI_ISL_2345436, EPI_ISL_2345543, EPI_ISL_2345545, EPI_ISL_2345548, EPI_ISL_2345550 | | | |
| see above | PRONTO SOCORRO MUNICIPAL TAMBAU | Instituto Butantan / ESALQ-Piracicaba | Antonio Jorge Martins; Bianca Cechetto Carlos. Mendelics: Bibiana Santos; Claudia Renata dos Santos Barros; David Schlesinger; David Schlesinger. Hemocentro Ribeirão Preto: Simone Kashima; Debora Botequio Moretti; Debora Botequio Moretti. Centro de Genômica Funcional da ESALQ: Luiz Lehmann Coutinho; Dimas Tadeu Covas; Elaine Cristina Marqueze; Elaine Vieira Santos; Elaine Vieira dos Santos; Elisangela Chicaroni Mattos; Erika Freitas; Evandra Strazza Rodrigues; Felipe Allan da Silva da Costa; Flavia Aburjaile; Guilherme Targino Valente; Heidge Fukumasu; Heidge Fukumasu. USP-Botucatu: Rejane Maria Tommasini Grotto; Instituto Butantan: Alexander Roberto Precioso; Jayme A. Souza-Neto; Jayme Augusto de Souza-Neto; Jessika Cristina Chagas Lesbon; José Salvatore Leister Patané; João Paulo Kitajima; Luiz Alcantara; Luiz Carlos Junior de Alcantara; Luiz Lehmann Coutinho; Maria Carolina Elias; Marta Giovanetti; Maurício Lacerda Nogueira; Patricia Akemi Assato; Rafael dos Santos Bezerra; Raquel de Lello Rocha Campos Cassano. NGS Soluções Genômicas: Pilar Drummond Sampaio Corrêa Mariani. FZEA-USP Pirassununga: Mirele Daiana Poleti; Raul Machado Neto; Rejane Maria Tommasini Grotto; Ricardo Augusto Brassaloti; Ricardo Haddad; Rodrigo Tocantins Calado.; Sandra Coccuzzo Sampaio; Sandra Coccuzzo Sampaio Vessoni; Simone Kashima; Svetoslav Nanev Slavov; Vagner Fonseca; Vincent Louis Viala |
| EPI_ISL_1966736 | PRONTO SOCORRO PESTANA P S ANDRE SACCO | Instituto Butantan | Antonio Jorge Martins; Bianca Cechetto Carlos. Mendelics: Bibiana Santos; Claudia Renata dos Santos Barros; Cíntia Bittar; David Schlesinger. Hemocentro Ribeirão Preto: Simone Kashima; Debora Botequio Moretti; Elaine Cristina Marqueze; Elaine Vieira dos Santos; Elisangela Chicaroni Mattos; Erika Freitas; Evandra Strazza Rodrigues; Felipe Allan da Silva da Costa; Flavia Aburjaile; Fábio Sossai Possebon; Guilherme Campos; Guilherme Targino Valente; Heidge Fukumasu. USP-Botucatu: Rejane Maria Tommasini Grotto; Helena Lage Ferreira; Instituto Butantan: Dimas Tadeu Covas; Jardelina de Souza Todao Bernardino; Jayme A. Souza-Neto; Jessika Cristina Chagas Lesbon; Jorge A. Petroli Marchesi; José Salvatore Leister Patané; João Paulo Kitajima; João Pessoa Araújo Jr.; Leila Sabrina Ullmann; Loyze Paola Oliveira de Lima; Luiz Aurelio de Campos Crispin. Centro de Genômica Funcional da ESALQ: Luiz Lehmann Coutinho; Luiz Carlos Junior de Alcantara; Lívia Sacchetto; Maisa C. Pereira Parra; Maria Carolina Elias; Marta Giovanetti; Marília Moraes; Maurício Lacerda Nogueira. Prefeitura de Sao Paulo: Melissa Palmieri.; Patricia Akemi Assato; Paula Rahal; Paulo Inacio da Costa; Rafael dos Santos Bezerra; Raquel de Lello Rocha Campos Cassano. NGS Soluções Genômicas: Pilar Drummond Sampaio Corrêa Mariani. FZEA-USP Pirassununga: Mirele Daiana Poleti; Raul Machado Neto; Ricardo Augusto Brassaloti; Ricardo Haddad; Rodrigo Tocantins Calado. FAMERP-SJRP: Cecília Artico Banho; Sandra Coccuzzo Sampaio; Svetoslav Nanev Slavov; Vagner Fonseca; Vincent Louis Viala |
| EPI_ISL_2614568 | PS Jose Ibrahin | Instituto Adolfo Lutz, Interdiciplinary Procedures Center, Strategic Laboratory | Caio Vinicius Dias Lopes; Claudia Regina Gonçalves; Claudio Tavares Sacchi; Erica Valessa Ramos Gomes; Karoline Rodrigues Campos; Leonardo Jose Tadeu de Araujo |
| EPI_ISL_2614576, EPI_ISL_2614579 | PS Municipal Dr Alceu Lot | Instituto Adolfo Lutz, Interdiciplinary Procedures Center, Strategic Laboratory | Caio Vinicius Dias Lopes; Claudia Regina Gonçalves; Claudio Tavares Sacchi; Erica Valessa Ramos Gomes; Karoline Rodrigues Campos; Leonardo Jose Tadeu de Araujo |
| EPI_ISL_2209425 | PSF DR ANTONIO PIRES DE ALMEIDA PORTO FELIZ | Instituto Butantan | Antonio Jorge Martins; Claudia Renata dos Santos Barros; David Schlesinger; Debora Botequio Moretti; Dimas Tadeu Covas; Elaine Cristina Marqueze; Elaine Vieira Santos; Evandra Strazza Rodrigues; Heidge Fukumasu; Jayme Augusto de Souza-Neto; José Salvatore Leister Patané; Luiz Alcantara; Luiz Lehmann Coutinho; Maria Carolina Elias; Maurício Lacerda Nogueira; Rafael dos Santos Bezerra; Raul Machado Neto; Rejane Maria Tommasini Grotto; Ricardo Haddad; Sandra Coccuzzo Sampaio Vessoni; Simone Kashima; Svetoslav Nanev Slavov; Vincent Louis Viala. |
| EPI_ISL_1967020, EPI_ISL_1967021, EPI_ISL_1967022, EPI_ISL_1967023, EPI_ISL_1967024 | PSF DR ANTONIO PIRES DE ALMEIDA PORTO FELIZ | Instituto Butantan / ESALQ-USP (Piracicaba) | Antonio Jorge Martins; Bianca Cechetto Carlos. Mendelics: Bibiana Santos; Claudia Renata dos Santos Barros; Cíntia Bittar; David Schlesinger. Hemocentro Ribeirão Preto: Simone Kashima; Debora Botequio Moretti; Elaine Cristina Marqueze; Elaine Vieira dos Santos; Elisangela Chicaroni Mattos; Erika Freitas; Evandra Strazza Rodrigues; Felipe Allan da Silva da Costa; Flavia Aburjaile; Fábio Sossai Possebon; Guilherme Campos; Guilherme Targino Valente; Heidge Fukumasu. USP-Botucatu: Rejane Maria Tommasini Grotto; Helena Lage Ferreira; Instituto Butantan: Dimas Tadeu Covas; Jardelina de Souza Todao Bernardino; Jayme A. Souza-Neto; Jessika Cristina Chagas Lesbon; Jorge A. Petroli Marchesi; José Salvatore Leister Patané; João Paulo Kitajima; João Pessoa Araújo Jr.; Leila Sabrina Ullmann; Loyze Paola Oliveira de Lima; Luiz Aurelio de Campos Crispin. Centro de Genômica Funcional da ESALQ: Luiz Lehmann Coutinho; Luiz Carlos Junior de Alcantara; Lívia Sacchetto; Maisa C. Pereira Parra; Maria Carolina Elias; Marta Giovanetti; Marília Moraes; Maurício Lacerda Nogueira. Prefeitura de Sao Paulo: Melissa Palmieri.; Patricia Akemi Assato; Paula Rahal; Paulo Inacio da Costa; Rafael dos Santos Bezerra; Raquel de Lello Rocha Campos Cassano. NGS Soluções Genômicas: Pilar Drummond Sampaio Corrêa Mariani. FZEA-USP Pirassununga: Mirele Daiana Poleti; Raul Machado Neto; Ricardo Augusto Brassaloti; Ricardo Haddad; Rodrigo Tocantins Calado. FAMERP-SJRP: Cecília Artico Banho; Sandra Coccuzzo Sampaio; Svetoslav Nanev Slavov; Vagner Fonseca; Vincent Louis Viala |
| EPI_ISL_2344584 | PSF DR ANTONIO PIRES DE ALMEIDA PORTO FELIZ | Instituto Butantan / UNESP-Botucatu | Antonio Jorge Martins; Claudia Renata dos Santos Barros; David Schlesinger; Debora Botequio Moretti; Dimas Tadeu Covas; Elaine Cristina Marqueze; Elaine Vieira Santos; Evandra Strazza Rodrigues; Heidge Fukumasu; Jayme Augusto de Souza-Neto; José Salvatore Leister Patané; Luiz Alcantara; Luiz Lehmann Coutinho; Maria Carolina Elias; Maurício Lacerda Nogueira; Rafael dos Santos Bezerra; Raul Machado Neto; Rejane Maria Tommasini Grotto; Ricardo Haddad; Sandra Coccuzzo Sampaio Vessoni; Simone Kashima; Svetoslav Nanev Slavov; Vincent Louis Viala |
| EPI_ISL_2209433 | PSF MARIA JOSE SALTO DE PIRAPORA | Instituto Butantan | Antonio Jorge Martins; Claudia Renata dos Santos Barros; David Schlesinger; Debora Botequio Moretti; Dimas Tadeu Covas; Elaine Cristina Marqueze; Elaine Vieira Santos; Evandra Strazza Rodrigues; Heidge Fukumasu; Jayme Augusto de Souza-Neto; José Salvatore Leister Patané; Luiz Alcantara; Luiz Lehmann Coutinho; Maria Carolina Elias; Maurício Lacerda Nogueira; Rafael dos Santos Bezerra; Raul Machado Neto; Rejane Maria Tommasini Grotto; Ricardo Haddad; Sandra Coccuzzo Sampaio Vessoni; Simone Kashima; Svetoslav Nanev Slavov; Vincent Louis Viala. |
| EPI_ISL_1967033, | PSF MARIA JOSE | Instituto Butantan / | Antonio Jorge Martins; Bianca Cechetto Carlos. Mendelics: Bibiana Santos; Claudia Renata dos Santos Barros; Cíntia Bittar; David Schlesinger. Hemocentro Ribeirão Preto: Simone Kashima; Debora Botequio Moretti; Elaine Cristina Marqueze; Elaine Vieira dos Santos; Elisangela Chicaroni Mattos; Erika Freitas; Evandra Strazza Rodrigues; |

| | | | |
|---|---|---|---|
| EPI_ISL_1967034, EPI_ISL_1967037, EPI_ISL_1967038, EPI_ISL_1967039 | SALTO DE PIRAPORA | ESALQ-USP (Piracicaba) | Felipe Allan da Silva da Costa; Flavia Aburjaile; Fábio Sossai Possebon; Guilherme Campos; Guilherme Targino Valente; Heidge Fukumasu. USP-Botucatu: Rejane Maria Tommasini Grotto; Helena Lage Ferreira; Instituto Butantan: Dimas Tadeu Covas; Jardelina de Souza Todao Bernardino; Jayme A. Souza-Neto; Jessika Cristina Chagas Lesbon; Jorge A. Petroli Marchesi; José Salvatore Leister Patané; João Paulo Kitajima; João Pessoa Araújo Jr.; Leila Sabrina Ullmann; Loyze Paola Oliveira de Lima; Luiz Aurelio de Campos Crispin. Centro de Genômica Funcional da ESALQ: Luiz Lehmann Coutinho; Luiz Carlos Junior de Alcantara; Lívia Sacchetto; Maisa C. Pereira Parra; Maria Carolina Elias; Marta Giovanetti; Marília Moraes; Maurício Lacerda Nogueira. Prefeitura de Sao Paulo: Melissa Palmieri.; Patricia Akemi Assato; Paula Rahal; Paulo Inacio da Costa; Rafael dos Santos Bezerra; Raquel de Lello Rocha Campos Cassano. NGS Soluções Genômicas: Pilar Drummond Sampaio Corrêa Mariani. FZEA-USP Pirassununga: Mirele Daiana Poleti; Raul Machado Neto; Ricardo Augusto Brassaloti; Ricardo Haddad; Rodrigo Tocantins Calado. FAMERP-SJRP: Cecília Artico Banho; Sandra Coccuzzo Sampaio; Svetoslav Nanev Slavov; Vagner Fonseca; Vincent Louis Viala |
| EPI_ISL_2344659 | PSF MARIA JOSE SALTO DE PIRAPORA | Instituto Butantan / UNESP-Botucatu | Antonio Jorge Martins; Claudia Renata dos Santos Barros; David Schlesinger; Debora Botequio Moretti; Dimas Tadeu Covas; Elaine Cristina Marqueze; Elaine Vieira Santos; Evandra Strazza Rodrigues; Heidge Fukumasu; Jayme Augusto de Souza-Neto; José Salvatore Leister Patané; Luiz Alcantara; Luiz Lehmann Coutinho; Maria Carolina Elias; Maurício Lacerda Nogueira; Rafael dos Santos Bezerra; Raul Machado Neto; Rejane Maria Tommasini Grotto; Ricardo Haddad; Sandra Coccuzzo Sampaio Vessoni; Simone Kashima; Svetoslav Nanev Slavov; Vincent Louis Viala |
| EPI_ISL_2345960, EPI_ISL_2346010, EPI_ISL_2346021, EPI_ISL_2346032 | PSF PARQUE EMILIA | Instituto Butantan | Antonio Jorge Martins; Claudia Renata dos Santos Barros; David Schlesinger; Debora Botequio Moretti; Dimas Tadeu Covas; Elaine Cristina Marqueze; Elaine Vieira Santos; Evandra Strazza Rodrigues; Heidge Fukumasu; Jayme Augusto de Souza-Neto; José Salvatore Leister Patané; Luiz Alcantara; Luiz Lehmann Coutinho; Maria Carolina Elias; Maurício Lacerda Nogueira; Rafael dos Santos Bezerra; Raul Machado Neto; Rejane Maria Tommasini Grotto; Ricardo Haddad; Sandra Coccuzzo Sampaio Vessoni; Simone Kashima; Svetoslav Nanev Slavov; Vincent Louis Viala |
| EPI_ISL_1966786 | PSF SETOR LESTE NHANDEARA | Instituto Butantan / Mendelics | Antonio Jorge Martins; Bianca Cechetto Carlos. Mendelics: Bibiana Santos; Claudia Renata dos Santos Barros; Cíntia Bittar; David Schlesinger. Hemocentro Ribeirão Preto: Simone Kashima; Debora Botequio Moretti; Elaine Cristina Marqueze; Elaine Vieira dos Santos; Elisangela Chicaroni Mattos; Erika Freitas; Evandra Strazza Rodrigues; Felipe Allan da Silva da Costa; Flavia Aburjaile; Fábio Sossai Possebon; Guilherme Campos; Guilherme Targino Valente; Heidge Fukumasu. USP-Botucatu: Rejane Maria Tommasini Grotto; Helena Lage Ferreira; Instituto Butantan: Dimas Tadeu Covas; Jardelina de Souza Todao Bernardino; Jayme A. Souza-Neto; Jessika Cristina Chagas Lesbon; Jorge A. Petroli Marchesi; José Salvatore Leister Patané; João Paulo Kitajima; João Pessoa Araújo Jr.; Leila Sabrina Ullmann; Loyze Paola Oliveira de Lima; Luiz Aurelio de Campos Crispin. Centro de Genômica Funcional da ESALQ: Luiz Lehmann Coutinho; Luiz Carlos Junior de Alcantara; Lívia Sacchetto; Maisa C. Pereira Parra; Maria Carolina Elias; Marta Giovanetti; Marília Moraes; Maurício Lacerda Nogueira. Prefeitura de Sao Paulo: Melissa Palmieri.; Patricia Akemi Assato; Paula Rahal; Paulo Inacio da Costa; Rafael dos Santos Bezerra; Raquel de Lello Rocha Campos Cassano. NGS Soluções Genômicas: Pilar Drummond Sampaio Corrêa Mariani. FZEA-USP Pirassununga: Mirele Daiana Poleti; Raul Machado Neto; Ricardo Augusto Brassaloti; Ricardo Haddad; Rodrigo Tocantins Calado. FAMERP-SJRP: Cecília Artico Banho; Sandra Coccuzzo Sampaio; Svetoslav Nanev Slavov; Vagner Fonseca; Vincent Louis Viala |
| EPI_ISL_1385576, EPI_ISL_1385656, EPI_ISL_1385758, EPI_ISL_1385771, EPI_ISL_1470859, EPI_ISL_1470879, EPI_ISL_1470898, EPI_ISL_1471160, EPI_ISL_1471230, EPI_ISL_1471500, EPI_ISL_1471828, EPI_ISL_1542302, EPI_ISL_1542380, EPI_ISL_1542458, EPI_ISL_1542585, EPI_ISL_1542647, EPI_ISL_1542830, EPI_ISL_1542899, EPI_ISL_1543185, EPI_ISL_1543223, EPI_ISL_1543243, EPI_ISL_1543367, EPI_ISL_1543423, EPI_ISL_1543579, EPI_ISL_1543615, EPI_ISL_1543808, EPI_ISL_1543907, EPI_ISL_1633723, EPI_ISL_1633810, EPI_ISL_1633841, EPI_ISL_1633867, EPI_ISL_1633869, EPI_ISL_1633981, EPI_ISL_1634062, EPI_ISL_1634309, EPI_ISL_1635529, EPI_ISL_1635603, EPI_ISL_1635732, EPI_ISL_1635954, EPI_ISL_1636166, EPI_ISL_1636172, EPI_ISL_1717112, EPI_ISL_1717136, EPI_ISL_1717153, EPI_ISL_1717244, EPI_ISL_1717309, EPI_ISL_1717530, EPI_ISL_1717997 | | | |
| see above | Pandemic Response Lab - NYC | Pandemic Response Lab, R&D | Cybill del Castillo; Dylan Law; Haiping Hao; Henry Lee; Jon Laurent; Katharine Nelson; Melissa Hopkins; Michael Hammerling; Pradeep Bugga; Shinyoung Clair Kang; Sol Rey; William Ward |
| EPI_ISL_3982751, EPI_ISL_3982761, EPI_ISL_3982769, EPI_ISL_3982771 | Parana | LACEN PR | Guilherme Nardi Becker; Irina Nastassja Riediger; Maria do Carmo Debur Rossa; Mayra Presibella Giacomini; Nelson Quallio Marques |
| EPI_ISL_3982759, EPI_ISL_3982768 | Parana | Lacen-PR | Guilherme Nardi Becker; Irina Nastassja Riediger; Maria do Carmo Debur Rossa; Mayra Presibella Giacomini; Nelson Quallio Marques |
| EPI_ISL_3982749 | Parana | Lacen/PR | Guilherme Nardi Becker; Irina Nastassja Riediger; Maria do Carmo Debur Rossa; Mayra Presibella Giacomini; Nelson Quallio Marques |
| EPI_ISL_3982742, EPI_ISL_3982745, EPI_ISL_3982763 | Parana | Parana | Guilherme Nardi Becker; Irina Nastassja Riediger; Maria do Carmo Debur Rossa; Mayra Presibella Giacomini; Nelson Quallio Marques |
| EPI_ISL_792680, EPI_ISL_792681, EPI_ISL_792682, EPI_ISL_792683, EPI_ISL_1428639, EPI_ISL_1929419 | Pathogen Genomics Center, National Institute of Infectious Diseases | Pathogen Genomics Center, National Institute of Infectious Diseases | Hidemasa Izumiya; Ken-ichi Lee; Kentaro Itokawa; Makoto Kuroda; Masanori Hashino; Masatomo Morita; Nobuo Koizumi; Rina Tanaka; Shouji Yamamoto; Sunao Iyoda; Tsuyoshi Sekizuka |
| EPI_ISL_2993480 | Pathology, UAB | Pathology, UAB | E.J.; Hendrickson; Hernandez, J.; Leal; Lefkowitz; Moates, D.; R.C.; S.M. |
| EPI_ISL_2663257, EPI_ISL_2663260, EPI_ISL_2663262, EPI_ISL_2663263, EPI_ISL_2663265, EPI_ISL_2663266, EPI_ISL_2663270, EPI_ISL_2663272, EPI_ISL_2663273, EPI_ISL_2663274, EPI_ISL_2663275, EPI_ISL_2663276, EPI_ISL_2663278, EPI_ISL_2663282, EPI_ISL_2663283, EPI_ISL_2663285, EPI_ISL_2663287, EPI_ISL_2663288, EPI_ISL_2663289, EPI_ISL_2663291, EPI_ISL_2663292, EPI_ISL_2663294, EPI_ISL_2663295, EPI_ISL_2663296, EPI_ISL_2663297, EPI_ISL_2663298, EPI_ISL_2663299, EPI_ISL_2663300, EPI_ISL_2663301, EPI_ISL_2663302, EPI_ISL_2663303, EPI_ISL_2663304, EPI_ISL_2663305, EPI_ISL_2663306, EPI_ISL_2663307, EPI_ISL_2663308, EPI_ISL_2663309, EPI_ISL_2663310, EPI_ISL_2663312, EPI_ISL_2663313, EPI_ISL_2663315 | | | |
| see above | Plataforma de Vigilancia Molecular (PVM) - FIOCRUZ/BA | Plataforma de Vigilancia Molecular (PVM) - FIOCRUZ/BA | Bruno Bezerril Andrade; Camila I. de Oliveira on behalf of the Fiocruz COVID-19 Genomic Surveillance Network.; Clarissa Araújo Gurgel; Leonardo Paiva Farias; Marina Cucco; Ricardo Khouri; Tiago Graf |
| EPI_ISL_2629157, EPI_ISL_2629160, EPI_ISL_2629161, EPI_ISL_2629162 | Plateforme Covid IDF, site Broussais | HEGP - Laboratoire de Virologie | David Veyer; Hélène Péré; Julien Puech; Maxime Wack; Nabil Gastli; Nicolas Robillard; Sandrine Imbeaud |
| EPI_ISL_1498361 | Platform BIS UZA/UAntwerpen | Labo Klinische Biologie, UZA | Basil Britto Xavier; Christine Lammens; Herman Goossens; Jasmine Coppens; Marie Le Mercier; Veerle Matheeussen |
| EPI_ISL_1336265, EPI_ISL_1336269, EPI_ISL_1337455 | Platform BIS UZA/UAntwerpen | UAntwerp, Laboratory of Medical Microbiology | Basil Britto Xavier; Christine Lammens; Herman Goossens; Jasmine Coppens; Marie Le Mercier; Veerle Matheeussen |
| EPI_ISL_1533719 | Policlinica Munic da Est Turistica de Holambra | Instituto Adolfo Lutz, Interdiciplinary Procedures Center, Strategic Laboratory | Caio Vinicius Dias Lopes; Claudia Regina Gonçalves; Claudio Tavares Sacchi; Erica Valessa Ramos Gomes; Karoline Rodrigues Campos; Leonardo Jose Tadeu de Araujo |
| EPI_ISL_2344594, EPI_ISL_2344595, EPI_ISL_2344596, EPI_ISL_2344597, EPI_ISL_2344598, EPI_ISL_2344599, EPI_ISL_2344602, EPI_ISL_2344603, EPI_ISL_2344604, EPI_ISL_2344606, EPI_ISL_2344607, EPI_ISL_2344608, EPI_ISL_2344613, EPI_ISL_2344616, EPI_ISL_2344617, EPI_ISL_2344625, EPI_ISL_2344626, EPI_ISL_2344628, EPI_ISL_2344629, EPI_ISL_2344631, EPI_ISL_2344632, EPI_ISL_2344633, EPI_ISL_2344635, EPI_ISL_2344636, EPI_ISL_2344637, EPI_ISL_2344639, EPI_ISL_2344640, EPI_ISL_2344641, EPI_ISL_2344642, EPI_ISL_2344644, EPI_ISL_2344646, EPI_ISL_2344648, EPI_ISL_2344649, EPI_ISL_2344650, EPI_ISL_2344651, EPI_ISL_2344653, EPI_ISL_2345912, EPI_ISL_2345914, EPI_ISL_2345925, EPI_ISL_2345931, EPI_ISL_2346085, EPI_ISL_2346086, EPI_ISL_2346087, EPI_ISL_2346089, EPI_ISL_2346091, EPI_ISL_2346093 | | | |
| see above | Prefeirtura de SP | Instituto Butantan | Antonio Jorge Martins; Claudia Renata dos Santos Barros; David Schlesinger; Debora Botequio Moretti; Dimas Tadeu Covas; Elaine Cristina Marqueze; Elaine Vieira Santos; Evandra Strazza Rodrigues; Heidge Fukumasu; Jayme Augusto de Souza-Neto; José Salvatore Leister Patané; Luiz Alcantara; Luiz Lehmann Coutinho; Maria Carolina Elias; Maurício Lacerda Nogueira; Rafael dos Santos Bezerra; Raul Machado Neto; Rejane Maria Tommasini Grotto; Ricardo Haddad; Sandra Coccuzzo Sampaio Vessoni; Simone Kashima; Svetoslav Nanev Slavov; Vincent Louis Viala |
| EPI_ISL_1533714 | Pronto Socorro Dr Osmar Mesquita | Instituto Adolfo Lutz, Interdiciplinary Procedures Center, Strategic Laboratory | Caio Vinicius Dias Lopes; Claudia Regina Gonçalves; Claudio Tavares Sacchi; Erica Valessa Ramos Gomes; Karoline Rodrigues Campos; Leonardo Jose Tadeu de Araujo |
| EPI_ISL_2295734, EPI_ISL_2698785 | Public Health Laboratory, Minnesota Department of Health | University of Minnesota Genomics Center | Corbin Dirkx; Daryl M. Gohl; Jaquelyn Kuriger-Laber; John Garbe; and Sean Wang |
| EPI_ISL_2688283 | Puerto Rico Department of Health | Centers for Disease Control and Prevention, Dengue Branch | Betzabel Flores; Gabriela Paz-Bailey; Gilberto A. Santiago; Glenda Gonzalez; Jorge L. Munoz-Jordan; Keyla Charriez |
| EPI_ISL_1559414, EPI_ISL_1559585, EPI_ISL_1582010, EPI_ISL_1582020, EPI_ISL_1694689, EPI_ISL_1694691, EPI_ISL_1694693, EPI_ISL_1694950, EPI_ISL_1694952, EPI_ISL_1694953, EPI_ISL_1694954, EPI_ISL_1694955, EPI_ISL_1694956, EPI_ISL_1695037, EPI_ISL_1695039, EPI_ISL_1695050, EPI_ISL_1695051, EPI_ISL_1695052, EPI_ISL_1695053, EPI_ISL_1695056, EPI_ISL_1695061, EPI_ISL_1695063, EPI_ISL_1695069, EPI_ISL_1695071, EPI_ISL_1695072, EPI_ISL_1695074, EPI_ISL_1695075, EPI_ISL_1695079, EPI_ISL_1695081, EPI_ISL_1695082, EPI_ISL_1695083, EPI_ISL_1695086, EPI_ISL_1695088, EPI_ISL_1695090, EPI_ISL_1695092, EPI_ISL_1695093, EPI_ISL_1695094, EPI_ISL_1695095, EPI_ISL_1695096, EPI_ISL_1695097, EPI_ISL_1695098, EPI_ISL_1695099, EPI_ISL_1695100, EPI_ISL_1695101, EPI_ISL_1695103, EPI_ISL_1695104, EPI_ISL_1695105, EPI_ISL_1695106, EPI_ISL_1695110, EPI_ISL_1695619, EPI_ISL_1695622, EPI_ISL_1695625, EPI_ISL_1695628, EPI_ISL_1695631, EPI_ISL_1695634, EPI_ISL_1695637, EPI_ISL_1695640, EPI_ISL_1695643, EPI_ISL_1695650, EPI_ISL_1695653, EPI_ISL_1695659, EPI_ISL_1695665, EPI_ISL_1695668, EPI_ISL_1695674, EPI_ISL_1695677, EPI_ISL_1695690, EPI_ISL_1695696, EPI_ISL_1695699, EPI_ISL_1695702, EPI_ISL_1695705, EPI_ISL_1695708, EPI_ISL_1695714, EPI_ISL_1695717, EPI_ISL_1695721, EPI_ISL_1695724, EPI_ISL_1695727, EPI_ISL_1695729, EPI_ISL_1695732, EPI_ISL_1695739, EPI_ISL_1695741, EPI_ISL_1695744, EPI_ISL_1695747, EPI_ISL_1695750, EPI_ISL_1695753, EPI_ISL_1695756, EPI_ISL_1695759, EPI_ISL_1695762, EPI_ISL_1695766, EPI_ISL_1695769, EPI_ISL_1702163, EPI_ISL_1702164, EPI_ISL_1702165, EPI_ISL_1702166, EPI_ISL_1702167, EPI_ISL_1702168, EPI_ISL_1702169, EPI_ISL_1702170, EPI_ISL_1702171, EPI_ISL_1702172, EPI_ISL_1702173, EPI_ISL_1702174, EPI_ISL_1702175, EPI_ISL_1702176, EPI_ISL_1702177, EPI_ISL_1702178, EPI_ISL_1711970, EPI_ISL_1711971, EPI_ISL_1711973, EPI_ISL_1753613, EPI_ISL_1753614, EPI_ISL_1753615, EPI_ISL_1753616, EPI_ISL_1753617, EPI_ISL_1753618, EPI_ISL_1753619, EPI_ISL_1753624, EPI_ISL_1753626, EPI_ISL_1753627, EPI_ISL_1753633, EPI_ISL_1753634, EPI_ISL_1753635, EPI_ISL_1753636, EPI_ISL_1753639, EPI_ISL_1753641, EPI_ISL_1753642, EPI_ISL_1753644, EPI_ISL_1753648, EPI_ISL_1753649, EPI_ISL_1753652, EPI_ISL_1753658, EPI_ISL_1753659, EPI_ISL_1753661, EPI_ISL_1761871, EPI_ISL_1761872, EPI_ISL_1761874, EPI_ISL_1761875, EPI_ISL_1761876, EPI_ISL_1761877, EPI_ISL_1761878, EPI_ISL_1761879, EPI_ISL_1761881, EPI_ISL_1761883, EPI_ISL_1761887, EPI_ISL_1761889, EPI_ISL_1761890, EPI_ISL_1761891, EPI_ISL_1761894, EPI_ISL_1761895, EPI_ISL_1804813, EPI_ISL_1804814, EPI_ISL_1804815, EPI_ISL_1804818, EPI_ISL_1804819, EPI_ISL_1818053, EPI_ISL_1818054, EPI_ISL_1818055, EPI_ISL_1818056, EPI_ISL_1818058, EPI_ISL_1818062, EPI_ISL_1818063, EPI_ISL_1818064, EPI_ISL_1818065, EPI_ISL_1818070, EPI_ISL_1818071, EPI_ISL_1900043, EPI_ISL_1900046, EPI_ISL_1900048, EPI_ISL_1900051, EPI_ISL_1900054, EPI_ISL_1900057, EPI_ISL_1900059, EPI_ISL_1900062, EPI_ISL_1900064, EPI_ISL_1900067, EPI_ISL_1900070, EPI_ISL_1900077, EPI_ISL_1900079, EPI_ISL_1900083, EPI_ISL_1900084, EPI_ISL_1900086, EPI_ISL_1900089, EPI_ISL_1900092, EPI_ISL_1900094, EPI_ISL_1900097, EPI_ISL_1900100, EPI_ISL_1900102, EPI_ISL_1900105, EPI_ISL_1900121, EPI_ISL_1900124, EPI_ISL_1900128, EPI_ISL_1900134, EPI_ISL_1900139, EPI_ISL_1900152, EPI_ISL_1900155, EPI_ISL_1900427, EPI_ISL_1900433, EPI_ISL_1900434, EPI_ISL_1900435, EPI_ISL_1900437, EPI_ISL_1900438, EPI_ISL_1900439, EPI_ISL_1900441, EPI_ISL_1900442, EPI_ISL_1900443, EPI_ISL_1900444, EPI_ISL_1900445, EPI_ISL_1900446, EPI_ISL_1900447, EPI_ISL_1900448, EPI_ISL_1900450, EPI_ISL_1900451, EPI_ISL_1900452, EPI_ISL_1900455, EPI_ISL_1900457, EPI_ISL_1900458, EPI_ISL_1900460, EPI_ISL_1900462, EPI_ISL_1900464, EPI_ISL_1935556, EPI_ISL_1935557, EPI_ISL_1935558, EPI_ISL_1935561, EPI_ISL_1935562, EPI_ISL_1935563, EPI_ISL_1935569, EPI_ISL_1935570, EPI_ISL_1935571, EPI_ISL_1935573, EPI_ISL_1935574, EPI_ISL_1935582, EPI_ISL_1935583, EPI_ISL_1942169, EPI_ISL_1942170, EPI_ISL_1942171, EPI_ISL_1942173, EPI_ISL_1942174, EPI_ISL_1942176, EPI_ISL_1942177, EPI_ISL_1942178, EPI_ISL_1942179, EPI_ISL_1942181, EPI_ISL_1942182, EPI_ISL_1942183, EPI_ISL_1942184, EPI_ISL_1942185, EPI_ISL_1942188, EPI_ISL_1942190, EPI_ISL_1942192, EPI_ISL_1942193, EPI_ISL_1942194, EPI_ISL_1942195, EPI_ISL_1942196, EPI_ISL_1942198, EPI_ISL_1942200, EPI_ISL_1942201, EPI_ISL_1942202, EPI_ISL_1942203, EPI_ISL_1942205, EPI_ISL_1942206, EPI_ISL_1942207, EPI_ISL_1942208, EPI_ISL_1942211, EPI_ISL_1942213, EPI_ISL_1942214, EPI_ISL_1942215, EPI_ISL_1942216, EPI_ISL_1989980, EPI_ISL_1989985, EPI_ISL_1989991, EPI_ISL_1989992, EPI_ISL_1989993, EPI_ISL_1989995, EPI_ISL_1989996, EPI_ISL_1989997, EPI_ISL_1989998, EPI_ISL_1990000, EPI_ISL_1990001, EPI_ISL_1990002, EPI_ISL_1990003, EPI_ISL_2034714, EPI_ISL_2034716, EPI_ISL_2034722, EPI_ISL_2034729, EPI_ISL_2034732, EPI_ISL_2034739, EPI_ISL_2034742, EPI_ISL_2034746, EPI_ISL_2034749, EPI_ISL_2034751, EPI_ISL_2034755, EPI_ISL_2034762, EPI_ISL_2034766, EPI_ISL_2034769, EPI_ISL_2034779, EPI_ISL_2034785, EPI_ISL_2034789, EPI_ISL_2034793, EPI_ISL_2034811, EPI_ISL_2034825, EPI_ISL_2034833, EPI_ISL_2035308, EPI_ISL_2035327, EPI_ISL_2035328, EPI_ISL_2035332, EPI_ISL_2035333, EPI_ISL_2035339, EPI_ISL_2035341, EPI_ISL_2035342, EPI_ISL_2035345, EPI_ISL_2132580, EPI_ISL_2140759, EPI_ISL_2140761, EPI_ISL_2140762, EPI_ISL_2140765, EPI_ISL_2140771, EPI_ISL_2140772, EPI_ISL_2140773, EPI_ISL_2140794 | | | |
| see above | Quest Diagnostics Incorporated | Centers for Disease Control and Prevention Division of Viral Diseases, Pathogen Discovery | A. Gerasimova; A. Perez; Adrian Paskey; B. Anderson; Benjamin Rambo-Martin; Christopher Gulvick; Clinton R. Paden; Dakota Howard; Darlene Wagner; Dhwani Batra; Duncan MacCannell; F. Lacbawan; I. A. Shlyakhter; Jason Caravas; K.E. Livingston; Kara Moser; L.E. Bernstein; M. Hua; Matthew Schmerer; P. Tanpaiboon; Peter W. Cook; R. M. Kagan; R. Owen; R. V. Rolando; S. H. Rosenthal; Scott Sammons; Shatavia Morrison; Y. Liu; Yvette Unoarumhi |
| EPI_ISL_1798708, EPI_ISL_1798713, EPI_ISL_1798732, EPI_ISL_1798741, EPI_ISL_2018331, EPI_ISL_2018343 | RI State Health Laboratories | Centers for Disease Control and Prevention Division of Viral Diseases, Pathogen Discovery | Alison Laufer Halpin; Ben L. Rambo-Martin; Clinton R. Paden; Dakota Howard; Darlene Wagner; Dave Wentworth; Dhwani Batra; Jasmine Padilla; Justin Lee; Katie Dillon; Krista Queen; Kristen Knipe; Kristine Lacek; Mark Burroughs; Matthew Schmerer; Mili Sheth; Peter Cook; Sam Shepard; Sarah Nobles; Shoshona Le; Suxiang Tong; Vivien Dugan; Yvette Unoarumhi |
| EPI_ISL_2628503 | Rady Children's Hospital - San Diego | Andersen lab at Scripps Research | Christina Clarke; David Dimmock; Denise Malicki; Kathryn Bouic; Linda Luo; SEARCH Alliance San Diego with Nanda Radamchar; Teresa Mueller |

| | | | |
|---|---|---|---|
| EPI_ISL_1374280 | Randox Laboratories | Wellcome Sanger Institute for the COVID-19 Genomics UK (COG-UK) Consortium | Cordelia Langford; David K. Jackson; Dominic Kwiatkowski; Ewan Harrison; Ian Johnston; Jeffrey Barrett; John Sillitoe on behalf of the Wellcome Sanger Institute COVID-19 Surveillance Team; Randox Laboratories and Alex Alderton; Roberto Amato; Sonia Goncalves |
| EPI_ISL_1580755, EPI_ISL_1580758, EPI_ISL_1580768, EPI_ISL_1580775, EPI_ISL_1580781, EPI_ISL_1580789, EPI_ISL_1580790, EPI_ISL_1580796, EPI_ISL_2249922, EPI_ISL_2249934, EPI_ISL_2249945, EPI_ISL_2249948, EPI_ISL_2249953, EPI_ISL_2249967, EPI_ISL_2249984, EPI_ISL_2249998, EPI_ISL_2250004, EPI_ISL_2319634, EPI_ISL_2319636, EPI_ISL_2319664, EPI_ISL_2319674, EPI_ISL_2319682, EPI_ISL_2319683, EPI_ISL_2319685, EPI_ISL_2319696, EPI_ISL_2319701, EPI_ISL_2319715, EPI_ISL_2319738, EPI_ISL_2319756, EPI_ISL_2319757, EPI_ISL_2319766, EPI_ISL_2319805, EPI_ISL_2319810, EPI_ISL_2474142, EPI_ISL_2662115, EPI_ISL_2662128, EPI_ISL_2662131, EPI_ISL_2662132, EPI_ISL_2662137, EPI_ISL_2662145, EPI_ISL_2662154, EPI_ISL_2662159, EPI_ISL_2662164, EPI_ISL_2662166, EPI_ISL_2662168 | | | |
| see above | Reditus Laboratories | Reditus Laboratories | Alexa Eichelberger; Cassy Philips; Joshua J. Geltz; M.S.; Ph.D.; Rex Dyer; Robert M. Sgambelluri |
| EPI_ISL_1976763 | Regional Virus Laboratory, Belfast Health and Social Care Trust | COVID-19 Genomics UK (COG-UK) Consortium | Alison Watt; Ciara Cox; Conall McCaughey; David Simpson; Derek Fairley; James McKenna; Mairead Connor; Susan Feeney; Tanya Curran; Zoltan Molnar |
| EPI_ISL_2293423 | Research | National Reference Center for Viruses of Respiratory Infections, Institut Pasteur, Paris | Angela Brisebarre; Camille Capel; Christophe Malabat; Corinne Maufrais; Etienne Simon-Lorière; Frédéric Lemoine; Louise Lefrançois; Marianne Leruez-Ville; Marion Barbet; Maud Vanpeene; Méline Bizard; Sylvie Behillil; Sylvie Van der Werf; Vincent Enouf |
| EPI_ISL_3776837 | Respiratory Virus Unit, Microbiology Services Colindale, Public Health England | COVID-19 Genomics UK (COG-UK) Consortium | PHE Covid Sequencing Team |
| EPI_ISL_2253143, EPI_ISL_2253148, EPI_ISL_2253152, EPI_ISL_2253155, EPI_ISL_2253157, EPI_ISL_2253163, EPI_ISL_2254335, EPI_ISL_2254419, EPI_ISL_2254436, EPI_ISL_2254469 | | | |
| see above | Respiratory Viruses Branch, Centers for Disease Control and Prevention | Respiratory Viruses Branch, Centers for Disease Control and Prevention | Anderson, B.; Batra, D.; Bernstein, L.; C.R.; Campbell, P.; Caravas, J.; Case, R.; Clark, C.; Cook; Gerasimova, A.; Ghorpade, V.; Gulvick, C.; Hardison, M.; Houdeshell, H.; Howard, D.; Hua, M.; I.A.; Kagan; Kvalvaag, O.; Lacbawan, F.; Liu, Y.; Livingston, K.; MacCannell, D.; Morrison, S.; Moser, K.; Nall, D.; Owen, R.; P.W.; Paden; Paskey, A.; Perez, A.; R.M.; R.V.; Rambo-Martin, B.; Rolando; Rosenthal; S.H.; Sammons, S.; Sanders, E.; Schmerer, M.; Shlyakhter; Tanpaiboon, P.; Unoarumhi, Y.; Vest, A.; Wagner, D.; Westlund, S. |
| EPI_ISL_1578349, EPI_ISL_1710191, EPI_ISL_1710230, EPI_ISL_1919142, EPI_ISL_1919165, EPI_ISL_1919167, EPI_ISL_2016507, EPI_ISL_2133632 | | | |
| see above | Rhode Island Department of Health | Infectious Disease Program, Broad Institute of Harvard and MIT | Adams, G.; Azevedo, K.; B.L.; B.W.; Bauer, M.; Birren; Carter, A.; Chaluvadi, S.; D.J.; DeRuff, K.; Gladden-Young, A.; Huard, R.; J.E.; K.J.; King, E.; Lagerborg, K.; Lemieux; Loreth, C.; Miller, A.; Normandin, E.; P.C.; Park; Pearlman, L.; Reilly, S.; Rudy, M.; Sabeti; Siddle; Tomkins-Tinch, C.; and MacInnis |
| EPI_ISL_2603858 | Rhode Island State Health Laboratory | Rhode Island State Health Laboratory | Ewa King; Kristin Carpenter-Azevedo; Richard C. Huard |
| EPI_ISL_3982694, EPI_ISL_3982698, EPI_ISL_3982700, EPI_ISL_3982704, EPI_ISL_3982708, EPI_ISL_3982715 | Ribeirao Preto | Laboratorio de Biologia Molecular Hemocentro de Ribeirao Preto | Antonio Jorge Martins; Claudia Renata dos Santos Barros; David Schlesinger; Debora Botequio Moretti; Dimas Tadeu Covas; Elaine Cristina Marqueze; Elaine Vieira Santos; Evandra Strazza Rodrigues; Heidge Fukumasu; Jayme Augusto de Souza-Neto; José Salvatore Leister Patané; Luiz Alcantara; Luiz Lehmann Coutinho; Maria Carolina Elias; Maurício Lacerda Nogueira; Rafael dos Santos Bezerra; Raul Machado Neto; Rejane Maria Tommasini Grotto; Ricardo Haddad; Sandra Coccuzzo Sampaio Vessoni; Simone Kashima; Svetoslav Nanev Slavov; Vincent Louis Viala |
| EPI_ISL_2344739, EPI_ISL_2344742, EPI_ISL_2344745 | SAE CTA SERV ATEND ESPEC TESTAGEM E ACONSELHAMENTO | Instituto Butantan / ESALQ-Piracicaba | Antonio Jorge Martins; Claudia Renata dos Santos Barros; David Schlesinger; Debora Botequio Moretti; Dimas Tadeu Covas; Elaine Cristina Marqueze; Elaine Vieira Santos; Evandra Strazza Rodrigues; Heidge Fukumasu; Jayme Augusto de Souza-Neto; José Salvatore Leister Patané; Luiz Alcantara; Luiz Lehmann Coutinho; Maria Carolina Elias; Maurício Lacerda Nogueira; Rafael dos Santos Bezerra; Raul Machado Neto; Rejane Maria Tommasini Grotto; Ricardo Haddad; Sandra Coccuzzo Sampaio Vessoni; Simone Kashima; Svetoslav Nanev Slavov; Vincent Louis Viala |
| EPI_ISL_1966126, EPI_ISL_1966128, EPI_ISL_1966518 | SAE SERVICO DE ATENDIMENTO ESPECIALIZADO | Instituto Butantan / Mendelics | Antonio Jorge Martins; Bianca Cechetto Carlos. Mendelics: Bibiana Santos; Claudia Renata dos Santos Barros; Cíntia Bittar; David Schlesinger. Hemocentro Ribeirão Preto: Simone Kashima; Debora Botequio Moretti; Elaine Cristina Marqueze; Elaine Vieira dos Santos; Elisangela Chicaroni Mattos; Erika Freitas; Evandra Strazza Rodrigues; Felipe Allan da Silva da Costa; Flavia Aburjaile; Fábio Sossai Possebon; Guilherme Campos; Guilherme Targino Valente; Heidge Fukumasu. USP-Botucatu: Rejane Maria Tommasini Grotto; Helena Lage Ferreira; Instituto Butantan: Dimas Tadeu Covas; Jardelina de Souza Todao Bernardino; Jayme A. Souza-Neto; Jessika Cristina Chagas Lesbon; Jorge A. Petroli Marchesi; José Salvatore Leister Patané; João Paulo Kitajima; João Pessoa Araújo Jr.; Leila Sabrina Ullmann; Loyze Paola Oliveira de Lima; Luiz Aurelio de Campos Crispin. Centro de Genômica Funcional da ESALQ: Luiz Lehmann Coutinho; Luiz Carlos Junior de Alcantara; Lívia Sacchetto; Maisa C. Pereira Parra; Maria Carolina Elias; Marta Giovanetti; Marília Moraes; Maurício Lacerda Nogueira. Prefeitura de Sao Paulo: Melissa Palmieri.; Patricia Akemi Assato; Paula Rahal; Paulo Inacio da Costa; Rafael dos Santos Bezerra; Raquel de Lello Rocha Campos Cassano. NGS Soluções Genômicas: Pilar Drummond Sampaio Corrêa Mariani. FZEA-USP Pirassununga: Mirele Daiana Poleti; Raul Machado Neto; Ricardo Augusto Brassaloti; Ricardo Haddad; Rodrigo Tocantins Calado. FAMERP-SJRP: Cecília Artico Banho; Sandra Coccuzzo Sampaio; Svetoslav Nanev Slavov; Vagner Fonseca; Vincent Louis Viala |
| EPI_ISL_1468428 | SAE Servico de Atendimento Especializado | Instituto Adolfo Lutz, Interdiciplinary Procedures Center, Strategic Laboratory | Caio Vinicius Dias Lopes; Claudia Regina Gonçalves; Claudio Tavares Sacchi; Erica Valessa Ramos Gomes; Karoline Rodrigues Campos |
| EPI_ISL_2345889 | SANTA CASA DE GUARAREMA | Instituto Butantan / FZEA-USP-Pirassununga | Antonio Jorge Martins; Claudia Renata dos Santos Barros; David Schlesinger; Debora Botequio Moretti; Dimas Tadeu Covas; Elaine Cristina Marqueze; Elaine Vieira Santos; Evandra Strazza Rodrigues; Heidge Fukumasu; Jayme Augusto de Souza-Neto; José Salvatore Leister Patané; Luiz Alcantara; Luiz Lehmann Coutinho; Maria Carolina Elias; Maurício Lacerda Nogueira; Rafael dos Santos Bezerra; Raul Machado Neto; Rejane Maria Tommasini Grotto; Ricardo Haddad; Sandra Coccuzzo Sampaio Vessoni; Simone Kashima; Svetoslav Nanev Slavov; Vincent Louis Viala |
| EPI_ISL_2209475, EPI_ISL_2209480 | SANTA CASA DE MISERICORDIA DE TIETE | Instituto Butantan | Antonio Jorge Martins; Claudia Renata dos Santos Barros; David Schlesinger; Debora Botequio Moretti; Dimas Tadeu Covas; Elaine Cristina Marqueze; Elaine Vieira Santos; Evandra Strazza Rodrigues; Heidge Fukumasu; Jayme Augusto de Souza-Neto; José Salvatore Leister Patané; Luiz Alcantara; Luiz Lehmann Coutinho; Maria Carolina Elias; Maurício Lacerda Nogueira; Rafael dos Santos Bezerra; Raul Machado Neto; Rejane Maria Tommasini Grotto; Ricardo Haddad; Sandra Coccuzzo Sampaio Vessoni; Simone Kashima; Svetoslav Nanev Slavov; Vincent Louis Viala. |
| EPI_ISL_1967193 | SANTA CASA DE MISERICORDIA DE TIETE | Instituto Butantan / ESALQ-USP (Piracicaba) | Antonio Jorge Martins; Bianca Cechetto Carlos. Mendelics: Bibiana Santos; Claudia Renata dos Santos Barros; Cíntia Bittar; David Schlesinger. Hemocentro Ribeirão Preto: Simone Kashima; Debora Botequio Moretti; Elaine Cristina Marqueze; Elaine Vieira dos Santos; Elisangela Chicaroni Mattos; Erika Freitas; Evandra Strazza Rodrigues; Felipe Allan da Silva da Costa; Flavia Aburjaile; Fábio Sossai Possebon; Guilherme Campos; Guilherme Targino Valente; Heidge Fukumasu. USP-Botucatu: Rejane Maria Tommasini Grotto; Helena Lage Ferreira; Instituto Butantan: Dimas Tadeu Covas; Jardelina de Souza Todao Bernardino; Jayme A. Souza-Neto; Jessika Cristina Chagas Lesbon; Jorge A. Petroli Marchesi; José Salvatore Leister Patané; João Paulo Kitajima; João Pessoa Araújo Jr.; Leila Sabrina Ullmann; Loyze Paola Oliveira de Lima; Luiz Aurelio de Campos Crispin. Centro de Genômica Funcional da ESALQ: Luiz Lehmann Coutinho; Luiz Carlos Junior de Alcantara; Lívia Sacchetto; Maisa C. Pereira Parra; Maria Carolina Elias; Marta Giovanetti; Marília Moraes; Maurício Lacerda Nogueira. Prefeitura de Sao Paulo: Melissa Palmieri.; Patricia Akemi Assato; Paula Rahal; Paulo Inacio da Costa; Rafael dos Santos Bezerra; Raquel de Lello Rocha Campos Cassano. NGS Soluções Genômicas: Pilar Drummond Sampaio Corrêa Mariani. FZEA-USP Pirassununga: Mirele Daiana Poleti; Raul Machado Neto; Ricardo Augusto Brassaloti; Ricardo Haddad; Rodrigo Tocantins Calado. FAMERP-SJRP: Cecília Artico Banho; Sandra Coccuzzo Sampaio; Svetoslav Nanev Slavov; Vagner Fonseca; Vincent Louis Viala |
| EPI_ISL_2344656, EPI_ISL_2344681 | SANTA CASA DE MISERICORDIA DE TIETE | Instituto Butantan / UNESP-Botucatu | Antonio Jorge Martins; Claudia Renata dos Santos Barros; David Schlesinger; Debora Botequio Moretti; Dimas Tadeu Covas; Elaine Cristina Marqueze; Elaine Vieira Santos; Evandra Strazza Rodrigues; Heidge Fukumasu; Jayme Augusto de Souza-Neto; José Salvatore Leister Patané; Luiz Alcantara; Luiz Lehmann Coutinho; Maria Carolina Elias; Maurício Lacerda Nogueira; Rafael dos Santos Bezerra; Raul Machado Neto; Rejane Maria Tommasini Grotto; Ricardo Haddad; Sandra Coccuzzo Sampaio Vessoni; Simone Kashima; Svetoslav Nanev Slavov; Vincent Louis Viala |
| EPI_ISL_2209484 | SANTA CASA DE MISERICORDIA DE UBATUBA | Instituto Butantan | Antonio Jorge Martins; Claudia Renata dos Santos Barros; David Schlesinger; Debora Botequio Moretti; Dimas Tadeu Covas; Elaine Cristina Marqueze; Elaine Vieira Santos; Evandra Strazza Rodrigues; Heidge Fukumasu; Jayme Augusto de Souza-Neto; José Salvatore Leister Patané; Luiz Alcantara; Luiz Lehmann Coutinho; Maria Carolina Elias; Maurício Lacerda Nogueira; Rafael dos Santos Bezerra; Raul Machado Neto; Rejane Maria Tommasini Grotto; Ricardo Haddad; Sandra Coccuzzo Sampaio Vessoni; Simone Kashima; Svetoslav Nanev Slavov; Vincent Louis Viala. |
| EPI_ISL_1795361, EPI_ISL_2345636 | SANTA CASA DE MISERICORDIA DE UBATUBA | Instituto Butantan / ESALQ-Piracicaba | Antonio Jorge Martins; Bianca Cechetto Carlos. Mendelics: Bibiana Santos; Claudia Renata dos Santos Barros; David Schlesinger; David Schlesinger. Hemocentro Ribeirão Preto: Simone Kashima; Debora Botequio Moretti; Debora Botequio Moretti. Centro de Genômica Funcional da ESALQ: Luiz Lehmann Coutinho; Dimas Tadeu Covas; Elaine Cristina Marqueze; Elaine Vieira Santos; Elaine Vieira dos Santos; Elisangela Chicaroni Mattos; Erika Freitas; Evandra Strazza Rodrigues; Felipe Allan da Silva da Costa; Flavia Aburjaile; Guilherme Targino Valente; Heidge Fukumasu; Heidge Fukumasu. USP-Botucatu: Rejane Maria Tommasini Grotto; Instituto Butantan: Alexander Roberto Precioso; Jayme A. Souza-Neto; Jayme Augusto de Souza-Neto; Jessika Cristina Chagas Lesbon; José Salvatore Leister Patané; João Paulo Kitajima; Luiz Alcantara; Luiz Carlos Junior de Alcantara; Luiz Lehmann Coutinho; Maria Carolina Elias; Marta Giovanetti; Maurício Lacerda Nogueira; Patricia Akemi Assato; Rafael dos Santos Bezerra; Raquel de Lello Rocha Campos Cassano. NGS Soluções Genômicas: Pilar Drummond Sampaio Corrêa Mariani. FZEA-USP Pirassununga: Mirele Daiana Poleti; Raul Machado Neto; Rejane Maria Tommasini Grotto; Ricardo Augusto Brassaloti; Ricardo Haddad; Rodrigo Tocantins Calado.; Sandra Coccuzzo Sampaio; Sandra Coccuzzo Sampaio Vessoni; Simone Kashima; Svetoslav Nanev Slavov; Vagner Fonseca; Vincent Louis Viala |
| EPI_ISL_2345299 | SANTA CASA DE MISERICORDIA DE UBATUBA | Instituto Butantan / UNESP-Botucatu | Antonio Jorge Martins; Claudia Renata dos Santos Barros; David Schlesinger; Debora Botequio Moretti; Dimas Tadeu Covas; Elaine Cristina Marqueze; Elaine Vieira Santos; Evandra Strazza Rodrigues; Heidge Fukumasu; Jayme Augusto de Souza-Neto; José Salvatore Leister Patané; Luiz Alcantara; Luiz Lehmann Coutinho; Maria Carolina Elias; Maurício Lacerda Nogueira; Rafael dos Santos Bezerra; Raul Machado Neto; Rejane Maria Tommasini Grotto; Ricardo Haddad; Sandra Coccuzzo Sampaio Vessoni; Simone Kashima; Svetoslav Nanev Slavov; Vincent Louis Viala |
| EPI_ISL_2345648 | SANTA CASA DE MISERICORDIA SAO JOSE | Instituto Butantan / Mendelics | Antonio Jorge Martins; Claudia Renata dos Santos Barros; David Schlesinger; Debora Botequio Moretti; Dimas Tadeu Covas; Elaine Cristina Marqueze; Elaine Vieira Santos; Evandra Strazza Rodrigues; Heidge Fukumasu; Jayme Augusto de Souza-Neto; José Salvatore Leister Patané; Luiz Alcantara; Luiz Lehmann Coutinho; Maria Carolina Elias; Maurício Lacerda Nogueira; Rafael dos Santos Bezerra; Raul Machado Neto; Rejane Maria Tommasini Grotto; Ricardo Haddad; Sandra Coccuzzo Sampaio Vessoni; Simone Kashima; Svetoslav Nanev Slavov; Vincent Louis Viala |
| EPI_ISL_2209487 | SANTA CASA DE PORTO FELIZ | Instituto Butantan | Antonio Jorge Martins; Claudia Renata dos Santos Barros; David Schlesinger; Debora Botequio Moretti; Dimas Tadeu Covas; Elaine Cristina Marqueze; Elaine Vieira Santos; Evandra Strazza Rodrigues; Heidge Fukumasu; Jayme Augusto de Souza-Neto; José Salvatore Leister Patané; Luiz Alcantara; Luiz Lehmann Coutinho; Maria Carolina Elias; Maurício Lacerda Nogueira; Rafael dos Santos Bezerra; Raul Machado Neto; Rejane Maria Tommasini Grotto; Ricardo Haddad; Sandra Coccuzzo Sampaio Vessoni; Simone Kashima; Svetoslav Nanev Slavov; Vincent Louis Viala. |
| EPI_ISL_1967016, EPI_ISL_1967017, EPI_ISL_1967019 | SANTA CASA DE PORTO FELIZ | Instituto Butantan / ESALQ-USP (Piracicaba) | Antonio Jorge Martins; Bianca Cechetto Carlos. Mendelics: Bibiana Santos; Claudia Renata dos Santos Barros; Cíntia Bittar; David Schlesinger. Hemocentro Ribeirão Preto: Simone Kashima; Debora Botequio Moretti; Elaine Cristina Marqueze; Elaine Vieira dos Santos; Elisangela Chicaroni Mattos; Erika Freitas; Evandra Strazza Rodrigues; Felipe Allan da Silva da Costa; Flavia Aburjaile; Fábio Sossai Possebon; Guilherme Campos; Guilherme Targino Valente; Heidge Fukumasu. USP-Botucatu: Rejane Maria Tommasini Grotto; Helena Lage Ferreira; Instituto Butantan: Dimas Tadeu Covas; Jardelina de Souza Todao Bernardino; Jayme A. Souza-Neto; Jessika Cristina Chagas Lesbon; Jorge A. Petroli Marchesi; José Salvatore Leister Patané; João Paulo Kitajima; João Pessoa Araújo Jr.; Leila Sabrina Ullmann; Loyze Paola Oliveira de Lima; Luiz Aurelio de Campos Crispin. Centro de Genômica Funcional da ESALQ: Luiz Lehmann Coutinho; Luiz Carlos Junior de Alcantara; Lívia Sacchetto; Maisa C. Pereira Parra; Maria Carolina Elias; Marta Giovanetti; Marília Moraes; Maurício Lacerda Nogueira. Prefeitura de Sao Paulo: Melissa Palmieri.; Patricia Akemi Assato; Paula Rahal; Paulo Inacio da Costa; Rafael dos Santos Bezerra; Raquel de Lello Rocha Campos Cassano. NGS Soluções Genômicas: Pilar Drummond Sampaio Corrêa Mariani. FZEA-USP Pirassununga: Mirele Daiana Poleti; Raul Machado Neto; Ricardo Augusto Brassaloti; Ricardo Haddad; Rodrigo Tocantins Calado. FAMERP-SJRP: Cecília Artico Banho; Sandra Coccuzzo Sampaio; Svetoslav Nanev Slavov; Vagner Fonseca; Vincent Louis Viala |
| EPI_ISL_2344588 | SANTA CASA DE PORTO FELIZ | Instituto Butantan / UNESP-Botucatu | Antonio Jorge Martins; Claudia Renata dos Santos Barros; David Schlesinger; Debora Botequio Moretti; Dimas Tadeu Covas; Elaine Cristina Marqueze; Elaine Vieira Santos; Evandra Strazza Rodrigues; Heidge Fukumasu; Jayme Augusto de Souza-Neto; José Salvatore Leister Patané; Luiz Alcantara; Luiz Lehmann Coutinho; Maria Carolina Elias; Maurício Lacerda Nogueira; Rafael dos Santos Bezerra; Raul Machado Neto; Rejane Maria Tommasini Grotto; Ricardo Haddad; Sandra Coccuzzo Sampaio Vessoni; Simone Kashima; Svetoslav Nanev Slavov; Vincent Louis Viala |
| EPI_ISL_2648769 | SARS-CoV-2 Sequencing Castilla y Leon-Spain Consortium | SARS-CoV-2 Sequencing Castilla y Leon-Spain Consortium | Antonio Orduña-Domingo; Carlos Fuster Foz; Carmen Aldea-Mansilla; Carmen Gimeno Crespo; David Abad; Gregoria Megías Lobón; Jose María Eiros Bouza; Laura Sánchez de Prada; M. Isabel Fernandez-Natal; Marta Dominguez-Gil; Marta Hernandez; María Antonia García Castro; Mª Fe Brezmes-Valdivieso; Noelia Arenal Andrés; Silvia Rojo; Sonsoles Garcinuño Pérez |
| EPI_ISL_1046769, EPI_ISL_1428640, EPI_ISL_1927238, EPI_ISL_1927289, EPI_ISL_1927291 | SARS-CoV-2 testing team, National Institute of Infectious Diseases | Pathogen Genomics Center, National Institute of Infectious Diseases | Chang-Kweng Lim; Eri Nakayama; Hazuka Y Furihata; Hidemasa Izumiya; Ken Shimuta; Kentaro Itokawa; Koichi Ishikawa; Makoto Kuroda; Masanori Hashino; Masumichi Saito; Midori Nakamura-Hoshi; Motohiko Ogawa; Naomi Nojiri; Nobuo Koizumi; Nozomu Hanaoka; Rina Tanaka; Sana Uchikoba; Shigeru Kusagawa; Shigeru Tanjima; Sunao Iyoda; Takahiro Maeki; Tsuguto Fujimoto; Tsuyoshi Sekizuka |
| EPI_ISL_2209490 | SAUDE COLETIVA CAPAO BONITO | Instituto Butantan | Antonio Jorge Martins; Claudia Renata dos Santos Barros; David Schlesinger; Debora Botequio Moretti; Dimas Tadeu Covas; Elaine Cristina Marqueze; Elaine Vieira Santos; Evandra Strazza Rodrigues; Heidge Fukumasu; Jayme Augusto de Souza-Neto; José Salvatore Leister Patané; Luiz Alcantara; Luiz Lehmann Coutinho; Maria Carolina Elias; Maurício Lacerda Nogueira; Rafael dos Santos Bezerra; Raul Machado Neto; Rejane Maria Tommasini Grotto; Ricardo Haddad; Sandra Coccuzzo Sampaio Vessoni; Simone Kashima; Svetoslav Nanev Slavov; Vincent Louis Viala. |

| | | | |
|---|---|---|---|
| EPI_ISL_2344657 | SAUDE COLETIVA CAPAO BONITO | Instituto Butantan / UNESP-Botucatu | Antonio Jorge Martins; Claudia Renata dos Santos Barros; David Schlesinger; Debora Botequio Moretti; Dimas Tadeu Covas; Elaine Cristina Marqueze; Elaine Vieira Santos; Evandra Strazza Rodrigues; Heidge Fukumasu; Jayme Augusto de Souza-Neto; José Salvatore Leister Patané; Luiz Alcantara; Luiz Lehmann Coutinho; Maria Carolina Elias; Maurício Lacerda Nogueira; Rafael dos Santos Bezerra; Raul Machado Neto; Rejane Maria Tommasini Grotto; Ricardo Haddad; Sandra Coccuzzo Sampaio Vessoni; Simone Kashima; Svetoslav Nanev Slavov; Vincent Louis Viala |
| EPI_ISL_1711927 | SC Dept of Health and Env. Control-Bureau of Laboratories | Centers for Disease Control and Prevention Division of Viral Diseases, Pathogen Discovery | Alison Laufer Halpin; Ben L. Rambo-Martin; Clinton R. Paden; Dakota Howard; Darlene Wagner; Dave Wentworth; Dhwani Batra; Jasmine Padilla; Justin Lee; Katie Dillon; Krista Queen; Kristen Knipe; Kristine Lacek; Mark Burroughs; Matthew Schmerer; Mili Sheth; Peter Cook; Sam Shepard; Sarah Nobles; Shoshona Le; Suxiang Tong; Vivien Dugan; Yvette Unoarumhi |
| EPI_ISL_1445238, EPI_ISL_1966077, EPI_ISL_1966189, EPI_ISL_1966484, EPI_ISL_1966487, EPI_ISL_1966491, EPI_ISL_1967260, EPI_ISL_1967265, EPI_ISL_1967270 | | | |
| see above | SECAO CENTRO DE DIAGNOSTICO SECEDI | Instituto Butantan / Mendelics | Antonio Jorge Martins; Bianca Cechetto Carlos. Mendelics: Bibiana Santos; Bibiana Santos; Claudia Renata dos Santos Barros; Cíntia Bittar; David Schlesinger; David Schlesinger. Hemocentro Ribeirão Preto: Simone Kashima; Debora Botequio Moretti; Dimas Tadeu Covas; Elaine Cristina Marqueze; Elaine Vieira dos Santos; Elisangela Chicaroni Mattos; Erika Freitas; Evandra Strazza Rodrigues; Felipe Allan da Silva da Costa; Flavia Aburjaile; Fábio Sossai Possebon; Guilherme Campos; Guilherme Targino Valente; Heidge Fukumasu. USP-Botucatu: Rejane Maria Tommasini Grotto; Instituto Butantan: Dimas Tadeu Covas; Jardelina de Souza Todao Bernardino; Jayme A. Souza-Neto; Jessika Cristina Chagas Lesbon; Jorge A. Petroli Marchesi; José Salvatore Leister Patané; João Paulo Kitajima; João Pessoa Araújo Jr.; Leila Sabrina Ullmann; Loyze Paola Oliveira de Lima; Luiz Aurelio de Campos Crispin. Centro de Genômica Funcional da ESALQ: Luiz Lehmann Coutinho; Luiz Carlos Junior de Alcantara; Lívia Sacchetto; Maisa C. Pereira Parra; Maria Carolina Elias; Marta Giovanetti; Marília Moraes; Maurício Lacerda Nogueira. Prefeitura de Sao Paulo: Melissa Palmieri.; Patricia Akemi Assato; Paula Rahal; Paulo Inacio da Costa; Rafael dos Santos Bezerra; Raquel de Lello Rocha Campos Cassano. NGS Soluções Genômicas: Pilar Drummond Sampaio Corrêa Mariani. FZEA-USP Pirassununga: Mirele Daiana Poleti; Raul Machado Neto; Ricardo Augusto Brassaloti; Ricardo Haddad; Rodrigo Tocantins Calado.; Rodrigo Tocantins Calado. FAMERP-SJRP: Cecília Artico Banho; Sandra Coccuzzo Sampaio; Simone Kashima; Svetoslav Nanev Slavov; Vagner Fonseca; Vincent Louis Viala |
| EPI_ISL_1795364, EPI_ISL_2345639 | SECRETARIA DE SAUDE | Instituto Butantan / ESALQ-Piracicaba | Antonio Jorge Martins; Bianca Cechetto Carlos. Mendelics: Bibiana Santos; Claudia Renata dos Santos Barros; David Schlesinger; David Schlesinger. Hemocentro Ribeirão Preto: Simone Kashima; Debora Botequio Moretti; Debora Botequio Moretti. Centro de Genômica Funcional da ESALQ: Luiz Lehmann Coutinho; Dimas Tadeu Covas; Elaine Cristina Marqueze; Elaine Vieira Santos; Elaine Vieira dos Santos; Elisangela Chicaroni Mattos; Erika Freitas; Evandra Strazza Rodrigues; Felipe Allan da Silva da Costa; Flavia Aburjaile; Guilherme Targino Valente; Heidge Fukumasu; Heidge Fukumasu. USP-Botucatu: Rejane Maria Tommasini Grotto; Instituto Butantan: Alexander Roberto Precioso; Jayme A. Souza-Neto; Jayme Augusto de Souza-Neto; Jessika Cristina Chagas Lesbon; José Salvatore Leister Patané; João Paulo Kitajima; Luiz Alcantara; Luiz Carlos Junior de Alcantara; Luiz Lehmann Coutinho; Maria Carolina Elias; Maurício Lacerda Nogueira; Patricia Akemi Assato; Rafael dos Santos Bezerra; Raquel de Lello Rocha Campos Cassano. NGS Soluções Genômicas: Pilar Drummond Sampaio Corrêa Mariani. FZEA-USP Pirassununga: Mirele Daiana Poleti; Raul Machado Neto; Rejane Maria Tommasini Grotto; Ricardo Augusto Brassaloti; Ricardo Haddad; Rodrigo Tocantins Calado.; Sandra Coccuzzo Sampaio; Sandra Coccuzzo Sampaio Vessoni; Simone Kashima; Svetoslav Nanev Slavov; Vagner Fonseca; Vincent Louis Viala |
| EPI_ISL_2345750 | SECRETARIA DE SAUDE | Instituto Butantan / Mendelics | Antonio Jorge Martins; Claudia Renata dos Santos Barros; David Schlesinger; Debora Botequio Moretti; Dimas Tadeu Covas; Elaine Cristina Marqueze; Elaine Vieira Santos; Evandra Strazza Rodrigues; Heidge Fukumasu; Jayme Augusto de Souza-Neto; José Salvatore Leister Patané; Luiz Alcantara; Luiz Lehmann Coutinho; Maria Carolina Elias; Maurício Lacerda Nogueira; Rafael dos Santos Bezerra; Raul Machado Neto; Rejane Maria Tommasini Grotto; Ricardo Haddad; Sandra Coccuzzo Sampaio Vessoni; Simone Kashima; Svetoslav Nanev Slavov; Vincent Louis Viala |
| EPI_ISL_2209496 | SECRETARIA DE SAUDE DE RAFARD | Instituto Butantan | Antonio Jorge Martins; Claudia Renata dos Santos Barros; David Schlesinger; Debora Botequio Moretti; Dimas Tadeu Covas; Elaine Cristina Marqueze; Elaine Vieira Santos; Evandra Strazza Rodrigues; Heidge Fukumasu; Jayme Augusto de Souza-Neto; José Salvatore Leister Patané; Luiz Alcantara; Luiz Lehmann Coutinho; Maria Carolina Elias; Maurício Lacerda Nogueira; Rafael dos Santos Bezerra; Raul Machado Neto; Rejane Maria Tommasini Grotto; Ricardo Haddad; Sandra Coccuzzo Sampaio Vessoni; Simone Kashima; Svetoslav Nanev Slavov; Vincent Louis Viala. |
| EPI_ISL_2344726, EPI_ISL_2344727 | SECRETARIA DE SAUDE DE RAFARD | Instituto Butantan / FZEA-USP-Pirassununga | Antonio Jorge Martins; Claudia Renata dos Santos Barros; David Schlesinger; Debora Botequio Moretti; Dimas Tadeu Covas; Elaine Cristina Marqueze; Elaine Vieira Santos; Evandra Strazza Rodrigues; Heidge Fukumasu; Jayme Augusto de Souza-Neto; José Salvatore Leister Patané; Luiz Alcantara; Luiz Lehmann Coutinho; Maria Carolina Elias; Maurício Lacerda Nogueira; Rafael dos Santos Bezerra; Raul Machado Neto; Rejane Maria Tommasini Grotto; Ricardo Haddad; Sandra Coccuzzo Sampaio Vessoni; Simone Kashima; Svetoslav Nanev Slavov; Vincent Louis Viala |
| EPI_ISL_2209505, EPI_ISL_2209522 | SECRETARIA DE SAUDE DE SANTA GERTRUDES | Instituto Butantan | Antonio Jorge Martins; Claudia Renata dos Santos Barros; David Schlesinger; Debora Botequio Moretti; Dimas Tadeu Covas; Elaine Cristina Marqueze; Elaine Vieira Santos; Evandra Strazza Rodrigues; Heidge Fukumasu; Jayme Augusto de Souza-Neto; José Salvatore Leister Patané; Luiz Alcantara; Luiz Lehmann Coutinho; Maria Carolina Elias; Maurício Lacerda Nogueira; Rafael dos Santos Bezerra; Raul Machado Neto; Rejane Maria Tommasini Grotto; Ricardo Haddad; Sandra Coccuzzo Sampaio Vessoni; Simone Kashima; Svetoslav Nanev Slavov; Vincent Louis Viala. |
| EPI_ISL_2209255, EPI_ISL_2209937, EPI_ISL_2344703, EPI_ISL_2344730 | SECRETARIA DE SAUDE DE SANTA GERTRUDES | Instituto Butantan / FZEA-USP-Pirassununga | Antonio Jorge Martins; Claudia Renata dos Santos Barros; David Schlesinger; Debora Botequio Moretti; Dimas Tadeu Covas; Elaine Cristina Marqueze; Elaine Vieira Santos; Evandra Strazza Rodrigues; Heidge Fukumasu; Jayme Augusto de Souza-Neto; José Salvatore Leister Patané; Luiz Alcantara; Luiz Lehmann Coutinho; Maria Carolina Elias; Maurício Lacerda Nogueira; Rafael dos Santos Bezerra; Raul Machado Neto; Rejane Maria Tommasini Grotto; Ricardo Haddad; Sandra Coccuzzo Sampaio Vessoni; Simone Kashima; Svetoslav Nanev Slavov; Vincent Louis Viala |
| EPI_ISL_2209537 | SECRETARIA DE SAUDE DE SAO PEDRO | Instituto Butantan | Antonio Jorge Martins; Claudia Renata dos Santos Barros; David Schlesinger; Debora Botequio Moretti; Dimas Tadeu Covas; Elaine Cristina Marqueze; Elaine Vieira Santos; Evandra Strazza Rodrigues; Heidge Fukumasu; Jayme Augusto de Souza-Neto; José Salvatore Leister Patané; Luiz Alcantara; Luiz Lehmann Coutinho; Maria Carolina Elias; Maurício Lacerda Nogueira; Rafael dos Santos Bezerra; Raul Machado Neto; Rejane Maria Tommasini Grotto; Ricardo Haddad; Sandra Coccuzzo Sampaio Vessoni; Simone Kashima; Svetoslav Nanev Slavov; Vincent Louis Viala. |
| EPI_ISL_2209212, EPI_ISL_2344731 | SECRETARIA DE SAUDE DE SAO PEDRO | Instituto Butantan / FZEA-USP-Pirassununga | Antonio Jorge Martins; Claudia Renata dos Santos Barros; David Schlesinger; Debora Botequio Moretti; Dimas Tadeu Covas; Elaine Cristina Marqueze; Elaine Vieira Santos; Evandra Strazza Rodrigues; Heidge Fukumasu; Jayme Augusto de Souza-Neto; José Salvatore Leister Patané; Luiz Alcantara; Luiz Lehmann Coutinho; Maria Carolina Elias; Maurício Lacerda Nogueira; Rafael dos Santos Bezerra; Raul Machado Neto; Rejane Maria Tommasini Grotto; Ricardo Haddad; Sandra Coccuzzo Sampaio Vessoni; Simone Kashima; Svetoslav Nanev Slavov; Vincent Louis Viala |
| EPI_ISL_1966247 | SECRETARIA DE SAUDE DO MUNICIPIO DE IPUA | Instituto Butantan / Mendelics | Antonio Jorge Martins; Bianca Cechetto Carlos. Mendelics: Bibiana Santos; Claudia Renata dos Santos Barros; Cíntia Bittar; David Schlesinger. Hemocentro Ribeirão Preto: Simone Kashima; Debora Botequio Moretti; Elaine Cristina Marqueze; Elaine Vieira dos Santos; Elisangela Chicaroni Mattos; Erika Freitas; Evandra Strazza Rodrigues; Felipe Allan da Silva da Costa; Flavia Aburjaile; Fábio Sossai Possebon; Guilherme Campos; Guilherme Targino Valente; Heidge Fukumasu. USP-Botucatu: Rejane Maria Tommasini Grotto; Helena Lage Ferreira; Instituto Butantan: Dimas Tadeu Covas; Jardelina de Souza Todao Bernardino; Jayme A. Souza-Neto; Jessika Cristina Chagas Lesbon; Jorge A. Petroli Marchesi; José Salvatore Leister Patané; João Paulo Kitajima; João Pessoa Araújo Jr.; Leila Sabrina Ullmann; Loyze Paola Oliveira de Lima; Luiz Aurelio de Campos Crispin. Centro de Genômica Funcional da ESALQ: Luiz Lehmann Coutinho; Luiz Carlos Junior de Alcantara; Lívia Sacchetto; Maisa C. Pereira Parra; Maria Carolina Elias; Marta Giovanetti; Marília Moraes; Maurício Lacerda Nogueira. Prefeitura de Sao Paulo: Melissa Palmieri.; Patricia Akemi Assato; Paula Rahal; Paulo Inacio da Costa; Rafael dos Santos Bezerra; Raquel de Lello Rocha Campos Cassano. NGS Soluções Genômicas: Pilar Drummond Sampaio Corrêa Mariani. FZEA-USP Pirassununga: Mirele Daiana Poleti; Raul Machado Neto; Ricardo Augusto Brassaloti; Ricardo Haddad; Rodrigo Tocantins Calado. FAMERP-SJRP: Cecília Artico Banho; Sandra Coccuzzo Sampaio; Svetoslav Nanev Slavov; Vagner Fonseca; Vincent Louis Viala |
| EPI_ISL_1966713 | SECRETARIA MUNCIPAL DE SAUDE DE SANTA BARBARA D OESTE | Instituto Butantan / Mendelics | Antonio Jorge Martins; Bianca Cechetto Carlos. Mendelics: Bibiana Santos; Claudia Renata dos Santos Barros; Cíntia Bittar; David Schlesinger. Hemocentro Ribeirão Preto: Simone Kashima; Debora Botequio Moretti; Elaine Cristina Marqueze; Elaine Vieira dos Santos; Elisangela Chicaroni Mattos; Erika Freitas; Evandra Strazza Rodrigues; Felipe Allan da Silva da Costa; Flavia Aburjaile; Fábio Sossai Possebon; Guilherme Campos; Guilherme Targino Valente; Heidge Fukumasu. USP-Botucatu: Rejane Maria Tommasini Grotto; Helena Lage Ferreira; Instituto Butantan: Dimas Tadeu Covas; Jardelina de Souza Todao Bernardino; Jayme A. Souza-Neto; Jessika Cristina Chagas Lesbon; Jorge A. Petroli Marchesi; José Salvatore Leister Patané; João Paulo Kitajima; João Pessoa Araújo Jr.; Leila Sabrina Ullmann; Loyze Paola Oliveira de Lima; Luiz Aurelio de Campos Crispin. Centro de Genômica Funcional da ESALQ: Luiz Lehmann Coutinho; Luiz Carlos Junior de Alcantara; Lívia Sacchetto; Maisa C. Pereira Parra; Maria Carolina Elias; Marta Giovanetti; Marília Moraes; Maurício Lacerda Nogueira. Prefeitura de Sao Paulo: Melissa Palmieri.; Patricia Akemi Assato; Paula Rahal; Paulo Inacio da Costa; Rafael dos Santos Bezerra; Raquel de Lello Rocha Campos Cassano. NGS Soluções Genômicas: Pilar Drummond Sampaio Corrêa Mariani. FZEA-USP Pirassununga: Mirele Daiana Poleti; Raul Machado Neto; Ricardo Augusto Brassaloti; Ricardo Haddad; Rodrigo Tocantins Calado. FAMERP-SJRP: Cecília Artico Banho; Sandra Coccuzzo Sampaio; Svetoslav Nanev Slavov; Vagner Fonseca; Vincent Louis Viala |
| EPI_ISL_2209551, EPI_ISL_2209557 | SECRETARIA MUNICIPAL DA SAUDE DE GUARATINGUETA | Instituto Butantan | Antonio Jorge Martins; Claudia Renata dos Santos Barros; David Schlesinger; Debora Botequio Moretti; Dimas Tadeu Covas; Elaine Cristina Marqueze; Elaine Vieira Santos; Evandra Strazza Rodrigues; Heidge Fukumasu; Jayme Augusto de Souza-Neto; José Salvatore Leister Patané; Luiz Alcantara; Luiz Lehmann Coutinho; Maria Carolina Elias; Maurício Lacerda Nogueira; Rafael dos Santos Bezerra; Raul Machado Neto; Rejane Maria Tommasini Grotto; Ricardo Haddad; Sandra Coccuzzo Sampaio Vessoni; Simone Kashima; Svetoslav Nanev Slavov; Vincent Louis Viala. |
| EPI_ISL_2345300, EPI_ISL_2345305 | SECRETARIA MUNICIPAL DA SAUDE DE GUARATINGUETA | Instituto Butantan / UNESP-Botucatu | Antonio Jorge Martins; Claudia Renata dos Santos Barros; David Schlesinger; Debora Botequio Moretti; Dimas Tadeu Covas; Elaine Cristina Marqueze; Elaine Vieira Santos; Evandra Strazza Rodrigues; Heidge Fukumasu; Jayme Augusto de Souza-Neto; José Salvatore Leister Patané; Luiz Alcantara; Luiz Lehmann Coutinho; Maria Carolina Elias; Maurício Lacerda Nogueira; Rafael dos Santos Bezerra; Raul Machado Neto; Rejane Maria Tommasini Grotto; Ricardo Haddad; Sandra Coccuzzo Sampaio Vessoni; Simone Kashima; Svetoslav Nanev Slavov; Vincent Louis Viala |
| EPI_ISL_1795111, EPI_ISL_1795112, EPI_ISL_2345310, EPI_ISL_2345311 | SECRETARIA MUNICIPAL DA SAUDE DE GUARIBA | Instituto Butantan / ESALQ-Piracicaba | Antonio Jorge Martins; Bianca Cechetto Carlos. Mendelics: Bibiana Santos; Claudia Renata dos Santos Barros; David Schlesinger; David Schlesinger. Hemocentro Ribeirão Preto: Simone Kashima; Debora Botequio Moretti; Debora Botequio Moretti. Centro de Genômica Funcional da ESALQ: Luiz Lehmann Coutinho; Dimas Tadeu Covas; Elaine Cristina Marqueze; Elaine Vieira Santos; Elaine Vieira dos Santos; Elisangela Chicaroni Mattos; Erika Freitas; Evandra Strazza Rodrigues; Felipe Allan da Silva da Costa; Flavia Aburjaile; Guilherme Targino Valente; Heidge Fukumasu; Heidge Fukumasu. USP-Botucatu: Rejane Maria Tommasini Grotto; Instituto Butantan: Alexander Roberto Precioso; Jayme A. Souza-Neto; Jayme Augusto de Souza-Neto; Jessika Cristina Chagas Lesbon; José Salvatore Leister Patané; João Paulo Kitajima; Luiz Alcantara; Luiz Carlos Junior de Alcantara; Luiz Lehmann Coutinho; Maria Carolina Elias; Marta Giovanetti; Maurício Lacerda Nogueira; Patricia Akemi Assato; Rafael dos Santos Bezerra; Raquel de Lello Rocha Campos Cassano. NGS Soluções Genômicas: Pilar Drummond Sampaio Corrêa Mariani. FZEA-USP Pirassununga: Mirele Daiana Poleti; Raul Machado Neto; Rejane Maria Tommasini Grotto; Ricardo Augusto Brassaloti; Ricardo Haddad; Rodrigo Tocantins Calado.; Sandra Coccuzzo Sampaio; Sandra Coccuzzo Sampaio Vessoni; Simone Kashima; Svetoslav Nanev Slavov; Vagner Fonseca; Vincent Louis Viala |
| EPI_ISL_2170910 | SECRETARIA MUNICIPAL DA SAUDE DE GUARIBA | Instituto Butantan / Mendelics | Antonio Jorge Martins; Bianca Cechetto Carlos. Mendelics: Bibiana Santos; Claudia Renata dos Santos Barros; Cíntia Bittar; David Schlesinger. Hemocentro Ribeirão Preto: Simone Kashima; Debora Botequio Moretti; Elaine Cristina Marqueze; Elaine Vieira dos Santos; Elisangela Chicaroni Mattos; Erika Freitas; Evandra Strazza Rodrigues; Felipe Allan da Silva da Costa; Flavia Aburjaile; Fábio Sossai Possebon; Guilherme Campos; Guilherme Targino Valente; Heidge Fukumasu. USP-Botucatu: Rejane Maria Tommasini Grotto; Helena Lage Ferreira; Instituto Butantan: Dimas Tadeu Covas; Jardelina de Souza Todao Bernardino; Jayme A. Souza-Neto; Jessika Cristina Chagas Lesbon; Jorge A. Petroli Marchesi; José Salvatore Leister Patané; João Paulo Kitajima; João Pessoa Araújo Jr.; Leila Sabrina Ullmann; Loyze Paola Oliveira de Lima; Luiz Aurelio de Campos Crispin. Centro de Genômica Funcional da ESALQ: Luiz Lehmann Coutinho; Luiz Carlos Junior de Alcantara; Lívia Sacchetto; Maisa C. Pereira Parra; Maria Carolina Elias; Marta Giovanetti; Marília Moraes; Maurício Lacerda Nogueira. Prefeitura de Sao Paulo: Melissa Palmieri.; Patricia Akemi Assato; Paula Rahal; Paulo Inacio da Costa; Rafael dos Santos Bezerra; Raquel de Lello Rocha Campos Cassano. NGS Soluções Genômicas: Pilar Drummond Sampaio Corrêa Mariani. FZEA-USP Pirassununga: Mirele Daiana Poleti; Raul Machado Neto; Ricardo Augusto Brassaloti; Ricardo Haddad; Rodrigo Tocantins Calado. FAMERP-SJRP: Cecília Artico Banho; Sandra Coccuzzo Sampaio; Svetoslav Nanev Slavov; Vagner Fonseca; Vincent Louis Viala |
| EPI_ISL_1966950, EPI_ISL_1966951 | SECRETARIA MUNICIPAL DA SAUDE DE ITARARE | Instituto Butantan / ESALQ-USP (Piracicaba) | Antonio Jorge Martins; Bianca Cechetto Carlos. Mendelics: Bibiana Santos; Claudia Renata dos Santos Barros; Cíntia Bittar; David Schlesinger. Hemocentro Ribeirão Preto: Simone Kashima; Debora Botequio Moretti; Elaine Cristina Marqueze; Elaine Vieira dos Santos; Elisangela Chicaroni Mattos; Erika Freitas; Evandra Strazza Rodrigues; Felipe Allan da Silva da Costa; Flavia Aburjaile; Fábio Sossai Possebon; Guilherme Campos; Guilherme Targino Valente; Heidge Fukumasu. USP-Botucatu: Rejane Maria Tommasini Grotto; Helena Lage Ferreira; Instituto Butantan: Dimas Tadeu Covas; Jardelina de Souza Todao Bernardino; Jayme A. Souza-Neto; Jessika Cristina Chagas Lesbon; Jorge A. Petroli Marchesi; José Salvatore Leister Patané; João Paulo Kitajima; João Pessoa Araújo Jr.; Leila Sabrina Ullmann; Loyze Paola Oliveira de Lima; Luiz Aurelio de Campos Crispin. Centro de Genômica Funcional da ESALQ: Luiz Lehmann Coutinho; Luiz Carlos Junior de Alcantara; Lívia Sacchetto; Maisa C. Pereira Parra; Maria Carolina Elias; Marta Giovanetti; Marília Moraes; Maurício Lacerda Nogueira. Prefeitura de Sao Paulo: Melissa Palmieri.; Patricia Akemi Assato; Paula Rahal; Paulo Inacio da Costa; Rafael dos Santos Bezerra; Raquel de Lello Rocha Campos Cassano. NGS Soluções Genômicas: Pilar Drummond Sampaio Corrêa Mariani. FZEA-USP Pirassununga: Mirele Daiana Poleti; Raul Machado Neto; Ricardo Augusto Brassaloti; Ricardo Haddad; Rodrigo Tocantins Calado. FAMERP-SJRP: Cecília Artico Banho; Sandra Coccuzzo Sampaio; Svetoslav Nanev Slavov; Vagner Fonseca; Vincent Louis Viala |
| EPI_ISL_2344748 | SECRETARIA MUNICIPAL DE SAUDE | Instituto Butantan / ESALQ-Piracicaba | Antonio Jorge Martins; Claudia Renata dos Santos Barros; David Schlesinger; Debora Botequio Moretti; Dimas Tadeu Covas; Elaine Cristina Marqueze; Elaine Vieira Santos; Evandra Strazza Rodrigues; Heidge Fukumasu; Jayme Augusto de Souza-Neto; José Salvatore Leister Patané; Luiz Alcantara; Luiz Lehmann Coutinho; Maria Carolina Elias; Maurício Lacerda Nogueira; Rafael dos Santos Bezerra; Raul Machado Neto; Rejane Maria Tommasini Grotto; Ricardo Haddad; Sandra Coccuzzo Sampaio Vessoni; Simone Kashima; Svetoslav Nanev Slavov; Vincent Louis Viala |
| EPI_ISL_1967301, EPI_ISL_1967303, EPI_ISL_2171024 | SECRETARIA MUNICIPAL DE SAUDE | Instituto Butantan / Mendelics | Antonio Jorge Martins; Bianca Cechetto Carlos. Mendelics: Bibiana Santos; Claudia Renata dos Santos Barros; Cíntia Bittar; David Schlesinger. Hemocentro Ribeirão Preto: Simone Kashima; Debora Botequio Moretti; Elaine Cristina Marqueze; Elaine Vieira dos Santos; Elisangela Chicaroni Mattos; Erika Freitas; Evandra Strazza Rodrigues; Felipe Allan da Silva da Costa; Flavia Aburjaile; Fábio Sossai Possebon; Guilherme Campos; Guilherme Targino Valente; Heidge Fukumasu. USP-Botucatu: Rejane Maria Tommasini Grotto; Helena Lage Ferreira; Instituto Butantan: Dimas Tadeu Covas; Jardelina de Souza Todao Bernardino; Jayme A. Souza-Neto; Jessika Cristina Chagas Lesbon; Jorge A. Petroli Marchesi; José Salvatore Leister Patané; João Paulo Kitajima; João Pessoa Araújo Jr.; Leila Sabrina Ullmann; Loyze Paola Oliveira de Lima; Luiz Aurelio de Campos Crispin. Centro de Genômica Funcional da ESALQ: Luiz Lehmann Coutinho; Luiz Carlos Junior de Alcantara; Lívia Sacchetto; Maisa C. Pereira Parra; Maria Carolina Elias; Marta Giovanetti; Marília Moraes; Maurício Lacerda Nogueira. Prefeitura de Sao Paulo: Melissa Palmieri.; Patricia Akemi Assato; Paula Rahal; Paulo Inacio da Costa; Rafael dos Santos Bezerra; Raquel de Lello Rocha Campos Cassano. NGS Soluções Genômicas: Pilar Drummond Sampaio Corrêa Mariani. FZEA-USP Pirassununga: Mirele Daiana Poleti; Raul Machado Neto; Ricardo Augusto Brassaloti; Ricardo Haddad; Rodrigo Tocantins Calado. FAMERP-SJRP: Cecília Artico Banho; Sandra Coccuzzo Sampaio; Svetoslav Nanev Slavov; Vagner Fonseca; Vincent Louis Viala |
| EPI_ISL_1966272 | SECRETARIA MUNICIPAL DE SAUDE DE ANDRADINA | Instituto Butantan / Mendelics | Antonio Jorge Martins; Bianca Cechetto Carlos. Mendelics: Bibiana Santos; Claudia Renata dos Santos Barros; Cíntia Bittar; David Schlesinger. Hemocentro Ribeirão Preto: Simone Kashima; Debora Botequio Moretti; Elaine Cristina Marqueze; Elaine Vieira dos Santos; Elisangela Chicaroni Mattos; Erika Freitas; Evandra Strazza Rodrigues; Felipe Allan da Silva da Costa; Flavia Aburjaile; Fábio Sossai Possebon; Guilherme Campos; Guilherme Targino Valente; Heidge Fukumasu. USP-Botucatu: Rejane Maria Tommasini Grotto; Helena Lage Ferreira; Instituto Butantan: Dimas Tadeu Covas; Jardelina de Souza Todao Bernardino; Jayme A. Souza-Neto; Jessika Cristina Chagas Lesbon; Jorge A. Petroli Marchesi; José Salvatore Leister Patané; João Paulo Kitajima; João Pessoa Araújo Jr.; Leila Sabrina Ullmann; Loyze Paola Oliveira de Lima; Luiz Aurelio de Campos Crispin. Centro de Genômica Funcional da ESALQ: Luiz Lehmann Coutinho; Luiz Carlos Junior de Alcantara; Lívia Sacchetto; Maisa C. Pereira Parra; Maria Carolina Elias; Marta Giovanetti; Marília Moraes; Maurício Lacerda Nogueira. Prefeitura de Sao Paulo: Melissa Palmieri.; Patricia Akemi Assato; Paula Rahal; Paulo Inacio da Costa; Rafael dos Santos Bezerra; Raquel de Lello Rocha Campos Cassano. NGS Soluções Genômicas: Pilar Drummond Sampaio Corrêa Mariani. FZEA-USP Pirassununga: Mirele Daiana Poleti; Raul Machado Neto; Ricardo Augusto Brassaloti; Ricardo Haddad; Rodrigo Tocantins Calado. FAMERP-SJRP: Cecília Artico Banho; Sandra Coccuzzo Sampaio; Svetoslav Nanev Slavov; Vagner Fonseca; Vincent Louis Viala |
| EPI_ISL_3912238 | SECRETARIA MUNICIPAL DE SAUDE DE CATUNDA | Analytical Competence Molecular Epidemiology Lab/ACME, Oswaldo Cruz Foundation, Ceara (FIOCRUZ CE) | Cleber Furtado Aksenen; Fabio Miyajima; Fernando Braga Stehling; Francisco Eder de Moura Lopes; Jamille Maria Mendes Bezerra; Joaquim Cesar do Nascimento Sousa Junior; Pedro Miguel Carneiro Jeronimo; Suzana Porto Almeida & Lucas Delerino on behalf of COVID-19 FIOCRUZ Genomic Network; Thais Ferreira de Oliveira; Thais de Oliveira Costa; Ticiane Cavalcante de Souza; Veridiana Pessoa Miyajima |
| EPI_ISL_2209610 | SECRETARIA MUNICIPAL DE SAUDE DE CORDEIROPOLIS | Instituto Butantan | Antonio Jorge Martins; Claudia Renata dos Santos Barros; David Schlesinger; Debora Botequio Moretti; Dimas Tadeu Covas; Elaine Cristina Marqueze; Elaine Vieira Santos; Evandra Strazza Rodrigues; Heidge Fukumasu; Jayme Augusto de Souza-Neto; José Salvatore Leister Patané; Luiz Alcantara; Luiz Lehmann Coutinho; Maria Carolina Elias; Maurício Lacerda Nogueira; Rafael dos Santos Bezerra; Raul Machado Neto; Rejane Maria Tommasini Grotto; Ricardo Haddad; Sandra Coccuzzo Sampaio Vessoni; Simone Kashima; Svetoslav Nanev Slavov; Vincent Louis Viala. |
| EPI_ISL_2344737 | SECRETARIA MUNICIPAL DE SAUDE DE CORDEIROPOLIS | Instituto Butantan / ESALQ-Piracicaba | Antonio Jorge Martins; Claudia Renata dos Santos Barros; David Schlesinger; Debora Botequio Moretti; Dimas Tadeu Covas; Elaine Cristina Marqueze; Elaine Vieira Santos; Evandra Strazza Rodrigues; Heidge Fukumasu; Jayme Augusto de Souza-Neto; José Salvatore Leister Patané; Luiz Alcantara; Luiz Lehmann Coutinho; Maria Carolina Elias; Maurício Lacerda Nogueira; Rafael dos Santos Bezerra; Raul Machado Neto; Rejane Maria Tommasini Grotto; Ricardo Haddad; Sandra Coccuzzo Sampaio Vessoni; Simone Kashima; Svetoslav Nanev Slavov; Vincent Louis Viala |
| EPI_ISL_2209145, EPI_ISL_2209457, EPI_ISL_2210037, EPI_ISL_2210082, EPI_ISL_2344672, EPI_ISL_2344675 | SECRETARIA MUNICIPAL DE SAUDE DE CORDEIROPOLIS | Instituto Butantan / FZEA-USP-Pirassununga | Antonio Jorge Martins; Claudia Renata dos Santos Barros; David Schlesinger; Debora Botequio Moretti; Dimas Tadeu Covas; Elaine Cristina Marqueze; Elaine Vieira Santos; Evandra Strazza Rodrigues; Heidge Fukumasu; Jayme Augusto de Souza-Neto; José Salvatore Leister Patané; Luiz Alcantara; Luiz Lehmann Coutinho; Maria Carolina Elias; Maurício Lacerda Nogueira; Rafael dos Santos Bezerra; Raul Machado Neto; Rejane Maria Tommasini Grotto; Ricardo Haddad; Sandra Coccuzzo Sampaio Vessoni; Simone Kashima; Svetoslav Nanev Slavov; Vincent Louis Viala |
| EPI_ISL_1966693 | SECRETARIA MUNICIPAL DE SAUDE DE JARINU | Instituto Butantan / Mendelics | Antonio Jorge Martins; Bianca Cechetto Carlos. Mendelics: Bibiana Santos; Claudia Renata dos Santos Barros; Cíntia Bittar; David Schlesinger. Hemocentro Ribeirão Preto: Simone Kashima; Debora Botequio Moretti; Elaine Cristina Marqueze; Elaine Vieira dos Santos; Elisangela Chicaroni Mattos; Erika Freitas; Evandra Strazza Rodrigues; Felipe Allan da Silva da Costa; Flavia Aburjaile; Fábio Sossai Possebon; Guilherme Campos; Guilherme Targino Valente; Heidge Fukumasu. USP-Botucatu: Rejane Maria Tommasini Grotto; Helena Lage Ferreira; Instituto Butantan: Dimas Tadeu Covas; Jardelina de Souza Todao Bernardino; Jayme A. Souza-Neto; Jessika Cristina Chagas Lesbon; Jorge A. Petroli Marchesi; José Salvatore Leister Patané; João Paulo Kitajima; João Pessoa Araújo Jr.; Leila Sabrina Ullmann; Loyze Paola Oliveira de Lima; Luiz Aurelio de Campos Crispin. Centro de Genômica Funcional da ESALQ: Luiz Lehmann Coutinho; Luiz Carlos Junior de Alcantara; Lívia Sacchetto; Maisa C. Pereira Parra; Maria |

| | | | |
|---|---|---|---|
| | | | Carolina Elias; Marta Giovanetti; Marília Moraes; Maurício Lacerda Nogueira. Prefeitura de Sao Paulo: Melissa Palmieri.; Patricia Akemi Assato; Paula Rahal; Paulo Inacio da Costa; Rafael dos Santos Bezerra; Raquel de Lello Rocha Campos Cassano. NGS Soluções Genômicas: Pilar Drummond Sampaio Corrêa Mariani. FZEA-USP Pirassununga: Mirele Daiana Poleti; Raul Machado Neto; Ricardo Augusto Brassaloti; Ricardo Haddad; Rodrigo Tocantins Calado. FAMERP-SJRP: Cecília Artico Banho; Sandra Coccuzzo Sampaio; Svetoslav Nanev Slavov; Vagner Fonseca; Vincent Louis Viala |
| EPI_ISL_3102282, EPI_ISL_3102297, EPI_ISL_3102527, EPI_ISL_3102534, EPI_ISL_3102540, EPI_ISL_3102541 | SECRETARIA MUNICIPAL DE SAUDE DE TIANGUA | Analytical Competence Molecular Epidemiology Lab/ACME, Oswaldo Cruz Foundation, Ceara (FIOCRUZ CE) | Cleber Furtado Aksenen; Fabio Miyajima; Fernando Braga Stehling; Francisco Eder de Moura Lopes; Jamille Maria Mendes Bezerra; Joaquim César do Nascimento Sousa Junior; Pedro Miguel Carneiro Jeronimo; Suzana Porto Almeida e Lucas Delerino; Thais Ferreira de Oliveira; Thais de Oliveira Costa; Ticiane Cavalcante de Souza; Veridiana Pessoa Miyajima |
| EPI_ISL_1967255, EPI_ISL_1967256, EPI_ISL_1967257 | SECRETARIA MUNICIPAL DE SAUDE PORTO FERREIRA | Instituto Butantan / Mendelics | Antonio Jorge Martins; Bianca Cechetto Carlos. Mendelics: Bibiana Santos; Claudia Renata dos Santos Barros; Cíntia Bittar; David Schlesinger. Hemocentro Ribeirão Preto: Simone Kashima; Debora Botequio Moretti; Elaine Cristina Marqueze; Elaine Vieira dos Santos; Elisangela Chicaroni Mattos; Erika Freitas; Evandra Strazza Rodrigues; Felipe Allan da Silva da Costa; Flavia Aburjaile; Fábio Sossai Possebon; Guilherme Campos; Guilherme Targino Valente; Heidge Fukumasu. USP-Botucatu: Rejane Maria Tommasini Grotto; Helena Lage Ferreira; Instituto Butantan: Dimas Tadeu Covas; Jardelina de Souza Todao Bernardino; Jayme A. Souza-Neto; Jessika Cristina Chagas Lesbon; Jorge A. Petroli Marchesi; José Salvatore Leister Patané; João Paulo Kitajima; João Pessoa Araújo Jr.; Leila Sabrina Ullmann; Loyze Paola Oliveira de Lima; Luiz Aurelio de Campos Crispin. Centro de Genômica Funcional da ESALQ: Luiz Lehmann Coutinho; Luiz Carlos Junior de Alcantara; Lívia Sacchetto; Maisa C. Pereira Parra; Maria Carolina Elias; Marta Giovanetti; Marília Moraes; Maurício Lacerda Nogueira. Prefeitura de Sao Paulo: Melissa Palmieri.; Patricia Akemi Assato; Paula Rahal; Paulo Inacio da Costa; Rafael dos Santos Bezerra; Raquel de Lello Rocha Campos Cassano. NGS Soluções Genômicas: Pilar Drummond Sampaio Corrêa Mariani. FZEA-USP Pirassununga: Mirele Daiana Poleti; Raul Machado Neto; Ricardo Augusto Brassaloti; Ricardo Haddad; Rodrigo Tocantins Calado. FAMERP-SJRP: Cecília Artico Banho; Sandra Coccuzzo Sampaio; Svetoslav Nanev Slavov; Vagner Fonseca; Vincent Louis Viala |
| EPI_ISL_1967098, EPI_ISL_1967100, EPI_ISL_1967101, EPI_ISL_1967102, EPI_ISL_1967103, EPI_ISL_1967104, EPI_ISL_1967105, EPI_ISL_1967106, EPI_ISL_1967108, EPI_ISL_1967109, EPI_ISL_1967111, EPI_ISL_1967114, EPI_ISL_1967115, EPI_ISL_1967116, EPI_ISL_1967117, EPI_ISL_1967118, EPI_ISL_1967121, EPI_ISL_1967122, EPI_ISL_1967126, EPI_ISL_1967127, EPI_ISL_1967128, EPI_ISL_1967129, EPI_ISL_1967130, EPI_ISL_1967132, EPI_ISL_1967133, EPI_ISL_1967134, EPI_ISL_1967135, EPI_ISL_1967136, EPI_ISL_1967138, EPI_ISL_1967140, EPI_ISL_1967142, EPI_ISL_1967143, EPI_ISL_1967144, EPI_ISL_1967145, EPI_ISL_1967146, EPI_ISL_1967147 | | | |
| see above | SECRETARIA MUNICIPAL DE SAUDE SOROCABA | Instituto Butantan / ESALQ-USP (Piracicaba) | Antonio Jorge Martins; Bianca Cechetto Carlos. Mendelics: Bibiana Santos; Claudia Renata dos Santos Barros; Cíntia Bittar; David Schlesinger. Hemocentro Ribeirão Preto: Simone Kashima; Debora Botequio Moretti; Elaine Cristina Marqueze; Elaine Vieira dos Santos; Elisangela Chicaroni Mattos; Erika Freitas; Evandra Strazza Rodrigues; Felipe Allan da Silva da Costa; Flavia Aburjaile; Fábio Sossai Possebon; Guilherme Campos; Guilherme Targino Valente; Heidge Fukumasu. USP-Botucatu: Rejane Maria Tommasini Grotto; Helena Lage Ferreira; Instituto Butantan: Dimas Tadeu Covas; Jardelina de Souza Todao Bernardino; Jayme A. Souza-Neto; Jessika Cristina Chagas Lesbon; Jorge A. Petroli Marchesi; José Salvatore Leister Patané; João Paulo Kitajima; João Pessoa Araújo Jr.; Leila Sabrina Ullmann; Loyze Paola Oliveira de Lima; Luiz Aurelio de Campos Crispin. Centro de Genômica Funcional da ESALQ: Luiz Lehmann Coutinho; Luiz Carlos Junior de Alcantara; Lívia Sacchetto; Maisa C. Pereira Parra; Maria Carolina Elias; Marta Giovanetti; Marília Moraes; Maurício Lacerda Nogueira. Prefeitura de Sao Paulo: Melissa Palmieri.; Patricia Akemi Assato; Paula Rahal; Paulo Inacio da Costa; Rafael dos Santos Bezerra; Raquel de Lello Rocha Campos Cassano. NGS Soluções Genômicas: Pilar Drummond Sampaio Corrêa Mariani. FZEA-USP Pirassununga: Mirele Daiana Poleti; Raul Machado Neto; Ricardo Augusto Brassaloti; Ricardo Haddad; Rodrigo Tocantins Calado. FAMERP-SJRP: Cecília Artico Banho; Sandra Coccuzzo Sampaio; Svetoslav Nanev Slavov; Vagner Fonseca; Vincent Louis Viala |
| EPI_ISL_1445176, EPI_ISL_1966125, EPI_ISL_1966154, EPI_ISL_1966177, EPI_ISL_1966199, EPI_ISL_2170909 | SECRETARIA MUNICIPAL DE SAUDE SOROCABA | Instituto Butantan / Mendelics | Antonio Jorge Martins; Bianca Cechetto Carlos. Mendelics: Bibiana Santos; Bibiana Santos; Claudia Renata dos Santos Barros; Cíntia Bittar; David Schlesinger; David Schlesinger. Hemocentro Ribeirão Preto: Simone Kashima; Debora Botequio Moretti; Dimas Tadeu Covas; Elaine Cristina Marqueze; Elaine Vieira Santos; Elaine Vieira dos Santos; Elisangela Chicaroni Mattos; Erika Freitas; Evandra Strazza Rodrigues; Felipe Allan da Silva da Costa; Flavia Aburjaile; Fábio Sossai Possebon; Guilherme Campos; Guilherme Targino Valente; Heidge Fukumasu; Heidge Fukumasu. USP-Botucatu: Rejane Maria Tommasini Grotto; Helena Lage Ferreira; Instituto Butantan: Dimas Tadeu Covas; Jardelina de Souza Todao Bernardino; Jayme A. Souza-Neto; Jayme Augusto de Souza-Neto; Jessika Cristina Chagas Lesbon; Jorge A. Petroli Marchesi; José Salvatore Leister Patané; João Paulo Kitajima; João Pessoa Araújo Jr.; Leila Sabrina Ullmann; Loyze Paola Oliveira de Lima; Luiz Alcantara; Luiz Aurelio de Campos Crispin. Centro de Genômica Funcional da ESALQ: Luiz Lehmann Coutinho; Luiz Carlos Junior de Alcantara; Luiz Lehmann Coutinho; Lívia Sacchetto; Maisa C. Pereira Parra; Maria Carolina Elias; Marta Giovanetti; Marília Moraes; Maurício Lacerda Nogueira; Maurício Lacerda Nogueira. Prefeitura de Sao Paulo: Melissa Palmieri.; Patricia Akemi Assato; Paula Rahal; Paulo Inacio da Costa; Rafael dos Santos Bezerra; Raquel de Lello Rocha Campos Cassano. NGS Soluções Genômicas: Pilar Drummond Sampaio Corrêa Mariani. FZEA-USP Pirassununga: Mirele Daiana Poleti; Raul Machado Neto; Rejane Maria Tommasini Grotto; Ricardo Augusto Brassaloti; Ricardo Haddad; Ricardo Haddad; Rodrigo Tocantins Calado.; Rodrigo Tocantins Calado. FAMERP-SJRP: Cecília Artico Banho; Sandra Coccuzzo Sampaio; Sandra Coccuzzo Sampaio Vessoni; Simone Kashima; Svetoslav Nanev Slavov; Vagner Fonseca; Vincent Louis Viala |
| EPI_ISL_2346069, EPI_ISL_2346072 | SERRANA | Instituto Butantan / Mendelics | Antonio Jorge Martins; Claudia Renata dos Santos Barros; David Schlesinger; Debora Botequio Moretti; Dimas Tadeu Covas; Elaine Cristina Marqueze; Elaine Vieira Santos; Evandra Strazza Rodrigues; Heidge Fukumasu; Jayme Augusto de Souza-Neto; José Salvatore Leister Patané; Luiz Alcantara; Luiz Lehmann Coutinho; Maria Carolina Elias; Maurício Lacerda Nogueira; Rafael dos Santos Bezerra; Raul Machado Neto; Rejane Maria Tommasini Grotto; Ricardo Haddad; Sandra Coccuzzo Sampaio Vessoni; Simone Kashima; Svetoslav Nanev Slavov; Vincent Louis Viala |
| EPI_ISL_1966184, EPI_ISL_1966185 | SERV DE VIG SANITARIA EPIDEMIO E CTRL DE ZOONOZES GUARUJA | Instituto Butantan / Mendelics | Antonio Jorge Martins; Bianca Cechetto Carlos. Mendelics: Bibiana Santos; Claudia Renata dos Santos Barros; Cíntia Bittar; David Schlesinger. Hemocentro Ribeirão Preto: Simone Kashima; Debora Botequio Moretti; Elaine Cristina Marqueze; Elaine Vieira dos Santos; Elisangela Chicaroni Mattos; Erika Freitas; Evandra Strazza Rodrigues; Felipe Allan da Silva da Costa; Flavia Aburjaile; Fábio Sossai Possebon; Guilherme Campos; Guilherme Targino Valente; Heidge Fukumasu. USP-Botucatu: Rejane Maria Tommasini Grotto; Helena Lage Ferreira; Instituto Butantan: Dimas Tadeu Covas; Jardelina de Souza Todao Bernardino; Jayme A. Souza-Neto; Jessika Cristina Chagas Lesbon; Jorge A. Petroli Marchesi; José Salvatore Leister Patané; João Paulo Kitajima; João Pessoa Araújo Jr.; Leila Sabrina Ullmann; Loyze Paola Oliveira de Lima; Luiz Aurelio de Campos Crispin. Centro de Genômica Funcional da ESALQ: Luiz Lehmann Coutinho; Luiz Carlos Junior de Alcantara; Lívia Sacchetto; Maisa C. Pereira Parra; Maria Carolina Elias; Marta Giovanetti; Marília Moraes; Maurício Lacerda Nogueira. Prefeitura de Sao Paulo: Melissa Palmieri.; Patricia Akemi Assato; Paula Rahal; Paulo Inacio da Costa; Rafael dos Santos Bezerra; Raquel de Lello Rocha Campos Cassano. NGS Soluções Genômicas: Pilar Drummond Sampaio Corrêa Mariani. FZEA-USP Pirassununga: Mirele Daiana Poleti; Raul Machado Neto; Ricardo Augusto Brassaloti; Ricardo Haddad; Rodrigo Tocantins Calado. FAMERP-SJRP: Cecília Artico Banho; Sandra Coccuzzo Sampaio; Svetoslav Nanev Slavov; Vagner Fonseca; Vincent Louis Viala |
| EPI_ISL_875566, EPI_ISL_875567, EPI_ISL_875568 | SIESP L'AQUILA | Istituto Zooprofilattico Sperimentale dell'Abruzzo e Molise "G. Caporale" | Ancora M; Calistri P; Cammà C; Curini V; Di Domenico M; Di Pasquale A; Lorusso A; Mangone I; Marcacci M; Puglia I; Rinaldi A; Savini G; Scialabba S |
| EPI_ISL_2209632, EPI_ISL_2209641 | SMS IPERO | Instituto Butantan | Antonio Jorge Martins; Claudia Renata dos Santos Barros; David Schlesinger; Debora Botequio Moretti; Dimas Tadeu Covas; Elaine Cristina Marqueze; Elaine Vieira Santos; Evandra Strazza Rodrigues; Heidge Fukumasu; Jayme Augusto de Souza-Neto; José Salvatore Leister Patané; Luiz Alcantara; Luiz Lehmann Coutinho; Maria Carolina Elias; Maurício Lacerda Nogueira; Rafael dos Santos Bezerra; Raul Machado Neto; Rejane Maria Tommasini Grotto; Ricardo Haddad; Sandra Coccuzzo Sampaio Vessoni; Simone Kashima; Svetoslav Nanev Slavov; Vincent Louis Viala. |
| EPI_ISL_2344586, EPI_ISL_2345642 | SMS IPERO | Instituto Butantan / UNESP-Botucatu | Antonio Jorge Martins; Claudia Renata dos Santos Barros; David Schlesinger; Debora Botequio Moretti; Dimas Tadeu Covas; Elaine Cristina Marqueze; Elaine Vieira Santos; Evandra Strazza Rodrigues; Heidge Fukumasu; Jayme Augusto de Souza-Neto; José Salvatore Leister Patané; Luiz Alcantara; Luiz Lehmann Coutinho; Maria Carolina Elias; Maurício Lacerda Nogueira; Rafael dos Santos Bezerra; Raul Machado Neto; Rejane Maria Tommasini Grotto; Ricardo Haddad; Sandra Coccuzzo Sampaio Vessoni; Simone Kashima; Svetoslav Nanev Slavov; Vincent Louis Viala |
| EPI_ISL_2209653, EPI_ISL_2209661 | SMS SECRETARIA MUNICIPAL DE SAUDE DE BOITUVA | Instituto Butantan | Antonio Jorge Martins; Claudia Renata dos Santos Barros; David Schlesinger; Debora Botequio Moretti; Dimas Tadeu Covas; Elaine Cristina Marqueze; Elaine Vieira Santos; Evandra Strazza Rodrigues; Heidge Fukumasu; Jayme Augusto de Souza-Neto; José Salvatore Leister Patané; Luiz Alcantara; Luiz Lehmann Coutinho; Maria Carolina Elias; Maurício Lacerda Nogueira; Rafael dos Santos Bezerra; Raul Machado Neto; Rejane Maria Tommasini Grotto; Ricardo Haddad; Sandra Coccuzzo Sampaio Vessoni; Simone Kashima; Svetoslav Nanev Slavov; Vincent Louis Viala. |
| EPI_ISL_1966840, EPI_ISL_1966841, EPI_ISL_1966842 | SMS SECRETARIA MUNICIPAL DE SAUDE DE BOITUVA | Instituto Butantan / ESALQ-USP (Piracicaba) | Antonio Jorge Martins; Bianca Cechetto Carlos. Mendelics: Bibiana Santos; Claudia Renata dos Santos Barros; Cíntia Bittar; David Schlesinger. Hemocentro Ribeirão Preto: Simone Kashima; Debora Botequio Moretti; Elaine Cristina Marqueze; Elaine Vieira dos Santos; Elisangela Chicaroni Mattos; Erika Freitas; Evandra Strazza Rodrigues; Felipe Allan da Silva da Costa; Flavia Aburjaile; Fábio Sossai Possebon; Guilherme Campos; Guilherme Targino Valente; Heidge Fukumasu. USP-Botucatu: Rejane Maria Tommasini Grotto; Helena Lage Ferreira; Instituto Butantan: Dimas Tadeu Covas; Jardelina de Souza Todao Bernardino; Jayme A. Souza-Neto; Jessika Cristina Chagas Lesbon; Jorge A. Petroli Marchesi; José Salvatore Leister Patané; João Paulo Kitajima; João Pessoa Araújo Jr.; Leila Sabrina Ullmann; Loyze Paola Oliveira de Lima; Luiz Aurelio de Campos Crispin. Centro de Genômica Funcional da ESALQ: Luiz Lehmann Coutinho; Luiz Carlos Junior de Alcantara; Lívia Sacchetto; Maisa C. Pereira Parra; Maria Carolina Elias; Marta Giovanetti; Marília Moraes; Maurício Lacerda Nogueira. Prefeitura de Sao Paulo: Melissa Palmieri.; Patricia Akemi Assato; Paula Rahal; Paulo Inacio da Costa; Rafael dos Santos Bezerra; Raquel de Lello Rocha Campos Cassano. NGS Soluções Genômicas: Pilar Drummond Sampaio Corrêa Mariani. FZEA-USP Pirassununga: Mirele Daiana Poleti; Raul Machado Neto; Ricardo Augusto Brassaloti; Ricardo Haddad; Rodrigo Tocantins Calado. FAMERP-SJRP: Cecília Artico Banho; Sandra Coccuzzo Sampaio; Svetoslav Nanev Slavov; Vagner Fonseca; Vincent Louis Viala |
| EPI_ISL_2344591, EPI_ISL_2345306 | SMS SECRETARIA MUNICIPAL DE SAUDE DE BOITUVA | Instituto Butantan / UNESP-Botucatu | Antonio Jorge Martins; Claudia Renata dos Santos Barros; David Schlesinger; Debora Botequio Moretti; Dimas Tadeu Covas; Elaine Cristina Marqueze; Elaine Vieira Santos; Evandra Strazza Rodrigues; Heidge Fukumasu; Jayme Augusto de Souza-Neto; José Salvatore Leister Patané; Luiz Alcantara; Luiz Lehmann Coutinho; Maria Carolina Elias; Maurício Lacerda Nogueira; Rafael dos Santos Bezerra; Raul Machado Neto; Rejane Maria Tommasini Grotto; Ricardo Haddad; Sandra Coccuzzo Sampaio Vessoni; Simone Kashima; Svetoslav Nanev Slavov; Vincent Louis Viala |
| EPI_ISL_1671329 | STONY BROOK UNIVERSITY HOSPITAL | Wadsworth Center, New York State Department of Health | Alexis Russell; Catharine Prussing; Daryl M. Lamson; Erasmus Schneider; Erica Lasek-Nesselquist; John Kelly; Jonathan Plitnick; Kirsten St. George; Matthew Shudt; Melissa A Leisner; Navjot Singh |
| EPI_ISL_1260858, EPI_ISL_1399628, EPI_ISL_1738830, EPI_ISL_1739193, EPI_ISL_1854238, EPI_ISL_2004325 | SYNLAB | Instituto Nacional de Saude (INSA) | Borges et al |
| EPI_ISL_3355506 | SYNLAB - ANGEL DIAGNOSTICA | Corporacion CorpoGen-Universidad de los Andes-Universidad Central | Christian Romero; Jorge Duitama; Juan Manuel Anzola; Laura González; Maryam Chaib De Mares; María Mercedes Zambrano; Nelly Díaz; Patricia Del Portillo; Silvia Restrepo |
| EPI_ISL_2155037 | SYNLAB -ANGEL DIAGNOSTICA | Instituto Nacional de Salud- Dirección de Investigación en Salud Pública | Carlos Franco-Muñoz; Carmen Osorio; Diana Malo; Diego A. Álvarez-Díaz; Diego Andrés Prada; Gerardo Santamaría; Hector Alejandro Ruiz-Moreno; Jhonnatan Reales-González; Jorge Rivera; Juan Camilo Martinez; Julian Naizaque; Katherine Laiton-Donato; Lisseth Pardo; Magdalena Wiesner; Marcela Mercado-Reyes; María T. Herrera-Sepúlveda; Marta Lopez Blanco; Martha Lucia Ospina Martinez; Paola Rojas; Sergio Gomez; Sheryll Corchuelo; Ángela Alarcon Cruz |
| EPI_ISL_1571685 | SYNLAB Labor MÃ¼nchen Zentrum LMZ | Robert Koch Institute | |
| EPI_ISL_1432466 | SYNLAB MVZ Ettlingen | Robert Koch Institute | |
| EPI_ISL_1348076 | SYNLAB MVZ Leinfelden-Echterdingen | Robert Koch Institute | |
| EPI_ISL_1150858 | SYNLAB MVZ Weiden | Robert Koch Institute | |
| EPI_ISL_1628370, EPI_ISL_1715139 | Sae Servico De Atendimento Especializado | Instituto Adolfo Lutz, Interdiciplinary Procedures Center, Strategic Laboratory | Caio Vinicius Dias Lopes; Claudia Regina Gonçalves; Claudio Tavares Sacchi; Erica Valessa Ramos Gomes; Karoline Rodrigues Campos; Katia Correa de Oliveira Santos; Leonardo Jose Tadeu de Araujo |
| EPI_ISL_1821206 | Sae Servico de Atendimento Especializado | Instituto Adolfo Lutz, Interdiciplinary Procedures Center, Strategic Laboratory | Caio Vinicius Dias Lopes; Claudia Regina Gonçalves; Claudio Tavares Sacchi; Erica Valessa Ramos Gomes; Karoline Rodrigues Campos; Leonardo Jose Tadeu de Araujo |
| EPI_ISL_1493581 | Sae servico de Atendimento Especializado | Instituto Adolfo Lutz, Interdiciplinary Procedures Center, Strategic Laboratory | Caio Vinicius Dias Lopes; Claudia Regina Gonçalves; Claudio Tavares Sacchi; Erica Valessa Ramos Gomes; Karoline Rodrigues Campos |
| EPI_ISL_1315073, EPI_ISL_1468923, EPI_ISL_1468924, EPI_ISL_1468925, EPI_ISL_1468926, EPI_ISL_1468927, EPI_ISL_1468928, EPI_ISL_1468929, EPI_ISL_1468930, EPI_ISL_1468931, EPI_ISL_1468932, EPI_ISL_1468933, EPI_ISL_1468934, EPI_ISL_1468935, EPI_ISL_1468937, EPI_ISL_1468938, EPI_ISL_1468939, EPI_ISL_1468940, EPI_ISL_1468941, EPI_ISL_1468942, EPI_ISL_1468943, EPI_ISL_1468945, EPI_ISL_1468946, EPI_ISL_1468948, EPI_ISL_1791414, EPI_ISL_1791415, EPI_ISL_1791416, EPI_ISL_1791420, EPI_ISL_1791421, EPI_ISL_1791422, EPI_ISL_1791455, EPI_ISL_1794617, EPI_ISL_1794626, EPI_ISL_1794631, EPI_ISL_1794644, EPI_ISL_1794979, EPI_ISL_1795017, EPI_ISL_1795022, EPI_ISL_1969253, EPI_ISL_1969267, EPI_ISL_1969269, EPI_ISL_1969382, EPI_ISL_1969388, EPI_ISL_1969417, EPI_ISL_1969438, EPI_ISL_1969442, EPI_ISL_1969443, EPI_ISL_1969444, EPI_ISL_1969445, EPI_ISL_1969446, EPI_ISL_1969447, EPI_ISL_1969448, EPI_ISL_1969450, EPI_ISL_2329538, EPI_ISL_2329542, EPI_ISL_2329547, EPI_ISL_2329584, EPI_ISL_2329589, EPI_ISL_2329600, EPI_ISL_2329602, EPI_ISL_2422984, EPI_ISL_2423010 | | | |
| see above | San Diego County Public Health Laboratory | Andersen lab at Scripps Research | Brett Austin; Jovan Shephard; SEARCH Alliance San Diego with Ashleigh Murphy; SEARCH Alliance San Diego with Tracy Basler |
| EPI_ISL_1805403, EPI_ISL_1805404, EPI_ISL_1970952 | San Juan Regional Medical Center | New Mexico Department of Health Scientific Laboratory | Anastacia Griego-Fisher; D'eldra Malone; Ellie Johnson; Jennifer Benoit; Linda Salazar; Mark Willmon; Ratheesh Rajan |

| | | | |
|---|---|---|---|
| EPI_ISL_1628374 | Santa Casa De Cravinhos | Instituto Adolfo Lutz, Interdiciplinary Procedures Center, Strategic Laboratory | Caio Vinicius Dias Lopes; Claudia Regina Gonçalves; Claudio Tavares Sacchi; Erica Valessa Ramos Gomes; Karoline Rodrigues Campos; Katia Correa de Oliveira Santos; Leonardo Jose Tadeu de Araujo |
| EPI_ISL_1628380 | Santa Casa De Pitangueiras | Instituto Adolfo Lutz, Interdiciplinary Procedures Center, Strategic Laboratory | Caio Vinicius Dias Lopes; Claudia Regina Gonçalves; Claudio Tavares Sacchi; Erica Valessa Ramos Gomes; Karoline Rodrigues Campos; Katia Correa de Oliveira Santos; Leonardo Jose Tadeu de Araujo |
| EPI_ISL_1468417, EPI_ISL_1468419, EPI_ISL_1468424, EPI_ISL_1533715, EPI_ISL_1625974 | Santa Casa de Aracatuba Hospital Sagrado Coracao de Jesus | Instituto Adolfo Lutz, Interdiciplinary Procedures Center, Strategic Laboratory | Caio Vinicius Dias Lopes; Claudia Regina Gonçalves; Claudio Tavares Sacchi; Erica Valessa Ramos Gomes; Karoline Rodrigues Campos; Katia Correa de Oliveira Santos; Leonardo Jose Tadeu de Araujo |
| EPI_ISL_1533704 | Santa Casa de Atibaia Pro Saude | Instituto Adolfo Lutz, Interdiciplinary Procedures Center, Strategic Laboratory | Caio Vinicius Dias Lopes; Claudia Regina Gonçalves; Claudio Tavares Sacchi; Erica Valessa Ramos Gomes; Karoline Rodrigues Campos; Leonardo Jose Tadeu de Araujo |
| EPI_ISL_1468420, EPI_ISL_1468421 | Santa Casa de Birigui | Instituto Adolfo Lutz, Interdiciplinary Procedures Center, Strategic Laboratory | Caio Vinicius Dias Lopes; Claudia Regina Gonçalves; Claudio Tavares Sacchi; Erica Valessa Ramos Gomes; Karoline Rodrigues Campos |
| EPI_ISL_1821204 | Santa Casa de Cravinhos | Instituto Adolfo Lutz, Interdiciplinary Procedures Center, Strategic Laboratory | Caio Vinicius Dias Lopes; Claudia Regina Gonçalves; Claudio Tavares Sacchi; Erica Valessa Ramos Gomes; Karoline Rodrigues Campos; Leonardo Jose Tadeu de Araujo |
| EPI_ISL_1628367 | Santa Casa de Guaira | Instituto Adolfo Lutz, Interdiciplinary Procedures Center, Strategic Laboratory | Caio Vinicius Dias Lopes; Claudia Regina Gonçalves; Claudio Tavares Sacchi; Erica Valessa Ramos Gomes; Karoline Rodrigues Campos; Katia Correa de Oliveira Santos; Leonardo Jose Tadeu de Araujo |
| EPI_ISL_1533720 | Santa Casa de Penapolis | Instituto Adolfo Lutz, Interdiciplinary Procedures Center, Strategic Laboratory | Caio Vinicius Dias Lopes; Claudia Regina Gonçalves; Claudio Tavares Sacchi; Erica Valessa Ramos Gomes; Karoline Rodrigues Campos; Leonardo Jose Tadeu de Araujo |
| EPI_ISL_2500207 | Santa Clara County Public Health Laboratory | Chan-Zuckerberg Biohub | CZB Cliahub Consortium |
| EPI_ISL_1664279 | Santa Clara Valley Medical Center | Santa Clara County Public Health Laboratory | Santa Clara County Public Health Department |
| EPI_ISL_1794727, EPI_ISL_1969535, EPI_ISL_1969536, EPI_ISL_1969544 | Scripps Medical Laboratory | Andersen lab at Scripps Research | Ellen Stefanski; Ian Mchardy; SEARCH Alliance San Diego with Michael Quigley |
| EPI_ISL_2692507 | Seattle Flu Study | Seattle Flu Study | Amanda Adler; Barry R. Lutz; Benjamin Pelle; Caitlin R. Wolf; Chris D. Frazar; Deborah A. Nickerson; Elisabeth Brandstetter; Erica Ryke; Helen Y. Chu; Janet A. Englund; Jay Shendure; Jover Lee; Kairsten Fay; Kirsten Lacombe; Lea M. Starita; Mark J. Rieder; Matthew Richardson; Matthew Thompson; Melissa Truong; Michael Boeckh; Michael Famulare; Misja Ilcisin; Peter D. Han; Thomas R. Sibley; Trevor Bedford |
| EPI_ISL_1715137 | Secretaria Municipal De Saude | Instituto Adolfo Lutz, Interdiciplinary Procedures Center, Strategic Laboratory | Caio Vinicius Dias Lopes; Claudia Regina Gonçalves; Claudio Tavares Sacchi; Erica Valessa Ramos Gomes; Karoline Rodrigues Campos; Katia Correa de Oliveira Santos; Leonardo Jose Tadeu de Araujo |
| EPI_ISL_1715143 | Secretaria Municipal de Saude De Guariba | Instituto Adolfo Lutz, Interdiciplinary Procedures Center, Strategic Laboratory | Caio Vinicius Dias Lopes; Claudia Regina Gonçalves; Claudio Tavares Sacchi; Erica Valessa Ramos Gomes; Karoline Rodrigues Campos; Katia Correa de Oliveira Santos; Leonardo Jose Tadeu de Araujo |
| EPI_ISL_1533722 | Secretaria Municipal de Saude De Piracaia | Instituto Adolfo Lutz, Interdiciplinary Procedures Center, Strategic Laboratory | Caio Vinicius Dias Lopes; Claudia Regina Gonçalves; Claudio Tavares Sacchi; Erica Valessa Ramos Gomes; Karoline Rodrigues Campos; Leonardo Jose Tadeu de Araujo |
| EPI_ISL_1628375, EPI_ISL_1628376 | Secretaria Municipal de Saude de Guariba | Instituto Adolfo Lutz, Interdiciplinary Procedures Center, Strategic Laboratory | Caio Vinicius Dias Lopes; Claudia Regina Gonçalves; Claudio Tavares Sacchi; Erica Valessa Ramos Gomes; Karoline Rodrigues Campos; Katia Correa de Oliveira Santos; Leonardo Jose Tadeu de Araujo |
| EPI_ISL_1821207 | Secretaria Municipal de Saúde de Lins | Instituto Adolfo Lutz, Interdiciplinary Procedures Center, Strategic Laboratory | Caio Vinicius Dias Lopes; Claudia Regina Gonçalves; Claudio Tavares Sacchi; Erica Valessa Ramos Gomes; Karoline Rodrigues Campos; Leonardo Jose Tadeu de Araujo |
| EPI_ISL_1381214 | Sentinelles Paris | National Reference Center for Viruses of Respiratory Infections, Institut Pasteur, Paris | Angela Brisebarre; Camille Capel; Etienne Simon-Lorière; Louise Lefrançois; Marion Barbet; Maud Vanpeene; Méline Bizard; Rousset Dominique; Sylvie Behillil; Sylvie van der Werf; Vincent Enouf |
| EPI_ISL_2179707, EPI_ISL_2179713, EPI_ISL_2179717, EPI_ISL_2179718, EPI_ISL_2179719, EPI_ISL_2179721, EPI_ISL_2179722, EPI_ISL_2179723, EPI_ISL_2179724, EPI_ISL_2179725, EPI_ISL_2179726, EPI_ISL_2179727, EPI_ISL_2179756 | | | |
| see above | Servicio Microbiología Hospital La Paz | Servicio Microbiología Hospital La Paz | Elie Dahdouh; Fernando Lázaro; Jesús Mingorance Cruz; Rubén Cáceres |
| EPI_ISL_2105572, EPI_ISL_2105574, EPI_ISL_2135142, EPI_ISL_2135254, EPI_ISL_2135271, EPI_ISL_2135281, EPI_ISL_2135339, EPI_ISL_2135692, EPI_ISL_2135703, EPI_ISL_2135706, EPI_ISL_2135726, EPI_ISL_2135988, EPI_ISL_2135989, EPI_ISL_2135990, EPI_ISL_2135991, EPI_ISL_2135992, EPI_ISL_2135993, EPI_ISL_2136000, EPI_ISL_2136012, EPI_ISL_2136013, EPI_ISL_2136014, EPI_ISL_2136063, EPI_ISL_2136066, EPI_ISL_2136105, EPI_ISL_2136113, EPI_ISL_2136114, EPI_ISL_2140066, EPI_ISL_2158828 | | | |
| see above | Servicio Virosis Respiratorias-Departamento Virología-INEI | Instituto Nacional Enfermedades Infecciosas C.G.Malbran | Avaro M.; Baumeister E.; Benedetti E.; Campos J.; Cisterna D.; Dattero ME; Lorenzo F.; Molina V.; Perandones C.; Poklepovich T.; Pontoriero A.; Russo M.; Tuduri E. |
| EPI_ISL_3149732 | Servicio de Inmunología del Hospital Julio Perrando | Laboratorio de Biología Molecular, Instituto de Medicina Regional on behalf of 'Proyecto Argentino Interinstitucional de genomica de SARS-CoV-2' (PAIS Consortium) | Bettina Brusés; Florencia Vallejos Schulze; Griselda Oria; Horacio Lucero.; Javier Mussin; Laura Formichelli; María Delia Foussal; Melina Lorenzini Campos; Raúl Maximiliano Acevedo |
| EPI_ISL_1746476 | Servicio de Microbiología Clínica (Complejo Hospitalario de Navarra, Pamplona) | Centro de Secuenciación NASERTIC | Ana Miqueleiz; Ana Navascués; Carmen Ezpeleta Baquedano |
| EPI_ISL_1665136, EPI_ISL_1665139 | Servicio de Microbiología Hospital Ramón y Cajal | Servicio de Microbiología Hospital Ramón y Cajal | JC Galán; JM González-Alba; Laura Martínez. Melanie Abreu; Manuel Ponce |
| EPI_ISL_1970054, EPI_ISL_1970073, EPI_ISL_1970078, EPI_ISL_1970079, EPI_ISL_2081669, EPI_ISL_2271594, EPI_ISL_2271616 | | | |
| see above | Servicio de Microbiología. Consorcio Hospital | SeqCOVID-SPAIN consortium/IBV(CSIC) | Begoña Fuster Escrivá; Carme Salvador García; Concepción Gimeno Cardona and SeqCOVID-SPAIN consortium; María Dolores Ocete; Rafael Medina González |

| | | | |
|---|---|---|---|
| | General Universitario de Valencia | | |
| EPI_ISL_1967806, EPI_ISL_2004015 | Servicio de Microbiología. Hospital Clínico Universitario de Valencia | SeqCOVID-SPAIN consortium/IBV(CSIC) | David Navarro Ortega; Eliseo Albert Vicent; Ignacio Torres and SeqCOVID-SPAIN consortium |
| EPI_ISL_1916260 | Servicio de Microbiología. Hospital General Universitario de Castellón | SeqCOVID-SPAIN consortium/IBV(CSIC) | María Dolores Tirado Balaguer and SeqCOVID-SPAIN consortium; Rosario Moreno Muñoz |
| EPI_ISL_2000725, EPI_ISL_2016680, EPI_ISL_2016681 | Servicio de Microbiología. Hospital Universitario Doctor Peset | SeqCOVID-SPAIN consortium/IBV(CSIC) | José Miguel Nogueira Coito and SeqCOVID-SPAIN consortium; Juan Alberola Enguídanos; Juan José Camarena Miñana; Rosa González Pellicer |
| EPI_ISL_2438680, EPI_ISL_2600562, EPI_ISL_2600567, EPI_ISL_2600575, EPI_ISL_2600576 | Servizo de Microbioloxía. Complexo Hospitalario de Santiago de Compostela | Servizo de Microbioloxía. Complexo Hospitalario de Santiago de Compostela | Amparo Coira; Antonio Aguilera; Carlos García_Riestra; Daniel Navarro; Gema Barbeito; Iria Roca; José Llovo; Laura Millán; Manuela Hernández; María Luisa Pérez_del_Molino; Mercedes Treviño; Rocío trastoy; Teresa Lopez_Valiño; Xosé Costa |
| EPI_ISL_1657403 | Servizo de Microbioloxía. Hospital Lucus Augusti. Lugo. | Servizo de Microbioloxía. Complexo Hospitalario de Santiago de Compostela | Amparo Coira; Ana Rodríguez Macias; Antonio Aguilera; Antonio Moreno Flores; Carlos García_Riestra; Daniel Navarro; Gema Barbeito; Iria Roca; Javier Alba Domínguez; José Llovo; Julia Pita Carretero; Laura Millán; Laura Sante Fernández; Manuela Hernández; María Luisa Pérez_del_Molino; Mercedes Treviño; Mª José Gude González; Patricia Capón González; Rocío Trastoy; Teresa Lopez_Valiño; Xosé Costa |
| EPI_ISL_2545233, EPI_ISL_2545263, EPI_ISL_2545271, EPI_ISL_2545281 | Shamir Medical Center (Asaf Harofe) | Shamir Medical Center (Asaf Harofe) | Abu Hamad Ramzia; Adina Bar Chaim; Anna Vishnevsky; Chen Weiner; Netta Zuckerman; Nir Rainy; Patricia Benveniste-Lekovitz; Reut Sorek Abramovich; Yevgeni Yegorov |
| EPI_ISL_1969590, EPI_ISL_1969598 | Sharp HealthCare Laboratory | Andersen lab at Scripps Research | Art Mendoza; Cathy Woerle; Jacquelyn Berumen; Liam McGinnis; Omid Bakhtar; SEARCH Alliance San Diego with Aaron Harding |
| EPI_ISL_1652500, EPI_ISL_1652509, EPI_ISL_1652510, EPI_ISL_1652511, EPI_ISL_1652514, EPI_ISL_2080165, EPI_ISL_2080174, EPI_ISL_2080175, EPI_ISL_2080189, EPI_ISL_2295927, EPI_ISL_2295973, EPI_ISL_2295981, EPI_ISL_2295988, EPI_ISL_2295989, EPI_ISL_2295992, EPI_ISL_2295994 | | | |
| see above | Simple Laboratories | Gagnon Lab, Southern Illinois University | Keith Gagnon |
| EPI_ISL_1909752 | Sonora Quest Laboratories | TGen North | "Jolene Bowers; Ashlyn Pfeiffer; Chris French; Darrin Lemmer; Dave Engelthaler; Hayley Yaglom; Heather Centner; The Arizona COVID Genomics Union (ACGU)" |
| EPI_ISL_1739286, EPI_ISL_1739287, EPI_ISL_1739288 | Southern Nevada Public Health Laboratory | Southern Nevada Public Health Laboratory | Michael Picker |
| EPI_ISL_1620462, EPI_ISL_1620463 | Stanford Clinical Virology Laboratory | Santa Clara County Public Health Laboratory | Santa Clara County Public Health Department |
| EPI_ISL_3050285, EPI_ISL_3387311, EPI_ISL_3387340, EPI_ISL_3916217, EPI_ISL_3916254 | Stanford Health Care | Stanford University School of Medicine, Clinical Virology Laboratory | Becky Jiang; Bernadette Troung; Daniel Solis; Fiona Yamamoto; Fumiko Yamamoto; James Zehnder; Malaya K. Sahoo; Mamdouh Sibai; Nathan Hammond; Selamawit Bihon; and Benjamin A. Pinsky |
| EPI_ISL_1770730, EPI_ISL_1771158, EPI_ISL_1771219, EPI_ISL_2008024 | State Laboratories Division, Hawaii State Department of Health | State Laboratories Division, Hawaii State Department of Health | Ayana Garnet; Drew Kuwazaki; Edward Desmond; Pamela O'Brien; Razvan Sultana |
| EPI_ISL_2378234 | State Testing Facility | Altius Institute for Biomedical Research | Alex Isner; Alex Nguyen; Amanda Gale; Audra Johnson; Clem Green; Daniel Bates; Eric Thorland; Jacob Rodriguez; Jean Robinston; Jemma Nelson; Jessica Kunder; John Stamatoyannopoulos; Joshua Richards; Julia Wald; Kneshay Harper; Lauren Mitchell; Mark Frerker; Matt Hartman; Michael Buckley; Muhammad Halimun; Rebecca Bruders; Sadie Patraw; Sofia Olsson; Tobias Ragoczy |
| EPI_ISL_1198303, EPI_ISL_1198304, EPI_ISL_1199970, EPI_ISL_1200011, EPI_ISL_1200012, EPI_ISL_1200015, EPI_ISL_1364274, EPI_ISL_1364318, EPI_ISL_1364406, EPI_ISL_1418135, EPI_ISL_1418716, EPI_ISL_1418718, EPI_ISL_1605767, EPI_ISL_1606420, EPI_ISL_1617477, EPI_ISL_1617677, EPI_ISL_1617925, EPI_ISL_1618017, EPI_ISL_1618026, EPI_ISL_1618549, EPI_ISL_1807431, EPI_ISL_1807642, EPI_ISL_1808214, EPI_ISL_1808549, EPI_ISL_1808650, EPI_ISL_1808675, EPI_ISL_1808983, EPI_ISL_1809023, EPI_ISL_1831938, EPI_ISL_1897801, EPI_ISL_1898036, EPI_ISL_1898075, EPI_ISL_1898077, EPI_ISL_1902110, EPI_ISL_1903726, EPI_ISL_1903767, EPI_ISL_1903787, EPI_ISL_2208312, EPI_ISL_2208428, EPI_ISL_2214976, EPI_ISL_2217247, EPI_ISL_2218106, EPI_ISL_2220583, EPI_ISL_2221389, EPI_ISL_2221847, EPI_ISL_2222188, EPI_ISL_2255538, EPI_ISL_2255651, EPI_ISL_2255694, EPI_ISL_2257212, EPI_ISL_2257214, EPI_ISL_2257516, EPI_ISL_2416020, EPI_ISL_2416023, EPI_ISL_3781496, EPI_ISL_3790127 | | | |
| see above | Swedish national genomic surveillance program of SARS-CoV-2 | The Public Health Agency of Sweden | Alma Brolund; Maria Lind Karlberg; Maximilian Riess; Swedish national genomic surveillance program of SARS-CoV-2 |
| EPI_ISL_1499585, EPI_ISL_1499588 | Synlab | Instituto Nacional de Saude (INSA) | Borges et al |
| EPI_ISL_2443058, EPI_ISL_2443059, EPI_ISL_2443062, EPI_ISL_2443064 | São Jose do Rio Preto | Instituto Butantan / FAMERP | Antonio Jorge Martins; Claudia Renata dos Santos Barros; David Schlesinger; Debora Botequio Moretti; Dimas Tadeu Covas; Elaine Cristina Marqueze; Elaine Vieira Santos; Evandra Strazza Rodrigues; Heidge Fukumasu; Jayme Augusto de Souza-Neto; José Salvatore Leister Patané; Luiz Alcantara; Luiz Lehmann Coutinho; Maria Carolina Elias; Maurício Lacerda Nogueira; Rafael dos Santos Bezerra; Raul Machado Neto; Rejane Maria Tommasini Grotto; Ricardo Haddad; Sandra Coccuzzo Sampaio Vessoni; Simone Kashima; Svetoslav Nanev Slavov; Vincent Louis Viala |
| EPI_ISL_2042410 | TN Division of Laboratory Services | Pathogen Discovery, Respiratory Viruses Branch, Division of Viral Diseases, Centers for Disease Control and Prevention | Adam Retchless; Anna Kelleher; Anna Uehara; Brian Lynch; Clinton R. Paden; Dhwani Batra; Haibin Wang; Han Jia Justin Ng; Jasmine Padilla; Jing Zhang; Justin Lee; Krista Queen; Mark Burroughs; Mili Sheth; Morgan Davis; Peter Cook; Rachel Marine; Sarah Nobles; Suxiang Tong; Tara Coalter; Yan Li; Ying Tao |
| EPI_ISL_1760594 | TXDSHS | TXDSHS | Anita Pokharel; Bonnie Oh; Chun Wang; Grace Kubin; Jenny Zhang; Lorraine Rodriguez; Maliha Rahman; Mayela Pedrueza; Myong Koag; Rachel Lee; Rashmi Tuladhar |
| EPI_ISL_1710724, EPI_ISL_1754154, EPI_ISL_1754156, EPI_ISL_1754158, EPI_ISL_1754160, EPI_ISL_1754161, EPI_ISL_1754163, EPI_ISL_1754164, EPI_ISL_1754174, EPI_ISL_2081360, EPI_ISL_2081379, EPI_ISL_2383082, EPI_ISL_2383084, EPI_ISL_2484171, EPI_ISL_2626628, EPI_ISL_2626631, EPI_ISL_2930433, EPI_ISL_2930435, EPI_ISL_2930438 | | | |
| see above | Tampa General Hospital Esoteric Lab | Tampa General Hospital Esoteric Research & Development Lab | Amorce Lima; Deanna Becker; Dominic Uy; Elaine Vendrone; Grant Vestal; Jon Faughn; Jorge Mecias-Frias; Suzane Silbert; Vicki Healer |
| EPI_ISL_1540817, EPI_ISL_1540831, EPI_ISL_1625680, EPI_ISL_1625681, EPI_ISL_1739426, EPI_ISL_1739478 | The Jackson Laboratory | The Jackson Laboratory | Adams M; Bergeron D; Kelly K; Li L; Long J; Omerza G; Renzette N |
| EPI_ISL_3930265, EPI_ISL_3930266, EPI_ISL_3930270, EPI_ISL_3930271, EPI_ISL_3930272, EPI_ISL_3930278, EPI_ISL_3930286 | | | |
| see above | The National University Hospital of Iceland | deCODE genetics | Agnar Helgason; Alma Moller; Arna B Agustsdottir; Arnaldur Gylfason; Asgeir Sigurdsson; Aslaug Jonasdottir; Berglind Eiriksdottir; Bjarni Thorbjornsson; Brynjar O Jensson; Daniel F Gudbjartsson; Droplaug N Magnusdottir; Elisabet E Gardarsdottir; Emil A Thorarensen; Gardar Sveinbjornsson; Gisli Masson; Gudmundur Georgsson; Gudmundur L Norddahl; Gudrun Sigmundsdottir; Hakon Jonsson; Hannes Eggertsson; Hilma Holm; Ingileif Jonsdottir; Jona Saemundsdottir; Kamilla S Josefsdottir; Kari Stefansson; Karl G Kristinsson; Kjartan R Gudmundsson; Kristin E Sveinsdottir; Kristjan E Hjorleifsson; Louise le Roux; Maney Sveinsdottir; Olafia S Gretarsdottir; Olafur T Magnusson; Pall Melsted; Patrick Sulem; Run Fridriksdottir; Solvi Rognvaldsson; Thora R Gunnarsdottir; Thordur Kristjansson; Thorolfur Gudnason; Unnur Thorsteinsdottir |
| EPI_ISL_2153744, EPI_ISL_2153745, EPI_ISL_2153767, EPI_ISL_2153800, EPI_ISL_2200101, EPI_ISL_2200110, EPI_ISL_2200133, EPI_ISL_2200145, EPI_ISL_2341359, EPI_ISL_2341362, EPI_ISL_2341378, EPI_ISL_2341386, EPI_ISL_2341395, EPI_ISL_2382473, EPI_ISL_2382486, EPI_ISL_2382783 | | | |
| see above | The Ohio State University Applied Microbiology Services Laboratory | The Ohio State University Applied Microbiology Services Laboratory | Seth A. Faith PhD |
| EPI_ISL_2228215 | TriCore Reference Laboratories | Center for Global Health, University of New Mexico Health Sciences Center | Cecilia Thompson; Darrell Dinwiddie; Daryl Domman; Karissa Culbreath; Kendra Pesko; Kurt Schwalm; Valerie Morley |
| EPI_ISL_2227542 | TriCore Reference | Center for Global | Cecilia Thompson; Darrell Dinwiddie; Daryl Domman; Karissa Culbreath; Kendra Pesko; Kurt Schwalm; Valerie Morley |

| | | | |
|---|---|---|---|
| | Laboratories | Health, University of New Mexico Health Sciences Center | |
| EPI_ISL_3062201 | U.O. Microbiologia Laboratorio Unico Centro Servizi - AUSL della Romagna | U.O. Microbiologia, Laboratorio Unico Centro Servizi - AUSL della Romagna | Giorgio Dirani |
| EPI_ISL_1960604 | UAB Diagnostikos laboratorija | National Public Health Surveillance Laboratory | Ana Steponkiene; Danas Baksa; Jelena Razmuk; Lukas Vasionis; Lukas Zemaitis; Migle Gabrielaite; Svajune Muralyte |
| EPI_ISL_3912246, EPI_ISL_3912247 | UAPS ANASTACIO MAGALHAES | Analytical Competence Molecular Epidemiology Lab/ACME, Oswaldo Cruz Foundation, Ceara (FIOCRUZ CE) | Cleber Furtado Aksenen; Fabio Miyajima; Fernando Braga Stehling; Francisco Eder de Moura Lopes; Jamille Maria Mendes Bezerra; Joaquim Cesar do Nascimento Sousa Junior; Pedro Miguel Carneiro Jeronimo; Suzana Porto Almeida & Lucas Delerino on behalf of COVID-19 FIOCRUZ Genomic Network; Thais Ferreira de Oliveira; Thais de Oliveira Costa; Ticiane Cavalcante de Souza; Veridiana Pessoa Miyajima |
| EPI_ISL_3912248 | UAPS ARGEU HERBSTER | Analytical Competence Molecular Epidemiology Lab/ACME, Oswaldo Cruz Foundation, Ceara (FIOCRUZ CE) | Cleber Furtado Aksenen; Fabio Miyajima; Fernando Braga Stehling; Francisco Eder de Moura Lopes; Jamille Maria Mendes Bezerra; Joaquim Cesar do Nascimento Sousa Junior; Pedro Miguel Carneiro Jeronimo; Suzana Porto Almeida & Lucas Delerino on behalf of COVID-19 FIOCRUZ Genomic Network; Thais Ferreira de Oliveira; Thais de Oliveira Costa; Ticiane Cavalcante de Souza; Veridiana Pessoa Miyajima |
| EPI_ISL_3912249 | UAPS BENEDITO ARTHUR DE CARVALHO | Analytical Competence Molecular Epidemiology Lab/ACME, Oswaldo Cruz Foundation, Ceara (FIOCRUZ CE) | Cleber Furtado Aksenen; Fabio Miyajima; Fernando Braga Stehling; Francisco Eder de Moura Lopes; Jamille Maria Mendes Bezerra; Joaquim Cesar do Nascimento Sousa Junior; Pedro Miguel Carneiro Jeronimo; Suzana Porto Almeida & Lucas Delerino on behalf of COVID-19 FIOCRUZ Genomic Network; Thais Ferreira de Oliveira; Thais de Oliveira Costa; Ticiane Cavalcante de Souza; Veridiana Pessoa Miyajima |
| EPI_ISL_3912252 | UAPS FRANCISCO MONTEIRO | Analytical Competence Molecular Epidemiology Lab/ACME, Oswaldo Cruz Foundation, Ceara (FIOCRUZ CE) | Cleber Furtado Aksenen; Fabio Miyajima; Fernando Braga Stehling; Francisco Eder de Moura Lopes; Jamille Maria Mendes Bezerra; Joaquim Cesar do Nascimento Sousa Junior; Pedro Miguel Carneiro Jeronimo; Suzana Porto Almeida & Lucas Delerino on behalf of COVID-19 FIOCRUZ Genomic Network; Thais Ferreira de Oliveira; Thais de Oliveira Costa; Ticiane Cavalcante de Souza; Veridiana Pessoa Miyajima |
| EPI_ISL_3912253 | UAPS GOTHARDO PEIXOTO | Analytical Competence Molecular Epidemiology Lab/ACME, Oswaldo Cruz Foundation, Ceara (FIOCRUZ CE) | Cleber Furtado Aksenen; Fabio Miyajima; Fernando Braga Stehling; Francisco Eder de Moura Lopes; Jamille Maria Mendes Bezerra; Joaquim Cesar do Nascimento Sousa Junior; Pedro Miguel Carneiro Jeronimo; Suzana Porto Almeida & Lucas Delerino on behalf of COVID-19 FIOCRUZ Genomic Network; Thais Ferreira de Oliveira; Thais de Oliveira Costa; Ticiane Cavalcante de Souza; Veridiana Pessoa Miyajima |
| EPI_ISL_3912255 | UAPS LICINIO NUNES DE MIRANDA | Analytical Competence Molecular Epidemiology Lab/ACME, Oswaldo Cruz Foundation, Ceara (FIOCRUZ CE) | Cleber Furtado Aksenen; Fabio Miyajima; Fernando Braga Stehling; Francisco Eder de Moura Lopes; Jamille Maria Mendes Bezerra; Joaquim Cesar do Nascimento Sousa Junior; Pedro Miguel Carneiro Jeronimo; Suzana Porto Almeida & Lucas Delerino on behalf of COVID-19 FIOCRUZ Genomic Network; Thais Ferreira de Oliveira; Thais de Oliveira Costa; Ticiane Cavalcante de Souza; Veridiana Pessoa Miyajima |
| EPI_ISL_3912256, EPI_ISL_3912258 | UAPS MACIEL DE BRITO | Analytical Competence Molecular Epidemiology Lab/ACME, Oswaldo Cruz Foundation, Ceara (FIOCRUZ CE) | Cleber Furtado Aksenen; Fabio Miyajima; Fernando Braga Stehling; Francisco Eder de Moura Lopes; Jamille Maria Mendes Bezerra; Joaquim Cesar do Nascimento Sousa Junior; Pedro Miguel Carneiro Jeronimo; Suzana Porto Almeida & Lucas Delerino on behalf of COVID-19 FIOCRUZ Genomic Network; Thais Ferreira de Oliveira; Thais de Oliveira Costa; Ticiane Cavalcante de Souza; Veridiana Pessoa Miyajima |
| EPI_ISL_3912259, EPI_ISL_3912260 | UAPS RIGOBERTO ROMERO | Analytical Competence Molecular Epidemiology Lab/ACME, Oswaldo Cruz Foundation, Ceara (FIOCRUZ CE) | Cleber Furtado Aksenen; Fabio Miyajima; Fernando Braga Stehling; Francisco Eder de Moura Lopes; Jamille Maria Mendes Bezerra; Joaquim Cesar do Nascimento Sousa Junior; Pedro Miguel Carneiro Jeronimo; Suzana Porto Almeida & Lucas Delerino on behalf of COVID-19 FIOCRUZ Genomic Network; Thais Ferreira de Oliveira; Thais de Oliveira Costa; Ticiane Cavalcante de Souza; Veridiana Pessoa Miyajima |
| EPI_ISL_3912261 | UAPS ROBERTO BRUNO | Analytical Competence Molecular Epidemiology Lab/ACME, Oswaldo Cruz Foundation, Ceara (FIOCRUZ CE) | Cleber Furtado Aksenen; Fabio Miyajima; Fernando Braga Stehling; Francisco Eder de Moura Lopes; Jamille Maria Mendes Bezerra; Joaquim Cesar do Nascimento Sousa Junior; Pedro Miguel Carneiro Jeronimo; Suzana Porto Almeida & Lucas Delerino on behalf of COVID-19 FIOCRUZ Genomic Network; Thais Ferreira de Oliveira; Thais de Oliveira Costa; Ticiane Cavalcante de Souza; Veridiana Pessoa Miyajima |
| EPI_ISL_3912262, EPI_ISL_3912263 | UAPS TEREZINHA PARENTE | Analytical Competence Molecular Epidemiology Lab/ACME, Oswaldo Cruz Foundation, Ceara (FIOCRUZ CE) | Cleber Furtado Aksenen; Fabio Miyajima; Fernando Braga Stehling; Francisco Eder de Moura Lopes; Jamille Maria Mendes Bezerra; Joaquim Cesar do Nascimento Sousa Junior; Pedro Miguel Carneiro Jeronimo; Suzana Porto Almeida & Lucas Delerino on behalf of COVID-19 FIOCRUZ Genomic Network; Thais Ferreira de Oliveira; Thais de Oliveira Costa; Ticiane Cavalcante de Souza; Veridiana Pessoa Miyajima |
| EPI_ISL_3912266 | UBASF DE MANGABEIRA | Analytical Competence Molecular Epidemiology Lab/ACME, Oswaldo Cruz Foundation, Ceara (FIOCRUZ CE) | Cleber Furtado Aksenen; Fabio Miyajima; Fernando Braga Stehling; Francisco Eder de Moura Lopes; Jamille Maria Mendes Bezerra; Joaquim Cesar do Nascimento Sousa Junior; Pedro Miguel Carneiro Jeronimo; Suzana Porto Almeida & Lucas Delerino on behalf of COVID-19 FIOCRUZ Genomic Network; Thais Ferreira de Oliveira; Thais de Oliveira Costa; Ticiane Cavalcante de Souza; Veridiana Pessoa Miyajima |
| EPI_ISL_3102509 | UBASF PLANALTO | Analytical Competence Molecular Epidemiology Lab/ACME, Oswaldo Cruz Foundation, Ceara (FIOCRUZ CE) | Cleber Furtado Aksenen; Fabio Miyajima; Fernando Braga Stehling; Francisco Eder de Moura Lopes; Jamille Maria Mendes Bezerra; Joaquim César do Nascimento Sousa Junior; Pedro Miguel Carneiro Jeronimo; Suzana Porto Almeida e Lucas Delerino; Thais Ferreira de Oliveira; Thais de Oliveira Costa; Ticiane Cavalcante de Souza; Veridiana Pessoa Miyajima |
| EPI_ISL_1731574, EPI_ISL_1752637 | UBDS Dr Marco Antonio Sahao Vila Virginia | Instituto Adolfo Lutz, Interdiciplinary Procedures Center, Strategic Laboratory | Caio Vinicius Dias Lopes; Claudia Regina Gonçalves; Claudio Tavares Sacchi; Erica Valessa Ramos Gomes; Karoline Rodrigues Campos; Katia Correa de Oliveira Santos; Leonardo Jose Tadeu de Araujo |
| EPI_ISL_1795287, EPI_ISL_1795289, EPI_ISL_2345525, EPI_ISL_2345527 | UBS ALCIMINIO DE ASSIS LOURENCO BADY BASSITT | Instituto Butantan / ESALQ-Piracicaba | Antonio Jorge Martins; Bianca Cechetto Carlos. Mendelics: Bibiana Santos; Claudia Renata dos Santos Barros; David Schlesinger; David Schlesinger. Hemocentro Ribeirão Preto: Simone Kashima; Debora Botequio Moretti; Debora Botequio Moretti. Centro de Genômica Funcional da ESALQ: Luiz Lehmann Coutinho; Dimas Tadeu Covas; Elaine Cristina Marqueze; Elaine Vieira Santos; Elaine Vieira dos Santos; Elisangela Chicaroni Mattos; Erika Freitas; Evandra Strazza Rodrigues; Felipe Allan da Silva da Costa; Flavia Aburjaile; Guilherme Targino Valente; Heidge Fukumasu; Heidge Fukumasu. USP-Botucatu: Rejane Maria Tommasini Grotto; Instituto Butantan: Alexander Roberto Precioso; Jayme A. Souza-Neto; Jayme Augusto de Souza-Neto; Jessika Cristina Chagas Lesbon; José Salvatore Leister Patané; João Paulo Kitajima; Luiz Alcantara; Luiz Carlos Junior de Alcantara; Luiz Lehmann Coutinho; Maria Carolina Elias; Marta Giovanetti; Maurício Lacerda Nogueira; Patricia Akemi Assato; Rafael dos Santos Bezerra; Raquel de Lello Rocha Campos Cassano. NGS Soluções Genômicas: Pilar Drummond Sampaio Corrêa Mariani. FZEA-USP Pirassununga: Mirele Daiana Poleti; Raul Machado Neto; Rejane Maria Tommasini Grotto; Ricardo Augusto Brassaloti; Ricardo Haddad; Rodrigo Tocantins Calado.; Sandra Coccuzzo Sampaio; Sandra Coccuzzo Sampaio Vessoni; Simone Kashima; Svetoslav Nanev Slavov; Vagner Fonseca; Vincent Louis Viala |
| EPI_ISL_1966870, EPI_ISL_1966875, EPI_ISL_1966878, EPI_ISL_1966879, EPI_ISL_1966882, EPI_ISL_1966885, EPI_ISL_1966886, EPI_ISL_1966888, EPI_ISL_1966890 | | | |
| see above | UBS ATALAIA DR MARIO AGUIAR FILHO | Instituto Butantan / ESALQ-USP (Piracicaba) | Antonio Jorge Martins; Bianca Cechetto Carlos. Mendelics: Bibiana Santos; Claudia Renata dos Santos Barros; Cíntia Bittar; David Schlesinger. Hemocentro Ribeirão Preto: Simone Kashima; Debora Botequio Moretti; Elaine Cristina Marqueze; Elaine Vieira dos Santos; Elisangela Chicaroni Mattos; Erika Freitas; Evandra Strazza Rodrigues; Felipe Allan da Silva da Costa; Flavia Aburjaile; Fábio Sossai Possebon; Guilherme Campos; Guilherme Targino Valente; Heidge Fukumasu. USP-Botucatu: Rejane Maria Tommasini Grotto; Helena Lage Ferreira; Instituto Butantan: Dimas Tadeu Covas; Jardelina de Souza Todao Bernardino; Jayme A. Souza-Neto; Jessika Cristina Chagas Lesbon; Jorge A. Petroli Marchesi; José Salvatore Leister Patané; João Paulo Kitajima; João Pessoa Araújo Jr.; Leila Sabrina Ullmann; Loyze Paola Oliveira de Lima; Luiz Aurelio de Campos Crispin. Centro de Genômica Funcional da ESALQ: Luiz Lehmann Coutinho; Luiz Carlos Junior de Alcantara; Lívia Sacchetto; Maisa C. Pereira Parra; Maria Carolina Elias; Marta Giovanetti; Marília Moraes; Maurício Lacerda Nogueira. Prefeitura de Sao Paulo: Melissa Palmieri.; Patricia Akemi Assato; Paula Rahal; Paulo Inacio da Costa; Rafael dos Santos Bezerra; Raquel de Lello Rocha Campos Cassano. NGS Soluções Genômicas: Pilar Drummond Sampaio Corrêa Mariani. FZEA-USP Pirassununga: Mirele Daiana Poleti; Raul Machado Neto; Ricardo Augusto Brassaloti; Ricardo Haddad; Rodrigo Tocantins Calado. FAMERP-SJRP: Cecília Artico Banho; Sandra Coccuzzo Sampaio; Svetoslav Nanev Slavov; Vagner Fonseca; Vincent Louis Viala |
| EPI_ISL_2614596 | UBS Atalaia | Instituto Adolfo Lutz, Interdiciplinary Procedures Center, Strategic Laboratory | Caio Vinicius Dias Lopes; Claudia Regina Gonçalves; Claudio Tavares Sacchi; Erica Valessa Ramos Gomes; Karoline Rodrigues Campos; Leonardo Jose Tadeu de Araujo |
| EPI_ISL_3912279 | UBS DE SITIO ALEGRE I | Analytical Competence Molecular Epidemiology Lab/ACME, Oswaldo Cruz Foundation, Ceara (FIOCRUZ CE) | Cleber Furtado Aksenen; Fabio Miyajima; Fernando Braga Stehling; Francisco Eder de Moura Lopes; Jamille Maria Mendes Bezerra; Joaquim Cesar do Nascimento Sousa Junior; Pedro Miguel Carneiro Jeronimo; Suzana Porto Almeida & Lucas Delerino on behalf of COVID-19 FIOCRUZ Genomic Network; Thais Ferreira de Oliveira; Thais de Oliveira Costa; Ticiane Cavalcante de Souza; Veridiana Pessoa Miyajima |
| EPI_ISL_1967159, EPI_ISL_1967160, EPI_ISL_1967161, EPI_ISL_1967162 | UBS DE TABAPUA | Instituto Butantan / ESALQ-USP (Piracicaba) | Antonio Jorge Martins; Bianca Cechetto Carlos. Mendelics: Bibiana Santos; Claudia Renata dos Santos Barros; Cíntia Bittar; David Schlesinger. Hemocentro Ribeirão Preto: Simone Kashima; Debora Botequio Moretti; Elaine Cristina Marqueze; Elaine Vieira dos Santos; Elisangela Chicaroni Mattos; Erika Freitas; Evandra Strazza Rodrigues; Felipe Allan da Silva da Costa; Flavia Aburjaile; Fábio Sossai Possebon; Guilherme Campos; Guilherme Targino Valente; Heidge Fukumasu. USP-Botucatu: Rejane Maria Tommasini Grotto; Helena Lage Ferreira; Instituto Butantan: Dimas Tadeu Covas; Jardelina de Souza Todao Bernardino; Jayme A. Souza-Neto; Jessika Cristina Chagas Lesbon; Jorge A. Petroli Marchesi; José Salvatore Leister Patané; João Paulo Kitajima; João Pessoa Araújo Jr.; Leila Sabrina Ullmann; Loyze Paola Oliveira de Lima; Luiz Aurelio de Campos Crispin. Centro de Genômica Funcional da ESALQ: Luiz Lehmann Coutinho; Luiz Carlos Junior de Alcantara; Lívia Sacchetto; Maisa C. Pereira Parra; Maria Carolina Elias; Marta Giovanetti; Marília Moraes; Maurício Lacerda Nogueira. Prefeitura de Sao Paulo: Melissa Palmieri.; Patricia Akemi Assato; Paula Rahal; Paulo Inacio da Costa; Rafael dos Santos Bezerra; Raquel de Lello Rocha Campos Cassano. NGS Soluções Genômicas: Pilar Drummond Sampaio Corrêa Mariani. FZEA-USP Pirassununga: Mirele Daiana Poleti; Raul Machado Neto; Ricardo Augusto Brassaloti; Ricardo Haddad; Rodrigo Tocantins Calado. FAMERP-SJRP: Cecília Artico Banho; Sandra Coccuzzo Sampaio; Svetoslav Nanev Slavov; Vagner Fonseca; Vincent Louis Viala |
| EPI_ISL_2345892, EPI_ISL_2345893, | UBS DO CIPO EMBU GUACU | Instituto Butantan / FZEA-USP-Pirassununga | Antonio Jorge Martins; Claudia Renata dos Santos Barros; David Schlesinger; Debora Botequio Moretti; Dimas Tadeu Covas; Elaine Cristina Marqueze; Elaine Vieira Santos; Evandra Strazza Rodrigues; Heidge Fukumasu; Jayme Augusto de Souza-Neto; José Salvatore Leister Patané; Luiz Alcantara; Luiz Lehmann Coutinho; Maria Carolina Elias; Maurício Lacerda Nogueira; Rafael dos Santos Bezerra; Raul Machado Neto; Rejane Maria Tommasini Grotto; Ricardo Haddad; Sandra Coccuzzo Sampaio Vessoni; Simone Kashima; Svetoslav Nanev Slavov; Vincent Louis Viala |

| | | | |
|---|---|---|---|
| EPI_ISL_2345900 | | | |
| EPI_ISL_1966543, EPI_ISL_1966544 | UBS DR ABELARDO PINHEIRO GUIMARAES SCRPARDO | Instituto Butantan / FZEA-USP (Pirassununga) | Antonio Jorge Martins; Bianca Cechetto Carlos. Mendelics: Bibiana Santos; Claudia Renata dos Santos Barros; Cíntia Bittar; David Schlesinger. Hemocentro Ribeirão Preto: Simone Kashima; Debora Botequio Moretti; Elaine Cristina Marqueze; Elaine Vieira dos Santos; Elisangela Chicaroni Mattos; Erika Freitas; Evandra Strazza Rodrigues; Felipe Allan da Silva da Costa; Flavia Aburjaile; Fábio Sossai Possebon; Guilherme Campos; Guilherme Targino Valente; Heidge Fukumasu. USP-Botucatu: Rejane Maria Tommasini Grotto; Helena Lage Ferreira; Instituto Butantan: Dimas Tadeu Covas; Jardelina de Souza Todao Bernardino; Jayme A. Souza-Neto; Jessika Cristina Chagas Lesbon; Jorge A. Petroli Marchesi; José Salvatore Leister Patané; João Paulo Kitajima; João Pessoa Araújo Jr.; Leila Sabrina Ullmann; Loyze Paola Oliveira de Lima; Luiz Aurelio de Campos Crispin. Centro de Genômica Funcional da ESALQ: Luiz Lehmann Coutinho; Luiz Carlos Junior de Alcantara; Lívia Sacchetto; Maisa C. Pereira Parra; Maria Carolina Elias; Marta Giovanetti; Marília Moraes; Maurício Lacerda Nogueira. Prefeitura de Sao Paulo: Melissa Palmieri.; Patricia Akemi Assato; Paula Rahal; Paulo Inacio da Costa; Rafael dos Santos Bezerra; Raquel de Lello Rocha Campos Cassano. NGS Soluções Genômicas: Pilar Drummond Sampaio Corrêa Mariani. FZEA-USP Pirassununga: Mirele Daiana Poleti; Raul Machado Neto; Ricardo Augusto Brassaloti; Ricardo Haddad; Rodrigo Tocantins Calado. FAMERP-SJRP: Cecília Artico Banho; Sandra Coccuzzo Sampaio; Svetoslav Nanev Slavov; Vagner Fonseca; Vincent Louis Viala |
| EPI_ISL_1966809, EPI_ISL_1966811 | UBS DR ABELARDO PINHEIRO GUIMARAES SCRPARDO | Instituto Butantan / Mendelics | Antonio Jorge Martins; Bianca Cechetto Carlos. Mendelics: Bibiana Santos; Claudia Renata dos Santos Barros; Cíntia Bittar; David Schlesinger. Hemocentro Ribeirão Preto: Simone Kashima; Debora Botequio Moretti; Elaine Cristina Marqueze; Elaine Vieira dos Santos; Elisangela Chicaroni Mattos; Erika Freitas; Evandra Strazza Rodrigues; Felipe Allan da Silva da Costa; Flavia Aburjaile; Fábio Sossai Possebon; Guilherme Campos; Guilherme Targino Valente; Heidge Fukumasu. USP-Botucatu: Rejane Maria Tommasini Grotto; Helena Lage Ferreira; Instituto Butantan: Dimas Tadeu Covas; Jardelina de Souza Todao Bernardino; Jayme A. Souza-Neto; Jessika Cristina Chagas Lesbon; Jorge A. Petroli Marchesi; José Salvatore Leister Patané; João Paulo Kitajima; João Pessoa Araújo Jr.; Leila Sabrina Ullmann; Loyze Paola Oliveira de Lima; Luiz Aurelio de Campos Crispin. Centro de Genômica Funcional da ESALQ: Luiz Lehmann Coutinho; Luiz Carlos Junior de Alcantara; Lívia Sacchetto; Maisa C. Pereira Parra; Maria Carolina Elias; Marta Giovanetti; Marília Moraes; Maurício Lacerda Nogueira. Prefeitura de Sao Paulo: Melissa Palmieri.; Patricia Akemi Assato; Paula Rahal; Paulo Inacio da Costa; Rafael dos Santos Bezerra; Raquel de Lello Rocha Campos Cassano. NGS Soluções Genômicas: Pilar Drummond Sampaio Corrêa Mariani. FZEA-USP Pirassununga: Mirele Daiana Poleti; Raul Machado Neto; Ricardo Augusto Brassaloti; Ricardo Haddad; Rodrigo Tocantins Calado. FAMERP-SJRP: Cecília Artico Banho; Sandra Coccuzzo Sampaio; Svetoslav Nanev Slavov; Vagner Fonseca; Vincent Louis Viala |
| EPI_ISL_1966289 | UBS DR ALLY ALAHMAR DE BARRETOS | Instituto Butantan / Mendelics | Antonio Jorge Martins; Bianca Cechetto Carlos. Mendelics: Bibiana Santos; Claudia Renata dos Santos Barros; Cíntia Bittar; David Schlesinger. Hemocentro Ribeirão Preto: Simone Kashima; Debora Botequio Moretti; Elaine Cristina Marqueze; Elaine Vieira dos Santos; Elisangela Chicaroni Mattos; Erika Freitas; Evandra Strazza Rodrigues; Felipe Allan da Silva da Costa; Flavia Aburjaile; Fábio Sossai Possebon; Guilherme Campos; Guilherme Targino Valente; Heidge Fukumasu. USP-Botucatu: Rejane Maria Tommasini Grotto; Helena Lage Ferreira; Instituto Butantan: Dimas Tadeu Covas; Jardelina de Souza Todao Bernardino; Jayme A. Souza-Neto; Jessika Cristina Chagas Lesbon; Jorge A. Petroli Marchesi; José Salvatore Leister Patané; João Paulo Kitajima; João Pessoa Araújo Jr.; Leila Sabrina Ullmann; Loyze Paola Oliveira de Lima; Luiz Aurelio de Campos Crispin. Centro de Genômica Funcional da ESALQ: Luiz Lehmann Coutinho; Luiz Carlos Junior de Alcantara; Lívia Sacchetto; Maisa C. Pereira Parra; Maria Carolina Elias; Marta Giovanetti; Marília Moraes; Maurício Lacerda Nogueira. Prefeitura de Sao Paulo: Melissa Palmieri.; Patricia Akemi Assato; Paula Rahal; Paulo Inacio da Costa; Rafael dos Santos Bezerra; Raquel de Lello Rocha Campos Cassano. NGS Soluções Genômicas: Pilar Drummond Sampaio Corrêa Mariani. FZEA-USP Pirassununga: Mirele Daiana Poleti; Raul Machado Neto; Ricardo Augusto Brassaloti; Ricardo Haddad; Rodrigo Tocantins Calado. FAMERP-SJRP: Cecília Artico Banho; Sandra Coccuzzo Sampaio; Svetoslav Nanev Slavov; Vagner Fonseca; Vincent Louis Viala |
| EPI_ISL_1966990, EPI_ISL_1966994 | UBS DR HELIO MIGLIARI | Instituto Butantan / ESALQ-USP (Piracicaba) | Antonio Jorge Martins; Bianca Cechetto Carlos. Mendelics: Bibiana Santos; Claudia Renata dos Santos Barros; Cíntia Bittar; David Schlesinger. Hemocentro Ribeirão Preto: Simone Kashima; Debora Botequio Moretti; Elaine Cristina Marqueze; Elaine Vieira dos Santos; Elisangela Chicaroni Mattos; Erika Freitas; Evandra Strazza Rodrigues; Felipe Allan da Silva da Costa; Flavia Aburjaile; Fábio Sossai Possebon; Guilherme Campos; Guilherme Targino Valente; Heidge Fukumasu. USP-Botucatu: Rejane Maria Tommasini Grotto; Helena Lage Ferreira; Instituto Butantan: Dimas Tadeu Covas; Jardelina de Souza Todao Bernardino; Jayme A. Souza-Neto; Jessika Cristina Chagas Lesbon; Jorge A. Petroli Marchesi; José Salvatore Leister Patané; João Paulo Kitajima; João Pessoa Araújo Jr.; Leila Sabrina Ullmann; Loyze Paola Oliveira de Lima; Luiz Aurelio de Campos Crispin. Centro de Genômica Funcional da ESALQ: Luiz Lehmann Coutinho; Luiz Carlos Junior de Alcantara; Lívia Sacchetto; Maisa C. Pereira Parra; Maria Carolina Elias; Marta Giovanetti; Marília Moraes; Maurício Lacerda Nogueira. Prefeitura de Sao Paulo: Melissa Palmieri.; Patricia Akemi Assato; Paula Rahal; Paulo Inacio da Costa; Rafael dos Santos Bezerra; Raquel de Lello Rocha Campos Cassano. NGS Soluções Genômicas: Pilar Drummond Sampaio Corrêa Mariani. FZEA-USP Pirassununga: Mirele Daiana Poleti; Raul Machado Neto; Ricardo Augusto Brassaloti; Ricardo Haddad; Rodrigo Tocantins Calado. FAMERP-SJRP: Cecília Artico Banho; Sandra Coccuzzo Sampaio; Svetoslav Nanev Slavov; Vagner Fonseca; Vincent Louis Viala |
| EPI_ISL_1966534 | UBS DR HELIO MIGLIARI | Instituto Butantan / FZEA-USP (Pirassununga) | Antonio Jorge Martins; Bianca Cechetto Carlos. Mendelics: Bibiana Santos; Claudia Renata dos Santos Barros; Cíntia Bittar; David Schlesinger. Hemocentro Ribeirão Preto: Simone Kashima; Debora Botequio Moretti; Elaine Cristina Marqueze; Elaine Vieira dos Santos; Elisangela Chicaroni Mattos; Erika Freitas; Evandra Strazza Rodrigues; Felipe Allan da Silva da Costa; Flavia Aburjaile; Fábio Sossai Possebon; Guilherme Campos; Guilherme Targino Valente; Heidge Fukumasu. USP-Botucatu: Rejane Maria Tommasini Grotto; Helena Lage Ferreira; Instituto Butantan: Dimas Tadeu Covas; Jardelina de Souza Todao Bernardino; Jayme A. Souza-Neto; Jessika Cristina Chagas Lesbon; Jorge A. Petroli Marchesi; José Salvatore Leister Patané; João Paulo Kitajima; João Pessoa Araújo Jr.; Leila Sabrina Ullmann; Loyze Paola Oliveira de Lima; Luiz Aurelio de Campos Crispin. Centro de Genômica Funcional da ESALQ: Luiz Lehmann Coutinho; Luiz Carlos Junior de Alcantara; Lívia Sacchetto; Maisa C. Pereira Parra; Maria Carolina Elias; Marta Giovanetti; Marília Moraes; Maurício Lacerda Nogueira. Prefeitura de Sao Paulo: Melissa Palmieri.; Patricia Akemi Assato; Paula Rahal; Paulo Inacio da Costa; Rafael dos Santos Bezerra; Raquel de Lello Rocha Campos Cassano. NGS Soluções Genômicas: Pilar Drummond Sampaio Corrêa Mariani. FZEA-USP Pirassununga: Mirele Daiana Poleti; Raul Machado Neto; Ricardo Augusto Brassaloti; Ricardo Haddad; Rodrigo Tocantins Calado. FAMERP-SJRP: Cecília Artico Banho; Sandra Coccuzzo Sampaio; Svetoslav Nanev Slavov; Vagner Fonseca; Vincent Louis Viala |
| EPI_ISL_1966252, EPI_ISL_1966253 | UBS DR MOHANNA ADAS | Instituto Butantan / Mendelics | Antonio Jorge Martins; Bianca Cechetto Carlos. Mendelics: Bibiana Santos; Claudia Renata dos Santos Barros; Cíntia Bittar; David Schlesinger. Hemocentro Ribeirão Preto: Simone Kashima; Debora Botequio Moretti; Elaine Cristina Marqueze; Elaine Vieira dos Santos; Elisangela Chicaroni Mattos; Erika Freitas; Evandra Strazza Rodrigues; Felipe Allan da Silva da Costa; Flavia Aburjaile; Fábio Sossai Possebon; Guilherme Campos; Guilherme Targino Valente; Heidge Fukumasu. USP-Botucatu: Rejane Maria Tommasini Grotto; Helena Lage Ferreira; Instituto Butantan: Dimas Tadeu Covas; Jardelina de Souza Todao Bernardino; Jayme A. Souza-Neto; Jessika Cristina Chagas Lesbon; Jorge A. Petroli Marchesi; José Salvatore Leister Patané; João Paulo Kitajima; João Pessoa Araújo Jr.; Leila Sabrina Ullmann; Loyze Paola Oliveira de Lima; Luiz Aurelio de Campos Crispin. Centro de Genômica Funcional da ESALQ: Luiz Lehmann Coutinho; Luiz Carlos Junior de Alcantara; Lívia Sacchetto; Maisa C. Pereira Parra; Maria Carolina Elias; Marta Giovanetti; Marília Moraes; Maurício Lacerda Nogueira. Prefeitura de Sao Paulo: Melissa Palmieri.; Patricia Akemi Assato; Paula Rahal; Paulo Inacio da Costa; Rafael dos Santos Bezerra; Raquel de Lello Rocha Campos Cassano. NGS Soluções Genômicas: Pilar Drummond Sampaio Corrêa Mariani. FZEA-USP Pirassununga: Mirele Daiana Poleti; Raul Machado Neto; Ricardo Augusto Brassaloti; Ricardo Haddad; Rodrigo Tocantins Calado. FAMERP-SJRP: Cecília Artico Banho; Sandra Coccuzzo Sampaio; Svetoslav Nanev Slavov; Vagner Fonseca; Vincent Louis Viala |
| EPI_ISL_2362270 | UBS GUILHERMINA DE NOBREGA ABREU | Instituto Butantan / Mendelics | Antonio Jorge Martins; Claudia Renata dos Santos Barros; David Schlesinger; Debora Botequio Moretti; Dimas Tadeu Covas; Elaine Cristina Marqueze; Elaine Vieira Santos; Evandra Strazza Rodrigues; Heidge Fukumasu; Jayme Augusto de Souza-Neto; José Salvatore Leister Patané; Luiz Alcantara; Luiz Lehmann Coutinho; Maria Carolina Elias; Maurício Lacerda Nogueira; Rafael dos Santos Bezerra; Raul Machado Neto; Rejane Maria Tommasini Grotto; Ricardo Haddad; Sandra Coccuzzo Sampaio Vessoni; Simone Kashima; Svetoslav Nanev Slavov; Vincent Louis Viala |
| EPI_ISL_1795324, EPI_ISL_2345585 | UBS II DE NARANDIBA | Instituto Butantan / ESALQ-Piracicaba | Antonio Jorge Martins; Bianca Cechetto Carlos. Mendelics: Bibiana Santos; Claudia Renata dos Santos Barros; David Schlesinger; David Schlesinger. Hemocentro Ribeirão Preto: Simone Kashima; Debora Botequio Moretti; Debora Botequio Moretti. Centro de Genômica Funcional da ESALQ: Luiz Lehmann Coutinho; Dimas Tadeu Covas; Elaine Cristina Marqueze; Elaine Vieira Santos; Elaine Vieira dos Santos; Elisangela Chicaroni Mattos; Erika Freitas; Evandra Strazza Rodrigues; Felipe Allan da Silva da Costa; Flavia Aburjaile; Guilherme Targino Valente; Heidge Fukumasu; Heidge Fukumasu. USP-Botucatu: Rejane Maria Tommasini Grotto; Instituto Butantan: Alexander Roberto Precioso; Jayme A. Souza-Neto; Jayme Augusto de Souza-Neto; Jessika Cristina Chagas Lesbon; José Salvatore Leister Patané; João Paulo Kitajima; Luiz Alcantara; Luiz Carlos Junior de Alcantara; Luiz Lehmann Coutinho; Maria Carolina Elias; Marta Giovanetti; Maurício Lacerda Nogueira; Patricia Akemi Assato; Rafael dos Santos Bezerra; Raquel de Lello Rocha Campos Cassano. NGS Soluções Genômicas: Pilar Drummond Sampaio Corrêa Mariani. FZEA-USP Pirassununga: Mirele Daiana Poleti; Raul Machado Neto; Rejane Maria Tommasini Grotto; Ricardo Augusto Brassaloti; Ricardo Haddad; Rodrigo Tocantins Calado.; Sandra Coccuzzo Sampaio; Sandra Coccuzzo Sampaio Vessoni; Simone Kashima; Svetoslav Nanev Slavov; Vagner Fonseca; Vincent Louis Viala |
| EPI_ISL_2344743 | UBS II DE PIRAPOZINHO C SERVICO DE EACS E ESF | Instituto Butantan / ESALQ-Piracicaba | Antonio Jorge Martins; Claudia Renata dos Santos Barros; David Schlesinger; Debora Botequio Moretti; Dimas Tadeu Covas; Elaine Cristina Marqueze; Elaine Vieira Santos; Evandra Strazza Rodrigues; Heidge Fukumasu; Jayme Augusto de Souza-Neto; José Salvatore Leister Patané; Luiz Alcantara; Luiz Lehmann Coutinho; Maria Carolina Elias; Maurício Lacerda Nogueira; Rafael dos Santos Bezerra; Raul Machado Neto; Rejane Maria Tommasini Grotto; Ricardo Haddad; Sandra Coccuzzo Sampaio Vessoni; Simone Kashima; Svetoslav Nanev Slavov; Vincent Louis Viala |
| EPI_ISL_1795326, EPI_ISL_2345587 | UBS II DE REGENTE FEIJO | Instituto Butantan / ESALQ-Piracicaba | Antonio Jorge Martins; Bianca Cechetto Carlos. Mendelics: Bibiana Santos; Claudia Renata dos Santos Barros; David Schlesinger; David Schlesinger. Hemocentro Ribeirão Preto: Simone Kashima; Debora Botequio Moretti; Debora Botequio Moretti. Centro de Genômica Funcional da ESALQ: Luiz Lehmann Coutinho; Dimas Tadeu Covas; Elaine Cristina Marqueze; Elaine Vieira Santos; Elaine Vieira dos Santos; Elisangela Chicaroni Mattos; Erika Freitas; Evandra Strazza Rodrigues; Felipe Allan da Silva da Costa; Flavia Aburjaile; Guilherme Targino Valente; Heidge Fukumasu; Heidge Fukumasu. USP-Botucatu: Rejane Maria Tommasini Grotto; Instituto Butantan: Alexander Roberto Precioso; Jayme A. Souza-Neto; Jayme Augusto de Souza-Neto; Jessika Cristina Chagas Lesbon; José Salvatore Leister Patané; João Paulo Kitajima; Luiz Alcantara; Luiz Carlos Junior de Alcantara; Luiz Lehmann Coutinho; Maria Carolina Elias; Marta Giovanetti; Maurício Lacerda Nogueira; Patricia Akemi Assato; Rafael dos Santos Bezerra; Raquel de Lello Rocha Campos Cassano. NGS Soluções Genômicas: Pilar Drummond Sampaio Corrêa Mariani. FZEA-USP Pirassununga: Mirele Daiana Poleti; Raul Machado Neto; Rejane Maria Tommasini Grotto; Ricardo Augusto Brassaloti; Ricardo Haddad; Rodrigo Tocantins Calado.; Sandra Coccuzzo Sampaio; Sandra Coccuzzo Sampaio Vessoni; Simone Kashima; Svetoslav Nanev Slavov; Vagner Fonseca; Vincent Louis Viala |
| EPI_ISL_2344751 | UBS II DE TACIBA | Instituto Butantan / ESALQ-Piracicaba | Antonio Jorge Martins; Claudia Renata dos Santos Barros; David Schlesinger; Debora Botequio Moretti; Dimas Tadeu Covas; Elaine Cristina Marqueze; Elaine Vieira Santos; Evandra Strazza Rodrigues; Heidge Fukumasu; Jayme Augusto de Souza-Neto; José Salvatore Leister Patané; Luiz Alcantara; Luiz Lehmann Coutinho; Maria Carolina Elias; Maurício Lacerda Nogueira; Rafael dos Santos Bezerra; Raul Machado Neto; Rejane Maria Tommasini Grotto; Ricardo Haddad; Sandra Coccuzzo Sampaio Vessoni; Simone Kashima; Svetoslav Nanev Slavov; Vincent Louis Viala |
| EPI_ISL_2209680 | UBS II DE TANABI MILTON MARTINS PERCHES | Instituto Butantan | Antonio Jorge Martins; Claudia Renata dos Santos Barros; David Schlesinger; Debora Botequio Moretti; Dimas Tadeu Covas; Elaine Cristina Marqueze; Elaine Vieira Santos; Evandra Strazza Rodrigues; Heidge Fukumasu; Jayme Augusto de Souza-Neto; José Salvatore Leister Patané; Luiz Alcantara; Luiz Lehmann Coutinho; Maria Carolina Elias; Maurício Lacerda Nogueira; Rafael dos Santos Bezerra; Raul Machado Neto; Rejane Maria Tommasini Grotto; Ricardo Haddad; Sandra Coccuzzo Sampaio Vessoni; Simone Kashima; Svetoslav Nanev Slavov; Vincent Louis Viala. |
| EPI_ISL_1795251, EPI_ISL_1795252, EPI_ISL_1795253, EPI_ISL_1795254, EPI_ISL_1795255, EPI_ISL_1795256, EPI_ISL_1795257, EPI_ISL_2345485, EPI_ISL_2345486, EPI_ISL_2345487, EPI_ISL_2345488, EPI_ISL_2345489, EPI_ISL_2345490, EPI_ISL_2345491 | | | |
| see above | UBS II DE TANABI MILTON MARTINS PERCHES | Instituto Butantan / ESALQ-Piracicaba | Antonio Jorge Martins; Bianca Cechetto Carlos. Mendelics: Bibiana Santos; Claudia Renata dos Santos Barros; David Schlesinger; David Schlesinger. Hemocentro Ribeirão Preto: Simone Kashima; Debora Botequio Moretti; Debora Botequio Moretti. Centro de Genômica Funcional da ESALQ: Luiz Lehmann Coutinho; Dimas Tadeu Covas; Elaine Cristina Marqueze; Elaine Vieira Santos; Elaine Vieira dos Santos; Elisangela Chicaroni Mattos; Erika Freitas; Evandra Strazza Rodrigues; Felipe Allan da Silva da Costa; Flavia Aburjaile; Guilherme Targino Valente; Heidge Fukumasu; Heidge Fukumasu. USP-Botucatu: Rejane Maria Tommasini Grotto; Instituto Butantan: Alexander Roberto Precioso; Jayme A. Souza-Neto; Jayme Augusto de Souza-Neto; Jessika Cristina Chagas Lesbon; José Salvatore Leister Patané; João Paulo Kitajima; Luiz Alcantara; Luiz Carlos Junior de Alcantara; Luiz Lehmann Coutinho; Maria Carolina Elias; Marta Giovanetti; Maurício Lacerda Nogueira; Patricia Akemi Assato; Rafael dos Santos Bezerra; Raquel de Lello Rocha Campos Cassano. NGS Soluções Genômicas: Pilar Drummond Sampaio Corrêa Mariani. FZEA-USP Pirassununga: Mirele Daiana Poleti; Raul Machado Neto; Rejane Maria Tommasini Grotto; Ricardo Augusto Brassaloti; Ricardo Haddad; Rodrigo Tocantins Calado.; Sandra Coccuzzo Sampaio; Sandra Coccuzzo Sampaio Vessoni; Simone Kashima; Svetoslav Nanev Slavov; Vagner Fonseca; Vincent Louis Viala |
| EPI_ISL_2344590 | UBS II DE TANABI MILTON MARTINS PERCHES | Instituto Butantan / UNESP-Botucatu | Antonio Jorge Martins; Claudia Renata dos Santos Barros; David Schlesinger; Debora Botequio Moretti; Dimas Tadeu Covas; Elaine Cristina Marqueze; Elaine Vieira Santos; Evandra Strazza Rodrigues; Heidge Fukumasu; Jayme Augusto de Souza-Neto; José Salvatore Leister Patané; Luiz Alcantara; Luiz Lehmann Coutinho; Maria Carolina Elias; Maurício Lacerda Nogueira; Rafael dos Santos Bezerra; Raul Machado Neto; Rejane Maria Tommasini Grotto; Ricardo Haddad; Sandra Coccuzzo Sampaio Vessoni; Simone Kashima; Svetoslav Nanev Slavov; Vincent Louis Viala |
| EPI_ISL_1795328, EPI_ISL_1795330, EPI_ISL_2345589, EPI_ISL_2345591 | UBS II DR EXPEDITO SHIZUO KUROCE | Instituto Butantan / ESALQ-Piracicaba | Antonio Jorge Martins; Bianca Cechetto Carlos. Mendelics: Bibiana Santos; Claudia Renata dos Santos Barros; David Schlesinger; David Schlesinger. Hemocentro Ribeirão Preto: Simone Kashima; Debora Botequio Moretti; Debora Botequio Moretti. Centro de Genômica Funcional da ESALQ: Luiz Lehmann Coutinho; Dimas Tadeu Covas; Elaine Cristina Marqueze; Elaine Vieira Santos; Elaine Vieira dos Santos; Elisangela Chicaroni Mattos; Erika Freitas; Evandra Strazza Rodrigues; Felipe Allan da Silva da Costa; Flavia Aburjaile; Guilherme Targino Valente; Heidge Fukumasu; Heidge Fukumasu. USP-Botucatu: Rejane Maria Tommasini Grotto; Instituto Butantan: Alexander Roberto Precioso; Jayme A. Souza-Neto; Jayme Augusto de Souza-Neto; Jessika Cristina Chagas Lesbon; José Salvatore Leister Patané; João Paulo Kitajima; Luiz Alcantara; Luiz Carlos Junior de Alcantara; Luiz Lehmann Coutinho; Maria Carolina Elias; Marta Giovanetti; Maurício Lacerda Nogueira; Patricia Akemi Assato; Rafael dos Santos Bezerra; Raquel de Lello Rocha Campos Cassano. NGS Soluções Genômicas: Pilar Drummond Sampaio Corrêa Mariani. FZEA-USP Pirassununga: Mirele Daiana Poleti; Raul Machado Neto; Rejane Maria Tommasini Grotto; Ricardo Augusto Brassaloti; Ricardo Haddad; Rodrigo Tocantins Calado.; Sandra Coccuzzo Sampaio; Sandra Coccuzzo Sampaio Vessoni; Simone Kashima; Svetoslav Nanev Slavov; Vagner Fonseca; Vincent Louis Viala |
| EPI_ISL_1966995 | UBS III DE PARIQUERA ACU PARIQUERA ACU | Instituto Butantan / ESALQ-USP (Piracicaba) | Antonio Jorge Martins; Bianca Cechetto Carlos. Mendelics: Bibiana Santos; Claudia Renata dos Santos Barros; Cíntia Bittar; David Schlesinger. Hemocentro Ribeirão Preto: Simone Kashima; Debora Botequio Moretti; Elaine Cristina Marqueze; Elaine Vieira dos Santos; Elisangela Chicaroni Mattos; Erika Freitas; Evandra Strazza Rodrigues; Felipe Allan da Silva da Costa; Flavia Aburjaile; Fábio Sossai Possebon; Guilherme Campos; Guilherme Targino Valente; Heidge Fukumasu. USP-Botucatu: Rejane Maria Tommasini Grotto; Helena Lage Ferreira; Instituto Butantan: Dimas Tadeu Covas; Jardelina de Souza Todao Bernardino; Jayme A. Souza-Neto; Jessika Cristina Chagas Lesbon; Jorge A. Petroli Marchesi; José Salvatore Leister Patané; João Paulo Kitajima; João Pessoa Araújo Jr.; Leila Sabrina Ullmann; Loyze Paola Oliveira de Lima; Luiz Aurelio de Campos Crispin. Centro de Genômica Funcional da ESALQ: Luiz Lehmann Coutinho; Luiz Carlos Junior de Alcantara; Lívia Sacchetto; Maisa C. Pereira Parra; Maria Carolina Elias; Marta Giovanetti; Marília Moraes; Maurício Lacerda Nogueira. Prefeitura de Sao Paulo: Melissa Palmieri.; Patricia Akemi Assato; Paula Rahal; Paulo Inacio da Costa; Rafael dos Santos Bezerra; Raquel de Lello Rocha Campos Cassano. NGS Soluções Genômicas: Pilar Drummond Sampaio Corrêa Mariani. FZEA-USP Pirassununga: Mirele Daiana Poleti; Raul Machado Neto; Ricardo Augusto Brassaloti; Ricardo Haddad; Rodrigo Tocantins Calado. FAMERP-SJRP: Cecília Artico Banho; Sandra Coccuzzo Sampaio; Svetoslav Nanev Slavov; Vagner Fonseca; Vincent Louis Viala |
| EPI_ISL_1795332, EPI_ISL_1795335, EPI_ISL_2345593, EPI_ISL_2345599 | UBS III DE RANCHARIA | Instituto Butantan / ESALQ-Piracicaba | Antonio Jorge Martins; Bianca Cechetto Carlos. Mendelics: Bibiana Santos; Claudia Renata dos Santos Barros; David Schlesinger; David Schlesinger. Hemocentro Ribeirão Preto: Simone Kashima; Debora Botequio Moretti; Debora Botequio Moretti. Centro de Genômica Funcional da ESALQ: Luiz Lehmann Coutinho; Dimas Tadeu Covas; Elaine Cristina Marqueze; Elaine Vieira Santos; Elaine Vieira dos Santos; Elisangela Chicaroni Mattos; Erika Freitas; Evandra Strazza Rodrigues; Felipe Allan da Silva da Costa; Flavia Aburjaile; Guilherme Targino Valente; Heidge Fukumasu; Heidge Fukumasu. USP-Botucatu: Rejane Maria Tommasini Grotto; Instituto Butantan: Alexander Roberto Precioso; Jayme A. Souza-Neto; Jayme Augusto de Souza-Neto; Jessika Cristina Chagas Lesbon; José Salvatore Leister Patané; João Paulo Kitajima; Luiz Alcantara; Luiz Carlos Junior de Alcantara; Luiz Lehmann Coutinho; Maria Carolina Elias; Marta Giovanetti; Maurício Lacerda Nogueira; Patricia Akemi Assato; Rafael dos Santos Bezerra; Raquel de Lello Rocha Campos Cassano. NGS Soluções Genômicas: Pilar Drummond Sampaio Corrêa Mariani. FZEA-USP Pirassununga: Mirele Daiana Poleti; Raul Machado Neto; Rejane Maria Tommasini Grotto; Ricardo Augusto Brassaloti; Ricardo Haddad; Rodrigo Tocantins Calado.; Sandra Coccuzzo Sampaio; Sandra Coccuzzo Sampaio Vessoni; Simone Kashima; Svetoslav Nanev Slavov; Vagner Fonseca; Vincent Louis Viala |
| EPI_ISL_2345970 | UBS IRMA AGUEDA MARIA JAIME | Instituto Butantan | Antonio Jorge Martins; Claudia Renata dos Santos Barros; David Schlesinger; Debora Botequio Moretti; Dimas Tadeu Covas; Elaine Cristina Marqueze; Elaine Vieira Santos; Evandra Strazza Rodrigues; Heidge Fukumasu; Jayme Augusto de Souza-Neto; José Salvatore Leister Patané; Luiz Alcantara; Luiz Lehmann Coutinho; Maria Carolina Elias; Maurício Lacerda Nogueira; Rafael dos Santos Bezerra; Raul Machado Neto; Rejane Maria Tommasini Grotto; Ricardo Haddad; Sandra Coccuzzo Sampaio Vessoni; Simone Kashima; Svetoslav Nanev Slavov; Vincent Louis Viala |
| EPI_ISL_2345982 | UBS JARDIM ACACIO | Instituto Butantan | Antonio Jorge Martins; Claudia Renata dos Santos Barros; David Schlesinger; Debora Botequio Moretti; Dimas Tadeu Covas; Elaine Cristina Marqueze; Elaine Vieira Santos; Evandra Strazza Rodrigues; Heidge Fukumasu; Jayme Augusto de Souza-Neto; José Salvatore Leister Patané; Luiz Alcantara; Luiz Lehmann Coutinho; Maria Carolina Elias; Maurício Lacerda Nogueira; Rafael dos Santos Bezerra; Raul Machado Neto; Rejane Maria Tommasini Grotto; Ricardo Haddad; Sandra Coccuzzo Sampaio Vessoni; Simone Kashima; Svetoslav Nanev Slavov; Vincent Louis Viala |
| EPI_ISL_2346025, EPI_ISL_2346026, EPI_ISL_2346034, EPI_ISL_2346039, EPI_ISL_2346042, EPI_ISL_2346044 | UBS JARDIM CAIUBY | Instituto Butantan | Antonio Jorge Martins; Claudia Renata dos Santos Barros; David Schlesinger; Debora Botequio Moretti; Dimas Tadeu Covas; Elaine Cristina Marqueze; Elaine Vieira Santos; Evandra Strazza Rodrigues; Heidge Fukumasu; Jayme Augusto de Souza-Neto; José Salvatore Leister Patané; Luiz Alcantara; Luiz Lehmann Coutinho; Maria Carolina Elias; Maurício Lacerda Nogueira; Rafael dos Santos Bezerra; Raul Machado Neto; Rejane Maria Tommasini Grotto; Ricardo Haddad; Sandra Coccuzzo Sampaio Vessoni; Simone Kashima; Svetoslav Nanev Slavov; Vincent Louis Viala |
| EPI_ISL_1795160, EPI_ISL_2345371 | UBS JARDIM ITAMARATY | Instituto Butantan / ESALQ-Piracicaba | Antonio Jorge Martins; Bianca Cechetto Carlos. Mendelics: Bibiana Santos; Claudia Renata dos Santos Barros; David Schlesinger; David Schlesinger. Hemocentro Ribeirão Preto: Simone Kashima; Debora Botequio Moretti; Debora Botequio Moretti. Centro de Genômica Funcional da ESALQ: Luiz Lehmann Coutinho; Dimas Tadeu Covas; Elaine Cristina Marqueze; Elaine Vieira Santos; Elaine Vieira dos Santos; Elisangela Chicaroni Mattos; Erika Freitas; Evandra Strazza Rodrigues; Felipe Allan da Silva da Costa; Flavia Aburjaile; Guilherme Targino Valente; Heidge Fukumasu; Heidge Fukumasu. USP-Botucatu: Rejane Maria Tommasini Grotto; Instituto Butantan: Alexander Roberto Precioso; Jayme A. Souza-Neto; Jayme Augusto de Souza-Neto; Jessika Cristina Chagas Lesbon; José Salvatore Leister Patané; João Paulo Kitajima; Luiz Alcantara; Luiz Carlos Junior de Alcantara; Luiz Lehmann Coutinho; Maria Carolina Elias; Marta Giovanetti; Maurício Lacerda Nogueira; Patricia Akemi Assato; Rafael dos Santos Bezerra; Raquel de Lello Rocha Campos Cassano. NGS Soluções Genômicas: Pilar Drummond Sampaio Corrêa Mariani. FZEA-USP Pirassununga: Mirele Daiana Poleti; Raul Machado Neto; Rejane Maria Tommasini Grotto; Ricardo Augusto Brassaloti; Ricardo Haddad; Rodrigo Tocantins Calado.; Sandra Coccuzzo Sampaio; Sandra Coccuzzo Sampaio Vessoni; Simone Kashima; Svetoslav Nanev Slavov; Vagner Fonseca; Vincent Louis Viala |
| EPI_ISL_2345986, EPI_ISL_2345987 | UBS JARDIM ODETE | Instituto Butantan | Antonio Jorge Martins; Claudia Renata dos Santos Barros; David Schlesinger; Debora Botequio Moretti; Dimas Tadeu Covas; Elaine Cristina Marqueze; Elaine Vieira Santos; Evandra Strazza Rodrigues; Heidge Fukumasu; Jayme Augusto de Souza-Neto; José Salvatore Leister Patané; Luiz Alcantara; Luiz Lehmann Coutinho; Maria Carolina Elias; Maurício Lacerda Nogueira; Rafael dos Santos Bezerra; Raul Machado Neto; Rejane Maria Tommasini Grotto; Ricardo Haddad; Sandra Coccuzzo Sampaio Vessoni; Simone Kashima; Svetoslav Nanev Slavov; Vincent Louis Viala |
| EPI_ISL_2345993, EPI_ISL_2346023 | UBS JARDIM PAINEIRA | Instituto Butantan | Antonio Jorge Martins; Claudia Renata dos Santos Barros; David Schlesinger; Debora Botequio Moretti; Dimas Tadeu Covas; Elaine Cristina Marqueze; Elaine Vieira Santos; Evandra Strazza Rodrigues; Heidge Fukumasu; Jayme Augusto de Souza-Neto; José Salvatore Leister Patané; Luiz Alcantara; Luiz Lehmann Coutinho; Maria Carolina Elias; Maurício Lacerda Nogueira; Rafael dos Santos Bezerra; Raul Machado Neto; Rejane Maria Tommasini Grotto; Ricardo Haddad; Sandra Coccuzzo Sampaio Vessoni; Simone Kashima; Svetoslav Nanev Slavov; Vincent Louis Viala |
| EPI_ISL_2345976 | UBS JARDIM PRIMAVERA | Instituto Butantan | Antonio Jorge Martins; Claudia Renata dos Santos Barros; David Schlesinger; Debora Botequio Moretti; Dimas Tadeu Covas; Elaine Cristina Marqueze; Elaine Vieira Santos; Evandra Strazza Rodrigues; Heidge Fukumasu; Jayme Augusto de Souza-Neto; José Salvatore Leister Patané; Luiz Alcantara; Luiz Lehmann Coutinho; Maria Carolina Elias; Maurício Lacerda Nogueira; Rafael dos Santos Bezerra; Raul Machado Neto; Rejane Maria Tommasini Grotto; Ricardo Haddad; Sandra Coccuzzo Sampaio Vessoni; Simone Kashima; Svetoslav Nanev Slavov; Vincent Louis Viala |
| EPI_ISL_1966918 | UBS JARDIM VITORINO IPERO | Instituto Butantan / ESALQ-USP (Piracicaba) | Antonio Jorge Martins; Bianca Cechetto Carlos. Mendelics: Bibiana Santos; Claudia Renata dos Santos Barros; Cíntia Bittar; David Schlesinger. Hemocentro Ribeirão Preto: Simone Kashima; Debora Botequio Moretti; Elaine Cristina Marqueze; Elaine Vieira dos Santos; Elisangela Chicaroni Mattos; Erika Freitas; Evandra Strazza Rodrigues; Felipe Allan da Silva da Costa; Flavia Aburjaile; Fábio Sossai Possebon; Guilherme Campos; Guilherme Targino Valente; Heidge Fukumasu. USP-Botucatu: Rejane Maria Tommasini Grotto; Helena Lage Ferreira; Instituto Butantan: Dimas Tadeu Covas; Jardelina de Souza Todao Bernardino; Jayme A. Souza-Neto; Jessika Cristina Chagas |

| | | | |
|---|---|---|---|
| | | | Lesbon; Jorge A. Petroli Marchesi; José Salvatore Leister Patané; João Paulo Kitajima; João Pessoa Araújo Jr.; Leila Sabrina Ullmann; Loyze Paola Oliveira de Lima; Luiz Aurelio de Campos Crispin. Centro de Genômica Funcional da ESALQ: Luiz Lehmann Coutinho; Luiz Carlos Junior de Alcantara; Lívia Sacchetto; Maisa C. Pereira Parra; Maria Carolina Elias; Marta Giovanetti; Marília Moraes; Maurício Lacerda Nogueira. Prefeitura de Sao Paulo: Melissa Palmieri.; Patricia Akemi Assato; Paula Rahal; Paulo Inacio da Costa; Rafael dos Santos Bezerra; Raquel de Lello Rocha Campos Cassano. NGS Soluções Genômicas: Pilar Drummond Sampaio Corrêa Mariani. FZEA-USP Pirassununga: Mirele Daiana Poleti; Raul Machado Neto; Ricardo Augusto Brassaloti; Ricardo Haddad; Rodrigo Tocantins Calado. FAMERP-SJRP: Cecília Artico Banho; Sandra Coccuzzo Sampaio; Svetoslav Nanev Slavov; Vagner Fonseca; Vincent Louis Viala |
| EPI_ISL_1966728, EPI_ISL_1966730 | UBS JOSE SABINO FERREIRA | Instituto Butantan | Antonio Jorge Martins; Bianca Cechetto Carlos. Mendelics: Bibiana Santos; Claudia Renata dos Santos Barros; Cíntia Bittar; David Schlesinger. Hemocentro Ribeirão Preto: Simone Kashima; Debora Botequio Moretti; Elaine Cristina Marqueze; Elaine Vieira dos Santos; Elisangela Chicaroni Mattos; Erika Freitas; Evandra Strazza Rodrigues; Felipe Allan da Silva da Costa; Flavia Aburjaile; Fábio Sossai Possebon; Guilherme Campos; Guilherme Targino Valente; Heidge Fukumasu. USP-Botucatu: Rejane Maria Tommasini Grotto; Helena Lage Ferreira; Instituto Butantan: Dimas Tadeu Covas; Jardelina de Souza Todao Bernardino; Jayme A. Souza-Neto; Jessika Cristina Chagas Lesbon; Jorge A. Petroli Marchesi; José Salvatore Leister Patané; João Paulo Kitajima; João Pessoa Araújo Jr.; Leila Sabrina Ullmann; Loyze Paola Oliveira de Lima; Luiz Aurelio de Campos Crispin. Centro de Genômica Funcional da ESALQ: Luiz Lehmann Coutinho; Luiz Carlos Junior de Alcantara; Lívia Sacchetto; Maisa C. Pereira Parra; Maria Carolina Elias; Marta Giovanetti; Marília Moraes; Maurício Lacerda Nogueira. Prefeitura de Sao Paulo: Melissa Palmieri.; Patricia Akemi Assato; Paula Rahal; Paulo Inacio da Costa; Rafael dos Santos Bezerra; Raquel de Lello Rocha Campos Cassano. NGS Soluções Genômicas: Pilar Drummond Sampaio Corrêa Mariani. FZEA-USP Pirassununga: Mirele Daiana Poleti; Raul Machado Neto; Ricardo Augusto Brassaloti; Ricardo Haddad; Rodrigo Tocantins Calado. FAMERP-SJRP: Cecília Artico Banho; Sandra Coccuzzo Sampaio; Svetoslav Nanev Slavov; Vagner Fonseca; Vincent Louis Viala |
| EPI_ISL_2614584 | UBS Jardim Jovaia | Instituto Adolfo Lutz, Interdiciplinary Procedures Center, Strategic Laboratory | Caio Vinicius Dias Lopes; Claudia Regina Gonçalves; Claudio Tavares Sacchi; Erica Valessa Ramos Gomes; Karoline Rodrigues Campos; Leonardo Jose Tadeu de Araujo |
| EPI_ISL_1121309 | UBS Jose Francisco Rezende | Instituto Adolfo Lutz, Interdiciplinary Procedures Center, Strategic Laboratory | Caio Vinicius Dias Lopes; Claudia Regina Gonçalves; Claudio Tavares Sacchi; Erica Valessa Ramos Gomes; Karoline Rodrigues Campos |
| EPI_ISL_1795266, EPI_ISL_1795267, EPI_ISL_1795268, EPI_ISL_2345500, EPI_ISL_2345501, EPI_ISL_2345502 | UBS LUIS FACHIN IPIGUA | Instituto Butantan / ESALQ-Piracicaba | Antonio Jorge Martins; Bianca Cechetto Carlos. Mendelics: Bibiana Santos; Claudia Renata dos Santos Barros; David Schlesinger; David Schlesinger. Hemocentro Ribeirão Preto: Simone Kashima; Debora Botequio Moretti; Debora Botequio Moretti. Centro de Genômica Funcional da ESALQ: Luiz Lehmann Coutinho; Dimas Tadeu Covas; Elaine Cristina Marqueze; Elaine Vieira Santos; Elaine Vieira dos Santos; Elisangela Chicaroni Mattos; Erika Freitas; Evandra Strazza Rodrigues; Felipe Allan da Silva da Costa; Flavia Aburjaile; Guilherme Targino Valente; Heidge Fukumasu; Heidge Fukumasu. USP-Botucatu: Rejane Maria Tommasini Grotto; Instituto Butantan: Alexander Roberto Precioso; Jayme A. Souza-Neto; Jayme Augusto de Souza-Neto; Jessika Cristina Chagas Lesbon; José Salvatore Leister Patané; João Paulo Kitajima; Luiz Alcantara; Luiz Carlos Junior de Alcantara; Luiz Lehmann Coutinho; Maria Carolina Elias; Marta Giovanetti; Maurício Lacerda Nogueira; Patricia Akemi Assato; Rafael dos Santos Bezerra; Raquel de Lello Rocha Campos Cassano. NGS Soluções Genômicas: Pilar Drummond Sampaio Corrêa Mariani. FZEA-USP Pirassununga: Mirele Daiana Poleti; Raul Machado Neto; Rejane Maria Tommasini Grotto; Ricardo Augusto Brassaloti; Ricardo Haddad; Rodrigo Tocantins Calado.; Sandra Coccuzzo Sampaio; Sandra Coccuzzo Sampaio Vessoni; Simone Kashima; Svetoslav Nanev Slavov; Vagner Fonseca; Vincent Louis Viala |
| EPI_ISL_1795334, EPI_ISL_2345596 | UBS MARCIA CRISTIANE DA SILVA DE OURO VERDE | Instituto Butantan / ESALQ-Piracicaba | Antonio Jorge Martins; Bianca Cechetto Carlos. Mendelics: Bibiana Santos; Claudia Renata dos Santos Barros; David Schlesinger; David Schlesinger. Hemocentro Ribeirão Preto: Simone Kashima; Debora Botequio Moretti. Centro de Genômica Funcional da ESALQ: Luiz Lehmann Coutinho; Dimas Tadeu Covas; Elaine Cristina Marqueze; Elaine Vieira Santos; Elaine Vieira dos Santos; Elisangela Chicaroni Mattos; Erika Freitas; Evandra Strazza Rodrigues; Felipe Allan da Silva da Costa; Flavia Aburjaile; Guilherme Targino Valente; Heidge Fukumasu. USP-Botucatu: Rejane Maria Tommasini Grotto; Instituto Butantan: Alexander Roberto Precioso; Jayme A. Souza-Neto; Jayme Augusto de Souza-Neto; Jessika Cristina Chagas Lesbon; José Salvatore Leister Patané; João Paulo Kitajima; Luiz Alcantara; Luiz Carlos Junior de Alcantara; Luiz Lehmann Coutinho; Maria Carolina Elias; Marta Giovanetti; Maurício Lacerda Nogueira; Patricia Akemi Assato; Rafael dos Santos Bezerra; Raquel de Lello Rocha Campos Cassano. NGS Soluções Genômicas: Pilar Drummond Sampaio Corrêa Mariani. FZEA-USP Pirassununga: Mirele Daiana Poleti; Raul Machado Neto; Rejane Maria Tommasini Grotto; Ricardo Augusto Brassaloti; Ricardo Haddad; Rodrigo Tocantins Calado.; Sandra Coccuzzo Sampaio; Sandra Coccuzzo Sampaio Vessoni; Simone Kashima; Svetoslav Nanev Slavov; Vagner Fonseca; Vincent Louis Viala |
| EPI_ISL_3912290 | UBS MARIA FRANCINA DE ARAUJO | Analytical Competence Molecular Epidemiology Lab/ACME, Oswaldo Cruz Foundation, Ceara (FIOCRUZ CE) | Cleber Furtado Aksenen; Fabio Miyajima; Fernando Braga Stehling; Francisco Eder de Moura Lopes; Jamille Maria Mendes Bezerra; Joaquim Cesar do Nascimento Sousa Junior; Pedro Miguel Carneiro Jeronimo; Suzana Porto Almeida & Lucas Delerino on behalf of COVID-19 FIOCRUZ Genomic Network; Thais Ferreira de Oliveira; Thais de Oliveira Costa; Ticiane Cavalcante de Souza; Veridiana Pessoa Miyajima |
| EPI_ISL_2345983, EPI_ISL_2346048 | UBS MARIA PIA DE OLIVEIRA | Instituto Butantan | Antonio Jorge Martins; Claudia Renata dos Santos Barros; David Schlesinger; Debora Botequio Moretti; Dimas Tadeu Covas; Elaine Cristina Marqueze; Elaine Vieira Santos; Evandra Strazza Rodrigues; Heidge Fukumasu; Jayme Augusto de Souza-Neto; José Salvatore Leister Patané; Luiz Alcantara; Luiz Lehmann Coutinho; Maria Carolina Elias; Maurício Lacerda Nogueira; Rafael dos Santos Bezerra; Raul Machado Neto; Rejane Maria Tommasini Grotto; Ricardo Haddad; Sandra Coccuzzo Sampaio Vessoni; Simone Kashima; Svetoslav Nanev Slavov; Vincent Louis Viala |
| EPI_ISL_2345977 | UBS MARINOPOLIS | Instituto Butantan | Antonio Jorge Martins; Claudia Renata dos Santos Barros; David Schlesinger; Debora Botequio Moretti; Dimas Tadeu Covas; Elaine Cristina Marqueze; Elaine Vieira Santos; Evandra Strazza Rodrigues; Heidge Fukumasu; Jayme Augusto de Souza-Neto; José Salvatore Leister Patané; Luiz Alcantara; Luiz Lehmann Coutinho; Maria Carolina Elias; Maurício Lacerda Nogueira; Rafael dos Santos Bezerra; Raul Machado Neto; Rejane Maria Tommasini Grotto; Ricardo Haddad; Sandra Coccuzzo Sampaio Vessoni; Simone Kashima; Svetoslav Nanev Slavov; Vincent Louis Viala |
| EPI_ISL_2346028 | UBS MORRO BRANCO | Instituto Butantan | Antonio Jorge Martins; Claudia Renata dos Santos Barros; David Schlesinger; Debora Botequio Moretti; Dimas Tadeu Covas; Elaine Cristina Marqueze; Elaine Vieira Santos; Evandra Strazza Rodrigues; Heidge Fukumasu; Jayme Augusto de Souza-Neto; José Salvatore Leister Patané; Luiz Alcantara; Luiz Lehmann Coutinho; Maria Carolina Elias; Maurício Lacerda Nogueira; Rafael dos Santos Bezerra; Raul Machado Neto; Rejane Maria Tommasini Grotto; Ricardo Haddad; Sandra Coccuzzo Sampaio Vessoni; Simone Kashima; Svetoslav Nanev Slavov; Vincent Louis Viala |
| EPI_ISL_1445209 | UBS OTACILIO FIRMINO LOPES | Instituto Butantan / Mendelics | Antonio Jorge Martins; Bibiana Santos; Claudia Renata dos Santos Barros; David Schlesinger; Debora Botequio Moretti; Dimas Tadeu Covas; Elaine Cristina Marqueze; Elaine Vieira dos Santos; Erika Freitas; Evandra Strazza Rodrigues; Flavia Aburjaile; José Salvatore Leister Patané; João Paulo Kitajima; Luiz Carlos Junior de Alcantara; Maria Carolina Elias; Marta Giovanetti; Rafael dos Santos Bezerra; Raul Machado Neto; Ricardo Haddad; Rodrigo Tocantins Calado.; Sandra Coccuzzo Sampaio; Simone Kashima; Svetoslav Nanev Slavov; Vagner Fonseca; Vincent Louis Viala |
| EPI_ISL_1966731, EPI_ISL_1966733, EPI_ISL_1966734 | UBS RAIMUNDA CAVALCANTE DE SOUZA | Instituto Butantan | Antonio Jorge Martins; Bianca Cechetto Carlos. Mendelics: Bibiana Santos; Claudia Renata dos Santos Barros; Cíntia Bittar; David Schlesinger. Hemocentro Ribeirão Preto: Simone Kashima; Debora Botequio Moretti; Elaine Cristina Marqueze; Elaine Vieira dos Santos; Elisangela Chicaroni Mattos; Erika Freitas; Evandra Strazza Rodrigues; Felipe Allan da Silva da Costa; Flavia Aburjaile; Fábio Sossai Possebon; Guilherme Campos; Guilherme Targino Valente; Heidge Fukumasu. USP-Botucatu: Rejane Maria Tommasini Grotto; Helena Lage Ferreira; Instituto Butantan: Dimas Tadeu Covas; Jardelina de Souza Todao Bernardino; Jayme A. Souza-Neto; Jessika Cristina Chagas Lesbon; Jorge A. Petroli Marchesi; José Salvatore Leister Patané; João Paulo Kitajima; João Pessoa Araújo Jr.; Leila Sabrina Ullmann; Loyze Paola Oliveira de Lima; Luiz Aurelio de Campos Crispin. Centro de Genômica Funcional da ESALQ: Luiz Lehmann Coutinho; Luiz Carlos Junior de Alcantara; Lívia Sacchetto; Maisa C. Pereira Parra; Maria Carolina Elias; Marta Giovanetti; Marília Moraes; Maurício Lacerda Nogueira. Prefeitura de Sao Paulo: Melissa Palmieri.; Patricia Akemi Assato; Paula Rahal; Paulo Inacio da Costa; Rafael dos Santos Bezerra; Raquel de Lello Rocha Campos Cassano. NGS Soluções Genômicas: Pilar Drummond Sampaio Corrêa Mariani. FZEA-USP Pirassununga: Mirele Daiana Poleti; Raul Machado Neto; Ricardo Augusto Brassaloti; Ricardo Haddad; Rodrigo Tocantins Calado. FAMERP-SJRP: Cecília Artico Banho; Sandra Coccuzzo Sampaio; Svetoslav Nanev Slavov; Vagner Fonseca; Vincent Louis Viala |
| EPI_ISL_2362255 | UBS RAIMUNDA CAVALCANTE DE SOUZA | Instituto Butantan / Mendelics | Antonio Jorge Martins; Claudia Renata dos Santos Barros; David Schlesinger; Debora Botequio Moretti; Dimas Tadeu Covas; Elaine Cristina Marqueze; Elaine Vieira Santos; Evandra Strazza Rodrigues; Heidge Fukumasu; Jayme Augusto de Souza-Neto; José Salvatore Leister Patané; Luiz Alcantara; Luiz Lehmann Coutinho; Maria Carolina Elias; Maurício Lacerda Nogueira; Rafael dos Santos Bezerra; Raul Machado Neto; Rejane Maria Tommasini Grotto; Ricardo Haddad; Sandra Coccuzzo Sampaio Vessoni; Simone Kashima; Svetoslav Nanev Slavov; Vincent Louis Viala |
| EPI_ISL_2346015 | UBS RECANTO MONICA | Instituto Butantan | Antonio Jorge Martins; Claudia Renata dos Santos Barros; David Schlesinger; Debora Botequio Moretti; Dimas Tadeu Covas; Elaine Cristina Marqueze; Elaine Vieira Santos; Evandra Strazza Rodrigues; Heidge Fukumasu; Jayme Augusto de Souza-Neto; José Salvatore Leister Patané; Luiz Alcantara; Luiz Lehmann Coutinho; Maria Carolina Elias; Maurício Lacerda Nogueira; Rafael dos Santos Bezerra; Raul Machado Neto; Rejane Maria Tommasini Grotto; Ricardo Haddad; Sandra Coccuzzo Sampaio Vessoni; Simone Kashima; Svetoslav Nanev Slavov; Vincent Louis Viala |
| EPI_ISL_1966535 | UBS REGIAO OESTE DE OURINHOS | Instituto Butantan / FZEA-USP (Pirassununga) | Antonio Jorge Martins; Bianca Cechetto Carlos. Mendelics: Bibiana Santos; Claudia Renata dos Santos Barros; Cíntia Bittar; David Schlesinger. Hemocentro Ribeirão Preto: Simone Kashima; Debora Botequio Moretti; Elaine Cristina Marqueze; Elaine Vieira dos Santos; Elisangela Chicaroni Mattos; Erika Freitas; Evandra Strazza Rodrigues; Felipe Allan da Silva da Costa; Flavia Aburjaile; Fábio Sossai Possebon; Guilherme Campos; Guilherme Targino Valente; Heidge Fukumasu. USP-Botucatu: Rejane Maria Tommasini Grotto; Helena Lage Ferreira; Instituto Butantan: Dimas Tadeu Covas; Jardelina de Souza Todao Bernardino; Jayme A. Souza-Neto; Jessika Cristina Chagas Lesbon; Jorge A. Petroli Marchesi; José Salvatore Leister Patané; João Paulo Kitajima; João Pessoa Araújo Jr.; Leila Sabrina Ullmann; Loyze Paola Oliveira de Lima; Luiz Aurelio de Campos Crispin. Centro de Genômica Funcional da ESALQ: Luiz Lehmann Coutinho; Luiz Carlos Junior de Alcantara; Lívia Sacchetto; Maisa C. Pereira Parra; Maria Carolina Elias; Marta Giovanetti; Marília Moraes; Maurício Lacerda Nogueira. Prefeitura de Sao Paulo: Melissa Palmieri.; Patricia Akemi Assato; Paula Rahal; Paulo Inacio da Costa; Rafael dos Santos Bezerra; Raquel de Lello Rocha Campos Cassano. NGS Soluções Genômicas: Pilar Drummond Sampaio Corrêa Mariani. FZEA-USP Pirassununga: Mirele Daiana Poleti; Raul Machado Neto; Ricardo Augusto Brassaloti; Ricardo Haddad; Rodrigo Tocantins Calado. FAMERP-SJRP: Cecília Artico Banho; Sandra Coccuzzo Sampaio; Svetoslav Nanev Slavov; Vagner Fonseca; Vincent Louis Viala |
| EPI_ISL_1966188 | UBS SALTO DE SAO JOSE | Instituto Butantan / Mendelics | Antonio Jorge Martins; Bianca Cechetto Carlos. Mendelics: Bibiana Santos; Claudia Renata dos Santos Barros; Cíntia Bittar; David Schlesinger. Hemocentro Ribeirão Preto: Simone Kashima; Debora Botequio Moretti; Elaine Cristina Marqueze; Elaine Vieira dos Santos; Elisangela Chicaroni Mattos; Erika Freitas; Evandra Strazza Rodrigues; Felipe Allan da Silva da Costa; Flavia Aburjaile; Fábio Sossai Possebon; Guilherme Campos; Guilherme Targino Valente; Heidge Fukumasu. USP-Botucatu: Rejane Maria Tommasini Grotto; Helena Lage Ferreira; Instituto Butantan: Dimas Tadeu Covas; Jardelina de Souza Todao Bernardino; Jayme A. Souza-Neto; Jessika Cristina Chagas Lesbon; Jorge A. Petroli Marchesi; José Salvatore Leister Patané; João Paulo Kitajima; João Pessoa Araújo Jr.; Leila Sabrina Ullmann; Loyze Paola Oliveira de Lima; Luiz Aurelio de Campos Crispin. Centro de Genômica Funcional da ESALQ: Luiz Lehmann Coutinho; Luiz Carlos Junior de Alcantara; Lívia Sacchetto; Maisa C. Pereira Parra; Maria Carolina Elias; Marta Giovanetti; Marília Moraes; Maurício Lacerda Nogueira. Prefeitura de Sao Paulo: Melissa Palmieri.; Patricia Akemi Assato; Paula Rahal; Paulo Inacio da Costa; Rafael dos Santos Bezerra; Raquel de Lello Rocha Campos Cassano. NGS Soluções Genômicas: Pilar Drummond Sampaio Corrêa Mariani. FZEA-USP Pirassununga: Mirele Daiana Poleti; Raul Machado Neto; Ricardo Augusto Brassaloti; Ricardo Haddad; Rodrigo Tocantins Calado. FAMERP-SJRP: Cecília Artico Banho; Sandra Coccuzzo Sampaio; Svetoslav Nanev Slavov; Vagner Fonseca; Vincent Louis Viala |
| EPI_ISL_2345967, EPI_ISL_2345969, EPI_ISL_2345971 | UBS SANTA MARIA | Instituto Butantan | Antonio Jorge Martins; Claudia Renata dos Santos Barros; David Schlesinger; Debora Botequio Moretti; Dimas Tadeu Covas; Elaine Cristina Marqueze; Elaine Vieira Santos; Evandra Strazza Rodrigues; Heidge Fukumasu; Jayme Augusto de Souza-Neto; José Salvatore Leister Patané; Luiz Alcantara; Luiz Lehmann Coutinho; Maria Carolina Elias; Maurício Lacerda Nogueira; Rafael dos Santos Bezerra; Raul Machado Neto; Rejane Maria Tommasini Grotto; Ricardo Haddad; Sandra Coccuzzo Sampaio Vessoni; Simone Kashima; Svetoslav Nanev Slavov; Vincent Louis Viala |
| EPI_ISL_1966847, EPI_ISL_1966848, EPI_ISL_1966849, EPI_ISL_1966850, EPI_ISL_1966851, EPI_ISL_1966852 | UBS SAO JUDAS | Instituto Butantan / ESALQ-USP (Piracicaba) | Antonio Jorge Martins; Bianca Cechetto Carlos. Mendelics: Bibiana Santos; Claudia Renata dos Santos Barros; Cíntia Bittar; David Schlesinger. Hemocentro Ribeirão Preto: Simone Kashima; Debora Botequio Moretti; Elaine Cristina Marqueze; Elaine Vieira dos Santos; Elisangela Chicaroni Mattos; Erika Freitas; Evandra Strazza Rodrigues; Felipe Allan da Silva da Costa; Flavia Aburjaile; Fábio Sossai Possebon; Guilherme Campos; Guilherme Targino Valente; Heidge Fukumasu. USP-Botucatu: Rejane Maria Tommasini Grotto; Helena Lage Ferreira; Instituto Butantan: Dimas Tadeu Covas; Jardelina de Souza Todao Bernardino; Jayme A. Souza-Neto; Jessika Cristina Chagas Lesbon; Jorge A. Petroli Marchesi; José Salvatore Leister Patané; João Paulo Kitajima; João Pessoa Araújo Jr.; Leila Sabrina Ullmann; Loyze Paola Oliveira de Lima; Luiz Aurelio de Campos Crispin. Centro de Genômica Funcional da ESALQ: Luiz Lehmann Coutinho; Luiz Carlos Junior de Alcantara; Lívia Sacchetto; Maisa C. Pereira Parra; Maria Carolina Elias; Marta Giovanetti; Marília Moraes; Maurício Lacerda Nogueira. Prefeitura de Sao Paulo: Melissa Palmieri.; Patricia Akemi Assato; Paula Rahal; Paulo Inacio da Costa; Rafael dos Santos Bezerra; Raquel de Lello Rocha Campos Cassano. NGS Soluções Genômicas: Pilar Drummond Sampaio Corrêa Mariani. FZEA-USP Pirassununga: Mirele Daiana Poleti; Raul Machado Neto; Ricardo Augusto Brassaloti; Ricardo Haddad; Rodrigo Tocantins Calado. FAMERP-SJRP: Cecília Artico Banho; Sandra Coccuzzo Sampaio; Svetoslav Nanev Slavov; Vagner Fonseca; Vincent Louis Viala |
| EPI_ISL_2345897, EPI_ISL_2345901, EPI_ISL_2345903 | UBS SAO LOURENCO DA SERRA | Instituto Butantan / FZEA-USP-Pirassununga | Antonio Jorge Martins; Claudia Renata dos Santos Barros; David Schlesinger; Debora Botequio Moretti; Dimas Tadeu Covas; Elaine Cristina Marqueze; Elaine Vieira Santos; Evandra Strazza Rodrigues; Heidge Fukumasu; Jayme Augusto de Souza-Neto; José Salvatore Leister Patané; Luiz Alcantara; Luiz Lehmann Coutinho; Maria Carolina Elias; Maurício Lacerda Nogueira; Rafael dos Santos Bezerra; Raul Machado Neto; Rejane Maria Tommasini Grotto; Ricardo Haddad; Sandra Coccuzzo Sampaio Vessoni; Simone Kashima; Svetoslav Nanev Slavov; Vincent Louis Viala |
| EPI_ISL_2614592, EPI_ISL_2841605 | UBS Silvio Joao Luiz de Lucia | Instituto Adolfo Lutz, Interdiciplinary Procedures Center, Strategic Laboratory | Caio Vinicius Dias Lopes; Claudia Regina Gonçalves; Claudio Tavares Sacchi; Erica Valessa Ramos Gomes; Karoline Rodrigues Campos; Leonardo Jose Tadeu de Araujo |
| EPI_ISL_1795270, EPI_ISL_1795272, EPI_ISL_1795273, EPI_ISL_2345504, EPI_ISL_2345507, EPI_ISL_2345509 | UBS TEREZA GALLO UCHOA | Instituto Butantan / ESALQ-Piracicaba | Antonio Jorge Martins; Bianca Cechetto Carlos. Mendelics: Bibiana Santos; Claudia Renata dos Santos Barros; David Schlesinger; David Schlesinger. Hemocentro Ribeirão Preto: Simone Kashima; Debora Botequio Moretti; Debora Botequio Moretti. Centro de Genômica Funcional da ESALQ: Luiz Lehmann Coutinho; Dimas Tadeu Covas; Elaine Cristina Marqueze; Elaine Vieira Santos; Elaine Vieira dos Santos; Elisangela Chicaroni Mattos; Erika Freitas; Evandra Strazza Rodrigues; Felipe Allan da Silva da Costa; Flavia Aburjaile; Guilherme Targino Valente; Heidge Fukumasu; Heidge Fukumasu. USP-Botucatu: Rejane Maria Tommasini Grotto; Instituto Butantan: Alexander Roberto Precioso; Jayme A. Souza-Neto; Jayme Augusto de Souza-Neto; Jessika Cristina Chagas Lesbon; José Salvatore Leister Patané; João Paulo Kitajima; Luiz Alcantara; Luiz Carlos Junior de Alcantara; Luiz Lehmann Coutinho; Maria Carolina Elias; Marta Giovanetti; Maurício Lacerda Nogueira; Patricia Akemi Assato; Rafael dos Santos Bezerra; Raquel de Lello Rocha Campos Cassano. NGS Soluções Genômicas: Pilar Drummond Sampaio Corrêa Mariani. FZEA-USP Pirassununga: Mirele Daiana Poleti; Raul Machado Neto; Rejane Maria Tommasini Grotto; Ricardo Augusto Brassaloti; Ricardo Haddad; Rodrigo Tocantins Calado.; Sandra Coccuzzo Sampaio; Sandra Coccuzzo Sampaio Vessoni; Simone Kashima; Svetoslav Nanev Slavov; Vagner Fonseca; Vincent Louis Viala |
| EPI_ISL_1509639, EPI_ISL_1509720 | UBS de Auriflama | Instituto Adolfo Lutz, Interdiciplinary Procedures Center, Strategic Laboratory | Caio Vinicius Dias Lopes; Claudia Regina Gonçalves; Claudio Tavares Sacchi; Erica Valessa Ramos Gomes; Karoline Rodrigues Campos; Leonardo Jose Tadeu de Araujo |
| EPI_ISL_3419561, EPI_ISL_3419562, EPI_ISL_3419575 | UC Davis Genome Center and Healthy Davis Together | UC Davis Genome Center | UC Davis |
| EPI_ISL_3321293 | UC-Christus Clinical Hospital Laboratory | Laboratory of Molecular Virology, School of Medicine, Pontificia Universidad Catolica de Chile | Ana Maira Guzman; Andres E. Munoz-Marcos; Catalina Pardo-Roa; Eileen Serrano; Erick Salinas; Estefany Poblete; Francisco Melo; Jorge Levican; Leonardo I. Almonacid; Maria Jose Avendano; Maria Patricia Vega; Rafael A. Medina; Ricardo Enrique de la Barra; Tamara Garcia-Salum |
| EPI_ISL_2758715, EPI_ISL_2758723, EPI_ISL_2758728, EPI_ISL_2758732, EPI_ISL_2758738, EPI_ISL_2758741 | UEL | IPEC Guarapuava | NAPI-Genômica (Novos Arranjo de Pesquisa e Inovação em Genômica): Ademar Dantas da Cunha Júnior Adriano Ferrasa Adriano Mondini Aldo Przybysz Alessandra Lourenço Cecchini Armani Alex Sandro Jorge Alexandra Ivo de Medeiros Alexandre Maller Aline Cristina Batista Rodrigues Johann Ana Lucia Ferreira Ana Marisa Fusco Almeida Anderson Joel Martino Andrade André Luís Laforga Vanzela Andrea Duarte Doetzer Andrea Name Colado Simao Andressa Pereira de Souza Anelisa Ramão Angelica Beate Winter Boldt Anna Hermínia Castro Gomes de Amorim Anna Silvia Penteado Setti da Rocha Antonio Camilo da Silva Filho Antonio Stabelini Neto Arthur Hirata Bertachi Barbara Mendes Paz Chao Betty Cristiane Kuhn Bruno Ambrozio Galindo Bruno Ribeiro Cruz Camilla Reginatto De Pierri Carla Fredrichsen Moya Araujo Carla Fredrichsen Moya Araujo Carlos Alberto Oliveira de Biagi Junior Carlos Augusto Nassar Carlos Eduardo Buss Carlos Gilberto Carlotti Junior Carlos Henrique Schneider Carolina Panis Carolina Weigert Galvão Caroline de Jesus Coelho Donha Caroline Guisantes de Salvo Toni Caryna Eurich Mazur Catiuscie Cabreira da Silva Tortorella Celso F. D. Doliveira Cesar Luiz Boguszewski Christiane Pienna Soares Chung Man Chin Claudia Moro Cleverson Busso Cristiane Cominetti Daiane Priscila Simão-Silva Dalila Luciola Zanette Daniel de Paula Daniel de Paula Daniel Rech Daniela Fiori Gradia Daniela Pretti da Cunha Tirapelli Daniela Viganó Zanoti Jeronymo Daniele Ukan Danielle Malheiros Ferreira Danielle Venturini Deborah Catharine de Assis Leite Deivid Calebe de Souza Dennis Armando Bertolini Edenir Inez Pamero Edna Maria Vissoci Reiche Edson Roberto Arpini Miguel Eduardo José de Almeida Araújo Eliana Carolina Vespero Eliandro Reis Tavares Elza Kimura Grimshaw Emanuel Maltempi de Souza Emanuele Cristina Gustani Buss Émerson Carraro Emiliana Cristina Melo ENILze Maria de Souza Fonseca Ribeiro Enilze Maria de Souza Fonseca Ribeiro Erika Izumi Erika Seki |

| | | | |
|---|---|---|---|
| | | | Kioshima Cotica Evani Marques Pereira Fabio Negretti Fábio Rodrigues Ferreira Seiva Felipe Dunin dos Santos Felipe Tuon Fernanda Andreia Rosa Fernanda Cestaro Prado Cortez Fernanda Ivanski Fernanda Maris Peria Flavia Regina Oliveira de Barros Franciele Aní Caovilla Follador Franciele Mara Lucca Zanardo Bohm Francinete Ramos Campos Fulviana Silva Nishiyama GABRIEL RIBEIRO CORDEIRO Gabriela Datsch Bennemann Gisele Santos de Oliveira Glaucio Valdameri Glauco Akelinghton Freire Vitiello Glauco Vieira Miranda Glaura Scantamburlo ALves Fernandes Guilherme Ferreira Silveira Gustavo Bianchini Porfírio Gustavo Lenci Marques Hélito Volpato Hildebrando Masshiro Nagai Huei Diana Lee Ilce Mara de Syllos Cólus Iris Rabinovich Israel Gomy Jackson Kawakami Jacques Duílio Brancher Jaime Luís Lopes Rocha Jaqueline Carvalho de Oliveira Jean Henrique da Silva Rodrigues Jean Leandro dos Santos Jeane Eliete Laguila Visentainer João Paulo Bianchi Ximenez Joaquim Manoel da Silva Jociani Ascari Joel Donazzolo Jorge Luis Maria Ruiz Jose Knopfholz José Luis da Conceição Silva José Sebastião dos Santos Joseane Carla Schabarum Juliana Cheleski Wiggers Juliana Mara Serpeloni Juliana Morini Küpper Cardoso Perseguini Karen Brajão de Oliveira Karin Braun Prado Karine Aparecida de Lima Katiany Rizzieri Caleffi Ferracioli Katiuscia de Oliveira Francisco Gabriel Kelvinson Fernandes Viana Larissa Beatriz Cossalter Larissa Danielle Bahls Pinto Laurival Antonio Vilas Boas Léia Carolina Lucio Líbero Mezzadri Neto Ligia Carla Faccin Galhardi Lirane Elize Defante Ferreto Luciana Furlaneto Maia Luciana Oliveira de Fariña Luciana Reis Azevedo Alanis Luciane Regina Cavalli Lucy Megumi Yamauchi Lioni Luis Paulo Gomes Mascarenhas Luis Paulo Gomes Mascarenhas Luís Paulo Mascarenhas Lupe Furtado Alle Lyvia Regina Biagi Silva Bertachi Mara Antonia Ramos Costa Mara L. Cordeiro Marcela Maria Birolim Marcelo Ricardo Vicari Marcia Edilaine Lopes Consolaro Marcia Holsbach Beltrame Marcia Regina Eches Perugini Marcos Abdo Arbex Marcos Pileggi MARCOS TADEU GRZELCZAK Marcus Peikriszwili Tartaruga Maria Angelica Ehara Watanabe Maria Antonia Ramos Costa Maria Claudia Gross Maria José Soares Mendes Giannini Maria Leandra Terencio Maria Lúcia Bonfleur Maria Luiza Guimarães de Oliveira Maria Luiza Petzl-Erler Mariana Abe Vicente Cavagnari Marina Kimiko Kadowaki Marise Fonseca dos Santos Marla Karine Amarante Maurício Turkiewicz Mauro Antonio Alves Castro Michel Rodrigo Zambrano Passarini Michele Potrich Michelle Orane Schemberger Milena Massumi Kozonoe Mônica Degraf Cavallin Monica Tereza Suldofski Mucio Luiz de Assis Cirino Nadia Graciele Krohn Najeh Maissar Khalil Nédia de Castilhos Ghisi Neide Tomimura Costa Neiva Leite Neyva Maria Lopes Romeiro Patrícia Amâncio da Rosa Patricia Dayane Carvalho Schaker Patricia Oehlmeyer Nassar Patrícia Savio de Araújo-Souza Patricia Silva Lucio Paulo Henrique Couto Souza Paulo Roberto Donadio Percy Nohama Quirino Alves de Lima Neto Rafael Deminice Rafael dos Santos Bezerra Raquel Alves dos Santos Renan Manozzo Galante Renata Ernlund Freitas de Macedo Rita de Cássia Garcia Simão Roberta Losi Guembarovski Roberto H. Herai Roberto Rosati Rodrigo Ferreira Rodrigo Rodrigues Matiello Rogério Neri Shinsato Rogério Pincela Mateus Rosane Aparecida Ribeiro Rosilene Fressatti Cardoso Rosilene Fressatti Cardoso Sandra Mara Guse Scós Venske Selene Elifio Esposito Sérgio Ossamu Ioshii Silvana Giuliatti Silvia Mara de Souza Halick Silvio Henrique Maia de Almeida Simone Neumann Wendt Spencer Luiz Marques Payão Stefan Wolanski Negrão Stephane Janaina de Moura Escobar Sueli Fumie Yamada Ogatta SUELI PERCIO QUINAIA Taciane Finatto Tatiana Mayumi Veiga Iriyoda Tayza Katelline Danilau Ostroski Tony Alexander Hild Valeria Valente Vanessa Nascimento Kozak Vanessa Santos Sotomaior Victor Breno Pedrosa Victoria Zeghbi Cochenski Borba Vivian Rotuno Moure Valdameri Wander Rogerio Pavanelli Weber Cláudio Francisco Nunes da Silva Willian Augusto de Melo Yohandra Reyes Torres |
| EPI_ISL_3369439, EPI_ISL_3369440, EPI_ISL_3369441 | UFPA- Federal University of Pará | ITV-Vale Institute of Technology | Amanda Vidal; Guilherme Oliveira; Tatianne Costa Negri; Ândrea Kely Campos Ribeiro dos Santos |
| EPI_ISL_2448437 | ULSS 2 Marca Trevigiana | Istituto Zooprofilattico Sperimentale delle Venezie | Adelaide Milani; Alessia Schivo; Alice Fusaro; Ambra Pastori; Annalisa Salviato; Antonia Ricci; Calogero Terregino; Edoardo Giussani; Elisa Palumbo; Erika Giorgia Quaranta; Isabella Monne; Luca Tassoni |
| EPI_ISL_3409408, EPI_ISL_3410073, EPI_ISL_3410074, EPI_ISL_3410075, EPI_ISL_3410076, EPI_ISL_3410079, EPI_ISL_3410083 | | | |
| see above | UMass Memorial Medical Center | Center for Microbiome Research | Doyle V. Ward; Karl J. Simon; Richard T. Ellison III |
| EPI_ISL_1373457, EPI_ISL_1854880, EPI_ISL_1854884 | UNC-CH COVID Surveillance Lab | Jeremy Wang | Alexander Rubinsteyn; Amir Barzin; Amy James Loftis; Corbin Jones; Jeremy Wang; Melissa Miller; Susan Fiscus |
| EPI_ISL_3912291 | UNID MISTA DE MORAUJO | Analytical Competence Molecular Epidemiology Lab/ACME, Oswaldo Cruz Foundation, Ceara (FIOCRUZ CE) | Cleber Furtado Aksenen; Fabio Miyajima; Fernando Braga Stehling; Francisco Eder de Moura Lopes; Jamille Maria Mendes Bezerra; Joaquim Cesar do Nascimento Sousa Junior; Pedro Miguel Carneiro Jeronimo; Suzana Porto Almeida & Lucas Delerino on behalf of COVID-19 FIOCRUZ Genomic Network; Thais Ferreira de Oliveira; Thais de Oliveira Costa; Ticiane Cavalcante de Souza; Veridiana Pessoa Miyajima |
| EPI_ISL_2344562 | UNIDADE BASICA DE SAUDE ALBERTO CARLOS BOINTONI | Instituto Butantan / FZEA-USP-Pirassununga | Antonio Jorge Martins; Claudia Renata dos Santos Barros; David Schlesinger; Debora Botequio Moretti; Dimas Tadeu Covas; Elaine Cristina Marqueze; Elaine Vieira Santos; Evandra Strazza Rodrigues; Heidge Fukumasu; Jayme Augusto de Souza-Neto; José Salvatore Leister Patané; Luiz Alcantara; Luiz Lehmann Coutinho; Maria Carolina Elias; Maurício Lacerda Nogueira; Rafael dos Santos Bezerra; Raul Machado Neto; Rejane Maria Tommasini Grotto; Ricardo Haddad; Sandra Coccuzzo Sampaio Vessoni; Simone Kashima; Svetoslav Nanev Slavov; Vincent Louis Viala |
| EPI_ISL_1966201 | UNIDADE BASICA DE SAUDE DA FAMILIA BILAC | Instituto Butantan / Mendelics | Antonio Jorge Martins; Bianca Cechetto Carlos. Mendelics: Bibiana Santos; Claudia Renata dos Santos Barros; Cíntia Bittar; David Schlesinger. Hemocentro Ribeirão Preto: Simone Kashima; Debora Botequio Moretti; Elaine Cristina Marqueze; Elaine Vieira dos Santos; Elisangela Chicaroni Mattos; Erika Freitas; Evandra Strazza Rodrigues; Felipe Allan da Silva da Costa; Flavia Aburjaile; Fábio Sossai Possebon; Guilherme Campos; Guilherme Targino Valente; Heidge Fukumasu. USP-Botucatu: Rejane Maria Tommasini Grotto; Helena Lage Ferreira; Instituto Butantan: Dimas Tadeu Covas; Jardelina de Souza Todao Bernardino; Jayme A. Souza-Neto; Jessika Cristina Chagas Lesbon; Jorge A. Petroli Marchesi; José Salvatore Leister Patané; João Paulo Kitajima; João Pessoa Araújo Jr.; Leila Sabrina Ullmann; Loyze Paola Oliveira de Lima; Luiz Aurelio de Campos Crispin. Centro de Genômica Funcional da ESALQ: Luiz Lehmann Coutinho; Luiz Carlos Junior de Alcantara; Lívia Sacchetto; Maisa C. Pereira Parra; Maria Carolina Elias; Marta Giovanetti; Marília Moraes; Maurício Lacerda Nogueira. Prefeitura de Sao Paulo: Melissa Palmieri.; Patricia Akemi Assato; Paula Rahal; Paulo Inacio da Costa; Rafael dos Santos Bezerra; Raquel de Lello Rocha Campos Cassano. NGS Soluções Genômicas: Pilar Drummond Sampaio Corrêa Mariani. FZEA-USP Pirassununga: Mirele Daiana Poleti; Raul Machado Neto; Ricardo Augusto Brassaloti; Ricardo Haddad; Rodrigo Tocantins Calado. FAMERP-SJRP: Cecília Artico Banho; Sandra Coccuzzo Sampaio; Svetoslav Nanev Slavov; Vagner Fonseca; Vincent Louis Viala |
| EPI_ISL_1795362, EPI_ISL_1795363, EPI_ISL_2345637, EPI_ISL_2345638 | UNIDADE BASICA DE SAUDE DE PIQUETE | Instituto Butantan / ESALQ-Piracicaba | Antonio Jorge Martins; Bianca Cechetto Carlos. Mendelics: Bibiana Santos; Claudia Renata dos Santos Barros; David Schlesinger; David Schlesinger. Hemocentro Ribeirão Preto: Simone Kashima; Debora Botequio Moretti; Debora Botequio Moretti. Centro de Genômica Funcional da ESALQ: Luiz Lehmann Coutinho; Dimas Tadeu Covas; Elaine Cristina Marqueze; Elaine Vieira Santos; Elaine Vieira dos Santos; Elisangela Chicaroni Mattos; Erika Freitas; Evandra Strazza Rodrigues; Felipe Allan da Silva da Costa; Flavia Aburjaile; Guilherme Targino Valente; Heidge Fukumasu; Heidge Fukumasu. USP-Botucatu: Rejane Maria Tommasini Grotto; Instituto Butantan: Alexander Roberto Precioso; Jayme A. Souza-Neto; Jayme Augusto de Souza-Neto; Jessika Cristina Chagas Lesbon; José Salvatore Leister Patané; João Paulo Kitajima; Luiz Alcantara; Luiz Carlos Junior de Alcantara; Luiz Lehmann Coutinho; Maria Carolina Elias; Marta Giovanetti; Maurício Lacerda Nogueira; Patricia Akemi Assato; Rafael dos Santos Bezerra; Raquel de Lello Rocha Campos Cassano. NGS Soluções Genômicas: Pilar Drummond Sampaio Corrêa Mariani. FZEA-USP Pirassununga: Mirele Daiana Poleti; Raul Machado Neto; Rejane Maria Tommasini Grotto; Ricardo Augusto Brassaloti; Ricardo Haddad; Rodrigo Tocantins Calado.; Sandra Coccuzzo Sampaio; Sandra Coccuzzo Sampaio Vessoni; Simone Kashima; Svetoslav Nanev Slavov; Vagner Fonseca; Vincent Louis Viala |
| EPI_ISL_1966515 | UNIDADE BASICA DE SAUDE DE TAQUARAL CAETANO PITELLI | Instituto Butantan / Mendelics | Antonio Jorge Martins; Bianca Cechetto Carlos. Mendelics: Bibiana Santos; Claudia Renata dos Santos Barros; Cíntia Bittar; David Schlesinger. Hemocentro Ribeirão Preto: Simone Kashima; Debora Botequio Moretti; Elaine Cristina Marqueze; Elaine Vieira dos Santos; Elisangela Chicaroni Mattos; Erika Freitas; Evandra Strazza Rodrigues; Felipe Allan da Silva da Costa; Flavia Aburjaile; Fábio Sossai Possebon; Guilherme Campos; Guilherme Targino Valente; Heidge Fukumasu. USP-Botucatu: Rejane Maria Tommasini Grotto; Helena Lage Ferreira; Instituto Butantan: Dimas Tadeu Covas; Jardelina de Souza Todao Bernardino; Jayme A. Souza-Neto; Jessika Cristina Chagas Lesbon; Jorge A. Petroli Marchesi; José Salvatore Leister Patané; João Paulo Kitajima; João Pessoa Araújo Jr.; Leila Sabrina Ullmann; Loyze Paola Oliveira de Lima; Luiz Aurelio de Campos Crispin. Centro de Genômica Funcional da ESALQ: Luiz Lehmann Coutinho; Luiz Carlos Junior de Alcantara; Lívia Sacchetto; Maisa C. Pereira Parra; Maria Carolina Elias; Marta Giovanetti; Marília Moraes; Maurício Lacerda Nogueira. Prefeitura de Sao Paulo: Melissa Palmieri.; Patricia Akemi Assato; Paula Rahal; Paulo Inacio da Costa; Rafael dos Santos Bezerra; Raquel de Lello Rocha Campos Cassano. NGS Soluções Genômicas: Pilar Drummond Sampaio Corrêa Mariani. FZEA-USP Pirassununga: Mirele Daiana Poleti; Raul Machado Neto; Ricardo Augusto Brassaloti; Ricardo Haddad; Rodrigo Tocantins Calado. FAMERP-SJRP: Cecília Artico Banho; Sandra Coccuzzo Sampaio; Svetoslav Nanev Slavov; Vagner Fonseca; Vincent Louis Viala |
| EPI_ISL_1795224, EPI_ISL_2345455 | UNIDADE BASICA DE SAUDE DR APOLONIO MORAES E SOUZA | Instituto Butantan / ESALQ-Piracicaba | Antonio Jorge Martins; Bianca Cechetto Carlos. Mendelics: Bibiana Santos; Claudia Renata dos Santos Barros; David Schlesinger; David Schlesinger. Hemocentro Ribeirão Preto: Simone Kashima; Debora Botequio Moretti; Debora Botequio Moretti. Centro de Genômica Funcional da ESALQ: Luiz Lehmann Coutinho; Dimas Tadeu Covas; Elaine Cristina Marqueze; Elaine Vieira Santos; Elaine Vieira dos Santos; Elisangela Chicaroni Mattos; Erika Freitas; Evandra Strazza Rodrigues; Felipe Allan da Silva da Costa; Flavia Aburjaile; Guilherme Targino Valente; Heidge Fukumasu; Heidge Fukumasu. USP-Botucatu: Rejane Maria Tommasini Grotto; Instituto Butantan: Alexander Roberto Precioso; Jayme A. Souza-Neto; Jayme Augusto de Souza-Neto; Jessika Cristina Chagas Lesbon; José Salvatore Leister Patané; João Paulo Kitajima; Luiz Alcantara; Luiz Carlos Junior de Alcantara; Luiz Lehmann Coutinho; Maria Carolina Elias; Marta Giovanetti; Maurício Lacerda Nogueira; Patricia Akemi Assato; Rafael dos Santos Bezerra; Raquel de Lello Rocha Campos Cassano. NGS Soluções Genômicas: Pilar Drummond Sampaio Corrêa Mariani. FZEA-USP Pirassununga: Mirele Daiana Poleti; Raul Machado Neto; Rejane Maria Tommasini Grotto; Ricardo Augusto Brassaloti; Ricardo Haddad; Rodrigo Tocantins Calado.; Sandra Coccuzzo Sampaio; Sandra Coccuzzo Sampaio Vessoni; Simone Kashima; Svetoslav Nanev Slavov; Vagner Fonseca; Vincent Louis Viala |
| EPI_ISL_3102311 | UNIDADE BASICA DE SAUDE DR FCO MARCIO FERNANDES | Analytical Competence Molecular Epidemiology Lab/ACME, Oswaldo Cruz Foundation, Ceara (FIOCRUZ CE) | Cleber Furtado Aksenen; Fabio Miyajima; Fernando Braga Stehling; Francisco Eder de Moura Lopes; Jamille Maria Mendes Bezerra; Joaquim César do Nascimento Sousa Junior; Pedro Miguel Carneiro Jeronimo; Suzana Porto Almeida e Lucas Delerino; Thais Ferreira de Oliveira; Thais de Oliveira Costa; Ticiane Cavalcante de Souza; Veridiana Pessoa Miyajima |
| EPI_ISL_1795227, EPI_ISL_2345458 | UNIDADE BASICA DE SAUDE DR LOTFALLAH MIZIARA | Instituto Butantan / ESALQ-Piracicaba | Antonio Jorge Martins; Bianca Cechetto Carlos. Mendelics: Bibiana Santos; Claudia Renata dos Santos Barros; David Schlesinger; David Schlesinger. Hemocentro Ribeirão Preto: Simone Kashima; Debora Botequio Moretti; Debora Botequio Moretti. Centro de Genômica Funcional da ESALQ: Luiz Lehmann Coutinho; Dimas Tadeu Covas; Elaine Cristina Marqueze; Elaine Vieira Santos; Elaine Vieira dos Santos; Elisangela Chicaroni Mattos; Erika Freitas; Evandra Strazza Rodrigues; Felipe Allan da Silva da Costa; Flavia Aburjaile; Guilherme Targino Valente; Heidge Fukumasu; Heidge Fukumasu. USP-Botucatu: Rejane Maria Tommasini Grotto; Instituto Butantan: Alexander Roberto Precioso; Jayme A. Souza-Neto; Jayme Augusto de Souza-Neto; Jessika Cristina Chagas Lesbon; José Salvatore Leister Patané; João Paulo Kitajima; Luiz Alcantara; Luiz Carlos Junior de Alcantara; Luiz Lehmann Coutinho; Maria Carolina Elias; Marta Giovanetti; Maurício Lacerda Nogueira; Patricia Akemi Assato; Rafael dos Santos Bezerra; Raquel de Lello Rocha Campos Cassano. NGS Soluções Genômicas: Pilar Drummond Sampaio Corrêa Mariani. FZEA-USP Pirassununga: Mirele Daiana Poleti; Raul Machado Neto; Rejane Maria Tommasini Grotto; Ricardo Augusto Brassaloti; Ricardo Haddad; Rodrigo Tocantins Calado.; Sandra Coccuzzo Sampaio; Sandra Coccuzzo Sampaio Vessoni; Simone Kashima; Svetoslav Nanev Slavov; Vagner Fonseca; Vincent Louis Viala |
| EPI_ISL_2209769 | UNIDADE BASICA DE SAUDE DR MATHEUS GABRIEL BONASSA | Instituto Butantan | Antonio Jorge Martins; Claudia Renata dos Santos Barros; David Schlesinger; Debora Botequio Moretti; Dimas Tadeu Covas; Elaine Cristina Marqueze; Elaine Vieira Santos; Evandra Strazza Rodrigues; Heidge Fukumasu; Jayme Augusto de Souza-Neto; José Salvatore Leister Patané; Luiz Alcantara; Luiz Lehmann Coutinho; Maria Carolina Elias; Maurício Lacerda Nogueira; Rafael dos Santos Bezerra; Raul Machado Neto; Rejane Maria Tommasini Grotto; Ricardo Haddad; Sandra Coccuzzo Sampaio Vessoni; Simone Kashima; Svetoslav Nanev Slavov; Vincent Louis Viala. |
| EPI_ISL_2209183 | UNIDADE BASICA DE SAUDE DR MATHEUS GABRIEL BONASSA | Instituto Butantan / FZEA-USP-Pirassununga | Antonio Jorge Martins; Claudia Renata dos Santos Barros; David Schlesinger; Debora Botequio Moretti; Dimas Tadeu Covas; Elaine Cristina Marqueze; Elaine Vieira Santos; Evandra Strazza Rodrigues; Heidge Fukumasu; Jayme Augusto de Souza-Neto; José Salvatore Leister Patané; Luiz Alcantara; Luiz Lehmann Coutinho; Maria Carolina Elias; Maurício Lacerda Nogueira; Rafael dos Santos Bezerra; Raul Machado Neto; Rejane Maria Tommasini Grotto; Ricardo Haddad; Sandra Coccuzzo Sampaio Vessoni; Simone Kashima; Svetoslav Nanev Slavov; Vincent Louis Viala |
| EPI_ISL_1966245 | UNIDADE BASICA DE SAUDE DR ORESTES DE MOURA PINTO | Instituto Butantan / Mendelics | Antonio Jorge Martins; Bianca Cechetto Carlos. Mendelics: Bibiana Santos; Claudia Renata dos Santos Barros; Cíntia Bittar; David Schlesinger. Hemocentro Ribeirão Preto: Simone Kashima; Debora Botequio Moretti; Elaine Cristina Marqueze; Elaine Vieira dos Santos; Elisangela Chicaroni Mattos; Erika Freitas; Evandra Strazza Rodrigues; Felipe Allan da Silva da Costa; Flavia Aburjaile; Fábio Sossai Possebon; Guilherme Campos; Guilherme Targino Valente; Heidge Fukumasu. USP-Botucatu: Rejane Maria Tommasini Grotto; Helena Lage Ferreira; Instituto Butantan: Dimas Tadeu Covas; Jardelina de Souza Todao Bernardino; Jayme A. Souza-Neto; Jessika Cristina Chagas Lesbon; Jorge A. Petroli Marchesi; José Salvatore Leister Patané; João Paulo Kitajima; João Pessoa Araújo Jr.; Leila Sabrina Ullmann; Loyze Paola Oliveira de Lima; Luiz Aurelio de Campos Crispin. Centro de Genômica Funcional da ESALQ: Luiz Lehmann Coutinho; Luiz Carlos Junior de Alcantara; Lívia Sacchetto; Maisa C. Pereira Parra; Maria Carolina Elias; Marta Giovanetti; Marília Moraes; Maurício Lacerda Nogueira. Prefeitura de Sao Paulo: Melissa Palmieri.; Patricia Akemi Assato; Paula Rahal; Paulo Inacio da Costa; Rafael dos Santos Bezerra; Raquel de Lello Rocha Campos Cassano. NGS Soluções Genômicas: Pilar Drummond Sampaio Corrêa Mariani. FZEA-USP Pirassununga: Mirele Daiana Poleti; Raul Machado Neto; Ricardo Augusto Brassaloti; Ricardo Haddad; Rodrigo Tocantins Calado. FAMERP-SJRP: Cecília Artico Banho; Sandra Coccuzzo Sampaio; Svetoslav Nanev Slavov; Vagner Fonseca; Vincent Louis Viala |
| EPI_ISL_1795225, EPI_ISL_2345456 | UNIDADE BASICA DE SAUDE DR WILSON HAYEK SAIHG | Instituto Butantan / ESALQ-Piracicaba | Antonio Jorge Martins; Bianca Cechetto Carlos. Mendelics: Bibiana Santos; Claudia Renata dos Santos Barros; David Schlesinger; David Schlesinger. Hemocentro Ribeirão Preto: Simone Kashima; Debora Botequio Moretti; Debora Botequio Moretti. Centro de Genômica Funcional da ESALQ: Luiz Lehmann Coutinho; Dimas Tadeu Covas; Elaine Cristina Marqueze; Elaine Vieira Santos; Elaine Vieira dos Santos; Elisangela Chicaroni Mattos; Erika Freitas; Evandra Strazza Rodrigues; Felipe Allan da Silva da Costa; Flavia Aburjaile; Guilherme Targino Valente; Heidge Fukumasu; Heidge Fukumasu. USP-Botucatu: Rejane Maria Tommasini Grotto; Instituto Butantan: Alexander Roberto Precioso; Jayme A. Souza-Neto; Jayme Augusto de Souza-Neto; Jessika Cristina Chagas Lesbon; José Salvatore Leister Patané; João Paulo Kitajima; Luiz Alcantara; Luiz Carlos Junior de Alcantara; Luiz Lehmann Coutinho; Maria Carolina Elias; Marta Giovanetti; Maurício Lacerda Nogueira; Patricia Akemi Assato; Rafael dos Santos Bezerra; Raquel de Lello Rocha Campos Cassano. NGS Soluções Genômicas: Pilar Drummond Sampaio Corrêa Mariani. FZEA-USP Pirassununga: Mirele Daiana Poleti; Raul Machado Neto; Rejane Maria Tommasini Grotto; Ricardo Augusto Brassaloti; Ricardo Haddad; Rodrigo Tocantins Calado.; Sandra Coccuzzo Sampaio; Sandra Coccuzzo Sampaio Vessoni; Simone Kashima; Svetoslav Nanev Slavov; Vagner Fonseca; Vincent Louis Viala |
| EPI_ISL_1966555 | UNIDADE BASICA DE SAUDE II DE TARABAI | Instituto Butantan / FZEA-USP (Pirassununga) | Antonio Jorge Martins; Bianca Cechetto Carlos. Mendelics: Bibiana Santos; Claudia Renata dos Santos Barros; Cíntia Bittar; David Schlesinger. Hemocentro Ribeirão Preto: Simone Kashima; Debora Botequio Moretti; Elaine Cristina Marqueze; Elaine Vieira dos Santos; Elisangela Chicaroni Mattos; Erika Freitas; Evandra Strazza Rodrigues; Felipe Allan da Silva da Costa; Flavia Aburjaile; Fábio Sossai Possebon; Guilherme Campos; Guilherme Targino Valente; Heidge Fukumasu. USP-Botucatu: Rejane Maria Tommasini Grotto; Helena Lage Ferreira; Instituto Butantan: Dimas Tadeu Covas; Jardelina de Souza Todao Bernardino; Jayme A. Souza-Neto; Jessika Cristina Chagas Lesbon; Jorge A. Petroli Marchesi; José Salvatore Leister Patané; João Paulo Kitajima; João Pessoa Araújo Jr.; Leila Sabrina Ullmann; Loyze Paola Oliveira de Lima; Luiz Aurelio de Campos Crispin. Centro de Genômica Funcional da ESALQ: Luiz Lehmann Coutinho; Luiz Carlos Junior de Alcantara; Lívia Sacchetto; Maisa C. Pereira Parra; Maria Carolina Elias; Marta Giovanetti; Marília Moraes; Maurício Lacerda Nogueira. Prefeitura de Sao Paulo: Melissa Palmieri.; Patricia Akemi Assato; Paula Rahal; Paulo Inacio da Costa; Rafael dos Santos Bezerra; Raquel de Lello Rocha Campos Cassano. NGS Soluções Genômicas: Pilar Drummond Sampaio Corrêa Mariani. FZEA-USP Pirassununga: Mirele Daiana Poleti; Raul Machado Neto; Ricardo Augusto Brassaloti; Ricardo Haddad; Rodrigo Tocantins Calado. FAMERP-SJRP: Cecília Artico Banho; Sandra Coccuzzo Sampaio; Svetoslav Nanev Slavov; Vagner Fonseca; Vincent Louis Viala |
| EPI_ISL_2344760 | UNIDADE BASICA DE SAUDE III DE ALFREDO MARCONDES | Instituto Butantan / ESALQ-Piracicaba | Antonio Jorge Martins; Claudia Renata dos Santos Barros; David Schlesinger; Debora Botequio Moretti; Dimas Tadeu Covas; Elaine Cristina Marqueze; Elaine Vieira Santos; Evandra Strazza Rodrigues; Heidge Fukumasu; Jayme Augusto de Souza-Neto; José Salvatore Leister Patané; Luiz Alcantara; Luiz Lehmann Coutinho; Maria Carolina Elias; Maurício Lacerda Nogueira; Rafael dos Santos Bezerra; Raul Machado Neto; Rejane Maria Tommasini Grotto; Ricardo Haddad; Sandra Coccuzzo Sampaio Vessoni; Simone Kashima; Svetoslav Nanev Slavov; Vincent Louis Viala |
| EPI_ISL_3912302 | UNIDADE BASICA DE SAUDE MUNICIPAL | Analytical Competence Molecular Epidemiology Lab/ACME, Oswaldo Cruz Foundation, Ceara (FIOCRUZ CE) | Cleber Furtado Aksenen; Fabio Miyajima; Fernando Braga Stehling; Francisco Eder de Moura Lopes; Jamille Maria Mendes Bezerra; Joaquim Cesar do Nascimento Sousa Junior; Pedro Miguel Carneiro Jeronimo; Suzana Porto Almeida & Lucas Delerino on behalf of COVID-19 FIOCRUZ Genomic Network; Thais Ferreira de Oliveira; Thais de Oliveira Costa; Ticiane Cavalcante de Souza; Veridiana Pessoa Miyajima |
| EPI_ISL_3102256, EPI_ISL_3102325, EPI_ISL_3102327 | UNIDADE BASICA DE SAUDE RAIMUNDO FONTELE GOMES | Analytical Competence Molecular Epidemiology Lab/ACME, Oswaldo Cruz Foundation, Ceara (FIOCRUZ CE) | Cleber Furtado Aksenen; Fabio Miyajima; Fernando Braga Stehling; Francisco Eder de Moura Lopes; Jamille Maria Mendes Bezerra; Joaquim César do Nascimento Sousa Junior; Pedro Miguel Carneiro Jeronimo; Suzana Porto Almeida e Lucas Delerino; Thais Ferreira de Oliveira; Thais de Oliveira Costa; Ticiane Cavalcante de Souza; Veridiana Pessoa Miyajima |
| EPI_ISL_3912305 | UNIDADE BASICA DE SAUDE SEDE I | Analytical Competence Molecular Epidemiology Lab/ACME, Oswaldo | Cleber Furtado Aksenen; Fabio Miyajima; Fernando Braga Stehling; Francisco Eder de Moura Lopes; Jamille Maria Mendes Bezerra; Joaquim Cesar do Nascimento Sousa Junior; Pedro Miguel Carneiro Jeronimo; Suzana Porto Almeida & Lucas Delerino on behalf of COVID-19 FIOCRUZ Genomic Network; Thais Ferreira de Oliveira; Thais de Oliveira Costa; Ticiane Cavalcante de Souza; Veridiana Pessoa Miyajima |

| | | | |
|---|---|---|---|
| | | Cruz Foundation, Ceara (FIOCRUZ CE) | |
| EPI_ISL_3912306 | UNIDADE BASICA DE SAUDE SEDE II | Analytical Competence Molecular Epidemiology Lab/ACME, Oswaldo Cruz Foundation, Ceara (FIOCRUZ CE) | Cleber Furtado Aksenen; Fabio Miyajima; Fernando Braga Stehling; Francisco Eder de Moura Lopes; Jamille Maria Mendes Bezerra; Joaquim Cesar do Nascimento Sousa Junior; Pedro Miguel Carneiro Jeronimo; Suzana Porto Almeida & Lucas Delerino on behalf of COVID-19 FIOCRUZ Genomic Network; Thais Ferreira de Oliveira; Thais de Oliveira Costa; Ticiane Cavalcante de Souza; Veridiana Pessoa Miyajima |
| EPI_ISL_1795226, EPI_ISL_2345457 | UNIDADE BASICA DR JOSE PARASSU CARVALHO | Instituto Butantan / ESALQ-Piracicaba | Antonio Jorge Martins; Bianca Cechetto Carlos. Mendelics: Bibiana Santos; Claudia Renata dos Santos Barros; David Schlesinger; David Schlesinger. Hemocentro Ribeirão Preto: Simone Kashima; Debora Botequio Moretti; Debora Botequio Moretti. Centro de Genômica Funcional da ESALQ: Luiz Lehmann Coutinho; Dimas Tadeu Covas; Elaine Cristina Marqueze; Elaine Vieira Santos; Elaine Vieira dos Santos; Elisangela Chicaroni Mattos; Erika Freitas; Evandra Strazza Rodrigues; Felipe Allan da Silva da Costa; Flavia Aburjaile; Guilherme Targino Valente; Heidge Fukumasu; Heidge Fukumasu. USP-Botucatu: Rejane Maria Tommasini Grotto; Instituto Butantan: Alexander Roberto Precioso; Jayme A. Souza-Neto; Jayme Augusto de Souza-Neto; Jessika Cristina Chagas Lesbon; José Salvatore Leister Patané; João Paulo Kitajima; Luiz Alcantara; Luiz Carlos Junior de Alcantara; Luiz Lehmann Coutinho; Maria Carolina Elias; Marta Giovanetti; Maurício Lacerda Nogueira; Patricia Akemi Assato; Rafael dos Santos Bezerra; Raquel de Lello Rocha Campos Cassano. NGS Soluções Genômicas: Pilar Drummond Sampaio Corrêa Mariani. FZEA-USP Pirassununga: Mirele Daiana Poleti; Raul Machado Neto; Rejane Maria Tommasini Grotto; Ricardo Augusto Brassaloti; Ricardo Haddad; Rodrigo Tocantins Calado.; Sandra Coccuzzo Sampaio; Sandra Coccuzzo Sampaio Vessoni; Simone Kashima; Svetoslav Nanev Slavov; Vincent Louis Viala |
| EPI_ISL_1795305, EPI_ISL_2345553 | UNIDADE DA SAUDE DO ADULTO CASA BRANCA PREFEITURA | Instituto Butantan / ESALQ-Piracicaba | Antonio Jorge Martins; Bianca Cechetto Carlos. Mendelics: Bibiana Santos; Claudia Renata dos Santos Barros; David Schlesinger; David Schlesinger. Hemocentro Ribeirão Preto: Simone Kashima; Debora Botequio Moretti; Debora Botequio Moretti. Centro de Genômica Funcional da ESALQ: Luiz Lehmann Coutinho; Dimas Tadeu Covas; Elaine Cristina Marqueze; Elaine Vieira Santos; Elaine Vieira dos Santos; Elisangela Chicaroni Mattos; Erika Freitas; Evandra Strazza Rodrigues; Felipe Allan da Silva da Costa; Flavia Aburjaile; Guilherme Targino Valente; Heidge Fukumasu; Heidge Fukumasu. USP-Botucatu: Rejane Maria Tommasini Grotto; Instituto Butantan: Alexander Roberto Precioso; Jayme A. Souza-Neto; Jayme Augusto de Souza-Neto; Jessika Cristina Chagas Lesbon; José Salvatore Leister Patané; João Paulo Kitajima; Luiz Alcantara; Luiz Carlos Junior de Alcantara; Luiz Lehmann Coutinho; Maria Carolina Elias; Marta Giovanetti; Maurício Lacerda Nogueira; Patricia Akemi Assato; Rafael dos Santos Bezerra; Raquel de Lello Rocha Campos Cassano. NGS Soluções Genômicas: Pilar Drummond Sampaio Corrêa Mariani. FZEA-USP Pirassununga: Mirele Daiana Poleti; Raul Machado Neto; Rejane Maria Tommasini Grotto; Ricardo Augusto Brassaloti; Ricardo Haddad; Rodrigo Tocantins Calado.; Sandra Coccuzzo Sampaio; Sandra Coccuzzo Sampaio Vessoni; Simone Kashima; Svetoslav Nanev Slavov; Vagner Fonseca; Vincent Louis Viala |
| EPI_ISL_2801319, EPI_ISL_3912310 | UNIDADE DE PRONTO ATENDIMENTO ARACATI UPA ARACATI | Analytical Competence Molecular Epidemiology Lab/ACME, Oswaldo Cruz Foundation, Ceara (FIOCRUZ CE) | Cleber Furtado Aksenen; Cleber Furtado Aksenen e Suzana Porto Almeida; Fabio Miyajima; Fernando Braga Stehling; Francisco Eder de Moura Lopes; Jamille Maria Mendes Bezerra; Joaquim Cesar do Nascimento Sousa Junior; Joaquim César do Nascimento Sousa Junior; Pedro Miguel Carneiro Jeronimo; Suzana Porto Almeida & Lucas Delerino on behalf of COVID-19 FIOCRUZ Genomic Network; Thais Ferreira de Oliveira; Thais de Oliveira Costa; Ticiane Cavalcante de Souza; Veridiana Pessoa Miyajima |
| EPI_ISL_2209786, EPI_ISL_2209801, EPI_ISL_2209802, EPI_ISL_2209807, EPI_ISL_2209809, EPI_ISL_2479920 | UNIDADE DE PRONTO ATENDIMENTO CENTRAL DE CARAGUATATUBA | Instituto Butantan | Antonio Jorge Martins; Claudia Renata dos Santos Barros; David Schlesinger; Debora Botequio Moretti; Dimas Tadeu Covas; Elaine Cristina Marqueze; Elaine Vieira Santos; Evandra Strazza Rodrigues; Heidge Fukumasu; Jayme Augusto de Souza-Neto; José Salvatore Leister Patané; Luiz Alcantara; Luiz Lehmann Coutinho; Maria Carolina Elias; Maurício Lacerda Nogueira; Rafael dos Santos Bezerra; Raul Machado Neto; Rejane Maria Tommasini Grotto; Ricardo Haddad; Sandra Coccuzzo Sampaio Vessoni; Simone Kashima; Svetoslav Nanev Slavov; Vincent Louis Viala; Vincent Louis Viala. |
| EPI_ISL_2344684, EPI_ISL_2344707, EPI_ISL_2344717, EPI_ISL_2344732, EPI_ISL_2345302 | UNIDADE DE PRONTO ATENDIMENTO CENTRAL DE CARAGUATATUBA | Instituto Butantan / UNESP-Botucatu | Antonio Jorge Martins; Claudia Renata dos Santos Barros; David Schlesinger; Debora Botequio Moretti; Dimas Tadeu Covas; Elaine Cristina Marqueze; Elaine Vieira Santos; Evandra Strazza Rodrigues; Heidge Fukumasu; Jayme Augusto de Souza-Neto; José Salvatore Leister Patané; Luiz Alcantara; Luiz Lehmann Coutinho; Maria Carolina Elias; Maurício Lacerda Nogueira; Rafael dos Santos Bezerra; Raul Machado Neto; Rejane Maria Tommasini Grotto; Ricardo Haddad; Sandra Coccuzzo Sampaio Vessoni; Simone Kashima; Svetoslav Nanev Slavov; Vincent Louis Viala |
| EPI_ISL_3912312, EPI_ISL_3912313 | UNIDADE DE PRONTO ATENDIMENTO DE CAUCAIA | Analytical Competence Molecular Epidemiology Lab/ACME, Oswaldo Cruz Foundation, Ceara (FIOCRUZ CE) | Cleber Furtado Aksenen; Fabio Miyajima; Fernando Braga Stehling; Francisco Eder de Moura Lopes; Jamille Maria Mendes Bezerra; Joaquim Cesar do Nascimento Sousa Junior; Pedro Miguel Carneiro Jeronimo; Suzana Porto Almeida & Lucas Delerino on behalf of COVID-19 FIOCRUZ Genomic Network; Thais Ferreira de Oliveira; Thais de Oliveira Costa; Ticiane Cavalcante de Souza; Veridiana Pessoa Miyajima |
| EPI_ISL_3912096, EPI_ISL_3912319 | UNIDADE DE PRONTO ATENDIMENTO DE EUSEBIO | Analytical Competence Molecular Epidemiology Lab/ACME, Oswaldo Cruz Foundation, Ceara (FIOCRUZ CE) | Cleber Furtado Aksenen; Fabio Miyajima; Fernando Braga Stehling; Francisco Eder de Moura Lopes; Jamille Maria Mendes Bezerra; Joaquim Cesar do Nascimento Sousa Junior; Pedro Miguel Carneiro Jeronimo; Suzana Porto Almeida & Lucas Delerino on behalf of COVID-19 FIOCRUZ Genomic Network; Thais Ferreira de Oliveira; Thais de Oliveira Costa; Ticiane Cavalcante de Souza; Veridiana Pessoa Miyajima |
| EPI_ISL_1966248 | UNIDADE DE PRONTO ATENDIMENTO DE JALES | Instituto Butantan / Mendelics | Antonio Jorge Martins; Bianca Cechetto Carlos. Mendelics: Bibiana Santos; Claudia Renata dos Santos Barros; Cíntia Bittar; David Schlesinger. Hemocentro Ribeirão Preto: Simone Kashima; Debora Botequio Moretti; Elaine Cristina Marqueze; Elaine Vieira dos Santos; Elisangela Chicaroni Mattos; Erika Freitas; Evandra Strazza Rodrigues; Felipe Allan da Silva da Costa; Flavia Aburjaile; Fábio Sossai Possebon; Guilherme Campos; Guilherme Targino Valente; Heidge Fukumasu. USP-Botucatu: Rejane Maria Tommasini Grotto; Helena Lage Ferreira; Instituto Butantan: Dimas Tadeu Covas; Jardelina de Souza Todao Bernardino; Jayme A. Souza-Neto; Jessika Cristina Chagas Lesbon; Jorge A. Petroli Marchesi; José Salvatore Leister Patané; João Paulo Kitajima; João Pessoa Araújo Jr.; Leila Sabrina Ullmann; Loyze Paola Oliveira de Lima; Luiz Aurelio de Campos Crispin. Centro de Genômica Funcional da ESALQ: Luiz Lehmann Coutinho; Luiz Carlos Junior de Alcantara; Lívia Sacchetto; Maisa C. Pereira Parra; Maria Carolina Elias; Marta Giovanetti; Marília Moraes; Maurício Lacerda Nogueira. Prefeitura de Sao Paulo: Melissa Palmieri.; Patricia Akemi Assato; Paula Rahal; Paulo Inacio da Costa; Rafael dos Santos Bezerra; Raquel de Lello Rocha Campos Cassano. NGS Soluções Genômicas: Pilar Drummond Sampaio Corrêa Mariani. FZEA-USP Pirassununga: Mirele Daiana Poleti; Raul Machado Neto; Ricardo Augusto Brassaloti; Ricardo Haddad; Rodrigo Tocantins Calado. FAMERP-SJRP: Cecília Artico Banho; Sandra Coccuzzo Sampaio; Svetoslav Nanev Slavov; Vagner Fonseca; Vincent Louis Viala |
| EPI_ISL_2345988 | UNIDADE DE PRONTO ATENDIMENTO DE VARZEA PAULISTA UPA II | Instituto Butantan | Antonio Jorge Martins; Claudia Renata dos Santos Barros; David Schlesinger; Debora Botequio Moretti; Dimas Tadeu Covas; Elaine Cristina Marqueze; Elaine Vieira Santos; Evandra Strazza Rodrigues; Heidge Fukumasu; Jayme Augusto de Souza-Neto; José Salvatore Leister Patané; Luiz Alcantara; Luiz Lehmann Coutinho; Maria Carolina Elias; Maurício Lacerda Nogueira; Rafael dos Santos Bezerra; Raul Machado Neto; Rejane Maria Tommasini Grotto; Ricardo Haddad; Sandra Coccuzzo Sampaio Vessoni; Simone Kashima; Svetoslav Nanev Slavov; Vincent Louis Viala |
| EPI_ISL_1966721, EPI_ISL_1966723, EPI_ISL_2171033, EPI_ISL_2171035, EPI_ISL_2171039 | UNIDADE DE PRONTO ATENDIMENTO DE VARZEA PAULISTA UPA II | Instituto Butantan / Mendelics | Antonio Jorge Martins; Bianca Cechetto Carlos. Mendelics: Bibiana Santos; Claudia Renata dos Santos Barros; Cíntia Bittar; David Schlesinger. Hemocentro Ribeirão Preto: Simone Kashima; Debora Botequio Moretti; Elaine Cristina Marqueze; Elaine Vieira dos Santos; Elisangela Chicaroni Mattos; Erika Freitas; Evandra Strazza Rodrigues; Felipe Allan da Silva da Costa; Flavia Aburjaile; Fábio Sossai Possebon; Guilherme Campos; Guilherme Targino Valente; Heidge Fukumasu. USP-Botucatu: Rejane Maria Tommasini Grotto; Helena Lage Ferreira; Instituto Butantan: Dimas Tadeu Covas; Jardelina de Souza Todao Bernardino; Jayme A. Souza-Neto; Jessika Cristina Chagas Lesbon; Jorge A. Petroli Marchesi; José Salvatore Leister Patané; João Paulo Kitajima; João Pessoa Araújo Jr.; Leila Sabrina Ullmann; Loyze Paola Oliveira de Lima; Luiz Aurelio de Campos Crispin. Centro de Genômica Funcional da ESALQ: Luiz Lehmann Coutinho; Luiz Carlos Junior de Alcantara; Lívia Sacchetto; Maisa C. Pereira Parra; Maria Carolina Elias; Marta Giovanetti; Marília Moraes; Maurício Lacerda Nogueira. Prefeitura de Sao Paulo: Melissa Palmieri.; Patricia Akemi Assato; Paula Rahal; Paulo Inacio da Costa; Rafael dos Santos Bezerra; Raquel de Lello Rocha Campos Cassano. NGS Soluções Genômicas: Pilar Drummond Sampaio Corrêa Mariani. FZEA-USP Pirassununga: Mirele Daiana Poleti; Raul Machado Neto; Ricardo Augusto Brassaloti; Ricardo Haddad; Rodrigo Tocantins Calado. FAMERP-SJRP: Cecília Artico Banho; Sandra Coccuzzo Sampaio; Svetoslav Nanev Slavov; Vagner Fonseca; Vincent Louis Viala |
| EPI_ISL_1967042, EPI_ISL_1967047, EPI_ISL_1967048, EPI_ISL_1967049 | UNIDADE DE PRONTO ATENDIMENTO UPA I SANTA ISABEL | Instituto Butantan / ESALQ-USP (Piracicaba) | Antonio Jorge Martins; Bianca Cechetto Carlos. Mendelics: Bibiana Santos; Claudia Renata dos Santos Barros; Cíntia Bittar; David Schlesinger. Hemocentro Ribeirão Preto: Simone Kashima; Debora Botequio Moretti; Elaine Cristina Marqueze; Elaine Vieira dos Santos; Elisangela Chicaroni Mattos; Erika Freitas; Evandra Strazza Rodrigues; Felipe Allan da Silva da Costa; Flavia Aburjaile; Fábio Sossai Possebon; Guilherme Campos; Guilherme Targino Valente; Heidge Fukumasu. USP-Botucatu: Rejane Maria Tommasini Grotto; Helena Lage Ferreira; Instituto Butantan: Dimas Tadeu Covas; Jardelina de Souza Todao Bernardino; Jayme A. Souza-Neto; Jessika Cristina Chagas Lesbon; Jorge A. Petroli Marchesi; José Salvatore Leister Patané; João Paulo Kitajima; João Pessoa Araújo Jr.; Leila Sabrina Ullmann; Loyze Paola Oliveira de Lima; Luiz Aurelio de Campos Crispin. Centro de Genômica Funcional da ESALQ: Luiz Lehmann Coutinho; Luiz Carlos Junior de Alcantara; Lívia Sacchetto; Maisa C. Pereira Parra; Maria Carolina Elias; Marta Giovanetti; Marília Moraes; Maurício Lacerda Nogueira. Prefeitura de Sao Paulo: Melissa Palmieri.; Patricia Akemi Assato; Paula Rahal; Paulo Inacio da Costa; Rafael dos Santos Bezerra; Raquel de Lello Rocha Campos Cassano. NGS Soluções Genômicas: Pilar Drummond Sampaio Corrêa Mariani. FZEA-USP Pirassununga: Mirele Daiana Poleti; Raul Machado Neto; Ricardo Augusto Brassaloti; Ricardo Haddad; Rodrigo Tocantins Calado. FAMERP-SJRP: Cecília Artico Banho; Sandra Coccuzzo Sampaio; Svetoslav Nanev Slavov; Vagner Fonseca; Vincent Louis Viala |
| EPI_ISL_1795230, EPI_ISL_1795231, EPI_ISL_2345461, EPI_ISL_2345463 | UNIDADE DE SAUDE DA FAMILIA JOSE ADALBERTO LELLIS GARCIA | Instituto Butantan / ESALQ-Piracicaba | Antonio Jorge Martins; Bianca Cechetto Carlos. Mendelics: Bibiana Santos; Claudia Renata dos Santos Barros; David Schlesinger; David Schlesinger. Hemocentro Ribeirão Preto: Simone Kashima; Debora Botequio Moretti; Debora Botequio Moretti. Centro de Genômica Funcional da ESALQ: Luiz Lehmann Coutinho; Dimas Tadeu Covas; Elaine Cristina Marqueze; Elaine Vieira Santos; Elaine Vieira dos Santos; Elisangela Chicaroni Mattos; Erika Freitas; Evandra Strazza Rodrigues; Felipe Allan da Silva da Costa; Flavia Aburjaile; Guilherme Targino Valente; Heidge Fukumasu; Heidge Fukumasu. USP-Botucatu: Rejane Maria Tommasini Grotto; Instituto Butantan: Alexander Roberto Precioso; Jayme A. Souza-Neto; Jayme Augusto de Souza-Neto; Jessika Cristina Chagas Lesbon; José Salvatore Leister Patané; João Paulo Kitajima; Luiz Alcantara; Luiz Carlos Junior de Alcantara; Luiz Lehmann Coutinho; Maria Carolina Elias; Marta Giovanetti; Maurício Lacerda Nogueira; Patricia Akemi Assato; Rafael dos Santos Bezerra; Raquel de Lello Rocha Campos Cassano. NGS Soluções Genômicas: Pilar Drummond Sampaio Corrêa Mariani. FZEA-USP Pirassununga: Mirele Daiana Poleti; Raul Machado Neto; Rejane Maria Tommasini Grotto; Ricardo Augusto Brassaloti; Ricardo Haddad; Rodrigo Tocantins Calado.; Sandra Coccuzzo Sampaio; Sandra Coccuzzo Sampaio Vessoni; Simone Kashima; Svetoslav Nanev Slavov; Vagner Fonseca; Vincent Louis Viala |
| EPI_ISL_1795293, EPI_ISL_1795294, EPI_ISL_2345534, EPI_ISL_2345535 | UNIDADE DE SAUDE DE ITOBI ALCIBIADES PIRES | Instituto Butantan / ESALQ-Piracicaba | Antonio Jorge Martins; Bianca Cechetto Carlos. Mendelics: Bibiana Santos; Claudia Renata dos Santos Barros; David Schlesinger; David Schlesinger. Hemocentro Ribeirão Preto: Simone Kashima; Debora Botequio Moretti; Debora Botequio Moretti. Centro de Genômica Funcional da ESALQ: Luiz Lehmann Coutinho; Dimas Tadeu Covas; Elaine Cristina Marqueze; Elaine Vieira Santos; Elaine Vieira dos Santos; Elisangela Chicaroni Mattos; Erika Freitas; Evandra Strazza Rodrigues; Felipe Allan da Silva da Costa; Flavia Aburjaile; Guilherme Targino Valente; Heidge Fukumasu; Heidge Fukumasu. USP-Botucatu: Rejane Maria Tommasini Grotto; Instituto Butantan: Alexander Roberto Precioso; Jayme A. Souza-Neto; Jayme Augusto de Souza-Neto; Jessika Cristina Chagas Lesbon; José Salvatore Leister Patané; João Paulo Kitajima; Luiz Alcantara; Luiz Carlos Junior de Alcantara; Luiz Lehmann Coutinho; Maria Carolina Elias; Marta Giovanetti; Maurício Lacerda Nogueira; Patricia Akemi Assato; Rafael dos Santos Bezerra; Raquel de Lello Rocha Campos Cassano. NGS Soluções Genômicas: Pilar Drummond Sampaio Corrêa Mariani. FZEA-USP Pirassununga: Mirele Daiana Poleti; Raul Machado Neto; Rejane Maria Tommasini Grotto; Ricardo Augusto Brassaloti; Ricardo Haddad; Rodrigo Tocantins Calado.; Sandra Coccuzzo Sampaio; Sandra Coccuzzo Sampaio Vessoni; Simone Kashima; Svetoslav Nanev Slavov; Vagner Fonseca; Vincent Louis Viala |
| EPI_ISL_1795122, EPI_ISL_1795124, EPI_ISL_1795128, EPI_ISL_1795129, EPI_ISL_1795130, EPI_ISL_1795131, EPI_ISL_1795133, EPI_ISL_1795135, EPI_ISL_1795385, EPI_ISL_1795386, EPI_ISL_2345322, EPI_ISL_2345323, EPI_ISL_2345326, EPI_ISL_2345330, EPI_ISL_2345331, EPI_ISL_2345332, EPI_ISL_2345333, EPI_ISL_2345335, EPI_ISL_2345336, EPI_ISL_2345338 | | | |
| see above | UNIDADE DE VIGILANCIA EM SAUDE | Instituto Butantan / ESALQ-Piracicaba | Antonio Jorge Martins; Bianca Cechetto Carlos. Mendelics: Bibiana Santos; Claudia Renata dos Santos Barros; David Schlesinger; David Schlesinger. Hemocentro Ribeirão Preto: Simone Kashima; Debora Botequio Moretti; Debora Botequio Moretti. Centro de Genômica Funcional da ESALQ: Luiz Lehmann Coutinho; Dimas Tadeu Covas; Elaine Cristina Marqueze; Elaine Vieira Santos; Elaine Vieira dos Santos; Elisangela Chicaroni Mattos; Erika Freitas; Evandra Strazza Rodrigues; Felipe Allan da Silva da Costa; Flavia Aburjaile; Guilherme Targino Valente; Heidge Fukumasu; Heidge Fukumasu. USP-Botucatu: Rejane Maria Tommasini Grotto; Instituto Butantan: Alexander Roberto Precioso; Jayme A. Souza-Neto; Jayme Augusto de Souza-Neto; Jessika Cristina Chagas Lesbon; José Salvatore Leister Patané; João Paulo Kitajima; Luiz Alcantara; Luiz Carlos Junior de Alcantara; Luiz Lehmann Coutinho; Maria Carolina Elias; Marta Giovanetti; Maurício Lacerda Nogueira; Patricia Akemi Assato; Rafael dos Santos Bezerra; Raquel de Lello Rocha Campos Cassano. NGS Soluções Genômicas: Pilar Drummond Sampaio Corrêa Mariani. FZEA-USP Pirassununga: Mirele Daiana Poleti; Raul Machado Neto; Rejane Maria Tommasini Grotto; Ricardo Augusto Brassaloti; Ricardo Haddad; Rodrigo Tocantins Calado.; Sandra Coccuzzo Sampaio; Sandra Coccuzzo Sampaio Vessoni; Simone Kashima; Svetoslav Nanev Slavov; Vagner Fonseca; Vincent Louis Viala |
| EPI_ISL_1967053, EPI_ISL_1967057 | UNIDADE DE VIGILANCIA EM SAUDE | Instituto Butantan / ESALQ-USP (Piracicaba) | Antonio Jorge Martins; Bianca Cechetto Carlos. Mendelics: Bibiana Santos; Claudia Renata dos Santos Barros; Cíntia Bittar; David Schlesinger. Hemocentro Ribeirão Preto: Simone Kashima; Debora Botequio Moretti; Elaine Cristina Marqueze; Elaine Vieira dos Santos; Elisangela Chicaroni Mattos; Erika Freitas; Evandra Strazza Rodrigues; Felipe Allan da Silva da Costa; Flavia Aburjaile; Fábio Sossai Possebon; Guilherme Campos; Guilherme Targino Valente; Heidge Fukumasu. USP-Botucatu: Rejane Maria Tommasini Grotto; Helena Lage Ferreira; Instituto Butantan: Dimas Tadeu Covas; Jardelina de Souza Todao Bernardino; Jayme A. Souza-Neto; Jessika Cristina Chagas Lesbon; Jorge A. Petroli Marchesi; José Salvatore Leister Patané; João Paulo Kitajima; João Pessoa Araújo Jr.; Leila Sabrina Ullmann; Loyze Paola Oliveira de Lima; Luiz Aurelio de Campos Crispin. Centro de Genômica Funcional da ESALQ: Luiz Lehmann Coutinho; Luiz Carlos Junior de Alcantara; Lívia Sacchetto; Maisa C. Pereira Parra; Maria Carolina Elias; Marta Giovanetti; Marília Moraes; Maurício Lacerda Nogueira. Prefeitura de Sao Paulo: Melissa Palmieri.; Patricia Akemi Assato; Paula Rahal; Paulo Inacio da Costa; Rafael dos Santos Bezerra; Raquel de Lello Rocha Campos Cassano. NGS Soluções Genômicas: Pilar Drummond Sampaio Corrêa Mariani. FZEA-USP Pirassununga: Mirele Daiana Poleti; Raul Machado Neto; Ricardo Augusto Brassaloti; Ricardo Haddad; Rodrigo Tocantins Calado. FAMERP-SJRP: Cecília Artico Banho; Sandra Coccuzzo Sampaio; Svetoslav Nanev Slavov; Vagner Fonseca; Vincent Louis Viala |
| EPI_ISL_1966549, EPI_ISL_1966550 | UNIDADE DE VIGILANCIA EM SAUDE | Instituto Butantan / FZEA-USP (Pirassununga) | Antonio Jorge Martins; Bianca Cechetto Carlos. Mendelics: Bibiana Santos; Claudia Renata dos Santos Barros; Cíntia Bittar; David Schlesinger. Hemocentro Ribeirão Preto: Simone Kashima; Debora Botequio Moretti; Elaine Cristina Marqueze; Elaine Vieira dos Santos; Elisangela Chicaroni Mattos; Erika Freitas; Evandra Strazza Rodrigues; Felipe Allan da Silva da Costa; Flavia Aburjaile; Fábio Sossai Possebon; Guilherme Campos; Guilherme Targino Valente; Heidge Fukumasu. USP-Botucatu: Rejane Maria Tommasini Grotto; Helena Lage Ferreira; Instituto Butantan: Dimas Tadeu Covas; Jardelina de Souza Todao Bernardino; Jayme A. Souza-Neto; Jessika Cristina Chagas Lesbon; Jorge A. Petroli Marchesi; José Salvatore Leister Patané; João Paulo Kitajima; João Pessoa Araújo Jr.; Leila Sabrina Ullmann; Loyze Paola Oliveira de Lima; Luiz Aurelio de Campos Crispin. Centro de Genômica Funcional da ESALQ: Luiz Lehmann Coutinho; Luiz Carlos Junior de Alcantara; Lívia Sacchetto; Maisa C. Pereira Parra; Maria Carolina Elias; Marta Giovanetti; Marília Moraes; Maurício Lacerda Nogueira. Prefeitura de Sao Paulo: Melissa Palmieri.; Patricia Akemi Assato; Paula Rahal; Paulo Inacio da Costa; Rafael dos Santos Bezerra; Raquel de Lello Rocha Campos Cassano. NGS Soluções Genômicas: Pilar Drummond Sampaio Corrêa Mariani. FZEA-USP Pirassununga: Mirele Daiana Poleti; Raul Machado Neto; Ricardo Augusto Brassaloti; Ricardo Haddad; Rodrigo Tocantins Calado. FAMERP-SJRP: Cecília Artico Banho; Sandra Coccuzzo Sampaio; Svetoslav Nanev Slavov; Vagner Fonseca; Vincent Louis Viala |
| EPI_ISL_2209856 | UNIDADE DE VIGILANCIA EPIDEMIOLOGICA | Instituto Butantan | Antonio Jorge Martins; Claudia Renata dos Santos Barros; David Schlesinger; Debora Botequio Moretti; Dimas Tadeu Covas; Elaine Cristina Marqueze; Elaine Vieira Santos; Evandra Strazza Rodrigues; Heidge Fukumasu; Jayme Augusto de Souza-Neto; José Salvatore Leister Patané; Luiz Alcantara; Luiz Lehmann Coutinho; Maria Carolina Elias; Maurício Lacerda Nogueira; Rafael dos Santos Bezerra; Raul Machado Neto; Rejane Maria Tommasini Grotto; Ricardo Haddad; Sandra Coccuzzo Sampaio Vessoni; Simone Kashima; Svetoslav Nanev Slavov; Vincent Louis Viala. |
| EPI_ISL_2209129, EPI_ISL_2209261, EPI_ISL_2344688 | UNIDADE DE VIGILANCIA EPIDEMIOLOGICA | Instituto Butantan / FZEA-USP-Pirassununga | Antonio Jorge Martins; Claudia Renata dos Santos Barros; David Schlesinger; Debora Botequio Moretti; Dimas Tadeu Covas; Elaine Cristina Marqueze; Elaine Vieira Santos; Evandra Strazza Rodrigues; Heidge Fukumasu; Jayme Augusto de Souza-Neto; José Salvatore Leister Patané; Luiz Alcantara; Luiz Lehmann Coutinho; Maria Carolina Elias; Maurício Lacerda Nogueira; Rafael dos Santos Bezerra; Raul Machado Neto; Rejane Maria Tommasini Grotto; Ricardo Haddad; Sandra Coccuzzo Sampaio Vessoni; Simone Kashima; Svetoslav Nanev Slavov; Vincent Louis Viala |
| EPI_ISL_1795110, EPI_ISL_2345309 | UNIDADE ESF DR LUIZ SPINA | Instituto Butantan / ESALQ-Piracicaba | Antonio Jorge Martins; Bianca Cechetto Carlos. Mendelics: Bibiana Santos; Claudia Renata dos Santos Barros; David Schlesinger; David Schlesinger. Hemocentro Ribeirão Preto: Simone Kashima; Debora Botequio Moretti; Debora Botequio Moretti. Centro de Genômica Funcional da ESALQ: Luiz Lehmann Coutinho; Dimas Tadeu Covas; Elaine Cristina Marqueze; Elaine Vieira Santos; Elaine Vieira dos Santos; Elisangela Chicaroni Mattos; Erika Freitas; Evandra Strazza Rodrigues; Felipe Allan da Silva da Costa; Flavia Aburjaile; Guilherme Targino Valente; Heidge Fukumasu; Heidge Fukumasu. USP-Botucatu: Rejane Maria Tommasini Grotto; Instituto Butantan: Alexander Roberto Precioso; Jayme A. Souza-Neto; Jayme Augusto de Souza-Neto; Jessika Cristina Chagas Lesbon; José Salvatore Leister Patané; João Paulo Kitajima; Luiz Alcantara; Luiz Carlos Junior de Alcantara; Luiz Lehmann Coutinho; Maria Carolina Elias; Marta Giovanetti; Maurício Lacerda Nogueira; Patricia Akemi Assato; Rafael dos Santos Bezerra; Raquel de Lello Rocha Campos Cassano. NGS Soluções Genômicas: Pilar Drummond Sampaio Corrêa Mariani. FZEA-USP Pirassununga: Mirele Daiana Poleti; Raul Machado Neto; Rejane Maria Tommasini Grotto; Ricardo Augusto Brassaloti; Ricardo Haddad; Rodrigo Tocantins Calado.; Sandra Coccuzzo Sampaio; Sandra Coccuzzo Sampaio Vessoni; Simone Kashima; Svetoslav Nanev Slavov; Vagner Fonseca; Vincent Louis Viala |
| EPI_ISL_1795222, | UNIDADE MISTA | Instituto Butantan / | Antonio Jorge Martins; Bianca Cechetto Carlos. Mendelics: Bibiana Santos; Claudia Renata dos Santos Barros; David Schlesinger; David Schlesinger. Hemocentro Ribeirão Preto: Simone Kashima; Debora Botequio Moretti; Debora Botequio Moretti. Centro de Genômica Funcional da ESALQ: Luiz Lehmann Coutinho; Dimas Tadeu Covas; |

| | | | |
|---|---|---|---|
| EPI_ISL_2345452 | ARACOIABA DA SERRA | ESALQ-Piracicaba | Elaine Cristina Marqueze; Elaine Vieira Santos; Elaine Vieira dos Santos; Elisangela Chicaroni Mattos; Erika Freitas; Evandra Strazza Rodrigues; Felipe Allan da Silva da Costa; Flavia Aburjaile; Guilherme Targino Valente; Heidge Fukumasu; Heidge Fukumasu. USP-Botucatu: Rejane Maria Tommasini Grotto; Instituto Butantan: Alexander Roberto Precioso; Jayme A. Souza-Neto; Jayme Augusto de Souza-Neto; Jessika Cristina Chagas Lesbon; José Salvatore Leister Patané; João Paulo Kitajima; Luiz Alcantara; Luiz Carlos Junior de Alcantara; Luiz Lehmann Coutinho; Maria Carolina Elias; Marta Giovanetti; Maurício Lacerda Nogueira; Patricia Akemi Assato; Rafael dos Santos Bezerra; Raquel de Lello Rocha Campos Cassano. NGS Soluções Genômicas: Pilar Drummond Sampaio Corrêa Mariani. FZEA-USP Pirassununga: Mirele Daiana Poleti; Raul Machado Neto; Rejane Maria Tommasini Grotto; Ricardo Augusto Brassaloti; Ricardo Haddad; Rodrigo Tocantins Calado.; Sandra Coccuzzo Sampaio; Sandra Coccuzzo Sampaio Vessoni; Simone Kashima; Svetoslav Nanev Slavov; Vagner Fonseca; Vincent Louis Viala |
| EPI_ISL_1966649 | UNIDADE MISTA DE BOM JESUS DOS PERDOES | Instituto Butantan / Mendelics | Antonio Jorge Martins; Bianca Cechetto Carlos. Mendelics: Bibiana Santos; Claudia Renata dos Santos Barros; Cíntia Bittar; David Schlesinger. Hemocentro Ribeirão Preto: Simone Kashima; Debora Botequio Moretti; Elaine Cristina Marqueze; Elaine Vieira dos Santos; Elisangela Chicaroni Mattos; Erika Freitas; Evandra Strazza Rodrigues; Felipe Allan da Silva da Costa; Flavia Aburjaile; Fábio Sossai Possebon; Guilherme Campos; Guilherme Targino Valente; Heidge Fukumasu. USP-Botucatu: Rejane Maria Tommasini Grotto; Helena Lage Ferreira; Instituto Butantan: Dimas Tadeu Covas; Jardelina de Souza Todao Bernardino; Jayme A. Souza-Neto; Jessika Cristina Chagas Lesbon; Jorge A. Petroli Marchesi; José Salvatore Leister Patané; João Paulo Kitajima; João Pessoa Araújo Jr.; Leila Sabrina Ullmann; Loyze Paola Oliveira de Lima; Luiz Aurelio de Campos Crispin. Centro de Genômica Funcional da ESALQ: Luiz Lehmann Coutinho; Luiz Carlos Junior de Alcantara; Lívia Sacchetto; Maisa C. Pereira Parra; Maria Carolina Elias; Marta Giovanetti; Marília Moraes; Maurício Lacerda Nogueira. Prefeitura de Sao Paulo: Melissa Palmieri.; Patricia Akemi Assato; Paula Rahal; Paulo Inacio da Costa; Rafael dos Santos Bezerra; Raquel de Lello Rocha Campos Cassano. NGS Soluções Genômicas: Pilar Drummond Sampaio Corrêa Mariani. FZEA-USP Pirassununga: Mirele Daiana Poleti; Raul Machado Neto; Ricardo Augusto Brassaloti; Ricardo Haddad; Rodrigo Tocantins Calado. FAMERP-SJRP: Cecília Artico Banho; Sandra Coccuzzo Sampaio; Svetoslav Nanev Slavov; Vagner Fonseca; Vincent Louis Viala |
| EPI_ISL_1966915 | UNIDADE MISTA DE IGUAPE | Instituto Butantan / ESALQ-USP (Piracicaba) | Antonio Jorge Martins; Bianca Cechetto Carlos. Mendelics: Bibiana Santos; Claudia Renata dos Santos Barros; Cíntia Bittar; David Schlesinger. Hemocentro Ribeirão Preto: Simone Kashima; Debora Botequio Moretti; Elaine Cristina Marqueze; Elaine Vieira dos Santos; Elisangela Chicaroni Mattos; Erika Freitas; Evandra Strazza Rodrigues; Felipe Allan da Silva da Costa; Flavia Aburjaile; Fábio Sossai Possebon; Guilherme Campos; Guilherme Targino Valente; Heidge Fukumasu. USP-Botucatu: Rejane Maria Tommasini Grotto; Helena Lage Ferreira; Instituto Butantan: Dimas Tadeu Covas; Jardelina de Souza Todao Bernardino; Jayme A. Souza-Neto; Jessika Cristina Chagas Lesbon; Jorge A. Petroli Marchesi; José Salvatore Leister Patané; João Paulo Kitajima; João Pessoa Araújo Jr.; Leila Sabrina Ullmann; Loyze Paola Oliveira de Lima; Luiz Aurelio de Campos Crispin. Centro de Genômica Funcional da ESALQ: Luiz Lehmann Coutinho; Luiz Carlos Junior de Alcantara; Lívia Sacchetto; Maisa C. Pereira Parra; Maria Carolina Elias; Marta Giovanetti; Marília Moraes; Maurício Lacerda Nogueira. Prefeitura de Sao Paulo: Melissa Palmieri.; Patricia Akemi Assato; Paula Rahal; Paulo Inacio da Costa; Rafael dos Santos Bezerra; Raquel de Lello Rocha Campos Cassano. NGS Soluções Genômicas: Pilar Drummond Sampaio Corrêa Mariani. FZEA-USP Pirassununga: Mirele Daiana Poleti; Raul Machado Neto; Ricardo Augusto Brassaloti; Ricardo Haddad; Rodrigo Tocantins Calado. FAMERP-SJRP: Cecília Artico Banho; Sandra Coccuzzo Sampaio; Svetoslav Nanev Slavov; Vagner Fonseca; Vincent Louis Viala |
| EPI_ISL_1966246 | UNIDADE MISTA DE IGUAPE | Instituto Butantan / Mendelics | Antonio Jorge Martins; Bianca Cechetto Carlos. Mendelics: Bibiana Santos; Claudia Renata dos Santos Barros; Cíntia Bittar; David Schlesinger. Hemocentro Ribeirão Preto: Simone Kashima; Debora Botequio Moretti; Elaine Cristina Marqueze; Elaine Vieira dos Santos; Elisangela Chicaroni Mattos; Erika Freitas; Evandra Strazza Rodrigues; Felipe Allan da Silva da Costa; Flavia Aburjaile; Fábio Sossai Possebon; Guilherme Campos; Guilherme Targino Valente; Heidge Fukumasu. USP-Botucatu: Rejane Maria Tommasini Grotto; Helena Lage Ferreira; Instituto Butantan: Dimas Tadeu Covas; Jardelina de Souza Todao Bernardino; Jayme A. Souza-Neto; Jessika Cristina Chagas Lesbon; Jorge A. Petroli Marchesi; José Salvatore Leister Patané; João Paulo Kitajima; João Pessoa Araújo Jr.; Leila Sabrina Ullmann; Loyze Paola Oliveira de Lima; Luiz Aurelio de Campos Crispin. Centro de Genômica Funcional da ESALQ: Luiz Lehmann Coutinho; Luiz Carlos Junior de Alcantara; Lívia Sacchetto; Maisa C. Pereira Parra; Maria Carolina Elias; Marta Giovanetti; Marília Moraes; Maurício Lacerda Nogueira. Prefeitura de Sao Paulo: Melissa Palmieri.; Patricia Akemi Assato; Paula Rahal; Paulo Inacio da Costa; Rafael dos Santos Bezerra; Raquel de Lello Rocha Campos Cassano. NGS Soluções Genômicas: Pilar Drummond Sampaio Corrêa Mariani. FZEA-USP Pirassununga: Mirele Daiana Poleti; Raul Machado Neto; Ricardo Augusto Brassaloti; Ricardo Haddad; Rodrigo Tocantins Calado. FAMERP-SJRP: Cecília Artico Banho; Sandra Coccuzzo Sampaio; Svetoslav Nanev Slavov; Vagner Fonseca; Vincent Louis Viala |
| EPI_ISL_1795116, EPI_ISL_1795117, EPI_ISL_1795118, EPI_ISL_1795119, EPI_ISL_2345315, EPI_ISL_2345317, EPI_ISL_2345318, EPI_ISL_2345319 | | | |
| see above | UNIDADE MISTA DE LUIZ ANTONIO | Instituto Butantan / ESALQ-Piracicaba | Antonio Jorge Martins; Bianca Cechetto Carlos. Mendelics: Bibiana Santos; Claudia Renata dos Santos Barros; David Schlesinger; David Schlesinger. Hemocentro Ribeirão Preto: Simone Kashima; Debora Botequio Moretti; Debora Botequio Moretti. Centro de Genômica Funcional da ESALQ: Luiz Lehmann Coutinho; Dimas Tadeu Covas; Elaine Cristina Marqueze; Elaine Vieira Santos; Elaine Vieira dos Santos; Elisangela Chicaroni Mattos; Erika Freitas; Evandra Strazza Rodrigues; Felipe Allan da Silva da Costa; Flavia Aburjaile; Guilherme Targino Valente; Heidge Fukumasu; Heidge Fukumasu. USP-Botucatu: Rejane Maria Tommasini Grotto; Instituto Butantan: Alexander Roberto Precioso; Jayme A. Souza-Neto; Jayme Augusto de Souza-Neto; Jessika Cristina Chagas Lesbon; José Salvatore Leister Patané; João Paulo Kitajima; Luiz Alcantara; Luiz Carlos Junior de Alcantara; Luiz Lehmann Coutinho; Maria Carolina Elias; Marta Giovanetti; Maurício Lacerda Nogueira; Patricia Akemi Assato; Rafael dos Santos Bezerra; Raquel de Lello Rocha Campos Cassano. NGS Soluções Genômicas: Pilar Drummond Sampaio Corrêa Mariani. FZEA-USP Pirassununga: Mirele Daiana Poleti; Raul Machado Neto; Rejane Maria Tommasini Grotto; Ricardo Augusto Brassaloti; Ricardo Haddad; Rodrigo Tocantins Calado.; Sandra Coccuzzo Sampaio; Sandra Coccuzzo Sampaio Vessoni; Simone Kashima; Svetoslav Nanev Slavov; Vagner Fonseca; Vincent Louis Viala |
| EPI_ISL_1966864, EPI_ISL_1966865, EPI_ISL_1966867 | UNIDADE MISTA DE SAUDE CAPELA DO ALTO | Instituto Butantan / ESALQ-USP (Piracicaba) | Antonio Jorge Martins; Bianca Cechetto Carlos. Mendelics: Bibiana Santos; Claudia Renata dos Santos Barros; Cíntia Bittar; David Schlesinger. Hemocentro Ribeirão Preto: Simone Kashima; Debora Botequio Moretti; Elaine Cristina Marqueze; Elaine Vieira dos Santos; Elisangela Chicaroni Mattos; Erika Freitas; Evandra Strazza Rodrigues; Felipe Allan da Silva da Costa; Flavia Aburjaile; Fábio Sossai Possebon; Guilherme Campos; Guilherme Targino Valente; Heidge Fukumasu. USP-Botucatu: Rejane Maria Tommasini Grotto; Helena Lage Ferreira; Instituto Butantan: Dimas Tadeu Covas; Jardelina de Souza Todao Bernardino; Jayme A. Souza-Neto; Jessika Cristina Chagas Lesbon; Jorge A. Petroli Marchesi; José Salvatore Leister Patané; João Paulo Kitajima; João Pessoa Araújo Jr.; Leila Sabrina Ullmann; Loyze Paola Oliveira de Lima; Luiz Aurelio de Campos Crispin. Centro de Genômica Funcional da ESALQ: Luiz Lehmann Coutinho; Luiz Carlos Junior de Alcantara; Lívia Sacchetto; Maisa C. Pereira Parra; Maria Carolina Elias; Marta Giovanetti; Marília Moraes; Maurício Lacerda Nogueira. Prefeitura de Sao Paulo: Melissa Palmieri.; Patricia Akemi Assato; Paula Rahal; Paulo Inacio da Costa; Rafael dos Santos Bezerra; Raquel de Lello Rocha Campos Cassano. NGS Soluções Genômicas: Pilar Drummond Sampaio Corrêa Mariani. FZEA-USP Pirassununga: Mirele Daiana Poleti; Raul Machado Neto; Ricardo Augusto Brassaloti; Ricardo Haddad; Rodrigo Tocantins Calado. FAMERP-SJRP: Cecília Artico Banho; Sandra Coccuzzo Sampaio; Svetoslav Nanev Slavov; Vagner Fonseca; Vincent Louis Viala |
| EPI_ISL_3102324, EPI_ISL_3102528 | UNIDADE MISTA DE SAUDE DE MISSAO VELHA | Analytical Competence Molecular Epidemiology Lab/ACME, Oswaldo Cruz Foundation, Ceara (FIOCRUZ CE) | Cleber Furtado Aksenen; Fabio Miyajima; Fernando Braga Stehling; Francisco Eder de Moura Lopes; Jamille Maria Mendes Bezerra; Joaquim César do Nascimento Sousa Junior; Pedro Miguel Carneiro Jeronimo; Suzana Porto Almeida e Lucas Delerino; Thais Ferreira de Oliveira; Thais de Oliveira Costa; Ticiane Cavalcante de Souza; Veridiana Pessoa Miyajima |
| EPI_ISL_1966986, EPI_ISL_1966987 | UNIDADE MISTA DE SAUDE DE ORIENTE | Instituto Butantan / ESALQ-USP (Piracicaba) | Antonio Jorge Martins; Bianca Cechetto Carlos. Mendelics: Bibiana Santos; Claudia Renata dos Santos Barros; Cíntia Bittar; David Schlesinger. Hemocentro Ribeirão Preto: Simone Kashima; Debora Botequio Moretti; Elaine Cristina Marqueze; Elaine Vieira dos Santos; Elisangela Chicaroni Mattos; Erika Freitas; Evandra Strazza Rodrigues; Felipe Allan da Silva da Costa; Flavia Aburjaile; Fábio Sossai Possebon; Guilherme Campos; Guilherme Targino Valente; Heidge Fukumasu. USP-Botucatu: Rejane Maria Tommasini Grotto; Helena Lage Ferreira; Instituto Butantan: Dimas Tadeu Covas; Jardelina de Souza Todao Bernardino; Jayme A. Souza-Neto; Jessika Cristina Chagas Lesbon; Jorge A. Petroli Marchesi; José Salvatore Leister Patané; João Paulo Kitajima; João Pessoa Araújo Jr.; Leila Sabrina Ullmann; Loyze Paola Oliveira de Lima; Luiz Aurelio de Campos Crispin. Centro de Genômica Funcional da ESALQ: Luiz Lehmann Coutinho; Luiz Carlos Junior de Alcantara; Lívia Sacchetto; Maisa C. Pereira Parra; Maria Carolina Elias; Marta Giovanetti; Marília Moraes; Maurício Lacerda Nogueira. Prefeitura de Sao Paulo: Melissa Palmieri.; Patricia Akemi Assato; Paula Rahal; Paulo Inacio da Costa; Rafael dos Santos Bezerra; Raquel de Lello Rocha Campos Cassano. NGS Soluções Genômicas: Pilar Drummond Sampaio Corrêa Mariani. FZEA-USP Pirassununga: Mirele Daiana Poleti; Raul Machado Neto; Ricardo Augusto Brassaloti; Ricardo Haddad; Rodrigo Tocantins Calado. FAMERP-SJRP: Cecília Artico Banho; Sandra Coccuzzo Sampaio; Svetoslav Nanev Slavov; Vagner Fonseca; Vincent Louis Viala |
| EPI_ISL_2209900 | UNIDADE MISTA DE SAUDE MOMBUCA | Instituto Butantan | Antonio Jorge Martins; Claudia Renata dos Santos Barros; David Schlesinger; Debora Botequio Moretti; Dimas Tadeu Covas; Elaine Cristina Marqueze; Elaine Vieira Santos; Evandra Strazza Rodrigues; Heidge Fukumasu; Jayme Augusto de Souza-Neto; José Salvatore Leister Patané; Luiz Alcantara; Luiz Lehmann Coutinho; Maria Carolina Elias; Maurício Lacerda Nogueira; Rafael dos Santos Bezerra; Raul Machado Neto; Rejane Maria Tommasini Grotto; Ricardo Haddad; Sandra Coccuzzo Sampaio Vessoni; Simone Kashima; Svetoslav Nanev Slavov; Vincent Louis Viala. |
| EPI_ISL_2209990, EPI_ISL_2344725 | UNIDADE MISTA DE SAUDE MOMBUCA | Instituto Butantan / FZEA-USP-Pirassununga | Antonio Jorge Martins; Claudia Renata dos Santos Barros; David Schlesinger; Debora Botequio Moretti; Dimas Tadeu Covas; Elaine Cristina Marqueze; Elaine Vieira Santos; Evandra Strazza Rodrigues; Heidge Fukumasu; Jayme Augusto de Souza-Neto; José Salvatore Leister Patané; Luiz Alcantara; Luiz Lehmann Coutinho; Maria Carolina Elias; Maurício Lacerda Nogueira; Rafael dos Santos Bezerra; Raul Machado Neto; Rejane Maria Tommasini Grotto; Ricardo Haddad; Sandra Coccuzzo Sampaio Vessoni; Simone Kashima; Svetoslav Nanev Slavov; Vincent Louis Viala |
| EPI_ISL_1795136, EPI_ISL_1795137, EPI_ISL_1795138, EPI_ISL_1795139, EPI_ISL_1795140, EPI_ISL_1795149, EPI_ISL_1795150, EPI_ISL_2345339, EPI_ISL_2345340, EPI_ISL_2345341, EPI_ISL_2345342, EPI_ISL_2345343, EPI_ISL_2345358, EPI_ISL_2345359 | | | |
| see above | UNIDADE MISTA DE VISTA ALEGRE DO ALTO VISTA ALEGRE DO ALTO | Instituto Butantan / ESALQ-Piracicaba | Antonio Jorge Martins; Bianca Cechetto Carlos. Mendelics: Bibiana Santos; Claudia Renata dos Santos Barros; David Schlesinger; David Schlesinger. Hemocentro Ribeirão Preto: Simone Kashima; Debora Botequio Moretti; Debora Botequio Moretti. Centro de Genômica Funcional da ESALQ: Luiz Lehmann Coutinho; Dimas Tadeu Covas; Elaine Cristina Marqueze; Elaine Vieira Santos; Elaine Vieira dos Santos; Elisangela Chicaroni Mattos; Erika Freitas; Evandra Strazza Rodrigues; Felipe Allan da Silva da Costa; Flavia Aburjaile; Guilherme Targino Valente; Heidge Fukumasu; Heidge Fukumasu. USP-Botucatu: Rejane Maria Tommasini Grotto; Instituto Butantan: Alexander Roberto Precioso; Jayme A. Souza-Neto; Jayme Augusto de Souza-Neto; Jessika Cristina Chagas Lesbon; José Salvatore Leister Patané; João Paulo Kitajima; Luiz Alcantara; Luiz Carlos Junior de Alcantara; Luiz Lehmann Coutinho; Maria Carolina Elias; Marta Giovanetti; Maurício Lacerda Nogueira; Patricia Akemi Assato; Rafael dos Santos Bezerra; Raquel de Lello Rocha Campos Cassano. NGS Soluções Genômicas: Pilar Drummond Sampaio Corrêa Mariani. FZEA-USP Pirassununga: Mirele Daiana Poleti; Raul Machado Neto; Rejane Maria Tommasini Grotto; Ricardo Augusto Brassaloti; Ricardo Haddad; Rodrigo Tocantins Calado.; Sandra Coccuzzo Sampaio; Sandra Coccuzzo Sampaio Vessoni; Simone Kashima; Svetoslav Nanev Slavov; Vagner Fonseca; Vincent Louis Viala |
| EPI_ISL_2209908 | UNIDADE MISTA OSVALDO P CAMARGO ELIAS FAUSTO | Instituto Butantan | Antonio Jorge Martins; Claudia Renata dos Santos Barros; David Schlesinger; Debora Botequio Moretti; Dimas Tadeu Covas; Elaine Cristina Marqueze; Elaine Vieira Santos; Evandra Strazza Rodrigues; Heidge Fukumasu; Jayme Augusto de Souza-Neto; José Salvatore Leister Patané; Luiz Alcantara; Luiz Lehmann Coutinho; Maria Carolina Elias; Maurício Lacerda Nogueira; Rafael dos Santos Bezerra; Raul Machado Neto; Rejane Maria Tommasini Grotto; Ricardo Haddad; Sandra Coccuzzo Sampaio Vessoni; Simone Kashima; Svetoslav Nanev Slavov; Vincent Louis Viala. |
| EPI_ISL_2344750 | UNIDADE MISTA OSVALDO P CAMARGO ELIAS FAUSTO | Instituto Butantan / ESALQ-Piracicaba | Antonio Jorge Martins; Claudia Renata dos Santos Barros; David Schlesinger; Debora Botequio Moretti; Dimas Tadeu Covas; Elaine Cristina Marqueze; Elaine Vieira Santos; Evandra Strazza Rodrigues; Heidge Fukumasu; Jayme Augusto de Souza-Neto; José Salvatore Leister Patané; Luiz Alcantara; Luiz Lehmann Coutinho; Maria Carolina Elias; Maurício Lacerda Nogueira; Rafael dos Santos Bezerra; Raul Machado Neto; Rejane Maria Tommasini Grotto; Ricardo Haddad; Sandra Coccuzzo Sampaio Vessoni; Simone Kashima; Svetoslav Nanev Slavov; Vincent Louis Viala |
| EPI_ISL_2209248, EPI_ISL_2344701, EPI_ISL_2362239 | UNIDADE MISTA OSVALDO P CAMARGO ELIAS FAUSTO | Instituto Butantan / FZEA-USP-Pirassununga | Antonio Jorge Martins; Claudia Renata dos Santos Barros; David Schlesinger; Debora Botequio Moretti; Dimas Tadeu Covas; Elaine Cristina Marqueze; Elaine Vieira Santos; Evandra Strazza Rodrigues; Heidge Fukumasu; Jayme Augusto de Souza-Neto; José Salvatore Leister Patané; Luiz Alcantara; Luiz Lehmann Coutinho; Maria Carolina Elias; Maurício Lacerda Nogueira; Rafael dos Santos Bezerra; Raul Machado Neto; Rejane Maria Tommasini Grotto; Ricardo Haddad; Sandra Coccuzzo Sampaio Vessoni; Simone Kashima; Svetoslav Nanev Slavov; Vincent Louis Viala |
| EPI_ISL_3102488, EPI_ISL_3912321, EPI_ISL_3912322, EPI_ISL_3912323, EPI_ISL_3912324 | UNIDADE PRONTO ATENDIMENTO AUTRAN NUNES | Analytical Competence Molecular Epidemiology Lab/ACME, Oswaldo Cruz Foundation, Ceara (FIOCRUZ CE) | Cleber Furtado Aksenen; Fabio Miyajima; Fernando Braga Stehling; Francisco Eder de Moura Lopes; Jamille Maria Mendes Bezerra; Joaquim Cesar do Nascimento Sousa Junior; Joaquim César do Nascimento Sousa Junior; Pedro Miguel Carneiro Jeronimo; Suzana Porto Almeida & Lucas Delerino on behalf of COVID-19 FIOCRUZ Genomic Network; Suzana Porto Almeida e Lucas Delerino; Thais Ferreira de Oliveira; Thais de Oliveira Costa; Ticiane Cavalcante de Souza; Veridiana Pessoa Miyajima |
| EPI_ISL_3912325, EPI_ISL_3912326, EPI_ISL_3912327, EPI_ISL_3912328 | UNIDADE PRONTO ATENDIMENTO CANINDEZINHO | Analytical Competence Molecular Epidemiology Lab/ACME, Oswaldo Cruz Foundation, Ceara (FIOCRUZ CE) | Cleber Furtado Aksenen; Fabio Miyajima; Fernando Braga Stehling; Francisco Eder de Moura Lopes; Jamille Maria Mendes Bezerra; Joaquim Cesar do Nascimento Sousa Junior; Pedro Miguel Carneiro Jeronimo; Suzana Porto Almeida & Lucas Delerino on behalf of COVID-19 FIOCRUZ Genomic Network; Thais Ferreira de Oliveira; Thais de Oliveira Costa; Ticiane Cavalcante de Souza; Veridiana Pessoa Miyajima |
| EPI_ISL_3102266 | UNIDADE PRONTO ATENDIMENTO CONJUNTO CEARA | Analytical Competence Molecular Epidemiology Lab/ACME, Oswaldo Cruz Foundation, Ceara (FIOCRUZ CE) | Cleber Furtado Aksenen; Fabio Miyajima; Fernando Braga Stehling; Francisco Eder de Moura Lopes; Jamille Maria Mendes Bezerra; Joaquim César do Nascimento Sousa Junior; Pedro Miguel Carneiro Jeronimo; Suzana Porto Almeida e Lucas Delerino; Thais Ferreira de Oliveira; Thais de Oliveira Costa; Ticiane Cavalcante de Souza; Veridiana Pessoa Miyajima |
| EPI_ISL_3102260, EPI_ISL_3102279, EPI_ISL_3102281, EPI_ISL_3102300, EPI_ISL_3102480, EPI_ISL_3912329, EPI_ISL_3912330 | | | |
| see above | UNIDADE PRONTO ATENDIMENTO JANGURUSSU | Analytical Competence Molecular Epidemiology Lab/ACME, Oswaldo Cruz Foundation, Ceara (FIOCRUZ CE) | Cleber Furtado Aksenen; Fabio Miyajima; Fernando Braga Stehling; Francisco Eder de Moura Lopes; Jamille Maria Mendes Bezerra; Joaquim Cesar do Nascimento Sousa Junior; Joaquim César do Nascimento Sousa Junior; Pedro Miguel Carneiro Jeronimo; Suzana Porto Almeida & Lucas Delerino on behalf of COVID-19 FIOCRUZ Genomic Network; Suzana Porto Almeida e Lucas Delerino; Thais Ferreira de Oliveira; Thais de Oliveira Costa; Ticiane Cavalcante de Souza; Veridiana Pessoa Miyajima |
| EPI_ISL_3102286 | UNIDADE PRONTO ATENDIMENTO JOSE WALTER | Analytical Competence Molecular Epidemiology Lab/ACME, Oswaldo Cruz Foundation, Ceara (FIOCRUZ CE) | Cleber Furtado Aksenen; Fabio Miyajima; Fernando Braga Stehling; Francisco Eder de Moura Lopes; Jamille Maria Mendes Bezerra; Joaquim César do Nascimento Sousa Junior; Pedro Miguel Carneiro Jeronimo; Suzana Porto Almeida e Lucas Delerino; Thais Ferreira de Oliveira; Thais de Oliveira Costa; Ticiane Cavalcante de Souza; Veridiana Pessoa Miyajima |
| EPI_ISL_3912331, EPI_ISL_3912332 | UNIDADE PRONTO ATENDIMENTO MESSEJANA | Analytical Competence Molecular Epidemiology Lab/ACME, Oswaldo Cruz Foundation, Ceara (FIOCRUZ CE) | Cleber Furtado Aksenen; Fabio Miyajima; Fernando Braga Stehling; Francisco Eder de Moura Lopes; Jamille Maria Mendes Bezerra; Joaquim Cesar do Nascimento Sousa Junior; Pedro Miguel Carneiro Jeronimo; Suzana Porto Almeida & Lucas Delerino on behalf of COVID-19 FIOCRUZ Genomic Network; Thais Ferreira de Oliveira; Thais de Oliveira Costa; Ticiane Cavalcante de Souza; Veridiana Pessoa Miyajima |
| EPI_ISL_3102476, EPI_ISL_3912333, EPI_ISL_3912334 | UNIDADE PRONTO ATENDIMENTO PRAIA DO FUTURO | Analytical Competence Molecular Epidemiology Lab/ACME, Oswaldo Cruz Foundation, Ceara (FIOCRUZ CE) | Cleber Furtado Aksenen; Fabio Miyajima; Fernando Braga Stehling; Francisco Eder de Moura Lopes; Jamille Maria Mendes Bezerra; Joaquim Cesar do Nascimento Sousa Junior; Joaquim César do Nascimento Sousa Junior; Pedro Miguel Carneiro Jeronimo; Suzana Porto Almeida & Lucas Delerino on behalf of COVID-19 FIOCRUZ Genomic Network; Suzana Porto Almeida e Lucas Delerino; Thais Ferreira de Oliveira; Thais de Oliveira Costa; Ticiane Cavalcante de Souza; Veridiana Pessoa Miyajima |
| EPI_ISL_3102284, EPI_ISL_3102303, EPI_ISL_3102315, | UNIDADE REFERENCIA COVID 19 | Analytical Competence Molecular Epidemiology Lab/ACME, Oswaldo | Cleber Furtado Aksenen; Fabio Miyajima; Fernando Braga Stehling; Francisco Eder de Moura Lopes; Jamille Maria Mendes Bezerra; Joaquim Cesar do Nascimento Sousa Junior; Joaquim César do Nascimento Sousa Junior; Pedro Miguel Carneiro Jeronimo; Suzana Porto Almeida & Lucas Delerino on behalf of COVID-19 FIOCRUZ Genomic Network; Suzana Porto Almeida e Lucas Delerino; Thais Ferreira de Oliveira; Thais de Oliveira Costa; Ticiane Cavalcante de Souza; Veridiana Pessoa Miyajima |

| | | | |
|---|---|---|---|
| EPI_ISL_3102483, EPI_ISL_3102484, EPI_ISL_3912381 | | Cruz Foundation, Ceara (FIOCRUZ CE) | |
| EPI_ISL_2801318 | UNIDADE REFERENCIA COVID 19 CRATO | Analytical Competence Molecular Epidemiology Lab/ACME, Oswaldo Cruz Foundation, Ceara (FIOCRUZ CE) | Cleber Furtado Aksenen e Suzana Porto Almeida; Fabio Miyajima; Fernando Braga Stehling; Francisco Eder de Moura Lopes; Jamille Maria Mendes Bezerra; Joaquim César do Nascimento Sousa Junior; Pedro Miguel Carneiro Jeronimo; Thais Ferreira de Oliveira; Thais de Oliveira Costa; Ticiane Cavalcante de Souza; Veridiana Pessoa Miyajima |
| EPI_ISL_2344580 | UNIDADE RESPIRATORIA NOVA HORTOLANDIA | Instituto Butantan / FZEA-USP-Pirassununga | Antonio Jorge Martins; Claudia Renata dos Santos Barros; David Schlesinger; Debora Botequio Moretti; Dimas Tadeu Covas; Elaine Cristina Marqueze; Elaine Vieira Santos; Evandra Strazza Rodrigues; Heidge Fukumasu; Jayme Augusto de Souza-Neto; José Salvatore Leister Patané; Luiz Alcantara; Luiz Lehmann Coutinho; Maria Carolina Elias; Maurício Lacerda Nogueira; Rafael dos Santos Bezerra; Raul Machado Neto; Rejane Maria Tommasini Grotto; Ricardo Haddad; Sandra Coccuzzo Sampaio Vessoni; Simone Kashima; Svetoslav Nanev Slavov; Vincent Louis Viala |
| EPI_ISL_1966166, EPI_ISL_1966603, EPI_ISL_1966606, EPI_ISL_1967289, EPI_ISL_1967291, EPI_ISL_1967292, EPI_ISL_1967293 | | | |
| see above | UNIDADE RESPIRATORIA NOVA HORTOLANDIA | Instituto Butantan / Mendelics | Antonio Jorge Martins; Bianca Cechetto Carlos. Mendelics: Bibiana Santos; Claudia Renata dos Santos Barros; Cíntia Bittar; David Schlesinger. Hemocentro Ribeirão Preto: Simone Kashima; Debora Botequio Moretti; Elaine Cristina Marqueze; Elaine Vieira dos Santos; Elisangela Chicaroni Mattos; Erika Freitas; Evandra Strazza Rodrigues; Felipe Allan da Silva da Costa; Flavia Aburjaile; Fábio Sossai Possebon; Guilherme Campos; Guilherme Targino Valente; Heidge Fukumasu. USP-Botucatu: Rejane Maria Tommasini Grotto; Helena Lage Ferreira; Instituto Butantan: Dimas Tadeu Covas; Jardelina de Souza Todao Bernardino; Jayme A. Souza-Neto; Jessika Cristina Chagas Lesbon; Jorge A. Petroli Marchesi; José Salvatore Leister Patané; João Paulo Kitajima; João Pessoa Araújo Jr.; Leila Sabrina Ullmann; Loyze Paola Oliveira de Lima; Luiz Aurelio de Campos Crispin. Centro de Genômica Funcional da ESALQ: Luiz Lehmann Coutinho; Luiz Carlos Junior de Alcantara; Lívia Sacchetto; Maisa C. Pereira Parra; Maria Carolina Elias; Marta Giovanetti; Marília Moraes; Maurício Lacerda Nogueira. Prefeitura de Sao Paulo: Melissa Palmieri.; Patricia Akemi Assato; Paula Rahal; Paulo Inacio da Costa; Rafael dos Santos Bezerra; Raquel de Lello Rocha Campos Cassano. NGS Soluções Genômicas: Pilar Drummond Sampaio Corrêa Mariani. FZEA-USP Pirassununga: Mirele Daiana Poleti; Raul Machado Neto; Ricardo Augusto Brassaloti; Ricardo Haddad; Rodrigo Tocantins Calado. FAMERP-SJRP: Cecília Artico Banho; Sandra Coccuzzo Sampaio; Svetoslav Nanev Slavov; Vagner Fonseca; Vincent Louis Viala |
| EPI_ISL_1966250 | UNIDADE SAUDE DA FAMILIA DRA ZILDA ARNS NEUMANN JALES | Instituto Butantan / Mendelics | Antonio Jorge Martins; Bianca Cechetto Carlos. Mendelics: Bibiana Santos; Claudia Renata dos Santos Barros; Cíntia Bittar; David Schlesinger. Hemocentro Ribeirão Preto: Simone Kashima; Debora Botequio Moretti; Elaine Cristina Marqueze; Elaine Vieira dos Santos; Elisangela Chicaroni Mattos; Erika Freitas; Evandra Strazza Rodrigues; Felipe Allan da Silva da Costa; Flavia Aburjaile; Fábio Sossai Possebon; Guilherme Campos; Guilherme Targino Valente; Heidge Fukumasu. USP-Botucatu: Rejane Maria Tommasini Grotto; Helena Lage Ferreira; Instituto Butantan: Dimas Tadeu Covas; Jardelina de Souza Todao Bernardino; Jayme A. Souza-Neto; Jessika Cristina Chagas Lesbon; Jorge A. Petroli Marchesi; José Salvatore Leister Patané; João Paulo Kitajima; João Pessoa Araújo Jr.; Leila Sabrina Ullmann; Loyze Paola Oliveira de Lima; Luiz Aurelio de Campos Crispin. Centro de Genômica Funcional da ESALQ: Luiz Lehmann Coutinho; Luiz Carlos Junior de Alcantara; Lívia Sacchetto; Maisa C. Pereira Parra; Maria Carolina Elias; Marta Giovanetti; Marília Moraes; Maurício Lacerda Nogueira. Prefeitura de Sao Paulo: Melissa Palmieri.; Patricia Akemi Assato; Paula Rahal; Paulo Inacio da Costa; Rafael dos Santos Bezerra; Raquel de Lello Rocha Campos Cassano. NGS Soluções Genômicas: Pilar Drummond Sampaio Corrêa Mariani. FZEA-USP Pirassununga: Mirele Daiana Poleti; Raul Machado Neto; Ricardo Augusto Brassaloti; Ricardo Haddad; Rodrigo Tocantins Calado. FAMERP-SJRP: Cecília Artico Banho; Sandra Coccuzzo Sampaio; Svetoslav Nanev Slavov; Vagner Fonseca; Vincent Louis Viala |
| EPI_ISL_1795220, EPI_ISL_2345450 | UNIDADE SENTINELA COVID19 | Instituto Butantan / ESALQ-Piracicaba | Antonio Jorge Martins; Bianca Cechetto Carlos. Mendelics: Bibiana Santos; Claudia Renata dos Santos Barros; David Schlesinger; David Schlesinger. Hemocentro Ribeirão Preto: Simone Kashima; Debora Botequio Moretti; Debora Botequio Moretti. Centro de Genômica Funcional da ESALQ: Luiz Lehmann Coutinho; Dimas Tadeu Covas; Elaine Cristina Marqueze; Elaine Vieira Santos; Elaine Vieira dos Santos; Elisangela Chicaroni Mattos; Erika Freitas; Evandra Strazza Rodrigues; Felipe Allan da Silva da Costa; Flavia Aburjaile; Guilherme Targino Valente; Heidge Fukumasu; Heidge Fukumasu. USP-Botucatu: Rejane Maria Tommasini Grotto; Instituto Butantan: Alexander Roberto Precioso; Jayme A. Souza-Neto; Jayme Augusto de Souza-Neto; Jessika Cristina Chagas Lesbon; José Salvatore Leister Patané; João Paulo Kitajima; Luiz Alcantara; Luiz Carlos Junior de Alcantara; Luiz Lehmann Coutinho; Maria Carolina Elias; Marta Giovanetti; Maurício Lacerda Nogueira; Patricia Akemi Assato; Rafael dos Santos Bezerra; Raquel de Lello Rocha Campos Cassano. NGS Soluções Genômicas: Pilar Drummond Sampaio Corrêa Mariani. FZEA-USP Pirassununga: Mirele Daiana Poleti; Raul Machado Neto; Rejane Maria Tommasini Grotto; Ricardo Augusto Brassaloti; Ricardo Haddad; Rodrigo Tocantins Calado.; Sandra Coccuzzo Sampaio; Sandra Coccuzzo Sampaio Vessoni; Simone Kashima; Svetoslav Nanev Slavov; Vagner Fonseca; Vincent Louis Viala |
| EPI_ISL_1966854, EPI_ISL_1966857 | UNIDADE SENTINELA COVID19 | Instituto Butantan / ESALQ-USP (Piracicaba) | Antonio Jorge Martins; Bianca Cechetto Carlos. Mendelics: Bibiana Santos; Claudia Renata dos Santos Barros; Cíntia Bittar; David Schlesinger. Hemocentro Ribeirão Preto: Simone Kashima; Debora Botequio Moretti; Elaine Cristina Marqueze; Elaine Vieira dos Santos; Elisangela Chicaroni Mattos; Erika Freitas; Evandra Strazza Rodrigues; Felipe Allan da Silva da Costa; Flavia Aburjaile; Fábio Sossai Possebon; Guilherme Campos; Guilherme Targino Valente; Heidge Fukumasu. USP-Botucatu: Rejane Maria Tommasini Grotto; Helena Lage Ferreira; Instituto Butantan: Dimas Tadeu Covas; Jardelina de Souza Todao Bernardino; Jayme A. Souza-Neto; Jessika Cristina Chagas Lesbon; Jorge A. Petroli Marchesi; José Salvatore Leister Patané; João Paulo Kitajima; João Pessoa Araújo Jr.; Leila Sabrina Ullmann; Loyze Paola Oliveira de Lima; Luiz Aurelio de Campos Crispin. Centro de Genômica Funcional da ESALQ: Luiz Lehmann Coutinho; Luiz Carlos Junior de Alcantara; Lívia Sacchetto; Maisa C. Pereira Parra; Maria Carolina Elias; Marta Giovanetti; Marília Moraes; Maurício Lacerda Nogueira. Prefeitura de Sao Paulo: Melissa Palmieri.; Patricia Akemi Assato; Paula Rahal; Paulo Inacio da Costa; Rafael dos Santos Bezerra; Raquel de Lello Rocha Campos Cassano. NGS Soluções Genômicas: Pilar Drummond Sampaio Corrêa Mariani. FZEA-USP Pirassununga: Mirele Daiana Poleti; Raul Machado Neto; Ricardo Augusto Brassaloti; Ricardo Haddad; Rodrigo Tocantins Calado. FAMERP-SJRP: Cecília Artico Banho; Sandra Coccuzzo Sampaio; Svetoslav Nanev Slavov; Vagner Fonseca; Vincent Louis Viala |
| EPI_ISL_3102294 | UNIDADE SENTINELA DE JUAZEIRO DO NORTE | Analytical Competence Molecular Epidemiology Lab/ACME, Oswaldo Cruz Foundation, Ceara (FIOCRUZ CE) | Cleber Furtado Aksenen; Fabio Miyajima; Fernando Braga Stehling; Francisco Eder de Moura Lopes; Jamille Maria Mendes Bezerra; Joaquim César do Nascimento Sousa Junior; Pedro Miguel Carneiro Jeronimo; Suzana Porto Almeida e Lucas Delerino; Thais Ferreira de Oliveira; Thais de Oliveira Costa; Ticiane Cavalcante de Souza; Veridiana Pessoa Miyajima |
| EPI_ISL_1966859, EPI_ISL_1966860 | UNIDADE SENTINELA VILA APARECIDA | Instituto Butantan / ESALQ-USP (Piracicaba) | Antonio Jorge Martins; Bianca Cechetto Carlos. Mendelics: Bibiana Santos; Claudia Renata dos Santos Barros; Cíntia Bittar; David Schlesinger. Hemocentro Ribeirão Preto: Simone Kashima; Debora Botequio Moretti; Elaine Cristina Marqueze; Elaine Vieira dos Santos; Elisangela Chicaroni Mattos; Erika Freitas; Evandra Strazza Rodrigues; Felipe Allan da Silva da Costa; Flavia Aburjaile; Fábio Sossai Possebon; Guilherme Campos; Guilherme Targino Valente; Heidge Fukumasu. USP-Botucatu: Rejane Maria Tommasini Grotto; Helena Lage Ferreira; Instituto Butantan: Dimas Tadeu Covas; Jardelina de Souza Todao Bernardino; Jayme A. Souza-Neto; Jessika Cristina Chagas Lesbon; Jorge A. Petroli Marchesi; José Salvatore Leister Patané; João Paulo Kitajima; João Pessoa Araújo Jr.; Leila Sabrina Ullmann; Loyze Paola Oliveira de Lima; Luiz Aurelio de Campos Crispin. Centro de Genômica Funcional da ESALQ: Luiz Lehmann Coutinho; Luiz Carlos Junior de Alcantara; Lívia Sacchetto; Maisa C. Pereira Parra; Maria Carolina Elias; Marta Giovanetti; Marília Moraes; Maurício Lacerda Nogueira. Prefeitura de Sao Paulo: Melissa Palmieri.; Patricia Akemi Assato; Paula Rahal; Paulo Inacio da Costa; Rafael dos Santos Bezerra; Raquel de Lello Rocha Campos Cassano. NGS Soluções Genômicas: Pilar Drummond Sampaio Corrêa Mariani. FZEA-USP Pirassununga: Mirele Daiana Poleti; Raul Machado Neto; Ricardo Augusto Brassaloti; Ricardo Haddad; Rodrigo Tocantins Calado. FAMERP-SJRP: Cecília Artico Banho; Sandra Coccuzzo Sampaio; Svetoslav Nanev Slavov; Vagner Fonseca; Vincent Louis Viala |
| EPI_ISL_2136324, EPI_ISL_2136329, EPI_ISL_2136334 | UNIGEM | Universidad Nacional de Colombia - Laboratorio Genómico One Health | Andres F. Cardona-Rios; Carlos Franco-Muñoz; Carolina Muñoz-Arango; Celeny Ortiz; Daniel O. Maldonado-Perez; Diego A. Álvarez-Díaz; Hector Alejandro Ruiz-Moreno; Idabely Betancur Ortiz; Jorge E. Osorio; Juan P. Hernandez-Ortiz; Karl A Ciuoderis; Katherine Laiton-Donato; Laura Silvana Perez; Lina M. Hurtado; Marcela Mercado-Reyes; Maria Angélica Maya; Maria Stella López; Rita Almanza Payares; Sandra Ines Cano; Simón Villegas Velásquez |
| EPI_ISL_1399629, EPI_ISL_1738859, EPI_ISL_1738922, EPI_ISL_1738949, EPI_ISL_1738952, EPI_ISL_1738962, EPI_ISL_1739269 | | | |
| see above | UNILABS | Instituto Nacional de Saude (INSA) | Borges et al |
| EPI_ISL_2157423 | UNIVERSIDADE FEDERAL DE VIÇOSA | Laboratory of Respiratory Viruses and Measles, Oswaldo Cruz Institute, FIOCRUZ | Alice Sampaio Rocha; Ana Carolina Mendonca; Anna Carolina Paixao; Elisa Cavalcante Pereira; Fernando Motta; Luciana Appolinario; Marilda Siqueira on behalf of the Fiocruz COVID-19 Genomic Surveillance Network; Paola Resende; Renata Serrano Lopes; Rubens Pasa; Taina Venas |
| EPI_ISL_2209401, EPI_ISL_2209404, EPI_ISL_2344754, EPI_ISL_2344764, EPI_ISL_2344768, EPI_ISL_2344770, EPI_ISL_2344773 | | | |
| see above | UPA 24 HORAS ANA JACINTA | Instituto Butantan / ESALQ-Piracicaba | Antonio Jorge Martins; Claudia Renata dos Santos Barros; David Schlesinger; Debora Botequio Moretti; Dimas Tadeu Covas; Elaine Cristina Marqueze; Elaine Vieira Santos; Evandra Strazza Rodrigues; Heidge Fukumasu; Jayme Augusto de Souza-Neto; José Salvatore Leister Patané; Luiz Alcantara; Luiz Lehmann Coutinho; Maria Carolina Elias; Maurício Lacerda Nogueira; Rafael dos Santos Bezerra; Raul Machado Neto; Rejane Maria Tommasini Grotto; Ricardo Haddad; Sandra Coccuzzo Sampaio Vessoni; Simone Kashima; Svetoslav Nanev Slavov; Vincent Louis Viala |
| EPI_ISL_2209379, EPI_ISL_2344736, EPI_ISL_2344758, EPI_ISL_2344762, EPI_ISL_2344766 | UPA 24 HORAS DR ALOISIO MUNIZ DE ANDRADE | Instituto Butantan / ESALQ-Piracicaba | Antonio Jorge Martins; Claudia Renata dos Santos Barros; David Schlesinger; Debora Botequio Moretti; Dimas Tadeu Covas; Elaine Cristina Marqueze; Elaine Vieira Santos; Evandra Strazza Rodrigues; Heidge Fukumasu; Jayme Augusto de Souza-Neto; José Salvatore Leister Patané; Luiz Alcantara; Luiz Lehmann Coutinho; Maria Carolina Elias; Maurício Lacerda Nogueira; Rafael dos Santos Bezerra; Raul Machado Neto; Rejane Maria Tommasini Grotto; Ricardo Haddad; Sandra Coccuzzo Sampaio Vessoni; Simone Kashima; Svetoslav Nanev Slavov; Vincent Louis Viala |
| EPI_ISL_3912383 | UPA 24 HORAS MARACANAU | Analytical Competence Molecular Epidemiology Lab/ACME, Oswaldo Cruz Foundation, Ceara (FIOCRUZ CE) | Cleber Furtado Aksenen; Fabio Miyajima; Fernando Braga Stehling; Francisco Eder de Moura Lopes; Jamille Maria Mendes Bezerra; Joaquim Cesar do Nascimento Sousa Junior; Pedro Miguel Carneiro Jeronimo; Suzana Porto Almeida & Lucas Delerino on behalf of COVID-19 FIOCRUZ Genomic Network; Thais Ferreira de Oliveira; Thais de Oliveira Costa; Ticiane Cavalcante de Souza; Veridiana Pessoa Miyajima |
| EPI_ISL_1966891, EPI_ISL_1966892, EPI_ISL_1966894 | UPA ATALAIA DRA RITA DE CASSIA SORIO | Instituto Butantan / ESALQ-USP (Piracicaba) | Antonio Jorge Martins; Bianca Cechetto Carlos. Mendelics: Bibiana Santos; Claudia Renata dos Santos Barros; Cíntia Bittar; David Schlesinger. Hemocentro Ribeirão Preto: Simone Kashima; Debora Botequio Moretti; Elaine Cristina Marqueze; Elaine Vieira dos Santos; Elisangela Chicaroni Mattos; Erika Freitas; Evandra Strazza Rodrigues; Felipe Allan da Silva da Costa; Flavia Aburjaile; Fábio Sossai Possebon; Guilherme Campos; Guilherme Targino Valente; Heidge Fukumasu. USP-Botucatu: Rejane Maria Tommasini Grotto; Helena Lage Ferreira; Instituto Butantan: Dimas Tadeu Covas; Jardelina de Souza Todao Bernardino; Jayme A. Souza-Neto; Jessika Cristina Chagas Lesbon; Jorge A. Petroli Marchesi; José Salvatore Leister Patané; João Paulo Kitajima; João Pessoa Araújo Jr.; Leila Sabrina Ullmann; Loyze Paola Oliveira de Lima; Luiz Aurelio de Campos Crispin. Centro de Genômica Funcional da ESALQ: Luiz Lehmann Coutinho; Luiz Carlos Junior de Alcantara; Lívia Sacchetto; Maisa C. Pereira Parra; Maria Carolina Elias; Marta Giovanetti; Marília Moraes; Maurício Lacerda Nogueira. Prefeitura de Sao Paulo: Melissa Palmieri.; Patricia Akemi Assato; Paula Rahal; Paulo Inacio da Costa; Rafael dos Santos Bezerra; Raquel de Lello Rocha Campos Cassano. NGS Soluções Genômicas: Pilar Drummond Sampaio Corrêa Mariani. FZEA-USP Pirassununga: Mirele Daiana Poleti; Raul Machado Neto; Ricardo Augusto Brassaloti; Ricardo Haddad; Rodrigo Tocantins Calado. FAMERP-SJRP: Cecília Artico Banho; Sandra Coccuzzo Sampaio; Svetoslav Nanev Slavov; Vagner Fonseca; Vincent Louis Viala |
| EPI_ISL_2614574, EPI_ISL_2614601 | UPA Akira Tada | Instituto Adolfo Lutz, Interdiciplinary Procedures Center, Strategic Laboratory | Caio Vinicius Dias Lopes; Claudia Regina Gonçalves; Claudio Tavares Sacchi; Erica Valessa Ramos Gomes; Karoline Rodrigues Campos; Leonardo Jose Tadeu de Araujo |
| EPI_ISL_1966595, EPI_ISL_1966596, EPI_ISL_1966601, EPI_ISL_1967218, EPI_ISL_1967222, EPI_ISL_1967225, EPI_ISL_1967226 | | | |
| see above | UPA DE BEBEDOURO | Instituto Butantan / Mendelics | Antonio Jorge Martins; Bianca Cechetto Carlos. Mendelics: Bibiana Santos; Claudia Renata dos Santos Barros; Cíntia Bittar; David Schlesinger. Hemocentro Ribeirão Preto: Simone Kashima; Debora Botequio Moretti; Elaine Cristina Marqueze; Elaine Vieira dos Santos; Elisangela Chicaroni Mattos; Erika Freitas; Evandra Strazza Rodrigues; Felipe Allan da Silva da Costa; Flavia Aburjaile; Fábio Sossai Possebon; Guilherme Campos; Guilherme Targino Valente; Heidge Fukumasu. USP-Botucatu: Rejane Maria Tommasini Grotto; Helena Lage Ferreira; Instituto Butantan: Dimas Tadeu Covas; Jardelina de Souza Todao Bernardino; Jayme A. Souza-Neto; Jessika Cristina Chagas Lesbon; Jorge A. Petroli Marchesi; José Salvatore Leister Patané; João Paulo Kitajima; João Pessoa Araújo Jr.; Leila Sabrina Ullmann; Loyze Paola Oliveira de Lima; Luiz Aurelio de Campos Crispin. Centro de Genômica Funcional da ESALQ: Luiz Lehmann Coutinho; Luiz Carlos Junior de Alcantara; Lívia Sacchetto; Maisa C. Pereira Parra; Maria Carolina Elias; Marta Giovanetti; Marília Moraes; Maurício Lacerda Nogueira. Prefeitura de Sao Paulo: Melissa Palmieri.; Patricia Akemi Assato; Paula Rahal; Paulo Inacio da Costa; Rafael dos Santos Bezerra; Raquel de Lello Rocha Campos Cassano. NGS Soluções Genômicas: Pilar Drummond Sampaio Corrêa Mariani. FZEA-USP Pirassununga: Mirele Daiana Poleti; Raul Machado Neto; Ricardo Augusto Brassaloti; Ricardo Haddad; Rodrigo Tocantins Calado. FAMERP-SJRP: Cecília Artico Banho; Sandra Coccuzzo Sampaio; Svetoslav Nanev Slavov; Vagner Fonseca; Vincent Louis Viala |
| EPI_ISL_1966436, EPI_ISL_2209040, EPI_ISL_2345725 | UPA DR AKIRA TADA | Instituto Butantan / Mendelics | Antonio Jorge Martins; Bianca Cechetto Carlos. Mendelics: Bibiana Santos; Claudia Renata dos Santos Barros; Cíntia Bittar; David Schlesinger; David Schlesinger. Hemocentro Ribeirão Preto: Simone Kashima; Debora Botequio Moretti; Dimas Tadeu Covas; Elaine Cristina Marqueze; Elaine Vieira Santos; Elaine Vieira dos Santos; Elisangela Chicaroni Mattos; Erika Freitas; Evandra Strazza Rodrigues; Felipe Allan da Silva da Costa; Flavia Aburjaile; Fábio Sossai Possebon; Guilherme Campos; Guilherme Targino Valente; Heidge Fukumasu; Heidge Fukumasu. USP-Botucatu: Rejane Maria Tommasini Grotto; Helena Lage Ferreira; Instituto Butantan: Dimas Tadeu Covas; Jardelina de Souza Todao Bernardino; Jayme A. Souza-Neto; Jayme Augusto de Souza-Neto; Jessika Cristina Chagas Lesbon; Jorge A. Petroli Marchesi; José Salvatore Leister Patané; João Paulo Kitajima; João Pessoa Araújo Jr.; Leila Sabrina Ullmann; Loyze Paola Oliveira de Lima; Luiz Alcantara; Luiz Aurelio de Campos Crispin. Centro de Genômica Funcional da ESALQ: Luiz Lehmann Coutinho; Luiz Carlos Junior de Alcantara; Luiz Lehmann Coutinho; Lívia Sacchetto; Maisa C. Pereira Parra; Maria Carolina Elias; Marta Giovanetti; Marília Moraes; Maurício Lacerda Nogueira; Maurício Lacerda Nogueira. Prefeitura de Sao Paulo: Melissa Palmieri.; Patricia Akemi Assato; Paula Rahal; Paulo Inacio da Costa; Rafael dos Santos Bezerra; Raquel de Lello Rocha Campos Cassano. NGS Soluções Genômicas: Pilar Drummond Sampaio Corrêa Mariani. FZEA-USP Pirassununga: Mirele Daiana Poleti; Raul Machado Neto; Rejane Maria Tommasini Grotto; Ricardo Augusto Brassaloti; Ricardo Haddad; Rodrigo Tocantins Calado. FAMERP-SJRP: Cecília Artico Banho; Sandra Coccuzzo Sampaio; Sandra Coccuzzo Sampaio Vessoni; Simone Kashima; Svetoslav Nanev Slavov; Vagner Fonseca; Vincent Louis Viala |
| EPI_ISL_2345996, EPI_ISL_2346000, EPI_ISL_2346008, EPI_ISL_2346009, EPI_ISL_2346011, EPI_ISL_2346019, EPI_ISL_2346020, EPI_ISL_2346029, EPI_ISL_2346038 | | | |
| see above | UPA DR FRANCO DA ROCHA | Instituto Butantan | Antonio Jorge Martins; Claudia Renata dos Santos Barros; David Schlesinger; Debora Botequio Moretti; Dimas Tadeu Covas; Elaine Cristina Marqueze; Elaine Vieira Santos; Evandra Strazza Rodrigues; Heidge Fukumasu; Jayme Augusto de Souza-Neto; José Salvatore Leister Patané; Luiz Alcantara; Luiz Lehmann Coutinho; Maria Carolina Elias; Maurício Lacerda Nogueira; Rafael dos Santos Bezerra; Raul Machado Neto; Rejane Maria Tommasini Grotto; Ricardo Haddad; Sandra Coccuzzo Sampaio Vessoni; Simone Kashima; Svetoslav Nanev Slavov; Vincent Louis Viala |
| EPI_ISL_2208904, EPI_ISL_2345716, EPI_ISL_2345812, EPI_ISL_2345862, EPI_ISL_2362256 | UPA DR FRANCO DA ROCHA | Instituto Butantan / Mendelics | Antonio Jorge Martins; Claudia Renata dos Santos Barros; David Schlesinger; Debora Botequio Moretti; Dimas Tadeu Covas; Elaine Cristina Marqueze; Elaine Vieira Santos; Evandra Strazza Rodrigues; Heidge Fukumasu; Jayme Augusto de Souza-Neto; José Salvatore Leister Patané; Luiz Alcantara; Luiz Lehmann Coutinho; Maria Carolina Elias; Maurício Lacerda Nogueira; Rafael dos Santos Bezerra; Raul Machado Neto; Rejane Maria Tommasini Grotto; Ricardo Haddad; Sandra Coccuzzo Sampaio Vessoni; Simone Kashima; Svetoslav Nanev Slavov; Vincent Louis Viala |
| EPI_ISL_1628373 | UPA De Bebedouro | Instituto Adolfo Lutz, Interdiciplinary Procedures Center, Strategic Laboratory | Caio Vinicius Dias Lopes; Claudia Regina Gonçalves; Claudio Tavares Sacchi; Erica Valessa Ramos Gomes; Karoline Rodrigues Campos; Katia Correa de Oliveira Santos; Leonardo Jose Tadeu de Araujo |

| | | | |
|---|---|---|---|
| EPI_ISL_1628369, EPI_ISL_1715140, EPI_ISL_1731576, EPI_ISL_1752639 | UPA Dr Luis Atilio Losi Viana Ribeirao Preto | Instituto Adolfo Lutz, Interdiciplinary Procedures Center, Strategic Laboratory | Caio Vinicius Dias Lopes; Claudia Regina Gonçalves; Claudio Tavares Sacchi; Erica Valessa Ramos Gomes; Karoline Rodrigues Campos; Katia Correa de Oliveira Santos; Leonardo Jose Tadeu de Araujo |
| EPI_ISL_3912384 | UPA I UNIDADE DE PRONTO ATENDIMENTO | Analytical Competence Molecular Epidemiology Lab/ACME, Oswaldo Cruz Foundation, Ceara (FIOCRUZ CE) | Cleber Furtado Aksenen; Fabio Miyajima; Fernando Braga Stehling; Francisco Eder de Moura Lopes; Jamille Maria Mendes Bezerra; Joaquim Cesar do Nascimento Sousa Junior; Pedro Miguel Carneiro Jeronimo; Suzana Porto Almeida & Lucas Delerino on behalf of COVID-19 FIOCRUZ Genomic Network; Thais Ferreira de Oliveira; Thais de Oliveira Costa; Ticiane Cavalcante de Souza; Veridiana Pessoa Miyajima |
| EPI_ISL_2171011 | UPA II JUNDIAI | Instituto Butantan / Mendelics | Antonio Jorge Martins; Bianca Cechetto Carlos. Mendelics: Bibiana Santos; Claudia Renata dos Santos Barros; Cíntia Bittar; David Schlesinger. Hemocentro Ribeirão Preto: Simone Kashima; Debora Botequio Moretti; Elaine Cristina Marqueze; Elaine Vieira dos Santos; Elisangela Chicaroni Mattos; Erika Freitas; Evandra Strazza Rodrigues; Felipe Allan da Silva da Costa; Flavia Aburjaile; Fábio Sossai Possebon; Guilherme Campos; Guilherme Targino Valente; Heidge Fukumasu. USP-Botucatu: Rejane Maria Tommasini Grotto; Helena Lage Ferreira; Instituto Butantan: Dimas Tadeu Covas; Jardelina de Souza Todao Bernardino; Jayme A. Souza-Neto; Jessika Cristina Chagas Lesbon; Jorge A. Petroli Marchesi; José Salvatore Leister Patané; João Paulo Kitajima; João Pessoa Araújo Jr.; Leila Sabrina Ullmann; Loyze Paola Oliveira de Lima; Luiz Aurelio de Campos Crispin. Centro de Genômica Funcional da ESALQ: Luiz Lehmann Coutinho; Luiz Carlos Junior de Alcantara; Lívia Sacchetto; Maisa C. Pereira Parra; Maria Carolina Elias; Marta Giovanetti; Marília Moraes; Maurício Lacerda Nogueira. Prefeitura de Sao Paulo: Melissa Palmieri.; Patricia Akemi Assato; Paula Rahal; Paulo Inacio da Costa; Rafael dos Santos Bezerra; Raquel de Lello Rocha Campos Cassano. NGS Soluções Genômicas: Pilar Drummond Sampaio Corrêa Mariani. FZEA-USP Pirassununga: Mirele Daiana Poleti; Raul Machado Neto; Ricardo Augusto Brassaloti; Ricardo Haddad; Rodrigo Tocantins Calado. FAMERP-SJRP: Cecília Artico Banho; Sandra Coccuzzo Sampaio; Svetoslav Nanev Slavov; Vagner Fonseca; Vincent Louis Viala |
| EPI_ISL_1966729 | UPA JOSE CAMPOS BARRETO ZEQUINHA BARRETO | Instituto Butantan | Antonio Jorge Martins; Bianca Cechetto Carlos. Mendelics: Bibiana Santos; Claudia Renata dos Santos Barros; Cíntia Bittar; David Schlesinger. Hemocentro Ribeirão Preto: Simone Kashima; Debora Botequio Moretti; Elaine Cristina Marqueze; Elaine Vieira dos Santos; Elisangela Chicaroni Mattos; Erika Freitas; Evandra Strazza Rodrigues; Felipe Allan da Silva da Costa; Flavia Aburjaile; Fábio Sossai Possebon; Guilherme Campos; Guilherme Targino Valente; Heidge Fukumasu. USP-Botucatu: Rejane Maria Tommasini Grotto; Helena Lage Ferreira; Instituto Butantan: Dimas Tadeu Covas; Jardelina de Souza Todao Bernardino; Jayme A. Souza-Neto; Jessika Cristina Chagas Lesbon; Jorge A. Petroli Marchesi; José Salvatore Leister Patané; João Paulo Kitajima; João Pessoa Araújo Jr.; Leila Sabrina Ullmann; Loyze Paola Oliveira de Lima; Luiz Aurelio de Campos Crispin. Centro de Genômica Funcional da ESALQ: Luiz Lehmann Coutinho; Luiz Carlos Junior de Alcantara; Lívia Sacchetto; Maisa C. Pereira Parra; Maria Carolina Elias; Marta Giovanetti; Marília Moraes; Maurício Lacerda Nogueira. Prefeitura de Sao Paulo: Melissa Palmieri.; Patricia Akemi Assato; Paula Rahal; Paulo Inacio da Costa; Rafael dos Santos Bezerra; Raquel de Lello Rocha Campos Cassano. NGS Soluções Genômicas: Pilar Drummond Sampaio Corrêa Mariani. FZEA-USP Pirassununga: Mirele Daiana Poleti; Raul Machado Neto; Ricardo Augusto Brassaloti; Ricardo Haddad; Rodrigo Tocantins Calado. FAMERP-SJRP: Cecília Artico Banho; Sandra Coccuzzo Sampaio; Svetoslav Nanev Slavov; Vagner Fonseca; Vincent Louis Viala |
| EPI_ISL_2344576 | UPA UNIDADE DE PRONTO ATENDIMENTO 24 HORAS BOM JESUS | Instituto Butantan / FZEA-USP-Pirassununga | Antonio Jorge Martins; Claudia Renata dos Santos Barros; David Schlesinger; Debora Botequio Moretti; Dimas Tadeu Covas; Elaine Cristina Marqueze; Elaine Vieira Santos; Evandra Strazza Rodrigues; Heidge Fukumasu; Jayme Augusto de Souza-Neto; José Salvatore Leister Patané; Luiz Alcantara; Luiz Lehmann Coutinho; Maria Carolina Elias; Maurício Lacerda Nogueira; Rafael dos Santos Bezerra; Raul Machado Neto; Rejane Maria Tommasini Grotto; Ricardo Haddad; Sandra Coccuzzo Sampaio Vessoni; Simone Kashima; Svetoslav Nanev Slavov; Vincent Louis Viala |
| EPI_ISL_1966650, EPI_ISL_1966669 | UPA UNIDADE DE PRONTO ATENDIMENTO 24 HORAS BOM JESUS | Instituto Butantan / Mendelics | Antonio Jorge Martins; Bianca Cechetto Carlos. Mendelics: Bibiana Santos; Claudia Renata dos Santos Barros; Cíntia Bittar; David Schlesinger. Hemocentro Ribeirão Preto: Simone Kashima; Debora Botequio Moretti; Elaine Cristina Marqueze; Elaine Vieira dos Santos; Elisangela Chicaroni Mattos; Erika Freitas; Evandra Strazza Rodrigues; Felipe Allan da Silva da Costa; Flavia Aburjaile; Fábio Sossai Possebon; Guilherme Campos; Guilherme Targino Valente; Heidge Fukumasu. USP-Botucatu: Rejane Maria Tommasini Grotto; Helena Lage Ferreira; Instituto Butantan: Dimas Tadeu Covas; Jardelina de Souza Todao Bernardino; Jayme A. Souza-Neto; Jessika Cristina Chagas Lesbon; Jorge A. Petroli Marchesi; José Salvatore Leister Patané; João Paulo Kitajima; João Pessoa Araújo Jr.; Leila Sabrina Ullmann; Loyze Paola Oliveira de Lima; Luiz Aurelio de Campos Crispin. Centro de Genômica Funcional da ESALQ: Luiz Lehmann Coutinho; Luiz Carlos Junior de Alcantara; Lívia Sacchetto; Maisa C. Pereira Parra; Maria Carolina Elias; Marta Giovanetti; Marília Moraes; Maurício Lacerda Nogueira. Prefeitura de Sao Paulo: Melissa Palmieri.; Patricia Akemi Assato; Paula Rahal; Paulo Inacio da Costa; Rafael dos Santos Bezerra; Raquel de Lello Rocha Campos Cassano. NGS Soluções Genômicas: Pilar Drummond Sampaio Corrêa Mariani. FZEA-USP Pirassununga: Mirele Daiana Poleti; Raul Machado Neto; Ricardo Augusto Brassaloti; Ricardo Haddad; Rodrigo Tocantins Calado. FAMERP-SJRP: Cecília Artico Banho; Sandra Coccuzzo Sampaio; Svetoslav Nanev Slavov; Vagner Fonseca; Vincent Louis Viala |
| EPI_ISL_3912385 | UPA UNIDADE DE PRONTO ATENDIMENTO BOM JARDIM | Analytical Competence Molecular Epidemiology Lab/ACME, Oswaldo Cruz Foundation, Ceara (FIOCRUZ CE) | Cleber Furtado Aksenen; Fabio Miyajima; Fernando Braga Stehling; Francisco Eder de Moura Lopes; Jamille Maria Mendes Bezerra; Joaquim Cesar do Nascimento Sousa Junior; Pedro Miguel Carneiro Jeronimo; Suzana Porto Almeida & Lucas Delerino on behalf of COVID-19 FIOCRUZ Genomic Network; Thais Ferreira de Oliveira; Thais de Oliveira Costa; Ticiane Cavalcante de Souza; Veridiana Pessoa Miyajima |
| EPI_ISL_3102307 | UPA UNIDADE DE PRONTO ATENDIMENTO EDSON QUEIROZ | Analytical Competence Molecular Epidemiology Lab/ACME, Oswaldo Cruz Foundation, Ceara (FIOCRUZ CE) | Cleber Furtado Aksenen; Fabio Miyajima; Fernando Braga Stehling; Francisco Eder de Moura Lopes; Jamille Maria Mendes Bezerra; Joaquim César do Nascimento Sousa Junior; Pedro Miguel Carneiro Jeronimo; Suzana Porto Almeida e Lucas Delerino; Thais Ferreira de Oliveira; Thais de Oliveira Costa; Ticiane Cavalcante de Souza; Veridiana Pessoa Miyajima |
| EPI_ISL_1303547, EPI_ISL_1358318, EPI_ISL_1358321, EPI_ISL_2614587, EPI_ISL_2614588 | UPA Vila Santa Catarina | Instituto Adolfo Lutz, Interdiciplinary Procedures Center, Strategic Laboratory | Caio Vinicius Dias Lopes; Claudia Regina Gonçalves; Claudio Tavares Sacchi; Erica Valessa Ramos Gomes; Karoline Rodrigues Campos; Leonardo Jose Tadeu de Araujo |
| EPI_ISL_1715145 | UPA de Bebedouro | Instituto Adolfo Lutz, Interdiciplinary Procedures Center, Strategic Laboratory | Caio Vinicius Dias Lopes; Claudia Regina Gonçalves; Claudio Tavares Sacchi; Erica Valessa Ramos Gomes; Karoline Rodrigues Campos; Katia Correa de Oliveira Santos; Leonardo Jose Tadeu de Araujo |
| EPI_ISL_2614567 | UPA de Jandira | Instituto Adolfo Lutz, Interdiciplinary Procedures Center, Strategic Laboratory | Caio Vinicius Dias Lopes; Claudia Regina Gonçalves; Claudio Tavares Sacchi; Erica Valessa Ramos Gomes; Karoline Rodrigues Campos; Leonardo Jose Tadeu de Araujo |
| EPI_ISL_1855230 | URMC LABS | Wadsworth Center, New York State Department of Health | Alexis Russell; Catharine Prussing; Daryl M. Lamson; Erasmus Schneider; Erica Lasek-Nesselquist; John Kelly; Jonathan Plitnick; Kirsten St. George; Matthew Shudt; Melissa A Leisner; Navjot Singh |
| EPI_ISL_1711128, EPI_ISL_2402238 | US Air Force School of Aerospace Medicine | US Air Force School of Aerospace Medicine | Amanda Javorina; Anthony Fries; Carol Garrett; Clarise Starr; Elizabeth Macias; Jennifer Meyer; Sarah Purves; William Buggele; William Gruner |
| EPI_ISL_1966542 | USF DR APARECIDO RODRIGUES MOUCO SCRPARDO | Instituto Butantan / FZEA-USP (Pirassununga) | Antonio Jorge Martins; Bianca Cechetto Carlos. Mendelics: Bibiana Santos; Claudia Renata dos Santos Barros; Cíntia Bittar; David Schlesinger. Hemocentro Ribeirão Preto: Simone Kashima; Debora Botequio Moretti; Elaine Cristina Marqueze; Elaine Vieira dos Santos; Elisangela Chicaroni Mattos; Erika Freitas; Evandra Strazza Rodrigues; Felipe Allan da Silva da Costa; Flavia Aburjaile; Fábio Sossai Possebon; Guilherme Campos; Guilherme Targino Valente; Heidge Fukumasu. USP-Botucatu: Rejane Maria Tommasini Grotto; Helena Lage Ferreira; Instituto Butantan: Dimas Tadeu Covas; Jardelina de Souza Todao Bernardino; Jayme A. Souza-Neto; Jessika Cristina Chagas Lesbon; Jorge A. Petroli Marchesi; José Salvatore Leister Patané; João Paulo Kitajima; João Pessoa Araújo Jr.; Leila Sabrina Ullmann; Loyze Paola Oliveira de Lima; Luiz Aurelio de Campos Crispin. Centro de Genômica Funcional da ESALQ: Luiz Lehmann Coutinho; Luiz Carlos Junior de Alcantara; Lívia Sacchetto; Maisa C. Pereira Parra; Maria Carolina Elias; Marta Giovanetti; Marília Moraes; Maurício Lacerda Nogueira. Prefeitura de Sao Paulo: Melissa Palmieri.; Patricia Akemi Assato; Paula Rahal; Paulo Inacio da Costa; Rafael dos Santos Bezerra; Raquel de Lello Rocha Campos Cassano. NGS Soluções Genômicas: Pilar Drummond Sampaio Corrêa Mariani. FZEA-USP Pirassununga: Mirele Daiana Poleti; Raul Machado Neto; Ricardo Augusto Brassaloti; Ricardo Haddad; Rodrigo Tocantins Calado. FAMERP-SJRP: Cecília Artico Banho; Sandra Coccuzzo Sampaio; Svetoslav Nanev Slavov; Vagner Fonseca; Vincent Louis Viala |
| EPI_ISL_1966908 | USF I DE GUAIMBE | Instituto Butantan / ESALQ-USP (Piracicaba) | Antonio Jorge Martins; Bianca Cechetto Carlos. Mendelics: Bibiana Santos; Claudia Renata dos Santos Barros; Cíntia Bittar; David Schlesinger. Hemocentro Ribeirão Preto: Simone Kashima; Debora Botequio Moretti; Elaine Cristina Marqueze; Elaine Vieira dos Santos; Elisangela Chicaroni Mattos; Erika Freitas; Evandra Strazza Rodrigues; Felipe Allan da Silva da Costa; Flavia Aburjaile; Fábio Sossai Possebon; Guilherme Campos; Guilherme Targino Valente; Heidge Fukumasu. USP-Botucatu: Rejane Maria Tommasini Grotto; Helena Lage Ferreira; Instituto Butantan: Dimas Tadeu Covas; Jardelina de Souza Todao Bernardino; Jayme A. Souza-Neto; Jessika Cristina Chagas Lesbon; Jorge A. Petroli Marchesi; José Salvatore Leister Patané; João Paulo Kitajima; João Pessoa Araújo Jr.; Leila Sabrina Ullmann; Loyze Paola Oliveira de Lima; Luiz Aurelio de Campos Crispin. Centro de Genômica Funcional da ESALQ: Luiz Lehmann Coutinho; Luiz Carlos Junior de Alcantara; Lívia Sacchetto; Maisa C. Pereira Parra; Maria Carolina Elias; Marta Giovanetti; Marília Moraes; Maurício Lacerda Nogueira. Prefeitura de Sao Paulo: Melissa Palmieri.; Patricia Akemi Assato; Paula Rahal; Paulo Inacio da Costa; Rafael dos Santos Bezerra; Raquel de Lello Rocha Campos Cassano. NGS Soluções Genômicas: Pilar Drummond Sampaio Corrêa Mariani. FZEA-USP Pirassununga: Mirele Daiana Poleti; Raul Machado Neto; Ricardo Augusto Brassaloti; Ricardo Haddad; Rodrigo Tocantins Calado. FAMERP-SJRP: Cecília Artico Banho; Sandra Coccuzzo Sampaio; Svetoslav Nanev Slavov; Vagner Fonseca; Vincent Louis Viala |
| EPI_ISL_2344763 | USF II ALADIA ARAN RODRIGUES | Instituto Butantan / ESALQ-Piracicaba | Antonio Jorge Martins; Claudia Renata dos Santos Barros; David Schlesinger; Debora Botequio Moretti; Dimas Tadeu Covas; Elaine Cristina Marqueze; Elaine Vieira Santos; Evandra Strazza Rodrigues; Heidge Fukumasu; Jayme Augusto de Souza-Neto; José Salvatore Leister Patané; Luiz Alcantara; Luiz Lehmann Coutinho; Maria Carolina Elias; Maurício Lacerda Nogueira; Rafael dos Santos Bezerra; Raul Machado Neto; Rejane Maria Tommasini Grotto; Ricardo Haddad; Sandra Coccuzzo Sampaio Vessoni; Simone Kashima; Svetoslav Nanev Slavov; Vincent Louis Viala |
| EPI_ISL_2345959 | USF JARDIM DO LAGO ARTUR NOGUEIRA | Instituto Butantan | Antonio Jorge Martins; Claudia Renata dos Santos Barros; David Schlesinger; Debora Botequio Moretti; Dimas Tadeu Covas; Elaine Cristina Marqueze; Elaine Vieira Santos; Evandra Strazza Rodrigues; Heidge Fukumasu; Jayme Augusto de Souza-Neto; José Salvatore Leister Patané; Luiz Alcantara; Luiz Lehmann Coutinho; Maria Carolina Elias; Maurício Lacerda Nogueira; Rafael dos Santos Bezerra; Raul Machado Neto; Rejane Maria Tommasini Grotto; Ricardo Haddad; Sandra Coccuzzo Sampaio Vessoni; Simone Kashima; Svetoslav Nanev Slavov; Vincent Louis Viala |
| EPI_ISL_2210010 | USF PAULISTA FERNANDOPOLIS ANTONIO PIVATO | Instituto Butantan | Antonio Jorge Martins; Claudia Renata dos Santos Barros; David Schlesinger; Debora Botequio Moretti; Dimas Tadeu Covas; Elaine Cristina Marqueze; Elaine Vieira Santos; Evandra Strazza Rodrigues; Heidge Fukumasu; Jayme Augusto de Souza-Neto; José Salvatore Leister Patané; Luiz Alcantara; Luiz Lehmann Coutinho; Maria Carolina Elias; Maurício Lacerda Nogueira; Rafael dos Santos Bezerra; Raul Machado Neto; Rejane Maria Tommasini Grotto; Ricardo Haddad; Sandra Coccuzzo Sampaio Vessoni; Simone Kashima; Svetoslav Nanev Slavov; Vincent Louis Viala. |
| EPI_ISL_1966526, EPI_ISL_1966527, EPI_ISL_1966528, EPI_ISL_1966529 | USF PAULISTA FERNANDOPOLIS ANTONIO PIVATO | Instituto Butantan / FZEA-USP (Pirassununga) | Antonio Jorge Martins; Bianca Cechetto Carlos. Mendelics: Bibiana Santos; Claudia Renata dos Santos Barros; Cíntia Bittar; David Schlesinger. Hemocentro Ribeirão Preto: Simone Kashima; Debora Botequio Moretti; Elaine Cristina Marqueze; Elaine Vieira dos Santos; Elisangela Chicaroni Mattos; Erika Freitas; Evandra Strazza Rodrigues; Felipe Allan da Silva da Costa; Flavia Aburjaile; Fábio Sossai Possebon; Guilherme Campos; Guilherme Targino Valente; Heidge Fukumasu. USP-Botucatu: Rejane Maria Tommasini Grotto; Helena Lage Ferreira; Instituto Butantan: Dimas Tadeu Covas; Jardelina de Souza Todao Bernardino; Jayme A. Souza-Neto; Jessika Cristina Chagas Lesbon; Jorge A. Petroli Marchesi; José Salvatore Leister Patané; João Paulo Kitajima; João Pessoa Araújo Jr.; Leila Sabrina Ullmann; Loyze Paola Oliveira de Lima; Luiz Aurelio de Campos Crispin. Centro de Genômica Funcional da ESALQ: Luiz Lehmann Coutinho; Luiz Carlos Junior de Alcantara; Lívia Sacchetto; Maisa C. Pereira Parra; Maria Carolina Elias; Marta Giovanetti; Marília Moraes; Maurício Lacerda Nogueira. Prefeitura de Sao Paulo: Melissa Palmieri.; Patricia Akemi Assato; Paula Rahal; Paulo Inacio da Costa; Rafael dos Santos Bezerra; Raquel de Lello Rocha Campos Cassano. NGS Soluções Genômicas: Pilar Drummond Sampaio Corrêa Mariani. FZEA-USP Pirassununga: Mirele Daiana Poleti; Raul Machado Neto; Ricardo Augusto Brassaloti; Ricardo Haddad; Rodrigo Tocantins Calado. FAMERP-SJRP: Cecília Artico Banho; Sandra Coccuzzo Sampaio; Svetoslav Nanev Slavov; Vagner Fonseca; Vincent Louis Viala |
| EPI_ISL_2344540 | USF PAULISTA FERNANDOPOLIS ANTONIO PIVATO | Instituto Butantan / UNESP-Botucatu | Antonio Jorge Martins; Claudia Renata dos Santos Barros; David Schlesinger; Debora Botequio Moretti; Dimas Tadeu Covas; Elaine Cristina Marqueze; Elaine Vieira Santos; Evandra Strazza Rodrigues; Heidge Fukumasu; Jayme Augusto de Souza-Neto; José Salvatore Leister Patané; Luiz Alcantara; Luiz Lehmann Coutinho; Maria Carolina Elias; Maurício Lacerda Nogueira; Rafael dos Santos Bezerra; Raul Machado Neto; Rejane Maria Tommasini Grotto; Ricardo Haddad; Sandra Coccuzzo Sampaio Vessoni; Simone Kashima; Svetoslav Nanev Slavov; Vincent Louis Viala |
| EPI_ISL_2346046 | USF PEQUENO CORACAO | Instituto Butantan | Antonio Jorge Martins; Claudia Renata dos Santos Barros; David Schlesinger; Debora Botequio Moretti; Dimas Tadeu Covas; Elaine Cristina Marqueze; Elaine Vieira Santos; Evandra Strazza Rodrigues; Heidge Fukumasu; Jayme Augusto de Souza-Neto; José Salvatore Leister Patané; Luiz Alcantara; Luiz Lehmann Coutinho; Maria Carolina Elias; Maurício Lacerda Nogueira; Rafael dos Santos Bezerra; Raul Machado Neto; Rejane Maria Tommasini Grotto; Ricardo Haddad; Sandra Coccuzzo Sampaio Vessoni; Simone Kashima; Svetoslav Nanev Slavov; Vincent Louis Viala |
| EPI_ISL_2345999 | USF PQ DOS TRABALHADORES ARTUR NOGUEIRA | Instituto Butantan | Antonio Jorge Martins; Claudia Renata dos Santos Barros; David Schlesinger; Debora Botequio Moretti; Dimas Tadeu Covas; Elaine Cristina Marqueze; Elaine Vieira Santos; Evandra Strazza Rodrigues; Heidge Fukumasu; Jayme Augusto de Souza-Neto; José Salvatore Leister Patané; Luiz Alcantara; Luiz Lehmann Coutinho; Maria Carolina Elias; Maurício Lacerda Nogueira; Rafael dos Santos Bezerra; Raul Machado Neto; Rejane Maria Tommasini Grotto; Ricardo Haddad; Sandra Coccuzzo Sampaio Vessoni; Simone Kashima; Svetoslav Nanev Slavov; Vincent Louis Viala |
| EPI_ISL_1795391, EPI_ISL_2345528 | USF ROSA CRUZ | Instituto Butantan / ESALQ-Piracicaba | Antonio Jorge Martins; Bianca Cechetto Carlos. Mendelics: Bibiana Santos; Claudia Renata dos Santos Barros; David Schlesinger; David Schlesinger. Hemocentro Ribeirão Preto: Simone Kashima; Debora Botequio Moretti; Debora Botequio Moretti. Centro de Genômica Funcional da ESALQ: Luiz Lehmann Coutinho; Dimas Tadeu Covas; Elaine Cristina Marqueze; Elaine Vieira Santos; Elaine Vieira dos Santos; Elisangela Chicaroni Mattos; Erika Freitas; Evandra Strazza Rodrigues; Felipe Allan da Silva da Costa; Flavia Aburjaile; Guilherme Targino Valente; Heidge Fukumasu; Heidge Fukumasu. USP-Botucatu: Rejane Maria Tommasini Grotto; Instituto Butantan: Alexander Roberto Precioso; Jayme A. Souza-Neto; Jayme Augusto de Souza-Neto; Jessika Cristina Chagas Lesbon; José Salvatore Leister Patané; João Paulo Kitajima; Luiz Alcantara; Luiz Carlos Junior de Alcantara; Luiz Lehmann Coutinho; Maria Carolina Elias; Marta Giovanetti; Maurício Lacerda Nogueira; Patricia Akemi Assato; Rafael dos Santos Bezerra; Raquel de Lello Rocha Campos Cassano. NGS Soluções Genômicas: Pilar Drummond Sampaio Corrêa Mariani. FZEA-USP Pirassununga: Mirele Daiana Poleti; Raul Machado Neto; Rejane Maria Tommasini Grotto; Ricardo Augusto Brassaloti; Ricardo Haddad; Rodrigo Tocantins Calado.; Sandra Coccuzzo Sampaio; Sandra Coccuzzo Sampaio Vessoni; Simone Kashima; Svetoslav Nanev Slavov; Vagner Fonseca; Vincent Louis Viala |
| EPI_ISL_2345979 | USF SACILOTTO ARTUR NOGUEIRA | Instituto Butantan | Antonio Jorge Martins; Claudia Renata dos Santos Barros; David Schlesinger; Debora Botequio Moretti; Dimas Tadeu Covas; Elaine Cristina Marqueze; Elaine Vieira Santos; Evandra Strazza Rodrigues; Heidge Fukumasu; Jayme Augusto de Souza-Neto; José Salvatore Leister Patané; Luiz Alcantara; Luiz Lehmann Coutinho; Maria Carolina Elias; Maurício Lacerda Nogueira; Rafael dos Santos Bezerra; Raul Machado Neto; Rejane Maria Tommasini Grotto; Ricardo Haddad; Sandra Coccuzzo Sampaio Vessoni; Simone Kashima; Svetoslav Nanev Slavov; Vincent Louis Viala |
| EPI_ISL_1795175, EPI_ISL_1795189, EPI_ISL_1795192, EPI_ISL_1795193, EPI_ISL_1795384, EPI_ISL_2345392, EPI_ISL_2345406, EPI_ISL_2345410, EPI_ISL_2345413, EPI_ISL_2345415 | | | |
| see above | USF SALERNO | Instituto Butantan / ESALQ-Piracicaba | Antonio Jorge Martins; Bianca Cechetto Carlos. Mendelics: Bibiana Santos; Claudia Renata dos Santos Barros; David Schlesinger; David Schlesinger. Hemocentro Ribeirão Preto: Simone Kashima; Debora Botequio Moretti; Debora Botequio Moretti. Centro de Genômica Funcional da ESALQ: Luiz Lehmann Coutinho; Dimas Tadeu Covas; Elaine Cristina Marqueze; Elaine Vieira Santos; Elaine Vieira dos Santos; Elisangela Chicaroni Mattos; Erika Freitas; Evandra Strazza Rodrigues; Felipe Allan da Silva da Costa; Flavia Aburjaile; Guilherme Targino Valente; Heidge Fukumasu; Heidge Fukumasu. USP-Botucatu: Rejane Maria Tommasini Grotto; Instituto Butantan: Alexander Roberto Precioso; Jayme A. Souza-Neto; Jayme Augusto de Souza-Neto; Jessika Cristina Chagas Lesbon; José Salvatore Leister Patané; João Paulo Kitajima; Luiz Alcantara; Luiz Carlos Junior de Alcantara; Luiz Lehmann Coutinho; Maria Carolina Elias; Marta Giovanetti; Maurício Lacerda Nogueira; Patricia Akemi Assato; Rafael dos Santos Bezerra; Raquel de Lello Rocha Campos Cassano. NGS Soluções Genômicas: Pilar Drummond Sampaio Corrêa Mariani. FZEA-USP Pirassununga: Mirele Daiana Poleti; Raul Machado Neto; Rejane Maria Tommasini Grotto; Ricardo Augusto Brassaloti; Ricardo Haddad; Rodrigo Tocantins Calado.; Sandra Coccuzzo Sampaio; Sandra Coccuzzo Sampaio Vessoni; Simone Kashima; Svetoslav Nanev Slavov; Vagner Fonseca; Vincent Louis Viala |

| | | | |
|---|---|---|---|
| EPI_ISL_1678288, EPI_ISL_3354276, EPI_ISL_3354285 | UT-Unified State Labs: Public Health Utah DOH | Centers for Disease Control and Prevention Division of Viral Diseases, Pathogen Discovery | Alex Burgin; Alison Laufer Halpin; Ben L. Rambo-Martin; Ben Rambo-Martin; Clinton Paden; Clinton R. Paden; Dakota Howard; Darlene Wagner; Dave Wentworth; Dhwani Batra; Jasmine Padilla; Justin Lee; Katie Dillon; Krista Queen; Kristen Knipe; Kristine Lacek; Mark Burroughs; Matthew Schmerer; Meghan Bentz; Mili Sheth; Peter Cook; Sam Shepard; Sarah Nobles; Shoshona Le; Suxiang Tong; Vivien Dugan; Yvette Unoarumhi |
| EPI_ISL_1821703 | UTMG Pathology LLC | UTMG Pathology LLC | Abdallah Azouz; Kelsey Matande; Parasakthy Kumaravelu; Tiara Hymon; Timothy Hodge; Vickie Baselski |
| EPI_ISL_2295545 | UTMG Pathology LLC, University Clinical Health | UTMG Pathology LLC, University Clinical Health | Azouz, A.; Baselski, V.; Hodge, T.; Hymon, T.; Kumaravelu, P.; Matande, K. |
| EPI_ISL_1209089, EPI_ISL_1209127, EPI_ISL_1601379, EPI_ISL_1601541, EPI_ISL_1616687, EPI_ISL_1616688, EPI_ISL_1616727, EPI_ISL_1616825, EPI_ISL_1620958, EPI_ISL_1620967, EPI_ISL_1628169, EPI_ISL_1628172, EPI_ISL_1679827, EPI_ISL_1679836, EPI_ISL_1679848, EPI_ISL_1680170, EPI_ISL_1680187, EPI_ISL_1680202, EPI_ISL_1680265, EPI_ISL_1680343, EPI_ISL_1680344, EPI_ISL_1680359, EPI_ISL_1715555, EPI_ISL_1785673, EPI_ISL_1785688, EPI_ISL_1785761, EPI_ISL_1785764, EPI_ISL_1785765, EPI_ISL_1785766, EPI_ISL_1785831, EPI_ISL_1785842, EPI_ISL_1785934, EPI_ISL_1785941, EPI_ISL_1785951, EPI_ISL_1789247, EPI_ISL_1789302, EPI_ISL_1789318, EPI_ISL_1789452, EPI_ISL_1789457, EPI_ISL_1789459, EPI_ISL_1789461, EPI_ISL_1826087, EPI_ISL_1826100, EPI_ISL_1826101, EPI_ISL_1826107, EPI_ISL_1826115, EPI_ISL_1826116, EPI_ISL_1826117, EPI_ISL_1826125, EPI_ISL_1849074, EPI_ISL_1849262, EPI_ISL_1849267, EPI_ISL_1849277, EPI_ISL_1849438, EPI_ISL_1849659, EPI_ISL_1849723, EPI_ISL_1849737, EPI_ISL_1849739, EPI_ISL_1849833, EPI_ISL_1850126, EPI_ISL_1850135, EPI_ISL_2002160, EPI_ISL_2002168, EPI_ISL_2002177, EPI_ISL_2002182, EPI_ISL_2002188, EPI_ISL_2002244, EPI_ISL_2002510, EPI_ISL_2002753, EPI_ISL_2002789, EPI_ISL_2002838, EPI_ISL_2002907, EPI_ISL_2002915 | | | |
| see above | UW Virology Lab | UW Virology Lab | Alexander Greninger; Hong Xie; Keith R Jerome; Lasata Shrestha; Meei-Li Huang; Michelle Lin; Noah Baker; Noah R. Baker; Pavitra Roychoudhury; Saraswathi Sathees; Sean Ellis; Shah Mohamed Bakhash; Tien V. Nguyen |
| EPI_ISL_2382297 | Uconn Health Clinical Microbiology | Brenton Graveley Lab, UConn Health | Brenton Graveley; Christina Caron; Kimberly Hayes; Lori Avery; Michael Senh; Sara Olson; Stacey Ferraro; Xintao Wei |
| EPI_ISL_1008714 | Unidad de Patología Clínica | Instituto de diagnóstico y Referencia Epidemiologicos (INDRE) Departamento de Virología | Abril Rodriguez-Maldonado; Claudia Wong-Arambula; Ernesto Ramirez-Gonzalez.; Fabiola Garces-Ayala; Gisela Barrera-Badillo; Irma Lopez-Martinez; Lucia Hernandez-Rivas; Natividad Cruz-Ortiz; Tatiana Nunez-Garcia |
| EPI_ISL_2614582 | Unidade Mista de Saude | Instituto Adolfo Lutz, Interdiciplinary Procedures Center, Strategic Laboratory | Caio Vinicius Dias Lopes; Claudia Regina Gonçalves; Claudio Tavares Sacchi; Erica Valessa Ramos Gomes; Karoline Rodrigues Campos; Leonardo Jose Tadeu de Araujo |
| EPI_ISL_1533713, EPI_ISL_1533717 | Unidade de Pronto Atendimento Jd Amanda | Instituto Adolfo Lutz, Interdiciplinary Procedures Center, Strategic Laboratory | Caio Vinicius Dias Lopes; Claudia Regina Gonçalves; Claudio Tavares Sacchi; Erica Valessa Ramos Gomes; Karoline Rodrigues Campos; Leonardo Jose Tadeu de Araujo |
| EPI_ISL_1858758, EPI_ISL_1858761, EPI_ISL_1858763, EPI_ISL_1858765, EPI_ISL_1858767, EPI_ISL_1858771, EPI_ISL_1858775, EPI_ISL_1858777, EPI_ISL_1858779, EPI_ISL_1858781, EPI_ISL_1858783, EPI_ISL_1858786, EPI_ISL_1858788, EPI_ISL_1858790, EPI_ISL_1858792, EPI_ISL_1858794, EPI_ISL_1858796, EPI_ISL_1858798, EPI_ISL_1858800, EPI_ISL_1858802, EPI_ISL_1858803, EPI_ISL_1858805, EPI_ISL_1858807, EPI_ISL_1858809, EPI_ISL_1858811, EPI_ISL_1858814, EPI_ISL_1858816, EPI_ISL_1858818, EPI_ISL_1858819, EPI_ISL_1858822, EPI_ISL_1858824, EPI_ISL_1858826, EPI_ISL_1858828, EPI_ISL_1858830, EPI_ISL_1858832, EPI_ISL_1858834, EPI_ISL_1858836, EPI_ISL_1858838, EPI_ISL_1858842, EPI_ISL_1858843, EPI_ISL_1858846, EPI_ISL_1858847, EPI_ISL_1858850, EPI_ISL_1858852, EPI_ISL_1858853, EPI_ISL_1858856, EPI_ISL_1858858, EPI_ISL_1858860, EPI_ISL_1858862, EPI_ISL_1858864, EPI_ISL_1858866, EPI_ISL_1858868, EPI_ISL_1858870, EPI_ISL_1858872, EPI_ISL_1858878, EPI_ISL_1858880, EPI_ISL_1858882, EPI_ISL_1858884, EPI_ISL_1858886, EPI_ISL_1858888, EPI_ISL_1858890, EPI_ISL_1858892, EPI_ISL_1858894, EPI_ISL_1858896, EPI_ISL_1858898, EPI_ISL_1858900, EPI_ISL_1858902, EPI_ISL_1858904, EPI_ISL_1858906, EPI_ISL_1858908, EPI_ISL_1858910, EPI_ISL_1858912, EPI_ISL_1858914, EPI_ISL_1858916, EPI_ISL_1858918, EPI_ISL_1858920, EPI_ISL_1858922, EPI_ISL_1858924, EPI_ISL_1858926, EPI_ISL_1858928, EPI_ISL_1858930, EPI_ISL_1858933, EPI_ISL_1858934, EPI_ISL_1858936, EPI_ISL_1858938, EPI_ISL_1858940, EPI_ISL_1858942, EPI_ISL_1858944, EPI_ISL_1858946, EPI_ISL_1858948, EPI_ISL_1858950, EPI_ISL_1858952, EPI_ISL_1858954, EPI_ISL_1858957, EPI_ISL_1858960, EPI_ISL_1858963, EPI_ISL_1858964, EPI_ISL_1858966, EPI_ISL_1858968, EPI_ISL_1858970, EPI_ISL_1858973, EPI_ISL_1858975, EPI_ISL_1858977, EPI_ISL_1858979, EPI_ISL_1858981, EPI_ISL_1858984, EPI_ISL_1858986, EPI_ISL_1858988, EPI_ISL_1858990, EPI_ISL_1858992, EPI_ISL_1858994, EPI_ISL_1858996, EPI_ISL_1858998, EPI_ISL_1859000, EPI_ISL_1859002, EPI_ISL_1859005, EPI_ISL_1859007, EPI_ISL_1859008, EPI_ISL_2101429, EPI_ISL_2101430, EPI_ISL_2101431, EPI_ISL_2101432, EPI_ISL_2101433, EPI_ISL_2101443, EPI_ISL_2101455, EPI_ISL_2101456, EPI_ISL_2101457, EPI_ISL_2101458, EPI_ISL_2101459, EPI_ISL_2101460, EPI_ISL_2101461, EPI_ISL_2101462, EPI_ISL_2101463, EPI_ISL_2101470, EPI_ISL_2101471, EPI_ISL_2101472, EPI_ISL_2101473, EPI_ISL_2101483, EPI_ISL_2101484, EPI_ISL_2101494, EPI_ISL_2101495, EPI_ISL_2101505, EPI_ISL_2101506, EPI_ISL_2101510, EPI_ISL_2101512, EPI_ISL_2101520, EPI_ISL_2101522, EPI_ISL_2101523, EPI_ISL_2101524, EPI_ISL_2101525, EPI_ISL_2101526, EPI_ISL_2101527, EPI_ISL_2101566, EPI_ISL_2101567, EPI_ISL_2101568, EPI_ISL_2101569, EPI_ISL_2101570, EPI_ISL_2101571, EPI_ISL_2101572, EPI_ISL_2101612, EPI_ISL_2101638, EPI_ISL_2101639, EPI_ISL_2101640, EPI_ISL_2101641, EPI_ISL_2101642, EPI_ISL_2101643, EPI_ISL_2101644, EPI_ISL_2101645, EPI_ISL_2101646, EPI_ISL_2101647, EPI_ISL_2101648, EPI_ISL_2101649, EPI_ISL_2101650, EPI_ISL_2101651, EPI_ISL_2101652, EPI_ISL_2101653, EPI_ISL_2101654, EPI_ISL_2101655, EPI_ISL_2101656, EPI_ISL_2101657, EPI_ISL_2101658, EPI_ISL_2101659, EPI_ISL_2101660, EPI_ISL_2101661, EPI_ISL_2101662, EPI_ISL_2101663, EPI_ISL_2101664, EPI_ISL_2101665, EPI_ISL_2101666, EPI_ISL_2101667, EPI_ISL_2101668, EPI_ISL_2101669, EPI_ISL_2101670, EPI_ISL_2101676, EPI_ISL_2101677, EPI_ISL_2101678, EPI_ISL_2101679, EPI_ISL_2101680, EPI_ISL_2101681, EPI_ISL_2101682, EPI_ISL_2101683, EPI_ISL_2101684, EPI_ISL_2101685, EPI_ISL_2101686, EPI_ISL_2101687, EPI_ISL_2101688, EPI_ISL_2101692, EPI_ISL_2101693, EPI_ISL_2101694, EPI_ISL_2101695, EPI_ISL_2101715, EPI_ISL_2101722, EPI_ISL_2101724, EPI_ISL_2101731, EPI_ISL_2101734, EPI_ISL_2101740, EPI_ISL_2101741, EPI_ISL_2101744 | | | |
| see above | Unidade de apoio ao diagnóstico da COVID – UNADIG | Bioinformatics Laboratory / LNCC | Alessandra P Lamarca; Alexandra L Gerber; Amilcar Tanuri; Ana Paula de C Guimaraes; Ana Paula de C Guimarães; Ana Tereza R Vasconcelos; Andrea Cony Cavalcanti; Andréa Cony Cavalcanti; Caio Luiz Pereira Ribeiro; Cassia Alves; Cintia Policarpo; Claudia Maria Braga de Mello; Cristiane Gomes da Silva; Diana Mariani; Douglas Terra Machado; Flavio Dias da Silva; Flávio Dias da Silva; Gleidson da Silva de Oliveira; Leandro Magalhaes de Souza; Leandro Magalhães de Souza; Liliane Cavalcante; Luiz G P de Almeida; Marcio Henrique de Oliveira Garcia; Mario Sergio Ribeiro; Ronaldo da Silva F Jr; Silvia Carvalho; Thais Felix Cruz |
| EPI_ISL_2248718 | Univeristy of New Mexico Hospital | Center for Global Health, University of New Mexico Health Sciences Center | Darrell Dinwiddie; Daryl Domman; Jon Femling; Justin Bacca; Kurt Schwalm; Valerie Morley |
| EPI_ISL_1540707, EPI_ISL_1656146 | Universidad Tecnica Particular de Loja | Institute of Microbiology, Universidad San Francisco de Quito | Belén Prado-Vivar; Bernardo Gutiérrez; David Zuñiga; Fernando Serrano; Gabriel Trueba; Juan José Guadalupe; Katherine Ojeda; Luis Flores; Melissa Ortega; Michelle Grunauer; Monica Becerra-Wong; Paola Dalgo; Patricio Rojas-Silva; Paúl Cárdenas; Raiza Briceño; Raiza Briceño; Sully Márquez; Verónica Barragán |
| EPI_ISL_2660543, EPI_ISL_2660544, EPI_ISL_2660547, EPI_ISL_2660548, EPI_ISL_2660549, EPI_ISL_2660550, EPI_ISL_2660551, EPI_ISL_2660552, EPI_ISL_2660554, EPI_ISL_2660555, EPI_ISL_2660556, EPI_ISL_2660557, EPI_ISL_2660558, EPI_ISL_2660559, EPI_ISL_2660560, EPI_ISL_2660561 | | | |
| see above | Universidade Federal de Viçosa (UFV) | Laboratory of Respiratory Viruses and Measles, Oswaldo Cruz Institute, FIOCRUZ | Alice Sampaio Rocha; Ana Carolina Mendonca; Anna Carolina Paixao; Elisa Cavalcante Pereira; Fernando Motta; Luciana Appolinario; Marilda Siqueira on behalf of the Fiocruz COVID-19 Genomic Surveillance Network; Paola Resende; Renata Serrano Lopes; Rubens Pasa; Taina Venas |
| EPI_ISL_1472387, EPI_ISL_1472400, EPI_ISL_1714760, EPI_ISL_1714761, EPI_ISL_1714762, EPI_ISL_1714763, EPI_ISL_1714764, EPI_ISL_1714765, EPI_ISL_1714766, EPI_ISL_1714767, EPI_ISL_1714768, EPI_ISL_1714769 | | | |
| see above | University Hospitals Translational Laboratory (UHTL), University Hospitals | University Hospitals Translational Laboratory (UHTL), University Hospitals | Alouani, D.; Sadri, N.; Song, X. |
| EPI_ISL_981706, EPI_ISL_981707, EPI_ISL_981708, EPI_ISL_981709, EPI_ISL_981710, EPI_ISL_981711, EPI_ISL_981712, EPI_ISL_981713, EPI_ISL_981714, EPI_ISL_1040362, EPI_ISL_1040363, EPI_ISL_1084930, EPI_ISL_1084931, EPI_ISL_1166788, EPI_ISL_1296218, EPI_ISL_1296219, EPI_ISL_1296220, EPI_ISL_1369506, EPI_ISL_1369512, EPI_ISL_1369531, EPI_ISL_1369541, EPI_ISL_1448560, EPI_ISL_1448613, EPI_ISL_1533404, EPI_ISL_1533405, EPI_ISL_1533406, EPI_ISL_1708132 | | | |
| see above | University Hospitals of Geneva, Laboratory of Virology | HUG, Laboratory of Virology and the Health2030 Genome Center | Ana Rita Goncalves; Deborah Penet; Emmanouil Dermitzakis; Henri Pegeot; Ioannis Xenarios; Keith Harshman; Laurent Kaiser; Lorenzo Cerutti; Melyssa Elies; Samuel Cordey |
| EPI_ISL_2939098, EPI_ISL_2939100 | University of Arkansas for Medical Sciences | Center for Global Health, University of New Mexico Health Sciences Center | Catherine Kirkpatrick; Darrell Dinwiddie; Daryl Domman; David Ussery; Jennifer Laudadio; Joshua L. Kennedy; Kurt Schwalm; Susan Harley; Uday Chalwadi; Valerie Morley |
| EPI_ISL_2246579, EPI_ISL_2246581, EPI_ISL_2246585, EPI_ISL_2246590, EPI_ISL_2246605, EPI_ISL_2246606, EPI_ISL_2246607, EPI_ISL_2246622, EPI_ISL_2692017 | | | |
| see above | University of Chicago Medicine | RIPHL at Rush University Medical Center | Cindy Bethel; Felix Araujo Perez; Kevin Kunstman; Laura Furtado; Lauren Megger; Marieta Hyde; Max Kolton; Stefan Green |
| EPI_ISL_1491429, EPI_ISL_1491508, EPI_ISL_1580479, EPI_ISL_1651560, EPI_ISL_1855929, EPI_ISL_1855930, EPI_ISL_2598264 | | | |
| see above | University of Michigan Clinical Microbiology Laboratory | Lauring Lab, University of Michigan, Department of Microbiology and Immunology | Valesano |
| EPI_ISL_1711070 | University of Mississippi Medical Center, Department of Pathology | University of Mississippi Medical Center, Molecular and Genomics Core Facility | Ashley C. Johnson; D. Ashley Robinson; Ithiel J. Frame; Krishna K. Ayyalasomayajula; Michael R. Garrett |
| EPI_ISL_1791529, EPI_ISL_2007829 | University of Oregon COVID-19 MAP Laboratory | University of Oregon Genomics and Cell Characterization Core Facility (GC3F) | Ariana White; Douglas Turnbull; Jason Carriere; Jason Sydes; Jeff Bishop; Peter Batzel |
| EPI_ISL_1366655, EPI_ISL_1366657, EPI_ISL_1743716, EPI_ISL_1972943, EPI_ISL_1972944, EPI_ISL_1972945, EPI_ISL_1972946, EPI_ISL_1972947, EPI_ISL_1972948 | | | |
| see above | Usansolo-Galdakao University Hospital | Cruces University Hospital | Ana Belén de la Hoz; Ana Belén de la Oz; Ana Gual-de-Torrella; Izaskun Alejo-Cancho; Mikel Gallego; mikel Gallego |
| EPI_ISL_1403867, EPI_ISL_1483098, EPI_ISL_1483099, EPI_ISL_2081706, EPI_ISL_2081799, EPI_ISL_2290229, EPI_ISL_2290289, EPI_ISL_2290348, EPI_ISL_2290361, EPI_ISL_2290459, EPI_ISL_2292392, EPI_ISL_2458488, EPI_ISL_2496714, EPI_ISL_2677862, EPI_ISL_2677893, EPI_ISL_2677894, EPI_ISL_2679011, EPI_ISL_2679026, EPI_ISL_2679231, EPI_ISL_3144683, EPI_ISL_3269709 | | | |
| see above | Utah Public Health Laboratory | Utah Public Health Laboratory | Erin L. Young; Kelly F. Oakeson; Olinto Linares-Perdomo; Tara Gallagher |
| EPI_ISL_2283946 | VA Connecticut Healthcare System | Yale Center for Genomic Analysis | Brooke Sullivan; Curt Scharfe; Irina Tikhonova; Kaya Bilguvar; Shrikant Mane |

| | | | |
|---|---|---|---|
| EPI_ISL_3045958 | VIDOH (Virgin Islands Department of Health) | Grubaugh Lab - Yale School of Public Health | Anderson Brito; Annie Watkins; Brett Ellis; Chaney Kalinich; Chantal Vogels; Esther Ellis; Isabel Ott; Jendai Richards; Jessica Rothman; Joseph Fauver; Kendall Billig; Mallery Breban; Marlon Lawrence; Mary Petrone; Nathan Grubaugh; TaLesa Aderohunmu; Tara Alpert; Tobias Koch |
| EPI_ISL_1370399 | VIDYMED EPALINGES | Laboratory of genomics and metagenomics, Institute of Microbiology, University Hospital Centre and University of Lausanne, Switzerland | Claire Bertelli; Damien Jacot; Gilbert Greub; Sébastien Aeby; Trestan Pillonel |
| EPI_ISL_1795339, EPI_ISL_1795340, EPI_ISL_1795343, EPI_ISL_2345605, EPI_ISL_2345606, EPI_ISL_2345609 | VIGILANCIA EM SAUDE | Instituto Butantan / ESALQ-Piracicaba | Antonio Jorge Martins; Bianca Cechetto Carlos. Mendelics: Bibiana Santos; Claudia Renata dos Santos Barros; David Schlesinger; David Schlesinger. Hemocentro Ribeirão Preto: Simone Kashima; Debora Botequio Moretti; Debora Botequio Moretti. Centro de Genômica Funcional da ESALQ: Luiz Lehmann Coutinho; Dimas Tadeu Covas; Elaine Cristina Marqueze; Elaine Vieira Santos; Elaine Vieira dos Santos; Elisangela Chicaroni Mattos; Erika Freitas; Evandra Strazza Rodrigues; Felipe Allan da Silva da Costa; Flavia Aburjaile; Guilherme Targino Valente; Heidge Fukumasu; Heidge Fukumasu. USP-Botucatu: Rejane Maria Tommasini Grotto; Instituto Butantan: Alexander Roberto Precioso; Jayme A. Souza-Neto; Jayme Augusto de Souza-Neto; Jessika Cristina Chagas Lesbon; José Salvatore Leister Patané; João Paulo Kitajima; Luiz Alcantara; Luiz Carlos Junior de Alcantara; Luiz Lehmann Coutinho; Maria Carolina Elias; Marta Giovanetti; Maurício Lacerda Nogueira; Patricia Akemi Assato; Rafael dos Santos Bezerra; Raquel de Lello Rocha Campos Cassano. NGS Soluções Genômicas: Pilar Drummond Sampaio Corrêa Mariani. FZEA-USP Pirassununga: Mirele Daiana Poleti; Raul Machado Neto; Rejane Maria Tommasini Grotto; Ricardo Augusto Brassaloti; Ricardo Haddad; Rodrigo Tocantins Calado.; Sandra Coccuzzo Sampaio; Sandra Coccuzzo Sampaio Vessoni; Simone Kashima; Svetoslav Nanev Slavov; Vagner Fonseca; Vincent Louis Viala |
| EPI_ISL_1966525 | VIGILANCIA EM SAUDE | Instituto Butantan / FZEA-USP (Pirassununga) | Antonio Jorge Martins; Bianca Cechetto Carlos. Mendelics: Bibiana Santos; Claudia Renata dos Santos Barros; Cíntia Bittar; David Schlesinger. Hemocentro Ribeirão Preto: Simone Kashima; Debora Botequio Moretti; Elaine Cristina Marqueze; Elaine Vieira dos Santos; Elisangela Chicaroni Mattos; Erika Freitas; Evandra Strazza Rodrigues; Felipe Allan da Silva da Costa; Flavia Aburjaile; Fábio Sossai Possebon; Guilherme Campos; Guilherme Targino Valente; Heidge Fukumasu. USP-Botucatu: Rejane Maria Tommasini Grotto; Helena Lage Ferreira; Instituto Butantan: Dimas Tadeu Covas; Jardelina de Souza Todao Bernardino; Jayme A. Souza-Neto; Jessika Cristina Chagas Lesbon; Jorge A. Petroli Marchesi; José Salvatore Leister Patané; João Paulo Kitajima; João Pessoa Araújo Jr.; Leila Sabrina Ullmann; Loyze Paola Oliveira de Lima; Luiz Aurelio de Campos Crispin. Centro de Genômica Funcional da ESALQ: Luiz Lehmann Coutinho; Luiz Carlos Junior de Alcantara; Lívia Sacchetto; Maisa C. Pereira Parra; Maria Carolina Elias; Marta Giovanetti; Marília Moraes; Maurício Lacerda Nogueira. Prefeitura de Sao Paulo: Melissa Palmieri.; Patricia Akemi Assato; Paula Rahal; Paulo Inacio da Costa; Rafael dos Santos Bezerra; Raquel de Lello Rocha Campos Cassano. NGS Soluções Genômicas: Pilar Drummond Sampaio Corrêa Mariani. FZEA-USP Pirassununga: Mirele Daiana Poleti; Raul Machado Neto; Ricardo Augusto Brassaloti; Ricardo Haddad; Rodrigo Tocantins Calado. FAMERP-SJRP: Cecília Artico Banho; Sandra Coccuzzo Sampaio; Svetoslav Nanev Slavov; Vagner Fonseca; Vincent Louis Viala |
| EPI_ISL_1966556, EPI_ISL_1967199, EPI_ISL_1967200, EPI_ISL_1967202 | VIGILANCIA EM SAUDE SAE VIGILANCIA EPIDEMIOLOGICA | Instituto Butantan / Mendelics | Antonio Jorge Martins; Bianca Cechetto Carlos. Mendelics: Bibiana Santos; Claudia Renata dos Santos Barros; Cíntia Bittar; David Schlesinger. Hemocentro Ribeirão Preto: Simone Kashima; Debora Botequio Moretti; Elaine Cristina Marqueze; Elaine Vieira dos Santos; Elisangela Chicaroni Mattos; Erika Freitas; Evandra Strazza Rodrigues; Felipe Allan da Silva da Costa; Flavia Aburjaile; Fábio Sossai Possebon; Guilherme Campos; Guilherme Targino Valente; Heidge Fukumasu. USP-Botucatu: Rejane Maria Tommasini Grotto; Helena Lage Ferreira; Instituto Butantan: Dimas Tadeu Covas; Jardelina de Souza Todao Bernardino; Jayme A. Souza-Neto; Jessika Cristina Chagas Lesbon; Jorge A. Petroli Marchesi; José Salvatore Leister Patané; João Paulo Kitajima; João Pessoa Araújo Jr.; Leila Sabrina Ullmann; Loyze Paola Oliveira de Lima; Luiz Aurelio de Campos Crispin. Centro de Genômica Funcional da ESALQ: Luiz Lehmann Coutinho; Luiz Carlos Junior de Alcantara; Lívia Sacchetto; Maisa C. Pereira Parra; Maria Carolina Elias; Marta Giovanetti; Marília Moraes; Maurício Lacerda Nogueira. Prefeitura de Sao Paulo: Melissa Palmieri.; Patricia Akemi Assato; Paula Rahal; Paulo Inacio da Costa; Rafael dos Santos Bezerra; Raquel de Lello Rocha Campos Cassano. NGS Soluções Genômicas: Pilar Drummond Sampaio Corrêa Mariani. FZEA-USP Pirassununga: Mirele Daiana Poleti; Raul Machado Neto; Ricardo Augusto Brassaloti; Ricardo Haddad; Rodrigo Tocantins Calado. FAMERP-SJRP: Cecília Artico Banho; Sandra Coccuzzo Sampaio; Svetoslav Nanev Slavov; Vagner Fonseca; Vincent Louis Viala |
| EPI_ISL_1966084 | VIGILANCIA EPIDEMIOLOGICA | Instituto Butantan / Mendelics | Antonio Jorge Martins; Bianca Cechetto Carlos. Mendelics: Bibiana Santos; Claudia Renata dos Santos Barros; Cíntia Bittar; David Schlesinger. Hemocentro Ribeirão Preto: Simone Kashima; Debora Botequio Moretti; Elaine Cristina Marqueze; Elaine Vieira dos Santos; Elisangela Chicaroni Mattos; Erika Freitas; Evandra Strazza Rodrigues; Felipe Allan da Silva da Costa; Flavia Aburjaile; Fábio Sossai Possebon; Guilherme Campos; Guilherme Targino Valente; Heidge Fukumasu. USP-Botucatu: Rejane Maria Tommasini Grotto; Helena Lage Ferreira; Instituto Butantan: Dimas Tadeu Covas; Jardelina de Souza Todao Bernardino; Jayme A. Souza-Neto; Jessika Cristina Chagas Lesbon; Jorge A. Petroli Marchesi; José Salvatore Leister Patané; João Paulo Kitajima; João Pessoa Araújo Jr.; Leila Sabrina Ullmann; Loyze Paola Oliveira de Lima; Luiz Aurelio de Campos Crispin. Centro de Genômica Funcional da ESALQ: Luiz Lehmann Coutinho; Luiz Carlos Junior de Alcantara; Lívia Sacchetto; Maisa C. Pereira Parra; Maria Carolina Elias; Marta Giovanetti; Marília Moraes; Maurício Lacerda Nogueira. Prefeitura de Sao Paulo: Melissa Palmieri.; Patricia Akemi Assato; Paula Rahal; Paulo Inacio da Costa; Rafael dos Santos Bezerra; Raquel de Lello Rocha Campos Cassano. NGS Soluções Genômicas: Pilar Drummond Sampaio Corrêa Mariani. FZEA-USP Pirassununga: Mirele Daiana Poleti; Raul Machado Neto; Ricardo Augusto Brassaloti; Ricardo Haddad; Rodrigo Tocantins Calado. FAMERP-SJRP: Cecília Artico Banho; Sandra Coccuzzo Sampaio; Svetoslav Nanev Slavov; Vagner Fonseca; Vincent Louis Viala |
| EPI_ISL_1966296, EPI_ISL_1966300 | VIGILANCIA EPIDEMIOLOGICA DE IBATE | Instituto Butantan / Mendelics | Antonio Jorge Martins; Bianca Cechetto Carlos. Mendelics: Bibiana Santos; Claudia Renata dos Santos Barros; Cíntia Bittar; David Schlesinger. Hemocentro Ribeirão Preto: Simone Kashima; Debora Botequio Moretti; Elaine Cristina Marqueze; Elaine Vieira dos Santos; Elisangela Chicaroni Mattos; Erika Freitas; Evandra Strazza Rodrigues; Felipe Allan da Silva da Costa; Flavia Aburjaile; Fábio Sossai Possebon; Guilherme Campos; Guilherme Targino Valente; Heidge Fukumasu. USP-Botucatu: Rejane Maria Tommasini Grotto; Helena Lage Ferreira; Instituto Butantan: Dimas Tadeu Covas; Jardelina de Souza Todao Bernardino; Jayme A. Souza-Neto; Jessika Cristina Chagas Lesbon; Jorge A. Petroli Marchesi; José Salvatore Leister Patané; João Paulo Kitajima; João Pessoa Araújo Jr.; Leila Sabrina Ullmann; Loyze Paola Oliveira de Lima; Luiz Aurelio de Campos Crispin. Centro de Genômica Funcional da ESALQ: Luiz Lehmann Coutinho; Luiz Carlos Junior de Alcantara; Lívia Sacchetto; Maisa C. Pereira Parra; Maria Carolina Elias; Marta Giovanetti; Marília Moraes; Maurício Lacerda Nogueira. Prefeitura de Sao Paulo: Melissa Palmieri.; Patricia Akemi Assato; Paula Rahal; Paulo Inacio da Costa; Rafael dos Santos Bezerra; Raquel de Lello Rocha Campos Cassano. NGS Soluções Genômicas: Pilar Drummond Sampaio Corrêa Mariani. FZEA-USP Pirassununga: Mirele Daiana Poleti; Raul Machado Neto; Ricardo Augusto Brassaloti; Ricardo Haddad; Rodrigo Tocantins Calado. FAMERP-SJRP: Cecília Artico Banho; Sandra Coccuzzo Sampaio; Svetoslav Nanev Slavov; Vagner Fonseca; Vincent Louis Viala |
| EPI_ISL_2344746, EPI_ISL_2344809 | VIGILANCIA EPIDEMIOLOGICA DE LEME | Instituto Butantan / ESALQ-Piracicaba | Antonio Jorge Martins; Claudia Renata dos Santos Barros; David Schlesinger; Debora Botequio Moretti; Dimas Tadeu Covas; Elaine Cristina Marqueze; Elaine Vieira Santos; Evandra Strazza Rodrigues; Heidge Fukumasu; Jayme Augusto de Souza-Neto; José Salvatore Leister Patané; Luiz Alcantara; Luiz Lehmann Coutinho; Maria Carolina Elias; Maurício Lacerda Nogueira; Rafael dos Santos Bezerra; Raul Machado Neto; Rejane Maria Tommasini Grotto; Ricardo Haddad; Sandra Coccuzzo Sampaio Vessoni; Simone Kashima; Svetoslav Nanev Slavov; Vincent Louis Viala |
| EPI_ISL_2210045, EPI_ISL_2344694, EPI_ISL_2344713, EPI_ISL_2344714, EPI_ISL_2344715, EPI_ISL_2344720 | VIGILANCIA EPIDEMIOLOGICA DE LEME | Instituto Butantan / FZEA-USP-Pirassununga | Antonio Jorge Martins; Claudia Renata dos Santos Barros; David Schlesinger; Debora Botequio Moretti; Dimas Tadeu Covas; Elaine Cristina Marqueze; Elaine Vieira Santos; Evandra Strazza Rodrigues; Heidge Fukumasu; Jayme Augusto de Souza-Neto; José Salvatore Leister Patané; Luiz Alcantara; Luiz Lehmann Coutinho; Maria Carolina Elias; Maurício Lacerda Nogueira; Rafael dos Santos Bezerra; Raul Machado Neto; Rejane Maria Tommasini Grotto; Ricardo Haddad; Sandra Coccuzzo Sampaio Vessoni; Simone Kashima; Svetoslav Nanev Slavov; Vincent Louis Viala |
| EPI_ISL_2210165, EPI_ISL_2210189, EPI_ISL_2210231, EPI_ISL_2210243, EPI_ISL_2210257, EPI_ISL_2210281 | VIGILANCIA EPIDEMIOLOGICA E CONTROLE DE VETORES PIRASSUNUN | Instituto Butantan | Antonio Jorge Martins; Claudia Renata dos Santos Barros; David Schlesinger; Debora Botequio Moretti; Dimas Tadeu Covas; Elaine Cristina Marqueze; Elaine Vieira Santos; Evandra Strazza Rodrigues; Heidge Fukumasu; Jayme Augusto de Souza-Neto; José Salvatore Leister Patané; Luiz Alcantara; Luiz Lehmann Coutinho; Maria Carolina Elias; Maurício Lacerda Nogueira; Rafael dos Santos Bezerra; Raul Machado Neto; Rejane Maria Tommasini Grotto; Ricardo Haddad; Sandra Coccuzzo Sampaio Vessoni; Simone Kashima; Svetoslav Nanev Slavov; Vincent Louis Viala. |
| EPI_ISL_2344771 | VIGILANCIA EPIDEMIOLOGICA E CONTROLE DE VETORES PIRASSUNUN | Instituto Butantan / ESALQ-Piracicaba | Antonio Jorge Martins; Claudia Renata dos Santos Barros; David Schlesinger; Debora Botequio Moretti; Dimas Tadeu Covas; Elaine Cristina Marqueze; Elaine Vieira Santos; Evandra Strazza Rodrigues; Heidge Fukumasu; Jayme Augusto de Souza-Neto; José Salvatore Leister Patané; Luiz Alcantara; Luiz Lehmann Coutinho; Maria Carolina Elias; Maurício Lacerda Nogueira; Rafael dos Santos Bezerra; Raul Machado Neto; Rejane Maria Tommasini Grotto; Ricardo Haddad; Sandra Coccuzzo Sampaio Vessoni; Simone Kashima; Svetoslav Nanev Slavov; Vincent Louis Viala |
| EPI_ISL_2209443, EPI_ISL_2209448, EPI_ISL_2210019, EPI_ISL_2210030, EPI_ISL_2344555, EPI_ISL_2344585, EPI_ISL_2344663, EPI_ISL_2344664, EPI_ISL_2344667, EPI_ISL_2344670, EPI_ISL_2344695, EPI_ISL_2344697, EPI_ISL_2344698, EPI_ISL_2344704, EPI_ISL_2344706, EPI_ISL_2344719 | | | |
| see above | VIGILANCIA EPIDEMIOLOGICA E CONTROLE DE VETORES PIRASSUNUN | Instituto Butantan / FZEA-USP-Pirassununga | Antonio Jorge Martins; Claudia Renata dos Santos Barros; David Schlesinger; Debora Botequio Moretti; Dimas Tadeu Covas; Elaine Cristina Marqueze; Elaine Vieira Santos; Evandra Strazza Rodrigues; Heidge Fukumasu; Jayme Augusto de Souza-Neto; José Salvatore Leister Patané; Luiz Alcantara; Luiz Lehmann Coutinho; Maria Carolina Elias; Maurício Lacerda Nogueira; Rafael dos Santos Bezerra; Raul Machado Neto; Rejane Maria Tommasini Grotto; Ricardo Haddad; Sandra Coccuzzo Sampaio Vessoni; Simone Kashima; Svetoslav Nanev Slavov; Vincent Louis Viala |
| EPI_ISL_1795217, EPI_ISL_2345445 | VIGILANCIA EPIDEMIOLOGICA JARDINOPOLIS SP | Instituto Butantan / ESALQ-Piracicaba | Antonio Jorge Martins; Bianca Cechetto Carlos. Mendelics: Bibiana Santos; Claudia Renata dos Santos Barros; David Schlesinger; David Schlesinger. Hemocentro Ribeirão Preto: Simone Kashima; Debora Botequio Moretti; Debora Botequio Moretti. Centro de Genômica Funcional da ESALQ: Luiz Lehmann Coutinho; Dimas Tadeu Covas; Elaine Cristina Marqueze; Elaine Vieira Santos; Elaine Vieira dos Santos; Elisangela Chicaroni Mattos; Erika Freitas; Evandra Strazza Rodrigues; Felipe Allan da Silva da Costa; Flavia Aburjaile; Guilherme Targino Valente; Heidge Fukumasu; Heidge Fukumasu. USP-Botucatu: Rejane Maria Tommasini Grotto; Instituto Butantan: Alexander Roberto Precioso; Jayme A. Souza-Neto; Jayme Augusto de Souza-Neto; Jessika Cristina Chagas Lesbon; José Salvatore Leister Patané; João Paulo Kitajima; Luiz Alcantara; Luiz Carlos Junior de Alcantara; Luiz Lehmann Coutinho; Maria Carolina Elias; Marta Giovanetti; Maurício Lacerda Nogueira; Patricia Akemi Assato; Rafael dos Santos Bezerra; Raquel de Lello Rocha Campos Cassano. NGS Soluções Genômicas: Pilar Drummond Sampaio Corrêa Mariani. FZEA-USP Pirassununga: Mirele Daiana Poleti; Raul Machado Neto; Rejane Maria Tommasini Grotto; Ricardo Augusto Brassaloti; Ricardo Haddad; Rodrigo Tocantins Calado.; Sandra Coccuzzo Sampaio; Sandra Coccuzzo Sampaio Vessoni; Simone Kashima; Svetoslav Nanev Slavov; Vagner Fonseca; Vincent Louis Viala |
| EPI_ISL_1795218, EPI_ISL_2345447 | VIGILANCIA EPIDIMIOLOGICA DE ARACARIGUAMA | Instituto Butantan / ESALQ-Piracicaba | Antonio Jorge Martins; Bianca Cechetto Carlos. Mendelics: Bibiana Santos; Claudia Renata dos Santos Barros; David Schlesinger; David Schlesinger. Hemocentro Ribeirão Preto: Simone Kashima; Debora Botequio Moretti; Debora Botequio Moretti. Centro de Genômica Funcional da ESALQ: Luiz Lehmann Coutinho; Dimas Tadeu Covas; Elaine Cristina Marqueze; Elaine Vieira Santos; Elaine Vieira dos Santos; Elisangela Chicaroni Mattos; Erika Freitas; Evandra Strazza Rodrigues; Felipe Allan da Silva da Costa; Flavia Aburjaile; Guilherme Targino Valente; Heidge Fukumasu; Heidge Fukumasu. USP-Botucatu: Rejane Maria Tommasini Grotto; Instituto Butantan: Alexander Roberto Precioso; Jayme A. Souza-Neto; Jayme Augusto de Souza-Neto; Jessika Cristina Chagas Lesbon; José Salvatore Leister Patané; João Paulo Kitajima; Luiz Alcantara; Luiz Carlos Junior de Alcantara; Luiz Lehmann Coutinho; Maria Carolina Elias; Marta Giovanetti; Maurício Lacerda Nogueira; Patricia Akemi Assato; Rafael dos Santos Bezerra; Raquel de Lello Rocha Campos Cassano. NGS Soluções Genômicas: Pilar Drummond Sampaio Corrêa Mariani. FZEA-USP Pirassununga: Mirele Daiana Poleti; Raul Machado Neto; Rejane Maria Tommasini Grotto; Ricardo Augusto Brassaloti; Ricardo Haddad; Rodrigo Tocantins Calado.; Sandra Coccuzzo Sampaio; Sandra Coccuzzo Sampaio Vessoni; Simone Kashima; Svetoslav Nanev Slavov; Vagner Fonseca; Vincent Louis Viala |
| EPI_ISL_1966830, EPI_ISL_1966831 | VIGILANCIA EPIDIMIOLOGICA DE ARACARIGUAMA | Instituto Butantan / ESALQ-USP (Piracicaba) | Antonio Jorge Martins; Bianca Cechetto Carlos. Mendelics: Bibiana Santos; Claudia Renata dos Santos Barros; Cíntia Bittar; David Schlesinger. Hemocentro Ribeirão Preto: Simone Kashima; Debora Botequio Moretti; Elaine Cristina Marqueze; Elaine Vieira dos Santos; Elisangela Chicaroni Mattos; Erika Freitas; Evandra Strazza Rodrigues; Felipe Allan da Silva da Costa; Flavia Aburjaile; Fábio Sossai Possebon; Guilherme Campos; Guilherme Targino Valente; Heidge Fukumasu. USP-Botucatu: Rejane Maria Tommasini Grotto; Helena Lage Ferreira; Instituto Butantan: Dimas Tadeu Covas; Jardelina de Souza Todao Bernardino; Jayme A. Souza-Neto; Jessika Cristina Chagas Lesbon; Jorge A. Petroli Marchesi; José Salvatore Leister Patané; João Paulo Kitajima; João Pessoa Araújo Jr.; Leila Sabrina Ullmann; Loyze Paola Oliveira de Lima; Luiz Aurelio de Campos Crispin. Centro de Genômica Funcional da ESALQ: Luiz Lehmann Coutinho; Luiz Carlos Junior de Alcantara; Lívia Sacchetto; Maisa C. Pereira Parra; Maria Carolina Elias; Marta Giovanetti; Marília Moraes; Maurício Lacerda Nogueira. Prefeitura de Sao Paulo: Melissa Palmieri.; Patricia Akemi Assato; Paula Rahal; Paulo Inacio da Costa; Rafael dos Santos Bezerra; Raquel de Lello Rocha Campos Cassano. NGS Soluções Genômicas: Pilar Drummond Sampaio Corrêa Mariani. FZEA-USP Pirassununga: Mirele Daiana Poleti; Raul Machado Neto; Ricardo Augusto Brassaloti; Ricardo Haddad; Rodrigo Tocantins Calado. FAMERP-SJRP: Cecília Artico Banho; Sandra Coccuzzo Sampaio; Svetoslav Nanev Slavov; Vagner Fonseca; Vincent Louis Viala |
| EPI_ISL_2210285, EPI_ISL_2210295, EPI_ISL_2345973, EPI_ISL_2345975 | VIGILANCIA SANITARIA E VIG EPIDEMIOLOGICA DE ITAPEVI | Instituto Butantan | Antonio Jorge Martins; Claudia Renata dos Santos Barros; David Schlesinger; Debora Botequio Moretti; Dimas Tadeu Covas; Elaine Cristina Marqueze; Elaine Vieira Santos; Evandra Strazza Rodrigues; Heidge Fukumasu; Jayme Augusto de Souza-Neto; José Salvatore Leister Patané; Luiz Alcantara; Luiz Lehmann Coutinho; Maria Carolina Elias; Maurício Lacerda Nogueira; Rafael dos Santos Bezerra; Raul Machado Neto; Rejane Maria Tommasini Grotto; Ricardo Haddad; Sandra Coccuzzo Sampaio Vessoni; Simone Kashima; Svetoslav Nanev Slavov; Vincent Louis Viala; Vincent Louis Viala. |
| EPI_ISL_1966935, EPI_ISL_1966936, EPI_ISL_1966937, EPI_ISL_1966938, EPI_ISL_1966940 | VIGILANCIA SANITARIA E VIG EPIDEMIOLOGICA DE ITAPEVI | Instituto Butantan / ESALQ-USP (Piracicaba) | Antonio Jorge Martins; Bianca Cechetto Carlos. Mendelics: Bibiana Santos; Claudia Renata dos Santos Barros; Cíntia Bittar; David Schlesinger. Hemocentro Ribeirão Preto: Simone Kashima; Debora Botequio Moretti; Elaine Cristina Marqueze; Elaine Vieira dos Santos; Elisangela Chicaroni Mattos; Erika Freitas; Evandra Strazza Rodrigues; Felipe Allan da Silva da Costa; Flavia Aburjaile; Fábio Sossai Possebon; Guilherme Campos; Guilherme Targino Valente; Heidge Fukumasu. USP-Botucatu: Rejane Maria Tommasini Grotto; Helena Lage Ferreira; Instituto Butantan: Dimas Tadeu Covas; Jardelina de Souza Todao Bernardino; Jayme A. Souza-Neto; Jessika Cristina Chagas Lesbon; Jorge A. Petroli Marchesi; José Salvatore Leister Patané; João Paulo Kitajima; João Pessoa Araújo Jr.; Leila Sabrina Ullmann; Loyze Paola Oliveira de Lima; Luiz Aurelio de Campos Crispin. Centro de Genômica Funcional da ESALQ: Luiz Lehmann Coutinho; Luiz Carlos Junior de Alcantara; Lívia Sacchetto; Maisa C. Pereira Parra; Maria Carolina Elias; Marta Giovanetti; Marília Moraes; Maurício Lacerda Nogueira. Prefeitura de Sao Paulo: Melissa Palmieri.; Patricia Akemi Assato; Paula Rahal; Paulo Inacio da Costa; Rafael dos Santos Bezerra; Raquel de Lello Rocha Campos Cassano. NGS Soluções Genômicas: Pilar Drummond Sampaio Corrêa Mariani. FZEA-USP Pirassununga: Mirele Daiana Poleti; Raul Machado Neto; Ricardo Augusto Brassaloti; Ricardo Haddad; Rodrigo Tocantins Calado. FAMERP-SJRP: Cecília Artico Banho; Sandra Coccuzzo Sampaio; Svetoslav Nanev Slavov; Vagner Fonseca; Vincent Louis Viala |
| EPI_ISL_2345881, EPI_ISL_2345885, EPI_ISL_2345887 | VIGILANCIA SANITARIA E VIG EPIDEMIOLOGICA DE ITAPEVI | Instituto Butantan / FZEA-USP-Pirassununga | Antonio Jorge Martins; Claudia Renata dos Santos Barros; David Schlesinger; Debora Botequio Moretti; Dimas Tadeu Covas; Elaine Cristina Marqueze; Elaine Vieira Santos; Evandra Strazza Rodrigues; Heidge Fukumasu; Jayme Augusto de Souza-Neto; José Salvatore Leister Patané; Luiz Alcantara; Luiz Lehmann Coutinho; Maria Carolina Elias; Maurício Lacerda Nogueira; Rafael dos Santos Bezerra; Raul Machado Neto; Rejane Maria Tommasini Grotto; Ricardo Haddad; Sandra Coccuzzo Sampaio Vessoni; Simone Kashima; Svetoslav Nanev Slavov; Vincent Louis Viala |
| EPI_ISL_2036221, EPI_ISL_2036229, EPI_ISL_2036236, EPI_ISL_2036257, EPI_ISL_2036258, EPI_ISL_2036259, EPI_ISL_2036260, EPI_ISL_2109691, EPI_ISL_2293028, EPI_ISL_3667127, EPI_ISL_3667131 | | | |
| see above | VIROLOGY LABORATORY-CHU NICE | VIROLOGY LABORATORY-CHU NICE | Aicha El Yakine; Geraldine Gonfrier; Jean Machowiak; Sebastien Vitale; Valerie Giordanengo; Virginie Flahou |
| EPI_ISL_913587 | Vault Health | Minnesota Department of Health, Public Health Laboratory | Alexandra Lorentz; Jacob Garfin; Matt Plumb; and Xiong Wang |
| EPI_ISL_1533716 | Vigilancia Em Saude | Instituto Adolfo Lutz, Interdiciplinary Procedures Center, Strategic Laboratory | Caio Vinicius Dias Lopes; Claudia Regina Gonçalves; Claudio Tavares Sacchi; Erica Valessa Ramos Gomes; Karoline Rodrigues Campos; Leonardo Jose Tadeu de Araujo |

| | | | |
|---|---|---|---|
| EPI_ISL_1628378 | Vigilancia Epidemiologica Jardinopolis | Instituto Adolfo Lutz, Interdiciplinary Procedures Center, Strategic Laboratory | Caio Vinicius Dias Lopes; Claudia Regina Gonçalves; Claudio Tavares Sacchi; Erica Valessa Ramos Gomes; Karoline Rodrigues Campos; Katia Correa de Oliveira Santos; Leonardo Jose Tadeu de Araujo |
| EPI_ISL_2614586 | Vigilância Sanitaria E Epidemiologica | Instituto Adolfo Lutz, Interdiciplinary Procedures Center, Strategic Laboratory | Caio Vinicius Dias Lopes; Claudia Regina Gonçalves; Claudio Tavares Sacchi; Erica Valessa Ramos Gomes; Karoline Rodrigues Campos; Leonardo Jose Tadeu de Araujo |
| EPI_ISL_1747467, EPI_ISL_1747931 | Viollier AG | Clinical Bacteriology | Adrian Egli; Alfredo Mari; Christiane Beckmann; Hans Hirsch; Helena MB Seth-Smith; Julia Bielicki; Karoline Leuzinger; Madlen Stange; Manuel Battegay; Tim Roloff |
| EPI_ISL_1659173, EPI_ISL_1914208, EPI_ISL_1914209 | Viollier AG | Department of Biosystems Science and Engineering, ETH Zurich | Andrea Patrignani; Andreia Cabral de Gouvea; Catharine Aquino; Chaoran Chen; Christiane Beckmann; Christoph Noppen; David Dreifuss; Deborah Penet; Doris Popovic; Emmanouil Dermitzakis; Griffin White; Henri Pegeot; Ioannis Xenarios; Ivan Topolsky; Jay Tracy; Katharina Jahn; Keith Harshman; Lara Fuhrmann; Laura Neff; Lennart Opitz; Lorenzo Cerutti; Maria Domenica Moccia; Maurice Redondo; Niko Beerenwinkel; Noemie Santamaria de Souza; Olivier Kobel; Philipp Jablonski; Ralph Schlapbach; Sarah Nadeau; Simon Gruter; Sophie Seidel; Tanja Stadler; Timothy Sykes |
| EPI_ISL_1119344, EPI_ISL_1361446, EPI_ISL_1750646 | Viollier AG | Department of Biosystems Science and Engineering, ETH Zürich | Chaoran Chen; Christian Beisel; Christiane Beckmann; Christoph Noppen; David Dreifuss; Elodie Burcklen; Ina Nissen; Ivan Topolsky; Katharina Jahn; Lara Fuhrmann; Maurice Redondo; Mirjam Feldkamp; Natascha Santacroce; Niko Beerenwinkel; Noemie Santamaria de Souza; Olivier Kobel; Philipp Jablonski; Rebecca Denes; Sarah Nadeau; Sophie Seidel; Tanja Stadler |
| EPI_ISL_1805663, EPI_ISL_2362006 | Virginia Division of Consolidated Laboratory Services | Virginia Division of Consolidated Laboratory Services | Virginia DCLS |
| EPI_ISL_1104214, EPI_ISL_1104216, EPI_ISL_1636703 | Virology Department, Royal Infirmary of Edinburgh, NHS Lothian / School of Biological Sciences, University of Edinburgh | COVID-19 Genomics UK (COG-UK) Consortium | Colquhoun R; Cotton S; Dewar R; Hill V; Jackson B; McCrone JT; McHugh M; O'Toole Á; Rambaut A; Rooke S; Scher E; Templeton K; Yu X |
| EPI_ISL_1823830 | WA State Department of Health | Centers for Disease Control and Prevention Division of Viral Diseases, Pathogen Discovery | Alison Laufer Halpin; Ben L. Rambo-Martin; Clinton R. Paden; Dakota Howard; Darlene Wagner; Dave Wentworth; Dhwani Batra; Jasmine Padilla; Justin Lee; Katie Dillon; Krista Queen; Kristen Knipe; Kristine Lacek; Mark Burroughs; Matthew Schmerer; Mili Sheth; Peter Cook; Sam Shepard; Sarah Nobles; Shoshona Le; Suxiang Tong; Vivien Dugan; Yvette Unoarumhi |
| EPI_ISL_1671475 | WESTCHESTER MEDICAL CENTER | Wadsworth Center, New York State Department of Health | Alexis Russell; Catharine Prussing; Daryl M. Lamson; Erasmus Schneider; Erica Lasek-Nesselquist; John Kelly; Jonathan Plitnick; Kirsten St. George; Matthew Shudt; Melissa A Leisner; Navjot Singh |
| EPI_ISL_1653885, EPI_ISL_1747415, EPI_ISL_1747425, EPI_ISL_1747435, EPI_ISL_1747450, EPI_ISL_1747455, EPI_ISL_1897420, EPI_ISL_1897431 | | | |
| see above | Washington State Department of Health Public Health Laboratories | Washington State Department of Health Public Health Laboratories | Avi Singh; Darren Lucas; Denny Russell; Drew MacKellar; Geoff Melly; Hannah Gray; Joenice Gonzalez; JohnAric Peterson; Philip Dykema; Rebecca Cao; Vanessa De Los Santos |
| EPI_ISL_1971097 | Weill Cornell Medicine | New York Genome Center | Andre Corvelo; Arryn Craney; Chris Mason; Dayna M. Oschwald; Hanna Rennert; Lars F Westblade; Margaret Elizabeth Ross; Melissa Cushing; Michael Zody; Olivier Elemento; Priya Velu; Samantha Fennessey; Tom Maniatis |
| EPI_ISL_2400232, EPI_ISL_2454296, EPI_ISL_2454549 | Wexner Medical Center | The Ohio State University College of Medicine | Koenig, S.; Seminetta, J. |
| EPI_ISL_1760191, EPI_ISL_1760196, EPI_ISL_1760198 | Wichita State University - Molecular Diagnostics Lab | Kansas Health and Environmental Lab | Ben Olsen; Jonathan Barnell; Mike Grose; and Phil Adam |
| EPI_ISL_1420978, EPI_ISL_1626455, EPI_ISL_1626464, EPI_ISL_1793915, EPI_ISL_1793994, EPI_ISL_1794087, EPI_ISL_1794124, EPI_ISL_1794239, EPI_ISL_1794445, EPI_ISL_1794519, EPI_ISL_1794549, EPI_ISL_2018546, EPI_ISL_2018581, EPI_ISL_2018614, EPI_ISL_2018675, EPI_ISL_2018676, EPI_ISL_2018692, EPI_ISL_2018733, EPI_ISL_2018802, EPI_ISL_2018804, EPI_ISL_2279053 | | | |
| see above | Wisconsin State Laboratory of Hygiene Communicable Disease Division | Wisconsin State Laboratory of Hygiene Communicable Disease Division | Abigail C. Shockey; Alicia J. Mooney; Erika M. Hanson; Kelsey R. Florek; Richard Griesser; Sara Wagner; Tonya Danz |
| EPI_ISL_3722611 | WolfTRACS Surveillance Lab | Harris/Rasmussen Lab | Angela Harris; David Rasmussen; Gina Chaput; Megan Jacob; Temi Ibitoye |
| EPI_ISL_1293215, EPI_ISL_1378750, EPI_ISL_1447557, EPI_ISL_1447603, EPI_ISL_1447620, EPI_ISL_1674553, EPI_ISL_1792971, EPI_ISL_1793089, EPI_ISL_1963711, EPI_ISL_1963744, EPI_ISL_2023066 | | | |
| see above | Yale Clinical Virology Lab | Grubaugh Lab - Yale School of Public Health | Anderson Brito; Annie Watkins; Chaney Kalinich; Chantal Vogels; Isabel Ott; Isabell Ott; Jessica Rothman; Joseph Fauver; Mallery Breban; Marie L. Landry; Mary Petrone; Nathan Grubaugh; Tara Alpert |
| EPI_ISL_1793506, EPI_ISL_2034772 | Yale Clinical Virology Lab | Yale Center for Genomic Analysis | Brooke Sullivan; Christopher Castaldi; Curt Scharfe; Irina Tikhonova; Kaya Bilguvar; Shrikant Mane |
| EPI_ISL_1147719 | amedes MVZ für Laboratoriumsmedizin Fürstenfeldbruck | Robert Koch Institute | |
| EPI_ISL_2613707 | cerballiance-IDF | Cerba lab | Aude Lessenne; Bénédicte Roquebert; Emmanuel Lecorche; Kader Merah; Laura Verdurme; Patrice Herisson; Sabine Trombert-Paolantoni; Stéphanie Haïm-Boukobza; Thierry Collin |
| EPI_ISL_1262813 | hopital | National Reference Center for Viruses of Respiratory Infections, Institut Pasteur, Paris | Angela Brisebarre; Camille Capel; Etienne Simon-Lorière; Leruez-Ville Marianne; Marion Barbet; Maud Vanpeene; Méline Bizard; Sylvie Behillil; Sylvie van der Werf; Vincent Enouf |
| EPI_ISL_1085359 | laboratoire Belle Epine | Department of Virology, Henri Mondor University Hospital, Assistance Publique Hôpitaux de Paris, Université Paris-Est Créteil, INSERM U955 | Alexandre Soulier; Christophe Rodriguez; Elisabeth Trawinski; Guillaume Gricourt; Jean-Michel Pawlotsky; Melissa N'Debi; Slim Fourati; Vanessa Demontant |
| EPI_ISL_1799013 | unknown | Instituto Nacional de Saude (INSA) | Borges et al |
| EPI_ISL_2488795, EPI_ISL_2488796 | Evandro Chagas Institute | Evandro Chagas Institute | A.M.; Barbagelata; E.C.; E.M.A.; Ferreira; J.A.; Junior; K.C.; L.C.; L.S.; M.C.; P.S.; Pinheiro; Santos; Silva; Sousa; Sousa Junior; W.D.C.; da Silva |
