## Supplementary File 3 for "Comparative genomics and characterization of SARS-CoV-2 P.1 (Gamma) Variant of Concern (VOC) from Amazonas, Brazil"

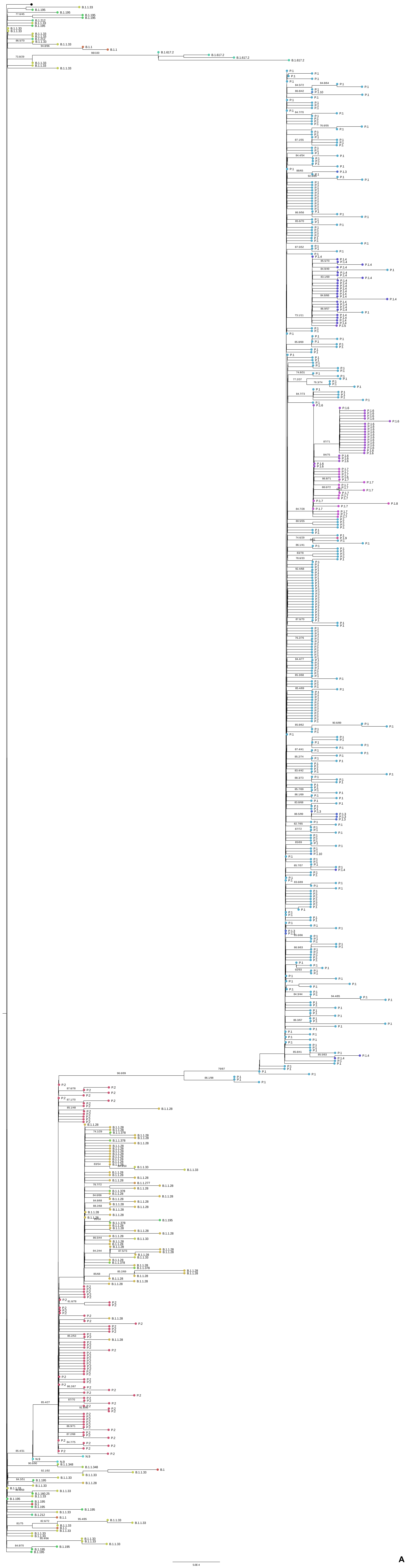
A

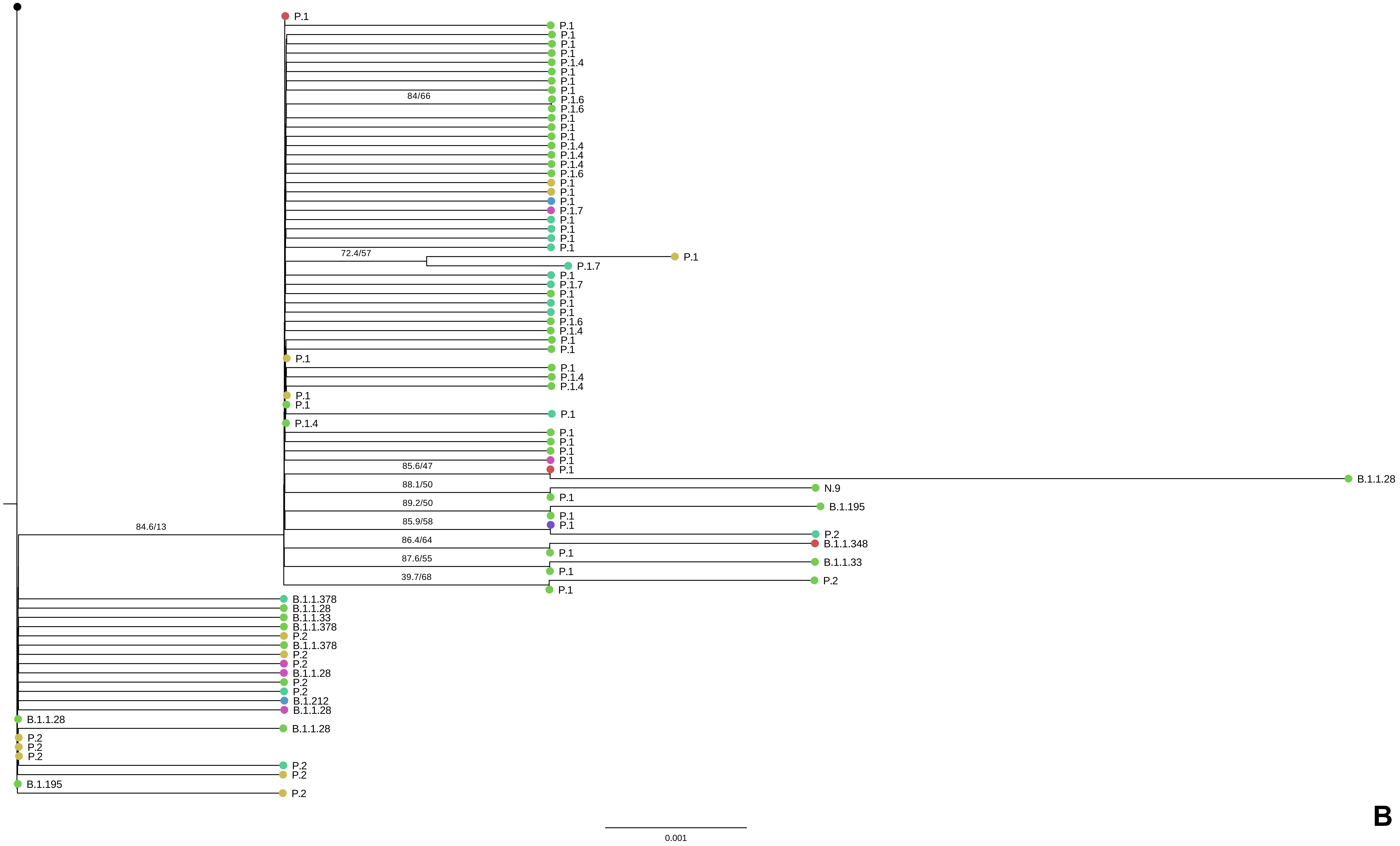

P.1
P.1
P.1
P.1
P.1
P.1.4
P.1
P.1
P.1
84/66
P.1.6
P.1.6
P.1
P.1
P.1
P.1.4
P.1.4
P.1.4
P.1.6
P.1
P.1
P.1
P.1.7
P.1
P.1
P.1
P.1
72.4/57
P.1
P.1.7
P.1
P.1.7
P.1
P.1
P.1
P.1.6
P.1.4
P.1
P.1
P.1
P.1
P.1.4
P.1.4
P.1
P.1
P.1
P.1.4
P.1
P.1
P.1
P.1
85.6/47
P.1
B.1.1.28
88.1/50
N.9
P.1
89.2/50
B.1.195
P.1
85.9/58
P.1
84.6/13
P.2
86.4/64
B.1.1.348
P.1
87.6/55
B.1.1.33
P.1
39.7/68
P.2
P.1
B.1.1.378
B.1.1.28
B.1.1.33
B.1.1.378
P.2
B.1.1.378
P.2
P.2
B.1.1.28
P.2
P.2
B.1.212
B.1.1.28
B.1.1.28
B.1.1.28
P.2
P.2
P.2
P.2
P.2
B.1.195
P.2
0.001
B

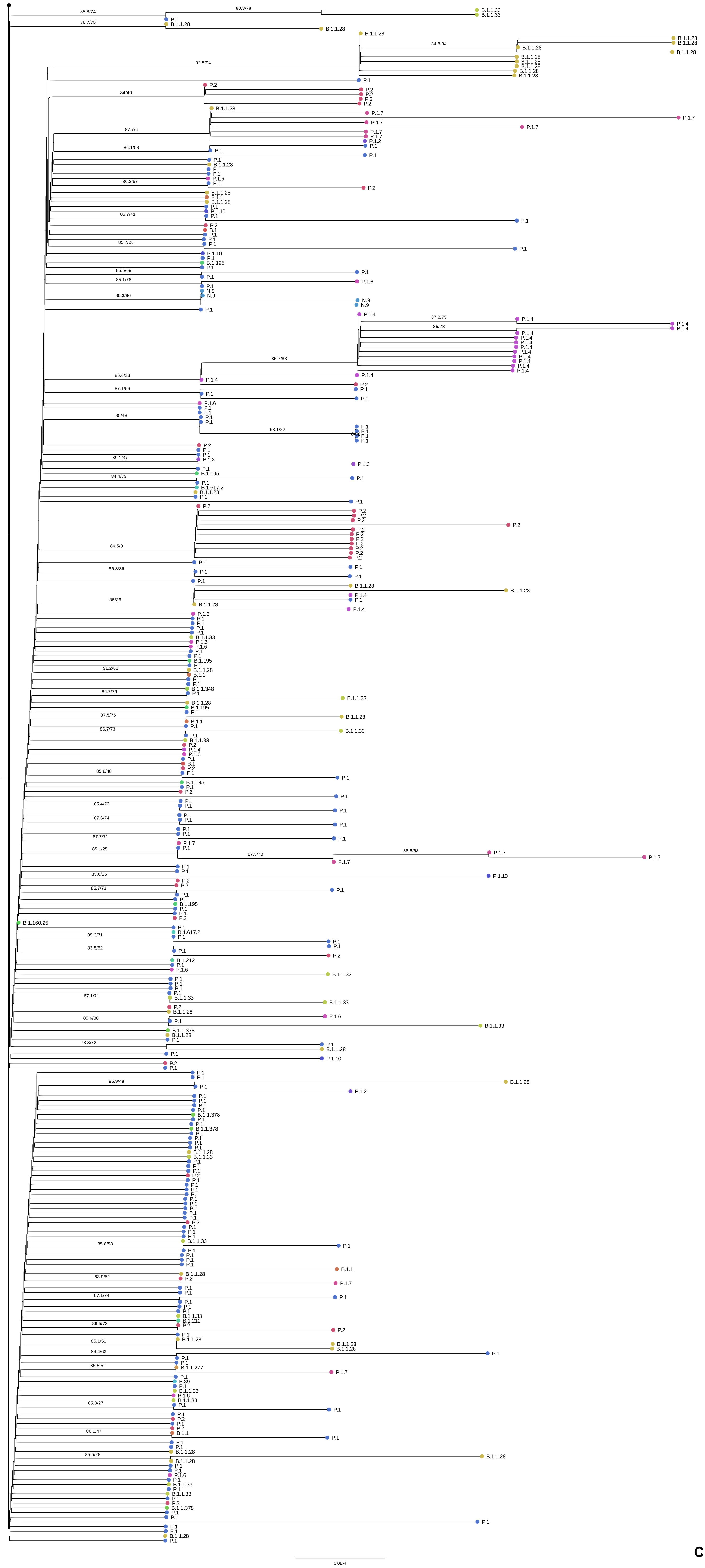


C

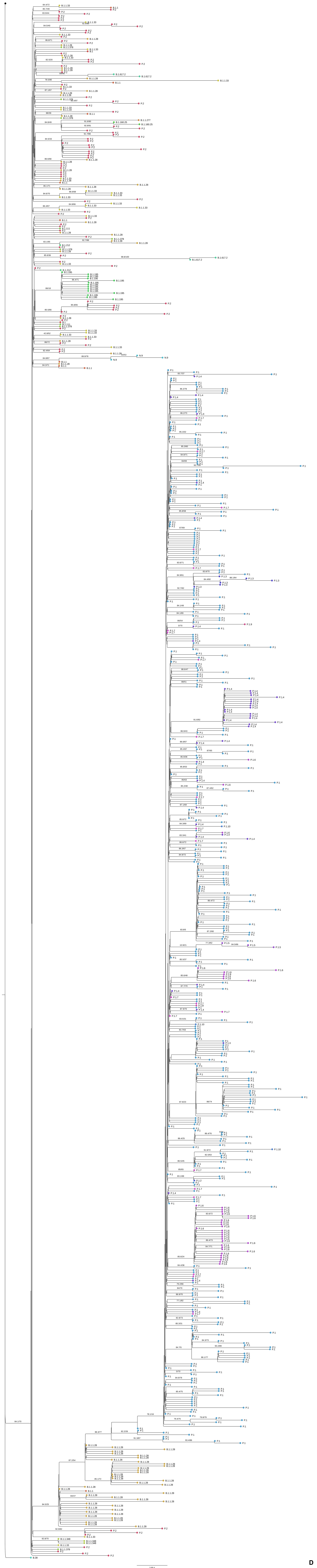

D

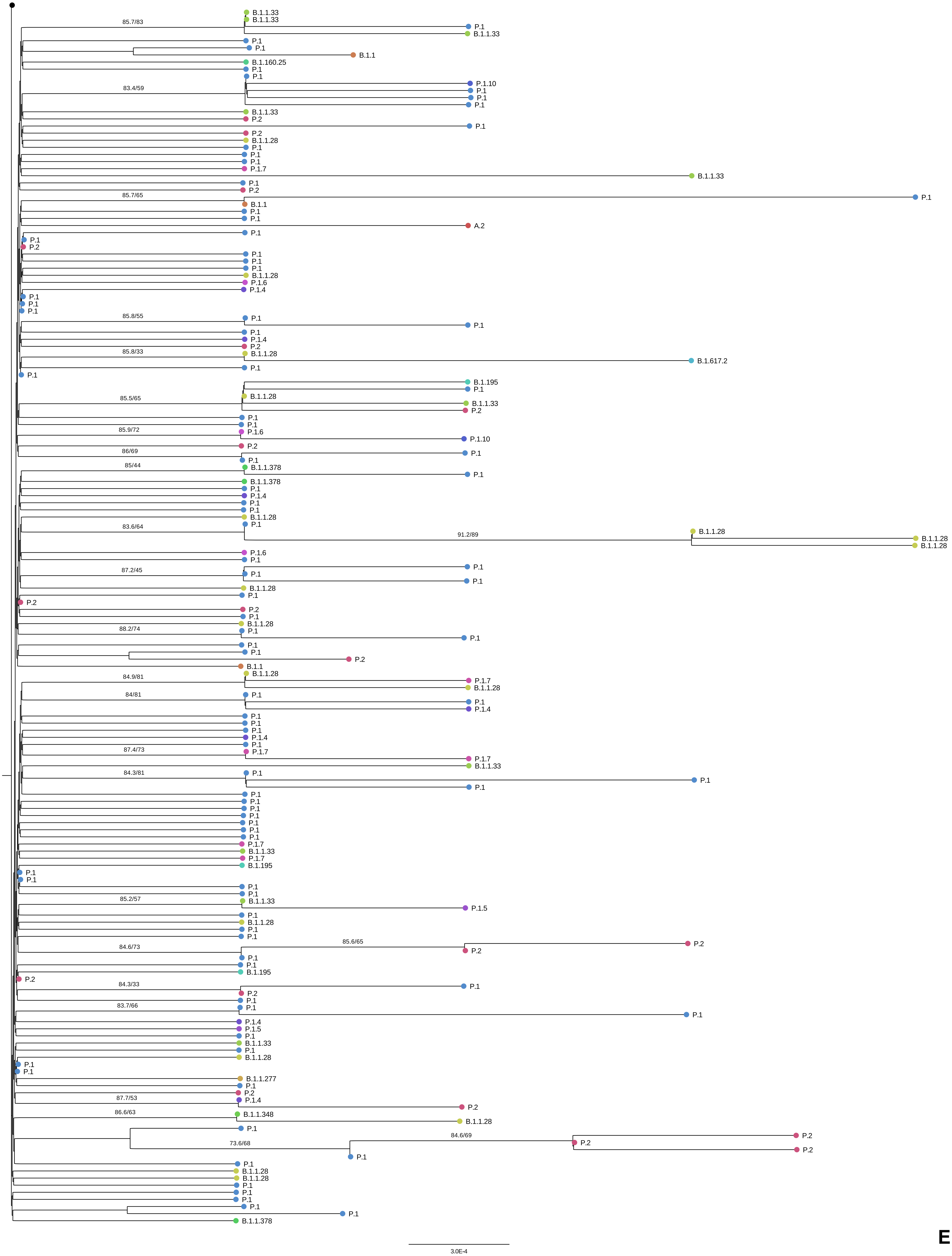
B.1.1.33
B.1.1.33
85.7/83
P.1
B.1.1.33
P.1
P.1
B.1.1
B.1.160.25
P.1
P.1
83.4/59
P.1.10
P.1
P.1
P.1
B.1.1.33
P.2
P.1
P.2
B.1.1.28
P.1
P.1
P.1
P.1.7
B.1.1.33
P.1
P.2
85.7/65
P.1
B.1.1
P.1
P.1
A.2
P.1
P.1
P.2
P.1
P.1
P.1
B.1.1.28
P.1.6
P.1.4
P.1
P.1
P.1
85.8/55
P.1
P.1
P.1
P.1.4
P.2
85.8/33
B.1.1.28
B.1.617.2
P.1
P.1
B.1.195
P.1
85.5/65
B.1.1.28
B.1.1.33
P.2
P.1
P.1
85.9/72
P.1.6
P.1.10
P.2
86/69
P.1
P.1
85/44
B.1.1.378
P.1
B.1.1.378
P.1
P.1.4
P.1
P.1
B.1.1.28
83.6/64
P.1
B.1.1.28
91.2/89
B.1.1.28
B.1.1.28
P.1.6
P.1
P.1
87.2/45
P.1
P.1
B.1.1.28
P.1
P.2
P.2
P.1
B.1.1.28
88.2/74
P.1
P.1
P.1
P.1
P.2
B.1.1
84.9/81
B.1.1.28
P.1.7
B.1.1.28
84/81
P.1
P.1
P.1.4
P.1
P.1
P.1
P.1.4
P.1
87.4/73
P.1.7
P.1.7
B.1.1.33
84.3/81
P.1
P.1
P.1
P.1
P.1
P.1
P.1
P.1
P.1
P.1
P.1.7
B.1.1.33
P.1.7
B.1.195
P.1
P.1
P.1
P.1
85.2/57
B.1.1.33
P.1.5
P.1
B.1.1.28
P.1
P.1
85.6/65
P.2
84.6/73
P.2
P.1
P.1
B.1.195
P.2
84.3/33
P.1
P.2
P.1
83.7/66
P.1
P.1
P.1.4
P.1.5
P.1
B.1.1.33
P.1
B.1.1.28
P.1
P.1
B.1.1.277
P.1
P.2
87.7/53
P.1.4
P.2
86.6/63
B.1.1.348
B.1.1.28
P.1
84.6/69
P.2
73.6/68
P.2
P.2
P.1
P.1
B.1.1.28
B.1.1.28
P.1
P.1
P.1
P.1
P.1
B.1.1.378
3.0E-4

**E**

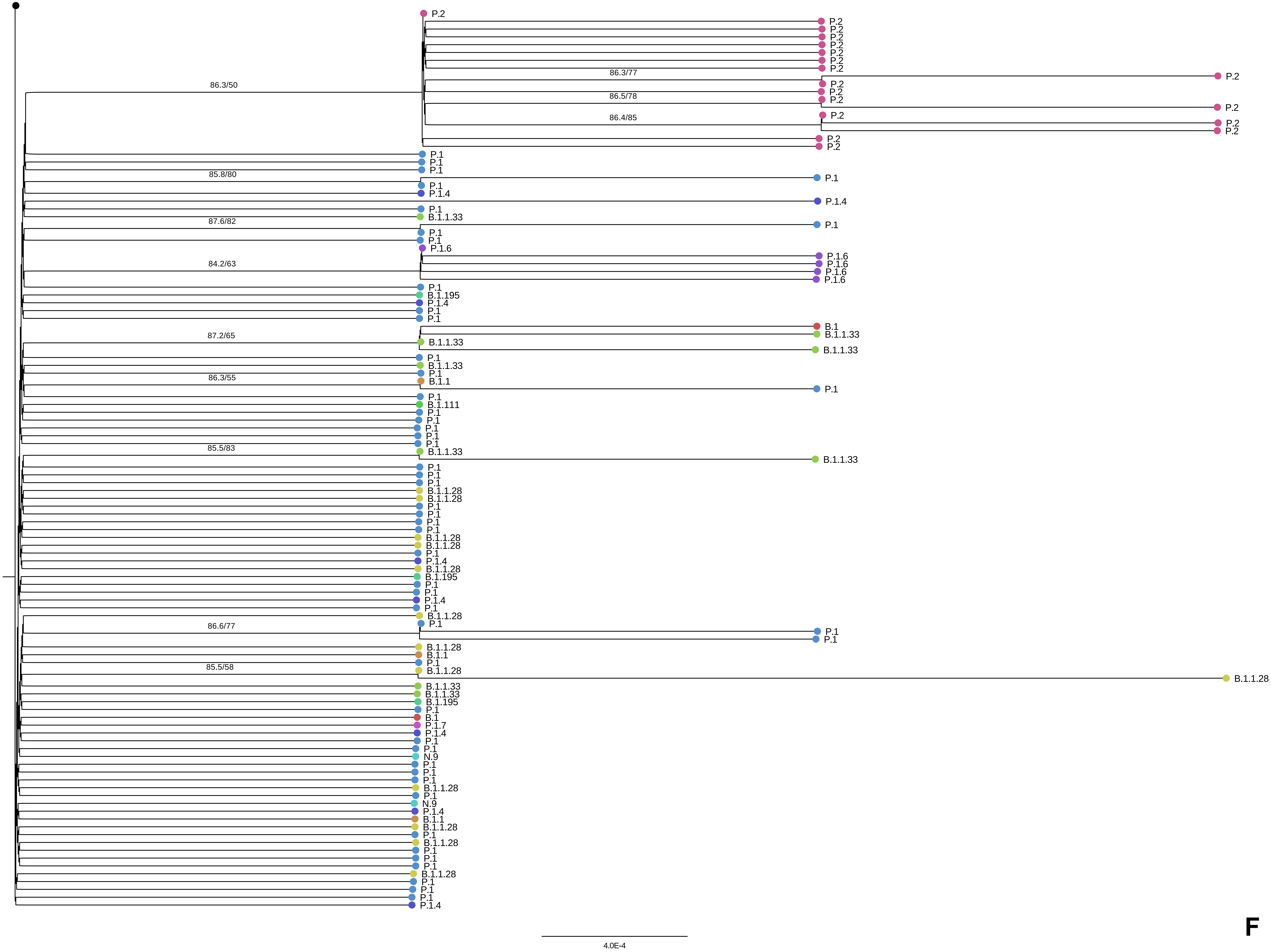
P.2
P.2
P.2
P.2
P.2
P.2
P.2
P.2
86.3/77
P.2
P.2
P.2
86.5/78
P.2
P.2
86.4/85
P.2
P.2
P.2
P.2
P.2
86.3/50
P.1
P.1
P.1
85.8/80
P.1
P.1
P.1.4
P.1.4
P.1
B.1.1.33
87.6/82
P.1
P.1
P.1
P.1.6
84.2/63
P.1.6
P.1.6
P.1.6
P.1.6
P.1
B.1.195
P.1.4
P.1
P.1
B.1
B.1.1.33
87.2/65
B.1.1.33
B.1.1.33
P.1
B.1.1.33
P.1
86.3/55
B.1.1
P.1
P.1
B.1.111
P.1
P.1
P.1
P.1
P.1
85.5/83
B.1.1.33
B.1.1.33
P.1
P.1
P.1
B.1.1.28
B.1.1.28
P.1
P.1
P.1
P.1
B.1.1.28
B.1.1.28
P.1
P.1.4
B.1.1.28
B.1.195
P.1
P.1
P.1.4
P.1
B.1.1.28
86.6/77
P.1
P.1
P.1
B.1.1.28
B.1.1
P.1
85.5/58
B.1.1.28
B.1.1.28
B.1.1.33
B.1.1.33
B.1.195
P.1
B.1
P.1.7
P.1.4
P.1
P.1
N.9
P.1
P.1
P.1
B.1.1.28
P.1
N.9
P.1.4
B.1.1
B.1.1.28
P.1
B.1.1.28
P.1
P.1
P.1
B.1.1.28
P.1
P.1
P.1
P.1.4
4.0E-4
F

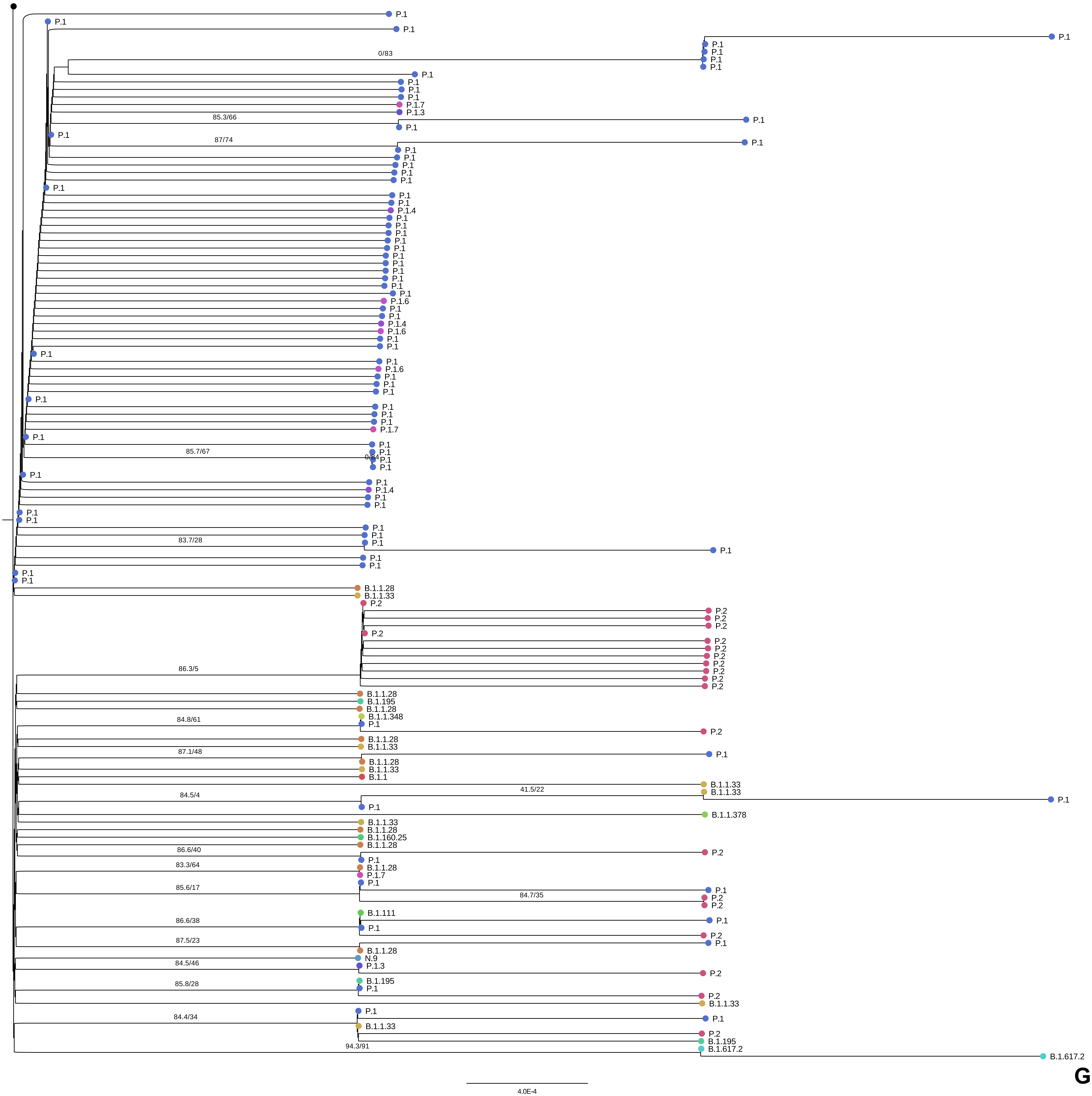

P.1
P.1
P.1
P.1
0/83
P.1
P.1
P.1
P.1
P.1
P.1
P.1
P.1.7
P.1.3
85.3/66
P.1
P.1
P.1
87/74
P.1
P.1
P.1
P.1
P.1
P.1
P.1
P.1
P.1
P.1.4
P.1
P.1
P.1
P.1
P.1
P.1
P.1
P.1
P.1
P.1
P.1
P.1.6
P.1
P.1
P.1.4
P.1.6
P.1
P.1
P.1
P.1
P.1.6
P.1
P.1
P.1
P.1
P.1
P.1
P.1
P.1.7
P.1
P.1
85.7/67
P.1
0/84
P.1
P.1
P.1
P.1
P.1.4
P.1
P.1
P.1
P.1
P.1
P.1
83.7/28
P.1
P.1
P.1
P.1
P.1
P.1
B.1.1.28
B.1.1.33
P.2
P.2
P.2
P.2
P.2
P.2
P.2
P.2
P.2
P.2
86.3/5
P.2
P.2
B.1.1.28
B.1.195
B.1.1.28
84.8/61
B.1.1.348
P.1
P.2
B.1.1.28
B.1.1.33
87.1/48
P.1
B.1.1.28
B.1.1.33
B.1.1
B.1.1.33
41.5/22
B.1.1.33
84.5/4
P.1
P.1
B.1.1.378
B.1.1.33
B.1.1.28
B.1.160.25
B.1.1.28
86.6/40
P.2
P.1
83.3/64
B.1.1.28
P.1.7
85.6/17
P.1
P.1
84.7/35
P.2
P.2
B.1.111
86.6/38
P.1
P.1
P.2
87.5/23
P.1
B.1.1.28
N.9
84.5/46
P.1.3
P.2
85.8/28
B.1.195
P.1
P.2
B.1.1.33
P.1
84.4/34
P.1
B.1.1.33
P.2
B.1.195
94.3/91
B.1.617.2
B.1.617.2
4.0E-4


G

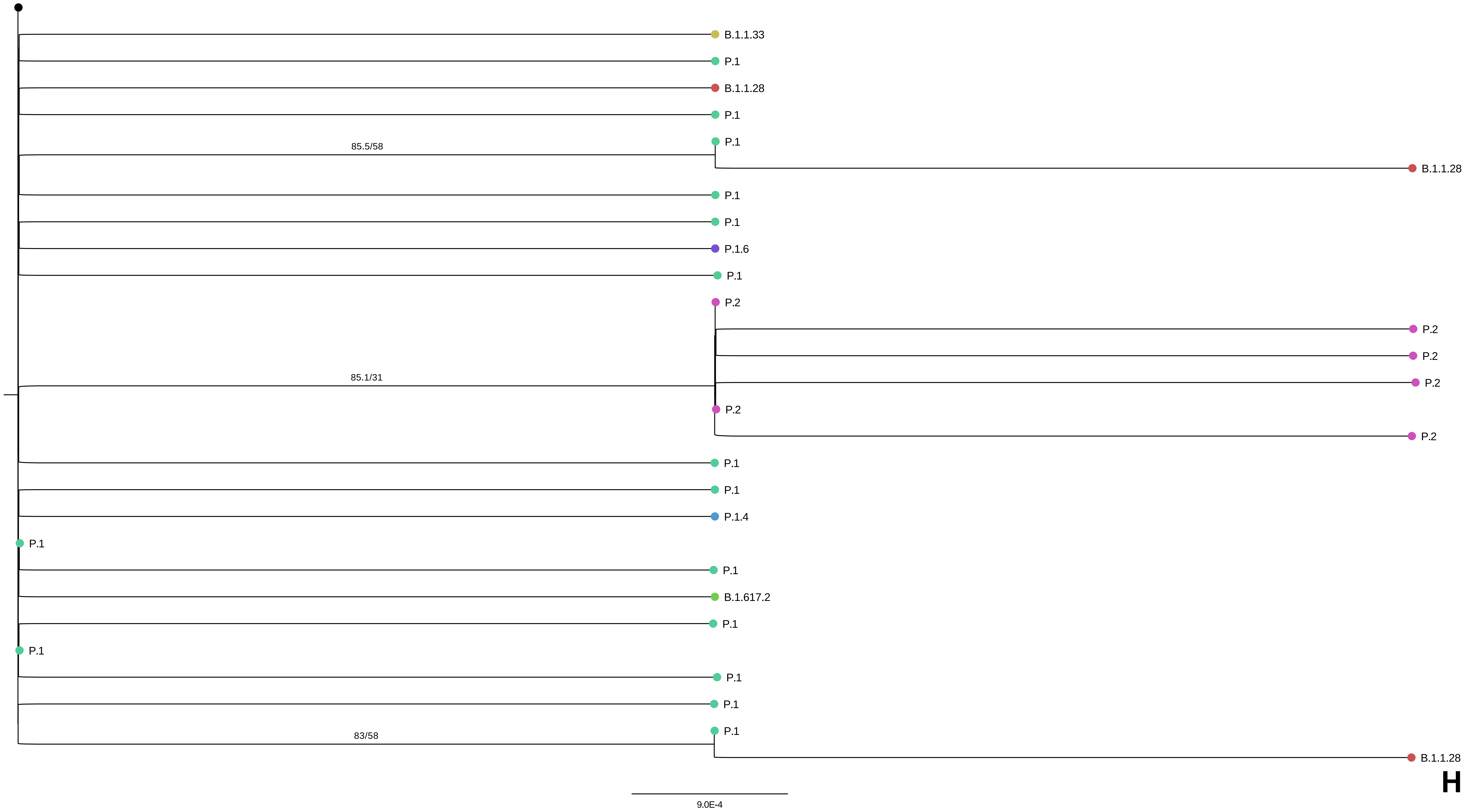

B.1.1.33
P.1
B.1.1.28
P.1
85.5/58
P.1
B.1.1.28
P.1
P.1
P.1.6
P.1
P.2
P.2
P.2
85.1/31
P.2
P.2
P.2
P.1
P.1
P.1.4
P.1
P.1
B.1.617.2
P.1
P.1
P.1
P.1
83/58
P.1
B.1.1.28
H
9.0E-4

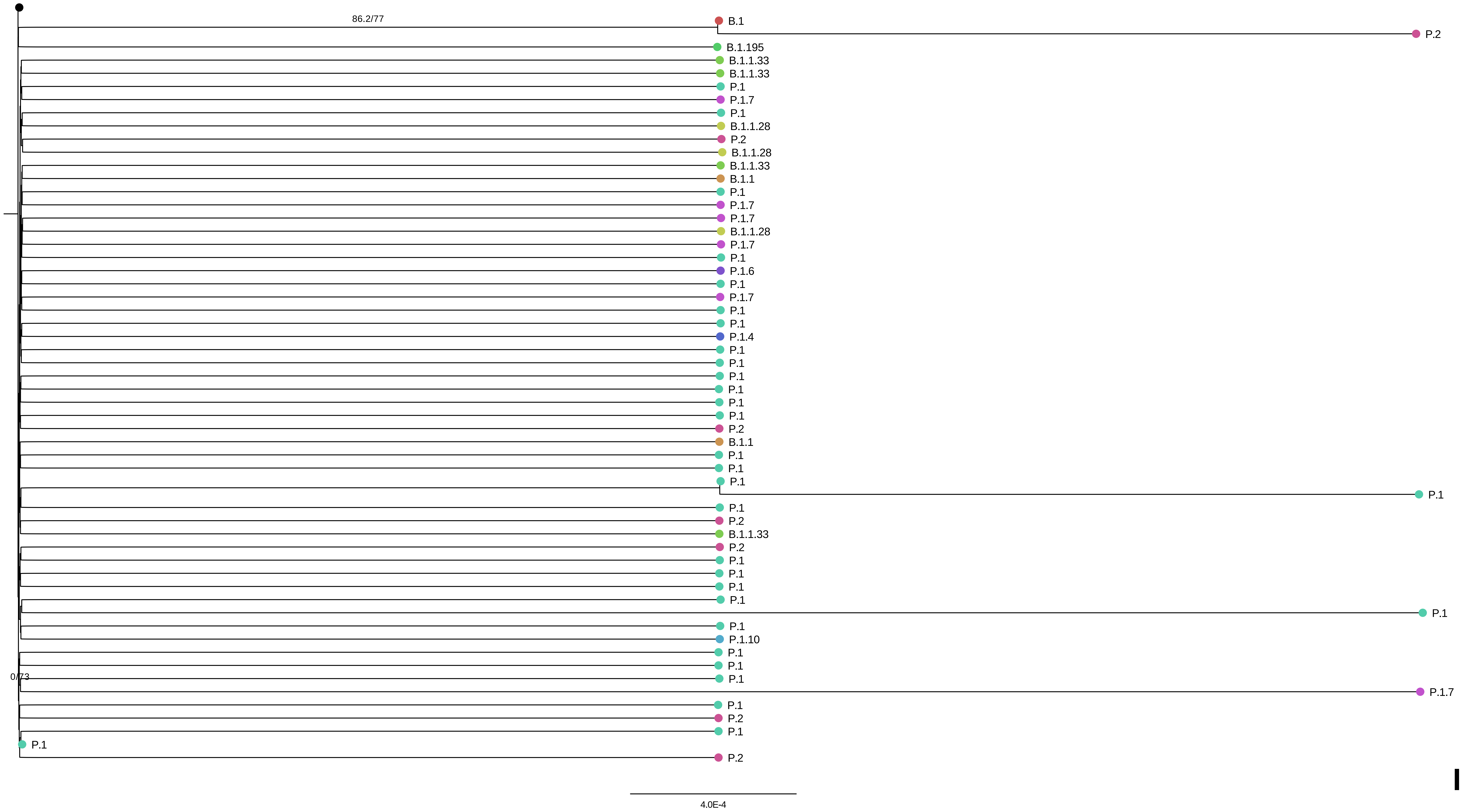
86.2/77
B.1
P.2
B.1.195
B.1.1.33
B.1.1.33
P.1
P.1.7
P.1
B.1.1.28
P.2
B.1.1.28
B.1.1.33
B.1.1
P.1
P.1.7
P.1.7
B.1.1.28
P.1.7
P.1
P.1.6
P.1
P.1.7
P.1
P.1
P.1.4
P.1
P.1
P.1
P.1
P.1
P.1
P.2
B.1.1
P.1
P.1
P.1
P.1
P.1
P.2
B.1.1.33
P.2
P.1
P.1
P.1
P.1
P.1
P.1
P.1.10
P.1
P.1
0/73
P.1
P.1.7
P.1
P.2
P.1
P.1
P.2
4.0E-4

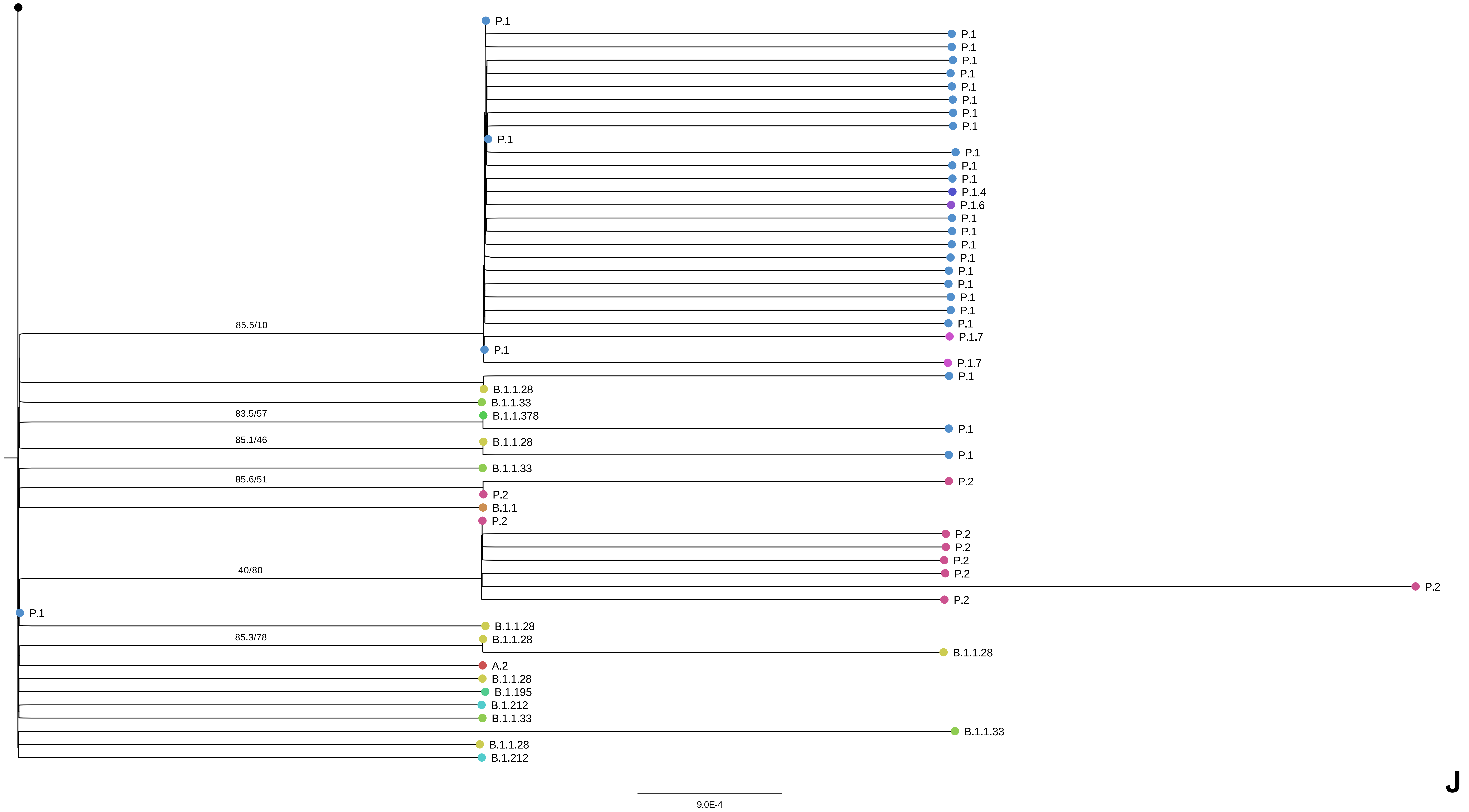

P.1
P.1.4
P.1.6
P.1.7
85.5/10
B.1.1.28
B.1.1.33
83.5/57
B.1.1.378
85.1/46
85.6/51
P.2
B.1.1
40/80
85.3/78
A.2
B.1.195
B.1.212
9.0E-4
J

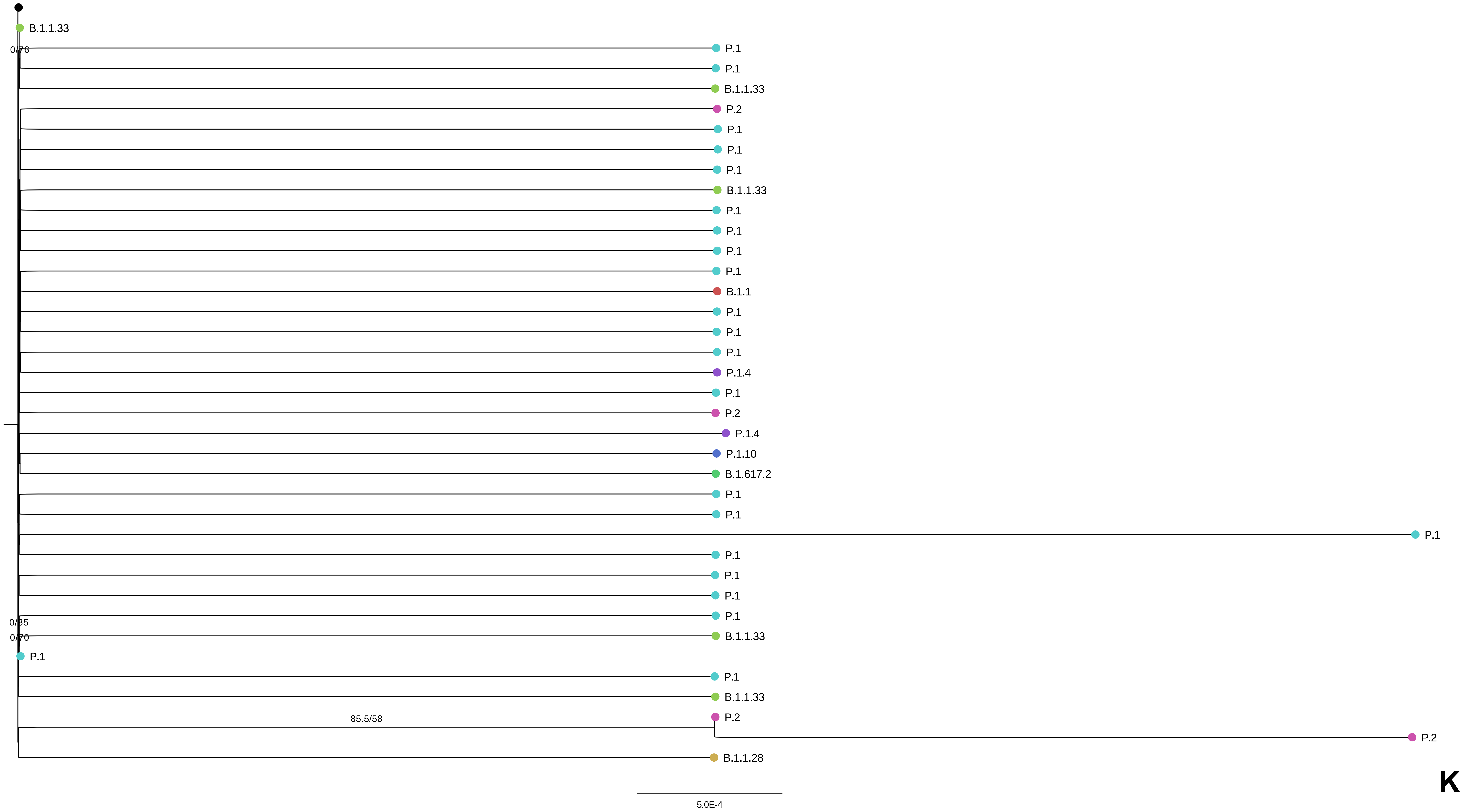

B.1.1.33
0/76
P.1
P.1
B.1.1.33
P.2
P.1
P.1
P.1
B.1.1.33
P.1
P.1
P.1
P.1
B.1.1
P.1
P.1
P.1
P.1.4
P.1
P.2
P.1.4
P.1.10
B.1.617.2
P.1
P.1
P.1
P.1
P.1
P.1
P.1
0/35
0/70
B.1.1.33
P.1
P.1
B.1.1.33
85.5/58
P.2
P.2
B.1.1.28
5.0E-4
K
