## Supplementary File 1 for "Comparative genomics and characterization of SARS-CoV-2 P.1 (Gamma) Variant of Concern (VOC) from Amazonas, Brazil"

# 38966

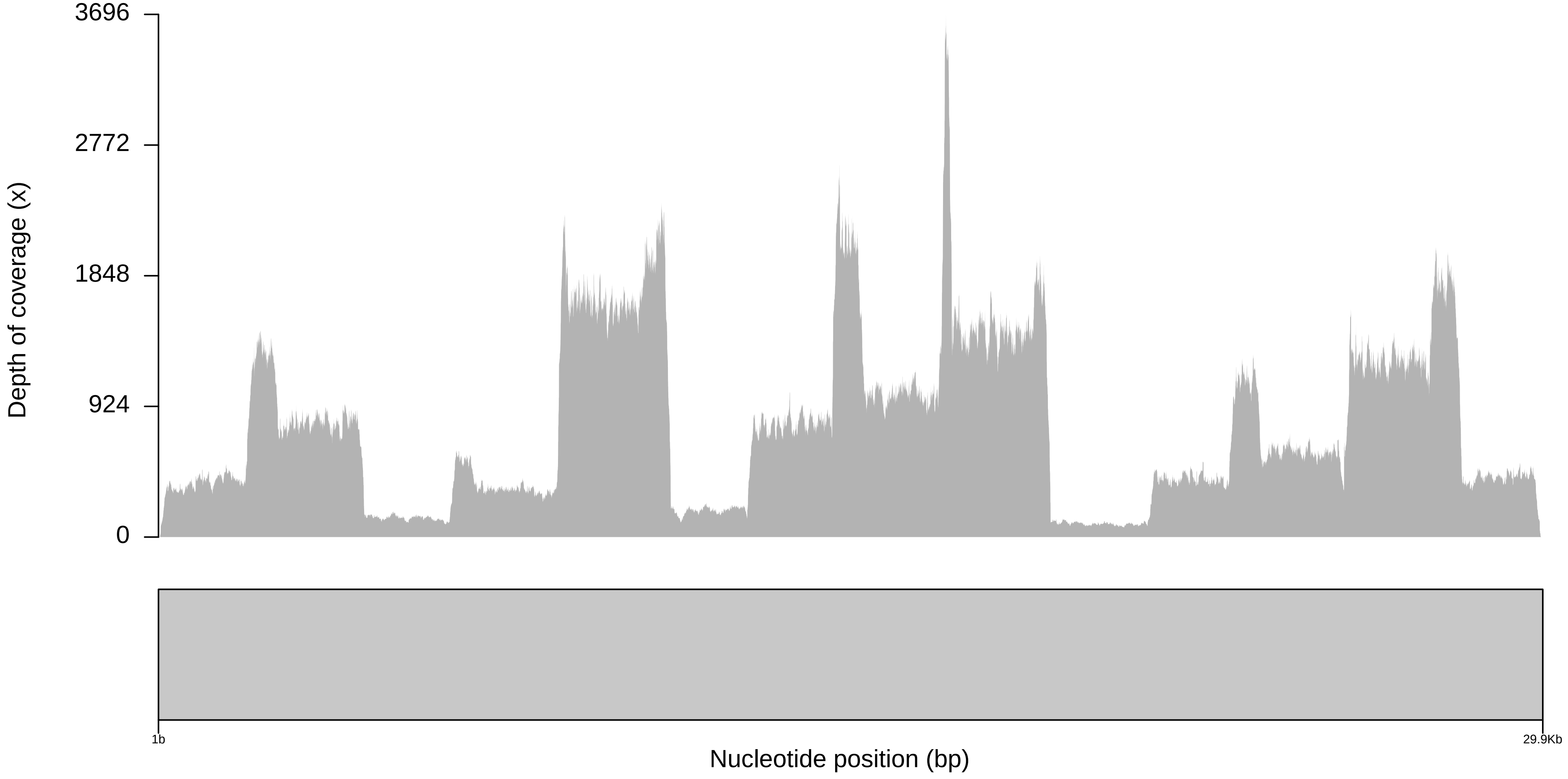

3696
2772
1848
924
0
Depth of coverage (x)
1b
29.9Kb
Nucleotide position (bp)

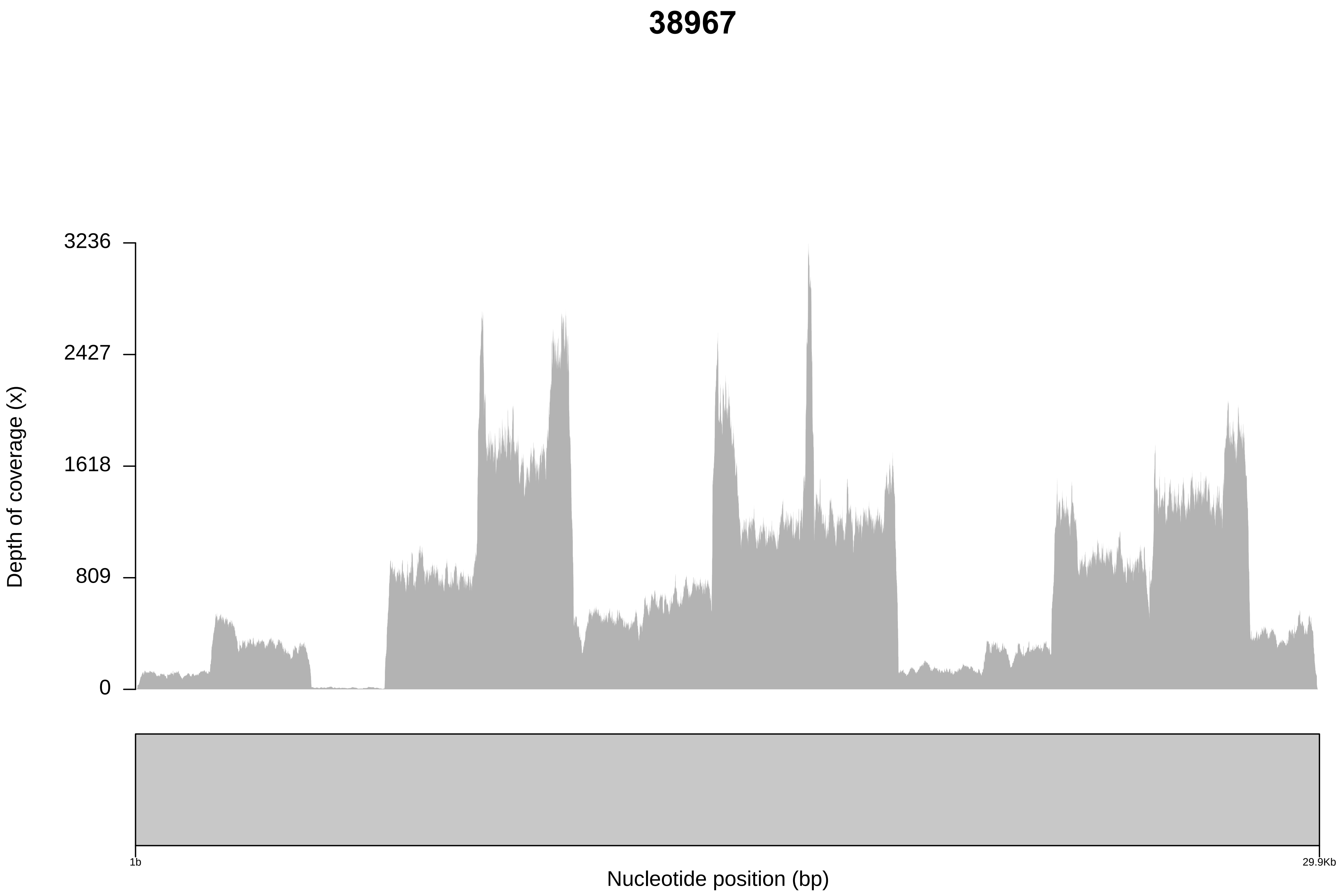
38967
3236
2427
1618
809
0
Depth of coverage (x)
1b
29.9Kb
Nucleotide position (bp)

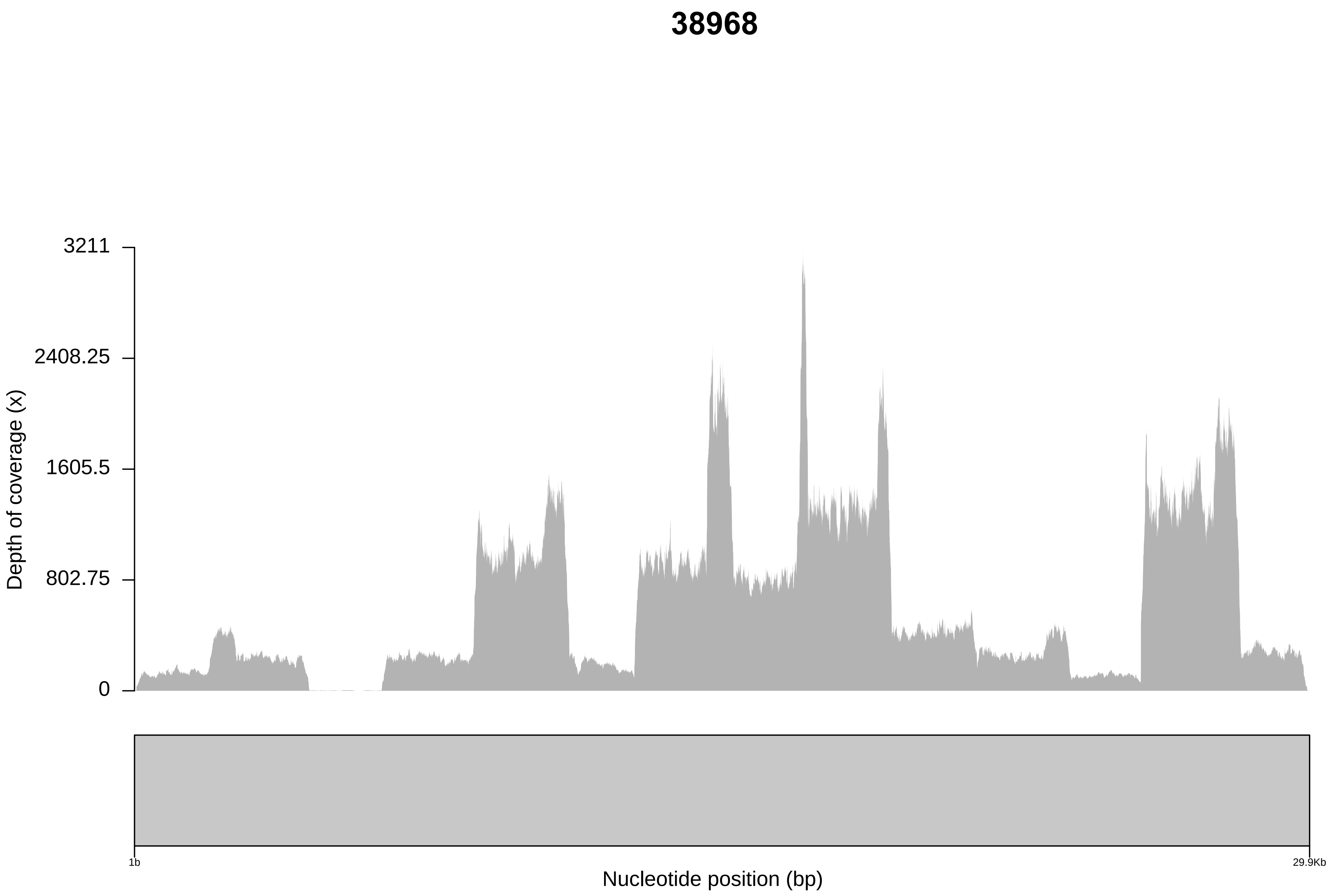
38968
3211
2408.25
1605.5
802.75
0
Depth of coverage (x)
1b
29.9Kb
Nucleotide position (bp)

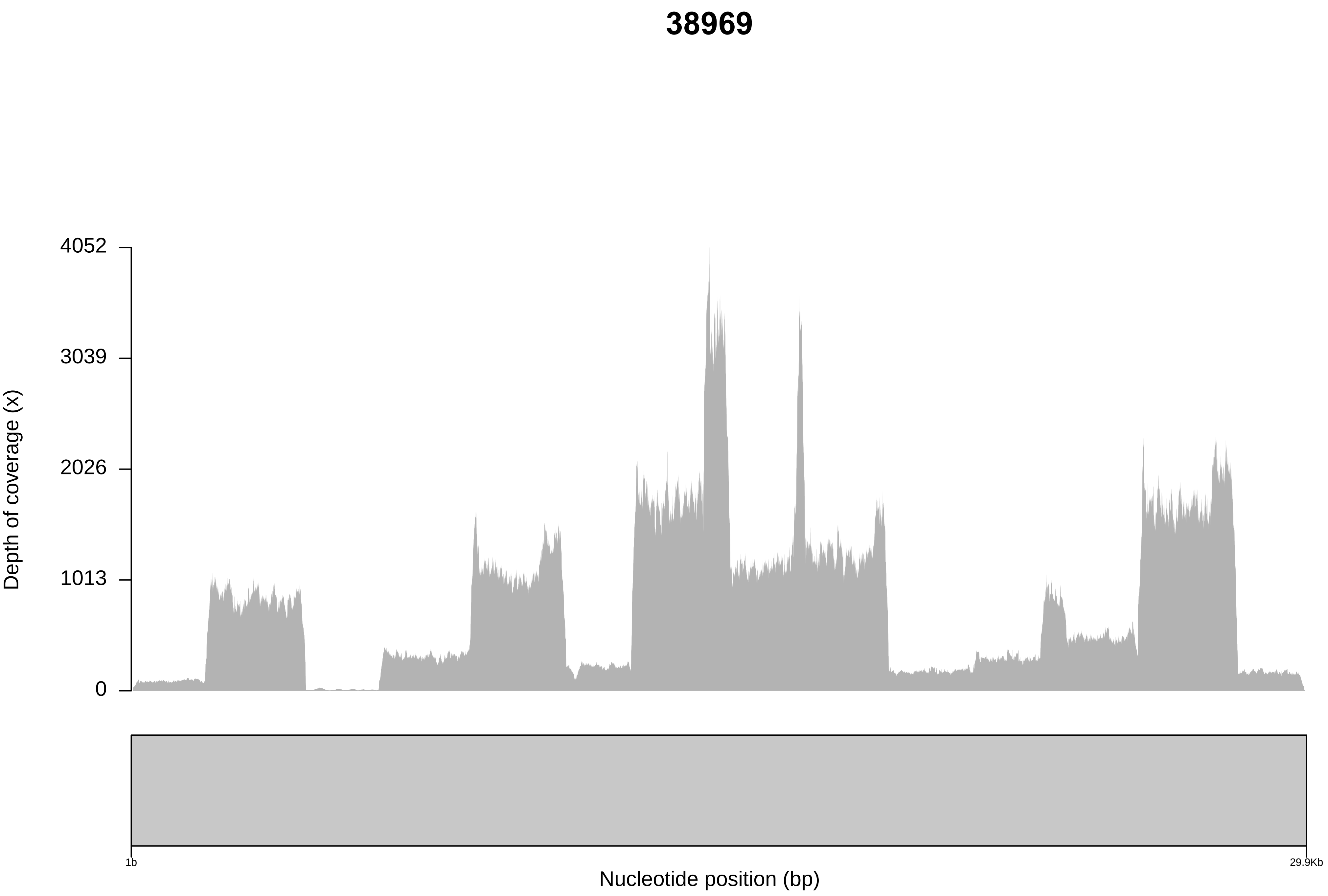
38969
4052
3039
2026
1013
0
Depth of coverage (x)
1b
29.9Kb
Nucleotide position (bp)

# 39428

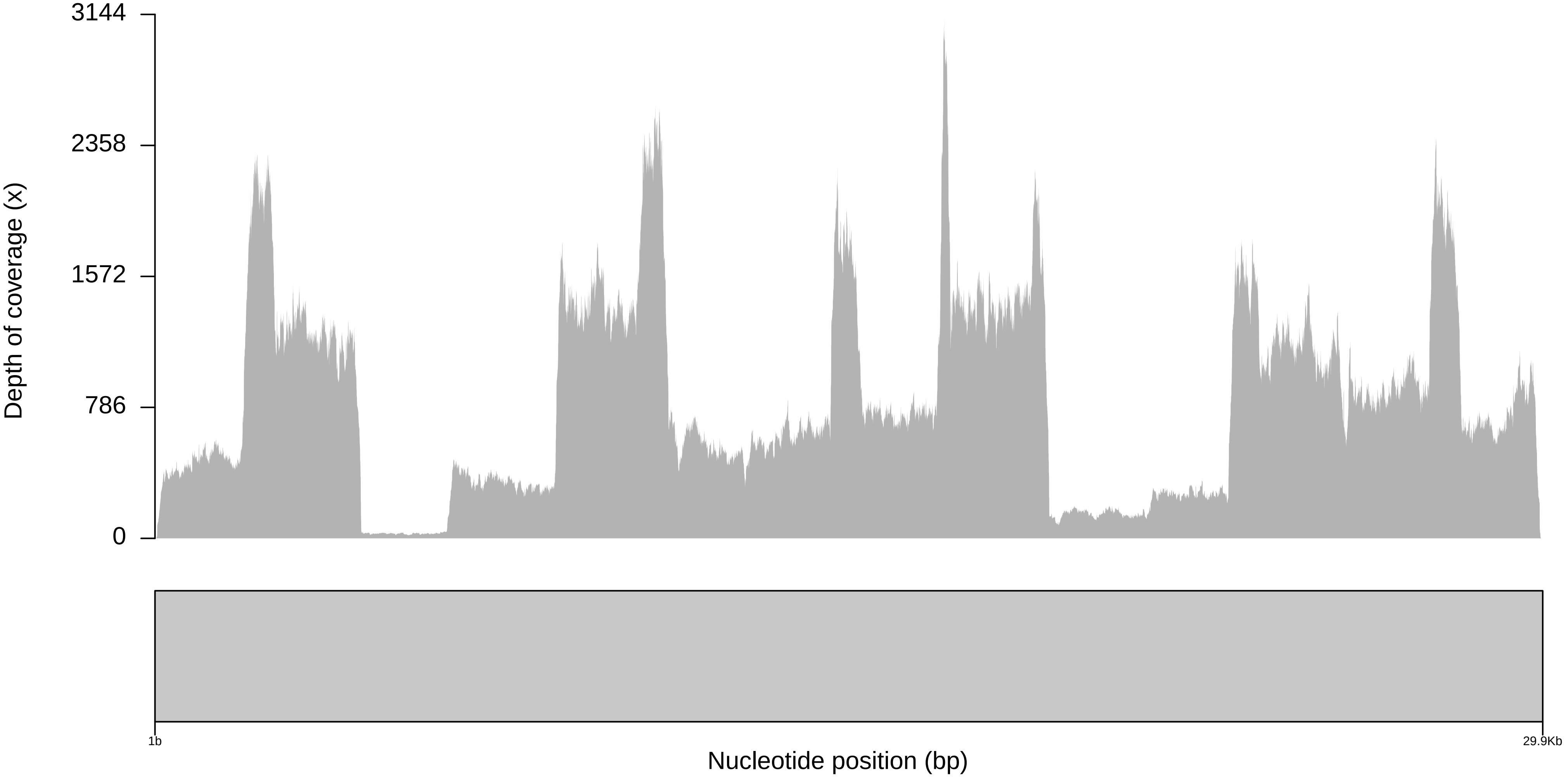
3144
2358
1572
786
0
Depth of coverage (x)
1b
29.9Kb
Nucleotide position (bp)

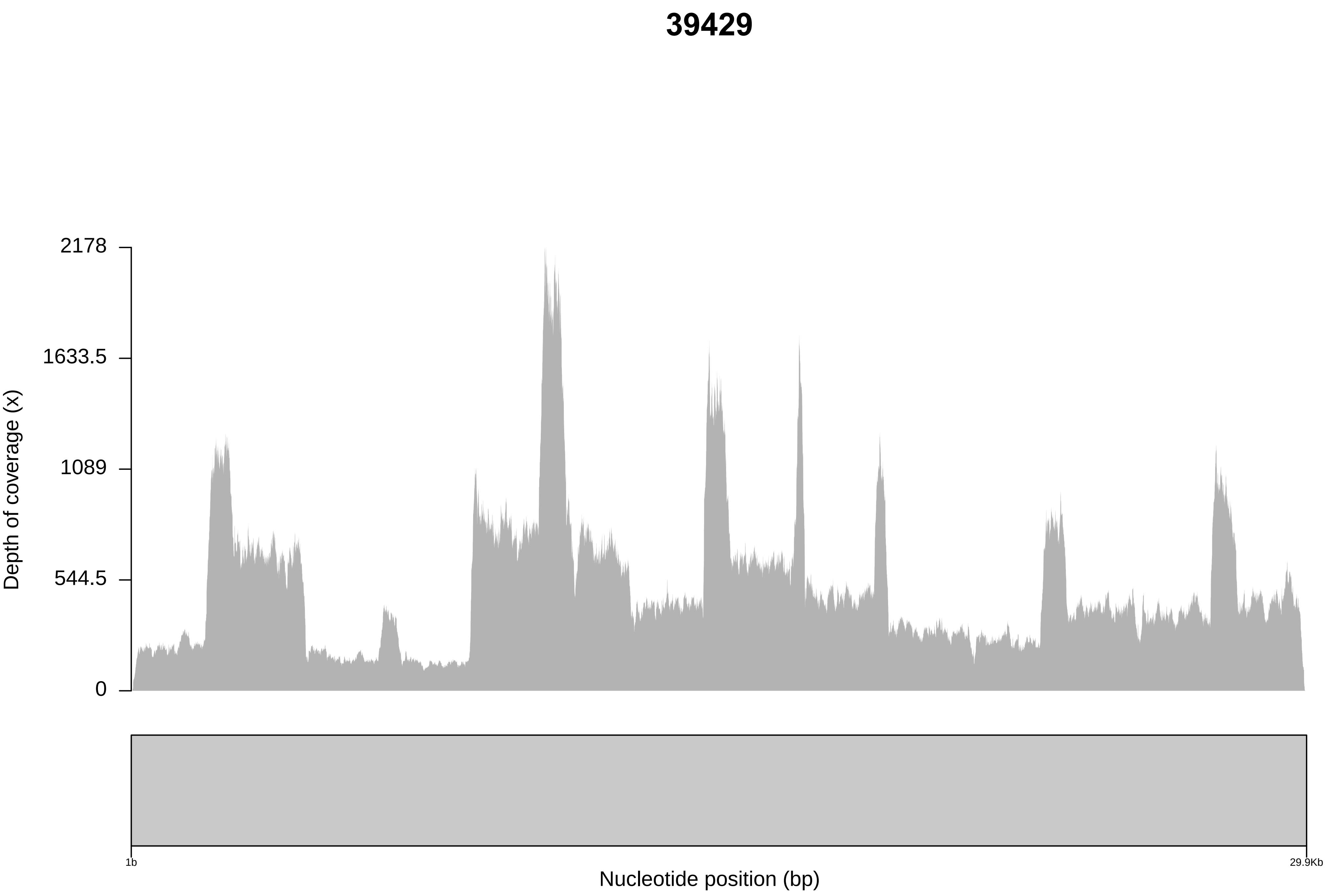

39429
2178
1633.5
1089
544.5
0
Depth of coverage (x)
1b
29.9Kb
Nucleotide position (bp)

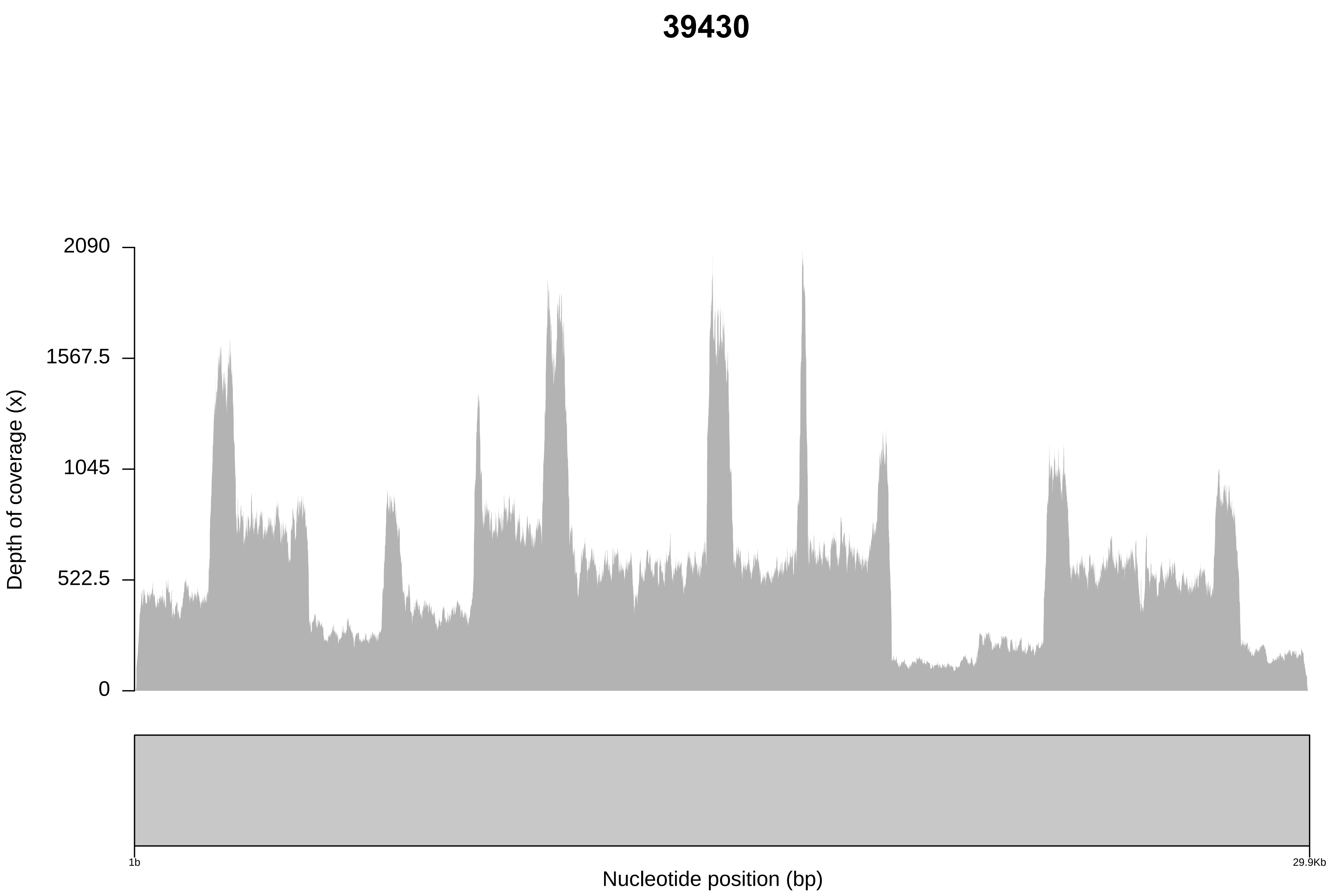
39430
Depth of coverage (x)
2090
1567.5
1045
522.5
0
1b
29.9Kb
Nucleotide position (bp)

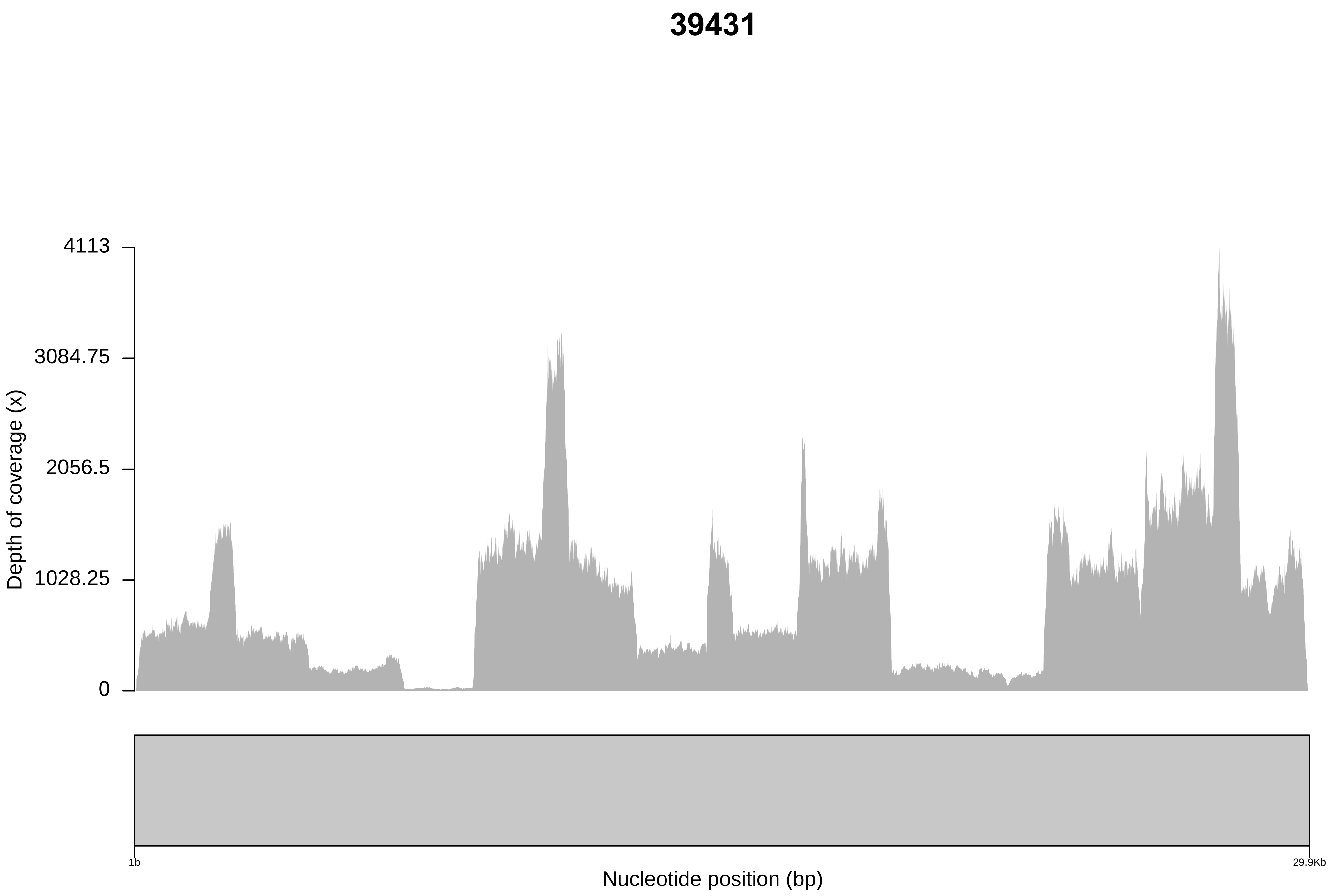
39431
4113
3084.75
2056.5
1028.25
0
Depth of coverage (x)
1b
29.9Kb
Nucleotide position (bp)

39432

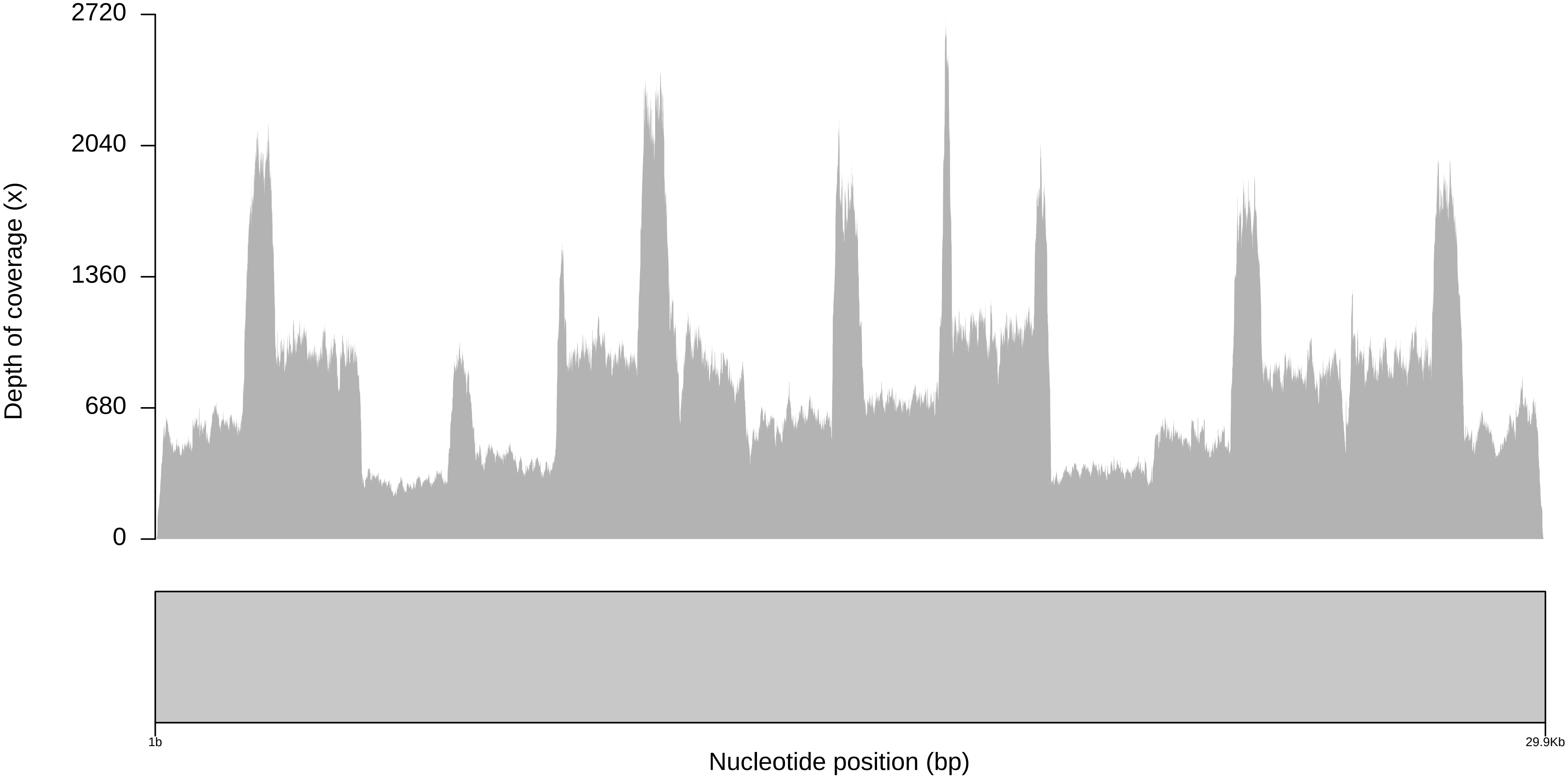

2720
2040
1360
680
0
Depth of coverage (x)
1b
29.9Kb
Nucleotide position (bp)

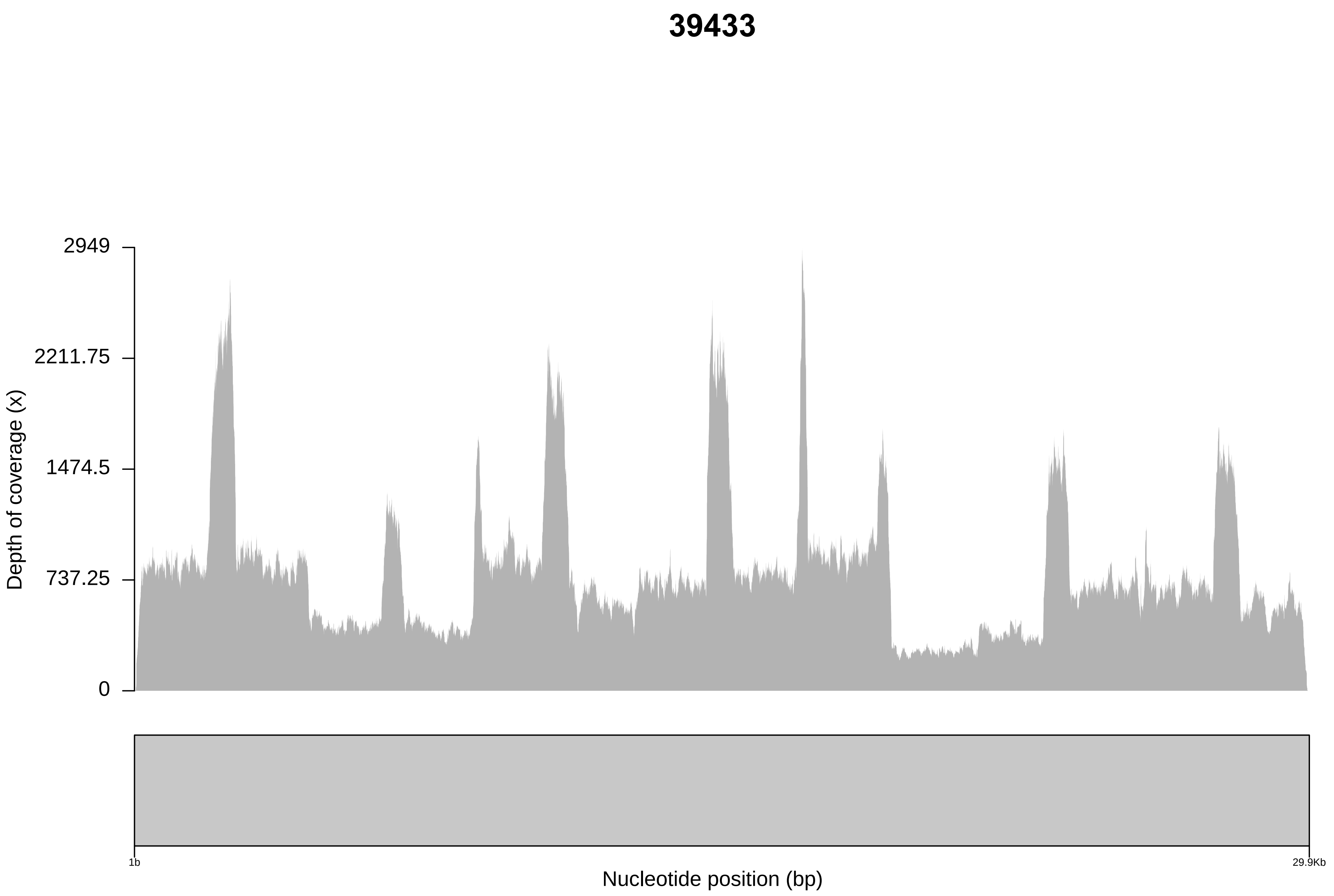

39433
2949
2211.75
1474.5
737.25
0
Depth of coverage (x)
1b
29.9Kb
Nucleotide position (bp)

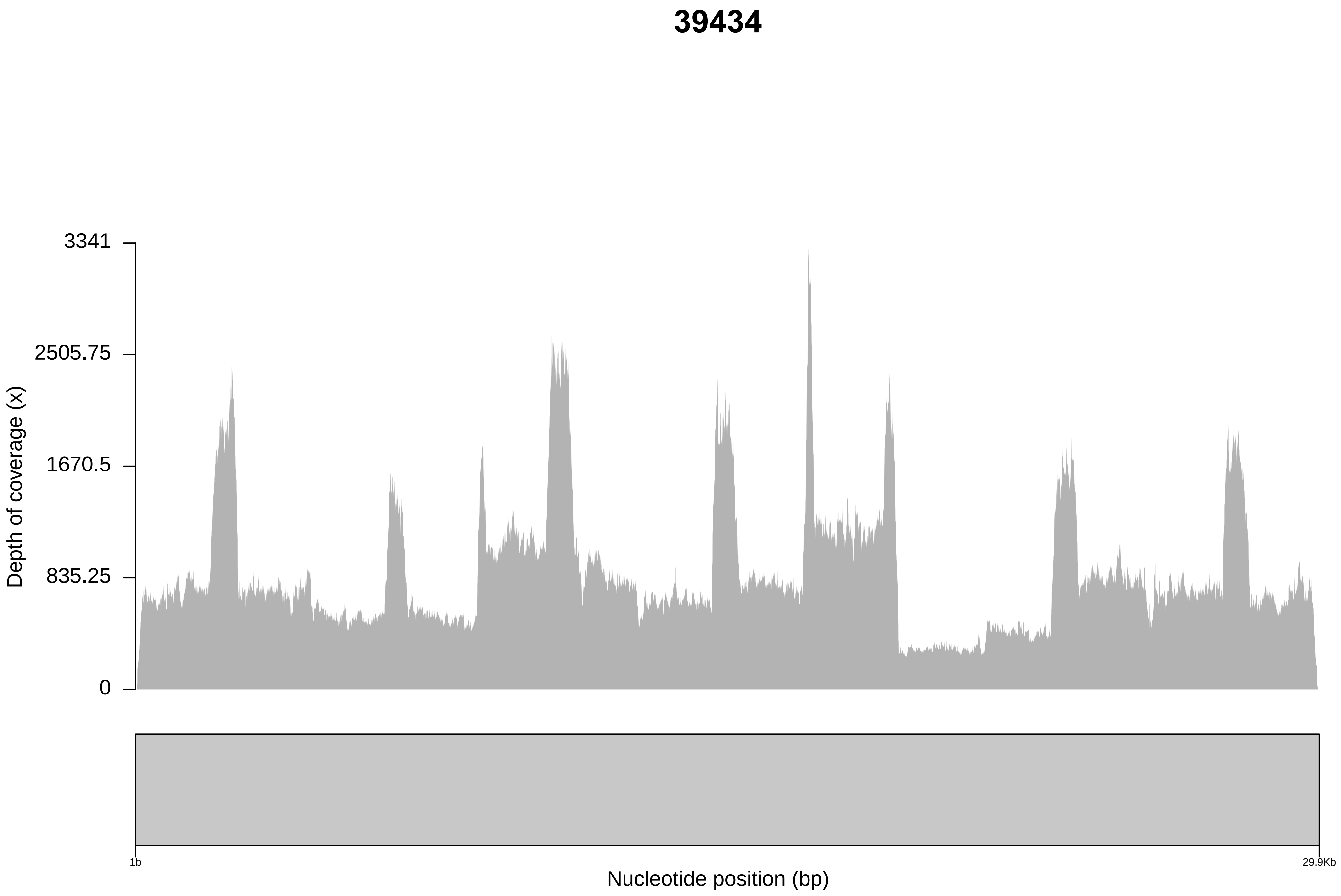

39434
3341
2505.75
1670.5
835.25
0
Depth of coverage (x)
1b
29.9Kb
Nucleotide position (bp)

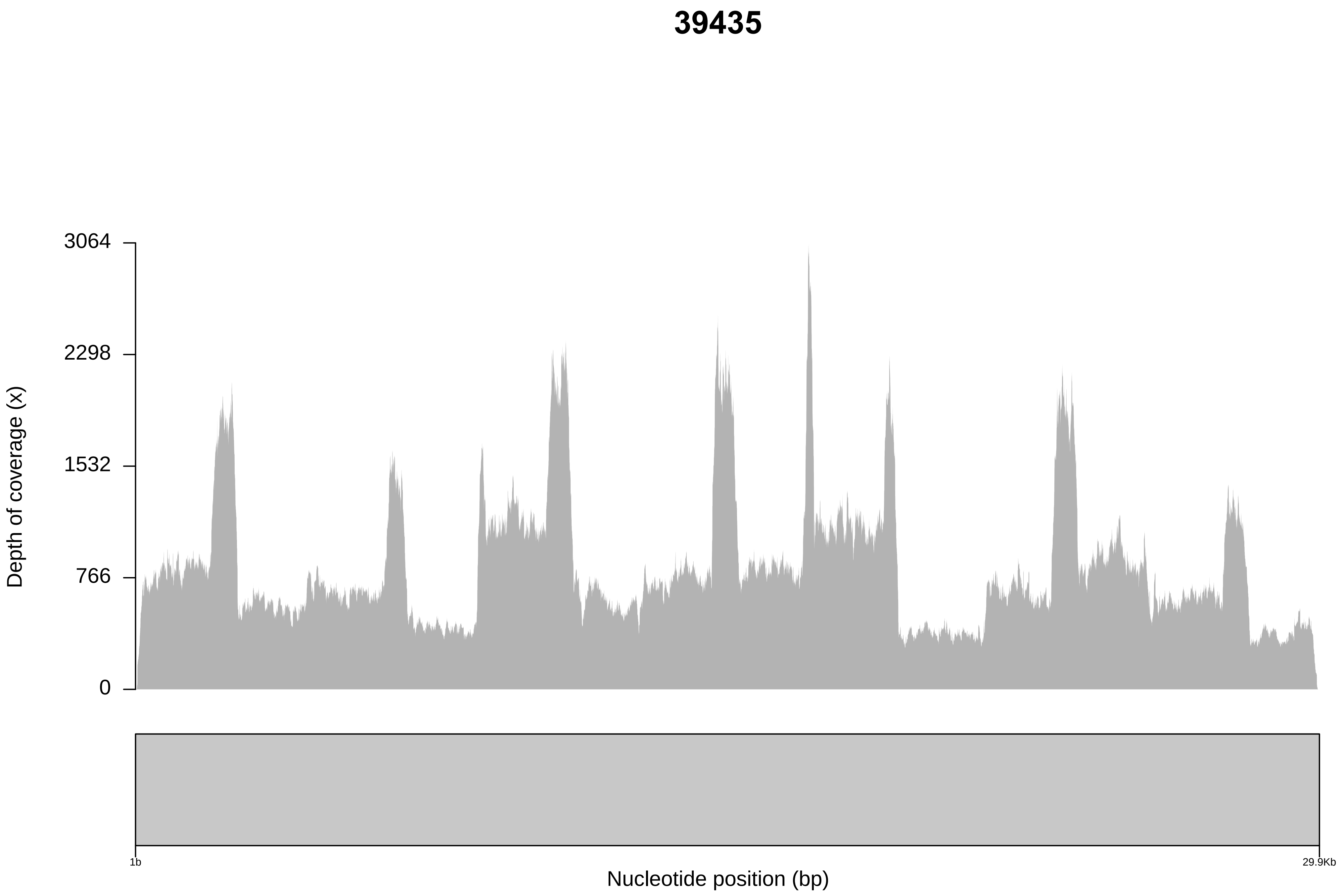

39435
3064
2298
1532
766
0
Depth of coverage (x)
1b
29.9Kb
Nucleotide position (bp)

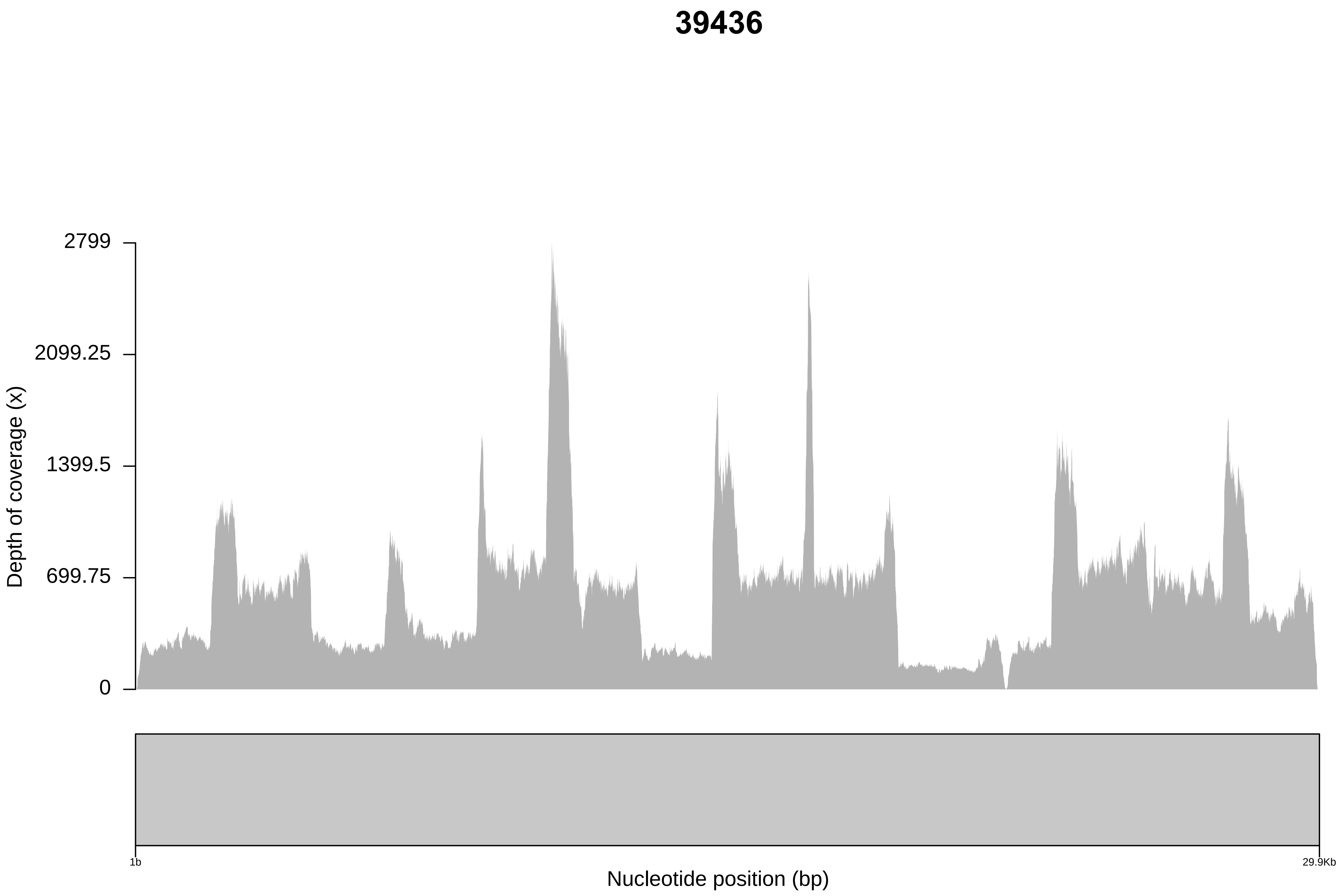
39436
2799
2099.25
1399.5
699.75
0
Depth of coverage (x)
1b
29.9Kb
Nucleotide position (bp)

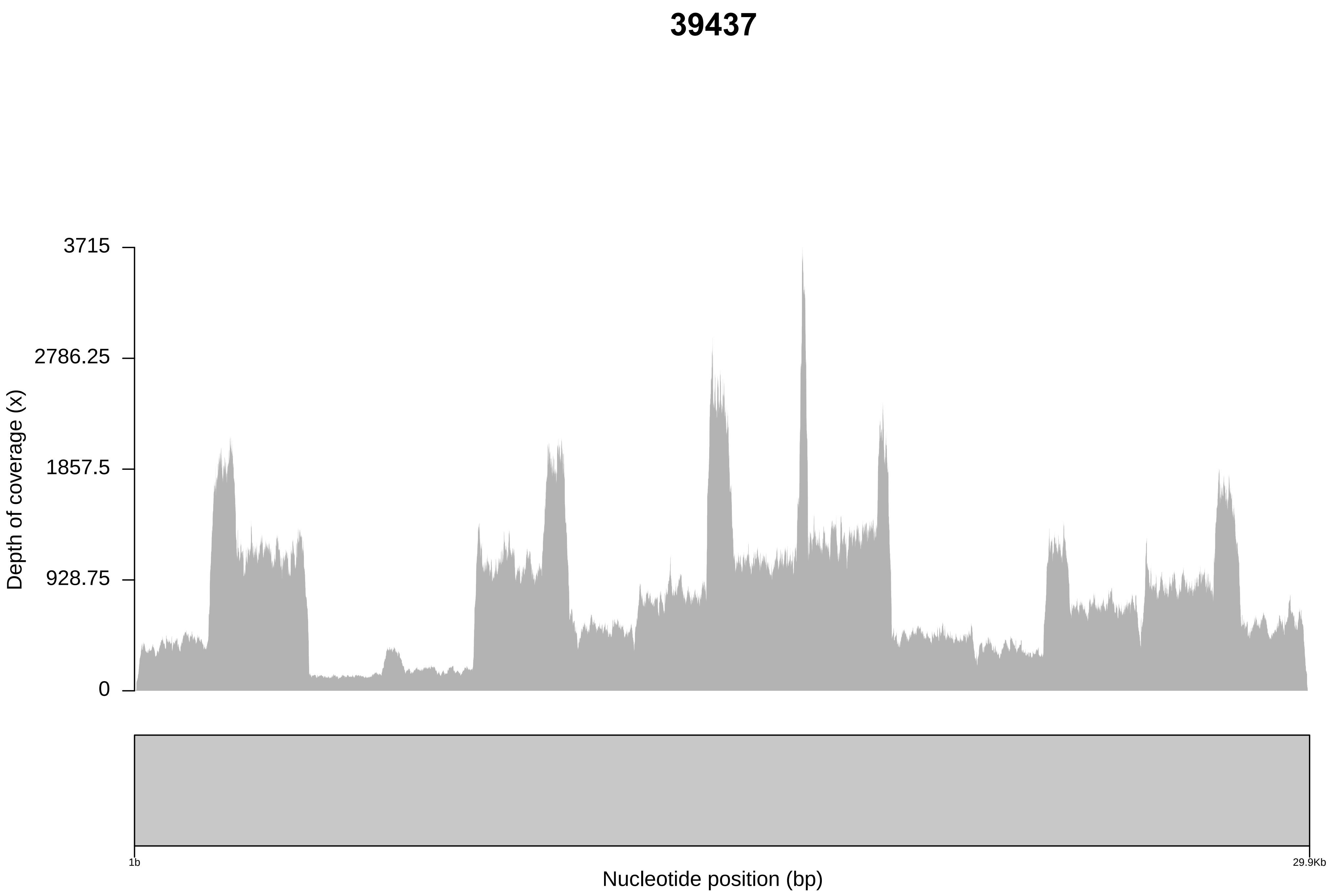
39437
Depth of coverage (x)
3715
2786.25
1857.5
928.75
0
1b
29.9Kb
Nucleotide position (bp)

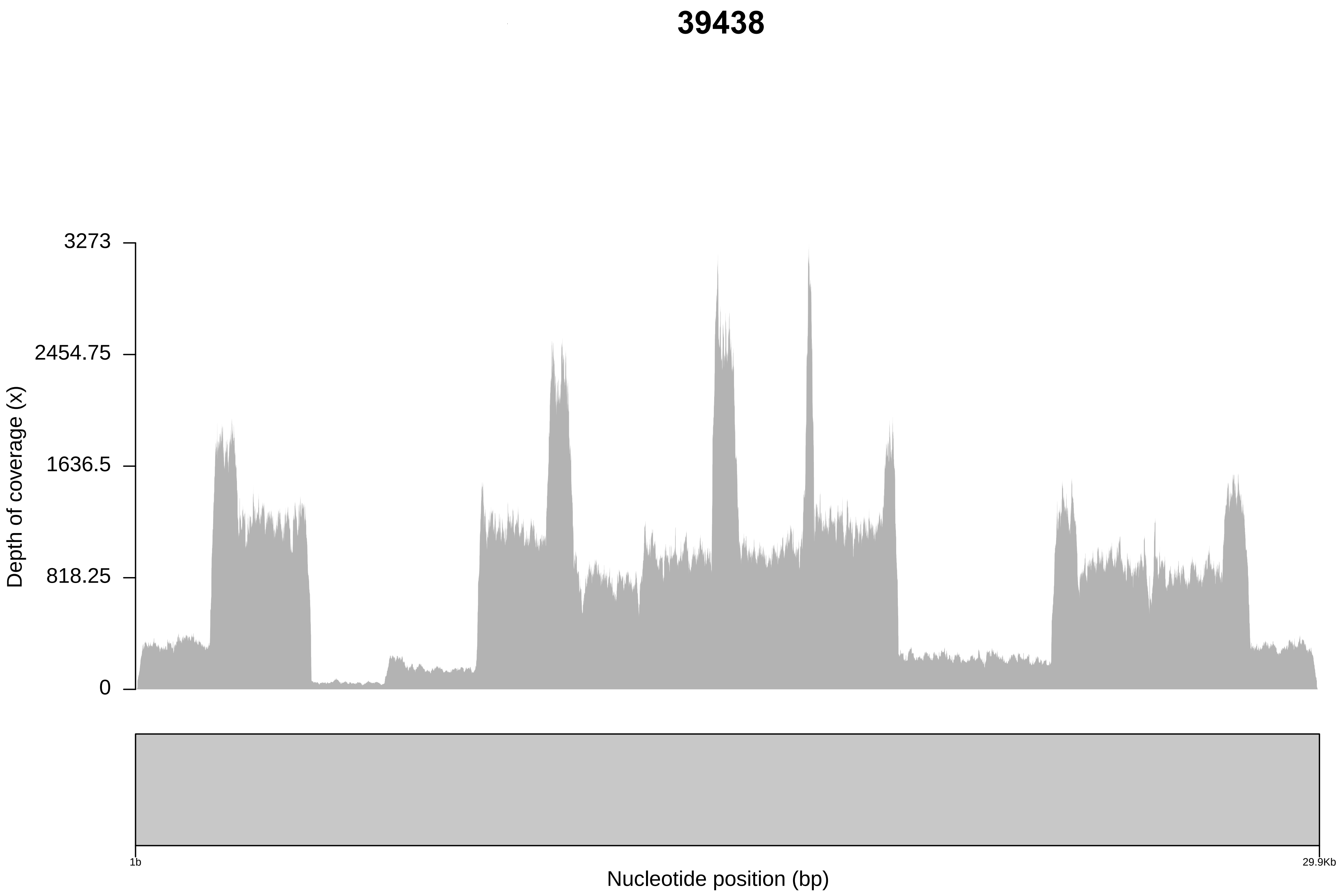
39438
3273
2454.75
1636.5
818.25
0
Depth of coverage (x)
1b
29.9Kb
Nucleotide position (bp)

# 39439

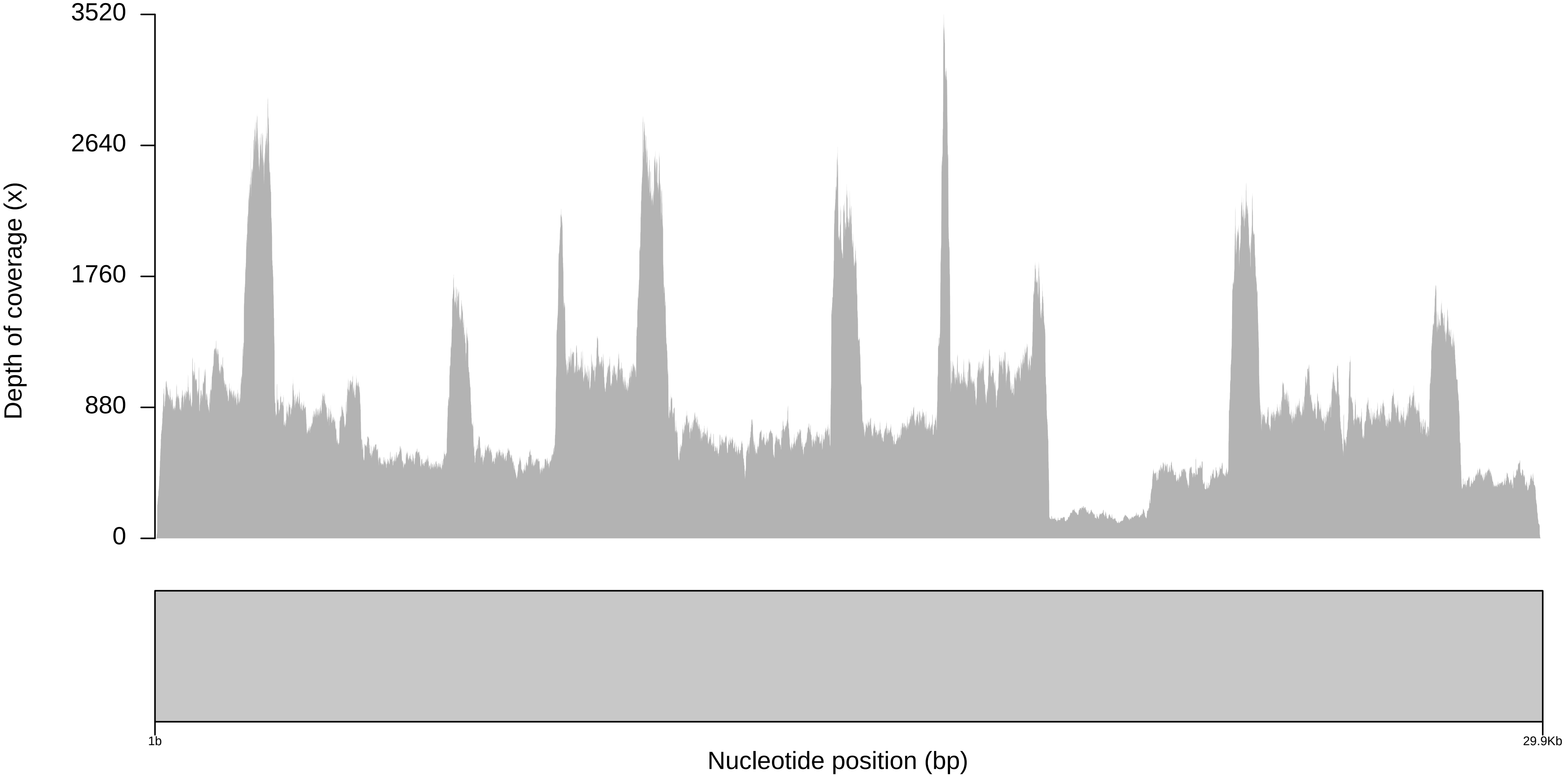

3520
2640
1760
880
0
Depth of coverage (x)
1b
29.9Kb
Nucleotide position (bp)

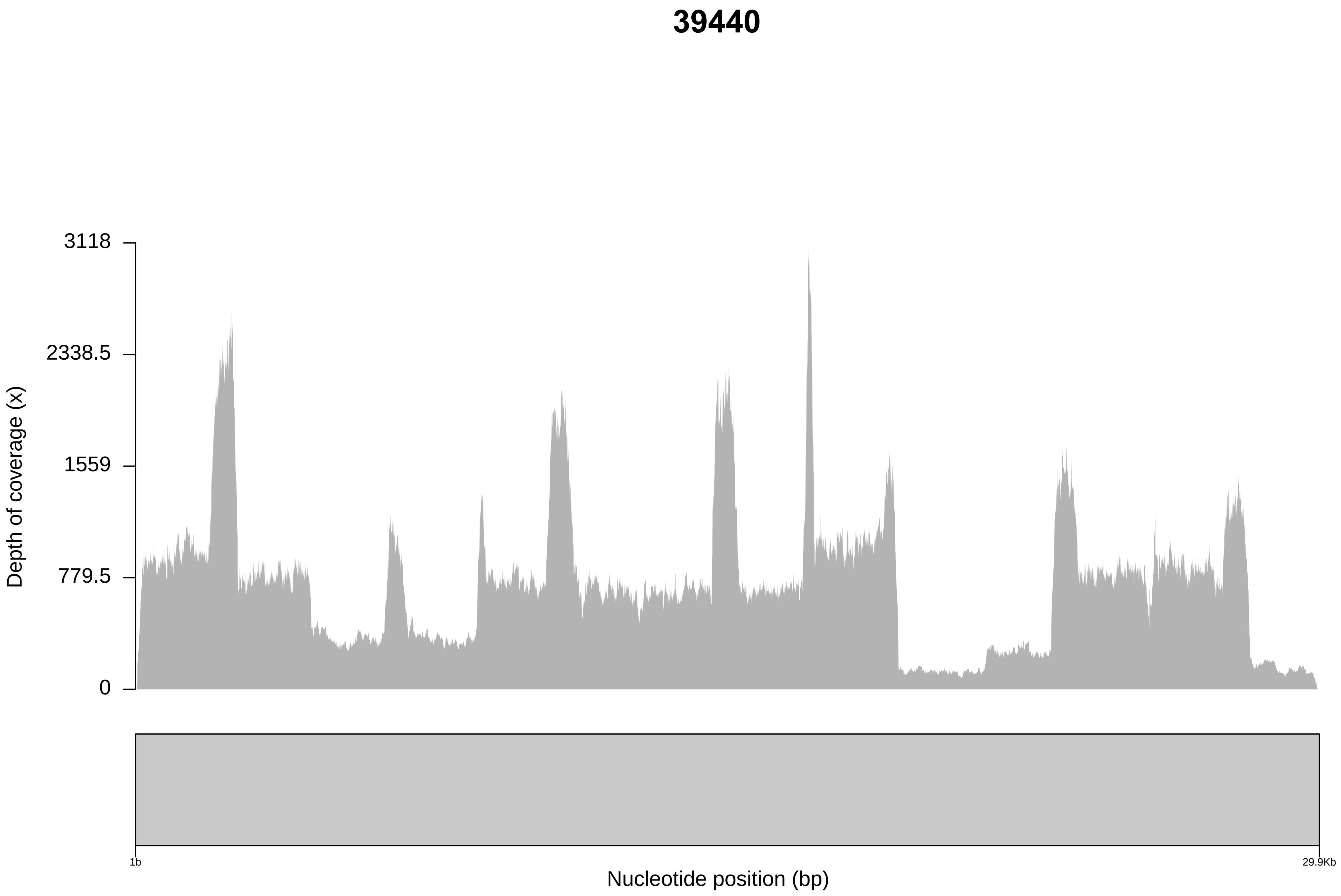
39440
3118
2338.5
1559
779.5
0
Depth of coverage (x)
1b
29.9Kb
Nucleotide position (bp)

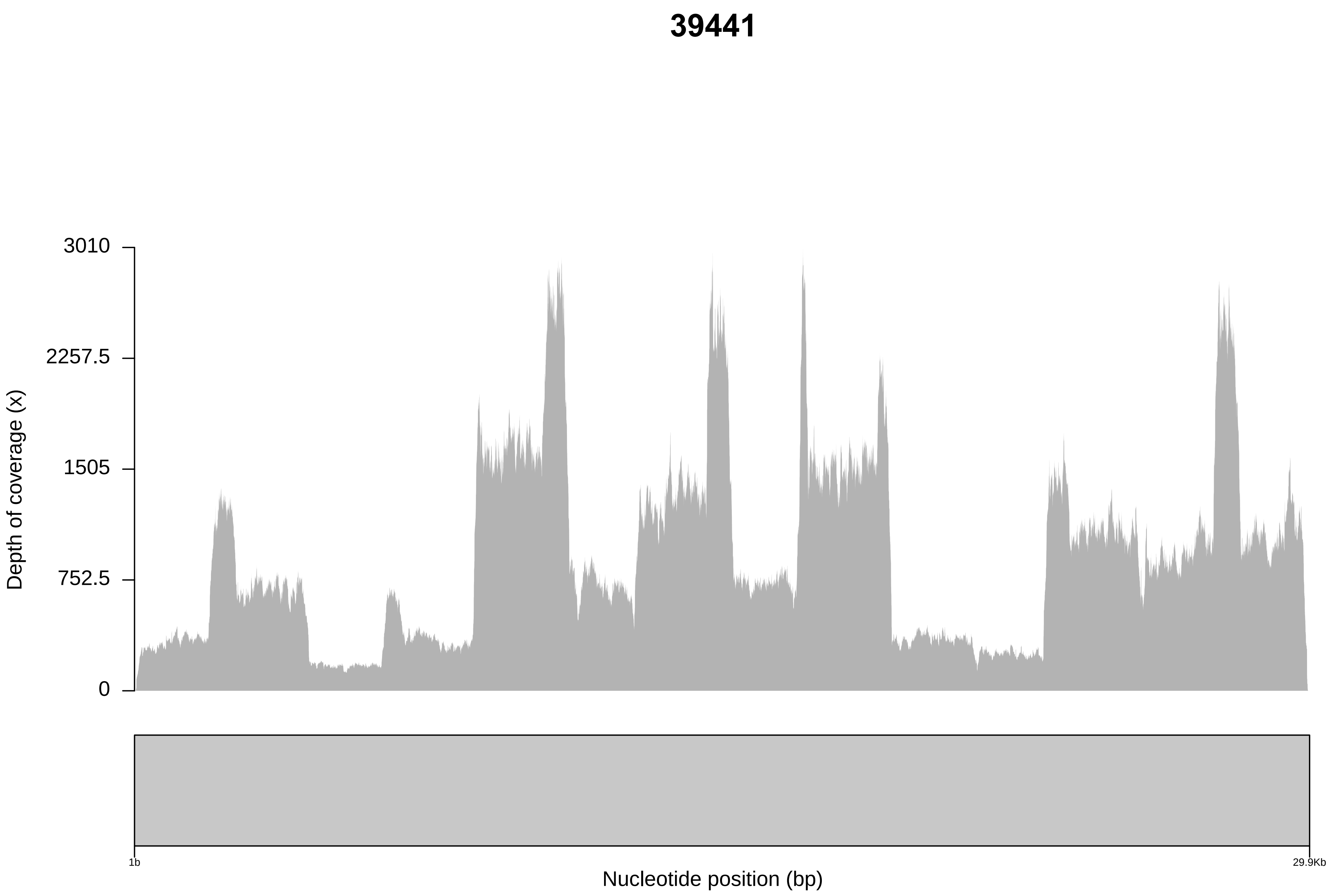

39441
3010
2257.5
1505
752.5
0
Depth of coverage (x)
1b
29.9Kb
Nucleotide position (bp)

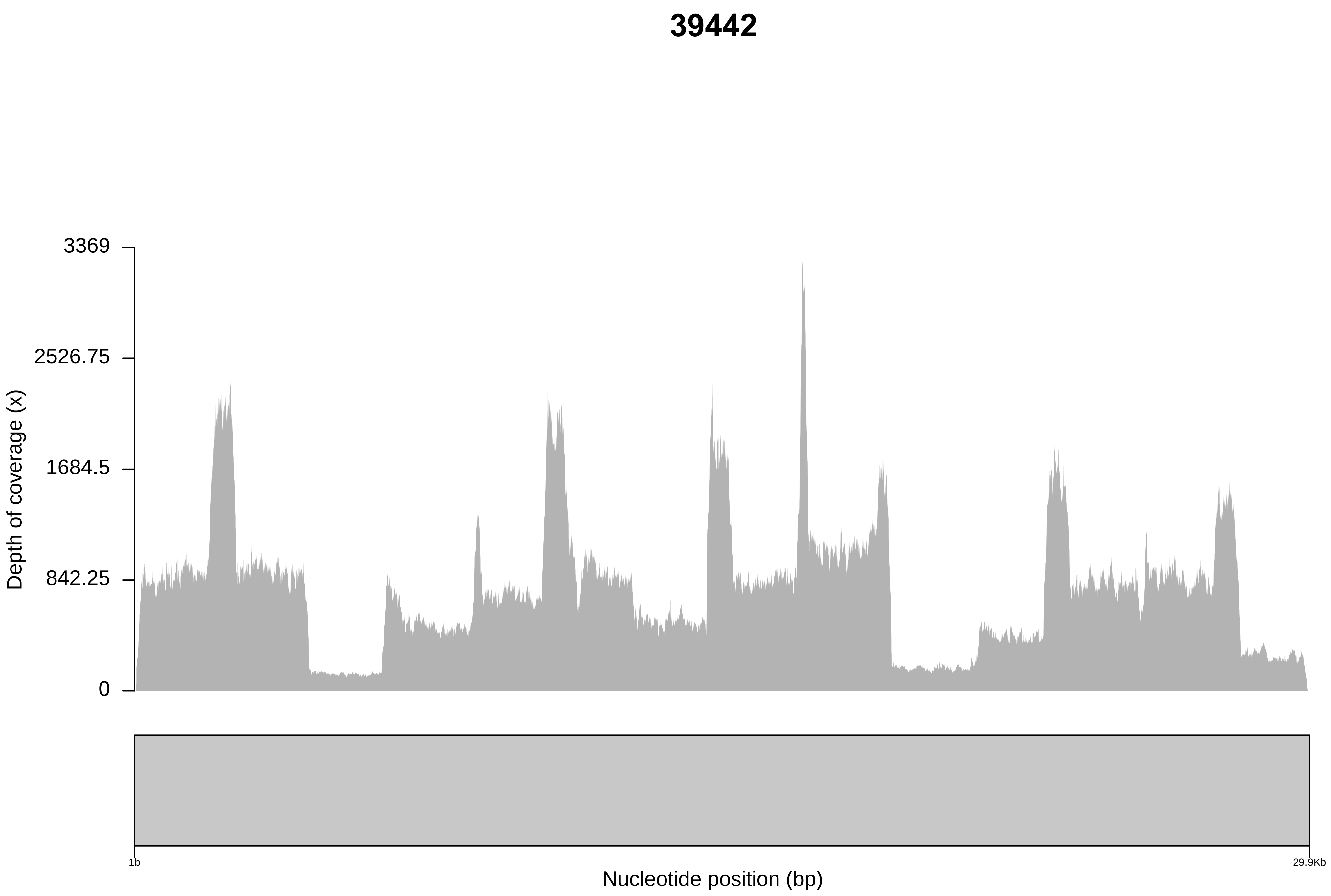
39442
3369
2526.75
1684.5
842.25
0
Depth of coverage (x)
1b
29.9Kb
Nucleotide position (bp)

# 39443

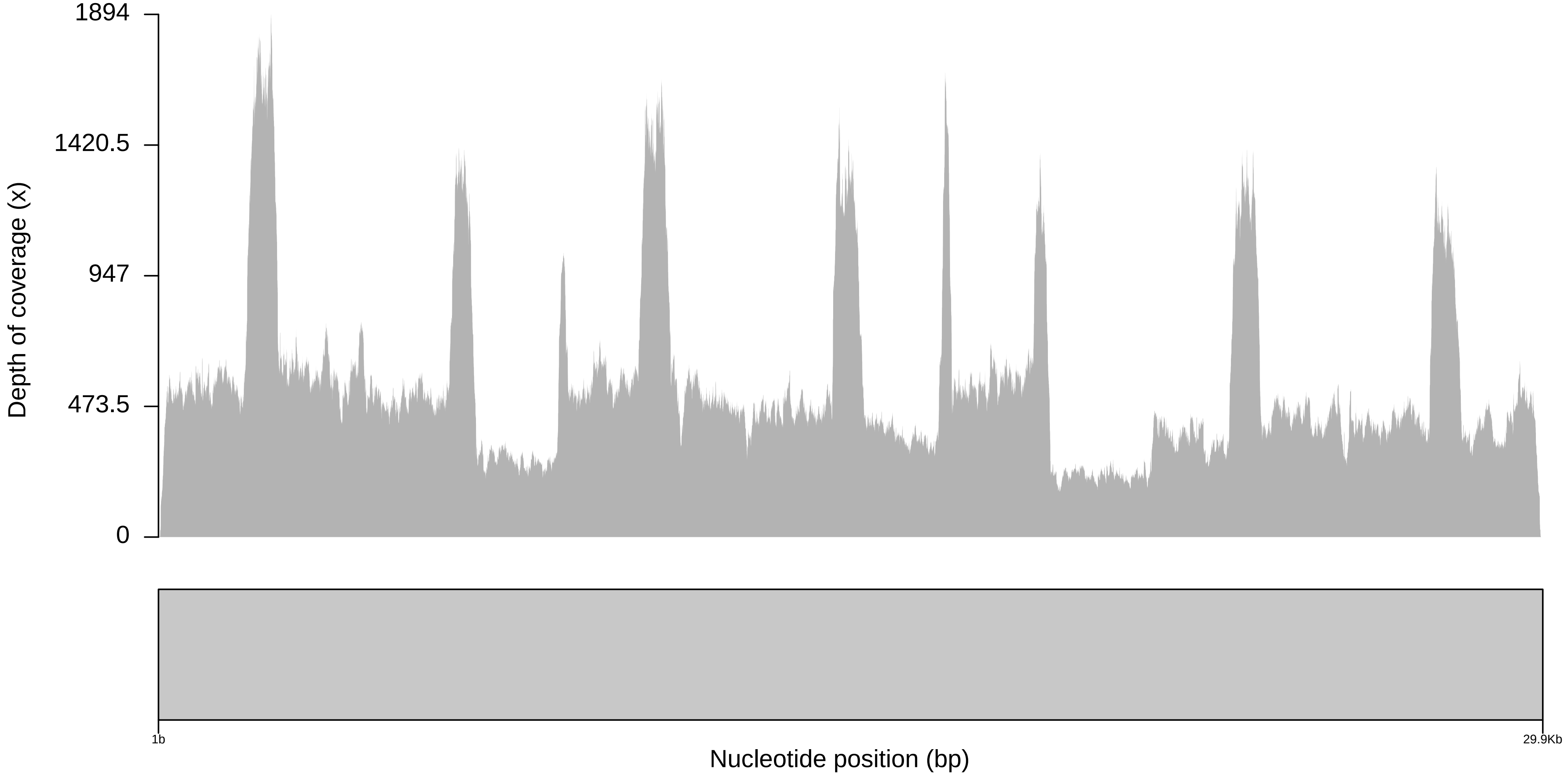

1894
1420.5
947
473.5
0
Depth of coverage (x)
1b
29.9Kb
Nucleotide position (bp)

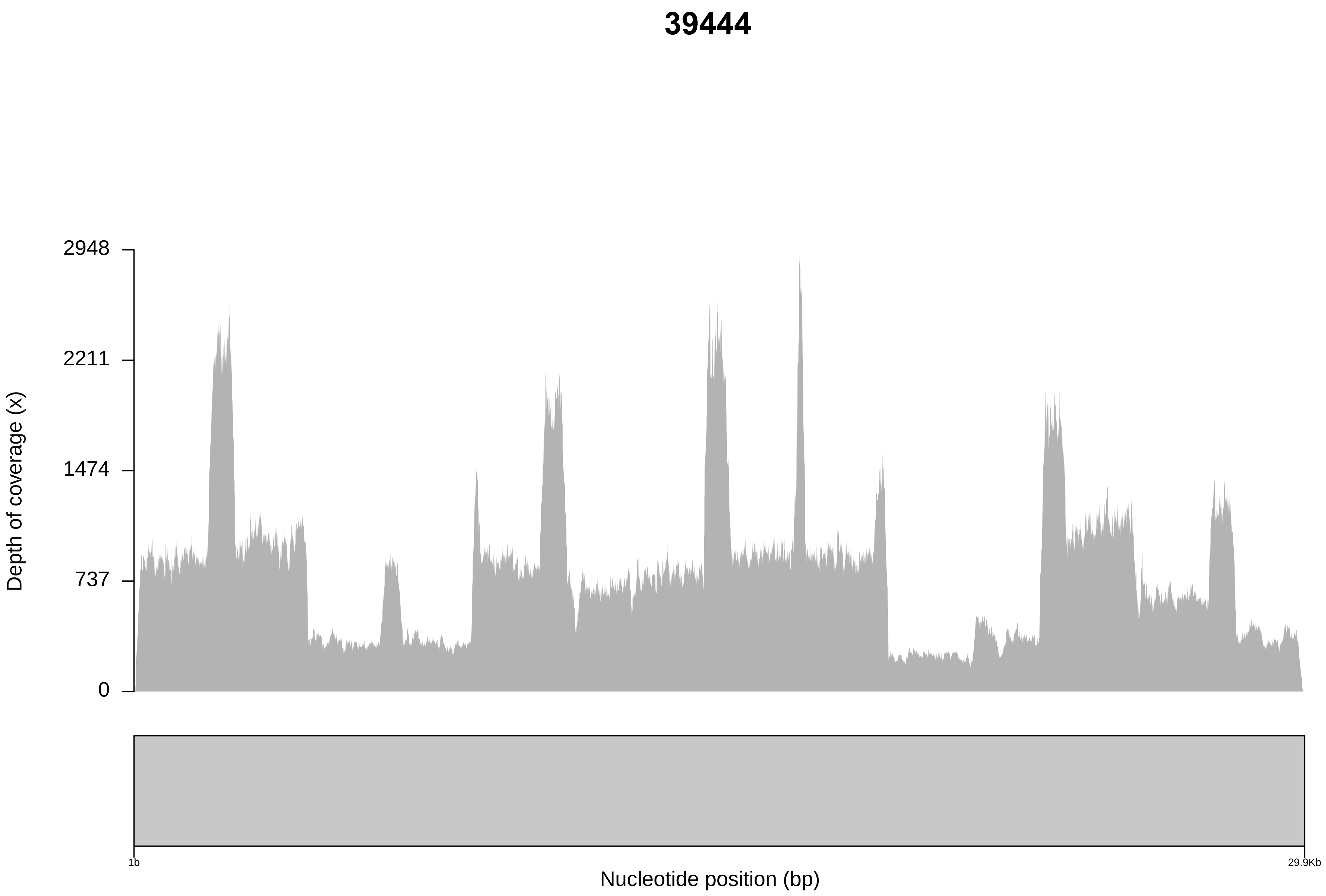

39444
Depth of coverage (x)
2948
2211
1474
737
0
1b
29.9Kb
Nucleotide position (bp)

# 39445

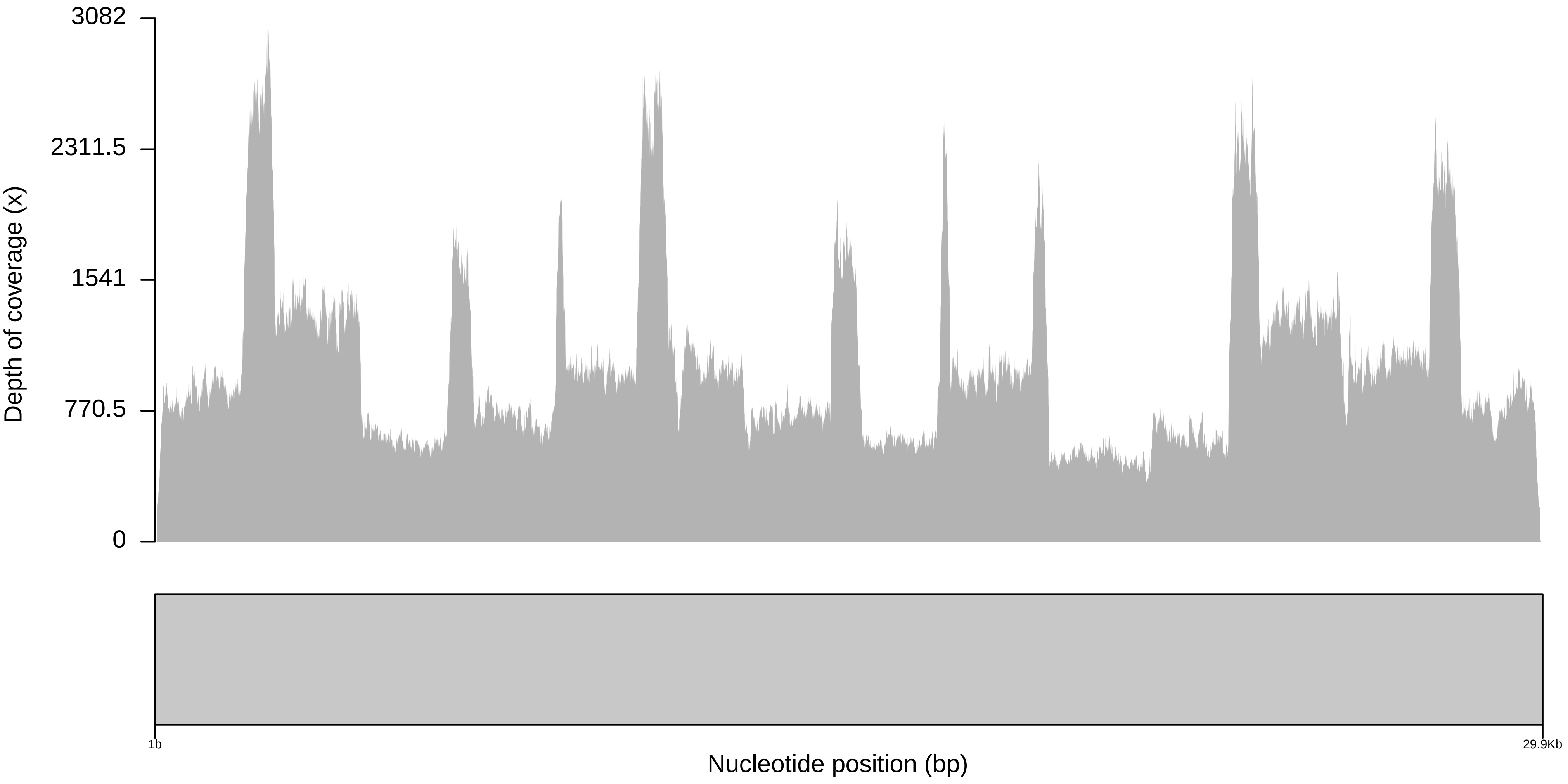
3082
2311.5
1541
770.5
0
Depth of coverage (x)
1b
29.9Kb
Nucleotide position (bp)

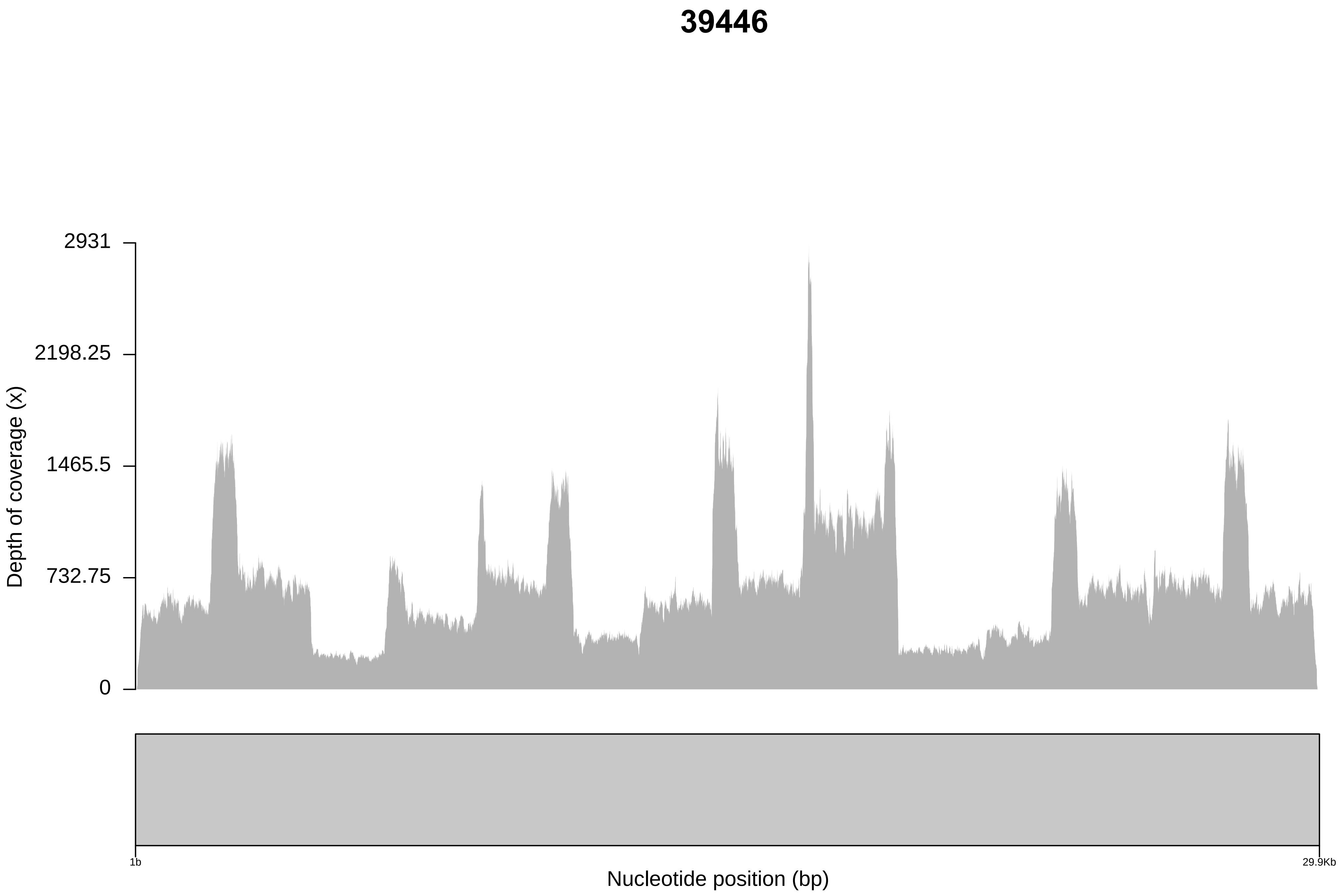

39446
2931
2198.25
1465.5
732.75
0
Depth of coverage (x)
1b
29.9Kb
Nucleotide position (bp)

## 39447

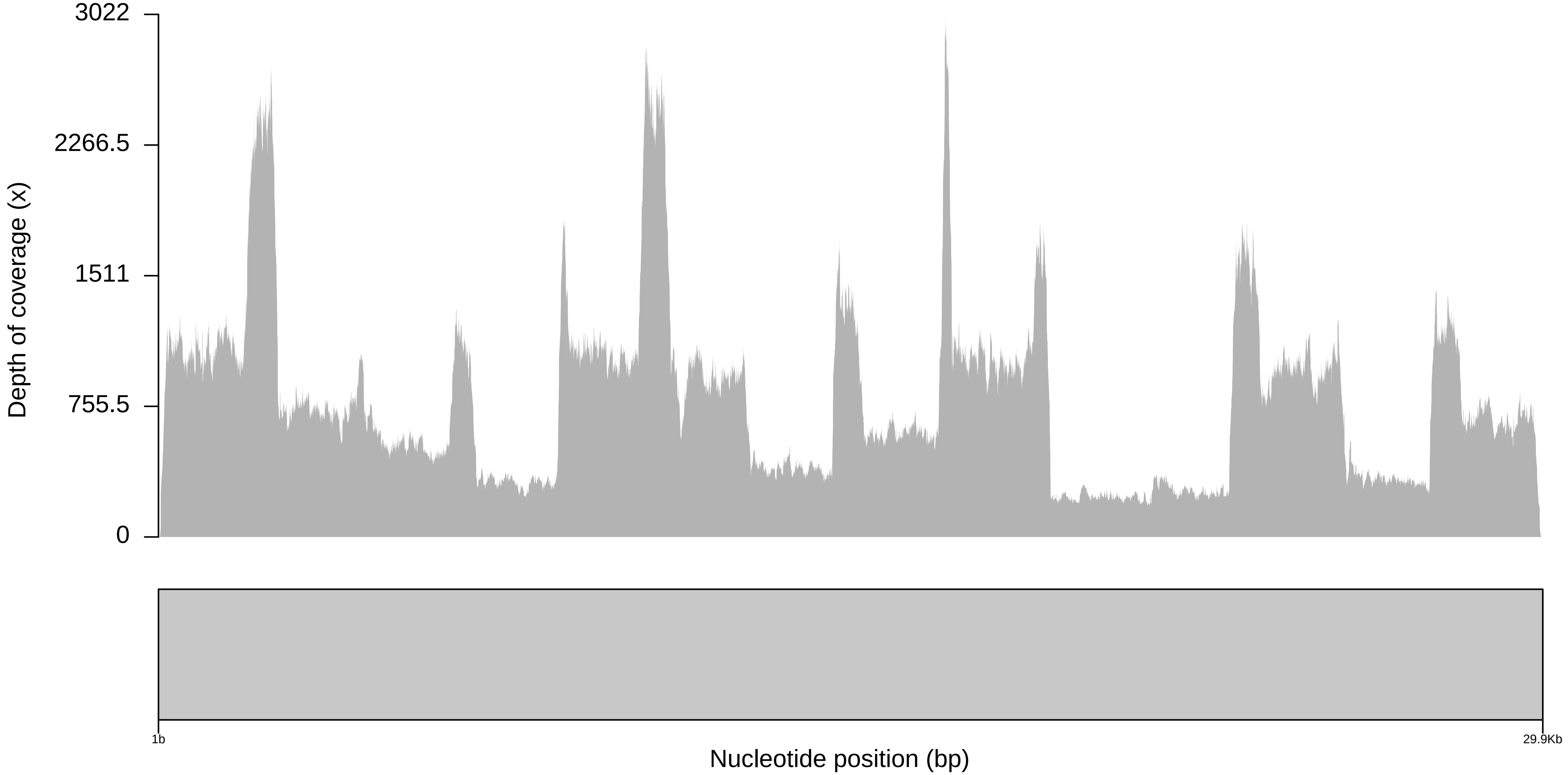

3022
2266.5
1511
755.5
0
Depth of coverage (x)
1b
29.9Kb
Nucleotide position (bp)

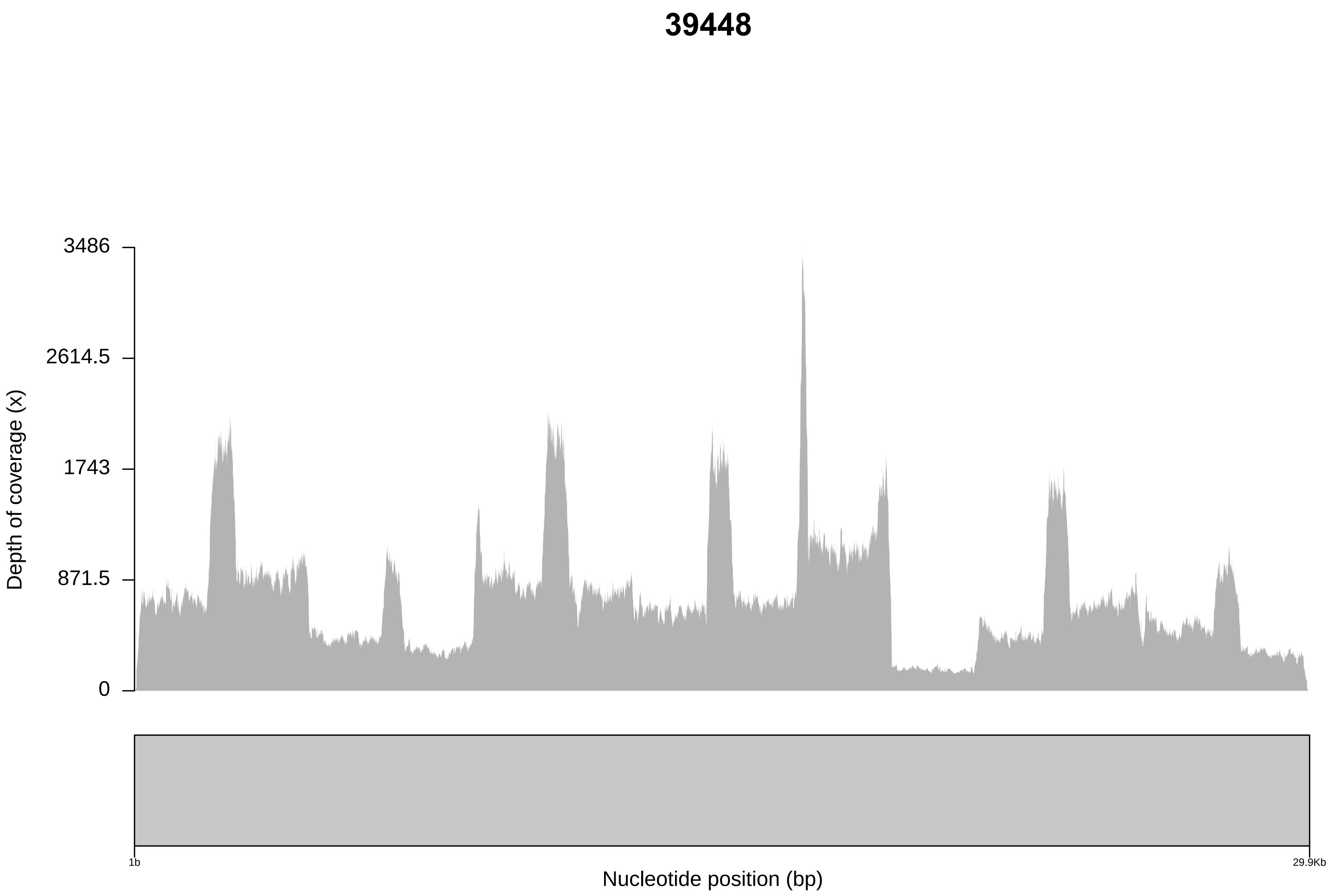

39448
3486
2614.5
1743
871.5
0
Depth of coverage (x)
1b
29.9Kb
Nucleotide position (bp)

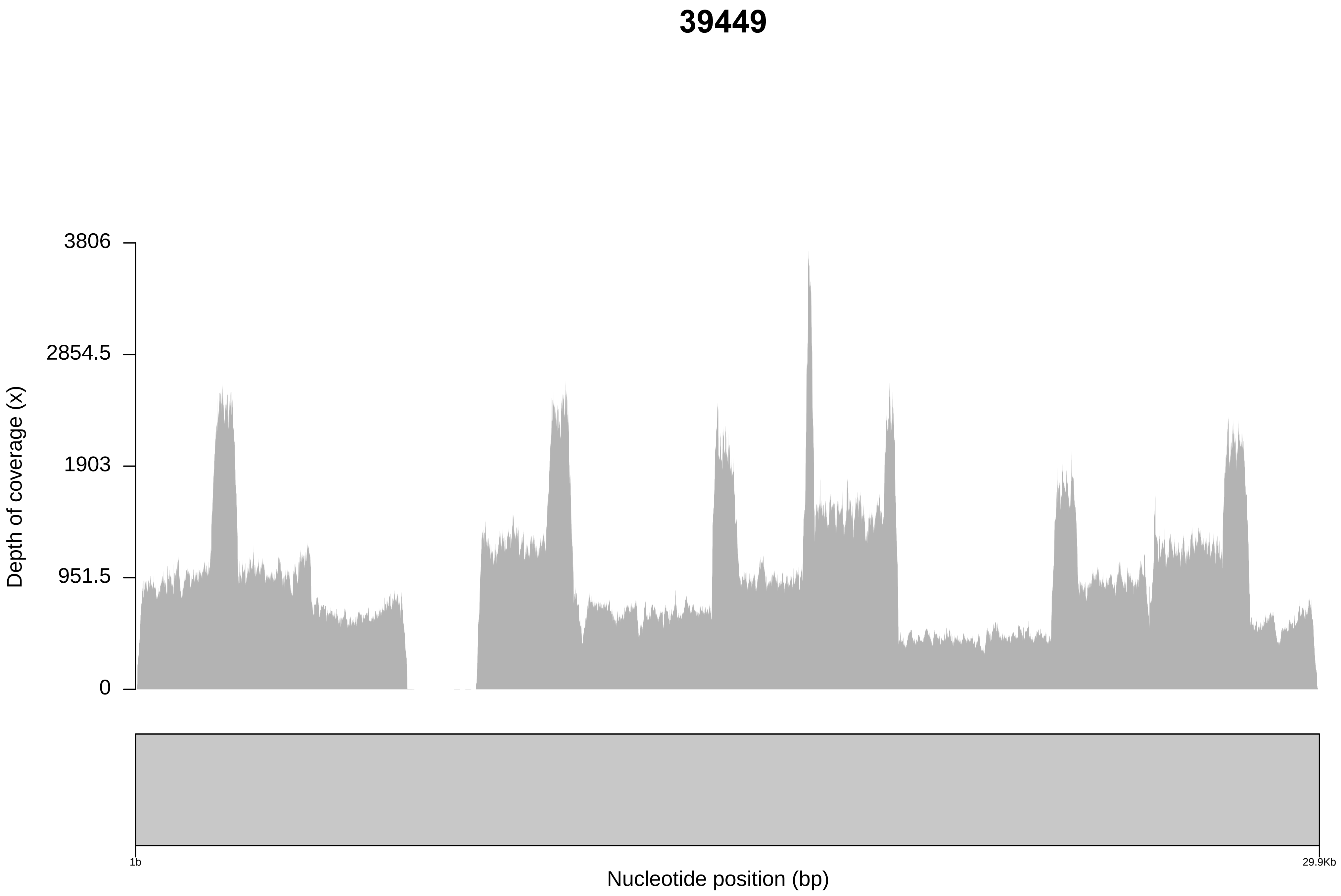

39449
3806
2854.5
1903
951.5
0
Depth of coverage (x)
1b
29.9Kb
Nucleotide position (bp)

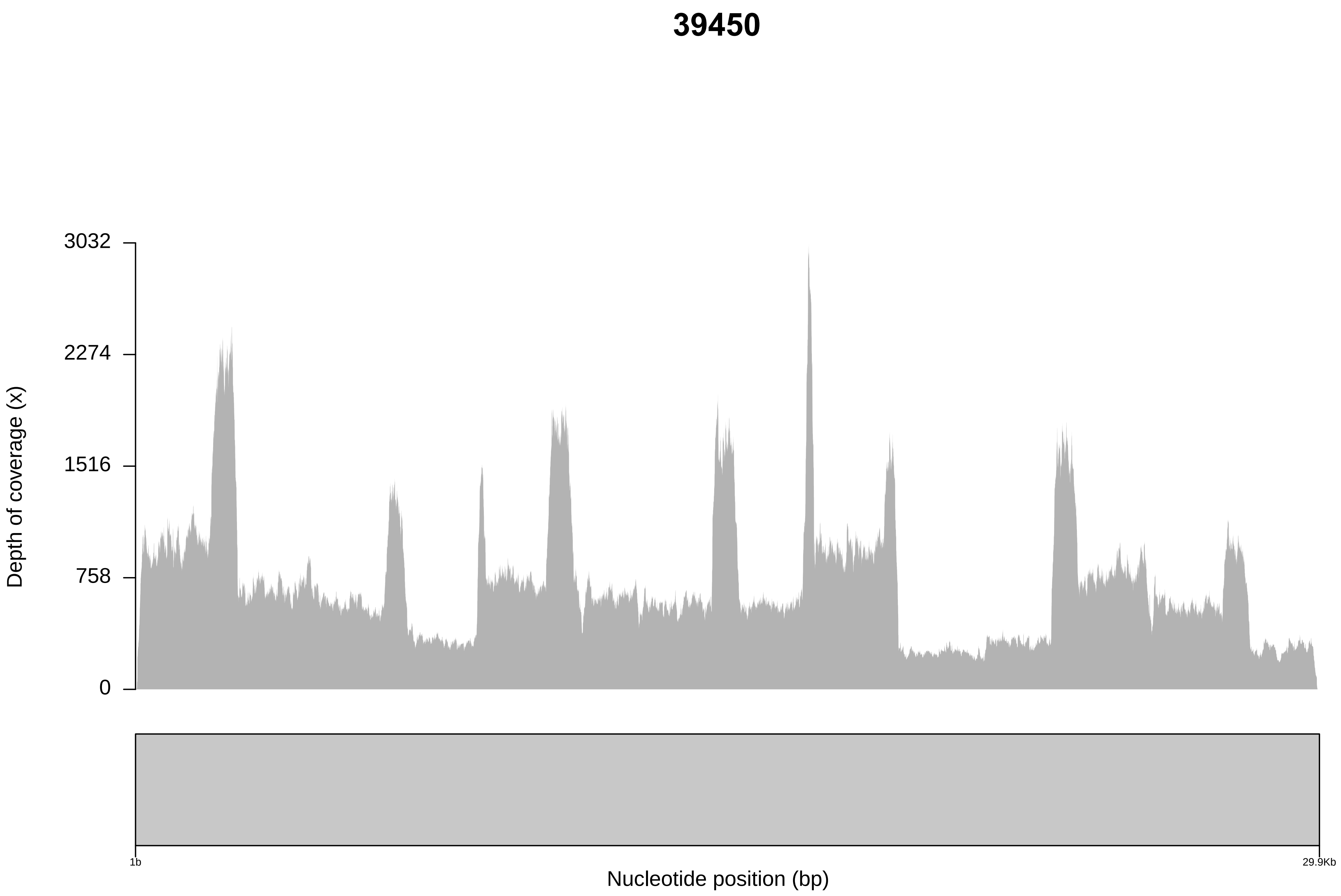

39450
3032
2274
1516
758
0
Depth of coverage (x)
1b
29.9Kb
Nucleotide position (bp)

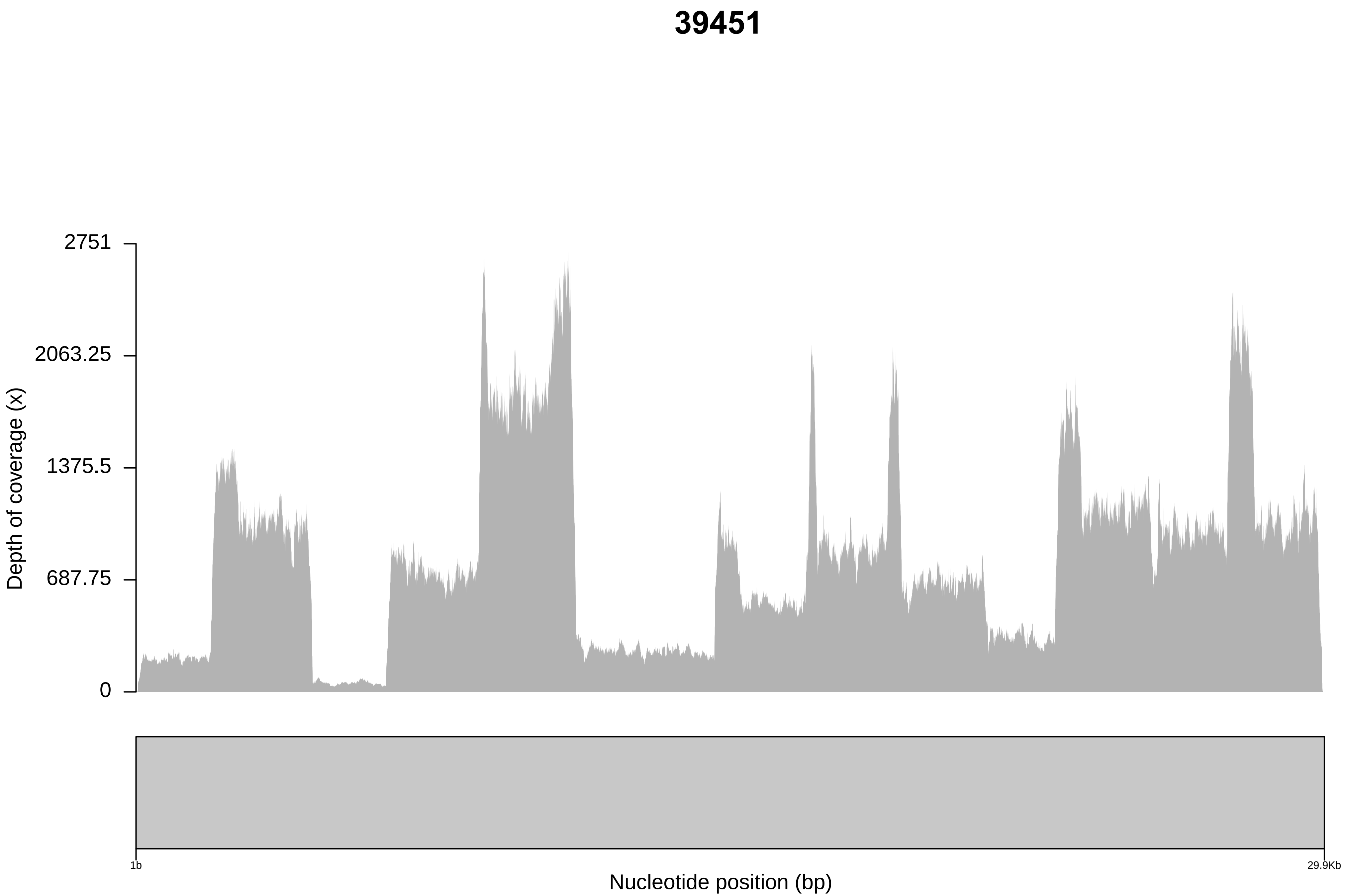
39451
Depth of coverage (x)
2751
2063.25
1375.5
687.75
0
1b
29.9Kb
Nucleotide position (bp)

# 39452

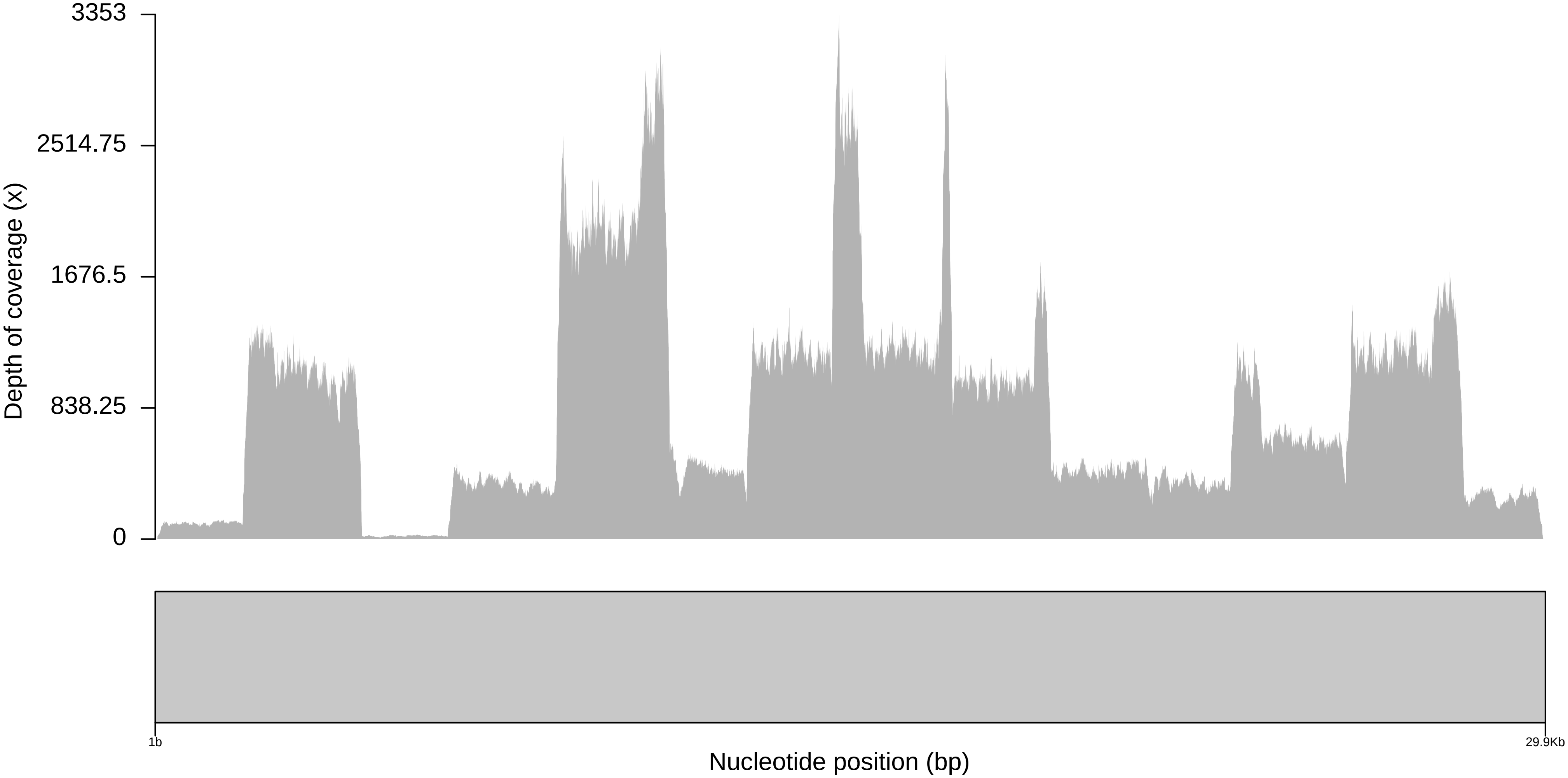

3353
2514.75
1676.5
838.25
0
Depth of coverage (x)
1b
29.9Kb
Nucleotide position (bp)

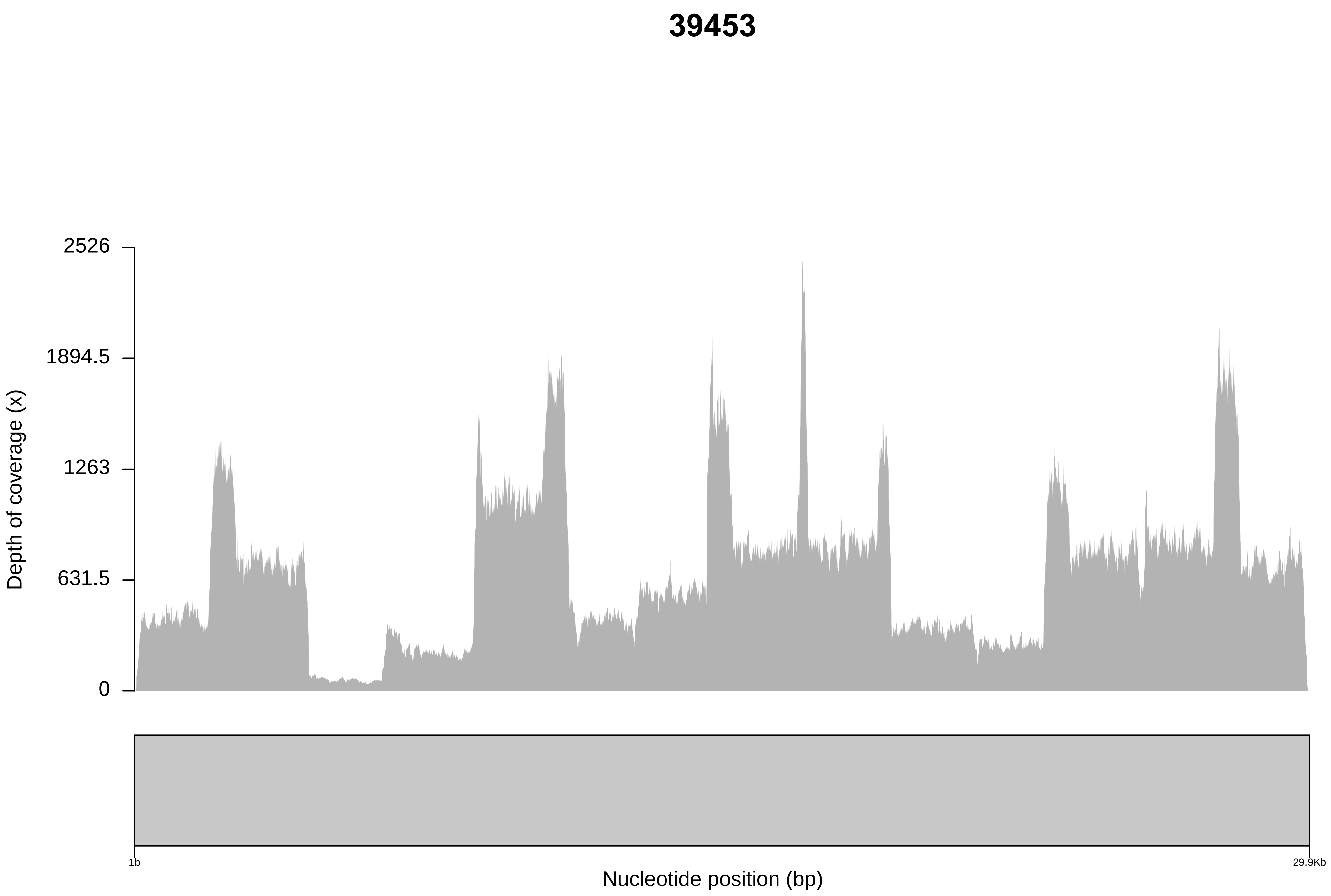

39453
2526
1894.5
1263
631.5
0
Depth of coverage (x)
1b
29.9Kb
Nucleotide position (bp)

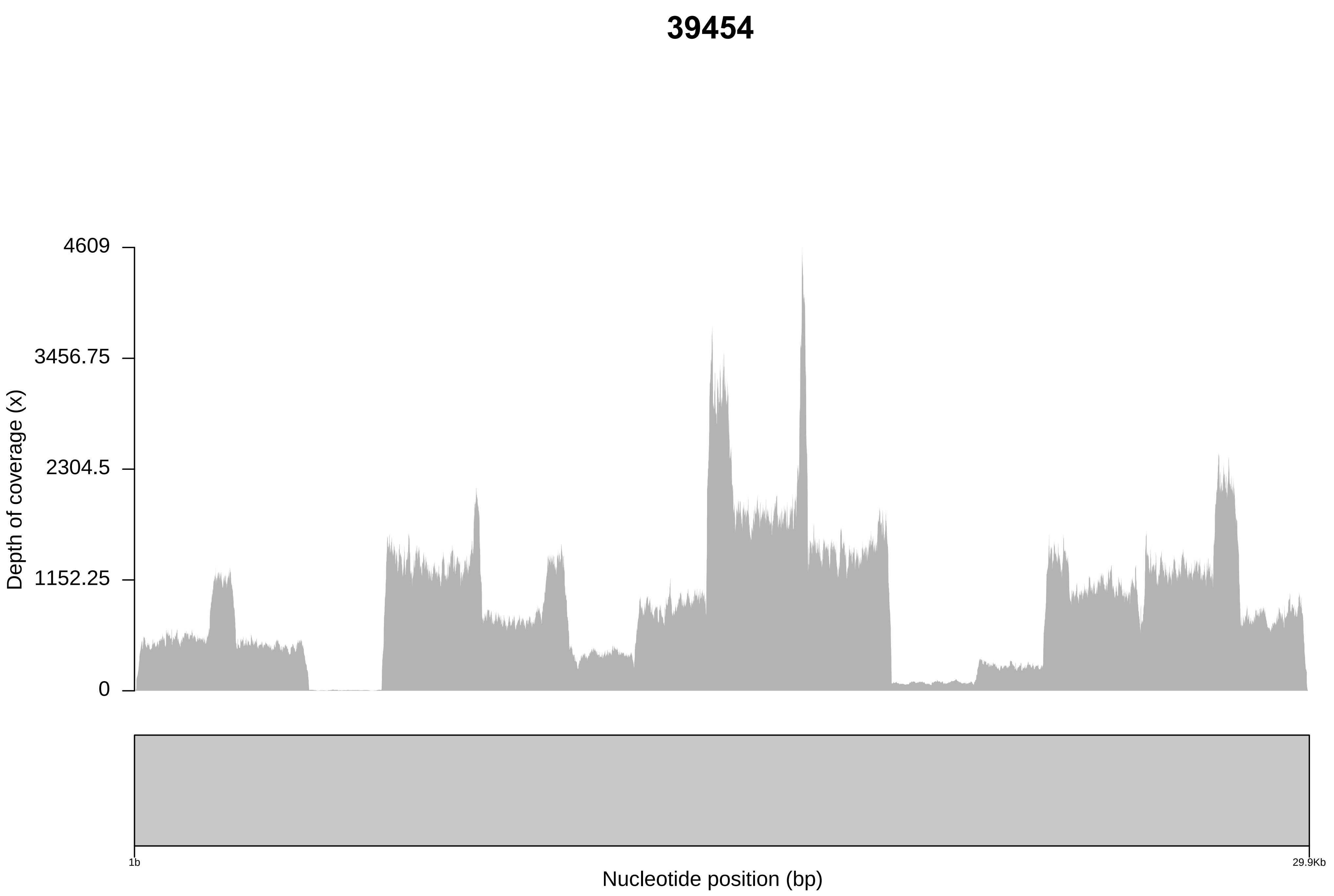

39454
4609
3456.75
2304.5
1152.25
0
Depth of coverage (x)
1b
29.9Kb
Nucleotide position (bp)

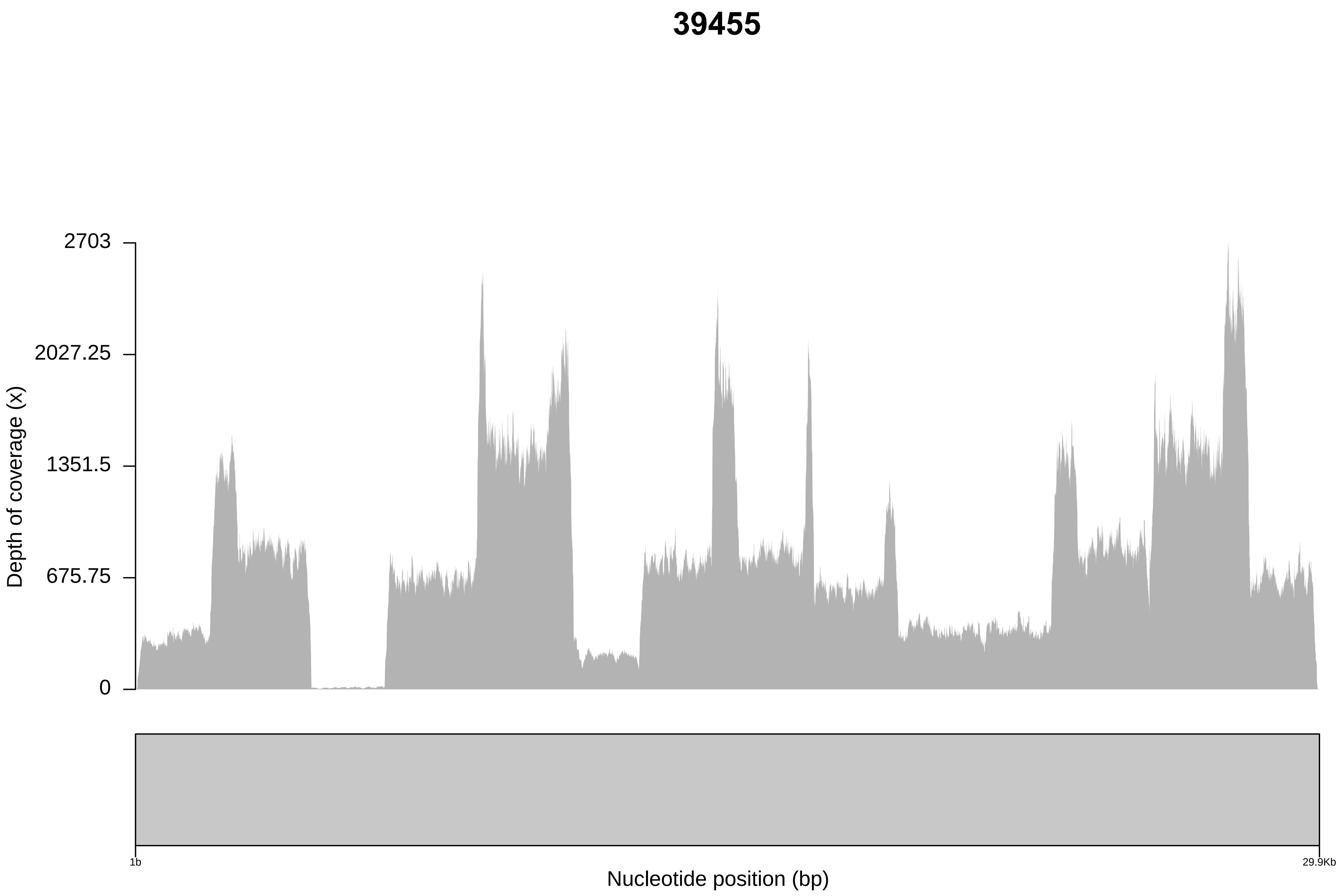

39455
2703
2027.25
1351.5
675.75
0
Depth of coverage (x)
1b
29.9Kb
Nucleotide position (bp)

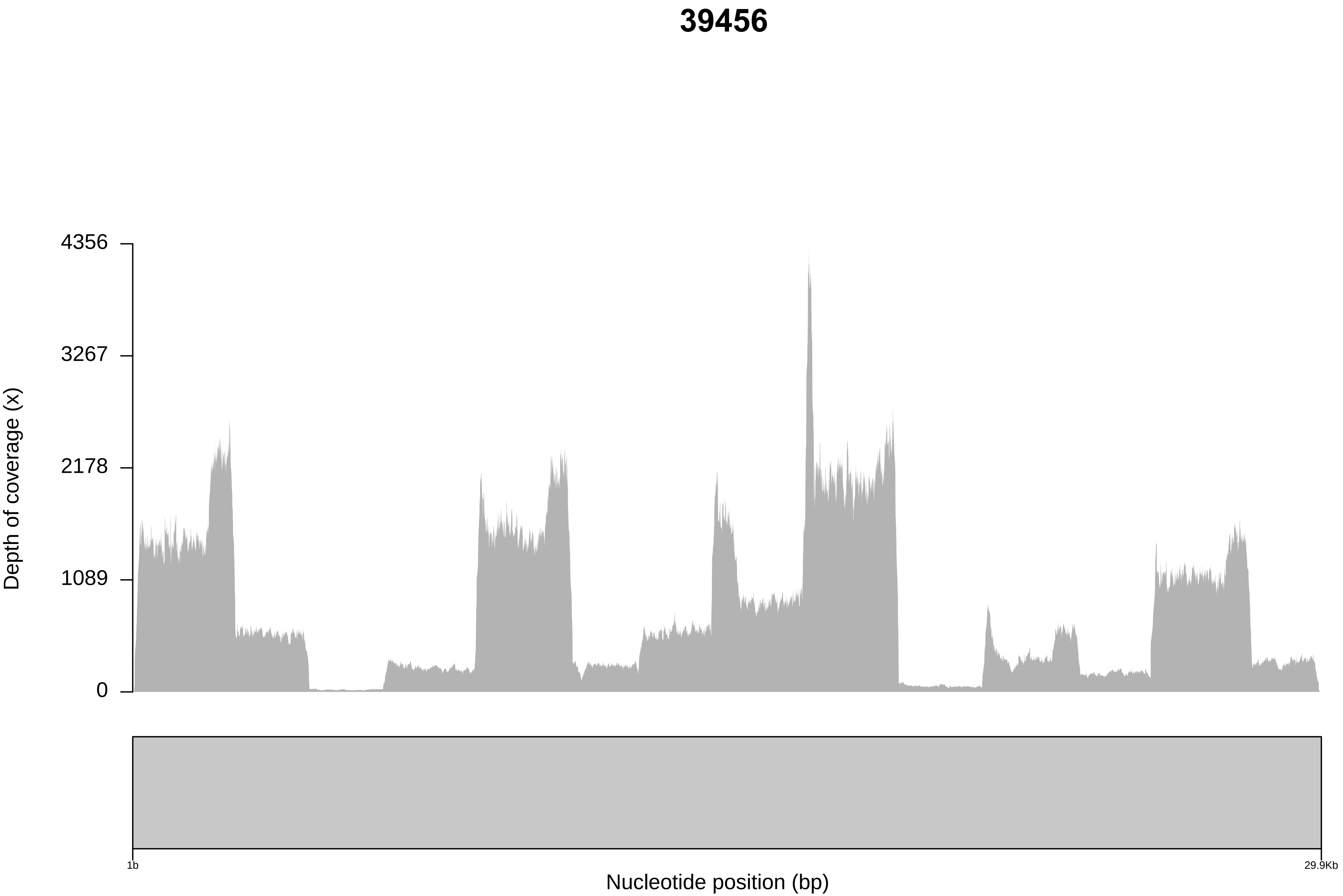

39456
4356
3267
2178
1089
0
Depth of coverage (x)
1b
29.9Kb
Nucleotide position (bp)

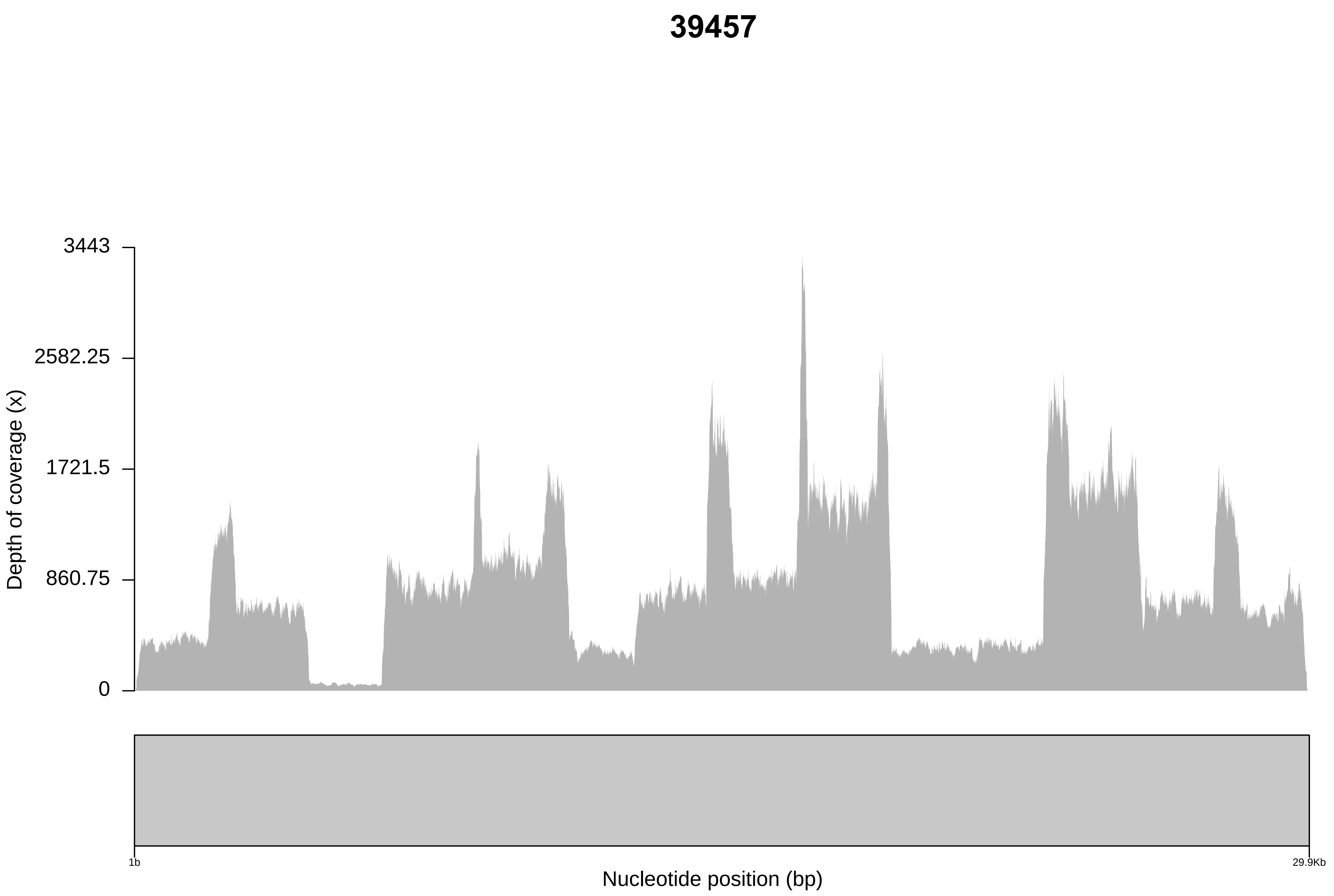

39457
3443
2582.25
1721.5
860.75
0
Depth of coverage (x)
1b
29.9Kb
Nucleotide position (bp)

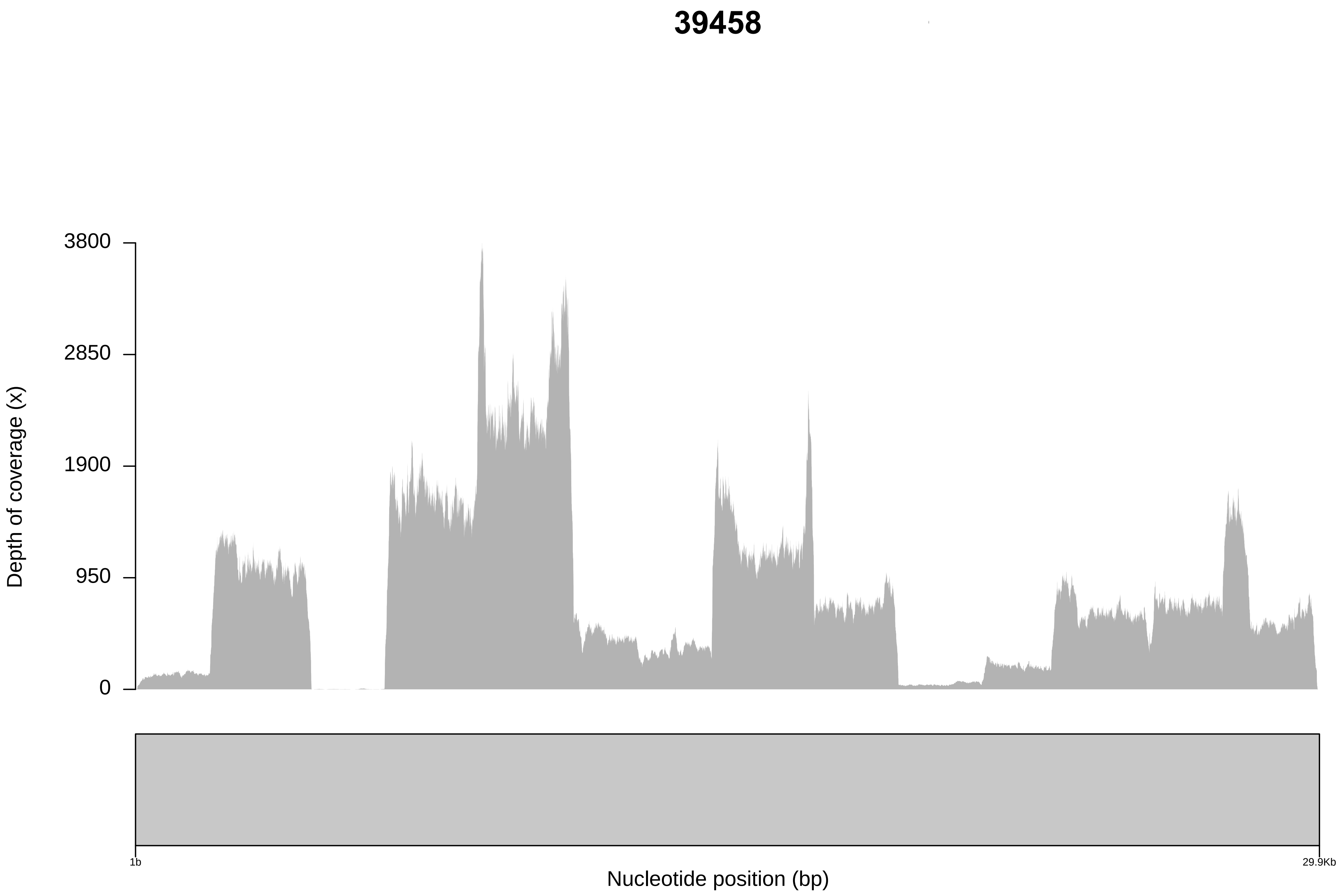

39458
Depth of coverage (x)
3800
2850
1900
950
0
1b
29.9Kb
Nucleotide position (bp)

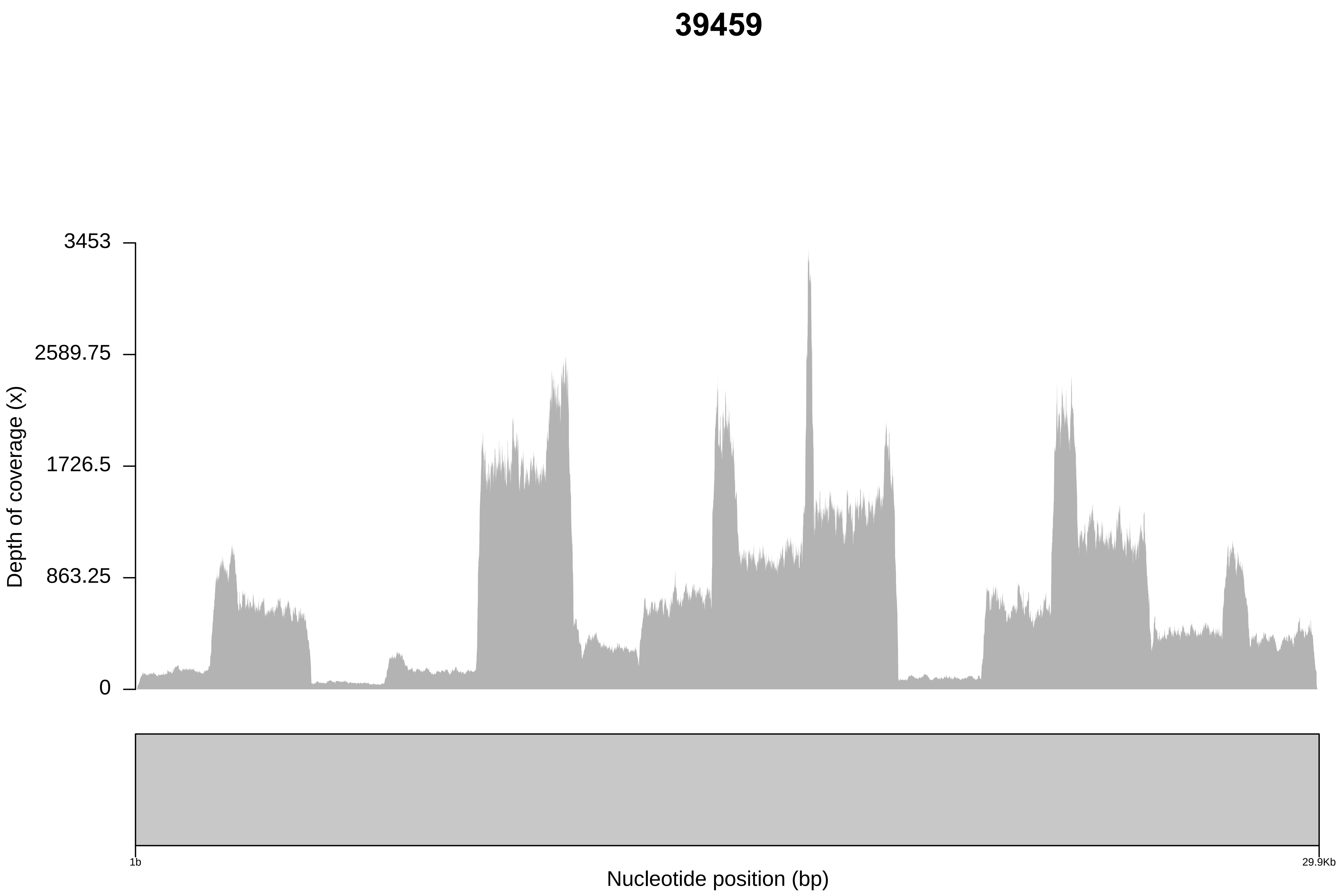

39459
3453
2589.75
1726.5
863.25
0
Depth of coverage (x)
1b
29.9Kb
Nucleotide position (bp)

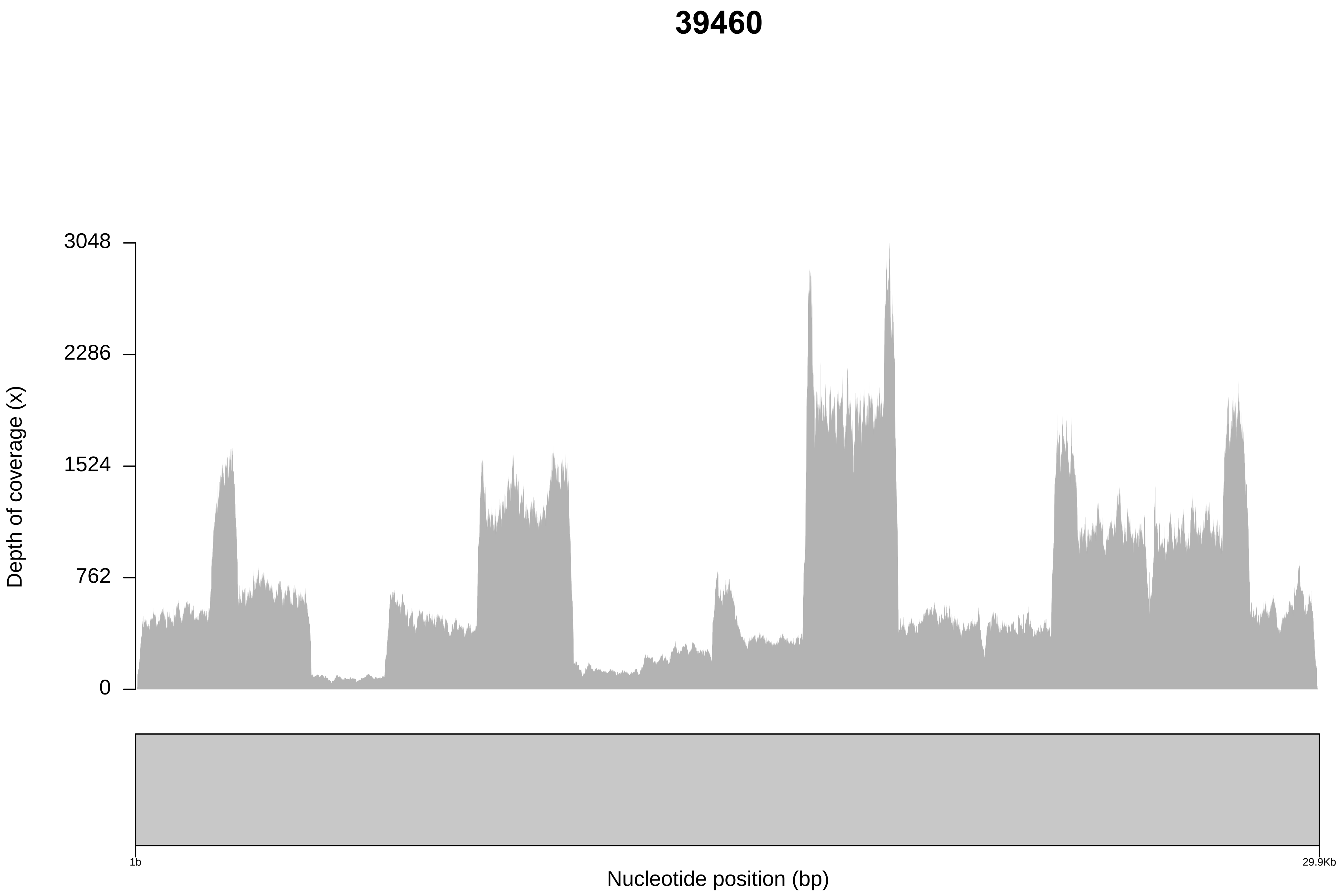

39460
Depth of coverage (x)
3048
2286
1524
762
0
1b
29.9Kb
Nucleotide position (bp)

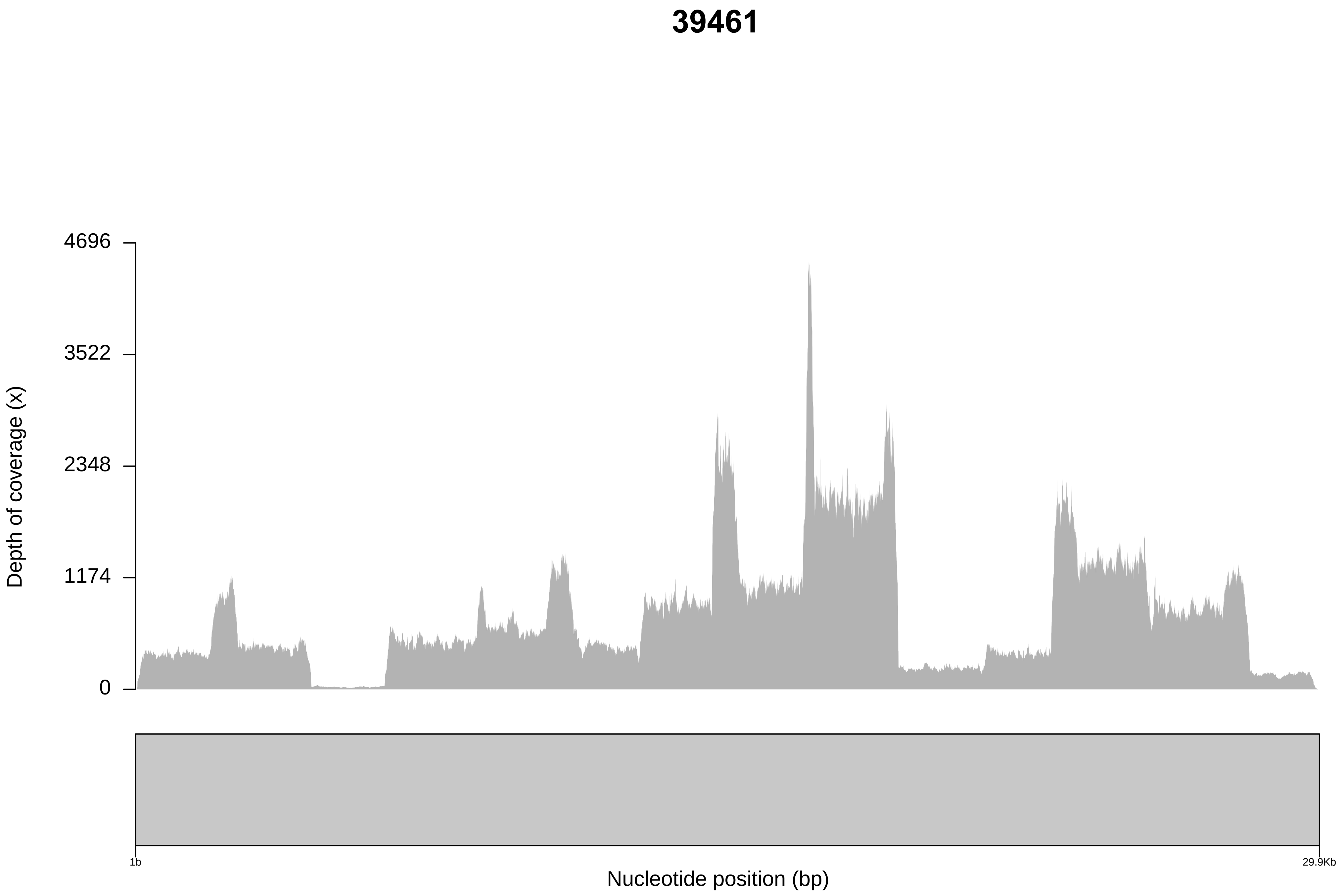

39461
Depth of coverage (x)
4696
3522
2348
1174
0
1b
29.9Kb
Nucleotide position (bp)

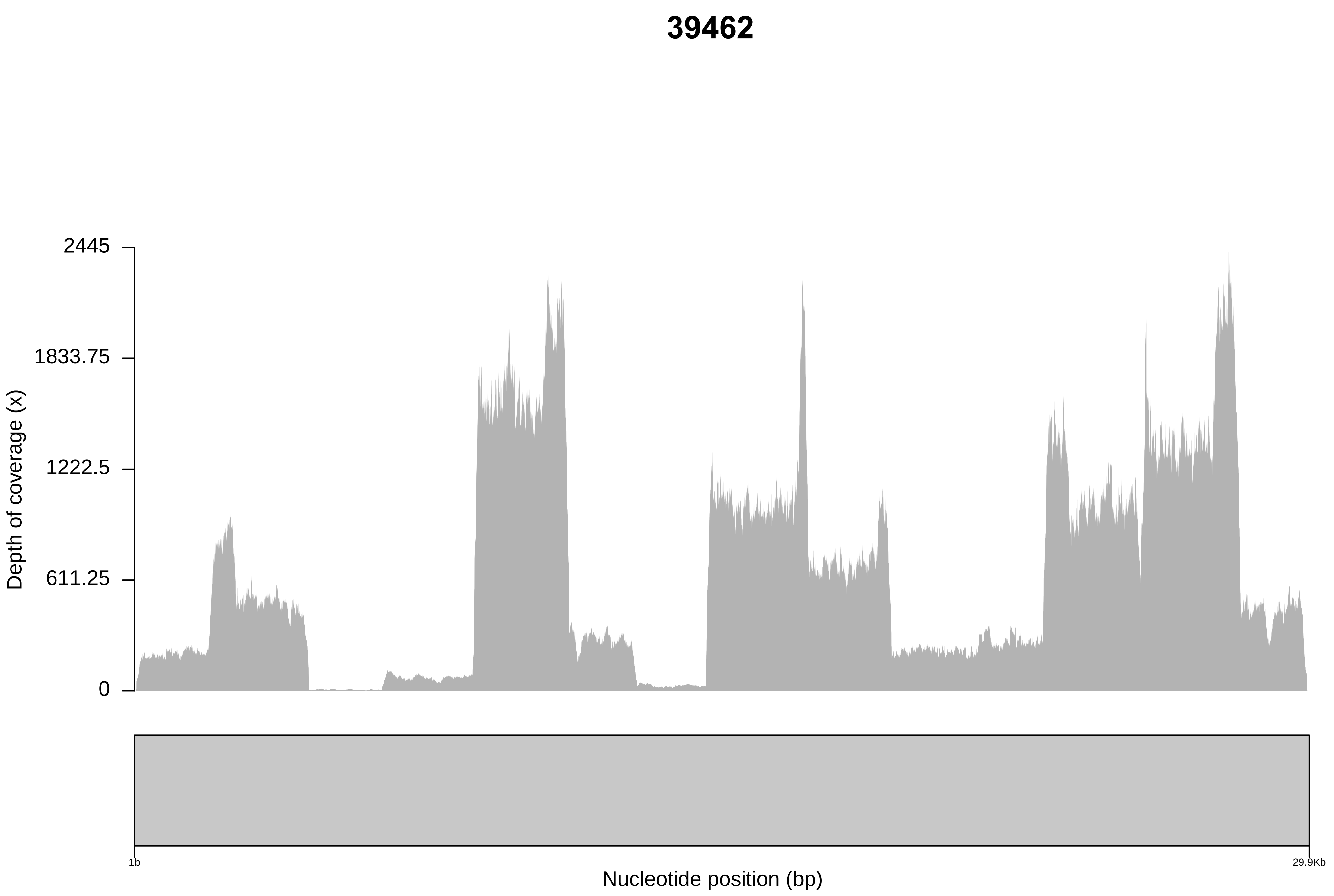

39462
Depth of coverage (x)
2445
1833.75
1222.5
611.25
0
1b
29.9Kb
Nucleotide position (bp)

# 39463

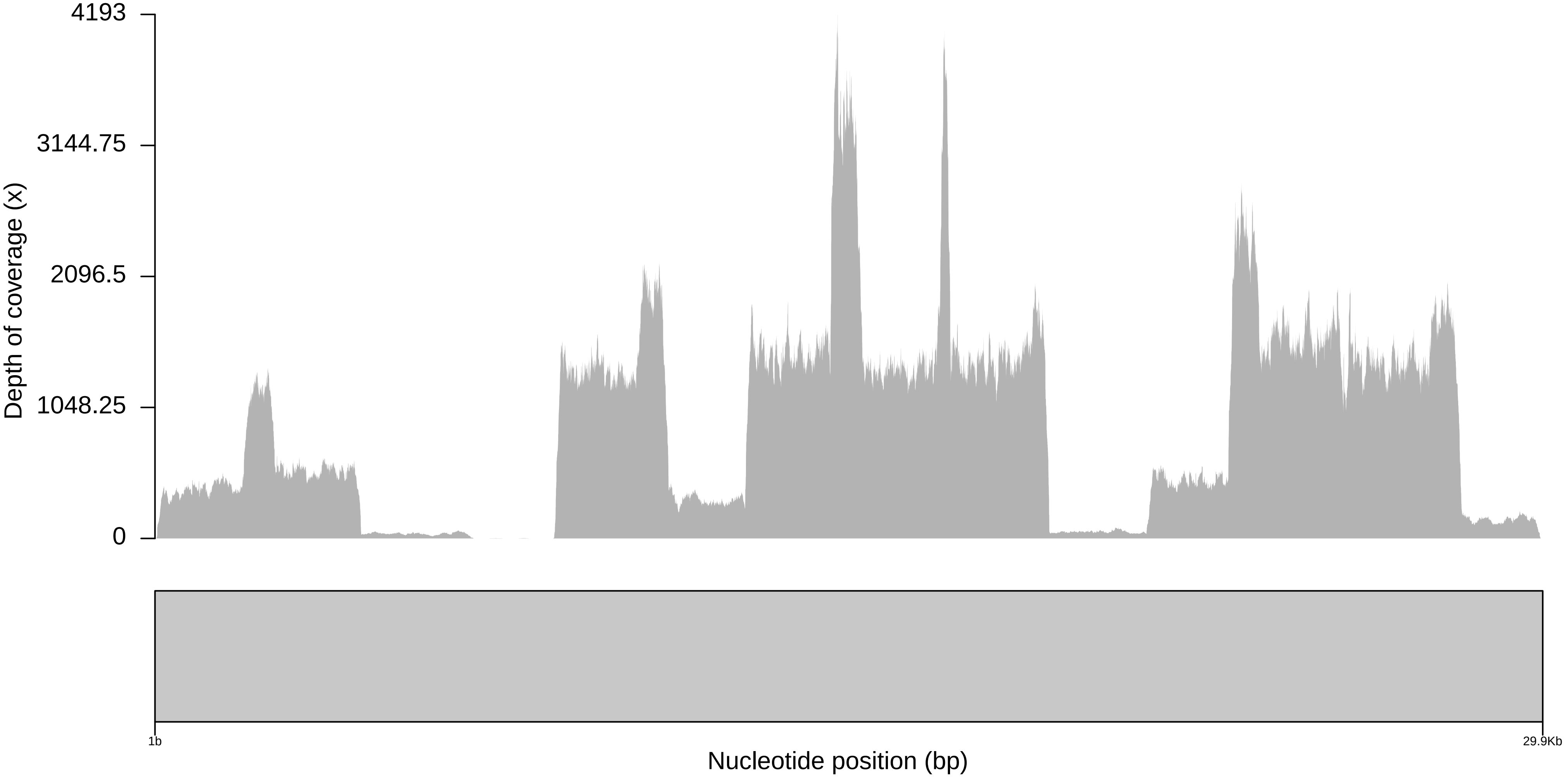

4193
3144.75
2096.5
1048.25
0
Depth of coverage (x)
1b
29.9Kb
Nucleotide position (bp)

## 39464

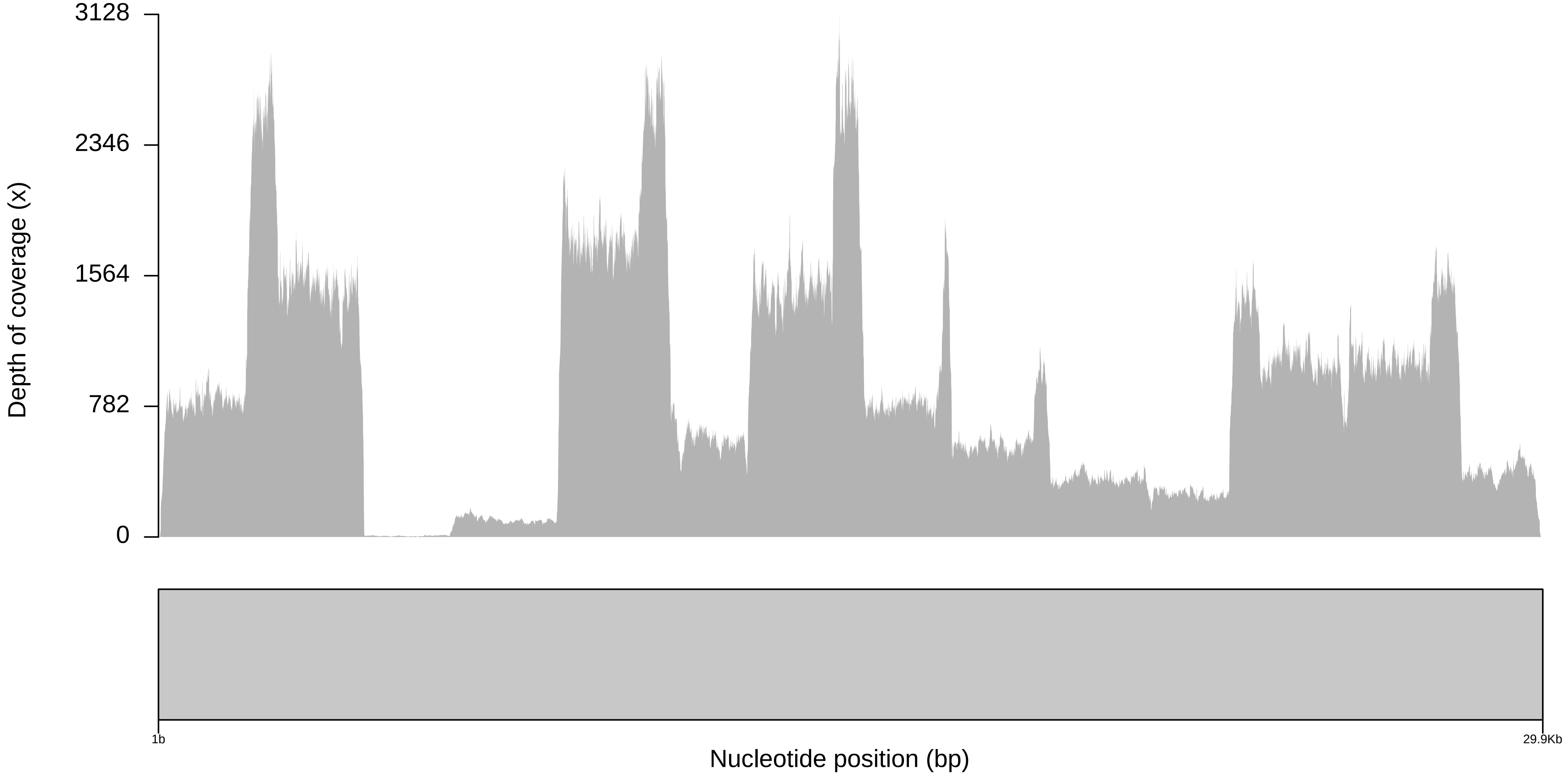

3128
2346
1564
782
0
Depth of coverage (x)
1b
29.9Kb
Nucleotide position (bp)

# 39465

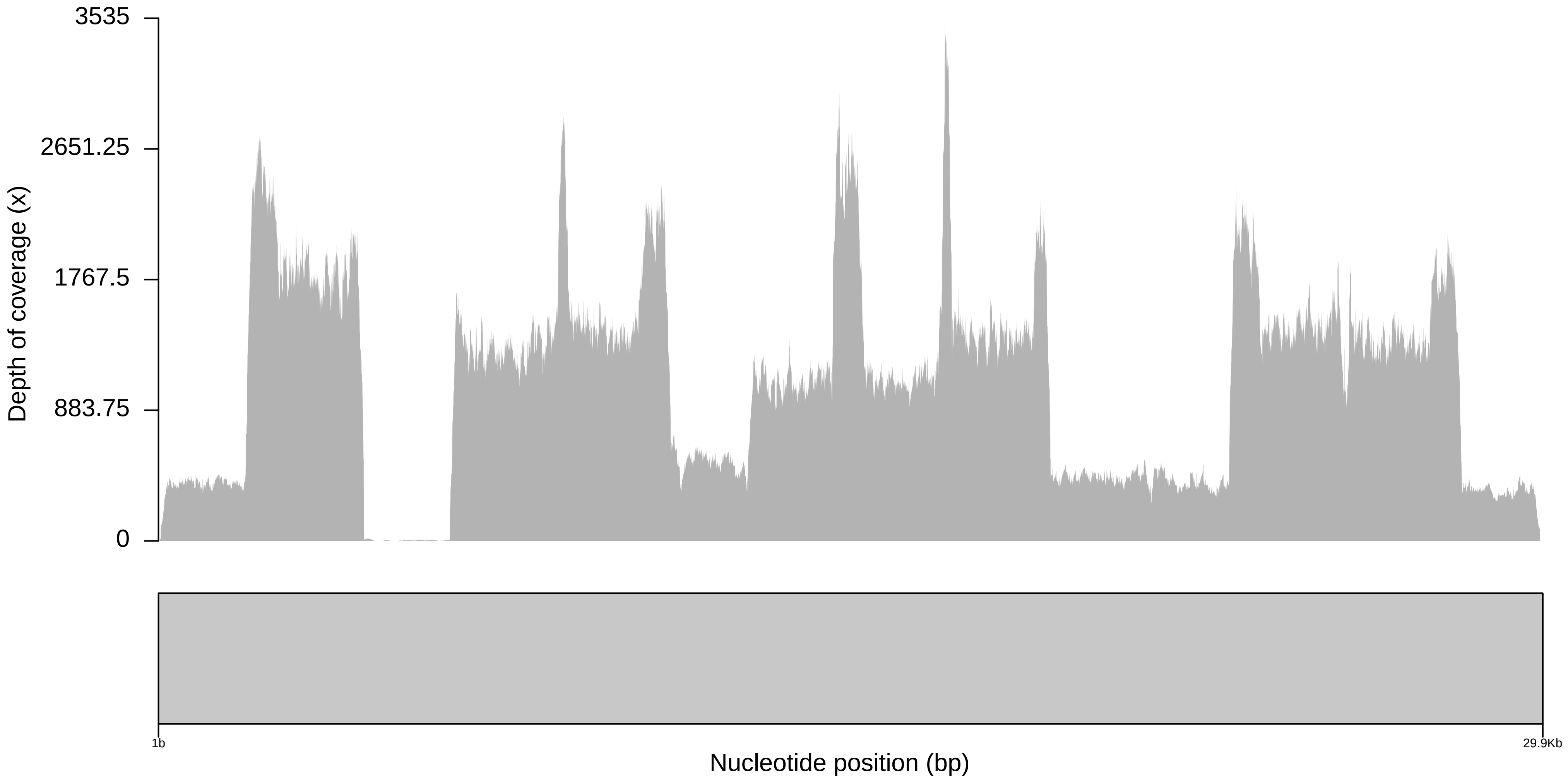

3535
2651.25
1767.5
883.75
0
Depth of coverage (x)
1b
29.9Kb
Nucleotide position (bp)

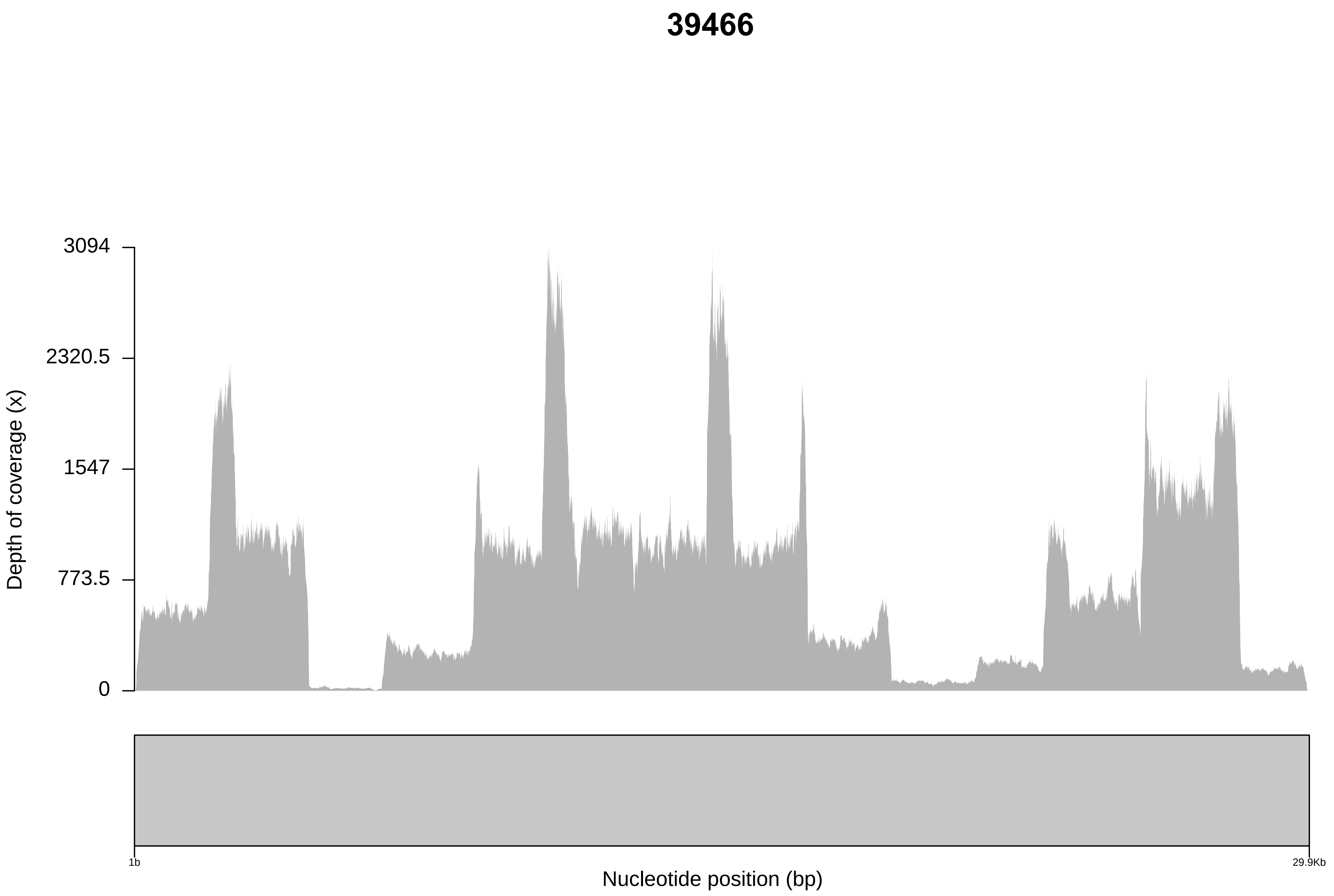

39466
3094
2320.5
1547
773.5
0
Depth of coverage (x)
1b
29.9Kb
Nucleotide position (bp)

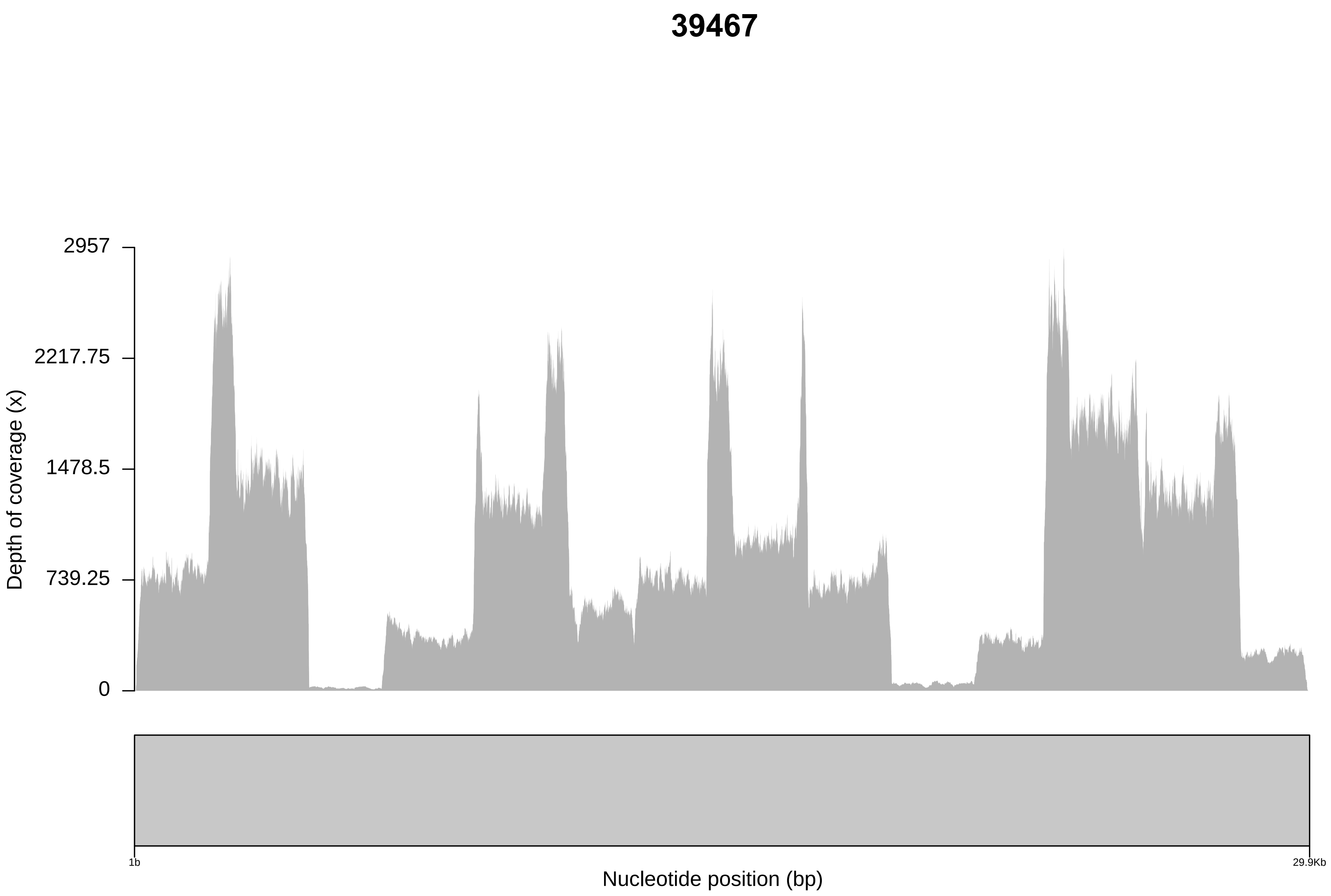

39467
2957
2217.75
1478.5
739.25
0
Depth of coverage (x)
1b
29.9Kb
Nucleotide position (bp)
